# Development and Implementation of a scalable and versatile test for COVID-19 diagnostics in rural communities

**DOI:** 10.1101/2021.03.01.21252679

**Authors:** A. Ceci, C. Muñoz-Ballester, A. Tegge, K.L. Brown, R.A. Umans, F.M. Michel, D. Patel, B. Tewari, J. Martin, O. Alcoreza, T. Maynard, D. Martinez-Martinez, P. Bordwine, N. Bissell, M. Friedlander, H. Sontheimer, C.V. Finkielstein

## Abstract

Rapid and widespread testing of severe acute respiratory coronavirus 2 (SARS-CoV-2) is essential for an effective public health response aimed at containing and mitigating the coronavirus disease 2019 (COVID-19) pandemic. Successful health policy implementation relies on early identification of infected individuals and extensive contact tracing.However, rural communities, where resources for testing are sparse or simply absent, face distinctive challenges to achieving this success. Accordingly, we report the development of an academic, public land grant University laboratory-based detection assay for the identification of SARS-CoV-2 in samples from various clinical specimens that can be readily deployed in areas where access to testing is limited. The test, which is a quantitative reverse transcription polymerase chain reaction (RT-qPCR)-based procedure, was validated on samples provided by the state laboratory and submitted for FDA Emergency Use Authorization. Our test exhibits comparable sensitivity and exceeds specificity and inclusivity values compared to other molecular assays. Additionally, this test can be re-configured to meet supply chain shortages, modified for scale up demands, and is amenable to several clinical specimens. Test development also involved 3D engineering critical supplies and formulating a stable collection media that allowed samples to be transported for hours over a dispersed rural region without the need for a cold-chain. These two elements that were critical when shortages impacted testing and when personnel needed to reach areas that were geographically isolated from the testing center. Overall, using a robust, easy-to-adapt methodology, we show that an academic laboratory can supplement COVID-19 testing needs and help local health departments assess and manage outbreaks. This additional testing capacity is particularly germane for smaller cities and rural regions that would otherwise be unable to meet the testing demand.

## INTRODUCTION

The coronavirus disease 2019 (COVID-19) outbreak caused by the severe acute respiratory syndrome coronavirus 2 (SARS-CoV-2) surprised health care systems and diagnostic facilities worldwide. First reported as pneumonia cases of unknown origin in the province of Wuhan, China early in December 2019^1^; SARS-CoV-2 infections rapidly moved to the status of a public health emergency of international concern (PHEIC) by the World Health Organization (WHO) Director-General in late January 2020. On March 11^th^, 2020, the WHO declared the novel coronavirus outbreak a global pandemic and urged countries to implement large scale testing as a strategy for controlling the spread of SARS-CoV-2^2^.

Three months after public disclosure of the first COVID-19 case, the international community faced a challenging public health scenario: *i*) widespread availability of testing was almost nonexistent, *ii*) confusion about what would suffice as a reliable read out for diagnostics, and *iii*) public health policies were inconsistent, rarely implemented, and enforced. This scenario was particularly challenging in the United States (U.S.), at least in part, due to a lack of coordination among federal and state agencies, inadequate acknowledgement of the crisis and few sufficiently meaningful steps taken to control it. These decisions resulted, as of February 11^th^, 2021 in 27+ million people being infected and 473,873 deaths in the U.S. alone^3^.

At the time of the latest U.S. decennial census, approximately 60 million people (roughly 19% of the U.S. population) live in rural communities, an area comprising more than 95% of the total U.S. land mass (United States Census Bureau). Communities in rural areas continue to face unique challenges that stem from social inequities, literacy levels below national standards^4^, and limited access to quality health care. Consequently, rural Americans are facing the COVID-19 pandemic from a precarious vantage point, which is aggravated by the lack of healthcare infrastructure and the presence of an adult population with disproportionate comorbidities and disabilities compared to their non-rural counterpart^5-7^. A case in point is southwest Virginia (SWVA), the state’s westernmost region including all of Virginia’s seven coal-producing counties. According to the latest U.S. census data, SWVA is poorer, older, less diverse, and has a slower population growth than most regions in the State of Virginia^8^ and yet, it was in this region where testing capabilities for COVID-19 were scarce or absent when COVID-19 was declared a national emergency.

As a response to the national demand for testing, the U.S. Centers for Disease Control and Prevention (CDC) developed a COVID-19 assay for analyzing patient specimens in laboratories, which was certified under the Clinical Laboratory Improvement Amendments (CLIA) and received Emergency Use Authorization (EUA) permission by the Food and Drug Administration (FDA). However, chemical shortages for reagents placed pressure on laboratories wanting to run COVID-19 tests. Testing laboratories ultimately faced the same roadblock in the reagent supply chain, which affected their diagnostic capabilities. These limitations also increased the wait time for test results, posing stressful uncertainty and constraints on patients and their families. Despite a fragmented national approach within the U.S., academic laboratories have become an integral element in tracking the spread of COVID-19 across local communities^9,10^.

With over 300 academic laboratories within the U.S. alone, many of these institutions possess not just the technical capacity but also the expertise required to expand COVID-19 testing. RT-qPCR assays are routinely performed in biomedical research programs and researchers were thus positioned to implement modified versions of these assays to detect SARS-CoV-2. Therefore, the scientific community could serve a specialized function and strengthen its connection to local communities and public health programs by generating its own COVID-19 pipeline operation. In this article, we report the development of an academic, SARS-CoV-2 RT-qPCR molecular diagnostic test that was validated against samples provided by the state laboratory and submitted for EUA approval (EUA# 200383). Our test has aided multiple local Health Districts of southwestern Virginia (SWVA) to track and manage the spread of COVID-19 in the general population and at potential outbreak sites across the region. Accordingly, our test allows for the *in vitro* detection of RNA from SARS-CoV-2 in respiratory (nasopharyngeal swabs, oropharyngeal swabs, anterior nasal swabs, mid-turbinate nasal swabs, bronchoalveolar lavage, nasopharyngeal wash/aspirates, and nasal aspirates) and saliva samples obtained from individuals suspected of having COVID-19. Particularly appealing aspects of this molecular test include its flexibility for implementation in various experimental settings, easy incorporation into high-throughput platforms to scale up capacity, and the versatility to maneuver around supply chain deficiencies. As of February 2021, 78,000+ individuals from seven health districts in SWVA, including more than 650 businesses, nursing homes, physician and dentistry offices, construction sites and all public schools in the New River Valley (NRV) have been tested with this newly developed assay where specimens were processed and reported within 24 hours of sample arrival.

## RESULTS

In response to the COVID-19 pandemic, there arose, and continues to be, an urgent need for obtaining timely, accurate test results from large numbers of people throughout the Commonwealth of Virginia. This need is twofold: *i*) to be able to test and provide appropriate medical care and quarantine recommendations for patients who are symptomatic, including those who may currently be hospitalized or may soon need to be hospitalized, and *ii*) more wide-scale surveillance testing to identify asymptomatic and pre-symptomatic individuals to facilitate public health mediated contact tracing and associated mitigation strategies. As testing availability was initially unable to meet demands for quick turnaround times and throughput ability were limited, there was considerable need for enhanced testing availability throughout the Commonwealth of Virginia and the country. As a result, our academic institution developed a test and a comprehensive program, in partnership with the Virginia Department of Health and Virginia Department of General Services, to support testing of personnel, students, and the community-at-large in SWVA.

### Test principle and optimization

Our assay is a real-time reverse transcription polymerase chain reaction (RT-qPCR)-based test designed to amplify three distinct SARS-CoV-2 genes, *i.e*., *N, E*, and *S*, in the context of the expression of a housekeeping control gene named *RPP30*. A flow chart of the general procedure is presented in Fig. 1.A. Briefly, clinical specimens (*e.g*., mid-turbinate) were collected using a commercial sterile synthetic rayon/Dacron®/polyester/nylon tipped applicator and transported in a specially formulated transport media. As swabs became a limiting factor during the pandemic, in-house manufacturing of 3D-printed swabs became a stopgap alternative to replace traditional flocked swabs (Fig. 1.B-C). Accordingly, nasopharyngeal swabs were fabricated by inverse stereolithography following a design process developed by University of South Florida Health (patent is pending) with materials commonly used for clinical printing that were FDA-approved for biocompatibility ^11^. This design, already evaluated in a clinical trial ^11^, was duplicated in our laboratory as described in the Materials and Methods section. The mechanical properties of the two printed swabs, an adult and pediatric version, exhibit comparable tensile and torsional strength within the variances observed (Fig. 1.D). These results were further compared to the testing values of commercially available swabs previously reported by Formlabs (Copan and Puritan, Swab Verification Summary Report #SR-0010 rev#00). In general, the performance, toughness, and torsional testing results of our 3D-printed swabs fell within those approved to be the standard of care [Copan (highest) and Purtan (lowest)].

**Figure 1.**
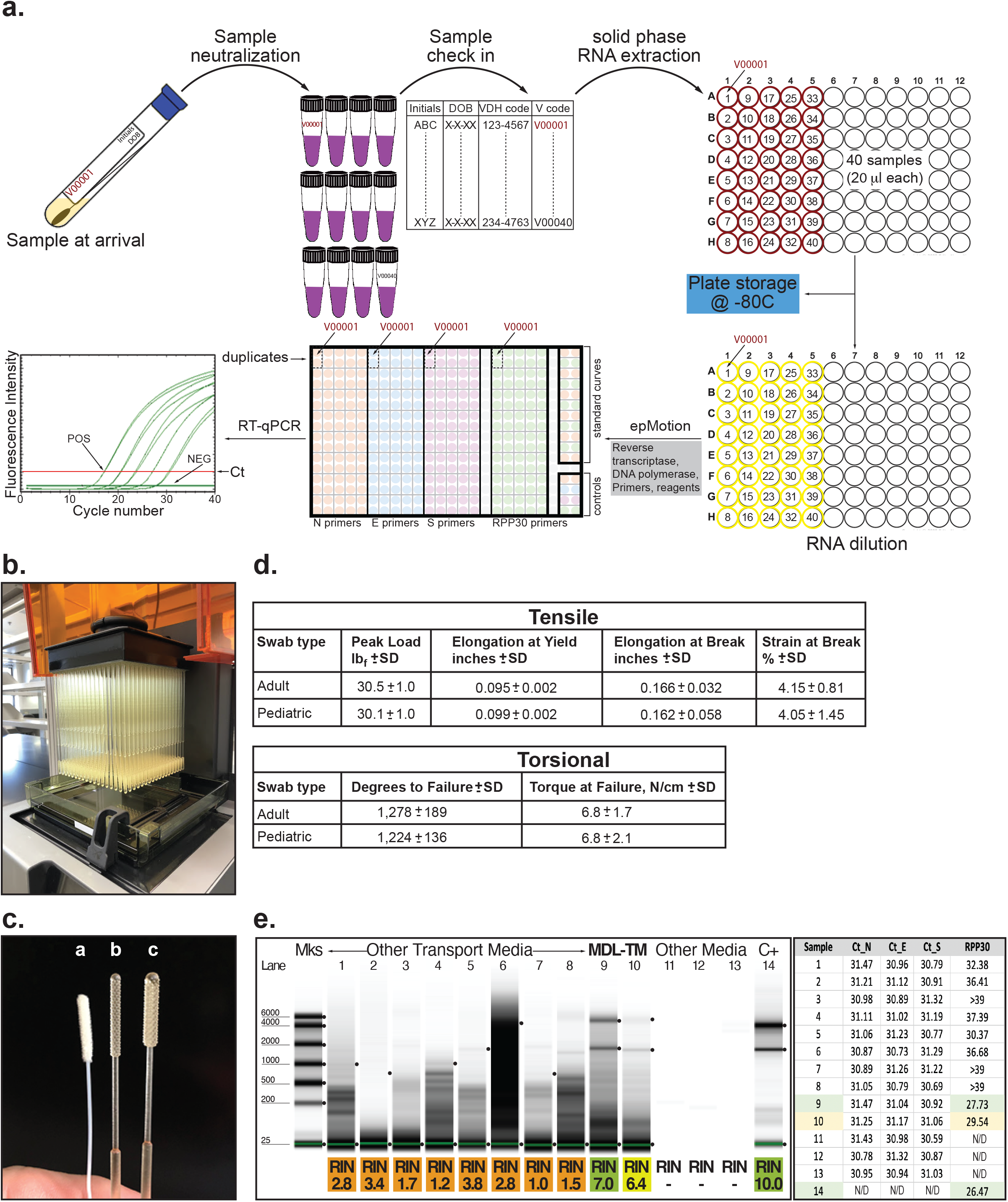
**a**. SARS-CoV-2 testing flow chart. Briefly, tubes containing MDL-TM and swab were checked in the system, a matrix created to follow the sample throughout the whole process, and RNA was extracted as described in the Materials and Methods section. Plates containing RNA from all 40 samples were either stored at −80°C until needed or, an aliquot of each sample was diluted and subjected to RT-qPCR amplification. Each sample was tested in duplicate for 4 genes (*N, E*, and *S* from SARS-CoV-2 and *RPP30*) in a 384-well format plate. In addition, each plate included positive and negative controls and standard curves for the *N* and *RPP30* genes as specified in the Materials and Methods section. **b**. Completed print of 324 adult swabs using Formlabs Form2 with Surgical Guide resin. **c**. PurFlock (Puritan Medical Products, LLC) test swab (a) compared to 3D-printed pediatric (b) and adult (c) swabs. **d**. Summary of average tensile (*top*) and torsional (*bottom*) testing results of 3D-printed adult and pediatric swabs (n=10 samples of each type), respectively. **e**. *Left panel*, RNA integrity analysis (RIN) of clinical samples collected using various transport media (lanes 1 to 8) or the transport media developed by our laboratory (lanes 9-10, MDL-TM). Duplicate RNA samples were purified the day of delivery or ten days after collection, respectively (lanes 9 and 10). C+: total RNA sample purified from human cells. *Right panel*, a suspension of negative nasopharyngeal swabs collected in various formulations of transport media were spiked with inactive SARS-CoV-2 virus (∼300 copies, ATCC) (samples 1 to 14). Samples were extracted and RNA amplified as described in Materials and Methods. Matching samples (9 and 10) were processed immediately or maintained at 4°C for 10 days before being analyzed. Sample 14 was total RNA purified from cultured mammalian cells. N/A = not amplification observed. *N*/*S*/*E* gene cutoff: 37.079, 95%CI[35.75, 36.67]; *RPP30* gene cutoff: 38.119, 95%CI[36.02, 36.46].

Nasopharyngeal swabs were placed in a specific formulated transport media (MDL-TM) that *i*) inactivates infectious agents, *ii*) stabilizes RNA molecules, and *iii*) does not require uninterrupted cold-chain logistics. We initially demonstrated the efficacy of the MDL transport media in stabilizing nucleic acids by performing RNA integrity analyses (Fig. 1.E). Accordingly, inactive SARS-CoV-2 virus was spiked into various formulations of TM consisting of a suspension of a negative nasopharyngeal (NP) swab. Viral particles were released from the swab into the MDL media using mechanical stirring. An aliquot of the eluate was then subjected to a TRIzol/EtOH-based nucleic acid extraction protocol in a 96-well column format system following various washing/elution procedures as described in the Materials and Methods section. The integrity of purified RNA was evaluated by microcapillary electrophoretic RNA separation and assigned an RNA integrity number (RIN) ranging from 1 (worst) to 10 (best) (Fig. 1.B, left panel). The RIN results validated our transport media as an effective diluent as it was the one formulation that best preserved the quality of the RNA even 10 days after collection (Fig. 1.E, *left panel*). Next, we evaluated whether components of the TM might inhibit the amplification reaction. Consequently, total RNA was purified from each transport media sample and used for reverse transcription and amplification in a formulation that contains RNase inhibitor and additives to reduce formation of the primer’s secondary structure. Primers for the amplification of SARS-CoV-2 (*N, E*, and *S*) and human housekeeping (*RPP30*) genes were included in four independent reaction mixtures. Under these conditions, maximum sensitivity and reliability was ensured as false positives resulting from primer dimers were less likely to occur. Reactions were, then, assembled in a 384-well plate format and amplified as described in the Materials and Methods section. Results show that the composition of the MDL-TM did not inhibit gene amplification and preserves the quality of the sample, seen by the comparable Ct values obtained for the viral genes, even 10 days after collection (Fig. 1.E, right panel).

Clinical specimens were expected to exhibit fluorescence amplification curves when using the *Hs_RPP30* primers with Ct values that cross the threshold line below a cutoff determined by a standard curve, thus, confirming the presence of human material *via* the *RPP30* gene. Samples from presumptive positive COVID-19 patients were expected to amplify at least two out of the three genes from SARS-CoV-2, (*i.e*., *N, E*, and *S*), a criterion that allows for higher specificity and reduces the likelihood of false negative results (see Supplementary Table II for complete criteria). Standard curves for *RPP30* were included in each run and for each plate. Failure to detect *RPP30* in any clinical specimen may indicate: *i*) improper extraction of RNA from the clinical sample, *ii*) absence of sufficient human cellular material due to poor collection or loss of specimen integrity, *iii*) improper set-up and execution, or *iv*) equipment malfunction. Interpretation of results when a negative *RPP30* was reported was as follows: *i*) if at least two of the viral genes (*N, E, S*) were positive even in the absence of a positive *RPP30*, the result was considered valid. A negative *RPP30* signal did not preclude the detection of viral RNA in a clinical specimen, and *ii*) if viral genes <u>and</u> *RPP30* were negative for the specimen, the result was considered invalid. In this last scenario, if a residual clinical sample was available, it was recommended to repeat the extraction procedure as well as the test. However, if all markers remained negative after a re-test, the result was reported as “invalid” and a new specimen needed to be collected (Supplementary Table II).

The analytical sensitivity of the assay was determined by limiting dilution studies (limit of detection, LOD). Accordingly, assays were designed for the detection of SARS-CoV-2 using stocks of inactive virus (ATCC) spiked into a diluent consisting of a suspension of a previously characterized negative nasopharyngeal swab maintained in viral transport media. Ten-fold serial dilutions of characterized stocks of viral SARS-CoV-2 were tested in triplicate with each primer set. Of note, the number of copies of SARS-CoV-2 virus was batch-specific and was provided by ATCC on its Certificate of Analysis. Samples were manually extracted and RT-qPCR assays performed following the procedure described in the Materials and Methods section.

To determine the preliminary LOD, we used a batch of virus with a copy number of 1.2 × 10^3 copies/µl as determined by digital PCR (ddPCR). Per FDA guidelines, the number of viral copies spiked in the original sample for LOD determination varied from 10^4^ to 10^−5^. For each dilution, and after sample processing and amplification, the number of viral copies detected using each set of primers in the assay were reported per reaction volume (Table I). Validation of the LOD was carried out by testing the system’s sensitivity for detection in 20 samples, each spiked with a concentration of inactive virus such that the final number of copies in the reaction was 300, 20, or 1.3 (*i.e*., thirty- and two-times above and seven-times below the LOD determined in Table I, respectively, Supplementary Tables III, IV, V). As a result, the LOD (10 copies per reaction) was determined as the lowest concentration where ≧ 95% of the replicates were positive.

**Table I.** Estimation of the Limit of Detection (LOD). Ct values were determined using the CFX Maestro 2.0 software, (Bio-Rad) using the default single threshold, baseline subtracted curve fitting model. *Indicates that the raw data curve did not reach a plateau. Cut-off value for *N* gene: 37.29, 95% CI: [36.91, 37.67] as determined using the 2019-nCoV_*N*_Positive control (IDT) as the template; N/A = not amplification observed.

| **N primer** |  |  |  |  |  |
| --- | --- | --- | --- | --- | --- |
| **Viral copies/10µl reaction** | **Ct Replicate 1** | **Ct Replicate 2** | **Ct Replicate 3** | **Mean CT** | **% Positive** |
| 10,000 | 23.7 | 23.49 | 23.72 | 23.64 | 100 |
| 1,000 | 27.15 | 27.05 | 27.05 | 27.08 | 100 |
| 100 | 30.7 | 30.4 | 30.6 | 30.6 | 100 |
| 10 | 34.34 | 34.75 | 33.95 | 34.35 | 100 |
| 1 | 39.32* | 37.21* | 37.27* | 37.93 | 0 |
| 0.1 | N/A | N/A | N/A | N/A | 0 |
| 0.01 | N/A | N/A | N/A | N/A | 0 |
| 0.001 | N/A | N/A | N/A | N/A | 0 |
| 0.0001 | N/A | N/A | N/A | N/A | 0 |
| 0.00001 | N/A | N/A | N/A | N/A | 0 |
| **E primer** |  |  |  |  |  |
| **Viral copies/10µl reaction** | **Ct Replicate 1** | **Ct Replicate 2** | **Ct Replicate 3** | **Mean CT** | **% Positive** |
| 10,000 | 23.22 | 23.25 | 23.33 | 23.27 | 100 |
| 1,000 | 26.93 | 26.73 | 26.85 | 26.84 | 100 |
| 100 | 30.4 | 30.45 | 30.49 | 30.45 | 100 |
| 10 | 34.42 | 33.72 | 33.98 | 34.04 | 100 |
| 1 | 36.29* | 37.17* | 36.24* | 36.56 | 66.66 |
| 0.1 | N/A | N/A | N/A | N/A | 0 |
| 0.01 | N/A | N/A | N/A | N/A | 0 |
| 0.001 | N/A | N/A | N/A | N/A | 0 |
| 0.0001 | N/A | N/A | N/A | N/A | 0 |
| 0.00001 | N/A | N/A | N/A | N/A | 0 |
| **S primer** |  |  |  |  |  |
| **Viral copies/10µl reaction** | **Ct Replicate 1** | **Ct Replicate 2** | **Ct Replicate 3** | **Mean CT** | **% Positive** |
| 10,000 | 23.1 | 23.12 | 23.26 | 23.16 | 100 |
| 1,000 | 26.59 | 26.54 | 26.64 | 26.59 | 100 |
| 100 | 30.01 | 30.14 | 29.84 | 30.0 | 100 |
| 10 | 33.66 | 33.72 | 33.07 | 33.48 | 100 |
| 1 | N/A | 35.4* | N/A | 35.4 | 33.33 |
| 0.1 | N/A | N/A | N/A | N/A | 0 |
| 0.01 | N/A | N/A | N/A | N/A | 0 |
| 0.001 | N/A | N/A | N/A | N/A | 0 |
| 0.0001 | N/A | N/A | N/A | N/A | 0 |
| 0.00001 | N/A | N/A | N/A | N/A | 0 |

### Inclusivity analysis (Analytical Sensitivity)

*In silico* analysis using published sequences was performed to determine the predicted inclusivity of our SARS-CoV-2 assay. 32,911 SARS-CoV-2 sequences were downloaded from NCBI (Supplementary Table VI). The exclusion criteria removed 536 sequences from further analysis as they were incomplete (535 sequences) or of poor quality (1 sequence). Of the remaining 32,375 sequences, 27 sequences contained mismatches near the 3’ end of the primer binding site that might affect amplification by one of the primers and 60 sequences contained multiple mismatches over the length of a primer binding region that might affect amplification. None of the 32,375 SARS-CoV-2 sequences obtained from NBCI contained mismatches that could affect amplification in more than one primer set. Therefore, our SARS-CoV-2 assay was predicted to have 100% inclusivity by *in silico* analysis of all SARS-CoV-2 sequences published in NCBI as of January 13, 2021.

240,791 SARS-CoV-2 sequences were downloaded from GISAID (gisaid.org) (Supplementary Table VII). The exclusion criteria removed 11,453 sequences from further analysis as they were of poor quality. All sequences completely covered the six primer binding regions. Of the remaining 229,338 sequences, 919 sequences contained mismatches near the 3’ end of the primer binding site that might affect amplification by one of the primers and 378 sequences contained multiple mismatches over the length of a primer binding region that might affect amplification. Two of the 229,338 SARS-CoV-2 sequences contained mismatches that could affect amplification by two different primer sets. Thus, our SARS-CoV-2 assay was predicted to have 99.9991% inclusivity by *in silico* analysis of all SARS-CoV-2 sequences published in GISAID as of January 7, 2021.

### Cross-reactivity analysis (Analytical Specificity)

To validate our RT-qPCR assay for specificity towards SARS-CoV-2 *versus* other common respiratory flora and viral pathogens, we performed *in silico* cross-reactivity analyses. Per FDA guidelines, *in silico* cross-reactivity is defined when greater than 80% homology between one of the primers/probes and any sequence is present in the targeted microorganism. Respiratory pathogens were separated into two main categories including those that belong to the same family as SARS-CoV-2 (*e.g*., human coronavirus 229E, OC43, HKU1, NL63 and SARS-CoV-1 and MERS-coronavirus) and others pathogens considered high priority and likely circulating in our area (*e.g*., influenza A and B, rhinoviruses, *Streptococcus pneumoniae*, and *Pseudomonas aeruginosa*). The criteria for potentially affecting specificity of our assay included having greater than 80% homology to at least two of the three primer sets.

BLAST analysis queries of our SARS-CoV-2 qRT-PCR assay primers were performed against the public domain nucleotide sequences listed in Table II. Among all *in silico* tested organisms, the *N* and *E* primers showed >80% homology for the bat SARS-like coronavirus genome (Supplementary Table VIII). Our findings also indicated that *N*, but not *E* or *S*, forward and reverse primers could amplify bat coronavirus CoVZXC21 and CoVZC45 as these sequences perfectly match our N primers, thus may generate a positive result for the N gene in our test. However, the S primers showed no significant homology with any non-target sequence, including bat coronaviruses. For the E primer sets, there is predicted to be no significant amplification for SARS-CoV-1 viruses as only one of the E primers had homology to these viruses. Lastly, sequences of the *N, E, S* primers did not show significant homology with other pathogens that most commonly cause respiratory infections (Table II).

**Table II.** *In silico* analysis of primers used for amplification compared to common respiratory flora and other respiratory pathogens. Per FDA definition, cross-reactivity occurs when homology is greater than 80% between the primer and the template sequence in the targeted microorganism. Sequences were retrieved from the National Center for Biotechnology Information, https://www.ncbi.nlm.nih.gov/taxonomy/

|  | Taxonomy ID | Number of sequences with % identity 80-100, E value ≤10, Query coverage 80-100 |  |  |  |  |  |
| --- | --- | --- | --- | --- | --- | --- | --- |
|  |  | N F | N R | E F | E R | S F | S R |
| Human coronavirus 229E | 11137 | 0 | 0 | 0 | 0 | 0 | 0 |
| Human coronavirus OC43 | 31631 | 0 | 0 | 0 | 0 | 0 | 0 |
| Human coronavirus HKU1 | 290028 | 0 | 0 | 0 | 0 | 0 | 0 |
| Human coronavirus NL63 | 277944 | 0 | 0 | 0 | 0 | 0 | 0 |
| HCoV-SARS | 694009 | 3 | 3 | 21 | 3 | 0 | 0 |
| MERS-CoV | 1335626 | 0 | 0 | 0 | 0 | 0 | 0 |
| Human adenovirus | 1907210 | 0 | 0 | 0 | 0 | 0 | 0 |
| Human metapneumovirus | 162145 | 0 | 0 | 0 | 0 | 0 | 0 |
| parainfluenza virus 1 | 12730 | 0 | 0 | 0 | 0 | 0 | 0 |
| Human rubalavirus 2 (Parainfluenza virus 2) | 1979160 | 0 | 0 | 0 | 0 | 0 | 0 |
| parainfluenza virus 3 | 11216 | 0 | 0 | 0 | 0 | 0 | 0 |
| Human rubalavirus 4 (Parainfluenza virus 4) | 1979161 | 0 | 0 | 0 | 0 | 0 | 0 |
| influenza A | 2072034 | 0 | 0 | 0 | 0 | 0 | 0 |
| influenza B | 2072149 | 0 | 0 | 0 | 0 | 0 | 0 |
| enterovirus (e.g. EV68) | 42789 | 0 | 0 | 0 | 0 | 0 | 0 |
| respiratory syncytial virus | 12814 | 0 | 0 | 0 | 0 | 0 | 0 |
| rhinovirus (Human rhinovirus A1) | 573824 | 0 | 0 | 0 | 0 | 0 | 0 |
| *Chlamydia pneumoniae* TW-183 | 182082 | 0 | 0 | 0 | 0 | 0 | 0 |
| *Haemophilus influenzae* | 727 | 0 | 0 | 0 | 0 | 0 | 0 |
| *Legionella pneumophila* | 446 | 0 | 0 | 0 | 0 | 0 | 0 |
| *Mycobacterium tuberculosis* | 1773 | 0 | 0 | 0 | 0 | 0 | 0 |
| *Streptococcus pneumoniae* | 1313 | 0 | 0 | 0 | 0 | 0 | 0 |
| *Streptococcus pyogenes* | 1314 | 0 | 0 | 0 | 0 | 0 | 0 |
| *Bordetella pertussis* | 520 | 0 | 0 | 0 | 0 | 0 | 0 |
| *Mycoplasma pneumoniae* | 2104 | 0 | 0 | 0 | 0 | 0 | 0 |
| *Pneumocystis jirovecii* (PJP) | 42068 | 0 | 0 | 0 | 0 | 0 | 0 |
| *Candida albicans* | 5476 | 0 | 0 | 0 | 0 | 0 | 0 |
| *Pseudomonas aeruginosa* | 287 | 0 | 0 | 0 | 0 | 0 | 0 |
| *Staphylococcus epidermis* | 1282 | 0 | 0 | 0 | 0 | 0 | 0 |
| *Staphylococcus salivarius* | 1304 | 0 | 0 | 0 | 0 | 0 | 0 |

### Test validation and clinical evaluation

Initial experiments were devoted to validating our test using 66 contrived specimen samples (33 positive and 33 negative) in a pilot study. Samples were contrived by spiking a known concentration of inactive SARS-CoV-2 virus, or a genetically related virus with similar tropism towards the respiratory and gastrointestinal track as SARS-CoV-2 but that was undetectable by our primers (Bovine coronavirus, BCoVs) as the negative control. In either case, samples were loaded with viral particles (analyte) equal to two-times the LOD as established in Table I. Different amounts of analyte (samples 1-27 and 28-33) was added into matrices previously determined to be negative by the *Power* SYBR™ Green RNA-to-CT™ *1-Step* Kit. Results from positive samples are summarized in Supplementary Table IX. A single plate-specific cutoff value of 36.25, 95%CI [35.60, 36.90] was used for all viral primer sets. Our analyses indicated that the Ct value corresponding to the inferior point of the 95% interval of confidence for the LOD for the *E* and *S* genes were smaller (*i.e*., larger number of viral copies) than that observed for the *N* gene. To be conservative, a threshold approach was utilized and all sample Ct values for the *E, N*, and *S* genes were compared to the Ct value of the inferior point of the 95% interval of confidence calculated for gene *N*.

Next, to establish the clinical assay performance, 81 participant samples, each consisting of a NP swab, were collected in MDL-TM and tested for the presence of SARS-CoV-2 using our RT-qPCR test. Comparable aliquots of all clinical specimens were submitted to the Virginia Division of Consolidated Laboratory Services (DCLS) for validation. The comparator assay performed by DCLS followed the CDC EUA IFU (<u>CDC DOC 006-0099 rev. 03)</u> “CDC 2019-Novel Coronavirus (2019-nCoV) Real-Time RT-PCR Diagnostic Panel” and used probes and a primer kit previously approved “IDT-2019-nCoV CDC EUA Kit (primers) cat # 10006606.” Results from our test were reported in accordance with the criteria summarized in Supplementary Table II in which amplification of, at least two viral genes (*N, E*, or *S*) below the lowest 95% IC value threshold were required to report a positive result. In agreement with CDC guidelines, reports from DCLS were based on the detection of only one viral gene, *N*, with two different primer sets named N1 and N2. Ct values for all genes and corresponding cut-offs for each clinical sample are summarized in Supplementary Table X. Results show 100% agreement among samples compared from both laboratories with 45 positive and 33 negative NP swab specimens evaluated in both locations (Positive Percent Agreement: 100%, 95% CI [92.1, 100]; Negative Percent Agreement: 100%, 95% CI [90.3-100]). Overall, our results show the robustness, specificity, and sensitivity of our test for timely identification of the SARS-CoV-2 virus in clinical specimens.

### Increasing testing capacity by sample pooling

As infection rates in the population and shortages in the supply chain increased, it was necessary to reallocate resources towards those individuals in need of immediate testing (*e.g*., health professionals, patients, individuals in nursing homes) versus others for which screening would be advantageous (*e.g*., family groups, athletic teams, group workers, students). Therefore, it was determined that a pooling of samples strategy would be useful, for example, in areas with low prevalence of SARS-CoV-2 as was the case for SWVA at specific windows of time during the pandemic.

Initial experiments were devoted to determining the sensitivity of the pooling method and in defining the optimal number of samples per pool. Accordingly, we used our protocol to pool test archived individual samples with n-sample pools each consisting of 1 positive sample and n-1 negative samples. In all cases, samples were from NP swabs taken from patients and tested in accordance with the protocol described in the Materials and Methods section. A sample’s status (*i.e*., positive or negative) was determined based on cut-off values and the 95% CI for the *N* and *RPP30* genes as explained earlier. Once tested, samples were separated into two groups: positives and negatives. To generate the negative pools for testing, 4 samples from the negative group were used in each of the pooled samples. No negative sample was used in more than one negative pooled sample. Some, but not all, of the negative samples had enough volume to be included in the positive pool. To generate the positive pools for testing, 3 negative samples were used in combination with one positive sample. All positive pooled samples contained a different set of negative samples; no two positive pooled samples contained the same three negative samples.

First, we validated pooling studies using positive samples with at least 25% of them being within 2 – 3 Cts of the cut-off, and no more than within 2 −4 Cts overall. Validation was performed in pools of 2, 3, and 4 samples (Supplementary Table XI, XII, and XIII). For pools containing n=2 samples [one positive and one negative (P1+N1) or two negative samples (N1+N2)], we analyzed 20 negative and positive pools containing 2 samples in each pool. Supplementary Table XI summarizes the results of the Cts for the *N, E, S*, and *Hs_RPP30* genes and cut-offs for *N* and *Hs_RPP30* for each of the two samples in the pool (columns F-L and P-V, Pool_raw_data tab) and for the pool itself (columns Z-AF, Pool_raw_data tab). Note that the assay cut-off for each sample varied since thresholds were determined on a plate-by-plate basis. When Ct values for each gene of the positive sample were compared with the corresponding n=2 pool, we found that Ct differences were less than two in 22 samples analyzed (Supplementary Table XI, Positives_tab). Regression analysis of individual *versus* pooled Cts showed an R^2^ of 0.94, 0.92, and 0.91 for the *N, E, S*, genes, respectively [LOD individual N: 38.54, 95%CI (38.12, 38.97); LOD pool N: 37.29, 95% CI (36.91, 37.67)] (Supplementary Table XI, Regression_tab). These results indicated that, following this pooling procedure, samples with Cts for the N gene between 38.12 and 36.91 would have been detected using individual testing but not when pooling.

We next evaluated the result of 20 negative and 22 positive pooled samples containing 3 individual specimens (P2+N3+N4 and N3+N4+N5, Supplementary Table XII). As was the case when pooling 2 samples, deviation of Ct values was within 2 units when comparing individual versus 3 pooled samples (pool_raw_data tab and positives tab). Regression analysis for individual *versus* pooled Cts showed that the R^2^ values for the *N, E*, and *S* genes were 0.96, 0.95, and 0.96, respectively [LOD individual N: 36.17, 95%CI (35.79, 36.56); LOD pool N: 37.29, 95% CI (36.91, 37.67)] (Supplementary Table XII, Regression_tab). Thus, samples with Cts between 35.79 and 36.91 would have been detected when testing individual samples but would have been missed when pooled. In a set of 20 negative (N6+N7+N8+N9) and 21 positive pooled samples of n=4 (P3+N6+N7+N8), 4 (19%) patient samples were within 4 Ct values of the threshold. We used the inferior limit from the individual test on the regression analysis tab of Supplementary Table XIII. With that value, we identified how many samples had *N*/*E*/*S* within a 4Ct range. Three samples had all 3 genes within the 4Ct value and 1 sample had 2 genes within the 4Ct value. Therefore, 4 samples were in the “weak” category and were likely missed when pooling. Based on the validation test (20 negative and 21 positive pooled samples), we observed 100% Positive Percent Agreement (PPA) and 100% Negative Percent Agreement (NPA). Per FDA recommendation, results were reported in a two-by-two table (Supplementary Table XIV.A). There was no difference in processing for pooled and single samples.

Finally, to assess the impact of pooling on individually tested samples evaluated in our laboratory, the FDA recommends comparing pooled Cts versus individual Ct values for the corresponding samples using Passing-Bablok regression analysis. The estimates for the slope and 95% confidence intervals of the slopes for the *N, E*, and *S* genes are summarized in Supplementary Table XIV.B. These results suggest that the relationship between pooled and individual samples are similar,as indicated by all confidence intervals, including a slope of 1.

We then evaluated the sensitivity of pooled testing. For this, we considered the most recent 100 positive samples as determined through individual testing. We evaluated their observed Ct values compared to theoretical and estimated Ct shifts for n=2, n=3, and n=4 (Supplementary Table XV). Using the theoretical Ct shift as a result of pooling 2 samples (log2(2)=1Ct shift), we observed a sensitivity of 82%. In addition, we estimated the pooled Ct threshold for each individual assay using the estimated observed shift in Ct value as a result of pooling (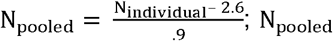 is the estimated Ct threshold for an individual plate and N_individual_ is the observed Ct threshold for the respective plate). From this analysis, we observed a sensitivity of 100%. Using the theoretical Ct shift as a result of pooling 3 samples (log2(3)=1.58Ct shift), we observed a sensitivity of 76%. In addition, we estimated the pooled Ct threshold for each individual assay using the estimated observed shift in Ct value as a result of pooling (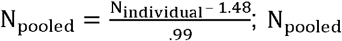 is the estimated Ct threshold for an individual plate and N_individual_ is the observed Ct threshold for the respective plate). From this analysis, we observed a sensitivity of 82%. Using the theoretical Ct shift as a result of pooling 4 samples (log2(4)=2Ct shift), we observed a sensitivity of 71%. In addition, we estimated the pooled Ct threshold for each individual assay using the estimated observed shift in Ct value as a result of pooling (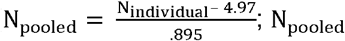 is the estimated Ct threshold for an individual plate and N_individual_ is the observed Ct threshold for the respective plate). From this analysis, we observed a sensitivity of 82%.

### Pooling as screening strategy in rural areas

Next, we implemented pooled testing as strategy for screening individuals suspected of COVID-19 within a region of SWVA. To determine whether pooling versus individual testing was a better choice for COVID-19 screening, we needed to evaluate the prevalence of positive cases within a certain time frame in the area. As a proof-of-concept, we chose to monitor the positivity rate among 3,807 individuals tested between July 10^th^ to July 30^th^, 2020 as follows: *i*) July 10^th^ to 16^th^, 1,288 individuals, *ii*) July 17^th^ to 23^rd^, 1,136 individuals, and *iii*) July 24^th^ to 30^th^, 1,383 individuals living in the New River Valley (Table III). The highest positivity rate within that time-frame was 4.4%; thus, using this positivity rate, pooling (n=4) would reduce the number of expected tests by 59% compared to individual sample testing. This was calculated using the Shiny application for pooled testing as described by Abdalhamid *et al*. [^12^, https://www.chrisbilder.com/shiny]. Here, we used estimates of 95% sensitivity and 100% specificity for our calculations. These estimates also showed that when the positivity rate in the sample reaches above 25%, individual sample testing would be a more efficient approach than pooling. If the positivity rate exceeds 25%, then we will return to testing samples individually. As positivity rate is an important variable for consideration when deciding to use pooled versus individual sample testing, it became important to monitor its value and the number of weak positives in the region by individual sample testing using a 7-day moving average (*i.e*., the average positive testing rate of the prior 7 days). Comparison among weekly positivity rates and the 25% positivity threshold were used to adjust our testing strategy.

**Table III.** Weekly positivity rate among NRVHD samples as monitored for pool testing. NEG: Negative, POS: Positive, Inc: Inconclusive, Inv: Invalid.

| Dates | # POS | % POS | # NEG | % NEG | # Inc/Inv | % Inc/Inv | Total |
| --- | --- | --- | --- | --- | --- | --- | --- |
| July 10<sup>th</sup> to 16<sup>th</sup> | 48 | 3.7% | 1221 | 94.8% | 19 | 1.5% | 1,288 |
| July 17<sup>th</sup> to 23<sup>rd</sup> | 50 | 4.4% | 1074 | 94.5% | 12 | 1.1% | 1,136 |
| July 24<sup>th</sup> to 30<sup>th</sup> | 58 | 4.2% | 1286 | 93.0% | 39 | 2.8% | 1,383 |

Samples from participants located in two health districts in SWVA were used for initial pooled testing - the New River Valley and the Mt. Rogers Health Districts (NRVHD and MRHD, respectively) (Fig. 2.A). These districts are considered “at-risk” regions as defined by the Appalachian Regional Commission based on economic indicators such as unemployment rate, income per capita, and poverty rate. People in rural areas face multiple challenges associated with coverage and access to health care as a result of unemployment, low annual income, limited number of providers and hospitals in the region, as well as long travel distances to access care ^4^. As a result, individuals in rural areas are less likely to have private health coverage than those in urbanized districts of the same state (Supplementary Table XVI). In fact, enrollment in Medicaid and Medicare are higher in the NRVHD (9.08% and 12.20%, respectively) and the MRHD (15.60% and 18%, respectively) than in urban health districts in Virginia [*e.g*., Fairfax Health District (FHD) (6.03% and 8.36%, respectively), Supplementary Table XVI]

**Figure 2.**
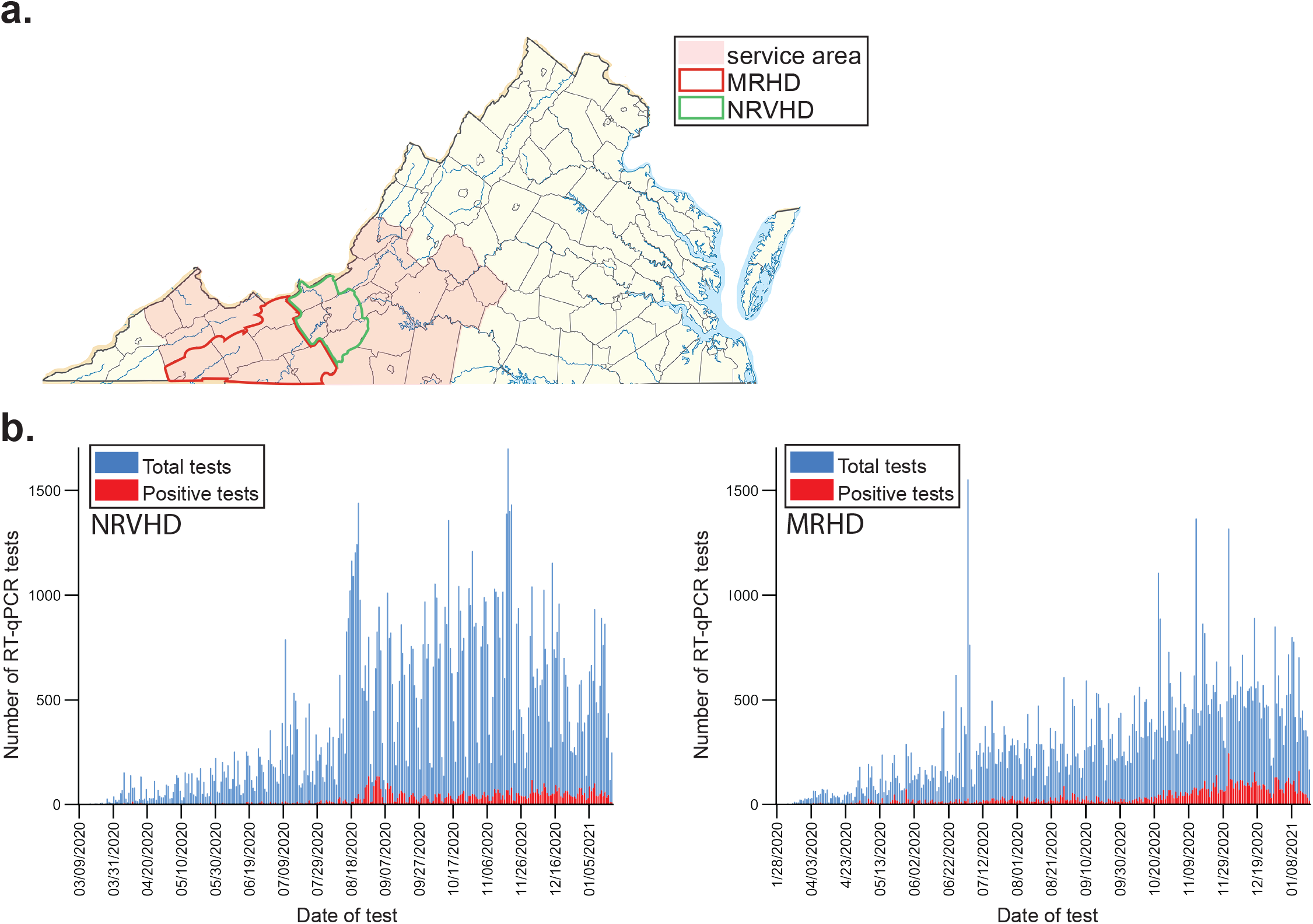
**a**. Map of the State of Virginia, U.S. with all 95 counties and 38 independent cities, considered county-equivalents, on display. Our laboratory’s service area is shaded in orange. The limits of the New River Valley Health District (NRVHD) and Mt. Rogers Health Districts (MRHD) are indicated in green and red, respectively. **b**. Graphs indicate the daily number of RT-qPCR tests processed (blue) in NRVDH (left) and MRHD (right) along with the number of positive results (red).

Of these, samples from NRVHD and MRHD were from a cohort of 7,792 and 648 participants, respectively. Positivity rates in NRV and MRHD were <5% at times of collection (July 29 to August 18, 2020, Fig. 2.B). Nasopharyngeal swabs were pooled in groups of 4 and samples were processed as described in Materials and Methods. The Cts from pooled samples were compared against the cut-off and deemed positive, negative, inconclusive (and therefore a repeat), or invalid (Fig. 3.A-B). Results for NRVDH showed that 96.71% of pool samples were negative (7,536 unique samples) and only 3.29% (256 unique samples) were below the threshold and, therefore, individually retested (Fig. 3.A). Of those 256 samples, 64.06% were negative (164 samples), 26.17% were positive (67 samples), 6.64% were inconclusive (17 samples), and 3.13% invalid (8 samples) (Fig. 3.A). Interestingly, the analysis of MRDH pooled samples was comparable to that of NRVHD, this is 95.06% were negative (616 samples) and 4.94% were retested (32 samples) (Fig. 3.B). Of the retest of the MRHD pooled samples, 70.97% were negative (22 samples) and 29.03% were positive (9 samples) and 1 invalid (Fig. 3.B). These results indicate that pooling is a reliable and effective approach for large screening of participants when positivity rates are below the threshold.

**Figure 3.**
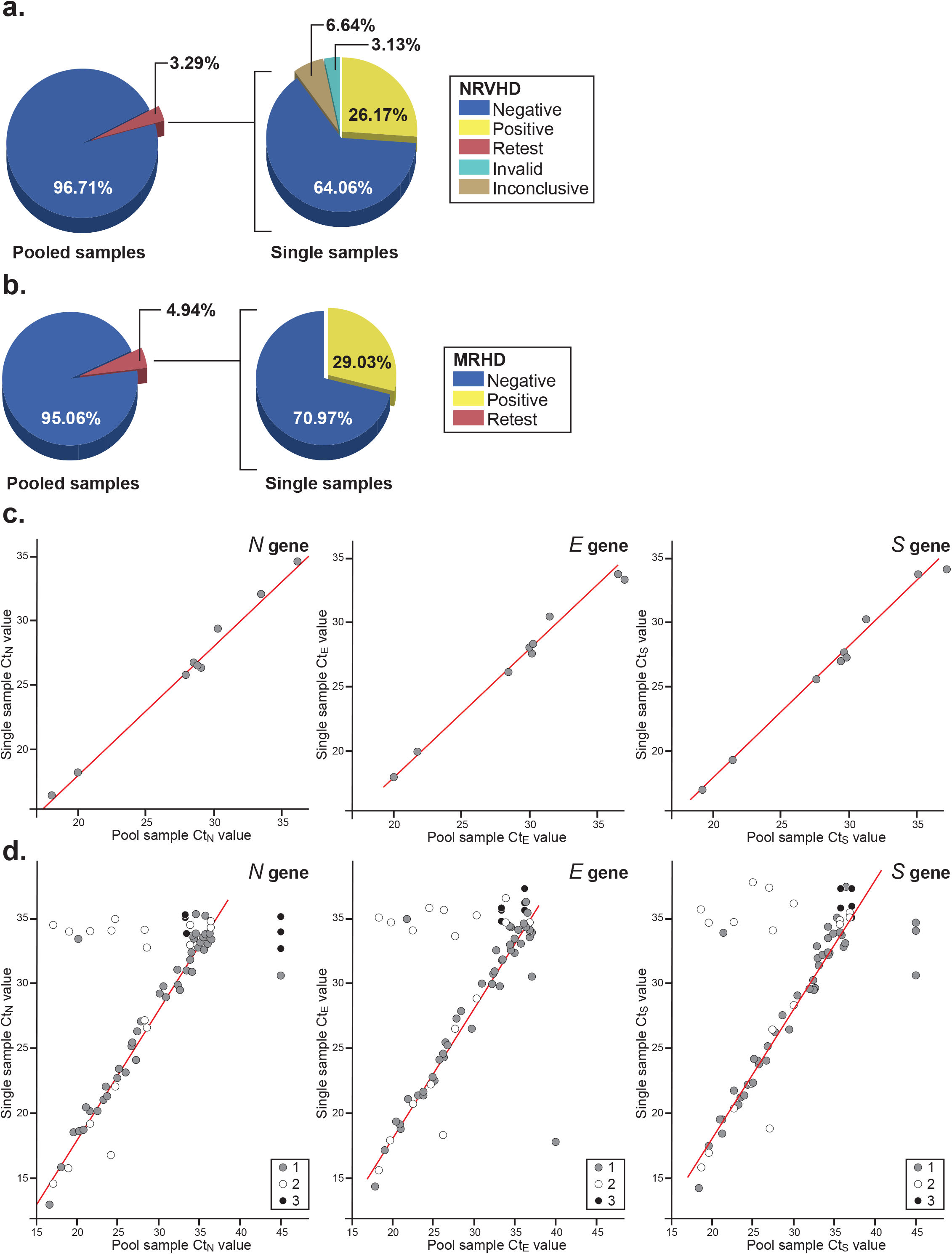
Samples were collected from participants at two locations MRHD (648 participants) and NRVHD (7,792 participants) (**a** and **b**, respectively) and used for pooling studies. Briefly, pools of 4 samples from each district were analyzed by RT-qPCR and deemed negative based on our threshold criteria for Ct values or “retest,” if positive or inconclusive. Samples from “retest pools” were analyzed as singles. Accordingly, the RNA from each of the original samples in the pool was re-extracted before amplification and the results grouped as negative, positive, invalid, and inconclusive. **c-d**. Scatter plot showing the association between the Ct value for the pooled sample compared to the Ct value for the positive sample(s) within the pool from the MRHD (C) and NRVHD (D). Points were colored based on the total number of positive samples in the pool of 4 samples (gray: 1 positive sample, white: 2 positive samples, black: 3 positive samples). Red lines indicate the theoretical Ct value shift (2 Ct units) expected when pooling together 1 positive sample with 3 negative samples.

Next, we asked whether the Ct values of the 4-pooled samples that were retested correlate with the Ct value of the positive sample in that pool. Since a high correlation and the theoretical shift in Ct values assume that only one sample in the pool is positive, we chose for comparison pools that had only 1 positive sample (Fig. 3.C-D). We found correlation values of 0.877 (p-value: <0.001), 0.908 (p-value: <0.001), 0.846 (p-value: <0.001) for the *N, E*, and *S* genes from NRVHD samples, respectively whereas those from the MRHD showed stronger correlations with values of 0.996 (p-value: <0.001), 0.992 (p-value: <0.001), and 0.994 (p-value: <0.001) for the *N, E*, and *S* genes, respectively. Thus, pooling of samples is a relevant public health alternative for large scale screening and for the detection of pre-symptomatic and asymptomatic individuals. Pooling of as low as 1 positive sample with up to 3 negatives prior to single sample testing using our assay did not adversely impact the detection of positives. At positivity rates of less than 5% or 10%, this approach could save up to 57% and 42% reagents and kits used for testing, respectively. This is in addition to saving processing time, speeding up the reporting of results, and aiding in the implementation of safe reopening plans.

### Expanding specimen collection and clinical evaluation

Next, we focused our efforts on developing a collection strategy for a noninvasive alternative to upper respiratory swabbing for testing. The need for this development rested in three important factors: *i*) swab sampling causes the patients to cough, sneeze, or have other uncomfortable reactions that makes collection more challenging, especially in children and the elderly people, while exposing healthcare workers to an increased risk of virus transmission, *ii*) swabbing requires a level of technical proficiency that could affect the quality of the sample collected if not properly done, and *iii*) continuous disruptions in the production of operational material with testing swabs were the weakest link in the supply chain.

To address these underlying problems, we expanded our test to include saliva as a noninvasive alternative to respiratory swabbing. Saliva is easy to collect, is presumably preferred by the participant, and does not require any particular technical proficiency for collection. Therefore, we proceeded to compare paired self-collected saliva samples with NP swabs collected by a healthcare worker (comparator assay) from 122 suspected COVID-19 patients. Participants provided written informed consent and had COVID-19 symptoms. To minimize the occurrence of discordant results, both clinical specimens were simultaneously collected, transported into MDL-TM, and processed using the protocol for extraction and amplification described in the Materials and Methods section. Of the 122 participants, 92 were negative and 30 were positive for SARS-CoV-2 as determined by our RT-qPCR test. Per FDA criteria, a minimum of 95% positive and negative agreement with similar Ct values for paired specimen types was considered an acceptable clinical performance. As shown in Supplementary Table XVII, Ct values for the *N* gene in positive swab samples ranged from ∼15 to 35 and, thus, covered a range of clinically relevant values for comparison. Our findings showed that matching saliva samples were 100% in agreement with swab reports resulting in 100% PPA and 100% NPA. Thus, the use of saliva specimens in our assay can provide a reliable read-out for those patients for which swabbing is not the preferred option.

## CONCLUSIONS

Widespread diagnostic testing and contact tracing are of utmost importance for slowing and, in some cases, containing the COVID-19 pandemic while vaccines are being developed, distributed, and administered. Implementation of aggressive testing schemes, quick reporting of test results, effective public health interventions, and adequate policymaking have contributed to limiting COVID-related death and SARS-CoV-2 spread in countries such as Germany, South Korea, and Singapore. The U.S., however, lags behind these countries in testing capability, resources, and organization, particularly in rural areas.

As the number of COVID-19 cases increased in the U.S., the Health and Human Services Secretary determined, based on the potential risk to national security and U.S. citizens, that circumstances justified the authorization of emergency use (EUA) *in vitro* diagnostics for the detection/identification of SARS-CoV-2. By the end of February 2020, the Food and Drug Administration (FDA) issued a guidance, which was updated in March and May 2020, with specifics for the development of *in vitro* tests. Since then, 314 molecular- and antibody-based tests, with wide differences in analytical performances, have been granted or are under review by the FDA for EUA authorization ^13^.

Diagnostic tests can be catalogued based on their intended use, (*e.g*., detection of viral RNA, antigens, or antibodies developed by the host, early-stage diagnostics of COVID-19 disease, screening), detection method (*e.g*., RT-qPCR, RT-LAMP, RT-RPA, ELISA, ddPCR, Neutralization test), and sensitivity (number of viral copies detected), to mention a few. As is the case for other molecular and immunological tests used for viral diagnostics, factors such as the quality/type of the clinical specimen, the material used for collection, the composition of the transport media, and storage conditions directly impact test performance. On top of these technical hurdles, large scale testing faced logistical, implementation, and turnaround time issues that were critical to the implementation of efficient contact tracing strategies and for containing the spread of the virus. These additional hurdles are particularly exacerbated in rural areas where access to public health resources, healthcare preparedness, and testing capabilities are limited or scattered under the best scenario, or, in most cases, are simply absent. It is at this juncture, where technical necessity meets rapid implementation, that innovation is required to overcome a broken supply chain and academic laboratories can fill the gap.

Our molecular test is technically designed to be sensitive and specific for the detection of SARS-CoV-2 using various clinical specimens (nasopharyngeal and saliva (in this paper), oropharyngeal, anterior nasal, mid-turbinate, bronchoalveolar, nasal aspirates in our EUA #200883 application) and strategies (single *versus* pooled samples) and for test results to be reported within hours of receiving a patient’s sample. Our RT-qPCR-based test has an analytical sensitivity of 10 particles, a predicted inclusivity of 99.9991% over 240,000+ SARS-CoV-2 sequences analyzed, and a specificity of 100% for the SARS-CoV-2 virus. Combined, these statistics make our test comparable or better than other molecular-based technologies for SARS-CoV-2 detection, including ddPCR, isothermal amplification (RRT-LAMP, RT-NEAR, RT-RPA), and CRISPR-based (DETECTR, STOP) methods. It definitely outperforms serological tests for which specificities have been reported to range between 67-98.7% [Abbott Laboratories, SinoBiological, and ^14-19^].

A notable difference between our assay and other molecular diagnostic technologies lies in our design strategy for identifying and classifying a clinical sample as either positive or negative. Many molecular diagnostics rely on the detection of a single gene in the SARS-CoV-2 virus, *i.e*., the *N* gene. Our assay, however, reports the result of the amplification of three SARS-CoV-2 genes (*N, E*, and *S*). The rationale behind this considers genetic variations of SARS-CoV-2 among circulating variants for which the sole amplification of a single gene could result in a false negative result if the mutation takes place in a region of the genome assessed by the test. Thus, a molecular diagnostics test developed to detect multiple genetic targets of SARS-CoV-2 is less susceptible to genetic variation. The FDA provides regular alerts to health care providers of potential false negative results that might result as a consequence of using commercial kits where detection could be compromised by a prevalent new variant of the virus. False positive results are equally concerning and arise from different sources, some of which are related to the format of the test and others to the sensitive nature of its amplification. The most notorious, and certainly unfortunate, case of false positive results appeared early in the pandemic and resulted from both contamination of the U.S. CDC test kits during the production phase ^20^ and cross-reactivity of a primer-probe set ^21^. As such, we developed a multi-level approach to address these issues starting with an efficient laboratory design where processing stations are physically segregated, included an array of controls in each plate that flags sources of cross- and carryover-contaminations, monitored for human error using cross-analysis, and developed a criterion for which at least two SARS-CoV-2 genes needed to amplify below a threshold cut-off for a sample to be reported positive.

The evolving nature of SARS-CoV-2 brings an additional consideration for test development and implementation that is of particular concern to commercial enterprises as they struggle to add new laboratory tests into routine use for the detection of new variants of the virus. Unlike large clinical laboratories, the operational flow for testing in laboratories within academic institutions facilitates just-in-time adaptations to the evolving nature of the SARS-CoV-2 virus. A case-in-point is the variant B.1.1.7 (VUI202012/01) of SARS-CoV-2, which was first identified in the United Kingdom in September 2020, and has spread to over 30 countries by the end of January 2021^22,23^. Implementation of procedures for the detection of new genomic variants of SARS-CoV-2 would need to rely on *de novo* production of specific supply kits and scale up operations. Thus, high-throughput academic laboratories with customized procedures can easily substitute individual components and be rapidly operational. A resilient test system like ours would prompt rapid public health responses and assess SARS-CoV-2 variant transmissibility and severity in the population. For instance, the D614G variant of SARS-CoV-2, which emerged in late January 2020, replaced the original SARS-CoV-2 variant identified in Wuhan, China in a period of several months to become the dominant variant globally ^24^, could not be specifically identified by any single commercially available kit ^25^. Under this circumstance, the only tool for public health officials to rely on for decision-making in response to an outbreak from a new variant virus was real-time whole-genome sequencing (WGS). Although a cutting-edge technology, WGS presents some logistical and technical challenges that make it difficult to broadly implement. This includes the need for streamlined sample preparation and a bioinformatics pipeline that ensures access to highly accurate and complete sequence platforms, access to metadata for interpretation, and automated tools for data analysis. A customized in-house screening test for a new variant can easily be developed by generating mutations of interest within an oligonucleotide sequence; thus, a new alternative screening test can be developed and implemented in a matter of hours.

In addition to this technical nimbleness, an academic institution such as ours (Virginia Tech) that is a public land-grant university with a motto of *Ut Prosim* (that I may serve), has demonstrated that is able to nimbly pivot its daily operations and support for its academic research community from standard activities to a service-oriented model that in cooperation with public agencies can rapidly deploy leading edge science in the provision of service for the immediate public good.

Therefore, the ability of academic institutions to establish reliable SARS-CoV-2 detection assays is paramount to mitigating the devastating health and economic toll triggered by the COVID-19 pandemic. This is particularly important in rural communities where the burden associated with a lack of testing capacity adds to the stigma of being diagnosed with a positive result, causing a dangerous downward spiral where people might choose not to be tested even when showing symptoms of the disease^26^. If there has been one thing this pandemic has taught us, it is that academic institutions have the desire, expertise, and resources to step up when the diagnostic system is overwhelmed.

## METHODS

### Specimen collection

The testing protocol was submitted for Emergency Use Authorization from the Federal Drug Administration (EUA# 200383) and approved by the Virginia Tech Institutional Review Board. Clinical specimens were collected from individuals suspected of having COVID-19 using nasopharyngeal swabbing (NP) by trained health professionals ^27^. When indicated, paired saliva (∼ 1 ml) and NP samples were collected from the same patient and analyzed simultaneously for comparison. Swabbing was performed using an ultrafine, flocked, polyester/rayon/Dacron™ bicomponent fiber tipped swab (Puritan) and placed into 100mm round-bottom, polystyrene collection tubes (Corning) containing 1.5 mL of transport media. Transport media (named MDL, Molecular Diagnostics Laboratory, media) was formulated to inactivate the virus shortly after collection while preserving the integrity of the sample at room temperature during transport. Thus, the media consisted of 1:1 volume of Dulbecco’s Modified Eagle’s Medium (DMEM) containing low glucose (1g/l), sodium pyruvate and L-glutamine (Corning) media and 2x DNA/RNA Shield™ (Zymo Research).

### Analytical laboratory layout for in-house SARS-CoV-2 testing

Our academic laboratory assembles the sample collection kits for distribution, receives and neutralizes clinical specimens, purifies the nucleic acid, amplifies the SARS-Cov-2 virus and human genes, and performs data analysis and reporting. As a result, there was a need to design a specific laboratory layout that would allow for manipulation of a large number of samples (*i.e*., ∼1,600 samples/day) and high-throughput processing without risking contamination in 8,780 sq.ft. (∼ 816 m^2^) of space. Accordingly, sample processing and analysis were physically separated in “stations,” each of which was color-coded and represented the following processes: sample neutralization (red), sample check in (black), RNA extraction (yellow), kit assembly (orange), RT-qPCR master mix assembly (green), and RNA dilution/standard curve preparation (blue). An additional station, data analysis and reporting, takes place remotely. Importantly, red/yellow/black/orange stations are physically separated from the blue/green stations by a buffer area of 300 sq.ft. (∼ 28 m^2^) to reduce any potential aerosol contamination. In addition, personnel entry to the red/yellow/black/orange stations is segregated from that of the blue/green entry to avoid cross-circulation. Supplementary Fig. 1 shows a blueprint of our current facility and a description of major instrumentation in place for sample analysis.

### Fabrication and quality control testing of 3D-printed nasopharyngeal swabs

Two types of nasopharyngeal swabs for COVID-19 testing were fabricated by inverse stereolithography desktop 3D printing. The two swab designs, one adult and one pediatric, were developed and released by University of South Florida (USF) and Northwell Health in collaboration with Formlabs, Inc. (Boston, MA). Swabs were printed in batches of 324 per build using a Formlabs Form 2 printer with Surgical Guide (v1) resin. The build platform with swabs still attached were washed for 20 min in molecular biology grade isopropanol (Fisher Bioreagents) using a Form Wash (Formlabs). The swabs were allowed to dry in air for 30 min and then removed using a scraper. An initial inspection was performed to look for uncured resin and remove defective parts. The swabs were then mounted in groups of 80 onto a custom fixture design (Formlabs). The fixture with swabs was loaded in a Form Cure (Formlabs) and cured for 30 min at 60°C. After cooling in air to room temperature, the swabs were removed from the rack and again inspected for defects, ragging (loosely attached bits of cured resin, especially around the handle tab), and excessive bending (>10°). Several swabs from each batch were randomly selected for destructive mechanical testing. The remaining swabs were inserted tip-first and in groups of 20 or 25 along with an additional sterilization indicator into 5¼” x 10” strip self-sealing autoclavable pouches (Fisherbrand™). Sterilization was done according to the CDC recommended cycle of 30 min at 121°C / 250°F in a gravity displacement autoclave. Labels showing the manufacturer, swab type (adult or pediatric), reference and lot number, and contact and other information were applied subsequent to autoclaving to avoid damage to the label.

Mechanical testing of ten pediatric and ten adult swabs was conducted by a third party, SGS Polymer Solutions Incorporated (SGS PSI, Christiansburg, VA). As reported by the company, the samples were conditioned in accordance with AS™ D618-33 for >40 h at 23 ± 1 °C and 50 ± 10% relative humidity. Tensile testing was performed using a custom method which is congruent with AS™ D638-14+. Tensile tests used an MTS Insight 30 test frame with a 250 N load cell and TestWorks 4 software. Torsion testing was also performed using a custom method. In brief, a torque wrench was fitted with an adapter holding a chuck into which the swab (with end tab removed) could be inserted and held. The full length of the swab was used except for the tab at the end. The swab was rotated until failure. The rotations were counted manually every 90° and the max torque was recorded at failure.

### Molecular diagnostic testing for SARS-CoV-2

The tube containing the transport media and flocked swab was vigorously vortexed for 30 sec (∼1,300 rpm) and 300 µl of the solution was transferred to a 5 ml RNase-free tube (Eppendorf) containing 900 µl TRIzol^™^ reagent (Thermo Fisher Scientific). Isolation of total RNA was carried out using a 96-well spin column plate using a protocol readily adaptable to any acid-guanidinium-phenol based reagent. All steps of the purification protocol were carried out at room temperature and the initial transfer of the transport media to the TRIzol-containing tube was the sole step carried out inside a Biosafety Type A2 Cabinet. A volume of 1.2 ml of ethanol (95-100%) was added to the specimen/TRIzol mixture, vortexed for 10 sec, and transferred (600 µl) into a well of a 96-well spin plate (Zymo Research). Plates were centrifuged at 3,600xg for 6 min and the procedure was repeated, as needed, until the specimen/TRIzol/EtOH (∼2.4 ml) volume was centrifuged through the column and the flow-through was discarded. Washes were carried out following manufacturer’s instructions (Zymo Research). A final spin was included in the protocol in which unloaded plates were centrifuged at 3,600xg for 2 min to ensure complete removal of residual volume before eluting the RNA. Elution of total RNA from spin plates was carried out using 20 µl of DNase/RNase-free water and quantification was carried out using a Nanodrop 2000^™^ system. Based on our experience of processing more than 64,000 samples, total RNA concentrations varied from as low as 5 ng/μl to 280 ng/μl. Plates were stored at −80°C until RT-qPCR reactions were ready for assembly.

Total RNA samples were diluted 1:20 in nuclease-free water and a master mix for each primer set was prepared using the *Power* SYBR^™^ Green RNA-to-CT^™^ *1-Step* Kit (Applied Biosystems). The RT-qPCR assay was developed to amplify three distinct viral genes encoding the nucleocapsid (*N*), envelope (*E*), and spike (*S*) proteins, as well as a human ribonuclease (RNAseP) control gene (Hs_*RPP30*) using 2 µl of the diluted RNA sample as a template. Reagents were distributed into a 384-well format using an epMotion 5073m automated liquid handling system and RT-qPCR amplification was performed in a Bio-Rad CFX384 Touch Real-Time PCR Detection System. Detailed experimental conditions are summarized in Supplementary Table I.A.

### Cross-contamination and reaction set up controls

A number of controls, both positive and negative, were routinely included in each plate and aimed at monitoring the RT-qPCR reaction set-up, reagent integrity, cross-contamination during RNA extraction, and to detect the presence of a potential positive, yet asymptomatic, operator. As a result, controls were as follows: *i*) no template RT-qPCR reaction with each set of primers tested, *ii*) detection of *RNse P* gene in all samples using the Hs_*RPP30* primers, *iii*) source of plasmid for *RNAse P* and *N* genes (standard curves), *iv*) purified RNA from inactive virus (ATCC® VR-1986HKTM) as an amplification control for the E and S primers, and *v*) purified RNA from a previously confirmed negative patient.

### Primer design

Primers targeting the SARS-CoV-2 nucleocapsid and envelope were previously validated and published ^28^. A set of S primers was designed against the publicly available NCBI reference sequence NC_045512.2 “Severe acute respiratory syndrome coronavirus 2 isolate Wuhan-Hu-1, complete genome,” with the gene encoding for the spike protein being located between positions 21,563 - 25,384 nt. Housekeeping primers against human ribonuclease P gene (*RPP30*) were from the CDC 2019-nCoV Real-Time RT-PCR Diagnostic Panel (Division of Viral Diseases, National Center for Immunization and Respiratory Diseases, Centers for Disease Control and Prevention, Atlanta, GA, USA). A summary of the primers used in our studies is presented in Supplementary Table I.B.

### Assay results and interpretation

Cut-off threshold cycle (Ct) and 95% interval of confidence values were experimentally determined in each plate for the *N* and *RPP30* genes using a standard dilution curve with the 2019-nCoV_*N*_Positive (Integrated DNA Technologies) and Hs_*RPP30* Positive (Integrated DNA Technologies) control DNA. Specifically, a linear model is fit to determine the relationship between the Ct values for standard dilution concentrations of each control as a function of the concentration of virus in the log scale (see Supplementary Fig. 2). The intercept of the linear model (*i.e*., log10 (viral copy) = 0) estimates the expected Ct value when only one copy of the *N* gene is present in the sample, or the Limit of Detection (LOD) for the assay. To account for uncertainty, the 95% interval of confidence for this LOD was calculated as:

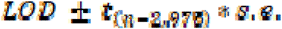

Where

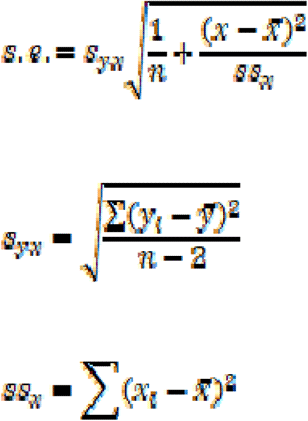

The final Ct threshold used to determine positivity corresponds to the inferior point of the 95% interval of confidence (*i.e*.,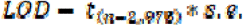). That is, if the Ct value reported for a sample is less than the inferior point for gene *N*, then the sample is determined to be positive for gene *N*; otherwise, the sample is determined to be negative for gene *N*. Preliminary analyses indicated that the Ct value corresponding to the inferior point of the 95% interval of confidence for the *E* and *S* genes were smaller (*i.e*., larger number of viral copies) than that observed for the *N* gene. To be conservative and to reduce the false positive rate, a one threshold approach was utilized and all sample Ct values for the *E, N*, and *S* genes were compared to the Ct value of the inferior point of the 95% interval of confidence calculated for gene *N*. A similar procedure was completed for the *RPP30* gene. The inferior point of the 95% interval of confidence for the LOD was computed, and if the observed Ct value for RPP30 was less than the inferior point of the 95% interval of confidence value for a sample, the RPP30 gene was determined to be positive for the sample; otherwise, the gene was determined to be negative. A sample was determined to be positive if at least two genes were determined positive. Supplementary Table II summarizes the criteria used to report results.

### Sequence analysis

All available complete SARS-CoV-2 sequences (taxid: 2697049) were downloaded on January 13, 2021 from NCBI (https://www.ncbi.nlm.nih.gov/labs/virus/vssi/#/virus?SeqType_s=Nucleotide&VirusLineage_ss=SARS-CoV-2,%20taxid:2697049&Completeness_s=complete). In addition, all SARS-CoV-2 sequences that were identified as being complete and having high coverage were downloaded from GISAID (gisaid.org) to include sequences available up to January 7, 2021. GISAID sequences were downloaded from https://www.gisaid.org/ in batches of less than 10,000 sequences. The two sets of SARS-CoV-2 sequences were aligned separately using the DECIPHER software toolset ^29^ in R studio (rstudio.com). To mitigate limitations due to computing power, sequence alignments were performed using subsets of 1,000 sequences at a time. Each of these aligned subsets were merged to include up to 10,000 sequences. These merged alignments were merged with 3 other alignment sets of up to 10,000 sequences to generate alignments of up to 40,000 sequences total. This resulted in seven alignment groups for the downloaded GISAID sequences and one for the downloaded NCBI sequences. These sequence alignment groups were used to analyze each of the primer binding regions. Aligned SARS-CoV-2 sequences were included for further analysis if sequence information was available for all six primer binding regions and if the sequences contained less than four mixed bases in any one primer binding region. For the remaining sequences, the alignments for each primer binding region were analyzed for mismatches that could affect amplification. The criteria for mismatches that could affect amplification included having any mismatch within three nucleotides of the 3’ end of a primer binding region or having three or more mismatches over the length of the primer binding region. The criteria for potentially affecting inclusivity of the assay included having mismatches that could affect amplification for at least two different primer sets.

## Supporting information

Sup_Table III

Sup_Table VIII

Sup_Table XIV

Sup_Table XVI

Sup_Table XIII

Sup_Table XVII

Sup_Table_VII

Sup_Table_XII

## Data Availability

All data related to the analysis in our manuscript is included as supplementary material and available to the scientific community.

## ACKNOWLEDGEMENTS

Implementation of this program included important contributions from not only the researchers listed in this article, but also from the leadership at our University (Virginia Tech). Establishment of the testing laboratory was through a highly collaborative approach under the auspices of the university’s student health center, local health districts and state health department epidemiologists and information technology teams. This project was supported by a Go Virginia Economic Resilience and Recovery Grant (20-GOVA-ERR-02A and 20-GOVA-ERR-02D), the Department of General Services of the Commonwealth of Virginia (DGS-201020-UVT), and funds from Virginia Tech, the Fralin Biomedical Research Institute at VTC, and the Fralin Life Sciences Institute. F.M.M. acknowledges support from NSF through CAREER-1652237 and the Virginia Tech Center for Earth and Environmental Nanotechnology “NanoEarth” (NNCI-1542100).

## AUTHOR CONTRIBUTIONS

A.C. analyzed the data summarized in Figs. 2.B,3.A and 3.B, Tables I and III, Supplementary Tables IV, IX, X, XI, XII, XIII, XV, and XVII. C.M.B contributed to Fig. 2.B, Table I, Supplementary Tables I, III, V, and XVI, and developed the statistical framework used for data analyses throughout the paper. A.T. contributed to Figs. 3.C and 3.D, Table III, Supplementary Fig. 2, Supplementary Tables X, XI, XII, XIII, XIV, XV, and XVII. K.L.B. contributed to Supplementary Tables VI, VII, and VIII. R.A.U. contributed to Table I and Supplementary Table I, III, and V. F.M.M. contributed to Figs. 1.B, 1.C, and 1.D. D.M.M. contributed to Table II. T.M. contributed to Supplementary Table I. C.V.F. contributed to Fig. 1.E and Table III. P.B. and N.B. contributed to sample collection. C.M.B., R.A.U., D.P., B.T., J.M., O.A., D.M.M., and T.M. worked out the technical details to optimize the experimental protocols used for sample analysis. T.M. and C.V.F aided in interpreting results. M.F. contributed to the implementation of the research. C.V.F. designed all figures and wrote the paper with input from all authors. C.V.F. and H.S. were involved in planning and co-supervising the project. All authors are indebted to R.F.C.B.

## SUPPLEMENTARY LEGENDS

**Supplementary Figure 1**. Diagram of the COVID-19 testing laboratory indicating the various work stations and locations of major equipment with color-coded markings along the floor indicating paths for specifically assigned laboratory personnel. Neutralization room (red station): Clinical samples are received, checked for compliance, neutralized, and aliquots separated for freezing in this location. Scribe section (black station): Neutralized samples are checked into the system and keys are assembled to follow samples throughout the pipeline. RNA extraction (yellow station): Technicians process plate samples in dedicated workstations to avoid contamination. Once the procedure is completed and samples eluted, plates are sealed and stored at −80°C for RT-qPCR amplification. PCR setup (Green station): Reaction mixture for RT-qPCR amplifications are set up in a PCR-clean workstation located opposite from where RNA samples were processed and in a dedicated alcove. The assembled PCR master mix is loaded into an automated epMotion liquid handler and dispensed in a 384-well plate format. Standard curves and sample amplification (blue station): A separate station is allocated to the manual loading of the standard curves for the N and Hs_RPP30 genes as well as the positive (inactive virus) and negative (no template reaction) controls that are added to each plate. Amplification is carried out using 18 Bio-Rad CFX384 Touch Real-Time PCR detection systems placed in parallel sides of an island, connected to power backup systems, and remotely monitored during the run and for data retrieval.

**Supplementary Figure 2**. Real time RT-qPCR efficiencies for SARS-CoV-2 viral genes.

**Supplementary Table I. a**. Description of the RT-qPCR experimental conditions. **b**. (*) Primers were published by Won *et al*. (2020) Exp. Neurobiol. April 30; 29(2):107-119. Doi: 10.5607/en20009 and were based on information deposited in the NCBI site and i) viral sequence access number: GenBank: MT039890.1, ii) S gene access number: ACCESSION MT039890 REGION: 21563..25384, https://www.ncbi.nlm.nih.gov/nuccore/MT039890.1?from=21563&to=25384, iii) E gene access number: ACCESSION MT039890 REGION: 26245..26472, https://www.ncbi.nlm.nih.gov/nuccore/MT039890.1?from=26245&to=26472, iv) N gene access number: MT039890 REGION: 28274..29533, v) https://www.ncbi.nlm.nih.gov/nuccore/MT039890.1?from=28274&to=29533. The sequence of the human RNA specific primer sets that serve as internal control were retrieved from the bulletin published by the Department of Health & Human Services, Centers for Disease Control and Prevention (CDC), Atlanta, GA on 24 Ja. 2020 in the “2019-Novel Coronavirus (2019-nCoV) Real-time rRT-PCR Panel Primers and Probes” document from the Division of Viral Diseases (https://www.cdc.gov/coronavirus/2019-ncov/lab/rt-pcr-panel-primer-probes.html).

**Supplementary Table II**. Interpretation of results. Summary of possible result outcomes and their reporting. NEG: Negative (no amplification), POS: Positive (amplification below the threshold).

**Supplementary Table III**. Validation of the LOD using 300 copies of viral particles per reaction. The cut-off value for the SARS-CoV-2 *N* gene was 37.29, 95% CI: [36.91, 37.67].

**Supplementary Table IV**. Validation of the LOD using 20 copies of viral particles per reaction. The cut-off value for the SARS-CoV-2 *N* gene was 37.7747, 95% CI: [37.04, 38.51].

**Supplementary Table V**. Validation of the LOD using 1.3 copies of viral particles per reaction. The cut-off value for the SARS-CoV-2 *N* gene was 37.29, 95% CI: [36.91, 37.67].

**Supplementary Table VI**. Predicted analytical sensitivity of the COVID-19 assay primers using SARS-CoV-2 sequences downloaded from NCBI. Virus sequences were sorted by their DECIPHER database number. Sequences are shown 5’ to 3’. For each virus sequence, a dot is shown if the same nucleotide is found in the COVID-19 primer at that position. The IUPAC nucleotide code is given when there is a difference at that position compared to the assay primer. Virus sequences with mismatches that may impact assay performance are identified by gray shading and are tabulated in the appropriate column. Poor quality virus sequences were separated into their own spreadsheet and the reason for their exclusion is tabulated.

**Supplementary Table VII**. Predicted analytical sensitivity of the COVID-19 assay primers using SARS-CoV-2 sequences downloaded from GISAID. Virus sequences were divided into seven groups based on submission date and placed into seven different spreadsheets. Each group was further divided into four DECIPHER databases. Virus sequences are sorted by their DECIPHER database and database number. Sequences are shown 5’ to 3’. For each virus sequence, a dot is shown if the same nucleotide is found in the COVID-19 primer at that position. The IUPAC nucleotide code is given when there is a difference at that position compared to the assay primer. Virus sequences with mismatches that may impact assay performance are identified by gray shading and are tabulated in the appropriate column. Poor quality virus sequences were separated into their own spreadsheet and the reason for their exclusion is tabulated. An accounting of the GISAID sequences is given in the last spreadsheet.

**Supplementary Table VIII**. SARS-Coronavirus (Tax ID: 694009) alignments using BLAST for customized N and E sets of primers. Alignment of S primers did not result in any significant homology.

**Supplementary Table IX**. Previously reported negative samples were contrived with either inactive SARS-CoV-2 or BCoVs virus (33 samples each) at concentrations of more than two-times the LOD as determined in Table I. Following, RNA was purified from all samples and amplified using primers to detect *N, E*, and *S* genes. Cut-off: Ct: 36.25, 95%CI [35.60, 36.90].

**Supplementary Table X**. Samples from participants were separated in two aliquots. One aliquot was analyzed using our test and its duplicate was submitted for confirmation to the Virginia Division of Consolidated Lab Services (DCLS) in Virginia. For each sample, Ct_N_, Ct_E_, Ct_S_, and Ct_RPP30_ indicate Ct values when obtained using our test. CDC N1 and N2 indicate Ct values obtained by DCLS. Under “Agreement”, “Yes” indicates consensus between our test and the DCLS result.

**Supplementary Table XI**. Pool contained n=2 samples, *i.e*., two negative (named “N”) or one positive + one negative (named “P”). Tab “Pool_Raw_Data:” Ct values for *N, E, S*, and *RPP30* genes are indicated for each sample (columns F-L and P-V) and the pool (column Z-AF). Tab “Positives:” Indicated the Ct difference for all genes of each positive sample compared to the pool value for that sample. Tab “Regression:” Regression analysis Pool (Ct) values *versus* single Ct values for N, E, and S genes. N/A: No amplification.

**Supplementary Table XII**. Pool contained n=3 samples, *i.e*., three negative (named “N”) or one positive + two negative (named “P”). Tab “Pool_Raw_Data:” Ct values for *N, E, S*, and *RPP30* genes are indicated for each sample (columns F-L, P-V, and Z-AF) and the pool (column AJ-AP). Tab “Positives:” Indicated the Ct difference for all genes of each positive sample compared to the pool value for that sample. Tab “Regression:” Regression analysis Pool (Ct) values *versus* single Ct values for N, E, and S genes. N/A: No amplification.

**Supplementary Table XIII**. Pool contained n=4 samples, *i.e*., four negative (named “N”) or one positive + three negative (named “P”). Tab “Pool_Raw_Data:” Ct values for *N, E, S*, and *RPP30* genes are indicated for each sample (columns C-H, J-O, Q-V, and X-AC) and the pool (column AE-AJ). Tab “Positives:” Indicated the Ct difference for all genes of each positive sample compared to the pool value for that sample. Tab “Regression:” Regression analysis Pool (Ct) values *versus* single Ct values for N, E, and S genes. N/A: No amplification.

**Supplementary Table XIV. a**. Two-by-two report of positive percent agreement (PPA) and negative percent agreement (NPA). **b**. Passing-Bablok regression analysis for the *N, E*, and *S* genes.

**Supplementary Table XV**. Theoretical results from pooling experiments using 100 positive samples diluted into pools of n=2, n=3, and n=4. Each worksheet includes the observed Ct values for the *N, E, S*, and *RPP30* genes (columns E-H), the observed (column I), theoretical (column K), and estimated (column L) plate specific Ct_N_ thresholds used for determining positivity, and the theoretical (columns M-O) and estimated (columns P-R) results for the *N, E*, and *S* genes. Theoretical Ct values were defined using a log2(n) shift in the Ct value of the threshold. Estimated Ct values were defined empirically, as described in the text.

**Supplementary Table XVI**. Summary of demographic and clinical data from three health districts in Virginia. MRHD: Mt. Rogers Health District, NRVHD: New River Valley Health District, FHD: Fairfax Health District. Data was obtained from * https://datausa.io/, ** U.S. Department of Health and Human Services (https://healthdata.gov/dataset/covid-19-reported-patient-impact-and-hospital-capacity-facility) and https://www.vdh.virginia.gov/, *** population in health district divided by the number of ICU beds.

**Supplementary Table XVII**. Matching saliva and swab samples were collected from 122 participants and analyzed by RT-qPCR. Of these, 92 and 30 were reported negative and positive, respectively. Columns E-H show Ct values for the *RPP30, N, E*, and *S* genes obtained from swab samples. Columns I-J show cut-off values for the *RPP30* and *N* genes for each swab sample. Columns O-R show Ct values for the *RPP30, N, E*, and *S* genes obtained from saliva samples. Columns S-T show cut-off values for the *RPP30* and *N* genes for each saliva sample. Columns K and U have the test results and the column V states “True” when the results from both sample collection methods for each patient match.

