## Supplementary figures and images for "Development and Implementation of a scalable and versatile test for COVID-19 diagnostics in rural communities"

### Sup_Fig_1

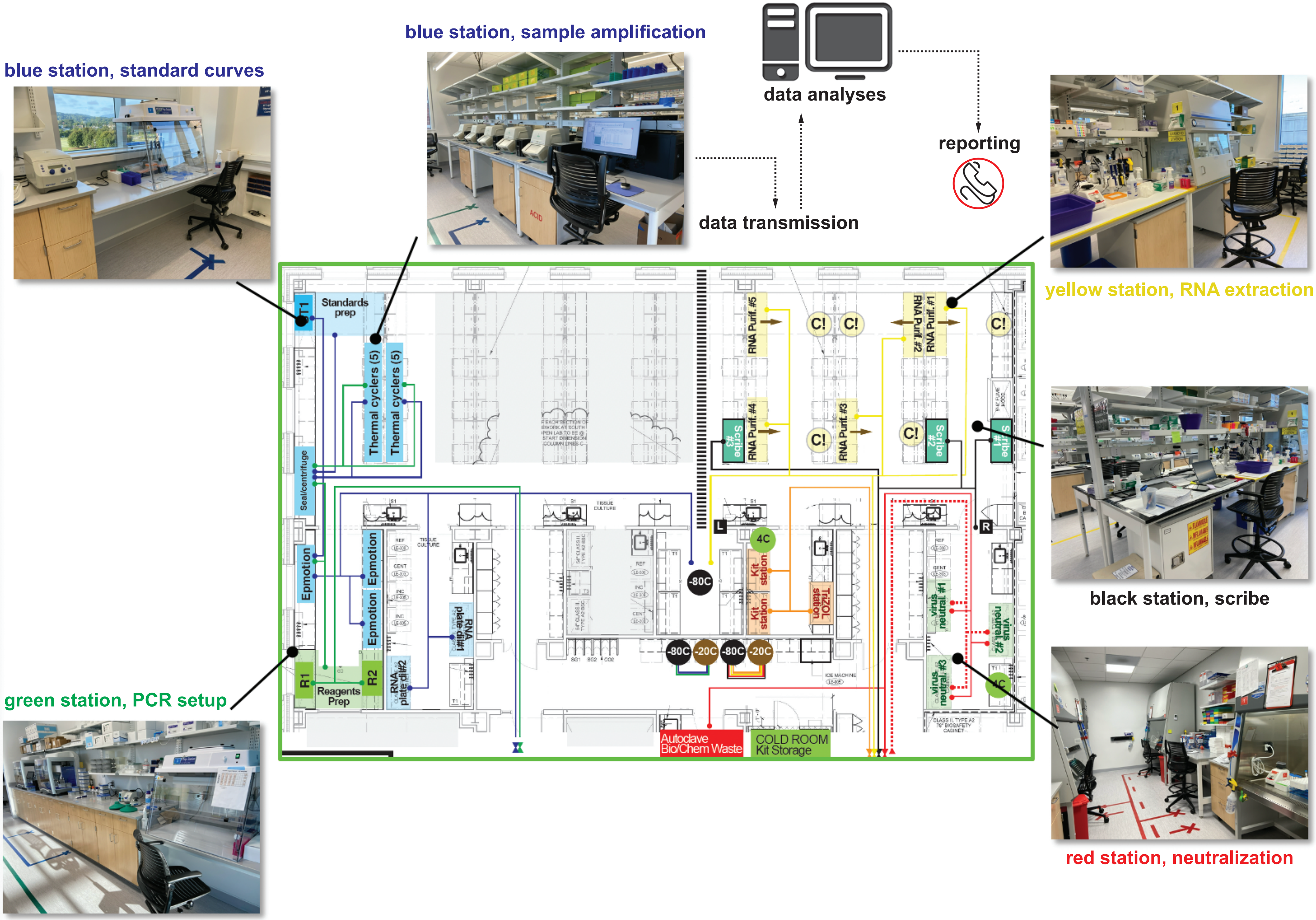

### Sup_Fig_2

Sup_Figure_2

| Copy # ($10^X$) | N | E | S |
|---|---|---|---|
| 0 | 37.93333 | 36.56667 | - |
| 1 | 34.34667 | 34.04000 | 33.48333 |
| 2 | 30.56667 | 30.44667 | 29.99667 |
| 3 | 27.08333 | 26.83667 | 26.59000 |
| 4 | 23.63667 | 23.26667 | 23.16000 |

| | | | |
|---|---|---|---|
| slope | -3.58570 | -3.38030 | -3.43770 |
| efficiency | 1.90058 | 1.97620 | 1.95385 |
| percentage | 90.05775 | 97.62038 | 95.3854 |

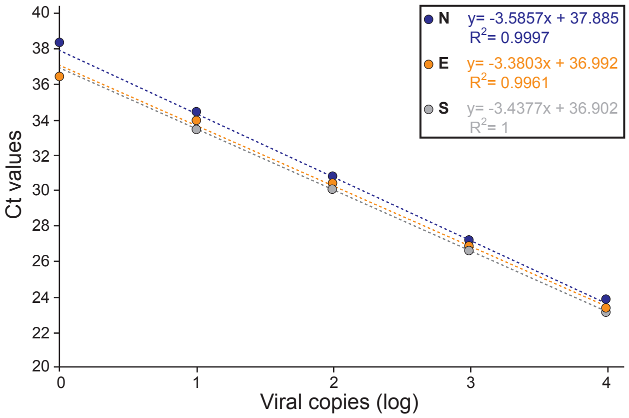
