## Supplementary material for "Development and Implementation of a scalable and versatile test for COVID-19 diagnostics in rural communities": Sup_Table I

**Supplementary Table I**

**A.**

PCR Master Mix:

| **Master mix components** | **Volume per reaction (µl)/well** |
|---|---|
| RT-PCR enzyme mix (125x) | 0.08 |
| SYBR™ master mix (2x) | 5 |
| 4µM Forward Primer | 0.5 |
| 4µM Reverse Primer | 0.5 |
| DNase/RNase free water | 1.92 |
| Total | 8 |

+ 2 µl (1:20 dilution) of RNA template, positive control plasmid, mon-template control water.

Cyclin parameters:

| **Cycling Stage** | **Step** | **Temperature** | **Time** |
|---|---|---|---|
| Holding | 1 | 48°C | 30:00 |
| | 2 | 95°C | 10:00 |
| Cycling | 3 | 95°C | 00:15 |
| | 4 | 60°C | 01:00 |
| | 5 | Go to Step 3, 44 more times | |
| Melt Curve, increment 0.5°C | 6 | 95°C | 00:15 |
| | | 65°C | 00:15 |

**B.**

| **Primer name** | **Forward sequence (5' to 3')** | **Reverse sequence (5' to 3')** | **Target gene** | **Gene product** |
|---|---|---|---|---|
| N* | GCTGCAATCGTGCTACAACT | TGAACTGTTGCGACTACGTG | *N* gene | SARS-CoV-2 nucleocapsid protein |
| E* | TTCGGAAGAGACAGGTACGTT | CACACAATCGATGCGCAGTA | *E* gene | SARS-CoV-2 Small envelope protein |
| S | GCTGGTGCTGCAGCTTATTA | AGGGTCAAGTGCACAGTCTA | *S* gene | SARS-CoV-2 Spike surface glycoprotein |
| CDC_RNAseP (from CDC) | AGATTTGGACCTGCGAGCG | GAGCGGCTGTCTCCACAAGT | *RPP30* gene | Human Ribonuclease P (RNase P) protein |
