## Supplementary material for "Development and Implementation of a scalable and versatile test for COVID-19 diagnostics in rural communities": Sup_Table II

**Supplementary Table II**

| *N* gene | *S* gene | *E* gene | *RPP30* gene | Status | Result | Action |
|---|---|---|---|---|---|---|
| NEG | NEG | NEG | NEG | Invalid | N/A | Repeat test. If the repeat results remain invalid, a new sample would need to be collected. |
| NEG | NEG | NEG | POS | Valid | SARS-CoV-2 no detected | Report results. |
| Only one SARS-CoV-2 target gene is POS | | | POS | Valid | SARS-CoV-2 inconclusive | Repeat test. If the repeat result remains inconclusive, additional confirmation testing should be conducted if clinically indicated. |
| Two or more SARS-CoV-2 target gene are POS | | | POS | Valid | SARS-CoV-2 positive | Report results. |
