## Supplementary material for "Development and Implementation of a scalable and versatile test for COVID-19 diagnostics in rural communities": Sup_Table III

### Supplementary Table III

| cut-off value for N: 37.29, 95% CI: [36.91, 37.67] | | | | | | | |
|---|---|---|---|---|---|---|---|
| **Viral RNA copies/reaction** | **Replicate** | **$C_T$_N** | **N Conclusion** | **$C_T$_E** | **E Conclusion** | **$C_T$_S** | **S Conclusion** |
| 300 copies | 1 | 31.36 | Positive | 31.45 | Positive | 31.39 | Positive |
| 300 copies | 2 | 31.25 | Positive | 31.15 | Positive | 31.79 | Positive |
| 300 copies | 3 | 31.02 | Positive | 31.23 | Positive | 31.59 | Positive |
| 300 copies | 4 | 31.32 | Positive | 31.55 | Positive | 31.55 | Positive |
| 300 copies | 5 | 31.53 | Positive | 31.25 | Positive | 31.41 | Positive |
| 300 copies | 6 | 31.28 | Positive | 30.74 | Positive | 31.34 | Positive |
| 300 copies | 7 | 31.13 | Positive | 30.97 | Positive | 31.35 | Positive |
| 300 copies | 8 | 31.04 | Positive | 31.03 | Positive | 31.31 | Positive |
| 300 copies | 9 | 31.19 | Positive | 31.31 | Positive | 31.43 | Positive |
| 300 copies | 10 | 31.31 | Positive | 31.45 | Positive | 31.63 | Positive |
| 300 copies | 11 | 31.44 | Positive | 31.09 | Positive | 30.98 | Positive |
| 300 copies | 12 | 31 | Positive | 30.73 | Positive | 30.46 | Positive |
| 300 copies | 13 | 30.92 | Positive | 30.75 | Positive | 31.1 | Positive |
| 300 copies | 14 | 31.09 | Positive | 31.04 | Positive | 30.74 | Positive |
| 300 copies | 15 | 31.38 | Positive | 30.94 | Positive | 31.09 | Positive |
| 300 copies | 16 | 31.2 | Positive | 30.57 | Positive | 30.53 | Positive |
| 300 copies | 17 | 31.15 | Positive | 30.94 | Positive | 31.03 | Positive |
| 300 copies | 18 | 31.1 | Positive | 30.79 | Positive | 30.97 | Positive |
| 300 copies | 19 | 31.47 | Positive | 31.04 | Positive | 31.06 | Positive |
| 300 copies | 20 | 31.36 | Positive | 31.26 | Positive | 30.97 | Positive |
| | **SD** | **0.17** | | **0.27** | | **0.36** | |
| | **% Positive** | | **20/20** | | **20/20** | | **20/20** |
