## Supplementary material for "Development and Implementation of a scalable and versatile test for COVID-19 diagnostics in rural communities": Sup_Table IV

### Supplementary Table IV

| cut-off value for N: 37.7747, 95% CI: [37.04, 38.51] | | | | | | | |
|---|---|---|---|---|---|---|---|
| **Viral RNA copies/reaction** | **Replicate** | **$C_T$_N** | **N Conclusion** | **$C_T$_E** | **E Conclusion** | **$C_T$_S** | **S Conclusion** |
| 20 copies | 1 | 35 | Positive | 33.6 | Positive | 33.11 | Positive |
| 20 copies | 2 | 33.39 | Positive | 33.72 | Positive | 33.42 | Positive |
| 20 copies | 3 | 33.92 | Positive | 33.4 | Positive | 33.07 | Positive |
| 20 copies | 4 | 33.38 | Positive | 33.09 | Positive | 32.59 | Positive |
| 20 copies | 5 | 33.87 | Positive | 33.6 | Positive | 34.2 | Positive |
| 20 copies | 6 | 33.2 | Positive | 33.18 | Positive | 33.43 | Positive |
| 20 copies | 7 | 33.21 | Positive | 33.47 | Positive | 32.95 | Positive |
| 20 copies | 8 | 33.81 | Positive | 33.8 | Positive | 34.51 | Positive |
| 20 copies | 9 | 34.14 | Positive | 33.69 | Positive | 33.28 | Positive |
| 20 copies | 10 | 34.04 | Positive | 34.41 | Positive | 33.67 | Positive |
| 20 copies | 11 | 33.75 | Positive | 32.96 | Positive | 33.25 | Positive |
| 20 copies | 12 | 34.06 | Positive | 33.83 | Positive | 33.2 | Positive |
| 20 copies | 13 | 33.67 | Positive | 33.63 | Positive | 33.18 | Positive |
| 20 copies | 14 | 33.68 | Positive | 33 | Positive | 33.27 | Positive |
| 20 copies | 15 | 34.35 | Positive | 33.74 | Positive | 33.69 | Positive |
| 20 copies | 16 | 33.67 | Positive | 33.35 | Positive | 33.32 | Positive |
| 20 copies | 17 | 33.85 | Positive | 33.85 | Positive | 33.02 | Positive |
| 20 copies | 18 | 33.71 | Positive | 33.61 | Positive | 32.5 | Positive |
| 20 copies | 19 | 35.33 | Positive | 33.44 | Positive | 32.94 | Positive |
| 20 copies | 20 | 33.66 | Positive | 34.27 | Positive | 32.82 | Positive |
| | **SD** | **0.5298607** | | **0.3733293** | | **0.4812363** | |
| | **% Positive** | | **20/20** | | **20/20** | | **20/20** |
