## Supplementary material for "Development and Implementation of a scalable and versatile test for COVID-19 diagnostics in rural communities": Sup_Table V

### Supplementary Table V

| cut-off value for N: 37.29, 95% CI: [36.91, 37.67] | | | | | | | |
|---|---|---|---|---|---|---|---|
| Viral RNA copies/reaction | Replicate | $C_T$_N | N Conclusion | $C_T$_E | E Conclusion | $C_T$_S | S Conclusion |
| 1.3 copies | 1 | 37.17 | Negative | N/A | Negative | N/A | Negative |
| 1.3 copies | 2 | N/A | Negative | 37.14 | Negative | N/A | Negative |
| 1.3 copies | 3 | N/A | Negative | N/A | Negative | 37.49 | Negative |
| 1.3 copies | 4 | N/A | Negative | N/A | Negative | 40.63 | Negative |
| 1.3 copies | 5 | N/A | Negative | N/A | Negative | N/A | Negative |
| 1.3 copies | 6 | 37.64 | Negative | 37.14 | Negative | N/A | Negative |
| 1.3 copies | 7 | N/A | Negative | 37.23 | Negative | N/A | Negative |
| 1.3 copies | 8 | N/A | Negative | N/A | Negative | N/A | Negative |
| 1.3 copies | 9 | N/A | Negative | N/A | Negative | 37.5 | Negative |
| 1.3 copies | 10 | N/A | Negative | 37.29 | Negative | N/A | Negative |
| 1.3 copies | 11 | N/A | Negative | N/A | Negative | N/A | Negative |
| 1.3 copies | 12 | 37.07 | Negative | N/A | Negative | N/A | Negative |
| 1.3 copies | 13 | N/A | Negative | N/A | Negative | 36.6 | Negative |
| 1.3 copies | 14 | N/A | Negative | N/A | Negative | 38.38 | Negative |
| 1.3 copies | 15 | N/A | Negative | 37.42 | Negative | N/A | Negative |
| 1.3 copies | 16 | N/A | Negative | N/A | Negative | N/A | Negative |
| 1.3 copies | 17 | 37.35 | Negative | 37.25 | Negative | 37.55 | Negative |
| 1.3 copies | 18 | N/A | Negative | N/A | Negative | N/A | Negative |
| 1.3 copies | 19 | N/A | Negative | N/A | Negative | 44.65 | Negative |
| 1.3 copies | 20 | N/A | Negative | 42.59 | Negative | N/A | Negative |
| | **SD** | 0.25 | | 2.02 | | 2.8 | |
| | **% Positive** | | **0/20** | | **0/20** | | **0/20** |
