## Supplementary material for "Development and Implementation of a scalable and versatile test for COVID-19 diagnostics in rural communities": Sup_Table VIII

### Supplementary Table VIII

The following alignments refer to sequences that have filtered results of ≥80% Identity, ≥80% query coverage, and an E value ≤10 for the N forward primer; Total of 3 hits.

**Bat coronavirus RaTG13, complete genome**

**Sequence ID:** MN996532.1 **Length:** 29855 **Number of Matches:** 1

**Range 1: 28702 to 28721** GenBank Graphics ▼ Next Match ▲ Previous Match

| Score | Expect | Identities | Gaps | Strand |
|---|---|---|---|---|
| 40.1 bits(20) | 0.47 | 20/20(100%) | 0/20(0%) | Plus/Plus |

```
Query  1      GCTGCAATCGTGCTACAACT  20
              ||||||||||||||||||||
Sbjct  28702  GCTGCAATCGTGCTACAACT  28721
```

**Bat SARS-like coronavirus isolate bat-SL-CoVZXC21, complete genome**

**Sequence ID:** MG772934.1 **Length:** 29732 **Number of Matches:** 1

**Range 1: 28572 to 28591** GenBank Graphics ▼ Next Match ▲ Previous Match

| Score | Expect | Identities | Gaps | Strand |
|---|---|---|---|---|
| 40.1 bits(20) | 0.47 | 20/20(100%) | 0/20(0%) | Plus/Plus |

```
Query  1      GCTGCAATCGTGCTACAACT  20
              ||||||||||||||||||||
Sbjct  28572  GCTGCAATCGTGCTACAACT  28591
```

**Bat SARS-like coronavirus isolate bat-SL-CoVZC45, complete genome**

**Sequence ID:** MG772933.1 **Length:** 29802 **Number of Matches:** 1

**Range 1: 28641 to 28660** GenBank Graphics ▼ Next Match ▲ Previous Match

| Score | Expect | Identities | Gaps | Strand |
|---|---|---|---|---|
| 40.1 bits(20) | 0.47 | 20/20(100%) | 0/20(0%) | Plus/Plus |

```
Query  1      GCTGCAATCGTGCTACAACT  20
              ||||||||||||||||||||
Sbjct  28641  GCTGCAATCGTGCTACAACT  28660
```

---

The following alignments refer to sequences that have filtered results of ≥80% Identity, ≥80% query coverage, and an E value ≤10 for the N reverse primer; Total of 3 hits.

**Bat coronavirus RaTG13, complete genome**

**Sequence ID:** MN996532.1 **Length:** 29855 **Number of Matches:** 1

**Range 1: 28802 to 28821** GenBank Graphics ▼ Next Match ▲ Previous Match

| Score | Expect | Identities | Gaps | Strand |
|---|---|---|---|---|
| 40.1 bits(20) | 0.47 | 20/20(100%) | 0/20(0%) | Plus/Minus |

```
Query  1      TGAACTGTTGCGACTACGTG  20
              ||||||||||||||||||||
Sbjct  28821  TGAACTGTTGCGACTACGTG  28802
```

**Bat SARS-like coronavirus isolate bat-SL-CoVZXC21, complete genome**

**Sequence ID:** MG772934.1 **Length:** 29732 **Number of Matches:** 1

**Range 1: 28672 to 28691** GenBank Graphics ▼ Next Match ▲ Previous Match

| Score | Expect | Identities | Gaps | Strand |
|---|---|---|---|---|
| 40.1 bits(20) | 0.47 | 20/20(100%) | 0/20(0%) | Plus/Minus |

```
Query  1      TGAACTGTTGCGACTACGTG  20
              ||||||||||||||||||||
Sbjct  28691  TGAACTGTTGCGACTACGTG  28672
```

**Bat SARS-like coronavirus isolate bat-SL-CoVZC45, complete genome**

**Sequence ID:** MG772933.1 **Length:** 29802 **Number of Matches:** 1

**Range 1: 28741 to 28760** GenBank Graphics ▼ Next Match ▲ Previous Match

| Score | Expect | Identities | Gaps | Strand |
|---|---|---|---|---|
| 40.1 bits(20) | 0.47 | 20/20(100%) | 0/20(0%) | Plus/Minus |

```
Query  1      TGAACTGTTGCGACTACGTG  20
              ||||||||||||||||||||
Sbjct  28760  TGAACTGTTGCGACTACGTG  28741
```

---

The following alignments refer to sequences that have filtered results of ≥80% Identity, ≥80% query coverage, and an E value ≤10 for the E forward primer; Total of 21 hits.

#### Bat coronavirus RaTG13, complete genome

**Sequence ID:** MN996532.1 **Length:** 29855 **Number of Matches:** 1

**Range 1: 26229 to 26249** GenBank Graphics ▼ Next Match ▲ Previous Match

| Score | Expect | Identities | Gaps | Strand |
|---|---|---|---|---|
| 42.1 bits(21) | 0.12 | 21/21(100%) | 0/21(0%) | Plus/Plus |

```
Query  1      TTCGGAAGAGACAGGTACGTT  21
              |||||||||||||||||||||
Sbjct  26229  TTCGGAAGAGACAGGTACGTT  26249
```

#### Bat SARS-like coronavirus isolate bat-SL-CoVZXC21, complete genome

**Sequence ID:** MG772934.1 **Length:** 29732 **Number of Matches:** 1

**Range 1: 26095 to 26115** GenBank Graphics ▼ Next Match ▲ Previous Match

| Score | Expect | Identities | Gaps | Strand |
|---|---|---|---|---|
| 42.1 bits(21) | 0.12 | 21/21(100%) | 0/21(0%) | Plus/Plus |

```
Query  1      TTCGGAAGAGACAGGTACGTT  21
              |||||||||||||||||||||
Sbjct  26095  TTCGGAAGAGACAGGTACGTT  26115
```

#### Bat SARS-like coronavirus isolate bat-SL-CoVZC45, complete genome

**Sequence ID:** MG772933.1 **Length:** 29802 **Number of Matches:** 1

**Range 1: 26164 to 26184** GenBank Graphics ▼ Next Match ▲ Previous Match

| Score | Expect | Identities | Gaps | Strand |
|---|---|---|---|---|
| 42.1 bits(21) | 0.12 | 21/21(100%) | 0/21(0%) | Plus/Plus |

```
Query  1      TTCGGAAGAGACAGGTACGTT  21
              |||||||||||||||||||||
Sbjct  26164  TTCGGAAGAGACAGGTACGTT  26184
```

#### BtRs-BetaCoV/YN2013, complete genome

**Sequence ID:** KJ473816.1 **Length:** 29142 **Number of Matches:** 1

**Range 1: 25831 to 25851** GenBank Graphics ▼ Next Match ▲ Previous Match

| Score | Expect | Identities | Gaps | Strand |
|---|---|---|---|---|
| 42.1 bits(21) | 0.12 | 21/21(100%) | 0/21(0%) | Plus/Plus |

```
Query  1      TTCGGAAGAGACAGGTACGTT  21
              |||||||||||||||||||||
Sbjct  25831  TTCGGAAGAGACAGGTACGTT  25851
```

#### BtRs-BetaCoV/GX2013, complete genome

**Sequence ID:** KJ473815.1 **Length:** 29161 **Number of Matches:** 1

**Range 1: 25847 to 25867** GenBank Graphics ▼ Next Match ▲ Previous Match

| Score | Expect | Identities | Gaps | Strand |
|---|---|---|---|---|
| 42.1 bits(21) | 0.12 | 21/21(100%) | 0/21(0%) | Plus/Plus |

```
Query  1      TTCGGAAGAGACAGGTACGTT  21
              |||||||||||||||||||||
Sbjct  25847  TTCGGAAGAGACAGGTACGTT  25867
```

#### BtRs-BetaCoV/HuB2013, complete genome

**Sequence ID:** KJ473814.1 **Length:** 29658 **Number of Matches:** 1

**Range 1: 26021 to 26041** GenBank Graphics ▼ Next Match ▲ Previous Match

| Score | Expect | Identities | Gaps | Strand |
|---|---|---|---|---|
| 42.1 bits(21) | 0.12 | 21/21(100%) | 0/21(0%) | Plus/Plus |

```
Query  1      TTCGGAAGAGACAGGTACGTT  21
              |||||||||||||||||||||
Sbjct  26021  TTCGGAAGAGACAGGTACGTT  26041
```

#### SARS-related bat coronavirus isolate Longquan-140 orf1ab polyprotein, spike glycoprotein, envelope protein, membrane protein, and nucleocapsid protein genes, complete cds

**Sequence ID:** KF294457.1 **Length:** 29676 **Number of Matches:** 1

**Range 1: 26061 to 26081** GenBank Graphics ▼ Next Match ▲ Previous Match

| Score | Expect | Identities | Gaps | Strand |
|---|---|---|---|---|
| 42.1 bits(21) | 0.12 | 21/21(100%) | 0/21(0%) | Plus/Plus |

```
Query  1      TTCGGAAGAGACAGGTACGTT  21
              |||||||||||||||||||||
Sbjct  26061  TTCGGAAGAGACAGGTACGTT  26081
```

#### Bat coronavirus Cp/Yunnan2011, complete genome

**Sequence ID:** JX993988.1 **Length:** 29452 **Number of Matches:** 1

**Range 1: 25989 to 26009** GenBank Graphics ▼ Next Match ▲ Previous Match

| Score | Expect | Identities | Gaps | Strand |
|---|---|---|---|---|
| 42.1 bits(21) | 0.12 | 21/21(100%) | 0/21(0%) | Plus/Plus |

```
Query  1      TTCGGAAGAGACAGGTACGTT  21
              |||||||||||||||||||||
Sbjct  25989  TTCGGAAGAGACAGGTACGTT  26009
```

#### Bat SARS coronavirus HKU3-13, complete genome

**Sequence ID:** GQ153548.1 **Length:** 29677 **Number of Matches:** 1

**Range 1: 26045 to 26065** GenBank Graphics ▼ Next Match ▲ Previous Match

| Score | Expect | Identities | Gaps | Strand |
|---|---|---|---|---|
| 42.1 bits(21) | 0.12 | 21/21(100%) | 0/21(0%) | Plus/Plus |

```
Query  1      TTCGGAAGAGACAGGTACGTT  21
              |||||||||||||||||||||
Sbjct  26045  TTCGGAAGAGACAGGTACGTT  26065
```

#### Bat SARS coronavirus HKU3-12, complete genome

**Sequence ID:** GQ153547.1 **Length:** 29704 **Number of Matches:** 1

**Range 1: 26072 to 26092** GenBank Graphics ▼ Next Match ▲ Previous Match

| Score | Expect | Identities | Gaps | Strand |
|---|---|---|---|---|
| 42.1 bits(21) | 0.12 | 21/21(100%) | 0/21(0%) | Plus/Plus |

```
Query  1      TTCGGAAGAGACAGGTACGTT  21
              |||||||||||||||||||||
Sbjct  26072  TTCGGAAGAGACAGGTACGTT  26092
```

#### Bat SARS coronavirus HKU3-11, complete genome

**Sequence ID:** GQ153546.1 **Length:** 29695 **Number of Matches:** 1

**Range 1: 26063 to 26083** GenBank Graphics ▼ Next Match ▲ Previous Match

| Score | Expect | Identities | Gaps | Strand |
|---|---|---|---|---|
| 42.1 bits(21) | 0.12 | 21/21(100%) | 0/21(0%) | Plus/Plus |

```
Query  1      TTCGGAAGAGACAGGTACGTT  21
              |||||||||||||||||||||
Sbjct  26063  TTCGGAAGAGACAGGTACGTT  26083
```

#### Bat SARS coronavirus HKU3-10, complete genome

**Sequence ID:** GQ153545.1 **Length:** 29695 **Number of Matches:** 1

**Range 1: 26063 to 26083** GenBank Graphics ▼ Next Match ▲ Previous Match

| Score | Expect | Identities | Gaps | Strand |
|---|---|---|---|---|
| 42.1 bits(21) | 0.12 | 21/21(100%) | 0/21(0%) | Plus/Plus |

```
Query  1      TTCGGAAGAGACAGGTACGTT  21
              |||||||||||||||||||||
Sbjct  26063  TTCGGAAGAGACAGGTACGTT  26083
```

#### Bat SARS coronavirus HKU3-9, complete genome

**Sequence ID:** GQ153544.1 **Length:** 29695 **Number of Matches:** 1

**Range 1: 26063 to 26083** GenBank Graphics ▼ Next Match ▲ Previous Match

| Score | Expect | Identities | Gaps | Strand |
|---|---|---|---|---|
| 42.1 bits(21) | 0.12 | 21/21(100%) | 0/21(0%) | Plus/Plus |

```
Query  1      TTCGGAAGAGACAGGTACGTT  21
              |||||||||||||||||||||
Sbjct  26063  TTCGGAAGAGACAGGTACGTT  26083
```

#### Bat SARS coronavirus HKU3-8, complete genome

**Sequence ID: GQ153543.1** **Length: 29681** **Number of Matches: 1**

**Range 1: 26075 to 26095** GenBank Graphics ▼ Next Match ▲ Previous Match

| Score | Expect | Identities | Gaps | Strand |
|---|---|---|---|---|
| 42.1 bits(21) | 0.12 | 21/21(100%) | 0/21(0%) | Plus/Plus |

```
Query  1      TTCGGAAGAGACAGGTACGTT  21
              |||||||||||||||||||||
Sbjct  26075  TTCGGAAGAGACAGGTACGTT  26095
```

#### Bat SARS coronavirus HKU3-7, complete genome

**Sequence ID: GQ153542.1** **Length: 29716** **Number of Matches: 1**

**Range 1: 26084 to 26104** GenBank Graphics ▼ Next Match ▲ Previous Match

| Score | Expect | Identities | Gaps | Strand |
|---|---|---|---|---|
| 42.1 bits(21) | 0.12 | 21/21(100%) | 0/21(0%) | Plus/Plus |

```
Query  1      TTCGGAAGAGACAGGTACGTT  21
              |||||||||||||||||||||
Sbjct  26084  TTCGGAAGAGACAGGTACGTT  26104
```

#### Bat SARS coronavirus HKU3-6, complete genome

**Sequence ID: GQ153541.1** **Length: 29704** **Number of Matches: 1**

**Range 1: 26072 to 26092** GenBank Graphics ▼ Next Match ▲ Previous Match

| Score | Expect | Identities | Gaps | Strand |
|---|---|---|---|---|
| 42.1 bits(21) | 0.12 | 21/21(100%) | 0/21(0%) | Plus/Plus |

```
Query  1      TTCGGAAGAGACAGGTACGTT  21
              |||||||||||||||||||||
Sbjct  26072  TTCGGAAGAGACAGGTACGTT  26092
```

#### Bat SARS coronavirus HKU3-5, complete genome

**Sequence ID: GQ153540.1** **Length: 29704** **Number of Matches: 1**

**Range 1: 26072 to 26092** GenBank Graphics ▼ Next Match ▲ Previous Match

| Score | Expect | Identities | Gaps | Strand |
|---|---|---|---|---|
| 42.1 bits(21) | 0.12 | 21/21(100%) | 0/21(0%) | Plus/Plus |

```
Query  1      TTCGGAAGAGACAGGTACGTT  21
              |||||||||||||||||||||
Sbjct  26072  TTCGGAAGAGACAGGTACGTT  26092
```

#### Bat SARS coronavirus HKU3-4, complete genome

**Sequence ID: GQ153539.1** **Length: 29704** **Number of Matches: 1**

**Range 1: 26072 to 26092** GenBank Graphics ▼ Next Match ▲ Previous Match

| Score | Expect | Identities | Gaps | Strand |
|---|---|---|---|---|
| 42.1 bits(21) | 0.12 | 21/21(100%) | 0/21(0%) | Plus/Plus |

```
Query  1      TTCGGAAGAGACAGGTACGTT  21
              |||||||||||||||||||||
Sbjct  26072  TTCGGAAGAGACAGGTACGTT  26092
```

#### bat SARS coronavirus HKU3-3, complete genome

**Sequence ID: DQ084200.1** **Length: 29711** **Number of Matches: 1**

**Range 1: 26055 to 26075** GenBank Graphics ▼ Next Match ▲ Previous Match

| Score | Expect | Identities | Gaps | Strand |
|---|---|---|---|---|
| 42.1 bits(21) | 0.12 | 21/21(100%) | 0/21(0%) | Plus/Plus |

```
Query  1      TTCGGAAGAGACAGGTACGTT  21
              |||||||||||||||||||||
Sbjct  26055  TTCGGAAGAGACAGGTACGTT  26075
```

**Bat SARS coronavirus HKU3-1, complete genome**

**Sequence ID:** DQ022305.2 **Length:** 29728 **Number of Matches:** 1

**Range 1: 26072 to 26092** GenBank Graphics ▼ Next Match ▲ Previous Match

| Score | Expect | Identities | Gaps | Strand |
|---|---|---|---|---|
| 42.1 bits(21) | 0.12 | 21/21(100%) | 0/21(0%) | Plus/Plus |

```
Query  1      TTCGGAAGAGACAGGTACGTT  21
              |||||||||||||||||||||
Sbjct  26072  TTCGGAAGAGACAGGTACGTT  26092
```

**bat SARS coronavirus HKU3-2, complete genome**

**Sequence ID:** DQ084199.1 **Length:** 29687 **Number of Matches:** 1

**Range 1: 26055 to 26075** GenBank Graphics ▼ Next Match ▲ Previous Match

| Score | Expect | Identities | Gaps | Strand |
|---|---|---|---|---|
| 42.1 bits(21) | 0.12 | 21/21(100%) | 0/21(0%) | Plus/Plus |

```
Query  1      TTCGGAAGAGACAGGTACGTT  21
              |||||||||||||||||||||
Sbjct  26055  TTCGGAAGAGACAGGTACGTT  26075
```

---

The following alignments refer to sequences that have filtered results of ≥80% Identity, ≥80% query coverage, and an E value ≤10 for the <u>E reverse primer</u>; Total of 3 hits.

**SARS coronavirus ExoN1 strain SARS/VeroE6_lab/USA/ExoN1_c5.7P20/2010, complete genome**

**Sequence ID:** KF514407.1 **Length:** 29689 **Number of Matches:** 1

**Range 1: 26198 to 26217** GenBank Graphics ▼ Next Match ▲ Previous Match

| Score | Expect | Identities | Gaps | Strand |
|---|---|---|---|---|
| 40.1 bits(20) | 0.47 | 20/20(100%) | 0/20(0%) | Plus/Minus |

```
Query  1      CACACAATCGATGCGCAGTA  20
              ||||||||||||||||||||
Sbjct  26217  CACACAATCGATGCGCAGTA  26198
```

**SARS coronavirus ExoN1 isolate c5P10, complete genome**

**Sequence ID:** JX162087.1 **Length:** 29688 **Number of Matches:** 1

**Range 1: 26198 to 26217** GenBank Graphics ▼ Next Match ▲ Previous Match

| Score | Expect | Identities | Gaps | Strand |
|---|---|---|---|---|
| 40.1 bits(20) | 0.47 | 20/20(100%) | 0/20(0%) | Plus/Minus |

```
Query  1      CACACAATCGATGCGCAGTA  20
              ||||||||||||||||||||
Sbjct  26217  CACACAATCGATGCGCAGTA  26198
```

**SARS coronavirus ExoN1 isolate P3pp53, complete genome**

**Sequence ID:** FJ882956.1 **Length:** 29644 **Number of Matches:** 1

**Range 1: 26178 to 26197** GenBank Graphics ▼ Next Match ▲ Previous Match

| Score | Expect | Identities | Gaps | Strand |
|---|---|---|---|---|
| 40.1 bits(20) | 0.47 | 20/20(100%) | 0/20(0%) | Plus/Minus |

```
Query  1      CACACAATCGATGCGCAGTA  20
              ||||||||||||||||||||
Sbjct  26197  CACACAATCGATGCGCAGTA  26178
```
