## Supplementary material for "Development and Implementation of a scalable and versatile test for COVID-19 diagnostics in rural communities": Sup_Table X

**Supplementary Table X**

| Sample ID | $Ct_N$ | $Ct_E$ | $Ct_S$ | $Ct_{RPP30}$ | Cut-off N | Cut-off RPP30 | CDC N1 Ct | CDC N2 Ct | Agreement? |
|---|---|---|---|---|---|---|---|---|---|
| V0001455 | 18.82 | 20.85 | 21.45 | 28.45 | 36.51 | 38.26 | 17.3 | 17.2 | YES |
| V0001531 | 42.23 | -- | 38.93 | 27.64 | 37.07 | 38.34 | -- | -- | YES |
| V0001533 | -- | 40.97 | 40.09 | 27.44 | 37.07 | 38.34 | -- | -- | YES |
| V0001537 | 37.86 | -- | 39.73 | 27.11 | 37.07 | 38.34 | -- | -- | YES |
| V0001547 | 44.16 | -- | 40.36 | 26.01 | 37.07 | 38.34 | -- | -- | YES |
| V0001550 | -- | 44.64 | 41.39 | 27.80 | 37.07 | 38.34 | -- | -- | YES |
| V0001730 | -- | -- | -- | 29.32 | 35.60 | 35.42 | -- | -- | YES |
| V0001792 | 24.59 | 25.37 | 25.74 | 32.37 | 33.94 | 36.13 | 18.5 | 18.5 | YES |
| V0001939 | -- | -- | -- | 26.94 | 37.07 | 38.34 | -- | -- | YES |
| V0001947 | 34.55 | 34.63 | 33.77 | 29.87 | 36.61 | 34.36 | 30.8 | 31.2 | YES |
| V0002420 | -- | -- | 36.12 | 27.13 | 37.07 | 38.34 | -- | -- | YES |
| V0002422 | -- | -- | -- | 29.47 | 37.07 | 38.34 | -- | -- | YES |
| V0002449 | 38.09 | 42.18 | 43.74 | 27.50 | 37.07 | 38.34 | -- | -- | YES |
| V0002450 | 43.12 | -- | 38.56 | 26.24 | 37.07 | 38.34 | -- | -- | YES |
| V0002455 | 39.45 | -- | 44.77 | 27.56 | 37.07 | 38.34 | -- | -- | YES |
| V0002456 | -- | 43.03 | 39.87 | 27.0 | 37.07 | 38.34 | -- | -- | YES |
| V0002457 | -- | -- | -- | 29.77 | 37.07 | 38.34 | -- | -- | YES |
| V0002458 | -- | 42.44 | 39.81 | 28.20 | 37.07 | 38.34 | -- | -- | YES |
| V0002459 | 43.94 | -- | 41.70 | 27.75 | 37.07 | 38.34 | -- | -- | YES |
| V0002460 | 42.24 | -- | -- | 28.31 | 37.07 | 38.34 | -- | -- | YES |
| V0002463 | 40.38 | -- | 38.49 | 27.16 | 37.07 | 38.34 | -- | -- | YES |
| V0002466 | 43.73 | -- | 43.25 | 27.04 | 37.07 | 38.34 | -- | -- | YES |
| V0002467 | 43.59 | 42.54 | 39.57 | 26.37 | 37.07 | 38.34 | -- | -- | YES |
| V0002470 | -- | 41.07 | 39.11 | 28.07 | 37.07 | 38.34 | -- | -- | YES |
| V0002471 | -- | -- | -- | 27.54 | 37.07 | 38.34 | -- | -- | YES |
| V0002472 | -- | -- | 44.41 | 27.97 | 37.07 | 38.34 | -- | -- | YES |
| V0002474 | 37.34 | -- | 40.06 | 28.02 | 37.07 | 38.34 | -- | -- | YES |
| V0002475 | -- | -- | 42.54 | 28.14 | 37.07 | 38.34 | -- | -- | YES |
| V0002478 | 41.09 | -- | -- | 27.57 | 37.07 | 38.34 | -- | 40.6 | YES |
| V0002479 | 41.29 | -- | 40.19 | 26.76 | 37.07 | 38.34 | -- | -- | YES |
| V0002481 | -- | -- | 43.63 | 28.02 | 37.07 | 38.34 | -- | -- | YES |
| V0002482 | -- | -- | 43.37 | 28.40 | 37.07 | 38.34 | -- | -- | YES |
| V0002483 | -- | -- | 37.84 | 27.52 | 37.07 | 38.34 | -- | -- | YES |
| V0002487 | -- | -- | 40.31 | 27.73 | 37.07 | 38.34 | -- | -- | YES |
| V0002489 | -- | -- | 39.34 | 27.80 | 37.07 | 38.34 | -- | -- | YES |
| V0002510 | -- | 37.3 | -- | 29.31 | 35.60 | 35.42 | -- | -- | YES |
| V0002520 | -- | 36.45 | -- | 27.37 | 35.60 | 35.42 | -- | -- | YES |
| V0003324 | 41.12 | -- | -- | 28.02 | 35.60 | 35.42 | -- | -- | YES |
| V0003341 | -- | -- | -- | 29.35 | 35.60 | 35.42 | -- | -- | YES |
| V0003371 | 31.62 | 31.51 | 31.89 | 31.11 | 33.65 | 35.31 | 27.6 | 27.6 | YES |
| V0003376 | 27.54 | 27.62 | 27.84 | 29.89 | 33.65 | 35.31 | 23.5 | 23.5 | YES |
| V0004170 | 24.56 | 24.79 | 24.88 | 32.41 | 34.36 | 34.94 | 15.6 | 16.0 | YES |
| V0004173 | 20.57 | 21.52 | 21.40 | 29.37 | 34.34 | 35.47 | 16.6 | 16.6 | YES |
| V0004179 | 25.08 | 25.83 | 25.28 | 29.48 | 34.34 | 35.47 | 20.1 | 20.2 | YES |
| V0004182 | 24.92 | 27.03 | 26.47 | 33.53 | 36.77 | 37.91 | 17.2 | 17.2 | YES |
| V0004184 | 30.47 | 34.66 | 33.28 | 34.83 | 36.77 | 37.91 | 21.5 | 21.5 | YES |
| V0004191 | 26.14 | 27.90 | 27.32 | 35.26 | 36.77 | 37.91 | 17.1 | 16.8 | YES |
| V0004195 | 31.10 | 32.23 | 31.44 | 34.49 | 36.77 | 37.91 | 22.3 | 21.8 | YES |
| V0004196 | 27.93 | 28.04 | 27.70 | 33.62 | 34.34 | 35.47 | 21.9 | 21.5 | YES |
| V0004197 | 32.24 | 32.65 | 32.40 | 28.32 | 34.34 | 35.47 | 27.2 | 27.7 | YES |

| | | | | | | | | | |
|---|---|---|---|---|---|---|---|---|---|
| V0004198 | 34.38 | 36.20 | 35.54 | 35.49 | 36.77 | 37.91 | 23.5 | 23.4 | YES |
| V0004199 | 31.72 | 31.90 | 32.85 | 38.66 | 34.36 | 34.94 | 16.4 | 16.7 | YES |
| V0004200 | 28.88 | 31.17 | 30.35 | 36.05 | 36.77 | 37.91 | 18.3 | 18.3 | YES |
| V0004201 | 26.63 | 28.49 | 28.30 | 35.37 | 36.77 | 37.91 | 15.8 | 16.3 | YES |
| V0004202 | 28.61 | 30.30 | 29.32 | 34.53 | 36.77 | 37.91 | 20.2 | 19.9 | YES |
| V0004205 | 23.02 | 25.07 | 23.98 | 35.68 | 36.77 | 37.91 | 15.7 | 16.1 | YES |
| V0004222 | 25.38 | 25.19 | 24.93 | 30.17 | 34.34 | 35.47 | 19.7 | 19.4 | YES |
| V0004230 | 30.65 | 30.97 | 30.80 | 36.19 | 34.36 | 34.94 | 19.9 | 19.0 | YES |
| V0004251 | 24.59 | 25.98 | 25.88 | 30.82 | 34.34 | 35.47 | 19.6 | 19.5 | YES |
| V0004258 | 27.57 | 28.01 | 27.68 | 29.81 | 34.34 | 35.47 | 19.8 | 21.0 | YES |
| V0004309 | 27.01 | 29.15 | 29.49 | 31.88 | 34.36 | 34.94 | 18.7 | 18.9 | YES |
| V0004312 | 32.22 | 32.34 | 32.40 | 36.22 | 34.36 | 34.94 | 22.3 | 22.0 | YES |
| V0004313 | 30.03 | 30.15 | 29.97 | 26.41 | 34.34 | 35.47 | 26.2 | 25.9 | YES |
| V0004318 | 21.42 | 24.36 | 23.99 | 35.36 | 36.77 | 37.91 | 15.5 | 16.3 | YES |
| V0004319 | 20.52 | 22.66 | 21.72 | 26.81 | 34.36 | 34.94 | 16.6 | 17.4 | YES |
| V0004323 | 24.92 | 25.96 | 26.55 | 34.11 | 34.36 | 34.94 | 16.1 | 16.3 | YES |
| V0004324 | 26.08 | 28.82 | 28.41 | 34.26 | 36.77 | 37.91 | 16.3 | 17.0 | YES |
| V0004329 | 30.18 | 30.68 | 30.17 | 30.80 | 34.34 | 35.47 | 35.1 | 38.1 | YES |
| V0004338 | 29.38 | 31.72 | 31.61 | 34.37 | 36.77 | 37.91 | 20.1 | 20.2 | YES |
| V0004342 | 21.86 | 24.98 | 24.99 | 35.27 | 36.77 | 37.91 | 15.6 | 16.4 | YES |
| V0004344 | 27.40 | 29.58 | 28.63 | 35.93 | 36.77 | 37.91 | 16.7 | 17.5 | YES |
| V0004349 | 27.18 | 30.11 | 29.70 | 36.25 | 36.77 | 37.91 | 17.4 | 17.6 | YES |
| V0004352 | 25.46 | 26.81 | 27.03 | 35.10 | 34.36 | 34.94 | 15.9 | 15.9 | YES |
| V0004360 | 32.69 | 34.27 | 35.14 | 34.44 | 34.36 | 34.94 | 23.0 | 22.8 | YES |
| V0004364 | 26.04 | 28.54 | 28.02 | 34.92 | 36.77 | 37.91 | 16.4 | 16.7 | YES |
| V0004365 | 28.43 | 29.83 | 29.05 | 36.5 | 36.77 | 37.91 | 18.9 | 18.6 | YES |
| V0004366 | 32.46 | 31.49 | 31.89 | 36.64 | 34.36 | 34.94 | 16.4 | 16.9 | YES |
| V0004368 | 23.71 | 24.54 | 24.66 | 30.76 | 34.34 | 35.47 | 17.4 | 17.3 | YES |
| V0004380 | 26.66 | 27.33 | 27.07 | 37.34 | 34.36 | 34.94 | 15.8 | 16.0 | YES |
| V0004565 | 29.14 | 31.81 | 31.29 | 35.13 | 36.77 | 37.91 | 17.7 | 17.9 | YES |
| V0004567 | 22.30 | 22.64 | 22.43 | 28.68 | 34.34 | 35.47 | 18.2 | 18.2 | YES |
