## Supplementary material for "Development and Implementation of a scalable and versatile test for COVID-19 diagnostics in rural communities": Sup_Table XIV

**Supplementary Table XIV**

**A.**

| | | Pooled Result | |
|---|---|---|---|
| | | - | + |
| Individual Result | - | 20 | 0 |
| | + | 0 | 21 |

**B.**

| Gene | Slope | Lower 95% CI | Upper 95% CI |
|---|---|---|---|
| *N* | 0.94 | 0.86 | 1.11 |
| *E* | 0.96 | 0.83 | 1.10 |
| *S* | 0.98 | 0.84 | 1.17 |
