## Supplementary material for "Development and Implementation of a scalable and versatile test for COVID-19 diagnostics in rural communities": Sup_Table XVI

**Supplementary Table XVI**

| | | MRHD | NRVHD | FHD |
|---|---|---|---|---|
| Demographics* | Population | 187,324 | 182,147 | 1,147,532 |
| | Median age | 47.4 | 34.4 | 38.4 |
| | Population under 18 | 19% | 16% | 23% |
| | Population 18-64 | 58% | 67% | 63% |
| | Population +65 | 23% | 16% | 14% |
| | Per capita income | 25073 | 28,210 | 57,492 |
| | Median household income | 44600 | 55825 | 128374 |
| | Persons below poverty line | 14.80% | 21.1 | 6% |
| | High school or higher | 84.6% | 91.90% | 92.70% |
| | Bachelor's degree or higher | 21% | 34.80% | 62.40% |
| | Veteran status | 8.80% | 5.90% | 8.50% |
| Clinical data | | | | |
| | Uninsured* | 10.50% | 6.97% | 8.18% |
| | Medicaid* | 15.60% | 9.08% | 6.03% |
| | Medicare* | 18% | 12.20% | 8.36% |
| | Employer coverage* | 40.9 | 54.60% | 58.40% |
| | Military or VA* | 2.09% | 2.07% | 3.81% |
| | ICU beds** | 16 | 35 | 333 |
| | ICU bed per capita*** | 11,708 people to 1 bed | 5,204 people to 1 bed | 3,446 people to 1 bed |
