## Supplementary material for "Development and Implementation of a scalable and versatile test for COVID-19 diagnostics in rural communities": Sup_Table XIII

**Pool_raw_data**

| A | B | CtRPP | D | E | F | G | H | I | J | K | L | M | N | O |
|---|---|---|---|---|---|---|---|---|---|---|---|---|---|---|
| | Sample 1 | | | | | | | Sample 2 | | | | | | |
| **Pool name** | **Original test date** | **$Ct_{RPP}$** | **$Ct_{N2}$** | **$Ct_{E1}$** | **$Ct_{S6}$** | **cut-off N2** | **cut-off RPP30** | **Original test date** | **$Ct_{RPP}$** | **$Ct_{N2}$** | **$Ct_{E1}$** | **$Ct_{S6}$** | **cut-off N2** | **cut-off RPP30** |
| 1N | | 29.65 | N/A | N/A | N/A | 34.78 | 35.37 | | 30.66 | N/A | N/A | N/A | 34.78 | 35.37 |
| 2N | | 32.04 | N/A | N/A | N/A | 34.78 | 35.37 | | 29.48 | 38.87 | N/A | N/A | 34.78 | 35.37 |
| 3N | | 29.14 | N/A | N/A | N/A | 34.78 | 35.37 | | 30.17 | 44.61 | N/A | N/A | 34.78 | 35.37 |
| 4N | | 29.08 | 43.23 | N/A | N/A | 34.78 | 35.37 | | 28.23 | N/A | N/A | 40.97 | 34.78 | 35.37 |
| 5N | | 28.40 | N/A | 43.83 | 43.58 | 34.78 | 35.37 | | 29.67 | N/A | N/A | 40.24 | 34.78 | 35.37 |
| 6N | | 32.08 | N/A | N/A | N/A | 34.78 | 35.37 | | 30.35 | N/A | N/A | 40.00 | 34.78 | 35.37 |
| 7N | | 29.17 | N/A | N/A | N/A | 34.86 | 34.75 | | 30.81 | N/A | N/A | N/A | 34.86 | 34.75 |
| 8N | | 27.52 | N/A | 43.27 | N/A | 34.86 | 34.75 | | 30.64 | N/A | N/A | N/A | 34.86 | 34.75 |
| 9N | | 30.52 | N/A | N/A | N/A | 34.86 | 34.75 | | 30.88 | N/A | N/A | N/A | 34.86 | 34.75 |
| 10N | | 28.02 | N/A | N/A | N/A | 34.86 | 34.75 | | 28.38 | N/A | N/A | 43.41 | 34.86 | 34.75 |
| 11N | | 27.38 | N/A | N/A | N/A | 34.78 | 35.37 | | 31.39 | N/A | N/A | N/A | 34.86 | 34.75 |
| 12N | | 31.82 | N/A | 37.02 | N/A | 34.86 | 34.75 | | 31.16 | N/A | N/A | N/A | 34.86 | 34.75 |
| 13N | | 30.12 | N/A | N/A | 41.78 | 34.78 | 35.37 | | 28.47 | 39.42 | N/A | N/A | 34.78 | 35.37 |
| 14N | | 30.14 | N/A | N/A | N/A | 34.78 | 35.37 | | 29.53 | 42.71 | N/A | N/A | 34.78 | 35.37 |
| 15N | | 30.59 | N/A | N/A | N/A | 34.86 | 34.75 | | 28.02 | N/A | N/A | 42.06 | 34.57 | 35.53 |
| 16N | | 34.93 | N/A | N/A | N/A | 34.78 | 35.37 | | 29.22 | N/A | 36.63 | N/A | 34.78 | 35.37 |
| 17N | | 28.97 | N/A | N/A | N/A | 34.86 | 34.75 | | 28.22 | N/A | N/A | 41.10 | 34.86 | 34.75 |
| 18N | | 30.30 | 41.06 | N/A | N/A | 34.62 | 35.53 | | 31.55 | N/A | N/A | N/A | 34.86 | 34.75 |
| 19N | | 28.61 | N/A | N/A | 40.87 | 34.83 | 36.97 | | 29.38 | 41.11 | N/A | N/A | 34.83 | 36.97 |
| 20N | | 29.54 | N/A | N/A | 38.64 | 34.83 | 36.97 | | 28.95 | N/A | N/A | 43.42 | 34.83 | 36.97 |
| 1P | | 27.99 | 25.93 | 28.19 | 28.31 | 34.85 | 36.50 | | 30.66 | N/A | N/A | N/A | 34.83 | 36.97 |
| 2P | | 30.23 | 19.81 | 20.86 | 21.52 | 35.05 | 36.98 | | 30.64 | N/A | N/A | N/A | 34.83 | 36.97 |
| 3P | | 30.50 | 20.67 | 20.84 | 20.54 | 35.45 | 36.50 | | 28.38 | N/A | N/A | 43.41 | 34.83 | 36.97 |
| 4P | | 27.63 | 32.52 | 33.79 | 34.24 | 34.75 | 34.73 | | 25.68 | 36.67 | N/A | 36.33 | 34.75 | 34.73 |
| 5P | | 28.43 | 15.32 | 16.49 | 16.58 | 34.82 | 38.17 | | 26.46 | 41.00 | 41.41 | 40.55 | 34.82 | 38.17 |
| 6P | | 29.03 | 26.65 | 28.78 | 29.38 | 34.82 | 38.17 | | 27.17 | 36.47 | 36.01 | 42.53 | 34.82 | 38.17 |
| 7P | | 29.10 | 21.48 | 23.29 | 23.16 | 35.36 | 37.96 | | 29.99 | N/A | N/A | N/A | 35.36 | 37.96 |
| 8P | | 29.60 | 32.91 | 33.68 | 33.48 | 35.36 | 37.96 | | 29.50 | N/A | N/A | 39.25 | 35.36 | 37.96 |
| 9P | | 28.41 | 21.68 | 22.80 | 22.99 | 35.36 | 37.96 | | 28.99 | N/A | N/A | 37.58 | 35.36 | 37.96 |
| 10P | | 28.27 | 20.07 | 21.38 | 21.71 | 35.14 | 38.32 | | 29.83 | N/A | N/A | N/A | 35.14 | 38.32 |
| 11P | | 29.45 | 21.49 | 23.53 | 24.17 | 35.49 | 38.34 | | 27.67 | 43.23 | N/A | N/A | 35.49 | 38.34 |
| 12P | | 28.25 | 17.45 | 19.10 | 19.55 | 35.36 | 37.96 | | 32.04 | N/A | N/A | N/A | 35.36 | 37.96 |
| 13P | | 28.56 | 20.85 | 21.81 | 21.73 | 34.72 | 37.90 | | 28.70 | 37.18 | N/A | N/A | 34.72 | 37.90 |
| 14P | | 29.29 | 17.10 | 19.07 | 19.20 | 34.72 | 37.90 | | 29.42 | N/A | N/A | N/A | 33.76 | 38.43 |
| 15P | | 28.49 | 23.06 | 24.23 | 24.17 | 34.72 | 37.90 | | 27.28 | 38.62 | N/A | 37.70 | 34.89 | 37.21 |
| 16P | | 28.01 | 21.08 | 22.30 | 22.20 | 34.72 | 37.90 | | 27.91 | 43.50 | N/A | 44.66 | 34.72 | 37.90 |
| 17P | | 27.86 | 22.61 | 24.66 | 24.47 | 34.72 | 37.90 | | 27.56 | N/A | 42.57 | 41.58 | 34.72 | 37.90 |
| 18P | | 27.79 | 31.22 | 31.20 | 30.91 | 34.72 | 37.90 | | 29.22 | N/A | 36.63 | N/A | 34.72 | 37.90 |
| 19P | | 27.81 | 33.73 | 32.82 | 33.39 | 33.76 | 38.43 | | 28.22 | N/A | N/A | 41.10 | 33.76 | 38.43 |
| 20P | | 28.46 | 21.16 | 22.26 | 22.16 | 34.89 | 37.21 | | 31.55 | N/A | N/A | N/A | 34.89 | 37.21 |
| 21P | | 27.06 | 33.26 | 33.12 | 32.61 | 37.16 | 38.01 | | 27.56 | N/A | N/A | 41.85 | 37.16 | 38.01 |

| A | P | Q | R | S | T | U | V | W | X | Y | Z | AA | AB | AC | AD | AE | AF | AG | AH | AI | AJ |
|---|---|---|---|---|---|---|---|---|---|---|---|---|---|---|---|---|---|---|---|---|---|
| | Sample 3 | | | | | | | Sample 4 | | | | | | | Pool | | | | | | |
| **Pool name** | **Original test date** | **$Ct_{RPP}$** | **$Ct_{N2}$** | **$Ct_{E1}$** | **$Ct_{S6}$** | **cut-off N2** | **cut-off RPP30** | **Original test date** | **$Ct_{RPP}$** | **$Ct_{N2}$** | **$Ct_{E1}$** | **$Ct_{S6}$** | **cut-off N2** | **cut-off RPP30** | **Original test date** | **$Ct_{RPP}$** | **$Ct_{N2}$** | **$Ct_{E1}$** | **$Ct_{S6}$** | **cut-off N2** | **cut-off RPP30** |
| 1N | | 26.43 | N/A | N/A | 44.16 | 34.78 | 35.37 | | 23.62 | N/A | N/A | 41.22 | 34.78 | 35.37 | | 27.49 | 27.49 | N/A | N/A | 35.45 | 36.50 |
| 2N | | 27.93 | 39.49 | N/A | 42.52 | 34.78 | 35.37 | | 28.55 | N/A | N/A | N/A | 34.78 | 35.37 | | 28.20 | 43.22 | N/A | 38.33 | 35.45 | 36.50 |
| 3N | | 29.06 | N/A | N/A | N/A | 34.78 | 35.37 | | 29.13 | N/A | N/A | 37.27 | 34.78 | 35.37 | | 28.99 | N/A | N/A | N/A | 35.45 | 36.50 |
| 4N | | 31.01 | N/A | N/A | N/A | 34.78 | 35.37 | | 29.73 | N/A | N/A | 41.21 | 34.78 | 35.37 | | 29.10 | N/A | N/A | 44.81 | 35.45 | 36.50 |
| 5N | | 29.14 | N/A | N/A | N/A | 34.78 | 35.37 | | 29.15 | N/A | N/A | 38.58 | 34.78 | 35.37 | | 28.54 | N/A | N/A | 37.38 | 35.45 | 36.50 |
| 6N | | 29.44 | 38.40 | 40.34 | N/A | 34.78 | 35.37 | | 29.45 | N/A | N/A | 41.71 | 34.86 | 34.75 | | 29.97 | N/A | N/A | N/A | 35.45 | 36.50 |
| 7N | | 29.33 | N/A | N/A | N/A | 34.86 | 34.75 | | 30.38 | N/A | 37.10 | 39.72 | 34.86 | 34.75 | | 29.28 | 38.82 | N/A | N/A | 35.45 | 36.50 |
| 8N | | 27.81 | N/A | N/A | 40.70 | 34.86 | 34.75 | | 28.70 | N/A | N/A | 40.85 | 34.86 | 34.75 | | 29.26 | N/A | N/A | N/A | 35.45 | 36.50 |
| 9N | | 30.29 | 37.16 | N/A | N/A | 34.86 | 34.75 | | 31.64 | N/A | N/A | N/A | 34.86 | 34.75 | | 29.84 | N/A | N/A | 40.30 | 35.45 | 36.50 |
| 10N | | 29.17 | N/A | N/A | N/A | 34.86 | 34.75 | | 31.14 | N/A | N/A | 40.94 | 34.86 | 34.75 | | 28.10 | N/A | N/A | 37.62 | 35.45 | 36.50 |
| 11N | | 29.46 | N/A | N/A | N/A | 34.86 | 34.75 | | 29.62 | 36.24 | 37.70 | 41.67 | 34.86 | 34.75 | | 28.37 | N/A | N/A | 43.20 | 35.45 | 36.50 |
| 12N | | 30.27 | N/A | N/A | 39.80 | 34.86 | 34.75 | | 30.44 | N/A | N/A | N/A | 34.86 | 34.75 | | 30.37 | 42.20 | N/A | 37.71 | 35.45 | 36.50 |
| 13N | | 27.05 | N/A | N/A | 38.24 | 34.78 | 35.37 | | 28.80 | N/A | N/A | 44.72 | 34.78 | 35.37 | | 28.10 | N/A | N/A | N/A | 35.45 | 36.50 |
| 14N | | 28.44 | N/A | N/A | 44.75 | 34.78 | 35.37 | | 27.16 | N/A | N/A | 42.08 | 34.78 | 35.37 | | 28.14 | N/A | N/A | 41.68 | 35.45 | 36.50 |
| 15N | | 27.35 | N/A | N/A | 40.98 | 34.57 | 35.53 | | 28.56 | 44.14 | N/A | N/A | 34.57 | 35.53 | | 27.48 | N/A | N/A | 38.61 | 35.45 | 36.50 |
| 16N | | 27.68 | N/A | N/A | 43.37 | 34.78 | 35.37 | | 28.81 | N/A | N/A | N/A | 34.78 | 35.37 | | 28.43 | N/A | 41.70 | N/A | 34.84 | 37.00 |
| 17N | | 28.56 | N/A | N/A | 39.98 | 34.86 | 34.75 | | 28.42 | N/A | N/A | N/A | 34.86 | 34.75 | | 28.13 | N/A | N/A | N/A | 34.84 | 37.00 |
| 18N | | 29.71 | 36.96 | N/A | N/A | 34.86 | 34.75 | | 29.12 | N/A | N/A | N/A | 34.86 | 34.75 | | 29.32 | N/A | N/A | N/A | 34.84 | 37.00 |
| 19N | | 28.51 | N/A | N/A | N/A | 34.83 | 36.97 | | 29.18 | N/A | N/A | 44.22 | 34.83 | 36.97 | | 28.64 | N/A | N/A | 42.16 | 34.84 | 37.00 |
| 20N | | 31.05 | N/A | 39.29 | N/A | 34.83 | 36.97 | | 29.57 | N/A | N/A | N/A | 34.83 | 36.97 | | 30.06 | N/A | N/A | N/A | 34.84 | 37.00 |
| 1P | | 26.43 | N/A | N/A | 44.16 | 34.83 | 36.97 | | 23.62 | N/A | N/A | 41.22 | 34.83 | 36.97 | | 27.66 | 28.73 | 30.42 | 30.70 | 35.45 | 36.50 |
| 2P | | 27.81 | N/A | N/A | 40.70 | 34.83 | 36.97 | | 28.70 | N/A | N/A | 40.85 | 34.83 | 36.97 | | 28.87 | 23.25 | 24.25 | 25.45 | 35.45 | 36.50 |
| 3P | | 29.17 | N/A | N/A | N/A | 34.83 | 36.97 | | 31.14 | N/A | N/A | 40.94 | 34.83 | 36.97 | | 29.01 | 20.31 | 20.61 | 20.46 | 35.45 | 36.50 |
| 4P | | 27.20 | 36.60 | 44.43 | 44.24 | 34.75 | 34.73 | | 27.27 | 44.23 | N/A | 41.82 | 34.75 | 34.73 | | 27.08 | 33.09 | 34.22 | 34.70 | 35.82 | 34.13 |
| 5P | | 29.24 | N/A | N/A | N/A | 34.82 | 38.17 | | 27.30 | 42.96 | 44.99 | 41.84 | 34.82 | 38.17 | | 28.82 | 18.01 | 18.74 | 18.38 | 35.82 | 34.13 |
| 6P | | 27.35 | N/A | N/A | 38.54 | 34.82 | 38.17 | | 29.59 | N/A | N/A | N/A | 34.82 | 38.17 | | 28.07 | 29.18 | 30.90 | 31.17 | 35.82 | 34.13 |
| 7P | | 28.23 | 42.18 | N/A | N/A | 35.36 | 37.96 | | 27.83 | N/A | 42.17 | 37.90 | 35.36 | 37.96 | | 29.04 | 23.68 | 25.06 | 24.96 | 35.82 | 34.13 |
| 8P | | 28.65 | 35.15 | 37.22 | 44.27 | 35.36 | 37.96 | | 29.08 | N/A | N/A | 42.11 | 35.36 | 37.96 | | 28.73 | 33.72 | 34.38 | 34.91 | 35.82 | 34.13 |
| 9P | | 28.80 | N/A | N/A | 37.14 | 35.36 | 37.96 | | 30.19 | N/A | N/A | 43.22 | 35.36 | 37.96 | | 28.09 | 23.05 | 24.41 | 24.19 | 34.22 | 34.24 |
| 10P | | 28.76 | N/A | N/A | N/A | 35.14 | 38.32 | | 28.11 | 36.28 | 36.04 | 37.87 | 35.14 | 38.32 | | 30.80 | 25.15 | 26.27 | 26.49 | 35.14 | 37.18 |
| 11P | | 28.29 | N/A | N/A | N/A | 35.49 | 38.34 | | 29.39 | 37.08 | N/A | N/A | 35.49 | 38.34 | | 30.11 | 24.67 | 26.74 | 27.12 | 35.14 | 37.18 |
| 12P | | 27.98 | N/A | N/A | 40.87 | 35.36 | 37.96 | | 28.57 | N/A | N/A | 42.20 | 35.36 | 37.96 | | 31.54 | 21.92 | 23.31 | 23.86 | 35.14 | 37.18 |
| 13P | | 27.68 | 37.09 | N/A | 38.44 | 34.72 | 37.90 | | 28.85 | N/A | N/A | N/A | 34.72 | 37.90 | | 29.62 | 24.14 | 24.72 | 24.63 | 35.14 | 37.18 |
| 14P | | 28.22 | N/A | N/A | 41.53 | 33.76 | 38.43 | | 26.30 | N/A | N/A | N/A | 33.76 | 38.43 | | 28.85 | 19.40 | 20.95 | 21.14 | 35.14 | 37.18 |
| 15P | | 28.20 | 44.63 | 37.09 | 36.69 | 34.89 | 37.21 | | 28.32 | 41.72 | N/A | 42.70 | 34.89 | 37.21 | | 30.08 | 26.01 | 27.06 | 27.09 | 35.14 | 37.18 |
| 16P | | 27.89 | N/A | N/A | 42.15 | 34.72 | 37.90 | | 28.25 | 35.86 | 36.22 | 36.62 | 34.72 | 37.90 | | 29.42 | 23.36 | 24.73 | 24.56 | 35.14 | 37.18 |
| 17P | | 28.17 | N/A | N/A | 39.26 | 34.72 | 37.90 | | 27.19 | N/A | N/A | 39.64 | 34.72 | 37.90 | | 29.61 | 28.27 | 30.14 | 30.08 | 35.14 | 37.18 |
| 18P | | 27.68 | N/A | N/A | 43.37 | 34.72 | 37.90 | | 28.81 | N/A | N/A | N/A | 34.72 | 37.90 | | 28.34 | 32.06 | 32.87 | 32.60 | 34.84 | 37.00 |
| 19P | | 28.56 | N/A | N/A | 39.98 | 33.76 | 38.43 | | 28.42 | N/A | N/A | N/A | 33.76 | 38.43 | | 28.20 | 35.50 | 35.10 | 36.11 | 34.84 | 37.00 |
| 20P | | 29.71 | 36.96 | N/A | N/A | 34.89 | 37.21 | | 29.12 | N/A | N/A | N/A | 34.89 | 37.21 | | 28.60 | 22.62 | 24.04 | 24.09 | 34.84 | 37.00 |
| 21P | | 29.65 | 39.62 | 44.09 | N/A | 37.16 | 38.01 | | 29.24 | N/A | 40.00 | 44.69 | 37.16 | 38.01 | | 28.06 | 35.91 | 35.93 | 34.74 | 34.84 | 37.00 |

**Positives**

| A | B | C | D | E | F | G | H | I | J | K | L | M |
|---|---|---|---|---|---|---|---|---|---|---|---|---|
| | $Ct_{RPP}$ | | | $Ct_{N2}$ | | | $Ct_{E1}$ | | | $Ct_{S6}$ | | |
| **Pool name** | **Individual** | **Pool** | **Ct diffrence** | **Individual** | **Pool** | **Ct diffrence** | **Individual** | **Pool** | **Ct diffrence** | **Individual** | **Pool** | **Ct diffrence** |
| 1P | 27.99 | 27.66 | -0.33 | 25.93 | 28.73 | 2.80 | 28.19 | 30.42 | 2.23 | 28.31 | 30.70 | 2.39 |
| 2P | 30.23 | 28.87 | -1.36 | 19.81 | 23.25 | 3.43 | 20.86 | 24.25 | 3.39 | 21.52 | 25.45 | 3.93 |
| 3P | 30.50 | 29.01 | -1.50 | 20.67 | 20.31 | -0.36 | 20.84 | 20.61 | -0.22 | 20.54 | 20.46 | -0.08 |
| 4P | 27.63 | 27.08 | -0.55 | 32.52 | 33.09 | 0.57 | 33.79 | 34.22 | 0.43 | 34.24 | 34.70 | 0.45 |
| 5P | 28.43 | 28.82 | 0.39 | 15.32 | 18.01 | 2.69 | 16.49 | 18.74 | 2.25 | 16.58 | 18.38 | 1.79 |
| 6P | 29.03 | 28.07 | -0.95 | 26.65 | 29.18 | 2.52 | 28.78 | 30.90 | 2.12 | 29.38 | 31.17 | 1.79 |
| 7P | 29.10 | 29.04 | -0.05 | 21.48 | 23.68 | 2.20 | 23.29 | 25.06 | 1.77 | 23.16 | 24.96 | 1.80 |
| 8P | 29.60 | 28.73 | -0.88 | 32.91 | 33.72 | 0.81 | 33.68 | 34.38 | 0.70 | 33.48 | 34.91 | 1.43 |
| 9P | 28.41 | 28.09 | -0.31 | 21.68 | 23.05 | 1.37 | 22.80 | 24.41 | 1.61 | 22.99 | 24.19 | 1.20 |
| 10P | 28.27 | 30.80 | 2.53 | 20.07 | 25.15 | 5.08 | 21.38 | 26.27 | 4.89 | 21.71 | 26.49 | 4.78 |
| 11P | 29.45 | 30.11 | 0.66 | 21.49 | 24.67 | 3.17 | 23.53 | 26.74 | 3.21 | 24.17 | 27.12 | 2.95 |
| 12P | 28.25 | 31.54 | 3.28 | 17.45 | 21.92 | 4.48 | 19.10 | 23.31 | 4.21 | 19.55 | 23.86 | 4.31 |
| 13P | 28.56 | 29.62 | 1.06 | 20.85 | 24.14 | 3.29 | 21.81 | 24.72 | 2.90 | 21.73 | 24.63 | 2.90 |
| 14P | 29.29 | 28.85 | -0.44 | 17.10 | 19.40 | 2.31 | 19.07 | 20.95 | 1.88 | 19.20 | 21.14 | 1.95 |
| 15P | 28.49 | 30.08 | 1.59 | 23.06 | 26.01 | 2.95 | 24.23 | 27.06 | 2.83 | 24.17 | 27.09 | 2.92 |
| 16P | 28.01 | 29.42 | 1.41 | 21.08 | 23.36 | 2.28 | 22.30 | 24.73 | 2.43 | 22.20 | 24.56 | 2.35 |
| 17P | 27.86 | 29.61 | 1.75 | 22.61 | 28.27 | 5.66 | 24.66 | 30.14 | 5.48 | 24.47 | 30.08 | 5.61 |
| 18P | 27.79 | 28.34 | 0.54 | 31.22 | 32.06 | 0.84 | 31.20 | 32.87 | 1.67 | 30.91 | 32.60 | 1.70 |
| 19P | 27.81 | 28.20 | 0.39 | 33.73 | 35.50 | 1.78 | 32.82 | 35.10 | 2.28 | 33.39 | 36.11 | 2.72 |
| 20P | 28.46 | 28.60 | 0.13 | 21.16 | 22.62 | 1.46 | 22.26 | 24.04 | 1.78 | 22.16 | 24.09 | 1.93 |
| 21P | 27.06 | 28.06 | 1.00 | 33.26 | 35.91 | 2.65 | 33.12 | 35.93 | 2.81 | 32.61 | 34.74 | 2.13 |

**Regression**

| | $Ct_N$ | | $Ct_E$ | | $Ct_S$ | |
|---|---|---|---|---|---|---|
| **Pool name** | **Individual** | **Pool** | **Individual** | **Pool** | **Individual** | **Pool** |
| 1P | 25.93 | 28.73 | 28.19 | 30.42 | 28.31 | 30.70 |
| 2P | 19.81 | 23.25 | 20.86 | 24.25 | 21.52 | 25.45 |
| 3P | 20.67 | 20.31 | 20.84 | 20.61 | 20.54 | 20.46 |
| 4P | 32.52 | 33.09 | 33.79 | 34.22 | 34.24 | 34.70 |
| 5P | 15.32 | 18.01 | 16.49 | 18.74 | 16.58 | 18.38 |
| 6P | 26.65 | 29.18 | 28.78 | 30.90 | 29.38 | 31.17 |
| 7P | 21.48 | 23.68 | 23.29 | 25.06 | 23.16 | 24.96 |
| 8P | 32.91 | 33.72 | 33.68 | 34.38 | 33.48 | 34.91 |
| 9P | 21.68 | 23.05 | 22.80 | 24.41 | 22.99 | 24.19 |
| 10P | 20.07 | 25.15 | 21.38 | 26.27 | 21.71 | 26.49 |
| 11P | 21.49 | 24.67 | 23.53 | 26.74 | 24.17 | 27.12 |
| 12P | 17.45 | 21.92 | 19.10 | 23.31 | 19.55 | 23.86 |
| 13P | 20.85 | 24.14 | 21.81 | 24.72 | 21.73 | 24.63 |
| 14P | 17.10 | 19.40 | 19.07 | 20.95 | 19.20 | 21.14 |
| 15P | 23.06 | 26.01 | 24.23 | 27.06 | 24.17 | 27.09 |
| 16P | 21.08 | 23.36 | 22.30 | 24.73 | 22.20 | 24.56 |
| 17P | 22.61 | 28.27 | 24.66 | 30.14 | 24.47 | 30.08 |
| 18P | 31.22 | 32.06 | 31.20 | 32.87 | 30.91 | 32.60 |
| 19P | 33.73 | 35.50 | 32.82 | 35.10 | 33.39 | 36.11 |
| 20P | 21.16 | 22.62 | 22.26 | 24.04 | 22.16 | 24.09 |
| 21P | 33.26 | 35.91 | 33.12 | 35.93 | 32.61 | 34.74 |

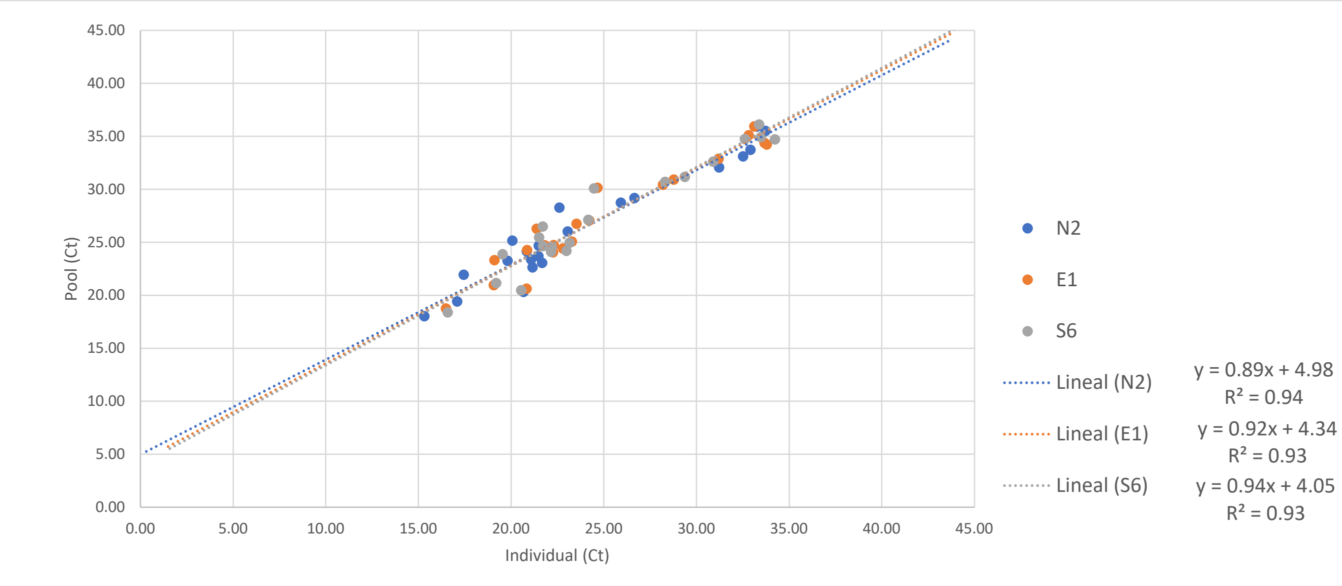


| | Pool | | | Individual | | |
|---|---|---|---|---|---|---|
| **Gene** | **LOD** | **95% CI lower** | **95% CI upper** | **LOD** | **95% CI lower** | **95% CI upper** |
| **N** | 37.29 | 36.91 | 37.67 | 36.30 | 35.88 | 36.73 |
| **E** | 37.29 | 36.91 | 37.67 | 35.82 | 35.40 | 36.23 |
| **S** | 37.29 | 36.91 | 37.67 | 35.36 | 34.96 | 35.77 |

For a pool which is at the limit of detection, the individual sample would have had a Ct around 36.30 for N2. This means that during the pooling procedure samples with Cts between 36.30 and 37.29 would have been detected using individual testing but will not be due to pooling.
