## Supplementary material for "Development and Implementation of a scalable and versatile test for COVID-19 diagnostics in rural communities": Sup_Table XV

Cts for FDA (n=4)

| A | B | C | D | E | F | G | H | I | J | K | L | M | N | O | P | Q | R |
|---|---|---|---|---|---|---|---|---|---|---|---|---|---|---|---|---|---|
| Positive | Specimen code | Date qPCR run | Plate number | $Ct_{RPP}$ | $Ct_{N2}$ | $Ct_{E1}$ | $Ct_{S6}$ | cut-off N | cut-off RPP30 | Theoretical Ct cutoff | Estimated CT cutoff | Theoretical Result N | Theoretical Result E | Theoretical Result S | Estimated Result N | Estimated Result E | Estimated Result S |
| 1 | V0010590 | | | 28.22 | 15.73 | 17.65 | 18.22 | 35.95 | 35.46 | 33.95 | 34.81 | Positive | Positive | Positive | Positive | Positive | Positive |
| 2 | V0012870 | | | 29.06 | 17.38 | 18.63 | 19.21 | 35.95 | 35.46 | 33.95 | 34.81 | Positive | Positive | Positive | Positive | Positive | Positive |
| 3 | V0014312 | | | 27.97 | 14.82 | 16.26 | 16.66 | 35.95 | 35.46 | 33.95 | 34.81 | Positive | Positive | Positive | Positive | Positive | Positive |
| 4 | V0015270 | | | 28.23 | 23.00 | 23.36 | 23.08 | 35.95 | 35.46 | 33.95 | 34.81 | Positive | Positive | Positive | Positive | Positive | Positive |
| 5 | V0015941 | | | 29.27 | 16.67 | 18.43 | 18.47 | 35.95 | 35.46 | 33.95 | 34.81 | Positive | Positive | Positive | Positive | Positive | Positive |
| 6 | V0016364 | | | 27.74 | 36.70 | 33.60 | 35.19 | 35.45 | 35.37 | 33.45 | 34.24 | Negative | Negative | Negative | Negative | Positive | Negative |
| 7 | 258011 | | | 35.23 | 23.38 | 23.41 | 22.96 | 34.75 | 35.43 | 32.75 | 33.47 | Positive | Positive | Positive | Positive | Positive | Positive |
| 8 | 266249 | | | 29.77 | 24.76 | 25.29 | 24.76 | 34.75 | 35.43 | 32.75 | 33.47 | Positive | Positive | Positive | Positive | Positive | Positive |
| 9 | V0016301 | | | 29.11 | 34.33 | 33.48 | 35.10 | 34.75 | 35.43 | 32.75 | 33.47 | Negative | Negative | Negative | Negative | Negative | Negative |
| 10 | V0010606 | | | 28.22 | 30.24 | 30.20 | 30.03 | 35.48 | 35.63 | 33.48 | 34.28 | Positive | Positive | Positive | Positive | Positive | Positive |
| 11 | V0015699 | | | 29.76 | 18.06 | 19.57 | 20.18 | 35.48 | 35.63 | 33.48 | 34.28 | Positive | Positive | Positive | Positive | Positive | Positive |
| 12 | V0014597 | | | 30.07 | 18.06 | 19.20 | 20.00 | 35.26 | 36.36 | 33.26 | 34.03 | Positive | Positive | Positive | Positive | Positive | Positive |
| 13 | V0015158 | | | 28.52 | 32.77 | 32.66 | 34.00 | 35.31 | 36.34 | 33.31 | 34.09 | Positive | Positive | Negative | Positive | Positive | Positive |
| 14 | V0015163 | | | 29.45 | 31.35 | 30.63 | 31.00 | 35.31 | 36.34 | 33.31 | 34.09 | Positive | Positive | Positive | Positive | Positive | Positive |
| 15 | V0010589 | | | 29.76 | 33.66 | 33.57 | 33.70 | 36.21 | 36.58 | 34.21 | 35.10 | Positive | Positive | Positive | Positive | Positive | Positive |
| 16 | V0016168 | | | 29.38 | 34.07 | 33.15 | 33.75 | 35.15 | 35.61 | 33.15 | 33.91 | Negative | Positive | Negative | Negative | Positive | Positive |
| 17 | V0014934 | | | 28.25 | 27.92 | 28.42 | 28.62 | 35.12 | 35.85 | 33.12 | 33.88 | Positive | Positive | Positive | Positive | Positive | Positive |
| 18 | V0015461 | | | 29.25 | 33.51 | 32.90 | 33.58 | 35.12 | 35.85 | 33.12 | 33.88 | Negative | Positive | Negative | Positive | Positive | Positive |
| 19 | V0014582 | | | 28.98 | 30.32 | 30.12 | 30.35 | 35.21 | 36.42 | 33.21 | 33.98 | Positive | Positive | Positive | Positive | Positive | Positive |
| 20 | V0014587 | | | 27.31 | 34.65 | 34.36 | 33.93 | 35.21 | 36.42 | 33.21 | 33.98 | Negative | Negative | Negative | Negative | Negative | Positive |
| 21 | V0015145 | | | 28.12 | 31.64 | 31.54 | 32.14 | 35.21 | 36.42 | 33.21 | 33.98 | Positive | Positive | Positive | Positive | Positive | Positive |
| 22 | V0014584 | | | 28.19 | 21.33 | 23.24 | 24.14 | 36.07 | 35.23 | 34.07 | 34.94 | Positive | Positive | Positive | Positive | Positive | Positive |
| 23 | V0014589 | | | 28.86 | 20.21 | 21.28 | 21.60 | 36.07 | 35.23 | 34.07 | 34.94 | Positive | Positive | Positive | Positive | Positive | Positive |
| 24 | V0014595 | | | 29.05 | 18.88 | 20.66 | 21.64 | 36.07 | 35.23 | 34.07 | 34.94 | Positive | Positive | Positive | Positive | Positive | Positive |
| 25 | V0014616 | | | 28.45 | 31.51 | 31.73 | 32.37 | 35.80 | 35.45 | 33.80 | 34.65 | Positive | Positive | Positive | Positive | Positive | Positive |
| 26 | V0014903 | | | 28.57 | 32.77 | 33.42 | 34.04 | 35.80 | 35.45 | 33.80 | 34.65 | Positive | Positive | Negative | Positive | Positive | Positive |
| 27 | V0014995 | | | 28.96 | 35.25 | 33.92 | 34.60 | 35.80 | 35.45 | 33.80 | 34.65 | Negative | Negative | Negative | Negative | Positive | Positive |
| 28 | 249008 | | | 29.30 | 21.14 | 22.74 | 23.00 | 34.38 | 36.43 | 32.38 | 33.04 | Positive | Positive | Positive | Positive | Positive | Positive |
| 29 | V0014349 | | | 27.23 | 33.86 | 33.41 | 35.71 | 35.26 | 35.46 | 33.26 | 34.04 | Negative | Negative | Negative | Positive | Positive | Negative |
| 30 | 239215 | | | 29.84 | 18.68 | 19.70 | 20.25 | 36.19 | 36.27 | 34.19 | 35.08 | Positive | Positive | Positive | Positive | Positive | Positive |
| 31 | V0014367 | | | 29.57 | 19.07 | 20.62 | 21.66 | 36.19 | 36.27 | 34.19 | 35.08 | Positive | Positive | Positive | Positive | Positive | Positive |
| 32 | V0013598 | | | 30.79 | 20.50 | 22.21 | 22.95 | 36.16 | 36.06 | 34.16 | 35.04 | Positive | Positive | Positive | Positive | Positive | Positive |
| 33 | V0013672 | | | 29.30 | 27.35 | 28.91 | 29.54 | 36.16 | 36.06 | 34.16 | 35.04 | Positive | Positive | Positive | Positive | Positive | Positive |
| 34 | V0244808 | | | 32.05 | 31.01 | 32.18 | 33.34 | 35.95 | 35.40 | 33.95 | 34.81 | Positive | Positive | Positive | Positive | Positive | Positive |
| 35 | V0249008 | | | 30.80 | 22.04 | 23.67 | 24.34 | 35.95 | 35.40 | 33.95 | 34.81 | Positive | Positive | Positive | Positive | Positive | Positive |
| 36 | V0267038 | | | 30.37 | 16.99 | 19.28 | 19.82 | 35.95 | 35.40 | 33.95 | 34.81 | Positive | Positive | Positive | Positive | Positive | Positive |
| 37 | V0014502 | | | 29.25 | 19.77 | 19.55 | 20.06 | 35.68 | 35.84 | 33.68 | 34.51 | Positive | Positive | Positive | Positive | Positive | Positive |
| 38 | V0014521 | | | 30.47 | 25.08 | 24.55 | 24.81 | 35.68 | 35.84 | 33.68 | 34.51 | Positive | Positive | Positive | Positive | Positive | Positive |
| 39 | V0013602 | | | 29.41 | 17.20 | 18.65 | 19.01 | 35.57 | 36.89 | 33.57 | 34.38 | Positive | Positive | Positive | Positive | Positive | Positive |
| 40 | V0013659 | | | 29.99 | 13.49 | 16.05 | 16.61 | 35.80 | 36.81 | 33.80 | 34.64 | Positive | Positive | Positive | Positive | Positive | Positive |
| 41 | V0013660 | | | 28.91 | 19.76 | 20.31 | 20.84 | 35.80 | 36.81 | 33.80 | 34.64 | Positive | Positive | Positive | Positive | Positive | Positive |
| 42 | V0013691 | | | 28.96 | 24.06 | 24.09 | 24.1 | 35.14 | 36.38 | 33.14 | 33.90 | Positive | Positive | Positive | Positive | Positive | Positive |
| 43 | V0013695 | | | 29.90 | 26.89 | 26.79 | 26.9 | 35.14 | 36.38 | 33.14 | 33.90 | Positive | Positive | Positive | Positive | Positive | Positive |
| 44 | V0014147 | | | 27.70 | 24.30 | 24.99 | 25.18 | 36.02 | 36.37 | 34.02 | 34.89 | Positive | Positive | Positive | Positive | Positive | Positive |
| 45 | V0011683 | | | 29.73 | 17.58 | 19.49 | 19.37 | 36.63 | 36.94 | 34.63 | 35.57 | Positive | Positive | Positive | Positive | Positive | Positive |
| 46 | V0013950 | | | 27.19 | 19.63 | 20.55 | 20.56 | 36.63 | 36.94 | 34.63 | 35.57 | Positive | Positive | Positive | Positive | Positive | Positive |
| 47 | V0014047 | | | 27.99 | 24.22 | 25.13 | 25.22 | 36.63 | 36.94 | 34.63 | 35.57 | Positive | Positive | Positive | Positive | Positive | Positive |
| 48 | V0013209 | | | 28.73 | 35.81 | 35.48 | 35.51 | 36.18 | 35.46 | 34.18 | 35.07 | Negative | Negative | Negative | Negative | Negative | Negative |
| 49 | V0014172 | | | 28.39 | 26.94 | 27.60 | 27.08 | 36.18 | 35.46 | 34.18 | 35.07 | Positive | Positive | Positive | Positive | Positive | Positive |
| 50 | V0014208 | | | 28.28 | 22.75 | 23.47 | 23.50 | 36.18 | 35.46 | 34.18 | 35.07 | Positive | Positive | Positive | Positive | Positive | Positive |
| 51 | V0011423 | | | 27.97 | 26.64 | 28.07 | 28.40 | 36.53 | 35.77 | 34.53 | 35.46 | Positive | Positive | Positive | Positive | Positive | Positive |
| 52 | V0014057 | | | 28.89 | 21.48 | 22.35 | 22.57 | 36.53 | 35.77 | 34.53 | 35.46 | Positive | Positive | Positive | Positive | Positive | Positive |

| | | | | | | | | | | | | | | | |
|---|---|---|---|---|---|---|---|---|---|---|---|---|---|---|---|
| 53 | V0011438 | 28.28 | 19.32 | 21.47 | 22.33 | 36.94 | 36.95 | 34.94 | 35.92 | Positive | Positive | Positive | Positive | Positive | Positive |
| 54 | V0012919 | 27.95 | 22.28 | 22.81 | 22.95 | 36.94 | 36.95 | 34.94 | 35.92 | Positive | Positive | Positive | Positive | Positive | Positive |
| 55 | V0014055 | 29.30 | 27.99 | 28.54 | 28.45 | 36.94 | 36.95 | 34.94 | 35.92 | Positive | Positive | Positive | Positive | Positive | Positive |
| 56 | V0014061 | 28.23 | 26.56 | 27.50 | 27.86 | 36.94 | 36.95 | 34.94 | 35.92 | Positive | Positive | Positive | Positive | Positive | Positive |
| 57 | V0012907 | 29.70 | 15.19 | 16.42 | 17.14 | 34.83 | 36.30 | 32.83 | 33.55 | Positive | Positive | Positive | Positive | Positive | Positive |
| 58 | V0003662 | 29.15 | 33.54 | 34.72 | 35.70 | 34.83 | 36.30 | 32.83 | 33.55 | Negative | Negative | Negative | Positive | Negative | Negative |
| 59 | V0012806 | 29.20 | 27.11 | 26.94 | 26.93 | 37.03 | 35.40 | 35.03 | 36.02 | Positive | Positive | Positive | Positive | Positive | Positive |
| 60 | V0012116 | 29.97 | 35.21 | 36.56 | 36.21 | 37.03 | 35.40 | 35.03 | 36.02 | Negative | Negative | Negative | Positive | Negative | Negative |
| 61 | V0012153 | 29.33 | 34.55 | 37.44 | 35.54 | 37.03 | 35.40 | 35.03 | 36.02 | Positive | Negative | Negative | Positive | Negative | Positive |
| 62 | V0012513 | 32.64 | 35.03 | 35.09 | 36.12 | 36.60 | 36.96 | 34.60 | 35.54 | Negative | Negative | Negative | Positive | Positive | Negative |
| 63 | V0014030 | 29.40 | 36.55 | 36.39 | 36.83 | 36.60 | 36.96 | 34.60 | 35.54 | Negative | Negative | Negative | Negative | Negative | Negative |
| 64 | V0011709 | 28.99 | 34.29 | 34.75 | 36.70 | 36.60 | 36.96 | 34.60 | 35.54 | Positive | Negative | Negative | Positive | Positive | Negative |
| 65 | V0012038 | 27.46 | 27.07 | 27.58 | 27.68 | 36.46 | 35.56 | 34.46 | 35.39 | Positive | Positive | Positive | Positive | Positive | Positive |
| 66 | V0012076 | 30.36 | 20.23 | 21.60 | 21.85 | 36.46 | 35.56 | 34.46 | 35.39 | Positive | Positive | Positive | Positive | Positive | Positive |
| 67 | V0013487 | 30.13 | 34.67 | 34.95 | 35.82 | 36.46 | 35.56 | 34.46 | 35.39 | Negative | Negative | Negative | Positive | Positive | Negative |
| 68 | V0012539 | 32.13 | 36.04 | 36.52 | 36.30 | 36.37 | 36.90 | 34.37 | 35.28 | Negative | Negative | Negative | Negative | Negative | Negative |
| 69 | V0013218 | 29.54 | 20.20 | 20.24 | 20.29 | 36.37 | 36.90 | 34.37 | 35.28 | Positive | Positive | Positive | Positive | Positive | Positive |
| 70 | V0013385 | 28.72 | 27.71 | 27.52 | 27.85 | 36.37 | 36.90 | 34.37 | 35.28 | Positive | Positive | Positive | Positive | Positive | Positive |
| 71 | V0013210 | 28.33 | 29.53 | 29.88 | 29.79 | 36.09 | 35.41 | 34.09 | 34.97 | Positive | Positive | Positive | Positive | Positive | Positive |
| 72 | V0012016 | 28.18 | 25.03 | 23.84 | 23.99 | 36.04 | 36.13 | 34.04 | 34.91 | Positive | Positive | Positive | Positive | Positive | Positive |
| 73 | V0012096 | 27.94 | 29.64 | 30.30 | 30.29 | 36.04 | 36.13 | 34.04 | 34.91 | Positive | Positive | Positive | Positive | Positive | Positive |
| 74 | V0012100 | 28.21 | 25.32 | 25.51 | 25.89 | 36.04 | 36.13 | 34.04 | 34.91 | Positive | Positive | Positive | Positive | Positive | Positive |
| 75 | V0012315 | 30.14 | 36.14 | 34.94 | 35.90 | 35.98 | 35.56 | 33.98 | 34.84 | Negative | Negative | Negative | Negative | Negative | Negative |
| 76 | V0010757 | 28.60 | 16.49 | 17.23 | 17.09 | 36.11 | 36.43 | 34.11 | 34.99 | Positive | Positive | Positive | Positive | Positive | Positive |
| 77 | V0010968 | 28.45 | 34.05 | 35.83 | 35.88 | 36.24 | 36.28 | 34.24 | 35.14 | Positive | Negative | Negative | Positive | Negative | Negative |
| 78 | V0012065 | 29.89 | 35.11 | 34.55 | 36.57 | 36.24 | 36.28 | 34.24 | 35.14 | Negative | Negative | Negative | Positive | Positive | Negative |
| 79 | V0012208 | 28.20 | 32.33 | 32.87 | 33.42 | 36.24 | 36.28 | 34.24 | 35.14 | Positive | Positive | Positive | Positive | Positive | Positive |
| 80 | V0012303 | 30.40 | 18.75 | 20.23 | 20.66 | 34.72 | 36.43 | 32.72 | 33.43 | Positive | Positive | Positive | Positive | Positive | Positive |
| 81 | V0012307 | 31.77 | 21.62 | 23.33 | 24.10 | 34.72 | 36.43 | 32.72 | 33.43 | Positive | Positive | Positive | Positive | Positive | Positive |
| 82 | V0011361 | 30.28 | 34.42 | 36.63 | 35.55 | 35.76 | 35.17 | 33.76 | 34.59 | Negative | Negative | Negative | Positive | Negative | Negative |
| 83 | V0012002 | 29.06 | 17.45 | 17.82 | 18.21 | 35.70 | 35.19 | 33.70 | 34.53 | Positive | Positive | Positive | Positive | Positive | Positive |
| 84 | V0012305 | 29.46 | 23.81 | 25.37 | 26.50 | 35.70 | 35.19 | 33.70 | 34.53 | Positive | Positive | Positive | Positive | Positive | Positive |
| 85 | V0012311 | 25.18 | 36.73 | 34.84 | 34.49 | 35.70 | 35.19 | 33.70 | 34.53 | Negative | Negative | Negative | Negative | Negative | Positive |
| 86 | V0009459 | 28.97 | 35.91 | 35.35 | 35.86 | 35.89 | 35.44 | 33.89 | 34.75 | Negative | Negative | Negative | Negative | Negative | Negative |
| 87 | V0010733 | 29.56 | 16.35 | 18.29 | 18.17 | 36.45 | 34.90 | 34.45 | 35.37 | Positive | Positive | Positive | Positive | Positive | Positive |
| 88 | V0010756 | 29.71 | 34.39 | 37.02 | 36.20 | 36.45 | 34.90 | 34.45 | 35.37 | Positive | Negative | Negative | Positive | Negative | Negative |
| 89 | V0011595 | 28.00 | 21.16 | 23.21 | 23.50 | 36.70 | 34.78 | 34.70 | 35.65 | Positive | Positive | Positive | Positive | Positive | Positive |
| 90 | V0011727 | 28.48 | 27.96 | 29.20 | 29.03 | 36.70 | 34.78 | 34.70 | 35.65 | Positive | Positive | Positive | Positive | Positive | Positive |
| 91 | V0011736 | 28.71 | 34.72 | 34.87 | 35.54 | 36.70 | 34.78 | 34.70 | 35.65 | Negative | Negative | Negative | Positive | Positive | Positive |
| 92 | V0011057 | 27.14 | 34.89 | 35.73 | 35.67 | 36.12 | 36.22 | 34.12 | 35.00 | Negative | Negative | Negative | Positive | Negative | Negative |
| 93 | V0011710 | 28.08 | 35.74 | 34.20 | 36.01 | 36.12 | 36.22 | 34.12 | 35.00 | Negative | Negative | Negative | Negative | Positive | Negative |
| 94 | V0010958 | 27.83 | 37.29 | 37.23 | 45.00 | 37.33 | 36.09 | 35.33 | 36.36 | Negative | Negative | Negative | Negative | Negative | Negative |
| 95 | V0011446 | 27.37 | 23.50 | 25.28 | 25.97 | 37.33 | 36.09 | 35.33 | 36.36 | Positive | Positive | Positive | Positive | Positive | Positive |
| 96 | V0009529 | 28.67 | 33.80 | 36.04 | 36.63 | 37.33 | 36.09 | 35.33 | 36.36 | Positive | Negative | Negative | Positive | Positive | Negative |
| 97 | V0009535 | 29.38 | 15.97 | 17.74 | 18.86 | 37.33 | 36.09 | 35.33 | 36.36 | Positive | Positive | Positive | Positive | Positive | Positive |
| 98 | V0009548 | 28.25 | 36.10 | 37.26 | 37.60 | 37.33 | 36.09 | 35.33 | 36.36 | Negative | Negative | Negative | Positive | Negative | Negative |
| 99 | V0012405 | 28.69 | 33.34 | 32.72 | 33.96 | 36.87 | 35.30 | 34.87 | 35.84 | Positive | Positive | Positive | Positive | Positive | Positive |
| 100 | V0010641 | 26.97 | 33.80 | 34.16 | 33.61 | 35.84 | 35.60 | 33.84 | 34.68 | Positive | Negative | Positive | Positive | Positive | Positive |

Cts for FDA (n=3)

| A | B | C | D | E | F | G | H | I | J | K | L | M | N | O | P | Q | R |
|---|---|---|---|---|---|---|---|---|---|---|---|---|---|---|---|---|---|
| Positive | Specimen code | Date qPCR run | Plate number | $Ct_{RPP}$ | $Ct_{N2}$ | $Ct_{E1}$ | $Ct_{S6}$ | cut-off N | cut-off RPP30 | Theoretical Ct cutoff | Estimated CT cutoff | Theoretical Result N | Theoretical Result E | Theoretical Result S | Estimated Result N | Estimated Result E | Estimated Result S |
| 1 | V0010590 | | | 28.22 | 15.73 | 17.65 | 18.22 | 35.95 | 35.46 | 34.36 | 34.82 | Positive | Positive | Positive | Positive | Positive | Positive |
| 2 | V0012870 | | | 29.06 | 17.38 | 18.63 | 19.21 | 35.95 | 35.46 | 34.36 | 34.82 | Positive | Positive | Positive | Positive | Positive | Positive |
| 3 | V0014312 | | | 27.97 | 14.82 | 16.26 | 16.66 | 35.95 | 35.46 | 34.36 | 34.82 | Positive | Positive | Positive | Positive | Positive | Positive |
| 4 | V0015270 | | | 28.23 | 23.00 | 23.36 | 23.08 | 35.95 | 35.46 | 34.36 | 34.82 | Positive | Positive | Positive | Positive | Positive | Positive |
| 5 | V0015941 | | | 29.27 | 16.67 | 18.43 | 18.47 | 35.95 | 35.46 | 34.36 | 34.82 | Positive | Positive | Positive | Positive | Positive | Positive |
| 6 | V0016364 | | | 27.74 | 36.70 | 33.60 | 35.19 | 35.45 | 35.37 | 33.86 | 34.31 | Negative | Positive | Negative | Negative | Positive | Negative |
| 7 | 258011 | | | 35.23 | 23.38 | 23.41 | 22.96 | 34.75 | 35.43 | 33.17 | 33.61 | Positive | Positive | Positive | Positive | Positive | Positive |
| 8 | 266249 | | | 29.77 | 24.76 | 25.29 | 24.76 | 34.75 | 35.43 | 33.17 | 33.61 | Positive | Positive | Positive | Positive | Positive | Positive |
| 9 | V0016301 | | | 29.11 | 34.33 | 33.48 | 35.10 | 34.75 | 35.43 | 33.17 | 33.61 | Negative | Negative | Negative | Negative | Positive | Negative |
| 10 | V0010606 | | | 28.22 | 30.24 | 30.20 | 30.03 | 35.48 | 35.63 | 33.90 | 34.35 | Positive | Positive | Positive | Positive | Positive | Positive |
| 11 | V0015699 | | | 29.76 | 18.06 | 19.57 | 20.18 | 35.48 | 35.63 | 33.90 | 34.35 | Positive | Positive | Positive | Positive | Positive | Positive |
| 12 | V0014597 | | | 30.07 | 18.06 | 19.20 | 20.00 | 35.26 | 36.36 | 33.67 | 34.12 | Positive | Positive | Positive | Positive | Positive | Positive |
| 13 | V0015158 | | | 28.52 | 32.77 | 32.66 | 34.00 | 35.31 | 36.34 | 33.73 | 34.18 | Positive | Positive | Negative | Positive | Positive | Positive |
| 14 | V0015163 | | | 29.45 | 31.35 | 30.63 | 31.00 | 35.31 | 36.34 | 33.73 | 34.18 | Positive | Positive | Positive | Positive | Positive | Positive |
| 15 | V0010589 | | | 29.76 | 33.66 | 33.57 | 33.70 | 36.21 | 36.58 | 34.62 | 35.08 | Positive | Positive | Positive | Positive | Positive | Positive |
| 16 | V0016168 | | | 29.38 | 34.07 | 33.15 | 33.75 | 35.15 | 35.61 | 33.56 | 34.01 | Negative | Positive | Negative | Negative | Positive | Positive |
| 17 | V0014934 | | | 28.25 | 27.92 | 28.42 | 28.62 | 35.12 | 35.85 | 33.54 | 33.98 | Positive | Positive | Positive | Positive | Positive | Positive |
| 18 | V0015461 | | | 29.25 | 33.51 | 32.90 | 33.58 | 35.12 | 35.85 | 33.54 | 33.98 | Positive | Positive | Negative | Positive | Positive | Positive |
| 19 | V0014582 | | | 28.98 | 30.32 | 30.12 | 30.35 | 35.21 | 36.42 | 33.62 | 34.07 | Positive | Positive | Positive | Positive | Positive | Positive |
| 20 | V0014587 | | | 27.31 | 34.65 | 34.36 | 33.93 | 35.21 | 36.42 | 33.62 | 34.07 | Negative | Negative | Negative | Negative | Negative | Positive |
| 21 | V0015145 | | | 28.12 | 31.64 | 31.54 | 32.14 | 35.21 | 36.42 | 33.62 | 34.07 | Positive | Positive | Positive | Positive | Positive | Positive |
| 22 | V0014584 | | | 28.19 | 21.33 | 23.24 | 24.14 | 36.07 | 35.23 | 34.48 | 34.94 | Positive | Positive | Positive | Positive | Positive | Positive |
| 23 | V0014589 | | | 28.86 | 20.21 | 21.28 | 21.60 | 36.07 | 35.23 | 34.48 | 34.94 | Positive | Positive | Positive | Positive | Positive | Positive |
| 24 | V0014595 | | | 29.05 | 18.88 | 20.66 | 21.64 | 36.07 | 35.23 | 34.48 | 34.94 | Positive | Positive | Positive | Positive | Positive | Positive |
| 25 | V0014616 | | | 28.45 | 31.51 | 31.73 | 32.37 | 35.80 | 35.45 | 34.22 | 34.67 | Positive | Positive | Positive | Positive | Positive | Positive |
| 26 | V0014903 | | | 28.57 | 32.77 | 33.42 | 34.04 | 35.80 | 35.45 | 34.22 | 34.67 | Positive | Positive | Positive | Positive | Positive | Positive |
| 27 | V0014995 | | | 28.96 | 35.25 | 33.92 | 34.60 | 35.80 | 35.45 | 34.22 | 34.67 | Negative | Positive | Negative | Negative | Positive | Positive |
| 28 | 249008 | | | 29.30 | 21.14 | 22.74 | 23.00 | 34.38 | 36.43 | 32.79 | 33.23 | Positive | Positive | Positive | Positive | Positive | Positive |
| 29 | V0014349 | | | 27.23 | 33.86 | 33.41 | 35.71 | 35.26 | 35.46 | 33.68 | 34.12 | Negative | Positive | Negative | Positive | Positive | Negative |
| 30 | 239215 | | | 29.84 | 18.68 | 19.70 | 20.25 | 36.19 | 36.27 | 34.60 | 35.06 | Positive | Positive | Positive | Positive | Positive | Positive |
| 31 | V0014367 | | | 29.57 | 19.07 | 20.62 | 21.66 | 36.19 | 36.27 | 34.60 | 35.06 | Positive | Positive | Positive | Positive | Positive | Positive |
| 32 | V0013598 | | | 30.79 | 20.50 | 22.21 | 22.95 | 36.16 | 36.06 | 34.57 | 35.03 | Positive | Positive | Positive | Positive | Positive | Positive |
| 33 | V0013672 | | | 29.30 | 27.35 | 28.91 | 29.54 | 36.16 | 36.06 | 34.57 | 35.03 | Positive | Positive | Positive | Positive | Positive | Positive |
| 34 | V0244808 | | | 32.05 | 31.01 | 32.18 | 33.34 | 35.95 | 35.40 | 34.37 | 34.82 | Positive | Positive | Positive | Positive | Positive | Positive |
| 35 | V0249008 | | | 30.80 | 22.04 | 23.67 | 24.34 | 35.95 | 35.40 | 34.37 | 34.82 | Positive | Positive | Positive | Positive | Positive | Positive |
| 36 | V0267038 | | | 30.37 | 16.99 | 19.28 | 19.82 | 35.95 | 35.40 | 34.37 | 34.82 | Positive | Positive | Positive | Positive | Positive | Positive |
| 37 | V0014502 | | | 29.25 | 19.77 | 19.55 | 20.06 | 35.68 | 35.84 | 34.09 | 34.55 | Positive | Positive | Positive | Positive | Positive | Positive |
| 38 | V0014521 | | | 30.47 | 25.08 | 24.55 | 24.81 | 35.68 | 35.84 | 34.09 | 34.55 | Positive | Positive | Positive | Positive | Positive | Positive |
| 39 | V0013602 | | | 29.41 | 17.20 | 18.65 | 19.01 | 35.57 | 36.89 | 33.99 | 34.43 | Positive | Positive | Positive | Positive | Positive | Positive |
| 40 | V0013659 | | | 29.99 | 13.49 | 16.05 | 16.61 | 35.80 | 36.81 | 34.21 | 34.66 | Positive | Positive | Positive | Positive | Positive | Positive |
| 41 | V0013660 | | | 28.91 | 19.76 | 20.31 | 20.84 | 35.80 | 36.81 | 34.21 | 34.66 | Positive | Positive | Positive | Positive | Positive | Positive |
| 42 | V0013691 | | | 28.96 | 24.06 | 24.09 | 24.1 | 35.14 | 36.38 | 33.55 | 34.00 | Positive | Positive | Positive | Positive | Positive | Positive |
| 43 | V0013695 | | | 29.90 | 26.89 | 26.79 | 26.9 | 35.14 | 36.38 | 33.55 | 34.00 | Positive | Positive | Positive | Positive | Positive | Positive |
| 44 | V0014147 | | | 27.70 | 24.30 | 24.99 | 25.18 | 36.02 | 36.37 | 34.44 | 34.89 | Positive | Positive | Positive | Positive | Positive | Positive |
| 45 | V0011683 | | | 29.73 | 17.58 | 19.49 | 19.37 | 36.63 | 36.94 | 35.04 | 35.50 | Positive | Positive | Positive | Positive | Positive | Positive |
| 46 | V0013950 | | | 27.19 | 19.63 | 20.55 | 20.56 | 36.63 | 36.94 | 35.04 | 35.50 | Positive | Positive | Positive | Positive | Positive | Positive |
| 47 | V0014047 | | | 27.99 | 24.22 | 25.13 | 25.22 | 36.63 | 36.94 | 35.04 | 35.50 | Positive | Positive | Positive | Positive | Positive | Positive |
| 48 | V0013209 | | | 28.73 | 35.81 | 35.48 | 35.51 | 36.18 | 35.46 | 34.60 | 35.05 | Negative | Negative | Negative | Negative | Negative | Negative |
| 49 | V0014172 | | | 28.39 | 26.94 | 27.60 | 27.08 | 36.18 | 35.46 | 34.60 | 35.05 | Positive | Positive | Positive | Positive | Positive | Positive |
| 50 | V0014208 | | | 28.28 | 22.75 | 23.47 | 23.50 | 36.18 | 35.46 | 34.60 | 35.05 | Positive | Positive | Positive | Positive | Positive | Positive |
| 51 | V0011423 | | | 27.97 | 26.64 | 28.07 | 28.40 | 36.53 | 35.77 | 34.95 | 35.41 | Positive | Positive | Positive | Positive | Positive | Positive |
| 52 | V0014057 | | | 28.89 | 21.48 | 22.35 | 22.57 | 36.53 | 35.77 | 34.95 | 35.41 | Positive | Positive | Positive | Positive | Positive | Positive |

| | | | | | | | | | | | | | | | |
|---|---|---|---|---|---|---|---|---|---|---|---|---|---|---|---|
| 53 | V0011438 | 28.28 | 19.32 | 21.47 | 22.33 | 36.94 | 36.95 | 35.35 | 35.81 | Positive | Positive | Positive | Positive | Positive | Positive |
| 54 | V0012919 | 27.95 | 22.28 | 22.81 | 22.95 | 36.94 | 36.95 | 35.35 | 35.81 | Positive | Positive | Positive | Positive | Positive | Positive |
| 55 | V0014055 | 29.30 | 27.99 | 28.54 | 28.45 | 36.94 | 36.95 | 35.35 | 35.81 | Positive | Positive | Positive | Positive | Positive | Positive |
| 56 | V0014061 | 28.23 | 26.56 | 27.50 | 27.86 | 36.94 | 36.95 | 35.35 | 35.81 | Positive | Positive | Positive | Positive | Positive | Positive |
| 57 | V0012907 | 29.70 | 15.19 | 16.42 | 17.14 | 34.83 | 36.30 | 33.24 | 33.69 | Positive | Positive | Positive | Positive | Positive | Positive |
| 58 | V0003662 | 29.15 | 33.54 | 34.72 | 35.70 | 34.83 | 36.30 | 33.24 | 33.69 | Negative | Negative | Negative | Positive | Negative | Negative |
| 59 | V0012806 | 29.20 | 27.11 | 26.94 | 26.93 | 37.03 | 35.40 | 35.44 | 35.90 | Positive | Positive | Positive | Positive | Positive | Positive |
| 60 | V0012116 | 29.97 | 35.21 | 36.56 | 36.21 | 37.03 | 35.40 | 35.44 | 35.90 | Positive | Negative | Negative | Positive | Negative | Negative |
| 61 | V0012153 | 29.33 | 34.55 | 37.44 | 35.54 | 37.03 | 35.40 | 35.44 | 35.90 | Positive | Negative | Negative | Positive | Negative | Positive |
| 62 | V0012513 | 32.64 | 35.03 | 35.09 | 36.12 | 36.60 | 36.96 | 35.02 | 35.48 | Negative | Negative | Negative | Positive | Positive | Negative |
| 63 | V0014030 | 29.40 | 36.55 | 36.39 | 36.83 | 36.60 | 36.96 | 35.02 | 35.48 | Negative | Negative | Negative | Negative | Negative | Negative |
| 64 | V0011709 | 28.99 | 34.29 | 34.75 | 36.70 | 36.60 | 36.96 | 35.02 | 35.48 | Positive | Positive | Negative | Positive | Positive | Negative |
| 65 | V0012038 | 27.46 | 27.07 | 27.58 | 27.68 | 36.46 | 35.56 | 34.88 | 35.34 | Positive | Positive | Positive | Positive | Positive | Positive |
| 66 | V0012076 | 30.36 | 20.23 | 21.60 | 21.85 | 36.46 | 35.56 | 34.88 | 35.34 | Positive | Positive | Positive | Positive | Positive | Positive |
| 67 | V0013487 | 30.13 | 34.67 | 34.95 | 35.82 | 36.46 | 35.56 | 34.88 | 35.34 | Positive | Negative | Negative | Positive | Positive | Negative |
| 68 | V0012539 | 32.13 | 36.04 | 36.52 | 36.30 | 36.37 | 36.90 | 34.78 | 35.24 | Negative | Negative | Negative | Negative | Negative | Negative |
| 69 | V0013218 | 29.54 | 20.20 | 20.24 | 20.29 | 36.37 | 36.90 | 34.78 | 35.24 | Positive | Positive | Positive | Positive | Positive | Positive |
| 70 | V0013385 | 28.72 | 27.71 | 27.52 | 27.85 | 36.37 | 36.90 | 34.78 | 35.24 | Positive | Positive | Positive | Positive | Positive | Positive |
| 71 | V0013210 | 28.33 | 29.53 | 29.88 | 29.79 | 36.09 | 35.41 | 34.51 | 34.96 | Positive | Positive | Positive | Positive | Positive | Positive |
| 72 | V0012016 | 28.18 | 25.03 | 23.84 | 23.99 | 36.04 | 36.13 | 34.46 | 34.91 | Positive | Positive | Positive | Positive | Positive | Positive |
| 73 | V0012096 | 27.94 | 29.64 | 30.30 | 30.29 | 36.04 | 36.13 | 34.46 | 34.91 | Positive | Positive | Positive | Positive | Positive | Positive |
| 74 | V0012100 | 28.21 | 25.32 | 25.51 | 25.89 | 36.04 | 36.13 | 34.46 | 34.91 | Positive | Positive | Positive | Positive | Positive | Positive |
| 75 | V0012315 | 30.14 | 36.14 | 34.94 | 35.90 | 35.98 | 35.56 | 34.39 | 34.84 | Negative | Negative | Negative | Negative | Negative | Negative |
| 76 | V0010757 | 28.60 | 16.49 | 17.23 | 17.09 | 36.11 | 36.43 | 34.52 | 34.98 | Positive | Positive | Positive | Positive | Positive | Positive |
| 77 | V0010968 | 28.45 | 34.05 | 35.83 | 35.88 | 36.24 | 36.28 | 34.66 | 35.11 | Positive | Negative | Negative | Positive | Negative | Negative |
| 78 | V0012065 | 29.89 | 35.11 | 34.55 | 36.57 | 36.24 | 36.28 | 34.66 | 35.11 | Negative | Positive | Negative | Positive | Positive | Negative |
| 79 | V0012208 | 28.20 | 32.33 | 32.87 | 33.42 | 36.24 | 36.28 | 34.66 | 35.11 | Positive | Positive | Positive | Positive | Positive | Positive |
| 80 | V0012303 | 30.40 | 18.75 | 20.23 | 20.66 | 34.72 | 36.43 | 33.14 | 33.58 | Positive | Positive | Positive | Positive | Positive | Positive |
| 81 | V0012307 | 31.77 | 21.62 | 23.33 | 24.10 | 34.72 | 36.43 | 33.14 | 33.58 | Positive | Positive | Positive | Positive | Positive | Positive |
| 82 | V0011361 | 30.28 | 34.42 | 36.63 | 35.55 | 35.76 | 35.17 | 34.17 | 34.63 | Negative | Negative | Negative | Positive | Negative | Negative |
| 83 | V0012002 | 29.06 | 17.45 | 17.82 | 18.21 | 35.70 | 35.19 | 34.11 | 34.56 | Positive | Positive | Positive | Positive | Positive | Positive |
| 84 | V0012305 | 29.46 | 23.81 | 25.37 | 26.50 | 35.70 | 35.19 | 34.11 | 34.56 | Positive | Positive | Positive | Positive | Positive | Positive |
| 85 | V0012311 | 25.18 | 36.73 | 34.84 | 34.49 | 35.70 | 35.19 | 34.11 | 34.56 | Negative | Negative | Negative | Negative | Negative | Positive |
| 86 | V0009459 | 28.97 | 35.91 | 35.35 | 35.86 | 35.89 | 35.44 | 34.31 | 34.76 | Negative | Negative | Negative | Negative | Negative | Negative |
| 87 | V0010733 | 29.56 | 16.35 | 18.29 | 18.17 | 36.45 | 34.90 | 34.86 | 35.32 | Positive | Positive | Positive | Positive | Positive | Positive |
| 88 | V0010756 | 29.71 | 34.39 | 37.02 | 36.20 | 36.45 | 34.90 | 34.86 | 35.32 | Positive | Negative | Negative | Positive | Negative | Negative |
| 89 | V0011595 | 28.00 | 21.16 | 23.21 | 23.50 | 36.70 | 34.78 | 35.11 | 35.57 | Positive | Positive | Positive | Positive | Positive | Positive |
| 90 | V0011727 | 28.48 | 27.96 | 29.20 | 29.03 | 36.70 | 34.78 | 35.11 | 35.57 | Positive | Positive | Positive | Positive | Positive | Positive |
| 91 | V0011736 | 28.71 | 34.72 | 34.87 | 35.54 | 36.70 | 34.78 | 35.11 | 35.57 | Positive | Positive | Negative | Positive | Positive | Positive |
| 92 | V0011057 | 27.14 | 34.89 | 35.73 | 35.67 | 36.12 | 36.22 | 34.53 | 34.99 | Negative | Negative | Negative | Positive | Negative | Negative |
| 93 | V0011710 | 28.08 | 35.74 | 34.20 | 36.01 | 36.12 | 36.22 | 34.53 | 34.99 | Negative | Positive | Negative | Negative | Positive | Negative |
| 94 | V0010958 | 27.83 | 37.29 | 37.23 | 45.00 | 37.33 | 36.09 | 35.75 | 36.21 | Negative | Negative | Negative | Negative | Negative | Negative |
| 95 | V0011446 | 27.37 | 23.50 | 25.28 | 25.97 | 37.33 | 36.09 | 35.75 | 36.21 | Positive | Positive | Positive | Positive | Positive | Positive |
| 96 | V0009529 | 28.67 | 33.80 | 36.04 | 36.63 | 37.33 | 36.09 | 35.75 | 36.21 | Positive | Negative | Negative | Positive | Positive | Negative |
| 97 | V0009535 | 29.38 | 15.97 | 17.74 | 18.86 | 37.33 | 36.09 | 35.75 | 36.21 | Positive | Positive | Positive | Positive | Positive | Positive |
| 98 | V0009548 | 28.25 | 36.10 | 37.26 | 37.60 | 37.33 | 36.09 | 35.75 | 36.21 | Negative | Negative | Negative | Positive | Negative | Negative |
| 99 | V0012405 | 28.69 | 33.34 | 32.72 | 33.96 | 36.87 | 35.30 | 35.28 | 35.75 | Positive | Positive | Positive | Positive | Positive | Positive |
| 100 | V0010641 | 26.97 | 33.80 | 34.16 | 33.61 | 35.84 | 35.60 | 34.25 | 34.71 | Positive | Positive | Positive | Positive | Positive | Positive |

Cts for FDA (n=2)

| A | B | C | D | E | F | G | H | I | J | K | L | M | N | O | P | Q | R |
|---|---|---|---|---|---|---|---|---|---|---|---|---|---|---|---|---|---|
| Positive | Specimen code | Date qPCR run | Plate number | $Ct_{RPP}$ | $Ct_{N2}$ | $Ct_{E1}$ | $Ct_{S6}$ | cut-off N | cut-off RPP30 | Theoretical Ct cutoff | Estimated CT cutoff | Theoretical Result N | Theoretical Result E | Theoretical Result S | Estimated Result N | Estimated Result E | Estimated Result S |
| 1 | V0010590 | | | 28.22 | 15.73 | 17.65 | 18.22 | 35.95 | 35.46 | 34.95 | 37.05 | Positive | Positive | Positive | Positive | Positive | Positive |
| 2 | V0012870 | | | 29.06 | 17.38 | 18.63 | 19.21 | 35.95 | 35.46 | 34.95 | 37.05 | Positive | Positive | Positive | Positive | Positive | Positive |
| 3 | V0014312 | | | 27.97 | 14.82 | 16.26 | 16.66 | 35.95 | 35.46 | 34.95 | 37.05 | Positive | Positive | Positive | Positive | Positive | Positive |
| 4 | V0015270 | | | 28.23 | 23.00 | 23.36 | 23.08 | 35.95 | 35.46 | 34.95 | 37.05 | Positive | Positive | Positive | Positive | Positive | Positive |
| 5 | V0015941 | | | 29.27 | 16.67 | 18.43 | 18.47 | 35.95 | 35.46 | 34.95 | 37.05 | Positive | Positive | Positive | Positive | Positive | Positive |
| 6 | V0016364 | | | 27.74 | 36.70 | 33.60 | 35.19 | 35.45 | 35.37 | 34.45 | 36.50 | Negative | Positive | Negative | Negative | Positive | Positive |
| 7 | ###011 | | | 35.23 | 23.38 | 23.41 | 22.96 | 34.75 | 35.43 | 33.75 | 35.73 | Positive | Positive | Positive | Positive | Positive | Positive |
| 8 | ###249 | | | 29.77 | 24.76 | 25.29 | 24.76 | 34.75 | 35.43 | 33.75 | 35.73 | Positive | Positive | Positive | Positive | Positive | Positive |
| 9 | V0016301 | | | 29.11 | 34.33 | 33.48 | 35.10 | 34.75 | 35.43 | 33.75 | 35.73 | Negative | Positive | Negative | Positive | Positive | Positive |
| 10 | V0010606 | | | 28.22 | 30.24 | 30.20 | 30.03 | 35.48 | 35.63 | 34.48 | 36.54 | Positive | Positive | Positive | Positive | Positive | Positive |
| 11 | V0015699 | | | 29.76 | 18.06 | 19.57 | 20.18 | 35.48 | 35.63 | 34.48 | 36.54 | Positive | Positive | Positive | Positive | Positive | Positive |
| 12 | V0014597 | | | 30.07 | 18.06 | 19.20 | 20.00 | 35.26 | 36.36 | 34.26 | 36.29 | Positive | Positive | Positive | Positive | Positive | Positive |
| 13 | V0015158 | | | 28.52 | 32.77 | 32.66 | 34.00 | 35.31 | 36.34 | 34.31 | 36.35 | Positive | Positive | Positive | Positive | Positive | Positive |
| 14 | V0015163 | | | 29.45 | 31.35 | 30.63 | 31.00 | 35.31 | 36.34 | 34.31 | 36.35 | Positive | Positive | Positive | Positive | Positive | Positive |
| 15 | V0010589 | | | 29.76 | 33.66 | 33.57 | 33.70 | 36.21 | 36.58 | 35.21 | 37.34 | Positive | Positive | Positive | Positive | Positive | Positive |
| 16 | V0016168 | | | 29.38 | 34.07 | 33.15 | 33.75 | 35.15 | 35.61 | 34.15 | 36.16 | Positive | Positive | Positive | Positive | Positive | Positive |
| 17 | V0014934 | | | 28.25 | 27.92 | 28.42 | 28.62 | 35.12 | 35.85 | 34.12 | 36.14 | Positive | Positive | Positive | Positive | Positive | Positive |
| 18 | V0015461 | | | 29.25 | 33.51 | 32.90 | 33.58 | 35.12 | 35.85 | 34.12 | 36.14 | Positive | Positive | Positive | Positive | Positive | Positive |
| 19 | V0014582 | | | 28.98 | 30.32 | 30.12 | 30.35 | 35.21 | 36.42 | 34.21 | 36.23 | Positive | Positive | Positive | Positive | Positive | Positive |
| 20 | V0014587 | | | 27.31 | 34.65 | 34.36 | 33.93 | 35.21 | 36.42 | 34.21 | 36.23 | Negative | Negative | Positive | Positive | Positive | Positive |
| 21 | V0015145 | | | 28.12 | 31.64 | 31.54 | 32.14 | 35.21 | 36.42 | 34.21 | 36.23 | Positive | Positive | Positive | Positive | Positive | Positive |
| 22 | V0014584 | | | 28.19 | 21.33 | 23.24 | 24.14 | 36.07 | 35.23 | 35.07 | 37.19 | Positive | Positive | Positive | Positive | Positive | Positive |
| 23 | V0014589 | | | 28.86 | 20.21 | 21.28 | 21.60 | 36.07 | 35.23 | 35.07 | 37.19 | Positive | Positive | Positive | Positive | Positive | Positive |
| 24 | V0014595 | | | 29.05 | 18.88 | 20.66 | 21.64 | 36.07 | 35.23 | 35.07 | 37.19 | Positive | Positive | Positive | Positive | Positive | Positive |
| 25 | V0014616 | | | 28.45 | 31.51 | 31.73 | 32.37 | 35.80 | 35.45 | 34.80 | 36.89 | Positive | Positive | Positive | Positive | Positive | Positive |
| 26 | V0014903 | | | 28.57 | 32.77 | 33.42 | 34.04 | 35.80 | 35.45 | 34.80 | 36.89 | Positive | Positive | Positive | Positive | Positive | Positive |
| 27 | V0014995 | | | 28.96 | 35.25 | 33.92 | 34.60 | 35.80 | 35.45 | 34.80 | 36.89 | Negative | Positive | Positive | Positive | Positive | Positive |
| 28 | ###008 | | | 29.30 | 21.14 | 22.74 | 23.00 | 34.38 | 36.43 | 33.38 | 35.31 | Positive | Positive | Positive | Positive | Positive | Positive |
| 29 | V0014349 | | | 27.23 | 33.86 | 33.41 | 35.71 | 35.26 | 35.46 | 34.26 | 36.29 | Positive | Positive | Negative | Positive | Positive | Positive |
| 30 | ###215 | | | 29.84 | 18.68 | 19.70 | 20.25 | 36.19 | 36.27 | 35.19 | 37.32 | Positive | Positive | Positive | Positive | Positive | Positive |
| 31 | V0014367 | | | 29.57 | 19.07 | 20.62 | 21.66 | 36.19 | 36.27 | 35.19 | 37.32 | Positive | Positive | Positive | Positive | Positive | Positive |
| 32 | V0013598 | | | 30.79 | 20.50 | 22.21 | 22.95 | 36.16 | 36.06 | 35.16 | 37.29 | Positive | Positive | Positive | Positive | Positive | Positive |
| 33 | V0013672 | | | 29.30 | 27.35 | 28.91 | 29.54 | 36.16 | 36.06 | 35.16 | 37.29 | Positive | Positive | Positive | Positive | Positive | Positive |
| 34 | V0244808 | | | 32.05 | 31.01 | 32.18 | 33.34 | 35.95 | 35.40 | 34.95 | 37.06 | Positive | Positive | Positive | Positive | Positive | Positive |
| 35 | V0249008 | | | 30.80 | 22.04 | 23.67 | 24.34 | 35.95 | 35.40 | 34.95 | 37.06 | Positive | Positive | Positive | Positive | Positive | Positive |
| 36 | V0267038 | | | 30.37 | 16.99 | 19.28 | 19.82 | 35.95 | 35.40 | 34.95 | 37.06 | Positive | Positive | Positive | Positive | Positive | Positive |
| 37 | V0014502 | | | 29.25 | 19.77 | 19.55 | 20.06 | 35.68 | 35.84 | 34.68 | 36.76 | Positive | Positive | Positive | Positive | Positive | Positive |
| 38 | V0014521 | | | 30.47 | 25.08 | 24.55 | 24.81 | 35.68 | 35.84 | 34.68 | 36.76 | Positive | Positive | Positive | Positive | Positive | Positive |
| 39 | V0013602 | | | 29.41 | 17.20 | 18.65 | 19.01 | 35.57 | 36.89 | 34.57 | 36.63 | Positive | Positive | Positive | Positive | Positive | Positive |
| 40 | V0013659 | | | 29.99 | 13.49 | 16.05 | 16.61 | 35.80 | 36.81 | 34.80 | 36.89 | Positive | Positive | Positive | Positive | Positive | Positive |
| 41 | V0013660 | | | 28.91 | 19.76 | 20.31 | 20.84 | 35.80 | 36.81 | 34.80 | 36.89 | Positive | Positive | Positive | Positive | Positive | Positive |
| 42 | V0013691 | | | 28.96 | 24.06 | 24.09 | 24.1 | 35.14 | 36.38 | 34.14 | 36.15 | Positive | Positive | Positive | Positive | Positive | Positive |
| 43 | V0013695 | | | 29.90 | 26.89 | 26.79 | 26.9 | 35.14 | 36.38 | 34.14 | 36.15 | Positive | Positive | Positive | Positive | Positive | Positive |
| 44 | V0014147 | | | 27.70 | 24.30 | 24.99 | 25.18 | 36.02 | 36.37 | 35.02 | 37.14 | Positive | Positive | Positive | Positive | Positive | Positive |
| 45 | V0011683 | | | 29.73 | 17.58 | 19.49 | 19.37 | 36.63 | 36.94 | 35.63 | 37.81 | Positive | Positive | Positive | Positive | Positive | Positive |
| 46 | V0013950 | | | 27.19 | 19.63 | 20.55 | 20.56 | 36.63 | 36.94 | 35.63 | 37.81 | Positive | Positive | Positive | Positive | Positive | Positive |
| 47 | V0014047 | | | 27.99 | 24.22 | 25.13 | 25.22 | 36.63 | 36.94 | 35.63 | 37.81 | Positive | Positive | Positive | Positive | Positive | Positive |
| 48 | V0013209 | | | 28.73 | 35.81 | 35.48 | 35.51 | 36.18 | 35.46 | 35.18 | 37.32 | Negative | Negative | Negative | Positive | Positive | Positive |
| 49 | V0014172 | | | 28.39 | 26.94 | 27.60 | 27.08 | 36.18 | 35.46 | 35.18 | 37.32 | Positive | Positive | Positive | Positive | Positive | Positive |
| 50 | V0014208 | | | 28.28 | 22.75 | 23.47 | 23.50 | 36.18 | 35.46 | 35.18 | 37.32 | Positive | Positive | Positive | Positive | Positive | Positive |

| | | | | | | | | | | | | | | | |
|---|---|---|---|---|---|---|---|---|---|---|---|---|---|---|---|
| 51 | V0011423 | 27.97 | 26.64 | 28.07 | 28.40 | 36.53 | 35.77 | 35.53 | 37.70 | Positive | Positive | Positive | Positive | Positive | Positive |
| 52 | V0014057 | 28.89 | 21.48 | 22.35 | 22.57 | 36.53 | 35.77 | 35.53 | 37.70 | Positive | Positive | Positive | Positive | Positive | Positive |
| 53 | V0011438 | 28.28 | 19.32 | 21.47 | 22.33 | 36.94 | 36.95 | 35.94 | 38.15 | Positive | Positive | Positive | Positive | Positive | Positive |
| 54 | V0012919 | 27.95 | 22.28 | 22.81 | 22.95 | 36.94 | 36.95 | 35.94 | 38.15 | Positive | Positive | Positive | Positive | Positive | Positive |
| 55 | V0014055 | 29.30 | 27.99 | 28.54 | 28.45 | 36.94 | 36.95 | 35.94 | 38.15 | Positive | Positive | Positive | Positive | Positive | Positive |
| 56 | V0014061 | 28.23 | 26.56 | 27.50 | 27.86 | 36.94 | 36.95 | 35.94 | 38.15 | Positive | Positive | Positive | Positive | Positive | Positive |
| 57 | V0012907 | 29.70 | 15.19 | 16.42 | 17.14 | 34.83 | 36.30 | 33.83 | 35.81 | Positive | Positive | Positive | Positive | Positive | Positive |
| 58 | V0003662 | 29.15 | 33.54 | 34.72 | 35.70 | 34.83 | 36.30 | 33.83 | 35.81 | Positive | Negative | Negative | Positive | Positive | Positive |
| 59 | V0012806 | 29.20 | 27.11 | 26.94 | 26.93 | 37.03 | 35.40 | 36.03 | 38.25 | Positive | Positive | Positive | Positive | Positive | Positive |
| 60 | V0012116 | 29.97 | 35.21 | 36.56 | 36.21 | 37.03 | 35.40 | 36.03 | 38.25 | Positive | Negative | Negative | Positive | Positive | Positive |
| 61 | V0012153 | 29.33 | 34.55 | 37.44 | 35.54 | 37.03 | 35.40 | 36.03 | 38.25 | Positive | Negative | Positive | Positive | Positive | Positive |
| 62 | V0012513 | 32.64 | 35.03 | 35.09 | 36.12 | 36.60 | 36.96 | 35.60 | 37.78 | Positive | Positive | Negative | Positive | Positive | Positive |
| 63 | V0014030 | 29.40 | 36.55 | 36.39 | 36.83 | 36.60 | 36.96 | 35.60 | 37.78 | Negative | Negative | Negative | Positive | Positive | Positive |
| 64 | V0011709 | 28.99 | 34.29 | 34.75 | 36.70 | 36.60 | 36.96 | 35.60 | 37.78 | Positive | Positive | Negative | Positive | Positive | Positive |
| 65 | V0012038 | 27.46 | 27.07 | 27.58 | 27.68 | 36.46 | 35.56 | 35.46 | 37.63 | Positive | Positive | Positive | Positive | Positive | Positive |
| 66 | V0012076 | 30.36 | 20.23 | 21.60 | 21.85 | 36.46 | 35.56 | 35.46 | 37.63 | Positive | Positive | Positive | Positive | Positive | Positive |
| 67 | V0013487 | 30.13 | 34.67 | 34.95 | 35.82 | 36.46 | 35.56 | 35.46 | 37.63 | Positive | Positive | Negative | Positive | Positive | Positive |
| 68 | V0012539 | 32.13 | 36.04 | 36.52 | 36.30 | 36.37 | 36.90 | 35.37 | 37.52 | Negative | Negative | Negative | Positive | Positive | Positive |
| 69 | V0013218 | 29.54 | 20.20 | 20.24 | 20.29 | 36.37 | 36.90 | 35.37 | 37.52 | Positive | Positive | Positive | Positive | Positive | Positive |
| 70 | V0013385 | 28.72 | 27.71 | 27.52 | 27.85 | 36.37 | 36.90 | 35.37 | 37.52 | Positive | Positive | Positive | Positive | Positive | Positive |
| 71 | V0013210 | 28.33 | 29.53 | 29.88 | 29.79 | 36.09 | 35.41 | 35.09 | 37.21 | Positive | Positive | Positive | Positive | Positive | Positive |
| 72 | V0012016 | 28.18 | 25.03 | 23.84 | 23.99 | 36.04 | 36.13 | 35.04 | 37.16 | Positive | Positive | Positive | Positive | Positive | Positive |
| 73 | V0012096 | 27.94 | 29.64 | 30.30 | 30.29 | 36.04 | 36.13 | 35.04 | 37.16 | Positive | Positive | Positive | Positive | Positive | Positive |
| 74 | V0012100 | 28.21 | 25.32 | 25.51 | 25.89 | 36.04 | 36.13 | 35.04 | 37.16 | Positive | Positive | Positive | Positive | Positive | Positive |
| 75 | V0012315 | 30.14 | 36.14 | 34.94 | 35.90 | 35.98 | 35.56 | 34.98 | 37.08 | Negative | Positive | Negative | Positive | Positive | Positive |
| 76 | V0010757 | 28.60 | 16.49 | 17.23 | 17.09 | 36.11 | 36.43 | 35.11 | 37.23 | Positive | Positive | Positive | Positive | Positive | Positive |
| 77 | V0010968 | 28.45 | 34.05 | 35.83 | 35.88 | 36.24 | 36.28 | 35.24 | 37.38 | Positive | Negative | Negative | Positive | Positive | Positive |
| 78 | V0012065 | 29.89 | 35.11 | 34.55 | 36.57 | 36.24 | 36.28 | 35.24 | 37.38 | Positive | Positive | Negative | Positive | Positive | Positive |
| 79 | V0012208 | 28.20 | 32.33 | 32.87 | 33.42 | 36.24 | 36.28 | 35.24 | 37.38 | Positive | Positive | Positive | Positive | Positive | Positive |
| 80 | V0012303 | 30.40 | 18.75 | 20.23 | 20.66 | 34.72 | 36.43 | 33.72 | 35.69 | Positive | Positive | Positive | Positive | Positive | Positive |
| 81 | V0012307 | 31.77 | 21.62 | 23.33 | 24.10 | 34.72 | 36.43 | 33.72 | 35.69 | Positive | Positive | Positive | Positive | Positive | Positive |
| 82 | V0011361 | 30.28 | 34.42 | 36.63 | 35.55 | 35.76 | 35.17 | 34.76 | 36.84 | Positive | Negative | Negative | Positive | Positive | Positive |
| 83 | V0012002 | 29.06 | 17.45 | 17.82 | 18.21 | 35.70 | 35.19 | 34.70 | 36.78 | Positive | Positive | Positive | Positive | Positive | Positive |
| 84 | V0012305 | 29.46 | 23.81 | 25.37 | 26.50 | 35.70 | 35.19 | 34.70 | 36.78 | Positive | Positive | Positive | Positive | Positive | Positive |
| 85 | V0012311 | 25.18 | 36.73 | 34.84 | 34.49 | 35.70 | 35.19 | 34.70 | 36.78 | Negative | Negative | Positive | Positive | Positive | Positive |
| 86 | V0009459 | 28.97 | 35.91 | 35.35 | 35.86 | 35.89 | 35.44 | 34.89 | 36.99 | Negative | Negative | Negative | Positive | Positive | Positive |
| 87 | V0010733 | 29.56 | 16.35 | 18.29 | 18.17 | 36.45 | 34.90 | 35.45 | 37.61 | Positive | Positive | Positive | Positive | Positive | Positive |
| 88 | V0010756 | 29.71 | 34.39 | 37.02 | 36.20 | 36.45 | 34.90 | 35.45 | 37.61 | Positive | Negative | Negative | Positive | Positive | Positive |
| 89 | V0011595 | 28.00 | 21.16 | 23.21 | 23.50 | 36.70 | 34.78 | 35.70 | 37.89 | Positive | Positive | Positive | Positive | Positive | Positive |
| 90 | V0011727 | 28.48 | 27.96 | 29.20 | 29.03 | 36.70 | 34.78 | 35.70 | 37.89 | Positive | Positive | Positive | Positive | Positive | Positive |
| 91 | V0011736 | 28.71 | 34.72 | 34.87 | 35.54 | 36.70 | 34.78 | 35.70 | 37.89 | Positive | Positive | Positive | Positive | Positive | Positive |
| 92 | V0011057 | 27.14 | 34.89 | 35.73 | 35.67 | 36.12 | 36.22 | 35.12 | 37.24 | Positive | Negative | Negative | Positive | Positive | Positive |
| 93 | V0011710 | 28.08 | 35.74 | 34.20 | 36.01 | 36.12 | 36.22 | 35.12 | 37.24 | Negative | Positive | Negative | Positive | Positive | Positive |
| 94 | V0010958 | 27.83 | 37.29 | 37.23 | 45.00 | 37.33 | 36.09 | 36.33 | 38.59 | Negative | Negative | Negative | Positive | Positive | Negative |
| 95 | V0011446 | 27.37 | 23.50 | 25.28 | 25.97 | 37.33 | 36.09 | 36.33 | 38.59 | Positive | Positive | Positive | Positive | Positive | Positive |
| 96 | V0009529 | 28.67 | 33.80 | 36.04 | 36.63 | 37.33 | 36.09 | 36.33 | 38.59 | Positive | Positive | Negative | Positive | Positive | Positive |
| 97 | V0009535 | 29.38 | 15.97 | 17.74 | 18.86 | 37.33 | 36.09 | 36.33 | 38.59 | Positive | Positive | Positive | Positive | Positive | Positive |
| 98 | V0009548 | 28.25 | 36.10 | 37.26 | 37.60 | 37.33 | 36.09 | 36.33 | 38.59 | Positive | Negative | Negative | Positive | Positive | Positive |
| 99 | V0012405 | 28.69 | 33.34 | 32.72 | 33.96 | 36.87 | 35.30 | 35.87 | 38.08 | Positive | Positive | Positive | Positive | Positive | Positive |
| 100 | V0010641 | 26.97 | 33.80 | 34.16 | 33.61 | 35.84 | 35.60 | 34.84 | 36.93 | Positive | Positive | Positive | Positive | Positive | Positive |
