## Supplementary material for "Development and Implementation of a scalable and versatile test for COVID-19 diagnostics in rural communities": Sup_Table XVII

**Saliva samples**

| A | B | C | D | E | F | G | H | I | J | K | L | M | N | O | P | Q | R | S | T | U | V | W |
|---|---|---|---|---|---|---|---|---|---|---|---|---|---|---|---|---|---|---|---|---|---|---|
| Patient number | Swab VT ID | Swab date received | Swab plate number | Sw $Ct_{RPP}$ | Sw $Ct_N$ | Sw $Ct_E$ | Sw $Ct_S$ | Sw-RPP cutoff | Sw-N cutoff | Swab result | Saliva VT ID | Saliva date received | Saliva plate number | Sl $Ct_{RPP}$ | Sl $Ct_N$ | Sl $Ct_E$ | Sl $Ct_S$ | Sl-RPP cutoff | Sl-N cuttoff | Saliva result | Swab and Saliva match | Final call |
| | V0008867 | | | 29.62 | N/A | N/A | N/A | 36.94 | 35.36 | Negative | V0012691 | | | 29.93 | N/A | N/A | 44.43 | 36.94 | 35.36 | Negative | true | True negative |
| | V0009268 | | | 28.60 | 41.33 | N/A | 41.85 | 36.94 | 35.36 | Negative | V0012713 | | | 35.69 | N/A | N/A | N/A | 36.94 | 35.36 | Negative | true | True negative |
| | V0009379 | | | 30.53 | N/A | N/A | N/A | 36.94 | 35.36 | Negative | V0012726 | | | 34.98 | N/A | N/A | N/A | 36.94 | 35.36 | Negative | true | True negative |
| | V0010862 | | | 28.46 | N/A | N/A | 38.63 | 36.94 | 35.36 | Negative | V0012701 | | | 30.38 | N/A | N/A | N/A | 36.94 | 35.36 | Negative | true | True negative |
| | V0010888 | | | 28.49 | N/A | N/A | 38.94 | 36.94 | 35.36 | Negative | V0012707 | | | 30.64 | 37.43 | N/A | 41.53 | 36.94 | 35.36 | Negative | true | True negative |
| | V0010750 | | | 30.04 | 36.63 | 36.73 | 36.17 | 38.28 | 35.38 | Negative | V0012728 | | | 30.76 | N/A | 43.92 | N/A | 38.28 | 35.38 | Negative | true | True negative |
| | V0011036 | | | 28.97 | 44.53 | 39.92 | 43.86 | 38.28 | 35.38 | Negative | V0012689 | | | 34.96 | N/A | N/A | N/A | 37.60 | 35.92 | Negative | true | True negative |
| | V0010899 | | | 29.34 | N/A | N/A | N/A | 37.60 | 35.92 | Negative | V0012492 | | | 33.14 | N/A | N/A | N/A | 37.60 | 35.92 | Negative | true | True negative |
| | V0011029 | | | 28.76 | N/A | N/A | N/A | 37.60 | 35.92 | Negative | V0012685 | | | 36.11 | N/A | N/A | N/A | 37.60 | 35.92 | Negative | true | True negative |
| | V0011039 | | | 28.91 | N/A | N/A | 38.45 | 37.60 | 35.92 | Negative | V0012682 | | | 33.15 | N/A | N/A | N/A | 37.60 | 35.92 | Negative | true | True negative |
| | V0011045 | | | 28.74 | N/A | N/A | N/A | 37.60 | 35.92 | Negative | V0012725 | | | 32.57 | 43.35 | N/A | 42.64 | 37.60 | 35.92 | Negative | true | True negative |
| | V0011054 | | | 27.90 | N/A | 42.16 | 39.60 | 37.60 | 35.92 | Negative | V0012719 | | | 34.13 | N/A | N/A | N/A | 37.60 | 35.92 | Negative | true | True negative |
| | V0011767 | | | 29.27 | N/A | N/A | N/A | 37.60 | 35.92 | Negative | V0012489 | | | 32.37 | N/A | N/A | N/A | 37.60 | 35.92 | Negative | true | True negative |
| | V0012985 | | | 29.12 | 37.91 | N/A | N/A | 36.22 | 36.12 | Negative | V0012720 | | | 32.44 | N/A | N/A | N/A | 36.13 | 36.95 | Negative | true | True negative |
| | V0011484 | | | 28.79 | N/A | N/A | N/A | 39.12 | 37.98 | Negative | V0012490 | | | 32.08 | N/A | N/A | N/A | 39.12 | 37.98 | Negative | true | True negative |
| | V0010674 | | | 28.13 | 43.35 | N/A | 43.10 | 37.30 | 37.12 | Negative | V0012487 | | | 38.06 | N/A | N/A | N/A | 37.30 | 37.12 | Negative | true | True negative |
| | V0011742 | | | 28.20 | 44.75 | N/A | N/A | 37.30 | 37.12 | Negative | V0012493 | | | 32.66 | 42.60 | 41.67 | N/A | 37.30 | 37.12 | Negative | true | True negative |
| | V0010710 | | | 26.69 | N/A | N/A | N/A | 37.30 | 37.12 | Negative | V0012496 | | | 35.36 | N/A | N/A | 42.02 | 37.30 | 37.12 | Negative | true | True negative |
| | V0010702 | | | 28.03 | N/A | N/A | 41.83 | 37.30 | 37.12 | Negative | V0012575 | | | 31.87 | 41.56 | 41.20 | N/A | 37.30 | 37.12 | Negative | true | True negative |
| | V0011658 | | | 28.37 | N/A | 38.07 | N/A | 37.30 | 37.12 | Negative | V0012683 | | | 32.42 | N/A | N/A | N/A | 37.30 | 37.12 | Negative | true | True negative |
| | V0010644 | | | 29.18 | N/A | N/A | 43.52 | 37.30 | 37.12 | Negative | V0012690 | | | 30.53 | N/A | N/A | 43.73 | 37.30 | 37.12 | Negative | true | True negative |
| | V0013010 | | | 28.21 | 43.26 | N/A | 37.98 | 34.78 | 36.70 | Negative | V0012579 | | | 34.78 | N/A | N/A | N/A | 35.17 | 35.76 | Negative | true | True negative |
| | V0010762 | | | 29.17 | N/A | 43.66 | N/A | 36.09 | 37.33 | Negative | V0012574 | | | 29.33 | N/A | 39.45 | 38.15 | 35.17 | 35.76 | Negative | true | True negative |
| | V0011433 | | | 28.97 | 40.69 | 39.36 | 42.44 | 36.09 | 37.33 | Negative | V0012518 | | | 31.47 | N/A | N/A | 43.68 | 35.17 | 35.76 | Negative | true | True negative |
| | V0011650 | | | 31.02 | N/A | N/A | N/A | 35.44 | 35.89 | Negative | V0012521 | | | 32.85 | N/A | N/A | N/A | 35.17 | 35.76 | Negative | true | True negative |
| | V0011678 | | | 29.09 | 42.88 | 42.01 | N/A | 36.22 | 36.12 | Negative | V0012567 | | | 34.98 | N/A | N/A | N/A | 35.17 | 35.76 | Negative | true | True negative |
| | V0013241 | | | 28.14 | 39.25 | N/A | 40.34 | 34.89 | 35.60 | Negative | V0012532 | | | 34.64 | N/A | N/A | N/A | 34.89 | 35.60 | Negative | true | True negative |
| | V0012786 | | | 28.25 | N/A | N/A | 42.23 | 36.30 | 34.83 | Negative | V0012540 | | | 34.40 | N/A | N/A | N/A | 36.91 | 35.28 | Negative | true | True negative |
| | V0012926 | | | 29.32 | N/A | N/A | N/A | 36.30 | 34.83 | Negative | V0012537 | | | 36.21 | N/A | N/A | N/A | 36.91 | 35.28 | Negative | true | True negative |
| | V0012921 | | | 28.25 | N/A | N/A | 44.53 | 36.30 | 34.83 | Negative | V0012511 | | | 32.21 | N/A | N/A | N/A | 36.91 | 35.28 | Negative | true | True negative |
| | V0013398 | | | 28.26 | N/A | N/A | 41.01 | 36.90 | 36.37 | Negative | V0012500 | | | 29.79 | N/A | N/A | N/A | 36.91 | 35.28 | Negative | true | True negative |
| | V0013014 | | | 28.99 | N/A | N/A | N/A | 36.95 | 36.94 | Negative | V0012556 | | | 34.46 | N/A | N/A | N/A | 35.83 | 36.17 | Negative | true | True negative |
| | V0014305 | | | 29.01 | N/A | N/A | N/A | 36.95 | 36.94 | Negative | V0012555 | | | 34.12 | N/A | N/A | N/A | 35.83 | 36.17 | Negative | true | True negative |
| | V0013231 | | | 27.38 | 39.57 | 38.67 | 39.17 | 36.95 | 36.94 | Negative | V0010599 | | | 33.16 | N/A | N/A | N/A | 35.83 | 36.17 | Negative | true | True negative |
| | V0013204 | | | 29.18 | N/A | N/A | N/A | 35.77 | 36.53 | Negative | V0012550 | | | 34.94 | N/A | N/A | N/A | 35.83 | 36.17 | Negative | true | True negative |
| | V0013948 | | | 27.53 | N/A | N/A | N/A | 36.95 | 36.94 | Negative | V0012531 | | | 31.28 | N/A | 37.99 | N/A | 37.06 | 35.68 | Negative | true | True negative |
| | V0014222 | | | 27.04 | 37.62 | N/A | 41.66 | 36.95 | 36.94 | Negative | V0012495 | | | 31.73 | N/A | 44.85 | 41.52 | 37.06 | 35.68 | Negative | true | True negative |
| | V0014249 | | | 28.19 | 39.93 | N/A | 43.67 | 35.77 | 36.53 | Negative | V0012508 | | | 33.84 | N/A | N/A | N/A | 37.06 | 35.68 | Negative | true | True negative |
| | V0014118 | | | 27.31 | N/A | N/A | N/A | 35.77 | 36.53 | Negative | V0012524 | | | 31.10 | N/A | N/A | N/A | 37.06 | 35.68 | Negative | true | True negative |
| | V0014227 | | | 27.88 | 43.83 | N/A | N/A | 35.77 | 36.53 | Negative | V0012560 | | | 35.77 | 37.27 | N/A | N/A | 37.06 | 35.68 | Negative | true | True negative |
| | V0013205 | | | 29.39 | N/A | N/A | N/A | 35.77 | 36.53 | Negative | V0012565 | | | 31.81 | N/A | 39.16 | N/A | 37.06 | 35.68 | Negative | true | True negative |
| | V0014243 | | | 27.88 | 43.07 | N/A | 43.56 | 36.95 | 36.94 | Negative | V0012534 | | | 28.86 | N/A | N/A | N/A | 37.06 | 35.68 | Negative | true | True negative |
| | V0014177 | | | 29.15 | N/A | N/A | 42.86 | 36.95 | 36.94 | Negative | V0012563 | | | 29.85 | N/A | N/A | N/A | 37.06 | 35.68 | Negative | true | True negative |
| | V0014221 | | | 29.22 | N/A | N/A | 40.19 | 36.95 | 36.94 | Negative | V0012519 | | | 31.50 | N/A | 38.10 | N/A | 37.06 | 35.68 | Negative | true | True negative |
| | V0014341 | | | 28.22 | N/A | N/A | 38.89 | 36.95 | 36.94 | Negative | V0012549 | | | 31.26 | N/A | N/A | N/A | 37.06 | 35.68 | Negative | true | True negative |
| | V0013980 | | | 28.42 | 43.10 | 44.80 | N/A | 35.46 | 36.18 | Negative | V0012498 | | | 34.32 | N/A | N/A | 38.65 | 37.06 | 35.68 | Negative | true | True negative |
| | V0014256 | | | 27.56 | 42.38 | N/A | 42.74 | 36.95 | 36.94 | Negative | V0012559 | | | 33.44 | N/A | N/A | N/A | 37.06 | 35.68 | Negative | true | True negative |
| | V0010569 | | | 28.56 | N/A | 41.00 | 43.48 | 36.95 | 36.94 | Negative | V0010597 | | | 31.21 | N/A | N/A | N/A | 37.06 | 35.68 | Negative | true | True negative |
| | V0014168 | | | 26.07 | 44.53 | N/A | N/A | 35.77 | 36.53 | Negative | V0012497 | | | 30.33 | N/A | N/A | N/A | 37.06 | 35.68 | Negative | true | True negative |
| | V0014258 | | | 29.34 | 43.46 | N/A | 38.76 | 35.77 | 36.53 | Negative | V0012527 | | | 30.15 | N/A | N/A | N/A | 37.06 | 35.68 | Negative | true | True negative |
| | V0014022 | | | 30.42 | 41.57 | N/A | N/A | 35.77 | 36.53 | Negative | V0012514 | | | 35.05 | N/A | N/A | 37.47 | 37.06 | 35.68 | Negative | true | True negative |
| | V0014174 | | | 28.14 | N/A | N/A | N/A | 35.77 | 36.53 | Negative | V0012516 | | | 32.72 | N/A | N/A | 39.05 | 37.06 | 35.68 | Negative | true | True negative |
| | V0012894 | | | 27.49 | N/A | 42.45 | N/A | 35.37 | 35.45 | Negative | V0012552 | | | 32.39 | N/A | N/A | N/A | 35.37 | 35.45 | Negative | true | True negative |
| | V0015894 | | | 27.65 | N/A | N/A | N/A | 36.42 | 34.78 | Negative | V0012485 | | | 34.40 | N/A | N/A | N/A | 36.42 | 34.78 | Negative | true | True negative |
| | V0015689 | | | 29.92 | N/A | N/A | N/A | 36.42 | 34.78 | Negative | V0012545 | | | 31.33 | N/A | N/A | N/A | 36.42 | 34.78 | Negative | true | True negative |
| | V0015939 | | | 29.46 | N/A | 39.31 | N/A | 36.42 | 34.78 | Negative | V0012551 | | | 38.14 | N/A | N/A | N/A | 36.42 | 34.78 | Negative | true | True negative |
| | V0015700 | | | 28.72 | 37.21 | N/A | N/A | 35.46 | 35.95 | Negative | V0012568 | | | 31.09 | N/A | N/A | N/A | 36.42 | 34.78 | Negative | true | True negative |
| | V0015931 | | | 29.33 | 37.19 | N/A | N/A | 35.46 | 35.95 | Negative | V0012488 | | | 34.00 | N/A | N/A | N/A | 36.10 | 36.36 | Negative | true | True negative |
| | V0015917 | | | 28.31 | N/A | N/A | N/A | 36.42 | 34.78 | Negative | V0012491 | | | 33.63 | N/A | N/A | N/A | 36.10 | 36.36 | Negative | true | True negative |

| | | | | | | | | | | | | | | | | | | | | | | |
|---|---|---|---|---|---|---|---|---|---|---|---|---|---|---|---|---|---|---|---|---|---|---|
| | V0015298 | | | 28.77 | 37.21 | 36.07 | N/A | 35.46 | 35.95 | Negative | V0012501 | | | 32.64 | N/A | N/A | N/A | 36.10 | 36.36 | Negative | true | True negative |
| | V0015687 | | | 32.34 | N/A | N/A | N/A | 35.46 | 35.95 | Negative | V0012572 | | | 30.97 | N/A | N/A | N/A | 36.10 | 36.36 | Negative | true | True negative |
| | V0010574 | | | 26.68 | 40.65 | N/A | 42.30 | 36.42 | 34.78 | Negative | V0012573 | | | 35.09 | N/A | N/A | N/A | 36.10 | 36.36 | Negative | true | True negative |
| | V0015982 | | | 29.10 | N/A | N/A | 41.40 | 36.20 | 36.35 | Negative | V0012576 | | | 33.84 | N/A | N/A | N/A | 36.10 | 36.36 | Negative | true | True negative |
| | V0016413 | | | 29.20 | N/A | N/A | 38.00 | 37.03 | 36.52 | Negative | V0016982 | | | 32.90 | N/A | N/A | N/A | 36.39 | 37.05 | Negative | true | True negative |
| | V0016407 | | | 29.90 | N/A | 37.10 | 37.70 | 37.03 | 36.52 | Negative | V0016984 | | | 35.00 | N/A | N/A | N/A | 36.39 | 37.05 | Negative | true | True negative |
| | V0016754 | | | 29.01 | N/A | N/A | 37.68 | 36.95 | 35.95 | Negative | V0016986 | | | 34.70 | N/A | N/A | N/A | 36.39 | 37.05 | Negative | true | True negative |
| | V0016764 | | | 29.07 | N/A | N/A | N/A | 36.39 | 37.05 | Negative | V0016991 | | | 32.10 | N/A | N/A | N/A | 36.39 | 37.05 | Negative | true | True negative |
| | V0015373 | | | 28.90 | N/A | N/A | N/A | 37.03 | 36.52 | Negative | V0017001 | | | 31.30 | N/A | N/A | N/A | 36.39 | 37.05 | Negative | true | True negative |
| | V0016400 | | | 29.30 | N/A | N/A | N/A | 37.03 | 36.52 | Negative | V0017011 | | | 30.10 | N/A | N/A | N/A | 36.39 | 37.05 | Negative | true | True negative |
| | V0015694 | | | 28.80 | N/A | N/A | N/A | 37.03 | 36.52 | Negative | V0017014 | | | 31.90 | N/A | N/A | N/A | 36.39 | 37.05 | Negative | true | True negative |
| | V0015820 | | | 28.82 | 36.22 | 36.51 | N/A | 34.85 | 35.12 | Negative | V0016792 | | | 34.04 | N/A | N/A | N/A | 37.78 | 36.02 | Negative | true | True negative |
| | V0015407 | | | 28.39 | N/A | N/A | N/A | 35.41 | 36.85 | Negative | V0016806 | | | 37.55 | N/A | N/A | N/A | 37.78 | 36.02 | Negative | true | True negative |
| | V0009668 | | | 30.19 | N/A | N/A | N/A | 37.03 | 36.11 | Negative | V0016820 | | | 33.40 | N/A | N/A | N/A | 37.78 | 36.02 | Negative | true | True negative |
| | V0015406 | | | 29.07 | N/A | N/A | 38.14 | 35.70 | 36.11 | Negative | V0016782 | | | 33.98 | 36.52 | 37.58 | 36.25 | 35.54 | 35.88 | Negative | true | True negative |
| | V0016864 | | | 29.26 | N/A | N/A | N/A | 35.91 | 36.21 | Negative | V0016987 | | | 32.20 | 45.00 | 44.60 | 45.00 | 36.46 | 36.01 | Negative | true | True negative |
| | V0015380 | | | 27.13 | N/A | N/A | N/A | 36.76 | 35.76 | Negative | V0016988 | | | 32.70 | 45.00 | 45.00 | 45.00 | 36.46 | 36.01 | Negative | true | true negative |
| | V0015378 | | | 29.19 | N/A | N/A | N/A | 37.51 | 36.06 | Negative | V0016993 | | | 34.80 | 45.00 | 45.00 | 45.00 | 36.46 | 36.01 | Negative | true | True negative |
| | V0017052 | | | 28.90 | N/A | N/A | 37.42 | 36.76 | 35.76 | Negative | V0016994 | | | 31.10 | 45.00 | 45.00 | 45.00 | 36.46 | 36.01 | Negative | true | True negative |
| | V0015374 | | | 29.14 | N/A | N/A | N/A | 36.76 | 35.76 | Negative | V0016998 | | | 32.50 | 45.00 | 45.00 | 45.00 | 36.46 | 36.01 | Negative | true | True negative |
| | V0016877 | | | 30.27 | N/A | N/A | N/A | 36.76 | 35.76 | Negative | V0017000 | | | 32.30 | 45.00 | 41.20 | 45.00 | 36.46 | 36.01 | Negative | true | True negative |
| | V0017193 | | | 28.28 | N/A | N/A | N/A | 36.76 | 35.76 | Negative | V0017009 | | | 32.40 | 45.00 | 45.00 | 45.00 | 36.46 | 36.01 | Negative | true | True negative |
| | V0017194 | | | 27.28 | N/A | N/A | 38.31 | 36.76 | 35.76 | Negative | V0017012 | | | 32.80 | 45.00 | 45.00 | 40.30 | 36.46 | 36.01 | Negative | true | True negative |
| | V0016791 | | | 28.05 | N/A | N/A | 44.88 | 36.76 | 35.76 | Negative | V0017017 | | | 33.00 | 45.00 | 45.00 | 45.00 | 36.46 | 36.01 | Negative | true | True negative |
| | V0017262 | | | 27.02 | N/A | N/A | 38.76 | 36.76 | 35.76 | Negative | V0017018 | | | 28.70 | 45.00 | 45.00 | 45.00 | 36.46 | 36.01 | Negative | true | True negative |
| | V0016803 | | | 27.13 | N/A | N/A | 38.01 | 36.76 | 35.76 | Negative | V0017021 | | | 33.70 | 45.00 | 45.00 | 45.00 | 36.46 | 36.01 | Negative | true | true negative |
| | V0017354 | | | 31.31 | N/A | N/A | N/A | 35.78 | 36.30 | Negative | V0015057 | | | 34.93 | N/A | N/A | N/A | 35.16 | 36.00 | Negative | true | True negative |
| | V0016985 | | | 33.43 | N/A | N/A | N/A | 35.78 | 36.30 | Negative | V0018455 | | | 34.42 | N/A | N/A | N/A | 35.67 | 36.11 | Negative | true | True negative |
| | V0018463 | | | 29.11 | 37.02 | 37.13 | 38.30 | 35.78 | 36.30 | Negative | V0018462 | | | 32.88 | N/A | N/A | N/A | 35.67 | 36.11 | Negative | true | True negative |
| | V0018465 | | | 30.14 | 37.06 | N/A | 38.03 | 35.78 | 36.30 | Negative | V0018439 | | | 34.08 | N/A | N/A | N/A | 35.67 | 36.11 | Negative | true | True negative |
| | V0018411 | | | 28.94 | N/A | N/A | N/A | 36.02 | 35.87 | Negative | V0019258 | | | 33.90 | N/A | N/A | N/A | 35.10 | 35.65 | Negative | true | True negative |
| | V0018364 | | | 27.85 | 36.92 | N/A | 38.24 | 36.13 | 35.17 | Negative | V0019262 | | | 34.00 | 36.70 | N/A | N/A | 35.10 | 35.65 | Negative | true | True negative |
| | V0015036 | | | 29.49 | N/A | N/A | N/A | 36.13 | 35.17 | Negative | V0019268 | | | 32.80 | N/A | N/A | N/A | 35.10 | 35.65 | Negative | true | True negative |
| | V0017794 | | | 27.27 | 23.65 | 24.36 | 24.57 | 34.93 | 36.37 | Positive | V0017791 | | | 34.40 | 34.80 | 35.00 | 34.90 | 34.82 | 36.02 | Positive | true | True positive |
| | V0017841 | | | 28.42 | 24.36 | 23.92 | 24.09 | 34.93 | 36.37 | Positive | V0017792 | | | 30.40 | 31.30 | 32.80 | 31.20 | 34.82 | 36.02 | Positive | true | True positive |
| | V0017807 | | | 28.94 | 18.49 | 19.18 | 19.46 | 34.93 | 36.37 | Positive | V0017798 | | | 33.80 | 23.70 | 24.20 | 23.00 | 34.82 | 36.02 | Positive | true | True positive |
| | V0017813 | | | 28.12 | 25.18 | 24.60 | 24.71 | 34.93 | 36.37 | Positive | V0017800 | | | 29.80 | 24.70 | 24.90 | 24.50 | 34.82 | 36.02 | Positive | true | True positive |
| | V0017831 | | | 28.62 | 16.86 | 18.08 | 18.51 | 34.93 | 36.37 | Positive | V0017804 | | | 29.30 | 27.20 | 27.90 | 27.00 | 34.82 | 36.02 | Positive | true | True positive |
| | V0017789 | | | 27.95 | 28.21 | 28.48 | 28.52 | 34.93 | 36.37 | Positive | V0017819 | | | 34.50 | 32.10 | 33.50 | 34.10 | 34.82 | 36.02 | Positive | true | True positive |
| | V0017796 | | | 28.63 | 18.81 | 18.84 | 18.91 | 34.93 | 36.37 | Positive | V0017825 | | | 33.10 | 25.70 | 26.90 | 26.90 | 34.82 | 36.02 | Positive | true | True positive |
| | V0017817 | | | 27.95 | 28.80 | 29.11 | 29.24 | 34.93 | 36.37 | Positive | V0017835 | | | 30.50 | 32.20 | 32.10 | 31.60 | 34.82 | 36.02 | Positive | true | True positive |
| | V0017821 | | | 28.15 | 24.05 | 24.25 | 24.43 | 34.93 | 36.37 | Positive | V0017836 | | | 33.80 | 30.20 | 30.40 | 30.20 | 34.82 | 36.02 | Positive | true | True positive |
| | V0017832 | | | 29.20 | 17.38 | 17.52 | 17.70 | 34.93 | 36.37 | Positive | V0017837 | | | 32.30 | 24.00 | 24.30 | 24.00 | 34.82 | 36.02 | Positive | true | True positive |
| | V0017801 | | | 25.49 | 16.73 | 16.39 | 15.97 | 34.93 | 36.37 | Positive | V0017839 | | | 34.10 | 31.70 | 31.50 | 31.90 | 34.82 | 36.02 | Positive | true | True positive |
| | V0017827 | | | 28.70 | 25.40 | 25.59 | 25.69 | 35.85 | 37.10 | Positive | V0017828 | | | 31.00 | 26.10 | 26.70 | 26.80 | 34.82 | 36.02 | Positive | true | True positive |
| | V0018461 | | | 29.18 | 30.68 | 32.15 | 32.93 | 35.78 | 36.30 | Positive | V0018451 | | | 32.44 | 26.27 | 25.99 | 25.69 | 35.67 | 36.11 | Positive | true | True positive |
| | V0018464 | | | 29.38 | 31.98 | 32.29 | 32.47 | 35.78 | 36.30 | Positive | V0018477 | | | 33.54 | 32.30 | 32.21 | 32.12 | 35.67 | 36.11 | Positive | true | True positive |
| | V0018374 | | | 29.62 | 13.39 | 14.94 | 15.31 | 36.02 | 35.87 | Positive | V0019280 | | | 32.50 | 27.90 | 28.90 | 29.30 | 35.10 | 35.65 | Positive | true | True positive |
| | V0017196 | | | 29.11 | 16.55 | 17.82 | 18.32 | 36.76 | 35.76 | Positive | V0016989 | | | 31.20 | 28.50 | 28.50 | 29.30 | 36.46 | 36.01 | Positive | true | True positive |
| | V0016771 | | | 26.50 | 23.10 | 19.20 | 19.10 | 37.03 | 36.52 | Positive | V0017016 | | | 33.00 | 34.30 | 29.10 | 29.00 | 36.39 | 37.05 | Positive | true | True positive |
| | V0015396 | | | 28.60 | 35.05 | 36.18 | 37.94 | 35.70 | 36.11 | Positive | V0016786 | | | 34.38 | 34.20 | 34.41 | 36.71 | 37.78 | 36.02 | Positive | true | True positive |
| | V0015398 | | | 26.60 | 14.19 | 15.77 | 15.94 | 36.90 | 35.71 | Positive | V0016811 | | | 33.70 | 34.00 | 35.15 | 34.85 | 37.78 | 36.02 | Positive | true | True positive |
| | V0007630 | | | 29.10 | 21.48 | 23.29 | 23.16 | 33.84 | 33.95 | Positive | V0007330 | | | 30.15 | 28.36 | 28.45 | 28.28 | 34.28 | 34.55 | Positive | true | True positive |
| | V0007197 | | | 28.49 | 23.06 | 24.23 | 24.17 | 34.92 | 34.18 | Positive | V0005904 | | | 35.78 | 31.98 | 31.96 | 32.17 | 34.75 | 34.35 | Positive | true | True positive |
| | V0008357 | | | 29.45 | 21.49 | 23.53 | 24.17 | 34.35 | 34.00 | Positive | V0003705 | | | 34.64 | 34.08 | 34.65 | 34.02 | 34.75 | 34.35 | Positive | true | True positive |
| | V0008371 | | | 29.29 | 17.10 | 19.07 | 19.20 | 34.89 | 34.49 | Positive | V0003697 | | | 35.26 | 27.79 | 27.85 | 27.71 | 34.75 | 34.35 | Positive | true | True positive |
| | V0010895 | | | 29.33 | 26.08 | 26.80 | 27.19 | 38.28 | 35.38 | Positive | V0012730 | | | 30.66 | 32.50 | 32.34 | 32.65 | 38.28 | 35.38 | Positive | true | True positive |
| | V0011736 | | | 28.71 | 34.73 | 34.87 | 35.54 | 34.78 | 36.70 | Positive | V0012535 | | | 31.80 | 30.11 | 30.51 | 30.59 | 35.17 | 35.76 | Positive | true | True positive |
| | V0011727 | | | 28.49 | 27.96 | 29.20 | 29.04 | 34.78 | 36.70 | Positive | V0012580 | | | 30.24 | 33.12 | 33.35 | 33.78 | 35.17 | 35.76 | Positive | true | True positive |
| | V0010757 | | | 28.60 | 16.49 | 17.23 | 17.09 | 36.43 | 36.11 | Positive | V0011478 | | | 34.59 | 30.42 | 31.01 | 31.38 | 36.43 | 36.11 | Positive | true | True positive |
| | V0015941 | | | 29.27 | 16.67 | 18.43 | 18.47 | 35.46 | 35.95 | Positive | V0012566 | | | 30.31 | 31.79 | 31.69 | 32.29 | 36.42 | 34.78 | Positive | true | True positive |
| | V0014312 | | | 27.97 | 14.82 | 16.26 | 16.66 | 35.46 | 35.95 | Positive | V0012571 | | | 32.21 | 26.46 | 27.46 | 27.35 | 36.10 | 36.36 | Positive | true | True positive |
| | V0012907 | | | 29.70 | 15.19 | 16.42 | 17.14 | 36.30 | 34.83 | Positive | V0012510 | | | 31.88 | 26.80 | 26.91 | 27.05 | 36.91 | 35.28 | Positive | true | True positive |
