## Supplementary material for "Development and Implementation of a scalable and versatile test for COVID-19 diagnostics in rural communities": Sup_Table VI

NCBI

| A | B | C | D | E | F | G | H | I | J | K |
|---|---|---|---|---|---|---|---|---|---|---|
| Database ID | NCBI Description | SK_N forward GCTGCAATCGTGCTACAACT | SK_N reverse TGAACTGTTGCGACTACGTG | SK_E forward TTCGGAAGAGACAGGTACGTT | SK_E reverse CACACAATCGAAGCGCAGTA | SK_S forward GCTGGTGCTGCAGCTTATTA | SK_S reverse AGGGTCAAGTGCACAGTCTA | 3' mismatches? | Multiple mismatches? | More than 1 primer set? |
| 1 | NC_045512.2 \|Severe acute respiratory syndrome cor | ···················· | ···················· | ···················· | ···················· | ···················· | ···················· | | | |
| 2 | MW466791.1 \|Severe acute respiratory syndrome coro | ···················· | ···················· | ···················· | ···················· | ···················· | ···················· | | | |
| 3 | MW466792.1 \|Severe acute respiratory syndrome coro | ···················· | ···················· | ···················· | ···················· | ···················· | ···················· | | | |
| 4 | MW466793.1 \|Severe acute respiratory syndrome coro | ···················· | ···················· | ···················· | ···················· | ···················· | ···················· | | | |
| 5 | MW466794.1 \|Severe acute respiratory syndrome coro | ···················· | ···················· | ···················· | ···················· | ···················· | ···················· | | | |
| 6 | MW466795.1 \|Severe acute respiratory syndrome coro | ···················· | ···················· | ···················· | ···················· | ···················· | ···················· | | | |
| 7 | MW466796.1 \|Severe acute respiratory syndrome coro | ···················· | ···················· | ···················· | ···················· | ···················· | ···················· | | | |
| 8 | MW466797.1 \|Severe acute respiratory syndrome coro | ···················· | ···················· | ···················· | ···················· | ···················· | ···················· | | | |
| 9 | MW466798.1 \|Severe acute respiratory syndrome coro | ···················· | ···················· | ···················· | ···················· | ···················· | ···················· | | | |
| 10 | MW466799.1 \|Severe acute respiratory syndrome coro | ···················· | ···················· | ···················· | ···················· | ···················· | ···················· | | | |
| 11 | MW466800.1 \|Severe acute respiratory syndrome coro | ···················· | ···················· | ···················· | ···················· | ···················· | ···················· | | | |
| 12 | MW467418.1 \|Severe acute respiratory syndrome coro | ···················· | ···················· | ···················· | ···················· | ···················· | ···················· | | | |
| 13 | MW467419.1 \|Severe acute respiratory syndrome coro | ···················· | ·A·················· | ···················· | ···················· | ···················· | ···················· | | | |
| 14 | MW467420.1 \|Severe acute respiratory syndrome coro | ···················· | ···················· | ···················· | ···················· | ···················· | ···················· | | | |
| 15 | MW467421.1 \|Severe acute respiratory syndrome coro | ···················· | ···················· | ···················· | ···················· | ···················· | ···················· | | | |
| 16 | MW467422.1 \|Severe acute respiratory syndrome coro | ···················· | ···················· | ···················· | ···················· | ···················· | ···················· | | | |
| 17 | MW467423.1 \|Severe acute respiratory syndrome coro | ···················· | ···················· | ···················· | ···················· | ···················· | ···················· | | | |
| 18 | MW467424.1 \|Severe acute respiratory syndrome coro | ···················· | ······A············· | ···················· | ···················· | ···················· | ···················· | | | |
| 19 | MW467425.1 \|Severe acute respiratory syndrome coro | ···················· | ······A············· | ···················· | ···················· | ···················· | ···················· | | | |
| 20 | MW467426.1 \|Severe acute respiratory syndrome coro | ···················· | ···················· | ···················· | ···················· | ···················· | ···················· | | | |
| 21 | MW467427.1 \|Severe acute respiratory syndrome coro | ···················· | ···················· | ···················· | ···················· | ···················· | ···················· | | | |
| 22 | MW467428.1 \|Severe acute respiratory syndrome coro | ···················· | ···················· | ···················· | ···················· | ···················· | ···················· | | | |
| 23 | MW467429.1 \|Severe acute respiratory syndrome coro | ···················· | ···················· | ···················· | ···················· | ···················· | ···················· | | | |
| 24 | MW467430.1 \|Severe acute respiratory syndrome coro | ···················· | ···················· | ···················· | ···················· | ···················· | ···················· | | | |
| 25 | MW467431.1 \|Severe acute respiratory syndrome coro | ···················· | ·········A·········· | ···················· | ···················· | ···················· | ···················· | | | |
| 26 | MW467432.1 \|Severe acute respiratory syndrome coro | ···················· | ···················· | ···················· | ···················· | ···················· | ···················· | | | |
| 27 | MW467433.1 \|Severe acute respiratory syndrome coro | ···················· | ···················· | ···················· | ···················· | ···················· | ···················· | | | |
| 28 | MW467434.1 \|Severe acute respiratory syndrome coro | ···················· | ···················· | ···················· | ···················· | ···················· | ···················· | | | |
| 29 | MW467435.1 \|Severe acute respiratory syndrome coro | ···················· | ···················· | ···················· | ···················· | ···················· | ···················· | | | |
| 30 | MW467436.1 \|Severe acute respiratory syndrome coro | ···················· | ···················· | ···················· | ···················· | ···················· | ···················· | | | |
| 31 | MW467437.1 \|Severe acute respiratory syndrome coro | ···················· | ···················· | ···················· | ···················· | ···················· | ···················· | | | |
| 32 | MW467438.1 \|Severe acute respiratory syndrome coro | ···················· | ···················· | ···················· | ···················· | ···················· | ···················· | | | |
| 33 | MW467439.1 \|Severe acute respiratory syndrome coro | ···················· | ···················· | ···················· | ···················· | ···················· | ···················· | | | |
| 34 | MW467440.1 \|Severe acute respiratory syndrome coro | ···················· | ···················· | ···················· | ···················· | ···················· | ···················· | | | |
| 35 | MW467441.1 \|Severe acute respiratory syndrome coro | ···················· | ···················· | ···················· | ···················· | ···················· | ···················· | | | |
| 36 | MW467442.1 \|Severe acute respiratory syndrome coro | ···················· | ···················· | ···················· | ···················· | ···················· | ···················· | | | |
| 37 | MW467443.1 \|Severe acute respiratory syndrome coro | ···················· | ···················· | ···················· | ···················· | ···················· | ···················· | | | |
| 38 | MW467444.1 \|Severe acute respiratory syndrome coro | ···················· | ···················· | ···················· | ···················· | ···················· | ···················· | | | |
| 39 | MW467445.1 \|Severe acute respiratory syndrome coro | ···················· | ···················· | ···················· | ···················· | ···················· | ···················· | | | |
| 40 | MW467446.1 \|Severe acute respiratory syndrome coro | ···················· | ···················· | ···················· | ···················· | ···················· | ···················· | | | |
| 41 | MW467447.1 \|Severe acute respiratory syndrome coro | ···················· | ···················· | ···················· | ···················· | ···················· | ···················· | | | |
| 42 | MW467448.1 \|Severe acute respiratory syndrome coro | ···················· | ···················· | ···················· | ···················· | ···················· | ···················· | | | |
| 43 | MW467449.1 \|Severe acute respiratory syndrome coro | ···················· | ···················· | ···················· | ···················· | ···················· | ···················· | | | |
| 44 | MW467450.1 \|Severe acute respiratory syndrome coro | ···················· | ···················· | ···················· | ···················· | ···················· | ···················· | | | |
| 45 | MW467451.1 \|Severe acute respiratory syndrome coro | ···················· | ·········A·········· | ···················· | ···················· | ···················· | ···················· | | | |
| 46 | MW467452.1 \|Severe acute respiratory syndrome coro | ···················· | ···················· | ···················· | ···················· | ···················· | ···················· | | | |
| 47 | MW467453.1 \|Severe acute respiratory syndrome coro | ···················· | ···················· | ···················· | ···················· | ···················· | ···················· | | | |
| 48 | MW467454.1 \|Severe acute respiratory syndrome coro | ···················· | ·A·················· | ···················· | ···················· | ···················· | ···················· | | | |
| 49 | MW467455.1 \|Severe acute respiratory syndrome coro | ···················· | ···················· | ···················· | ···················· | ···················· | ···················· | | | |
| 50 | MW467456.1 \|Severe acute respiratory syndrome coro | ···················· | ···················· | ···················· | ···················· | ···················· | ···················· | | | |
| 51 | MW467457.1 \|Severe acute respiratory syndrome coro | ···················· | ·········A·········· | ···················· | ···················· | ···················· | ···················· | | | |
| 52 | MW467458.1 \|Severe acute respiratory syndrome coro | ···················· | ···················· | ···················· | ···················· | ···················· | ···················· | | | |
| 53 | MW467459.1 \|Severe acute respiratory syndrome coro | ···················· | ·········A·········· | ···················· | ···················· | ···················· | ···················· | | | |
| 54 | MW467460.1 \|Severe acute respiratory syndrome coro | ···················· | ···················· | ···················· | ···················· | ···················· | ···················· | | | |
| 55 | MW467461.1 \|Severe acute respiratory syndrome coro | ···················· | ···················· | ···················· | ···················· | ···················· | ···················· | | | |
| 56 | MW467462.1 \|Severe acute respiratory syndrome coro | ···················· | ···················· | ···················· | ···················· | ···················· | ···················· | | | |
| 57 | MW467463.1 \|Severe acute respiratory syndrome coro | ···················· | ···················· | ···················· | ···················· | ···················· | ···················· | | | |
| 58 | MW467464.1 \|Severe acute respiratory syndrome coro | ···················· | ···················· | ···················· | ···················· | ···················· | ···················· | | | |
| 59 | MW467465.1 \|Severe acute respiratory syndrome coro | ···················· | ·········A·········· | ···················· | ···················· | ···················· | ···················· | | | |
| 60 | MW467466.1 \|Severe acute respiratory syndrome coro | ···················· | ···················· | ···················· | ···················· | ···················· | ···················· | | | |
| 61 | MW467467.1 \|Severe acute respiratory syndrome coro | ···················· | ···················· | ···················· | ··················C· | ···················· | ···················· | 1 | | |
| 62 | MW467468.1 \|Severe acute respiratory syndrome coro | ···················· | ···················· | ···················· | ···················· | ···················· | ···A················ | | | |
| 63 | MW467469.1 \|Severe acute respiratory syndrome coro | ···················· | ···················· | ···················· | ···················· | ···················· | ···················· | | | |
| 64 | MW467470.1 \|Severe acute respiratory syndrome coro | ···················· | ···················· | ···················· | ···················· | ···················· | ···················· | | | |
| 65 | MW467471.1 \|Severe acute respiratory syndrome coro | ···················· | ···················· | ···················· | ···················· | ···················· | ···················· | | | |
| 66 | MW467472.1 \|Severe acute respiratory syndrome coro | ···················· | ···················· | ···················· | ···················· | ···················· | ···················· | | | |
| 67 | MW467473.1 \|Severe acute respiratory syndrome coro | ···················· | ···················· | ···················· | ···················· | ···················· | ···················· | | | |

| | | | | | | | | | |
|---|---|---|---|---|---|---|---|---|---|
| 68 | MW467474.1 | \|Severe acute respiratory syndrome coro | .................... | .................... | .................... | .................... | .................... | .................... | |
| 69 | MW467475.1 | \|Severe acute respiratory syndrome coro | .................... | .................... | .................... | .................... | .................... | .................... | |
| 70 | MW467476.1 | \|Severe acute respiratory syndrome coro | .................... | .................... | .................... | .................... | .................... | .................... | |
| 71 | MW467477.1 | \|Severe acute respiratory syndrome coro | .................... | .................... | .................... | .................... | .................... | .................... | |
| 72 | MW467478.1 | \|Severe acute respiratory syndrome coro | .................... | ......A............. | .................... | .................... | .................... | .................... | |
| 73 | MW467479.1 | \|Severe acute respiratory syndrome coro | .................... | .................... | .................... | .................... | .................... | .................... | |
| 74 | MW467480.1 | \|Severe acute respiratory syndrome coro | .................... | .................... | .................... | .................... | .................... | .................... | |
| 75 | MW467481.1 | \|Severe acute respiratory syndrome coro | .................... | .................... | .................... | .................... | .................... | .................... | |
| 76 | MW467482.1 | \|Severe acute respiratory syndrome coro | .................... | .................... | .................... | .................... | .................... | .................... | |
| 77 | MW467483.1 | \|Severe acute respiratory syndrome coro | .................... | ........A........... | .................... | .................... | .................... | .................... | |
| 78 | MW467484.1 | \|Severe acute respiratory syndrome coro | .................... | ......A............. | .................... | .................... | .................... | .................... | |
| 79 | MW467485.1 | \|Severe acute respiratory syndrome coro | .................... | ......A............. | .................... | .................... | .................... | .................... | |
| 80 | MW467486.1 | \|Severe acute respiratory syndrome coro | .................... | .A.................. | .................... | .................... | .................... | ..A................. | |
| 81 | MW467487.1 | \|Severe acute respiratory syndrome coro | .................... | .................... | .................... | .................... | .................... | .................... | |
| 82 | MW467488.1 | \|Severe acute respiratory syndrome coro | .................... | .................... | .................... | .................... | .................... | .................... | |
| 83 | MW467489.1 | \|Severe acute respiratory syndrome coro | .................... | .A.................. | .................... | .................... | .................... | ..A................. | |
| 84 | MW467490.1 | \|Severe acute respiratory syndrome coro | .................... | .A.................. | .................... | .................... | ......A............. | .................... | |
| 85 | MW467491.1 | \|Severe acute respiratory syndrome coro | .................... | ......A............. | .................... | .................... | .................... | .................... | |
| 86 | MW467492.1 | \|Severe acute respiratory syndrome coro | .................... | .................... | .................... | .................... | .................... | .................... | |
| 87 | MW467493.1 | \|Severe acute respiratory syndrome coro | .................... | .................... | .................... | .................... | .................... | .................... | |
| 88 | MW467494.1 | \|Severe acute respiratory syndrome coro | .................... | .................... | .................... | .................... | .................... | .................... | |
| 89 | MW467495.1 | \|Severe acute respiratory syndrome coro | .................... | .................... | .................... | .................... | .................... | .................... | |
| 90 | MW467496.1 | \|Severe acute respiratory syndrome coro | .................... | .................... | .................... | .................... | .................... | .................... | |
| 91 | MW467497.1 | \|Severe acute respiratory syndrome coro | .................... | .................... | .................... | .................... | ...A................ | .................... | |
| 92 | MW467498.1 | \|Severe acute respiratory syndrome coro | .................... | .................... | .................... | .................... | .................... | .................... | |
| 93 | MW467499.1 | \|Severe acute respiratory syndrome coro | .................... | .................... | .................... | .................... | .................... | .................... | |
| 94 | MW467500.1 | \|Severe acute respiratory syndrome coro | .................... | .................... | .................... | .................... | .................... | .................... | |
| 95 | MW467501.1 | \|Severe acute respiratory syndrome coro | .................... | ........A........... | .................... | .................... | .................... | .................G.. | 1 |
| 96 | MW467502.1 | \|Severe acute respiratory syndrome coro | .................... | .................... | .................... | .................... | .................... | .................... | |
| 97 | MW467503.1 | \|Severe acute respiratory syndrome coro | .................... | .................... | .................... | .................... | .................... | .................... | |
| 98 | MW467504.1 | \|Severe acute respiratory syndrome coro | .................... | .................... | .................... | .................... | .................... | .................... | |
| 99 | MW467505.1 | \|Severe acute respiratory syndrome coro | .................... | .................... | .................... | .................... | .................... | .................... | |
| 100 | MW467506.1 | \|Severe acute respiratory syndrome coro | .................... | .................... | .................... | .................... | .................... | .................... | |
| 101 | MW467507.1 | \|Severe acute respiratory syndrome coro | .................... | ......A............. | .................... | .................... | .................... | .................... | |
| 102 | MW468415.1 | \|Severe acute respiratory syndrome coro | .................... | .................... | .................... | .................... | .................... | .................... | |
| 103 | MW471388.1 | \|Severe acute respiratory syndrome coro | .................... | .................... | .................... | .................... | .................... | .................... | |
| 104 | MW471389.1 | \|Severe acute respiratory syndrome coro | .................... | .................... | .................... | .................... | .................... | .................... | |
| 105 | MW471390.1 | \|Severe acute respiratory syndrome coro | .................... | .................... | .................... | .................... | .................... | .................... | |
| 106 | MW471391.1 | \|Severe acute respiratory syndrome coro | .................... | .................... | .................... | .................... | .................... | .................... | |
| 107 | MW471392.1 | \|Severe acute respiratory syndrome coro | .................... | .................... | .................... | .................... | .................... | .................... | |
| 108 | MW471393.1 | \|Severe acute respiratory syndrome coro | .................... | .................... | .................... | .................... | .................... | .................... | |
| 109 | MW471398.1 | \|Severe acute respiratory syndrome coro | .................... | .................... | .................... | .................... | .................... | .................... | |
| 110 | MW471399.1 | \|Severe acute respiratory syndrome coro | .................... | .................... | .................... | .................... | .................... | .................... | |
| 111 | MW471400.1 | \|Severe acute respiratory syndrome coro | .................... | .................... | .................... | .................... | .................... | .................... | |
| 112 | MW471401.1 | \|Severe acute respiratory syndrome coro | .................... | .................... | .................... | .................... | .................... | .................... | |
| 113 | MW473685.1 | \|Severe acute respiratory syndrome coro | .................... | .................... | .................... | .................... | .................... | .................... | |
| 114 | MW473686.1 | \|Severe acute respiratory syndrome coro | .................... | .................... | .................... | .................... | .................... | .................... | |
| 115 | MW473687.1 | \|Severe acute respiratory syndrome coro | .................... | .................... | .................... | .................... | .................... | .................... | |
| 116 | MW473688.1 | \|Severe acute respiratory syndrome coro | .................... | .................... | .................... | .................... | .................... | .................... | |
| 117 | MW473689.1 | \|Severe acute respiratory syndrome coro | .................... | .................... | .................... | .................... | .................... | .................... | |
| 118 | MW473690.1 | \|Severe acute respiratory syndrome coro | .................... | .................... | .................... | .................... | .................... | .................... | |
| 119 | MW473692.1 | \|Severe acute respiratory syndrome coro | .................... | .................... | .................... | .................... | .................... | .................... | |
| 120 | MW473694.1 | \|Severe acute respiratory syndrome coro | .................... | .................... | .................... | .................... | .................... | .................... | |
| 121 | MW473696.1 | \|Severe acute respiratory syndrome coro | .................... | .................... | .................... | .................... | .................... | .................... | |
| 122 | MW473697.1 | \|Severe acute respiratory syndrome coro | .................... | .................... | .................... | .................... | .................... | .................... | |
| 123 | MW473699.1 | \|Severe acute respiratory syndrome coro | .................... | .................... | .................... | .................... | .................... | .................... | |
| 124 | MW473700.1 | \|Severe acute respiratory syndrome coro | .................... | .................... | .................... | .................... | .................... | .................... | |
| 125 | MW474104.1 | \|Severe acute respiratory syndrome coro | .................... | .................... | .................... | .................... | .................... | .................... | |
| 126 | MW474105.1 | \|Severe acute respiratory syndrome coro | .................... | .A.................. | .................... | .................... | .................... | .................... | |
| 127 | MW474106.1 | \|Severe acute respiratory syndrome coro | .................... | .................... | .................... | .................... | .................... | .................... | |
| 128 | MW474107.1 | \|Severe acute respiratory syndrome coro | .................... | .................... | .................... | .................... | .................... | .................... | |
| 129 | MW474108.1 | \|Severe acute respiratory syndrome coro | .................... | .................... | .................... | .................... | .................... | .................... | |
| 130 | MW474109.1 | \|Severe acute respiratory syndrome coro | .................... | .A.................. | .................... | .................... | .................... | .................... | |
| 131 | MW474110.1 | \|Severe acute respiratory syndrome coro | .................... | .................... | .................... | .................... | .................... | .................... | |
| 132 | MW474111.1 | \|Severe acute respiratory syndrome coro | .................... | .................... | .................... | .................... | .................... | .................... | |
| 133 | MW474112.1 | \|Severe acute respiratory syndrome coro | .................... | .................... | .................... | .................... | .................... | .................... | |
| 134 | MW474113.1 | \|Severe acute respiratory syndrome coro | .................... | .A.................. | .................... | .................... | .................... | .................... | |
| 135 | MW474114.1 | \|Severe acute respiratory syndrome coro | .................... | .................... | .................... | .................... | .................... | .................... | |
| 136 | MW474115.1 | \|Severe acute respiratory syndrome coro | .................... | .................... | .................... | .................... | .................... | .................... | |
| 137 | MW474116.1 | \|Severe acute respiratory syndrome coro | .................... | .................... | .................... | .................... | .................... | .................... | |
| 138 | MW474117.1 | \|Severe acute respiratory syndrome coro | .................... | .................... | .................... | .................... | .................... | .................... | |
| 139 | MW474118.1 | \|Severe acute respiratory syndrome coro | .................... | .................... | .................... | .................... | .................... | .................... | |
| 140 | MW474119.1 | \|Severe acute respiratory syndrome coro | .................... | .A.................. | .................... | .................... | .................... | .................... | |
| 141 | MW474120.1 | \|Severe acute respiratory syndrome coro | .................... | .A.................. | .................... | .................... | .................... | .................... | |
| 142 | MW474121.1 | \|Severe acute respiratory syndrome coro | .................... | .................... | .................... | .................... | .................... | .................... | |
| 143 | MW474122.1 | \|Severe acute respiratory syndrome coro | .................... | .................... | .................... | .................... | .................... | .................... | |
| 144 | MW474123.1 | \|Severe acute respiratory syndrome coro | .................... | .................... | .................... | .................... | .................... | .................... | |
| 145 | MW474124.1 | \|Severe acute respiratory syndrome coro | .................... | .................... | .................... | .................... | .................... | .................... | |

146 MW474125.1 |Severe acute respiratory syndrome coro .................... .................... .................... .................... .................... ....................
147 MW474126.1 |Severe acute respiratory syndrome coro .................... .................... .................... .................... .................... ....................
148 MW474127.1 |Severe acute respiratory syndrome coro .................... .................... .................... .................... .................... ....................
149 MW474128.1 |Severe acute respiratory syndrome coro .................... .................... .................... .................... .................... ....................
150 MW474129.1 |Severe acute respiratory syndrome coro .................... .................... .................... .................... .................... ....................
151 MW474130.1 |Severe acute respiratory syndrome coro .................... .................... .................... .................... .................... ....................
152 MW474131.1 |Severe acute respiratory syndrome coro .................... .................... .................... .................... .................... ....................
153 MW474132.1 |Severe acute respiratory syndrome coro .................... .................... .................... .................... .................... ....................
154 MW474133.1 |Severe acute respiratory syndrome coro .................... .................... .................... .................... .................... ....................
155 MW474134.1 |Severe acute respiratory syndrome coro .................... .................... .................... .................... .................... ....................
156 MW474135.1 |Severe acute respiratory syndrome coro .................... .................... .................... .................... .................... ....................
157 MW474136.1 |Severe acute respiratory syndrome coro .................... .................... .................... .................... .................... ....................
158 MW474137.1 |Severe acute respiratory syndrome coro .................... .................... .................... .................... .................... ....................
159 MW474138.1 |Severe acute respiratory syndrome coro .................... .................... .................... .................... .................... ....................
160 MW474139.1 |Severe acute respiratory syndrome coro .................... .................... .................... .................... .................... ....................
161 MW474140.1 |Severe acute respiratory syndrome coro .................... .................... .................... .................... .................... ....................
162 MW474141.1 |Severe acute respiratory syndrome coro .................... .................... .................... .................... .................... ....................
163 MW474142.1 |Severe acute respiratory syndrome coro .................... .................... .................... .................... .................... ....................
164 MW474143.1 |Severe acute respiratory syndrome coro .................... .................... .................... .................... .................... ....................
165 MW474144.1 |Severe acute respiratory syndrome coro .................... .................... .................... .................... .................... ....................
166 MW474145.1 |Severe acute respiratory syndrome coro .................... .................... .................... .................... .................... ....................
167 MW474146.1 |Severe acute respiratory syndrome coro .................... .A.................. .................... .................... .................... ....................
168 MW474147.1 |Severe acute respiratory syndrome coro .................... .A.................. .................... .................... .................... ....................
169 MW474148.1 |Severe acute respiratory syndrome coro .................... .................... .................... .................... .................... ....................
170 MW474149.1 |Severe acute respiratory syndrome coro .................... .................... .................... .................... .................... ....................
171 MW474150.1 |Severe acute respiratory syndrome coro .................... .................... .................... .................... .................... ....................
172 MW474151.1 |Severe acute respiratory syndrome coro .................... .................... .................... .................... .................... ....................
173 MW474152.1 |Severe acute respiratory syndrome coro .................... .................... .................... .................... .................... ....................
174 MW474153.1 |Severe acute respiratory syndrome coro .................... .................... .................... .................... .................... ....................
175 MW474154.1 |Severe acute respiratory syndrome coro .................... .................... .................... .................... .................... ....................
176 MW474155.1 |Severe acute respiratory syndrome coro .................... .................... .................... .................... .................... ....................
177 MW474156.1 |Severe acute respiratory syndrome coro .................... .................... .................... .................... .................... ....................
178 MW474157.1 |Severe acute respiratory syndrome coro .................... .A.................. .................... .................... .................... ....................
179 MW474158.1 |Severe acute respiratory syndrome coro .................... .................... .................... .................... .................... ....................
180 MW474159.1 |Severe acute respiratory syndrome coro .................... .................... .................... .................... .................... ....................
181 MW474160.1 |Severe acute respiratory syndrome coro .................... .A.................. .................... .................... .................... ....................
182 MW474161.1 |Severe acute respiratory syndrome coro .................... .A.................. .................... .................... .................... ....................
183 MW474162.1 |Severe acute respiratory syndrome coro .................... .................... .................... .................... .................... ....................
184 MW474163.1 |Severe acute respiratory syndrome coro .................... .................... .................... .................... .................... ....................
185 MW474164.1 |Severe acute respiratory syndrome coro .................... .................... .................... .................... .................... ....................
186 MW474165.1 |Severe acute respiratory syndrome coro .................... .................... .................... .................... .................... ....................
187 MW474166.1 |Severe acute respiratory syndrome coro .................... .................... ..A................. .................... .................... ....................
188 MW474167.1 |Severe acute respiratory syndrome coro .................... .................... .................... .................... .................... ....................
189 MW474168.1 |Severe acute respiratory syndrome coro .................... .................... .................... .................... .................... ....................
190 MW474169.1 |Severe acute respiratory syndrome coro .................... .................... .................... .................... .................... ....................
191 MW474170.1 |Severe acute respiratory syndrome coro .................... .................... .................... .................... .................... ....................
192 MW474171.1 |Severe acute respiratory syndrome coro .................... .................... .................... .................... .................... ....................
193 MW474173.1 |Severe acute respiratory syndrome coro .................... .A.................. .................... .................... .................... ....................
194 MW474175.1 |Severe acute respiratory syndrome coro .................... .A.................. .................... .................... .................... ....................
195 MW474176.1 |Severe acute respiratory syndrome coro .................... .................... .................... .................... .................... ....................
196 MW474177.1 |Severe acute respiratory syndrome coro .................... .................... .................... .................... .................... ....................
197 MW474178.1 |Severe acute respiratory syndrome coro .................... .................... .................... .................... .................... ....................
198 MW474179.1 |Severe acute respiratory syndrome coro .................... .A.................. .................... .................... .................... ....................
199 MW474180.1 |Severe acute respiratory syndrome coro .................... .................... .................... .................... .................... ....................
200 MW474181.1 |Severe acute respiratory syndrome coro .................... .................... .................... .................... .................... ....................
201 MW474182.1 |Severe acute respiratory syndrome coro .................... .................... .................... .................... .................... ....................
202 MW474183.1 |Severe acute respiratory syndrome coro .................... .................... .................... .................... .................... ....................
203 MW474184.1 |Severe acute respiratory syndrome coro .................... .................... .................... .................... .................... ....................
204 MW474185.1 |Severe acute respiratory syndrome coro .................... .................... .................... .................... .................... ....................
205 MW474186.1 |Severe acute respiratory syndrome coro .................... .................... .................... .................... .................... ....................
206 MW474187.1 |Severe acute respiratory syndrome coro .................... .................... .................... .................... .................... ....................
207 MW474188.1 |Severe acute respiratory syndrome coro .................... .A.................. .................... .................... .................... ....................
208 MW474189.1 |Severe acute respiratory syndrome coro .................... .................... ..A................. .................... .................... ....................
209 MW474190.1 |Severe acute respiratory syndrome coro .................... .................... .................... .................... .................... ....................
210 MW474191.1 |Severe acute respiratory syndrome coro .................... .................... .................... .................... .................... ....................
211 MW474192.1 |Severe acute respiratory syndrome coro .................... .A.................. .................... .................... .................... ....................
212 MW474193.1 |Severe acute respiratory syndrome coro .................... .................... .................... .................... .................... ....................
213 MW474194.1 |Severe acute respiratory syndrome coro .................... .................... .................... .................... .................... ....................
214 MW474195.1 |Severe acute respiratory syndrome coro .................... .................... .................... .................... .................... ....................
215 MW474196.1 |Severe acute respiratory syndrome coro .................... .A.................. .................... .................... .................... ....................
216 MW474197.1 |Severe acute respiratory syndrome coro .................... .................... .................... .................... .................... ....................
217 MW474198.1 |Severe acute respiratory syndrome coro .................... .................... .................... .................... .................... ....................
218 MW474199.1 |Severe acute respiratory syndrome coro .................... .................... .................... .................... .................... ....................
219 MW474200.1 |Severe acute respiratory syndrome coro .................... .................... .................... .................... .................... ....................
220 MW474201.1 |Severe acute respiratory syndrome coro .................... .................... .................... .................... .................... ....................
221 MW474202.1 |Severe acute respiratory syndrome coro .................... .................... .................... .................... .................... ....................
222 MW474203.1 |Severe acute respiratory syndrome coro .................... .................... .................... .................... .................... ....................
223 MW474204.1 |Severe acute respiratory syndrome coro .................... .................... .................... .................... .................... ....................

```
224 MW474205.1 |Severe acute respiratory syndrome coro    ····················  ····················  ····················  ····················  ····················  ····················
225 MW474206.1 |Severe acute respiratory syndrome coro    ····················  ····················  ····················  ····················  ····················  ····················
226 MW474207.1 |Severe acute respiratory syndrome coro    ····················  ····················  ····················  ····················  ····················  ····················
227 MW474209.1 |Severe acute respiratory syndrome coro    ····················  ·A··················  ····················  ····················  ····················  ····················
228 MW474210.1 |Severe acute respiratory syndrome coro    ····················  ····················  ····················  ····················  ····················  ····················
229 MW474211.1 |Severe acute respiratory syndrome coro    ····················  ····················  ····················  ····················  ····················  ····················
230 MW474212.1 |Severe acute respiratory syndrome coro    ····················  ····················  ····················  ····················  ····················  ····················
231 MW474213.1 |Severe acute respiratory syndrome coro    ····················  ····················  ····················  ····················  ····················  ····················
232 MW474214.1 |Severe acute respiratory syndrome coro    ····················  ····················  ····················  ····················  ····················  ····················
233 MW474215.1 |Severe acute respiratory syndrome coro    ····················  ····················  ····················  ····················  ····················  ····················
234 MW474216.1 |Severe acute respiratory syndrome coro    ····················  ····················  ····················  ····················  ····················  ····················
235 MW474217.1 |Severe acute respiratory syndrome coro    ····················  ····················  ····················  ····················  ····················  ····················
236 MW474219.1 |Severe acute respiratory syndrome coro    ····················  ····················  ····················  ····················  ····················  ····················
237 MW474220.1 |Severe acute respiratory syndrome coro    ····················  ····················  ····················  ····················  ····················  ····················
238 MW474222.1 |Severe acute respiratory syndrome coro    ····················  ····················  ····················  ····················  ····················  ····················
239 MW474223.1 |Severe acute respiratory syndrome coro    ····················  ····················  ····················  ····················  ····················  ····················
240 MW474224.1 |Severe acute respiratory syndrome coro    ····················  ····················  ····················  ····················  ····················  ····················
241 MW474225.1 |Severe acute respiratory syndrome coro    ····················  ····················  ····················  ····················  ····················  ····················
242 MW474226.1 |Severe acute respiratory syndrome coro    ····················  ····················  ····················  ····················  ····················  ····················
243 MW474227.1 |Severe acute respiratory syndrome coro    ····················  ····················  ····················  ····················  ····················  ····················
244 MW474228.1 |Severe acute respiratory syndrome coro    ····················  ····················  ····················  ····················  ····················  ····················
245 MW474247.1 |Severe acute respiratory syndrome coro    ····················  ····················  ····················  ····················  ····················  ····················
246 MW474249.1 |Severe acute respiratory syndrome coro    ····················  ·A··················  ····················  ····················  ····················  ····················
247 MW474278.1 |Severe acute respiratory syndrome coro    ····················  ····················  ····················  ····················  ····················  ····················
248 MW474304.1 |Severe acute respiratory syndrome coro    ····················  ····················  ····················  ····················  ····················  ····················
249 MW474307.1 |Severe acute respiratory syndrome coro    ····················  ·A··················  ····················  ····················  ····················  ····················
250 MW474328.1 |Severe acute respiratory syndrome coro    ····················  ····················  ····················  ····················  ····················  ····················
251 MW474332.1 |Severe acute respiratory syndrome coro    ····················  ····················  ····················  ····················  ····················  ····················
252 MW474335.1 |Severe acute respiratory syndrome coro    ····················  ····················  ····················  ····················  ····················  ····················
253 MW474339.1 |Severe acute respiratory syndrome coro    ····················  ····················  ····················  ····················  ····················  ····················
254 MW474341.1 |Severe acute respiratory syndrome coro    ····················  ····················  ····················  ····················  ····················  ····················
255 MW474346.1 |Severe acute respiratory syndrome coro    ····················  ····················  ····················  ····················  ····················  ····················
256 MW474350.1 |Severe acute respiratory syndrome coro    ····················  ·A··················  ····················  ····················  ····················  ····················
257 MW474354.1 |Severe acute respiratory syndrome coro    ····················  ····················  ····················  ····················  ····················  ····················
258 MW474367.1 |Severe acute respiratory syndrome coro    ····················  ····················  ····················  ····················  ····················  ····················
259 MW474739.1 |Severe acute respiratory syndrome coro    ····················  ····················  ····················  ····················  ····················  ····················
260 MW474741.1 |Severe acute respiratory syndrome coro    ····················  ····················  ····················  ····················  ····················  ····················
261 MW474742.1 |Severe acute respiratory syndrome coro    ····················  ····················  ····················  ····················  ····················  ····················
262 MW463452.1 |Severe acute respiratory syndrome coro    ····················  ····················  ····················  ····················  ····················  ····················
263 MW463453.1 |Severe acute respiratory syndrome coro    ····················  ····················  ····················  ····················  ····················  ····················
264 MW463456.1 |Severe acute respiratory syndrome coro    ····················  ····················  ····················  ····················  ····················  ····················
265 MW463458.1 |Severe acute respiratory syndrome coro    ····················  ····················  ····················  ····················  ····················  ····················
266 MW466639.1 |Severe acute respiratory syndrome coro    ····················  ····················  ····················  ····················  ····················  ····················
267 MW454126.1 |Severe acute respiratory syndrome coro    ····················  ····················  ····················  ····················  ····················  ····················
268 MW454127.1 |Severe acute respiratory syndrome coro    ····················  ····················  ····················  ····················  ··A·················  ····················
269 MW454130.1 |Severe acute respiratory syndrome coro    ····················  ····················  ····················  ····················  ····················  ····················
270 MW454390.1 |Severe acute respiratory syndrome coro    ····················  ····················  ····················  ····················  ····················  ····················
271 MW454391.1 |Severe acute respiratory syndrome coro    ····················  ····················  ····················  ····················  ····················  ····················
272 MW454392.1 |Severe acute respiratory syndrome coro    ····················  ····················  ····················  ····················  ····················  ····················
273 MW454393.1 |Severe acute respiratory syndrome coro    ····················  ····················  ····················  ····················  ····················  ····················
274 MW454394.1 |Severe acute respiratory syndrome coro    ····················  ····················  ····················  ····················  ····················  ····················
275 MW454395.1 |Severe acute respiratory syndrome coro    ····················  ····················  ····················  ····················  ····················  ····················
276 MW454396.1 |Severe acute respiratory syndrome coro    ····················  ····················  ····················  ····················  ····················  ····················
277 MW454397.1 |Severe acute respiratory syndrome coro    ····················  ·A··················  ····················  ····················  ····················  ····················
278 MW454398.1 |Severe acute respiratory syndrome coro    ····················  ·A··················  ····················  ····················  ····················  ····················
279 MW454399.1 |Severe acute respiratory syndrome coro    ····················  ····················  ····················  ····················  ····················  ····················
280 MW454400.1 |Severe acute respiratory syndrome coro    ····················  ····················  ····················  ····················  ····················  ····················
281 MW454401.1 |Severe acute respiratory syndrome coro    ····················  ····················  ····················  ····················  ····················  ····················
282 MW454402.1 |Severe acute respiratory syndrome coro    ····················  ·A··················  ····················  ····················  ····················  ····················
283 MW454403.1 |Severe acute respiratory syndrome coro    ····················  ····················  ····················  ····················  ····················  ····················
284 MW454404.1 |Severe acute respiratory syndrome coro    ····················  ····················  ····················  ····················  ····················  ····················
285 MW454406.1 |Severe acute respiratory syndrome coro    ····················  ····················  ····················  ····················  ····················  ····················
286 MW454407.1 |Severe acute respiratory syndrome coro    ····················  ····················  ····················  ····················  ····················  ····················
287 MW454408.1 |Severe acute respiratory syndrome coro    ····················  ····················  ····················  ····················  ····················  ····················
288 MW454410.1 |Severe acute respiratory syndrome coro    ····················  ····················  ····················  ····················  ····················  ····················
289 MW454411.1 |Severe acute respiratory syndrome coro    ····················  ·A··················  ····················  ····················  ····················  ····················
290 MW454412.1 |Severe acute respiratory syndrome coro    ····················  ·A··················  ····················  ····················  ····················  ····················
291 MW454414.1 |Severe acute respiratory syndrome coro    ····················  ····················  ····················  ····················  ····················  ····················
292 MW454415.1 |Severe acute respiratory syndrome coro    ····················  ····················  ····················  ····················  ····················  ····················
293 MW454416.1 |Severe acute respiratory syndrome coro    ····················  ····················  ····················  ····················  ····················  ····················
294 MW454417.1 |Severe acute respiratory syndrome coro    ····················  ·A··················  ····················  ····················  ····················  ····················
295 MW454418.1 |Severe acute respiratory syndrome coro    ····················  ·A··················  ····················  ····················  ····················  ····················
296 MW454419.1 |Severe acute respiratory syndrome coro    ····················  ····················  ····················  ····················  ····················  ····················
297 MW454420.1 |Severe acute respiratory syndrome coro    ····················  ····················  ····················  ····················  ····················  ····················
298 MW454421.1 |Severe acute respiratory syndrome coro    ····················  ·A··················  ····················  ····················  ····················  ····················
299 MW454422.1 |Severe acute respiratory syndrome coro    ····················  ····················  ····················  ····················  ····················  ····················
300 MW454423.1 |Severe acute respiratory syndrome coro    ····················  ····················  ····················  ····················  ····················  ····················
301 MW454424.1 |Severe acute respiratory syndrome coro    ····················  ·A··················  ····················  ····················  ····················  ····················
```

| | | | | | | | | |
|---|---|---|---|---|---|---|---|---|
| 302 | MW454425.1 | \|Severe acute respiratory syndrome coro | .................... | .A.................. | .................... | .................... | .................... | .................... |
| 303 | MW454426.1 | \|Severe acute respiratory syndrome coro | .................... | .A.................. | .................... | .................... | .................... | .................... |
| 304 | MW454427.1 | \|Severe acute respiratory syndrome coro | .................... | .A.................. | .................... | .................... | .................... | .................... |
| 305 | MW454428.1 | \|Severe acute respiratory syndrome coro | .................... | .................... | .................... | .................... | .................... | .................... |
| 306 | MW454429.1 | \|Severe acute respiratory syndrome coro | .................... | .A.................. | .................... | .................... | .................... | .................... |
| 307 | MW454431.1 | \|Severe acute respiratory syndrome coro | .................... | .................... | .................... | .................... | .................... | .................... |
| 308 | MW454433.1 | \|Severe acute respiratory syndrome coro | .................... | .................... | ...T................ | .................... | .................... | .................... |
| 309 | MW454434.1 | \|Severe acute respiratory syndrome coro | .................... | .................... | .................... | .................... | .................... | .................... |
| 310 | MW454435.1 | \|Severe acute respiratory syndrome coro | .................... | .................... | .................... | .................... | .................... | .................... |
| 311 | MW454436.1 | \|Severe acute respiratory syndrome coro | .................... | .................... | .................... | .................... | .................... | .................... |
| 312 | MW454438.1 | \|Severe acute respiratory syndrome coro | .................... | .A.................. | .................... | .................... | .................... | .................... |
| 313 | MW454440.1 | \|Severe acute respiratory syndrome coro | .................... | .A.................. | .................... | .................... | .................... | .................... |
| 314 | MW454441.1 | \|Severe acute respiratory syndrome coro | .................... | .................... | .................... | .................... | .................... | .................... |
| 315 | MW454442.1 | \|Severe acute respiratory syndrome coro | .................... | .A.................. | .................... | .................... | .................... | .................... |
| 316 | MW454446.1 | \|Severe acute respiratory syndrome coro | .................... | .A.................. | .................... | .................... | .................... | .................... |
| 317 | MW454447.1 | \|Severe acute respiratory syndrome coro | .................... | .A.................. | .................... | .................... | .................... | .................... |
| 318 | MW454449.1 | \|Severe acute respiratory syndrome coro | .................... | .................... | .................... | .................... | .................... | .................... |
| 319 | MW454450.1 | \|Severe acute respiratory syndrome coro | .................... | .................... | .................... | .................... | .................... | .................... |
| 320 | MW454451.1 | \|Severe acute respiratory syndrome coro | .................... | .................... | .................... | .................... | .................... | .................... |
| 321 | MW454452.1 | \|Severe acute respiratory syndrome coro | .................... | .................... | .................... | .................... | .................... | .................... |
| 322 | MW454453.1 | \|Severe acute respiratory syndrome coro | .................... | .................... | .................... | .................... | .................... | .................... |
| 323 | MW454455.1 | \|Severe acute respiratory syndrome coro | .................... | .................... | .................... | .................... | .................... | .................... |
| 324 | MW454456.1 | \|Severe acute respiratory syndrome coro | .................... | .................... | .................... | .................... | .................... | .................... |
| 325 | MW454457.1 | \|Severe acute respiratory syndrome coro | .................... | .................... | .................... | .................... | .................... | .................... |
| 326 | MW454459.1 | \|Severe acute respiratory syndrome coro | .................... | .................... | .................... | .................... | .................... | .................... |
| 327 | MW454460.1 | \|Severe acute respiratory syndrome coro | .................... | .................... | .................... | .................... | .................... | .................... |
| 328 | MW454461.1 | \|Severe acute respiratory syndrome coro | .................... | .A.................. | .................... | .................... | .................... | .................... |
| 329 | MW454462.1 | \|Severe acute respiratory syndrome coro | .................... | .A.................. | .................... | .................... | .................... | .................... |
| 330 | MW454463.1 | \|Severe acute respiratory syndrome coro | .................... | .................... | .................... | .................... | .................... | .................... |
| 331 | MW454464.1 | \|Severe acute respiratory syndrome coro | .................... | .................... | .................... | .................... | .................... | .................... |
| 332 | MW454467.1 | \|Severe acute respiratory syndrome coro | .................... | .................... | .................... | .................... | .................... | .................... |
| 333 | MW454468.1 | \|Severe acute respiratory syndrome coro | .................... | .................... | .................... | .................... | .................... | .................... |
| 334 | MW454469.1 | \|Severe acute respiratory syndrome coro | .................... | .................... | .................... | .................... | .................... | .................... |
| 335 | MW454470.1 | \|Severe acute respiratory syndrome coro | .................... | .................... | .................... | .................... | .................... | .................... |
| 336 | MW454471.1 | \|Severe acute respiratory syndrome coro | .................... | .................... | .................... | .................... | .................... | .................... |
| 337 | MW454472.1 | \|Severe acute respiratory syndrome coro | .................... | .................... | .................... | .................... | .................... | .................... |
| 338 | MW454474.1 | \|Severe acute respiratory syndrome coro | .................... | .................... | .................... | .................... | .................... | .................... |
| 339 | MW454475.1 | \|Severe acute respiratory syndrome coro | .................... | .................... | .................... | .................... | .................... | .................... |
| 340 | MW454476.1 | \|Severe acute respiratory syndrome coro | .................... | .................... | .................... | .................... | .................... | .................... |
| 341 | MW454477.1 | \|Severe acute respiratory syndrome coro | .................... | .................... | .................... | .................... | .................... | .................... |
| 342 | MW454478.1 | \|Severe acute respiratory syndrome coro | .................... | .................... | .................... | .................... | .................... | .................... |
| 343 | MW454479.1 | \|Severe acute respiratory syndrome coro | .................... | .................... | .................... | .................... | .................... | .................... |
| 344 | MW454480.1 | \|Severe acute respiratory syndrome coro | .................... | .................... | .................... | .................... | .................... | .................... |
| 345 | MW454481.1 | \|Severe acute respiratory syndrome coro | .................... | .................... | .................... | .................... | .................... | .................... |
| 346 | MW454482.1 | \|Severe acute respiratory syndrome coro | .................... | .................... | .................... | .................... | .................... | .................... |
| 347 | MW454483.1 | \|Severe acute respiratory syndrome coro | .................... | .................... | .................... | .................... | .................... | .................... |
| 348 | MW454486.1 | \|Severe acute respiratory syndrome coro | .................... | .................... | .................... | .................... | .................... | .................... |
| 349 | MW454487.1 | \|Severe acute respiratory syndrome coro | .................... | .................... | .................... | .................... | .................... | .................... |
| 351 | MW454490.1 | \|Severe acute respiratory syndrome coro | .................... | .................... | .................... | .................... | .................... | .................... |
| 352 | MW454492.1 | \|Severe acute respiratory syndrome coro | .................... | .................... | .................... | .................... | .................... | .................... |
| 353 | MW454493.1 | \|Severe acute respiratory syndrome coro | .................... | .................... | .................... | .................... | .................... | .................... |
| 354 | MW454494.1 | \|Severe acute respiratory syndrome coro | .................... | .................... | .................... | .................... | .................... | .................... |
| 355 | MW454495.1 | \|Severe acute respiratory syndrome coro | .................... | .................... | .................... | .................... | .................... | .................... |
| 356 | MW454496.1 | \|Severe acute respiratory syndrome coro | .................... | .................... | .................... | .................... | .................... | .................... |
| 357 | MW454498.1 | \|Severe acute respiratory syndrome coro | .................... | .................... | .................... | .................... | .................... | .................... |
| 358 | MW454499.1 | \|Severe acute respiratory syndrome coro | .................... | .................... | .................... | .................... | .................... | .................... |
| 359 | MW454500.1 | \|Severe acute respiratory syndrome coro | .................... | .................... | .................... | .................... | .................... | .................... |
| 360 | MW454502.1 | \|Severe acute respiratory syndrome coro | .................... | .................... | .................... | .................... | .................... | .................... |
| 361 | MW454503.1 | \|Severe acute respiratory syndrome coro | .................... | .................... | .................... | .................... | .................... | .................... |
| 362 | MW454504.1 | \|Severe acute respiratory syndrome coro | .................... | .................... | .................... | .................... | .................... | .................... |
| 363 | MW454505.1 | \|Severe acute respiratory syndrome coro | .................... | .................... | .................... | .................... | .................... | .................... |
| 364 | MW454506.1 | \|Severe acute respiratory syndrome coro | .................... | .................... | .................... | .................... | .................... | .................... |
| 365 | MW454507.1 | \|Severe acute respiratory syndrome coro | .................... | .................... | .................... | .................... | .................... | .................... |
| 366 | MW454508.1 | \|Severe acute respiratory syndrome coro | .................... | .................... | .................... | .................... | ......T............. | .................... |
| 367 | MW454509.1 | \|Severe acute respiratory syndrome coro | .................... | .................... | .................... | .................... | .................... | .................... |
| 368 | MW454510.1 | \|Severe acute respiratory syndrome coro | .................... | .................... | .................... | .................... | .................... | .................... |
| 369 | MW454511.1 | \|Severe acute respiratory syndrome coro | .................... | .................... | .................... | .................... | .................... | .................... |
| 370 | MW454512.1 | \|Severe acute respiratory syndrome coro | .................... | .A.................. | .................... | .................... | .................... | .................... |
| 371 | MW454513.1 | \|Severe acute respiratory syndrome coro | .................... | .................... | .................... | .................... | .................... | .................... |
| 372 | MW454514.1 | \|Severe acute respiratory syndrome coro | .................... | .A.................. | .................... | .................... | .................... | .................... |
| 373 | MW454515.1 | \|Severe acute respiratory syndrome coro | .................... | .................... | .................... | .................... | .................... | .................... |
| 374 | MW454516.1 | \|Severe acute respiratory syndrome coro | .................... | .A.................. | .................... | .................... | .................... | .................... |
| 375 | MW454517.1 | \|Severe acute respiratory syndrome coro | .................... | .................... | .................... | .................... | .................... | .................... |
| 376 | MW454518.1 | \|Severe acute respiratory syndrome coro | .................... | .................... | .................... | .................... | .................... | .................... |
| 377 | MW454519.1 | \|Severe acute respiratory syndrome coro | .................... | .A.................. | .................... | .................... | .................... | .................... |
| 378 | MW454520.1 | \|Severe acute respiratory syndrome coro | .................... | .................... | .................... | .................... | .................... | .................... |
| 379 | MW454521.1 | \|Severe acute respiratory syndrome coro | .................... | .................... | .................... | .................... | .................... | .................... |
| 380 | MW454522.1 | \|Severe acute respiratory syndrome coro | .................... | .................... | .................... | .................... | .................... | .................... |

| | | | | | | | | |
|---|---|---|---|---|---|---|---|---|
| 381 | MW454523.1 | \|Severe acute respiratory syndrome coro | ···················· | ···················· | ····················· | ···················· | ···················· | ···················· |
| 382 | MW454524.1 | \|Severe acute respiratory syndrome coro | ···················· | ···················· | ····················· | ···················· | ···················· | ···················· |
| 383 | MW454525.1 | \|Severe acute respiratory syndrome coro | ···················· | ···················· | ····················· | ···················· | ···················· | ···················· |
| 384 | MW454526.1 | \|Severe acute respiratory syndrome coro | ···················· | ···················· | ····················· | ···················· | ···················· | ···················· |
| 385 | MW454527.1 | \|Severe acute respiratory syndrome coro | ···················· | ···················· | ····················· | ···················· | ···················· | ···················· |
| 386 | MW454528.1 | \|Severe acute respiratory syndrome coro | ···················· | ···················· | ····················· | ···················· | ···················· | ···················· |
| 387 | MW454529.1 | \|Severe acute respiratory syndrome coro | ···················· | ···················· | ····················· | ···················· | ···················· | ···················· |
| 388 | MW454530.1 | \|Severe acute respiratory syndrome coro | ···················· | ···················· | ····················· | ···················· | ···················· | ···················· |
| 389 | MW454531.1 | \|Severe acute respiratory syndrome coro | ···················· | ···················· | ····················· | ···················· | ···················· | ···················· |
| 390 | MW454532.1 | \|Severe acute respiratory syndrome coro | ···················· | ···················· | ····················· | ···················· | ···················· | ···················· |
| 391 | MW454533.1 | \|Severe acute respiratory syndrome coro | ···················· | ···················· | ····················· | ···················· | ···················· | ···················· |
| 392 | MW454534.1 | \|Severe acute respiratory syndrome coro | ···················· | ···················· | ····················· | ···················· | ···················· | ···················· |
| 393 | MW454535.1 | \|Severe acute respiratory syndrome coro | ···················· | ·A·················· | ····················· | ···················· | ···················· | ···················· |
| 394 | MW454536.1 | \|Severe acute respiratory syndrome coro | ···················· | ···················· | ····················· | ···················· | ···················· | ···················· |
| 395 | MW454537.1 | \|Severe acute respiratory syndrome coro | ···················· | ···················· | ····················· | ···················· | ···················· | ···················· |
| 396 | MW454538.1 | \|Severe acute respiratory syndrome coro | ···················· | ···················· | ····················· | ···················· | ···················· | ···················· |
| 397 | MW454539.1 | \|Severe acute respiratory syndrome coro | ···················· | ···················· | ····················· | ···················· | ···················· | ···················· |
| 398 | MW454541.1 | \|Severe acute respiratory syndrome coro | ···················· | ·A·················· | ····················· | ···················· | ···················· | ···················· |
| 399 | MW454542.1 | \|Severe acute respiratory syndrome coro | ···················· | ···················· | ····················· | ···················· | ···················· | ···················· |
| 400 | MW454543.1 | \|Severe acute respiratory syndrome coro | ···················· | ···················· | ····················· | ···················· | ···················· | ···················· |
| 401 | MW454544.1 | \|Severe acute respiratory syndrome coro | ···················· | ···················· | ····················· | ···················· | ···················· | ···················· |
| 402 | MW454545.1 | \|Severe acute respiratory syndrome coro | ···················· | ···················· | ····················· | ···················· | ···················· | ···················· |
| 403 | MW454546.1 | \|Severe acute respiratory syndrome coro | ···················· | ·A·················· | ····················· | ···················· | ···················· | ···················· |
| 404 | MW454547.1 | \|Severe acute respiratory syndrome coro | ···················· | ···················· | ····················· | ···················· | ···················· | ···················· |
| 405 | MW454548.1 | \|Severe acute respiratory syndrome coro | ···················· | ···················· | ····················· | ···················· | ···················· | ···················· |
| 406 | MW454549.1 | \|Severe acute respiratory syndrome coro | ···················· | ···················· | ····················· | ···················· | ···················· | ···················· |
| 407 | MW454550.1 | \|Severe acute respiratory syndrome coro | ···················· | ···················· | ····················· | ···················· | ···················· | ···················· |
| 408 | MW454551.1 | \|Severe acute respiratory syndrome coro | ···················· | ···················· | ····················· | ···················· | ···················· | ···················· |
| 409 | MW454552.1 | \|Severe acute respiratory syndrome coro | ···················· | ···················· | ····················· | ···················· | ···················· | ···················· |
| 410 | MW454553.1 | \|Severe acute respiratory syndrome coro | ···················· | ···················· | ····················· | ···················· | ···················· | ···················· |
| 411 | MW454554.1 | \|Severe acute respiratory syndrome coro | ···················· | ···················· | ····················· | ···················· | ···················· | ···················· |
| 412 | MW454555.1 | \|Severe acute respiratory syndrome coro | ···················· | ···················· | ····················· | ···················· | ···················· | ···················· |
| 413 | MW454556.1 | \|Severe acute respiratory syndrome coro | ···················· | ···················· | ····················· | ···················· | ···················· | ···················· |
| 414 | MW454557.1 | \|Severe acute respiratory syndrome coro | ···················· | ···················· | ····················· | ···················· | ···················· | ···················· |
| 415 | MW454558.1 | \|Severe acute respiratory syndrome coro | ···················· | ···················· | ····················· | ···················· | ···················· | ···················· |
| 416 | MW454559.1 | \|Severe acute respiratory syndrome coro | ···················· | ···················· | ····················· | ···················· | ···················· | ···················· |
| 417 | MW454560.1 | \|Severe acute respiratory syndrome coro | ···················· | ···················· | ····················· | ···················· | ···················· | ···················· |
| 418 | MW454561.1 | \|Severe acute respiratory syndrome coro | ···················· | ···················· | ····················· | ···················· | ···················· | ···················· |
| 419 | MW454562.1 | \|Severe acute respiratory syndrome coro | ···················· | ···················· | ····················· | ···················· | ···················· | ···················· |
| 420 | MW454563.1 | \|Severe acute respiratory syndrome coro | ···················· | ···················· | ····················· | ···················· | ···················· | ···················· |
| 421 | MW454564.1 | \|Severe acute respiratory syndrome coro | ···················· | ···················· | ····················· | ···················· | ···················· | ···················· |
| 422 | MW454565.1 | \|Severe acute respiratory syndrome coro | ···················· | ···················· | ····················· | ···················· | ···················· | ···················· |
| 423 | MW454566.1 | \|Severe acute respiratory syndrome coro | ···················· | ·········A·········· | ····················· | ···················· | ···················· | ···················· |
| 424 | MW454567.1 | \|Severe acute respiratory syndrome coro | ···················· | ···················· | ····················· | ···················· | ···················· | ···················· |
| 425 | MW454568.1 | \|Severe acute respiratory syndrome coro | ···················· | ···················· | ····················· | ···················· | ···················· | ···················· |
| 426 | MW454569.1 | \|Severe acute respiratory syndrome coro | ···················· | ···················· | ····················· | ···················· | ···················· | ···················· |
| 427 | MW454570.1 | \|Severe acute respiratory syndrome coro | ···················· | ···················· | ····················· | ···················· | ···················· | ···················· |
| 428 | MW454571.1 | \|Severe acute respiratory syndrome coro | ···················· | ···················· | ····················· | ···················· | ···················· | ···················· |
| 429 | MW454572.1 | \|Severe acute respiratory syndrome coro | ···················· | ···················· | ····················· | ···················· | ···················· | ···················· |
| 430 | MW454573.1 | \|Severe acute respiratory syndrome coro | ···················· | ···················· | ····················· | ···················· | ···················· | ···················· |
| 431 | MW454574.1 | \|Severe acute respiratory syndrome coro | ···················· | ···················· | ····················· | ···················· | ···················· | ···················· |
| 432 | MW454575.1 | \|Severe acute respiratory syndrome coro | ···················· | ···················· | ····················· | ···················· | ···················· | ···················· |
| 433 | MW454576.1 | \|Severe acute respiratory syndrome coro | ···················· | ·A·················· | ····················· | ···················· | ···················· | ···················· |
| 434 | MW454577.1 | \|Severe acute respiratory syndrome coro | ···················· | ···················· | ····················· | ···················· | ···················· | ···················· |
| 435 | MW454578.1 | \|Severe acute respiratory syndrome coro | ···················· | ···················· | ····················· | ···················· | ···················· | ···················· |
| 436 | MW454579.1 | \|Severe acute respiratory syndrome coro | ···················· | ···················· | ····················· | ···················· | ···················· | ···················· |
| 437 | MW454580.1 | \|Severe acute respiratory syndrome coro | ···················· | ···················· | ····················· | ···················· | ···················· | ···················· |
| 438 | MW454581.1 | \|Severe acute respiratory syndrome coro | ···················· | ···················· | ····················· | ···················· | ···················· | ···················· |
| 439 | MW454582.1 | \|Severe acute respiratory syndrome coro | ···················· | ···················· | ···········T········· | ···················· | ···················· | ···················· |
| 440 | MW454583.1 | \|Severe acute respiratory syndrome coro | ···················· | ···················· | ····················· | ···················· | ···················· | ···················· |
| 441 | MW454584.1 | \|Severe acute respiratory syndrome coro | ···················· | ···················· | ····················· | ···················· | ···················· | ···················· |
| 442 | MW454585.1 | \|Severe acute respiratory syndrome coro | ···················· | ···················· | ····················· | ···················· | ···················· | ···················· |
| 443 | MW454586.1 | \|Severe acute respiratory syndrome coro | ···················· | ···················· | ····················· | ···················· | ···················· | ···················· |
| 444 | MW454587.1 | \|Severe acute respiratory syndrome coro | ···················· | ···················· | ····················· | ···················· | ···················· | ···················· |
| 445 | MW454588.1 | \|Severe acute respiratory syndrome coro | ···················· | ···················· | ····················· | ···················· | ···················· | ···················· |
| 446 | MW454589.1 | \|Severe acute respiratory syndrome coro | ···················· | ···················· | ····················· | ···················· | ···················· | ···················· |
| 447 | MW454590.1 | \|Severe acute respiratory syndrome coro | ···················· | ···················· | ····················· | ···················· | ···················· | ···················· |
| 448 | MW454591.1 | \|Severe acute respiratory syndrome coro | ···················· | ···················· | ····················· | ···················· | ···················· | ···················· |
| 449 | MW454592.1 | \|Severe acute respiratory syndrome coro | ···················· | ···················· | ····················· | ···················· | ···················· | ···················· |
| 450 | MW454593.1 | \|Severe acute respiratory syndrome coro | ···················· | ···················· | ····················· | ···················· | ···················· | ···················· |
| 451 | MW454594.1 | \|Severe acute respiratory syndrome coro | ···················· | ···················· | ····················· | ···················· | ···················· | ···················· |
| 452 | MW454595.1 | \|Severe acute respiratory syndrome coro | ···················· | ···················· | ····················· | ···················· | ···················· | ···················· |
| 453 | MW454596.1 | \|Severe acute respiratory syndrome coro | ···················· | ···················· | ····················· | ·············A······ | ···················· | ···················· |
| 454 | MW454597.1 | \|Severe acute respiratory syndrome coro | ···················· | ···················· | ····················· | ···················· | ···················· | ···················· |
| 455 | MW454598.1 | \|Severe acute respiratory syndrome coro | ···················· | ···················· | ····················· | ···················· | ···················· | ···················· |
| 456 | MW454599.1 | \|Severe acute respiratory syndrome coro | ···················· | ···················· | ····················· | ···················· | ···················· | ···················· |
| 457 | MW454600.1 | \|Severe acute respiratory syndrome coro | ···················· | ···················· | ····················· | ···················· | ···················· | ···················· |
| 458 | MW454601.1 | \|Severe acute respiratory syndrome coro | ···················· | ···················· | ····················· | ···················· | ···················· | ···················· |

459 MW454602.1 |Severe acute respiratory syndrome coro .................... .................... .................... .................... .................... ....................
460 MW454603.1 |Severe acute respiratory syndrome coro .................... .................... .................... .................... .................... ....................
461 MW454604.1 |Severe acute respiratory syndrome coro .................... .................... .................... .................... .................... ....................
462 MW454605.1 |Severe acute respiratory syndrome coro .................... .................... .................... .................... .................... ....................
463 MW454606.1 |Severe acute respiratory syndrome coro .................... .................... .................... .................... .................... ....................
464 MW454607.1 |Severe acute respiratory syndrome coro .................... .................... .................... .................... .................... ....................
465 MW454608.1 |Severe acute respiratory syndrome coro .................... .A.................. .................... .................... .................... ....................
466 MW454609.1 |Severe acute respiratory syndrome coro .................... .................... .................... .................... .................... ....................
467 MW454610.1 |Severe acute respiratory syndrome coro .................... .................... .................... .................... .................... ....................
468 MW454611.1 |Severe acute respiratory syndrome coro .................... .................... .................... .................... .................... ....................
469 MW454612.1 |Severe acute respiratory syndrome coro .................... .................... .................... .................... .................... ....................
470 MW454613.1 |Severe acute respiratory syndrome coro .................... .................... .................... .................... .................... ....................
471 MW454614.1 |Severe acute respiratory syndrome coro .................... .................... .................... .................... .................... ....................
472 MW454615.1 |Severe acute respiratory syndrome coro .................... .................... .................... .................... .................... ....................
473 MW454617.1 |Severe acute respiratory syndrome coro .................... .................... .................... .................... .................... ....................
474 MW454618.1 |Severe acute respiratory syndrome coro .................... .................... .................... .................... .................... ....................
475 MW454619.1 |Severe acute respiratory syndrome coro .................... .................... .................... .................... .................... ....................
476 MW454620.1 |Severe acute respiratory syndrome coro .................... .................... .................... .................... .................... ....................
477 MW454621.1 |Severe acute respiratory syndrome coro .................... .................... .................... .................... .................... ....................
478 MW454622.1 |Severe acute respiratory syndrome coro .................... ...A................ .................... .................... .................... ....................
479 MW454623.1 |Severe acute respiratory syndrome coro .................... .................... .................... .................... .................... ....................
480 MW454624.1 |Severe acute respiratory syndrome coro .................... .................... .................... .................... .................... ....................
481 MW454625.1 |Severe acute respiratory syndrome coro .................... .................... .................... .................... .................... ....................
482 MW454626.1 |Severe acute respiratory syndrome coro .................... .................... .................... .................... .................... ....................
483 MW454628.1 |Severe acute respiratory syndrome coro .................... .................... .................... .................... .................... ....................
484 MW454629.1 |Severe acute respiratory syndrome coro .................... .................... .................... .................... .................... ....................
485 MW454630.1 |Severe acute respiratory syndrome coro .................... .................... .................... .................... .................... ....................
486 MW454631.1 |Severe acute respiratory syndrome coro .................... .A.................. .................... .................... .................... ....................
487 MW454632.1 |Severe acute respiratory syndrome coro .................... .................... .................... .................... .................... ....................
488 MW454633.1 |Severe acute respiratory syndrome coro .................... .................... .................... .................... .................... ....................
489 MW454634.1 |Severe acute respiratory syndrome coro .................... .................... .................... .................... .................... ....................
490 MW454635.1 |Severe acute respiratory syndrome coro .................... .................... .................... .................... .................... ....................
491 MW454636.1 |Severe acute respiratory syndrome coro .................... .................... .................... .................... .................... ....................
492 MW454637.1 |Severe acute respiratory syndrome coro .................... .................... .................... .................... .................... ....................
493 MW454638.1 |Severe acute respiratory syndrome coro .................... .................... .................... .................... .................... ....................
494 MW454639.1 |Severe acute respiratory syndrome coro .................... .................... .................... .................... .................... ....................
495 MW454640.1 |Severe acute respiratory syndrome coro .................... .................... .................... .................... .................... ....................
496 MW454641.1 |Severe acute respiratory syndrome coro .................... .................... .................... .................... .................... ....................
497 MW454642.1 |Severe acute respiratory syndrome coro .................... .................... .................... .................... .................... ....................
498 MW454643.1 |Severe acute respiratory syndrome coro .................... .................... .................... .................... .................... ....................
499 MW454644.1 |Severe acute respiratory syndrome coro .................... .................... .................... .................... .................... ....................
500 MW454645.1 |Severe acute respiratory syndrome coro .................... .................... .................... .................... .................... ....................
501 MW454646.1 |Severe acute respiratory syndrome coro .................... .................... .................... .................... .................... ....................
502 MW454647.1 |Severe acute respiratory syndrome coro .................... .................... .................... .................... .................... ....................
503 MW454648.1 |Severe acute respiratory syndrome coro .................... .................... .................... .................... .................... ....................
504 MW454649.1 |Severe acute respiratory syndrome coro .................... .A.................. .................... .................... .................... ....................
505 MW454650.1 |Severe acute respiratory syndrome coro .................... .................... .................... .................... .................... ....................
506 MW454651.1 |Severe acute respiratory syndrome coro .................... .................... .................... .................... .................... ....................
507 MW454652.1 |Severe acute respiratory syndrome coro .................... .................... .................... .................... ......T............. ....................
508 MW454653.1 |Severe acute respiratory syndrome coro .................... .................... .................... .................... .................... ....................
509 MW454654.1 |Severe acute respiratory syndrome coro .................... .................... .................... .................... .................... ....................
510 MW454655.1 |Severe acute respiratory syndrome coro .................... .................... .................... .................... .................... ....................
511 MW454656.1 |Severe acute respiratory syndrome coro .................... .................... .................... .................... .................... ....................
512 MW454657.1 |Severe acute respiratory syndrome coro .................... .................... .................... .................... .................... ....................
513 MW454658.1 |Severe acute respiratory syndrome coro .................... .................... .................... .................... .................... ....................
514 MW454659.1 |Severe acute respiratory syndrome coro .................... .................... .................... .................... .................... ....................
515 MW454660.1 |Severe acute respiratory syndrome coro .................... .A.................. .................... .................... .................... ....................
516 MW454661.1 |Severe acute respiratory syndrome coro .................... .................... .................... .................... .................... ....................
517 MW454662.1 |Severe acute respiratory syndrome coro .................... .................... .................... .................... .................... ....................
518 MW454663.1 |Severe acute respiratory syndrome coro .................... .A.................. .................... .................... .................... ....................
519 MW454664.1 |Severe acute respiratory syndrome coro .................... .................... .................... .................... .................... ....................
520 MW454665.1 |Severe acute respiratory syndrome coro .................... .................... .................... .................... .................... ....................
521 MW454666.1 |Severe acute respiratory syndrome coro .................... .................... .................... .................... .................... ....................
522 MW454667.1 |Severe acute respiratory syndrome coro .................... .................... .................... .................... .................... ....................
524 MW454669.1 |Severe acute respiratory syndrome coro .................... .................... .................... .................... .................... ....................
525 MW454670.1 |Severe acute respiratory syndrome coro .................... .................... .................... .................... .................... ....................
526 MW454671.1 |Severe acute respiratory syndrome coro .................... .A.................. .................... .................... .................... ....................
527 MW454672.1 |Severe acute respiratory syndrome coro .................... .................... .................... .................... .................... ....................
528 MW454674.1 |Severe acute respiratory syndrome coro .................... .................... .................... .................... .................... ....................
529 MW454675.1 |Severe acute respiratory syndrome coro .................... .................... .................... .................... .................... ....................
530 MW454676.1 |Severe acute respiratory syndrome coro .................... .................... .................... .................... .................... ....................
531 MW454677.1 |Severe acute respiratory syndrome coro .................... .................... .................... .................... .................... ....................
532 MW454678.1 |Severe acute respiratory syndrome coro .................... .................... .................... .................... .................... ....................
533 MW454679.1 |Severe acute respiratory syndrome coro .................... .................... .................... .................... .................... ....................
534 MW454680.1 |Severe acute respiratory syndrome coro .................... .................... .................... .................... .................... ....................
535 MW454681.1 |Severe acute respiratory syndrome coro .................... .................... .................... .................... .................... ....................
536 MW454682.1 |Severe acute respiratory syndrome coro .................... .................... .................... .................... .................... ....................
537 MW454683.1 |Severe acute respiratory syndrome coro .................... .................... .................... .................... .................... ....................

| | | | | | | | | |
|---|---|---|---|---|---|---|---|---|
| 538 | MW454684.1 | \|Severe acute respiratory syndrome coro | ···················· | ···················· | ···················· | ···················· | ···················· | ···················· |
| 539 | MW454685.1 | \|Severe acute respiratory syndrome coro | ···················· | ···················· | ···················· | ···················· | ···················· | ···················· |
| 540 | MW454686.1 | \|Severe acute respiratory syndrome coro | ···················· | ···················· | ···················· | ···················· | ···················· | ···················· |
| 541 | MW454687.1 | \|Severe acute respiratory syndrome coro | ···················· | ···················· | ···················· | ···················· | ···················· | ···················· |
| 542 | MW454688.1 | \|Severe acute respiratory syndrome coro | ···················· | ···················· | ···················· | ···················· | ···················· | ···················· |
| 544 | MW454690.1 | \|Severe acute respiratory syndrome coro | ···················· | ···················· | ···················· | ···················· | ···················· | ···················· |
| 545 | MW454691.1 | \|Severe acute respiratory syndrome coro | ···················· | ···················· | ···················· | ···················· | ···················· | ···················· |
| 546 | MW454692.1 | \|Severe acute respiratory syndrome coro | ···················· | ·A·················· | ···················· | ···················· | ···················· | ···················· |
| 547 | MW454693.1 | \|Severe acute respiratory syndrome coro | ···················· | ···················· | ···················· | ···················· | ···················· | ···················· |
| 548 | MW454694.1 | \|Severe acute respiratory syndrome coro | ···················· | ···················· | ···················· | ···················· | ···················· | ···················· |
| 549 | MW454695.1 | \|Severe acute respiratory syndrome coro | ···················· | ···················· | ···················· | ···················· | ···················· | ···················· |
| 550 | MW454696.1 | \|Severe acute respiratory syndrome coro | ···················· | ·A·················· | ···················· | ···················· | ···················· | ···················· |
| 551 | MW454697.1 | \|Severe acute respiratory syndrome coro | ···················· | ···················· | ···················· | ···················· | ···················· | ···················· |
| 552 | MW454698.1 | \|Severe acute respiratory syndrome coro | ···················· | ···················· | ···················· | ···················· | ···················· | ···················· |
| 553 | MW454699.1 | \|Severe acute respiratory syndrome coro | ···················· | ···················· | ···················· | ···················· | ···················· | ···················· |
| 554 | MW454700.1 | \|Severe acute respiratory syndrome coro | ···················· | ···················· | ···················· | ···················· | ···················· | ···················· |
| 555 | MW454701.1 | \|Severe acute respiratory syndrome coro | ···················· | ···················· | ···················· | ···················· | ···················· | ···················· |
| 556 | MW454702.1 | \|Severe acute respiratory syndrome coro | ···················· | ···················· | ···················· | ···················· | ···················· | ···················· |
| 557 | MW454704.1 | \|Severe acute respiratory syndrome coro | ···················· | ···················· | ···················· | ···················· | ···················· | ···················· |
| 558 | MW454707.1 | \|Severe acute respiratory syndrome coro | ···················· | ···················· | ···················· | ···················· | ···················· | ···················· |
| 559 | MW454708.1 | \|Severe acute respiratory syndrome coro | ···················· | ···················· | ···················· | ···················· | ···················· | ···················· |
| 560 | MW454709.1 | \|Severe acute respiratory syndrome coro | ···················· | ···················· | ···················· | ···················· | ···················· | ···················· |
| 561 | MW454710.1 | \|Severe acute respiratory syndrome coro | ···················· | ···················· | ···················· | ···················· | ···················· | ···················· |
| 562 | MW454711.1 | \|Severe acute respiratory syndrome coro | ···················· | ···················· | ···················· | ···················· | ···················· | ···················· |
| 563 | MW454712.1 | \|Severe acute respiratory syndrome coro | ···················· | ···················· | ···················· | ···················· | ···················· | ···················· |
| 564 | MW454713.1 | \|Severe acute respiratory syndrome coro | ···················· | ···················· | ···················· | ···················· | ···················· | ···················· |
| 565 | MW454714.1 | \|Severe acute respiratory syndrome coro | ···················· | ···················· | ···················· | ···················· | ···················· | ···················· |
| 566 | MW454715.1 | \|Severe acute respiratory syndrome coro | ···················· | ···················· | ···················· | ···················· | ···················· | ···················· |
| 567 | MW454716.1 | \|Severe acute respiratory syndrome coro | ···················· | ···················· | ···················· | ···················· | ···················· | ···················· |
| 568 | MW454717.1 | \|Severe acute respiratory syndrome coro | ···················· | ···················· | ···················· | ···················· | ···················· | ···················· |
| 569 | MW454718.1 | \|Severe acute respiratory syndrome coro | ···················· | ···················· | ···················· | ···················· | ···················· | ···················· |
| 570 | MW454719.1 | \|Severe acute respiratory syndrome coro | ···················· | ·A·················· | ···················· | ···················· | ···················· | ···················· |
| 571 | MW454720.1 | \|Severe acute respiratory syndrome coro | ···················· | ···················· | ···················· | ···················· | ···················· | ···················· |
| 572 | MW454721.1 | \|Severe acute respiratory syndrome coro | ···················· | ···················· | ···················· | ···················· | ···················· | ···················· |
| 573 | MW454722.1 | \|Severe acute respiratory syndrome coro | ···················· | ···················· | ···················· | ···················· | ···················· | ···················· |
| 574 | MW454723.1 | \|Severe acute respiratory syndrome coro | ···················· | ···················· | ···················· | ···················· | ···················· | ···················· |
| 575 | MW454724.1 | \|Severe acute respiratory syndrome coro | ···················· | ···················· | ···················· | ···················· | ···················· | ···················· |
| 576 | MW454725.1 | \|Severe acute respiratory syndrome coro | ···················· | ···················· | ···················· | ···················· | ···················· | ···················· |
| 577 | MW454726.1 | \|Severe acute respiratory syndrome coro | ···················· | ···················· | ···················· | ···················· | ···················· | ···················· |
| 578 | MW454727.1 | \|Severe acute respiratory syndrome coro | ···················· | ·A·················· | ···················· | ···················· | ···················· | ···················· |
| 579 | MW454728.1 | \|Severe acute respiratory syndrome coro | ···················· | ···················· | ···················· | ···················· | ···················· | ···················· |
| 580 | MW454729.1 | \|Severe acute respiratory syndrome coro | ···················· | ···················· | ···················· | ···················· | ···················· | ···················· |
| 581 | MW454730.1 | \|Severe acute respiratory syndrome coro | ···················· | ···················· | ···················· | ···················· | ···················· | ···················· |
| 582 | MW454731.1 | \|Severe acute respiratory syndrome coro | ···················· | ···················· | ···················· | ···················· | ···················· | ···················· |
| 583 | MW454732.1 | \|Severe acute respiratory syndrome coro | ···················· | ···················· | ···················· | ···················· | ···················· | ···················· |
| 584 | MW454733.1 | \|Severe acute respiratory syndrome coro | ···················· | ···················· | ···················· | ···················· | ···················· | ···················· |
| 585 | MW454734.1 | \|Severe acute respiratory syndrome coro | ···················· | ·A·················· | ···················· | ···················· | ···················· | ···················· |
| 586 | MW454735.1 | \|Severe acute respiratory syndrome coro | ···················· | ···················· | ···················· | ···················· | ···················· | ···················· |
| 587 | MW454736.1 | \|Severe acute respiratory syndrome coro | ···················· | ···················· | ···················· | ···················· | ···················· | ···················· |
| 588 | MW454737.1 | \|Severe acute respiratory syndrome coro | ···················· | ·A·················· | ···················· | ···················· | ···················· | ···················· |
| 589 | MW454738.1 | \|Severe acute respiratory syndrome coro | ···················· | ·A·················· | ···················· | ···················· | ···················· | ···················· |
| 590 | MW454739.1 | \|Severe acute respiratory syndrome coro | ···················· | ···················· | ···················· | ···················· | ···················· | ···················· |
| 591 | MW454740.1 | \|Severe acute respiratory syndrome coro | ···················· | ···················· | ···················· | ···················· | ···················· | ···················· |
| 592 | MW454741.1 | \|Severe acute respiratory syndrome coro | ···················· | ···················· | ···················· | ···················· | ···················· | ···················· |
| 593 | MW454742.1 | \|Severe acute respiratory syndrome coro | ···················· | ·············A······ | ···················· | ···················· | ···················· | ···················· |
| 594 | MW454743.1 | \|Severe acute respiratory syndrome coro | ···················· | ···················· | ···················· | ···················· | ···················· | ···················· |
| 595 | MW454744.1 | \|Severe acute respiratory syndrome coro | ···················· | ·A·················· | ···················· | ···················· | ···················· | ···················· |
| 596 | MW454745.1 | \|Severe acute respiratory syndrome coro | ···················· | ···················· | ···················· | ···················· | ···················· | ···················· |
| 597 | MW454746.1 | \|Severe acute respiratory syndrome coro | ···················· | ···················· | ···················· | ···················· | ···················· | ···················· |
| 598 | MW454747.1 | \|Severe acute respiratory syndrome coro | ···················· | ···················· | ···················· | ···················· | ···················· | ···················· |
| 599 | MW454748.1 | \|Severe acute respiratory syndrome coro | ···················· | ···················· | ···················· | ···················· | ···················· | ···················· |
| 600 | MW454749.1 | \|Severe acute respiratory syndrome coro | ···················· | ···················· | ···················· | ···················· | ···················· | ···················· |
| 601 | MW454750.1 | \|Severe acute respiratory syndrome coro | ···················· | ···················· | ···················· | ···················· | ···················· | ···················· |
| 602 | MW454751.1 | \|Severe acute respiratory syndrome coro | ···················· | ···················· | ···················· | ···················· | ···················· | ···················· |
| 603 | MW454752.1 | \|Severe acute respiratory syndrome coro | ···················· | ···················· | ···················· | ···················· | ···················· | ···················· |
| 604 | MW454753.1 | \|Severe acute respiratory syndrome coro | ···················· | ···················· | ···················· | ···················· | ···················· | ···················· |
| 605 | MW454754.1 | \|Severe acute respiratory syndrome coro | ···················· | ···················· | ···················· | ···················· | ···················· | ···················· |
| 606 | MW454755.1 | \|Severe acute respiratory syndrome coro | ···················· | ···················· | ···················· | ···················· | ···················· | ···················· |
| 607 | MW454756.1 | \|Severe acute respiratory syndrome coro | ···················· | ···················· | ···················· | ···················· | ···················· | ···················· |
| 608 | MW454757.1 | \|Severe acute respiratory syndrome coro | ···················· | ···················· | ···················· | ···················· | ···················· | ···················· |
| 609 | MW454758.1 | \|Severe acute respiratory syndrome coro | ···················· | ···················· | ···················· | ···················· | ···················· | ···················· |
| 610 | MW454759.1 | \|Severe acute respiratory syndrome coro | ···················· | ···················· | ···················· | ···················· | ···················· | ···················· |
| 611 | MW454760.1 | \|Severe acute respiratory syndrome coro | ···················· | ···················· | ···················· | ···················· | ···················· | ···················· |
| 612 | MW454761.1 | \|Severe acute respiratory syndrome coro | ···················· | ·A·················· | ···················· | ···················· | ···················· | ···················· |
| 613 | MW454762.1 | \|Severe acute respiratory syndrome coro | ···················· | ···················· | ···················· | ···················· | ···················· | ···················· |
| 614 | MW454763.1 | \|Severe acute respiratory syndrome coro | ···················· | ···················· | ···················· | ···················· | ···················· | ···················· |
| 615 | MW454764.1 | \|Severe acute respiratory syndrome coro | ···················· | ···················· | ···················· | ···················· | ···················· | ···················· |
| 616 | MW454765.1 | \|Severe acute respiratory syndrome coro | ···················· | ···················· | ···················· | ···················· | ···················· | ···················· |

| | | | | | | | | |
|---|---|---|---|---|---|---|---|---|
| 617 | MW454766.1 | \|Severe acute respiratory syndrome coro | ···················· | ···················· | ···················· | ···················· | ···················· | ···················· |
| 618 | MW454767.1 | \|Severe acute respiratory syndrome coro | ···················· | ·A·················· | ···················· | ···················· | ···················· | ···················· |
| 619 | MW454768.1 | \|Severe acute respiratory syndrome coro | ···················· | ···················· | ···················· | ···················· | ···················· | ···················· |
| 620 | MW454770.1 | \|Severe acute respiratory syndrome coro | ···················· | ···················· | ···················· | ···················· | ···················· | ···················· |
| 621 | MW454771.1 | \|Severe acute respiratory syndrome coro | ···················· | ···················· | ···················· | ···················· | ···················· | ···················· |
| 622 | MW456917.1 | \|Severe acute respiratory syndrome coro | ···················· | ···················· | ···················· | ···················· | ···················· | ···················· |
| 623 | MW460569.1 | \|Severe acute respiratory syndrome coro | ···················· | ···················· | ···················· | ···················· | ···················· | ···················· |
| 624 | MW460570.1 | \|Severe acute respiratory syndrome coro | ···················· | ···················· | ···················· | ···················· | ···················· | ···················· |
| 625 | MW460572.1 | \|Severe acute respiratory syndrome coro | ···················· | ···················· | ···················· | ···················· | ···················· | ···················· |
| 626 | MW460573.1 | \|Severe acute respiratory syndrome coro | ···················· | ···················· | ···················· | ···················· | ···········C········ | ···················· |
| 627 | MW460574.1 | \|Severe acute respiratory syndrome coro | ···················· | ···················· | ···················· | ···················· | ···················· | ···················· |
| 628 | MW460575.1 | \|Severe acute respiratory syndrome coro | ···················· | ···················· | ···················· | ···················· | ···················· | ···················· |
| 629 | MW460576.1 | \|Severe acute respiratory syndrome coro | ···················· | ···················· | ···················· | ···················· | ···················· | ···················· |
| 630 | MW460578.1 | \|Severe acute respiratory syndrome coro | ···················· | ···················· | ···················· | ···················· | ···················· | ···················· |
| 631 | MW460580.1 | \|Severe acute respiratory syndrome coro | ···················· | ·A·················· | ···················· | ···················· | ···················· | ···················· |
| 632 | MW460582.1 | \|Severe acute respiratory syndrome coro | ···················· | ···················· | ···················· | ···················· | ···················· | ···················· |
| 633 | MW460583.1 | \|Severe acute respiratory syndrome coro | ···················· | ···················· | ···················· | ···················· | ···················· | ···················· |
| 634 | MW460586.1 | \|Severe acute respiratory syndrome coro | ···················· | ···················· | ···················· | ···················· | ···················· | ···················· |
| 635 | MW460587.1 | \|Severe acute respiratory syndrome coro | ···················· | ···················· | ···················· | ···················· | ···················· | ···················· |
| 636 | MW460589.1 | \|Severe acute respiratory syndrome coro | ···················· | ·A·················· | ···················· | ···················· | ···················· | ···················· |
| 637 | MW460593.1 | \|Severe acute respiratory syndrome coro | ···················· | ·A·················· | ···················· | ···················· | ···················· | ···················· |
| 638 | MW460594.1 | \|Severe acute respiratory syndrome coro | ···················· | ·A·················· | ···················· | ···················· | ···················· | ···················· |
| 639 | MW460595.1 | \|Severe acute respiratory syndrome coro | ···················· | ·A·················· | ···················· | ···················· | ···················· | ···················· |
| 640 | MW460596.1 | \|Severe acute respiratory syndrome coro | ···················· | ·A·················· | ···················· | ···················· | ···················· | ···················· |
| 641 | MW460597.1 | \|Severe acute respiratory syndrome coro | ···················· | ···················· | ···················· | ···················· | ···················· | ···········T········ |
| 642 | MW460598.1 | \|Severe acute respiratory syndrome coro | ···················· | ·A·················· | ···················· | ···················· | ···················· | ···················· |
| 643 | MW460601.1 | \|Severe acute respiratory syndrome coro | ···················· | ···················· | ···················· | ···················· | ···················· | ···················· |
| 644 | MW460604.1 | \|Severe acute respiratory syndrome coro | ···················· | ···················· | ···················· | ···················· | ···················· | ···················· |
| 645 | MW460605.1 | \|Severe acute respiratory syndrome coro | ···················· | ···················· | ···················· | ···················· | ···················· | ···················· |
| 646 | MW460611.1 | \|Severe acute respiratory syndrome coro | ···················· | ·A·················· | ···················· | ···················· | ···················· | ···················· |
| 647 | MW460612.1 | \|Severe acute respiratory syndrome coro | ···················· | ·A·················· | ···················· | ···················· | ···················· | ···················· |
| 648 | MW460613.1 | \|Severe acute respiratory syndrome coro | ···················· | ···················· | ···················· | ···················· | ···················· | ···················· |
| 649 | MW460617.1 | \|Severe acute respiratory syndrome coro | ···················· | ···················· | ···················· | ···················· | ···················· | ···················· |
| 650 | MW460618.1 | \|Severe acute respiratory syndrome coro | ···················· | ···················· | ···················· | ···················· | ···················· | ···················· |
| 651 | MW460619.1 | \|Severe acute respiratory syndrome coro | ···················· | ·A·················· | ···················· | ···················· | ···················· | ···················· |
| 652 | MW460620.1 | \|Severe acute respiratory syndrome coro | ···················· | ···················· | ···················· | ···················· | ···················· | ···················· |
| 653 | MW460621.1 | \|Severe acute respiratory syndrome coro | ···················· | ···················· | ···················· | ···················· | ···················· | ···················· |
| 654 | MW460622.1 | \|Severe acute respiratory syndrome coro | ···················· | ···················· | ···················· | ···················· | ···················· | ···················· |
| 655 | MW460623.1 | \|Severe acute respiratory syndrome coro | ···················· | ···················· | ···················· | ···················· | ···········C········ | ···················· |
| 656 | MW460624.1 | \|Severe acute respiratory syndrome coro | ···················· | ···················· | ···················· | ···················· | ···················· | ···················· |
| 657 | MW460627.1 | \|Severe acute respiratory syndrome coro | ···················· | ···················· | ···················· | ···················· | ···················· | ···················· |
| 658 | MW460629.1 | \|Severe acute respiratory syndrome coro | ···················· | ···················· | ···················· | ···················· | ···················· | ···················· |
| 659 | MW460630.1 | \|Severe acute respiratory syndrome coro | ···················· | ···················· | ···················· | ···················· | ···················· | ···················· |
| 660 | MW460631.1 | \|Severe acute respiratory syndrome coro | ···················· | ···················· | ···················· | ···················· | ···················· | ···················· |
| 661 | MW460632.1 | \|Severe acute respiratory syndrome coro | ···················· | ···················· | ···················· | ···················· | ···················· | ···················· |
| 662 | MW460634.1 | \|Severe acute respiratory syndrome coro | ···················· | ···················· | ···················· | ···················· | ···················· | ···················· |
| 663 | MW460636.1 | \|Severe acute respiratory syndrome coro | ···················· | ···················· | ···················· | ···················· | ···················· | ···················· |
| 664 | MW460638.1 | \|Severe acute respiratory syndrome coro | ···················· | ···················· | ···················· | ···················· | ···················· | ···················· |
| 665 | MW460640.1 | \|Severe acute respiratory syndrome coro | ···················· | ···················· | ···················· | ···················· | ···················· | ···················· |
| 666 | MW460641.1 | \|Severe acute respiratory syndrome coro | ···················· | ···················· | ···················· | ···················· | ···················· | ···················· |
| 667 | MW460643.1 | \|Severe acute respiratory syndrome coro | ···················· | ···················· | ···················· | ···················· | ···················· | ···················· |
| 668 | MW460644.1 | \|Severe acute respiratory syndrome coro | ···················· | ···················· | ···················· | ···················· | ···················· | ···················· |
| 669 | MW460647.1 | \|Severe acute respiratory syndrome coro | ···················· | ···················· | ···················· | ···················· | ···················· | ···················· |
| 670 | MW460648.1 | \|Severe acute respiratory syndrome coro | ···················· | ···················· | ···················· | ···················· | ···················· | ···················· |
| 671 | MW460650.1 | \|Severe acute respiratory syndrome coro | ···················· | ···················· | ···················· | ···················· | ··········T········· | ···················· |
| 672 | MW460654.1 | \|Severe acute respiratory syndrome coro | ···················· | ···················· | ···················· | ···················· | ···················· | ···················· |
| 673 | MW460658.1 | \|Severe acute respiratory syndrome coro | ···················· | ···················· | ···················· | ···················· | ···················· | ···················· |
| 674 | MW460660.1 | \|Severe acute respiratory syndrome coro | ···················· | ···················· | ···················· | ···················· | ···················· | ···················· |
| 675 | MW460663.1 | \|Severe acute respiratory syndrome coro | ···················· | ·A·················· | ···················· | ···················· | ···················· | ···················· |
| 676 | MW460665.1 | \|Severe acute respiratory syndrome coro | ···················· | ···················· | ···················· | ···················· | ···················· | ···················· |
| 677 | MW460666.1 | \|Severe acute respiratory syndrome coro | ···················· | ···················· | ···················· | ···················· | ···················· | ···················· |
| 678 | MW460667.1 | \|Severe acute respiratory syndrome coro | ···················· | ···················· | ···················· | ···················· | ···················· | ···················· |
| 679 | MW460668.1 | \|Severe acute respiratory syndrome coro | ···················· | ·A·················· | ···················· | ···················· | ···················· | ···················· |
| 680 | MW460669.1 | \|Severe acute respiratory syndrome coro | ···················· | ·A·················· | ···················· | ···················· | ···················· | ···················· |
| 681 | MW460670.1 | \|Severe acute respiratory syndrome coro | ···················· | ···················· | ···················· | ···················· | ···················· | ···················· |
| 682 | MW460876.1 | \|Severe acute respiratory syndrome coro | ···················· | ···················· | ···················· | ···················· | ···················· | ···················· |
| 683 | MW460878.1 | \|Severe acute respiratory syndrome coro | ········T··········· | ·A·················· | ···················· | ···················· | ···················· | ···················· |
| 684 | MW460879.1 | \|Severe acute respiratory syndrome coro | ···················· | ·A·················· | ···················· | ···················· | ···················· | ···················· |
| 685 | MW460883.1 | \|Severe acute respiratory syndrome coro | ···················· | ···················· | ···················· | ···················· | ···················· | ···················· |
| 686 | MW460884.1 | \|Severe acute respiratory syndrome coro | ···················· | ···················· | ···················· | ···················· | ···················· | ···················· |
| 687 | MW460885.1 | \|Severe acute respiratory syndrome coro | ···················· | ···················· | ···················· | ···················· | ···················· | ···················· |
| 688 | MW462644.1 | \|Severe acute respiratory syndrome coro | ···················· | ·A·················· | ···················· | ···················· | ···················· | ···················· |
| 689 | MW462645.1 | \|Severe acute respiratory syndrome coro | ···················· | ·A·················· | ···················· | ···················· | ···················· | ···················· |
| 690 | MW462646.1 | \|Severe acute respiratory syndrome coro | ···················· | ·A·················· | ···················· | ···················· | ···················· | ···················· |
| 691 | MW462647.1 | \|Severe acute respiratory syndrome coro | ···················· | ···················· | ···················· | ···················· | ···················· | ···················· |
| 692 | MW462648.1 | \|Severe acute respiratory syndrome coro | ···················· | ·A·················· | ···················· | ···················· | ···················· | ···················· |
| 693 | MW462649.1 | \|Severe acute respiratory syndrome coro | ···················· | ···················· | ···················· | ···················· | ···················· | ···················· |
| 694 | MW462650.1 | \|Severe acute respiratory syndrome coro | ···················· | ···················· | ···················· | ···················· | ···················· | ···················· |

```
695 MW462651.1 |Severe acute respiratory syndrome coro  ····················  ····················  ·····················  ····················  ····················  ····················
696 MW462652.1 |Severe acute respiratory syndrome coro  ····················  ····················  ·····················  ····················  ····················  ··R·················
697 MW462654.1 |Severe acute respiratory syndrome coro  ····················  ····················  ·····················  ····················  ····················  ····················
698 MW462655.1 |Severe acute respiratory syndrome coro  ····················  ····················  ·····················  ····················  ····················  ····················
699 MW462656.1 |Severe acute respiratory syndrome coro  ····················  ····················  ·····················  ····················  ····················  ····················
700 MW462657.1 |Severe acute respiratory syndrome coro  ····················  ····················  ·····················  ····················  ····················  ····················
701 MW462658.1 |Severe acute respiratory syndrome coro  ····················  ····················  ·····················  ····················  ····················  ····················
702 MW462659.1 |Severe acute respiratory syndrome coro  ····················  ····················  ·····················  ····················  ····················  ····················
703 MW462660.1 |Severe acute respiratory syndrome coro  ····················  ····················  ·····················  ····················  ····················  ····················
704 MW462661.1 |Severe acute respiratory syndrome coro  ····················  ····················  ·····················  ····················  ····················  ····················
705 MW462662.1 |Severe acute respiratory syndrome coro  ····················  ····················  ·····················  ····················  ····················  ····················
706 MW462663.1 |Severe acute respiratory syndrome coro  ····················  ····················  ·····················  ····················  ····················  ····················
707 MW462664.1 |Severe acute respiratory syndrome coro  ····················  ····················  ·····················  ····················  ····················  ····················
708 MW462665.1 |Severe acute respiratory syndrome coro  ····················  ····················  ·····················  ····················  ····················  ····················
709 MW462666.1 |Severe acute respiratory syndrome coro  ····················  ·A··················  ·····················  ····················  ····················  ····················
710 MW462667.1 |Severe acute respiratory syndrome coro  ····················  ····················  ·····················  ····················  ····················  ····················
711 MW462668.1 |Severe acute respiratory syndrome coro  ····················  ····················  ·····················  ····················  ····················  ····················
712 MW462669.1 |Severe acute respiratory syndrome coro  ····················  ····················  ·····················  ····················  ····················  ····················
713 MW462670.1 |Severe acute respiratory syndrome coro  ····················  ·A··················  ·····················  ····················  ····················  ····················
714 MW462671.1 |Severe acute respiratory syndrome coro  ····················  ····················  ·····················  ····················  ····················  ····················
715 MW462672.1 |Severe acute respiratory syndrome coro  ····················  ····················  ·····················  ····················  ····················  ····················
716 MW462673.1 |Severe acute respiratory syndrome coro  ····················  ····················  ·····················  ····················  ····················  ····················
717 MW462674.1 |Severe acute respiratory syndrome coro  ····················  ·············A······  ·····················  ····················  ····················  ····················
718 MW462675.1 |Severe acute respiratory syndrome coro  ····················  ····················  ·····················  ····················  ····················  ····················
719 MW463056.1 |Severe acute respiratory syndrome coro  ····················  ····················  ·····················  ····················  ····················  ····················
720 MW453103.1 |Severe acute respiratory syndrome coro  ····················  ····················  ·····················  ····················  ····················  ····················
721 MW453106.1 |Severe acute respiratory syndrome coro  ····················  ····················  ·····················  ····················  ····················  ····················
722 MW453107.1 |Severe acute respiratory syndrome coro  ····················  ····················  ·····················  ····················  ····················  ····················
723 MW453108.1 |Severe acute respiratory syndrome coro  ····················  ····················  ·····················  ····················  ····················  ····················
724 MW453109.1 |Severe acute respiratory syndrome coro  ····················  ····················  ·····················  ····················  ····················  ····················
725 MW453110.1 |Severe acute respiratory syndrome coro  ····················  ····················  ·····················  ····················  ····················  ····················
726 MW453111.1 |Severe acute respiratory syndrome coro  ····················  ·A··················  ·····················  ····················  ····················  ····················
727 MW453112.1 |Severe acute respiratory syndrome coro  ····················  ····················  ·····················  ····················  ····················  ····················
728 MW453114.1 |Severe acute respiratory syndrome coro  ····················  ····················  ·····················  ····················  ····················  ····················
729 MW453115.1 |Severe acute respiratory syndrome coro  ····················  ·A··················  ·····················  ····················  ····················  ····················
730 MW453116.1 |Severe acute respiratory syndrome coro  ····················  ····················  ·····················  ····················  ····················  ····················
731 MW453117.1 |Severe acute respiratory syndrome coro  ····················  ·A··················  ·····················  ····················  ····················  ····················
732 MW453118.1 |Severe acute respiratory syndrome coro  ····················  ·A··················  ·····················  ····················  ····················  ····················
733 MW453119.1 |Severe acute respiratory syndrome coro  ····················  ····················  ·····················  ····················  ····················  ····················
734 MW453120.1 |Severe acute respiratory syndrome coro  ····················  ····················  ·····················  ····················  ····················  ····················
735 MW453121.1 |Severe acute respiratory syndrome coro  ····················  ·A··················  ·····················  ····················  ····················  ····················
736 MW453122.1 |Severe acute respiratory syndrome coro  ····················  ····················  ·····················  ····················  ····················  ····················
737 MW453123.1 |Severe acute respiratory syndrome coro  ····················  ····················  ·····················  ····················  ····················  ····················
738 MW453124.1 |Severe acute respiratory syndrome coro  ····················  ·A··················  ·····················  ····················  ····················  ····················
739 MW453126.1 |Severe acute respiratory syndrome coro  ····················  ·A··················  ·····················  ····················  ····················  ····················
740 MW453084.1 |Severe acute respiratory syndrome coro  ····················  ····················  ·····················  ····················  ····················  ····················
741 MW453085.1 |Severe acute respiratory syndrome coro  ····················  ····················  ·····················  ····················  ····················  ····················
742 LR991698.2 |Severe acute respiratory syndrome coro  ····················  ····················  ·····················  ····················  ····················  ····················
743 LR991699.2 |Severe acute respiratory syndrome coro  ····················  ····················  ·····················  ····················  ····················  ····················
744 MW450666.1 |Severe acute respiratory syndrome coro  ····················  ····················  ·····················  ····················  ····················  ····················
745 MW451205.1 |Severe acute respiratory syndrome coro  ····················  ····················  ·····················  ····················  ····················  ····················
746 MW451601.1 |Severe acute respiratory syndrome coro  ····················  ····················  ·····················  ····················  ····················  ····················
747 MW451602.1 |Severe acute respiratory syndrome coro  ····················  ····················  ·····················  ····················  ····················  ····················
748 MW452531.1 |Severe acute respiratory syndrome coro  ····················  ····················  ·····················  ····················  ····················  ····················
749 MW452532.1 |Severe acute respiratory syndrome coro  ····················  ····················  ·····················  ····················  ··········T·········  ····················
750 MW452534.1 |Severe acute respiratory syndrome coro  ····················  ····················  ·····················  ····················  ····················  ····················
751 MW452536.1 |Severe acute respiratory syndrome coro  ····················  ····················  ·····················  ····················  ····················  ····················
752 MW452537.1 |Severe acute respiratory syndrome coro  ····················  ····················  ·····················  ····················  ····················  ····················
753 MW452539.1 |Severe acute respiratory syndrome coro  ····················  ····················  ·····················  ····················  ····················  ····················
754 MW452542.1 |Severe acute respiratory syndrome coro  ····················  ····················  ·····················  ····················  ····················  ····················
755 MW452543.1 |Severe acute respiratory syndrome coro  ····················  ····················  ·····················  ····················  ····················  ····················
756 MW452544.1 |Severe acute respiratory syndrome coro  ····················  ····················  ·····················  ····················  ····················  ····················
757 MW446194.1 |Severe acute respiratory syndrome coro  ····················  ····················  ·····················  ····················  ····················  ····················
758 MW446195.1 |Severe acute respiratory syndrome coro  ····················  ····················  ·····················  ····················  ····················  ····················
759 MW446196.1 |Severe acute respiratory syndrome coro  ····················  ····················  ·····················  ····················  ····················  ····················
760 MW446197.1 |Severe acute respiratory syndrome coro  ····················  ····················  ·····················  ····················  ····················  ····················
761 MW446198.1 |Severe acute respiratory syndrome coro  ····················  ····················  ·····················  ····················  ····················  ····················
762 MW446199.1 |Severe acute respiratory syndrome coro  ····················  ····················  ·····················  ····················  ····················  ····················
763 MW446200.1 |Severe acute respiratory syndrome coro  ····················  ····················  ·····················  ····················  ····················  ····················
764 MW446201.1 |Severe acute respiratory syndrome coro  ····················  ····················  ·····················  ····················  ····················  ····················
765 MW446202.1 |Severe acute respiratory syndrome coro  ····················  ····················  ·····················  ····················  ····················  ····················
766 MW446203.1 |Severe acute respiratory syndrome coro  ····················  ····················  ·····················  ····················  ····················  ····················
767 MW446204.1 |Severe acute respiratory syndrome coro  ····················  ····················  ·····················  ····················  ····················  ····················
768 MW446205.1 |Severe acute respiratory syndrome coro  ····················  ····················  ·····················  ····················  ····················  ····················
769 MW446206.1 |Severe acute respiratory syndrome coro  ····················  ····················  ·····················  ····················  ····················  ····················
770 MW446207.1 |Severe acute respiratory syndrome coro  ····················  ····················  ·····················  ····················  ····················  ····················
771 MW446208.1 |Severe acute respiratory syndrome coro  ····················  ····················  ·····················  ····················  ····················  ····················
772 MW446209.1 |Severe acute respiratory syndrome coro  ····················  ·A··················  ·····················  ····················  ·NN·················  ····················
```

| | | | | | | | | |
|---|---|---|---|---|---|---|---|---|
| 773 | MW446210.1 | |Severe acute respiratory syndrome coro | ···················· | ·A·················· | ···················· | ···················· | ···················· | ···················· |
| 774 | MW446211.1 | |Severe acute respiratory syndrome coro | ···················· | ···················· | ···················· | ···················· | ···················· | ···················· |
| 775 | MW446212.1 | |Severe acute respiratory syndrome coro | ···················· | ···················· | ···················· | ···················· | ···················· | ···················· |
| 776 | MW446213.1 | |Severe acute respiratory syndrome coro | ···················· | ·A·················· | ···················· | ···················· | ···················· | ···················· |
| 777 | MW446214.1 | |Severe acute respiratory syndrome coro | ···················· | ···················· | ···················· | ···················· | ···················· | ···················· |
| 778 | MW446215.1 | |Severe acute respiratory syndrome coro | ···················· | ···················· | ···················· | ···················· | ···················· | ···················· |
| 779 | MW446216.1 | |Severe acute respiratory syndrome coro | ···················· | ···················· | ···················· | ···················· | ···················· | ···················· |
| 780 | MW446217.1 | |Severe acute respiratory syndrome coro | ···················· | ···················· | ···················· | ···················· | ···················· | ···················· |
| 781 | MW446218.1 | |Severe acute respiratory syndrome coro | ···················· | ···················· | ···················· | ···················· | ···················· | ···················· |
| 783 | MW446220.1 | |Severe acute respiratory syndrome coro | ···················· | ···················· | ···················· | ···················· | ···················· | ···················· |
| 784 | MW446221.1 | |Severe acute respiratory syndrome coro | ···················· | ·A·················· | ···················· | ···················· | ···················· | ···················· |
| 785 | MW446222.1 | |Severe acute respiratory syndrome coro | ···················· | ···················· | ···················· | ···················· | ···················· | ···················· |
| 786 | MW446223.1 | |Severe acute respiratory syndrome coro | ···················· | ···················· | ···················· | ···················· | ···················· | ···················· |
| 787 | MW446224.1 | |Severe acute respiratory syndrome coro | ···················· | ·A·················· | ···················· | ···················· | ···················· | ···················· |
| 788 | MW446225.1 | |Severe acute respiratory syndrome coro | ···················· | ·A·················· | ···················· | ···················· | ···················· | ···················· |
| 789 | MW446226.1 | |Severe acute respiratory syndrome coro | ···················· | ·A·················· | ···················· | ···················· | ···················· | ···················· |
| 790 | MW446227.1 | |Severe acute respiratory syndrome coro | ···················· | ···················· | ···················· | ···················· | ···················· | ···················· |
| 791 | MW446228.1 | |Severe acute respiratory syndrome coro | ···················· | ·A·················· | ···················· | ···················· | ···················· | ···················· |
| 792 | MW446229.1 | |Severe acute respiratory syndrome coro | ···················· | ···················· | ···················· | ···················· | ···················· | ···················· |
| 793 | MW446230.1 | |Severe acute respiratory syndrome coro | ···················· | ···················· | ···················· | ···················· | ···················· | ···················· |
| 794 | MW446231.1 | |Severe acute respiratory syndrome coro | ···················· | ···················· | ···················· | ···················· | ···················· | ···················· |
| 795 | MW446232.1 | |Severe acute respiratory syndrome coro | ···················· | ···················· | ···················· | ···················· | ···················· | ···················· |
| 796 | MW446233.1 | |Severe acute respiratory syndrome coro | ···················· | ···················· | ···················· | ···················· | ···················· | ···················· |
| 797 | MW446234.1 | |Severe acute respiratory syndrome coro | ···················· | ···················· | ···················· | ···················· | ···················· | ···················· |
| 798 | MW446235.1 | |Severe acute respiratory syndrome coro | ···················· | ···················· | ···················· | ···················· | ···················· | ···················· |
| 799 | MW446821.1 | |Severe acute respiratory syndrome coro | ···················· | ···················· | ···················· | ···················· | ···················· | ···················· |
| 800 | MW446822.1 | |Severe acute respiratory syndrome coro | ···················· | ·A·················· | ···················· | ···················· | ···················· | ···················· |
| 801 | MW446823.1 | |Severe acute respiratory syndrome coro | ···················· | ···················· | ···················· | ···················· | ···················· | ···················· |
| 802 | MW446824.1 | |Severe acute respiratory syndrome coro | ···················· | ···················· | ···················· | ···················· | ···················· | ···················· |
| 803 | MW446825.1 | |Severe acute respiratory syndrome coro | ···················· | ···················· | ···················· | ···················· | ···················· | ···················· |
| 804 | MW446826.1 | |Severe acute respiratory syndrome coro | ···················· | ···················· | ···················· | ···················· | ···················· | ···················· |
| 805 | MW446827.1 | |Severe acute respiratory syndrome coro | ···················· | ···················· | ···················· | ···················· | ···················· | ···················· |
| 806 | MW446828.1 | |Severe acute respiratory syndrome coro | ···················· | ···················· | ···················· | ···················· | ···················· | ···················· |
| 807 | MW446829.1 | |Severe acute respiratory syndrome coro | ···················· | ···················· | ···················· | ···················· | ···················· | ···················· |
| 808 | MW446830.1 | |Severe acute respiratory syndrome coro | ···················· | ···················· | ···················· | ···················· | ···················· | ···················· |
| 809 | MW446831.1 | |Severe acute respiratory syndrome coro | ···················· | ···················· | ···················· | ···················· | ···················· | ···················· |
| 810 | MW446832.1 | |Severe acute respiratory syndrome coro | ···················· | ·A·················· | ···················· | ···················· | ···················· | ···················· |
| 811 | MW446833.1 | |Severe acute respiratory syndrome coro | ···················· | ···················· | ···················· | ···················· | ···················· | ···················· |
| 812 | MW446834.1 | |Severe acute respiratory syndrome coro | ···················· | ···················· | ···················· | ···················· | ···················· | ···················· |
| 813 | MW446835.1 | |Severe acute respiratory syndrome coro | ···················· | ···················· | ···················· | ···················· | ···················· | ···················· |
| 814 | MW446836.1 | |Severe acute respiratory syndrome coro | ···················· | ···················· | ···················· | ···················· | ···················· | ···················· |
| 815 | MW446837.1 | |Severe acute respiratory syndrome coro | ···················· | ·A·················· | ···················· | ···················· | ···················· | ···················· |
| 816 | MW446838.1 | |Severe acute respiratory syndrome coro | ···················· | ···················· | ···················· | ···················· | ···················· | ···················· |
| 817 | MW446839.1 | |Severe acute respiratory syndrome coro | ···················· | ···················· | ···················· | ···················· | ···················· | ···················· |
| 818 | MW446840.1 | |Severe acute respiratory syndrome coro | ···················· | ···················· | ···················· | ···················· | ···················· | ···················· |
| 819 | MW446843.1 | |Severe acute respiratory syndrome coro | ···················· | ···················· | ···················· | ···················· | ···················· | ···················· |
| 820 | MW446844.1 | |Severe acute respiratory syndrome coro | ···················· | ···················· | ···················· | ···················· | ···················· | ···················· |
| 821 | MW446846.1 | |Severe acute respiratory syndrome coro | ···················· | ···················· | ···················· | ···················· | ···················· | ···················· |
| 822 | MW446847.1 | |Severe acute respiratory syndrome coro | ···················· | ············C······· | ···················· | ···················· | ···················· | ···················· |
| 823 | MW446848.1 | |Severe acute respiratory syndrome coro | ···················· | ············C······· | ···················· | ···················· | ···················· | ···················· |
| 824 | MW446849.1 | |Severe acute respiratory syndrome coro | ···················· | ···················· | ···················· | ···················· | ···················· | ···················· |
| 825 | MW446850.1 | |Severe acute respiratory syndrome coro | ···················· | ···················· | ···················· | ···················· | ···················· | ···················· |
| 826 | MW446851.1 | |Severe acute respiratory syndrome coro | ···················· | ·A·················· | ···················· | ···················· | ···················· | ···················· |
| 827 | MW446852.1 | |Severe acute respiratory syndrome coro | ···················· | ···················· | ···················· | ···················· | ···················· | ···················· |
| 828 | MW446854.1 | |Severe acute respiratory syndrome coro | ···················· | ············C······· | ···················· | ···················· | ···················· | ···················· |
| 829 | MW446855.1 | |Severe acute respiratory syndrome coro | ···················· | ············C······· | ···················· | ···················· | ···················· | ···················· |
| 830 | MW446856.1 | |Severe acute respiratory syndrome coro | ···················· | ···················· | ···················· | ···················· | ···················· | ···················· |
| 831 | MW446857.1 | |Severe acute respiratory syndrome coro | ···················· | ···················· | ···················· | ···················· | ···················· | ···················· |
| 832 | MW446858.1 | |Severe acute respiratory syndrome coro | ···················· | ············C······· | ···················· | ···················· | ···················· | ···················· |
| 833 | MW446859.1 | |Severe acute respiratory syndrome coro | ···················· | ···················· | ···················· | ···················· | ···················· | ···················· |
| 834 | MW446860.1 | |Severe acute respiratory syndrome coro | ···················· | ·A·················· | ···················· | ···················· | ···················· | ···················· |
| 835 | MW446861.1 | |Severe acute respiratory syndrome coro | ···················· | ·A·················· | ···················· | ···················· | ···················· | ···················· |
| 836 | MW446862.1 | |Severe acute respiratory syndrome coro | ···················· | ···················· | ···················· | ···················· | ···················· | ···················· |
| 837 | MW446863.1 | |Severe acute respiratory syndrome coro | ···················· | ···················· | ···················· | ···················· | ···················· | ···················· |
| 838 | MW446864.1 | |Severe acute respiratory syndrome coro | ···················· | ···················· | ···················· | ···················· | ···················· | ···················· |
| 839 | MW446865.1 | |Severe acute respiratory syndrome coro | ···················· | ···················· | ···················· | ···················· | ···················· | ···················· |
| 840 | MW446866.1 | |Severe acute respiratory syndrome coro | ···················· | ·A·················· | ···················· | ···················· | ···················· | ···················· |
| 841 | MW446867.1 | |Severe acute respiratory syndrome coro | ···················· | ···················· | ···················· | ···················· | ···················· | ···················· |
| 842 | MW446868.1 | |Severe acute respiratory syndrome coro | ···················· | ·A·················· | ···················· | ···················· | ···················· | ···················· |
| 843 | MW446869.1 | |Severe acute respiratory syndrome coro | ···················· | ···················· | ···················· | ···················· | ···················· | ···················· |
| 844 | MW446870.1 | |Severe acute respiratory syndrome coro | ···················· | ···················· | ···················· | ···················· | ···················· | ···················· |
| 845 | MW446871.1 | |Severe acute respiratory syndrome coro | ···················· | ···················· | ···················· | ···················· | ···················· | ···················· |
| 846 | MW446872.1 | |Severe acute respiratory syndrome coro | ···················· | ············C······· | ···················· | ···················· | ···················· | ···················· |
| 847 | MW446873.1 | |Severe acute respiratory syndrome coro | ···················· | ···················· | ···················· | ···················· | ···················· | ···················· |
| 848 | MW446874.1 | |Severe acute respiratory syndrome coro | ···················· | ···················· | ···················· | ···················· | ···················· | ···················· |
| 849 | MW446875.1 | |Severe acute respiratory syndrome coro | ···················· | ···················· | ···················· | ···················· | ···················· | ···················· |
| 850 | MW446876.1 | |Severe acute respiratory syndrome coro | ···················· | ···················· | ···················· | ···················· | ···················· | ···················· |
| 851 | MW446877.1 | |Severe acute respiratory syndrome coro | ···················· | ···················· | ···················· | ···················· | ···················· | ···················· |

| | | | | | | | | |
|---|---|---|---|---|---|---|---|---|
| 852 | MW447609.1 | \|Severe acute respiratory syndrome coro | ···················· | ···················· | ···················· | ···················· | ···················· | ···················· |
| 853 | MW447610.1 | \|Severe acute respiratory syndrome coro | ···················· | ···················· | ···················· | ···················· | ···················· | ···················· |
| 854 | MW447611.1 | \|Severe acute respiratory syndrome coro | ···················· | ···················· | ···················· | ···················· | ···················· | ···················· |
| 855 | MW447612.1 | \|Severe acute respiratory syndrome coro | ···················· | ···················· | ···················· | ···················· | ···················· | ···················· |
| 856 | MW447614.1 | \|Severe acute respiratory syndrome coro | ···················· | ·A·················· | ···················· | ···················· | ···················· | ··A················· |
| 857 | MW447616.1 | \|Severe acute respiratory syndrome coro | ···················· | ·A·················· | ···················· | ···················· | ···················· | ··A················· |
| 858 | MW447618.1 | \|Severe acute respiratory syndrome coro | ···················· | ···················· | ···················· | ···················· | ···················· | ···················· |
| 859 | MW447619.1 | \|Severe acute respiratory syndrome coro | ···················· | ·A·················· | ···················· | ···················· | ···················· | ··A················· |
| 860 | MW447620.1 | \|Severe acute respiratory syndrome coro | ···················· | ···················· | ···················· | ···················· | ···················· | ···················· |
| 861 | MW447621.1 | \|Severe acute respiratory syndrome coro | ···················· | ···················· | ···················· | ···················· | ···················· | ···················· |
| 862 | MW447622.1 | \|Severe acute respiratory syndrome coro | ···················· | ···················· | ···················· | ···················· | ···················· | ···················· |
| 863 | MW447623.1 | \|Severe acute respiratory syndrome coro | ···················· | ···················· | ···················· | ···················· | ···················· | ···················· |
| 864 | MW447624.1 | \|Severe acute respiratory syndrome coro | ···················· | ·A·················· | ···················· | ···················· | ···················· | ··A················· |
| 865 | MW447626.1 | \|Severe acute respiratory syndrome coro | ···················· | ···················· | ···················· | ···················· | ···················· | ···················· |
| 866 | MW447627.1 | \|Severe acute respiratory syndrome coro | ···················· | ···················· | ···················· | ···················· | ···················· | ···················· |
| 867 | MW447630.1 | \|Severe acute respiratory syndrome coro | ···················· | ···················· | ···················· | ···················· | ···················· | ···················· |
| 868 | MW447632.1 | \|Severe acute respiratory syndrome coro | ···················· | ·A·················· | ···················· | ···················· | ···················· | ··A················· |
| 869 | MW447634.1 | \|Severe acute respiratory syndrome coro | ···················· | ···················· | ···················· | ···················· | ···················· | ···················· |
| 870 | MW447635.1 | \|Severe acute respiratory syndrome coro | ···················· | ···················· | ···················· | ···················· | ···················· | ···················· |
| 871 | MW447636.1 | \|Severe acute respiratory syndrome coro | ···················· | ···················· | ···················· | ···················· | ···················· | ···················· |
| 872 | MW447637.1 | \|Severe acute respiratory syndrome coro | ···················· | ···················· | ···················· | ···················· | ···················· | ···················· |
| 873 | MW447638.1 | \|Severe acute respiratory syndrome coro | ···················· | ·A·················· | ···················· | ···················· | ···················· | ··A················· |
| 874 | MW447639.1 | \|Severe acute respiratory syndrome coro | ···················· | ···················· | ···················· | ···················· | ···················· | ···················· |
| 875 | MW447640.1 | \|Severe acute respiratory syndrome coro | ···················· | ·A·················· | ···················· | ···················· | ···················· | ··A················· |
| 876 | MW447641.1 | \|Severe acute respiratory syndrome coro | ···················· | ···················· | ···················· | ···················· | ···················· | ···················· |
| 877 | MW447642.1 | \|Severe acute respiratory syndrome coro | ···················· | ···················· | ···················· | ···················· | ···················· | ···················· |
| 878 | MW447643.1 | \|Severe acute respiratory syndrome coro | ···················· | ···················· | ···················· | ···················· | ···················· | ···················· |
| 879 | MW447644.1 | \|Severe acute respiratory syndrome coro | ···················· | ···················· | ···················· | ···················· | ···················· | ···················· |
| 880 | MW447645.1 | \|Severe acute respiratory syndrome coro | ···················· | ·A·················· | ···················· | ···················· | ···················· | ··A················· |
| 881 | MW449268.1 | \|Severe acute respiratory syndrome coro | ···················· | ···················· | ···················· | ···················· | ···················· | ···················· |
| 882 | MW449277.1 | \|Severe acute respiratory syndrome coro | ···················· | ·A·················· | ···················· | ···················· | ···················· | ···················· |
| 883 | MW449278.1 | \|Severe acute respiratory syndrome coro | ···················· | ···················· | ···················· | ···················· | ···················· | ···················· |
| 884 | MW449279.1 | \|Severe acute respiratory syndrome coro | ···················· | ···················· | ···················· | ···················· | ···················· | ···················· |
| 885 | MW449281.1 | \|Severe acute respiratory syndrome coro | ···················· | ·A·················· | ···················· | ···················· | ···················· | ···················· |
| 886 | MW449283.1 | \|Severe acute respiratory syndrome coro | ···················· | ·A·················· | ···················· | ···················· | ···················· | ···················· |
| 887 | MW449287.1 | \|Severe acute respiratory syndrome coro | ···················· | ···················· | ···················· | ···················· | ···················· | ···················· |
| 888 | MW449289.1 | \|Severe acute respiratory syndrome coro | ···················· | ···················· | ···················· | ···················· | ···················· | ···················· |
| 889 | MW449290.1 | \|Severe acute respiratory syndrome coro | ···················· | ·A·················· | ···················· | ···················· | ···················· | ···················· |
| 890 | MW449292.1 | \|Severe acute respiratory syndrome coro | ···················· | ·A·················· | ···················· | ···················· | ···················· | ···················· |
| 891 | MW449293.1 | \|Severe acute respiratory syndrome coro | ···················· | ···················· | ···················· | ···················· | ···················· | ···················· |
| 892 | MW449298.1 | \|Severe acute respiratory syndrome coro | ···················· | ·A·················· | ···················· | ···················· | ···················· | ···················· |
| 893 | MW449300.1 | \|Severe acute respiratory syndrome coro | ···················· | ·A·················· | ···················· | ···················· | ···················· | ···················· |
| 894 | MW449301.1 | \|Severe acute respiratory syndrome coro | ···················· | ·A·················· | ···················· | ···················· | ···················· | ··A················· |
| 895 | MW449303.1 | \|Severe acute respiratory syndrome coro | ···················· | ···················· | ··T················· | ···················· | ···················· | ···················· |
| 896 | MW449305.1 | \|Severe acute respiratory syndrome coro | ···················· | ·A·················· | ···················· | ···················· | ···················· | ···················· |
| 897 | MW449310.1 | \|Severe acute respiratory syndrome coro | ···················· | ·A·················· | ···················· | ···················· | ···················· | ···················· |
| 898 | MW449312.1 | \|Severe acute respiratory syndrome coro | ···················· | ···················· | ···················· | ···················· | ···················· | ···················· |
| 899 | MW449317.1 | \|Severe acute respiratory syndrome coro | ···················· | ···················· | ···················· | ···················· | ···················· | ···················· |
| 900 | MW449319.1 | \|Severe acute respiratory syndrome coro | ···················· | ·A·················· | ···················· | ···················· | ···················· | ···················· |
| 901 | MW449321.1 | \|Severe acute respiratory syndrome coro | ···················· | ···················· | ···················· | ···················· | ···················· | ···················· |
| 902 | MW449323.1 | \|Severe acute respiratory syndrome coro | ···················· | ·A·················· | ···················· | ···················· | ···················· | ···················· |
| 903 | MW449325.1 | \|Severe acute respiratory syndrome coro | ···················· | ·A·················· | ···················· | ···················· | ···················· | ···················· |
| 904 | MW449326.1 | \|Severe acute respiratory syndrome coro | ···················· | ···················· | ···················· | ···················· | ···················· | ···················· |
| 905 | MW449329.1 | \|Severe acute respiratory syndrome coro | ···················· | ·A·················· | ···················· | ···················· | ···················· | ···················· |
| 906 | MW449331.1 | \|Severe acute respiratory syndrome coro | ···················· | ···················· | ···················· | ···················· | ···················· | ···················· |
| 907 | MW449339.1 | \|Severe acute respiratory syndrome coro | ···················· | ···················· | ···················· | ···················· | ···················· | ···················· |
| 908 | MW449343.1 | \|Severe acute respiratory syndrome coro | ···················· | ···················· | ···················· | ···················· | ···················· | ···················· |
| 909 | MW449346.1 | \|Severe acute respiratory syndrome coro | ···················· | ·A·················· | ···················· | ···················· | ···················· | ···················· |
| 910 | MW449349.1 | \|Severe acute respiratory syndrome coro | ···················· | ·A·················· | ···················· | ···················· | ···················· | ···················· |
| 911 | MW449351.1 | \|Severe acute respiratory syndrome coro | ···················· | ···················· | ···················· | ···················· | ···················· | ···················· |
| 912 | MW449355.1 | \|Severe acute respiratory syndrome coro | ···················· | ···················· | ···················· | ···················· | ···················· | ···················· |
| 913 | MW449356.1 | \|Severe acute respiratory syndrome coro | ···················· | ·A·················· | ···················· | ···················· | ···················· | ···················· |
| 914 | MW449357.1 | \|Severe acute respiratory syndrome coro | ···················· | ·A·················· | ···················· | ···················· | ···················· | ···················· |
| 915 | MW449360.1 | \|Severe acute respiratory syndrome coro | ···················· | ···················· | ···················· | ···················· | ···················· | ···················· |
| 916 | MW449368.1 | \|Severe acute respiratory syndrome coro | ···················· | ···················· | ···················· | ···················· | ···················· | ···················· |
| 917 | MW449369.1 | \|Severe acute respiratory syndrome coro | ···················· | ···················· | ···················· | ···················· | ···················· | ···················· |
| 918 | MW449370.1 | \|Severe acute respiratory syndrome coro | ···················· | ·A·················· | ···················· | ···················· | ···················· | ···················· |
| 919 | MW449372.1 | \|Severe acute respiratory syndrome coro | ···················· | ···················· | ···················· | ···················· | ···················· | ···················· |
| 920 | MW449373.1 | \|Severe acute respiratory syndrome coro | ···················· | ···················· | ···················· | ···················· | ···················· | ···················· |
| 921 | MW449374.1 | \|Severe acute respiratory syndrome coro | ···················· | ···················· | ···················· | ···················· | ···················· | ···················· |
| 922 | MW449376.1 | \|Severe acute respiratory syndrome coro | ···················· | ···················· | ···················· | ···················· | ···················· | ···················· |
| 923 | MW449377.1 | \|Severe acute respiratory syndrome coro | ···················· | ···················· | ···················· | ···················· | ···················· | ···················· |
| 924 | MW449383.1 | \|Severe acute respiratory syndrome coro | ···················· | ···················· | ···················· | ···················· | ···················· | ···················· |
| 925 | MW449386.1 | \|Severe acute respiratory syndrome coro | ···················· | ···················· | ···················· | ···················· | ···················· | ···················· |
| 926 | MW449387.1 | \|Severe acute respiratory syndrome coro | ···················· | ···················· | ···················· | ···················· | ···················· | ···················· |
| 927 | MW449389.1 | \|Severe acute respiratory syndrome coro | ···················· | ···················· | ···················· | ···················· | ···················· | ···················· |
| 928 | MW449392.1 | \|Severe acute respiratory syndrome coro | ···················· | ·A·················· | ···················· | ···················· | ···················· | ···················· |
| 929 | MW449399.1 | \|Severe acute respiratory syndrome coro | ···················· | ·A·················· | ···················· | ···················· | ···················· | ···················· |

```
930 MW449403.1 |Severe acute respiratory syndrome coro   ····················  ·A··················  ····················  ····················  ····················  ····················
931 MW449406.1 |Severe acute respiratory syndrome coro   ····················  ·A··················  ····················  ····················  ····················  ····················
932 MW449407.1 |Severe acute respiratory syndrome coro   ····················  ·A··················  ····················  ····················  ····················  ····················
933 MW449408.1 |Severe acute respiratory syndrome coro   ····················  ····················  ····················  ····················  ····················  ····················
934 MW449412.1 |Severe acute respiratory syndrome coro   ····················  ·A··················  ····················  ····················  ····················  ····················
935 MW449414.1 |Severe acute respiratory syndrome coro   ····················  ····················  ····················  ····················  ····················  ····················
936 MW449416.1 |Severe acute respiratory syndrome coro   ····················  ····················  ····················  ····················  ····················  ····················
937 MW449419.1 |Severe acute respiratory syndrome coro   ····················  ····················  ····················  ····················  ····················  ····················
938 MW449421.1 |Severe acute respiratory syndrome coro   ····················  ····················  ····················  ····················  ····················  ····················
939 MW449422.1 |Severe acute respiratory syndrome coro   ····················  ····················  ····················  ····················  ····················  ····················
940 MW449428.1 |Severe acute respiratory syndrome coro   ····················  ·A··················  ····················  ····················  ····················  ····················
941 MW449429.1 |Severe acute respiratory syndrome coro   ····················  ·A··················  ····················  ····················  ····················  ····················
942 MW449431.1 |Severe acute respiratory syndrome coro   ········T···········  ·A··················  ····················  ····················  ····················  ····················
943 MW449432.1 |Severe acute respiratory syndrome coro   ····················  ····················  ····················  ····················  ····················  ····················
944 MW449434.1 |Severe acute respiratory syndrome coro   ····················  ·A··················  ····················  ····················  ····················  ····················
945 MW449438.1 |Severe acute respiratory syndrome coro   ····················  ····················  ····················  ····················  ····················  ····················
946 MW449440.1 |Severe acute respiratory syndrome coro   ····················  ·A··················  ····················  ····················  ····················  ····················
947 MW449448.1 |Severe acute respiratory syndrome coro   ····················  ·A··················  ····················  ····················  ····················  ····················
948 MW449449.1 |Severe acute respiratory syndrome coro   ····················  ····················  ····················  ····················  ····················  ····················
949 MW449452.1 |Severe acute respiratory syndrome coro   ····················  ····················  ····················  ····················  ····················  ····················
950 MW449453.1 |Severe acute respiratory syndrome coro   ····················  ·A··················  ····················  ····················  ····················  ····················
951 MW449454.1 |Severe acute respiratory syndrome coro   ····················  ····················  ····················  ····················  ····················  ····················
952 MW449455.1 |Severe acute respiratory syndrome coro   ····················  ·A··················  ····················  ····················  ····················  ····················
953 MW449457.1 |Severe acute respiratory syndrome coro   ····················  ·A··················  ····················  ····················  ····················  ····················
954 MW449459.1 |Severe acute respiratory syndrome coro   ····················  ·A··················  ····················  ····················  ····················  ····················
955 MW449466.1 |Severe acute respiratory syndrome coro   ····················  ····················  ····················  ····················  ····················  ····················
956 MW449467.1 |Severe acute respiratory syndrome coro   ····················  ·A··················  ····················  ····················  ····················  ····················
957 MW449468.1 |Severe acute respiratory syndrome coro   ····················  ····················  ····················  ····················  ····················  ····················
958 MW449470.1 |Severe acute respiratory syndrome coro   ····················  ····················  ····················  ····················  ····················  ····················
959 MW449477.1 |Severe acute respiratory syndrome coro   ····················  ····················  ····················  ····················  ····················  ····················
960 MW449478.1 |Severe acute respiratory syndrome coro   ····················  ····················  ····················  ····················  ····················  ····················
961 MW449482.1 |Severe acute respiratory syndrome coro   ····················  ····················  ····················  ····················  ····················  ····················
962 MW449494.1 |Severe acute respiratory syndrome coro   ········T···········  ·A··················  ····················  ····················  ····················  ····················
963 MW449499.1 |Severe acute respiratory syndrome coro   ····················  ·A··················  ····················  ····················  ····················  ····················
964 MW449514.1 |Severe acute respiratory syndrome coro   ····················  ·············A······  ····················  ····················  ····················  ····················
965 MW322968.1 |Severe acute respiratory syndrome coro   ····················  ····················  ····················  ····················  ····················  ····················
966 MW441903.1 |Severe acute respiratory syndrome coro   ····················  ····················  ····················  ····················  ····················  ····················
967 MW441904.1 |Severe acute respiratory syndrome coro   ····················  ····················  ····················  ····················  ····················  ····················
968 MW441905.1 |Severe acute respiratory syndrome coro   ····················  ····················  ····················  ····················  ····················  ····················
969 MW441907.1 |Severe acute respiratory syndrome coro   ····················  ····················  ····················  ····················  ····················  ····················
970 MW441908.1 |Severe acute respiratory syndrome coro   ····················  ····················  ····················  ····················  ····················  ····················
971 MW441909.1 |Severe acute respiratory syndrome coro   ····················  ····················  ····················  ····················  ····················  ····················
972 MW441910.1 |Severe acute respiratory syndrome coro   ····················  ····················  ····················  ····················  ····················  ····················
973 MW441911.1 |Severe acute respiratory syndrome coro   ····················  ····················  ····················  ····················  ····················  ····················
974 MW441912.1 |Severe acute respiratory syndrome coro   ····················  ····················  ····················  ····················  ····················  ····················
975 MW441913.1 |Severe acute respiratory syndrome coro   ····················  ····················  ····················  ····················  ····················  ····················
976 MW441914.1 |Severe acute respiratory syndrome coro   ····················  ····················  ····················  ····················  ····················  ····················
977 MW441915.1 |Severe acute respiratory syndrome coro   ····················  ····················  ····················  ····················  ····················  ····················
978 MW441916.1 |Severe acute respiratory syndrome coro   ····················  ····················  ····················  ····················  ····················  ····················
979 MW441917.1 |Severe acute respiratory syndrome coro   ····················  ····················  ····················  ····················  ····················  ····················
980 MW441918.1 |Severe acute respiratory syndrome coro   ····················  ····················  ····················  ····················  ····················  ····················
981 MW441919.1 |Severe acute respiratory syndrome coro   ····················  ····················  ····················  ····················  ····················  ····················
982 MW441920.1 |Severe acute respiratory syndrome coro   ····················  ····················  ····················  ····················  ····················  ····················
983 MW433744.1 |Severe acute respiratory syndrome coro   ····················  ····················  ····················  ····················  ····················  ····················
984 MW433746.1 |Severe acute respiratory syndrome coro   ····················  ····················  ····················  ····················  ····················  ····················
985 MW433749.1 |Severe acute respiratory syndrome coro   ····················  ····················  ····················  ····················  ····················  ····················
986 MW433750.1 |Severe acute respiratory syndrome coro   ····················  ····················  ····················  ····················  ····················  ····················
987 MW433752.1 |Severe acute respiratory syndrome coro   ····················  ····················  ····················  ····················  ····················  ····················
988 MW433757.1 |Severe acute respiratory syndrome coro   ····················  ····················  ····················  ····················  ····················  ····················
989 MW433758.1 |Severe acute respiratory syndrome coro   ····················  ····················  ····················  ····················  ····················  ····················
990 MW433759.1 |Severe acute respiratory syndrome coro   ····················  ····················  ····················  ····················  ····················  ····················
991 MW433762.1 |Severe acute respiratory syndrome coro   ····················  ·A··················  ····················  ····················  ····················  ····················
992 MW433763.1 |Severe acute respiratory syndrome coro   ····················  ····················  ····················  ····················  ····················  ····················
993 MW433764.1 |Severe acute respiratory syndrome coro   ····················  ·A··················  ····················  ····················  ····················  ····················
995 MW433769.1 |Severe acute respiratory syndrome coro   ····················  ····················  ····················  ····················  ····················  ····················
996 MW433771.1 |Severe acute respiratory syndrome coro   ····················  ····················  ····················  ····················  ····················  ····················
997 MW433772.1 |Severe acute respiratory syndrome coro   ····················  ····················  ····················  ····················  ····················  ····················
998 MW435626.1 |Severe acute respiratory syndrome coro   ····················  ·A··················  ····················  ····················  ····················  ····················
999 MW435630.1 |Severe acute respiratory syndrome coro   ····················  ····················  ····················  ····················  ····················  ····················
1000 MW435638.1 |Severe acute respiratory syndrome coro   ····················  ····················  ····················  ····················  ····················  ····················
1001 MW435642.1 |Severe acute respiratory syndrome coro   ····················  ····················  ····················  ····················  ····················  ····················
1002 MW435649.1 |Severe acute respiratory syndrome coro   ····················  ·A··················  ····················  ····················  ····················  ····················
1003 MW435658.1 |Severe acute respiratory syndrome coro   ····················  ·A··················  ····················  ····················  ····················  ····················
1004 MW435661.1 |Severe acute respiratory syndrome coro   ····················  ····················  ····················  ····················  ····················  ····················
1005 MW435666.1 |Severe acute respiratory syndrome coro   ····················  ····················  ····················  ····················  ····················  ····················
1006 MW439257.1 |Severe acute respiratory syndrome coro   ····················  ····················  ····················  ····················  ····················  ····················
1007 MW440397.1 |Severe acute respiratory syndrome coro   ····················  ·A··················  ····················  ····················  ····················  ····················
1008 MW440410.1 |Severe acute respiratory syndrome coro   ····················  ·A··················  ····················  ····················  ····················  ····················
```

```
1009 MW440433.1 |Severe acute respiratory syndrome coro  ···················· ···················· ···················· ···················· ···················· ····················
1010 MW426406.1 |Severe acute respiratory syndrome coro  ···················· ···················· ···················· ···················· ···················· ····················
1011 MW428253.1 |Severe acute respiratory syndrome coro  ···················· ···················· ···················· ···················· ···················· ····················
1012 MW430964.1 |Severe acute respiratory syndrome coro  ···················· ···················· ···················· ···················· ···················· ····················
1013 MW430965.1 |Severe acute respiratory syndrome coro  ···················· ···················· ···················· ···················· ···················· ····················
1014 MW430966.1 |Severe acute respiratory syndrome coro  ···················· ···················· ···················· ···················· ···················· ····················
1015 MW430970.1 |Severe acute respiratory syndrome coro  ···················· ···················· ···················· ···················· ···················· ····················
1016 MW430971.1 |Severe acute respiratory syndrome coro  ···················· ···················· ···················· ···················· ···················· ····················
1017 MW430972.1 |Severe acute respiratory syndrome coro  ···················· ···················· ···················· ···················· ···················· ····················
1018 MW430973.1 |Severe acute respiratory syndrome coro  ···················· ···················· ···················· ···················· ···················· ····················
1019 MW430974.1 |Severe acute respiratory syndrome coro  ···················· ···················· ···················· ···················· ···················· ····················
1020 MW430975.1 |Severe acute respiratory syndrome coro  ···················· ···················· ···················· ···················· ···················· ····················
1021 MW430976.1 |Severe acute respiratory syndrome coro  ···················· ···················· ···················· ···················· ···················· ····················
1022 MW430977.1 |Severe acute respiratory syndrome coro  ···················· ···················· ···················· ···················· ···················· ····················
1023 MW430981.1 |Severe acute respiratory syndrome coro  ···················· ···················· ···················· ···················· ···················· ····················
1024 MW430983.1 |Severe acute respiratory syndrome coro  ···················· ···················· ···················· ···················· ···················· ····················
1025 MW430984.1 |Severe acute respiratory syndrome coro  ···················· ···················· ···················· ···················· ···················· ····················
1026 MW430985.1 |Severe acute respiratory syndrome coro  ···················· ···················· ···················· ···················· ···················· ····················
1027 MW430988.1 |Severe acute respiratory syndrome coro  ···················· ···················· ···················· ···················· ···················· ····················
1028 MW430989.1 |Severe acute respiratory syndrome coro  ···················· ···················· ···················· ···················· ···················· ····················
1029 MW430990.1 |Severe acute respiratory syndrome coro  ···················· ···················· ···················· ···················· ···················· ····················
1030 MW430991.1 |Severe acute respiratory syndrome coro  ···················· ···················· ···················· ···················· ···················· ····················
1031 MT628700.1 |Severe acute respiratory syndrome coro  ···················· ···················· ···················· ···················· ···················· ····················
1032 MT628701.1 |Severe acute respiratory syndrome coro  ···················· ···················· ···················· ···················· ···················· ····················
1033 MW305250.1 |Severe acute respiratory syndrome coro  ···················· ···················· ···················· ···················· ···················· ····················
1034 MW305251.1 |Severe acute respiratory syndrome coro  ···················· ···················· ···················· ···················· ···················· ····················
1035 MW425563.1 |Severe acute respiratory syndrome coro  ···················· ···················· ···················· ···················· ···················· ····················
1036 MW425708.1 |Severe acute respiratory syndrome coro  ···················· ·A·················· ···················· ···················· ···················· ····················
1037 MW425715.1 |Severe acute respiratory syndrome coro  ···················· ···················· ···················· ···················· ···················· ····················
1038 MW425716.1 |Severe acute respiratory syndrome coro  ···················· ·············A······ ···················· ···················· ···················· ····················
1039 MW425719.1 |Severe acute respiratory syndrome coro  ···················· ···················· ···················· ···················· ···················· ····················
1040 MW425720.1 |Severe acute respiratory syndrome coro  ···················· ····T··············· ···················· ···················· ···················· ····················
1041 MW425722.1 |Severe acute respiratory syndrome coro  ···················· ···················· ···················· ···················· ···················· ····················
1042 MW425727.1 |Severe acute respiratory syndrome coro  ···················· ···················· ···················· ···················· ···················· ····················
1043 MW425735.1 |Severe acute respiratory syndrome coro  ···················· ···················· ···················· ···················· ···················· ····················
1044 MW425741.1 |Severe acute respiratory syndrome coro  ···················· ·A·················· ···················· ···················· ···················· ····················
1045 MW425743.1 |Severe acute respiratory syndrome coro  ···················· ···················· ···················· ···················· ···················· ····················
1046 MW425747.1 |Severe acute respiratory syndrome coro  ···················· ···················· ···················· ···················· ···················· ····················
1047 MW425749.1 |Severe acute respiratory syndrome coro  ···················· ···················· ···················· ···················· ···················· ····················
1048 MW425750.1 |Severe acute respiratory syndrome coro  ···················· ···················· ···················· ···················· ···················· ····················
1049 MW425751.1 |Severe acute respiratory syndrome coro  ···················· ···················· ···················· ···················· ···················· ····················
1050 MW425837.1 |Severe acute respiratory syndrome coro  ···················· ···················· ···················· ···················· ···················· ····················
1051 MW425851.1 |Severe acute respiratory syndrome coro  ···················· ···················· ···················· ···················· ···················· ····················
1052 MW425852.1 |Severe acute respiratory syndrome coro  ···················· ···················· ···················· ···················· ···················· ····················
1053 MW425853.1 |Severe acute respiratory syndrome coro  ···················· ···················· ···················· ···················· ···················· ····················
1054 MW425854.1 |Severe acute respiratory syndrome coro  ···················· ···················· ···················· ···················· ···················· ····················
1055 MW425855.1 |Severe acute respiratory syndrome coro  ···················· ···················· ···················· ···················· ···················· ····················
1056 MW425856.1 |Severe acute respiratory syndrome coro  ···················· ···················· ···················· ···················· ···················· ····················
1057 MW422884.1 |Severe acute respiratory syndrome coro  ···················· ···················· ···················· ···················· ···················· ····················
1058 MW423684.1 |Severe acute respiratory syndrome coro  ···················· ···················· ···················· ···················· ···················· ····················
1059 MW423685.1 |Severe acute respiratory syndrome coro  ···················· ···················· ···················· ···················· ···················· ····················
1060 MW423686.1 |Severe acute respiratory syndrome coro  ···················· ···················· ···················· ···················· ···················· ····················
1061 MW424824.1 |Severe acute respiratory syndrome coro  ···················· ···················· ···················· ···················· ···················· ····················
1062 MW424827.1 |Severe acute respiratory syndrome coro  ···················· ···················· ···················· ···················· ···················· ····················
1063 MW424832.1 |Severe acute respiratory syndrome coro  ···················· ···················· ···················· ···················· ···················· ····················
1064 MW424835.1 |Severe acute respiratory syndrome coro  ···················· ···················· ···················· ···················· ···················· ····················
1065 MW424836.1 |Severe acute respiratory syndrome coro  ···················· ···················· ···················· ···················· ···················· ····················
1066 MW424837.1 |Severe acute respiratory syndrome coro  ···················· ···················· ···················· ···················· ···················· ····················
1067 MW424842.1 |Severe acute respiratory syndrome coro  ···················· ···················· ···················· ···················· ···················· ····················
1068 MW424854.1 |Severe acute respiratory syndrome coro  ···················· ···················· ···················· ···················· ···················· ····················
1069 MW424857.1 |Severe acute respiratory syndrome coro  ···················· ···················· ···················· ···················· ···················· ····················
1070 MW424860.1 |Severe acute respiratory syndrome coro  ···················· ···················· ···················· ···················· ···················· ····················
1071 MW424861.1 |Severe acute respiratory syndrome coro  ···················· ·A·················· ···················· ···················· ···················· ····················
1072 MW424862.1 |Severe acute respiratory syndrome coro  ···················· ···················· ···················· ···················· ···················· ····················
1073 MW424863.1 |Severe acute respiratory syndrome coro  ···················· ···················· ···················· ···················· ···················· ····················
1074 MW424864.1 |Severe acute respiratory syndrome coro  ···················· ···················· ···················· ···················· ···················· ····················
1075 MW424865.1 |Severe acute respiratory syndrome coro  ···················· ···················· ···················· ···················· ···················· ····················
1076 MW424867.1 |Severe acute respiratory syndrome coro  ···················· ···················· ···················· ···················· ···················· ····················
1077 MW424868.1 |Severe acute respiratory syndrome coro  ···················· ···················· ···················· ···················· ···················· ····················
1078 MW424869.1 |Severe acute respiratory syndrome coro  ···················· ···················· ···················· ···················· ···················· ····················
1079 MW424871.1 |Severe acute respiratory syndrome coro  ···················· ···················· ···················· ···················· ···················· ····················
1080 MW424872.1 |Severe acute respiratory syndrome coro  ···················· ·A·················· ···················· ···················· ···················· ····················
1081 MW424873.1 |Severe acute respiratory syndrome coro  ···················· ·A·················· ···················· ···················· ···················· ····················
1082 MW424874.1 |Severe acute respiratory syndrome coro  ··A················· ···················· ···················· ···················· ···················· ····················
1083 MW424875.1 |Severe acute respiratory syndrome coro  ···················· ···················· ···················· ···················· ···················· ····················
1084 MW424876.1 |Severe acute respiratory syndrome coro  ···················· ···················· ···················· ···················· ···················· ····················
1085 MW424877.1 |Severe acute respiratory syndrome coro  ···················· ···················· ···················· ···················· ···················· ····················
1086 MW424878.1 |Severe acute respiratory syndrome coro  ···················· ···················· ···················· ···················· ···················· ····················
```

```
1087 MW424879.1 |Severe acute respiratory syndrome coro    ····················  ····················  ····················  ····················  ····················  ····················
1088 MW424882.1 |Severe acute respiratory syndrome coro    ····················  ····················  ····················  ····················  ····················  ····················
1089 MW424883.1 |Severe acute respiratory syndrome coro    ····················  ····················  ····················  ····················  ····················  ····················
1090 MW424884.1 |Severe acute respiratory syndrome coro    ····················  ····················  ····················  ····················  ····················  ····················
1091 MW424886.1 |Severe acute respiratory syndrome coro    ····················  ····················  ····················  ····················  ····················  ····················
1092 MW424892.1 |Severe acute respiratory syndrome coro    ····················  ····················  ····················  ····················  ····················  ····················
1093 MW424895.1 |Severe acute respiratory syndrome coro    ····················  ·A··················  ····················  ····················  ····················  ····················
1094 MW424897.1 |Severe acute respiratory syndrome coro    ····················  ····················  ····················  ····················  ····················  ····················
1095 MW424898.1 |Severe acute respiratory syndrome coro    ····················  ····················  ····················  ····················  ····················  ····················
1096 MW424900.1 |Severe acute respiratory syndrome coro    ····················  ····················  ····················  ····················  ····················  ····················
1097 MW424901.1 |Severe acute respiratory syndrome coro    ····················  ····················  ····················  ····················  ····················  ····················
1098 MW424903.1 |Severe acute respiratory syndrome coro    ····················  ····················  ····················  ····················  ····················  ····················
1099 MW424907.1 |Severe acute respiratory syndrome coro    ····················  ·A··················  ····················  ····················  ····················  ····················
1100 MW424908.1 |Severe acute respiratory syndrome coro    ····················  ·A··················  ····················  ····················  ····················  ····················
1101 MW424909.1 |Severe acute respiratory syndrome coro    ····················  ····················  ····················  ····················  ····················  ····················
1102 MW424912.1 |Severe acute respiratory syndrome coro    ····················  ····················  ····················  ····················  ····················  ····················
1103 MW424915.1 |Severe acute respiratory syndrome coro    ····················  ·A··················  ····················  ····················  ····················  ····················
1104 MW424916.1 |Severe acute respiratory syndrome coro    ····················  ····················  ····················  ····················  ····················  ····················
1105 MW424917.1 |Severe acute respiratory syndrome coro    ····················  ····················  ····················  ····················  ····················  ····················
1106 MW424923.1 |Severe acute respiratory syndrome coro    ····················  ····················  ····················  ····················  ····················  ····················
1107 MW424924.1 |Severe acute respiratory syndrome coro    ····················  ····················  ····················  ····················  ····················  ····················
1108 MW424926.1 |Severe acute respiratory syndrome coro    ····················  ······A·············  ····················  ····················  ····················  ····················
1109 MW424929.1 |Severe acute respiratory syndrome coro    ····················  ····················  ····················  ····················  ····················  ····················
1110 MW424936.1 |Severe acute respiratory syndrome coro    ····················  ·A··················  ····················  ····················  ····················  ····················
1111 MW424937.1 |Severe acute respiratory syndrome coro    ····················  ····················  ····················  ····················  ····················  ····················
1112 MW424939.1 |Severe acute respiratory syndrome coro    ····················  ·A··················  ····················  ····················  ····················  ····················
1113 MW424940.1 |Severe acute respiratory syndrome coro    ····················  ····················  ····················  ····················  ····················  ····················
1114 MW424941.1 |Severe acute respiratory syndrome coro    ····················  ····················  ····················  ····················  ····················  ····················
1115 MW424945.1 |Severe acute respiratory syndrome coro    ····················  ····················  ····················  ····················  ····················  ····················
1116 MW424946.1 |Severe acute respiratory syndrome coro    ····················  ····················  ····················  ····················  ····················  ····················
1117 MW424947.1 |Severe acute respiratory syndrome coro    ····················  ·A··················  ····················  ····················  ····················  ····················
1118 MW424948.1 |Severe acute respiratory syndrome coro    ····················  ····················  ····················  ····················  ····················  ····················
1119 MW424949.1 |Severe acute respiratory syndrome coro    ····················  ····················  ····················  ····················  ····················  ····················
1120 MW424951.1 |Severe acute respiratory syndrome coro    ····················  ····················  ····················  ····················  ····················  ····················
1121 MW424953.1 |Severe acute respiratory syndrome coro    ····················  ····················  ····················  ····················  ····················  ····················
1122 MW424954.1 |Severe acute respiratory syndrome coro    ····················  ····················  ····················  ····················  ····················  ····················
1123 MW424955.1 |Severe acute respiratory syndrome coro    ····················  ·A··················  ····················  ····················  ····················  ····················
1124 MW424957.1 |Severe acute respiratory syndrome coro    ····················  ····················  ····················  ····················  ····················  ····················
1125 MW424958.1 |Severe acute respiratory syndrome coro    ····················  ·A··················  ····················  ····················  ····················  ····················
1126 MW424959.1 |Severe acute respiratory syndrome coro    ····················  ····················  ····················  ····················  ····················  ····················
1127 MW424963.1 |Severe acute respiratory syndrome coro    ····················  ····················  ····················  ····················  ····················  ····················
1128 MW424967.1 |Severe acute respiratory syndrome coro    ····················  ····················  ····················  ····················  ····················  ····················
1129 MW424968.1 |Severe acute respiratory syndrome coro    ····················  ····················  ····················  ····················  ····················  ····················
1130 MW424969.1 |Severe acute respiratory syndrome coro    ····················  ·A··················  ····················  ····················  ····················  ····················
1131 MW424971.1 |Severe acute respiratory syndrome coro    ····················  ····················  ····················  ····················  ····················  ····················
1132 MW424973.1 |Severe acute respiratory syndrome coro    ····················  ····················  ····················  ····················  ····················  ····················
1133 MW424977.1 |Severe acute respiratory syndrome coro    ····················  ····················  ····················  ····················  ····················  ····················
1134 MW424978.1 |Severe acute respiratory syndrome coro    ····················  ····················  ····················  ····················  ····················  ····················
1135 MW424980.1 |Severe acute respiratory syndrome coro    ····················  ····················  ····················  ····················  ····················  ····················
1136 MW424981.1 |Severe acute respiratory syndrome coro    ····················  ····················  ····················  ····················  ····················  ····················
1137 MW424983.1 |Severe acute respiratory syndrome coro    ····················  ····················  ····················  ····················  ····················  ····················
1138 MW424984.1 |Severe acute respiratory syndrome coro    ····················  ····················  ····················  ····················  ····················  ····················
1139 MW419981.1 |Severe acute respiratory syndrome coro    ····················  ····················  ····················  ····················  ····················  ····················
1140 MW419982.1 |Severe acute respiratory syndrome coro    ····················  ····················  ····················  ····················  ····················  ····················
1141 MW419983.1 |Severe acute respiratory syndrome coro    ····················  ····················  ····················  ····················  ····················  ····················
1142 MW419984.1 |Severe acute respiratory syndrome coro    ····················  ····················  ····················  ····················  ····················  ····················
1143 MW419985.1 |Severe acute respiratory syndrome coro    ····················  ····················  ····················  ····················  ····················  ····················
1144 MW419986.1 |Severe acute respiratory syndrome coro    ····················  ····················  ····················  ····················  ····················  ····················
1145 MW419987.1 |Severe acute respiratory syndrome coro    ····················  ····················  ····················  ····················  ····················  ····················
1146 MW419988.1 |Severe acute respiratory syndrome coro    ····················  ····················  ····················  ····················  ····················  ····················
1147 MW419997.1 |Severe acute respiratory syndrome coro    ····················  ····················  ····················  ····················  ····················  ····················
1148 MW419999.1 |Severe acute respiratory syndrome coro    ····················  ····················  ····················  ····················  ····················  ····················
1149 MW420006.1 |Severe acute respiratory syndrome coro    ····················  ····················  ····················  ····················  ····················  ····················
1150 MW420012.1 |Severe acute respiratory syndrome coro    ····················  ····················  ····················  ····················  ····················  ····················
1151 MW420023.1 |Severe acute respiratory syndrome coro    ····················  ····················  ····················  ····················  ····················  ····················
1152 MW420026.1 |Severe acute respiratory syndrome coro    ····················  ····················  ····················  ····················  ····················  ····················
1153 MW420028.1 |Severe acute respiratory syndrome coro    ····················  ····················  ····················  ····················  ····················  ····················
1154 MW420032.1 |Severe acute respiratory syndrome coro    ····················  ····················  ····················  ····················  ····················  ····················
1155 MW420033.1 |Severe acute respiratory syndrome coro    ····················  ····················  ····················  ····················  ····················  ····················
1156 MW420035.1 |Severe acute respiratory syndrome coro    ····················  ····················  ····················  ····················  ····················  ····················
1157 MW420038.1 |Severe acute respiratory syndrome coro    ····················  ·A··················  ····················  ····················  ····················  ····················
1158 MW420039.1 |Severe acute respiratory syndrome coro    ····················  ·A··················  ····················  ····················  ····················  ····················
1159 MW420041.1 |Severe acute respiratory syndrome coro    ····················  ····················  ····················  ····················  ····················  ····················
1160 MW420043.1 |Severe acute respiratory syndrome coro    ····················  ····················  ····················  ····················  ····················  ····················
1161 MW420047.1 |Severe acute respiratory syndrome coro    ····················  ·A··················  ····················  ····················  ····················  ····················
1162 MW420048.1 |Severe acute respiratory syndrome coro    ····················  ·A··················  ····················  ····················  ····················  ····················
1163 MW420049.1 |Severe acute respiratory syndrome coro    ····················  ····················  ····················  ····················  ····················  ····················
1164 MW420050.1 |Severe acute respiratory syndrome coro    ····················  ····················  ····················  ····················  ····················  ····················
```

```
1165 MW420051.1 |Severe acute respiratory syndrome coro    ···················· ···················· ···················· ···················· ···················· ····················
1166 MW420054.1 |Severe acute respiratory syndrome coro    ···················· ···················· ···················· ···················· ···················· ····················
1167 MW420055.1 |Severe acute respiratory syndrome coro    ···················· ···················· ···················· ···················· ···················· ····················
1168 MW420060.1 |Severe acute respiratory syndrome coro    ···················· ···················· ···················· ···················· ···················· ····················
1169 MW420061.1 |Severe acute respiratory syndrome coro    ···················· ·A·················· ···················· ···················· ···················· ····················
1170 MW420065.1 |Severe acute respiratory syndrome coro    ···················· ···················· ···················· ···················· ···················· ····················
1171 MW420066.1 |Severe acute respiratory syndrome coro    ···················· ···················· ···················· ···················· ···················· ····················
1172 MW420068.1 |Severe acute respiratory syndrome coro    ···················· ···················· ···················· ···················· ···················· ····················
1173 MW420069.1 |Severe acute respiratory syndrome coro    ···················· ·A·················· ···················· ···················· ···················· ····················
1174 MW420071.1 |Severe acute respiratory syndrome coro    ···················· ···················· ···················· ···················· ·········A·········· ····················
1175 MW420072.1 |Severe acute respiratory syndrome coro    ···················· ···················· ···················· ···················· ···················· ····················
1176 MW420073.1 |Severe acute respiratory syndrome coro    ···················· ···················· ···················· ···················· ···················· ····················
1177 MW420074.1 |Severe acute respiratory syndrome coro    ···················· ···················· ···················· ···················· ···················· ····················
1178 MW420076.1 |Severe acute respiratory syndrome coro    ···················· ···················· ···················· ···················· ···················· ····················
1179 MW420077.1 |Severe acute respiratory syndrome coro    ···················· ···················· ···················· ···················· ···················· ····················
1180 MW420079.1 |Severe acute respiratory syndrome coro    ···················· ···················· ···················· ···················· ···················· ····················
1181 MW420080.1 |Severe acute respiratory syndrome coro    ···················· ···················· ···················· ···················· ···················· ····················
1182 MW420081.1 |Severe acute respiratory syndrome coro    ···················· ···················· ···················· ···················· ···················· ····················
1183 MW420082.1 |Severe acute respiratory syndrome coro    ···················· ···················· ···················· ···················· ···················· ····················
1184 MW420084.1 |Severe acute respiratory syndrome coro    ···················· ···················· ···················· ···················· ···················· ····················
1185 MW420085.1 |Severe acute respiratory syndrome coro    ···················· ···················· ···················· ···················· ···················· ····················
1186 MW420086.1 |Severe acute respiratory syndrome coro    ···················· ·A·················· ···················· ···················· ···················· ····················
1187 MW420088.1 |Severe acute respiratory syndrome coro    ···················· ···················· ···················· ···················· ···················· ····················
1188 MW420091.1 |Severe acute respiratory syndrome coro    ···················· ·A·················· ···················· ···················· ···················· ····················
1189 MW420092.1 |Severe acute respiratory syndrome coro    ···················· ···················· ···················· ···················· ···················· ····················
1190 MW420095.1 |Severe acute respiratory syndrome coro    ···················· ···················· ···················· ···················· ···················· ····················
1191 MW420096.1 |Severe acute respiratory syndrome coro    ···················· ···················· ···················· ···················· ···················· ····················
1192 MW420097.1 |Severe acute respiratory syndrome coro    ···················· ···················· ···················· ···················· ···················· ····················
1193 MW420098.1 |Severe acute respiratory syndrome coro    ···················· ·A·················· ···················· ···················· ···················· ····················
1194 MW420099.1 |Severe acute respiratory syndrome coro    ············T······· ···················· ···················· ···················· ···················· ····················
1195 MW420100.1 |Severe acute respiratory syndrome coro    ···················· ···················· ···················· ···················· ···················· ····················
1196 MW420103.1 |Severe acute respiratory syndrome coro    ···················· ···················· ···················· ···················· ···················· ····················
1197 MW420104.1 |Severe acute respiratory syndrome coro    ···················· ·A·················· ···················· ···················· ···················· ····················
1198 MW420105.1 |Severe acute respiratory syndrome coro    ···················· ···················· ···················· ···················· ···················· ····················
1199 MW420109.1 |Severe acute respiratory syndrome coro    ···················· ·A·················· ···················· ···················· ···················· ····················
1200 MW420110.1 |Severe acute respiratory syndrome coro    ···················· ···················· ···················· ···················· ···················· ····················
1201 MW420111.1 |Severe acute respiratory syndrome coro    ···················· ·A·················· ···················· ···················· ···················· ····················
1202 MW420115.1 |Severe acute respiratory syndrome coro    ···················· ···················· ···················· ···················· ···················· ····················
1203 MW420121.1 |Severe acute respiratory syndrome coro    ···················· ·A·················· ···················· ···················· ···················· ····················
1204 MW420122.1 |Severe acute respiratory syndrome coro    ···················· ·A·················· ···················· ···················· ···················· ····················
1205 MW420123.1 |Severe acute respiratory syndrome coro    ···················· ···················· ···················· ···················· ···················· ····················
1206 MW420127.1 |Severe acute respiratory syndrome coro    ···················· ···················· ···················· ···················· ···················· ····················
1207 MW420128.1 |Severe acute respiratory syndrome coro    ···················· ···················· ···················· ···················· ···················· ····················
1208 MW420130.1 |Severe acute respiratory syndrome coro    ···················· ···················· ···················· ···················· ···················· ····················
1209 MW420131.1 |Severe acute respiratory syndrome coro    ···················· ·A·················· ···················· ···················· ···················· ····················
1210 MW420137.1 |Severe acute respiratory syndrome coro    ···················· ·A·················· ···················· ···················· ···················· ····················
1211 MW420138.1 |Severe acute respiratory syndrome coro    ···················· ···················· ···················· ···················· ···················· ····················
1212 MW420140.1 |Severe acute respiratory syndrome coro    ···················· ···················· ···················· ···················· ···················· ····················
1213 MW420141.1 |Severe acute respiratory syndrome coro    ···················· ·A·················· ···················· ···················· ···················· ····················
1214 MW420143.1 |Severe acute respiratory syndrome coro    ···················· ···················· ···················· ···················· ···················· ····················
1215 MW420145.1 |Severe acute respiratory syndrome coro    ···················· ···················· ···················· ···················· ···················· ····················
1216 MW420146.1 |Severe acute respiratory syndrome coro    ···················· ···················· ···················· ···················· ···················· ····················
1217 MW420147.1 |Severe acute respiratory syndrome coro    ···················· ···················· ···················· ···················· ···················· ····················
1218 MW420148.1 |Severe acute respiratory syndrome coro    ···················· ·A·················· ···················· ···················· ···················· ····················
1219 MW420149.1 |Severe acute respiratory syndrome coro    ···················· ···················· ···················· ···················· ···················· ···············A····
1220 MW420150.1 |Severe acute respiratory syndrome coro    ···················· ···················· ···················· ···················· ···················· ····················
1221 MW420151.1 |Severe acute respiratory syndrome coro    ···················· ·A·················· ···················· ···················· ···················· ····················
1222 MW420154.1 |Severe acute respiratory syndrome coro    ···················· ·A···A·············· ···················· ···················· ···················· ····················
1223 MW420155.1 |Severe acute respiratory syndrome coro    ···················· ···················· ···················· ···················· ···················· ····················
1224 MW420156.1 |Severe acute respiratory syndrome coro    ···················· ···················· ···················· ···················· ···················· ····················
1225 MW420157.1 |Severe acute respiratory syndrome coro    ···················· ·A·················· ···················· ···················· ···················· ····················
1226 MW420158.1 |Severe acute respiratory syndrome coro    ···················· ···················· ···················· ···················· ···················· ····················
1227 MW420160.1 |Severe acute respiratory syndrome coro    ···················· ···················· ···················· ···················· ···················· ····················
1228 MW420161.1 |Severe acute respiratory syndrome coro    ···················· ···················· ···················· ···················· ···················· ····················
1229 MW420163.1 |Severe acute respiratory syndrome coro    ···················· ·A·················· ···················· ···················· ···················· ····················
1230 MW420164.1 |Severe acute respiratory syndrome coro    ···················· ·A·················· ···················· ···················· ···················· ····················
1231 MW420165.1 |Severe acute respiratory syndrome coro    ···················· ···················· ···················· ···················· ···················· ····················
1232 MW420167.1 |Severe acute respiratory syndrome coro    ···················· ···················· ···················· ···················· ···················· ····················
1233 MW420168.1 |Severe acute respiratory syndrome coro    ···················· ·A·················· ···················· ···················· ···················· ····················
1234 MW420172.1 |Severe acute respiratory syndrome coro    ···················· ···················· ···················· ···················· ···················· ····················
1235 MW420175.1 |Severe acute respiratory syndrome coro    ···················· ···················· ···················· ···················· ···················· ····················
1236 MW420179.1 |Severe acute respiratory syndrome coro    ···················· ·A·················· ···················· ···················· ···················· ····················
1237 MW420180.1 |Severe acute respiratory syndrome coro    ···················· ···················· ···················· ···················· ···················· ····················
1238 MW420181.1 |Severe acute respiratory syndrome coro    ···················· ···················· ···················· ···················· ···················· ····················
1239 MW420182.1 |Severe acute respiratory syndrome coro    ···················· ···················· ···················· ···················· ···················· ····················
1240 MW420184.1 |Severe acute respiratory syndrome coro    ···················· ···················· ···················· ···················· ···················· ····················
1241 MW420187.1 |Severe acute respiratory syndrome coro    ···················· ···················· ···················· ···················· ···················· ····················
1242 MW420189.1 |Severe acute respiratory syndrome coro    ···················· ···················· ···················· ···················· ···················· ···············A····
```

```
1243 MW420190.1 |Severe acute respiratory syndrome coro   ····················  ····················  ····················  ····················  ····················  ····················
1244 MW420192.1 |Severe acute respiratory syndrome coro   ····················  ·A··················  ····················  ····················  ····················  ····················
1245 MW420194.1 |Severe acute respiratory syndrome coro   ····················  ····················  ····················  ····················  ····················  ····················
1246 MW420195.1 |Severe acute respiratory syndrome coro   ····················  ····················  ····················  ····················  ····················  ····················
1247 MW420196.1 |Severe acute respiratory syndrome coro   ····················  ····················  ····················  ····················  ····················  ····················
1248 MW420198.1 |Severe acute respiratory syndrome coro   ····················  ····················  ····················  ····················  ····················  ····················
1249 MW420200.1 |Severe acute respiratory syndrome coro   ····················  ····················  ····················  ····················  ····················  ····················
1250 MW420201.1 |Severe acute respiratory syndrome coro   ····················  ····················  ····················  ····················  ····················  ····················
1251 MW420202.1 |Severe acute respiratory syndrome coro   ····················  ····················  ····················  ····················  ····················  ····················
1252 MW420204.1 |Severe acute respiratory syndrome coro   ····················  ····················  ····················  ····················  ····················  ····················
1253 MW420211.1 |Severe acute respiratory syndrome coro   ····················  ····················  ····················  ····················  ····················  ····················
1254 MW420214.1 |Severe acute respiratory syndrome coro   ····················  ·A··················  ····················  ····················  ····················  ····················
1255 MW420215.1 |Severe acute respiratory syndrome coro   ····················  ····················  ····················  ····················  ····················  ····················
1256 MW420217.1 |Severe acute respiratory syndrome coro   ····················  ····················  ····················  ····················  ····················  ····················
1257 MW420218.1 |Severe acute respiratory syndrome coro   ····················  ·A··················  ····················  ····················  ····················  ····················
1258 MW420219.1 |Severe acute respiratory syndrome coro   ····················  ····················  ····················  ····················  ····················  ····················
1259 MW420220.1 |Severe acute respiratory syndrome coro   ····················  ····················  ····················  ····················  ····················  ····················
1260 MW420221.1 |Severe acute respiratory syndrome coro   ····················  ····················  ····················  ····················  ····················  ····················
1261 MW420223.1 |Severe acute respiratory syndrome coro   ····················  ·A··················  ····················  ····················  ····················  ····················
1262 MW420226.1 |Severe acute respiratory syndrome coro   ····················  ····················  ····················  ····················  ····················  ····················
1263 MW420227.1 |Severe acute respiratory syndrome coro   ····················  ····················  ····················  ····················  ·····T··············  ····················
1264 MW420228.1 |Severe acute respiratory syndrome coro   ····················  ·A··················  ····················  ····················  ····················  ····················
1265 MW420229.1 |Severe acute respiratory syndrome coro   ····················  ····················  ····················  ····················  ····················  ····················
1266 MW420231.1 |Severe acute respiratory syndrome coro   ····················  ····················  ····················  ····················  ····················  ····················
1267 MW420232.1 |Severe acute respiratory syndrome coro   ····················  ·A··················  ····················  ····················  ····················  ····················
1268 MW420234.1 |Severe acute respiratory syndrome coro   ····················  ····················  ····················  ····················  ····················  ····················
1269 MW420235.1 |Severe acute respiratory syndrome coro   ····················  ·A··················  ····················  ····················  ····················  ····················
1270 MW420237.1 |Severe acute respiratory syndrome coro   ····················  ····················  ····················  ····················  ····················  ····················
1271 MW420238.1 |Severe acute respiratory syndrome coro   ····················  ····················  ····················  ····················  ····················  ····················
1272 MW420240.1 |Severe acute respiratory syndrome coro   ····················  ····················  ····················  ····················  ····················  ····················
1273 MW420241.1 |Severe acute respiratory syndrome coro   ····················  ····················  ····················  ····················  ····················  ····················
1274 MW420242.1 |Severe acute respiratory syndrome coro   ····················  ····················  ····················  ····················  ····················  ····················
1275 MW420243.1 |Severe acute respiratory syndrome coro   ····················  ····················  ····················  ····················  ····················  ····················
1276 MW420244.1 |Severe acute respiratory syndrome coro   ····················  ····················  ····················  ····················  ····················  ····················
1277 MW420245.1 |Severe acute respiratory syndrome coro   ····················  ·A··················  ····················  ····················  ····················  ····················
1278 MW420246.1 |Severe acute respiratory syndrome coro   ····················  ····················  ····················  ····················  ····················  ····················
1279 MW420247.1 |Severe acute respiratory syndrome coro   ····················  ····················  ····················  ····················  ····················  ····················
1280 MW420248.1 |Severe acute respiratory syndrome coro   ····················  ····················  ····················  ····················  ····················  ····················
1281 MW420249.1 |Severe acute respiratory syndrome coro   ····················  ····················  ····················  ····················  ····················  ····················
1282 MW420251.1 |Severe acute respiratory syndrome coro   ····················  ····················  ····················  ····················  ····················  ····················
1283 MW420252.1 |Severe acute respiratory syndrome coro   ····················  ····················  ····················  ····················  ····················  ····················
1284 MW420254.1 |Severe acute respiratory syndrome coro   ····················  ····················  ····················  ····················  ····················  ····················
1285 MW420256.1 |Severe acute respiratory syndrome coro   ····················  ·A··················  ····················  ····················  ····················  ····················
1286 MW420257.1 |Severe acute respiratory syndrome coro   ····················  ····················  ····················  ····················  ····················  ····················
1287 MW420258.1 |Severe acute respiratory syndrome coro   ····················  ····················  ····················  ····················  ····················  ····················
1288 MW420261.1 |Severe acute respiratory syndrome coro   ····················  ····················  ····················  ····················  ····················  ····················
1289 MW420262.1 |Severe acute respiratory syndrome coro   ····················  ····················  ····················  ····················  ····················  ····················
1290 MW420264.1 |Severe acute respiratory syndrome coro   ····················  ····················  ····················  ····················  ····················  ····················
1291 MW420266.1 |Severe acute respiratory syndrome coro   ····················  ····················  ····················  ····················  ····················  ····················
1292 MW420267.1 |Severe acute respiratory syndrome coro   ····················  ····················  ····················  ····················  ····················  ····················
1293 MW420270.1 |Severe acute respiratory syndrome coro   ····················  ·A··················  ····················  ····················  ····················  ····················
1294 MW420271.1 |Severe acute respiratory syndrome coro   ····················  ····················  ····················  ····················  ····················  ····················
1295 MW420272.1 |Severe acute respiratory syndrome coro   ····················  ····················  ····················  ····················  ····················  ····················
1296 MW420273.1 |Severe acute respiratory syndrome coro   ····················  ····················  ····················  ····················  ····················  ····················
1297 MW420274.1 |Severe acute respiratory syndrome coro   ····················  ·A··················  ····················  ····················  ····················  ····················
1298 MW420275.1 |Severe acute respiratory syndrome coro   ····················  ·A··················  ····················  ····················  ····················  ····················
1299 MW420276.1 |Severe acute respiratory syndrome coro   ····················  ····················  ····················  ····················  ····················  ····················
1300 MW420278.1 |Severe acute respiratory syndrome coro   ····················  ·A··················  ····················  ····················  ····················  ····················
1301 MW420279.1 |Severe acute respiratory syndrome coro   ····················  ····················  ····················  ····················  ····················  ····················
1302 MW420280.1 |Severe acute respiratory syndrome coro   ····················  ·A··················  ····················  ····················  ····················  ····················
1303 MW420282.1 |Severe acute respiratory syndrome coro   ····················  ····················  ····················  ····················  ····················  ····················
1304 MW420283.1 |Severe acute respiratory syndrome coro   ····················  ····················  ····················  ····················  ····················  ····················
1305 MW420284.1 |Severe acute respiratory syndrome coro   ····················  ····················  ····················  ····················  ····················  ····················
1306 MW420285.1 |Severe acute respiratory syndrome coro   ····················  ····················  ····················  ····················  ····················  ····················
1307 MW420286.1 |Severe acute respiratory syndrome coro   ····················  ·A··················  ····················  ····················  ····················  ····················
1308 MW420288.1 |Severe acute respiratory syndrome coro   ····················  ····················  ····················  ····················  ····················  ····················
1309 MW420291.1 |Severe acute respiratory syndrome coro   ····················  ····················  ····················  ····················  ····················  ····················
1310 MW420294.1 |Severe acute respiratory syndrome coro   ····················  ····················  ····················  ····················  ····················  ····················
1311 MW420295.1 |Severe acute respiratory syndrome coro   ····················  ····················  ····················  ····················  ····················  ····················
1312 MW420296.1 |Severe acute respiratory syndrome coro   ····················  ·A··················  ····················  ····················  ····················  ····················
1313 MW420297.1 |Severe acute respiratory syndrome coro   ····················  ····················  ····················  ····················  ····················  ····················
1314 MW420298.1 |Severe acute respiratory syndrome coro   ····················  ····················  ····················  ····················  ····················  ····················
1315 MW420300.1 |Severe acute respiratory syndrome coro   ····················  ····················  ····················  ····················  ····················  ····················
1316 MW420304.1 |Severe acute respiratory syndrome coro   ····················  ····················  ····················  ····················  ····················  ····················
1317 MW420309.1 |Severe acute respiratory syndrome coro   ····················  ····················  ····················  ····················  ····················  ····················
1318 MW420310.1 |Severe acute respiratory syndrome coro   ····················  ····················  ····················  ····················  ····················  ····················
1319 MW420312.1 |Severe acute respiratory syndrome coro   ····················  ····················  ····················  ····················  ····················  ····················
1320 MW420314.1 |Severe acute respiratory syndrome coro   ····················  ····················  ····················  ····················  ····················  ····················
```

| | | | | | | | | |
|---|---|---|---|---|---|---|---|---|
| 1321 | MW420316.1 | \|Severe acute respiratory syndrome coro | .................... | .................... | .................... | .................... | .................... | .................... |
| 1322 | MW420319.1 | \|Severe acute respiratory syndrome coro | .................... | .................... | .................... | .................... | .................... | .................... |
| 1323 | MW420325.1 | \|Severe acute respiratory syndrome coro | .................... | .A.................. | .................... | .................... | .................... | .................... |
| 1324 | MW420327.1 | \|Severe acute respiratory syndrome coro | .................... | .................... | .................... | .................... | .................... | .................... |
| 1325 | MW420328.1 | \|Severe acute respiratory syndrome coro | .................... | .A.................. | .................... | .................... | .................... | .................... |
| 1326 | MW420329.1 | \|Severe acute respiratory syndrome coro | .................... | .................... | .................... | .................... | .................... | .................... |
| 1327 | MW420330.1 | \|Severe acute respiratory syndrome coro | .................... | .................... | .................... | .................... | .................... | .................... |
| 1328 | MW420334.1 | \|Severe acute respiratory syndrome coro | .................... | .................... | .................... | .................... | .................... | .................... |
| 1329 | MW420336.1 | \|Severe acute respiratory syndrome coro | .................... | .................... | .................... | .................... | .................... | .................... |
| 1330 | MW420337.1 | \|Severe acute respiratory syndrome coro | .................... | .................... | .................... | .................... | .................... | .................... |
| 1331 | MW420339.1 | \|Severe acute respiratory syndrome coro | .................... | .A.................. | .................... | .................... | .................... | .................... |
| 1332 | MW420342.1 | \|Severe acute respiratory syndrome coro | .................... | .................... | .................... | .................... | .................... | .................... |
| 1333 | MW420343.1 | \|Severe acute respiratory syndrome coro | .................... | .................... | .................... | .................... | .................... | .................... |
| 1334 | MW420346.1 | \|Severe acute respiratory syndrome coro | .................... | .A.................. | .................... | .................... | .................... | .................... |
| 1335 | MW420347.1 | \|Severe acute respiratory syndrome coro | .................... | .................... | .................... | .................... | .................... | .................... |
| 1336 | MW420349.1 | \|Severe acute respiratory syndrome coro | .................... | .................... | .................... | .................... | .................... | .................... |
| 1337 | MW420350.1 | \|Severe acute respiratory syndrome coro | .................... | .................... | .................... | .................... | .................... | .................... |
| 1338 | MW420355.1 | \|Severe acute respiratory syndrome coro | .................... | .................... | .................... | .................... | .................... | .................... |
| 1339 | MW420356.1 | \|Severe acute respiratory syndrome coro | .................... | .................... | .................... | .................... | .................... | .................... |
| 1340 | MW420357.1 | \|Severe acute respiratory syndrome coro | .................... | .................... | .................... | .................... | .................... | .................... |
| 1341 | MW420361.1 | \|Severe acute respiratory syndrome coro | .................... | .A.................. | .................... | .................... | .................... | .................... |
| 1342 | MW420362.1 | \|Severe acute respiratory syndrome coro | .................... | .................... | .................... | .................... | .................... | .................... |
| 1343 | MW420366.1 | \|Severe acute respiratory syndrome coro | .................... | .................... | .................... | .................... | .................... | .................... |
| 1344 | MW420367.1 | \|Severe acute respiratory syndrome coro | .................... | .................... | .................... | .................... | .................... | .................... |
| 1345 | MW420368.1 | \|Severe acute respiratory syndrome coro | .................... | .................... | .................... | .................... | .................... | .................... |
| 1346 | MW420370.1 | \|Severe acute respiratory syndrome coro | .................... | .................... | .................... | .................... | .................... | .................... |
| 1347 | MW420371.1 | \|Severe acute respiratory syndrome coro | .................... | .................... | .................... | .................... | .................... | .................... |
| 1348 | MW420372.1 | \|Severe acute respiratory syndrome coro | .................... | .................... | .................... | .................... | .................... | .................... |
| 1349 | MW420373.1 | \|Severe acute respiratory syndrome coro | .................... | .................... | .................... | .................... | .................... | .................... |
| 1350 | MW420374.1 | \|Severe acute respiratory syndrome coro | .................... | .................... | .................... | .................... | .................... | .................... |
| 1351 | MW420375.1 | \|Severe acute respiratory syndrome coro | .................... | .................... | .................... | .................... | .................... | .................... |
| 1352 | MW420377.1 | \|Severe acute respiratory syndrome coro | .................... | .................... | .................... | .................... | .................... | .................... |
| 1353 | MW420378.1 | \|Severe acute respiratory syndrome coro | .................... | .................... | .................... | .................... | .................... | .................... |
| 1354 | MW420379.1 | \|Severe acute respiratory syndrome coro | .................... | .A.................. | .................... | .................... | .................... | .................... |
| 1355 | MW420381.1 | \|Severe acute respiratory syndrome coro | .................... | .................... | .................... | .................... | .................... | .................... |
| 1356 | MW420382.1 | \|Severe acute respiratory syndrome coro | .................... | .A.................. | .................... | .................... | .................... | .................... |
| 1357 | MW420383.1 | \|Severe acute respiratory syndrome coro | .................... | .................... | .................... | .................... | .................... | .................... |
| 1358 | MW420385.1 | \|Severe acute respiratory syndrome coro | .................... | .................... | .................... | .................... | .................... | .................... |
| 1359 | MW420386.1 | \|Severe acute respiratory syndrome coro | .................... | .................... | .................... | .................... | .................... | .................... |
| 1360 | MW420387.1 | \|Severe acute respiratory syndrome coro | .................... | .A.................. | .................... | .................... | .................... | .................... |
| 1361 | MW420388.1 | \|Severe acute respiratory syndrome coro | .................... | .A.................. | .................... | .................... | .................... | .................... |
| 1362 | MW420390.1 | \|Severe acute respiratory syndrome coro | .................... | .................... | .................... | .................... | .................... | .................... |
| 1363 | MW420392.1 | \|Severe acute respiratory syndrome coro | .................... | .................... | .................... | .................... | .................... | .................... |
| 1364 | MW420393.1 | \|Severe acute respiratory syndrome coro | .................... | .................... | .................... | .................... | .................... | .................... |
| 1365 | MW420395.1 | \|Severe acute respiratory syndrome coro | .................... | .................... | .................... | .................... | .................... | .................... |
| 1366 | MW420396.1 | \|Severe acute respiratory syndrome coro | .................... | .................... | .................... | .................... | .................... | .................... |
| 1367 | MW420398.1 | \|Severe acute respiratory syndrome coro | .................... | .................... | .................... | .................... | .................... | .................... |
| 1368 | MW420399.1 | \|Severe acute respiratory syndrome coro | .................... | .................... | .................... | .................... | .................... | .................... |
| 1369 | MW420401.1 | \|Severe acute respiratory syndrome coro | .................... | .A.................. | .................... | .................... | .................... | .................... |
| 1370 | MW420402.1 | \|Severe acute respiratory syndrome coro | .................... | .................... | .................... | .................... | .................... | .................... |
| 1371 | MW420403.1 | \|Severe acute respiratory syndrome coro | .................... | .................... | .................... | .................... | .................... | .................... |
| 1372 | MW420404.1 | \|Severe acute respiratory syndrome coro | .................... | .................... | .................... | .................... | .................... | .................... |
| 1373 | MW420405.1 | \|Severe acute respiratory syndrome coro | .................... | .................... | .................... | .................... | .................... | .................... |
| 1374 | MW420406.1 | \|Severe acute respiratory syndrome coro | .................... | .................... | .................... | .................... | .................... | .................... |
| 1375 | MW420407.1 | \|Severe acute respiratory syndrome coro | .................... | .................... | .................... | .................... | .................... | .................... |
| 1376 | MW420408.1 | \|Severe acute respiratory syndrome coro | .................... | .................... | .................... | .................... | .................... | .................... |
| 1377 | MW420409.1 | \|Severe acute respiratory syndrome coro | .................... | .................... | .................... | .................... | .................... | .................... |
| 1378 | MW420410.1 | \|Severe acute respiratory syndrome coro | .................... | .................... | .................... | .................... | .................... | .................... |
| 1379 | MW420411.1 | \|Severe acute respiratory syndrome coro | .................... | .................... | .................... | ............A....... | .................... | .................... |
| 1380 | MW420414.1 | \|Severe acute respiratory syndrome coro | .................... | .................... | .................... | .................... | .................... | .................... |
| 1381 | MW420415.1 | \|Severe acute respiratory syndrome coro | .................... | .A.................. | .................... | .................... | .................... | .................... |
| 1382 | MW420416.1 | \|Severe acute respiratory syndrome coro | .................... | .A.................. | .................... | .................... | .................... | .................... |
| 1383 | MW420417.1 | \|Severe acute respiratory syndrome coro | .................... | .A.................. | .................... | .................... | .................... | .................... |
| 1384 | MW420418.1 | \|Severe acute respiratory syndrome coro | .................... | .................... | .................... | .................... | .................... | .................... |
| 1385 | MW420419.1 | \|Severe acute respiratory syndrome coro | .................... | .A.................. | .................... | .................... | .................... | .................... |
| 1386 | MW420422.1 | \|Severe acute respiratory syndrome coro | .................... | .................... | .................... | .................... | .................... | .................... |
| 1387 | MW420425.1 | \|Severe acute respiratory syndrome coro | .................... | .................... | .................... | .................... | .................... | .................... |
| 1388 | MW420427.1 | \|Severe acute respiratory syndrome coro | .................... | .................... | .................... | .................... | .................... | .................... |
| 1389 | MW420428.1 | \|Severe acute respiratory syndrome coro | .................... | .................... | .................... | .................... | .................... | .................... |
| 1390 | MW420429.1 | \|Severe acute respiratory syndrome coro | .................... | .................... | .................... | .................... | .................... | .................... |
| 1391 | MW420431.1 | \|Severe acute respiratory syndrome coro | .................... | .................... | .................... | .................... | .................... | .................... |
| 1392 | MW420433.1 | \|Severe acute respiratory syndrome coro | .................... | .A.................. | .................... | .................... | .................... | .................... |
| 1393 | MW420434.1 | \|Severe acute respiratory syndrome coro | .................... | .................... | .................... | .................... | .................... | .................... |
| 1394 | MW420435.1 | \|Severe acute respiratory syndrome coro | .................... | .................... | .................... | .................... | .................... | .................... |
| 1395 | MW420441.1 | \|Severe acute respiratory syndrome coro | .................... | .................... | .................... | .................... | .................... | .................... |
| 1396 | MW420442.1 | \|Severe acute respiratory syndrome coro | .................... | .A...A.............. | .................... | .................... | .................... | .................... |
| 1397 | MW420443.1 | \|Severe acute respiratory syndrome coro | .................... | .................... | .................... | .................... | .................... | .................... |
| 1398 | MW420444.1 | \|Severe acute respiratory syndrome coro | .................... | .................... | .................... | .................... | .................... | .................... |

| | | | | | | | | |
|---|---|---|---|---|---|---|---|---|
| 1399 | MW420445.1 | \|Severe acute respiratory syndrome coro | ···················· | ·A·················· | ···················· | ···················· | ···················· | ···················· |
| 1400 | MW420447.1 | \|Severe acute respiratory syndrome coro | ···················· | ·A·················· | ···················· | ···················· | ···················· | ···················· |
| 1401 | MW420449.1 | \|Severe acute respiratory syndrome coro | ···················· | ·A·················· | ···················· | ···················· | ···················· | ···················· |
| 1402 | MW420451.1 | \|Severe acute respiratory syndrome coro | ···················· | ···················· | ···················· | ···················· | ···················· | ···················· |
| 1403 | MW420452.1 | \|Severe acute respiratory syndrome coro | ···················· | ···················· | ···················· | ···················· | ···················· | ···················· |
| 1404 | MW420457.1 | \|Severe acute respiratory syndrome coro | ···················· | ···················· | ···················· | ···················· | ···················· | ···················· |
| 1405 | MW420458.1 | \|Severe acute respiratory syndrome coro | ···················· | ···················· | ···················· | ···················· | ···················· | ···················· |
| 1406 | MW420459.1 | \|Severe acute respiratory syndrome coro | ···················· | ·A·················· | ···················· | ···················· | ···················· | ···················· |
| 1407 | MW420460.1 | \|Severe acute respiratory syndrome coro | ···················· | ·A·················· | ···················· | ···················· | ···················· | ···················· |
| 1408 | MW420462.1 | \|Severe acute respiratory syndrome coro | ···················· | ·A·················· | ···················· | ···················· | ···················· | ···················· |
| 1409 | MW420463.1 | \|Severe acute respiratory syndrome coro | ···················· | ···················· | ···················· | ···················· | ···················· | ···················· |
| 1410 | MW420464.1 | \|Severe acute respiratory syndrome coro | ···················· | ···················· | ···················· | ···················· | ···················· | ···················· |
| 1411 | MW420465.1 | \|Severe acute respiratory syndrome coro | ···················· | ·A·················· | ···················· | ···················· | ···················· | ···················· |
| 1412 | MW420467.1 | \|Severe acute respiratory syndrome coro | ···················· | ···················· | ···················· | ···················· | ···················· | ···················· |
| 1413 | MW420468.1 | \|Severe acute respiratory syndrome coro | ···················· | ···················· | ···················· | ···················· | ···················· | ···················· |
| 1414 | MW420470.1 | \|Severe acute respiratory syndrome coro | ···················· | ···················· | ···················· | ···················· | ···················· | ···················· |
| 1415 | MW420471.1 | \|Severe acute respiratory syndrome coro | ···················· | ···················· | ···················· | ···················· | ···················· | ···················· |
| 1416 | MW420473.1 | \|Severe acute respiratory syndrome coro | ···················· | ···················· | ···················· | ···················· | ···················· | ···················· |
| 1417 | MW420477.1 | \|Severe acute respiratory syndrome coro | ···················· | ···················· | ···················· | ···················· | ···················· | ···················· |
| 1418 | MW420479.1 | \|Severe acute respiratory syndrome coro | ···················· | ·A·················· | ···················· | ···················· | ···················· | ···················· |
| 1419 | MW420480.1 | \|Severe acute respiratory syndrome coro | ···················· | ···················· | ···················· | ···················· | ···················· | ···················· |
| 1420 | MW420481.1 | \|Severe acute respiratory syndrome coro | ···················· | ···················· | ···················· | ···················· | ···················· | ···················· |
| 1421 | MW420482.1 | \|Severe acute respiratory syndrome coro | ···················· | ·A·················· | ···················· | ···················· | ···················· | ···················· |
| 1422 | MW420483.1 | \|Severe acute respiratory syndrome coro | ···················· | ···················· | ···················· | ···················· | ···················· | ···················· |
| 1423 | MW420487.1 | \|Severe acute respiratory syndrome coro | ···················· | ·A·················· | ···················· | ···················· | ···················· | ···················· |
| 1424 | MW420488.1 | \|Severe acute respiratory syndrome coro | ···················· | ···················· | ···················· | ···················· | ···················· | ···················· |
| 1425 | MW420491.1 | \|Severe acute respiratory syndrome coro | ···················· | ···················· | ···················· | ···················· | ···················· | ···················· |
| 1426 | MW420492.1 | \|Severe acute respiratory syndrome coro | ···················· | ···················· | ···················· | ···················· | ···················· | ···················· |
| 1427 | MW420494.1 | \|Severe acute respiratory syndrome coro | ···················· | ···················· | ···················· | ···················· | ···················· | ···················· |
| 1428 | MW420495.1 | \|Severe acute respiratory syndrome coro | ···················· | ···················· | ···················· | ···················· | ···················· | ···················· |
| 1429 | MW420497.1 | \|Severe acute respiratory syndrome coro | ···················· | ···················· | ···················· | ···················· | ···················· | ···················· |
| 1430 | MW420498.1 | \|Severe acute respiratory syndrome coro | ···················· | ···················· | ···················· | ···················· | ···················· | ···················· |
| 1431 | MW420501.1 | \|Severe acute respiratory syndrome coro | ···················· | ···················· | ···················· | ···················· | ···················· | ···················· |
| 1432 | MW420503.1 | \|Severe acute respiratory syndrome coro | ···················· | ···················· | ···················· | ···················· | ···················· | ···················· |
| 1433 | MW420505.1 | \|Severe acute respiratory syndrome coro | ···················· | ···················· | ···················· | ···················· | ···················· | ···················· |
| 1434 | MW420506.1 | \|Severe acute respiratory syndrome coro | ···················· | ···················· | ···················· | ···················· | ···················· | ···················· |
| 1435 | MW420507.1 | \|Severe acute respiratory syndrome coro | ···················· | ···················· | ···················· | ···················· | ···················· | ···················· |
| 1436 | MW420512.1 | \|Severe acute respiratory syndrome coro | ···················· | ·A···A·············· | ···················· | ···················· | ···················· | ···················· |
| 1437 | MW420513.1 | \|Severe acute respiratory syndrome coro | ···················· | ·A·················· | ···················· | ···················· | ···················· | ···················· |
| 1438 | MW420514.1 | \|Severe acute respiratory syndrome coro | ···················· | ···················· | ···················· | ···················· | ···················· | ···················· |
| 1439 | MW420517.1 | \|Severe acute respiratory syndrome coro | ···················· | ···················· | ···················· | ···················· | ···················· | ···················· |
| 1440 | MW420521.1 | \|Severe acute respiratory syndrome coro | ···················· | ·A·················· | ···················· | ···················· | ···················· | ···················· |
| 1441 | MW420526.1 | \|Severe acute respiratory syndrome coro | ···················· | ·A·················· | ···················· | ···················· | ···················· | ···················· |
| 1442 | MW420530.1 | \|Severe acute respiratory syndrome coro | ···················· | ···················· | ···················· | ···················· | ···················· | ···················· |
| 1443 | MW420531.1 | \|Severe acute respiratory syndrome coro | ···················· | ·A·················· | ···················· | ···················· | ···················· | ···················· |
| 1444 | MW420532.1 | \|Severe acute respiratory syndrome coro | ···················· | ···················· | ···················· | ···················· | ···················· | ···················· |
| 1445 | MW420533.1 | \|Severe acute respiratory syndrome coro | ···················· | ·A·················· | ···················· | ···················· | ···················· | ···················· |
| 1446 | MW420535.1 | \|Severe acute respiratory syndrome coro | ···················· | ···················· | ···················· | ···················· | ···················· | ···················· |
| 1447 | MW420537.1 | \|Severe acute respiratory syndrome coro | ···················· | ···················· | ···················· | ···················· | ···················· | ···················· |
| 1448 | MW420539.1 | \|Severe acute respiratory syndrome coro | ···················· | ·A·················· | ···················· | ···················· | ···················· | ···················· |
| 1449 | MW420540.1 | \|Severe acute respiratory syndrome coro | ···················· | ···················· | ···················· | ···················· | ···················· | ···················· |
| 1450 | MW420541.1 | \|Severe acute respiratory syndrome coro | ···················· | ···················· | ···················· | ···················· | ···················· | ···················· |
| 1451 | MW420542.1 | \|Severe acute respiratory syndrome coro | ···················· | ·A·················· | ···················· | ···················· | ···················· | ···················· |
| 1452 | MW420544.1 | \|Severe acute respiratory syndrome coro | ···················· | ···················· | ···················· | ···················· | ···················· | ···················· |
| 1453 | MW420546.1 | \|Severe acute respiratory syndrome coro | ···················· | ···················· | ···················· | ···················· | ···················· | ···················· |
| 1454 | MW420547.1 | \|Severe acute respiratory syndrome coro | ···················· | ···················· | ···················· | ···················· | ···················· | ···················· |
| 1455 | MW420548.1 | \|Severe acute respiratory syndrome coro | ···················· | ···················· | ···················· | ···················· | ···················· | ···················· |
| 1456 | MW420549.1 | \|Severe acute respiratory syndrome coro | ···················· | ···················· | ···················· | ···················· | ···················· | ···················· |
| 1457 | MW420550.1 | \|Severe acute respiratory syndrome coro | ···················· | ·A·················· | ···················· | ···················· | ···················· | ···················· |
| 1458 | MW420553.1 | \|Severe acute respiratory syndrome coro | ···················· | ···················· | ···················· | ···················· | ···················· | ···················· |
| 1459 | MW420554.1 | \|Severe acute respiratory syndrome coro | ···················· | ···················· | ···················· | ···················· | ···················· | ···················· |
| 1460 | MW420557.1 | \|Severe acute respiratory syndrome coro | ···················· | ·A·················· | ···················· | ···················· | ···················· | ···················· |
| 1461 | MW420559.1 | \|Severe acute respiratory syndrome coro | ···················· | ·A·················· | ···················· | ···················· | ···················· | ···················· |
| 1462 | MW420560.1 | \|Severe acute respiratory syndrome coro | ···················· | ·A·················· | ···················· | ···················· | ···················· | ···················· |
| 1463 | MW420562.1 | \|Severe acute respiratory syndrome coro | ···················· | ···················· | ···················· | ···················· | ···················· | ···················· |
| 1464 | MW420565.1 | \|Severe acute respiratory syndrome coro | ···················· | ···················· | ···················· | ···················· | ···················· | ···················· |
| 1465 | MW420568.1 | \|Severe acute respiratory syndrome coro | ···················· | ·A·················· | ···················· | ···················· | ···················· | ···················· |
| 1466 | MW420570.1 | \|Severe acute respiratory syndrome coro | ···················· | ···················· | ···················· | ···················· | ···················· | ···················· |
| 1467 | MW420571.1 | \|Severe acute respiratory syndrome coro | ···················· | ···················· | ···················· | ···················· | ···················· | ···················· |
| 1468 | MW420572.1 | \|Severe acute respiratory syndrome coro | ···················· | ···················· | ···················· | ···················· | ···················· | ···················· |
| 1469 | MW420573.1 | \|Severe acute respiratory syndrome coro | ···················· | ···················· | ···················· | ···················· | ···················· | ···················· |
| 1470 | MW420574.1 | \|Severe acute respiratory syndrome coro | ···················· | ···················· | ···················· | ···················· | ···················· | ···················· |
| 1471 | MW420575.1 | \|Severe acute respiratory syndrome coro | ···················· | ···················· | ···················· | ···················· | ···················· | ···················· |
| 1472 | MW420576.1 | \|Severe acute respiratory syndrome coro | ···················· | ···················· | ···················· | ···················· | ···················· | ···················· |
| 1473 | MW420579.1 | \|Severe acute respiratory syndrome coro | ···················· | ···················· | ···················· | ···················· | ···················· | ···················· |
| 1474 | MW420582.1 | \|Severe acute respiratory syndrome coro | ···················· | ···················· | ···················· | ···················· | ···················· | ···················· |
| 1475 | MW420588.1 | \|Severe acute respiratory syndrome coro | ···················· | ·A·················· | ···················· | ···················· | ···················· | ···················· |
| 1476 | MW420593.1 | \|Severe acute respiratory syndrome coro | ···················· | ·A·················· | ···················· | ···················· | ···················· | ···················· |

```
1477 MW420594.1 |Severe acute respiratory syndrome coro    ····················  ····················  ····················  ····················  ····················  ····················
1478 MW420595.1 |Severe acute respiratory syndrome coro    ····················  ·A··················  ····················  ····················  ····················  ····················
1479 MW420599.1 |Severe acute respiratory syndrome coro    ····················  ····················  ····················  ····················  ····················  ····················
1480 MW420601.1 |Severe acute respiratory syndrome coro    ····················  ·A··················  ····················  ····················  ····················  ····················
1481 MW420607.1 |Severe acute respiratory syndrome coro    ····················  ····················  ····················  ····················  ····················  ····················
1482 MW420609.1 |Severe acute respiratory syndrome coro    ····················  ·A··················  ····················  ····················  ····················  ····················
1483 MW420612.1 |Severe acute respiratory syndrome coro    ····················  ·A··················  ····················  ····················  ····················  ····················
1484 MW420617.1 |Severe acute respiratory syndrome coro    ····················  ····················  ····················  ····················  ····················  ····················
1485 MW420621.1 |Severe acute respiratory syndrome coro    ····················  ····················  ····················  ····················  ····················  ····················
1486 MW420624.1 |Severe acute respiratory syndrome coro    ····················  ····················  ····················  ····················  ····················  ····················
1487 MW420626.1 |Severe acute respiratory syndrome coro    ····················  ····················  ····················  ····················  ····················  ····················
1488 MW420628.1 |Severe acute respiratory syndrome coro    ····················  ····················  ····················  ····················  ····················  ····················
1489 MW420631.1 |Severe acute respiratory syndrome coro    ····················  ·A··················  ····················  ····················  ····················  ····················
1490 MW420634.1 |Severe acute respiratory syndrome coro    ····················  ····················  ····················  ····················  ····················  ····················
1491 MW420638.1 |Severe acute respiratory syndrome coro    ····················  ····················  ····················  ····················  ····················  ····················
1492 MW420640.1 |Severe acute respiratory syndrome coro    ····················  ····················  ····················  ····················  ····················  ····················
1493 MW420644.1 |Severe acute respiratory syndrome coro    ····················  ····················  ····················  ····················  ····················  ····················
1494 MW420645.1 |Severe acute respiratory syndrome coro    ····················  ····················  ····················  ····················  ····················  ····················
1495 MW420646.1 |Severe acute respiratory syndrome coro    ····················  ····················  ····················  ····················  ····················  ····················
1496 MW420651.1 |Severe acute respiratory syndrome coro    ····················  ·A··················  ····················  ····················  ····················  ····················
1497 MW420653.1 |Severe acute respiratory syndrome coro    ····················  ·A··················  ····················  ····················  ····················  ····················
1498 MW420654.1 |Severe acute respiratory syndrome coro    ····················  ····················  ····················  ····················  ····················  ····················
1499 MW420655.1 |Severe acute respiratory syndrome coro    ····················  ·A··················  ····················  ····················  ····················  ····················
1500 MW420656.1 |Severe acute respiratory syndrome coro    ····················  ····················  ····················  ····················  ····················  ····················
1501 MW420659.1 |Severe acute respiratory syndrome coro    ····················  ····················  ····················  ····················  ····················  ····················
1502 MW420663.1 |Severe acute respiratory syndrome coro    ····················  ····················  ····················  ····················  ····················  ····················
1503 MW420664.1 |Severe acute respiratory syndrome coro    ····················  ····················  ····················  ····················  ····················  ····················
1504 MW420666.1 |Severe acute respiratory syndrome coro    ····················  ····················  ····················  ····················  ····················  ····················
1505 MW420669.1 |Severe acute respiratory syndrome coro    ····················  ····················  ····················  ····················  ····················  ····················
1506 MW420671.1 |Severe acute respiratory syndrome coro    ····················  ·A··················  ····················  ····················  ····················  ····················
1507 MW420673.1 |Severe acute respiratory syndrome coro    ····················  ·A··················  ····················  ····················  ····················  ····················
1508 MW420675.1 |Severe acute respiratory syndrome coro    ····················  ····················  ····················  ····················  ····················  ····················
1509 MW420679.1 |Severe acute respiratory syndrome coro    ····················  ····················  ····················  ····················  ····················  ····················
1510 MW420681.1 |Severe acute respiratory syndrome coro    ····················  ····················  ····················  ····················  ····················  ····················
1511 MW420683.1 |Severe acute respiratory syndrome coro    ····················  ····················  ····················  ····················  ····················  ····················
1512 MW420684.1 |Severe acute respiratory syndrome coro    ····················  ····················  ····················  ····················  ····················  ····················
1513 MW420685.1 |Severe acute respiratory syndrome coro    ····················  ····················  ····················  ····················  ····················  ····················
1514 MW420686.1 |Severe acute respiratory syndrome coro    ····················  ····················  ····················  ····················  ····················  ····················
1515 MW420687.1 |Severe acute respiratory syndrome coro    ····················  ····················  ····················  ····················  ····················  ····················
1516 MW420689.1 |Severe acute respiratory syndrome coro    ····················  ····················  ····················  ····················  ····················  ····················
1517 MW420690.1 |Severe acute respiratory syndrome coro    ····················  ····················  ····················  ····················  ····················  ····················
1518 MW420691.1 |Severe acute respiratory syndrome coro    ····················  ····················  ····················  ····················  ····················  ····················
1519 MW420693.1 |Severe acute respiratory syndrome coro    ····················  ····················  ····················  ····················  ····················  ····················
1520 MW420694.1 |Severe acute respiratory syndrome coro    ····················  ····················  ····················  ····················  ····················  ····················
1521 MW420695.1 |Severe acute respiratory syndrome coro    ····················  ····················  ····················  ····················  ····················  ····················
1522 MW420696.1 |Severe acute respiratory syndrome coro    ····················  ····················  ····················  ····················  ····················  ····················
1523 MW420698.1 |Severe acute respiratory syndrome coro    ····················  ····················  ····················  ····················  ····················  ····················
1524 MW420701.1 |Severe acute respiratory syndrome coro    ····················  ····················  ····················  ····················  ····················  ····················
1525 MW420702.1 |Severe acute respiratory syndrome coro    ····················  ····················  ····················  ····················  ····················  ····················
1526 MW420704.1 |Severe acute respiratory syndrome coro    ····················  ····················  ····················  ····················  ····················  ····················
1527 MW420705.1 |Severe acute respiratory syndrome coro    ····················  ·A··················  ····················  ····················  ····················  ····················
1528 MW420707.1 |Severe acute respiratory syndrome coro    ····················  ····················  ····················  ····················  ····················  ····················
1529 MW420708.1 |Severe acute respiratory syndrome coro    ····················  ·A··················  ····················  ····················  ····················  ····················
1530 MW420709.1 |Severe acute respiratory syndrome coro    ····················  ·A··················  ····················  ····················  ····················  ····················
1531 MW420710.1 |Severe acute respiratory syndrome coro    ····················  ····················  ····················  ····················  ····················  ····················
1532 MW420712.1 |Severe acute respiratory syndrome coro    ····················  ····················  ····················  ····················  ····················  ····················
1533 MW420716.1 |Severe acute respiratory syndrome coro    ····················  ····················  ····················  ····················  ····················  ····················
1534 MW420719.1 |Severe acute respiratory syndrome coro    ····················  ····················  ····················  ····················  ····················  ····················
1535 MW420721.1 |Severe acute respiratory syndrome coro    ····················  ····················  ····················  ····················  ····················  ····················
1536 MW420722.1 |Severe acute respiratory syndrome coro    ····················  ····················  ····················  ····················  ····················  ····················
1537 MW420723.1 |Severe acute respiratory syndrome coro    ····················  ·A··················  ····················  ····················  ····················  ····················
1538 MW420727.1 |Severe acute respiratory syndrome coro    ····················  ····················  ····················  ····················  ····················  ····················
1539 MW420728.1 |Severe acute respiratory syndrome coro    ····················  ····················  ····················  ····················  ····················  ····················
1540 MW420729.1 |Severe acute respiratory syndrome coro    ····················  ····················  ····················  ····················  ····················  ····················
1541 MW420730.1 |Severe acute respiratory syndrome coro    ····················  ····················  ····················  ····················  ····················  ····················
1542 MW420731.1 |Severe acute respiratory syndrome coro    ····················  ····················  ····················  ····················  ····················  ····················
1543 MW420732.1 |Severe acute respiratory syndrome coro    ····················  ·A··················  ····················  ····················  ····················  ····················
1544 MW420734.1 |Severe acute respiratory syndrome coro    ····················  ····················  ····················  ····················  ····················  ····················
1545 MW420735.1 |Severe acute respiratory syndrome coro    ····················  ····················  ····················  ····················  ····················  ····················
1546 MW420737.1 |Severe acute respiratory syndrome coro    ····················  ·A··················  ····················  ····················  ····················  ····················
1547 MW420740.1 |Severe acute respiratory syndrome coro    ····················  ·A··················  ····················  ····················  ····················  ····················
1548 MW420741.1 |Severe acute respiratory syndrome coro    ····················  ·A··················  ····················  ····················  ····················  ····················
1549 MW420742.1 |Severe acute respiratory syndrome coro    ····················  ····················  ····················  ····················  ····················  ····················
1550 MW420746.1 |Severe acute respiratory syndrome coro    ····················  ·A··················  ····················  ····················  ····················  ····················
1551 MW420747.1 |Severe acute respiratory syndrome coro    ····················  ····················  ····················  ····················  ····················  ····················
1552 MW420748.1 |Severe acute respiratory syndrome coro    ····················  ····················  ····················  ····················  ····················  ····················
1553 MW420749.1 |Severe acute respiratory syndrome coro    ····················  ····················  ····················  ····················  ····················  ····················
1554 MW420751.1 |Severe acute respiratory syndrome coro    ····················  ····················  ····················  ····················  ····················  ····················
```

1555 MW420756.1 |Severe acute respiratory syndrome coro ···················· ···················· ···················· ···················· ···················· ····················
1556 MW420759.1 |Severe acute respiratory syndrome coro ···················· ···················· ···················· ···················· ···················· ····················
1557 MW420761.1 |Severe acute respiratory syndrome coro ···················· ·A·················· ···················· ···················· ···················· ····················
1558 MW420762.1 |Severe acute respiratory syndrome coro ···················· ···················· ···················· ···················· ···················· ····················
1559 MW420765.1 |Severe acute respiratory syndrome coro ···················· ···················· ···················· ···················· ···················· ····················
1560 MW420766.1 |Severe acute respiratory syndrome coro ···················· ·A·················· ···················· ···················· ···················· ····················
1561 MW420769.1 |Severe acute respiratory syndrome coro ···················· ···················· ···················· ···················· ···················· ····················
1562 MW420772.1 |Severe acute respiratory syndrome coro ···················· ···················· ···················· ···················· ···················· ····················
1563 MW420774.1 |Severe acute respiratory syndrome coro ···················· ···················· ···················· ···················· ···················· ····················
1564 MW420776.1 |Severe acute respiratory syndrome coro ···················· ···················· ···················· ···················· ···················· ····················
1565 MW420777.1 |Severe acute respiratory syndrome coro ···················· ·A·················· ···················· ···················· ···················· ····················
1566 MW420778.1 |Severe acute respiratory syndrome coro ···················· ···················· ···················· ···················· ···················· ····················
1567 MW420780.1 |Severe acute respiratory syndrome coro ···················· ···················· ···················· ···················· ···················· ····················
1568 MW420781.1 |Severe acute respiratory syndrome coro ···················· ···················· ···················· ···················· ···················· ····················
1569 MW420782.1 |Severe acute respiratory syndrome coro ···················· ···················· ···················· ···················· ···················· ····················
1570 MW420783.1 |Severe acute respiratory syndrome coro ···················· ···················· ···················· ···················· ···················· ····················
1571 MW420787.1 |Severe acute respiratory syndrome coro ···················· ···················· ···················· ···················· ···················· ····················
1572 MW420789.1 |Severe acute respiratory syndrome coro ···················· ·A·················· ···················· ···················· ···················· ····················
1573 MW420791.1 |Severe acute respiratory syndrome coro ···················· ···················· ···················· ···················· ···················· ····················
1574 MW420794.1 |Severe acute respiratory syndrome coro ···················· ·A·················· ···················· ···················· ···················· ····················
1575 MW420795.1 |Severe acute respiratory syndrome coro ···················· ···················· ···················· ···················· ···················· ····················
1576 MW420796.1 |Severe acute respiratory syndrome coro ···················· ···················· ···················· ···················· ···················· ····················
1577 MW420797.1 |Severe acute respiratory syndrome coro ···················· ···················· ···················· ···················· ···················· ····················
1578 MW420798.1 |Severe acute respiratory syndrome coro ···················· ···················· ···················· ···················· ···················· ····················
1579 MW420801.1 |Severe acute respiratory syndrome coro ···················· ···················· ···················· ···················· ···················· ····················
1580 MW420802.1 |Severe acute respiratory syndrome coro ···················· ···················· ···················· ···················· ···················· ····················
1581 MW420804.1 |Severe acute respiratory syndrome coro ···················· ···················· ···················· ···················· ···················· ····················
1582 MW420805.1 |Severe acute respiratory syndrome coro ···················· ···················· ···················· ···················· ···················· ····················
1583 MW420809.1 |Severe acute respiratory syndrome coro ···················· ···················· ···················· ···················· ···················· ····················
1584 MW420814.1 |Severe acute respiratory syndrome coro ···················· ···················· ···················· ···················· ···················· ····················
1585 MW420816.1 |Severe acute respiratory syndrome coro ···················· ···················· ···················· ···················· ···················· ····················
1586 MW420818.1 |Severe acute respiratory syndrome coro ···················· ···················· ···················· ···················· ···················· ····················
1587 MW420819.1 |Severe acute respiratory syndrome coro ···················· ···················· ···················· ···················· ···················· ····················
1588 MW420826.1 |Severe acute respiratory syndrome coro ···················· ···················· ···················· ···················· ···················· ····················
1589 MW420827.1 |Severe acute respiratory syndrome coro ···················· ·A·················· ···················· ···················· ···················· ····················
1590 MW420828.1 |Severe acute respiratory syndrome coro ···················· ···················· ···················· ···················· ···················· ····················
1591 MW420829.1 |Severe acute respiratory syndrome coro ···················· ···················· ···················· ···················· ···················· ····················
1592 MW420831.1 |Severe acute respiratory syndrome coro ···················· ···················· ···················· ···················· ···················· ····················
1593 MW420832.1 |Severe acute respiratory syndrome coro ···················· ···················· ···················· ···················· ···················· ····················
1594 MW420833.1 |Severe acute respiratory syndrome coro ···················· ···················· ···················· ···················· ···················· ····················
1595 MW420835.1 |Severe acute respiratory syndrome coro ···················· ···················· ···················· ···················· ···················· ····················
1596 MW420836.1 |Severe acute respiratory syndrome coro ···················· ···················· ···················· ···················· ···················· ····················
1597 MW420838.1 |Severe acute respiratory syndrome coro ···················· ···················· ···················· ···················· ···················· ····················
1598 MW420839.1 |Severe acute respiratory syndrome coro ···················· ···················· ···················· ···················· ···················· ····················
1599 MW420842.1 |Severe acute respiratory syndrome coro ···················· ···················· ···················· ···················· ···················· ····················
1600 MW420844.1 |Severe acute respiratory syndrome coro ···················· ···················· ···················· ···················· ···················· ····················
1601 MW420846.1 |Severe acute respiratory syndrome coro ···················· ·A·················· ···················· ···················· ···················· ····················
1602 MW420849.1 |Severe acute respiratory syndrome coro ···················· ·A·················· ···················· ···················· ···················· ····················
1603 MW420851.1 |Severe acute respiratory syndrome coro ···················· ···················· ···················· ···················· ···················· ····················
1604 MW420852.1 |Severe acute respiratory syndrome coro ···················· ···················· ···················· ···················· ···················· ····················
1605 MW420854.1 |Severe acute respiratory syndrome coro ···················· ·A·················· ···················· ···················· ···················· ····················
1606 MW420856.1 |Severe acute respiratory syndrome coro ···················· ···················· ···················· ···················· ···················· ····················
1607 MW420857.1 |Severe acute respiratory syndrome coro ···················· ···················· ···················· ···················· ···················· ····················
1608 MW420858.1 |Severe acute respiratory syndrome coro ···················· ···················· ···················· ···················· ···················· ····················
1609 MW420859.1 |Severe acute respiratory syndrome coro ···················· ···················· ···················· ···················· ···················· ····················
1610 MW420860.1 |Severe acute respiratory syndrome coro ···················· ···················· ···················· ···················· ···················· ····················
1611 MW420861.1 |Severe acute respiratory syndrome coro ···················· ·A·················· ···················· ···················· ···················· ····················
1612 MW420862.1 |Severe acute respiratory syndrome coro ···················· ···················· ···················· ···················· ···················· ····················
1613 MW420864.1 |Severe acute respiratory syndrome coro ···················· ·A·················· ···················· ···················· ···················· ····················
1614 MW420865.1 |Severe acute respiratory syndrome coro ···················· ···················· ···················· ···················· ···················· ····················
1615 MW420866.1 |Severe acute respiratory syndrome coro ···················· ·A·················· ···················· ···················· ···················· ····················
1616 MW420867.1 |Severe acute respiratory syndrome coro ···················· ···················· ···················· ···················· ···················· ····················
1617 MW420868.1 |Severe acute respiratory syndrome coro ···················· ···················· ···················· ···················· ···················· ····················
1618 MW420869.1 |Severe acute respiratory syndrome coro ···················· ···················· ···················· ···················· ···················· ····················
1619 MW420870.1 |Severe acute respiratory syndrome coro ···················· ···················· ···················· ···················· ···················· ····················
1620 MW420871.1 |Severe acute respiratory syndrome coro ···················· ···················· ···················· ···················· ···················· ····················
1621 MW420874.1 |Severe acute respiratory syndrome coro ···················· ···················· ···················· ···················· ···················· ····················
1622 MW420875.1 |Severe acute respiratory syndrome coro ···················· ···················· ···················· ···················· ···················· ····················
1623 MW420876.1 |Severe acute respiratory syndrome coro ···················· ·A·················· ···················· ···················· ···················· ····················
1624 MW420877.1 |Severe acute respiratory syndrome coro ···················· ···················· ···················· ···················· ···················· ····················
1625 MW420878.1 |Severe acute respiratory syndrome coro ···················· ···················· ···················· ···················· ···················· ····················
1626 MW420880.1 |Severe acute respiratory syndrome coro ···················· ···················· ···················· ···················· ···················· ····················
1627 MW420881.1 |Severe acute respiratory syndrome coro ···················· ···················· ···················· ···················· ···················· ····················
1628 MW420882.1 |Severe acute respiratory syndrome coro ···················· ···················· ···················· ···················· ···················· ····················
1629 MW420884.1 |Severe acute respiratory syndrome coro ···················· ·A·················· ···················· ···················· ···················· ····················
1630 MW420885.1 |Severe acute respiratory syndrome coro ···················· ·A·················· ···················· ···················· ···················· ····················
1631 MW420886.1 |Severe acute respiratory syndrome coro ···················· ···················· ···················· ···················· ···················· ····················
1632 MW420888.1 |Severe acute respiratory syndrome coro ···················· ···················· ···················· ···················· ···················· ····················

1633 MW420889.1 |Severe acute respiratory syndrome coro ···················· ···················· ···················· ···················· ···················· ····················
1634 MW420890.1 |Severe acute respiratory syndrome coro ···················· ···················· ···················· ···················· ···················· ····················
1635 MW420891.1 |Severe acute respiratory syndrome coro ···················· ···················· ···················· ···················· ···················· ····················
1636 MW420892.1 |Severe acute respiratory syndrome coro ···················· ···················· ···················· ···················· ···················· ····················
1637 MW420893.1 |Severe acute respiratory syndrome coro ···················· ···················· ···················· ···················· ···················· ····················
1638 MW420894.1 |Severe acute respiratory syndrome coro ···················· ···················· ···················· ···················· ···················· ····················
1639 MW420895.1 |Severe acute respiratory syndrome coro ···················· ···················· ···················· ···················· ···················· ····················
1640 MW420896.1 |Severe acute respiratory syndrome coro ···················· ·A·················· ···················· ···················· ···················· ····················
1641 MW420897.1 |Severe acute respiratory syndrome coro ···················· ···················· ···················· ···················· ···················· ····················
1642 MW420898.1 |Severe acute respiratory syndrome coro ···················· ···················· ···················· ···················· ···················· ····················
1643 MW420899.1 |Severe acute respiratory syndrome coro ···················· ·A·················· ···················· ···················· ···················· ····················
1644 MW420900.1 |Severe acute respiratory syndrome coro ···················· ···················· ···················· ···················· ···················· ····················
1645 MW420901.1 |Severe acute respiratory syndrome coro ···················· ····A··············· ···················· ···················· ···················· ····················
1646 MW420902.1 |Severe acute respiratory syndrome coro ···················· ···················· ···················· ···················· ···················· ····················
1647 MW420903.1 |Severe acute respiratory syndrome coro ···················· ···················· ···················· ···················· ···················· ····················
1648 MW420904.1 |Severe acute respiratory syndrome coro ···················· ·A·················· ···················· ···················· ···················· ····················
1649 MW420905.1 |Severe acute respiratory syndrome coro ···················· ···················· ···················· ···················· ···················· ····················
1650 MW420906.1 |Severe acute respiratory syndrome coro ···················· ···················· ···················· ···················· ···················· ····················
1651 MW420907.1 |Severe acute respiratory syndrome coro ···················· ·A·················· ···················· ···················· ···················· ····················
1652 MW420909.1 |Severe acute respiratory syndrome coro ···················· ···················· ···················· ···················· ···················· ····················
1653 MW420910.1 |Severe acute respiratory syndrome coro ···················· ···················· ···················· ···················· ···················· ····················
1654 MW420912.1 |Severe acute respiratory syndrome coro ···················· ·A·················· ···················· ···················· ···················· ····················
1655 MW420913.1 |Severe acute respiratory syndrome coro ···················· ···················· ···················· ···················· ···················· ····················
1656 MW420914.1 |Severe acute respiratory syndrome coro ···················· ···················· ···················· ···················· ···················· ····················
1657 MW420916.1 |Severe acute respiratory syndrome coro ···················· ···················· ···················· ···················· ···················· ····················
1658 MW420917.1 |Severe acute respiratory syndrome coro ···················· ···················· ···················· ···················· ···················· ····················
1659 MW421921.1 |Severe acute respiratory syndrome coro ···················· ···················· ···················· ···················· ···················· ····················
1660 MW421927.1 |Severe acute respiratory syndrome coro ···················· ·A·················· ···················· ···················· ···················· ····················
1661 MW421928.1 |Severe acute respiratory syndrome coro ···················· ···················· ···················· ···················· ···················· ····················
1662 MW421930.1 |Severe acute respiratory syndrome coro ···················· ···················· ···················· ···················· ···················· ····················
1663 MW421939.1 |Severe acute respiratory syndrome coro ···················· ···················· ···················· ···················· ···················· ····················
1664 MW421942.1 |Severe acute respiratory syndrome coro ···················· ·A·················· ···················· ···················· ···················· ····················
1665 MW421949.1 |Severe acute respiratory syndrome coro ···················· ···················· ···················· ···················· ···················· ····················
1666 MW421952.1 |Severe acute respiratory syndrome coro ···················· ···················· ···················· ···················· ···················· ····················
1667 MW421954.1 |Severe acute respiratory syndrome coro ···················· ···················· ···················· ···················· ···················· ····················
1668 MW421957.1 |Severe acute respiratory syndrome coro ···················· ···················· ···················· ···················· ···················· ····················
1669 MW421959.1 |Severe acute respiratory syndrome coro ···················· ···················· ···················· ···················· ···················· ····················
1670 MW421962.1 |Severe acute respiratory syndrome coro ···················· ···················· ···················· ···················· ···················· ····················
1671 MW421966.1 |Severe acute respiratory syndrome coro ···················· ·A·················· ···················· ···················· ···················· ····················
1672 MW421973.1 |Severe acute respiratory syndrome coro ···················· ···················· ···················· ···················· ···················· ····················
1673 MW421975.1 |Severe acute respiratory syndrome coro ···················· ···················· ···················· ···················· ···················· ····················
1674 MW421976.1 |Severe acute respiratory syndrome coro ···················· ···················· ···················· ···················· ···················· ····················
1675 MW421977.1 |Severe acute respiratory syndrome coro ···················· ···················· ···················· ···················· ···················· ····················
1676 MW422255.1 |Severe acute respiratory syndrome coro ···················· ···················· ···················· ···················· ···················· ····················
1677 MW422256.1 |Severe acute respiratory syndrome coro ···················· ···················· ···················· ···················· ···················· ····················
1678 MW406485.1 |Severe acute respiratory syndrome coro ···················· ···················· ···················· ···················· ···················· ····················
1679 MW406487.1 |Severe acute respiratory syndrome coro ···················· ·A·················· ···················· ···················· ···················· ····················
1680 MW406488.1 |Severe acute respiratory syndrome coro ···················· ·A·················· ···················· ···················· ···················· ····················
1681 MW406490.1 |Severe acute respiratory syndrome coro ···················· ·A·················· ···················· ···················· ···················· ····················
1682 MW406491.1 |Severe acute respiratory syndrome coro ···················· ···················· ···················· ···················· ···················· ····················
1683 MW406492.1 |Severe acute respiratory syndrome coro ···················· ···················· ···················· ···················· ···················· ····················
1684 MW406493.1 |Severe acute respiratory syndrome coro ···················· ·A·················· ···················· ···················· ···················· ····················
1685 MW406494.1 |Severe acute respiratory syndrome coro ···················· ···················· ···················· ···················· ···················· ····················
1686 MW406495.1 |Severe acute respiratory syndrome coro ···················· ·A·················· ···················· ···················· ···················· ····················
1687 MW406496.1 |Severe acute respiratory syndrome coro ···················· ···················· ···················· ···················· ···················· ····················
1688 MW406497.1 |Severe acute respiratory syndrome coro ···················· ···················· ···················· ···················· ···················· ····················
1689 MW406498.1 |Severe acute respiratory syndrome coro ···················· ···················· ···················· ···················· ···················· ····················
1690 MW406499.1 |Severe acute respiratory syndrome coro ···················· ···················· ···················· ···················· ···················· ····················
1691 MW406500.1 |Severe acute respiratory syndrome coro ···················· ···················· ···················· ···················· ···················· ····················
1692 MW406501.1 |Severe acute respiratory syndrome coro ···················· ···················· ···················· ···················· ···················· ····················
1693 MW406502.1 |Severe acute respiratory syndrome coro ···················· ·A·················· ···················· ···················· ···················· ····················
1694 MW406503.1 |Severe acute respiratory syndrome coro ···················· ···················· ···················· ···················· ···················· ····················
1695 MW406504.1 |Severe acute respiratory syndrome coro ···················· ···················· ···················· ···················· ···················· ····················
1696 MW406505.1 |Severe acute respiratory syndrome coro ···················· ···················· ···················· ···················· ···················· ····················
1697 MW406507.1 |Severe acute respiratory syndrome coro ···················· ···················· ···················· ···················· ···················· ····················
1698 MW406509.1 |Severe acute respiratory syndrome coro ···················· ···················· ···················· ···················· ···················· ····················
1699 MW406511.1 |Severe acute respiratory syndrome coro ···················· ·············A······ ···················· ···················· ···················· ····················
1700 MW406512.1 |Severe acute respiratory syndrome coro ···················· ···················· ···················· ···················· ···················· ····················
1701 MW406513.1 |Severe acute respiratory syndrome coro ···················· ···················· ···················· ···················· ···················· ····················
1702 MW406514.1 |Severe acute respiratory syndrome coro ···················· ···················· ···················· ···················· ···················· ····················
1703 MW406515.1 |Severe acute respiratory syndrome coro ···················· ···················· ···················· ···················· ···················· ····················
1704 MW406516.1 |Severe acute respiratory syndrome coro ···················· ···················· ···················· ···················· ···················· ····················
1705 MW406517.1 |Severe acute respiratory syndrome coro ···················· ···················· ···················· ···················· ···················· ····················
1706 MW406518.1 |Severe acute respiratory syndrome coro ···················· ···················· ···················· ···················· ···················· ····················
1707 MW406519.1 |Severe acute respiratory syndrome coro ···················· ·A·················· ···················· ···················· ···················· ····················
1708 MW406520.1 |Severe acute respiratory syndrome coro ···················· ···················· ···················· ···················· ···················· ····················
1709 MW406521.1 |Severe acute respiratory syndrome coro ···················· ···················· ···················· ···················· ···················· ····················
1710 MW406522.1 |Severe acute respiratory syndrome coro ···················· ···················· ···················· ···················· ···················· ····················

| | | | | | | | | |
|---|---|---|---|---|---|---|---|---|
| 1711 | MW406523.1 | \|Severe acute respiratory syndrome coro | ···················· | ···················· | ···················· | ···················· | ···················· | ···················· |
| 1712 | MW406524.1 | \|Severe acute respiratory syndrome coro | ···················· | ·A·················· | ···················· | ···················· | ···················· | ···················· |
| 1713 | MW406525.1 | \|Severe acute respiratory syndrome coro | ···················· | ·A·················· | ···················· | ···················· | ···················· | ···················· |
| 1714 | MW406526.1 | \|Severe acute respiratory syndrome coro | ···················· | ···················· | ···················· | ···················· | ···················· | ···················· |
| 1715 | MW406527.1 | \|Severe acute respiratory syndrome coro | ···················· | ···················· | ···················· | ···················· | ···················· | ···················· |
| 1716 | MW406528.1 | \|Severe acute respiratory syndrome coro | ···················· | ···················· | ···················· | ···················· | ···················· | ···················· |
| 1717 | MW406529.1 | \|Severe acute respiratory syndrome coro | ···················· | ···················· | ···················· | ···················· | ···················· | ···················· |
| 1718 | MW406531.1 | \|Severe acute respiratory syndrome coro | ···················· | ···················· | ···················· | ···················· | ···················· | ···················· |
| 1719 | MW406532.1 | \|Severe acute respiratory syndrome coro | ···················· | ···················· | ···················· | ···················· | ···················· | ···················· |
| 1720 | MW406533.1 | \|Severe acute respiratory syndrome coro | ···················· | ···················· | ···················· | ···················· | ···················· | ···················· |
| 1721 | MW406534.1 | \|Severe acute respiratory syndrome coro | ···················· | ···················· | ···················· | ···················· | ···················· | ···················· |
| 1722 | MW406535.1 | \|Severe acute respiratory syndrome coro | ···················· | ···················· | ···················· | ···················· | T··················· | ···················· |
| 1723 | MW406536.1 | \|Severe acute respiratory syndrome coro | ···················· | ···················· | ···················· | ···················· | ···················· | ···················· |
| 1724 | MW406537.1 | \|Severe acute respiratory syndrome coro | ···················· | ···················· | ···················· | ···················· | ···················· | ···················· |
| 1725 | MW406538.1 | \|Severe acute respiratory syndrome coro | ···················· | ···················· | ···················· | ···················· | ···················· | ···················· |
| 1726 | MW406539.1 | \|Severe acute respiratory syndrome coro | ···················· | ···················· | ···················· | ···················· | ···················· | ···················· |
| 1727 | MW406540.1 | \|Severe acute respiratory syndrome coro | ···················· | ···················· | ···················· | ···················· | ···················· | ···················· |
| 1728 | MW406542.1 | \|Severe acute respiratory syndrome coro | ···················· | ·A·················· | ···················· | ···················· | ···················· | ···················· |
| 1729 | MW406543.1 | \|Severe acute respiratory syndrome coro | ···················· | ···················· | ···················· | ···················· | ···················· | ···················· |
| 1730 | MW406544.1 | \|Severe acute respiratory syndrome coro | ···················· | ···················· | ···················· | ···················· | ···················· | ···················· |
| 1731 | MW406545.1 | \|Severe acute respiratory syndrome coro | ···················· | ···················· | ···················· | ···················· | ···················· | ···················· |
| 1732 | MW406546.1 | \|Severe acute respiratory syndrome coro | ···················· | ···················· | ···················· | ···················· | ···················· | ···················· |
| 1733 | MW406547.1 | \|Severe acute respiratory syndrome coro | ···················· | ·A·················· | ···················· | ···················· | ···················· | ···················· |
| 1734 | MW406548.1 | \|Severe acute respiratory syndrome coro | ···················· | ·A·················· | ···················· | ···················· | ···················· | ···················· |
| 1735 | MW406549.1 | \|Severe acute respiratory syndrome coro | ···················· | ···················· | ···················· | ···················· | ···················· | ···················· |
| 1736 | MW406550.1 | \|Severe acute respiratory syndrome coro | ···················· | ·A·················· | ···················· | ···················· | ···················· | ···················· |
| 1737 | MW406551.1 | \|Severe acute respiratory syndrome coro | ···················· | ············A······· | ···················· | ···················· | ···················· | ···················· |
| 1738 | MW406552.1 | \|Severe acute respiratory syndrome coro | ···················· | ···················· | ···················· | ···················· | ···················· | ···················· |
| 1739 | MW406553.1 | \|Severe acute respiratory syndrome coro | ···················· | ············A······· | ···················· | ···················· | ···················· | ···················· |
| 1740 | MW406554.1 | \|Severe acute respiratory syndrome coro | ···················· | ···················· | ···················· | ···················· | ···················· | ···················· |
| 1741 | MW406555.1 | \|Severe acute respiratory syndrome coro | ···················· | ···················· | ···················· | ···················· | ···················· | ···················· |
| 1742 | MW406556.1 | \|Severe acute respiratory syndrome coro | ···················· | ···················· | ···················· | ···················· | ···················· | ···················· |
| 1743 | MW406557.1 | \|Severe acute respiratory syndrome coro | ···················· | ···················· | ···················· | ···················· | ···················· | ···················· |
| 1744 | MW406558.1 | \|Severe acute respiratory syndrome coro | ···················· | ···················· | ···················· | ···················· | ·T·················· | ···················· |
| 1745 | MW406559.1 | \|Severe acute respiratory syndrome coro | ···················· | ···················· | ···················· | ···················· | ···················· | ···················· |
| 1746 | MW406560.1 | \|Severe acute respiratory syndrome coro | ···················· | ···················· | ···················· | ···················· | ···················· | ···················· |
| 1747 | MW406564.1 | \|Severe acute respiratory syndrome coro | ···················· | ···················· | ···················· | ···················· | ···················· | ···················· |
| 1748 | MW406565.1 | \|Severe acute respiratory syndrome coro | ···················· | ············A······· | ···················· | ···················· | ···················· | ···················· |
| 1749 | MW406566.1 | \|Severe acute respiratory syndrome coro | ···················· | ···················· | ···················· | ···················· | ···················· | ···················· |
| 1750 | MW406568.1 | \|Severe acute respiratory syndrome coro | ···················· | ············M······· | ···················· | ···················· | ···················· | ···················· |
| 1751 | MW406569.1 | \|Severe acute respiratory syndrome coro | ···················· | ···················· | ···················· | ···················· | ···················· | ···················· |
| 1752 | MW406570.1 | \|Severe acute respiratory syndrome coro | ···················· | ·A·················· | ···················· | ···················· | ···················· | ···················· |
| 1753 | MW406571.1 | \|Severe acute respiratory syndrome coro | ···················· | ···················· | ···················· | ···················· | ···················· | ···················· |
| 1754 | MW406572.1 | \|Severe acute respiratory syndrome coro | ···················· | ···················· | ···················· | ···················· | ···················· | ···················· |
| 1755 | MW406573.1 | \|Severe acute respiratory syndrome coro | ···················· | ·A·················· | ···················· | ···················· | ···················· | ···················· |
| 1756 | MW406574.1 | \|Severe acute respiratory syndrome coro | ···················· | ···················· | ···················· | ···················· | ···················· | ···················· |
| 1757 | MW406577.1 | \|Severe acute respiratory syndrome coro | ···················· | ···················· | ···················· | ···················· | ···················· | ···················· |
| 1758 | MW406580.1 | \|Severe acute respiratory syndrome coro | ···················· | ···················· | ···················· | ···················· | ···················· | ···················· |
| 1759 | MW406581.1 | \|Severe acute respiratory syndrome coro | ···················· | ···················· | ···················· | ···················· | ···················· | ···················· |
| 1760 | MW406584.1 | \|Severe acute respiratory syndrome coro | ···················· | ···················· | ···················· | ···················· | ···················· | ···················· |
| 1761 | MW406586.1 | \|Severe acute respiratory syndrome coro | ···················· | ···················· | ···················· | ···················· | ···················· | ···················· |
| 1762 | MW406587.1 | \|Severe acute respiratory syndrome coro | ···················· | ···················· | ···················· | ···················· | ···················· | ···················· |
| 1763 | MW406588.1 | \|Severe acute respiratory syndrome coro | ···················· | ···················· | ···················· | ···················· | ···················· | ···················· |
| 1764 | MW406590.1 | \|Severe acute respiratory syndrome coro | ···················· | ···················· | ···················· | ···················· | ···················· | ···················· |
| 1765 | MW406591.1 | \|Severe acute respiratory syndrome coro | ···················· | ···················· | ···················· | ···················· | ···················· | ···················· |
| 1766 | MW406594.1 | \|Severe acute respiratory syndrome coro | ···················· | ···················· | ···················· | ···················· | ···················· | ···················· |
| 1767 | MW406595.1 | \|Severe acute respiratory syndrome coro | ···················· | ···················· | ···················· | ···················· | ···················· | ···················· |
| 1768 | MW406597.1 | \|Severe acute respiratory syndrome coro | ···················· | ···················· | ···················· | ···················· | ···················· | ···················· |
| 1769 | MW406604.1 | \|Severe acute respiratory syndrome coro | ···················· | ···················· | ···················· | ···················· | ···················· | ···················· |
| 1770 | MW406608.1 | \|Severe acute respiratory syndrome coro | ···················· | ·R·················· | ···················· | ···················· | ···················· | ···················· |
| 1771 | MW406610.1 | \|Severe acute respiratory syndrome coro | ···················· | ·R·················· | ···················· | ···················· | ···················· | ···················· |
| 1772 | MW406613.1 | \|Severe acute respiratory syndrome coro | ···················· | ···················· | ···················· | ···················· | ···················· | ···················· |
| 1773 | MW406618.1 | \|Severe acute respiratory syndrome coro | ···················· | ·A·················· | ···················· | ···················· | ···················· | ···················· |
| 1774 | MW406619.1 | \|Severe acute respiratory syndrome coro | ···················· | ···················· | ···················· | ···················· | ···················· | ···················· |
| 1775 | MW406620.1 | \|Severe acute respiratory syndrome coro | ···················· | ···················· | ···················· | ···················· | ···················· | ···················· |
| 1776 | MW406627.1 | \|Severe acute respiratory syndrome coro | ···················· | ···················· | ···················· | ···················· | ···················· | ···················· |
| 1777 | MW406628.1 | \|Severe acute respiratory syndrome coro | ···················· | ···················· | ···················· | ···················· | ···················· | ···················· |
| 1778 | MW406629.1 | \|Severe acute respiratory syndrome coro | ···················· | ···················· | ···················· | ···················· | ···················· | ···················· |
| 1779 | MW406631.1 | \|Severe acute respiratory syndrome coro | ···················· | ···················· | ···················· | ···················· | ···················· | ···················· |
| 1780 | MW406633.1 | \|Severe acute respiratory syndrome coro | ···················· | ···················· | ···················· | ···················· | ···················· | ···················· |
| 1781 | MW406639.1 | \|Severe acute respiratory syndrome coro | ···················· | ···················· | ···················· | ···················· | ···················· | ···················· |
| 1782 | MW406642.1 | \|Severe acute respiratory syndrome coro | ···················· | ·R·················· | ···················· | ···················· | ···················· | ···················· |
| 1783 | MW406643.1 | \|Severe acute respiratory syndrome coro | ···················· | ···················· | ···················· | ···················· | ···················· | ···················· |
| 1784 | MW406644.1 | \|Severe acute respiratory syndrome coro | ···················· | ·R·················· | ···················· | ···················· | ···················· | ···················· |
| 1785 | MW411627.1 | \|Severe acute respiratory syndrome coro | ···················· | ···················· | ···················· | ···················· | ···················· | ···················· |
| 1786 | MW411628.1 | \|Severe acute respiratory syndrome coro | ···················· | ···················· | ···················· | ···················· | ···················· | ···················· |
| 1787 | MW411629.1 | \|Severe acute respiratory syndrome coro | ···················· | ···················· | ···················· | ···················· | ···················· | ···················· |
| 1788 | MW411630.1 | \|Severe acute respiratory syndrome coro | ···················· | ···················· | ···················· | ···················· | ···················· | ···················· |

```
1789 MW411631.1 |Severe acute respiratory syndrome coro  ····················  ····················  ····················  ····················  ····················  ····················
1790 MW411632.1 |Severe acute respiratory syndrome coro  ····················  ····················  ····················  ····················  ····················  ····················
1791 MW411633.1 |Severe acute respiratory syndrome coro  ····················  ····················  ····················  ····················  ····················  ····················
1792 MW411634.1 |Severe acute respiratory syndrome coro  ····················  ····················  ····················  ····················  ····················  ····················
1793 MW411635.1 |Severe acute respiratory syndrome coro  ····················  ·A··················  ····················  ····················  ····················  ····················
1794 MW411636.1 |Severe acute respiratory syndrome coro  ····················  ····················  ····················  ····················  ····················  ····················
1795 MW411637.1 |Severe acute respiratory syndrome coro  ····················  ·A··················  ····················  ····················  ····················  ····················
1796 MW411638.1 |Severe acute respiratory syndrome coro  ····················  ····················  ····················  ····················  ····················  ····················
1797 MW411639.1 |Severe acute respiratory syndrome coro  ····················  ····················  ····················  ····················  ····················  ····················
1798 MW411640.1 |Severe acute respiratory syndrome coro  ····················  ·A··················  ····················  ····················  ····················  ····················
1799 MW411641.1 |Severe acute respiratory syndrome coro  ····················  ····················  ····················  ····················  ·········T··········  ····················
1800 MW411642.1 |Severe acute respiratory syndrome coro  ····················  ·A··················  ····················  ····················  ····················  ····················
1801 MW411643.1 |Severe acute respiratory syndrome coro  ····················  ·A··················  ····················  ····················  ····················  ····················
1802 MW411644.1 |Severe acute respiratory syndrome coro  ····················  ·A··················  ····················  ····················  ····················  ····················
1803 MW411645.1 |Severe acute respiratory syndrome coro  ····················  ·A··················  ····················  ····················  ····················  ····················
1804 MW411646.1 |Severe acute respiratory syndrome coro  ····················  ····················  ····················  ····················  ····················  ····················
1805 MW411647.1 |Severe acute respiratory syndrome coro  ····················  ····················  ····················  ····················  ····················  ····················
1806 MW411648.1 |Severe acute respiratory syndrome coro  ····················  ·A··················  ····················  ····················  ····················  ····················
1807 MW411649.1 |Severe acute respiratory syndrome coro  ····················  ····A···············  ····················  ····················  ····················  ····················
1808 MW411650.1 |Severe acute respiratory syndrome coro  ····················  ····················  ····················  ····················  ····················  ····················
1809 MW411651.1 |Severe acute respiratory syndrome coro  ····················  ····················  ····················  ····················  ····················  ····················
1810 MW411652.1 |Severe acute respiratory syndrome coro  ····················  ····················  ····················  ····················  ····················  ····················
1811 MW411653.1 |Severe acute respiratory syndrome coro  ····················  ····················  ····················  ····················  ····················  ····················
1812 MW411654.1 |Severe acute respiratory syndrome coro  ····················  ····················  ····················  ····················  ····················  ····················
1813 MW411655.1 |Severe acute respiratory syndrome coro  ····················  ····················  ····················  ····················  ····················  ····················
1814 MW411656.1 |Severe acute respiratory syndrome coro  ····················  ····················  ····················  ····················  ····················  ····················
1815 MW411871.1 |Severe acute respiratory syndrome coro  ····················  ····················  ····················  ····················  ····················  ····················
1816 MW411872.1 |Severe acute respiratory syndrome coro  ····················  ·A··················  ····················  ····················  ····················  ····················
1817 MW411873.1 |Severe acute respiratory syndrome coro  ····················  ·A··················  ····················  ····················  ····················  ····················
1818 MW411874.1 |Severe acute respiratory syndrome coro  ····················  ····················  ····················  ····················  ····················  ····················
1819 MW411875.1 |Severe acute respiratory syndrome coro  ····················  ·A··················  ····················  ····················  ····················  ····················
1820 MW411876.1 |Severe acute respiratory syndrome coro  ····················  ·A··················  ····················  ····················  ····················  ···A················
1821 MW411877.1 |Severe acute respiratory syndrome coro  ····················  ·A··················  ····················  ····················  ····················  ···A················
1822 MW411878.1 |Severe acute respiratory syndrome coro  ····················  ·A··················  ····················  ····················  ····················  ····················
1823 MW411879.1 |Severe acute respiratory syndrome coro  ····················  ····················  ····················  ····················  ····················  ····················
1824 MW411880.1 |Severe acute respiratory syndrome coro  ····················  ····················  ····················  ····················  ····················  ····················
1825 MW411881.1 |Severe acute respiratory syndrome coro  ····················  ·A··················  ····················  ····················  ····················  ····················
1826 MW411882.1 |Severe acute respiratory syndrome coro  ····················  ·A··················  ····················  ····················  ····················  ···A················
1827 MW411883.1 |Severe acute respiratory syndrome coro  ····················  ····················  ····················  ····················  ····················  ····················
1828 MW411884.1 |Severe acute respiratory syndrome coro  ····················  ·A··················  ····················  ····················  ····················  ····················
1829 MW411885.1 |Severe acute respiratory syndrome coro  ····················  ····················  ····················  ····················  ····················  ····················
1830 MW411886.1 |Severe acute respiratory syndrome coro  ····················  ····················  ····················  ····················  ····················  ····················
1831 MW411887.1 |Severe acute respiratory syndrome coro  ··················A·  ····················  ····················  ····················  ····················  ····················  1
1832 MW411888.1 |Severe acute respiratory syndrome coro  ····················  ····················  ····················  ····················  ····················  ····················
1833 MW411889.1 |Severe acute respiratory syndrome coro  ····················  ····················  ····················  ····················  ····················  ····················
1834 MW411890.1 |Severe acute respiratory syndrome coro  ····················  ····················  ····················  ····················  ····················  ····················
1835 MW411891.1 |Severe acute respiratory syndrome coro  ····················  ·A··················  ····················  ····················  ····················  ····················
1836 MW411892.1 |Severe acute respiratory syndrome coro  ····················  ····················  ····················  ····················  ····················  ····················
1837 MW411893.1 |Severe acute respiratory syndrome coro  ····················  ····················  ····················  ····················  ····················  ····················
1838 MW411894.1 |Severe acute respiratory syndrome coro  ····················  ·A··················  ····················  ····················  ····················  ····················
1839 MW411895.1 |Severe acute respiratory syndrome coro  ····················  ····················  ····················  ····················  ····················  ····················
1840 MW411896.1 |Severe acute respiratory syndrome coro  ····················  ····················  ····················  ····················  ····················  ····················
1841 MW411897.1 |Severe acute respiratory syndrome coro  ····················  ·A··················  ····················  ····················  ····················  ····················
1842 MW411898.1 |Severe acute respiratory syndrome coro  ····················  ····················  ····················  ····················  ····················  ····················
1843 MW411899.1 |Severe acute respiratory syndrome coro  ····················  ····················  ····················  ····················  ····················  ····················
1844 MW411900.1 |Severe acute respiratory syndrome coro  ····················  ·A··················  ····················  ····················  ····················  ····················
1845 MW411901.1 |Severe acute respiratory syndrome coro  ····················  ····················  ····················  ····················  ····················  ····················
1846 MW411903.1 |Severe acute respiratory syndrome coro  ····················  ····················  ····················  ····················  ····················  ····················
1847 MW411904.1 |Severe acute respiratory syndrome coro  ····················  ····················  ····················  ····················  ····················  ····················
1848 MW411905.1 |Severe acute respiratory syndrome coro  ····················  ····················  ····················  ····················  ····················  ····················
1849 MW411906.1 |Severe acute respiratory syndrome coro  ····················  ····················  ····················  ····················  ····················  ····················
1850 MW411907.1 |Severe acute respiratory syndrome coro  ····················  ····················  ····················  ····················  ····················  ····················
1851 MW411908.1 |Severe acute respiratory syndrome coro  ····················  ····················  ····················  ····················  ····················  ····················
1852 MW411909.1 |Severe acute respiratory syndrome coro  ····················  ····················  ····················  ····················  ····················  ····················
1853 MW411910.1 |Severe acute respiratory syndrome coro  ····················  ····················  ····················  ····················  ····················  ····················
1854 MW411911.1 |Severe acute respiratory syndrome coro  ····················  ····················  ····················  ····················  ····················  ····················
1855 MW411912.1 |Severe acute respiratory syndrome coro  ····················  ·A··················  ····················  ····················  ····················  ····················
1856 MW411913.1 |Severe acute respiratory syndrome coro  ····················  ·A··················  ····················  ····················  ····················  ····················
1857 MW411914.1 |Severe acute respiratory syndrome coro  ····················  ·A··················  ····················  ····················  ····················  ····················
1858 MW411915.1 |Severe acute respiratory syndrome coro  ····················  ·A··················  ····················  ····················  ····················  ····················
1859 MW411916.1 |Severe acute respiratory syndrome coro  ····················  ·A··················  ····················  ····················  ····················  ····················
1860 MW411917.1 |Severe acute respiratory syndrome coro  ····················  ·A··················  ····················  ····················  ····················  ····················
1861 MW411918.1 |Severe acute respiratory syndrome coro  ····················  ·A··················  ····················  ····················  ····················  ····················
1862 MW411919.1 |Severe acute respiratory syndrome coro  ····················  ····················  ····················  ····················  ·········T··········  ····················
1863 MW411920.1 |Severe acute respiratory syndrome coro  ····················  ····················  ····················  ····················  ····················  ····················
1864 MW411921.1 |Severe acute respiratory syndrome coro  ····················  ····················  ····················  ····················  ····················  ····················
1865 MW411922.1 |Severe acute respiratory syndrome coro  ····················  ····················  ····················  ····················  ····················  ····················
1866 MW411923.1 |Severe acute respiratory syndrome coro  ····················  ····················  ····················  ····················  ····················  ····················
```

```
1867 MW411924.1 |Severe acute respiratory syndrome coro  ···················· ·A····A············· ···················· ···················· ···················· ····················
1868 MW411926.1 |Severe acute respiratory syndrome coro  ···················· ···················· ···················· ···················· ···················· ····················
1869 MW411928.1 |Severe acute respiratory syndrome coro  ···················· ·A·················· ···················· ···················· ···················· ····················
1870 MW411929.1 |Severe acute respiratory syndrome coro  ···················· ·A·················· ···················· ···················· ···················· ····················
1871 MW411931.1 |Severe acute respiratory syndrome coro  ···················· ···················· ···················· ···················· ···················· ····················
1872 MW411932.1 |Severe acute respiratory syndrome coro  ···················· ···················· ···················· ···················· ···················· ····················
1873 MW411933.1 |Severe acute respiratory syndrome coro  ···················· ···················· ···················· ···················· ···················· ····················
1874 MW411934.1 |Severe acute respiratory syndrome coro  ···················· ···················· ···················· ···················· ···················· ····················
1875 MW411937.1 |Severe acute respiratory syndrome coro  ···················· ·············A······ ···················· ···················· ···················· ····················
1876 MW411938.1 |Severe acute respiratory syndrome coro  ···················· ···················· ···················· ···················· ···················· ····················
1877 MW411939.1 |Severe acute respiratory syndrome coro  ···················· ···················· ···················· ···················· ···················· ····················
1878 MW411940.1 |Severe acute respiratory syndrome coro  ···················· ···················· ···················· ···················· ···················· ····················
1879 MW411941.1 |Severe acute respiratory syndrome coro  ···················· ···················· ···················· ···················· ···················· ····················
1880 MW411943.1 |Severe acute respiratory syndrome coro  ···················· ···················· ···················· ···················· ···················· ····················
1881 MW411944.1 |Severe acute respiratory syndrome coro  ···················· ···················· ···················· ···················· ···················· ····················
1882 MW411945.1 |Severe acute respiratory syndrome coro  ···················· ···················· ···················· ···················· ···················· ····················
1883 MW411947.1 |Severe acute respiratory syndrome coro  ···················· ···················· ···················· ···················· ···················· ····················
1884 MW411948.1 |Severe acute respiratory syndrome coro  ···················· ···················· ···················· ···················· ···················· ····················
1885 MW411949.1 |Severe acute respiratory syndrome coro  ···················· ···················· ···················· ···················· ···················· ····················
1886 MT232662.1 |Severe acute respiratory syndrome coro  ···················· ···················· ···················· ···················· ···················· ····················
1887 MT232664.1 |Severe acute respiratory syndrome coro  ···················· ···················· ···················· ···················· ···················· ····················
1888 MT232667.1 |Severe acute respiratory syndrome coro  ···················· ···················· ···················· ···················· ···················· ····················
1889 MT232671.1 |Severe acute respiratory syndrome coro  ···················· ···················· ···················· ···················· ···················· ····················
1890 MT232672.1 |Severe acute respiratory syndrome coro  ···················· ···················· ···················· ···················· ···················· ····················
1891 MT232673.1 |Severe acute respiratory syndrome coro  ···················· ···················· ···················· ···················· ···················· ····················
1892 MT232675.1 |Severe acute respiratory syndrome coro  ···················· ···················· ···················· ···················· ···················· ····················
1893 MT232677.1 |Severe acute respiratory syndrome coro  ···················· ···················· ···················· ···················· ···················· ····················
1894 MT232679.1 |Severe acute respiratory syndrome coro  ···················· ···················· ···················· ···················· ···················· ····················
1895 MT232681.1 |Severe acute respiratory syndrome coro  ···················· ···················· ···················· ···················· ···················· ····················
1896 MT232682.1 |Severe acute respiratory syndrome coro  ···················· ···················· ···················· ···················· ···················· ····················
1897 MT232683.1 |Severe acute respiratory syndrome coro  ···················· ···················· ···················· ···················· ···················· ····················
1898 MT232684.1 |Severe acute respiratory syndrome coro  ···················· ···················· ···················· ···················· ···················· ····················
1899 MT232686.1 |Severe acute respiratory syndrome coro  ···················· ···················· ···················· ···················· ···················· ····················
1900 MT232687.1 |Severe acute respiratory syndrome coro  ···················· ···················· ···················· ···················· ···················· ····················
1901 MT232688.1 |Severe acute respiratory syndrome coro  ···················· ···················· ···················· ···················· ···················· ····················
1902 MT232690.1 |Severe acute respiratory syndrome coro  ···················· ···················· ···················· ···················· ···················· ····················
1903 MT232692.1 |Severe acute respiratory syndrome coro  ···················· ···················· ···················· ···················· ···················· ··················C·          1
1904 MT232693.1 |Severe acute respiratory syndrome coro  ···················· ···················· ···················· ···················· ···················· ····················
1905 MT232694.1 |Severe acute respiratory syndrome coro  ···················· ···················· ···················· ···················· ···················· ····················
1906 MT232696.1 |Severe acute respiratory syndrome coro  ···················· ···················· ···················· ···················· ···················· ····················
1907 MT232697.1 |Severe acute respiratory syndrome coro  ···················· ···················· ···················· ···················· ···················· ····················
1908 MT232702.1 |Severe acute respiratory syndrome coro  ···················· ···················· ···················· ···················· ···················· ····················
1909 MT232704.1 |Severe acute respiratory syndrome coro  ···················· ···················· ···················· ···················· ···················· ····················
1910 MT232707.1 |Severe acute respiratory syndrome coro  ···················· ···················· ···················· ···················· ···················· ····················
1911 MT232709.1 |Severe acute respiratory syndrome coro  ···················· ···················· ···················· ···················· ···················· ····················
1912 MW406646.1 |Severe acute respiratory syndrome coro  ···················· ···················· ···················· ···················· ·········T·········· ····················
1913 MW406647.1 |Severe acute respiratory syndrome coro  ···················· ···················· ···················· ···················· ·········T·········· ····················
1914 MW406648.1 |Severe acute respiratory syndrome coro  ···················· ···················· ···················· ···················· ·········T·········· ····················
1915 MW406649.1 |Severe acute respiratory syndrome coro  ···················· ···················· ···················· ···················· ·········T·········· ····················
1916 MW406650.1 |Severe acute respiratory syndrome coro  ···················· ···················· ···················· ···················· ·········T·········· ····················
1917 MW406651.1 |Severe acute respiratory syndrome coro  ···················· ···················· ···················· ···················· ·········T·········· ····················
1918 MW406652.1 |Severe acute respiratory syndrome coro  ···················· ···················· ···················· ···················· ·········T·········· ····················
1919 MW406653.1 |Severe acute respiratory syndrome coro  ···················· ···················· ···················· ···················· ·········T·········· ····················
1920 MW406654.1 |Severe acute respiratory syndrome coro  ···················· ···················· ···················· ···················· ·········T·········· ····················
1921 MW406655.1 |Severe acute respiratory syndrome coro  ···················· ···················· ···················· ···················· ·········T·········· ····················
1922 MW406656.1 |Severe acute respiratory syndrome coro  ···················· ···················· ···················· ···················· ···················· ····················
1923 MW406657.1 |Severe acute respiratory syndrome coro  ···················· ···················· ···················· ···················· ···················· ····················
1924 MW406658.1 |Severe acute respiratory syndrome coro  ···················· ···················· ···················· ···················· ···················· ····················
1925 MW406659.1 |Severe acute respiratory syndrome coro  ···················· ···················· ···················· ···················· ···················· ····················
1926 MW406660.1 |Severe acute respiratory syndrome coro  ···················· ···················· ···················· ···················· ···················· ····················
1927 MW406661.1 |Severe acute respiratory syndrome coro  ···················· ···················· ···················· ···················· ···················· ····················
1928 MW406662.1 |Severe acute respiratory syndrome coro  ···················· ······A············· ···················· ···················· ···················· ····················
1929 MW406663.1 |Severe acute respiratory syndrome coro  ···················· ······A············· ···················· ···················· ···················· ····················
1930 MW406664.1 |Severe acute respiratory syndrome coro  ···················· ······A············· ···················· ···················· ···················· ····················
1931 MW406665.1 |Severe acute respiratory syndrome coro  ···················· ······A············· ···················· ···················· ···················· ····················
1932 MW406666.1 |Severe acute respiratory syndrome coro  ···················· ······A············· ···················· ···················· ···················· ····················
1933 MW406667.1 |Severe acute respiratory syndrome coro  ···················· ···················· ···················· ···················· ···················· ····················
1934 MW406668.1 |Severe acute respiratory syndrome coro  ···················· ······A············· ···················· ···················· ···················· ····················
1935 MW406669.1 |Severe acute respiratory syndrome coro  ···················· ···················· ···················· ···················· ···················· ····················
1936 MW406670.1 |Severe acute respiratory syndrome coro  ···················· ···················· ···················· ···················· ···················· ····················
1937 MW406671.1 |Severe acute respiratory syndrome coro  ···················· ···················· ···················· ···················· ···················· ····················
1938 MW406672.1 |Severe acute respiratory syndrome coro  ···················· ···················· ···················· ···················· ···················· ····················
1939 MW406673.1 |Severe acute respiratory syndrome coro  ···················· ···················· ···················· ···················· ···················· ····················
1940 MW406674.1 |Severe acute respiratory syndrome coro  ···················· ···················· ···················· ···················· ···················· ····················
1941 MW406675.1 |Severe acute respiratory syndrome coro  ···················· ···················· ···················· ···················· ···················· ····················
1942 MW406676.1 |Severe acute respiratory syndrome coro  ···················· ···················· ···················· ···················· ···················· ····················
1943 MW406677.1 |Severe acute respiratory syndrome coro  ···················· ···················· ···················· ···················· ···················· ····················
1944 MW406678.1 |Severe acute respiratory syndrome coro  ···················· ···················· ···················· ···················· ···················· ····················
```

1945 MW406679.1 |Severe acute respiratory syndrome coro ···················· ···················· ···················· ···················· ···················· ····················
1946 MW406680.1 |Severe acute respiratory syndrome coro ···················· ···················· ···················· ···················· ···················· ····················
1947 MW406681.1 |Severe acute respiratory syndrome coro ···················· ···················· ···················· ···················· ···················· ····················
1948 MW406682.1 |Severe acute respiratory syndrome coro ···················· ···················· ···················· ···················· ···················· ····················
1949 MW406731.1 |Severe acute respiratory syndrome coro ···················· ····T··············· ···················· ···················· ···················· ····················
1950 MW406732.1 |Severe acute respiratory syndrome coro ···················· ····T··············· ···················· ···················· ···················· ····················
1951 MW406733.1 |Severe acute respiratory syndrome coro ···················· ····T··············· ···················· ···················· ···················· ····················
1952 MW406735.1 |Severe acute respiratory syndrome coro ···················· ···················· ···················· ···················· ···················· ····················
1953 MW406736.1 |Severe acute respiratory syndrome coro ···················· ····T··············· ···················· ···················· ···················· ····················
1954 MW406737.1 |Severe acute respiratory syndrome coro ···················· ·A·················· ···················· ···················· ···················· ····················
1955 MW406738.1 |Severe acute respiratory syndrome coro ···················· ···················· ···················· ···················· ···················· ····················
1956 MW406739.1 |Severe acute respiratory syndrome coro ···················· ····T··············· ···················· ···················· ···················· ····················
1957 MW406740.1 |Severe acute respiratory syndrome coro ···················· ···················· ···················· ···················· ···················· ····················
1958 MW406781.1 |Severe acute respiratory syndrome coro ···················· ···················· ···················· ···················· ···················· ····················
1959 MW406794.1 |Severe acute respiratory syndrome coro ···················· ···················· ···················· ···················· ···················· ····················
1960 MW404672.1 |Severe acute respiratory syndrome coro ···················· ···················· ···················· ···················· ···················· ····················
1961 MW404673.1 |Severe acute respiratory syndrome coro ···················· ···················· ···················· ···················· ···················· ····················
1962 MW405462.1 |Severe acute respiratory syndrome coro ···················· ···················· ···················· ···················· ···················· ····················
1963 MW405463.1 |Severe acute respiratory syndrome coro ···················· ···················· ···················· ···················· ···················· ····················
1964 MW405464.1 |Severe acute respiratory syndrome coro ···················· ···················· ···················· ···················· ···················· ····················
1965 MW405465.1 |Severe acute respiratory syndrome coro ···················· ···················· ···················· ···················· ···················· ····················
1966 MW405466.1 |Severe acute respiratory syndrome coro ···················· ···················· ···················· ···················· ···················· ····················
1967 MW405467.1 |Severe acute respiratory syndrome coro ···················· ···················· ···················· ···················· ···················· ····················
1968 MW405468.1 |Severe acute respiratory syndrome coro ···················· ···················· ···················· ···················· ···················· ····················
1969 MW405469.1 |Severe acute respiratory syndrome coro ···················· ·A·················· ···················· ···················· ···················· ····················
1970 MW405470.1 |Severe acute respiratory syndrome coro ···················· ···················· ···················· ···················· ···················· ····················
1971 MW405471.1 |Severe acute respiratory syndrome coro ···················· ···················· ···················· ···················· ···················· ····················
1972 MW405472.1 |Severe acute respiratory syndrome coro ···················· ···················· ···················· ···················· ···················· ····················
1973 MW405473.1 |Severe acute respiratory syndrome coro ········T··········· ·A·················· ···················· ···················· ···················· ····················
1974 MW405474.1 |Severe acute respiratory syndrome coro ···················· ···················· ···················· ···················· ···················· ····················
1975 MW405475.1 |Severe acute respiratory syndrome coro ···················· ···················· ···················· ···················· ···················· ····················
1976 MW405476.1 |Severe acute respiratory syndrome coro ···················· ···················· ···················· ···················· ···················· ····················
1977 MW405477.1 |Severe acute respiratory syndrome coro ···················· ·A·················· ···················· ···················· ···················· ····················
1978 MW405478.1 |Severe acute respiratory syndrome coro ···················· ······A············· ···················· ···················· ···················· ····················
1979 MW405479.1 |Severe acute respiratory syndrome coro ···················· ···················· ···················· ···················· ···················· ····················
1980 MW405480.1 |Severe acute respiratory syndrome coro ···················· ···················· ···················· ···················· ···················· ····················
1981 MW405481.1 |Severe acute respiratory syndrome coro ···················· ···················· ···················· ···················· ···················· ····················
1982 MW405482.1 |Severe acute respiratory syndrome coro ···················· ····A··············· ···················· ···················· ···················· ····················
1983 MW405483.1 |Severe acute respiratory syndrome coro ···················· ····A··············· ···················· ···················· ···················· ····················
1984 MW405484.1 |Severe acute respiratory syndrome coro ···················· ···················· ···················· ···················· ···················· ····················
1985 MW405485.1 |Severe acute respiratory syndrome coro ···················· ···················· ···················· ···················· ···················· ····················
1986 MW405486.1 |Severe acute respiratory syndrome coro ···················· ···················· ···················· ···················· ···················· ····················
1987 MW403520.1 |Severe acute respiratory syndrome coro ···················· ···················· ···················· ···················· ···················· ····················
1988 MW403523.1 |Severe acute respiratory syndrome coro ···················· ···················· ···················· ···················· ···················· ····················
1989 MW403524.1 |Severe acute respiratory syndrome coro ···················· ···················· ···················· ···················· ···················· ····················
1990 MW403525.1 |Severe acute respiratory syndrome coro ···················· ···················· ···················· ···················· ···················· ····················
1991 MW403526.1 |Severe acute respiratory syndrome coro ···················· ···················· ···················· ···················· ···················· ····················
1992 MW403527.1 |Severe acute respiratory syndrome coro ···················· ···················· ···················· ···················· ···················· ····················
1993 MW403529.1 |Severe acute respiratory syndrome coro ···················· ···················· ···················· ···················· ···················· ····················
1994 MW403530.1 |Severe acute respiratory syndrome coro ···················· ···················· ···················· ···················· ···················· ····················
1995 MW403532.1 |Severe acute respiratory syndrome coro ···················· ···················· ···················· ···················· ···················· ····················
1996 MW403533.1 |Severe acute respiratory syndrome coro ···················· ···················· ···················· ···················· ···················· ····················
1997 MW403534.1 |Severe acute respiratory syndrome coro ···················· ···················· ···················· ···················· ···················· ····················
1998 MW403535.1 |Severe acute respiratory syndrome coro ···················· ···················· ···················· ···················· ···················· ····················
1999 MW403538.1 |Severe acute respiratory syndrome coro ···················· ···················· ···················· ···················· ···················· ····················
2000 MW403540.1 |Severe acute respiratory syndrome coro ···················· ···················· ···················· ···················· ···················· ····················
2001 MW403541.1 |Severe acute respiratory syndrome coro ···················· ···················· ···················· ···················· ···················· ····················
2002 MW403542.1 |Severe acute respiratory syndrome coro ···················· ···················· ···················· ···················· ···················· ····················
2003 MW403544.1 |Severe acute respiratory syndrome coro ···················· ···················· ···················· ···················· ···················· ····················
2004 MW403545.1 |Severe acute respiratory syndrome coro ···················· ···················· ···················· ···················· ···················· ····················
2005 MW403546.1 |Severe acute respiratory syndrome coro ···················· ···················· ···················· ···················· ···················· ····················
2006 MW403548.1 |Severe acute respiratory syndrome coro ···················· ···················· ···················· ···················· ···················· ····················
2007 MW403549.1 |Severe acute respiratory syndrome coro ···················· ···················· ···················· ···················· ···················· ····················
2008 MW403550.1 |Severe acute respiratory syndrome coro ···················· ···················· ···················· ···················· ···················· ····················
2009 MW403551.1 |Severe acute respiratory syndrome coro ···················· ···················· ···················· ···················· ···················· ····················
2010 MW403552.1 |Severe acute respiratory syndrome coro ···················· ···················· ···················· ···················· ···················· ····················
2011 MW403553.1 |Severe acute respiratory syndrome coro ···················· ···················· ···················· ···················· ···················· ····················
2012 MW403557.1 |Severe acute respiratory syndrome coro ···················· ···················· ···················· ···················· ···················· ····················
2013 MW403559.1 |Severe acute respiratory syndrome coro ···················· ···················· ···················· ···················· ···················· ····················
2014 MW403562.1 |Severe acute respiratory syndrome coro ···················· ···················· ···················· ···················· ···················· ····················
2015 MW403564.1 |Severe acute respiratory syndrome coro ···················· ···················· ···················· ···················· ···················· ····················
2016 MW403573.1 |Severe acute respiratory syndrome coro ···················· ···················· ···················· ···················· ···················· ····················
2017 MW403589.1 |Severe acute respiratory syndrome coro ···················· ···················· ···················· ···················· ···················· ····················
2018 MW403598.1 |Severe acute respiratory syndrome coro ···················· ···················· ···················· ···················· ···················· ····················
2019 MW403600.1 |Severe acute respiratory syndrome coro ···················· ·A·················· ···················· ···················· ···················· ····················
2020 MW403604.1 |Severe acute respiratory syndrome coro ···················· ···················· ···················· ···················· ···················· ····················
2021 MW403605.1 |Severe acute respiratory syndrome coro ···················· ···················· ···················· ···················· ···················· ····················
2022 MW403606.1 |Severe acute respiratory syndrome coro ···················· ···················· ···················· ···················· ···················· ····················

| | | | | | | | | |
|---|---|---|---|---|---|---|---|---|
| 2023 | MW403607.1 | Severe acute respiratory syndrome coro | ···················· | ···················· | ···················· | ···················· | ···················· | ···················· |
| 2024 | MW403608.1 | Severe acute respiratory syndrome coro | ···················· | ···················· | ···················· | ···················· | ···················· | ···················· |
| 2025 | MW403609.1 | Severe acute respiratory syndrome coro | ···················· | ···················· | ···················· | ···················· | ···················· | ···················· |
| 2026 | MW403610.1 | Severe acute respiratory syndrome coro | ···················· | ···················· | ···················· | ···················· | ···················· | ···················· |
| 2027 | MW403611.1 | Severe acute respiratory syndrome coro | ···················· | ·A·················· | ···················· | ···················· | ···················· | ···················· |
| 2028 | MW403612.1 | Severe acute respiratory syndrome coro | ···················· | ···················· | ···················· | ···················· | ···················· | ···················· |
| 2029 | MW403613.1 | Severe acute respiratory syndrome coro | ···················· | ···················· | ···················· | ···················· | ···················· | ···················· |
| 2030 | MW403615.1 | Severe acute respiratory syndrome coro | ···················· | ···················· | ···················· | ···················· | ···················· | ···················· |
| 2031 | MW403616.1 | Severe acute respiratory syndrome coro | ···················· | ···················· | ···················· | ···················· | ···················· | ···················· |
| 2032 | MW403617.1 | Severe acute respiratory syndrome coro | ···················· | ···················· | ···················· | ···················· | ···················· | ···················· |
| 2033 | MW403618.1 | Severe acute respiratory syndrome coro | ···················· | ·A·················· | ···················· | ···················· | ···················· | ···················· |
| 2034 | MW403625.1 | Severe acute respiratory syndrome coro | ···················· | ···················· | ···················· | ···················· | ···················· | ···················· |
| 2035 | MW403627.1 | Severe acute respiratory syndrome coro | ···················· | ···················· | ···················· | ···················· | ···················· | ···················· |
| 2036 | MW403628.1 | Severe acute respiratory syndrome coro | ···················· | ···················· | ···················· | ···················· | ···················· | ···················· |
| 2037 | MW403629.1 | Severe acute respiratory syndrome coro | ···················· | ···················· | ···················· | ···················· | ···················· | ···················· |
| 2038 | MW403631.1 | Severe acute respiratory syndrome coro | ···················· | ···················· | ···················· | ···················· | ···················· | ···················· |
| 2039 | MW403632.1 | Severe acute respiratory syndrome coro | ···················· | ···················· | ···················· | ···················· | ···················· | ···················· |
| 2040 | MW403636.1 | Severe acute respiratory syndrome coro | ···················· | ···················· | ···················· | ···················· | ···················· | ···················· |
| 2041 | MW403641.1 | Severe acute respiratory syndrome coro | ···················· | ·A·················· | ···················· | ···················· | ···················· | ···················· |
| 2042 | MW403642.1 | Severe acute respiratory syndrome coro | ···················· | ···················· | ···················· | ···················· | ···················· | ···················· |
| 2043 | MW403644.1 | Severe acute respiratory syndrome coro | ···················· | ···················· | ···················· | ···················· | ···················· | ···················· |
| 2044 | MW403646.1 | Severe acute respiratory syndrome coro | ···················· | ···················· | ···················· | ···················· | ···················· | ···················· |
| 2045 | MW403648.1 | Severe acute respiratory syndrome coro | ···················· | ···················· | ···················· | ···················· | ···················· | ···················· |
| 2046 | MW403649.1 | Severe acute respiratory syndrome coro | ···················· | ···················· | ···················· | ···················· | ···················· | ···················· |
| 2047 | MW403651.1 | Severe acute respiratory syndrome coro | ·····T·············· | ···················· | ···················· | ···················· | ···················· | ···················· |
| 2048 | MW403652.1 | Severe acute respiratory syndrome coro | ···················· | ···················· | ···················· | ···················· | ···················· | ···················· |
| 2049 | MW403653.1 | Severe acute respiratory syndrome coro | ···················· | ···················· | ···················· | ···················· | ···················· | ···················· |
| 2050 | MW403655.1 | Severe acute respiratory syndrome coro | ···················· | ···················· | ···················· | ···················· | ···················· | ···················· |
| 2051 | MW403656.1 | Severe acute respiratory syndrome coro | ···················· | ···················· | ···················· | ···················· | ···················· | ···················· |
| 2052 | MW403661.1 | Severe acute respiratory syndrome coro | ···················· | ···················· | ···················· | ···················· | ···················· | ···················· |
| 2053 | MW403662.1 | Severe acute respiratory syndrome coro | ···················· | ···················· | ···················· | ···················· | ···················· | ···················· |
| 2054 | MW403663.1 | Severe acute respiratory syndrome coro | ···················· | ····G··············· | ···················· | ···················· | ···················· | ···················· |
| 2055 | MW403664.1 | Severe acute respiratory syndrome coro | ···················· | ···················· | ···················· | ···················· | ···················· | ···················· |
| 2056 | MW403665.1 | Severe acute respiratory syndrome coro | ···················· | ···················· | ···················· | ···················· | ···················· | ···················· |
| 2057 | MW403668.1 | Severe acute respiratory syndrome coro | ···················· | ···················· | ···················· | ···················· | ···················· | ···················· |
| 2058 | MW403670.1 | Severe acute respiratory syndrome coro | ···················· | ···················· | ···················· | ···················· | ···················· | ···················· |
| 2059 | MW403671.1 | Severe acute respiratory syndrome coro | ···················· | ···················· | ···················· | ···················· | ···················· | ···················· |
| 2060 | MW403672.1 | Severe acute respiratory syndrome coro | ···················· | ···················· | ···················· | ···················· | ···················· | ···················· |
| 2061 | MW403673.1 | Severe acute respiratory syndrome coro | ···················· | ···················· | ···················· | ···················· | ·········T·········· | ···················· |
| 2062 | MW403674.1 | Severe acute respiratory syndrome coro | ···················· | ···················· | ···················· | ···················· | ·········T·········· | ···················· |
| 2063 | MW403675.1 | Severe acute respiratory syndrome coro | ···················· | ···················· | ···················· | ···················· | ·········T·········· | ···················· |
| 2064 | MW403676.1 | Severe acute respiratory syndrome coro | ···················· | ···················· | ···················· | ···················· | ·········T·········· | ···················· |
| 2065 | MW403677.1 | Severe acute respiratory syndrome coro | ···················· | ···················· | ···················· | ···················· | ·········T·········· | ···················· |
| 2066 | MW403678.1 | Severe acute respiratory syndrome coro | ···················· | ···················· | ···················· | ···················· | ·········T·········· | ···················· |
| 2067 | MW403679.1 | Severe acute respiratory syndrome coro | ···················· | ···················· | ···················· | ···················· | ·········T·········· | ···················· |
| 2068 | MW403681.1 | Severe acute respiratory syndrome coro | ···················· | ···················· | ···················· | ···················· | ·········T·········· | ···················· |
| 2069 | MW403682.1 | Severe acute respiratory syndrome coro | ···················· | ···················· | ···················· | ···················· | ·········T·········· | ···················· |
| 2070 | MW403683.1 | Severe acute respiratory syndrome coro | ···················· | ···················· | ···················· | ···················· | ·········T·········· | ···················· |
| 2071 | MW403684.1 | Severe acute respiratory syndrome coro | ···················· | ···················· | ···················· | ···················· | ·········T·········· | ···················· |
| 2072 | MW403685.1 | Severe acute respiratory syndrome coro | ···················· | ···················· | ···················· | ···················· | ·········T·········· | ···················· |
| 2073 | MW403686.1 | Severe acute respiratory syndrome coro | ···················· | ···················· | ···················· | ···················· | ·········T·········· | ···················· |
| 2074 | MW403687.1 | Severe acute respiratory syndrome coro | ···················· | ···················· | ···················· | ···················· | ·········T·········· | ···················· |
| 2075 | MW403688.1 | Severe acute respiratory syndrome coro | ···················· | ···················· | ···················· | ···················· | ·········T·········· | ···················· |
| 2076 | MW403689.1 | Severe acute respiratory syndrome coro | ···················· | ···················· | ···················· | ···················· | ·········T·········· | ···················· |
| 2077 | MW403690.1 | Severe acute respiratory syndrome coro | ···················· | ···················· | ···················· | ···················· | ·········T·········· | ···················· |
| 2078 | MW403691.1 | Severe acute respiratory syndrome coro | ···················· | ···················· | ···················· | ···················· | ·········T·········· | ···················· |
| 2079 | MW403692.1 | Severe acute respiratory syndrome coro | ···················· | ···················· | ···················· | ···················· | ···················· | ···················· |
| 2080 | MW403693.1 | Severe acute respiratory syndrome coro | ···················· | ···················· | ···················· | ···················· | ···················· | ···················· |
| 2081 | MW403694.1 | Severe acute respiratory syndrome coro | ···················· | ···················· | ···················· | ···················· | ···················· | ···················· |
| 2082 | MW403695.1 | Severe acute respiratory syndrome coro | ···················· | ···················· | ···················· | ···················· | ···················· | ···················· |
| 2083 | MW403696.1 | Severe acute respiratory syndrome coro | ···················· | ···················· | ···················· | ···················· | ···················· | ···················· |
| 2084 | MW403697.1 | Severe acute respiratory syndrome coro | ···················· | ···················· | ···················· | ···················· | ···················· | ···················· |
| 2085 | MW403698.1 | Severe acute respiratory syndrome coro | ···················· | ···················· | ···················· | ···················· | ···················· | ···················· |
| 2086 | MW403699.1 | Severe acute respiratory syndrome coro | ···················· | ···················· | ···················· | ···················· | ···················· | ···················· |
| 2087 | MW403700.1 | Severe acute respiratory syndrome coro | ···················· | ···················· | ···················· | ···················· | ···················· | ···················· |
| 2089 | MW403702.1 | Severe acute respiratory syndrome coro | ···················· | ···················· | ···················· | ···················· | ···················· | ···················· |
| 2090 | MW403703.1 | Severe acute respiratory syndrome coro | ···················· | ···················· | ···················· | ···················· | ···················· | ···················· |
| 2091 | MW403763.1 | Severe acute respiratory syndrome coro | ···················· | ···················· | ···················· | ···················· | ···················· | ···················· |
| 2092 | MW403764.1 | Severe acute respiratory syndrome coro | ···················· | ···················· | ···················· | ···················· | ···················· | ···················· |
| 2093 | MW403765.1 | Severe acute respiratory syndrome coro | ···················· | ···················· | ···················· | ···················· | ···················· | ···················· |
| 2094 | MW403766.1 | Severe acute respiratory syndrome coro | ···················· | ···················· | ···················· | ···················· | ···················· | ···················· |
| 2095 | MW403767.1 | Severe acute respiratory syndrome coro | ···················· | ···················· | ···················· | ···················· | ···················· | ···················· |
| 2096 | MW403768.1 | Severe acute respiratory syndrome coro | ···················· | ···················· | ···················· | ···················· | ···················· | ···················· |
| 2097 | MW403769.1 | Severe acute respiratory syndrome coro | ···················· | ···················· | ···················· | ···················· | ···················· | ···················· |
| 2098 | MW064259.1 | Severe acute respiratory syndrome coro | ···················· | ···················· | ···················· | ···················· | ···················· | ···················· |
| 2099 | MW064260.1 | Severe acute respiratory syndrome coro | ···················· | ···················· | ···················· | ···················· | ···················· | ···················· |
| 2100 | MW064263.1 | Severe acute respiratory syndrome coro | ···················· | ···················· | ···················· | ···················· | ···················· | ···················· |
| 2101 | MW064264.1 | Severe acute respiratory syndrome coro | ···················· | ···················· | ···················· | ···················· | ···················· | ···················· |

| | | | | | | | | |
|---|---|---|---|---|---|---|---|---|
| 2102 | MW389566.1 | \|Severe acute respiratory syndrome coro | ···················· | ···················· | ···················· | ···················· | ···················· | ···················· |
| 2103 | MW389567.1 | \|Severe acute respiratory syndrome coro | ···················· | ····T··············· | ···················· | ···················· | ···················· | ···················· |
| 2104 | MW389569.1 | \|Severe acute respiratory syndrome coro | ···················· | ····T··············· | ···················· | ···················· | ···················· | ···················· |
| 2105 | MW389570.1 | \|Severe acute respiratory syndrome coro | ···················· | ····T··············· | ···················· | ···················· | ···················· | ···················· |
| 2106 | MW389571.1 | \|Severe acute respiratory syndrome coro | ···················· | ···················· | ···················· | ···················· | ···················· | ···················· |
| 2107 | MW389572.1 | \|Severe acute respiratory syndrome coro | ···················· | ····T··············· | ···················· | ···················· | ···················· | ···················· |
| 2108 | MW389573.1 | \|Severe acute respiratory syndrome coro | ···················· | ···················· | ···················· | ···················· | ···················· | ···················· |
| 2109 | MW389574.1 | \|Severe acute respiratory syndrome coro | ···················· | ····T··············· | ···················· | ···················· | ···················· | ···················· |
| 2110 | MW389575.1 | \|Severe acute respiratory syndrome coro | ···················· | ····T··············· | ···················· | ···················· | ···················· | ···················· |
| 2111 | MW389576.1 | \|Severe acute respiratory syndrome coro | ···················· | ···················· | ···················· | ···················· | ···················· | ···················· |
| 2112 | MW389579.1 | \|Severe acute respiratory syndrome coro | ···················· | ···················· | ···················· | ···················· | ···················· | ···················· |
| 2113 | MW389583.1 | \|Severe acute respiratory syndrome coro | ···················· | ···················· | ···················· | ···················· | ···················· | ···················· |
| 2114 | MW389598.1 | \|Severe acute respiratory syndrome coro | ···················· | ···················· | ···················· | ···················· | ···················· | ···················· |
| 2115 | MW389599.1 | \|Severe acute respiratory syndrome coro | ···················· | ···················· | ···················· | ···················· | ···················· | ···················· |
| 2116 | MW389600.1 | \|Severe acute respiratory syndrome coro | ···················· | ···················· | ···················· | ···················· | ···················· | ···················· |
| 2117 | MW389602.1 | \|Severe acute respiratory syndrome coro | ···················· | ···················· | ···················· | ···················· | ···················· | ···················· |
| 2118 | MW389603.1 | \|Severe acute respiratory syndrome coro | ···················· | ···················· | ···················· | ···················· | ···················· | ···················· |
| 2119 | MW389604.1 | \|Severe acute respiratory syndrome coro | ···················· | ···················· | ···················· | ···················· | ···················· | ···················· |
| 2120 | MW389605.1 | \|Severe acute respiratory syndrome coro | ···················· | ···················· | ···················· | ···················· | ···················· | ···················· |
| 2121 | MW389606.1 | \|Severe acute respiratory syndrome coro | ···················· | ···················· | ···················· | ···················· | ···················· | ···················· |
| 2122 | MW389607.1 | \|Severe acute respiratory syndrome coro | ···················· | ···················· | ···················· | ···················· | ···················· | ···················· |
| 2123 | MW389608.1 | \|Severe acute respiratory syndrome coro | ···················· | ····T··············· | ···················· | ···················· | ···················· | ···················· |
| 2124 | MW389609.1 | \|Severe acute respiratory syndrome coro | ···················· | ···················· | ···················· | ···················· | ···················· | ···················· |
| 2125 | MW389611.1 | \|Severe acute respiratory syndrome coro | ···················· | ···················· | ···················· | ···················· | ···················· | ···················· |
| 2126 | MW389612.1 | \|Severe acute respiratory syndrome coro | ···················· | ···················· | ···················· | ···················· | ···················· | ···················· |
| 2127 | MW389613.1 | \|Severe acute respiratory syndrome coro | ···················· | ·A·················· | ···················· | ···················· | ···················· | ···················· |
| 2128 | MW389614.1 | \|Severe acute respiratory syndrome coro | ···················· | ·A·················· | ···················· | ···················· | ···················· | ···················· |
| 2129 | MW389615.1 | \|Severe acute respiratory syndrome coro | ···················· | ·A·················· | ···················· | ···················· | ···················· | ···················· |
| 2130 | MW389616.1 | \|Severe acute respiratory syndrome coro | ···················· | ···················· | ···················· | ···················· | ···················· | ···················· |
| 2131 | MW389618.1 | \|Severe acute respiratory syndrome coro | ···················· | ···················· | ···················· | ···················· | ···················· | ···················· |
| 2132 | MW389619.1 | \|Severe acute respiratory syndrome coro | ···················· | ·A·················· | ···················· | ···················· | ···················· | ···················· |
| 2133 | MW389621.1 | \|Severe acute respiratory syndrome coro | ···················· | ·A·················· | ···················· | ···················· | ···················· | ···················· |
| 2134 | MW389622.1 | \|Severe acute respiratory syndrome coro | ···················· | ···················· | ···················· | ···················· | ···················· | ···················· |
| 2135 | MW389623.1 | \|Severe acute respiratory syndrome coro | ···················· | ···················· | ···················· | ···················· | ···················· | ···················· |
| 2136 | MW389624.1 | \|Severe acute respiratory syndrome coro | ···················· | ·A·················· | ···················· | ···················· | ···················· | ···················· |
| 2137 | MW389625.1 | \|Severe acute respiratory syndrome coro | ···················· | ···················· | ···················· | ···················· | ···················· | ···················· |
| 2138 | MW389626.1 | \|Severe acute respiratory syndrome coro | ···················· | ····T··············· | ···················· | ···················· | ···················· | ···················· |
| 2139 | MW389627.1 | \|Severe acute respiratory syndrome coro | ···················· | ·A·················· | ···················· | ···················· | ···················· | ···················· |
| 2140 | MW389628.1 | \|Severe acute respiratory syndrome coro | ···················· | ···················· | ···················· | ···················· | ···················· | ···················· |
| 2141 | MW389630.1 | \|Severe acute respiratory syndrome coro | ···················· | ····T··············· | ···················· | ···················· | ···················· | ···················· |
| 2142 | MW389631.1 | \|Severe acute respiratory syndrome coro | ···················· | ···················· | ···················· | ···················· | ···················· | ···················· |
| 2143 | MW389635.1 | \|Severe acute respiratory syndrome coro | ···················· | ·A·················· | ···················· | ···················· | ···················· | ···················· |
| 2144 | MW389636.1 | \|Severe acute respiratory syndrome coro | ···················· | ···················· | ···················· | ···················· | ···················· | ···················· |
| 2145 | MW389638.1 | \|Severe acute respiratory syndrome coro | ···················· | ···················· | ···················· | ···················· | ···················· | ···················· |
| 2146 | MW389639.1 | \|Severe acute respiratory syndrome coro | ···················· | ···················· | ···················· | ···················· | ···················· | ···················· |
| 2147 | MW389640.1 | \|Severe acute respiratory syndrome coro | ···················· | ···················· | ···················· | ···················· | ···················· | ···················· |
| 2148 | MW389641.1 | \|Severe acute respiratory syndrome coro | ···················· | ·A·················· | ···················· | ···················· | ···················· | ···················· |
| 2149 | MW389644.1 | \|Severe acute respiratory syndrome coro | ···················· | ···················· | ···················· | ···················· | ···················· | ···················· |
| 2150 | MW389646.1 | \|Severe acute respiratory syndrome coro | ···················· | ·A·················· | ···················· | ···················· | ···················· | ···················· |
| 2151 | MW389649.1 | \|Severe acute respiratory syndrome coro | ···················· | ·A·················· | ···················· | ···················· | ···················· | ···················· |
| 2152 | MW390832.1 | \|Severe acute respiratory syndrome coro | ···················· | ···················· | ···················· | ···················· | ···················· | ···················· |
| 2153 | MW390833.1 | \|Severe acute respiratory syndrome coro | ···················· | ···················· | ···················· | ···················· | ···················· | ···················· |
| 2154 | MW390834.1 | \|Severe acute respiratory syndrome coro | ···················· | ···················· | ···················· | ···················· | ···················· | ···················· |
| 2155 | MW390835.1 | \|Severe acute respiratory syndrome coro | ···················· | ···················· | ···················· | ···················· | ···················· | ···················· |
| 2156 | MW390836.1 | \|Severe acute respiratory syndrome coro | ···················· | ·A·················· | ···················· | ···················· | ···················· | ···················· |
| 2157 | MW390837.1 | \|Severe acute respiratory syndrome coro | ···················· | ···················· | ···················· | ···················· | ···················· | ···················· |
| 2158 | MW390838.1 | \|Severe acute respiratory syndrome coro | ···················· | ···················· | ···················· | ···················· | ···················· | ···················· |
| 2159 | MW390839.1 | \|Severe acute respiratory syndrome coro | ···················· | ···················· | ···················· | ···················· | ···················· | ···················· |
| 2160 | MW390840.1 | \|Severe acute respiratory syndrome coro | ···················· | ···················· | ···················· | ···················· | ···················· | ···················· |
| 2161 | MW390841.1 | \|Severe acute respiratory syndrome coro | ···················· | ···················· | ···················· | ···················· | ···················· | ···················· |
| 2162 | MW390842.1 | \|Severe acute respiratory syndrome coro | ···················· | ···················· | ···················· | ···················· | ···················· | ···················· |
| 2163 | MW390843.1 | \|Severe acute respiratory syndrome coro | ···················· | ···················· | ···················· | ···················· | ···················· | ···················· |
| 2164 | MW390844.1 | \|Severe acute respiratory syndrome coro | ···················· | ···················· | ···················· | ···················· | ···················· | ···················· |
| 2165 | MW390845.1 | \|Severe acute respiratory syndrome coro | ···················· | ···················· | ···················· | ···················· | ···················· | ···················· |
| 2166 | MW390846.1 | \|Severe acute respiratory syndrome coro | ···················· | ···················· | ···················· | ···················· | ···················· | ···················· |
| 2167 | MW390847.1 | \|Severe acute respiratory syndrome coro | ···················· | ···················· | ···················· | ···················· | ···················· | ···················· |
| 2168 | MW390848.1 | \|Severe acute respiratory syndrome coro | ···················· | ···················· | ···················· | ···················· | ···················· | ···················· |
| 2169 | MW390849.1 | \|Severe acute respiratory syndrome coro | ···················· | ···················· | ···················· | ···················· | ···················· | ···················· |
| 2170 | MW390850.1 | \|Severe acute respiratory syndrome coro | ···················· | ···················· | ···················· | ···················· | ···················· | ···················· |
| 2171 | MW390851.1 | \|Severe acute respiratory syndrome coro | ···················· | ···················· | ···················· | ···················· | ···················· | ···················· |
| 2172 | MW390852.1 | \|Severe acute respiratory syndrome coro | ···················· | ···················· | ···················· | ···················· | ···················· | ···················· |
| 2173 | MW390853.1 | \|Severe acute respiratory syndrome coro | ···················· | ···················· | ···················· | ···················· | ···················· | ···················· |
| 2174 | MW390854.1 | \|Severe acute respiratory syndrome coro | ···················· | ···················· | ···················· | ···················· | ···················· | ···················· |
| 2175 | MW390855.1 | \|Severe acute respiratory syndrome coro | ···················· | ···················· | ···················· | ···················· | ···················· | ···················· |
| 2176 | MW390856.1 | \|Severe acute respiratory syndrome coro | ···················· | ···················· | ···················· | ···················· | ···················· | ···················· |
| 2177 | MW390857.1 | \|Severe acute respiratory syndrome coro | ···················· | ···················· | ···················· | ···················· | ···················· | ···················· |
| 2178 | MW390858.1 | \|Severe acute respiratory syndrome coro | ···················· | ···················· | ···················· | ···················· | ···················· | ···················· |
| 2179 | MW390859.1 | \|Severe acute respiratory syndrome coro | ···················· | ···················· | ···················· | ···················· | ···················· | ···················· |

```
2180 MW390860.1 |Severe acute respiratory syndrome coro    ....................  ....................  ....................  ....................  ....................  ....................
2181 MW390861.1 |Severe acute respiratory syndrome coro    ....................  ....................  ....................  ....................  ....................  ....................
2182 MW390862.1 |Severe acute respiratory syndrome coro    ....................  ....................  ....................  ....................  ....................  ....................
2183 MW390863.1 |Severe acute respiratory syndrome coro    ....................  ....................  ....................  ....................  ....................  ....................
2184 MW390864.1 |Severe acute respiratory syndrome coro    ....................  ....................  ....................  ....................  ....................  ....................
2185 MW390865.1 |Severe acute respiratory syndrome coro    ....................  ....................  ....................  ....................  ....................  ....................
2187 MW390867.1 |Severe acute respiratory syndrome coro    ....................  ....................  ....................  ....................  ....................  ....................
2188 MW390868.1 |Severe acute respiratory syndrome coro    ....................  ....................  ....................  ....................  ....................  ....................
2189 MW384414.1 |Severe acute respiratory syndrome coro    ....................  ....................  ....................  ....................  ....................  ....................
2190 MW384415.1 |Severe acute respiratory syndrome coro    ....................  ....................  ....................  ....................  ....................  ....................
2191 MW384417.1 |Severe acute respiratory syndrome coro    ....................  ....................  ....................  ....................  ....................  ....................
2192 MW384418.1 |Severe acute respiratory syndrome coro    ....................  ....................  ....................  ....................  ....................  ....................
2193 MW384419.1 |Severe acute respiratory syndrome coro    ....................  ....................  ....................  ....................  ....................  ....................
2194 MW384420.1 |Severe acute respiratory syndrome coro    ....................  ....................  ....................  ....................  ....................  ....................
2195 MW384421.1 |Severe acute respiratory syndrome coro    ....................  ....................  ....................  ....................  ....................  ....................
2196 MW384422.1 |Severe acute respiratory syndrome coro    ....................  ....................  ....................  ....................  ....................  ....................
2197 MW384423.1 |Severe acute respiratory syndrome coro    ....................  ....................  ....................  ....................  ....................  ....................
2198 MW384424.1 |Severe acute respiratory syndrome coro    ....................  ....................  ....................  ....................  ....................  ....................
2199 MW384425.1 |Severe acute respiratory syndrome coro    ....................  ....................  ....................  ....................  ....................  ....................
2200 MW384426.1 |Severe acute respiratory syndrome coro    ....................  ....................  ....................  ....................  ....................  ....................
2201 MW384427.1 |Severe acute respiratory syndrome coro    ....................  .A..................  ....................  ....................  ....................  ....................
2202 MW386175.1 |Severe acute respiratory syndrome coro    ....................  ....................  ....................  ....................  ....................  ....................
2203 MW386176.1 |Severe acute respiratory syndrome coro    ....................  ....................  ....................  ....................  ....................  ....................
2204 MW386177.1 |Severe acute respiratory syndrome coro    ....................  ....................  ....................  ....................  ....................  ....................
2205 MW375726.1 |Severe acute respiratory syndrome coro    ....................  ....................  ....................  ....................  ....................  ....................
2206 MW375727.1 |Severe acute respiratory syndrome coro    ....................  ....................  ....................  ....................  ....................  ....................
2207 MW375728.1 |Severe acute respiratory syndrome coro    ....................  ....................  ....................  ....................  ....................  ....................
2208 MW375729.1 |Severe acute respiratory syndrome coro    ....................  ....................  ....................  ....................  ....................  ....................
2209 MW375730.1 |Severe acute respiratory syndrome coro    ....................  ....................  ....................  ....................  ....................  ....................
2210 MW375731.1 |Severe acute respiratory syndrome coro    ....................  ....................  ....................  ....................  ....................  ....................
2211 MW375993.1 |Severe acute respiratory syndrome coro    ....................  ....................  ....................  ....................  ....................  ....................
2212 MW375994.1 |Severe acute respiratory syndrome coro    ....................  ....................  ....................  ....................  ....................  ....................
2213 MW368700.1 |Severe acute respiratory syndrome coro    ....................  .A..................  ....................  ....................  ....................  ..A.................
2214 MW374904.1 |Severe acute respiratory syndrome coro    ....................  ....................  ....................  ....................  ....................  ....................
2215 MW374905.1 |Severe acute respiratory syndrome coro    ....................  ....................  ....................  ....................  ....................  ....................
2216 MW374906.1 |Severe acute respiratory syndrome coro    ....................  ....................  ....................  ....................  ....................  ....................
2217 MW374907.1 |Severe acute respiratory syndrome coro    ....................  .A..................  ....................  ....................  ....................  ....................
2218 MW374908.1 |Severe acute respiratory syndrome coro    ....................  .A..................  ....................  ....................  ....................  ....................
2219 MW374909.1 |Severe acute respiratory syndrome coro    ....................  ....................  ....................  ....................  ....................  ....................
2220 MW374910.1 |Severe acute respiratory syndrome coro    ....................  ....................  ....................  ....................  ....................  ....................
2221 MW374911.1 |Severe acute respiratory syndrome coro    ....................  ....................  ....................  ....................  ....................  ....................
2222 MW374912.1 |Severe acute respiratory syndrome coro    ....................  ....................  ....................  ....................  ....................  ....................
2223 MW374913.1 |Severe acute respiratory syndrome coro    ....................  ....................  ....................  ....................  ....................  ....................
2224 MW374914.1 |Severe acute respiratory syndrome coro    ....................  ....................  ....................  ....................  ....................  ....................
2225 MW374915.1 |Severe acute respiratory syndrome coro    ....................  ....................  ....................  ....................  ....................  ....................
2226 MW374916.1 |Severe acute respiratory syndrome coro    ....................  ....................  ....................  ....................  ....................  ....................
2227 MW374917.1 |Severe acute respiratory syndrome coro    ....................  ..C.................  ....................  ....................  ....................  ....................
2228 MW374918.1 |Severe acute respiratory syndrome coro    ....................  ....................  ....................  ....................  ....................  ....................
2229 MW374919.1 |Severe acute respiratory syndrome coro    ....................  ....................  ....................  ....................  ....................  ....................
2230 MW374920.1 |Severe acute respiratory syndrome coro    ....................  ....................  ....................  ....................  ....................  ....................
2231 MW374921.1 |Severe acute respiratory syndrome coro    ....................  .A..................  ....................  ....................  ....................  ....................
2232 MW374922.1 |Severe acute respiratory syndrome coro    ....................  ....................  ....................  ....................  ....................  ....................
2233 MW374923.1 |Severe acute respiratory syndrome coro    ....................  ....................  ....................  ....................  ....................  ....................
2234 MW374924.1 |Severe acute respiratory syndrome coro    ....................  .A..................  ....................  ....................  ....................  ....................
2235 MW374925.1 |Severe acute respiratory syndrome coro    ....................  ....................  ....................  ....................  ....................  ....................
2236 MW374926.1 |Severe acute respiratory syndrome coro    ....................  .A..................  ....................  ....................  ....................  ....................
2237 MW374927.1 |Severe acute respiratory syndrome coro    ....................  ....................  ....................  ....................  ....................  ....................
2238 MW374928.1 |Severe acute respiratory syndrome coro    ....................  .A..................  ....................  ....................  ....................  ....................
2239 MW374929.1 |Severe acute respiratory syndrome coro    ....................  ....................  ....................  ....................  ....................  ....................
2240 MW374930.1 |Severe acute respiratory syndrome coro    ....................  ....................  ....................  ....................  ....................  ....................
2241 MW374931.1 |Severe acute respiratory syndrome coro    ....................  ....................  ....................  ....................  ....................  ....................
2242 MW374932.1 |Severe acute respiratory syndrome coro    ....................  ....................  ....................  ....................  ....................  ....................
2243 MW374933.1 |Severe acute respiratory syndrome coro    ....................  ....................  ....................  ....................  ....................  ....................
2244 MW374934.1 |Severe acute respiratory syndrome coro    ....................  ....................  ....................  ....................  ....................  ....................
2245 MW374935.1 |Severe acute respiratory syndrome coro    ....................  ....................  ....................  ....................  ....................  ....................
2246 MW374936.1 |Severe acute respiratory syndrome coro    ....................  ....................  ....................  ....................  ....................  ....................
2247 MW374937.1 |Severe acute respiratory syndrome coro    ....................  ....................  ....................  ....................  ....................  ....................
2248 MW374938.1 |Severe acute respiratory syndrome coro    ....................  ....................  ....................  ....................  ....................  ....................
2249 MW374939.1 |Severe acute respiratory syndrome coro    ....................  ....................  ....................  ....................  ....................  ....................
2250 MW374940.1 |Severe acute respiratory syndrome coro    ....................  ....................  ....................  ....................  ....................  ....................
2251 MW374941.1 |Severe acute respiratory syndrome coro    ....................  ....................  ....................  ....................  ....................  ....................
2252 MW374942.1 |Severe acute respiratory syndrome coro    ....................  ....................  ....................  ....................  ....................  ....................
2253 MW374943.1 |Severe acute respiratory syndrome coro    ....................  ....................  ....................  ....................  ....................  ....................
2254 MW374944.1 |Severe acute respiratory syndrome coro    ....................  ....................  ....................  ....................  ....................  ....................
2255 MW374945.1 |Severe acute respiratory syndrome coro    ....................  ....................  ....................  ....................  ....................  ....................
2256 MW374946.1 |Severe acute respiratory syndrome coro    ........T...........  .A..................  ....................  ....................  ....................  ....................
2257 MW374947.1 |Severe acute respiratory syndrome coro    ....................  ....................  ....................  ....................  ....................  ....................
2258 MW374948.1 |Severe acute respiratory syndrome coro    ....................  ....................  ....................  ....................  ....................  ....................
```

2259 MW365356.1 |Severe acute respiratory syndrome coro ···················· ···················· ···················· ···················· ···················· ····················
2260 MW365357.1 |Severe acute respiratory syndrome coro ···················· ···················· ···················· ···················· ···················· ····················
2261 MW365358.1 |Severe acute respiratory syndrome coro ···················· ···················· ···················· ···················· ···················· ····················
2262 MW365359.1 |Severe acute respiratory syndrome coro ···················· ···················· ···················· ···················· ···················· ····················
2263 MW366724.1 |Severe acute respiratory syndrome coro ···················· ···················· ···················· ···················· ···················· ····················
2264 MW366750.1 |Severe acute respiratory syndrome coro ···················· ···················· ···················· ···················· ···················· ····················
2265 MW368439.1 |Severe acute respiratory syndrome coro ···················· ···················· ···················· ···················· ···················· ····················
2266 MW368440.1 |Severe acute respiratory syndrome coro ···················· ···················· ···················· ···················· ···················· ····················
2267 MW362553.1 |Severe acute respiratory syndrome coro ···················· ···················· ···················· ···················· ···················· ····················
2268 MW362554.1 |Severe acute respiratory syndrome coro ···················· ···················· ···················· ···················· ···················· ····················
2269 MW263334.1 |Severe acute respiratory syndrome coro ···················· ·A·················· ···················· ···················· ···················· ····················
2270 MW263335.1 |Severe acute respiratory syndrome coro ···················· ···················· ···················· ···················· ···················· ····················
2271 MW263336.1 |Severe acute respiratory syndrome coro ···················· ···················· ···················· ···················· ···················· ····················
2272 MW263337.1 |Severe acute respiratory syndrome coro ···················· ···················· ···················· ···················· ···················· ····················
2273 MW356670.1 |Severe acute respiratory syndrome coro ···················· ···················· ···················· ···················· ···················· ····················
2274 MW356671.1 |Severe acute respiratory syndrome coro ···················· ···················· ···················· ···················· ···················· ····················
2275 MW356672.1 |Severe acute respiratory syndrome coro ···················· ·A·················· ···················· ···················· ···················· ···A················
2276 MW356784.1 |Severe acute respiratory syndrome coro ···················· ···················· ···················· ···················· ···················· ····················
2277 MW356785.1 |Severe acute respiratory syndrome coro ···················· ···················· ···················· ···················· ······T············· ····················
2278 MW356786.1 |Severe acute respiratory syndrome coro ···················· ···················· ···················· ···················· ···················· ····················
2279 MW345922.1 |Severe acute respiratory syndrome coro ···················· ···················· ···················· ···················· ···················· ····················
2280 MW345923.1 |Severe acute respiratory syndrome coro ·T·················· ···················· ···················· ···················· ···················· ····················
2281 MW345924.1 |Severe acute respiratory syndrome coro ···················· ·A·················· ···················· ···················· ···················· ···A················
2282 MW345925.1 |Severe acute respiratory syndrome coro ···················· ···················· ···················· ···················· ···················· ····················
2283 MW345926.1 |Severe acute respiratory syndrome coro ···················· ····A··············· ···················· ···················· ···················· ····················
2284 MW345927.1 |Severe acute respiratory syndrome coro ···················· ····A··············· ···················· ···················· ···················· ····················
2285 MW345928.1 |Severe acute respiratory syndrome coro ···················· ···················· ···················· ···················· ···················· ····················
2286 MW345929.1 |Severe acute respiratory syndrome coro ···················· ···················· ···················· ···················· ·······N············ ····················
2287 MW345930.1 |Severe acute respiratory syndrome coro ···················· ···················· ···················· ···················· ···················· ····················
2288 MW346179.1 |Severe acute respiratory syndrome coro ···················· ···················· ···················· ···················· ···················· ····················
2289 MW346315.1 |Severe acute respiratory syndrome coro ···················· ···················· ···················· ···················· ···················· ····················
2290 MW346329.1 |Severe acute respiratory syndrome coro ···················· ···················· ···················· ···················· ···················· ····················
2291 MW346331.1 |Severe acute respiratory syndrome coro ···················· ···················· ···················· ···················· ···················· ····················
2292 MW346340.1 |Severe acute respiratory syndrome coro ···················· ···················· ···················· ···················· ···················· ····················
2293 MW346345.1 |Severe acute respiratory syndrome coro ···················· ···················· ···················· ···················· ···················· ····················
2294 MW346348.1 |Severe acute respiratory syndrome coro ···················· ···················· ···················· ···················· ······T············· ····················
2295 MW346358.1 |Severe acute respiratory syndrome coro ···················· ···················· ···················· ···················· ···················· ····················
2296 MW346365.1 |Severe acute respiratory syndrome coro ···················· ···················· ···················· ···················· ···················· ····················
2297 MW346366.1 |Severe acute respiratory syndrome coro ···················· ···················· ···················· ···················· ···················· ····················
2298 MW346369.1 |Severe acute respiratory syndrome coro ···················· ···················· ···················· ···················· ···················· ····················
2299 MW346370.1 |Severe acute respiratory syndrome coro ···················· ···················· ···················· ···················· ···················· ····················
2300 MW346371.1 |Severe acute respiratory syndrome coro ···················· ···················· ···················· ···················· ···················· ····················
2301 MW349032.1 |Severe acute respiratory syndrome coro ···················· ···················· ···················· ···················· ···················· ····················
2302 MW349033.1 |Severe acute respiratory syndrome coro ···················· ···················· ···················· ···················· ···················· ····················
2303 MW349034.1 |Severe acute respiratory syndrome coro ···················· ···················· ···················· ···················· ···················· ····················
2304 MW349035.1 |Severe acute respiratory syndrome coro ···················· ···················· ···················· ···················· ···················· ····················
2305 MW349036.1 |Severe acute respiratory syndrome coro ···················· ···················· ···················· ···················· ···················· ····················
2306 MW349037.1 |Severe acute respiratory syndrome coro ···················· ···················· ···················· ···················· ···················· ····················
2307 MW349038.1 |Severe acute respiratory syndrome coro ···················· ···················· ···················· ···················· ···················· ····················
2308 MW349039.1 |Severe acute respiratory syndrome coro ···················· ···················· ···················· ···················· ···················· ····················
2309 MW349040.1 |Severe acute respiratory syndrome coro ···················· ···················· ···················· ···················· ···················· ····················
2310 MW349041.1 |Severe acute respiratory syndrome coro ···················· ···················· ···················· ···················· ···················· ····················
2311 MW349042.1 |Severe acute respiratory syndrome coro ···················· ···················· ···················· ···················· ···················· ····················
2312 MW349043.1 |Severe acute respiratory syndrome coro ···················· ···················· ···················· ···················· ···················· ····················
2313 MW349044.1 |Severe acute respiratory syndrome coro ···················· ···················· ···················· ···················· ···················· ····················
2314 MW349045.1 |Severe acute respiratory syndrome coro ···················· ···················· ···················· ···················· ···················· ····················
2315 MW349046.1 |Severe acute respiratory syndrome coro ···················· ···················· ···················· ···················· ···················· ····················
2316 MW349047.1 |Severe acute respiratory syndrome coro ···················· ···················· ···················· ···················· ···················· ····················
2317 MW349048.1 |Severe acute respiratory syndrome coro ···················· ···················· ···················· ···················· ···················· ····················
2318 MW349049.1 |Severe acute respiratory syndrome coro ···················· ···················· ···················· ···················· ···················· ····················
2319 MW349050.1 |Severe acute respiratory syndrome coro ···················· ···················· ···················· ···················· ···················· ····················
2320 MW349051.1 |Severe acute respiratory syndrome coro ···················· ···················· ···················· ···················· ··NN················ ····················
2321 MW349052.1 |Severe acute respiratory syndrome coro ···················· ···················· ···················· ···················· ···················· ····················
2322 MW349053.1 |Severe acute respiratory syndrome coro ···················· ···················· ···················· ···················· ···················· ····················
2323 MW349054.1 |Severe acute respiratory syndrome coro ···················· ···················· ···················· ···················· ···················· ····················
2324 MW349055.1 |Severe acute respiratory syndrome coro ···················· ···················· ···················· ···················· ···················· ····················
2325 MW349056.1 |Severe acute respiratory syndrome coro ···················· ···················· ···················· ···················· ···················· ····················
2326 MW349057.1 |Severe acute respiratory syndrome coro ···················· ···················· ···················· ···················· ···················· ····················
2327 MW349058.1 |Severe acute respiratory syndrome coro ···················· ···················· ···················· ···················· ···················· ····················
2328 MW349059.1 |Severe acute respiratory syndrome coro ···················· ···················· ···················· ···················· ···················· ····················
2329 MW349060.1 |Severe acute respiratory syndrome coro ···················· ···················· ···················· ···················· ···················· ····················
2330 MW349061.1 |Severe acute respiratory syndrome coro ···················· ···················· ···················· ···················· ···················· ····················
2331 MW349062.1 |Severe acute respiratory syndrome coro ···················· ·A·················· ···················· ···················· ···················· ····················
2332 MW349063.1 |Severe acute respiratory syndrome coro ···················· ···················· ···················· ···················· ···················· ····················
2333 MW349064.1 |Severe acute respiratory syndrome coro ···················· ···················· ···················· ···················· ···················· ····················
2334 MW349065.1 |Severe acute respiratory syndrome coro ···················· ···················· ···················· ···················· ···················· ····················
2335 MW349066.1 |Severe acute respiratory syndrome coro ···················· ···················· ···················· ···················· ···················· ····················
2336 MW349067.1 |Severe acute respiratory syndrome coro ···················· ···················· ···················· ···················· ···················· ····················

| | | | | | | | | |
|---|---|---|---|---|---|---|---|---|
| 2337 | MW349068.1 | \|Severe acute respiratory syndrome coro | ···················· | ···················· | ···················· | ···················· | ···················· | ···················· |
| 2338 | MW349069.1 | \|Severe acute respiratory syndrome coro | ···················· | ·A·················· | ···················· | ···················· | ···················· | ···················· |
| 2339 | MW349070.1 | \|Severe acute respiratory syndrome coro | ···················· | ·A·················· | ···················· | ···················· | ···················· | ···················· |
| 2340 | MW349071.1 | \|Severe acute respiratory syndrome coro | ···················· | ·A·················· | ···················· | ···················· | ···················· | ···················· |
| 2341 | MW349072.1 | \|Severe acute respiratory syndrome coro | ···················· | ·A·················· | ···················· | ···················· | ···················· | ···················· |
| 2342 | MW349073.1 | \|Severe acute respiratory syndrome coro | ···················· | ·A·················· | ···················· | ···················· | ···················· | ···················· |
| 2343 | MW349074.1 | \|Severe acute respiratory syndrome coro | ···················· | ·A·················· | ···················· | ···················· | ···················· | ···················· |
| 2344 | MW349075.1 | \|Severe acute respiratory syndrome coro | ···················· | ·A·················· | ···················· | ···················· | ···················· | ···················· |
| 2345 | MW349076.1 | \|Severe acute respiratory syndrome coro | ···················· | ·A·················· | ···················· | ···················· | ···················· | ···················· |
| 2346 | MW349077.1 | \|Severe acute respiratory syndrome coro | ···················· | ·A·················· | ···················· | ···················· | ···················· | ···················· |
| 2347 | MW349078.1 | \|Severe acute respiratory syndrome coro | ···················· | ·A·················· | ···················· | ···················· | ···················· | ···················· |
| 2348 | MW349079.1 | \|Severe acute respiratory syndrome coro | ···················· | ·A·················· | ···················· | ···················· | ···················· | ···················· |
| 2349 | MW349080.1 | \|Severe acute respiratory syndrome coro | ···················· | ·A·················· | ···················· | ···················· | ···················· | ···················· |
| 2350 | MW349081.1 | \|Severe acute respiratory syndrome coro | ···················· | ···················· | ···················· | ···················· | ···················· | ···················· |
| 2351 | MW349082.1 | \|Severe acute respiratory syndrome coro | ···················· | ·A·················· | ···················· | ···················· | ···················· | ···················· |
| 2352 | MW349083.1 | \|Severe acute respiratory syndrome coro | ···················· | ·A·················· | ···················· | ···················· | ···················· | ···················· |
| 2353 | MW349084.1 | \|Severe acute respiratory syndrome coro | ···················· | ·A·················· | ···················· | ···················· | ···················· | ···················· |
| 2354 | MW349085.1 | \|Severe acute respiratory syndrome coro | ···················· | ···················· | ···················· | ···················· | ···················· | ···················· |
| 2355 | MW349086.1 | \|Severe acute respiratory syndrome coro | ···················· | ·A·················· | ···················· | ···················· | ···················· | ···················· |
| 2356 | MW349087.1 | \|Severe acute respiratory syndrome coro | ···················· | ···················· | ···················· | ···················· | ···················· | ···················· |
| 2357 | MW349088.1 | \|Severe acute respiratory syndrome coro | ···················· | ·A·················· | ···················· | ···················· | ···················· | ···················· |
| 2358 | MW349089.1 | \|Severe acute respiratory syndrome coro | ···················· | ·A·················· | ···················· | ···················· | ···················· | ···················· |
| 2359 | MW349090.1 | \|Severe acute respiratory syndrome coro | ···················· | ·A·················· | ···················· | ···················· | ···················· | ···················· |
| 2360 | MW349091.1 | \|Severe acute respiratory syndrome coro | ···················· | ·A·················· | ···················· | ···················· | ···················· | ···················· |
| 2361 | MW349092.1 | \|Severe acute respiratory syndrome coro | ···················· | ···················· | ···················· | ···················· | ···················· | ···················· |
| 2362 | MW349093.1 | \|Severe acute respiratory syndrome coro | ···················· | ·A·················· | ···················· | ···················· | ···················· | ···················· |
| 2363 | MW349094.1 | \|Severe acute respiratory syndrome coro | ···················· | ···················· | ···················· | ···················· | ···················· | ···················· |
| 2364 | MW349095.1 | \|Severe acute respiratory syndrome coro | ···················· | ···················· | ···················· | ···················· | ···················· | ···················· |
| 2365 | MW349096.1 | \|Severe acute respiratory syndrome coro | ···················· | ···················· | ···················· | ···················· | ···················· | ···················· |
| 2366 | MW349097.1 | \|Severe acute respiratory syndrome coro | ···················· | ···················· | ···················· | ···················· | ···················· | ···R················ |
| 2367 | MW349098.1 | \|Severe acute respiratory syndrome coro | ···················· | ···················· | ···················· | ···················· | ···················· | ···················· |
| 2368 | MW349099.1 | \|Severe acute respiratory syndrome coro | ···················· | ···················· | ···················· | ···················· | ···················· | ···················· |
| 2369 | MW349100.1 | \|Severe acute respiratory syndrome coro | ···················· | ·A·················· | ···················· | ···················· | ···················· | ···················· |
| 2370 | MW349101.1 | \|Severe acute respiratory syndrome coro | ···················· | ···················· | ···················· | ···················· | ···················· | ···················· |
| 2371 | MW349102.1 | \|Severe acute respiratory syndrome coro | ···················· | ···················· | ···················· | ···················· | ···················· | ···················· |
| 2372 | MW349103.1 | \|Severe acute respiratory syndrome coro | ···················· | ···················· | ···················· | ···················· | ···················· | ···················· |
| 2373 | MW349104.1 | \|Severe acute respiratory syndrome coro | ···················· | ···················· | ···················· | ···················· | ···················· | ···················· |
| 2374 | MW341443.1 | \|Severe acute respiratory syndrome coro | ···················· | ···················· | ···················· | ···················· | ···················· | ···················· |
| 2375 | MW341462.1 | \|Severe acute respiratory syndrome coro | ···················· | ···················· | ···················· | ···················· | ···················· | ···················· |
| 2376 | MW341463.1 | \|Severe acute respiratory syndrome coro | ···················· | ···················· | ···················· | ···················· | ···················· | ···················· |
| 2377 | MW341464.1 | \|Severe acute respiratory syndrome coro | ···················· | ···················· | ···················· | ···················· | ···················· | ···················· |
| 2378 | MW341465.1 | \|Severe acute respiratory syndrome coro | ···················· | ···················· | ···················· | ···················· | ···················· | ···················· |
| 2379 | MW341466.1 | \|Severe acute respiratory syndrome coro | ···················· | ···················· | ···················· | ···················· | ···················· | ···················· |
| 2380 | MW341468.1 | \|Severe acute respiratory syndrome coro | ···················· | ···················· | ···················· | ···················· | ···················· | ···················· |
| 2381 | MW341469.1 | \|Severe acute respiratory syndrome coro | ···················· | ···················· | ···················· | ···················· | ···················· | ···················· |
| 2382 | MW341470.1 | \|Severe acute respiratory syndrome coro | ···················· | ···················· | ···················· | ···················· | ···················· | ···················· |
| 2383 | MW341471.1 | \|Severe acute respiratory syndrome coro | ···················· | ···················· | ···················· | ···················· | ···················· | ···················· |
| 2384 | MW341472.1 | \|Severe acute respiratory syndrome coro | ···················· | ···················· | ···················· | ···················· | ···················· | ···················· |
| 2385 | MW341473.1 | \|Severe acute respiratory syndrome coro | ···················· | ···················· | ···················· | ···················· | ···················· | ···················· |
| 2386 | MW341474.1 | \|Severe acute respiratory syndrome coro | ···················· | ···················· | ···················· | ···················· | ···················· | ···················· |
| 2387 | MW341475.1 | \|Severe acute respiratory syndrome coro | ···················· | ···················· | ···················· | ···················· | ···················· | ···················· |
| 2388 | MW341476.1 | \|Severe acute respiratory syndrome coro | ···················· | ···················· | ···················· | ···················· | ···················· | ···················· |
| 2389 | MW341477.1 | \|Severe acute respiratory syndrome coro | ···················· | ···················· | ···················· | ···················· | ···················· | ···················· |
| 2390 | MW341478.1 | \|Severe acute respiratory syndrome coro | ···················· | ···················· | ···················· | ···················· | ···················· | ···················· |
| 2391 | MW341781.1 | \|Severe acute respiratory syndrome coro | ···················· | ···················· | ···················· | ···················· | ···················· | ···················· |
| 2392 | MW341793.1 | \|Severe acute respiratory syndrome coro | ···················· | ···················· | ···················· | ···················· | ···················· | ···················· |
| 2393 | MW341795.1 | \|Severe acute respiratory syndrome coro | ···················· | ···················· | ···················· | ···················· | ···················· | ···················· |
| 2394 | MW341796.1 | \|Severe acute respiratory syndrome coro | ···················· | ···················· | ···················· | ···················· | ···················· | ···················· |
| 2395 | MW341798.1 | \|Severe acute respiratory syndrome coro | ···················· | ···················· | ···················· | ···················· | ···················· | ···················· |
| 2396 | MW341799.1 | \|Severe acute respiratory syndrome coro | ···················· | ···················· | ···················· | ···················· | ···················· | ···················· |
| 2397 | MW341801.1 | \|Severe acute respiratory syndrome coro | ···················· | ···················· | ···················· | ···················· | ···················· | ···················· |
| 2398 | MW341802.1 | \|Severe acute respiratory syndrome coro | ···················· | ···················· | ···················· | ···················· | ···················· | ···················· |
| 2399 | MW341803.1 | \|Severe acute respiratory syndrome coro | ···················· | ···················· | ···················· | ···················· | ···················· | ···················· |
| 2400 | MW341804.1 | \|Severe acute respiratory syndrome coro | ···················· | ···················· | ···················· | ···················· | ···················· | ···················· |
| 2401 | MW341805.1 | \|Severe acute respiratory syndrome coro | ···················· | ···················· | ···················· | ···················· | ···················· | ···················· |
| 2402 | MW341807.1 | \|Severe acute respiratory syndrome coro | ···················· | ···················· | ···················· | ···················· | ···················· | ···················· |
| 2403 | MW341808.1 | \|Severe acute respiratory syndrome coro | ···················· | ···················· | ···················· | ···················· | ···················· | ···················· |
| 2404 | MW341809.1 | \|Severe acute respiratory syndrome coro | ···················· | ···················· | ···················· | ···················· | ···················· | ···················· |
| 2405 | MW341810.1 | \|Severe acute respiratory syndrome coro | ···················· | ···················· | ···················· | ···················· | ···················· | ···················· |
| 2406 | MW341811.1 | \|Severe acute respiratory syndrome coro | ···················· | ···················· | ···················· | ···················· | ···················· | ···················· |
| 2407 | MW341812.1 | \|Severe acute respiratory syndrome coro | ···················· | ·A·················· | ···················· | ···················· | ···················· | ···················· |
| 2408 | MW341813.1 | \|Severe acute respiratory syndrome coro | ···················· | ···················· | ···················· | ···················· | ···················· | ···················· |
| 2409 | MW341814.1 | \|Severe acute respiratory syndrome coro | ···················· | ···················· | ···················· | ···················· | ···················· | ···················· |
| 2410 | MW341815.1 | \|Severe acute respiratory syndrome coro | ···················· | ···················· | ···················· | ···················· | ···················· | ···················· |
| 2411 | MW341818.1 | \|Severe acute respiratory syndrome coro | ···················· | ···················· | ···················· | ···················· | ···················· | ···················· |
| 2412 | MW341820.1 | \|Severe acute respiratory syndrome coro | ···················· | ···················· | ···················· | ···················· | ···················· | ···················· |
| 2413 | MW341822.1 | \|Severe acute respiratory syndrome coro | ···················· | ···················· | ···················· | ···················· | ···················· | ···················· |
| 2414 | MW341823.1 | \|Severe acute respiratory syndrome coro | ···················· | ···················· | ···················· | ···················· | ···················· | ···················· |

2415 MW341825.1 |Severe acute respiratory syndrome coro ···················· ···················· ····················· ···················· ···················· ····················
2416 MW341826.1 |Severe acute respiratory syndrome coro ···················· ···················· ····················· ···················· ···················· ····················
2417 MW341827.1 |Severe acute respiratory syndrome coro ···················· ·A·················· ····················· ···················· ···················· ····················
2418 MW341830.1 |Severe acute respiratory syndrome coro ···················· ···················· ····················· ···················· ···················· ····················
2419 MW341831.1 |Severe acute respiratory syndrome coro ···················· ···················· ····················· ···················· ···················· ····················
2420 MW341832.1 |Severe acute respiratory syndrome coro ···················· ··C················· ····················· ···················· ···················· ····················
2421 MW341834.1 |Severe acute respiratory syndrome coro ···················· ···················· ····················· ···················· ···················· ····················
2422 MW341835.1 |Severe acute respiratory syndrome coro ···················· ···················· ····················· ···················· ···················· ····················
2423 MW341836.1 |Severe acute respiratory syndrome coro ···················· ··N················· ····················· ···················· ···················· ····················
2424 MW341852.1 |Severe acute respiratory syndrome coro ···················· ···················· ····················· ···················· ···················· ····················
2425 MW341860.1 |Severe acute respiratory syndrome coro ···················· ···················· ····················· ···················· ···················· ····················
2426 MW341861.1 |Severe acute respiratory syndrome coro ···················· ···················· ····················· ···················· ···················· ····················
2427 MW341862.1 |Severe acute respiratory syndrome coro ···················· ···················· ····················· ···················· ···················· ····················
2428 MW341863.1 |Severe acute respiratory syndrome coro ···················· ···················· ····················· ···················· ···················· ····················
2429 MW341864.1 |Severe acute respiratory syndrome coro ···················· ···················· ····················· ···················· ···················· ····················
2430 MW341865.1 |Severe acute respiratory syndrome coro ···················· ···················· ····················· ···················· ···················· ····················
2431 MW341866.1 |Severe acute respiratory syndrome coro ···················· ···················· ····················· ···················· ···················· ····················
2432 MW341867.1 |Severe acute respiratory syndrome coro ···················· ···················· ····················· ···················· ···················· ····················
2433 MW341868.1 |Severe acute respiratory syndrome coro ···················· ···················· ····················· ···················· ···················· ····················
2434 MW341869.1 |Severe acute respiratory syndrome coro ···················· ·A·················· ····················· ···················· ···················· ····················
2435 MW341870.1 |Severe acute respiratory syndrome coro ···················· ···················· ····················· ···················· ···················· ····················
2436 MW341871.1 |Severe acute respiratory syndrome coro ···················· ···················· ····················· ···················· ···················· ····················
2437 MW341872.1 |Severe acute respiratory syndrome coro ···················· ···················· ····················· ···················· ···················· ····················
2438 MW341873.1 |Severe acute respiratory syndrome coro ···················· ···················· ····················· ···················· ···················· ····················
2439 MW341874.1 |Severe acute respiratory syndrome coro ···················· ···················· ····················· ···················· ···················· ····················
2440 MW341875.1 |Severe acute respiratory syndrome coro ···················· ···················· ····················· ···················· ···················· ····················
2441 MW341876.1 |Severe acute respiratory syndrome coro ···················· ·A·················· ····················· ···················· ···················· ····················
2442 MW341877.1 |Severe acute respiratory syndrome coro ···················· ···················· ····················· ···················· ···················· ····················
2443 MW341878.1 |Severe acute respiratory syndrome coro ···················· ···················· ····················· ···················· ···················· ····················
2444 MW341879.1 |Severe acute respiratory syndrome coro ···················· ···················· ····················· ···················· ···················· ····················
2445 MW341880.1 |Severe acute respiratory syndrome coro ···················· ···················· ····················· ···················· ···················· ····················
2446 MW341881.1 |Severe acute respiratory syndrome coro ···················· ···················· ····················· ···················· ···················· ····················
2447 MW341882.1 |Severe acute respiratory syndrome coro ···················· ···················· ····················· ···················· ···················· ····················
2448 MW341900.1 |Severe acute respiratory syndrome coro ···················· ···················· ····················· ···················· ···················· ····················
2449 MW341915.1 |Severe acute respiratory syndrome coro ···················· ···················· ····················· ···················· ···················· ····················
2450 MW341916.1 |Severe acute respiratory syndrome coro ···················· ···················· ····················· ···················· ···················· ····················
2451 MW341918.1 |Severe acute respiratory syndrome coro ···················· ···················· ····················· ···················· ···················· ····················
2452 MW341919.1 |Severe acute respiratory syndrome coro ···················· ···················· ····················· ···················· ···················· ····················
2453 MW341920.1 |Severe acute respiratory syndrome coro ···················· ···················· ····················· ···················· ···················· ····················
2454 MW341921.1 |Severe acute respiratory syndrome coro ···················· ···················· ····················· ···················· ···················· ····················
2455 MW341923.1 |Severe acute respiratory syndrome coro ···················· ···················· ····················· ···················· ···················· ····················
2456 MW341927.1 |Severe acute respiratory syndrome coro ···················· ···················· ····················· ···················· ···················· ····················
2457 MW341928.1 |Severe acute respiratory syndrome coro ···················· ···················· ····················· ···················· ···················· ····················
2458 MW341929.1 |Severe acute respiratory syndrome coro ···················· ···················· ····················· ···················· ···················· ····················
2459 MW341930.1 |Severe acute respiratory syndrome coro ···················· ···················· ····················· ···················· ···················· ····················
2460 MW341931.1 |Severe acute respiratory syndrome coro ···················· ···················· ····················· ···················· ···················· ····················
2461 MW341932.1 |Severe acute respiratory syndrome coro ···················· ···················· ····················· ···················· ···················· ····················
2462 MW341933.1 |Severe acute respiratory syndrome coro ···················· ···················· ····················· ···················· ···················· ····················
2463 MW341935.1 |Severe acute respiratory syndrome coro ···················· ···················· ····················· ···················· ···················· ····················
2464 MW341937.1 |Severe acute respiratory syndrome coro ···················· ···················· ····················· ···················· ···················· ····················
2465 MW341938.1 |Severe acute respiratory syndrome coro ···················· ···················· ····················· ···················· ···················· ····················
2466 MW341939.1 |Severe acute respiratory syndrome coro ···················· ···················· ····················· ···················· ···················· ····················
2467 MW341940.1 |Severe acute respiratory syndrome coro ···················· ···················· ····················· ···················· ···················· ····················
2468 MW341943.1 |Severe acute respiratory syndrome coro ···················· ···················· ····················· ···················· ···················· ····················
2469 MW341947.1 |Severe acute respiratory syndrome coro ···················· ···················· ····················· ···················· ···················· ····················
2470 MW341948.1 |Severe acute respiratory syndrome coro ···················· ···················· ····················· ···················· ···················· ····················
2471 MW341951.1 |Severe acute respiratory syndrome coro ···················· ···················· ····················· ···················· ···················· ····················
2472 MW341952.1 |Severe acute respiratory syndrome coro ···················· ···················· ····················· ···················· ···················· ····················
2473 MW341953.1 |Severe acute respiratory syndrome coro ···················· ···················· ····················· ···················· ···················· ····················
2474 MW341954.1 |Severe acute respiratory syndrome coro ···················· ···················· ····················· ···················· ···················· ····················
2475 MW341955.1 |Severe acute respiratory syndrome coro ···················· ·A·················· ····················· ···················· ···················· ····················
2476 MW341958.1 |Severe acute respiratory syndrome coro ···················· ···················· ····················· ···················· ···················· ····················
2477 MW341959.1 |Severe acute respiratory syndrome coro ···················· ···················· ····················· ···················· ···················· ····················
2478 MW341960.1 |Severe acute respiratory syndrome coro ···················· ···················· ····················· ···················· ···················· ····················
2479 MW341966.1 |Severe acute respiratory syndrome coro ···················· ···················· ····················· ···················· ···················· ····················
2480 MW341968.1 |Severe acute respiratory syndrome coro ···················· ···················· ····················· ···················· ···················· ····················
2481 MW341982.1 |Severe acute respiratory syndrome coro ···················· ···················· ····················· ···················· ···················· ····················
2482 MW341987.1 |Severe acute respiratory syndrome coro ···················· ···················· ····················· ···················· ···················· ····················
2483 MW341995.1 |Severe acute respiratory syndrome coro ···················· ···················· ····················· ···················· ···················· ····················
2484 MW342000.1 |Severe acute respiratory syndrome coro ···················· ···················· ····················· ···················· ···················· ····················
2485 MW342006.1 |Severe acute respiratory syndrome coro ···················· ···················· ····················· ···················· ···················· ····················
2486 MW342047.1 |Severe acute respiratory syndrome coro ···················· ···················· ····················· ···················· ···················· ····················
2487 MW342051.1 |Severe acute respiratory syndrome coro ···················· ···················· ····················· ···················· ···················· ····················
2488 MW342052.1 |Severe acute respiratory syndrome coro ···················· ···················· ····················· ···················· ···················· ····················
2489 MW342053.1 |Severe acute respiratory syndrome coro ···················· ···················· ····················· ···················· ···················· ····················
2490 MW342054.1 |Severe acute respiratory syndrome coro ···················· ···················· ····················· ···················· ···················· ····················
2491 MW342056.1 |Severe acute respiratory syndrome coro ···················· ···················· ····················· ···················· ···················· ····················
2492 MW342057.1 |Severe acute respiratory syndrome coro ···················· ···················· ····················· ···················· ···················· ····················

2493 MW342058.1 |Severe acute respiratory syndrome coro ···················· ···················· ···················· ···················· ···················· ····················
2494 MW342061.1 |Severe acute respiratory syndrome coro ···················· ···················· ···················· ···················· ···················· ····················
2495 MW342062.1 |Severe acute respiratory syndrome coro ···················· ···················· ···················· ···················· ···················· ····················
2496 MW342063.1 |Severe acute respiratory syndrome coro ···················· ···················· ···················· ···················· ···················· ····················
2497 MW342064.1 |Severe acute respiratory syndrome coro ···················· ···················· ···················· ···················· ···················· ····················
2498 MW342065.1 |Severe acute respiratory syndrome coro ···················· ···················· ···················· ···················· ···················· ····················
2499 MW342066.1 |Severe acute respiratory syndrome coro ···················· ···················· ···················· ···················· ···················· ····················
2500 MW342068.1 |Severe acute respiratory syndrome coro ···················· ···················· ···················· ···················· ···················· ····················
2501 MW342069.1 |Severe acute respiratory syndrome coro ···················· ···················· ···················· ···················· ···················· ····················
2502 MW342070.1 |Severe acute respiratory syndrome coro ···················· ·N·················· ···················· ···················· ···················· ····················
2503 MW342071.1 |Severe acute respiratory syndrome coro ···················· ···················· ···················· ···················· ···················· ····················
2504 MW342072.1 |Severe acute respiratory syndrome coro ···················· ···················· ···················· ···················· ···················· ····················
2505 MW342073.1 |Severe acute respiratory syndrome coro ···················· ···················· ···················· ···················· ···················· ····················
2506 MW342075.1 |Severe acute respiratory syndrome coro ···················· ···················· ···················· ···················· ···················· ····················
2507 MW342076.1 |Severe acute respiratory syndrome coro ···················· ···················· ···················· ···················· ···················· ····················
2508 MW342077.1 |Severe acute respiratory syndrome coro ···················· ···················· ···················· ···················· ···················· ····················
2509 MW342078.1 |Severe acute respiratory syndrome coro ···················· ···················· ···················· ···················· ···················· ····················
2510 MW342079.1 |Severe acute respiratory syndrome coro ···················· ···················· ···················· ···················· ···················· ····················
2511 MW342083.1 |Severe acute respiratory syndrome coro ···················· ···················· ···················· ···················· ···················· ····················
2512 MW342085.1 |Severe acute respiratory syndrome coro ···················· ···················· ···················· ···················· ···················· ····················
2513 MW342086.1 |Severe acute respiratory syndrome coro ···················· ···················· ···················· ···················· ···················· ····················
2514 MW342087.1 |Severe acute respiratory syndrome coro ···················· ···················· ···················· ···················· ···················· ····················
2515 MW342089.1 |Severe acute respiratory syndrome coro ·····N·············· ···················· ···················· ···················· ···················· ····················
2516 MW342090.1 |Severe acute respiratory syndrome coro ···················· ···················· ···················· ···················· ···················· ····················
2517 MW342091.1 |Severe acute respiratory syndrome coro ···················· ···················· ···················· ···················· ···················· ····················
2518 MW342092.1 |Severe acute respiratory syndrome coro ···················· ···················· ···················· ···················· ···················· ····················
2519 MW342093.1 |Severe acute respiratory syndrome coro ···················· ···················· ···················· ···················· ···················· ····················
2520 MW342095.1 |Severe acute respiratory syndrome coro ···················· ····A··············· ···················· ···················· ···················· ···A················
2521 MW342096.1 |Severe acute respiratory syndrome coro ···················· ···················· ···················· ···················· ···················· ····················
2522 MW342097.1 |Severe acute respiratory syndrome coro ···················· ···················· ···················· ···················· ···················· ····················
2523 MW342706.1 |Severe acute respiratory syndrome coro ···················· ···················· ···················· ···················· ···················· ····················
2524 MW342708.1 |Severe acute respiratory syndrome coro ···················· ···················· ···················· ···················· ···················· ····················
2525 MW343734.1 |Severe acute respiratory syndrome coro ···················· ···················· ···················· ···················· ···················· ····················
2526 MW343735.1 |Severe acute respiratory syndrome coro ···················· ···················· ···················· ···················· ···················· ····················
2527 MW343736.1 |Severe acute respiratory syndrome coro ···················· ···················· ···················· ···················· ···················· ····················
2528 MW343737.1 |Severe acute respiratory syndrome coro ···················· ···················· ···················· ···················· ···················· ····················
2529 MW343738.1 |Severe acute respiratory syndrome coro ···················· ···················· ···················· ···················· ···················· ····················
2530 MW343739.1 |Severe acute respiratory syndrome coro ···················· ···················· ···················· ···················· ···················· ····················
2531 MW343740.1 |Severe acute respiratory syndrome coro ···················· ···················· ···················· ···················· ···················· ····················
2532 MW343741.1 |Severe acute respiratory syndrome coro ···················· ···················· ···················· ···················· ···················· ····················
2533 MW343742.1 |Severe acute respiratory syndrome coro ···················· ···················· ···················· ···················· ···················· ····················
2534 MW343743.1 |Severe acute respiratory syndrome coro ···················· ···················· ···················· ···················· ···················· ····················
2535 MW343744.1 |Severe acute respiratory syndrome coro ···················· ···················· ···················· ···················· ···················· ····················
2536 MW343745.1 |Severe acute respiratory syndrome coro ···················· ···················· ···················· ···················· ···················· ····················
2537 MW343746.1 |Severe acute respiratory syndrome coro ···················· ···················· ···················· ···················· ···················· ····················
2538 MW343747.1 |Severe acute respiratory syndrome coro ···················· ···················· ···················· ···················· ···················· ····················
2539 MW343748.1 |Severe acute respiratory syndrome coro ···················· ···················· ···················· ···················· ···················· ····················
2540 MW343749.1 |Severe acute respiratory syndrome coro ···················· ···················· ···················· ···················· ···················· ····················
2541 MW343750.1 |Severe acute respiratory syndrome coro ···················· ···················· ···················· ···················· ···················· ····················
2542 MW343751.1 |Severe acute respiratory syndrome coro ···················· ···················· ···················· ···················· ···················· ····················
2543 MW343752.1 |Severe acute respiratory syndrome coro ···················· ···················· ···················· ···················· ···················· ····················
2544 MW343753.1 |Severe acute respiratory syndrome coro ···················· ···················· ···················· ···················· ···················· ····················
2545 MW343754.1 |Severe acute respiratory syndrome coro ···················· ···················· ···················· ···················· ···················· ····················
2546 MW343755.1 |Severe acute respiratory syndrome coro ···················· ···················· ···················· ···················· ···················· ····················
2547 MW343756.1 |Severe acute respiratory syndrome coro ···················· ···················· ···················· ···················· ···················· ····················
2548 MW343757.1 |Severe acute respiratory syndrome coro ···················· ···················· ···················· ···················· ···················· ····················
2549 MW343758.1 |Severe acute respiratory syndrome coro ···················· ···················· ···················· ···················· ···················· ····················
2550 MW343759.1 |Severe acute respiratory syndrome coro ···················· ···················· ···················· ···················· ···················· ····················
2551 MW343760.1 |Severe acute respiratory syndrome coro ···················· ···················· ···················· ···················· ···················· ····················
2552 MW343761.1 |Severe acute respiratory syndrome coro ···················· ···················· ···················· ···················· ···················· ····················
2553 MW343762.1 |Severe acute respiratory syndrome coro ···················· ···················· ···················· ···················· ···················· ····················
2554 MW343763.1 |Severe acute respiratory syndrome coro ···················· ···················· ···················· ···················· ···················· ····················
2555 MW343764.1 |Severe acute respiratory syndrome coro ···················· ···················· ···················· ···················· ···················· ····················
2556 MW343765.1 |Severe acute respiratory syndrome coro ···················· ···················· ···················· ···················· ···················· ····················
2557 MW343766.1 |Severe acute respiratory syndrome coro ···················· ···················· ···················· ···················· ···················· ····················
2558 MW343767.1 |Severe acute respiratory syndrome coro ···················· ···················· ···················· ···················· ···················· ····················
2559 MW343768.1 |Severe acute respiratory syndrome coro ···················· ···················· ···················· ···················· ···················· ····················
2560 MW343769.1 |Severe acute respiratory syndrome coro ···················· ···················· ···················· ···················· ···················· ····················
2561 MW343770.1 |Severe acute respiratory syndrome coro ···················· ···················· ···················· ···················· ···················· ····················
2562 MW343771.1 |Severe acute respiratory syndrome coro ···················· ···················· ···················· ···················· ···················· ····················
2563 MW343772.1 |Severe acute respiratory syndrome coro ···················· ···················· ···················· ···················· ···················· ····················
2564 MW343773.1 |Severe acute respiratory syndrome coro ···················· ···················· ···················· ···················· ···················· ····················
2565 MW343774.1 |Severe acute respiratory syndrome coro ···················· ···················· ···················· ···················· ···················· ····················
2566 MW343775.1 |Severe acute respiratory syndrome coro ···················· ···················· ···················· ···················· ···················· ····················
2567 MW343776.1 |Severe acute respiratory syndrome coro ···················· ···················· ···················· ···················· ···················· ····················
2568 MW343777.1 |Severe acute respiratory syndrome coro ···················· ···················· ···················· ···················· ···················· ····················
2569 MW343778.1 |Severe acute respiratory syndrome coro ···················· ···················· ···················· ···················· ···················· ····················
2570 MW343779.1 |Severe acute respiratory syndrome coro ···················· ···················· ···················· ···················· ···················· ····················

2571 MW343780.1 |Severe acute respiratory syndrome coro ···················· ···················· ····················· ···················· ···················· ····················
2572 MW343781.1 |Severe acute respiratory syndrome coro ···················· ···················· ····················· ···················· ···················· ····················
2573 MW343782.1 |Severe acute respiratory syndrome coro ···················· ···················· ····················· ···················· ···················· ····················
2574 MW343783.1 |Severe acute respiratory syndrome coro ···················· ···················· ····················· ···················· ···················· ····················
2575 MW343784.1 |Severe acute respiratory syndrome coro ···················· ···················· ····················· ···················· ···················· ····················
2576 MW343785.1 |Severe acute respiratory syndrome coro ···················· ···················· ····················· ···················· ···················· ····················
2577 MW343786.1 |Severe acute respiratory syndrome coro ···················· ···················· ····················· ···················· ···················· ····················
2578 MW345264.1 |Severe acute respiratory syndrome coro ···················· ·A·················· ····················· ···················· ···················· ····················
2579 MW345265.1 |Severe acute respiratory syndrome coro ···················· ···················· ····················· ···················· ···················· ····················
2580 MW345266.1 |Severe acute respiratory syndrome coro ···················· ···················· ····················· ···················· ···················· ····················
2581 MW345267.1 |Severe acute respiratory syndrome coro ···················· ···················· ····················· ···················· ···················· ····················
2582 MW345268.1 |Severe acute respiratory syndrome coro ···················· ·A·················· ····················· ···················· ···················· ····················
2583 MW345269.1 |Severe acute respiratory syndrome coro ···················· ···················· ····················· ···················· ···················· ····················
2584 MW345270.1 |Severe acute respiratory syndrome coro ···················· ···················· ····················· ···················· ···················· ····················
2585 MW345271.1 |Severe acute respiratory syndrome coro ···················· ···················· ····················· ···················· ···················· ····················
2586 MW345272.1 |Severe acute respiratory syndrome coro ···················· ···················· ····················· ···················· ···················· ····················
2587 MW345273.1 |Severe acute respiratory syndrome coro ···················· ···················· ····················· ···················· ···················· ····················
2588 MW345274.1 |Severe acute respiratory syndrome coro ···················· ···················· ····················· ···················· ···················· ····················
2589 MW345275.1 |Severe acute respiratory syndrome coro ···················· ···················· ····················· ···················· ···················· ····················
2590 MW345276.1 |Severe acute respiratory syndrome coro ···················· ···················· ····················· ···················· ···················· ····················
2591 MW345277.1 |Severe acute respiratory syndrome coro ···················· ············C······· ····················· ···················· ···················· ····················
2592 MW345278.1 |Severe acute respiratory syndrome coro ···················· ···················· ····················· ···················· ···················· ····················
2593 MW345279.1 |Severe acute respiratory syndrome coro ···················· ···················· ····················· ···················· ···················· ····················
2594 MW345280.1 |Severe acute respiratory syndrome coro ···················· ···················· ····················· ···················· ···················· ····················
2595 MW345281.1 |Severe acute respiratory syndrome coro ···················· ···················· ····················· ···················· ···················· ····················
2596 MW345283.1 |Severe acute respiratory syndrome coro ···················· ···················· ····················· ···················· ···················· ····················
2597 MW345284.1 |Severe acute respiratory syndrome coro ···················· ···················· ····················· ···················· ···················· ····················
2598 MW345285.1 |Severe acute respiratory syndrome coro ···················· ···················· ····················· ···················· ···················· ····················
2599 MW345824.1 |Severe acute respiratory syndrome coro ···················· ···················· ··T·················· ···················· ···················· ····················
2600 MW345825.1 |Severe acute respiratory syndrome coro ···················· ···················· ····················· ···················· ···················· ····················
2601 MW332531.1 |Severe acute respiratory syndrome coro ···················· ···················· ····················· ···················· ···················· ····················
2602 MW332532.1 |Severe acute respiratory syndrome coro ···················· ···················· ····················· ···················· ···················· ····················
2603 MW332533.1 |Severe acute respiratory syndrome coro ···················· ···················· ····················· ···················· ···················· ····················
2604 MW332534.1 |Severe acute respiratory syndrome coro ···················· ···················· ····················· ···················· ···················· ····················
2605 MW332535.1 |Severe acute respiratory syndrome coro ···················· ···················· ····················· ···················· ···················· ····················
2606 MW332536.1 |Severe acute respiratory syndrome coro ···················· ···················· ····················· ···················· ···················· ····················
2607 MW332537.1 |Severe acute respiratory syndrome coro ···················· ···················· ····················· ···················· ···················· ····················
2608 MW332538.1 |Severe acute respiratory syndrome coro ···················· ···················· ····················· ···················· ···················· ····················
2609 MW332539.1 |Severe acute respiratory syndrome coro ···················· ···················· ····················· ···················· ···················· ····················
2610 MW332540.1 |Severe acute respiratory syndrome coro ···················· ···················· ····················· ···················· ···················· ····················
2611 MW332551.1 |Severe acute respiratory syndrome coro ···················· ···················· ····················· ···················· ···················· ····················
2612 MW332552.1 |Severe acute respiratory syndrome coro ···················· ···················· ····················· ···················· ···················· ····················
2613 MW332553.1 |Severe acute respiratory syndrome coro ···················· ···················· ····················· ···················· ···················· ····················
2614 MW332554.1 |Severe acute respiratory syndrome coro ···················· ···················· ····················· ···················· ···················· ····················
2615 MW332555.1 |Severe acute respiratory syndrome coro ···················· ···················· ····················· ···················· ···················· ····················
2616 MW332558.1 |Severe acute respiratory syndrome coro ···················· ···················· ····················· ···················· ···················· ····················
2617 MW332559.1 |Severe acute respiratory syndrome coro ········T··········· ···················· ····················· ···················· ···················· ····················
2618 MW332560.1 |Severe acute respiratory syndrome coro ···················· ···················· ····················· ···················· ···················· ····················
2619 MW332561.1 |Severe acute respiratory syndrome coro ···················· ···················· ····················· ···················· ···················· ····················
2620 MW332562.1 |Severe acute respiratory syndrome coro ···················· ···················· ····················· ···················· ···················· ····················
2621 MW332563.1 |Severe acute respiratory syndrome coro ···················· ···················· ····················· ···················· ···················· ····················
2622 MW332564.1 |Severe acute respiratory syndrome coro ···········T········ ···················· ····················· ···················· ···················· ····················
2623 MW332652.1 |Severe acute respiratory syndrome coro ···················· ···················· ····················· ···················· ···················· ····················
2624 MW332653.1 |Severe acute respiratory syndrome coro ···················· ···················· ····················· ···················· ···················· ····················
2625 MW332655.1 |Severe acute respiratory syndrome coro ···················· ···················· ····················· ···················· ···················· ····················
2626 MW332656.1 |Severe acute respiratory syndrome coro ···················· ···················· ····················· ···················· ···················· ····················
2627 MW332657.1 |Severe acute respiratory syndrome coro ···················· ···················· ····················· ···················· ···················· ····················
2628 MW332658.1 |Severe acute respiratory syndrome coro ···················· ···················· ····················· ···················· ···················· ····················
2629 MW332659.1 |Severe acute respiratory syndrome coro ···················· ···················· ····················· ···················· ···················· ····················
2630 MW332662.1 |Severe acute respiratory syndrome coro ···················· ···················· ····················· ···················· ···················· ····················
2631 MW332664.1 |Severe acute respiratory syndrome coro ···················· ···················· ····················· ···················· ···················· ····················
2632 MW332665.1 |Severe acute respiratory syndrome coro ···················· ···················· ····················· ···················· ···················· ····················
2633 MW332666.1 |Severe acute respiratory syndrome coro ···················· ···················· ····················· ···················· ···················· ····················
2634 MW332667.1 |Severe acute respiratory syndrome coro ···················· ···················· ····················· ···················· ···················· ····················
2635 MW332668.1 |Severe acute respiratory syndrome coro ···················· ···················· ····················· ···················· ···················· ····················
2636 MW332669.1 |Severe acute respiratory syndrome coro ···················· ···················· ····················· ···················· ···················· ····················
2637 MW332670.1 |Severe acute respiratory syndrome coro ···················· ···················· ····················· ···················· ···················· ····················
2638 MW332671.1 |Severe acute respiratory syndrome coro ···················· ···················· ····················· ···················· ···················· ····················
2639 MW332672.1 |Severe acute respiratory syndrome coro ···················· ···················· ····················· ···················· ···················· ····················
2640 MW332673.1 |Severe acute respiratory syndrome coro ···················· ···················· ····················· ···················· ···················· ····················
2641 MW332674.1 |Severe acute respiratory syndrome coro ···················· ···················· ····················· ···················· ···················· ····················
2642 MW332675.1 |Severe acute respiratory syndrome coro ···················· ···················· ····················· ···················· ···················· ····················
2643 MW332679.1 |Severe acute respiratory syndrome coro ···················· ···················· ····················· ···················· ···················· ····················
2644 MW332686.1 |Severe acute respiratory syndrome coro ···················· ···················· ····················· ···················· ···················· ····················
2645 MW332688.1 |Severe acute respiratory syndrome coro ···················· ···················· ····················· ···················· ···················· ····················
2646 MW332691.1 |Severe acute respiratory syndrome coro ···················· ···················· ····················· ···················· ···················· ····················
2647 MW332694.1 |Severe acute respiratory syndrome coro ···················· ···················· ····················· ···················· ···················· ····················
2648 MW332712.1 |Severe acute respiratory syndrome coro ···················· ···················· ····················· ···················· ···················· ····················

| | | | | | | | | |
|---|---|---|---|---|---|---|---|---|
| 2649 | MW332713.1 | \|Severe acute respiratory syndrome coro | ···················· | ···················· | ···················· | ···················· | ···················· | ···················· |
| 2650 | MW332714.1 | \|Severe acute respiratory syndrome coro | ···················· | ···················· | ···················· | ···················· | ···················· | ···················· |
| 2651 | MW332715.1 | \|Severe acute respiratory syndrome coro | ···················· | ···················· | ···················· | ···················· | ···················· | ···················· |
| 2652 | MW332720.1 | \|Severe acute respiratory syndrome coro | ···················· | ···················· | ···················· | ···················· | ···················· | ···················· |
| 2653 | MW332721.1 | \|Severe acute respiratory syndrome coro | ···················· | ···················· | ···················· | ···················· | ···················· | ···················· |
| 2654 | MW332722.1 | \|Severe acute respiratory syndrome coro | ···················· | ···················· | ···················· | ···················· | ···················· | ···················· |
| 2655 | MW332723.1 | \|Severe acute respiratory syndrome coro | ···················· | ···················· | ···················· | ···················· | ···················· | ···················· |
| 2656 | MW332725.1 | \|Severe acute respiratory syndrome coro | ···················· | ···················· | ···················· | ···················· | ···················· | ···················· |
| 2657 | MW332726.1 | \|Severe acute respiratory syndrome coro | ···················· | ···················· | ···················· | ···················· | ···················· | ···················· |
| 2658 | MW332728.1 | \|Severe acute respiratory syndrome coro | ···················· | ···················· | ···················· | ···················· | ···················· | ···················· |
| 2659 | MW332729.1 | \|Severe acute respiratory syndrome coro | ···················· | ···················· | ···················· | ···················· | ···················· | ···················· |
| 2660 | MW332731.1 | \|Severe acute respiratory syndrome coro | ···················· | ···················· | ···················· | ···················· | ···················· | ···················· |
| 2661 | MW332732.1 | \|Severe acute respiratory syndrome coro | ···················· | ···················· | ···················· | ···················· | ···················· | ···················· |
| 2662 | MW332733.1 | \|Severe acute respiratory syndrome coro | ···················· | ···················· | ···················· | ···················· | ···················· | ···················· |
| 2663 | MW332735.1 | \|Severe acute respiratory syndrome coro | ···················· | ···················· | ···················· | ···················· | ···················· | ···················· |
| 2664 | MW332736.1 | \|Severe acute respiratory syndrome coro | ···················· | ···················· | ···················· | ···················· | ···················· | ···················· |
| 2665 | MW332738.1 | \|Severe acute respiratory syndrome coro | ···················· | ···················· | ···················· | ···················· | ···················· | ···················· |
| 2666 | MW332739.1 | \|Severe acute respiratory syndrome coro | ···················· | ···················· | ···················· | ···················· | ···················· | ···················· |
| 2667 | MW332740.1 | \|Severe acute respiratory syndrome coro | ···················· | ···················· | ···················· | ···················· | ···················· | ···················· |
| 2668 | MW332741.1 | \|Severe acute respiratory syndrome coro | ···················· | ···················· | ···················· | ···················· | ···················· | ···················· |
| 2669 | MW332742.1 | \|Severe acute respiratory syndrome coro | ···················· | ···················· | ···················· | ···················· | ···················· | ···················· |
| 2670 | MW332744.1 | \|Severe acute respiratory syndrome coro | ···················· | ···················· | ···················· | ···················· | ···················· | ···················· |
| 2671 | MW332748.1 | \|Severe acute respiratory syndrome coro | ···················· | ·A·················· | ···················· | ···················· | ···················· | ···················· |
| 2672 | MW332750.1 | \|Severe acute respiratory syndrome coro | ···················· | ···················· | ···················· | ···················· | ···················· | ···················· |
| 2673 | MW332751.1 | \|Severe acute respiratory syndrome coro | ···················· | ···················· | ···················· | ···················· | ···················· | ···················· |
| 2674 | MW332752.1 | \|Severe acute respiratory syndrome coro | ···················· | ···················· | ···················· | ···················· | ···················· | ···················· |
| 2675 | MW332753.1 | \|Severe acute respiratory syndrome coro | ···················· | ···················· | ···················· | ···················· | ···················· | ···················· |
| 2676 | MW332754.1 | \|Severe acute respiratory syndrome coro | ···················· | ···················· | ···················· | ···················· | ···················· | ···················· |
| 2677 | MW332755.1 | \|Severe acute respiratory syndrome coro | ···················· | ···················· | ···················· | ···················· | ···················· | ···················· |
| 2678 | MW332757.1 | \|Severe acute respiratory syndrome coro | ···················· | ···················· | ···················· | ···················· | ···················· | ···················· |
| 2679 | MW332758.1 | \|Severe acute respiratory syndrome coro | ···················· | ···················· | ···················· | ···················· | ···················· | ···················· |
| 2680 | MW332759.1 | \|Severe acute respiratory syndrome coro | ···················· | ···················· | ···················· | ···················· | ···················· | ···················· |
| 2681 | MW332761.1 | \|Severe acute respiratory syndrome coro | ···················· | ···················· | ···················· | ···················· | ···················· | ···················· |
| 2682 | MW332764.1 | \|Severe acute respiratory syndrome coro | ···················· | ···················· | ···················· | ···················· | ···················· | ···················· |
| 2683 | MW332766.1 | \|Severe acute respiratory syndrome coro | ···················· | ···················· | ···················· | ···················· | ···················· | ···················· |
| 2684 | MW332767.1 | \|Severe acute respiratory syndrome coro | ···················· | ···················· | ···················· | ···················· | ···················· | ···················· |
| 2685 | MW332768.1 | \|Severe acute respiratory syndrome coro | ···················· | ···················· | ···················· | ···················· | ···················· | ···················· |
| 2686 | MW332769.1 | \|Severe acute respiratory syndrome coro | ···················· | ···················· | ···················· | ···················· | ···················· | ···················· |
| 2687 | MW332770.1 | \|Severe acute respiratory syndrome coro | ···················· | ···················· | ···················· | ···················· | ···················· | ···················· |
| 2688 | MW332771.1 | \|Severe acute respiratory syndrome coro | ···················· | ···················· | ···················· | ···················· | ···················· | ···················· |
| 2689 | MW332772.1 | \|Severe acute respiratory syndrome coro | ···················· | ···················· | ···················· | ···················· | ···················· | ···················· |
| 2690 | MW332773.1 | \|Severe acute respiratory syndrome coro | ···················· | ···················· | ···················· | ···················· | ···················· | ···················· |
| 2691 | MW332775.1 | \|Severe acute respiratory syndrome coro | ···················· | ···················· | ···················· | ···················· | ···················· | ···················· |
| 2692 | MW332776.1 | \|Severe acute respiratory syndrome coro | ···················· | ···················· | ···················· | ···················· | ···················· | ···················· |
| 2693 | MW332777.1 | \|Severe acute respiratory syndrome coro | ···················· | ···················· | ···················· | ···················· | ···················· | ···················· |
| 2694 | MW332778.1 | \|Severe acute respiratory syndrome coro | ···················· | ···················· | ···················· | ···················· | ···················· | ···················· |
| 2695 | MW332779.1 | \|Severe acute respiratory syndrome coro | ···················· | ···················· | ···················· | ···················· | ···················· | ···················· |
| 2696 | MW332782.1 | \|Severe acute respiratory syndrome coro | ···················· | ···················· | ···················· | ···················· | ···················· | ···················· |
| 2697 | MW332784.1 | \|Severe acute respiratory syndrome coro | ···················· | ···················· | ···················· | ···················· | ···················· | ···················· |
| 2698 | MW332785.1 | \|Severe acute respiratory syndrome coro | ···················· | ···················· | ···················· | ···················· | ···················· | ···················· |
| 2699 | MW332786.1 | \|Severe acute respiratory syndrome coro | ···················· | ···················· | ···················· | ···················· | ···················· | ···················· |
| 2700 | MW332788.1 | \|Severe acute respiratory syndrome coro | ···················· | ···················· | ···················· | ···················· | ···················· | ···················· |
| 2701 | MW332790.1 | \|Severe acute respiratory syndrome coro | ···················· | ···················· | ···················· | ···················· | ···················· | ···················· |
| 2702 | MW332791.1 | \|Severe acute respiratory syndrome coro | ···················· | ···················· | ···················· | ···················· | ···················· | ···················· |
| 2703 | MW332793.1 | \|Severe acute respiratory syndrome coro | ···················· | ···················· | ···················· | ···················· | ···················· | ···················· |
| 2704 | MW332794.1 | \|Severe acute respiratory syndrome coro | ···················· | ···················· | ···················· | ···················· | ···················· | ···················· |
| 2705 | MW332796.1 | \|Severe acute respiratory syndrome coro | ···················· | ···················· | ···················· | ···················· | ···················· | ···················· |
| 2706 | MW332797.1 | \|Severe acute respiratory syndrome coro | ···················· | ···················· | ···················· | ···················· | ···················· | ···················· |
| 2707 | MW332803.1 | \|Severe acute respiratory syndrome coro | ···················· | ···················· | ···················· | ···················· | ···················· | ···················· |
| 2708 | MW332806.1 | \|Severe acute respiratory syndrome coro | ···················· | ···················· | ···················· | ···················· | ···················· | ···················· |
| 2709 | MW332808.1 | \|Severe acute respiratory syndrome coro | ···················· | ·············A······ | ···················· | ···················· | ···················· | ···················· |
| 2710 | MW332811.1 | \|Severe acute respiratory syndrome coro | ···················· | ···················· | ···················· | ···················· | ···················· | ···················· |
| 2711 | MW332812.1 | \|Severe acute respiratory syndrome coro | ···················· | ···················· | ···················· | ···················· | ···················· | ···················· |
| 2712 | MW332814.1 | \|Severe acute respiratory syndrome coro | ···················· | ···················· | ···················· | ···················· | ···················· | ···················· |
| 2713 | MW332817.1 | \|Severe acute respiratory syndrome coro | ···················· | ···················· | ···················· | ···················· | ···················· | ···················· |
| 2714 | MW332820.1 | \|Severe acute respiratory syndrome coro | ···················· | ···················· | ···················· | ···················· | ···················· | ···················· |
| 2715 | MW332822.1 | \|Severe acute respiratory syndrome coro | ···················· | ···················· | ···················· | ···················· | ···················· | ···················· |
| 2716 | MW332823.1 | \|Severe acute respiratory syndrome coro | ···················· | ···················· | ···················· | ···················· | ···················· | ···················· |
| 2717 | MW332825.1 | \|Severe acute respiratory syndrome coro | ···················· | ···················· | ···················· | ···················· | ···················· | ···················· |
| 2718 | MW332827.1 | \|Severe acute respiratory syndrome coro | ···················· | ···················· | ···················· | ···················· | ···················· | ···················· |
| 2719 | MW332828.1 | \|Severe acute respiratory syndrome coro | ···················· | ···················· | ···················· | ···················· | ···················· | ···················· |
| 2721 | MW332832.1 | \|Severe acute respiratory syndrome coro | ···················· | ···················· | ···················· | ···················· | ···················· | ···················· |
| 2722 | MW332833.1 | \|Severe acute respiratory syndrome coro | ···················· | ···················· | ···················· | ···················· | ···················· | ···················· |
| 2723 | MW332842.1 | \|Severe acute respiratory syndrome coro | ···················· | ···················· | ···················· | ···················· | ···················· | ···················· |
| 2724 | MW332857.1 | \|Severe acute respiratory syndrome coro | ···················· | ···················· | ···················· | ···················· | ···················· | ···················· |
| 2725 | MW332858.1 | \|Severe acute respiratory syndrome coro | ···················· | ·A·················· | ···················· | ···················· | ···················· | ···················· |
| 2726 | MW332859.1 | \|Severe acute respiratory syndrome coro | ···················· | ·············A······ | ···················· | ···················· | ···················· | ···················· |
| 2727 | MW332860.1 | \|Severe acute respiratory syndrome coro | ···················· | ···················· | ···················· | ···················· | ···················· | ···················· |

```
2728 MW332861.1 |Severe acute respiratory syndrome coro    ····················  ····················  ····················  ····················  ····················  ····················
2729 MW332862.1 |Severe acute respiratory syndrome coro    ····················  ·············A······  ····················  ····················  ····················  ····················
2730 MW332864.1 |Severe acute respiratory syndrome coro    ····················  ····················  ····················  ····················  ····················  ····················
2731 MW332865.1 |Severe acute respiratory syndrome coro    ····················  ····················  ····················  ····················  ····················  ····················
2732 MW332866.1 |Severe acute respiratory syndrome coro    ····················  ····················  ····················  ····················  ····················  ····················
2733 MW332868.1 |Severe acute respiratory syndrome coro    ····················  ····················  ····················  ····················  ····················  ····················
2734 MW332869.1 |Severe acute respiratory syndrome coro    ····················  ····················  ····················  ····················  ····················  ····················
2735 MW332870.1 |Severe acute respiratory syndrome coro    ····················  ····················  ····················  ····················  ····················  ····················
2736 MW332871.1 |Severe acute respiratory syndrome coro    ····················  ····················  ····················  ····················  ····················  ····················
2737 MW332872.1 |Severe acute respiratory syndrome coro    ····················  ····················  ····················  ····················  ····················  ····················
2738 MW332873.1 |Severe acute respiratory syndrome coro    ····················  ····················  ····················  ····················  ····················  ····················
2739 MW332874.1 |Severe acute respiratory syndrome coro    ····················  ····················  ····················  ····················  ····················  ····················
2740 MW332875.1 |Severe acute respiratory syndrome coro    ····················  ·············A······  ····················  ····················  ····················  ····················
2741 MW332876.1 |Severe acute respiratory syndrome coro    ····················  ····················  ····················  ····················  ····················  ····················
2742 MW332877.1 |Severe acute respiratory syndrome coro    ····················  ····················  ····················  ····················  ····················  ····················
2743 MW332878.1 |Severe acute respiratory syndrome coro    ····················  ····················  ····················  ····················  ····················  ····················
2744 MW332879.1 |Severe acute respiratory syndrome coro    ····················  ····················  ····················  ····················  ····················  ····················
2745 MW332880.1 |Severe acute respiratory syndrome coro    ····················  ····················  ····················  ····················  ····················  ····················
2746 MW332881.1 |Severe acute respiratory syndrome coro    ····················  ····················  ····················  ····················  ····················  ····················
2747 MW332882.1 |Severe acute respiratory syndrome coro    ····················  ····················  ····················  ····················  ····················  ····················
2748 MW332885.1 |Severe acute respiratory syndrome coro    ····················  ····················  ····················  ····················  ····················  ····················
2749 MW332886.1 |Severe acute respiratory syndrome coro    ····················  ·············A······  ····················  ····················  ····················  ····················
2750 MW332887.1 |Severe acute respiratory syndrome coro    ····················  ····················  ····················  ····················  ····················  ····················
2751 MW332890.1 |Severe acute respiratory syndrome coro    ····················  ····················  ····················  ····················  ····················  ····················
2752 MW332891.1 |Severe acute respiratory syndrome coro    ····················  ····················  ····················  ····················  ····················  ····················
2753 MW332892.1 |Severe acute respiratory syndrome coro    ····················  ····················  ····················  ····················  ····················  ····················
2754 MW332893.1 |Severe acute respiratory syndrome coro    ····················  ····················  ····················  ····················  ····················  ····················
2755 MW332895.1 |Severe acute respiratory syndrome coro    ····················  ····················  ····················  ····················  ····················  ····················
2756 MW332896.1 |Severe acute respiratory syndrome coro    ····················  ····················  ····················  ····················  ····················  ····················
2757 MW332897.1 |Severe acute respiratory syndrome coro    ····················  ····················  ····················  ····················  ····················  ····················
2758 MW332898.1 |Severe acute respiratory syndrome coro    ····················  ····················  ····················  ····················  ····················  ····················
2759 MW332899.1 |Severe acute respiratory syndrome coro    ····················  ····················  ····················  ····················  ····················  ····················
2760 MW332920.1 |Severe acute respiratory syndrome coro    ····················  ····················  ····················  ····················  ····················  ····················
2761 MW332921.1 |Severe acute respiratory syndrome coro    ····················  ····················  ····················  ····················  ····················  ····················
2762 MW332925.1 |Severe acute respiratory syndrome coro    ····················  ····················  ····················  ····················  ····················  ····················
2763 MW332934.1 |Severe acute respiratory syndrome coro    ····················  ····················  ····················  ····················  ····················  ····················
2764 MW332943.1 |Severe acute respiratory syndrome coro    ····················  ····················  ····················  ····················  ····················  ····················
2765 MW332945.1 |Severe acute respiratory syndrome coro    ····················  ····················  ····················  ····················  ····················  ····················
2766 MW332946.1 |Severe acute respiratory syndrome coro    ····················  ····················  ····················  ····················  ····················  ····················
2767 MW332948.1 |Severe acute respiratory syndrome coro    ····················  ····················  ····················  ····················  ····················  ····················
2768 MW332949.1 |Severe acute respiratory syndrome coro    ····················  ····················  ····················  ····················  ····················  ····················
2769 MW332951.1 |Severe acute respiratory syndrome coro    ····················  ····················  ····················  ····················  ····················  ····················
2770 MW332952.1 |Severe acute respiratory syndrome coro    ····················  ····················  ····················  ····················  ····················  ····················
2771 MW332953.1 |Severe acute respiratory syndrome coro    ····················  ····················  ····················  ····················  ····················  ····················
2772 MW332956.1 |Severe acute respiratory syndrome coro    ····················  ····················  ····················  ····················  ····················  ····················
2773 MW332957.1 |Severe acute respiratory syndrome coro    ····················  ····················  ····················  ····················  ····················  ····················
2774 MW332958.1 |Severe acute respiratory syndrome coro    ····················  ····················  ····················  ····················  ····················  ····················
2775 MW332959.1 |Severe acute respiratory syndrome coro    ····················  ····················  ····················  ····················  ····················  ····················
2776 MW332960.1 |Severe acute respiratory syndrome coro    ····················  ····················  ····················  ····················  ····················  ····················
2777 MW332961.1 |Severe acute respiratory syndrome coro    ····················  ····················  ····················  ····················  ····················  ····················
2778 MW332962.1 |Severe acute respiratory syndrome coro    ····················  ····················  ····················  ····················  ····················  ····················
2779 MW332963.1 |Severe acute respiratory syndrome coro    ····················  ····················  ····················  ····················  ····················  ····················
2780 MW332965.1 |Severe acute respiratory syndrome coro    ····················  ····················  ····················  ····················  ····················  ····················
2781 MW332970.1 |Severe acute respiratory syndrome coro    ····················  ·············A······  ····················  ····················  ····················  ····················
2782 MW332975.1 |Severe acute respiratory syndrome coro    ····················  ····················  ····················  ····················  ····················  ····················
2783 MW332979.1 |Severe acute respiratory syndrome coro    ····················  ····················  ····················  ····················  ····················  ····················
2784 MW332980.1 |Severe acute respiratory syndrome coro    ····················  ····················  ····················  ····················  ····················  ····················
2785 MW332989.1 |Severe acute respiratory syndrome coro    ····················  ····················  ····················  ····················  ····················  ····················
2786 MW332990.1 |Severe acute respiratory syndrome coro    ····················  ····················  ····················  ····················  ····················  ····················
2787 MW332992.1 |Severe acute respiratory syndrome coro    ····················  ····················  ····················  ····················  ····················  ····················
2788 MW332993.1 |Severe acute respiratory syndrome coro    ····················  ····················  ····················  ····················  ····················  ····················
2789 MW332995.1 |Severe acute respiratory syndrome coro    ····················  ····················  ····················  ····················  ····················  ····················
2790 MW332999.1 |Severe acute respiratory syndrome coro    ····················  ····················  ····················  ····················  ····················  ····················
2791 MW333000.1 |Severe acute respiratory syndrome coro    ····················  ····················  ····················  ····················  ····················  ····················
2792 MW333001.1 |Severe acute respiratory syndrome coro    ····················  ····················  ····················  ····················  ····················  ····················
2793 MW333002.1 |Severe acute respiratory syndrome coro    ····················  ····················  ····················  ····················  ····················  ····················
2794 MW333003.1 |Severe acute respiratory syndrome coro    ····················  ····················  ····················  ····················  ····················  ····················
2795 MW333004.1 |Severe acute respiratory syndrome coro    ····················  ····················  ····················  ····················  ····················  ····················
2796 MW333005.1 |Severe acute respiratory syndrome coro    ····················  ····················  ····················  ····················  ····················  ····················
2797 MW333007.1 |Severe acute respiratory syndrome coro    ····················  ····················  ····················  ····················  ····················  ····················
2798 MW333008.1 |Severe acute respiratory syndrome coro    ····················  ····················  ····················  ····················  ····················  ····················
2799 MW333009.1 |Severe acute respiratory syndrome coro    ····················  ····················  ····················  ····················  ····················  ····················
2800 MW333010.1 |Severe acute respiratory syndrome coro    ····················  ····················  ····················  ····················  ····················  ····················
2801 MW333012.1 |Severe acute respiratory syndrome coro    ····················  ····················  ····················  ····················  ····················  ····················
2802 MW333013.1 |Severe acute respiratory syndrome coro    ····················  ····················  ····················  ····················  ····················  ····················
2803 MW333014.1 |Severe acute respiratory syndrome coro    ····················  ····················  ····················  ····················  ····················  ····················
2804 MW333015.1 |Severe acute respiratory syndrome coro    ····················  ····················  ····················  ····················  ····················  ····················
2805 MW333016.1 |Severe acute respiratory syndrome coro    ····················  ····················  ····················  ····················  ····················  ····················
```

| | | | | | | | | |
|---|---|---|---|---|---|---|---|---|
| 2806 | MW333017.1 | \|Severe acute respiratory syndrome coro | ···················· | ···················· | ···················· | ···················· | ···················· | ···················· |
| 2807 | MW333018.1 | \|Severe acute respiratory syndrome coro | ···················· | ···················· | ···················· | ···················· | ···················· | ···················· |
| 2808 | MW333021.1 | \|Severe acute respiratory syndrome coro | ···················· | ···················· | ···················· | ···················· | ···················· | ···················· |
| 2809 | MW333022.1 | \|Severe acute respiratory syndrome coro | ···················· | ···················· | ···················· | ···················· | ···················· | ···················· |
| 2810 | MW333023.1 | \|Severe acute respiratory syndrome coro | ···················· | ···················· | ···················· | ···················· | ···················· | ···················· |
| 2811 | MW338760.1 | \|Severe acute respiratory syndrome coro | ···················· | ···················· | ···················· | ···················· | ···················· | ···················· |
| 2812 | MW338761.1 | \|Severe acute respiratory syndrome coro | ···················· | ···················· | ···················· | ···················· | ···················· | ···················· |
| 2813 | MW338762.1 | \|Severe acute respiratory syndrome coro | ···················· | ···················· | ···················· | ···················· | ···················· | ···················· |
| 2814 | MW338763.1 | \|Severe acute respiratory syndrome coro | ···················· | ···················· | ···················· | ···················· | ···················· | ···················· |
| 2815 | MW338764.1 | \|Severe acute respiratory syndrome coro | ···················· | ···················· | ···················· | ···················· | ···················· | ···················· |
| 2816 | MW338765.1 | \|Severe acute respiratory syndrome coro | ···················· | ···················· | ···················· | ···················· | ···················· | ···················· |
| 2817 | MW338766.1 | \|Severe acute respiratory syndrome coro | ···················· | ···················· | ···················· | ···················· | ···················· | ···················· |
| 2818 | MW338767.1 | \|Severe acute respiratory syndrome coro | ···················· | ···················· | ···················· | ···················· | ···················· | ···················· |
| 2819 | MW338768.1 | \|Severe acute respiratory syndrome coro | ···················· | ···················· | ···················· | ···················· | ···················· | ···················· |
| 2820 | MW338769.1 | \|Severe acute respiratory syndrome coro | ···················· | ···················· | ···················· | ···················· | ···················· | ···················· |
| 2821 | MW338770.1 | \|Severe acute respiratory syndrome coro | ···················· | ···················· | ···················· | ···················· | ···················· | ···················· |
| 2822 | MW338771.1 | \|Severe acute respiratory syndrome coro | ···················· | ···················· | ···················· | ···················· | ···················· | ···················· |
| 2823 | MW338772.1 | \|Severe acute respiratory syndrome coro | ···················· | ···················· | ···················· | ···················· | ···················· | ···················· |
| 2824 | MW338773.1 | \|Severe acute respiratory syndrome coro | ···················· | ···················· | ···················· | ···················· | ···················· | ···················· |
| 2825 | MW338774.1 | \|Severe acute respiratory syndrome coro | ···················· | ···················· | ···················· | ···················· | ···················· | ···················· |
| 2826 | MW338775.1 | \|Severe acute respiratory syndrome coro | ···················· | ···················· | ···················· | ···················· | ···················· | ···················· |
| 2827 | MW338776.1 | \|Severe acute respiratory syndrome coro | ···················· | ···················· | ···················· | ···················· | ···················· | ···················· |
| 2828 | MW338777.1 | \|Severe acute respiratory syndrome coro | ···················· | ···················· | ···················· | ···················· | ···················· | ···················· |
| 2829 | MW338778.1 | \|Severe acute respiratory syndrome coro | ···················· | ···················· | ···················· | ···················· | ···················· | ···················· |
| 2830 | MW338779.1 | \|Severe acute respiratory syndrome coro | ···················· | ···················· | ···················· | ···················· | ···················· | ···················· |
| 2831 | MW338780.1 | \|Severe acute respiratory syndrome coro | ···················· | ···················· | ···················· | ···················· | ···················· | ···················· |
| 2832 | MW338781.1 | \|Severe acute respiratory syndrome coro | ···················· | ···················· | ···················· | ···················· | ···················· | ···················· |
| 2833 | MW338782.1 | \|Severe acute respiratory syndrome coro | ···················· | ···················· | ···················· | ···················· | ···················· | ···················· |
| 2834 | MW338783.1 | \|Severe acute respiratory syndrome coro | ···················· | ·A·················· | ···················· | ···················· | ···················· | ···················· |
| 2835 | MW338784.1 | \|Severe acute respiratory syndrome coro | ···················· | ···················· | ···················· | ···················· | ···················· | ···················· |
| 2836 | MW338785.1 | \|Severe acute respiratory syndrome coro | ···················· | ···················· | ···················· | ···················· | ···················· | ···················· |
| 2837 | MW338786.1 | \|Severe acute respiratory syndrome coro | ···················· | ···················· | ···················· | ···················· | ···················· | ···················· |
| 2838 | MW338787.1 | \|Severe acute respiratory syndrome coro | ···················· | ···················· | ···················· | ···················· | ···················· | ···················· |
| 2839 | MW338788.1 | \|Severe acute respiratory syndrome coro | ···················· | ···················· | ···················· | ···················· | ···················· | ···················· |
| 2840 | MW338789.1 | \|Severe acute respiratory syndrome coro | ···················· | ···················· | ···················· | ···················· | ···················· | ···················· |
| 2841 | MW338790.1 | \|Severe acute respiratory syndrome coro | ···················· | ···················· | ···················· | ···················· | ···················· | ···················· |
| 2842 | MW338791.1 | \|Severe acute respiratory syndrome coro | ···················· | ···················· | ···················· | ···················· | ···················· | ···················· |
| 2843 | MW338792.1 | \|Severe acute respiratory syndrome coro | ···················· | ···················· | ···················· | ···················· | ···················· | ···················· |
| 2844 | MW338793.1 | \|Severe acute respiratory syndrome coro | ···················· | ···················· | ···················· | ···················· | ···················· | ···················· |
| 2845 | MW338794.1 | \|Severe acute respiratory syndrome coro | ···················· | ···················· | ···················· | ···················· | ···················· | ···················· |
| 2846 | MW338795.1 | \|Severe acute respiratory syndrome coro | ···················· | ···················· | ···················· | ···················· | ···················· | ···················· |
| 2847 | MW338796.1 | \|Severe acute respiratory syndrome coro | ···················· | ···················· | ···················· | ···················· | ···················· | ···················· |
| 2848 | MW338797.1 | \|Severe acute respiratory syndrome coro | ···················· | ···················· | ···················· | ···················· | ···················· | ···················· |
| 2849 | MW338798.1 | \|Severe acute respiratory syndrome coro | ···················· | ···················· | ···················· | ···················· | ···················· | ···················· |
| 2850 | MW338799.1 | \|Severe acute respiratory syndrome coro | ···················· | ···················· | ···················· | ···················· | ···················· | ···················· |
| 2851 | MW338800.1 | \|Severe acute respiratory syndrome coro | ···················· | ···················· | ···················· | ···················· | ···················· | ···················· |
| 2852 | MW338801.1 | \|Severe acute respiratory syndrome coro | ···················· | ···················· | ···················· | ···················· | ···················· | ···················· |
| 2853 | MW338802.1 | \|Severe acute respiratory syndrome coro | ···················· | ···················· | ···················· | ···················· | ···················· | ···················· |
| 2854 | MW338803.1 | \|Severe acute respiratory syndrome coro | ···················· | ···················· | ···················· | ···················· | ···················· | ···················· |
| 2855 | MW338804.1 | \|Severe acute respiratory syndrome coro | ···················· | ···················· | ···················· | ···················· | ···················· | ···················· |
| 2856 | MW338805.1 | \|Severe acute respiratory syndrome coro | ···················· | ···················· | ···················· | ···················· | ···················· | ···················· |
| 2857 | MW338806.1 | \|Severe acute respiratory syndrome coro | ···················· | ···················· | ···················· | ···················· | ···················· | ···················· |
| 2858 | MW338807.1 | \|Severe acute respiratory syndrome coro | ···················· | ·A·················· | ···················· | ···················· | ···················· | ···················· |
| 2859 | MW338808.1 | \|Severe acute respiratory syndrome coro | ···················· | ···················· | ···················· | ···················· | ···················· | ···················· |
| 2860 | MW338809.1 | \|Severe acute respiratory syndrome coro | ···················· | ·A·················· | ···················· | ···················· | ···················· | ···················· |
| 2861 | MW340733.1 | \|Severe acute respiratory syndrome coro | ···················· | ·A·················· | ···················· | ···················· | ···················· | ···················· |
| 2862 | MW340734.1 | \|Severe acute respiratory syndrome coro | ···················· | ···················· | ···················· | ···················· | ···················· | ···················· |
| 2863 | MW340735.1 | \|Severe acute respiratory syndrome coro | ···················· | ·A·················· | ···················· | ···················· | ···················· | ···················· |
| 2864 | MW340736.1 | \|Severe acute respiratory syndrome coro | ···················· | ···················· | ···················· | ···················· | ···················· | ···················· |
| 2865 | MW340737.1 | \|Severe acute respiratory syndrome coro | ···················· | ···················· | ···················· | ···················· | ···················· | ···················· |
| 2866 | MW340738.1 | \|Severe acute respiratory syndrome coro | ···················· | ···················· | ···················· | ···················· | ···················· | ···················· |
| 2867 | MW340739.1 | \|Severe acute respiratory syndrome coro | ···················· | ···················· | ···················· | ···················· | ···················· | ···················· |
| 2868 | MW340740.1 | \|Severe acute respiratory syndrome coro | ···················· | ···················· | ···················· | ···················· | ···················· | ···················· |
| 2869 | MW340741.1 | \|Severe acute respiratory syndrome coro | ···················· | ···················· | ···················· | ···················· | ···················· | ···················· |
| 2870 | MW340742.1 | \|Severe acute respiratory syndrome coro | ···················· | ···················· | ···················· | ···················· | ···················· | ···················· |
| 2871 | MW340743.1 | \|Severe acute respiratory syndrome coro | ···················· | ·A·················· | ···················· | ···················· | ···················· | ···················· |
| 2872 | MW340744.1 | \|Severe acute respiratory syndrome coro | ···················· | ···················· | ···················· | ···················· | ···················· | ···················· |
| 2873 | MW340745.1 | \|Severe acute respiratory syndrome coro | ···················· | ···················· | ···················· | ···················· | ···················· | ···················· |
| 2874 | MW340746.1 | \|Severe acute respiratory syndrome coro | ···················· | ···················· | ···················· | ···················· | ···················· | ···················· |
| 2875 | MW340747.1 | \|Severe acute respiratory syndrome coro | ···················· | ····A··············· | ···················· | ···················· | ···················· | ···················· |
| 2876 | MW340748.1 | \|Severe acute respiratory syndrome coro | ···················· | ···················· | ···················· | ···················· | ···················· | ···················· |
| 2877 | MW340749.1 | \|Severe acute respiratory syndrome coro | ···················· | ···················· | ···················· | ···················· | ···················· | ···················· |
| 2878 | MW340750.1 | \|Severe acute respiratory syndrome coro | ···················· | ···················· | ···················· | ···················· | ···················· | ···················· |
| 2879 | MW340751.1 | \|Severe acute respiratory syndrome coro | ···················· | ···················· | ···················· | ···················· | ···················· | ···················· |
| 2880 | MW340752.1 | \|Severe acute respiratory syndrome coro | ···················· | ···················· | ···················· | ···················· | ···················· | ···················· |
| 2881 | MW340753.1 | \|Severe acute respiratory syndrome coro | ···················· | ···················· | ···················· | ···················· | ···················· | ···················· |
| 2882 | MW340754.1 | \|Severe acute respiratory syndrome coro | ···················· | ···················· | ···················· | ···················· | ···················· | ···················· |
| 2883 | MW340755.1 | \|Severe acute respiratory syndrome coro | ···················· | ···················· | ···················· | ···················· | ···················· | ···················· |

```
2884 MW340756.1 |Severe acute respiratory syndrome coro   ····················  ····················  ····················  ····················  ····················  ····················
2885 MW340757.1 |Severe acute respiratory syndrome coro   ····················  ····················  ····················  ····················  ····················  ····················
2886 MW340758.1 |Severe acute respiratory syndrome coro   ····················  ····················  ····················  ····················  ····················  ····················
2887 MW340759.1 |Severe acute respiratory syndrome coro   ····················  ····················  ····················  ····················  ····················  ····················
2888 MW340760.1 |Severe acute respiratory syndrome coro   ····················  ····················  ····················  ····················  ····················  ····················
2889 MW340761.1 |Severe acute respiratory syndrome coro   ····················  ····················  ····················  ····················  ····················  ····················
2890 MW340763.1 |Severe acute respiratory syndrome coro   ····················  ····················  ····················  ····················  ····················  ····················
2891 MW340764.1 |Severe acute respiratory syndrome coro   ····················  ····················  ····················  ····················  ····················  ····················
2892 MW340765.1 |Severe acute respiratory syndrome coro   ····················  ····················  ····················  ····················  ····················  ····················
2893 MW340767.1 |Severe acute respiratory syndrome coro   ····················  ·A··················  ····················  ····················  ····················  ····················
2894 MW340768.1 |Severe acute respiratory syndrome coro   ····················  ·A··················  ····················  ····················  ····················  ····················
2895 MW340769.1 |Severe acute respiratory syndrome coro   ····················  ····C··············  ····················  ····················  ····················  ····················
2896 MW340770.1 |Severe acute respiratory syndrome coro   ····················  ····················  ····················  ····················  ····················  ····················
2897 MW340771.1 |Severe acute respiratory syndrome coro   ····················  ····················  ····················  ····················  ····················  ····················
2898 MW340772.1 |Severe acute respiratory syndrome coro   ····················  ····················  ····················  ····················  ····················  ····················
2899 MW340773.1 |Severe acute respiratory syndrome coro   ····················  ····················  ····················  ····················  ····················  ····················
2900 MW340775.1 |Severe acute respiratory syndrome coro   ····················  ····················  ····················  ····················  ····················  ····················
2901 MW340776.1 |Severe acute respiratory syndrome coro   ····················  ····················  ····················  ····················  ····················  ····················
2902 MW340777.1 |Severe acute respiratory syndrome coro   ····················  ····················  ····················  ····················  ····················  ····················
2903 MW340778.1 |Severe acute respiratory syndrome coro   ····················  ····················  ····················  ····················  ····················  ····················
2904 MW340779.1 |Severe acute respiratory syndrome coro   ····················  ····················  ····················  ····················  ····················  ····················
2905 MW340780.1 |Severe acute respiratory syndrome coro   ····················  ·A··················  ····················  ····················  ····················  ····················
2906 MW340781.1 |Severe acute respiratory syndrome coro   ····················  ····················  ····················  ····················  ····················  ····················
2907 MW340782.1 |Severe acute respiratory syndrome coro   ····················  ·A··················  ····················  ····················  ····················  ····················
2908 MW340783.1 |Severe acute respiratory syndrome coro   ····················  ·A··················  ····················  ····················  ····················  ····················
2909 MW340784.1 |Severe acute respiratory syndrome coro   ····················  ····················  ····················  ····················  ····················  ····················
2910 MW340785.1 |Severe acute respiratory syndrome coro   ····················  ····················  ····················  ····················  ····················  ····················
2911 MW340786.1 |Severe acute respiratory syndrome coro   ····················  ····················  ····················  ····················  ····················  ····················
2912 MW340787.1 |Severe acute respiratory syndrome coro   ····················  ····················  ····················  ····················  ····················  ····················
2913 MW340788.1 |Severe acute respiratory syndrome coro   ····················  ····················  ····················  ····················  ····················  ····················
2914 MW340789.1 |Severe acute respiratory syndrome coro   ····················  ····················  ····················  ····················  ····················  ····················
2915 MW340790.1 |Severe acute respiratory syndrome coro   ····················  ····················  ····················  ····················  ····················  ····················
2916 MW340791.1 |Severe acute respiratory syndrome coro   ····················  ····················  ····················  ····················  ····················  ····················
2917 MW340792.1 |Severe acute respiratory syndrome coro   ····················  ····················  ····················  ····················  ····················  ····················
2918 MW340793.1 |Severe acute respiratory syndrome coro   ····················  ····················  ····················  ····················  ····················  ····················
2919 MW340794.1 |Severe acute respiratory syndrome coro   ····················  ····················  ····················  ····················  ····················  ····················
2920 MW340795.1 |Severe acute respiratory syndrome coro   ····················  ····················  ····················  ····················  ····················  ····················
2921 MW340796.1 |Severe acute respiratory syndrome coro   ····················  ····················  ····················  ····················  ····················  ····················
2922 MW340797.1 |Severe acute respiratory syndrome coro   ····················  ····················  ····················  ····················  ····················  ····················
2923 MW340798.1 |Severe acute respiratory syndrome coro   ····················  ····················  ····················  ····················  ····················  ····················
2924 MW340799.1 |Severe acute respiratory syndrome coro   ····················  ····················  ····················  ····················  ····················  ····················
2925 MW341214.1 |Severe acute respiratory syndrome coro   ····················  ····················  ····················  ····················  ····················  ····················
2926 MW332213.1 |Severe acute respiratory syndrome coro   ····················  ·A··················  ····················  ····················  ····················  ··A·················
2927 MW332214.1 |Severe acute respiratory syndrome coro   ····················  ····················  ····················  ····················  ····················  ····················
2928 MW332215.1 |Severe acute respiratory syndrome coro   ····················  ····················  ····················  ····················  ····················  ····················
2929 MW332216.1 |Severe acute respiratory syndrome coro   ····················  ····················  ····················  ····················  ····················  ····················
2930 MW332217.1 |Severe acute respiratory syndrome coro   ····················  ····················  ····················  ····················  ····················  ····················
2931 MW332218.1 |Severe acute respiratory syndrome coro   ····················  ····················  ····················  ····················  ····················  ····················
2932 MW332219.1 |Severe acute respiratory syndrome coro   ····················  ····················  ····················  ····················  ····················  ····················
2933 MW332220.1 |Severe acute respiratory syndrome coro   ····················  ····················  ····················  ····················  ····················  ····················
2934 MW332221.1 |Severe acute respiratory syndrome coro   ····················  ····················  ····················  ····················  ····················  ····················
2935 MW332222.1 |Severe acute respiratory syndrome coro   ····················  ···A················  ····················  ····················  ····················  ··A·················
2936 MW332223.1 |Severe acute respiratory syndrome coro   ····················  ····················  ····················  ····················  ····················  ····················
2937 MW332224.1 |Severe acute respiratory syndrome coro   ····················  ····················  ····················  ····················  ····················  ····················
2938 MW332225.1 |Severe acute respiratory syndrome coro   ····················  ····················  ····················  ····················  ····················  ····················
2939 MW332226.1 |Severe acute respiratory syndrome coro   ····················  ····················  ····················  ····················  ····················  ····················
2940 MW332227.1 |Severe acute respiratory syndrome coro   ····················  ····················  ····················  ····················  ····················  ····················
2941 MW332228.1 |Severe acute respiratory syndrome coro   ····················  ····················  ····················  ····················  ····················  ····················
2942 MW332229.1 |Severe acute respiratory syndrome coro   ····················  ····················  ····················  ····················  ····················  ····················
2943 MW332230.1 |Severe acute respiratory syndrome coro   ····················  ····················  ····················  ····················  ····················  ····················
2944 MW332231.1 |Severe acute respiratory syndrome coro   ····················  ····················  ····················  ····················  ····················  ····················
2945 MW332232.1 |Severe acute respiratory syndrome coro   ····················  ····················  ····················  ····················  ····················  ····················
2946 MW332233.1 |Severe acute respiratory syndrome coro   ····················  ····················  ····················  ····················  ····················  ····················
2947 MW332235.1 |Severe acute respiratory syndrome coro   ····················  ····················  ····················  ····················  ····················  ····················
2948 MW332236.1 |Severe acute respiratory syndrome coro   ····················  ····················  ····················  ····················  ····················  ····················
2949 MW332237.1 |Severe acute respiratory syndrome coro   ····················  ····················  ····················  ····················  ····················  ····················
2950 MW332238.1 |Severe acute respiratory syndrome coro   ····················  ····················  ····················  ····················  ····················  ····················
2951 MW332239.1 |Severe acute respiratory syndrome coro   ····················  ····················  ····················  ····················  ····················  ····················
2952 MW332240.1 |Severe acute respiratory syndrome coro   ····················  ····················  ····················  ····················  ····················  ····················
2953 MW332241.1 |Severe acute respiratory syndrome coro   ····················  ····················  ····················  ····················  ····················  ····················
2954 MW327510.1 |Severe acute respiratory syndrome coro   ····················  ····················  ····················  ····················  ····················  ····················
2955 MW327511.1 |Severe acute respiratory syndrome coro   ····················  ····················  ····················  ····················  ····················  ····················
2956 MW327512.1 |Severe acute respiratory syndrome coro   ····················  ····················  ····················  ····················  ····················  ····················
2957 MW327513.1 |Severe acute respiratory syndrome coro   ····················  ····················  ····················  ····················  ····················  ····················
2958 MW327514.1 |Severe acute respiratory syndrome coro   ····················  ····················  ····················  ····················  ····················  ····················
2959 MW327515.1 |Severe acute respiratory syndrome coro   ····················  ····················  ····················  ····················  ····················  ····················
2960 MW327516.1 |Severe acute respiratory syndrome coro   ····················  ····················  ····················  ····················  ····················  ····················
2961 MW327517.1 |Severe acute respiratory syndrome coro   ····················  ····················  ····················  ····················  ····················  ····················
```

| | | | | | | | | |
|---|---|---|---|---|---|---|---|---|
| 2962 MW327518.1 | \|Severe acute respiratory syndrome coro | ···················· | ···················· | ···················· | ···················· | ···················· | ···················· | |
| 2963 MW327519.1 | \|Severe acute respiratory syndrome coro | ···················· | ···················· | ···················· | ···················· | ···················· | ···················· | |
| 2964 MW327520.1 | \|Severe acute respiratory syndrome coro | ···················· | ···················· | ···················· | ···················· | ···················· | ···················· | |
| 2965 MW327521.1 | \|Severe acute respiratory syndrome coro | ···················· | ···················· | ···················· | ···················· | ···················· | ···················· | |
| 2966 MW327522.1 | \|Severe acute respiratory syndrome coro | ···················· | ···················· | ···················· | ···················· | ···················· | ···················· | |
| 2967 MW327523.1 | \|Severe acute respiratory syndrome coro | ···················· | ···················· | ···················· | ···················· | ···················· | ···················· | |
| 2968 MW327524.1 | \|Severe acute respiratory syndrome coro | ···················· | ···················· | ···················· | ···················· | ···················· | ···················· | |
| 2969 MW327525.1 | \|Severe acute respiratory syndrome coro | ···················· | ···················· | ···················· | ···················· | ···················· | ···················· | |
| 2970 MW327526.1 | \|Severe acute respiratory syndrome coro | ···················· | ···················· | ···················· | ···················· | ···················· | ···················· | |
| 2971 MW327527.1 | \|Severe acute respiratory syndrome coro | ···················· | ···················· | ···················· | ···················· | ···················· | ···················· | |
| 2972 MW327528.1 | \|Severe acute respiratory syndrome coro | ···················· | ···················· | ···················· | ···················· | ···················· | ···················· | |
| 2973 MW327529.1 | \|Severe acute respiratory syndrome coro | ···················· | ···················· | ···················· | ···················· | ···················· | ···················· | |
| 2974 MW327530.1 | \|Severe acute respiratory syndrome coro | ···················· | ···················· | ···················· | ···················· | ···················· | ···················· | |
| 2975 MW327531.1 | \|Severe acute respiratory syndrome coro | ···················· | ···················· | ···················· | ···················· | ···················· | ···················· | |
| 2976 MW327532.1 | \|Severe acute respiratory syndrome coro | ···················· | ···················· | ···················· | ···················· | ···················· | ···················· | |
| 2977 MW327533.1 | \|Severe acute respiratory syndrome coro | ···················· | ···················· | ···················· | ···················· | ···················· | ···················· | |
| 2978 MW327534.1 | \|Severe acute respiratory syndrome coro | ···················· | ···················· | ···················· | ···················· | ···················· | ···················· | |
| 2979 MW327535.1 | \|Severe acute respiratory syndrome coro | ···················· | ···················· | ···················· | ···················· | ···················· | ···················· | |
| 2980 MW327536.1 | \|Severe acute respiratory syndrome coro | ···················· | ···················· | ···················· | ···················· | ···················· | ···················· | |
| 2981 MW327537.1 | \|Severe acute respiratory syndrome coro | ···················· | ···················· | ···················· | ···················· | ···················· | ···················· | |
| 2982 MW327538.1 | \|Severe acute respiratory syndrome coro | ···················· | ···················· | ···················· | ···················· | ···················· | ···················· | |
| 2983 MW327539.1 | \|Severe acute respiratory syndrome coro | ···················· | ···················· | ···················· | ···················· | ···················· | ···················· | |
| 2985 MW320730.1 | \|Severe acute respiratory syndrome coro | ···················· | ···················· | ···················· | ···················· | ···················· | ···················· | |
| 2986 MW320731.1 | \|Severe acute respiratory syndrome coro | ···················· | ···················· | ···················· | ···················· | ···················· | ···················· | |
| 2987 MW320733.1 | \|Severe acute respiratory syndrome coro | ···················· | ···················· | ···················· | ···················· | ···················· | ···················· | |
| 2988 MW320735.1 | \|Severe acute respiratory syndrome coro | ···················· | ···················· | ···················· | ···················· | ···················· | ···················· | |
| 2989 MW320740.1 | \|Severe acute respiratory syndrome coro | ···················· | ···················· | ···················· | ···················· | ···················· | ···················· | |
| 2990 MW320741.1 | \|Severe acute respiratory syndrome coro | ···················· | ···················· | ···················· | ···················· | ···················· | ···················· | |
| 2991 MW320746.1 | \|Severe acute respiratory syndrome coro | ···················· | ···················· | ···················· | ···················· | ···················· | ···················· | |
| 2993 MW320749.1 | \|Severe acute respiratory syndrome coro | ···················· | ···················· | ···················· | ···················· | ···················· | ···················· | |
| 2995 MW320752.1 | \|Severe acute respiratory syndrome coro | ···················· | ···················· | ···················· | ···················· | ···················· | ···················· | |
| 2996 MW320754.1 | \|Severe acute respiratory syndrome coro | ···················· | ···················· | ···················· | ···················· | ···················· | ···················· | |
| 2997 MW320755.1 | \|Severe acute respiratory syndrome coro | ···················· | ···················· | ···················· | ···················· | ···················· | ···················· | |
| 2998 MW320757.1 | \|Severe acute respiratory syndrome coro | ···················· | ···················· | ···················· | ···················· | ···················· | ···················· | |
| 2999 MW320758.1 | \|Severe acute respiratory syndrome coro | ···················· | ···················· | ···················· | ···················· | ···················· | ···················· | |
| 3000 MW320759.1 | \|Severe acute respiratory syndrome coro | ···················· | ···················· | ···················· | ···················· | ···················· | ···················· | |
| 3001 MW320760.1 | \|Severe acute respiratory syndrome coro | ···················· | ···················· | ···················· | ···················· | ···················· | ···················· | |
| 3002 MW320761.1 | \|Severe acute respiratory syndrome coro | ···················· | ···················· | ···················· | ···················· | ···················· | ···················· | |
| 3003 MW320762.1 | \|Severe acute respiratory syndrome coro | ···················· | ···················· | ···················· | ···················· | ···················· | ···················· | |
| 3004 MW320763.1 | \|Severe acute respiratory syndrome coro | ···················· | ···················· | ···················· | ···················· | ···················· | ···················· | |
| 3005 MW320765.1 | \|Severe acute respiratory syndrome coro | ···················· | ···················· | ···················· | ···················· | ···················· | ···················· | |
| 3006 MW320766.1 | \|Severe acute respiratory syndrome coro | ···················· | ···················· | ···················· | ···················· | ···················· | ···················· | |
| 3007 MW320767.1 | \|Severe acute respiratory syndrome coro | ···················· | ···················· | ···················· | ···················· | ···················· | ···················· | |
| 3008 MW320770.1 | \|Severe acute respiratory syndrome coro | ···················· | ···················· | ···················· | ···················· | NNN················· | ···················· | 1 |
| 3010 MW320772.1 | \|Severe acute respiratory syndrome coro | ···················· | ······A············· | ···················· | ···················· | ···················· | ···················· | |
| 3011 MW320775.1 | \|Severe acute respiratory syndrome coro | ···················· | ···················· | ···················· | ···················· | ···················· | ···················· | |
| 3012 MW320779.1 | \|Severe acute respiratory syndrome coro | ···················· | ···················· | ···················· | ···················· | ···················· | ···················· | |
| 3013 MW320782.1 | \|Severe acute respiratory syndrome coro | ···················· | ···················· | ···················· | ···················· | ···················· | ···················· | |
| 3014 MW320784.1 | \|Severe acute respiratory syndrome coro | ···················· | ···················· | ···················· | ···················· | ···················· | ···················· | |
| 3015 MW320786.1 | \|Severe acute respiratory syndrome coro | ···················· | ···················· | ···················· | ···················· | ···················· | ···················· | |
| 3016 MW320787.1 | \|Severe acute respiratory syndrome coro | ···················· | ···················· | ···················· | ···················· | ···················· | ···················· | |
| 3017 MW320790.1 | \|Severe acute respiratory syndrome coro | ···················· | ···················· | ···················· | ···················· | ···················· | ···················· | |
| 3018 MW320792.1 | \|Severe acute respiratory syndrome coro | ···················· | ···················· | ···················· | ···················· | ···················· | ···················· | |
| 3019 MW320793.1 | \|Severe acute respiratory syndrome coro | ···················· | ···················· | ···················· | ···················· | ···················· | ···················· | |
| 3020 MW320794.1 | \|Severe acute respiratory syndrome coro | ···················· | ···················· | ···················· | ···················· | ···················· | ···················· | |
| 3021 MW320795.1 | \|Severe acute respiratory syndrome coro | ···················· | ···················· | ···················· | ···················· | ···················· | ···················· | |
| 3022 MW320797.1 | \|Severe acute respiratory syndrome coro | ···················· | ···················· | ···················· | ···················· | ···················· | ···················· | |
| 3023 MW320798.1 | \|Severe acute respiratory syndrome coro | ···················· | ···················· | ···················· | ···················· | ···················· | ···················· | |
| 3024 MW320799.1 | \|Severe acute respiratory syndrome coro | ···················· | ···················· | ···················· | ···················· | ···················· | ···················· | |
| 3025 MW320800.1 | \|Severe acute respiratory syndrome coro | ···················· | ···················· | ···················· | ···················· | ···················· | ···················· | |
| 3026 MW320802.1 | \|Severe acute respiratory syndrome coro | ···················· | ···················· | ···················· | ···················· | ·····Y·············· | ···················· | |
| 3027 MW320805.1 | \|Severe acute respiratory syndrome coro | ···················· | ···················· | ···················· | ···················· | ···················· | ···················· | |
| 3028 MW320806.1 | \|Severe acute respiratory syndrome coro | ···················· | ···················· | ···················· | ···················· | ···················· | ···················· | |
| 3029 MW320808.1 | \|Severe acute respiratory syndrome coro | ···················· | ···················· | ···················· | ···················· | ···················· | ···················· | |
| 3030 MW320815.1 | \|Severe acute respiratory syndrome coro | ···················· | ···················· | ···················· | ···················· | ···················· | ···················· | |
| 3031 MW320818.1 | \|Severe acute respiratory syndrome coro | ···················· | ···················· | ···················· | ···················· | ···················· | ···················· | |
| 3032 MW320819.1 | \|Severe acute respiratory syndrome coro | ···················· | ···················· | ···················· | ···················· | ···················· | ···················· | |
| 3033 MW320820.1 | \|Severe acute respiratory syndrome coro | ···················· | ···················· | ···················· | ···················· | ···················· | ···················· | |
| 3034 MW320822.1 | \|Severe acute respiratory syndrome coro | ···················· | ···················· | ···················· | ···················· | ···················· | ···················· | |
| 3035 MW320823.1 | \|Severe acute respiratory syndrome coro | ···················· | ···················· | ···················· | ···················· | ···················· | ···················· | |
| 3036 MW320824.1 | \|Severe acute respiratory syndrome coro | ···················· | ···················· | ···················· | ···················· | ···················· | ···················· | |
| 3037 MW320825.1 | \|Severe acute respiratory syndrome coro | ···················· | ···················· | ···················· | ···················· | ···················· | ···················· | |
| 3038 MW320829.1 | \|Severe acute respiratory syndrome coro | ···················· | ···················· | ···················· | ···················· | ···················· | ···················· | |
| 3039 MW320830.1 | \|Severe acute respiratory syndrome coro | ···················· | ···················· | ···················· | ···················· | ···················· | ···················· | |
| 3040 MW320831.1 | \|Severe acute respiratory syndrome coro | ···················· | ···················· | ···················· | ···················· | ···················· | ···················· | |
| 3041 MW320834.1 | \|Severe acute respiratory syndrome coro | ···················· | ···················· | ···················· | ···················· | ···················· | ···················· | |
| 3042 MW320835.1 | \|Severe acute respiratory syndrome coro | ···················· | ···················· | ···················· | ···················· | ···················· | ···················· | |
| 3043 MW320838.1 | \|Severe acute respiratory syndrome coro | ···················· | ···················· | ···················· | ···················· | ···················· | ···················· | |

3044 MW320840.1 |Severe acute respiratory syndrome coro .................... .................... ..................... .................... .................... ....................
3045 MW320841.1 |Severe acute respiratory syndrome coro .................... .................... ..................... .................... .................... ....................
3046 MW320843.1 |Severe acute respiratory syndrome coro .................... .................... ..................... .................... .................... ....................
3047 MW320846.1 |Severe acute respiratory syndrome coro .................... .................... ..................... .................... .................... ....................
3048 MW320849.1 |Severe acute respiratory syndrome coro .................... .................... ..................... .................... .................... ....................
3049 MW320851.1 |Severe acute respiratory syndrome coro .................... .................... ..................... .................... .................... ....................
3050 MW320852.1 |Severe acute respiratory syndrome coro .................... .................... ..................... .................... .................... ....................
3051 MW320856.1 |Severe acute respiratory syndrome coro .................... .................... ..................... .................... .................... ....................
3052 MW320859.1 |Severe acute respiratory syndrome coro .................... .................... ..................... .................... .................... ....................
3053 MW320861.1 |Severe acute respiratory syndrome coro .................... .................... ..................... .................... .................... ....................
3055 MW320865.1 |Severe acute respiratory syndrome coro .................... .................... ..................... .................... .................... ....................
3056 MW320867.1 |Severe acute respiratory syndrome coro .................... .................... ..................... .................... .................... ....................
3057 MW320868.1 |Severe acute respiratory syndrome coro .................... .................... ..................... .................... .................... ....................
3058 MW320869.1 |Severe acute respiratory syndrome coro .................... .................... ..................... .................... .................... ....................
3059 MW320870.1 |Severe acute respiratory syndrome coro .................... .................... ..................... .................... .................... ....................
3060 MW320873.1 |Severe acute respiratory syndrome coro .................... .................... ..................... .................... .................... ....................
3061 MW320874.1 |Severe acute respiratory syndrome coro .................... .................... ..................... .................... .................... ....................
3062 MW320876.1 |Severe acute respiratory syndrome coro .................... .................... ..................... .................... .................... ....................
3063 MW320877.1 |Severe acute respiratory syndrome coro .................... .................... ..................... .................... .................... ....................
3064 MW320879.1 |Severe acute respiratory syndrome coro .................... .................... ..................... .................... .................... ....................
3065 MW320880.1 |Severe acute respiratory syndrome coro .................... .................... ..................... .................... .................... ....................
3067 MW320882.1 |Severe acute respiratory syndrome coro .................... .................... ..................... .................... .................... ....................
3068 MW320883.1 |Severe acute respiratory syndrome coro .................... .................... ..................... .................... .................... ....................
3070 MW320889.1 |Severe acute respiratory syndrome coro .................... .................... ..................... .................... .................... ....................
3071 MW320890.1 |Severe acute respiratory syndrome coro .................... .................... ..................... .................... .................... ....................
3073 MW320892.1 |Severe acute respiratory syndrome coro .................... .................... ..................... .................... .................... ....................
3074 MW320893.1 |Severe acute respiratory syndrome coro .................... .................... ..................... .................... .................... ....................
3075 MW320894.1 |Severe acute respiratory syndrome coro .................... .................... ..................... .................... .................... ....................
3076 MW320895.1 |Severe acute respiratory syndrome coro .................... .................... ..................... .................... .................... ....................
3077 MW320897.1 |Severe acute respiratory syndrome coro .................... .................... ..................... .................... .................... ....................
3078 MW320898.1 |Severe acute respiratory syndrome coro .................... .................... ..................... .................... .................... ....................
3079 MW320901.1 |Severe acute respiratory syndrome coro .................... .................... ..................... .................... .................... ....................
3080 MW320904.1 |Severe acute respiratory syndrome coro .................... .................... ..................... .................... .................... ....................
3081 MW320906.1 |Severe acute respiratory syndrome coro .................... .................... ..................... .................... .................... ....................
3083 MW320910.1 |Severe acute respiratory syndrome coro .................... .................... ..................... .................... .................... ....................
3084 MW320912.1 |Severe acute respiratory syndrome coro .................... .................... ..................... .................... .................... ....................
3086 MW320915.1 |Severe acute respiratory syndrome coro .................... .................... ..................... .................... .................... ....................
3087 MW320916.1 |Severe acute respiratory syndrome coro .................... .................... ..................... .................... .................... ....................
3088 MW320917.1 |Severe acute respiratory syndrome coro .................... .................... ..................... .................... .................... ....................
3089 MW320919.1 |Severe acute respiratory syndrome coro .................... .................... ..................... .................... .................... ....................
3091 MW320928.1 |Severe acute respiratory syndrome coro .................... .................... ..................... .................... .................... ....................
3092 MW320929.1 |Severe acute respiratory syndrome coro .................... .................... ..................... .................... .................... ....................
3095 MW320932.1 |Severe acute respiratory syndrome coro .................... .................... ..................... .................... .................... ....................
3096 MW320934.1 |Severe acute respiratory syndrome coro .................... .................... ..................... .................... .................... ....................
3098 MW320937.1 |Severe acute respiratory syndrome coro .................... .................... ..................... .................... .................... ....................
3099 MW320941.1 |Severe acute respiratory syndrome coro .................... .................... ..................... .................... .................... ....................
3100 MW320942.1 |Severe acute respiratory syndrome coro .................... .................... ..................... .................... .................... ....................
3101 MW320943.1 |Severe acute respiratory syndrome coro .................... .................... ..................... .................... .................... ....................
3102 MW320944.1 |Severe acute respiratory syndrome coro .................... .................... ..................... .................... .................... ....................
3103 MW320947.1 |Severe acute respiratory syndrome coro .................... .................... ..................... .................... .................... ....................
3104 MW320948.1 |Severe acute respiratory syndrome coro .................... .................... ..................... .................... .................... ....................
3105 MW320949.1 |Severe acute respiratory syndrome coro .................... .................... ..................... .................... .................... ....................
3106 MW320951.1 |Severe acute respiratory syndrome coro .................... .................... ..................... .................... .................... ....................
3107 MW320952.1 |Severe acute respiratory syndrome coro .................... .................... ..................... .................... .................... ....................
3109 MW320957.1 |Severe acute respiratory syndrome coro .................... .................... ..................... .................... .................... ....................
3110 MW320958.1 |Severe acute respiratory syndrome coro .................... .................... ..................... .................... .................... ....................
3112 MW320966.1 |Severe acute respiratory syndrome coro .................... .................... ..................... .................... .................... ....................
3114 MW320968.1 |Severe acute respiratory syndrome coro .................... .................... ..................... .................... .................... ....................
3115 MW320969.1 |Severe acute respiratory syndrome coro .................... .................... ..................... .................... .................... ....................
3116 MW320970.1 |Severe acute respiratory syndrome coro .................... .................... ..................... .................... .................... ....................
3117 MW320971.1 |Severe acute respiratory syndrome coro .................... .................... ..................... .................... .................... ....................
3118 MW320973.1 |Severe acute respiratory syndrome coro .................... .................... ..................... .................... .................... ....................
3119 MW320975.1 |Severe acute respiratory syndrome coro .................... .................... ..................... .................... .................... ....................
3120 MW320976.1 |Severe acute respiratory syndrome coro .................... .................... ..................... .................... .................... ....................
3122 MW320978.1 |Severe acute respiratory syndrome coro .................... .................... ..................... .................... .................... ....................
3123 MW320980.1 |Severe acute respiratory syndrome coro .................... .................... ..................... .................... .................... ....................
3124 MW320983.1 |Severe acute respiratory syndrome coro .................... .................... ..................... .................... .................... ....................
3125 MW320984.1 |Severe acute respiratory syndrome coro .................... .................... ..................... .................... .................... ....................
3126 MW320985.1 |Severe acute respiratory syndrome coro .................... .................... ..................... .................... .................... ....................
3127 MW320986.1 |Severe acute respiratory syndrome coro .................... .................... ..................... .................... .................... ....................
3128 MW320987.1 |Severe acute respiratory syndrome coro .................... .................... ..................... .................... .................... ....................
3129 MW320989.1 |Severe acute respiratory syndrome coro .................... .................... ..................... .................... .................... ....................
3130 MW320990.1 |Severe acute respiratory syndrome coro .................... .................... ..................... .................... .................... ....................
3131 MW320992.1 |Severe acute respiratory syndrome coro .................... .................... ..................... .................... .................... ....................
3132 MW320993.1 |Severe acute respiratory syndrome coro .................... .................... ..................... .................... .................... ....................
3134 MW320995.1 |Severe acute respiratory syndrome coro .................... .................... ..................... .................... .................... ....................
3135 MW320998.1 |Severe acute respiratory syndrome coro .................... .................... ..................... .................... .................... ....................
3136 MW320999.1 |Severe acute respiratory syndrome coro .................... .................... ..................... .................... .................... ....................

| | | | | | | | | | |
|---|---|---|---|---|---|---|---|---|---|
| 3138 | MW321006.1 | Severe acute respiratory syndrome coro | ···················· | ···················· | ···················· | ···················· | ···················· | ···················· | |
| 3139 | MW321008.1 | Severe acute respiratory syndrome coro | ···················· | ···················· | ···················· | ···················· | ···················· | ···················· | |
| 3140 | MW321009.1 | Severe acute respiratory syndrome coro | ···················· | ···················· | ···················· | ···················· | ···················· | ···················· | |
| 3141 | MW321010.1 | Severe acute respiratory syndrome coro | ···················· | ···················· | ···················· | ···················· | ···················· | ···················· | |
| 3142 | MW321011.1 | Severe acute respiratory syndrome coro | ···················· | ···················· | ···················· | ···················· | ···················· | ···················· | |
| 3143 | MW321014.1 | Severe acute respiratory syndrome coro | ···················· | ···················· | ···················· | ···················· | ···················· | ···················· | |
| 3144 | MW321019.1 | Severe acute respiratory syndrome coro | ···················· | ···················· | ···················· | ···················· | ···················· | ···················· | |
| 3145 | MW321022.1 | Severe acute respiratory syndrome coro | ···················· | ···················· | ···················· | ···················· | ···················· | ···················· | |
| 3146 | MW321024.1 | Severe acute respiratory syndrome coro | ···················· | ···················· | ···················· | ···················· | ···················· | ···················· | |
| 3147 | MW321025.1 | Severe acute respiratory syndrome coro | ···················· | ···················· | ···················· | ···················· | ···················· | ···················· | |
| 3148 | MW321026.1 | Severe acute respiratory syndrome coro | ···················· | ···················· | ···················· | ···················· | ···················· | ···················· | |
| 3149 | MW321030.1 | Severe acute respiratory syndrome coro | ···················· | ···················· | ···················· | ···················· | ···················· | ···················· | |
| 3150 | MW321033.1 | Severe acute respiratory syndrome coro | ···················· | ···················· | ···················· | ···················· | ···················· | ···················· | |
| 3151 | MW321036.1 | Severe acute respiratory syndrome coro | ···················· | ···················· | ···················· | ···················· | ···················· | ···················· | |
| 3154 | MW321042.1 | Severe acute respiratory syndrome coro | ···················· | ···················· | ···················· | ···················· | ···················· | ···················· | |
| 3155 | MW321043.1 | Severe acute respiratory syndrome coro | ···················· | ···················· | ···················· | ···················· | ···················· | ···················· | |
| 3156 | MW321044.1 | Severe acute respiratory syndrome coro | ···················· | ···················· | ···················· | ···················· | ···················· | ···················· | |
| 3158 | MW321046.1 | Severe acute respiratory syndrome coro | ···················· | ···················· | ···················· | ···················· | ···················· | ···················· | |
| 3159 | MW321048.1 | Severe acute respiratory syndrome coro | ···················· | ···················· | ···················· | ···················· | ···················· | ···················· | |
| 3161 | MW321052.1 | Severe acute respiratory syndrome coro | ···················· | ···················· | ···················· | ···················· | ···················· | ···················· | |
| 3162 | MW321053.1 | Severe acute respiratory syndrome coro | ···················· | ···················· | ···················· | ···················· | ···················· | ···················· | |
| 3163 | MW321054.1 | Severe acute respiratory syndrome coro | ···················· | ···················· | ···················· | ···················· | ···················· | ···················· | |
| 3164 | MW321056.1 | Severe acute respiratory syndrome coro | ···················· | ···················· | ···················· | ···················· | ···················· | ···················· | |
| 3165 | MW321057.1 | Severe acute respiratory syndrome coro | ···················· | ···················· | ···················· | ···················· | ···················· | ···················· | |
| 3166 | MW321058.1 | Severe acute respiratory syndrome coro | ···················· | ···················· | ···················· | ···················· | ···················· | ···················· | |
| 3167 | MW321059.1 | Severe acute respiratory syndrome coro | ···················· | ···················· | ···················· | ···················· | ···················· | ···················· | |
| 3168 | MW321060.1 | Severe acute respiratory syndrome coro | ···················· | ···················· | ···················· | ···················· | ···················· | ···················· | |
| 3169 | MW321062.1 | Severe acute respiratory syndrome coro | ···················· | ···················· | ···················· | ···················· | ···················· | ···················· | |
| 3170 | MW321063.1 | Severe acute respiratory syndrome coro | ···················· | ···················· | ···················· | ···················· | ···················· | ···················· | |
| 3171 | MW321064.1 | Severe acute respiratory syndrome coro | ···················· | ···················· | ···················· | ···················· | ···················· | ···················· | |
| 3173 | MW321066.1 | Severe acute respiratory syndrome coro | ···················· | ···················· | ···················· | ···················· | ···················· | ···················· | |
| 3174 | MW321067.1 | Severe acute respiratory syndrome coro | ···················· | ···················· | ···················· | ···················· | ···················· | ···················· | |
| 3175 | MW321068.1 | Severe acute respiratory syndrome coro | ···················· | ···················· | ···················· | ···················· | ···················· | ···················· | |
| 3176 | MW321070.1 | Severe acute respiratory syndrome coro | ···················· | ···················· | ···················· | ···················· | ···················· | ···················· | |
| 3177 | MW321071.1 | Severe acute respiratory syndrome coro | ···················· | ···················· | ···················· | ···················· | ···················· | ···················· | |
| 3180 | MW321080.1 | Severe acute respiratory syndrome coro | ···················· | ···················· | ···················· | ···················· | ···················· | ···················· | |
| 3181 | MW321081.1 | Severe acute respiratory syndrome coro | ···················· | ···················· | ···················· | ···················· | ···················· | ···················· | |
| 3182 | MW321082.1 | Severe acute respiratory syndrome coro | ···················· | ···················· | ···················· | ···················· | ···················· | ···················· | |
| 3183 | MW321085.1 | Severe acute respiratory syndrome coro | ···················· | ···················· | ···················· | ···················· | ···················· | ···················· | |
| 3184 | MW321086.1 | Severe acute respiratory syndrome coro | ···················· | ···················· | ···················· | ···················· | ···················· | ···················· | |
| 3186 | MW321090.1 | Severe acute respiratory syndrome coro | ···················· | ···················· | ···················· | ···················· | ···················· | ···················· | |
| 3187 | MW321091.1 | Severe acute respiratory syndrome coro | ···················· | ···················· | ···················· | ···················· | ···················· | ···················· | |
| 3189 | MW321093.1 | Severe acute respiratory syndrome coro | ···················· | ···················· | ···················· | ···················· | ···················· | ···················· | |
| 3191 | MW321098.1 | Severe acute respiratory syndrome coro | ···················· | ···················· | ···················· | ···················· | ···················· | ···················· | |
| 3192 | MW321099.1 | Severe acute respiratory syndrome coro | ···················· | ···················· | ···················· | ···················· | ···················· | ···················· | |
| 3193 | MW321101.1 | Severe acute respiratory syndrome coro | ···················· | ···················· | ···················· | ···················· | ···················· | ···················· | |
| 3194 | MW321103.1 | Severe acute respiratory syndrome coro | ···················· | ···················· | ···················· | ···················· | ···················· | ···················· | |
| 3195 | MW321104.1 | Severe acute respiratory syndrome coro | ···················· | ···················· | ···················· | ···················· | ···················· | ···················· | |
| 3196 | MW321107.1 | Severe acute respiratory syndrome coro | ···················· | ···················· | ···················· | ···················· | ···················· | ···················· | |
| 3198 | MW321111.1 | Severe acute respiratory syndrome coro | ···················· | ···················· | ···················· | ···················· | ···················· | ···················· | |
| 3199 | MW321112.1 | Severe acute respiratory syndrome coro | ···················· | ···················· | ···················· | ···················· | ···················· | ···················· | |
| 3200 | MW321113.1 | Severe acute respiratory syndrome coro | ···················· | ···················· | ···················· | ···················· | ···················· | ···················· | |
| 3201 | MW321114.1 | Severe acute respiratory syndrome coro | ···················· | ···················· | ···················· | ···················· | ···················· | ···················· | |
| 3202 | MW321115.1 | Severe acute respiratory syndrome coro | ···················· | ···················· | ···················· | ···················· | ···················· | ···················· | |
| 3203 | MW321116.1 | Severe acute respiratory syndrome coro | ···················· | ···················· | ···················· | ···················· | ···················· | ···················· | |
| 3205 | MW321119.1 | Severe acute respiratory syndrome coro | ···················· | ···················· | ···················· | ···················· | ···················· | ···················· | |
| 3206 | MW321120.1 | Severe acute respiratory syndrome coro | ···················· | ···················· | ···················· | ···················· | ···················· | ···················· | |
| 3207 | MW321121.1 | Severe acute respiratory syndrome coro | ···················· | ···················· | ···················· | ···················· | ···················· | ···················· | |
| 3208 | MW321122.1 | Severe acute respiratory syndrome coro | ···················· | ···················· | ···················· | ···················· | ···················· | ···················· | |
| 3209 | MW321123.1 | Severe acute respiratory syndrome coro | ···················· | ···················· | ···················· | ···················· | ···················· | ···················· | |
| 3210 | MW321124.1 | Severe acute respiratory syndrome coro | ···················· | ···················· | ···················· | ···················· | ···················· | ···················· | |
| 3211 | MW321125.1 | Severe acute respiratory syndrome coro | ···················· | ···················· | ···················· | ···················· | ···················· | ···················· | |
| 3212 | MW321127.1 | Severe acute respiratory syndrome coro | ···················· | ···················· | ···················· | ···················· | NNN················· | ···················· | 1 |
| 3213 | MW321128.1 | Severe acute respiratory syndrome coro | ···················· | ···················· | ···················· | ···················· | ···················· | ···················· | |
| 3214 | MW321132.1 | Severe acute respiratory syndrome coro | ···················· | ···················· | ···················· | ···················· | ···················· | ···················· | |
| 3215 | MW321134.1 | Severe acute respiratory syndrome coro | ···················· | ···················· | ···················· | ···················· | ···················· | ···················· | |
| 3216 | MW321136.1 | Severe acute respiratory syndrome coro | ···················· | ···················· | ···················· | ···················· | ···················· | ···················· | |
| 3217 | MW321137.1 | Severe acute respiratory syndrome coro | ···················· | ···················· | ···················· | ···················· | ···················· | ···················· | |
| 3218 | MW321139.1 | Severe acute respiratory syndrome coro | ···················· | ···················· | ···················· | ···················· | ···················· | ···················· | |
| 3219 | MW321143.1 | Severe acute respiratory syndrome coro | ···················· | ···················· | ···················· | ···················· | ···················· | ···················· | |
| 3220 | MW321148.1 | Severe acute respiratory syndrome coro | ···················· | ···················· | ···················· | ···················· | ···················· | ···················· | |
| 3222 | MW321150.1 | Severe acute respiratory syndrome coro | ···················· | ···················· | ···················· | ···················· | ···················· | ···················· | |
| 3223 | MW321155.1 | Severe acute respiratory syndrome coro | ···················· | ···················· | ···················· | ···················· | ···················· | ···················· | |
| 3224 | MW321157.1 | Severe acute respiratory syndrome coro | ·······Y············ | ···················· | ···················· | ···················· | ···················· | ···················· | |
| 3226 | MW321160.1 | Severe acute respiratory syndrome coro | ···················· | ···················· | ···················· | ···················· | ···················· | ···················· | |
| 3227 | MW321163.1 | Severe acute respiratory syndrome coro | ···················· | ···················· | ···················· | ···················· | ···················· | ···················· | |
| 3228 | MW321165.1 | Severe acute respiratory syndrome coro | ···················· | ···················· | ···················· | ···················· | ···················· | ···················· | |
| 3229 | MW321166.1 | Severe acute respiratory syndrome coro | ···················· | ···················· | ···················· | ···················· | ···················· | NN·················· | |

| | | | | | | | | | |
|---|---|---|---|---|---|---|---|---|---|
| 3230 | MW321167.1 | \|Severe acute respiratory syndrome coro | .................... | .................... | .................... | .................... | .................... | .................... | |
| 3231 | MW321171.1 | \|Severe acute respiratory syndrome coro | .................... | .................... | .................... | .................... | .................... | .................... | |
| 3233 | MW321173.1 | \|Severe acute respiratory syndrome coro | .................... | .................... | .................... | .................... | .................... | .................... | |
| 3234 | MW321174.1 | \|Severe acute respiratory syndrome coro | .................... | .................... | .................... | .................... | .................... | .................... | |
| 3235 | MW321176.1 | \|Severe acute respiratory syndrome coro | .................... | .................... | .................... | .................... | .................... | .................... | |
| 3236 | MW321177.1 | \|Severe acute respiratory syndrome coro | .................... | .................... | .................... | .................... | .................... | .................... | |
| 3237 | MW321179.1 | \|Severe acute respiratory syndrome coro | .................... | .................... | .................... | .................... | .................... | .................... | |
| 3238 | MW321184.1 | \|Severe acute respiratory syndrome coro | .................... | .................... | .................... | .................... | .................... | .................... | |
| 3239 | MW321186.1 | \|Severe acute respiratory syndrome coro | .................... | .................... | .................... | .................... | .................... | .................... | |
| 3240 | MW321189.1 | \|Severe acute respiratory syndrome coro | .................... | .................... | .................... | .................... | .................... | .................... | |
| 3241 | MW321193.1 | \|Severe acute respiratory syndrome coro | .................... | .................... | .................... | .................... | .................... | .................... | |
| 3242 | MW321196.1 | \|Severe acute respiratory syndrome coro | .................... | .................... | .................... | .................... | .................... | .................... | |
| 3243 | MW321197.1 | \|Severe acute respiratory syndrome coro | .................... | .................... | .................... | .................... | .................... | .................... | |
| 3244 | MW321201.1 | \|Severe acute respiratory syndrome coro | .................... | .................... | .................... | .................... | .................... | .................... | |
| 3245 | MW321202.1 | \|Severe acute respiratory syndrome coro | .................... | .................... | .................... | .................... | .................... | .................... | |
| 3246 | MW321203.1 | \|Severe acute respiratory syndrome coro | .................... | .................... | .................... | .................... | .................... | .................... | |
| 3247 | MW321206.1 | \|Severe acute respiratory syndrome coro | .................... | .................... | .................... | .................... | .................... | .................... | |
| 3248 | MW321207.1 | \|Severe acute respiratory syndrome coro | .................... | .................... | .................... | .................... | .................... | .................... | |
| 3249 | MW321208.1 | \|Severe acute respiratory syndrome coro | .................... | .................... | .................... | .................... | .................... | .................... | |
| 3250 | MW321211.1 | \|Severe acute respiratory syndrome coro | .................... | .................... | .................... | .................... | .................... | .................... | |
| 3251 | MW321212.1 | \|Severe acute respiratory syndrome coro | .................... | .................... | .................... | .................... | .................... | .................... | |
| 3252 | MW321214.1 | \|Severe acute respiratory syndrome coro | .................... | .................... | .................... | .................... | .................... | .................... | |
| 3253 | MW321216.1 | \|Severe acute respiratory syndrome coro | .................... | .................... | .................... | .................... | .................... | .................... | |
| 3254 | MW321217.1 | \|Severe acute respiratory syndrome coro | .................... | .................... | .................... | .................... | .................... | .................... | |
| 3256 | MW321220.1 | \|Severe acute respiratory syndrome coro | .................... | .................... | .................... | .................... | .................... | .................... | |
| 3257 | MW321221.1 | \|Severe acute respiratory syndrome coro | .................... | .................... | .................... | .................... | .................... | .................... | |
| 3258 | MW321225.1 | \|Severe acute respiratory syndrome coro | .................... | .................... | .................... | .................... | .................... | .................... | |
| 3259 | MW321226.1 | \|Severe acute respiratory syndrome coro | .................... | .................... | .................... | .................... | .................... | .................... | |
| 3260 | MW321228.1 | \|Severe acute respiratory syndrome coro | .................... | .................... | .................... | .................... | .................... | .................... | |
| 3261 | MW321229.1 | \|Severe acute respiratory syndrome coro | .................... | .................... | .................... | .................... | .................... | .................... | |
| 3262 | MW321230.1 | \|Severe acute respiratory syndrome coro | .................... | .................... | .................... | .................... | .................... | .................... | |
| 3263 | MW321233.1 | \|Severe acute respiratory syndrome coro | .................... | .................... | .................... | .................... | .................... | .................... | |
| 3264 | MW321238.1 | \|Severe acute respiratory syndrome coro | .................... | .................... | .................... | .................... | NNN................. | .................... | 1 |
| 3265 | MW321242.1 | \|Severe acute respiratory syndrome coro | .................... | .................... | .................... | .................... | .................... | .................... | |
| 3266 | MW321243.1 | \|Severe acute respiratory syndrome coro | .................... | .................... | .................... | .................... | .................... | .................... | |
| 3267 | MW321244.1 | \|Severe acute respiratory syndrome coro | .................... | .................... | .................... | .................... | .................... | .................... | |
| 3268 | MW321245.1 | \|Severe acute respiratory syndrome coro | .................... | .................... | .................... | .................... | .................... | .................... | |
| 3269 | MW321247.1 | \|Severe acute respiratory syndrome coro | .................... | .................... | .................... | .................... | .................... | .................... | |
| 3271 | MW321250.1 | \|Severe acute respiratory syndrome coro | .................... | .................... | .................... | .................... | .................... | .................... | |
| 3272 | MW321251.1 | \|Severe acute respiratory syndrome coro | .................... | .................... | .................... | .................... | .................... | .................... | |
| 3273 | MW321252.1 | \|Severe acute respiratory syndrome coro | .................... | .................... | .................... | .................... | .................... | .................... | |
| 3277 | MW321261.1 | \|Severe acute respiratory syndrome coro | .................... | .................... | .................... | .................... | .................... | .................... | |
| 3278 | MW321262.1 | \|Severe acute respiratory syndrome coro | .................... | .................... | .................... | .................... | .................... | .................... | |
| 3279 | MW321264.1 | \|Severe acute respiratory syndrome coro | .................... | .................... | .................... | .................... | .................... | .................... | |
| 3280 | MW321265.1 | \|Severe acute respiratory syndrome coro | .................... | .................... | .................... | .................... | .................... | .................... | |
| 3281 | MW321267.1 | \|Severe acute respiratory syndrome coro | .................... | .................... | .................... | .................... | .................... | .................... | |
| 3282 | MW321269.1 | \|Severe acute respiratory syndrome coro | .................... | .................... | .................... | .................... | .................... | .................... | |
| 3283 | MW321270.1 | \|Severe acute respiratory syndrome coro | .................... | .................... | .................... | .................... | .................... | .................... | |
| 3284 | MW321271.1 | \|Severe acute respiratory syndrome coro | .................... | .................... | .................... | .................... | .................... | .................... | |
| 3285 | MW321272.1 | \|Severe acute respiratory syndrome coro | .................... | .................... | .................... | .................... | .................... | .................... | |
| 3287 | MW321275.1 | \|Severe acute respiratory syndrome coro | .................... | .................... | .................... | .................... | .................... | .................... | |
| 3288 | MW321276.1 | \|Severe acute respiratory syndrome coro | .................... | .................... | .................... | .................... | .................... | .................... | |
| 3290 | MW321278.1 | \|Severe acute respiratory syndrome coro | .................... | .................... | .................... | .................... | .................... | .................... | |
| 3292 | MW321282.1 | \|Severe acute respiratory syndrome coro | .................... | .................... | .................... | .................... | .................... | .................... | |
| 3293 | MW321283.1 | \|Severe acute respiratory syndrome coro | .................... | .................... | .................... | .................... | .................... | .................... | |
| 3295 | MW321285.1 | \|Severe acute respiratory syndrome coro | .................... | .................... | .................... | .................... | .................... | .................... | |
| 3296 | MW321286.1 | \|Severe acute respiratory syndrome coro | .................... | .................... | .................... | .................... | .................... | .................... | |
| 3297 | MW321289.1 | \|Severe acute respiratory syndrome coro | .................... | .................... | .................... | .................... | .................... | .................... | |
| 3298 | MW321292.1 | \|Severe acute respiratory syndrome coro | .................... | .................... | .................... | .................... | .................... | .....W.............. | |
| 3299 | MW321295.1 | \|Severe acute respiratory syndrome coro | .................... | .................... | .................... | .................... | .................... | .................... | |
| 3301 | MW321298.1 | \|Severe acute respiratory syndrome coro | .................... | .................... | .................... | .................... | .................... | .................... | |
| 3302 | MW321300.1 | \|Severe acute respiratory syndrome coro | .................... | .................... | .................... | .................... | .................... | .................... | |
| 3303 | MW321303.1 | \|Severe acute respiratory syndrome coro | .................... | .................... | .................... | .................... | .................... | .................... | |
| 3304 | MW321304.1 | \|Severe acute respiratory syndrome coro | .................... | .................... | .................... | .................... | .................... | .................... | |
| 3306 | MW321316.1 | \|Severe acute respiratory syndrome coro | .................... | .................... | .................... | .................... | .................... | .................... | |
| 3307 | MW321320.1 | \|Severe acute respiratory syndrome coro | .................... | .................... | .................... | .................... | .................... | .................... | |
| 3308 | MW321321.1 | \|Severe acute respiratory syndrome coro | .................... | .................... | .................... | .................... | .................... | .................... | |
| 3309 | MW321322.1 | \|Severe acute respiratory syndrome coro | .................... | .................... | .................... | .................... | .................... | .................... | |
| 3310 | MW321326.1 | \|Severe acute respiratory syndrome coro | .................... | .................... | .................... | .................... | .................... | .................... | |
| 3311 | MW321327.1 | \|Severe acute respiratory syndrome coro | .................... | .................... | .................... | .................... | .................... | .................... | |
| 3313 | MW321329.1 | \|Severe acute respiratory syndrome coro | .................... | .................... | .................... | .................... | .................... | .................... | |
| 3314 | MW321332.1 | \|Severe acute respiratory syndrome coro | .................... | .................... | .................... | .................... | .................... | .................... | |
| 3315 | MW321336.1 | \|Severe acute respiratory syndrome coro | .................... | .................... | .................... | .................... | .................... | .................... | |
| 3316 | MW321337.1 | \|Severe acute respiratory syndrome coro | .................... | .................... | .................... | .................... | .................... | .................... | |
| 3318 | MW321343.1 | \|Severe acute respiratory syndrome coro | .................... | .................... | .................... | .................... | .................... | .................... | |
| 3319 | MW321344.1 | \|Severe acute respiratory syndrome coro | .................... | .................... | .................... | .................... | .................... | .................... | |
| 3320 | MW321345.1 | \|Severe acute respiratory syndrome coro | .................... | .................... | .................... | .................... | .................... | .................... | |
| 3321 | MW321346.1 | \|Severe acute respiratory syndrome coro | .................... | .................... | .................... | .................... | .................... | .................... | |

| | | | | | | | | |
|---|---|---|---|---|---|---|---|---|
| 3322 | MW321349.1 | \|Severe acute respiratory syndrome coro | ···················· | ···················· | ····················· | ···················· | ···················· | ···················· |
| 3323 | MW321350.1 | \|Severe acute respiratory syndrome coro | ···················· | ···················· | ····················· | ···················· | ···················· | ···················· |
| 3324 | MW321355.1 | \|Severe acute respiratory syndrome coro | ···················· | ···················· | ····················· | ···················· | ···················· | ···················· |
| 3325 | MW321359.1 | \|Severe acute respiratory syndrome coro | ···················· | ···················· | ····················· | ···················· | ···················· | ···················· |
| 3328 | MW321368.1 | \|Severe acute respiratory syndrome coro | ···················· | ···················· | ····················· | ···················· | ···················· | ···················· |
| 3329 | MW321369.1 | \|Severe acute respiratory syndrome coro | ···················· | ···················· | ····················· | ···················· | ···················· | ···················· |
| 3330 | MW321370.1 | \|Severe acute respiratory syndrome coro | ···················· | ···················· | ····················· | ···················· | ···················· | ···················· |
| 3331 | MW321371.1 | \|Severe acute respiratory syndrome coro | ···················· | ···················· | ····················· | ···················· | ···················· | ···················· |
| 3332 | MW321373.1 | \|Severe acute respiratory syndrome coro | ···················· | ···················· | ····················· | ···················· | ···················· | ···················· |
| 3333 | MW321374.1 | \|Severe acute respiratory syndrome coro | ···················· | ···················· | ····················· | ···················· | ···················· | ···················· |
| 3335 | MW321377.1 | \|Severe acute respiratory syndrome coro | ···················· | ···················· | ····················· | ···················· | ···················· | ···················· |
| 3336 | MW321382.1 | \|Severe acute respiratory syndrome coro | ···················· | ···················· | ····················· | ···················· | ···················· | ···················· |
| 3337 | MW321383.1 | \|Severe acute respiratory syndrome coro | ···················· | ···················· | ····················· | ···················· | ···················· | ···················· |
| 3338 | MW321385.1 | \|Severe acute respiratory syndrome coro | ···················· | ···················· | ····················· | ···················· | ···················· | ···················· |
| 3339 | MW321388.1 | \|Severe acute respiratory syndrome coro | ···················· | ···················· | ····················· | ···················· | ···················· | ···················· |
| 3340 | MW321390.1 | \|Severe acute respiratory syndrome coro | ···················· | ···················· | ····················· | ···················· | ···················· | ···················· |
| 3341 | MW321391.1 | \|Severe acute respiratory syndrome coro | ···················· | ···················· | ····················· | ···················· | ···················· | ···················· |
| 3342 | MW321396.1 | \|Severe acute respiratory syndrome coro | ···················· | ···················· | ····················· | ···················· | ···················· | ···················· |
| 3343 | MW321398.1 | \|Severe acute respiratory syndrome coro | ···················· | ···················· | ····················· | ···················· | ···················· | ···················· |
| 3344 | MW321399.1 | \|Severe acute respiratory syndrome coro | ···················· | ···················· | ····················· | ···················· | ···················· | ···················· |
| 3345 | MW321401.1 | \|Severe acute respiratory syndrome coro | ···················· | ···················· | ····················· | ···················· | ···················· | ···················· |
| 3346 | MW321402.1 | \|Severe acute respiratory syndrome coro | ···················· | ···················· | ····················· | ···················· | ···················· | ···················· |
| 3347 | MW321403.1 | \|Severe acute respiratory syndrome coro | ···················· | ···················· | ····················· | ···················· | ···················· | ···················· |
| 3348 | MW321404.1 | \|Severe acute respiratory syndrome coro | ···················· | ···················· | ····················· | ···················· | ···················· | ···················· |
| 3349 | MW321408.1 | \|Severe acute respiratory syndrome coro | ···················· | ···················· | ····················· | ···················· | ···················· | ···················· |
| 3350 | MW321411.1 | \|Severe acute respiratory syndrome coro | ···················· | ···················· | ····················· | ···················· | ···················· | ···················· |
| 3351 | MW321412.1 | \|Severe acute respiratory syndrome coro | ···················· | ···················· | ····················· | ···················· | ···················· | ···················· |
| 3354 | MW321415.1 | \|Severe acute respiratory syndrome coro | ···················· | ···················· | ····················· | ···················· | ···················· | ···················· |
| 3355 | MW321417.1 | \|Severe acute respiratory syndrome coro | ···················· | ···················· | ····················· | ···················· | ···················· | ···················· |
| 3356 | MW321420.1 | \|Severe acute respiratory syndrome coro | ···················· | ···················· | ····················· | ···················· | ···················· | ···················· |
| 3357 | MW321421.1 | \|Severe acute respiratory syndrome coro | ···················· | ···················· | ····················· | ···················· | ···················· | ···················· |
| 3358 | MW321425.1 | \|Severe acute respiratory syndrome coro | ···················· | ···················· | ····················· | ···················· | ···················· | ···················· |
| 3359 | MW321426.1 | \|Severe acute respiratory syndrome coro | ···················· | ···················· | ····················· | ···················· | ···················· | ···················· |
| 3361 | MW321429.1 | \|Severe acute respiratory syndrome coro | ···················· | ···················· | ····················· | ···················· | ·Y·················· | ·············S······ |
| 3362 | MW321430.1 | \|Severe acute respiratory syndrome coro | ···················· | ···················· | ····················· | ···················· | ···················· | ···················· |
| 3364 | MW315209.1 | \|Severe acute respiratory syndrome coro | ···················· | ·A·················· | ····················· | ···················· | ···················· | ···················· |
| 3365 | MW315210.1 | \|Severe acute respiratory syndrome coro | ···················· | ·A·················· | ····················· | ···················· | ···················· | ···················· |
| 3366 | MW315211.1 | \|Severe acute respiratory syndrome coro | ···················· | ·A·················· | ····················· | ···················· | ···················· | ···················· |
| 3367 | MW315212.1 | \|Severe acute respiratory syndrome coro | ···················· | ·A·················· | ····················· | ···················· | ···················· | ···················· |
| 3368 | MW315213.1 | \|Severe acute respiratory syndrome coro | ···················· | ·A·················· | ····················· | ···················· | ···················· | ···················· |
| 3369 | MW315214.1 | \|Severe acute respiratory syndrome coro | ···················· | ·A·················· | ····················· | ···················· | ···················· | ···················· |
| 3370 | MW315215.1 | \|Severe acute respiratory syndrome coro | ···················· | ·A·················· | ····················· | ···················· | ···················· | ···················· |
| 3371 | MW315965.1 | \|Severe acute respiratory syndrome coro | ···················· | ···················· | ····················· | ···················· | ···················· | ···················· |
| 3372 | MW315966.1 | \|Severe acute respiratory syndrome coro | ···················· | ···················· | ····················· | ···················· | ···················· | ···················· |
| 3373 | MW315967.1 | \|Severe acute respiratory syndrome coro | ···················· | ···················· | ····················· | ···················· | ···················· | ···················· |
| 3374 | MW315968.1 | \|Severe acute respiratory syndrome coro | ···················· | ···················· | ····················· | ···················· | ···················· | ···················· |
| 3375 | MW315969.1 | \|Severe acute respiratory syndrome coro | ···················· | ···················· | ····················· | ···················· | ···················· | ···················· |
| 3376 | MW315970.1 | \|Severe acute respiratory syndrome coro | ···················· | ···················· | ····················· | ···················· | ···················· | ···················· |
| 3377 | MW315971.1 | \|Severe acute respiratory syndrome coro | ···················· | ···················· | ····················· | ···················· | ···················· | ···················· |
| 3378 | MW315972.1 | \|Severe acute respiratory syndrome coro | ···················· | ···················· | ····················· | ···················· | ···················· | ···················· |
| 3379 | MW315973.1 | \|Severe acute respiratory syndrome coro | ···T················ | ···················· | ····················· | ···················· | ···················· | ···················· |
| 3380 | MW315974.1 | \|Severe acute respiratory syndrome coro | ···················· | ···················· | ····················· | ···················· | ···················· | ···················· |
| 3381 | MW315975.1 | \|Severe acute respiratory syndrome coro | ···················· | ···················· | ····················· | ···················· | ···················· | ···················· |
| 3382 | MW315976.1 | \|Severe acute respiratory syndrome coro | ···················· | ·A·················· | ····················· | ···················· | ···················· | ···················· |
| 3383 | MW315977.1 | \|Severe acute respiratory syndrome coro | ···················· | ···················· | ····················· | ···················· | ···················· | ···················· |
| 3384 | MW315978.1 | \|Severe acute respiratory syndrome coro | ···················· | ···················· | ····················· | ···················· | ···················· | ···················· |
| 3385 | MW315979.1 | \|Severe acute respiratory syndrome coro | ···················· | ···················· | ····················· | ···················· | ···················· | ···················· |
| 3386 | MW315980.1 | \|Severe acute respiratory syndrome coro | ···················· | ···················· | ····················· | ···················· | ···················· | ···················· |
| 3387 | MW315981.1 | \|Severe acute respiratory syndrome coro | ···················· | ···················· | ····················· | ···················· | ···················· | ···················· |
| 3388 | MW315982.1 | \|Severe acute respiratory syndrome coro | ···················· | ···················· | ····················· | ···················· | ···················· | ···················· |
| 3389 | MW315983.1 | \|Severe acute respiratory syndrome coro | ···················· | ···················· | ····················· | ···················· | ···················· | ···················· |
| 3390 | MW315984.1 | \|Severe acute respiratory syndrome coro | ···················· | ···················· | ····················· | ···················· | ···················· | ···················· |
| 3391 | MW315985.1 | \|Severe acute respiratory syndrome coro | ···················· | ···················· | ····················· | ···················· | ···················· | ···················· |
| 3392 | MW316741.1 | \|Severe acute respiratory syndrome coro | ···················· | ···················· | ····················· | ···················· | ···················· | ···················· |
| 3393 | MW309426.1 | \|Severe acute respiratory syndrome coro | ···················· | ···················· | ····················· | ···················· | ···················· | ···················· |
| 3394 | MW309429.1 | \|Severe acute respiratory syndrome coro | ···················· | ···················· | ····················· | ···················· | ···················· | ···················· |
| 3395 | MW309438.1 | \|Severe acute respiratory syndrome coro | ···················· | ···················· | ····················· | ···················· | ···················· | ···················· |
| 3396 | MW309443.1 | \|Severe acute respiratory syndrome coro | ···················· | ····A··············· | ····················· | ···················· | ···················· | ···················· |
| 3397 | MW309444.1 | \|Severe acute respiratory syndrome coro | ···················· | ···················· | ····················· | ···················· | ···················· | ···················· |
| 3398 | MW309446.1 | \|Severe acute respiratory syndrome coro | ···················· | ···················· | ····················· | ···················· | ···················· | ···················· |
| 3399 | MW309449.1 | \|Severe acute respiratory syndrome coro | ···················· | ···················· | ····················· | ···················· | ···················· | ···················· |
| 3400 | MW309463.1 | \|Severe acute respiratory syndrome coro | ···················· | ···················· | ····················· | ···················· | ···················· | ···················· |
| 3401 | MW309470.1 | \|Severe acute respiratory syndrome coro | ···················· | ···················· | ····················· | ···················· | ···················· | ···················· |
| 3402 | MW309474.1 | \|Severe acute respiratory syndrome coro | ···················· | ···················· | ····················· | ···················· | ···················· | ···················· |
| 3405 | MW309482.1 | \|Severe acute respiratory syndrome coro | ···················· | ···················· | ····················· | ···················· | ···················· | ···················· |
| 3407 | MW309485.1 | \|Severe acute respiratory syndrome coro | ···················· | ···················· | ····················· | ···················· | ···················· | ···················· |
| 3409 | MW309488.1 | \|Severe acute respiratory syndrome coro | ···················· | ···················· | ····················· | ···················· | ···················· | ···················· |
| 3410 | MW309493.1 | \|Severe acute respiratory syndrome coro | ···················· | ···················· | ····················· | ···················· | ···················· | ···················· |

| | | | | | | | | |
|---|---|---|---|---|---|---|---|---|
| 3411 | MW309496.1 | \|Severe acute respiratory syndrome coro | ···················· | ····A··············· | ····················· | ···················· | ···················· | ···················· |
| 3412 | MW309829.1 | \|Severe acute respiratory syndrome coro | ···················· | ···················· | ····················· | ···················· | ···················· | ···················· |
| 3413 | MW309830.1 | \|Severe acute respiratory syndrome coro | ···················· | ···················· | ····················· | ···················· | ···················· | ···················· |
| 3414 | MW309831.1 | \|Severe acute respiratory syndrome coro | ···················· | ···················· | ····················· | ···················· | ···················· | ···················· |
| 3415 | MW310413.1 | \|Severe acute respiratory syndrome coro | ···················· | ·A·················· | ····················· | ···················· | ···················· | ···················· |
| 3416 | MW310414.1 | \|Severe acute respiratory syndrome coro | ···················· | ···················· | ····················· | ···················· | ···················· | ···················· |
| 3417 | MW310415.1 | \|Severe acute respiratory syndrome coro | ···················· | ·A·················· | ····················· | ···················· | ···················· | ···················· |
| 3418 | MW310416.1 | \|Severe acute respiratory syndrome coro | ···················· | ···················· | ····················· | ···················· | ···················· | ···················· |
| 3419 | MW310417.1 | \|Severe acute respiratory syndrome coro | ···················· | ·A·················· | ····················· | ···················· | ···················· | ···················· |
| 3420 | MW310418.1 | \|Severe acute respiratory syndrome coro | ···················· | ··········A········· | ····················· | ···················· | ···················· | ···················· |
| 3421 | MW310419.1 | \|Severe acute respiratory syndrome coro | ···················· | ···················· | ····················· | ···················· | ···················· | ···················· |
| 3422 | MW310420.1 | \|Severe acute respiratory syndrome coro | ···················· | ············C······· | ····················· | ···················· | ···················· | ···················· |
| 3423 | MW310421.1 | \|Severe acute respiratory syndrome coro | ···················· | ············C······· | ····················· | ···················· | ···················· | ···················· |
| 3424 | MW310422.1 | \|Severe acute respiratory syndrome coro | ···················· | ···················· | ····················· | ···················· | ···················· | ···················· |
| 3425 | MW310423.1 | \|Severe acute respiratory syndrome coro | ···················· | ···················· | ····················· | ···················· | ···················· | ···················· |
| 3426 | MW310424.1 | \|Severe acute respiratory syndrome coro | ···················· | ···················· | ····················· | ···················· | ···················· | ···················· |
| 3427 | MW310425.1 | \|Severe acute respiratory syndrome coro | ···················· | ···················· | ····················· | ···················· | ···················· | ···················· |
| 3428 | MW310426.1 | \|Severe acute respiratory syndrome coro | ···················· | ·A·················· | ····················· | ···················· | ···················· | ···················· |
| 3429 | MW310427.1 | \|Severe acute respiratory syndrome coro | ···················· | ·············A······ | ····················· | ···················· | ···················· | ···················· |
| 3430 | MW310428.1 | \|Severe acute respiratory syndrome coro | ···················· | ·············A······ | ····················· | ···················· | ···················· | ···················· |
| 3431 | MW310429.1 | \|Severe acute respiratory syndrome coro | ···················· | ···················· | ···········T········· | ···················· | ···················· | ···················· |
| 3432 | MW310430.1 | \|Severe acute respiratory syndrome coro | ···················· | ···················· | ····················· | ···················· | ···················· | ···················· |
| 3433 | MW310431.1 | \|Severe acute respiratory syndrome coro | ···················· | ···················· | ····················· | ···················· | ···················· | ···················· |
| 3434 | MW310432.1 | \|Severe acute respiratory syndrome coro | ···················· | ·A·················· | ····················· | ···················· | ···················· | ···················· |
| 3435 | MW310433.1 | \|Severe acute respiratory syndrome coro | ···················· | ···················· | ····················· | ···················· | ···················· | ···················· |
| 3436 | MW310434.1 | \|Severe acute respiratory syndrome coro | ···················· | ···················· | ····················· | ···················· | ···················· | ···················· |
| 3437 | MW310435.1 | \|Severe acute respiratory syndrome coro | ···················· | ···················· | ····················· | ···················· | ···················· | ···················· |
| 3438 | MW310436.1 | \|Severe acute respiratory syndrome coro | ···················· | ·A·················· | ····················· | ···················· | ···················· | ···················· |
| 3439 | MW310437.1 | \|Severe acute respiratory syndrome coro | ···················· | ·A·················· | ····················· | ···················· | ···················· | ···················· |
| 3440 | MW310438.1 | \|Severe acute respiratory syndrome coro | ···················· | ·A·················· | ····················· | ···················· | ···················· | ···················· |
| 3441 | MW310439.1 | \|Severe acute respiratory syndrome coro | ···················· | ·A·················· | ····················· | ···················· | ···················· | ···················· |
| 3442 | MW310440.1 | \|Severe acute respiratory syndrome coro | ···················· | ···················· | ····················· | ···················· | ···················· | ···················· |
| 3443 | MW310441.1 | \|Severe acute respiratory syndrome coro | ···················· | ···················· | ····················· | ···················· | ···················· | ···················· |
| 3444 | MW310442.1 | \|Severe acute respiratory syndrome coro | ···················· | ···················· | ····················· | ···················· | ···················· | ···················· |
| 3445 | MW310443.1 | \|Severe acute respiratory syndrome coro | ···················· | ···················· | ····················· | ···················· | ···················· | ···················· |
| 3446 | MW310444.1 | \|Severe acute respiratory syndrome coro | ···················· | ···················· | ····················· | ···················· | ···················· | ···················· |
| 3447 | MW310445.1 | \|Severe acute respiratory syndrome coro | ···················· | ···················· | ····················· | ···················· | ···················· | ···················· |
| 3448 | MW310446.1 | \|Severe acute respiratory syndrome coro | ···················· | ···················· | ····················· | ···················· | ···················· | ···················· |
| 3449 | MW310447.1 | \|Severe acute respiratory syndrome coro | ···················· | ·A·················· | ····················· | ···················· | ···················· | ···················· |
| 3450 | MW310448.1 | \|Severe acute respiratory syndrome coro | ···················· | ···················· | ····················· | ···················· | ···················· | ···················· |
| 3451 | MW310449.1 | \|Severe acute respiratory syndrome coro | ···················· | ·A·················· | ····················· | ···················· | ···················· | ···················· |
| 3452 | MW310450.1 | \|Severe acute respiratory syndrome coro | ···················· | ···················· | ····················· | ···················· | ···················· | ···················· |
| 3453 | MW310451.1 | \|Severe acute respiratory syndrome coro | ···················· | ·A·················· | ····················· | ···················· | ···················· | ···················· |
| 3454 | MW310452.1 | \|Severe acute respiratory syndrome coro | ···················· | ·A·················· | ····················· | ···················· | ···················· | ···················· |
| 3455 | MW310453.1 | \|Severe acute respiratory syndrome coro | ···················· | ···················· | ····················· | ···················· | ···················· | ···················· |
| 3456 | MW310454.1 | \|Severe acute respiratory syndrome coro | ···················· | ···················· | ····················· | ···················· | ···················· | ···················· |
| 3457 | MW310455.1 | \|Severe acute respiratory syndrome coro | ···················· | ···················· | ····················· | ···················· | ···················· | ···················· |
| 3458 | MW310456.1 | \|Severe acute respiratory syndrome coro | ···················· | ···················· | ····················· | ···················· | ···················· | ···················· |
| 3459 | MW314605.1 | \|Severe acute respiratory syndrome coro | ···················· | ···················· | ····················· | ···················· | ···················· | ···················· |
| 3460 | MW302336.1 | \|Severe acute respiratory syndrome coro | ···················· | ···················· | ····················· | ···················· | ···················· | ···················· |
| 3461 | MW302341.1 | \|Severe acute respiratory syndrome coro | ···················· | ···················· | ····················· | ···················· | ···················· | ···················· |
| 3462 | MW306294.1 | \|Severe acute respiratory syndrome coro | ···················· | ·A·················· | ····················· | ···················· | ···················· | ···················· |
| 3463 | MW306295.1 | \|Severe acute respiratory syndrome coro | ···················· | ···················· | ····················· | ···················· | ···················· | ···················· |
| 3464 | MW306296.1 | \|Severe acute respiratory syndrome coro | ···················· | ···················· | ····················· | ···················· | ···················· | ···················· |
| 3465 | MW306297.1 | \|Severe acute respiratory syndrome coro | ···················· | ···················· | ····················· | ···················· | ···················· | ···················· |
| 3466 | MW306298.1 | \|Severe acute respiratory syndrome coro | ···················· | ···················· | ····················· | ···················· | ···················· | ···················· |
| 3467 | MW306299.1 | \|Severe acute respiratory syndrome coro | ···················· | ···················· | ····················· | ···················· | ··········Y········· | ···················· |
| 3468 | MW306300.1 | \|Severe acute respiratory syndrome coro | ···················· | ···················· | ····················· | ···················· | ···················· | ···················· |
| 3469 | MW306304.1 | \|Severe acute respiratory syndrome coro | ···················· | ·A·················· | ····················· | ···················· | ···················· | ···················· |
| 3470 | MW306306.1 | \|Severe acute respiratory syndrome coro | ···················· | ·A·················· | ····················· | ···················· | ···················· | ···················· |
| 3471 | MW306307.1 | \|Severe acute respiratory syndrome coro | ···················· | ·A·················· | ····················· | ···················· | ···················· | ···················· |
| 3472 | MW306308.1 | \|Severe acute respiratory syndrome coro | ···················· | ·A·················· | ····················· | ···················· | ···················· | ···················· |
| 3473 | MW306309.1 | \|Severe acute respiratory syndrome coro | ···················· | ·A·················· | ····················· | ···················· | ···················· | ···················· |
| 3474 | MW306315.1 | \|Severe acute respiratory syndrome coro | ···················· | ···················· | ····················· | ···················· | ···················· | ···················· |
| 3475 | MW306318.1 | \|Severe acute respiratory syndrome coro | ···················· | ·A·················· | ····················· | ···················· | ···················· | ···················· |
| 3476 | MW306319.1 | \|Severe acute respiratory syndrome coro | ···················· | ·A·················· | ····················· | ···················· | ···················· | ···················· |
| 3477 | MW306320.1 | \|Severe acute respiratory syndrome coro | ···················· | ·A·················· | ····················· | ···················· | N··················· | ···················· |
| 3480 | MW306323.1 | \|Severe acute respiratory syndrome coro | ···················· | ·A·················· | ····················· | ···················· | ···················· | ···················· |
| 3481 | MW306324.1 | \|Severe acute respiratory syndrome coro | ···················· | ···················· | ····················· | ···················· | ···················· | ···················· |
| 3482 | MW306325.1 | \|Severe acute respiratory syndrome coro | ···················· | ···················· | ····················· | ···················· | ···················· | ···················· |
| 3484 | MW306328.1 | \|Severe acute respiratory syndrome coro | ···················· | ·A·················· | ····················· | ···················· | ···················· | ···················· |
| 3485 | MW306329.1 | \|Severe acute respiratory syndrome coro | ···················· | ·A·················· | ····················· | ···················· | ···················· | ···················· |
| 3486 | MW306330.1 | \|Severe acute respiratory syndrome coro | ···················· | ·A·················· | ····················· | ···················· | ···················· | ···················· |
| 3487 | MW306332.1 | \|Severe acute respiratory syndrome coro | ···················· | ···················· | ····················· | ···················· | ···················· | ···················· |
| 3488 | MW306333.1 | \|Severe acute respiratory syndrome coro | ···················· | ·············A······ | ····················· | ···················· | ···················· | ···················· |
| 3489 | MW306334.1 | \|Severe acute respiratory syndrome coro | ···················· | ·A·················· | ····················· | ···················· | ···················· | ···················· |
| 3490 | MW306335.1 | \|Severe acute respiratory syndrome coro | ···················· | ·A·················· | ····················· | ···················· | ···················· | ···················· |
| 3491 | MW306336.1 | \|Severe acute respiratory syndrome coro | ···················· | ·A·················· | ····················· | ···················· | ···················· | ···················· |

| | | | | | | | | | |
|---|---|---|---|---|---|---|---|---|---|
| 3492 | MW306337.1 | \|Severe acute respiratory syndrome coro | ···················· | ·A·················· | ···················· | ···················· | ···················· | ···················· | |
| 3493 | MW306338.1 | \|Severe acute respiratory syndrome coro | ···················· | ·A·················· | ···················· | ···················· | ···················· | ···················· | |
| 3494 | MW306339.1 | \|Severe acute respiratory syndrome coro | ···················· | ·A·················· | ···················· | ···················· | ···················· | ···················· | |
| 3495 | MW306340.1 | \|Severe acute respiratory syndrome coro | ···················· | ·A·················· | ···················· | ···················· | ···················· | ···················· | |
| 3496 | MW306341.1 | \|Severe acute respiratory syndrome coro | ···················· | ·A·················· | ···················· | ···················· | ···················· | ···················· | |
| 3497 | MW306342.1 | \|Severe acute respiratory syndrome coro | ···················· | ·A·················· | ···················· | ···················· | ···················· | ···················· | |
| 3498 | MW306343.1 | \|Severe acute respiratory syndrome coro | ···················· | ·A·················· | ···················· | ···················· | ···················· | ···················· | |
| 3499 | MW306344.1 | \|Severe acute respiratory syndrome coro | ···················· | ·A·················· | ···················· | ···················· | ···················· | ···················· | |
| 3500 | MW306345.1 | \|Severe acute respiratory syndrome coro | ···················· | ·A·················· | ···················· | ···················· | ···················· | ···················· | |
| 3501 | MW306346.1 | \|Severe acute respiratory syndrome coro | ···················· | ·A·················· | ···················· | ···················· | ···················· | ···················· | |
| 3502 | MW306347.1 | \|Severe acute respiratory syndrome coro | ···················· | ·A·················· | ···················· | ···················· | ···················· | ···················· | |
| 3503 | MW306348.1 | \|Severe acute respiratory syndrome coro | ···················· | ···················· | ···················· | ···················· | ···················· | ···················· | |
| 3504 | MW306349.1 | \|Severe acute respiratory syndrome coro | ···················· | ···················· | ···················· | ···················· | ···················· | ···················· | |
| 3505 | MW306350.1 | \|Severe acute respiratory syndrome coro | ···················· | ·A·················· | ···················· | ···················· | ···················· | ···················· | |
| 3506 | MW306351.1 | \|Severe acute respiratory syndrome coro | ···················· | ···················· | ···················· | ···················· | ···················· | ···················· | |
| 3507 | MW306353.1 | \|Severe acute respiratory syndrome coro | ···················· | ·A·················· | ···················· | ···················· | ···················· | ···················· | |
| 3508 | MW306355.1 | \|Severe acute respiratory syndrome coro | ···················· | ·A·················· | ···················· | ···················· | ···················· | ···················· | |
| 3509 | MW306356.1 | \|Severe acute respiratory syndrome coro | ···················· | ···················· | ···················· | ···················· | ···················· | ···················· | |
| 3510 | MW306358.1 | \|Severe acute respiratory syndrome coro | ···················· | ·A·················· | ···················· | ···················· | ···················· | ···················· | |
| 3512 | MW306360.1 | \|Severe acute respiratory syndrome coro | ···················· | ·A·················· | ···················· | ···················· | ···················· | ···················· | |
| 3513 | MW306361.1 | \|Severe acute respiratory syndrome coro | ···················· | ·A·················· | ···················· | ···················· | ···················· | ···················· | |
| 3514 | MW306362.1 | \|Severe acute respiratory syndrome coro | ···················· | ·A·················· | ···················· | ···················· | ···················· | ···················· | |
| 3515 | MW306363.1 | \|Severe acute respiratory syndrome coro | ···················· | ·A·················· | ···················· | ···················· | ···················· | ···················· | |
| 3517 | MW306366.1 | \|Severe acute respiratory syndrome coro | ···················· | ·A·················· | ···················· | ···················· | ···················· | ···················· | |
| 3518 | MW306367.1 | \|Severe acute respiratory syndrome coro | ···················· | ···················· | ···················· | ···················· | ···················· | ···················· | |
| 3519 | MW306369.1 | \|Severe acute respiratory syndrome coro | ···················· | ···················· | ···················· | ···················· | ···················· | ···················· | |
| 3520 | MW306370.1 | \|Severe acute respiratory syndrome coro | ···················· | ···················· | ···················· | ···················· | ···················· | ···················· | |
| 3521 | MW306371.1 | \|Severe acute respiratory syndrome coro | ···················· | ···················· | ···················· | ···················· | NNN················· | ···················· | 1 |
| 3522 | MW306372.1 | \|Severe acute respiratory syndrome coro | ···················· | ···················· | ···················· | ···················· | ···················· | ···················· | |
| 3523 | MW306373.1 | \|Severe acute respiratory syndrome coro | ···················· | ···················· | ···················· | ···················· | ···················· | ···················· | |
| 3524 | MW306374.1 | \|Severe acute respiratory syndrome coro | ···················· | ···················· | ···················· | ···················· | ···················· | ···················· | |
| 3525 | MW306375.1 | \|Severe acute respiratory syndrome coro | ···················· | ···················· | ···················· | ···················· | ···················· | ···················· | |
| 3526 | MW306376.1 | \|Severe acute respiratory syndrome coro | ···················· | ···················· | ···················· | ···················· | ···················· | ···················· | |
| 3527 | MW306377.1 | \|Severe acute respiratory syndrome coro | ···················· | ···················· | ···················· | ···················· | ···················· | ···················· | |
| 3528 | MW306378.1 | \|Severe acute respiratory syndrome coro | ···················· | ···················· | ···················· | ···················· | ···················· | ···················· | |
| 3529 | MW306379.1 | \|Severe acute respiratory syndrome coro | ···················· | ···················· | ···················· | ···················· | ···················· | ···················· | |
| 3530 | MW306385.1 | \|Severe acute respiratory syndrome coro | ···················· | ·A·················· | ···················· | ···················· | ···················· | ···················· | |
| 3531 | MW306386.1 | \|Severe acute respiratory syndrome coro | ···················· | ·A·················· | ···················· | ···················· | ···················· | ···················· | |
| 3532 | MW306387.1 | \|Severe acute respiratory syndrome coro | ···················· | ·A·················· | ···················· | ···················· | ···················· | ···················· | |
| 3533 | MW306388.1 | \|Severe acute respiratory syndrome coro | ···················· | ·A·················· | ···················· | ···················· | ···················· | ···················· | |
| 3534 | MW306389.1 | \|Severe acute respiratory syndrome coro | ···················· | ·A·················· | ···················· | ···················· | ···················· | ···················· | |
| 3535 | MW306391.1 | \|Severe acute respiratory syndrome coro | ···················· | ·A·················· | ···················· | ···················· | ···················· | ···················· | |
| 3536 | MW306392.1 | \|Severe acute respiratory syndrome coro | ···················· | ·A·················· | ···················· | ···················· | ···················· | ···················· | |
| 3537 | MW306393.1 | \|Severe acute respiratory syndrome coro | ···················· | ·A·················· | ···················· | ···················· | ···················· | ···················· | |
| 3538 | MW306397.1 | \|Severe acute respiratory syndrome coro | ···················· | ·A·················· | ···················· | ···················· | ···················· | ···················· | |
| 3539 | MW306398.1 | \|Severe acute respiratory syndrome coro | ···················· | ·A·················· | ···················· | ···················· | ···················· | ···················· | |
| 3540 | MW306399.1 | \|Severe acute respiratory syndrome coro | ···················· | ·A·················· | ···················· | ···················· | ···················· | ···················· | |
| 3541 | MW306401.1 | \|Severe acute respiratory syndrome coro | ···················· | ·A·················· | ···················· | ···················· | ···················· | ···················· | |
| 3542 | MW306402.1 | \|Severe acute respiratory syndrome coro | ···················· | ·A·················· | ···················· | ···················· | ···················· | ···················· | |
| 3543 | MW306403.1 | \|Severe acute respiratory syndrome coro | ···················· | ·A·················· | ···················· | ···················· | ···················· | ···················· | |
| 3544 | MW306404.1 | \|Severe acute respiratory syndrome coro | ···················· | ·A·················· | ···················· | ···················· | ···················· | ···················· | |
| 3546 | MW306406.1 | \|Severe acute respiratory syndrome coro | ···················· | ···················· | ···················· | ···················· | ···················· | ···················· | |
| 3547 | MW306407.1 | \|Severe acute respiratory syndrome coro | ···················· | ···················· | ···················· | ···················· | ···················· | ···················· | |
| 3548 | MW306409.1 | \|Severe acute respiratory syndrome coro | ···················· | ···················· | ···················· | ···················· | ···················· | ···················· | |
| 3549 | MW306410.1 | \|Severe acute respiratory syndrome coro | ···················· | ···················· | ···················· | ···················· | ···················· | ···················· | |
| 3550 | MW306411.1 | \|Severe acute respiratory syndrome coro | ···················· | ···················· | ···················· | ···················· | ···················· | ···················· | |
| 3551 | MW306412.1 | \|Severe acute respiratory syndrome coro | ···················· | ···················· | ···················· | ···················· | ···················· | ···················· | |
| 3552 | MW306413.1 | \|Severe acute respiratory syndrome coro | ···················· | ···················· | ···················· | ···················· | ···················· | ···················· | |
| 3553 | MW306415.1 | \|Severe acute respiratory syndrome coro | ···················· | ···················· | ···················· | ···················· | ···················· | ···················· | |
| 3554 | MW306416.1 | \|Severe acute respiratory syndrome coro | ···················· | ···················· | ···················· | ···················· | ···················· | ···················· | |
| 3555 | MW306417.1 | \|Severe acute respiratory syndrome coro | ···················· | ···················· | ···················· | ···················· | ···················· | ···················· | |
| 3556 | MW306418.1 | \|Severe acute respiratory syndrome coro | ···················· | ···················· | ···················· | ···················· | ···················· | ···················· | |
| 3557 | MW306419.1 | \|Severe acute respiratory syndrome coro | ···················· | ···················· | ···················· | ···················· | ···················· | ···················· | |
| 3558 | MW306420.1 | \|Severe acute respiratory syndrome coro | ···················· | ·A·················· | ···················· | ···················· | ···················· | ···················· | |
| 3559 | MW306421.1 | \|Severe acute respiratory syndrome coro | ···················· | ···················· | ···················· | ···················· | ···················· | ···················· | |
| 3560 | MW306422.1 | \|Severe acute respiratory syndrome coro | ···················· | ···················· | ···················· | ···················· | ···················· | ···················· | |
| 3561 | MW306424.1 | \|Severe acute respiratory syndrome coro | ···················· | ···················· | ···················· | ···················· | ···················· | ···················· | |
| 3562 | MW306426.1 | \|Severe acute respiratory syndrome coro | ···················· | ···················· | ···················· | ···················· | ···················· | ···················· | |
| 3563 | MW306427.1 | \|Severe acute respiratory syndrome coro | ···················· | ···················· | ···················· | ···················· | ···················· | ···················· | |
| 3564 | MW306428.1 | \|Severe acute respiratory syndrome coro | ···················· | ···················· | ···················· | ···················· | ···················· | ···················· | |
| 3565 | MW306429.1 | \|Severe acute respiratory syndrome coro | ···················· | ···················· | ···················· | ···················· | ···················· | ···················· | |
| 3566 | MW306430.1 | \|Severe acute respiratory syndrome coro | ···················· | ···················· | ···················· | ···················· | ···················· | ···················· | |
| 3567 | MW306431.1 | \|Severe acute respiratory syndrome coro | ···················· | ···················· | ···················· | ···················· | ···················· | ···················· | |
| 3568 | MW306432.1 | \|Severe acute respiratory syndrome coro | ···················· | ···················· | ···················· | ···················· | ···················· | ···················· | |
| 3569 | MW306433.1 | \|Severe acute respiratory syndrome coro | ···················· | ···················· | ···················· | ···················· | ···················· | ···················· | |
| 3570 | MW306434.1 | \|Severe acute respiratory syndrome coro | ···················· | ···················· | ···················· | ···················· | ···················· | ···················· | |
| 3571 | MW306435.1 | \|Severe acute respiratory syndrome coro | ···················· | ···················· | ···················· | ···················· | ···················· | ···················· | |
| 3572 | MW306436.1 | \|Severe acute respiratory syndrome coro | ···················· | ···················· | ···················· | ···················· | ···················· | ···················· | |

3573 MW306437.1 |Severe acute respiratory syndrome coro ···················· ···················· ····················· ···················· ···················· ····················
3574 MW306438.1 |Severe acute respiratory syndrome coro ···················· ·A·················· ····················· ···················· ···················· ····················
3575 MW306439.1 |Severe acute respiratory syndrome coro ···················· ·A·················· ····················· ···················· ···················· ····················
3576 MW306440.1 |Severe acute respiratory syndrome coro ···················· ·A·················· ····················· ···················· ···················· ····················
3577 MW306441.1 |Severe acute respiratory syndrome coro ···················· ·A·················· ····················· ···················· ···················· ····················
3578 MW306442.1 |Severe acute respiratory syndrome coro ···················· ·A·················· ····················· ···················· ···················· ····················
3579 MW306443.1 |Severe acute respiratory syndrome coro ···················· ····A··············· ····················· ···················· ···················· ····················
3580 MW306444.1 |Severe acute respiratory syndrome coro ···················· ···················· ····················· ···················· ···················· ····················
3581 MW306445.1 |Severe acute respiratory syndrome coro ···················· ·A·················· ····················· ···················· ···················· ····················
3582 MW306446.1 |Severe acute respiratory syndrome coro ···················· ·A·················· ····················· ···················· ···················· ····················
3583 MW306447.1 |Severe acute respiratory syndrome coro ···················· ·A·················· ····················· ···················· ···················· ····················
3584 MW306448.1 |Severe acute respiratory syndrome coro ···················· ···················· ····················· ···················· ···················· ····················
3585 MW306449.1 |Severe acute respiratory syndrome coro ···················· ···················· ····················· ···················· ···················· ····················
3586 MW306450.1 |Severe acute respiratory syndrome coro ···················· ···················· ····················· ···················· ···················· ····················
3587 MW306666.1 |Severe acute respiratory syndrome coro ···················· ···················· ····················· ···················· ···················· ····················
3588 MW306667.1 |Severe acute respiratory syndrome coro ···················· ···················· ····················· ···················· ···················· ····················
3589 MW306668.1 |Severe acute respiratory syndrome coro ···················· ···················· ····················· ···················· ···················· ····················
3590 MW307809.1 |Severe acute respiratory syndrome coro ···················· ···················· ····················· ···················· ···················· ····················
3591 MW307831.1 |Severe acute respiratory syndrome coro ···················· ·A·················· ····················· ···················· ···················· ····················
3592 MW307889.1 |Severe acute respiratory syndrome coro ···················· ·A·················· ····················· ···················· ···················· ····················
3593 MW307977.1 |Severe acute respiratory syndrome coro ···················· ·A·················· ····················· ···················· ···················· ····················
3594 MW307979.1 |Severe acute respiratory syndrome coro ···················· ·A·················· ····················· ···················· ···················· ····················
3595 MW308137.1 |Severe acute respiratory syndrome coro ···················· ·A·················· ····················· ···················· ···················· ····················
3596 MW301121.1 |Severe acute respiratory syndrome coro ···················· ···················· ····················· ···················· ···················· ····················
3597 MW301192.1 |Severe acute respiratory syndrome coro ···················· ···················· ····················· ···················· ···················· ····················
3598 MW295556.1 |Severe acute respiratory syndrome coro ···················· ···················· ····················· ···················· ···················· ····················
3599 MW295640.1 |Severe acute respiratory syndrome coro ···················· ···················· ····················· ···················· ···················· ····················
3600 MW295643.1 |Severe acute respiratory syndrome coro ···················· ···················· ····················· ···················· ···················· ····················
3601 MW290937.1 |Severe acute respiratory syndrome coro ···················· ···················· ····················· ···················· ···················· ····················
3602 MW290938.1 |Severe acute respiratory syndrome coro ···················· ···················· ····················· ···················· ···················· ····················
3603 MW290940.1 |Severe acute respiratory syndrome coro ···················· ···················· ····················· ···················· ···················· ····················
3604 MW290941.1 |Severe acute respiratory syndrome coro ···················· ···················· ····················· ···················· ···················· ····················
3605 MW290942.1 |Severe acute respiratory syndrome coro ···················· ···················· ····················· ···················· ···················· ····················
3606 MW290947.1 |Severe acute respiratory syndrome coro ···················· ···················· ····················· ···················· ···················· ····················
3607 MW290948.1 |Severe acute respiratory syndrome coro ···················· ···················· ····················· ···················· ···················· ····················
3608 MW290951.1 |Severe acute respiratory syndrome coro ···················· ···················· ····················· ···················· ···················· ····················
3609 MW290952.1 |Severe acute respiratory syndrome coro ···················· ···················· ····················· ···················· ···················· ····················
3610 MW290960.1 |Severe acute respiratory syndrome coro ···················· ···················· ····················· ···················· ···················· ····················
3611 MW290964.1 |Severe acute respiratory syndrome coro ···················· ···················· ····················· ···················· ···················· ····················
3612 MW290965.1 |Severe acute respiratory syndrome coro ···················· ···················· ····················· ···················· ···················· ····················
3613 MW290966.1 |Severe acute respiratory syndrome coro ···················· ···················· ····················· ···················· ···················· ····················
3614 MW290969.1 |Severe acute respiratory syndrome coro ···················· ···················· ····················· ···················· ···················· ····················
3615 MW290970.1 |Severe acute respiratory syndrome coro ···················· ···················· ····················· ···················· ···················· ····················
3616 MW292629.1 |Severe acute respiratory syndrome coro ···················· ·A·················· ····················· ···················· ···················· ····················
3617 MW292630.1 |Severe acute respiratory syndrome coro ···················· ···················· ····················· ···················· ···················· ····················
3618 MW292632.1 |Severe acute respiratory syndrome coro ···················· ···················· ····················· ···················· ···················· ····················
3619 MW292633.1 |Severe acute respiratory syndrome coro ···················· ···················· ····················· ···················· ···················· ····················
3620 MW292634.1 |Severe acute respiratory syndrome coro ···················· ····A··············· ····················· ···················· ···················· ····················
3621 MW292635.1 |Severe acute respiratory syndrome coro ···················· ···················· ····················· ···················· ···················· ····················
3622 MW292636.1 |Severe acute respiratory syndrome coro ···················· ···················· ····················· ···················· ···················· ····················
3623 MW292637.1 |Severe acute respiratory syndrome coro ···················· ···················· ····················· ···················· ···················· ····················
3624 MW292638.1 |Severe acute respiratory syndrome coro ···················· ···················· ····················· ···················· ···················· ····················
3625 MW292639.1 |Severe acute respiratory syndrome coro ···················· ···················· ···········T········· ···················· ···················· ····················
3626 MW292640.1 |Severe acute respiratory syndrome coro ···················· ·A·················· ····················· ···················· ···················· ····················
3627 MW292641.1 |Severe acute respiratory syndrome coro ···················· ·A·················· ····················· ···················· ···················· ····················
3628 MW292642.1 |Severe acute respiratory syndrome coro ···················· ···················· ····················· ···················· ···················· ····················
3629 MW292644.1 |Severe acute respiratory syndrome coro ···················· ···················· ····················· ···················· ···················· ····················
3630 MW292645.1 |Severe acute respiratory syndrome coro ···················· ···················· ····················· ···················· ···················· ····················
3631 MW292646.1 |Severe acute respiratory syndrome coro ···················· ···················· ····················· ···················· ···················· ····················
3632 MW292647.1 |Severe acute respiratory syndrome coro ···················· ···················· ····················· ···················· ···················· ····················
3633 MW292648.1 |Severe acute respiratory syndrome coro ···················· ···················· ····················· ···················· ···················· ····················
3634 MW292649.1 |Severe acute respiratory syndrome coro ···················· ·A·················· ····················· ···················· ···················· ····················
3635 MW292650.1 |Severe acute respiratory syndrome coro ···················· ···················· ····················· ···················· ···················· ····················
3636 MW292651.1 |Severe acute respiratory syndrome coro ···················· ············C······· ····················· ···················· ···················· ····················
3637 MW292652.1 |Severe acute respiratory syndrome coro ···················· ············C······· ····················· ···················· ···················· ····················
3638 MW292653.1 |Severe acute respiratory syndrome coro ···················· ·A·················· ····················· ···················· ···················· ····················
3639 MW292654.1 |Severe acute respiratory syndrome coro ···················· ············C······· ····················· ···················· ···················· ····················
3640 MW292656.1 |Severe acute respiratory syndrome coro ···················· ···················· ····················· ···················· ···················· ····················
3641 MW292657.1 |Severe acute respiratory syndrome coro ···················· ···················· ····················· ···················· ···················· ····················
3642 MW292658.1 |Severe acute respiratory syndrome coro ···················· ···················· ····················· ···················· ···················· ····················
3643 MW292659.1 |Severe acute respiratory syndrome coro ···················· ·A·················· ····················· ···················· ···················· ····················
3644 MW292661.1 |Severe acute respiratory syndrome coro ···················· ·A·················· ····················· ···················· ···················· ····················
3645 MW292662.1 |Severe acute respiratory syndrome coro ···················· ·A·················· ····················· ···················· ···················· ····················
3646 MW292663.1 |Severe acute respiratory syndrome coro ···················· ···················· ····················· ···················· ···················· ····················
3647 MW292664.1 |Severe acute respiratory syndrome coro ···················· ···················· ····················· ···················· ···················· ····················
3648 MW292665.1 |Severe acute respiratory syndrome coro ···················· ···················· ····················· ···················· ···················· ····················
3649 MW294007.1 |Severe acute respiratory syndrome coro ···················· ···················· ····················· ···················· ···················· ····················
3650 MW294011.1 |Severe acute respiratory syndrome coro ···················· ···················· ····················· ···················· ···················· ····················

```
3651 MT994707.1 |Severe acute respiratory syndrome coro ···················· ···················· ···················· ···················· ···················· ····················
3652 MW288251.1 |Severe acute respiratory syndrome coro ···················· ···················· ···················· ···················· ···················· ····················
3653 MW288252.1 |Severe acute respiratory syndrome coro ···················· ···················· ···················· ···················· ···················· ····················
3654 MW288253.1 |Severe acute respiratory syndrome coro ···················· ···················· ···················· ···················· ···················· ····················
3655 MW288254.1 |Severe acute respiratory syndrome coro ···················· ···················· ···················· ···················· ···················· ····················
3656 MW288255.1 |Severe acute respiratory syndrome coro ···················· ···················· ···················· ···················· ···················· ····················
3657 MW288256.1 |Severe acute respiratory syndrome coro ···················· ···················· ···················· ···················· ···················· ····················
3658 MW288257.1 |Severe acute respiratory syndrome coro ···················· ···················· ···················· ···················· ···················· ····················
3659 MW288258.1 |Severe acute respiratory syndrome coro ···················· ···················· ···················· ···················· ···················· ····················
3660 MW288259.1 |Severe acute respiratory syndrome coro ···················· ·A·················· ···················· ···················· ···················· ····················
3661 MW288260.1 |Severe acute respiratory syndrome coro ···················· ···················· ···················· ···················· ···················· ····················
3662 MW288261.1 |Severe acute respiratory syndrome coro ···················· ···················· ···················· ···················· ···················· ····················
3663 MW288262.1 |Severe acute respiratory syndrome coro ···················· ···················· ···················· ···················· ···················· ····················
3664 MW288263.1 |Severe acute respiratory syndrome coro ···················· ···················· ···················· ···················· ···················· ····················
3665 MW288264.1 |Severe acute respiratory syndrome coro ···················· ···················· ···················· ···················· ···················· ····················
3666 MW288265.1 |Severe acute respiratory syndrome coro ···················· ···················· ···················· ···················· ···················· ····················
3667 MW288266.1 |Severe acute respiratory syndrome coro ···················· ···················· ···················· ···················· ···················· ····················
3668 MW288267.1 |Severe acute respiratory syndrome coro ···················· ···················· ···················· ···················· ···················· ····················
3669 MW288268.1 |Severe acute respiratory syndrome coro ···················· ···················· ···················· ···················· NNN················· ····················  1
3670 MW288269.1 |Severe acute respiratory syndrome coro ···················· ···················· ···················· ···················· ···················· ····················
3671 MW288270.1 |Severe acute respiratory syndrome coro ···················· ···················· ···················· ···················· ···················· ····················
3672 MW288271.1 |Severe acute respiratory syndrome coro ···················· ···················· ···················· ···················· ···················· ····················
3673 MW288272.1 |Severe acute respiratory syndrome coro ···················· ···················· ···················· ···················· ···················· ····················
3674 MW288273.1 |Severe acute respiratory syndrome coro ···················· ···················· ···················· ···················· ···················· ····················
3675 MW288274.1 |Severe acute respiratory syndrome coro ···················· ···················· ···················· ···················· ···················· ····················
3676 MW288275.1 |Severe acute respiratory syndrome coro ···················· ···················· ···················· ···················· ···················· ····················
3677 MW288276.1 |Severe acute respiratory syndrome coro ···················· ···················· ···················· ···················· ···················· ····················
3678 MW288277.1 |Severe acute respiratory syndrome coro ···················· ···················· ···················· ···················· ···················· ····················
3679 MW288278.1 |Severe acute respiratory syndrome coro ···················· ···················· ···················· ···················· ···················· ····················
3680 MW288279.1 |Severe acute respiratory syndrome coro ···················· ···················· ···················· ···················· ···················· ····················
3681 MW288280.1 |Severe acute respiratory syndrome coro ···················· ···················· ···················· ···················· ···················· ····················
3682 MW288281.1 |Severe acute respiratory syndrome coro ···················· ···················· ···················· ···················· ···················· ····················
3683 MW288282.1 |Severe acute respiratory syndrome coro ···················· ···················· ···················· ···················· ···················· ····················
3684 MW288283.1 |Severe acute respiratory syndrome coro ···················· ···················· ···················· ···················· ·····R·············· ····················
3685 MW288284.1 |Severe acute respiratory syndrome coro ···················· ···················· ···················· ···················· ···················· ····················
3686 MW288285.1 |Severe acute respiratory syndrome coro ···················· ···················· ···················· ···················· ···················· ········N···········
3687 MW286518.1 |Severe acute respiratory syndrome coro ···················· ···················· ···················· ···················· ···················· ····················
3688 MW286523.1 |Severe acute respiratory syndrome coro ···················· ···················· ···················· ···················· ···················· ····················
3689 MW286525.1 |Severe acute respiratory syndrome coro ···················· ···················· ···················· ···················· ···················· ····················
3690 MW286533.1 |Severe acute respiratory syndrome coro ···················· ···················· ···················· ···················· ···················· ····················
3691 MW286535.1 |Severe acute respiratory syndrome coro ···················· ···················· ···················· ···················· ···················· ····················
3692 MW286539.1 |Severe acute respiratory syndrome coro ···················· ···················· ···················· ···················· ···················· ····················
3693 MW286543.1 |Severe acute respiratory syndrome coro ···················· ···················· ···················· ···················· ···················· ····················
3695 MW286558.1 |Severe acute respiratory syndrome coro ···················· ···················· ···················· ···················· ···················· ····················
3696 MW286559.1 |Severe acute respiratory syndrome coro ···················· ···················· ···················· ···················· ···················· ····················
3697 MW286563.1 |Severe acute respiratory syndrome coro ···················· ···················· ···················· ···················· ···················· ····················
3698 MW286564.1 |Severe acute respiratory syndrome coro ···················· ···················· ···················· ···················· ···················· ····················
3699 MW286578.1 |Severe acute respiratory syndrome coro ···················· ···················· ···················· ···················· ···················· ····················
3700 MW286579.1 |Severe acute respiratory syndrome coro ···················· ···················· ···················· ···················· ···················· ····················
3702 MW286581.1 |Severe acute respiratory syndrome coro ···················· ···················· ···················· ···················· ···················· ····················
3703 MW286582.1 |Severe acute respiratory syndrome coro ···················· ···················· ···················· ···················· ···················· ····················
3704 MW286583.1 |Severe acute respiratory syndrome coro ···················· ···················· ···················· ···················· ···················· ····················
3705 MW286584.1 |Severe acute respiratory syndrome coro ···················· ···················· ···················· ···················· ···················· ····················
3706 MW286585.1 |Severe acute respiratory syndrome coro ···················· ···················· ···················· ···················· ···················· ····················
3707 MW286590.1 |Severe acute respiratory syndrome coro ···················· ···················· ···················· ···················· ···················· ····················
3708 MW286592.1 |Severe acute respiratory syndrome coro ···················· ···················· ···················· ···················· ···················· ····················
3709 MW286593.1 |Severe acute respiratory syndrome coro ···················· ···················· ···················· ···················· ···················· ····················
3710 MW286594.1 |Severe acute respiratory syndrome coro ···················· ···················· ···················· ···················· ···················· ····················
3711 MW286595.1 |Severe acute respiratory syndrome coro ···················· ···················· ···················· ···················· ···················· ····················
3712 MW286597.1 |Severe acute respiratory syndrome coro ···················· ···················· ···················· ···················· ···················· ····················
3713 MW286598.1 |Severe acute respiratory syndrome coro ···················· ···················· ···················· ···················· ···················· ····················
3714 MW286601.1 |Severe acute respiratory syndrome coro ···················· ···················· ···················· ···················· ···················· ····················
3716 MW286607.1 |Severe acute respiratory syndrome coro ···················· ···················· ···················· ···················· ···················· ····················
3717 MW286608.1 |Severe acute respiratory syndrome coro ···················· ···················· ···················· ···················· ···················· ····················
3718 MW286612.1 |Severe acute respiratory syndrome coro ···················· ···················· ···················· ···················· ···················· ····················
3719 MW286628.1 |Severe acute respiratory syndrome coro ···················· ···················· ···················· ···················· ···················· ····················
3720 MW286629.1 |Severe acute respiratory syndrome coro ···················· ···················· ···················· ···················· ···················· ····················
3721 MW286633.1 |Severe acute respiratory syndrome coro ···················· ···················· ···················· ···················· ···················· ····················
3722 MW286634.1 |Severe acute respiratory syndrome coro ···················· ···················· ···················· ···················· ···················· ····················
3723 MW286635.1 |Severe acute respiratory syndrome coro ···················· ···················· ···················· ···················· ···················· ····················
3724 MW286636.1 |Severe acute respiratory syndrome coro ···················· ···················· ···················· ···················· ···················· ····················
3725 MW286637.1 |Severe acute respiratory syndrome coro ···················· ···················· ···················· ···················· ···················· ····················
3726 MW286638.1 |Severe acute respiratory syndrome coro ···················· ···················· ···················· ···················· ···················· ····················
3727 MW286639.1 |Severe acute respiratory syndrome coro ···················· ···················· ···················· ···················· ···················· ····················
3728 MW286642.1 |Severe acute respiratory syndrome coro ···················· ···················· ···················· ···················· ···················· ····················
3729 MW286644.1 |Severe acute respiratory syndrome coro ···················· ···················· ···················· ···················· ···················· ····················
3730 MW286647.1 |Severe acute respiratory syndrome coro ···················· ···················· ···················· ···················· ···················· ····················
3731 MW286649.1 |Severe acute respiratory syndrome coro ···················· ···················· ···················· ···················· ···················· ····················
```

```
3732 MW286650.1 |Severe acute respiratory syndrome coro    ....................  ....................  .....................  ....................  ....................  ....................
3733 MW286652.1 |Severe acute respiratory syndrome coro    ....................  ....................  .....................  ....................  ....................  ....................
3734 MW286654.1 |Severe acute respiratory syndrome coro    ....................  ....................  .....................  ....................  ....................  ....................
3735 MW286655.1 |Severe acute respiratory syndrome coro    ....................  ....................  .....................  ....................  ....................  ....................
3736 MW286656.1 |Severe acute respiratory syndrome coro    ....................  ....................  .....................  ....................  ....................  ....................
3737 MW286661.1 |Severe acute respiratory syndrome coro    ....................  ....................  .....................  ....................  ....................  ....................
3738 MW286665.1 |Severe acute respiratory syndrome coro    ....................  ....................  .....................  ....................  ....................  ....................
3739 MW286668.1 |Severe acute respiratory syndrome coro    ....................  ....................  .....................  ....................  ....................  ....................
3740 MW286669.1 |Severe acute respiratory syndrome coro    ....................  ....................  .....................  ....................  ....................  ....................
3741 MW286670.1 |Severe acute respiratory syndrome coro    ....................  ....................  .....................  ....................  ....................  ....................
3742 MW286671.1 |Severe acute respiratory syndrome coro    ....................  ....................  .....................  ....................  ....................  ....................
3743 MW286676.1 |Severe acute respiratory syndrome coro    ....................  ....................  .....................  ....................  ....................  ....................
3744 MW286677.1 |Severe acute respiratory syndrome coro    ....................  ....................  .....................  ....................  ....................  ....................
3745 MW286678.1 |Severe acute respiratory syndrome coro    ....................  ....................  .....................  ....................  ....................  ....................
3746 MW286680.1 |Severe acute respiratory syndrome coro    ....................  ....................  .....................  ....................  ....................  ....................
3747 MW286682.1 |Severe acute respiratory syndrome coro    ....................  ....................  .....................  ....................  ....................  ....................
3748 MW286686.1 |Severe acute respiratory syndrome coro    ....................  ....................  .....................  ....................  ....................  ....................
3749 MW286690.1 |Severe acute respiratory syndrome coro    ....................  ....................  .....................  ....................  ....................  ....................
3750 MW286698.1 |Severe acute respiratory syndrome coro    ....................  ....................  .....................  ....................  ....................  ....................
3751 MW286700.1 |Severe acute respiratory syndrome coro    ....................  ....................  .....................  ....................  ....................  ....................
3752 MW286702.1 |Severe acute respiratory syndrome coro    ....................  ....................  .....................  ....................  ....................  ....................
3753 MW286705.1 |Severe acute respiratory syndrome coro    ....................  ....................  .....................  ....................  ....................  ....................
3754 MW286708.1 |Severe acute respiratory syndrome coro    ....................  ....................  .....................  ....................  ....................  ....................
3755 MW286712.1 |Severe acute respiratory syndrome coro    ....................  ....................  .....................  ....................  ....................  ....................
3756 MW286714.1 |Severe acute respiratory syndrome coro    ....................  ....................  .....................  ....................  ....................  ....................
3757 MW286715.1 |Severe acute respiratory syndrome coro    ....................  ....................  .....................  ....................  ....................  ....................
3758 MW286716.1 |Severe acute respiratory syndrome coro    ....................  ....................  .....................  ....................  ....................  ....................
3759 MW286717.1 |Severe acute respiratory syndrome coro    ....................  ....................  .....................  ....................  ....................  ....................
3760 MW286718.1 |Severe acute respiratory syndrome coro    ....................  ....................  .....................  ....................  ....................  ....................
3761 MW286719.1 |Severe acute respiratory syndrome coro    ....................  ....................  .....................  ....................  ....................  ....................
3762 MW286720.1 |Severe acute respiratory syndrome coro    ....................  ....................  .....................  ....................  ....................  ....................
3763 MW286721.1 |Severe acute respiratory syndrome coro    ....................  ....................  .....................  ....................  ....................  ....................
3764 MW286722.1 |Severe acute respiratory syndrome coro    ....................  ....................  .....................  ....................  ....................  ....................
3765 MW286725.1 |Severe acute respiratory syndrome coro    ....................  ....................  .....................  ....................  ....................  ....................
3766 MW286726.1 |Severe acute respiratory syndrome coro    ....................  ....................  .....................  ....................  ....................  ....................
3767 MW286727.1 |Severe acute respiratory syndrome coro    ....................  ....................  .....................  ....................  ....................  ....................
3768 MW286728.1 |Severe acute respiratory syndrome coro    ....................  ....................  .....................  ....................  ....................  ....................
3769 MW286729.1 |Severe acute respiratory syndrome coro    ....................  ....................  .....................  ....................  ....................  ....................
3770 MW286730.1 |Severe acute respiratory syndrome coro    ....................  ....................  .....................  ....................  ....................  ....................
3771 MW286732.1 |Severe acute respiratory syndrome coro    ....................  ....................  .....................  ....................  ....................  ....................
3772 MW286733.1 |Severe acute respiratory syndrome coro    ....................  ....................  .....................  ....................  ....................  ....................
3773 MW286734.1 |Severe acute respiratory syndrome coro    ....................  ....................  .....................  ....................  ....................  ....................
3774 MW286735.1 |Severe acute respiratory syndrome coro    ....................  ....................  .....................  ....................  ....................  ....................
3775 MW286737.1 |Severe acute respiratory syndrome coro    ....................  ....................  .....................  ....................  ....................  ....................
3776 MW286739.1 |Severe acute respiratory syndrome coro    ....................  ....................  .....................  ....................  ....................  ....................
3777 MW286740.1 |Severe acute respiratory syndrome coro    ....................  ....................  .....................  ....................  ....................  ....................
3778 MW286741.1 |Severe acute respiratory syndrome coro    ....................  ....................  .....................  ....................  ....................  ....................
3779 MW286742.1 |Severe acute respiratory syndrome coro    ....................  ....................  .....................  ....................  ....................  ....................
3780 MW286743.1 |Severe acute respiratory syndrome coro    ....................  ....................  .....................  ....................  ....................  ....................
3781 MW286745.1 |Severe acute respiratory syndrome coro    ....................  ....................  .....................  ....................  ....................  ....................
3782 MW286746.1 |Severe acute respiratory syndrome coro    ....................  ....................  .....................  ....................  ....................  ....................
3783 MW286749.1 |Severe acute respiratory syndrome coro    ....................  ....................  .....................  ....................  ....................  ....................
3784 MW286751.1 |Severe acute respiratory syndrome coro    ....................  ....................  .....................  ....................  ....................  ....................
3785 MW286752.1 |Severe acute respiratory syndrome coro    ....................  ....................  .....................  ....................  ....................  ....................
3786 MW286755.1 |Severe acute respiratory syndrome coro    ....................  ....................  .....................  ....................  ....................  ....................
3787 MW286757.1 |Severe acute respiratory syndrome coro    ....................  ....................  .....................  ....................  ....................  ....................
3788 MW286759.1 |Severe acute respiratory syndrome coro    ....................  ....................  .....................  ....................  ....................  ....................
3789 MW286760.1 |Severe acute respiratory syndrome coro    ....................  ....................  .....................  ....................  ....................  ....................
3790 MW286761.1 |Severe acute respiratory syndrome coro    ....................  ....................  .....................  ....................  ....................  ....................
3791 MW281864.1 |Severe acute respiratory syndrome coro    ....................  ....................  .....................  ....................  ....................  ....................
3792 LC593801.1 |Severe acute respiratory syndrome coro    ....................  ....................  .....................  ....................  ....................  ....................
3793 LC593802.1 |Severe acute respiratory syndrome coro    ....................  ....................  .....................  ....................  ....................  ....................
3794 LC593803.1 |Severe acute respiratory syndrome coro    ....................  ....................  .....................  ....................  ....................  ....................
3795 LC593804.1 |Severe acute respiratory syndrome coro    ....................  ....................  .....................  ....................  ....................  ....................
3796 LC593805.1 |Severe acute respiratory syndrome coro    ....................  ....................  .....................  ....................  ....................  ....................
3797 LC593806.1 |Severe acute respiratory syndrome coro    ....................  ....................  .....................  ....................  ....................  ....................
3798 LC593807.1 |Severe acute respiratory syndrome coro    ....................  ....................  .....................  ....................  ....................  ....................
3799 LC593808.1 |Severe acute respiratory syndrome coro    ....................  ....................  .....................  ....................  ....................  ....................
3800 LC593809.1 |Severe acute respiratory syndrome coro    ....................  ....................  .....................  ....................  ....................  ....................
3801 LC593810.1 |Severe acute respiratory syndrome coro    ....................  ....................  .....................  ....................  ....................  ....................
3802 LC593811.1 |Severe acute respiratory syndrome coro    ....................  ....................  .....................  ....................  ....................  ....................
3803 LC593812.1 |Severe acute respiratory syndrome coro    ....................  ....................  .....................  ....................  ....................  ....................
3804 LC593813.1 |Severe acute respiratory syndrome coro    ....................  ....................  .....................  ....................  ....................  ....................
3805 LC593814.1 |Severe acute respiratory syndrome coro    ....................  ....................  .....................  ....................  ....................  ....................
3806 LC593815.1 |Severe acute respiratory syndrome coro    ....................  ....................  .....................  ....................  ....................  ....................
3807 LC593816.1 |Severe acute respiratory syndrome coro    ....................  ....................  .....................  ....................  ....................  ....................
3808 LC593817.1 |Severe acute respiratory syndrome coro    ....................  ....................  .....................  ....................  ....................  ....................
3809 LC593818.1 |Severe acute respiratory syndrome coro    ....................  ....................  .....................  ....................  ....................  ....................
```

3810 LC593819.1 |Severe acute respiratory syndrome coro ···················· ···················· ···················· ···················· ···················· ····················
3811 MW276159.1 |Severe acute respiratory syndrome coro ···················· ·A·················· ···················· ···················· ···················· ····················
3812 MW276160.1 |Severe acute respiratory syndrome coro ···················· ·A·················· ···················· ···················· ···················· ····················
3813 MW276161.1 |Severe acute respiratory syndrome coro ···················· ·A·················· ···················· ···················· ···················· ····················
3814 MW276163.1 |Severe acute respiratory syndrome coro ···················· ·A·················· ···················· ···················· ···················· ····················
3815 MW276166.1 |Severe acute respiratory syndrome coro ···················· ·A·················· ···················· ···················· ···················· ····················
3816 MW276167.1 |Severe acute respiratory syndrome coro ···················· ·A·················· ···················· ···················· ···················· ····················
3817 MW276169.1 |Severe acute respiratory syndrome coro ···················· ·A·················· ···················· ···················· ···················· ····················
3818 MW276170.1 |Severe acute respiratory syndrome coro ···················· ·A·················· ···················· ···················· ···················· ····················
3819 MW276172.1 |Severe acute respiratory syndrome coro ···················· ·A·················· ···················· ···················· ···················· ····················
3820 MW276173.1 |Severe acute respiratory syndrome coro ···················· ·A·················· ···················· ···················· ···················· ····················
3821 MW276174.1 |Severe acute respiratory syndrome coro ···················· ·A·················· ···················· ···················· ···················· ····················
3822 MW276175.1 |Severe acute respiratory syndrome coro ···················· ·A·················· ···················· ···················· N··················· ····················
3823 MW276178.1 |Severe acute respiratory syndrome coro ···················· ·A·················· ···················· ···················· ···················· ····················
3824 MW276182.1 |Severe acute respiratory syndrome coro ···················· ·A·················· ···················· ···················· ···················· ····················
3825 MW276184.1 |Severe acute respiratory syndrome coro ···················· ·A·················· ···················· ···················· ···················· ····················
3826 MW276185.1 |Severe acute respiratory syndrome coro ···················· ·A·················· ···················· ···················· ···················· ····················
3827 MW276189.1 |Severe acute respiratory syndrome coro ···················· ···················· ···················· ···················· ···················· ····················
3828 MW276190.1 |Severe acute respiratory syndrome coro ···················· ···················· ···················· ···················· ···················· ····················
3829 MW276191.1 |Severe acute respiratory syndrome coro ···················· ···················· ···················· ···················· ···················· ····················
3830 MW276192.1 |Severe acute respiratory syndrome coro ···················· ·A·················· ···················· ···················· ···················· ····················
3831 MW276193.1 |Severe acute respiratory syndrome coro ···················· ···················· ···················· ···················· ···················· ····················
3832 MW276194.1 |Severe acute respiratory syndrome coro ···················· ·A·················· ···················· ···················· ···················· ····················
3833 MW276196.1 |Severe acute respiratory syndrome coro ···················· ·A·················· ···················· ···················· ···················· ····················
3834 MW276197.1 |Severe acute respiratory syndrome coro ···················· ·A·················· ···················· ···················· ···················· ····················
3835 MW276198.1 |Severe acute respiratory syndrome coro ···················· ···················· ···················· ···················· ···················· ····················
3836 MW276199.1 |Severe acute respiratory syndrome coro ···················· ·A·················· ···················· ···················· ···················· ····················
3837 MW276201.1 |Severe acute respiratory syndrome coro ···················· ·A·················· ···················· ···················· ···················· ····················
3838 MW276202.1 |Severe acute respiratory syndrome coro ···················· ·A·················· ···················· ···················· ···················· ····················
3839 MW276203.1 |Severe acute respiratory syndrome coro ···················· ·A·················· ···················· ···················· ···················· ····················
3840 MW276208.1 |Severe acute respiratory syndrome coro ···················· ·A·················· ···················· ···················· ···················· ····················
3841 MW276211.1 |Severe acute respiratory syndrome coro ···················· ···················· ···················· ···················· ···················· ····················
3842 MW276213.1 |Severe acute respiratory syndrome coro ···················· ···················· ···················· ···················· ···················· ····················
3843 MW276215.1 |Severe acute respiratory syndrome coro ···················· ···················· ···················· ···················· ···················· ····················
3844 MW276217.1 |Severe acute respiratory syndrome coro ···················· ···················· ···················· ···················· ···················· ····················
3845 MW276218.1 |Severe acute respiratory syndrome coro ···················· ···················· ···················· ···················· ···················· ····················
3846 MW276219.1 |Severe acute respiratory syndrome coro ···················· ···················· ···················· ···················· ···················· ····················
3847 MW276222.1 |Severe acute respiratory syndrome coro ···················· ·A·················· ···················· ···················· ···················· ····················
3848 MW276226.1 |Severe acute respiratory syndrome coro ···················· ···················· ···················· ···················· ···················· ····················
3849 MW276227.1 |Severe acute respiratory syndrome coro ···················· ···················· ···················· ···················· ···················· ····················
3850 MW276228.1 |Severe acute respiratory syndrome coro ···················· ·A·················· ···················· ···················· ···················· ····················
3851 MW276229.1 |Severe acute respiratory syndrome coro ···················· ···················· ···················· ···················· ···················· ····················
3852 MW276230.1 |Severe acute respiratory syndrome coro ···················· ···················· ···················· ···················· ···················· ····················
3853 MW276231.1 |Severe acute respiratory syndrome coro ···················· ···················· ···················· ···················· ···················· ····················
3854 MW276232.1 |Severe acute respiratory syndrome coro ···················· ·A·················· ···················· ···················· ···················· ····················
3855 MW276240.1 |Severe acute respiratory syndrome coro ···················· ···················· ···················· ···················· ···················· ····················
3856 MW276241.1 |Severe acute respiratory syndrome coro ···················· ·A·················· ···················· ···················· ···················· ····················
3857 MW276242.1 |Severe acute respiratory syndrome coro ···················· ···················· ···················· ···················· ···················· ····················
3858 MW276243.1 |Severe acute respiratory syndrome coro ···················· ···················· ···················· ···················· ···················· ····················
3859 MW276244.1 |Severe acute respiratory syndrome coro ···················· ·A·················· ···················· ···················· ···················· ····················
3860 MW276245.1 |Severe acute respiratory syndrome coro ···················· ···················· ···················· ···················· ···················· ····················
3861 MW276247.1 |Severe acute respiratory syndrome coro ···················· ···················· ···················· ···················· ···················· ····················
3862 MW276248.1 |Severe acute respiratory syndrome coro ···················· ·A·················· ···················· ···················· ···················· ····················
3863 MW276249.1 |Severe acute respiratory syndrome coro ···················· ·A·················· ···················· ···················· ···················· ····················
3864 MW276250.1 |Severe acute respiratory syndrome coro ···················· ·A·················· ···················· ···················· ···················· ····················
3865 MW276251.1 |Severe acute respiratory syndrome coro ···················· ···················· ···················· ···················· ···················· ····················
3866 MW276252.1 |Severe acute respiratory syndrome coro ···················· ···················· ···················· ···················· ···················· ····················
3867 MW276253.1 |Severe acute respiratory syndrome coro ···················· ···················· ···················· ···················· ···················· ····················
3869 MW276255.1 |Severe acute respiratory syndrome coro ···················· ···················· ···················· ···················· ···················· ····················
3870 MW276256.1 |Severe acute respiratory syndrome coro ···················· ···················· ···················· ···················· ···················· ····················
3871 MW276257.1 |Severe acute respiratory syndrome coro ···················· ···················· ···················· ···················· ···················· ····················
3872 MW276258.1 |Severe acute respiratory syndrome coro ···················· ···················· ···················· ···················· ···················· ····················
3873 MW276259.1 |Severe acute respiratory syndrome coro ···················· ···················· ···················· ···················· ···················· ····················
3874 MW276260.1 |Severe acute respiratory syndrome coro ···················· ···················· ···················· ···················· ···················· ····················
3875 MW276261.1 |Severe acute respiratory syndrome coro ···················· ···················· ···················· ···················· ···················· ····················
3876 MW276263.1 |Severe acute respiratory syndrome coro ···················· ·A·················· ···················· ···················· ···················· ····················
3877 MW276265.1 |Severe acute respiratory syndrome coro ···················· ···················· ···················· ···················· ···················· ····················
3878 MW276268.1 |Severe acute respiratory syndrome coro ···················· ···················· ···················· ···················· ···················· ····················
3879 MW276269.1 |Severe acute respiratory syndrome coro ···················· ···················· ···················· ···················· ···················· ····················
3880 MW276270.1 |Severe acute respiratory syndrome coro ···················· ···················· ···················· ···················· ···················· ····················
3881 MW276272.1 |Severe acute respiratory syndrome coro ···················· ···················· ···················· ···················· ···················· ····················
3882 MW276278.1 |Severe acute respiratory syndrome coro ···················· ·A·················· ···················· ···················· ···················· ····················
3883 MW276279.1 |Severe acute respiratory syndrome coro ···················· ···················· ···················· ···················· ···················· ····················
3884 MW276283.1 |Severe acute respiratory syndrome coro ···················· ·A·················· ···················· ···················· ···················· ····················
3885 MW276286.1 |Severe acute respiratory syndrome coro ···················· ·A·················· ···················· ···················· ···················· ····················
3886 MW276287.1 |Severe acute respiratory syndrome coro ···················· ···················· ···················· ···················· ···················· ····················
3887 MW276288.1 |Severe acute respiratory syndrome coro ···················· ···················· ···················· ···················· ···················· ····················
3888 MW276289.1 |Severe acute respiratory syndrome coro ···················· ·A·················· ···················· ···················· ···················· ····················

| | | | | | | | | |
|---|---|---|---|---|---|---|---|---|
| 3889 | MW276290.1 | \|Severe acute respiratory syndrome coro | ···················· | ·A·················· | ···················· | ···················· | ···················· | ···················· |
| 3890 | MW276292.1 | \|Severe acute respiratory syndrome coro | ···················· | ···················· | ···················· | ···················· | ···················· | ···················· |
| 3891 | MW276293.1 | \|Severe acute respiratory syndrome coro | ···················· | ···················· | ···················· | ···················· | ···················· | ···················· |
| 3892 | MW276294.1 | \|Severe acute respiratory syndrome coro | ···················· | ·A·················· | ···················· | ···················· | ···················· | ···················· |
| 3893 | MW276295.1 | \|Severe acute respiratory syndrome coro | ···················· | ·A·················· | ···················· | ···················· | ···················· | ···················· |
| 3894 | MW276296.1 | \|Severe acute respiratory syndrome coro | ···················· | ·A·················· | ···················· | ···················· | ···················· | ···················· |
| 3895 | MW276297.1 | \|Severe acute respiratory syndrome coro | ···················· | ·A·················· | ···················· | ···················· | ···················· | ···················· |
| 3896 | MW276298.1 | \|Severe acute respiratory syndrome coro | ···················· | ·A·················· | ···················· | ···················· | ···················· | ···················· |
| 3897 | MW276299.1 | \|Severe acute respiratory syndrome coro | ···················· | ···················· | ···················· | ···················· | ···················· | ···················· |
| 3898 | MW276300.1 | \|Severe acute respiratory syndrome coro | ···················· | ···················· | ···················· | ···················· | ···················· | ···················· |
| 3899 | MW276301.1 | \|Severe acute respiratory syndrome coro | ···················· | ···················· | ···················· | ···················· | ···················· | ···················· |
| 3900 | MW276302.1 | \|Severe acute respiratory syndrome coro | ···················· | ·A·················· | ···················· | ···················· | ···················· | ···················· |
| 3901 | MW276303.1 | \|Severe acute respiratory syndrome coro | ···················· | ·A·················· | ···················· | ···················· | ···················· | ···················· |
| 3902 | MW276304.1 | \|Severe acute respiratory syndrome coro | ···················· | ·A·················· | ···················· | ···················· | ···················· | ···················· |
| 3903 | MW276305.1 | \|Severe acute respiratory syndrome coro | ···················· | ·A·················· | ···················· | ···················· | ···················· | ···················· |
| 3904 | MW276306.1 | \|Severe acute respiratory syndrome coro | ···················· | ·A·················· | ···················· | ···················· | ···················· | ···················· |
| 3905 | MW276307.1 | \|Severe acute respiratory syndrome coro | ···················· | ·A·················· | ···················· | ···················· | ···················· | ···················· |
| 3906 | MW276308.1 | \|Severe acute respiratory syndrome coro | ···················· | ·A·················· | ···················· | ···················· | ···················· | ···················· |
| 3907 | MW276309.1 | \|Severe acute respiratory syndrome coro | ···················· | ·A·················· | ···················· | ···················· | ···················· | ···················· |
| 3908 | MW276310.1 | \|Severe acute respiratory syndrome coro | ···················· | ···················· | ···················· | ···················· | ···················· | ···················· |
| 3909 | MW276311.1 | \|Severe acute respiratory syndrome coro | ···················· | ·A·················· | ···················· | ···················· | ···················· | ···················· |
| 3910 | MW276312.1 | \|Severe acute respiratory syndrome coro | ···················· | ···················· | ···················· | ···················· | ···················· | ···················· |
| 3911 | MW276313.1 | \|Severe acute respiratory syndrome coro | ···················· | ·A·················· | ···················· | ···················· | ···················· | ···················· |
| 3912 | MW276314.1 | \|Severe acute respiratory syndrome coro | ···················· | ···················· | ···················· | ···················· | ···················· | ···················· |
| 3913 | MW276315.1 | \|Severe acute respiratory syndrome coro | ···················· | ·A·················· | ···················· | ···················· | ···················· | ···················· |
| 3914 | MW276317.1 | \|Severe acute respiratory syndrome coro | ···················· | ·A·················· | ···················· | ···················· | ···················· | ···················· |
| 3915 | MW276318.1 | \|Severe acute respiratory syndrome coro | ···················· | ·A·················· | ···················· | ···················· | ···················· | ···················· |
| 3916 | MW276319.1 | \|Severe acute respiratory syndrome coro | ···················· | ·A·················· | ···················· | ···················· | ···················· | ···················· |
| 3917 | MW276320.1 | \|Severe acute respiratory syndrome coro | ···················· | ···················· | ···················· | ···················· | ···················· | ···················· |
| 3918 | MW276321.1 | \|Severe acute respiratory syndrome coro | ···················· | ·A·················· | ···················· | ···················· | ···················· | ···················· |
| 3919 | MW276323.1 | \|Severe acute respiratory syndrome coro | ···················· | ···················· | ···················· | ···················· | ···················· | ···················· |
| 3920 | MW276324.1 | \|Severe acute respiratory syndrome coro | ···················· | ···················· | ···················· | ···················· | ···················· | ···················· |
| 3921 | MW276325.1 | \|Severe acute respiratory syndrome coro | ···················· | ···················· | ···················· | ···················· | ···················· | ···················· |
| 3922 | MW276326.1 | \|Severe acute respiratory syndrome coro | ···················· | ···················· | ···················· | ···················· | ···················· | ···················· |
| 3923 | MW276328.1 | \|Severe acute respiratory syndrome coro | ···················· | ···················· | ···················· | ···················· | ···················· | ···················· |
| 3924 | MW276330.1 | \|Severe acute respiratory syndrome coro | ···················· | ···················· | ···················· | ···················· | ···················· | ···················· |
| 3925 | MW276331.1 | \|Severe acute respiratory syndrome coro | ···················· | ···················· | ···················· | ···················· | ···················· | ···················· |
| 3926 | MW276332.1 | \|Severe acute respiratory syndrome coro | ···················· | ···················· | ···················· | ···················· | ···················· | ···················· |
| 3927 | MW276333.1 | \|Severe acute respiratory syndrome coro | ···················· | ···················· | ···················· | ···················· | ···················· | ···················· |
| 3928 | MW276334.1 | \|Severe acute respiratory syndrome coro | ···················· | ···················· | ···················· | ···················· | ···················· | ···················· |
| 3929 | MW276336.1 | \|Severe acute respiratory syndrome coro | ···················· | ···················· | ···················· | ···················· | ···················· | ···················· |
| 3930 | MW276337.1 | \|Severe acute respiratory syndrome coro | ···················· | ···················· | ···················· | ···················· | ···················· | ···················· |
| 3931 | MW276338.1 | \|Severe acute respiratory syndrome coro | ···················· | ···················· | ···················· | ···················· | ···················· | ···················· |
| 3932 | MW276339.1 | \|Severe acute respiratory syndrome coro | ···················· | ···················· | ···················· | ···················· | ···················· | ···················· |
| 3933 | MW276340.1 | \|Severe acute respiratory syndrome coro | ···················· | ···················· | ···················· | ···················· | ···················· | ···················· |
| 3934 | MW276341.1 | \|Severe acute respiratory syndrome coro | ···················· | ···················· | ···················· | ···················· | ···················· | ···················· |
| 3935 | MW276342.1 | \|Severe acute respiratory syndrome coro | ···················· | ···················· | ···················· | ···················· | ···················· | ···················· |
| 3936 | MW276343.1 | \|Severe acute respiratory syndrome coro | ···················· | ···················· | ···················· | ···················· | ···················· | ···················· |
| 3937 | MW276345.1 | \|Severe acute respiratory syndrome coro | ···················· | ···················· | ···················· | ···················· | ···················· | ···················· |
| 3938 | MW276346.1 | \|Severe acute respiratory syndrome coro | ···················· | ···················· | ···················· | ···················· | ···················· | ···················· |
| 3939 | MW276347.1 | \|Severe acute respiratory syndrome coro | ···················· | ·A·················· | ···················· | ···················· | ···················· | ···················· |
| 3940 | MW276348.1 | \|Severe acute respiratory syndrome coro | ···················· | ·A·················· | ···················· | ···················· | ···················· | ···················· |
| 3941 | MW276349.1 | \|Severe acute respiratory syndrome coro | ···················· | ·A·················· | ···················· | ···················· | ···················· | ···················· |
| 3942 | MW276350.1 | \|Severe acute respiratory syndrome coro | ···················· | ·A·················· | ···················· | ···················· | ···················· | ···················· |
| 3943 | MW276351.1 | \|Severe acute respiratory syndrome coro | ···················· | ·A·················· | ···················· | ···················· | ···················· | ···················· |
| 3944 | MW276352.1 | \|Severe acute respiratory syndrome coro | ···················· | ·A·················· | ···················· | ···················· | ···················· | ···················· |
| 3945 | MW276353.1 | \|Severe acute respiratory syndrome coro | ···················· | ···················· | ···················· | ···················· | ···················· | ···················· |
| 3946 | MW276356.1 | \|Severe acute respiratory syndrome coro | ···················· | ···················· | ···················· | ···················· | ···················· | ···················· |
| 3947 | MW276357.1 | \|Severe acute respiratory syndrome coro | ···················· | ···················· | ···················· | ···················· | ···················· | ···················· |
| 3948 | MW276358.1 | \|Severe acute respiratory syndrome coro | ···················· | ···················· | ···················· | ···················· | ···················· | ···················· |
| 3949 | MW276359.1 | \|Severe acute respiratory syndrome coro | ···················· | ···················· | ···················· | ···················· | ···················· | ···················· |
| 3951 | MW276361.1 | \|Severe acute respiratory syndrome coro | ···················· | ···················· | ···················· | ···················· | ···················· | ···················· |
| 3952 | MW276362.1 | \|Severe acute respiratory syndrome coro | ···················· | ·A·················· | ···················· | ···················· | ···················· | ···················· |
| 3953 | MW276363.1 | \|Severe acute respiratory syndrome coro | ···················· | ···················· | ···················· | ···················· | ···················· | ···················· |
| 3954 | MW276364.1 | \|Severe acute respiratory syndrome coro | ···················· | ···················· | ···················· | ···················· | ···················· | ···················· |
| 3955 | MW276365.1 | \|Severe acute respiratory syndrome coro | ···················· | ···················· | ···················· | ···················· | ···················· | ···················· |
| 3956 | MW276366.1 | \|Severe acute respiratory syndrome coro | ···················· | ···················· | ···················· | ···················· | ···················· | ···················· |
| 3957 | MW276367.1 | \|Severe acute respiratory syndrome coro | ···················· | ·A·················· | ···················· | ···················· | ···················· | ···················· |
| 3958 | MW276368.1 | \|Severe acute respiratory syndrome coro | ···················· | ···················· | ···················· | ···················· | ···················· | ···················· |
| 3959 | MW276369.1 | \|Severe acute respiratory syndrome coro | ···················· | ·A·················· | ···················· | ···················· | ···················· | ···················· |
| 3960 | MW276370.1 | \|Severe acute respiratory syndrome coro | ···················· | ···················· | ···················· | ···················· | ···················· | ···················· |
| 3961 | MW276371.1 | \|Severe acute respiratory syndrome coro | ···················· | ···················· | ···················· | ···················· | ···················· | ···················· |
| 3962 | MW276372.1 | \|Severe acute respiratory syndrome coro | ···················· | ···················· | ···················· | ···················· | ···················· | ···················· |
| 3963 | MW276373.1 | \|Severe acute respiratory syndrome coro | ···················· | ···················· | ···················· | ···················· | ···················· | ···················· |
| 3964 | MW276374.1 | \|Severe acute respiratory syndrome coro | ···················· | ·A·················· | ···················· | ···················· | ···················· | ···················· |
| 3965 | MW276375.1 | \|Severe acute respiratory syndrome coro | ···················· | ···················· | ···················· | ···················· | ···················· | ···················· |
| 3966 | MW276376.1 | \|Severe acute respiratory syndrome coro | ···················· | ···················· | ···················· | ···················· | ···················· | ···················· |
| 3967 | MW276377.1 | \|Severe acute respiratory syndrome coro | ···················· | ·A·················· | ···················· | ···················· | ···················· | ···················· |

| | | | | | | | | |
|---|---|---|---|---|---|---|---|---|
| 3968 | MW276380.1 | Severe acute respiratory syndrome coro | ···················· | ·A·················· | ···················· | ···················· | ···················· | ···················· |
| 3969 | MW276381.1 | Severe acute respiratory syndrome coro | ···················· | ·A·················· | ···················· | ···················· | ···················· | ···················· |
| 3970 | MW276382.1 | Severe acute respiratory syndrome coro | ···················· | ·A·················· | ···················· | ···················· | ···················· | ···················· |
| 3971 | MW276383.1 | Severe acute respiratory syndrome coro | ···················· | ·A·················· | ···················· | ···················· | ···················· | ···················· |
| 3972 | MW276384.1 | Severe acute respiratory syndrome coro | ···················· | ·A·················· | ···················· | ···················· | ···················· | ···················· |
| 3975 | MW276387.1 | Severe acute respiratory syndrome coro | ···················· | ···················· | ···················· | ···················· | ···················· | ···················· |
| 3976 | MW276388.1 | Severe acute respiratory syndrome coro | ···················· | ···················· | ···················· | ···················· | ···················· | ···················· |
| 3977 | MW276389.1 | Severe acute respiratory syndrome coro | ···················· | ···················· | ···················· | ···················· | ···················· | ···················· |
| 3978 | MW276390.1 | Severe acute respiratory syndrome coro | ···················· | ···················· | ···················· | ···················· | ···················· | ···················· |
| 3979 | MW276391.1 | Severe acute respiratory syndrome coro | ···················· | ···················· | ···················· | ···················· | ···················· | ···················· |
| 3980 | MW276393.1 | Severe acute respiratory syndrome coro | ···················· | ·A·················· | ···················· | ···················· | ··N················· | ···················· |
| 3981 | MW276394.1 | Severe acute respiratory syndrome coro | ···················· | ·A·················· | ···················· | ···················· | ···················· | ···················· |
| 3982 | MW276395.1 | Severe acute respiratory syndrome coro | ···················· | ·A·················· | ···················· | ···················· | ···················· | ···················· |
| 3983 | MW276396.1 | Severe acute respiratory syndrome coro | ···················· | ···················· | ···················· | ···················· | ···················· | ···················· |
| 3984 | MW276397.1 | Severe acute respiratory syndrome coro | ···················· | ···················· | ···················· | ···················· | ···················· | ···················· |
| 3985 | MW276398.1 | Severe acute respiratory syndrome coro | ···················· | ···················· | ···················· | ···················· | ···················· | ···················· |
| 3986 | MW276399.1 | Severe acute respiratory syndrome coro | ···················· | ···················· | ···················· | ···················· | ···················· | ···················· |
| 3987 | MW276400.1 | Severe acute respiratory syndrome coro | ···················· | ···················· | ···················· | ···················· | ···················· | ···················· |
| 3988 | MW276401.1 | Severe acute respiratory syndrome coro | ···················· | ···················· | ···················· | ···················· | ···················· | ···················· |
| 3989 | MW276402.1 | Severe acute respiratory syndrome coro | ···················· | ·A·················· | ···················· | ···················· | ···················· | ···················· |
| 3990 | MW276403.1 | Severe acute respiratory syndrome coro | ···················· | ·A·················· | ···················· | ···················· | ···················· | ···················· |
| 3991 | MW276404.1 | Severe acute respiratory syndrome coro | ···················· | ·A·················· | ···················· | ···················· | ···················· | ···················· |
| 3992 | MW276406.1 | Severe acute respiratory syndrome coro | ···················· | ···················· | ···················· | ···················· | ···················· | ···················· |
| 3993 | MW276407.1 | Severe acute respiratory syndrome coro | ···················· | ·A·················· | ···················· | ···················· | ···················· | ···················· |
| 3994 | MW276408.1 | Severe acute respiratory syndrome coro | ···················· | ···················· | ···················· | ···················· | ···················· | ···················· |
| 3995 | MW276409.1 | Severe acute respiratory syndrome coro | ···················· | ·A·················· | ···················· | ···················· | ···················· | ···················· |
| 3996 | MW276410.1 | Severe acute respiratory syndrome coro | ···················· | ·A·················· | ···················· | ···················· | ···················· | ···················· |
| 3997 | MW276411.1 | Severe acute respiratory syndrome coro | ···················· | ·A·················· | ···················· | ···················· | ···················· | ···················· |
| 3998 | MW276413.1 | Severe acute respiratory syndrome coro | ···················· | ·A·················· | ···················· | ···················· | ···················· | ···················· |
| 3999 | MW276415.1 | Severe acute respiratory syndrome coro | ···················· | ·A·················· | ···················· | ···················· | ···················· | ···················· |
| 4000 | MW276416.1 | Severe acute respiratory syndrome coro | ···················· | ·A·················· | ···················· | ···················· | ···················· | ···················· |
| 4001 | MW276417.1 | Severe acute respiratory syndrome coro | ···················· | ·A·················· | ···················· | ···················· | ···················· | ···················· |
| 4002 | MW276418.1 | Severe acute respiratory syndrome coro | ···················· | ·A·················· | ···················· | ···················· | ···················· | ···················· |
| 4003 | MW276419.1 | Severe acute respiratory syndrome coro | ···················· | ·A·················· | ···················· | ···················· | ···················· | ···················· |
| 4004 | MW276420.1 | Severe acute respiratory syndrome coro | ···················· | ·A·················· | ···················· | ···················· | ···················· | ···················· |
| 4005 | MW276421.1 | Severe acute respiratory syndrome coro | ···················· | ·A·················· | ···················· | ···················· | ···················· | ···················· |
| 4006 | MW276422.1 | Severe acute respiratory syndrome coro | ···················· | ·A·················· | ···················· | ···················· | ···················· | ···················· |
| 4007 | MW276423.1 | Severe acute respiratory syndrome coro | ···················· | ·A·················· | ···················· | ···················· | ···················· | ···················· |
| 4008 | MW276424.1 | Severe acute respiratory syndrome coro | ···················· | ·A·················· | ···················· | ···················· | ···················· | ···················· |
| 4009 | MW276425.1 | Severe acute respiratory syndrome coro | ···················· | ·A·················· | ···················· | ···················· | ···················· | ···················· |
| 4010 | MW276426.1 | Severe acute respiratory syndrome coro | ···················· | ·A·················· | ···················· | ···················· | ···················· | ···················· |
| 4011 | MW276427.1 | Severe acute respiratory syndrome coro | ···················· | ·A·················· | ···················· | ···················· | ···················· | ···················· |
| 4012 | MW276428.1 | Severe acute respiratory syndrome coro | ···················· | ·A·················· | ···················· | ···················· | ···················· | ···················· |
| 4013 | MW276429.1 | Severe acute respiratory syndrome coro | ···················· | ·A·················· | ···················· | ···················· | ···················· | ···················· |
| 4014 | MW276430.1 | Severe acute respiratory syndrome coro | ···················· | ···················· | ···················· | ···················· | ···················· | ···················· |
| 4015 | MW276431.1 | Severe acute respiratory syndrome coro | ···················· | ·A·················· | ···················· | ···················· | ···················· | ···················· |
| 4016 | MW276432.1 | Severe acute respiratory syndrome coro | ···················· | ·A·················· | ···················· | ···················· | ···················· | ···················· |
| 4017 | MW276433.1 | Severe acute respiratory syndrome coro | ···················· | ·A·················· | ···················· | ···················· | ···················· | ···················· |
| 4018 | MW276434.1 | Severe acute respiratory syndrome coro | ···················· | ·A·················· | ···················· | ···················· | ···················· | ···················· |
| 4020 | MW276437.1 | Severe acute respiratory syndrome coro | ···················· | ·A·················· | ···················· | ···················· | ···················· | ···················· |
| 4021 | MW276438.1 | Severe acute respiratory syndrome coro | ···················· | ···················· | ···················· | ···················· | ···················· | ···················· |
| 4022 | MW276440.1 | Severe acute respiratory syndrome coro | ···················· | ·A·················· | ···················· | ···················· | ···················· | ···················· |
| 4023 | MW276441.1 | Severe acute respiratory syndrome coro | ···················· | ·A·················· | ···················· | ···················· | ···················· | ···················· |
| 4024 | MW276442.1 | Severe acute respiratory syndrome coro | ···················· | ·A·················· | ···················· | ···················· | ···················· | ···················· |
| 4025 | MW276443.1 | Severe acute respiratory syndrome coro | ···················· | ·A·················· | ···················· | ···················· | ···················· | ···················· |
| 4026 | MW276444.1 | Severe acute respiratory syndrome coro | ···················· | ·A·················· | ···················· | ···················· | ···················· | ···················· |
| 4027 | MW276445.1 | Severe acute respiratory syndrome coro | ···················· | ·A·················· | ···················· | ···················· | ···················· | ···················· |
| 4028 | MW276446.1 | Severe acute respiratory syndrome coro | ···················· | ·A·················· | ···················· | ···················· | ···················· | ···················· |
| 4029 | MW276447.1 | Severe acute respiratory syndrome coro | ···················· | ·A·················· | ···················· | ···················· | ···················· | ···················· |
| 4030 | MW276448.1 | Severe acute respiratory syndrome coro | ···················· | ···················· | ···················· | ···················· | ···················· | ···················· |
| 4031 | MW276449.1 | Severe acute respiratory syndrome coro | ···················· | ···················· | ···················· | ···················· | ···················· | ···················· |
| 4032 | MW276450.1 | Severe acute respiratory syndrome coro | ···················· | ···················· | ···················· | ···················· | ···················· | ···················· |
| 4033 | MW276451.1 | Severe acute respiratory syndrome coro | ···················· | ···················· | ···················· | ···················· | ···················· | ···················· |
| 4034 | MW276452.1 | Severe acute respiratory syndrome coro | ···················· | ···················· | ···················· | ···················· | ···················· | ···················· |
| 4035 | MW276453.1 | Severe acute respiratory syndrome coro | ···················· | ···················· | ···················· | ···················· | ···················· | ···················· |
| 4036 | MW276454.1 | Severe acute respiratory syndrome coro | ···················· | ···················· | ···················· | ···················· | ···················· | ···················· |
| 4037 | MW276455.1 | Severe acute respiratory syndrome coro | ···················· | ···················· | ···················· | ···················· | ···················· | ···················· |
| 4038 | MW276456.1 | Severe acute respiratory syndrome coro | ···················· | ···················· | ···················· | ···················· | ···················· | ···················· |
| 4039 | MW276458.1 | Severe acute respiratory syndrome coro | ···················· | ···················· | ···················· | ···················· | ···················· | ···················· |
| 4040 | MW276463.1 | Severe acute respiratory syndrome coro | ···················· | ···················· | ···················· | ···················· | ···················· | ···················· |
| 4041 | MW276464.1 | Severe acute respiratory syndrome coro | ···················· | ·A·················· | ···················· | ···················· | ···················· | ···················· |
| 4042 | MW276465.1 | Severe acute respiratory syndrome coro | ···················· | ···················· | ···················· | ···················· | ···················· | ···················· |
| 4043 | MW276466.1 | Severe acute respiratory syndrome coro | ···················· | ···················· | ···················· | ···················· | ···················· | ···················· |
| 4044 | MW276467.1 | Severe acute respiratory syndrome coro | ···················· | ···················· | ···················· | ···················· | ···················· | ···················· |
| 4045 | MW276468.1 | Severe acute respiratory syndrome coro | ···················· | ···················· | ···················· | ···················· | ···················· | ···················· |
| 4046 | MW276469.1 | Severe acute respiratory syndrome coro | ···················· | ···················· | ···················· | ···················· | ···················· | ···················· |
| 4047 | MW276470.1 | Severe acute respiratory syndrome coro | ···················· | ···················· | ···················· | ···················· | ···················· | ···················· |
| 4048 | MW276471.1 | Severe acute respiratory syndrome coro | ···················· | ·A·················· | ···················· | ···················· | ···················· | ···················· |

| | | | | | | | | | |
|---|---|---|---|---|---|---|---|---|---|
| 4049 | MW276473.1 | \|Severe acute respiratory syndrome coro | ···················· | ···················· | ···················· | ···················· | ···················· | ···················· | |
| 4050 | MW276474.1 | \|Severe acute respiratory syndrome coro | ···················· | ···················· | ···················· | ···················· | ···················· | ···················· | |
| 4051 | MW276475.1 | \|Severe acute respiratory syndrome coro | ···················· | ·A·················· | ···················· | ···················· | ···················· | ···················· | |
| 4052 | MW276476.1 | \|Severe acute respiratory syndrome coro | ···················· | ·A·················· | ···················· | ···················· | ···················· | ···················· | |
| 4053 | MW276477.1 | \|Severe acute respiratory syndrome coro | ···················· | ···················· | ···················· | ···················· | ···················· | ···················· | |
| 4054 | MW276478.1 | \|Severe acute respiratory syndrome coro | ···················· | ···················· | ···················· | ···················· | ···················· | ···················· | |
| 4055 | MW276479.1 | \|Severe acute respiratory syndrome coro | ···················· | ···················· | ···················· | ···················· | ···················· | ···················· | |
| 4056 | MW276480.1 | \|Severe acute respiratory syndrome coro | ···················· | ···················· | ···················· | ···················· | ···················· | ···················· | |
| 4057 | MW276481.1 | \|Severe acute respiratory syndrome coro | ···················· | ···················· | ···················· | ···················· | ···················· | ···················· | |
| 4058 | MW276482.1 | \|Severe acute respiratory syndrome coro | ···················· | ···················· | ···················· | ···················· | ···················· | ···················· | |
| 4059 | MW276483.1 | \|Severe acute respiratory syndrome coro | ···················· | ···················· | ···················· | ···················· | ···················· | ···················· | |
| 4060 | MW276484.1 | \|Severe acute respiratory syndrome coro | ···················· | ···················· | ···················· | ···················· | ···················· | ···················· | |
| 4061 | MW276485.1 | \|Severe acute respiratory syndrome coro | ···················· | ·A·················· | ···················· | ···················· | ···················· | ···················· | |
| 4062 | MW276486.1 | \|Severe acute respiratory syndrome coro | ···················· | ···················· | ···················· | ···················· | ·············T······ | ···················· | |
| 4063 | MW276487.1 | \|Severe acute respiratory syndrome coro | ···················· | ···················· | ···················· | ···················· | ···················· | ···················· | |
| 4064 | MW276488.1 | \|Severe acute respiratory syndrome coro | ···················· | ···················· | ···················· | ···················· | ···················· | ···················· | |
| 4065 | MW276489.1 | \|Severe acute respiratory syndrome coro | ···················· | ···················· | ···················· | ···················· | ···················· | ···················· | |
| 4066 | MW276490.1 | \|Severe acute respiratory syndrome coro | ···················· | ·A·················· | ···················· | ···················· | ···················· | ···················· | |
| 4067 | MW276491.1 | \|Severe acute respiratory syndrome coro | ···················· | ·A·················· | ···················· | ···················· | ···················· | ···················· | |
| 4068 | MW276492.1 | \|Severe acute respiratory syndrome coro | ···················· | ·A·················· | ···················· | ···················· | ···················· | ···················· | |
| 4069 | MW276493.1 | \|Severe acute respiratory syndrome coro | ···················· | ···················· | ··T················· | ···················· | ···················· | ···················· | |
| 4070 | MW276494.1 | \|Severe acute respiratory syndrome coro | ···················· | ·A·················· | ···················· | ···················· | ···················· | ···················· | |
| 4071 | MW276495.1 | \|Severe acute respiratory syndrome coro | ···················· | ···················· | ···················· | ···················· | ···················· | ···················· | |
| 4072 | MW276496.1 | \|Severe acute respiratory syndrome coro | ···················· | ···················· | ···················· | ···················· | ···················· | ···················· | |
| 4073 | MW276498.1 | \|Severe acute respiratory syndrome coro | ···················· | ···················· | ···················· | ···················· | ···················· | ···················· | |
| 4074 | MW276499.1 | \|Severe acute respiratory syndrome coro | ···················· | ···················· | ···················· | ···················· | ···················· | ···················· | |
| 4075 | MW276504.1 | \|Severe acute respiratory syndrome coro | ···················· | ···················· | ···················· | ···················· | ···················· | ···················· | |
| 4076 | MW276505.1 | \|Severe acute respiratory syndrome coro | ···················· | ···················· | ···················· | ···················· | ···················· | ···················· | |
| 4077 | MW276506.1 | \|Severe acute respiratory syndrome coro | ···················· | ···················· | ···················· | ···················· | ···················· | ···················· | |
| 4078 | MW276507.1 | \|Severe acute respiratory syndrome coro | ···················· | ···················· | ···················· | ···················· | ···················· | ···················· | |
| 4079 | MW276508.1 | \|Severe acute respiratory syndrome coro | ···················· | ···················· | ···················· | ···················· | ···················· | ···················· | |
| 4080 | MW276509.1 | \|Severe acute respiratory syndrome coro | ···················· | ···················· | ···················· | ···················· | ···················· | ···················· | |
| 4081 | MW276510.1 | \|Severe acute respiratory syndrome coro | ···················· | ···················· | ···················· | ···················· | ···················· | ···················· | |
| 4082 | MW276511.1 | \|Severe acute respiratory syndrome coro | ···················· | ···················· | ···················· | ···················· | ···················· | ···················· | |
| 4083 | MW276512.1 | \|Severe acute respiratory syndrome coro | ···················· | ···················· | ···················· | ···················· | ···················· | ···················· | |
| 4084 | MW276514.1 | \|Severe acute respiratory syndrome coro | ···················· | ···················· | ···················· | ···················· | ···················· | ···················· | |
| 4086 | MW276519.1 | \|Severe acute respiratory syndrome coro | ···················· | ···················· | ···················· | ···················· | ···················· | ···················· | |
| 4088 | MW276521.1 | \|Severe acute respiratory syndrome coro | ···················· | ···················· | ···················· | ···················· | ···················· | ···················· | |
| 4091 | MW276524.1 | \|Severe acute respiratory syndrome coro | ···················· | ···················· | ···················· | ···················· | ···················· | ···················· | |
| 4093 | MW276527.1 | \|Severe acute respiratory syndrome coro | ···················· | ···················· | ···················· | ···················· | ···················· | ···················· | |
| 4094 | MW276528.1 | \|Severe acute respiratory syndrome coro | ···················· | ···················· | ···················· | ···················· | ···················· | ···················· | |
| 4095 | MW276529.1 | \|Severe acute respiratory syndrome coro | ···················· | ···················· | ···················· | ···················· | ···················· | ···················· | |
| 4096 | MW276533.1 | \|Severe acute respiratory syndrome coro | ···················· | ···················· | ···················· | ···················· | ···················· | ···················· | |
| 4097 | MW276536.1 | \|Severe acute respiratory syndrome coro | ···················· | ···················· | ···················· | ···················· | NNN················· | ···················· | 1 |
| 4098 | MW276537.1 | \|Severe acute respiratory syndrome coro | ···················· | ·A·················· | ···················· | ···················· | ···················· | ···················· | |
| 4099 | MW276538.1 | \|Severe acute respiratory syndrome coro | ···················· | ···················· | ···················· | ···················· | ···················· | ···················· | |
| 4100 | MW276539.1 | \|Severe acute respiratory syndrome coro | ···················· | ···················· | ···················· | ···················· | ···················· | ···················· | |
| 4101 | MW276540.1 | \|Severe acute respiratory syndrome coro | ···················· | ···················· | ···················· | ···················· | ···················· | ···················· | |
| 4102 | MW276542.1 | \|Severe acute respiratory syndrome coro | ···················· | ···················· | ···················· | ···················· | ···················· | ···················· | |
| 4103 | MW276543.1 | \|Severe acute respiratory syndrome coro | ···················· | ···················· | ···················· | ···················· | ···················· | ···················· | |
| 4104 | MW276544.1 | \|Severe acute respiratory syndrome coro | ···················· | ···················· | ···················· | ···················· | ···················· | ···················· | |
| 4105 | MW276545.1 | \|Severe acute respiratory syndrome coro | ···················· | ···················· | ···················· | ···················· | ···················· | ···················· | |
| 4106 | MW276546.1 | \|Severe acute respiratory syndrome coro | ···················· | ···················· | ···················· | ···················· | ···················· | ···················· | |
| 4109 | MW276549.1 | \|Severe acute respiratory syndrome coro | ···················· | ·A·················· | ···················· | ···················· | ···················· | ···················· | |
| 4110 | MW276550.1 | \|Severe acute respiratory syndrome coro | ···················· | ·A·················· | ···················· | ···················· | ···················· | ···················· | |
| 4111 | MW276551.1 | \|Severe acute respiratory syndrome coro | ···················· | ···················· | ···················· | ···················· | ···················· | ···················· | |
| 4112 | MW276552.1 | \|Severe acute respiratory syndrome coro | ···················· | ·A·················· | ···················· | ···················· | ···················· | ···················· | |
| 4113 | MW276553.1 | \|Severe acute respiratory syndrome coro | ···················· | ···················· | ···················· | ···················· | ···················· | ···················· | |
| 4114 | MW276554.1 | \|Severe acute respiratory syndrome coro | ···················· | ···················· | ···················· | ···················· | ···················· | ···················· | |
| 4115 | MW276555.1 | \|Severe acute respiratory syndrome coro | ···················· | ···················· | ···················· | ···················· | ···················· | ···················· | |
| 4116 | MW276556.1 | \|Severe acute respiratory syndrome coro | ···················· | ·A·················· | ···················· | ···················· | ···················· | ···················· | |
| 4117 | MW276557.1 | \|Severe acute respiratory syndrome coro | ···················· | ·A·················· | ···················· | ···················· | ···················· | ···················· | |
| 4118 | MW276558.1 | \|Severe acute respiratory syndrome coro | ···················· | ·A·················· | ···················· | ···················· | ···················· | ···················· | |
| 4119 | MW276559.1 | \|Severe acute respiratory syndrome coro | ···················· | ···················· | ···················· | ···················· | ···················· | ···················· | |
| 4120 | MW276560.1 | \|Severe acute respiratory syndrome coro | ···················· | ·A·················· | ···················· | ···················· | ···················· | ···················· | |
| 4121 | MW276561.1 | \|Severe acute respiratory syndrome coro | ···················· | ·A·················· | ···················· | ···················· | ···················· | ···················· | |
| 4122 | MW276562.1 | \|Severe acute respiratory syndrome coro | ···················· | ·A·················· | ···················· | ···················· | ···················· | ···················· | |
| 4123 | MW276563.1 | \|Severe acute respiratory syndrome coro | ···················· | ·A·················· | ···················· | ···················· | ···················· | ···················· | |
| 4124 | MW276564.1 | \|Severe acute respiratory syndrome coro | ···················· | ·A·················· | ···················· | ···················· | ···················· | ···················· | |
| 4126 | MW276566.1 | \|Severe acute respiratory syndrome coro | ···················· | ·A·················· | ···················· | ···················· | ···················· | ···················· | |
| 4127 | MW276567.1 | \|Severe acute respiratory syndrome coro | ···················· | ·A·················· | ···················· | ···················· | ···················· | ···················· | |
| 4128 | MW276568.1 | \|Severe acute respiratory syndrome coro | ···················· | ·A·················· | ···················· | ···················· | ···················· | ···················· | |
| 4129 | MW276569.1 | \|Severe acute respiratory syndrome coro | ···················· | ·A·················· | ···················· | ···················· | ···················· | ···················· | |
| 4130 | MW276570.1 | \|Severe acute respiratory syndrome coro | ···················· | ·A·················· | ···················· | ···················· | ···················· | ···················· | |
| 4131 | MW276571.1 | \|Severe acute respiratory syndrome coro | ···················· | ·A·················· | ···················· | ···················· | ···················· | ···················· | |
| 4132 | MW276572.1 | \|Severe acute respiratory syndrome coro | ···················· | ···················· | ···················· | ···················· | ···················· | ···················· | |
| 4133 | MW276573.1 | \|Severe acute respiratory syndrome coro | ···················· | ···················· | ···················· | ···················· | ···················· | ···················· | |
| 4134 | MW276574.1 | \|Severe acute respiratory syndrome coro | ···················· | ·A·················· | ···················· | ···················· | ···················· | ···················· | |

| | | | | | | | | |
|---|---|---|---|---|---|---|---|---|
| 4135 | MW276575.1 | \|Severe acute respiratory syndrome coro | ···················· | ·A·················· | ···················· | ···················· | ···················· | ···················· |
| 4136 | MW276577.1 | \|Severe acute respiratory syndrome coro | ···················· | ···················· | ···················· | ···················· | ···················· | ···················· |
| 4137 | MW276578.1 | \|Severe acute respiratory syndrome coro | ···················· | ···················· | ···················· | ···················· | ···················· | ···················· |
| 4138 | MW276579.1 | \|Severe acute respiratory syndrome coro | ···················· | ···················· | ···················· | ···················· | ···················· | ···················· |
| 4139 | MW276581.1 | \|Severe acute respiratory syndrome coro | ···················· | ·A·················· | ···················· | ···················· | ···················· | ···················· |
| 4140 | MW276582.1 | \|Severe acute respiratory syndrome coro | ···················· | ·A·················· | ···················· | ···················· | ···················· | ···················· |
| 4141 | MW276583.1 | \|Severe acute respiratory syndrome coro | ···················· | ·A·················· | ···················· | ···················· | ···················· | ···················· |
| 4142 | MW276584.1 | \|Severe acute respiratory syndrome coro | ···················· | ·A·················· | ···················· | ···················· | ···················· | ···················· |
| 4143 | MW276585.1 | \|Severe acute respiratory syndrome coro | ···················· | ·A·················· | ···················· | ···················· | ···················· | ···················· |
| 4144 | MW276586.1 | \|Severe acute respiratory syndrome coro | ···················· | ·A·················· | ···················· | ···················· | ···················· | ···················· |
| 4145 | MW276587.1 | \|Severe acute respiratory syndrome coro | ···················· | ·A·················· | ···················· | ···················· | ···················· | ···················· |
| 4147 | MW276589.1 | \|Severe acute respiratory syndrome coro | ···················· | ·A·················· | ···················· | ···················· | ···················· | ···················· |
| 4149 | MW276592.1 | \|Severe acute respiratory syndrome coro | ···················· | ·A·················· | ···················· | ···················· | ···················· | ···················· |
| 4150 | MW276593.1 | \|Severe acute respiratory syndrome coro | ···················· | ·A·················· | ···················· | ···················· | ···················· | ···················· |
| 4151 | MW276595.1 | \|Severe acute respiratory syndrome coro | ···················· | ·A·················· | ···················· | ···················· | ···················· | ···················· |
| 4152 | MW276596.1 | \|Severe acute respiratory syndrome coro | ···················· | ·A·················· | ···················· | ···················· | ···················· | ···················· |
| 4153 | MW276597.1 | \|Severe acute respiratory syndrome coro | ···················· | ·A·················· | ···················· | ···················· | ···················· | ···················· |
| 4154 | MW276599.1 | \|Severe acute respiratory syndrome coro | ···················· | ·A·················· | ···················· | ···················· | ···················· | ···················· |
| 4155 | MW276600.1 | \|Severe acute respiratory syndrome coro | ···················· | ···················· | ···················· | ···················· | ···················· | ···················· |
| 4156 | MW276601.1 | \|Severe acute respiratory syndrome coro | ···················· | ·A·················· | ···················· | ···················· | ···················· | ···················· |
| 4157 | MW276603.1 | \|Severe acute respiratory syndrome coro | ···················· | ·A·················· | ···················· | ···················· | ···················· | ···················· |
| 4159 | MW276606.1 | \|Severe acute respiratory syndrome coro | ···················· | ···················· | ···················· | ···················· | ···················· | ···················· |
| 4160 | MW276607.1 | \|Severe acute respiratory syndrome coro | ···················· | ···················· | ···················· | ···················· | ···················· | ···················· |
| 4161 | MW276608.1 | \|Severe acute respiratory syndrome coro | ···················· | ···················· | ···················· | ···················· | ···················· | ···················· |
| 4162 | MW276609.1 | \|Severe acute respiratory syndrome coro | ···················· | ···················· | ···················· | ···················· | ···················· | ···················· |
| 4163 | MW276610.1 | \|Severe acute respiratory syndrome coro | ···················· | ·············A······ | ···················· | ···················· | ···················· | ···················· |
| 4164 | MW276612.1 | \|Severe acute respiratory syndrome coro | ···················· | ·A·················· | ···················· | ···················· | ···················· | ···················· |
| 4165 | MW276613.1 | \|Severe acute respiratory syndrome coro | ···················· | ···················· | ···················· | ···················· | ···················· | ···················· |
| 4166 | MW276614.1 | \|Severe acute respiratory syndrome coro | ···················· | ·A·················· | ···················· | ···················· | ···················· | ···················· |
| 4167 | MW276615.1 | \|Severe acute respiratory syndrome coro | ···················· | ·A·················· | ···················· | ···················· | ···················· | ···················· |
| 4168 | MW276616.1 | \|Severe acute respiratory syndrome coro | ···················· | ·A·················· | ···················· | ···················· | ···················· | ···················· |
| 4169 | MW276617.1 | \|Severe acute respiratory syndrome coro | ···················· | ···················· | ···················· | ···················· | ···················· | ···················· |
| 4170 | MW276618.1 | \|Severe acute respiratory syndrome coro | ···················· | ·A·················· | ···················· | ···················· | ···················· | ···················· |
| 4171 | MW276621.1 | \|Severe acute respiratory syndrome coro | ···················· | ·A·················· | ···················· | ···················· | ···················· | ···················· |
| 4172 | MW276622.1 | \|Severe acute respiratory syndrome coro | ···················· | ···················· | ···················· | ···················· | ···················· | ···················· |
| 4173 | MW276623.1 | \|Severe acute respiratory syndrome coro | ···················· | ·A·················· | ···················· | ···················· | ···················· | ···················· |
| 4174 | MW276625.1 | \|Severe acute respiratory syndrome coro | ···················· | ···················· | ···········T········ | ···················· | ···················· | ···················· |
| 4175 | MW276626.1 | \|Severe acute respiratory syndrome coro | ···················· | ···················· | ···················· | ···················· | ···················· | ···················· |
| 4176 | MW276627.1 | \|Severe acute respiratory syndrome coro | ···················· | ·A·················· | ···················· | ···················· | ···················· | ···················· |
| 4177 | MW276629.1 | \|Severe acute respiratory syndrome coro | ···················· | ·A·················· | ···················· | ···················· | ···················· | ···················· |
| 4178 | MW276630.1 | \|Severe acute respiratory syndrome coro | ···················· | ·A·················· | ···················· | ···················· | ···················· | ···················· |
| 4179 | MW276631.1 | \|Severe acute respiratory syndrome coro | ···················· | ·A·················· | ···················· | ···················· | ···················· | ···················· |
| 4181 | MW276634.1 | \|Severe acute respiratory syndrome coro | ···················· | ·A·················· | ···················· | ···················· | ···················· | ···················· |
| 4182 | MW276635.1 | \|Severe acute respiratory syndrome coro | ···················· | ·A·················· | ···················· | ···················· | ···················· | ···················· |
| 4183 | MW276636.1 | \|Severe acute respiratory syndrome coro | ···T················ | ·A·················· | ···················· | ···················· | ···················· | ···················· |
| 4185 | MW276638.1 | \|Severe acute respiratory syndrome coro | ···················· | ···················· | ···················· | ···················· | ···················· | ···················· |
| 4186 | MW276639.1 | \|Severe acute respiratory syndrome coro | ···················· | ·A·················· | ···················· | ···················· | ···················· | ···················· |
| 4187 | MW276641.1 | \|Severe acute respiratory syndrome coro | ···················· | ·A·················· | ···················· | ···················· | ···················· | ···················· |
| 4189 | MW276644.1 | \|Severe acute respiratory syndrome coro | ···················· | ···················· | ···················· | ···················· | ···················· | ···················· |
| 4190 | MW276645.1 | \|Severe acute respiratory syndrome coro | ···················· | ···················· | ···················· | ···················· | ···················· | ···················· |
| 4191 | MW276646.1 | \|Severe acute respiratory syndrome coro | ···················· | ···················· | ···················· | ···················· | ···················· | ···················· |
| 4192 | MW276647.1 | \|Severe acute respiratory syndrome coro | ···················· | ···················· | ···················· | ···················· | ···················· | ···················· |
| 4193 | MW276648.1 | \|Severe acute respiratory syndrome coro | ···················· | ···················· | ···················· | ···················· | ···················· | ···················· |
| 4194 | MW276649.1 | \|Severe acute respiratory syndrome coro | ···················· | ·A·················· | ···················· | ···················· | ···················· | ···················· |
| 4195 | MW276651.1 | \|Severe acute respiratory syndrome coro | ···················· | ···················· | ···················· | ···················· | ···················· | ···················· |
| 4196 | MW276652.1 | \|Severe acute respiratory syndrome coro | ···················· | ···················· | ···················· | ···················· | ···················· | ···················· |
| 4197 | MW276653.1 | \|Severe acute respiratory syndrome coro | ···················· | ···················· | ···················· | ···················· | ···················· | ···················· |
| 4198 | MW276654.1 | \|Severe acute respiratory syndrome coro | ···················· | ···················· | ···················· | ···················· | ···················· | ···················· |
| 4199 | MW276656.1 | \|Severe acute respiratory syndrome coro | ···················· | ·A·················· | ···················· | ···················· | ···················· | ···················· |
| 4200 | MW276657.1 | \|Severe acute respiratory syndrome coro | ···················· | ···················· | ···················· | ···················· | ···················· | ···················· |
| 4201 | MW276658.1 | \|Severe acute respiratory syndrome coro | ···T················ | ·A·················· | ···················· | ···················· | ···················· | ···················· |
| 4202 | MW276659.1 | \|Severe acute respiratory syndrome coro | ···················· | ···················· | ···················· | ···················· | ···················· | ···················· |
| 4203 | MW276661.1 | \|Severe acute respiratory syndrome coro | ···················· | ···················· | ···················· | ···················· | ···················· | ···················· |
| 4204 | MW276663.1 | \|Severe acute respiratory syndrome coro | ···················· | ·A·················· | ···················· | ···················· | ···················· | ···················· |
| 4206 | MW276667.1 | \|Severe acute respiratory syndrome coro | ···················· | ···················· | ···················· | ···················· | ···················· | ···················· |
| 4207 | MW276669.1 | \|Severe acute respiratory syndrome coro | ···················· | ·A·················· | ···················· | ···················· | ···················· | ···················· |
| 4209 | MW276671.1 | \|Severe acute respiratory syndrome coro | ···················· | ···················· | ···················· | ···················· | ···················· | ···················· |
| 4210 | MW276672.1 | \|Severe acute respiratory syndrome coro | ···················· | ·A·················· | ···················· | ···················· | ···················· | ···················· |
| 4211 | MW276675.1 | \|Severe acute respiratory syndrome coro | ···················· | ···················· | ···················· | ···················· | ···················· | ···················· |
| 4212 | MW276679.1 | \|Severe acute respiratory syndrome coro | ···················· | ···················· | ···················· | ···················· | ···················· | ···················· |
| 4213 | MW276680.1 | \|Severe acute respiratory syndrome coro | ···················· | ···················· | ···················· | ···················· | ···················· | ···················· |
| 4214 | MW276681.1 | \|Severe acute respiratory syndrome coro | ···················· | ···················· | ···················· | ···················· | ···················· | ···················· |
| 4215 | MW276684.1 | \|Severe acute respiratory syndrome coro | ···················· | ···················· | ···················· | ···················· | ···················· | ···················· |
| 4216 | MW276686.1 | \|Severe acute respiratory syndrome coro | ···················· | ···················· | ···················· | ···················· | ···················· | ···················· |
| 4217 | MW276687.1 | \|Severe acute respiratory syndrome coro | ···················· | ···················· | ···················· | ···················· | ···················· | ···················· |
| 4218 | MW276688.1 | \|Severe acute respiratory syndrome coro | ···················· | ···················· | ···················· | ···················· | ···················· | ···················· |
| 4219 | MW276689.1 | \|Severe acute respiratory syndrome coro | ···················· | ···················· | ···················· | ···················· | ···················· | ···················· |
| 4220 | MW276690.1 | \|Severe acute respiratory syndrome coro | ···················· | ···················· | ···················· | ···················· | ···················· | ···················· |

| | | | | | | | | |
|---|---|---|---|---|---|---|---|---|
| 4221 | MW276691.1 | Severe acute respiratory syndrome coro | .................... | .................... | .................... | .................... | .................... | .................... |
| 4222 | MW276692.1 | Severe acute respiratory syndrome coro | .................... | .................... | .................... | .................... | .................... | .................... |
| 4223 | MW276693.1 | Severe acute respiratory syndrome coro | .................... | .................... | .................... | .................... | .................... | .................... |
| 4224 | MW276695.1 | Severe acute respiratory syndrome coro | .................... | .................... | .................... | .................... | .................... | .................... |
| 4225 | MW276696.1 | Severe acute respiratory syndrome coro | .................... | .................... | .................... | .................... | .................... | .................... |
| 4226 | MW276697.1 | Severe acute respiratory syndrome coro | .................... | .................... | .................... | .................... | .................... | .................... |
| 4227 | MW276699.1 | Severe acute respiratory syndrome coro | .................... | .................... | .................... | .................... | .................... | .................... |
| 4228 | MW276700.1 | Severe acute respiratory syndrome coro | .................... | .................... | .................... | .................... | .................... | .................... |
| 4229 | MW276702.1 | Severe acute respiratory syndrome coro | .................... | .................... | .................... | .................... | .................... | .................... |
| 4230 | MW276703.1 | Severe acute respiratory syndrome coro | .................... | .................... | .................... | .................... | .................... | .................... |
| 4231 | MW276704.1 | Severe acute respiratory syndrome coro | .................... | .................... | .................... | .................... | .................... | .................... |
| 4232 | MW276705.1 | Severe acute respiratory syndrome coro | .................... | .................... | .................... | .................... | .................... | .................... |
| 4233 | MW276707.1 | Severe acute respiratory syndrome coro | .................... | .................... | .................... | .................... | .................... | .................... |
| 4234 | MW276708.1 | Severe acute respiratory syndrome coro | .................... | .................... | .................... | .................... | .................... | .................... |
| 4235 | MW276709.1 | Severe acute respiratory syndrome coro | .................... | .................... | .................... | .................... | .................... | .................... |
| 4236 | MW276710.1 | Severe acute respiratory syndrome coro | .................... | .................... | .................... | .................... | .................... | .................... |
| 4237 | MW276711.1 | Severe acute respiratory syndrome coro | .................... | .................... | .................... | .................... | .................... | .................... |
| 4238 | MW276712.1 | Severe acute respiratory syndrome coro | .................... | .................... | .................... | .................... | .................... | .................... |
| 4239 | MW276713.1 | Severe acute respiratory syndrome coro | .................... | .................... | .................... | .................... | .................... | .................... |
| 4240 | MW276714.1 | Severe acute respiratory syndrome coro | .................... | .................... | .................... | .................... | .................... | .................... |
| 4241 | MW276717.1 | Severe acute respiratory syndrome coro | .................... | .................... | .................... | .................... | .................... | .................... |
| 4242 | MW276718.1 | Severe acute respiratory syndrome coro | .................... | .................... | .................... | .................... | .................... | .................... |
| 4243 | MW276720.1 | Severe acute respiratory syndrome coro | .................... | .................... | .................... | .................... | .................... | .................... |
| 4244 | MW276721.1 | Severe acute respiratory syndrome coro | .................... | .................... | .................... | .................... | .................... | .................... |
| 4245 | MW276723.1 | Severe acute respiratory syndrome coro | .................... | .................... | .................... | .................... | .................... | .................... |
| 4246 | MW276724.1 | Severe acute respiratory syndrome coro | .................... | .................... | .................... | .................... | .................... | .................... |
| 4247 | MW276725.1 | Severe acute respiratory syndrome coro | .................... | .................... | .................... | .................... | .................... | .................... |
| 4248 | MW276726.1 | Severe acute respiratory syndrome coro | .................... | .................... | .................... | .................... | .................... | .................... |
| 4249 | MW276727.1 | Severe acute respiratory syndrome coro | .................... | .................... | .................... | .................... | .................... | .................... |
| 4250 | MW276728.1 | Severe acute respiratory syndrome coro | .................... | .................... | .................... | .................... | .................... | .................... |
| 4251 | MW276731.1 | Severe acute respiratory syndrome coro | .................... | .................... | .................... | .................... | .................... | .................... |
| 4252 | MW276732.1 | Severe acute respiratory syndrome coro | .................... | .................... | .................... | .................... | .................... | .................... |
| 4253 | MW276733.1 | Severe acute respiratory syndrome coro | .................... | .................... | .................... | .................... | .................... | .................... |
| 4254 | MW276734.1 | Severe acute respiratory syndrome coro | .................... | .................... | .................... | .................... | .................... | .................... |
| 4255 | MW276735.1 | Severe acute respiratory syndrome coro | .................... | .................... | .................... | .................... | .................... | .................... |
| 4256 | MW276736.1 | Severe acute respiratory syndrome coro | .................... | .................... | .................... | .................... | .................... | .................... |
| 4257 | MW276737.1 | Severe acute respiratory syndrome coro | .................... | .................... | .................... | .................... | .................... | .................... |
| 4258 | MW276738.1 | Severe acute respiratory syndrome coro | .................... | .................... | .................... | .................... | .................... | .................... |
| 4259 | MW276740.1 | Severe acute respiratory syndrome coro | .................... | .................... | .................... | .................... | .................... | .................... |
| 4260 | MW276741.1 | Severe acute respiratory syndrome coro | .................... | .................... | .................... | .................... | .................... | .................... |
| 4261 | MW276742.1 | Severe acute respiratory syndrome coro | .................... | .................... | .................... | .................... | .................... | .................... |
| 4262 | MW276743.1 | Severe acute respiratory syndrome coro | .................... | .................... | .................... | .................... | .................... | .................... |
| 4263 | MW276744.1 | Severe acute respiratory syndrome coro | .................... | .................... | .................... | .................... | .................... | .................... |
| 4264 | MW276745.1 | Severe acute respiratory syndrome coro | .................... | .................... | .................... | .................... | .................... | .................... |
| 4266 | MW276748.1 | Severe acute respiratory syndrome coro | .................... | .................... | .................... | .................... | .................... | .................... |
| 4267 | MW276749.1 | Severe acute respiratory syndrome coro | .................... | .................... | .................... | .................... | .................... | .................... |
| 4268 | MW276750.1 | Severe acute respiratory syndrome coro | .................... | .................... | .................... | .................... | .................... | .................... |
| 4269 | MW276751.1 | Severe acute respiratory syndrome coro | .................... | .................... | .................... | .................... | .................... | .................... |
| 4270 | MW276752.1 | Severe acute respiratory syndrome coro | .................... | ......A............. | .................... | .................... | .................... | .................... |
| 4271 | MW276753.1 | Severe acute respiratory syndrome coro | .................... | .................... | .................... | .................... | .................... | .................... |
| 4272 | MW276754.1 | Severe acute respiratory syndrome coro | .................... | .................... | .................... | .................... | .................... | .................... |
| 4273 | MW276755.1 | Severe acute respiratory syndrome coro | .................... | .................... | .................... | .................... | .................... | .................... |
| 4274 | MW276756.1 | Severe acute respiratory syndrome coro | .................... | .................... | .................... | .................... | .................... | .................... |
| 4277 | MW276760.1 | Severe acute respiratory syndrome coro | .................... | .................... | .................... | .................... | .................... | .................... |
| 4278 | MW276762.1 | Severe acute respiratory syndrome coro | .................... | .................... | .................... | .................... | .................... | .................... |
| 4279 | MW276763.1 | Severe acute respiratory syndrome coro | .................... | .................... | .................... | .................... | .................... | .................... |
| 4280 | MW276764.1 | Severe acute respiratory syndrome coro | .................... | .................... | .................... | .................... | .................... | .................... |
| 4281 | MW276766.1 | Severe acute respiratory syndrome coro | .................... | .................... | .................... | .................... | .................... | .................... |
| 4282 | MW276767.1 | Severe acute respiratory syndrome coro | .................... | .................... | .................... | .................... | .................... | .................... |
| 4283 | MW276768.1 | Severe acute respiratory syndrome coro | .................... | .................... | .................... | .................... | .................... | .................... |
| 4284 | MW276769.1 | Severe acute respiratory syndrome coro | .................... | .................... | .................... | .................... | .................... | .................... |
| 4285 | MW276770.1 | Severe acute respiratory syndrome coro | .................... | .................... | .................... | .................... | .................... | .................... |
| 4286 | MW276772.1 | Severe acute respiratory syndrome coro | .................... | .................... | .................... | .................... | .................... | .................... |
| 4287 | MW276773.1 | Severe acute respiratory syndrome coro | .................... | .................... | .................... | .................... | .................... | .................... |
| 4288 | MW276774.1 | Severe acute respiratory syndrome coro | .................... | .................... | .................... | .................... | .................... | .................... |
| 4289 | MW276775.1 | Severe acute respiratory syndrome coro | .................... | .................... | .................... | .................... | .................... | .................... |
| 4290 | MW276776.1 | Severe acute respiratory syndrome coro | .................... | .................... | .................... | .................... | .................... | .................... |
| 4291 | MW276777.1 | Severe acute respiratory syndrome coro | .................... | .................... | .................... | .................... | .................... | .................... |
| 4292 | MW276778.1 | Severe acute respiratory syndrome coro | .................... | .................... | .................... | .................... | .................... | .................... |
| 4293 | MW276779.1 | Severe acute respiratory syndrome coro | .................... | .................... | .................... | .................... | .................... | .................... |
| 4294 | MW276780.1 | Severe acute respiratory syndrome coro | .................... | .................... | .................... | .................... | .................... | .................... |
| 4296 | MW276782.1 | Severe acute respiratory syndrome coro | .................... | .................... | .................... | .................... | .................... | .................... |
| 4297 | MW276783.1 | Severe acute respiratory syndrome coro | .................... | .................... | .................... | .................... | .................... | .................... |
| 4298 | MW276784.1 | Severe acute respiratory syndrome coro | .................... | .................... | .................... | .................... | .................... | .................... |
| 4299 | MW276785.1 | Severe acute respiratory syndrome coro | .................... | .................... | .................... | .................... | .................... | .................... |
| 4300 | MW276786.1 | Severe acute respiratory syndrome coro | .................... | .................... | .................... | .................... | .................... | .................... |
| 4301 | MW276787.1 | Severe acute respiratory syndrome coro | .................... | .................... | .................... | .................... | .................... | .................... |
| 4302 | MW276788.1 | Severe acute respiratory syndrome coro | .................... | .................... | .................... | .................... | .................... | .................... |

| | | | | | | | | | |
|---|---|---|---|---|---|---|---|---|---|
| 4303 | MW276789.1 | Severe acute respiratory syndrome coro | ···················· | ···················· | ···················· | ···················· | ···················· | ···················· | |
| 4304 | MW276791.1 | Severe acute respiratory syndrome coro | ···················· | ···················· | ···················· | ···················· | ···················· | ···················· | |
| 4305 | MW276792.1 | Severe acute respiratory syndrome coro | ···················· | ···················· | ···················· | ···················· | ···················· | ···················· | |
| 4306 | MW276793.1 | Severe acute respiratory syndrome coro | ···················· | ···················· | ···················· | ···················· | ···················· | ···················· | |
| 4307 | MW276794.1 | Severe acute respiratory syndrome coro | ···················· | ···················· | ···················· | ···················· | ···················· | ···················· | |
| 4308 | MW276795.1 | Severe acute respiratory syndrome coro | ···················· | ···················· | ···················· | ···················· | ···················· | ···················· | |
| 4309 | MW276796.1 | Severe acute respiratory syndrome coro | ···················· | ···················· | ···················· | ···················· | ···················· | ···················· | |
| 4310 | MW276797.1 | Severe acute respiratory syndrome coro | ···················· | ···················· | ···················· | ···················· | ···················· | ···················· | |
| 4311 | MW276798.1 | Severe acute respiratory syndrome coro | ···················· | ···················· | ···················· | ···················· | ···················· | ···················· | |
| 4312 | MW276799.1 | Severe acute respiratory syndrome coro | ···················· | ···················· | ···················· | ···················· | ···················· | ···················· | |
| 4313 | MW276800.1 | Severe acute respiratory syndrome coro | ···················· | ···················· | ···················· | ···················· | ···················· | ···················· | |
| 4314 | MW276802.1 | Severe acute respiratory syndrome coro | ···················· | ···················· | ···················· | ···················· | ···················· | ···················· | |
| 4315 | MW276804.1 | Severe acute respiratory syndrome coro | ···················· | ···················· | ···················· | ···················· | ···················· | ···················· | |
| 4316 | MW276805.1 | Severe acute respiratory syndrome coro | ···················· | ···················· | ···················· | ···················· | ···················· | ···················· | |
| 4317 | MW276806.1 | Severe acute respiratory syndrome coro | ···················· | ···················· | ···················· | ···················· | ···················· | ···················· | |
| 4318 | MW276807.1 | Severe acute respiratory syndrome coro | ···················· | ···················· | ···················· | ···················· | ···················· | ···················· | |
| 4319 | MW276808.1 | Severe acute respiratory syndrome coro | ···················· | ···················· | ···················· | ···················· | ···················· | ···················· | |
| 4320 | MW276809.1 | Severe acute respiratory syndrome coro | ···················· | ···················· | ···················· | ···················· | ···················· | ···················· | |
| 4321 | MW276811.1 | Severe acute respiratory syndrome coro | ···················· | ···················· | ···················· | ···················· | ···················· | ···················· | |
| 4322 | MW276812.1 | Severe acute respiratory syndrome coro | ···················· | ···················· | ···················· | ···················· | ···················· | ···················· | |
| 4323 | MW276813.1 | Severe acute respiratory syndrome coro | ···················· | ···················· | ···················· | ···················· | ···················· | ···················· | |
| 4324 | MW276814.1 | Severe acute respiratory syndrome coro | ···················· | ···················· | ···················· | ···················· | ···················· | ···················· | |
| 4325 | MW276816.1 | Severe acute respiratory syndrome coro | ···················· | ···················· | ···················· | ···················· | ···················· | ···················· | |
| 4326 | MW276817.1 | Severe acute respiratory syndrome coro | ···················· | ···················· | ···················· | ···················· | ···················· | ···················· | |
| 4327 | MW276818.1 | Severe acute respiratory syndrome coro | ···················· | ···················· | ···················· | ···················· | ···················· | ···················· | |
| 4328 | MW276820.1 | Severe acute respiratory syndrome coro | ···················· | ···················· | ···················· | ···················· | ···················· | ···················· | |
| 4329 | MW276821.1 | Severe acute respiratory syndrome coro | ···················· | ···················· | ···················· | ···················· | ···················· | ···················· | |
| 4330 | MW276822.1 | Severe acute respiratory syndrome coro | ···················· | ···················· | ···················· | ···················· | ···················· | ···················· | |
| 4331 | MW276823.1 | Severe acute respiratory syndrome coro | ···················· | ···················· | ···················· | ···················· | ···················· | ···················· | |
| 4332 | MW276824.1 | Severe acute respiratory syndrome coro | ···················· | ···················· | ···················· | ···················· | ···················· | ···················· | |
| 4333 | MW276826.1 | Severe acute respiratory syndrome coro | ···················· | ···················· | ···················· | ···················· | ···················· | ···················· | |
| 4334 | MW276827.1 | Severe acute respiratory syndrome coro | ···················· | ···················· | ···················· | ···················· | ···················· | ···················· | |
| 4335 | MW276828.1 | Severe acute respiratory syndrome coro | ···················· | ···················· | ···················· | ···················· | ···················· | ···················· | |
| 4336 | MW276829.1 | Severe acute respiratory syndrome coro | ···················· | ···················· | ···················· | ···················· | ···················· | ···················· | |
| 4337 | MW276830.1 | Severe acute respiratory syndrome coro | ···················· | ···················· | ···················· | ···················· | ···················· | ···················· | |
| 4338 | MW276831.1 | Severe acute respiratory syndrome coro | ···················· | ···················· | ···················· | ···················· | ···················· | ···················· | |
| 4339 | MW276832.1 | Severe acute respiratory syndrome coro | ···················· | ···················· | ···················· | ···················· | ···················· | ···················· | |
| 4340 | MW276833.1 | Severe acute respiratory syndrome coro | ···················· | ···················· | ···················· | ···················· | ···················· | ···················· | |
| 4341 | MW276834.1 | Severe acute respiratory syndrome coro | ···················· | ···················· | ···················· | ···················· | ···················· | ···················· | |
| 4342 | MW276835.1 | Severe acute respiratory syndrome coro | ···················· | ···················· | ···················· | ···················· | ···················· | ···················· | |
| 4343 | MW276836.1 | Severe acute respiratory syndrome coro | ···················· | ···················· | ···················· | ···················· | ···················· | ···················· | |
| 4344 | MW276837.1 | Severe acute respiratory syndrome coro | ···················· | ···················· | ···················· | ···················· | ···················· | ···················· | |
| 4345 | MW276839.1 | Severe acute respiratory syndrome coro | ···················· | ···················· | ···················· | ···················· | ···················· | ···················· | |
| 4346 | MW276840.1 | Severe acute respiratory syndrome coro | ···················· | ···················· | ···················· | ···················· | ···················· | ···················· | |
| 4347 | MW276841.1 | Severe acute respiratory syndrome coro | ···················· | ···················· | ···················· | ···················· | ···················· | ···················· | |
| 4348 | MW276842.1 | Severe acute respiratory syndrome coro | ···················· | ···················· | ···················· | ···················· | ···················· | ···················· | |
| 4349 | MW276844.1 | Severe acute respiratory syndrome coro | ···················· | ···················· | ···················· | ···················· | ···················· | ···················· | |
| 4350 | MW276845.1 | Severe acute respiratory syndrome coro | ···················· | ···················· | ···················· | ···················· | ···················· | ···················· | |
| 4351 | MW276847.1 | Severe acute respiratory syndrome coro | ···················· | ···················· | ···················· | ···················· | ···················· | ···················· | |
| 4352 | MW276850.1 | Severe acute respiratory syndrome coro | ···················· | ···················· | ···················· | ···················· | ···················· | ···················· | |
| 4353 | MW276851.1 | Severe acute respiratory syndrome coro | ···················· | ···················· | ···················· | ···················· | ···················· | ···················· | |
| 4354 | MW276852.1 | Severe acute respiratory syndrome coro | ···················· | ···················· | ···················· | ···················· | ···················· | ···················· | |
| 4355 | MW276854.1 | Severe acute respiratory syndrome coro | ···················· | ···················· | ···················· | ···················· | ···················· | ···················· | |
| 4356 | MW276856.1 | Severe acute respiratory syndrome coro | ···················· | ···················· | ···················· | ···················· | ···················· | ···················· | |
| 4357 | MW276857.1 | Severe acute respiratory syndrome coro | ···················· | ···················· | ···················· | ···················· | ···················· | ···················· | |
| 4358 | MW276858.1 | Severe acute respiratory syndrome coro | ···················· | ···················· | ···················· | ···················· | ···················· | ···················· | |
| 4359 | MW276861.1 | Severe acute respiratory syndrome coro | ···················· | ···················· | ···················· | ···················· | ···················· | ···················· | |
| 4360 | MW276863.1 | Severe acute respiratory syndrome coro | ···················· | ···················· | ···················· | ···················· | ···················· | ···················· | |
| 4361 | MW276865.1 | Severe acute respiratory syndrome coro | ···················· | ···················· | ···················· | ···················· | ···················· | ···················· | |
| 4362 | MW276866.1 | Severe acute respiratory syndrome coro | ···················· | ···················· | ···················· | ···················· | ···················· | ···················· | |
| 4363 | MW276867.1 | Severe acute respiratory syndrome coro | ···················· | ···················· | ···················· | ···················· | ···················· | ···················· | |
| 4364 | MW276868.1 | Severe acute respiratory syndrome coro | ···················· | ···················· | ···················· | ···················· | ···················· | ···················· | |
| 4365 | MW276869.1 | Severe acute respiratory syndrome coro | ···················· | ···················· | ···················· | ···················· | ···················· | ···················· | |
| 4366 | MW276870.1 | Severe acute respiratory syndrome coro | ···················· | ···················· | ···················· | ···················· | ···················· | ···················· | |
| 4367 | MW276871.1 | Severe acute respiratory syndrome coro | ···················· | ···················· | ···················· | ···················· | NNN················· | ···················· | 1 |
| 4368 | MW276872.1 | Severe acute respiratory syndrome coro | ···················· | ···················· | ···················· | ···················· | ···················· | ···················· | |
| 4369 | MW276873.1 | Severe acute respiratory syndrome coro | ···················· | ···················· | ···················· | ···················· | ···················· | ···················· | |
| 4370 | MW276874.1 | Severe acute respiratory syndrome coro | ···················· | ···················· | ···················· | ···················· | ···················· | ···················· | |
| 4371 | MW276875.1 | Severe acute respiratory syndrome coro | ···················· | ···················· | ···················· | ···················· | ···················· | ···················· | |
| 4372 | MW276876.1 | Severe acute respiratory syndrome coro | ···················· | ···················· | ···················· | ···················· | ···················· | ···················· | |
| 4373 | MW276877.1 | Severe acute respiratory syndrome coro | ···················· | ···················· | ···················· | ···················· | ···················· | ···················· | |
| 4374 | MW276879.1 | Severe acute respiratory syndrome coro | ···················· | ···················· | ···················· | ···················· | ···················· | ···················· | |
| 4375 | MW276880.1 | Severe acute respiratory syndrome coro | ···················· | ···················· | ···················· | ···················· | ···················· | ···················· | |
| 4376 | MW276882.1 | Severe acute respiratory syndrome coro | ···················· | ···················· | ···················· | ···················· | ···················· | ···················· | |
| 4377 | MW276883.1 | Severe acute respiratory syndrome coro | ···················· | ···················· | ···················· | ···················· | ···················· | ···················· | |
| 4378 | MW276885.1 | Severe acute respiratory syndrome coro | ···················· | ···················· | ···················· | ···················· | ···················· | ···················· | |
| 4379 | MW276886.1 | Severe acute respiratory syndrome coro | ···················· | ···················· | ···················· | ···················· | ···················· | ···················· | |
| 4382 | MW276889.1 | Severe acute respiratory syndrome coro | ···················· | ···················· | ···················· | ···················· | ···················· | ···················· | |

4383 MW276890.1 |Severe acute respiratory syndrome coro .................... .................... .................... .................... .................... ....................
4384 MW276891.1 |Severe acute respiratory syndrome coro .................... .................... .................... .................... .................... ....................
4385 MW276893.1 |Severe acute respiratory syndrome coro .................... .................... .................... .................... .................... ....................
4386 MW276895.1 |Severe acute respiratory syndrome coro .................... .................... .................... .................... .................... ....................
4387 MW276896.1 |Severe acute respiratory syndrome coro .................... .................... .................... .................... .................... ....................
4388 MW276901.1 |Severe acute respiratory syndrome coro .................... .................... .................... .................... .................... ....................
4389 MW276902.1 |Severe acute respiratory syndrome coro .................... .................... .................... .................... .................... ....................
4390 MW276903.1 |Severe acute respiratory syndrome coro .................... .................... .................... .................... .................... ....................
4391 MW276905.1 |Severe acute respiratory syndrome coro ........T........... .................... .................... .................... .................... ....................
4392 MW276907.1 |Severe acute respiratory syndrome coro .................... .................... .................... .................... .................... ....................
4393 MW276908.1 |Severe acute respiratory syndrome coro .................... .................... .................... .................... .................... ....................
4394 MW276909.1 |Severe acute respiratory syndrome coro .................... .................... .................... .................... .................... ....................
4395 MW276913.1 |Severe acute respiratory syndrome coro .................... .................... .................... .................... .................... ....................
4396 MW276916.1 |Severe acute respiratory syndrome coro .................... .................... .................... .................... .................... ....................
4397 MW276917.1 |Severe acute respiratory syndrome coro .................... .................... .................... .................... .................... ....................
4398 MW276918.1 |Severe acute respiratory syndrome coro .................... .................... .................... .................... .................... ....................
4399 MW276919.1 |Severe acute respiratory syndrome coro .................... .................... .................... .................... .................... ....................
4400 MW276920.1 |Severe acute respiratory syndrome coro .................... .................... .................... .................... .................... ....................
4401 MW276921.1 |Severe acute respiratory syndrome coro .................... .................... .................... .................... .................... ....................
4402 MW276922.1 |Severe acute respiratory syndrome coro .................... .................... .................... .................... .................... ....................
4404 MW276924.1 |Severe acute respiratory syndrome coro .................... .................... .................... .................... .................... ....................
4405 MW276925.1 |Severe acute respiratory syndrome coro .................... .................... .................... .................... .................... ....................
4406 MW276926.1 |Severe acute respiratory syndrome coro .................... .................... .................... .................... .................... ....................
4407 MW276927.1 |Severe acute respiratory syndrome coro .................... .................... .................... .................... .................... ....................
4408 MW276929.1 |Severe acute respiratory syndrome coro .................... .................... .................... .................... .................... ....................
4409 MW276930.1 |Severe acute respiratory syndrome coro .................... .................... .................... .................... .................... ....................
4410 MW276931.1 |Severe acute respiratory syndrome coro .................... .................... .................... .................... .................... ....................
4411 MW276932.1 |Severe acute respiratory syndrome coro .................... .................... .................... .................... .................... ....................
4412 MW276933.1 |Severe acute respiratory syndrome coro .................... .................... .................... .................... .................... ....................
4413 MW276936.1 |Severe acute respiratory syndrome coro .................... .................... .................... .................... .................... ....................
4414 MW276937.1 |Severe acute respiratory syndrome coro .................... .................... .................... .................... .................... ....................
4415 MW276938.1 |Severe acute respiratory syndrome coro .................... .................... .................... .................... .................... ....................
4416 MW276940.1 |Severe acute respiratory syndrome coro .................... .................... .................... .................... .................... ....................
4417 MW276941.1 |Severe acute respiratory syndrome coro .................... .................... .................... .................... .................... ....................
4418 MW276942.1 |Severe acute respiratory syndrome coro .................... .................... .................... .................... .................... ....................
4419 MW276943.1 |Severe acute respiratory syndrome coro .................... .................... .................... .................... .................... ....................
4420 MW276944.1 |Severe acute respiratory syndrome coro .................... .................... .................... .................... .................... ....................
4421 MW276945.1 |Severe acute respiratory syndrome coro .................... .................... .................... .................... .................... ....................
4422 MW276946.1 |Severe acute respiratory syndrome coro .................... .................... .................... .................... .................... ....................
4423 MW276947.1 |Severe acute respiratory syndrome coro .................... .................... .................... .................... .................... ....................
4424 MW276948.1 |Severe acute respiratory syndrome coro .................... .................... .................... .................... .................... ....................
4425 MW276950.1 |Severe acute respiratory syndrome coro .................... .................... .................... .................... .................... ....................
4426 MW276951.1 |Severe acute respiratory syndrome coro .................... .................... .................... .................... .................... ....................
4427 MW276952.1 |Severe acute respiratory syndrome coro .................... .................... .................... .................... .................... ....................
4428 MW276953.1 |Severe acute respiratory syndrome coro .................... .................... .................... .................... .................... ....................
4430 MW276955.1 |Severe acute respiratory syndrome coro .................... .................... .................... .................... .................... ....................
4431 MW276957.1 |Severe acute respiratory syndrome coro .................... .................... .................... .................... .................... ....................
4432 MW276958.1 |Severe acute respiratory syndrome coro .................... .................... .................... .................... .................... ....................
4433 MW276959.1 |Severe acute respiratory syndrome coro .................... .................... .................... .................... .................... ....................
4434 MW276960.1 |Severe acute respiratory syndrome coro .................... .................... .................... .................... .................... ....................
4435 MW276962.1 |Severe acute respiratory syndrome coro .................... .................... .................... .................... .................... ....................
4436 MW276963.1 |Severe acute respiratory syndrome coro .................... .................... .................... .................... .................... ....................
4437 MW276964.1 |Severe acute respiratory syndrome coro .................... .................... .................... .................... .................... ....................
4438 MW276965.1 |Severe acute respiratory syndrome coro .................... .................... .................... .................... .................... ....................
4439 MW276966.1 |Severe acute respiratory syndrome coro .................... .................... .................... .................... .................... ....................
4440 MW276967.1 |Severe acute respiratory syndrome coro .................... .................... .................... .................... .................... ....................
4441 MW276968.1 |Severe acute respiratory syndrome coro .................... .................... .................... .................... .................... ....................
4442 MW276969.1 |Severe acute respiratory syndrome coro .................... .................... .................... .................... .................... ....................
4443 MW276971.1 |Severe acute respiratory syndrome coro .................... .................... .................... .................... .................... ....................
4444 MW276972.1 |Severe acute respiratory syndrome coro .................... .................... .................... .................... .................... ....................
4445 MW276975.1 |Severe acute respiratory syndrome coro .................... .................... .................... .................... .................... ....................
4446 MW276976.1 |Severe acute respiratory syndrome coro .................... .................... .................... .................... .................... ....................
4447 MW276981.1 |Severe acute respiratory syndrome coro .................... .................... .................... .................... .................... ....................
4448 MW276982.1 |Severe acute respiratory syndrome coro .................... .................... .................... .................... .................... ....................
4449 MW276983.1 |Severe acute respiratory syndrome coro .................... .................... .................... .................... .................... ....................
4450 MW276985.1 |Severe acute respiratory syndrome coro .................... .................... .................... .................... .................... ....................
4451 MW276986.1 |Severe acute respiratory syndrome coro .................... .................... .................... .................... .................... ....................
4452 MW276987.1 |Severe acute respiratory syndrome coro .................... .................... .................... .................... .................... ....................
4453 MW276988.1 |Severe acute respiratory syndrome coro .................... .................... .................... .................... .................... ....................
4454 MW276989.1 |Severe acute respiratory syndrome coro .................... .................... .................... .................... .................... ....................
4455 MW276990.1 |Severe acute respiratory syndrome coro .................... .................... .................... .................... .................... ....................
4456 MW276991.1 |Severe acute respiratory syndrome coro .................... .................... .................... .................... .................... ....................
4457 MW276992.1 |Severe acute respiratory syndrome coro .................... .................... .................... .................... .................... ....................
4458 MW276993.1 |Severe acute respiratory syndrome coro .................... .................... .................... .................... .................... ....................
4459 MW276994.1 |Severe acute respiratory syndrome coro .................... .................... .................... .................... .................... ....................
4460 MW276995.1 |Severe acute respiratory syndrome coro .................... .................... .................... .................... .................... ....................
4461 MW277000.1 |Severe acute respiratory syndrome coro .................... .................... .................... .................... .................... ....................
4462 MW277001.1 |Severe acute respiratory syndrome coro .................... .................... .................... .................... .................... ....................

4463 MW277002.1 |Severe acute respiratory syndrome coro .................... .................... .................... .................... .................... ....................
4464 MW277003.1 |Severe acute respiratory syndrome coro .................... .................... .................... .................... .................... ....................
4465 MW277005.1 |Severe acute respiratory syndrome coro .................... .................... .................... .................... .................... ....................
4466 MW277006.1 |Severe acute respiratory syndrome coro .................... .................... .................... .................... .................... ....................
4467 MW277008.1 |Severe acute respiratory syndrome coro .................... .................... .................... .................... .................... ....................
4468 MW277009.1 |Severe acute respiratory syndrome coro .................... .................... .................... .................... .................... ....................
4469 MW277010.1 |Severe acute respiratory syndrome coro .................... .................... .................... .................... .................... ....................
4470 MW277012.1 |Severe acute respiratory syndrome coro .................... .................... .................... .................... .................... ....................
4471 MW277013.1 |Severe acute respiratory syndrome coro .................... .................... .................... .................... .................... ....................
4472 MW277016.1 |Severe acute respiratory syndrome coro .................... .................... .................... .................... .................... ....................
4473 MW277017.1 |Severe acute respiratory syndrome coro .................... .................... .................... .................... .................... ....................
4474 MW277018.1 |Severe acute respiratory syndrome coro .................... .................... .................... .................... .................... ....................
4475 MW277019.1 |Severe acute respiratory syndrome coro .................... .................... .................... .................... .................... ....................
4476 MW277021.1 |Severe acute respiratory syndrome coro .................... .................... .................... .................... .................... ....................
4477 MW277022.1 |Severe acute respiratory syndrome coro .................... .................... .................... .................... .................... ....................
4478 MW277023.1 |Severe acute respiratory syndrome coro .................... .................... .................... .................... .................... ....................
4479 MW277025.1 |Severe acute respiratory syndrome coro .................... ......A............. .................... .................... .................... ....................
4480 MW277026.1 |Severe acute respiratory syndrome coro .................... .................... .................... .................... .................... ....................
4481 MW277027.1 |Severe acute respiratory syndrome coro .................... .................... .................... .................... .................... ....................
4482 MW277029.1 |Severe acute respiratory syndrome coro .................... .................... .................... .................... .................... ....................
4484 MW277031.1 |Severe acute respiratory syndrome coro .................... .................... .................... .................... .................... ....................
4486 MW277033.1 |Severe acute respiratory syndrome coro .................... .................... .................... .................... .................... ....................
4487 MW277034.1 |Severe acute respiratory syndrome coro .................... .................... .................... .................... .................... ....................
4488 MW277035.1 |Severe acute respiratory syndrome coro .................... .................... .................... .................... .................... ....................
4489 MW277037.1 |Severe acute respiratory syndrome coro .................... .................... .................... .................... .................... ....................
4490 MW277038.1 |Severe acute respiratory syndrome coro .................... .................... .................... .................... .................... ....................
4492 MW277042.1 |Severe acute respiratory syndrome coro .................... .................... .................... .................... .................... ....................
4493 MW277043.1 |Severe acute respiratory syndrome coro .................... .................... .................... .................... .................... ....................
4494 MW277045.1 |Severe acute respiratory syndrome coro .................... .................... .................... .................... .................... ....................
4495 MW277047.1 |Severe acute respiratory syndrome coro .................... .................... .................... .................... .................... ....................
4496 MW277048.1 |Severe acute respiratory syndrome coro .................... .................... .................... .................... .................... ....................
4497 MW277049.1 |Severe acute respiratory syndrome coro .................... .................... .................... .................... .................... ....................
4498 MW277054.1 |Severe acute respiratory syndrome coro .................... .................... .................... .................... .................... ....................
4499 MW277055.1 |Severe acute respiratory syndrome coro .................... .................... .................... .................... .................... ....................
4500 MW277056.1 |Severe acute respiratory syndrome coro .................... .................... .................... .................... .................... ....................
4501 MW277057.1 |Severe acute respiratory syndrome coro .................... .................... .................... .................... .................... ....................
4502 MW277058.1 |Severe acute respiratory syndrome coro .................... .................... .................... .................... .................... ....................
4503 MW277059.1 |Severe acute respiratory syndrome coro .................... .................... .................... .................... .................... ....................
4504 MW277060.1 |Severe acute respiratory syndrome coro .................... .................... .................... .................... .................... ....................
4505 MW277061.1 |Severe acute respiratory syndrome coro .................... .................... .................... .................... .................... ....................
4506 MW277062.1 |Severe acute respiratory syndrome coro .................... .................... .................... .................... .................... ....................
4507 MW277064.1 |Severe acute respiratory syndrome coro .................... .................... .................... .................... .................... ....................
4508 MW277065.1 |Severe acute respiratory syndrome coro .................... .................... .................... .................... .................... ....................
4509 MW277067.1 |Severe acute respiratory syndrome coro .................... .................... .................... .................... .................... ....................
4510 MW277068.1 |Severe acute respiratory syndrome coro .................... .................... .................... .................... .................... ....................
4511 MW277069.1 |Severe acute respiratory syndrome coro .................... .................... .................... .................... .................... ....................
4512 MW277073.1 |Severe acute respiratory syndrome coro .................... .................... .................... .................... .................... ....................
4513 MW277074.1 |Severe acute respiratory syndrome coro .................... .................... .................... .................... .................... ....................
4514 MW277075.1 |Severe acute respiratory syndrome coro .................... .................... .................... .................... .................... ....................
4515 MW277076.1 |Severe acute respiratory syndrome coro .................... .................... .................... .................... .................... ....................
4516 MW277077.1 |Severe acute respiratory syndrome coro .................... .................... .................... .................... .................... ....................
4517 MW277078.1 |Severe acute respiratory syndrome coro .................... .................... .................... .................... .................... ....................
4518 MW277079.1 |Severe acute respiratory syndrome coro .................... .................... .................... .................... .................... ....................
4519 MW277080.1 |Severe acute respiratory syndrome coro .................... .................... .................... .................... .................... ....................
4520 MW277081.1 |Severe acute respiratory syndrome coro .................... .................... .................... .................... .................... ....................
4521 MW277083.1 |Severe acute respiratory syndrome coro .................... .................... .................... .................... .................... ....................
4522 MW277084.1 |Severe acute respiratory syndrome coro .................... .................... .................... .................... .................... ....................
4523 MW277085.1 |Severe acute respiratory syndrome coro .................... .................... .................... .................... .................... ....................
4524 MW277086.1 |Severe acute respiratory syndrome coro .................... .................... .................... .................... .................... ....................
4525 MW277087.1 |Severe acute respiratory syndrome coro .................... .................... .................... .................... .................... ....................
4526 MW277088.1 |Severe acute respiratory syndrome coro .................... .................... .................... .................... .................... ....................
4527 MW277089.1 |Severe acute respiratory syndrome coro .................... .................... .................... .................... .................... ....................
4528 MW277091.1 |Severe acute respiratory syndrome coro .................... .................... .................... .................... .................... ....................
4529 MW277093.1 |Severe acute respiratory syndrome coro .................... .................... .................... .................... .................... ....................
4530 MW277094.1 |Severe acute respiratory syndrome coro .................... .................... .................... .................... .................... ....................
4531 MW277095.1 |Severe acute respiratory syndrome coro .................... .................... .................... .................... .................... ....................
4532 MW277096.1 |Severe acute respiratory syndrome coro .................... .................... .................... .................... .................... ....................
4533 MW277097.1 |Severe acute respiratory syndrome coro .................... .................... .................... .................... .................... ....................
4534 MW277098.1 |Severe acute respiratory syndrome coro .................... .................... .................... .................... .................... ....................
4535 MW277099.1 |Severe acute respiratory syndrome coro .................... .................... .................... .................... .................... ....................
4536 MW277100.1 |Severe acute respiratory syndrome coro .................... .................... .................... .................... .................... ....................
4537 MW277101.1 |Severe acute respiratory syndrome coro .................... .................... .................... .................... .................... ....................
4538 MW277103.1 |Severe acute respiratory syndrome coro .................... .................... .................... .................... .................... ....................
4539 MW277105.1 |Severe acute respiratory syndrome coro .................... .................... .................... .................... .................... ....................
4540 MW277106.1 |Severe acute respiratory syndrome coro .................... .................... .................... .................... .................... ....................
4541 MW277107.1 |Severe acute respiratory syndrome coro .................... .................... .................... .................... .................... ....................
4542 MW277113.1 |Severe acute respiratory syndrome coro .................... .................... .................... .................... .................... ....................
4543 MW277114.1 |Severe acute respiratory syndrome coro .................... .................... .................... .................... .................... ....................

```
4544 MW277115.1 |Severe acute respiratory syndrome coro    ....................  ....................  .....................  ....................  ....................  ....................
4545 MW277116.1 |Severe acute respiratory syndrome coro    ....................  ....................  .....................  ....................  ....................  ....................
4546 MW277117.1 |Severe acute respiratory syndrome coro    ....................  ....................  .....................  ....................  ....................  ....................
4547 MW277119.1 |Severe acute respiratory syndrome coro    ....................  ....................  .....................  ....................  ....................  ....................
4548 MW277120.1 |Severe acute respiratory syndrome coro    ....................  ....................  .....................  ....................  ....................  ....................
4549 MW277123.1 |Severe acute respiratory syndrome coro    ....................  ....................  .....................  ....................  ....................  ....................
4550 MW277124.1 |Severe acute respiratory syndrome coro    ....................  ....................  .....................  ....................  ....................  ....................
4551 MW277125.1 |Severe acute respiratory syndrome coro    ....................  ....................  .....................  ....................  ....................  ....................
4552 MW277126.1 |Severe acute respiratory syndrome coro    ....................  ....................  .....................  ....................  ....................  ....................
4553 MW277127.1 |Severe acute respiratory syndrome coro    ....................  ....................  .....................  ....................  ....................  ....................
4554 MW277129.1 |Severe acute respiratory syndrome coro    ....................  ....................  .....................  ....................  ....................  ....................
4555 MW277131.1 |Severe acute respiratory syndrome coro    ....................  ....................  .....................  ....................  ....................  ....................
4556 MW277132.1 |Severe acute respiratory syndrome coro    ....................  ....................  .....................  ....................  ....................  ....................
4557 MW277133.1 |Severe acute respiratory syndrome coro    ....................  ....................  .....................  ....................  ....................  ....................
4558 MW277134.1 |Severe acute respiratory syndrome coro    ....................  ....................  .....................  ....................  ....................  ....................
4559 MW277135.1 |Severe acute respiratory syndrome coro    ....................  ....................  .....................  ....................  ....................  ....................
4560 MW277136.1 |Severe acute respiratory syndrome coro    ....................  ....................  .....................  ....................  ....................  ....................
4561 MW277138.1 |Severe acute respiratory syndrome coro    ....................  ....................  .....................  ....................  ....................  ....................
4562 MW277141.1 |Severe acute respiratory syndrome coro    ....................  ....................  .....................  ....................  ....................  ....................
4564 MW277145.1 |Severe acute respiratory syndrome coro    ....................  ....................  .....................  ....................  ....................  ....................
4565 MW277146.1 |Severe acute respiratory syndrome coro    ....................  ....................  .....................  ....................  ....................  ....................
4568 MW277150.1 |Severe acute respiratory syndrome coro    ....................  ....................  .....................  ....................  ....................  ....................
4569 MW277152.1 |Severe acute respiratory syndrome coro    ....................  ....................  .....................  ....................  ....................  ....................
4570 MW277153.1 |Severe acute respiratory syndrome coro    ....................  ....................  .....................  ....................  ....................  ....................
4571 MW277154.1 |Severe acute respiratory syndrome coro    ....................  ....................  .....................  ....................  ....................  ....................
4572 MW277156.1 |Severe acute respiratory syndrome coro    ....................  ....................  .....................  ....................  ....................  ....................
4573 MW277157.1 |Severe acute respiratory syndrome coro    ....................  ....................  .....................  ....................  ....................  ....................
4574 MW277158.1 |Severe acute respiratory syndrome coro    ....................  ....................  .....................  ....................  ....................  ....................
4575 MW277160.1 |Severe acute respiratory syndrome coro    ....................  ....................  .....................  ....................  ....................  ....................
4576 MW277161.1 |Severe acute respiratory syndrome coro    ....................  ....................  .....................  ....................  ....................  ....................
4577 MW277162.1 |Severe acute respiratory syndrome coro    ....................  ....................  .....................  ....................  ....................  ....................
4578 MW277164.1 |Severe acute respiratory syndrome coro    ....................  ....................  .....................  ....................  ....................  ....................
4579 MW277166.1 |Severe acute respiratory syndrome coro    ....................  ....................  .....................  ....................  ....................  ....................
4580 MW277167.1 |Severe acute respiratory syndrome coro    ....................  ....................  .....................  ....................  ....................  ....................
4581 MW277168.1 |Severe acute respiratory syndrome coro    ....................  ....................  .....................  ....................  ....................  ....................
4582 MW277169.1 |Severe acute respiratory syndrome coro    ....................  ....................  .....................  ....................  ....................  ....................
4583 MW277170.1 |Severe acute respiratory syndrome coro    ....................  ....................  .....................  ....................  ....................  ....................
4584 MW277173.1 |Severe acute respiratory syndrome coro    ....................  ....................  .....................  ....................  ....................  ....................
4585 MW277174.1 |Severe acute respiratory syndrome coro    ....................  ....................  .....................  ....................  ....................  ....................
4586 MW277175.1 |Severe acute respiratory syndrome coro    ....................  ....................  .....................  ....................  ....................  ....................
4587 MW277176.1 |Severe acute respiratory syndrome coro    ....................  ....................  .....................  ....................  ....................  ....................
4588 MW277177.1 |Severe acute respiratory syndrome coro    ....................  ....................  .....................  ....................  ....................  ....................
4589 MW277179.1 |Severe acute respiratory syndrome coro    ....................  ....................  .....................  ....................  ....................  ....................
4590 MW277180.1 |Severe acute respiratory syndrome coro    ....................  ....................  .....................  ....................  ....................  ....................
4591 MW277181.1 |Severe acute respiratory syndrome coro    ....................  ....................  .....................  ....................  ....................  ....................
4592 MW277184.1 |Severe acute respiratory syndrome coro    ....................  ....................  .....................  ....................  ....................  ....................
4593 MW277185.1 |Severe acute respiratory syndrome coro    ....................  ....................  .....................  ....................  ....................  ....................
4594 MW277187.1 |Severe acute respiratory syndrome coro    ....................  ....................  .....................  ....................  ....................  ....................
4595 MW277188.1 |Severe acute respiratory syndrome coro    ....................  ....................  .....................  ....................  ....................  ....................
4596 MW277189.1 |Severe acute respiratory syndrome coro    ....................  ....................  .....................  ....................  ....................  ....................
4597 MW277190.1 |Severe acute respiratory syndrome coro    ....................  ....................  .....................  ....................  ....................  ....................
4598 MW277191.1 |Severe acute respiratory syndrome coro    ....................  ....................  .....................  ....................  ....................  ....................
4599 MW277192.1 |Severe acute respiratory syndrome coro    ....................  ....................  .....................  ....................  ....................  ....................
4600 MW277193.1 |Severe acute respiratory syndrome coro    ....................  ....................  .....................  ....................  ....................  ....................
4601 MW277194.1 |Severe acute respiratory syndrome coro    ....................  ....................  .....................  ....................  ....................  ....................
4603 MW277198.1 |Severe acute respiratory syndrome coro    ....................  ....................  .....................  ....................  ....................  ....................
4604 MW277199.1 |Severe acute respiratory syndrome coro    ....................  ....................  .....................  ....................  ....................  ....................
4605 MW277200.1 |Severe acute respiratory syndrome coro    ....................  ....................  .....................  ....................  ....................  ....................
4606 MW277201.1 |Severe acute respiratory syndrome coro    ....................  ....................  .....................  ....................  ....................  ....................
4607 MW277202.1 |Severe acute respiratory syndrome coro    ....................  ....................  .....................  ....................  ....................  ....................
4608 MW277203.1 |Severe acute respiratory syndrome coro    ....................  ....................  .....................  ....................  ....................  ....................
4609 MW277205.1 |Severe acute respiratory syndrome coro    ....................  ....................  .....................  ....................  ....................  ....................
4610 MW277206.1 |Severe acute respiratory syndrome coro    ....................  ....................  .....................  ....................  ....................  ....................
4611 MW277207.1 |Severe acute respiratory syndrome coro    ....................  ....................  .....................  ....................  ....................  ....................
4612 MW277208.1 |Severe acute respiratory syndrome coro    ....................  ....................  .....................  ....................  ....................  ....................
4613 MW277209.1 |Severe acute respiratory syndrome coro    ....................  ....................  .....................  ....................  ....................  ....................
4614 MW277210.1 |Severe acute respiratory syndrome coro    ....................  ....................  .....................  ....................  ....................  ....................
4615 MW277211.1 |Severe acute respiratory syndrome coro    ....................  ....................  .....................  ....................  ....................  ....................
4616 MW277212.1 |Severe acute respiratory syndrome coro    ....................  ....................  .....................  ....................  ....................  ....................
4617 MW277213.1 |Severe acute respiratory syndrome coro    ....................  ....................  .....................  ....................  ....................  ....................
4618 MW277214.1 |Severe acute respiratory syndrome coro    ....................  ....................  .....................  ....................  ....................  ....................
4619 MW277215.1 |Severe acute respiratory syndrome coro    ....................  ....................  .....................  ....................  ....................  ....................
4620 MW277217.1 |Severe acute respiratory syndrome coro    ....................  ....................  .....................  ....................  ....................  ....................
4621 MW277218.1 |Severe acute respiratory syndrome coro    ....................  ....................  .....................  ....................  ....................  ....................
4622 MW277219.1 |Severe acute respiratory syndrome coro    ....................  ....................  .....................  ....................  ....................  ....................
4623 MW277220.1 |Severe acute respiratory syndrome coro    ....................  ....................  .....................  ....................  ....................  ....................
4624 MW277221.1 |Severe acute respiratory syndrome coro    ....................  ....................  .....................  ....................  ....................  ....................
4625 MW277224.1 |Severe acute respiratory syndrome coro    ....................  ....................  .....................  ....................  ....................  ....................
```

4626 MW277227.1 |Severe acute respiratory syndrome coro ···················· ···················· ····················· ···················· ···················· ····················
4627 MW277230.1 |Severe acute respiratory syndrome coro ···················· ···················· ····················· ···················· ···················· ····················
4628 MW277232.1 |Severe acute respiratory syndrome coro ···················· ···················· ····················· ···················· ···················· ····················
4629 MW277235.1 |Severe acute respiratory syndrome coro ···················· ···················· ····················· ···················· ···················· ····················
4630 MW277236.1 |Severe acute respiratory syndrome coro ···················· ···················· ····················· ···················· ···················· ····················
4631 MW277237.1 |Severe acute respiratory syndrome coro ···················· ···················· ····················· ···················· ···················· ····················
4632 MW277240.1 |Severe acute respiratory syndrome coro ···················· ···················· ····················· ···················· ···················· ····················
4633 MW277242.1 |Severe acute respiratory syndrome coro ···················· ···················· ····················· ···················· ···················· ····················
4634 MW277243.1 |Severe acute respiratory syndrome coro ···················· ···················· ····················· ···················· ···················· ····················
4635 MW277244.1 |Severe acute respiratory syndrome coro ···················· ···················· ····················· ···················· ···················· ····················
4636 MW277245.1 |Severe acute respiratory syndrome coro ···················· ···················· ····················· ···················· ···················· ····················
4637 MW277246.1 |Severe acute respiratory syndrome coro ···················· ···················· ····················· ···················· ···················· ····················
4638 MW277247.1 |Severe acute respiratory syndrome coro ···················· ···················· ····················· ···················· ···················· ····················
4639 MW277248.1 |Severe acute respiratory syndrome coro ···················· ···················· ····················· ···················· ···················· ····················
4640 MW277249.1 |Severe acute respiratory syndrome coro ···················· ···················· ····················· ···················· ···················· ····················
4641 MW277250.1 |Severe acute respiratory syndrome coro ···················· ···················· ····················· ···················· ···················· ····················
4642 MW277251.1 |Severe acute respiratory syndrome coro ···················· ···················· ····················· ···················· ···················· ····················
4643 MW277252.1 |Severe acute respiratory syndrome coro ···················· ···················· ····················· ···················· ···················· ····················
4644 MW277253.1 |Severe acute respiratory syndrome coro ···················· ···················· ····················· ···················· ···················· ····················
4645 MW277254.1 |Severe acute respiratory syndrome coro ···················· ···················· ····················· ···················· ···················· ····················
4646 MW277255.1 |Severe acute respiratory syndrome coro ···················· ···················· ····················· ···················· ···················· ····················
4647 MW277256.1 |Severe acute respiratory syndrome coro ···················· ···················· ····················· ···················· ···················· ····················
4648 MW277257.1 |Severe acute respiratory syndrome coro ···················· ···················· ····················· ···················· ···················· ····················
4649 MW277258.1 |Severe acute respiratory syndrome coro ···················· ···················· ····················· ···················· ···················· ····················
4650 MW277259.1 |Severe acute respiratory syndrome coro ···················· ···················· ····················· ···················· ···················· ····················
4651 MW277260.1 |Severe acute respiratory syndrome coro ···················· ···················· ····················· ···················· ···················· ····················
4652 MW277261.1 |Severe acute respiratory syndrome coro ···················· ···················· ····················· ···················· ···················· ····················
4653 MW277262.1 |Severe acute respiratory syndrome coro ···················· ···················· ····················· ···················· ···················· ····················
4654 MW277263.1 |Severe acute respiratory syndrome coro ···················· ···················· ····················· ···················· ···················· ····················
4655 MW277264.1 |Severe acute respiratory syndrome coro ···················· ···················· ····················· ···················· ···················· ····················
4656 MW277265.1 |Severe acute respiratory syndrome coro ···················· ···················· ····················· ···················· ···················· ····················
4657 MW277267.1 |Severe acute respiratory syndrome coro ···················· ···················· ····················· ···················· ···················· ····················
4658 MW277269.1 |Severe acute respiratory syndrome coro ···················· ···················· ····················· ···················· ···················· ····················
4659 MW277271.1 |Severe acute respiratory syndrome coro ···················· ···················· ····················· ···················· ···················· ····················
4660 MW277272.1 |Severe acute respiratory syndrome coro ···················· ······A············· ····················· ···················· ···················· ····················
4661 MW277273.1 |Severe acute respiratory syndrome coro ···················· ···················· ····················· ···················· ···················· ····················
4662 MW277274.1 |Severe acute respiratory syndrome coro ···················· ···················· ····················· ···················· ···················· ····················
4663 MW277275.1 |Severe acute respiratory syndrome coro ···················· ···················· ····················· ···················· ···················· ····················
4664 MW277276.1 |Severe acute respiratory syndrome coro ···················· ···················· ····················· ···················· ···················· ····················
4665 MW277277.1 |Severe acute respiratory syndrome coro ···················· ···················· ····················· ···················· ···················· ····················
4666 MW277279.1 |Severe acute respiratory syndrome coro ···················· ···················· ····················· ···················· ···················· ····················
4667 MW277281.1 |Severe acute respiratory syndrome coro ···················· ···················· ····················· ···················· ···················· ····················
4668 MW277282.1 |Severe acute respiratory syndrome coro ···················· ···················· ····················· ···················· ···················· ····················
4669 MW277283.1 |Severe acute respiratory syndrome coro ···················· ···················· ····················· ···················· ···················· ····················
4670 MW277284.1 |Severe acute respiratory syndrome coro ···················· ···················· ····················· ···················· ···················· ····················
4671 MW277285.1 |Severe acute respiratory syndrome coro ···················· ······A············· ····················· ···················· ···················· ····················
4672 MW277286.1 |Severe acute respiratory syndrome coro ···················· ···················· ····················· ···················· ···················· ····················
4673 MW277287.1 |Severe acute respiratory syndrome coro ···················· ···················· ····················· ···················· ···················· ····················
4674 MW277289.1 |Severe acute respiratory syndrome coro ···················· ···················· ····················· ···················· ···················· ····················
4675 MW277290.1 |Severe acute respiratory syndrome coro ···················· ···················· ····················· ···················· ···················· ····················
4676 MW277291.1 |Severe acute respiratory syndrome coro ···················· ···················· ····················· ···················· ···················· ····················
4677 MW277292.1 |Severe acute respiratory syndrome coro ···················· ···················· ····················· ···················· ···················· ····················
4678 MW277293.1 |Severe acute respiratory syndrome coro ···················· ···················· ····················· ···················· ···················· ····················
4679 MW277294.1 |Severe acute respiratory syndrome coro ···················· ···················· ····················· ···················· ···················· ····················
4680 MW277297.1 |Severe acute respiratory syndrome coro ···················· ···················· ····················· ···················· ···················· ····················
4681 MW277299.1 |Severe acute respiratory syndrome coro ···················· ···················· ····················· ···················· ···················· ····················
4682 MW277300.1 |Severe acute respiratory syndrome coro ···················· ···················· ····················· ···················· ···················· ····················
4683 MW277301.1 |Severe acute respiratory syndrome coro ···················· ···················· ····················· ···················· ···················· ····················
4684 MW277302.1 |Severe acute respiratory syndrome coro ···················· ···················· ····················· ···················· ···················· ····················
4685 MW277303.1 |Severe acute respiratory syndrome coro ···················· ···················· ····················· ···················· ···················· ····················
4686 MW277304.1 |Severe acute respiratory syndrome coro ···················· ···················· ····················· ···················· ···················· ····················
4687 MW277306.1 |Severe acute respiratory syndrome coro ···················· ···················· ····················· ···················· ···················· ····················
4688 MW277307.1 |Severe acute respiratory syndrome coro ···················· ···················· ····················· ···················· ···················· ····················
4689 MW277308.1 |Severe acute respiratory syndrome coro ···················· ···················· ····················· ···················· ···················· ····················
4690 MW277309.1 |Severe acute respiratory syndrome coro ···················· ···················· ····················· ···················· ···················· ····················
4691 MW277311.1 |Severe acute respiratory syndrome coro ···················· ······A············· ····················· ···················· ···················· ····················
4692 MW277312.1 |Severe acute respiratory syndrome coro ···················· ···················· ····················· ···················· ···················· ····················
4693 MW277313.1 |Severe acute respiratory syndrome coro ···················· ···················· ····················· ···················· ···················· ····················
4694 MW277314.1 |Severe acute respiratory syndrome coro ···················· ···················· ····················· ···················· ···················· ····················
4696 MW277316.1 |Severe acute respiratory syndrome coro ···················· ···················· ····················· ···················· ···················· ····················
4697 MW277317.1 |Severe acute respiratory syndrome coro ···················· ···················· ····················· ···················· ···················· ····················
4698 MW277318.1 |Severe acute respiratory syndrome coro ···················· ···················· ····················· ···················· ···················· ····················
4699 MW277319.1 |Severe acute respiratory syndrome coro ···················· ···················· ····················· ···················· ···················· ····················
4700 MW277320.1 |Severe acute respiratory syndrome coro ···················· ···················· ····················· ···················· ···················· ····················
4701 MW277321.1 |Severe acute respiratory syndrome coro ···················· ···················· ····················· ···················· ···················· ····················
4702 MW277322.1 |Severe acute respiratory syndrome coro ···················· ···················· ····················· ···················· ···················· ····················
4703 MW277323.1 |Severe acute respiratory syndrome coro ···················· ···················· ····················· ···················· ···················· ····················
4704 MW277324.1 |Severe acute respiratory syndrome coro ···················· ···················· ····················· ···················· ···················· ····················

| | | | | | | | |
|---|---|---|---|---|---|---|---|
| 4705 | MW277325.1 \|Severe acute respiratory syndrome coro | .................... | .................... | ..................... | .................... | .................... | .................... |
| 4706 | MW277327.1 \|Severe acute respiratory syndrome coro | .................... | .................... | ..................... | .................... | .................... | .................... |
| 4707 | MW277328.1 \|Severe acute respiratory syndrome coro | .................... | .................... | ..................... | .................... | .................... | .................... |
| 4708 | MW277329.1 \|Severe acute respiratory syndrome coro | .................... | .................... | ..................... | .................... | .................... | .................... |
| 4709 | MW277330.1 \|Severe acute respiratory syndrome coro | .................... | .................... | ..................... | .................... | .................... | .................... |
| 4710 | MW277331.1 \|Severe acute respiratory syndrome coro | .................... | .................... | ..................... | .................... | .................... | .................... |
| 4711 | MW277332.1 \|Severe acute respiratory syndrome coro | .................... | .................... | ..................... | .................... | .................... | .................... |
| 4712 | MW277334.1 \|Severe acute respiratory syndrome coro | .................... | .................... | ..................... | .................... | .................... | .................... |
| 4713 | MW277336.1 \|Severe acute respiratory syndrome coro | .................... | .................... | ..................... | .................... | .................... | .................... |
| 4714 | MW277337.1 \|Severe acute respiratory syndrome coro | .................... | .................... | ..................... | .................... | .................... | .................... |
| 4715 | MW277338.1 \|Severe acute respiratory syndrome coro | .................... | .................... | ..................... | .................... | .................... | .................... |
| 4716 | MW277339.1 \|Severe acute respiratory syndrome coro | .................... | .................... | ..................... | .................... | .................... | .................... |
| 4717 | MW277340.1 \|Severe acute respiratory syndrome coro | .................... | .................... | ..................... | .................... | .................... | .................... |
| 4718 | MW277341.1 \|Severe acute respiratory syndrome coro | .................... | .................... | ..................... | .................... | .................... | .................... |
| 4719 | MW277343.1 \|Severe acute respiratory syndrome coro | .................... | .................... | ..................... | .................... | .................... | .................... |
| 4720 | MW277344.1 \|Severe acute respiratory syndrome coro | .................... | .................... | ..................... | .................... | .................... | .................... |
| 4721 | MW277345.1 \|Severe acute respiratory syndrome coro | .................... | .................... | ..................... | .................... | .................... | .................... |
| 4722 | MW277346.1 \|Severe acute respiratory syndrome coro | .................... | .................... | ..................... | .................... | .................... | .................... |
| 4723 | MW277347.1 \|Severe acute respiratory syndrome coro | .................... | .................... | ..................... | .................... | .................... | .................... |
| 4724 | MW277348.1 \|Severe acute respiratory syndrome coro | .................... | .................... | ..................... | .................... | .................... | .................... |
| 4725 | MW277351.1 \|Severe acute respiratory syndrome coro | .................... | .................... | ..................... | .................... | .................... | .................... |
| 4726 | MW277352.1 \|Severe acute respiratory syndrome coro | .................... | .................... | ..................... | .................... | .................... | .................... |
| 4727 | MW277354.1 \|Severe acute respiratory syndrome coro | .................... | .................... | ..................... | .................... | .................... | .................... |
| 4728 | MW277355.1 \|Severe acute respiratory syndrome coro | .................... | .................... | ..................... | .................... | .................... | .................... |
| 4729 | MW277356.1 \|Severe acute respiratory syndrome coro | .................... | .................... | ..................... | .................... | .................... | .................... |
| 4730 | MW277357.1 \|Severe acute respiratory syndrome coro | .................... | .................... | ..................... | .................... | .................... | .................... |
| 4731 | MW277359.1 \|Severe acute respiratory syndrome coro | .................... | .................... | ..................... | .................... | .................... | .................... |
| 4732 | MW277361.1 \|Severe acute respiratory syndrome coro | .................... | .................... | ..................... | .................... | .................... | .................... |
| 4733 | MW277362.1 \|Severe acute respiratory syndrome coro | .................... | .................... | ..................... | .................... | .................... | .................... |
| 4734 | MW277363.1 \|Severe acute respiratory syndrome coro | .................... | .................... | ..................... | .................... | .................... | .................... |
| 4735 | MW277364.1 \|Severe acute respiratory syndrome coro | .................... | .................... | ..................... | .................... | .................... | .................... |
| 4736 | MW277367.1 \|Severe acute respiratory syndrome coro | .................... | .................... | ..................... | .................... | .................... | .................... |
| 4737 | MW277368.1 \|Severe acute respiratory syndrome coro | .................... | .................... | ..................... | .................... | .................... | .................... |
| 4738 | MW277369.1 \|Severe acute respiratory syndrome coro | .................... | .................... | ..................... | .................... | .................... | .................... |
| 4739 | MW277371.1 \|Severe acute respiratory syndrome coro | .................... | .................... | ..................... | .................... | .................... | .................... |
| 4740 | MW277372.1 \|Severe acute respiratory syndrome coro | .................... | .................... | ..................... | .................... | .................... | .................... |
| 4741 | MW277373.1 \|Severe acute respiratory syndrome coro | .................... | .................... | ..................... | .................... | .................... | .................... |
| 4742 | MW277376.1 \|Severe acute respiratory syndrome coro | .................... | .................... | ..................... | .................... | .................... | .................... |
| 4743 | MW277378.1 \|Severe acute respiratory syndrome coro | .................... | .................... | ..................... | .................... | .................... | .................... |
| 4744 | MW277379.1 \|Severe acute respiratory syndrome coro | .................... | .................... | ..................... | .................... | .................... | .................... |
| 4745 | MW277380.1 \|Severe acute respiratory syndrome coro | .................... | .................... | ..................... | .................... | .................... | .................... |
| 4746 | MW277381.1 \|Severe acute respiratory syndrome coro | .................... | .................... | ..................... | .................... | .................... | .................... |
| 4747 | MW277382.1 \|Severe acute respiratory syndrome coro | .................... | .................... | ..................... | .................... | .................... | .................... |
| 4748 | MW277385.1 \|Severe acute respiratory syndrome coro | .................... | .................... | ..................... | .................... | .................... | .................... |
| 4749 | MW277386.1 \|Severe acute respiratory syndrome coro | .................... | .................... | ..................... | .................... | .................... | .................... |
| 4751 | MW277388.1 \|Severe acute respiratory syndrome coro | .................... | .................... | ..................... | .................... | .................... | .................... |
| 4752 | MW277389.1 \|Severe acute respiratory syndrome coro | .................... | .................... | ..................... | .................... | .................... | .................... |
| 4753 | MW277390.1 \|Severe acute respiratory syndrome coro | .................... | .................... | ..................... | .................... | .................... | .................... |
| 4754 | MW277391.1 \|Severe acute respiratory syndrome coro | .................... | .................... | ..................... | .................... | .................... | .................... |
| 4755 | MW277392.1 \|Severe acute respiratory syndrome coro | .................... | .................... | ..................... | .................... | .................... | .................... |
| 4756 | MW277393.1 \|Severe acute respiratory syndrome coro | .................... | .................... | ..................... | .................... | .................... | .................... |
| 4757 | MW277394.1 \|Severe acute respiratory syndrome coro | .................... | .................... | ..................... | .................... | .................... | .................... |
| 4758 | MW277395.1 \|Severe acute respiratory syndrome coro | .................... | .................... | ..................... | .................... | .................... | .................... |
| 4759 | MW277396.1 \|Severe acute respiratory syndrome coro | .................... | .................... | ..................... | .................... | .................... | .................... |
| 4760 | MW277397.1 \|Severe acute respiratory syndrome coro | .................... | .................... | ..................... | .................... | .................... | .................... |
| 4761 | MW277398.1 \|Severe acute respiratory syndrome coro | .................... | .................... | ..................... | .................... | .................... | .................... |
| 4762 | MW277400.1 \|Severe acute respiratory syndrome coro | .................... | .................... | ..................... | .................... | .................... | .................... |
| 4763 | MW277402.1 \|Severe acute respiratory syndrome coro | .................... | .................... | ..................... | .................... | .................... | .................... |
| 4764 | MW277403.1 \|Severe acute respiratory syndrome coro | .................... | .................... | ..................... | .................... | .................... | .................... |
| 4765 | MW277404.1 \|Severe acute respiratory syndrome coro | .................... | .................... | ..................... | .................... | .................... | .................... |
| 4767 | MW277406.1 \|Severe acute respiratory syndrome coro | .................... | .................... | ..................... | .................... | .................... | .................... |
| 4768 | MW277408.1 \|Severe acute respiratory syndrome coro | .................... | .................... | ..................... | .................... | .................... | .................... |
| 4769 | MW277409.1 \|Severe acute respiratory syndrome coro | .................... | .................... | ..................... | .................... | .................... | .................... |
| 4770 | MW277410.1 \|Severe acute respiratory syndrome coro | .................... | .................... | ..................... | .................... | .................... | .................... |
| 4771 | MW277411.1 \|Severe acute respiratory syndrome coro | .................... | .................... | ..................... | .................... | .................... | .................... |
| 4772 | MW277413.1 \|Severe acute respiratory syndrome coro | .................... | .................... | ..................... | .................... | .................... | .................... |
| 4773 | MW277414.1 \|Severe acute respiratory syndrome coro | .................... | .................... | ..................... | .................... | .................... | .................... |
| 4774 | MW277416.1 \|Severe acute respiratory syndrome coro | .................... | .................... | ..................... | .................... | .................... | .................... |
| 4775 | MW277418.1 \|Severe acute respiratory syndrome coro | .................... | .................... | ..................... | .................... | .................... | .................... |
| 4776 | MW277419.1 \|Severe acute respiratory syndrome coro | .................... | .................... | ..................... | .................... | .................... | .................... |
| 4777 | MW277421.1 \|Severe acute respiratory syndrome coro | .................... | .................... | ..................... | .................... | .................... | .................... |
| 4778 | MW277422.1 \|Severe acute respiratory syndrome coro | .................... | .................... | ..................... | .................... | .................... | .................... |
| 4779 | MW277423.1 \|Severe acute respiratory syndrome coro | .................... | .................... | ..................... | .................... | .................... | .................... |
| 4780 | MW277424.1 \|Severe acute respiratory syndrome coro | .................... | .................... | ..................... | .................... | .................... | .................... |
| 4781 | MW277426.1 \|Severe acute respiratory syndrome coro | .................... | .................... | ..................... | .................... | .................... | .................... |
| 4782 | MW277427.1 \|Severe acute respiratory syndrome coro | .................... | .................... | ..................... | .................... | .................... | .................... |
| 4783 | MW277428.1 \|Severe acute respiratory syndrome coro | .................... | .................... | ..................... | .................... | .................... | .................... |
| 4784 | MW277429.1 \|Severe acute respiratory syndrome coro | .................... | .................... | ..................... | .................... | .................... | .................... |

| | | | | | | | |
|---|---|---|---|---|---|---|---|
| 4785 | MW277430.1 \|Severe acute respiratory syndrome coro | .................... | .................... | .................... | .................... | .................... | .................... |
| 4786 | MW277431.1 \|Severe acute respiratory syndrome coro | .................... | .................... | .................... | .................... | .................... | .................... |
| 4787 | MW277432.1 \|Severe acute respiratory syndrome coro | .................... | .................... | .................... | .................... | .................... | .................... |
| 4788 | MW277433.1 \|Severe acute respiratory syndrome coro | .................... | .................... | .................... | .................... | .................... | .................... |
| 4790 | MW277437.1 \|Severe acute respiratory syndrome coro | .................... | .................... | .................... | .................... | .................... | .................... |
| 4791 | MW277438.1 \|Severe acute respiratory syndrome coro | .................... | .................... | .................... | .................... | .................... | .................... |
| 4792 | MW277439.1 \|Severe acute respiratory syndrome coro | .................... | .................... | .................... | .................... | .................... | .................... |
| 4793 | MW277440.1 \|Severe acute respiratory syndrome coro | .................... | .................... | .................... | .................... | .................... | .................... |
| 4794 | MW277442.1 \|Severe acute respiratory syndrome coro | .................... | .................... | .................... | .................... | .................... | .................... |
| 4795 | MW277443.1 \|Severe acute respiratory syndrome coro | .................... | .................... | .................... | .................... | .................... | .................... |
| 4796 | MW277446.1 \|Severe acute respiratory syndrome coro | .................... | .................... | .................... | .................... | .................... | .................... |
| 4797 | MW277447.1 \|Severe acute respiratory syndrome coro | .................... | .................... | .................... | .................... | .................... | .................... |
| 4798 | MW277448.1 \|Severe acute respiratory syndrome coro | .................... | .................... | .................... | .................... | .................... | .................... |
| 4799 | MW277449.1 \|Severe acute respiratory syndrome coro | .................... | .................... | .................... | .................... | .................... | .................... |
| 4800 | MW277450.1 \|Severe acute respiratory syndrome coro | .................... | .................... | .................... | .................... | .................... | .................... |
| 4801 | MW277452.1 \|Severe acute respiratory syndrome coro | .................... | .................... | .................... | .................... | .................... | .................... |
| 4802 | MW277454.1 \|Severe acute respiratory syndrome coro | .................... | .................... | .................... | .................... | .................... | .................... |
| 4803 | MW277455.1 \|Severe acute respiratory syndrome coro | .................... | .................... | .................... | .................... | .................... | .................... |
| 4804 | MW277456.1 \|Severe acute respiratory syndrome coro | .................... | .................... | .................... | .................... | .................... | .................... |
| 4805 | MW277457.1 \|Severe acute respiratory syndrome coro | .................... | .................... | .................... | .................... | .................... | .................... |
| 4806 | MW277458.1 \|Severe acute respiratory syndrome coro | .................... | .................... | .................... | .................... | .................... | .................... |
| 4807 | MW277459.1 \|Severe acute respiratory syndrome coro | .................... | .................... | .................... | .................... | .................... | .................... |
| 4808 | MW277460.1 \|Severe acute respiratory syndrome coro | .................... | .................... | .................... | .................... | .................... | .................... |
| 4809 | MW277462.1 \|Severe acute respiratory syndrome coro | .................... | .................... | .................... | .................... | .................... | .................... |
| 4810 | MW277463.1 \|Severe acute respiratory syndrome coro | .................... | .................... | .................... | .................... | .................... | .................... |
| 4811 | MW277464.1 \|Severe acute respiratory syndrome coro | .................... | .................... | .................... | .................... | .................... | .................... |
| 4812 | MW277465.1 \|Severe acute respiratory syndrome coro | .................... | .................... | .................... | .................... | .................... | .................... |
| 4813 | MW277466.1 \|Severe acute respiratory syndrome coro | .................... | .................... | .................... | .................... | .................... | .................... |
| 4814 | MW277467.1 \|Severe acute respiratory syndrome coro | .................... | .................... | .................... | .................... | .................... | .................... |
| 4815 | MW277468.1 \|Severe acute respiratory syndrome coro | .................... | .................... | .................... | .................... | .................... | .................... |
| 4816 | MW277470.1 \|Severe acute respiratory syndrome coro | .................... | .................... | .................... | .................... | .................... | .................... |
| 4817 | MW277471.1 \|Severe acute respiratory syndrome coro | .................... | .................... | .................... | .................... | .................... | .................... |
| 4818 | MW277472.1 \|Severe acute respiratory syndrome coro | .................... | .................... | .................... | .................... | .................... | .................... |
| 4819 | MW277474.1 \|Severe acute respiratory syndrome coro | .................... | .................... | .................... | .................... | .................... | .................... |
| 4821 | MW277476.1 \|Severe acute respiratory syndrome coro | .................... | .................... | .................... | .................... | .................... | .................... |
| 4822 | MW277477.1 \|Severe acute respiratory syndrome coro | .................... | .................... | .................... | .................... | .................... | .................... |
| 4823 | MW277478.1 \|Severe acute respiratory syndrome coro | .................... | .................... | .................... | .................... | .................... | .................... |
| 4824 | MW277480.1 \|Severe acute respiratory syndrome coro | .................... | .................... | .................... | .................... | .................... | .................... |
| 4825 | MW277481.1 \|Severe acute respiratory syndrome coro | .................... | .................... | .................... | .................... | .................... | .................... |
| 4826 | MW277483.1 \|Severe acute respiratory syndrome coro | .................... | .................... | .................... | .................... | .................... | .................... |
| 4827 | MW277484.1 \|Severe acute respiratory syndrome coro | .................... | .................... | .................... | .................... | .................... | .................... |
| 4828 | MW277485.1 \|Severe acute respiratory syndrome coro | .................... | .................... | .................... | .................... | .................... | .................... |
| 4829 | MW277486.1 \|Severe acute respiratory syndrome coro | .................... | .................... | .................... | .................... | .................... | .................... |
| 4830 | MW277487.1 \|Severe acute respiratory syndrome coro | .................... | .................... | .................... | .................... | .................... | .................... |
| 4831 | MW277488.1 \|Severe acute respiratory syndrome coro | .................... | .................... | .................... | .................... | .................... | .................... |
| 4832 | MW277490.1 \|Severe acute respiratory syndrome coro | .................... | .................... | .................... | .................... | .................... | .................... |
| 4833 | MW277491.1 \|Severe acute respiratory syndrome coro | .................... | .................... | .................... | .................... | .................... | .................... |
| 4834 | MW277492.1 \|Severe acute respiratory syndrome coro | .................... | .................... | .................... | .................... | .................... | .................... |
| 4835 | MW277493.1 \|Severe acute respiratory syndrome coro | .................... | .................... | .................... | .................... | .................... | .................... |
| 4836 | MW277494.1 \|Severe acute respiratory syndrome coro | .................... | .................... | .................... | .................... | .................... | .................... |
| 4837 | MW277495.1 \|Severe acute respiratory syndrome coro | .................... | .................... | .................... | .................... | .................... | .................... |
| 4838 | MW277496.1 \|Severe acute respiratory syndrome coro | .................... | .................... | .................... | .................... | .................... | .................... |
| 4839 | MW277497.1 \|Severe acute respiratory syndrome coro | .................... | .................... | .................... | .................... | .................... | .................... |
| 4840 | MW277498.1 \|Severe acute respiratory syndrome coro | .................... | .................... | .................... | .................... | .................... | .................... |
| 4841 | MW277499.1 \|Severe acute respiratory syndrome coro | .................... | .................... | .................... | .................... | .................... | .................... |
| 4842 | MW277500.1 \|Severe acute respiratory syndrome coro | .................... | .................... | .................... | .................... | .................... | .................... |
| 4843 | MW277501.1 \|Severe acute respiratory syndrome coro | .................... | .................... | .................... | .................... | .................... | .................... |
| 4844 | MW277502.1 \|Severe acute respiratory syndrome coro | .................... | .................... | .................... | .................... | .................... | .................... |
| 4845 | MW277503.1 \|Severe acute respiratory syndrome coro | .................... | .................... | .................... | .................... | .................... | .................... |
| 4846 | MW277506.1 \|Severe acute respiratory syndrome coro | .................... | .................... | .................... | .................... | .................... | .................... |
| 4847 | MW277507.1 \|Severe acute respiratory syndrome coro | .................... | .................... | .................... | .................... | .................... | .................... |
| 4848 | MW277508.1 \|Severe acute respiratory syndrome coro | .................... | .................... | .................... | .................... | .................... | .................... |
| 4850 | MW277510.1 \|Severe acute respiratory syndrome coro | .................... | .................... | .................... | .................... | .................... | .................... |
| 4851 | MW277511.1 \|Severe acute respiratory syndrome coro | .................... | .................... | .................... | .................... | .................... | .................... |
| 4852 | MW277512.1 \|Severe acute respiratory syndrome coro | .................... | .................... | .................... | .................... | .................... | .................... |
| 4853 | MW277513.1 \|Severe acute respiratory syndrome coro | .................... | .................... | .................... | .................... | .................... | .................... |
| 4854 | MW277514.1 \|Severe acute respiratory syndrome coro | .................... | .................... | .................... | .................... | .................... | .................... |
| 4855 | MW277515.1 \|Severe acute respiratory syndrome coro | .................... | .................... | .................... | .................... | .................... | .................... |
| 4856 | MW277516.1 \|Severe acute respiratory syndrome coro | .................... | .................... | .................... | .................... | .................... | .................... |
| 4857 | MW277517.1 \|Severe acute respiratory syndrome coro | .................... | .................... | .................... | .................... | .................... | .................... |
| 4858 | MW277518.1 \|Severe acute respiratory syndrome coro | .................... | .................... | .................... | .................... | .................... | .................... |
| 4859 | MW277519.1 \|Severe acute respiratory syndrome coro | .................... | .................... | .................... | .................... | .................... | .................... |
| 4860 | MW277520.1 \|Severe acute respiratory syndrome coro | .................... | .................... | .................... | .................... | .................... | .................... |
| 4861 | MW277521.1 \|Severe acute respiratory syndrome coro | .................... | .................... | .................... | .................... | .................... | .................... |
| 4862 | MW277522.1 \|Severe acute respiratory syndrome coro | .................... | .................... | .................... | .................... | .................... | .................... |
| 4863 | MW277525.1 \|Severe acute respiratory syndrome coro | .................... | .................... | .................... | .................... | .................... | .................... |
| 4864 | MW277528.1 \|Severe acute respiratory syndrome coro | .................... | .................... | .................... | .................... | .................... | .................... |
| 4865 | MW277529.1 \|Severe acute respiratory syndrome coro | .................... | .................... | .................... | .................... | .................... | .................... |

4866 MW277530.1 |Severe acute respiratory syndrome coro ···················· ···················· ···················· ···················· ···················· ····················
4867 MW277532.1 |Severe acute respiratory syndrome coro ···················· ···················· ···················· ···················· ···················· ····················
4868 MW277534.1 |Severe acute respiratory syndrome coro ···················· ···················· ···················· ···················· ···················· ····················
4869 MW277535.1 |Severe acute respiratory syndrome coro ···················· ···················· ···················· ···················· ···················· ····················
4870 MW277536.1 |Severe acute respiratory syndrome coro ···················· ···················· ···················· ···················· ···················· ····················
4871 MW277537.1 |Severe acute respiratory syndrome coro ···················· ···················· ···················· ···················· ···················· ····················
4872 MW277538.1 |Severe acute respiratory syndrome coro ···················· ···················· ···················· ···················· ···················· ····················
4873 MW277539.1 |Severe acute respiratory syndrome coro ···················· ···················· ···················· ···················· ···················· ····················
4874 MW277540.1 |Severe acute respiratory syndrome coro ···················· ···················· ···················· ···················· ···················· ····················
4875 MW277544.1 |Severe acute respiratory syndrome coro ···················· ···················· ···················· ···················· ···················· ····················
4876 MW277546.1 |Severe acute respiratory syndrome coro ···················· ···················· ···················· ···················· ···················· ····················
4877 MW277547.1 |Severe acute respiratory syndrome coro ···················· ···················· ···················· ···················· ···················· ····················
4878 MW277548.1 |Severe acute respiratory syndrome coro ···················· ···················· ···················· ···················· ···················· ····················
4879 MW277550.1 |Severe acute respiratory syndrome coro ···················· ···················· ···················· ···················· ···················· ····················
4880 MW277551.1 |Severe acute respiratory syndrome coro ···················· ···················· ···················· ···················· ···················· ····················
4881 MW277553.1 |Severe acute respiratory syndrome coro ···················· ···················· ···················· ···················· ···················· ····················
4882 MW277554.1 |Severe acute respiratory syndrome coro ···················· ···················· ···················· ···················· ···················· ····················
4883 MW277555.1 |Severe acute respiratory syndrome coro ···················· ···················· ···················· ···················· ···················· ····················
4884 MW277556.1 |Severe acute respiratory syndrome coro ···················· ···················· ···················· ···················· ···················· ····················
4885 MW277557.1 |Severe acute respiratory syndrome coro ···················· ···················· ···················· ···················· ···················· ····················
4886 MW277558.1 |Severe acute respiratory syndrome coro ···················· ···················· ···················· ···················· ···················· ····················
4887 MW277559.1 |Severe acute respiratory syndrome coro ···················· ···················· ···················· ···················· ···················· ····················
4888 MW277560.1 |Severe acute respiratory syndrome coro ···················· ···················· ···················· ···················· ···················· ····················
4889 MW277561.1 |Severe acute respiratory syndrome coro ···················· ···················· ···················· ···················· ···················· ····················
4890 MW277562.1 |Severe acute respiratory syndrome coro ···················· ···················· ···················· ···················· ···················· ····················
4891 MW277563.1 |Severe acute respiratory syndrome coro ···················· ···················· ···················· ···················· ···················· ····················
4892 MW277567.1 |Severe acute respiratory syndrome coro ···················· ···················· ···················· ···················· ···················· ····················
4893 MW277568.1 |Severe acute respiratory syndrome coro ···················· ···················· ···················· ···················· ···················· ····················
4894 MW277569.1 |Severe acute respiratory syndrome coro ···················· ···················· ···················· ···················· ···················· ····················
4895 MW277571.1 |Severe acute respiratory syndrome coro ···················· ···················· ···················· ···················· ···················· ····················
4896 MW277572.1 |Severe acute respiratory syndrome coro ···················· ···················· ···················· ···················· ···················· ····················
4897 MW277574.1 |Severe acute respiratory syndrome coro ···················· ···················· ···················· ···················· ···················· ····················
4898 MW277576.1 |Severe acute respiratory syndrome coro ···················· ···················· ···················· ···················· ···················· ····················
4899 MW277577.1 |Severe acute respiratory syndrome coro ···················· ···················· ···················· ···················· ···················· ····················
4900 MW277579.1 |Severe acute respiratory syndrome coro ···················· ···················· ···················· ···················· ···················· ····················
4901 MW277581.1 |Severe acute respiratory syndrome coro ···················· ···················· ···················· ···················· ···················· ····················
4902 MW277582.1 |Severe acute respiratory syndrome coro ···················· ···················· ···················· ···················· ···················· ····················
4903 MW277583.1 |Severe acute respiratory syndrome coro ···················· ···················· ···················· ···················· ···················· ····················
4904 MW277585.1 |Severe acute respiratory syndrome coro ···················· ···················· ···················· ···················· ···················· ····················
4905 MW277586.1 |Severe acute respiratory syndrome coro ···················· ···················· ···················· ···················· ···················· ····················
4906 MW277587.1 |Severe acute respiratory syndrome coro ···················· ···················· ···················· ···················· ···················· ····················
4907 MW277588.1 |Severe acute respiratory syndrome coro ···················· ···················· ···················· ···················· ···················· ····················
4908 MW277589.1 |Severe acute respiratory syndrome coro ···················· ······A············· ···················· ···················· ···················· ····················
4909 MW277590.1 |Severe acute respiratory syndrome coro ···················· ···················· ···················· ···················· ···················· ····················
4910 MW277592.1 |Severe acute respiratory syndrome coro ···················· ···················· ···················· ···················· ···················· ····················
4911 MW277594.1 |Severe acute respiratory syndrome coro ···················· ···················· ···················· ···················· ···················· ····················
4912 MW277597.1 |Severe acute respiratory syndrome coro ···················· ···················· ···················· ···················· ···················· ····················
4913 MW277598.1 |Severe acute respiratory syndrome coro ···················· ···················· ···················· ···················· ···················· ····················
4914 MW277599.1 |Severe acute respiratory syndrome coro ···················· ···················· ···················· ···················· ···················· ····················
4915 MW277600.1 |Severe acute respiratory syndrome coro ···················· ···················· ···················· ···················· ···················· ····················
4916 MW277601.1 |Severe acute respiratory syndrome coro ···················· ···················· ···················· ···················· ···················· ····················
4917 MW277602.1 |Severe acute respiratory syndrome coro ···················· ···················· ···················· ···················· ···················· ····················
4919 MW277604.1 |Severe acute respiratory syndrome coro ···················· ···················· ···················· ···················· ···················· ····················
4920 MW277607.1 |Severe acute respiratory syndrome coro ···················· ···················· ···················· ···················· ···················· ····················
4921 MW277608.1 |Severe acute respiratory syndrome coro ···················· ···················· ···················· ···················· ···················· ····················
4922 MW277609.1 |Severe acute respiratory syndrome coro ···················· ···················· ···················· ···················· ···················· ····················
4923 MW277610.1 |Severe acute respiratory syndrome coro ···················· ···················· ···················· ···················· ···················· ····················
4924 MW277611.1 |Severe acute respiratory syndrome coro ···················· ···················· ···················· ···················· ···················· ····················
4925 MW277612.1 |Severe acute respiratory syndrome coro ···················· ···················· ···················· ···················· ···················· ····················
4926 MW277613.1 |Severe acute respiratory syndrome coro ···················· ···················· ···················· ···················· ···················· ····················
4927 MW277614.1 |Severe acute respiratory syndrome coro ···················· ···················· ···················· ···················· ···················· ····················
4928 MW277615.1 |Severe acute respiratory syndrome coro ···················· ···················· ···················· ···················· ···················· ····················
4929 MW277616.1 |Severe acute respiratory syndrome coro ···················· ···················· ···················· ···················· ···················· ····················
4930 MW277617.1 |Severe acute respiratory syndrome coro ···················· ···················· ···················· ···················· ···················· ····················
4931 MW277619.1 |Severe acute respiratory syndrome coro ···················· ···················· ···················· ···················· ···················· ····················
4932 MW277620.1 |Severe acute respiratory syndrome coro ···················· ···················· ···················· ···················· ···················· ····················
4933 MW277621.1 |Severe acute respiratory syndrome coro ···················· ···················· ···················· ···················· ···················· ····················
4934 MW277622.1 |Severe acute respiratory syndrome coro ···················· ···················· ···················· ···················· ···················· ····················
4935 MW277624.1 |Severe acute respiratory syndrome coro ···················· ···················· ···················· ···················· ···················· ····················
4936 MW277625.1 |Severe acute respiratory syndrome coro ···················· ······A············· ···················· ···················· ···················· ····················
4937 MW277626.1 |Severe acute respiratory syndrome coro ···················· ···················· ···················· ···················· ···················· ····················
4938 MW277628.1 |Severe acute respiratory syndrome coro ···················· ···················· ···················· ···················· ···················· ····················
4939 MW277629.1 |Severe acute respiratory syndrome coro ···················· ···················· ···················· ···················· ···················· ····················
4940 MW277630.1 |Severe acute respiratory syndrome coro ···················· ···················· ···················· ···················· ···················· ····················
4941 MW277631.1 |Severe acute respiratory syndrome coro ···················· ···················· ···················· ···················· ···················· ····················
4942 MW277632.1 |Severe acute respiratory syndrome coro ···················· ···················· ···················· ···················· ···················· ····················
4943 MW277633.1 |Severe acute respiratory syndrome coro ···················· ···················· ···················· ···················· ···················· ····················
4944 MW277635.1 |Severe acute respiratory syndrome coro ···················· ···················· ···················· ···················· ···················· ····················

| | | | | | | | | | |
|---|---|---|---|---|---|---|---|---|---|
| 4945 | MW277637.1 | Severe acute respiratory syndrome coro | .................... | .................... | .................... | .................... | .................... | .................... | |
| 4946 | MW277638.1 | Severe acute respiratory syndrome coro | .................... | .................... | .................... | .................... | .................... | .................... | |
| 4947 | MW277639.1 | Severe acute respiratory syndrome coro | .................... | .................... | .................... | .................... | .................... | .................... | |
| 4948 | MW277640.1 | Severe acute respiratory syndrome coro | .................... | .................... | .................... | .................... | .................... | .................... | |
| 4949 | MW277642.1 | Severe acute respiratory syndrome coro | .................... | .................... | .................... | .................... | .................... | .................... | |
| 4950 | MW277643.1 | Severe acute respiratory syndrome coro | .................... | .................... | .................... | .................... | .................... | .................... | |
| 4951 | MW277644.1 | Severe acute respiratory syndrome coro | .................... | .................... | .................... | .................... | .................... | .................... | |
| 4952 | MW277645.1 | Severe acute respiratory syndrome coro | .................... | .................... | .................... | .................... | .................... | .................... | |
| 4953 | MW277646.1 | Severe acute respiratory syndrome coro | .................... | .................... | .................... | .................... | .................... | .................... | |
| 4954 | MW277647.1 | Severe acute respiratory syndrome coro | .................... | .................... | .................... | .................... | .................... | .................... | |
| 4955 | MW277649.1 | Severe acute respiratory syndrome coro | .................... | .................... | .................... | .................... | .................... | .................... | |
| 4956 | MW277650.1 | Severe acute respiratory syndrome coro | .................... | .................... | .................... | .................... | .................... | .................... | |
| 4957 | MW277651.1 | Severe acute respiratory syndrome coro | .................... | .................... | .................... | .................... | .................... | .................... | |
| 4958 | MW277652.1 | Severe acute respiratory syndrome coro | .................... | .................... | .................... | .................... | .................... | .................... | |
| 4959 | MW277653.1 | Severe acute respiratory syndrome coro | .................... | .................... | .................... | .................... | .................... | .................... | |
| 4960 | MW277654.1 | Severe acute respiratory syndrome coro | .................... | .................... | .................... | .................... | .................... | .................... | |
| 4961 | MW277655.1 | Severe acute respiratory syndrome coro | .................... | .................... | .................... | .................... | .................... | .................... | |
| 4962 | MW277656.1 | Severe acute respiratory syndrome coro | .................... | .................... | .................... | .................... | .................... | .................... | |
| 4963 | MW277657.1 | Severe acute respiratory syndrome coro | .................... | .................... | .................... | .................... | .................... | .................... | |
| 4964 | MW277658.1 | Severe acute respiratory syndrome coro | .................... | .................... | .................... | .................... | .................... | .................... | |
| 4965 | MW277661.1 | Severe acute respiratory syndrome coro | .................... | .................... | .................... | .................... | .................... | .................... | |
| 4966 | MW277662.1 | Severe acute respiratory syndrome coro | .................... | .................... | .................... | .................... | .................... | .................... | |
| 4967 | MW279302.1 | Severe acute respiratory syndrome coro | .................... | .................... | .................... | .................... | .................... | ...............A.... | |
| 4968 | MW279303.1 | Severe acute respiratory syndrome coro | .................... | .................... | .................... | .................... | .................... | .................... | |
| 4969 | MW279314.1 | Severe acute respiratory syndrome coro | .................... | .................... | .................... | .................... | .................... | .................... | |
| 4970 | MW279315.1 | Severe acute respiratory syndrome coro | .................... | .................... | .................... | .................... | .................... | .................... | |
| 4971 | MW279316.1 | Severe acute respiratory syndrome coro | .................... | .................... | .................... | .................... | .................... | .................... | |
| 4972 | MW279317.1 | Severe acute respiratory syndrome coro | .................... | .................... | .................... | .................... | .................... | .................... | |
| 4973 | MW279318.1 | Severe acute respiratory syndrome coro | .................... | .................... | .................... | .................... | .................... | .................... | |
| 4974 | MW279319.1 | Severe acute respiratory syndrome coro | .................... | .................... | .................... | .................... | .................... | .................... | |
| 4975 | MW279320.1 | Severe acute respiratory syndrome coro | .................... | .................... | .................... | .................... | .................... | .................... | |
| 4976 | MW279321.1 | Severe acute respiratory syndrome coro | .................... | .................... | .................... | .................... | .................... | .................... | |
| 4977 | MW279322.1 | Severe acute respiratory syndrome coro | .................... | .................... | .................... | .................... | .................... | .................... | |
| 4978 | MW279323.1 | Severe acute respiratory syndrome coro | .................... | .................... | .................... | .................... | .................... | .................... | |
| 4979 | MW279324.1 | Severe acute respiratory syndrome coro | .................... | .................... | .................... | .................... | .................... | .................... | |
| 4980 | MW279325.1 | Severe acute respiratory syndrome coro | .................... | .................... | .................... | .................... | .................... | .................... | |
| 4981 | MW279326.1 | Severe acute respiratory syndrome coro | .................... | .A.................. | .................... | .................... | .................... | .................... | |
| 4982 | MW279327.1 | Severe acute respiratory syndrome coro | .................... | .A.................. | .................... | .................... | .................... | .................... | |
| 4983 | MW279328.1 | Severe acute respiratory syndrome coro | .................... | .A.................. | .................... | .................... | .................... | .................... | |
| 4984 | MW279329.1 | Severe acute respiratory syndrome coro | .................... | .A.................. | .................... | .................... | NNN................. | .................... | 1 |
| 4985 | MW279330.1 | Severe acute respiratory syndrome coro | .................... | .A.................. | .................... | .................... | .................... | .................... | |
| 4986 | MW279331.1 | Severe acute respiratory syndrome coro | .................... | .................... | .................... | .................... | .................... | .................... | |
| 4987 | MW279332.1 | Severe acute respiratory syndrome coro | .................... | .A.................. | .................... | .................... | .................... | .................... | |
| 4988 | MW279333.1 | Severe acute respiratory syndrome coro | .................... | .................... | .................... | .................... | .................... | .................... | |
| 4989 | MW279334.1 | Severe acute respiratory syndrome coro | .................... | .................... | .................... | .................... | .................... | .................... | |
| 4990 | MW279335.1 | Severe acute respiratory syndrome coro | .................... | .................... | .................... | .................... | .................... | .................... | |
| 4991 | MW279336.1 | Severe acute respiratory syndrome coro | .................... | .................... | .................... | .................... | .................... | .................... | |
| 4992 | MW279337.1 | Severe acute respiratory syndrome coro | .................... | .................... | .................... | .................... | .................... | .................... | |
| 4993 | MW279413.1 | Severe acute respiratory syndrome coro | .................... | .................... | .................... | .................... | .................... | .................... | |
| 4994 | MW279414.1 | Severe acute respiratory syndrome coro | .................... | .................... | .................... | .................... | .................... | .................... | |
| 4995 | MW279415.1 | Severe acute respiratory syndrome coro | .................... | .................... | .................... | .................... | .................... | .................... | |
| 4996 | MW279416.1 | Severe acute respiratory syndrome coro | .................... | .................... | .................... | .................... | .................... | .................... | |
| 4997 | MW279417.1 | Severe acute respiratory syndrome coro | .................... | .................... | .................... | .................... | .................... | .................... | |
| 4998 | MW279418.1 | Severe acute respiratory syndrome coro | .................... | .................... | .................... | .................... | .................... | .................... | |
| 4999 | MW279419.1 | Severe acute respiratory syndrome coro | .................... | .................... | .................... | .................... | .................... | .................... | |
| 5000 | MW279420.1 | Severe acute respiratory syndrome coro | .................... | .................... | .................... | .................... | .................... | .................... | |
| 5001 | MW279421.1 | Severe acute respiratory syndrome coro | .................... | .................... | .................... | .................... | .................... | .................... | |
| 5002 | MW279422.1 | Severe acute respiratory syndrome coro | .................... | .................... | .................... | .................... | .................... | .................... | |
| 5003 | MW279423.1 | Severe acute respiratory syndrome coro | .................... | .................... | .................... | .................... | .................... | .................... | |
| 5004 | MW279424.1 | Severe acute respiratory syndrome coro | .................... | .................... | .................... | .................... | .................... | .................... | |
| 5005 | MW279425.1 | Severe acute respiratory syndrome coro | .................... | .................... | .................... | .................... | .................... | .................... | |
| 5006 | MW279426.1 | Severe acute respiratory syndrome coro | .................... | .................... | .................... | .................... | .................... | .................... | |
| 5007 | MW279427.1 | Severe acute respiratory syndrome coro | .................... | .................... | .................... | .................... | .................... | .................... | |
| 5008 | MW279428.1 | Severe acute respiratory syndrome coro | .................... | .................... | .................... | .................... | .................... | .................... | |
| 5009 | MW279429.1 | Severe acute respiratory syndrome coro | .................... | .................... | .................... | .................... | .................... | .................... | |
| 5010 | MW279430.1 | Severe acute respiratory syndrome coro | .................... | .................... | .................... | .................... | .................... | .................... | |
| 5011 | MW279431.1 | Severe acute respiratory syndrome coro | .................... | .................... | .................... | .................... | .................... | .................... | |
| 5012 | MW279432.1 | Severe acute respiratory syndrome coro | .................... | .................... | .................... | .................... | .................... | .................... | |
| 5013 | MW279433.1 | Severe acute respiratory syndrome coro | .................... | .................... | .................... | .................... | .................... | .................... | |
| 5014 | MW279434.1 | Severe acute respiratory syndrome coro | .................... | .................... | .................... | .................... | .................... | .................... | |
| 5015 | MW279435.1 | Severe acute respiratory syndrome coro | .................... | .................... | .................... | .................... | .................... | .................... | |
| 5016 | MW279436.1 | Severe acute respiratory syndrome coro | .................... | .................... | .................... | .................... | .................... | .................... | |
| 5017 | MW279437.1 | Severe acute respiratory syndrome coro | .................... | .................... | .................... | .................... | .................... | .................... | |
| 5018 | MW279438.1 | Severe acute respiratory syndrome coro | .................... | .................... | .................... | .................... | .................... | .................... | |
| 5019 | MW279439.1 | Severe acute respiratory syndrome coro | .................... | .................... | .................... | .................... | .................... | .................... | |
| 5020 | MW279440.1 | Severe acute respiratory syndrome coro | .................... | .................... | .................... | .................... | .................... | .................... | |
| 5021 | MW279441.1 | Severe acute respiratory syndrome coro | .................... | .................... | .................... | .................... | .................... | .................... | |
| 5022 | MW279442.1 | Severe acute respiratory syndrome coro | .................... | .................... | .................... | .................... | .................... | .................... | |

```
5023 MW279443.1 |Severe acute respiratory syndrome coro    ····················  ····················  ·····················  ····················  ····················  ····················
5024 MW279444.1 |Severe acute respiratory syndrome coro    ····················  ····················  ·····················  ····················  ····················  ····················
5025 MW279445.1 |Severe acute respiratory syndrome coro    ····················  ····················  ·····················  ····················  ····················  ····················
5026 MW279446.1 |Severe acute respiratory syndrome coro    ····················  ····················  ·····················  ····················  ····················  ····················
5027 MW279447.1 |Severe acute respiratory syndrome coro    ····················  ····················  ·····················  ····················  ····················  ····················
5028 MW279448.1 |Severe acute respiratory syndrome coro    ····················  ····················  ·····················  ····················  ····················  ····················
5029 MW279449.1 |Severe acute respiratory syndrome coro    ····················  ····················  ·····················  ····················  ····················  ····················
5030 MW279450.1 |Severe acute respiratory syndrome coro    ····················  ····················  ·····················  ····················  ····················  ····················
5031 MW279451.1 |Severe acute respiratory syndrome coro    ····················  ····················  ·····················  ····················  ····················  ····················
5032 MW279452.1 |Severe acute respiratory syndrome coro    ····················  ····················  ·····················  ····················  ····················  ····················
5033 MW279453.1 |Severe acute respiratory syndrome coro    ····················  ····················  ·····················  ····················  ····················  ····················
5034 MW279454.1 |Severe acute respiratory syndrome coro    ····················  ····················  ·····················  ····················  ····················  ····················
5035 MW279455.1 |Severe acute respiratory syndrome coro    ····················  ·A··················  ·····················  ····················  ····················  ····················
5036 MW279456.1 |Severe acute respiratory syndrome coro    ····················  ····················  ·····················  ····················  ····················  ····················
5037 MW279457.1 |Severe acute respiratory syndrome coro    ····················  ····················  ·····················  ····················  ····················  ····················
5038 MW279458.1 |Severe acute respiratory syndrome coro    ····················  ····················  ·····················  ····················  ····················  ····················
5039 MW279459.1 |Severe acute respiratory syndrome coro    ····················  ····················  ·····················  ····················  ····················  ····················
5040 MW279460.1 |Severe acute respiratory syndrome coro    ····················  ····················  ·····················  ····················  ····················  ····················
5041 MW280149.1 |Severe acute respiratory syndrome coro    ····················  ····················  ·····················  ····················  ····················  ····················
5042 MW280150.1 |Severe acute respiratory syndrome coro    ····················  ·A··················  ·····················  ····················  ····················  ····················
5043 MW280151.1 |Severe acute respiratory syndrome coro    ····················  ····················  ·····················  ····················  ····················  ····················
5044 MW280152.1 |Severe acute respiratory syndrome coro    ····················  ····················  ·····················  ····················  ····················  ····················
5045 MW280153.1 |Severe acute respiratory syndrome coro    ····················  ····················  ·····················  ····················  ····················  ····················
5046 MW280154.1 |Severe acute respiratory syndrome coro    ····················  ·A··················  ·····················  ····················  ····················  ····················
5047 MW280155.1 |Severe acute respiratory syndrome coro    ····················  ····················  ·····················  ····················  ····················  ····················
5048 MW280156.1 |Severe acute respiratory syndrome coro    ····················  ····················  ·····················  ····················  ····················  ····················
5049 MW280157.1 |Severe acute respiratory syndrome coro    ····················  ····················  ·····················  ····················  ····················  ····················
5050 MW280158.1 |Severe acute respiratory syndrome coro    ····················  ············C·······  ·····················  ····················  ····················  ····················
5051 MW280159.1 |Severe acute respiratory syndrome coro    ····················  ····················  ·····················  ····················  ····················  ····················
5052 MW280162.1 |Severe acute respiratory syndrome coro    ····················  ····················  ·····················  ····················  ····················  ····················
5053 MW280163.1 |Severe acute respiratory syndrome coro    ····················  ····················  ·····················  ····················  ····················  ····················
5054 MW280165.1 |Severe acute respiratory syndrome coro    ····················  ····················  ·····················  ····················  ····················  ····················
5055 MW280166.1 |Severe acute respiratory syndrome coro    ····················  ····················  ·····················  ····················  ····················  ····················
5056 MW280167.1 |Severe acute respiratory syndrome coro    ····················  ····················  ·····················  ····················  ····················  ····················
5057 MW280168.1 |Severe acute respiratory syndrome coro    ····················  ····················  ·····················  ····················  ····················  ····················
5058 MW280169.1 |Severe acute respiratory syndrome coro    ····················  ····················  ·····················  ····················  ····················  ····················
5059 MW280170.1 |Severe acute respiratory syndrome coro    ····················  ····A···············  ·····················  ····················  ····················  ····················
5060 MW280171.1 |Severe acute respiratory syndrome coro    ····················  ····················  ·····················  ····················  ····················  ····················
5061 MW280172.1 |Severe acute respiratory syndrome coro    ····················  ····················  ·····················  ····················  ····················  ····················
5062 MW280173.1 |Severe acute respiratory syndrome coro    ····················  ····A···············  ·····················  ····················  ····················  ····················
5063 MW280174.1 |Severe acute respiratory syndrome coro    ····················  ····················  ·····················  ····················  ····················  ····················
5064 MW280175.1 |Severe acute respiratory syndrome coro    ····················  ····················  ·····················  ····················  ····················  ····················
5065 MW280176.1 |Severe acute respiratory syndrome coro    ····················  ····················  ·····················  ····················  ····················  ····················
5066 MW280177.1 |Severe acute respiratory syndrome coro    ····················  ····················  ·····················  ····················  ····················  ····················
5067 MW280178.1 |Severe acute respiratory syndrome coro    ····················  ····················  ·····················  ····················  ····················  ····················
5068 MW280179.1 |Severe acute respiratory syndrome coro    ····················  ·A··················  ·····················  ····················  ····················  ····················
5069 MW280180.1 |Severe acute respiratory syndrome coro    ····················  ····················  ·····················  ····················  ····················  ····················
5070 MW280181.1 |Severe acute respiratory syndrome coro    ····················  ····················  ·····················  ····················  ····················  ····················
5071 MW280182.1 |Severe acute respiratory syndrome coro    ····················  ····················  ·····················  ····················  ····················  ····················
5072 MW280183.1 |Severe acute respiratory syndrome coro    ····················  ····················  ·····················  ····················  ····················  ····················
5073 MW280184.1 |Severe acute respiratory syndrome coro    ····················  ····················  ·····················  ····················  ····················  ····················
5074 MW280185.1 |Severe acute respiratory syndrome coro    ····················  ····················  ·····················  ····················  ····················  ····················
5075 MW280187.1 |Severe acute respiratory syndrome coro    ····················  ····················  ·····················  ····················  ····················  ····················
5076 MW280450.1 |Severe acute respiratory syndrome coro    ····················  ····················  ·····················  ····················  ····················  ····················
5077 MW280451.1 |Severe acute respiratory syndrome coro    ····················  ·A··················  ·····················  ····················  ····················  ····················
5078 MW280452.1 |Severe acute respiratory syndrome coro    ····················  ·A··················  ·····················  ····················  ····················  ····················
5079 MW280453.1 |Severe acute respiratory syndrome coro    ····················  ····················  ·····················  ····················  ····················  ····················
5080 MW280454.1 |Severe acute respiratory syndrome coro    ····················  ····················  ·····················  ····················  ····················  ····················
5081 MW280456.1 |Severe acute respiratory syndrome coro    ····················  ····················  ·····················  ····················  ····················  ····················
5082 MW280457.1 |Severe acute respiratory syndrome coro    ····················  ····················  ·····················  ····················  ····················  ····················
5083 MW280458.1 |Severe acute respiratory syndrome coro    ····················  ····················  ·····················  ····················  ····················  ····················
5084 MW280459.1 |Severe acute respiratory syndrome coro    ····················  ·A··················  ·····················  ····················  ····················  ····················
5085 MW280460.1 |Severe acute respiratory syndrome coro    ····················  ·A··················  ·····················  ····················  ····················  ····················
5086 MW280461.1 |Severe acute respiratory syndrome coro    ····················  ····················  ·····················  ····················  ····················  ····················
5087 MW280462.1 |Severe acute respiratory syndrome coro    ····················  ····················  ·····················  ····················  ····················  ····················
5088 MW280464.1 |Severe acute respiratory syndrome coro    ····················  ····················  ·····················  ····················  ····················  ····················
5089 MW280465.1 |Severe acute respiratory syndrome coro    ····················  ····················  ·····················  ····················  ····················  ····················
5090 MW280467.1 |Severe acute respiratory syndrome coro    ····················  ·A··················  ···········T·········  ····················  ····················  ····················
5091 MW280468.1 |Severe acute respiratory syndrome coro    ····················  ·A··················  ·····················  ····················  ····················  ····················
5092 MW280469.1 |Severe acute respiratory syndrome coro    ····················  ····················  ·····················  ····················  ····················  ····················
5093 MW280470.1 |Severe acute respiratory syndrome coro    ····················  ····················  ·····················  ····················  ····················  ····················
5094 MW280471.1 |Severe acute respiratory syndrome coro    ····················  ····················  ·····················  ····················  ····················  ····················
5095 MW280472.1 |Severe acute respiratory syndrome coro    ····················  ····················  ·····················  ····················  ····················  ····················
5096 MW280473.1 |Severe acute respiratory syndrome coro    ····················  ····················  ·····················  ····················  ····················  ····················
5097 MW280474.1 |Severe acute respiratory syndrome coro    ····················  ····················  ·····················  ····················  ····················  ····················
5098 MW280475.1 |Severe acute respiratory syndrome coro    ····················  ····················  ·····················  ····················  ····················  ····················
5099 MW280476.1 |Severe acute respiratory syndrome coro    ····················  ····················  ·····················  ····················  ····················  ····················
5100 MW280478.1 |Severe acute respiratory syndrome coro    ····················  ····················  ·····················  ····················  ····················  ····················
```

| | | | | | | | | |
|---|---|---|---|---|---|---|---|---|
| 5101 | MW280479.1 | Severe acute respiratory syndrome coro | ···················· | ···················· | ···················· | ···················· | ···················· | ···················· |
| 5102 | MW280482.1 | Severe acute respiratory syndrome coro | ···················· | ·A·················· | ···················· | ···················· | ···················· | ···················· |
| 5103 | MW280483.1 | Severe acute respiratory syndrome coro | ···················· | ···················· | ···················· | ···················· | ···················· | ···················· |
| 5104 | MW280485.1 | Severe acute respiratory syndrome coro | ···················· | ···················· | ···················· | ···················· | ···················· | ···················· |
| 5105 | MW280486.1 | Severe acute respiratory syndrome coro | ···················· | ···················· | ···················· | ···················· | ···················· | ···················· |
| 5106 | MW280487.1 | Severe acute respiratory syndrome coro | ···················· | ···················· | ···················· | ···················· | ···················· | ···················· |
| 5107 | MW280488.1 | Severe acute respiratory syndrome coro | ···················· | ·A·················· | ···················· | ···················· | ···················· | ···················· |
| 5108 | MW280489.1 | Severe acute respiratory syndrome coro | ···················· | ·A·················· | ···················· | ···················· | ···················· | ···················· |
| 5109 | MW280490.1 | Severe acute respiratory syndrome coro | ···················· | ···················· | ···················· | ···················· | ···················· | ···················· |
| 5110 | MW280491.1 | Severe acute respiratory syndrome coro | ···················· | ···················· | ···················· | ···················· | ···················· | ···················· |
| 5111 | MW280492.1 | Severe acute respiratory syndrome coro | ···················· | ···················· | ···················· | ···················· | ···················· | ···················· |
| 5112 | MW280493.1 | Severe acute respiratory syndrome coro | ···················· | ···················· | ···················· | ···················· | ···················· | ···················· |
| 5113 | MW280494.1 | Severe acute respiratory syndrome coro | ···················· | ···················· | ···················· | ···················· | ···················· | ···················· |
| 5114 | MW280495.1 | Severe acute respiratory syndrome coro | ···················· | ·A·················· | ···················· | ···················· | ···················· | ···················· |
| 5115 | MW280496.1 | Severe acute respiratory syndrome coro | ···················· | ···················· | ···················· | ···················· | ···················· | ···················· |
| 5116 | MW280497.1 | Severe acute respiratory syndrome coro | ···················· | ···················· | ···················· | ···················· | ···················· | ···················· |
| 5117 | MW280498.1 | Severe acute respiratory syndrome coro | ···················· | ···················· | ···················· | ···················· | ···················· | ···················· |
| 5118 | MW280499.1 | Severe acute respiratory syndrome coro | ···················· | ·A·················· | ···················· | ···················· | ···················· | ···················· |
| 5119 | MW280501.1 | Severe acute respiratory syndrome coro | ···················· | ···················· | ···················· | ···················· | ···················· | ···················· |
| 5120 | MW280502.1 | Severe acute respiratory syndrome coro | ···················· | ·A·················· | ···················· | ···················· | ···················· | ···················· |
| 5121 | MW280503.1 | Severe acute respiratory syndrome coro | ···················· | ···················· | ···················· | ···················· | ···················· | ···················· |
| 5122 | MW280505.1 | Severe acute respiratory syndrome coro | ···················· | ···················· | ···················· | ···················· | ···················· | ···················· |
| 5123 | MW280507.1 | Severe acute respiratory syndrome coro | ···················· | ···················· | ···················· | ···················· | ···················· | ···················· |
| 5124 | MW280508.1 | Severe acute respiratory syndrome coro | ···················· | ···················· | ···················· | ···················· | ···················· | ···················· |
| 5125 | MW280509.1 | Severe acute respiratory syndrome coro | ···················· | ···················· | ···················· | ···················· | ···················· | ···················· |
| 5126 | MW280511.1 | Severe acute respiratory syndrome coro | ···················· | ···················· | ···················· | ···················· | ···················· | ···················· |
| 5127 | MW280512.1 | Severe acute respiratory syndrome coro | ···················· | ···················· | ···················· | ···················· | ···················· | ···················· |
| 5128 | MW280513.1 | Severe acute respiratory syndrome coro | ···················· | ···················· | ···················· | ···················· | ···················· | ···················· |
| 5129 | MW280516.1 | Severe acute respiratory syndrome coro | ···················· | ·A·················· | ···················· | ···················· | ···················· | ···················· |
| 5130 | MW280517.1 | Severe acute respiratory syndrome coro | ···················· | ···················· | ···················· | ···················· | ···················· | ···················· |
| 5131 | MW280518.1 | Severe acute respiratory syndrome coro | ···················· | ···················· | ···················· | ···················· | ···················· | ···················· |
| 5132 | MW280519.1 | Severe acute respiratory syndrome coro | ···················· | ···················· | ···················· | ···················· | ···················· | ···················· |
| 5133 | MW280520.1 | Severe acute respiratory syndrome coro | ···················· | ···················· | ···················· | ···················· | ···················· | ···················· |
| 5134 | MW280521.1 | Severe acute respiratory syndrome coro | ···················· | ···················· | ···················· | ···················· | ···················· | ···················· |
| 5135 | MW280522.1 | Severe acute respiratory syndrome coro | ···················· | ·A·················· | ···················· | ···················· | ···················· | ···················· |
| 5136 | MW280525.1 | Severe acute respiratory syndrome coro | ···················· | ···················· | ···················· | ···················· | ···················· | ···················· |
| 5137 | MW280527.1 | Severe acute respiratory syndrome coro | ···················· | ···················· | ···················· | ···················· | ···················· | ···················· |
| 5138 | MW280528.1 | Severe acute respiratory syndrome coro | ···················· | ···················· | ···················· | ···················· | ···················· | ···················· |
| 5139 | MW280529.1 | Severe acute respiratory syndrome coro | ···················· | ···················· | ···················· | ···················· | ···················· | ···················· |
| 5140 | MW280530.1 | Severe acute respiratory syndrome coro | ···················· | ···················· | ···················· | ···················· | ···················· | ···················· |
| 5141 | MW280531.1 | Severe acute respiratory syndrome coro | ···················· | ·············A······ | ···················· | ···················· | ···················· | ···················· |
| 5142 | MW280533.1 | Severe acute respiratory syndrome coro | ···················· | ···················· | ···················· | ···················· | ···················· | ···················· |
| 5143 | MW280534.1 | Severe acute respiratory syndrome coro | ···················· | ···················· | ···················· | ···················· | ···················· | ···················· |
| 5144 | MW280537.1 | Severe acute respiratory syndrome coro | ···················· | ···················· | ···················· | ···················· | ···················· | ···················· |
| 5145 | MW280538.1 | Severe acute respiratory syndrome coro | ···················· | ···················· | ···················· | ···················· | ···················· | ···················· |
| 5146 | MW280539.1 | Severe acute respiratory syndrome coro | ···················· | ···················· | ···················· | ···················· | ···················· | ···················· |
| 5147 | MW280541.1 | Severe acute respiratory syndrome coro | ···················· | ·············A······ | ···················· | ···················· | ···················· | ···················· |
| 5148 | MW280542.1 | Severe acute respiratory syndrome coro | ···················· | ·A·················· | ···················· | ···················· | ···················· | ···················· |
| 5149 | MW280545.1 | Severe acute respiratory syndrome coro | ···················· | ·A·················· | ···················· | ···················· | ···················· | ···················· |
| 5150 | MW280546.1 | Severe acute respiratory syndrome coro | ···················· | ···················· | ···················· | ···················· | ···················· | ···················· |
| 5151 | MW280547.1 | Severe acute respiratory syndrome coro | ···················· | ···················· | ···················· | ···················· | ···················· | ···················· |
| 5152 | MW280548.1 | Severe acute respiratory syndrome coro | ···················· | ···················· | ···················· | ···················· | ···················· | ···················· |
| 5153 | MW280549.1 | Severe acute respiratory syndrome coro | ···················· | ···················· | ···················· | ···················· | ···················· | ···················· |
| 5154 | MW280550.1 | Severe acute respiratory syndrome coro | ···················· | ···················· | ···················· | ···················· | ···················· | ···················· |
| 5155 | MW269882.1 | Severe acute respiratory syndrome coro | ···················· | ···················· | ···················· | ···················· | ···················· | ···················· |
| 5156 | MW269884.1 | Severe acute respiratory syndrome coro | ···················· | ···················· | ···················· | ···················· | ···················· | ···················· |
| 5157 | MW269885.1 | Severe acute respiratory syndrome coro | ···················· | ···················· | ···················· | ···················· | ···················· | ···················· |
| 5158 | MW269886.1 | Severe acute respiratory syndrome coro | ···················· | ···················· | ···················· | ···················· | ···················· | ···················· |
| 5159 | MW269887.1 | Severe acute respiratory syndrome coro | ···················· | ···················· | ···················· | ···················· | ···················· | ···················· |
| 5160 | MW269890.1 | Severe acute respiratory syndrome coro | ···················· | ···················· | ···················· | ···················· | ···················· | ···················· |
| 5161 | MW269891.1 | Severe acute respiratory syndrome coro | ···················· | ···················· | ···················· | ···················· | ···················· | ···················· |
| 5162 | MW269893.1 | Severe acute respiratory syndrome coro | ···················· | ···················· | ···················· | ···················· | ···················· | ···················· |
| 5163 | MW269895.1 | Severe acute respiratory syndrome coro | ···················· | ···················· | ···················· | ···················· | ···················· | ···················· |
| 5164 | MW269896.1 | Severe acute respiratory syndrome coro | ···················· | ···················· | ···················· | ···················· | ···················· | ···················· |
| 5165 | MW269898.1 | Severe acute respiratory syndrome coro | ···················· | ···················· | ···················· | ···················· | ···················· | ···················· |
| 5166 | MW269900.1 | Severe acute respiratory syndrome coro | ···················· | ···················· | ···················· | ···················· | ···················· | ···················· |
| 5167 | MW269902.1 | Severe acute respiratory syndrome coro | ···················· | ···················· | ···················· | ···················· | ···················· | ···················· |
| 5168 | MW269903.1 | Severe acute respiratory syndrome coro | ···················· | ···················· | ···················· | ···················· | ···················· | ···················· |
| 5169 | MW269904.1 | Severe acute respiratory syndrome coro | ···················· | ···················· | ···················· | ···················· | ···················· | ···················· |
| 5170 | MW273779.1 | Severe acute respiratory syndrome coro | ···················· | ···················· | ···················· | ···················· | ···················· | ···················· |
| 5171 | MW273783.1 | Severe acute respiratory syndrome coro | ···················· | ···················· | ···················· | ···················· | ···················· | ···················· |
| 5172 | MW273784.1 | Severe acute respiratory syndrome coro | ···················· | ···················· | ···················· | ···················· | ···················· | ···················· |
| 5173 | MW273785.1 | Severe acute respiratory syndrome coro | ···················· | ···················· | ···················· | ···················· | ···················· | ···················· |
| 5174 | MW273786.1 | Severe acute respiratory syndrome coro | ···················· | ···················· | ···················· | ···················· | ···················· | ···················· |
| 5175 | MW273787.1 | Severe acute respiratory syndrome coro | ···················· | ···················· | ···················· | ···················· | ···················· | ···················· |
| 5176 | MW273788.1 | Severe acute respiratory syndrome coro | ···················· | ···················· | ···················· | ···················· | ···················· | ···················· |
| 5177 | MW273789.1 | Severe acute respiratory syndrome coro | ···················· | ···················· | ···················· | ···················· | ···················· | ···················· |
| 5178 | MW273790.1 | Severe acute respiratory syndrome coro | ···················· | ···················· | ···················· | ···················· | ···················· | ···················· |

5179 MW273791.1 |Severe acute respiratory syndrome coro ···················· ···················· ···················· ···················· ···················· ····················
5180 MW273792.1 |Severe acute respiratory syndrome coro ···················· ···················· ···················· ···················· ···················· ····················
5181 MW273793.1 |Severe acute respiratory syndrome coro ···················· ···················· ···················· ···················· ···················· ····················
5182 MW273794.1 |Severe acute respiratory syndrome coro ···················· ···················· ···················· ···················· ···················· ····················
5183 MW273795.1 |Severe acute respiratory syndrome coro ···················· ···················· ···················· ···················· ···················· ····················
5184 MW273796.1 |Severe acute respiratory syndrome coro ···················· ···················· ···················· ···················· ···················· ····················
5185 MW273797.1 |Severe acute respiratory syndrome coro ···················· ···················· ···················· ···················· ···················· ····················
5186 MW273798.1 |Severe acute respiratory syndrome coro ···················· ···················· ···················· ···················· ···················· ····················
5187 MW273799.1 |Severe acute respiratory syndrome coro ···················· ···················· ···················· ···················· ···················· ····················
5188 MW273800.1 |Severe acute respiratory syndrome coro ···················· ···················· ···················· ···················· ···················· ····················
5189 MW273801.1 |Severe acute respiratory syndrome coro ···················· ···················· ···················· ···················· ···················· ····················
5190 MW273802.1 |Severe acute respiratory syndrome coro ···················· ···················· ···················· ···················· ···················· ····················
5191 MW273803.1 |Severe acute respiratory syndrome coro ···················· ···················· ···················· ···················· ···················· ····················
5192 MW273804.1 |Severe acute respiratory syndrome coro ···················· ···················· ···················· ···················· ···················· ····················
5193 MW273805.1 |Severe acute respiratory syndrome coro ···················· ···················· ···················· ···················· ···················· ····················
5194 MW273806.1 |Severe acute respiratory syndrome coro ···················· ···················· ···················· ···················· ···················· ····················
5195 MW273807.1 |Severe acute respiratory syndrome coro ···················· ···················· ···················· ···················· ···················· ····················
5196 MW273808.1 |Severe acute respiratory syndrome coro ···················· ·A·················· ···················· ···················· ···················· ····················
5197 MW273809.1 |Severe acute respiratory syndrome coro ···················· ·A·················· ···················· ···················· ···················· ····················
5198 MW273810.1 |Severe acute respiratory syndrome coro ···················· ···················· ···················· ···················· ···················· ····················
5199 MW273811.1 |Severe acute respiratory syndrome coro ···················· ···················· ···················· ···················· ···················· ····················
5200 MW273812.1 |Severe acute respiratory syndrome coro ···················· ···················· ···················· ···················· ···················· ····················
5201 MW273813.1 |Severe acute respiratory syndrome coro ···················· ···················· ···················· ···················· ···················· ····················
5202 MW273814.1 |Severe acute respiratory syndrome coro ···················· ···················· ···················· ···················· ···················· ····················
5203 MW273815.1 |Severe acute respiratory syndrome coro ···················· ···················· ···················· ···················· ···················· ····················
5204 MW273816.1 |Severe acute respiratory syndrome coro ···················· ···················· ···················· ···················· ···················· ····················
5205 MW273817.1 |Severe acute respiratory syndrome coro ···················· ···················· ···················· ···················· ···················· ····················
5206 MW273818.1 |Severe acute respiratory syndrome coro ···················· ···················· ···················· ···················· ···················· ····················
5207 MW273819.1 |Severe acute respiratory syndrome coro ···················· ···················· ···················· ···················· ···················· ····················
5208 MW273820.1 |Severe acute respiratory syndrome coro ···················· ···················· ···················· ···················· ···················· ····················
5209 MW273821.1 |Severe acute respiratory syndrome coro ···················· ···················· ···················· ···················· ···················· ····················
5210 MW273822.1 |Severe acute respiratory syndrome coro ···················· ···················· ···················· ···················· ···················· ····················
5211 MW273823.1 |Severe acute respiratory syndrome coro ···················· ···················· ···················· ···················· ···················· ····················
5212 MW273824.1 |Severe acute respiratory syndrome coro ···················· ···················· ···················· ···················· ···················· ····················
5213 MW273825.1 |Severe acute respiratory syndrome coro ···················· ···················· ···················· ···················· ···················· ····················
5214 MW273826.1 |Severe acute respiratory syndrome coro ···················· ·A·················· ···················· ···················· ···················· ····················
5215 MW273827.1 |Severe acute respiratory syndrome coro ···················· ···················· ···················· ···················· ···················· ····················
5216 MW273828.1 |Severe acute respiratory syndrome coro ···················· ···················· ···················· ···················· ···················· ····················
5217 MW273829.1 |Severe acute respiratory syndrome coro ···················· ···················· ···················· ···················· ···················· ····················
5218 MW273830.1 |Severe acute respiratory syndrome coro ···················· ···················· ···················· ···················· ···················· ····················
5219 MW273831.1 |Severe acute respiratory syndrome coro ···················· ···················· ···················· ···················· ···················· ····················
5220 MW273832.1 |Severe acute respiratory syndrome coro ···················· ···················· ···················· ···················· ···················· ····················
5221 MW273833.1 |Severe acute respiratory syndrome coro ···················· ···················· ···················· ···················· ···················· ····················
5222 MW273834.1 |Severe acute respiratory syndrome coro ···················· ···················· ···················· ···················· ···················· ····················
5223 MW273835.1 |Severe acute respiratory syndrome coro ···················· ···················· ···················· ···················· ···················· ····················
5224 MW273836.1 |Severe acute respiratory syndrome coro ···················· ···················· ···················· ···················· ···················· ····················
5225 MW273837.1 |Severe acute respiratory syndrome coro ···················· ···················· ···················· ···················· ···················· ····················
5226 MW273838.1 |Severe acute respiratory syndrome coro ···················· ···················· ···················· ···················· ···················· ····················
5227 MW273839.1 |Severe acute respiratory syndrome coro ···················· ···················· ···················· ···················· ···················· ····················
5228 MW273840.1 |Severe acute respiratory syndrome coro ···················· ···················· ···················· ···················· ···················· ····················
5229 MW273841.1 |Severe acute respiratory syndrome coro ···················· ···················· ···················· ···················· ···················· ····················
5230 MW273842.1 |Severe acute respiratory syndrome coro ···················· ···················· ···················· ···················· ···················· ····················
5231 MW273843.1 |Severe acute respiratory syndrome coro ···················· ···················· ···················· ···················· ···················· ····················
5232 MW273844.1 |Severe acute respiratory syndrome coro ···················· ···················· ···················· ···················· ···················· ····················
5233 MW273845.1 |Severe acute respiratory syndrome coro ···················· ···················· ···················· ···················· ···················· ····················
5234 MW273846.1 |Severe acute respiratory syndrome coro ···················· ···················· ···················· ···················· ···················· ····················
5235 MW273847.1 |Severe acute respiratory syndrome coro ···················· ···················· ···················· ···················· ···················· ····················
5236 MW273848.1 |Severe acute respiratory syndrome coro ···················· ···················· ···················· ···················· ···················· ····················
5237 MW273849.1 |Severe acute respiratory syndrome coro ···················· ···················· ···················· ···················· ···················· ····················
5238 MW273850.1 |Severe acute respiratory syndrome coro ···················· ···················· ···················· ···················· ···················· ····················
5239 MW273851.1 |Severe acute respiratory syndrome coro ···················· ···················· ···················· ···················· ···················· ····················
5240 MW273852.1 |Severe acute respiratory syndrome coro ···················· ···················· ···················· ···················· ···················· ····················
5241 MW273853.1 |Severe acute respiratory syndrome coro ···················· ···················· ···················· ···················· ···················· ····················
5242 MW273854.1 |Severe acute respiratory syndrome coro ···················· ···················· ···················· ···················· ···················· ····················
5243 MW273855.1 |Severe acute respiratory syndrome coro ···················· ···················· ···················· ···················· ···················· ····················
5244 MW273856.1 |Severe acute respiratory syndrome coro ···················· ···················· ···················· ···················· ···················· ····················
5245 MW273857.1 |Severe acute respiratory syndrome coro ···················· ···················· ···················· ···················· ···················· ····················
5246 MW273858.1 |Severe acute respiratory syndrome coro ···················· ···················· ···················· ···················· ···················· ····················
5247 MW273859.1 |Severe acute respiratory syndrome coro ···················· ···················· ···················· ···················· ···················· ····················
5248 MW273860.1 |Severe acute respiratory syndrome coro ···················· ···················· ···················· ···················· ···················· ····················
5249 MW273861.1 |Severe acute respiratory syndrome coro ···················· ···················· ···················· ···················· ···················· ····················
5250 MW273862.1 |Severe acute respiratory syndrome coro ···················· ···················· ···················· ···················· ···················· ····················
5251 MW273863.1 |Severe acute respiratory syndrome coro ···················· ···················· ···················· ···················· ···················· ····················
5252 MW273864.1 |Severe acute respiratory syndrome coro ···················· ···················· ···················· ···················· ···················· ····················
5253 MW273865.1 |Severe acute respiratory syndrome coro ···················· ···················· ···················· ···················· ···················· ····················
5254 MW273866.1 |Severe acute respiratory syndrome coro ···················· ···················· ···················· ···················· ···················· ····················
5255 MW273867.1 |Severe acute respiratory syndrome coro ···················· ···················· ···················· ···················· ···················· ····················
5256 MW273868.1 |Severe acute respiratory syndrome coro ···················· ···················· ···················· ···················· ···················· ····················

| | | | | | | | | |
|---|---|---|---|---|---|---|---|---|
| 5257 | MW273869.1 | \|Severe acute respiratory syndrome coro | ···················· | ···················· | ····················· | ···················· | ···················· | ···················· |
| 5258 | MW273870.1 | \|Severe acute respiratory syndrome coro | ···················· | ···················· | ····················· | ···················· | ···················· | ···················· |
| 5259 | MW273871.1 | \|Severe acute respiratory syndrome coro | ···················· | ···················· | ····················· | ···················· | ···················· | ···················· |
| 5260 | MW273872.1 | \|Severe acute respiratory syndrome coro | ···················· | ···················· | ····················· | ···················· | ···················· | ···················· |
| 5261 | MW273873.1 | \|Severe acute respiratory syndrome coro | ···················· | ···················· | ····················· | ···················· | ···················· | ···················· |
| 5262 | MW273874.1 | \|Severe acute respiratory syndrome coro | ···················· | ···················· | ····················· | ···················· | ···················· | ···················· |
| 5263 | MW273875.1 | \|Severe acute respiratory syndrome coro | ···················· | ···················· | ····················· | ···················· | ···················· | ···················· |
| 5264 | MW273876.1 | \|Severe acute respiratory syndrome coro | ···················· | ···················· | ····················· | ···················· | ···················· | ···················· |
| 5265 | MW273877.1 | \|Severe acute respiratory syndrome coro | ···················· | ···················· | ····················· | ···················· | ···················· | ···················· |
| 5266 | MW273878.1 | \|Severe acute respiratory syndrome coro | ···················· | ···················· | ····················· | ···················· | ···················· | ···················· |
| 5267 | MW273879.1 | \|Severe acute respiratory syndrome coro | ···················· | ···················· | ····················· | ···················· | ···················· | ···················· |
| 5268 | MW273880.1 | \|Severe acute respiratory syndrome coro | ···················· | ···················· | ····················· | ···················· | ···················· | ···················· |
| 5269 | MW273881.1 | \|Severe acute respiratory syndrome coro | ···················· | ···················· | ····················· | ···················· | ···················· | ···················· |
| 5270 | MW273882.1 | \|Severe acute respiratory syndrome coro | ···················· | ···················· | ····················· | ···················· | ···················· | ···················· |
| 5271 | MW273883.1 | \|Severe acute respiratory syndrome coro | ···················· | ···················· | ····················· | ···················· | ···················· | ···················· |
| 5272 | MW273884.1 | \|Severe acute respiratory syndrome coro | ···················· | ···················· | ····················· | ···················· | ···················· | ···················· |
| 5273 | MW273885.1 | \|Severe acute respiratory syndrome coro | ···················· | ···················· | ····················· | ···················· | ···················· | ···················· |
| 5274 | MW273886.1 | \|Severe acute respiratory syndrome coro | ···················· | ···················· | ····················· | ···················· | ···················· | ···················· |
| 5275 | MW273887.1 | \|Severe acute respiratory syndrome coro | ···················· | ···················· | ····················· | ···················· | ···················· | ···················· |
| 5276 | MW273888.1 | \|Severe acute respiratory syndrome coro | ···················· | ···················· | ····················· | ···················· | ···················· | ···················· |
| 5277 | MW273889.1 | \|Severe acute respiratory syndrome coro | ···················· | ···················· | ····················· | ···················· | ···················· | ···················· |
| 5278 | MW273890.1 | \|Severe acute respiratory syndrome coro | ···················· | ···················· | ····················· | ···················· | ···················· | ···················· |
| 5279 | MW273891.1 | \|Severe acute respiratory syndrome coro | ···················· | ···················· | ····················· | ···················· | ···················· | ···················· |
| 5280 | MW273892.1 | \|Severe acute respiratory syndrome coro | ···················· | ···················· | ····················· | ···················· | ···················· | ···················· |
| 5281 | MW265603.1 | \|Severe acute respiratory syndrome coro | ···················· | ···················· | ····················· | ···················· | ···················· | ···················· |
| 5282 | MW266938.1 | \|Severe acute respiratory syndrome coro | ···················· | ···················· | ····················· | ···················· | ···················· | ···················· |
| 5283 | MW266939.1 | \|Severe acute respiratory syndrome coro | ···················· | ···················· | ····················· | ···················· | ···················· | ···················· |
| 5284 | MW266940.1 | \|Severe acute respiratory syndrome coro | ···················· | ···················· | ····················· | ···················· | ···················· | ···················· |
| 5285 | MW266941.1 | \|Severe acute respiratory syndrome coro | ···················· | ···················· | ····················· | ···················· | ···················· | ···················· |
| 5286 | MW266942.1 | \|Severe acute respiratory syndrome coro | ···················· | ···················· | ····················· | ···················· | ···················· | ···················· |
| 5287 | MW266943.1 | \|Severe acute respiratory syndrome coro | ···················· | ···················· | ····················· | ···················· | ···················· | ···················· |
| 5288 | MW266944.1 | \|Severe acute respiratory syndrome coro | ···················· | ···················· | ····················· | ···················· | ···················· | ···················· |
| 5289 | MW266945.1 | \|Severe acute respiratory syndrome coro | ···················· | ···················· | ····················· | ···················· | ···················· | ···················· |
| 5290 | MW266946.1 | \|Severe acute respiratory syndrome coro | ···················· | ···················· | ····················· | ···················· | ···················· | ···················· |
| 5291 | MW266947.1 | \|Severe acute respiratory syndrome coro | ···················· | ···················· | ····················· | ···················· | ···················· | ···················· |
| 5292 | MW266948.1 | \|Severe acute respiratory syndrome coro | ···················· | ···················· | ····················· | ···················· | ···················· | ···················· |
| 5293 | MW266949.1 | \|Severe acute respiratory syndrome coro | ···················· | ···················· | ····················· | ···················· | ···················· | ···················· |
| 5294 | MW266950.1 | \|Severe acute respiratory syndrome coro | ···················· | ···················· | ····················· | ···················· | ···················· | ···················· |
| 5295 | MW266951.1 | \|Severe acute respiratory syndrome coro | ···················· | ···················· | ····················· | ···················· | ···················· | ···················· |
| 5296 | MW266952.1 | \|Severe acute respiratory syndrome coro | ···················· | ···················· | ····················· | ···················· | ···················· | ···················· |
| 5297 | MW266953.1 | \|Severe acute respiratory syndrome coro | ···················· | ···················· | ····················· | ···················· | ···················· | ···················· |
| 5298 | MW266954.1 | \|Severe acute respiratory syndrome coro | ···················· | ···················· | ····················· | ···················· | ···················· | ···················· |
| 5299 | MW266955.1 | \|Severe acute respiratory syndrome coro | ···················· | ···················· | ····················· | ···················· | ···················· | ···················· |
| 5300 | MW266956.1 | \|Severe acute respiratory syndrome coro | ···················· | ···················· | ····················· | ···················· | ···················· | ···················· |
| 5301 | MW267131.1 | \|Severe acute respiratory syndrome coro | ···················· | ···················· | ····················· | ···················· | ···················· | ···················· |
| 5302 | MW268703.1 | \|Severe acute respiratory syndrome coro | ···················· | ···················· | ····················· | ···················· | ···················· | ···················· |
| 5303 | MW268704.1 | \|Severe acute respiratory syndrome coro | ···················· | ···················· | ····················· | ···················· | ···················· | ···················· |
| 5304 | MW268705.1 | \|Severe acute respiratory syndrome coro | ···················· | ···················· | ····················· | ···················· | ···················· | ···················· |
| 5305 | MW268706.1 | \|Severe acute respiratory syndrome coro | ···················· | ···················· | ····················· | ···················· | ···················· | ···················· |
| 5306 | MW268707.1 | \|Severe acute respiratory syndrome coro | ···T················ | ···················· | ····················· | ···················· | ···················· | ···················· |
| 5307 | MW268708.1 | \|Severe acute respiratory syndrome coro | ···T················ | ···················· | ····················· | ···················· | ···················· | ···················· |
| 5308 | MW268709.1 | \|Severe acute respiratory syndrome coro | ···················· | ···················· | ····················· | ···················· | ···················· | ···················· |
| 5309 | MW268710.1 | \|Severe acute respiratory syndrome coro | ···················· | ·A·················· | ····················· | ···················· | ···················· | ···················· |
| 5310 | MW268711.1 | \|Severe acute respiratory syndrome coro | ···················· | ···················· | ····················· | ···················· | ···················· | ···················· |
| 5311 | MW268712.1 | \|Severe acute respiratory syndrome coro | ···················· | ···················· | ····················· | ···················· | ···················· | ···················· |
| 5312 | MW268713.1 | \|Severe acute respiratory syndrome coro | ···················· | ···················· | ····················· | ···················· | ···················· | ···················· |
| 5313 | MW268715.1 | \|Severe acute respiratory syndrome coro | ···················· | ···················· | ····················· | ···················· | ···················· | ···················· |
| 5314 | MW268716.1 | \|Severe acute respiratory syndrome coro | ···················· | ···················· | ····················· | ···················· | ···················· | ···················· |
| 5315 | MW268717.1 | \|Severe acute respiratory syndrome coro | ···················· | ···················· | ····················· | ···················· | ···················· | ···················· |
| 5316 | MW268718.1 | \|Severe acute respiratory syndrome coro | ···················· | ···················· | ····················· | ···················· | ···················· | ···················· |
| 5317 | MW268720.1 | \|Severe acute respiratory syndrome coro | ···················· | ···················· | ····················· | ···················· | ···················· | ···················· |
| 5318 | MW268721.1 | \|Severe acute respiratory syndrome coro | ···················· | ···················· | ····················· | ···················· | ···················· | ···················· |
| 5319 | MW268722.1 | \|Severe acute respiratory syndrome coro | ···················· | ···················· | ····················· | ···················· | ···················· | ···················· |
| 5320 | MW268723.1 | \|Severe acute respiratory syndrome coro | ···················· | ···················· | ····················· | ···················· | ···················· | ···················· |
| 5321 | MW268724.1 | \|Severe acute respiratory syndrome coro | ···················· | ···················· | ····················· | ···················· | ···················· | ···················· |
| 5322 | MW268725.1 | \|Severe acute respiratory syndrome coro | ···················· | ·············A······ | ····················· | ···················· | ···················· | ···················· |
| 5323 | MW268726.1 | \|Severe acute respiratory syndrome coro | ···················· | ···················· | ····················· | ···················· | ···················· | ···················· |
| 5324 | MW268727.1 | \|Severe acute respiratory syndrome coro | ···················· | ···················· | ····················· | ···················· | ···················· | ···················· |
| 5325 | MW268728.1 | \|Severe acute respiratory syndrome coro | ···················· | ···················· | ····················· | ···················· | ···················· | ···················· |
| 5326 | MW264424.1 | \|Severe acute respiratory syndrome coro | ···················· | ···················· | ····················· | ···················· | ···················· | ···················· |
| 5327 | MW264435.1 | \|Severe acute respiratory syndrome coro | ···················· | ···················· | ····················· | ···················· | ···················· | ···················· |
| 5328 | MT982401.1 | \|Severe acute respiratory syndrome coro | ···················· | ···················· | ····················· | ···················· | ···················· | ···················· |
| 5329 | MT982402.1 | \|Severe acute respiratory syndrome coro | ···················· | ···················· | ····················· | ···················· | ···················· | ···················· |
| 5330 | MT982403.1 | \|Severe acute respiratory syndrome coro | ···················· | ···················· | ····················· | ···················· | ···················· | ···················· |
| 5331 | MT982404.1 | \|Severe acute respiratory syndrome coro | ···················· | ···················· | ····················· | ···················· | ···················· | ···················· |
| 5332 | MT982405.1 | \|Severe acute respiratory syndrome coro | ···················· | ···················· | ····················· | ···················· | ···················· | ···················· |
| 5333 | MT982406.1 | \|Severe acute respiratory syndrome coro | ···················· | ···················· | ····················· | ···················· | ···················· | ···················· |
| 5334 | MW255832.1 | \|Severe acute respiratory syndrome coro | ···················· | ···················· | ····················· | ···········T········ | ···················· | ···················· |

| | | | | | | | | | |
|---|---|---|---|---|---|---|---|---|---|
| 5335 | MW261767.1 | Severe acute respiratory syndrome coro | ···················· | ···················· | ···················· | ···················· | ···················· | ···················· | |
| 5336 | MW261770.1 | Severe acute respiratory syndrome coro | ···················· | ···················· | ···················· | ···················· | ···················· | ···················· | |
| 5337 | MW250244.1 | Severe acute respiratory syndrome coro | ···················· | ···················· | ···················· | ···················· | ···················· | ···················· | |
| 5338 | MW250352.1 | Severe acute respiratory syndrome coro | ···················· | ···················· | ···················· | ···················· | ···················· | ···················· | |
| 5339 | MW251511.1 | Severe acute respiratory syndrome coro | ···················· | ···················· | ···················· | ···················· | ···················· | ···················· | |
| 5340 | MW242950.1 | Severe acute respiratory syndrome coro | ···················· | ·A·················· | ···················· | ···················· | ···················· | ···A················ | |
| 5341 | MW242960.1 | Severe acute respiratory syndrome coro | ···················· | ·A·················· | ···················· | ···················· | ···················· | ···A················ | |
| 5342 | MW242961.1 | Severe acute respiratory syndrome coro | ···················· | ·A·················· | ···················· | ···················· | ···················· | ···A················ | |
| 5343 | MW242962.1 | Severe acute respiratory syndrome coro | ···················· | ···················· | ···················· | ···················· | ···················· | ···················· | |
| 5344 | MW242963.1 | Severe acute respiratory syndrome coro | ···················· | ·A·················· | ···················· | ···················· | ···················· | ···A················ | |
| 5345 | MW242964.1 | Severe acute respiratory syndrome coro | ···················· | ·A·················· | ···················· | ···················· | ···················· | ···A················ | |
| 5346 | MW242965.1 | Severe acute respiratory syndrome coro | ···················· | ·A·················· | ···················· | ···················· | ···················· | ···A················ | |
| 5347 | MW242999.1 | Severe acute respiratory syndrome coro | ···················· | ···················· | ···················· | ···················· | ···················· | ···················· | |
| 5348 | MW243003.1 | Severe acute respiratory syndrome coro | ···················· | ···················· | ···················· | ···················· | ···················· | ···················· | |
| 5349 | MW243586.1 | Severe acute respiratory syndrome coro | ···················· | ·A·················· | ···················· | ···················· | ···················· | ···A················ | |
| 5350 | MW243587.1 | Severe acute respiratory syndrome coro | ···················· | ·A·················· | ···················· | ···················· | ···················· | ···A················ | |
| 5351 | MW243995.1 | Severe acute respiratory syndrome coro | ···················· | ···················· | ···················· | ···················· | ···················· | ···················· | |
| 5352 | MW243996.1 | Severe acute respiratory syndrome coro | ···················· | ···················· | ···················· | ···················· | ···················· | ···················· | |
| 5353 | MW243997.1 | Severe acute respiratory syndrome coro | ···················· | ···················· | ···················· | ···················· | ···················· | ···················· | |
| 5354 | MW243998.1 | Severe acute respiratory syndrome coro | ···················· | ···················· | ···················· | ···················· | ···················· | ···················· | |
| 5355 | MW243999.1 | Severe acute respiratory syndrome coro | ···················· | ···················· | ···················· | ···················· | ···················· | ···················· | |
| 5356 | MW244000.1 | Severe acute respiratory syndrome coro | ···················· | ···················· | ···················· | ···················· | ···················· | ···················· | |
| 5357 | MW244001.1 | Severe acute respiratory syndrome coro | ···················· | ···················· | ···················· | ···················· | ···················· | ···················· | |
| 5358 | MW244002.1 | Severe acute respiratory syndrome coro | ···················· | ···················· | ···················· | ···················· | ···················· | ···················· | |
| 5359 | MW244003.1 | Severe acute respiratory syndrome coro | ···················· | ···················· | ···················· | ···················· | ···················· | ···················· | |
| 5360 | MW244004.1 | Severe acute respiratory syndrome coro | ···················· | ···················· | ···················· | ···················· | ···················· | ···················· | |
| 5361 | MW244005.1 | Severe acute respiratory syndrome coro | ···················· | ···················· | ···················· | ···················· | ···················· | ···················· | |
| 5362 | MW244006.1 | Severe acute respiratory syndrome coro | ···················· | ···················· | ···················· | ···················· | ···················· | ···················· | |
| 5363 | MW244007.1 | Severe acute respiratory syndrome coro | ···················· | ···················· | ···················· | ···················· | ···················· | ···················· | |
| 5364 | MW244008.1 | Severe acute respiratory syndrome coro | ···················· | ···················· | ···················· | ···················· | ···················· | ···················· | |
| 5365 | MW244009.1 | Severe acute respiratory syndrome coro | ···················· | ···················· | ···················· | ···················· | ···················· | ···················· | |
| 5366 | MW244010.1 | Severe acute respiratory syndrome coro | ···················· | ···················· | ···················· | ···················· | ···················· | ···················· | |
| 5367 | MW244011.1 | Severe acute respiratory syndrome coro | ···················· | ·A·················· | ···················· | ···················· | ···················· | ···················· | |
| 5368 | MW244012.1 | Severe acute respiratory syndrome coro | ···················· | ·A·················· | ···················· | ···················· | ···················· | ···················· | |
| 5369 | MW244013.1 | Severe acute respiratory syndrome coro | ···················· | ···················· | ···················· | ···················· | ···················· | ···················· | |
| 5370 | MW244014.1 | Severe acute respiratory syndrome coro | ···················· | ······A············· | ···················· | ···················· | ···················· | ···················· | |
| 5371 | MW244015.1 | Severe acute respiratory syndrome coro | ···················· | ···················· | ···················· | ···················· | ···················· | ···················· | |
| 5372 | MW244016.1 | Severe acute respiratory syndrome coro | ···················· | ···················· | ···················· | ···················· | ···················· | ···················· | |
| 5373 | MW244017.1 | Severe acute respiratory syndrome coro | ···················· | ············C······· | ···················· | ···················· | ···················· | ···················· | |
| 5374 | MW244018.1 | Severe acute respiratory syndrome coro | ···················· | ·A·················· | ···················· | ···················· | ···················· | ···················· | |
| 5375 | MW244019.1 | Severe acute respiratory syndrome coro | ···················· | ···················· | ···················· | ···················· | ···················· | ···················· | |
| 5376 | MW246592.1 | Severe acute respiratory syndrome coro | ···················· | ···················· | ···················· | ···················· | ···················· | ···················· | |
| 5377 | MW246593.1 | Severe acute respiratory syndrome coro | ···················· | ···················· | ···················· | ···················· | ···················· | ···················· | |
| 5378 | MW246594.1 | Severe acute respiratory syndrome coro | ···················· | ···················· | ···················· | ···················· | ···················· | ···················· | |
| 5379 | MW246595.1 | Severe acute respiratory syndrome coro | ···················· | ·············A······ | ···················· | ···················· | ·NNN················ | ···················· | 1 |
| 5380 | MW246596.1 | Severe acute respiratory syndrome coro | ···················· | ·············A······ | ···················· | ···················· | ···················· | ···················· | |
| 5381 | MW246597.1 | Severe acute respiratory syndrome coro | ···················· | ·A·················· | ···················· | ···················· | ···················· | ···················· | |
| 5382 | MW246598.1 | Severe acute respiratory syndrome coro | ···················· | ·············A······ | ···················· | ···················· | ···················· | ···················· | |
| 5383 | MW246599.1 | Severe acute respiratory syndrome coro | ···················· | ·············A······ | ···················· | ···················· | ···················· | ···················· | |
| 5384 | MW246600.1 | Severe acute respiratory syndrome coro | ···················· | ·············A······ | ···················· | ···················· | ···················· | ···················· | |
| 5385 | MW246601.1 | Severe acute respiratory syndrome coro | ···················· | ·············A······ | ···················· | ···················· | ···················· | ···················· | |
| 5386 | MW246602.1 | Severe acute respiratory syndrome coro | ···················· | ···················· | ···················· | ···················· | ···················· | ···················· | |
| 5387 | MW246603.1 | Severe acute respiratory syndrome coro | ···················· | ···················· | ···················· | ···················· | ···················· | ···················· | |
| 5388 | MW246604.1 | Severe acute respiratory syndrome coro | ···················· | ···················· | ···················· | ···················· | ···················· | ···················· | |
| 5389 | MW246605.1 | Severe acute respiratory syndrome coro | ···················· | ···················· | ···················· | ···················· | ···················· | ···················· | |
| 5390 | MW246606.1 | Severe acute respiratory syndrome coro | ···················· | ···················· | ···················· | ···················· | ···················· | ···················· | |
| 5391 | MW246607.1 | Severe acute respiratory syndrome coro | ···················· | ···················· | ···················· | ···················· | ···················· | ···················· | |
| 5392 | MW246608.1 | Severe acute respiratory syndrome coro | ···················· | ···················· | ···················· | ···················· | ···················· | ···················· | |
| 5393 | MW246609.1 | Severe acute respiratory syndrome coro | ···················· | ·············A······ | ···················· | ···················· | ···················· | ···················· | |
| 5394 | MW246610.1 | Severe acute respiratory syndrome coro | ···················· | ···················· | ···················· | ···················· | ···················· | ···················· | |
| 5395 | MW246611.1 | Severe acute respiratory syndrome coro | ···················· | ···················· | ···················· | ···················· | ···················· | ···················· | |
| 5396 | MW246612.1 | Severe acute respiratory syndrome coro | ···················· | ···················· | ···················· | ···················· | ···················· | ···················· | |
| 5397 | MW246613.1 | Severe acute respiratory syndrome coro | ···················· | ···················· | ···················· | ···················· | ···················· | ···················· | |
| 5398 | MW246614.1 | Severe acute respiratory syndrome coro | ···················· | ·············A······ | ···················· | ···················· | ···················· | ···················· | |
| 5399 | MW246615.1 | Severe acute respiratory syndrome coro | ···················· | ·············A······ | ···················· | ···················· | ···················· | ···················· | |
| 5400 | MW246616.1 | Severe acute respiratory syndrome coro | ···················· | ·············A······ | ···················· | ···················· | ···················· | ···················· | |
| 5401 | MW246617.1 | Severe acute respiratory syndrome coro | ···················· | ·············A······ | ···················· | ···················· | ···················· | ···················· | |
| 5402 | MW246618.1 | Severe acute respiratory syndrome coro | ···················· | ·············A······ | ···················· | ···················· | ···················· | ···················· | |
| 5403 | MW246619.1 | Severe acute respiratory syndrome coro | ···················· | ·············A······ | ···················· | ···················· | ···················· | ···················· | |
| 5404 | MW246620.1 | Severe acute respiratory syndrome coro | ···················· | ·············A······ | ···················· | ···················· | ···················· | ···················· | |
| 5405 | MW246621.1 | Severe acute respiratory syndrome coro | ···················· | ·············A······ | ···················· | ···················· | ···················· | ···················· | |
| 5406 | MW246622.1 | Severe acute respiratory syndrome coro | ···················· | ·············A······ | ···················· | ···················· | ···················· | ···················· | |
| 5407 | MW246623.1 | Severe acute respiratory syndrome coro | ···················· | ·············A······ | ···················· | ···················· | ···················· | ···················· | |
| 5408 | MW246624.1 | Severe acute respiratory syndrome coro | ···················· | ·············A······ | ···················· | ···················· | ···················· | ···················· | |
| 5409 | MW246625.1 | Severe acute respiratory syndrome coro | ···················· | ·············A······ | ···················· | ···················· | ···················· | ···················· | |
| 5410 | MW246626.1 | Severe acute respiratory syndrome coro | ···················· | ·············A······ | ···················· | ···················· | ··N················· | ···················· | |
| 5411 | MW246627.1 | Severe acute respiratory syndrome coro | ···················· | ·············A······ | ···················· | ···················· | ···················· | ···················· | |
| 5412 | MW246628.1 | Severe acute respiratory syndrome coro | ···················· | ·············A······ | ···················· | ···················· | ···················· | ···················· | |

5413 MW246629.1 |Severe acute respiratory syndrome coro ···················· ·A·················· ····················· ···················· ···················· ····················
5414 MW242647.1 |Severe acute respiratory syndrome coro ···················· ·A·················· ····················· ···················· ···················· ····················
5415 MW242649.1 |Severe acute respiratory syndrome coro ···················· ···················· ····················· ···················· ···················· ····················
5416 MW242650.1 |Severe acute respiratory syndrome coro ···················· ···················· ····················· ···················· ···················· ····················
5417 MW242651.1 |Severe acute respiratory syndrome coro ···················· ···················· ····················· ···················· ···················· ····················
5418 MW242652.1 |Severe acute respiratory syndrome coro ···················· ···················· ····················· ···················· ···················· ····················
5419 MW242653.1 |Severe acute respiratory syndrome coro ···················· ·A·················· ····················· ···················· ···················· ··A·················
5420 MW242655.1 |Severe acute respiratory syndrome coro ···················· ···················· ····················· ···················· ···················· ····················
5421 MW242663.1 |Severe acute respiratory syndrome coro ···················· ·A·················· ····················· ···················· ···················· ··A·················
5422 MW242666.1 |Severe acute respiratory syndrome coro ···················· ·A·················· ····················· ···················· ···················· ··A·················
5423 MW242667.1 |Severe acute respiratory syndrome coro ···················· ···················· ····················· ···················· ···················· ····················
5424 MW242670.1 |Severe acute respiratory syndrome coro ···················· ·A·················· ····················· ···················· ···················· ··A·················
5425 MW242671.1 |Severe acute respiratory syndrome coro ···················· ·A·················· ····················· ···················· ···················· ··A·················
5426 MW242689.1 |Severe acute respiratory syndrome coro ···················· ·A·················· ····················· ···················· ···················· ····················
5427 MW242691.1 |Severe acute respiratory syndrome coro ···················· ···················· ····················· ···················· ···················· ····················
5428 MW242692.1 |Severe acute respiratory syndrome coro ···················· ···················· ····················· ···················· ···················· ····················
5429 MW242704.1 |Severe acute respiratory syndrome coro ···················· ···················· ····················· ···················· ···················· ····················
5430 MW242705.1 |Severe acute respiratory syndrome coro ···················· ···················· ····················· ···················· ···················· ····················
5431 MW242706.1 |Severe acute respiratory syndrome coro ···················· ·A·················· ····················· ···················· ···················· ····················
5432 MW242707.1 |Severe acute respiratory syndrome coro ···················· ···················· ····················· ···················· ···················· ····················
5433 MW242708.1 |Severe acute respiratory syndrome coro ···················· ···················· ····················· ···················· ···················· ··A·················
5434 MW242709.1 |Severe acute respiratory syndrome coro ···················· ···················· ····················· ···················· ···················· ····················
5435 MW242710.1 |Severe acute respiratory syndrome coro ···················· ···················· ····················· ···················· ···················· ····················
5436 MW242778.1 |Severe acute respiratory syndrome coro ···················· ·A·················· ····················· ···················· ···················· ··A·················
5437 MW242779.1 |Severe acute respiratory syndrome coro ···················· ·A·················· ····················· ···················· ···················· ··A·················
5438 MW242780.1 |Severe acute respiratory syndrome coro ···················· ···················· ····················· ···················· ···················· ····················
5439 MW242781.1 |Severe acute respiratory syndrome coro ···················· ···················· ····················· ···················· ···················· ····················
5440 MW242786.1 |Severe acute respiratory syndrome coro ···················· ···················· ····················· ···················· ···················· ····················
5441 MW185823.1 |Severe acute respiratory syndrome coro ···················· ···················· ····················· ···················· ···················· ····················
5442 MW185824.1 |Severe acute respiratory syndrome coro ···················· ···················· ····················· ···················· ···················· ····················
5443 MW185825.1 |Severe acute respiratory syndrome coro ···················· ···················· ····················· ···················· ···················· ····················
5444 MW185826.1 |Severe acute respiratory syndrome coro ···················· ···················· ····················· ···················· ···················· ····················
5445 MW185827.1 |Severe acute respiratory syndrome coro ···················· ···················· ····················· ···················· ···················· ····················
5446 MW237708.1 |Severe acute respiratory syndrome coro ···················· ···················· ····················· ···················· ···················· ····················
5447 MW237714.1 |Severe acute respiratory syndrome coro ···················· ···················· ····················· ···················· ···················· ····················
5448 MW237715.1 |Severe acute respiratory syndrome coro ···················· ···················· ····················· ···················· ···················· ····················
5449 MW237717.1 |Severe acute respiratory syndrome coro ···················· ···················· ····················· ···················· ···················· ····················
5450 MW237738.1 |Severe acute respiratory syndrome coro ···················· ···················· ····················· ···················· ···················· ····················
5451 MW237739.1 |Severe acute respiratory syndrome coro ···················· ···················· ····················· ···················· ···················· ····················
5452 MW237740.1 |Severe acute respiratory syndrome coro ···················· ···················· ····················· ···················· ···················· ····················
5453 MW237741.1 |Severe acute respiratory syndrome coro ···················· ···················· ····················· ···················· ···················· ····················
5454 MW237743.1 |Severe acute respiratory syndrome coro ···················· ···················· ····················· ···················· ···················· ····················
5455 MW237767.1 |Severe acute respiratory syndrome coro ···················· ···················· ····················· ···················· ···················· ····················
5456 MW237768.1 |Severe acute respiratory syndrome coro ···················· ···················· ····················· ···················· ···················· ····················
5457 MW240719.1 |Severe acute respiratory syndrome coro ···················· ···················· ····················· ···················· ···················· ····················
5458 MW240720.1 |Severe acute respiratory syndrome coro ···················· ···················· ····················· ···················· ···················· ····················
5459 MW240721.1 |Severe acute respiratory syndrome coro ···················· ···················· ····················· ···················· ···················· ····················
5460 MW240723.1 |Severe acute respiratory syndrome coro ···················· ·············A······ ····················· ···················· ···················· ····················
5461 MW240724.1 |Severe acute respiratory syndrome coro ···················· ·A·················· ····················· ···················· ···················· ····················
5462 MW240725.1 |Severe acute respiratory syndrome coro ···················· ·A·················· ····················· ···················· ···················· ····················
5463 MW240726.1 |Severe acute respiratory syndrome coro ···················· ·A·················· ····················· ···················· ···················· ····················
5464 MW240727.1 |Severe acute respiratory syndrome coro ···················· ·A·················· ····················· ···················· ···················· ····················
5465 MW240728.1 |Severe acute respiratory syndrome coro ···················· ·A·················· ····················· ···················· ···················· ····················
5466 MW240729.1 |Severe acute respiratory syndrome coro ···················· ·A·················· ····················· ···················· ···················· ····················
5467 MW240730.1 |Severe acute respiratory syndrome coro ···················· ·A·················· ····················· ···················· ···················· ····················
5468 MW240731.1 |Severe acute respiratory syndrome coro ···················· ·A·················· ····················· ···················· ···················· ····················
5469 MW240732.1 |Severe acute respiratory syndrome coro ···················· ·A·················· ····················· ···················· ···················· ····················
5470 MW240733.1 |Severe acute respiratory syndrome coro ···················· ·A·················· ····················· ···················· ···················· ····················
5471 MW240734.1 |Severe acute respiratory syndrome coro ···················· ·A·················· ····················· ···················· ···················· ····················
5472 MW240735.1 |Severe acute respiratory syndrome coro ···················· ·A·················· ····················· ···················· ···················· ····················
5473 MW240736.1 |Severe acute respiratory syndrome coro ···················· ·A·················· ····················· ···················· ···················· ····················
5474 MW240737.1 |Severe acute respiratory syndrome coro ···················· ·A·················· ····················· ···················· ···················· ····················
5475 MW240738.1 |Severe acute respiratory syndrome coro ···················· ·A·················· ····················· ···················· ···················· ····················
5476 MW240739.1 |Severe acute respiratory syndrome coro ···················· ·A·················· ····················· ···················· ···················· ····················
5477 MW240740.1 |Severe acute respiratory syndrome coro ···················· ·A·················· ····················· ···················· ···················· ····················
5478 MW240741.1 |Severe acute respiratory syndrome coro ···················· ·A·················· ····················· ···················· ···················· ····················
5479 MW240742.1 |Severe acute respiratory syndrome coro ···················· ·A·················· ····················· ···················· ···················· ····················
5480 MW240743.1 |Severe acute respiratory syndrome coro ···················· ···················· ····················· ···················· ···················· ····················
5481 MW240744.1 |Severe acute respiratory syndrome coro ···················· ···················· ····················· ···················· ···················· ····················
5482 MW240745.1 |Severe acute respiratory syndrome coro ···················· ···················· ····················· ···················· ···················· ····················
5483 MW240746.1 |Severe acute respiratory syndrome coro ···················· ···················· ····················· ···················· ···················· ····················
5484 MW240747.1 |Severe acute respiratory syndrome coro ···················· ·A·················· ····················· ···················· ···················· ····················
5485 MW240748.1 |Severe acute respiratory syndrome coro ···················· ···················· ····················· ···················· ···················· ····················
5486 MW240749.1 |Severe acute respiratory syndrome coro ···················· ···················· ····················· ···················· ···················· ····················
5487 MW240750.1 |Severe acute respiratory syndrome coro ···················· ···················· ····················· ···················· ···················· ····················
5488 MW240751.1 |Severe acute respiratory syndrome coro ···················· ···················· ····················· ···················· ···················· ····················
5489 MW240752.1 |Severe acute respiratory syndrome coro ···················· ···················· ····················· ···················· ···················· ····················
5490 MW240753.1 |Severe acute respiratory syndrome coro ···················· ············C······· ····················· ···················· ···················· ····················

| | | | | | | | |
|---|---|---|---|---|---|---|---|
| 5491 | MW240754.1 \|Severe acute respiratory syndrome coro | ···················· | ·A·················· | ···················· | ···················· | ···················· | ···················· |
| 5492 | MW240755.1 \|Severe acute respiratory syndrome coro | ···················· | ·A·················· | ···················· | ···················· | ···················· | ···················· |
| 5493 | MW240756.1 \|Severe acute respiratory syndrome coro | ···················· | ·A·················· | ···················· | ···················· | ···················· | ···················· |
| 5494 | MW240757.1 \|Severe acute respiratory syndrome coro | ···················· | ·A·················· | ···················· | ···················· | ···················· | ···················· |
| 5495 | MW240758.1 \|Severe acute respiratory syndrome coro | ···················· | ·A·················· | ···················· | ···················· | ···················· | ···················· |
| 5496 | MW240759.1 \|Severe acute respiratory syndrome coro | ···················· | ···················· | ···················· | ···················· | ···················· | ···················· |
| 5497 | MW240760.1 \|Severe acute respiratory syndrome coro | ···················· | ···················· | ···················· | ···················· | ···················· | ···················· |
| 5498 | MW240761.1 \|Severe acute respiratory syndrome coro | ···················· | ···················· | ···················· | ···················· | ···················· | ···················· |
| 5499 | MW240762.1 \|Severe acute respiratory syndrome coro | ···················· | ···················· | ···················· | ···················· | ···················· | ···················· |
| 5500 | MW240763.1 \|Severe acute respiratory syndrome coro | ···················· | ···················· | ···················· | ···················· | ···················· | ···················· |
| 5501 | MW241170.1 \|Severe acute respiratory syndrome coro | ···················· | ···················· | ···················· | ···················· | ···················· | ···················· |
| 5502 | MW241172.1 \|Severe acute respiratory syndrome coro | ···················· | ···················· | ···················· | ···················· | ···················· | ···················· |
| 5503 | MW241178.1 \|Severe acute respiratory syndrome coro | ···················· | ···················· | ···················· | ···················· | ···················· | ···················· |
| 5504 | MW241179.1 \|Severe acute respiratory syndrome coro | ···················· | ···················· | ···················· | ···················· | ···················· | ···················· |
| 5505 | MW241189.1 \|Severe acute respiratory syndrome coro | ···················· | ···················· | ···················· | ···················· | ···················· | ···················· |
| 5506 | MW241191.1 \|Severe acute respiratory syndrome coro | ···················· | ···················· | ···················· | ···················· | ···················· | ···················· |
| 5507 | MW241201.1 \|Severe acute respiratory syndrome coro | ···················· | ···················· | ···················· | ···················· | ···················· | ···················· |
| 5508 | MW241203.1 \|Severe acute respiratory syndrome coro | ···················· | ···················· | ···················· | ···················· | ···················· | ···················· |
| 5509 | MW241204.1 \|Severe acute respiratory syndrome coro | ···················· | ···················· | ···················· | ···················· | ···················· | ···················· |
| 5510 | MW241336.1 \|Severe acute respiratory syndrome coro | ···················· | ···················· | ···················· | ···················· | ···················· | ···················· |
| 5511 | MW241344.1 \|Severe acute respiratory syndrome coro | ···················· | ···················· | ···················· | ···················· | ···················· | ···················· |
| 5512 | MW241347.1 \|Severe acute respiratory syndrome coro | ···················· | ···················· | ···················· | ···················· | ···················· | ···················· |
| 5513 | MW241350.1 \|Severe acute respiratory syndrome coro | ···················· | ···················· | ···················· | ···················· | ···················· | ···················· |
| 5514 | MW241361.1 \|Severe acute respiratory syndrome coro | ···················· | ···················· | ···················· | ···················· | ···················· | ···················· |
| 5515 | MW241384.1 \|Severe acute respiratory syndrome coro | ···················· | ···················· | ···················· | ···················· | ···················· | ···················· |
| 5516 | MW228050.1 \|Severe acute respiratory syndrome coro | ···················· | ···················· | ···················· | ···················· | ···················· | ···················· |
| 5517 | MW228051.1 \|Severe acute respiratory syndrome coro | ···················· | ···················· | ···················· | ···················· | ···················· | ···················· |
| 5518 | MW228052.1 \|Severe acute respiratory syndrome coro | ···················· | ···················· | ···················· | ···················· | ···················· | ···················· |
| 5519 | MW228053.1 \|Severe acute respiratory syndrome coro | ···················· | ···················· | ···················· | ···················· | ···················· | ···················· |
| 5520 | MW228054.1 \|Severe acute respiratory syndrome coro | ···················· | ···················· | ···················· | ···················· | ···················· | ···················· |
| 5521 | MW228055.1 \|Severe acute respiratory syndrome coro | ···················· | ···················· | ···················· | ···················· | ···················· | ··A················· |
| 5522 | MW228056.1 \|Severe acute respiratory syndrome coro | ···················· | ···················· | ···················· | ···················· | ···················· | ··A················· |
| 5523 | MW228070.1 \|Severe acute respiratory syndrome coro | ···················· | ···A················ | ···················· | ···················· | ···················· | ···················· |
| 5524 | MW228218.1 \|Severe acute respiratory syndrome coro | ···················· | ···················· | ···················· | ···················· | ···················· | ···················· |
| 5525 | MW228219.1 \|Severe acute respiratory syndrome coro | ···················· | ···················· | ···················· | ···················· | ···················· | ···················· |
| 5526 | MW228221.1 \|Severe acute respiratory syndrome coro | ···················· | ···················· | ···················· | ···················· | ···················· | ···················· |
| 5527 | MW228222.1 \|Severe acute respiratory syndrome coro | ···················· | ·A·················· | ···················· | ···················· | ···················· | ···················· |
| 5528 | MW228223.1 \|Severe acute respiratory syndrome coro | ···················· | ···················· | ···················· | ···················· | ···················· | ···················· |
| 5529 | MW228224.1 \|Severe acute respiratory syndrome coro | ···················· | ···A················ | ···················· | ···················· | ···················· | ···················· |
| 5530 | MW228225.1 \|Severe acute respiratory syndrome coro | ···················· | ···················· | ···················· | ···················· | ···················· | ···················· |
| 5531 | MW228226.1 \|Severe acute respiratory syndrome coro | ···················· | ···················· | ···················· | ···················· | ···················· | ···················· |
| 5532 | MW228227.1 \|Severe acute respiratory syndrome coro | ···················· | ·A·················· | ···················· | ···················· | ···················· | ···················· |
| 5533 | MW228228.1 \|Severe acute respiratory syndrome coro | ···················· | ···················· | ···················· | ···················· | ···················· | ···················· |
| 5534 | MW228229.1 \|Severe acute respiratory syndrome coro | ···················· | ···················· | ···················· | ···················· | ···················· | ···················· |
| 5535 | MW228230.1 \|Severe acute respiratory syndrome coro | ···················· | ···················· | ···················· | ···················· | ···················· | ···················· |
| 5536 | MW228231.1 \|Severe acute respiratory syndrome coro | ···················· | ···················· | ···················· | ···················· | ···················· | ···················· |
| 5537 | MW228232.1 \|Severe acute respiratory syndrome coro | ···················· | ···················· | ···················· | ···················· | ···················· | ···················· |
| 5538 | MW228233.1 \|Severe acute respiratory syndrome coro | ···················· | ···················· | ···················· | ···················· | ···················· | ···················· |
| 5539 | MW228235.1 \|Severe acute respiratory syndrome coro | ···················· | ···················· | ···················· | ···················· | ···················· | ···················· |
| 5540 | MW228236.1 \|Severe acute respiratory syndrome coro | ···················· | ···················· | ···················· | ···················· | ···················· | ···················· |
| 5541 | MW228237.1 \|Severe acute respiratory syndrome coro | ···················· | ···················· | ···················· | ···················· | ···················· | ···················· |
| 5542 | MW228238.1 \|Severe acute respiratory syndrome coro | ···················· | ·A·················· | ···················· | ···················· | ···················· | ···················· |
| 5543 | MW228239.1 \|Severe acute respiratory syndrome coro | ···················· | ·A·················· | ···················· | ···················· | ···················· | ···················· |
| 5544 | MW228240.1 \|Severe acute respiratory syndrome coro | ···················· | ···················· | ···················· | ···················· | ···················· | ···················· |
| 5545 | MW228241.1 \|Severe acute respiratory syndrome coro | ···················· | ···················· | ···················· | ···················· | ···················· | ···················· |
| 5546 | MW228242.1 \|Severe acute respiratory syndrome coro | ···················· | ···················· | ···················· | ···················· | ···················· | ···················· |
| 5547 | MW228243.1 \|Severe acute respiratory syndrome coro | ···················· | ···················· | ···················· | ···················· | ···················· | ···················· |
| 5548 | MW228244.1 \|Severe acute respiratory syndrome coro | ···················· | ·A·················· | ···················· | ···················· | ···················· | ···················· |
| 5549 | MW228246.1 \|Severe acute respiratory syndrome coro | ···················· | ·A·················· | ···················· | ···················· | ···················· | ···················· |
| 5550 | MW228247.1 \|Severe acute respiratory syndrome coro | ···················· | ·A·················· | ···················· | ···················· | ···················· | ···················· |
| 5551 | MW228248.1 \|Severe acute respiratory syndrome coro | ···················· | ···················· | ···················· | ···················· | ···················· | ···················· |
| 5552 | MW228249.1 \|Severe acute respiratory syndrome coro | ···················· | ···················· | ···················· | ···················· | ···················· | ···················· |
| 5553 | MW228250.1 \|Severe acute respiratory syndrome coro | ···················· | ···················· | ···················· | ···················· | ···················· | ···················· |
| 5554 | MW228251.1 \|Severe acute respiratory syndrome coro | ···················· | ···················· | ···················· | ···················· | ···················· | ···················· |
| 5555 | MW228252.1 \|Severe acute respiratory syndrome coro | ···················· | ···················· | ···················· | ···················· | ···················· | ···················· |
| 5556 | MW228254.1 \|Severe acute respiratory syndrome coro | ···················· | ···················· | ···················· | ···················· | ···················· | ···················· |
| 5557 | MW228255.1 \|Severe acute respiratory syndrome coro | ···················· | ·A·················· | ···················· | ···················· | ···················· | ···················· |
| 5558 | MW228256.1 \|Severe acute respiratory syndrome coro | ···················· | ···················· | ···················· | ···················· | ···················· | ···················· |
| 5559 | MW228257.1 \|Severe acute respiratory syndrome coro | ···················· | ···················· | ···················· | ···················· | ···················· | ···················· |
| 5560 | MW228258.1 \|Severe acute respiratory syndrome coro | ···················· | ·A·················· | ···················· | ···················· | ···················· | ···················· |
| 5561 | MW228259.1 \|Severe acute respiratory syndrome coro | ···················· | ···················· | ···················· | ···················· | ···················· | ···················· |
| 5562 | MW228260.1 \|Severe acute respiratory syndrome coro | ···················· | ···················· | ···················· | ···················· | ···················· | ···················· |
| 5563 | MW228261.1 \|Severe acute respiratory syndrome coro | ···················· | ·A·················· | ···················· | ···················· | ···················· | ···················· |
| 5564 | MW228262.1 \|Severe acute respiratory syndrome coro | ···················· | ···················· | ···················· | ···················· | ···················· | ···················· |
| 5565 | MW228263.1 \|Severe acute respiratory syndrome coro | ···················· | ···················· | ···················· | ···················· | ···················· | ···················· |
| 5566 | MW228264.1 \|Severe acute respiratory syndrome coro | ···················· | ···················· | ···················· | ···················· | ···················· | ···················· |
| 5567 | MW228265.1 \|Severe acute respiratory syndrome coro | ···················· | ···················· | ···················· | ···················· | ···················· | ···················· |
| 5568 | MW228266.1 \|Severe acute respiratory syndrome coro | ···················· | ···················· | ···················· | ···················· | ···················· | ···················· |

| | | | | | | | | |
|---|---|---|---|---|---|---|---|---|
| 5569 | MW228267.1 | \|Severe acute respiratory syndrome coro | ···················· | ···················· | ···················· | ···················· | ···················· | ···················· |
| 5570 | MW228268.1 | \|Severe acute respiratory syndrome coro | ···················· | ···················· | ···················· | ···················· | ···················· | ···················· |
| 5571 | MW228269.1 | \|Severe acute respiratory syndrome coro | ···················· | ···················· | ···················· | ···················· | ···················· | ···················· |
| 5572 | MW228270.1 | \|Severe acute respiratory syndrome coro | ···················· | ···················· | ···················· | ···················· | ···················· | ···················· |
| 5573 | MW228271.1 | \|Severe acute respiratory syndrome coro | ···················· | ···················· | ···················· | ···················· | ···················· | ···················· |
| 5574 | MW228272.1 | \|Severe acute respiratory syndrome coro | ···················· | ···················· | ···················· | ···················· | ···················· | ···················· |
| 5575 | MW228273.1 | \|Severe acute respiratory syndrome coro | ···················· | ···················· | ···················· | ···················· | ···················· | ···················· |
| 5576 | MW228274.1 | \|Severe acute respiratory syndrome coro | ···················· | ···················· | ···················· | ···················· | ···················· | ···················· |
| 5577 | MW228275.1 | \|Severe acute respiratory syndrome coro | ···················· | ···················· | ···················· | ···················· | ···················· | ···················· |
| 5578 | MW228276.1 | \|Severe acute respiratory syndrome coro | ···················· | ···················· | ···················· | ···················· | ···················· | ···················· |
| 5579 | MW228277.1 | \|Severe acute respiratory syndrome coro | ···················· | ···················· | ···················· | ···················· | ···················· | ···················· |
| 5580 | MW228279.1 | \|Severe acute respiratory syndrome coro | ···················· | ···················· | ···················· | ···················· | ···················· | ···················· |
| 5581 | MW228280.1 | \|Severe acute respiratory syndrome coro | ···················· | ···················· | ···················· | ···················· | ···················· | ···················· |
| 5582 | MW229264.1 | \|Severe acute respiratory syndrome coro | ···················· | ···················· | ···················· | ···················· | ···················· | ···················· |
| 5583 | MW227482.1 | \|Severe acute respiratory syndrome coro | ···················· | ···················· | ···················· | ···················· | ···················· | ···················· |
| 5584 | MW227483.1 | \|Severe acute respiratory syndrome coro | ···················· | ···················· | ···················· | ···················· | ···················· | ···················· |
| 5585 | MW227484.1 | \|Severe acute respiratory syndrome coro | ···················· | ···················· | ···················· | ···················· | ···················· | ···················· |
| 5586 | MW227490.1 | \|Severe acute respiratory syndrome coro | ···················· | ···················· | ···················· | ···················· | ···················· | ···················· |
| 5587 | MW227491.1 | \|Severe acute respiratory syndrome coro | ···················· | ···················· | ···················· | ···················· | ···················· | ···················· |
| 5588 | MW227492.1 | \|Severe acute respiratory syndrome coro | ···················· | ···················· | ···················· | ···················· | ···················· | ···················· |
| 5589 | MW227568.1 | \|Severe acute respiratory syndrome coro | ···················· | ·A·················· | ···················· | ···················· | ······T············· | ···················· |
| 5590 | MT576563.1 | \|Severe acute respiratory syndrome coro | ···················· | ···················· | ···················· | ···················· | ···················· | ···················· |
| 5591 | MT835139.1 | \|Severe acute respiratory syndrome coro | ···················· | ···················· | ···················· | ···················· | ···················· | ···················· |
| 5592 | MT835140.1 | \|Severe acute respiratory syndrome coro | ···················· | ·A·················· | ···················· | ···················· | ···················· | ···················· |
| 5593 | MT835141.1 | \|Severe acute respiratory syndrome coro | ···················· | ·A·················· | ···················· | ···················· | ···················· | ···················· |
| 5594 | MT835142.1 | \|Severe acute respiratory syndrome coro | ···················· | ···················· | ···················· | ···················· | ···················· | ···················· |
| 5595 | MT835143.1 | \|Severe acute respiratory syndrome coro | ···················· | ···················· | ···················· | ···················· | ···················· | ···················· |
| 5596 | MW217299.1 | \|Severe acute respiratory syndrome coro | ···················· | ···················· | ···················· | ···················· | ···················· | ···················· |
| 5597 | MW217300.1 | \|Severe acute respiratory syndrome coro | ···················· | ···················· | ···················· | ···················· | ···················· | ···················· |
| 5598 | MW217301.1 | \|Severe acute respiratory syndrome coro | ···················· | ···················· | ···················· | ···················· | ···················· | ···················· |
| 5599 | MW217302.1 | \|Severe acute respiratory syndrome coro | ···················· | ···················· | ···················· | ···················· | ···················· | ···················· |
| 5600 | MW217303.1 | \|Severe acute respiratory syndrome coro | ···················· | ···················· | ···················· | ···················· | ···················· | ···················· |
| 5601 | MW217304.1 | \|Severe acute respiratory syndrome coro | ···················· | ···A················ | ···················· | ···················· | ···················· | ···················· |
| 5602 | MW217305.1 | \|Severe acute respiratory syndrome coro | ···················· | ···················· | ···················· | ···················· | ···················· | ···················· |
| 5603 | MW217306.1 | \|Severe acute respiratory syndrome coro | ···················· | ···················· | ···················· | ···················· | ···················· | ···················· |
| 5604 | MW217308.1 | \|Severe acute respiratory syndrome coro | ···················· | ···················· | ···················· | ···················· | ···················· | ···················· |
| 5605 | MW217309.1 | \|Severe acute respiratory syndrome coro | ···················· | ·A·················· | ···················· | ···················· | ···················· | ···················· |
| 5606 | MW217310.1 | \|Severe acute respiratory syndrome coro | ···················· | ···················· | ···················· | ···················· | ···················· | ···················· |
| 5607 | MW217311.1 | \|Severe acute respiratory syndrome coro | ···················· | ···················· | ···················· | ···················· | ···················· | ···················· |
| 5608 | MW217313.1 | \|Severe acute respiratory syndrome coro | ···················· | ···················· | ···················· | ···················· | ···················· | ···················· |
| 5609 | MW217314.1 | \|Severe acute respiratory syndrome coro | ···················· | ···················· | ···················· | ···················· | ···················· | ···················· |
| 5610 | MW217315.1 | \|Severe acute respiratory syndrome coro | ···················· | ···················· | ···················· | ···················· | ···················· | ···················· |
| 5611 | MW217316.1 | \|Severe acute respiratory syndrome coro | ···················· | ···················· | ···················· | ···················· | ···················· | ···················· |
| 5612 | MW217317.1 | \|Severe acute respiratory syndrome coro | ···················· | ···················· | ···················· | ···················· | ···················· | ···················· |
| 5613 | MW217318.1 | \|Severe acute respiratory syndrome coro | ···················· | ···················· | ···················· | ···················· | ···················· | ···················· |
| 5614 | MW217319.1 | \|Severe acute respiratory syndrome coro | ···················· | ···················· | ···················· | ···················· | ···················· | ···················· |
| 5615 | MW217320.1 | \|Severe acute respiratory syndrome coro | ···················· | ·A·················· | ···················· | ···················· | ···················· | ···················· |
| 5616 | MW217321.1 | \|Severe acute respiratory syndrome coro | ···················· | ·A·················· | ···················· | ···················· | ···················· | ···················· |
| 5617 | MW217322.1 | \|Severe acute respiratory syndrome coro | ···················· | ·A·················· | ···················· | ···················· | ···················· | ···················· |
| 5618 | MW217323.1 | \|Severe acute respiratory syndrome coro | ···················· | ·A·················· | ···················· | ···················· | ···················· | ···················· |
| 5619 | MW217324.1 | \|Severe acute respiratory syndrome coro | ···················· | ·A·················· | ···················· | ···················· | ···················· | ···················· |
| 5620 | MW217325.1 | \|Severe acute respiratory syndrome coro | ···················· | ·A·················· | ···················· | ···················· | ···················· | ···················· |
| 5621 | MW217326.1 | \|Severe acute respiratory syndrome coro | ···················· | ·A·················· | ···················· | ···················· | ···················· | ···················· |
| 5622 | MW217327.1 | \|Severe acute respiratory syndrome coro | ···················· | ···················· | ···················· | ···················· | ···················· | ···················· |
| 5623 | MW217328.1 | \|Severe acute respiratory syndrome coro | ···················· | ···················· | ···················· | ···················· | ···················· | ···················· |
| 5624 | MW217330.1 | \|Severe acute respiratory syndrome coro | ···················· | ·A·················· | ···················· | ···················· | ···················· | ···················· |
| 5625 | MW217332.1 | \|Severe acute respiratory syndrome coro | ···················· | ···················· | ···················· | ···················· | ···················· | ···················· |
| 5626 | MW217334.1 | \|Severe acute respiratory syndrome coro | ···················· | ···················· | ···················· | ···················· | ···················· | ···················· |
| 5627 | MW217335.1 | \|Severe acute respiratory syndrome coro | ···················· | ···················· | ···················· | ···················· | ···················· | ···················· |
| 5628 | MW217336.1 | \|Severe acute respiratory syndrome coro | ···················· | ···················· | ···················· | ···················· | ···················· | ···················· |
| 5629 | MW217337.1 | \|Severe acute respiratory syndrome coro | ···················· | ·A·················· | ···················· | ···················· | ···················· | ···················· |
| 5630 | MW217338.1 | \|Severe acute respiratory syndrome coro | ···················· | ···················· | ···················· | ···················· | ···················· | ···················· |
| 5631 | MW217339.1 | \|Severe acute respiratory syndrome coro | ···················· | ···················· | ···················· | ···················· | ···················· | ···················· |
| 5632 | MW217340.1 | \|Severe acute respiratory syndrome coro | ···················· | ···················· | ···················· | ···················· | ···················· | ···················· |
| 5633 | MW217341.1 | \|Severe acute respiratory syndrome coro | ···················· | ···················· | ···················· | ···················· | ···················· | ···················· |
| 5634 | MW217343.1 | \|Severe acute respiratory syndrome coro | ···················· | ···················· | ···················· | ···················· | ···················· | ···················· |
| 5635 | MW217344.1 | \|Severe acute respiratory syndrome coro | ···················· | ···················· | ···················· | ···················· | ···················· | ···················· |
| 5636 | MW217345.1 | \|Severe acute respiratory syndrome coro | ···················· | ·A·················· | ···················· | ···················· | ···················· | ···················· |
| 5637 | MW217346.1 | \|Severe acute respiratory syndrome coro | ···················· | ···················· | ···················· | ···················· | ···················· | ···················· |
| 5638 | MW217347.1 | \|Severe acute respiratory syndrome coro | ···················· | ···················· | ···················· | ···················· | ···················· | ···················· |
| 5639 | MW217348.1 | \|Severe acute respiratory syndrome coro | ···················· | ···················· | ···················· | ···················· | ···················· | ···················· |
| 5640 | MW217349.1 | \|Severe acute respiratory syndrome coro | ···················· | ···················· | ···················· | ···················· | ···················· | ···················· |
| 5641 | MW217350.1 | \|Severe acute respiratory syndrome coro | ···················· | ···················· | ···················· | ···················· | ···················· | ···················· |
| 5642 | MW217351.1 | \|Severe acute respiratory syndrome coro | ···················· | ···················· | ···················· | ···················· | ···················· | ···················· |
| 5643 | MW217352.1 | \|Severe acute respiratory syndrome coro | ···················· | ···················· | ···················· | ···················· | ···················· | ···················· |
| 5644 | MW217353.1 | \|Severe acute respiratory syndrome coro | ·T·················· | ···················· | ···················· | ···················· | ···················· | ···················· |
| 5645 | MW217354.1 | \|Severe acute respiratory syndrome coro | ···················· | ···················· | ···················· | ···················· | ···················· | ···················· |
| 5646 | MW217355.1 | \|Severe acute respiratory syndrome coro | ···················· | ···················· | ···················· | ···················· | ···················· | ···················· |

5647 MW217356.1 |Severe acute respiratory syndrome coro ···················· ···················· ···················· ···················· ···················· ····················
5648 MW217357.1 |Severe acute respiratory syndrome coro ···················· ···················· ···················· ···················· ···················· ····················
5649 MW217358.1 |Severe acute respiratory syndrome coro ···················· ···················· ···················· ···················· ···················· ····················
5650 MW217359.1 |Severe acute respiratory syndrome coro ···················· ·A·················· ···················· ···················· ···················· ····················
5651 MW217360.1 |Severe acute respiratory syndrome coro ···················· ···A················ ···················· ···················· ···················· ····················
5652 MW217361.1 |Severe acute respiratory syndrome coro ···················· ·A·················· ···················· ···················· ···················· ····················
5653 MW217362.1 |Severe acute respiratory syndrome coro ···················· ·A·················· ···················· ···················· ···················· ····················
5654 MW217363.1 |Severe acute respiratory syndrome coro ···················· ···A················ ···················· ···················· ···················· ····················
5655 MW217364.1 |Severe acute respiratory syndrome coro ···················· ·A·················· ···················· ···················· ···················· ····················
5656 MW217365.1 |Severe acute respiratory syndrome coro ···················· ···················· ···················· ···················· ···················· ····················
5657 MW217366.1 |Severe acute respiratory syndrome coro ···················· ···················· ···················· ···················· ···················· ····················
5658 MW217367.1 |Severe acute respiratory syndrome coro ···················· ·A·················· ···················· ···················· ···················· ····················
5659 MW217368.1 |Severe acute respiratory syndrome coro ···················· ···················· ···················· ···················· ···················· ····················
5660 MW217369.1 |Severe acute respiratory syndrome coro ···················· ···················· ···················· ···················· ···················· ····················
5661 MW217370.1 |Severe acute respiratory syndrome coro ···················· ···················· ···················· ···················· ···················· ····················
5662 MW217371.1 |Severe acute respiratory syndrome coro ···················· ···················· ···················· ···················· ···················· ····················
5663 MW217372.1 |Severe acute respiratory syndrome coro ···················· ···················· ···················· ···················· ···················· ····················
5664 MW217373.1 |Severe acute respiratory syndrome coro ···················· ···················· ···················· ···················· ···················· ····················
5665 MW217375.1 |Severe acute respiratory syndrome coro ···················· ···················· ···················· ···················· ···················· ····················
5666 MW217376.1 |Severe acute respiratory syndrome coro ···················· ···················· ···················· ···················· ···················· ····················
5667 MW217377.1 |Severe acute respiratory syndrome coro ···················· ···················· ···················· ···················· ···················· ····················
5668 MW217378.1 |Severe acute respiratory syndrome coro ···················· ···················· ···················· ···················· ···················· ····················
5669 MW217379.1 |Severe acute respiratory syndrome coro ···················· ···················· ···················· ···················· ···················· ····················
5670 MW217380.1 |Severe acute respiratory syndrome coro ···················· ···················· ···················· ···················· ···················· ····················
5671 MW219695.1 |Severe acute respiratory syndrome coro ···················· ···················· ···················· ···················· ···················· ····················
5672 MW221270.1 |Severe acute respiratory syndrome coro ···················· ··C················· ···················· ···················· ···················· ····················
5673 MW221275.1 |Severe acute respiratory syndrome coro ···················· ···················· ···················· ···················· ···················· ····················
5674 MW205968.1 |Severe acute respiratory syndrome coro ···················· ···················· ···················· ···················· ···················· ····················
5675 MW205969.1 |Severe acute respiratory syndrome coro ···················· ···················· ···················· ···················· ···················· ····················
5676 MW205970.1 |Severe acute respiratory syndrome coro ···················· ···················· ···················· ···················· ···················· ····················
5677 MW205971.1 |Severe acute respiratory syndrome coro ···················· ···················· ···················· ···················· ···················· ·········A··········
5678 MW205972.1 |Severe acute respiratory syndrome coro ···················· ···················· ···················· ···················· ···················· ····················
5679 MW205973.1 |Severe acute respiratory syndrome coro ···················· ···················· ···················· ···················· ···················· ····················
5680 MW205974.1 |Severe acute respiratory syndrome coro ···················· ···················· ···················· ···················· ···················· ····················
5681 MW205975.1 |Severe acute respiratory syndrome coro ···················· ···················· ···················· ···················· ···················· ····················
5682 MW205976.1 |Severe acute respiratory syndrome coro ···················· ···················· ···················· ···················· ···················· ····················
5683 MW205977.1 |Severe acute respiratory syndrome coro ···················· ···················· ···················· ···················· ···················· ····················
5684 MW205978.1 |Severe acute respiratory syndrome coro ···················· ···················· ···················· ···················· ···················· ····················
5685 MW205979.1 |Severe acute respiratory syndrome coro ···················· ···················· ···················· ···················· ···················· ····················
5686 MW205980.1 |Severe acute respiratory syndrome coro ···················· ···················· ···················· ···················· ···················· ····················
5687 MW205981.1 |Severe acute respiratory syndrome coro ···················· ···················· ···················· ···················· ···················· ····················
5688 MW205982.1 |Severe acute respiratory syndrome coro ···················· ···················· ···················· ···················· ···················· ····················
5689 MW205983.1 |Severe acute respiratory syndrome coro ···················· ···················· ···················· ···················· ···················· ····················
5690 MW205984.1 |Severe acute respiratory syndrome coro ···················· ···················· ···················· ···················· ···················· ····················
5691 MW205985.1 |Severe acute respiratory syndrome coro ···················· ···················· ···················· ···················· ···················· ····················
5692 MW205986.1 |Severe acute respiratory syndrome coro ···················· ···················· ···················· ···················· ···················· ····················
5693 MW205987.1 |Severe acute respiratory syndrome coro ···················· ···················· ···················· ···················· ···················· ····················
5694 MW205988.1 |Severe acute respiratory syndrome coro ···················· ···················· ···················· ···················· ···················· ····················
5695 MW205989.1 |Severe acute respiratory syndrome coro ···················· ···················· ···················· ···················· ···················· ····················
5696 MW205990.1 |Severe acute respiratory syndrome coro ···················· ···················· ···················· ···················· ···················· ····················
5697 MW205991.1 |Severe acute respiratory syndrome coro ···················· ···················· ···················· ···················· ···················· ····················
5698 MW205992.1 |Severe acute respiratory syndrome coro ···················· ·A·················· ···················· ···················· ···················· ····················
5699 MW205993.1 |Severe acute respiratory syndrome coro ···················· ···················· ···················· ···················· ···················· ····················
5700 MW205994.1 |Severe acute respiratory syndrome coro ···················· ···················· ···················· ···················· ···················· ····················
5701 MW205995.1 |Severe acute respiratory syndrome coro ···················· ···A················ ···················· ···················· ···················· ····················
5702 MW205996.1 |Severe acute respiratory syndrome coro ···················· ···················· ···················· ···················· ···················· ····················
5703 MW205997.1 |Severe acute respiratory syndrome coro ···················· ···················· ···················· ···················· ···················· ····················
5704 MW206007.1 |Severe acute respiratory syndrome coro ···················· ···················· ···················· ···················· ···················· ····················
5705 MW206008.1 |Severe acute respiratory syndrome coro ···················· ·A·················· ···················· ···················· ···················· ····················
5706 MW206009.1 |Severe acute respiratory syndrome coro ···················· ···················· ···················· ···················· ···················· ····················
5707 MW206010.1 |Severe acute respiratory syndrome coro ···················· ·A·················· ···················· ···················· ···················· ····················
5708 MW206011.1 |Severe acute respiratory syndrome coro ···················· ···················· ···················· ···················· ···················· ····················
5709 MW206012.1 |Severe acute respiratory syndrome coro ···················· ···················· ···················· ···················· ···················· ····················
5710 MW206013.1 |Severe acute respiratory syndrome coro ···················· ···················· ···················· ···················· ···················· ····················
5711 MW206014.1 |Severe acute respiratory syndrome coro ···················· ···················· ···················· ···················· ···················· ····················
5712 MW206015.1 |Severe acute respiratory syndrome coro ···················· ···················· ···················· ···················· ···················· ····················
5713 MW206016.1 |Severe acute respiratory syndrome coro ···················· ···················· ···················· ···················· ···················· ····················
5714 MW206017.1 |Severe acute respiratory syndrome coro ···················· ···················· ···················· ···················· ···················· ····················
5715 MW206018.1 |Severe acute respiratory syndrome coro ···················· ···················· ···················· ···················· ···················· ····················
5716 MW206019.1 |Severe acute respiratory syndrome coro ···················· ···················· ···················· ···················· ···················· ····················
5717 MW206020.1 |Severe acute respiratory syndrome coro ···················· ···················· ···················· ···················· ···················· ····················
5718 MW206021.1 |Severe acute respiratory syndrome coro ···················· ···················· ···················· ···················· ···················· ····················
5719 MW206022.1 |Severe acute respiratory syndrome coro ···················· ···················· ···················· ···················· ···················· ····················
5720 MW206023.1 |Severe acute respiratory syndrome coro ···················· ···················· ···················· ···················· ···················· ····················
5721 MW206024.1 |Severe acute respiratory syndrome coro ···················· ···················· ···················· ···················· ···················· ····················
5722 MW206025.1 |Severe acute respiratory syndrome coro ···················· ···················· ···················· ···················· ···················· ····················
5723 MW206026.1 |Severe acute respiratory syndrome coro ···················· ···················· ···················· ···················· ···················· ····················
5724 MW206027.1 |Severe acute respiratory syndrome coro ···················· ·A·················· ···················· ···················· ···················· ····················

```
5725 MW206028.1 |Severe acute respiratory syndrome coro ···················· ···················· ···················· ···················· ···················· ····················
5726 MW206029.1 |Severe acute respiratory syndrome coro ···················· ···················· ···················· ···················· ···················· ····················
5727 MW206030.1 |Severe acute respiratory syndrome coro ···················· ···················· ···················· ···················· ···················· ····················
5728 MW206031.1 |Severe acute respiratory syndrome coro ···················· ···················· ···················· ···················· ···················· ····················
5729 MW206032.1 |Severe acute respiratory syndrome coro ···················· ···················· ···················· ···················· ···················· ····················
5730 MW206033.1 |Severe acute respiratory syndrome coro ···················· ···················· ···················· ···················· ···················· ····················
5731 MW206034.1 |Severe acute respiratory syndrome coro ···················· ···················· ···················· ···················· ···················· ····················
5732 MW206035.1 |Severe acute respiratory syndrome coro ···················· ···················· ···················· ···················· ···················· ····················
5733 MW206036.1 |Severe acute respiratory syndrome coro ···················· ···················· ···················· ···················· ···················· ····················
5734 MW206037.1 |Severe acute respiratory syndrome coro ···················· ···················· ···················· ···················· ···················· ····················
5735 MW206038.1 |Severe acute respiratory syndrome coro ···················· ···················· ···················· ···················· ···················· ····················
5736 MW206039.1 |Severe acute respiratory syndrome coro ···················· ···················· ···················· ···················· ···················· ····················
5737 MW206040.1 |Severe acute respiratory syndrome coro ···················· ···················· ···················· ···················· ···················· ····················
5738 MW206041.1 |Severe acute respiratory syndrome coro ···················· ···················· ···················· ···················· ···················· ····················
5739 MW206042.1 |Severe acute respiratory syndrome coro ···················· ···················· ···················· ···················· ···················· ····················
5740 MW206043.1 |Severe acute respiratory syndrome coro ···················· ···················· ···················· ···················· ···················· ····················
5741 MW206044.1 |Severe acute respiratory syndrome coro ···················· ···················· ···················· ···················· ···················· ····················
5742 MW206045.1 |Severe acute respiratory syndrome coro ···················· ···················· ···················· ···················· ···················· ····················
5743 MW206046.1 |Severe acute respiratory syndrome coro ···················· ···················· ···················· ···················· ···················· ····················
5744 MW206047.1 |Severe acute respiratory syndrome coro ···················· ···················· ···················· ···················· ···················· ····················
5745 MW206048.1 |Severe acute respiratory syndrome coro ···················· ···················· ···················· ···················· ···················· ····················
5746 MW206049.1 |Severe acute respiratory syndrome coro ···················· ···················· ···················· ···················· ···················· ····················
5747 MW206050.1 |Severe acute respiratory syndrome coro ···················· ···················· ···················· ···················· ···················· ····················
5748 MW206051.1 |Severe acute respiratory syndrome coro ···················· ···················· ···················· ···················· ···················· ····················
5749 MW206052.1 |Severe acute respiratory syndrome coro ···················· ···················· ···················· ···················· ···················· ····················
5750 MW206053.1 |Severe acute respiratory syndrome coro ···················· ···················· ···················· ···················· ···················· ····················
5751 MW206054.1 |Severe acute respiratory syndrome coro ···················· ···················· ···················· ···················· ···················· ····················
5752 MW206055.1 |Severe acute respiratory syndrome coro ···················· ···················· ···················· ···················· ···················· ····················
5753 MW206056.1 |Severe acute respiratory syndrome coro ···················· ···················· ···················· ···················· ········A··········· ····················
5754 MW206057.1 |Severe acute respiratory syndrome coro ···················· ···················· ···················· ···················· ···················· ····················
5755 MW206058.1 |Severe acute respiratory syndrome coro ···················· ···················· ···················· ···················· ···················· ····················
5756 MW206059.1 |Severe acute respiratory syndrome coro ···················· ···················· ···················· ···················· ···················· ····················
5757 MW206060.1 |Severe acute respiratory syndrome coro ···················· ···················· ···················· ···················· ···················· ····················
5758 MW206061.1 |Severe acute respiratory syndrome coro ···················· ···················· ···················· ···················· ···················· ····················
5759 MW206062.1 |Severe acute respiratory syndrome coro ···················· ···················· ···················· ···················· ···················· ····················
5760 MW206063.1 |Severe acute respiratory syndrome coro ···················· ···················· ···················· ···················· ···················· ····················
5761 MW206064.1 |Severe acute respiratory syndrome coro ···················· ···················· ···················· ···················· ···················· ····················
5762 MW206065.1 |Severe acute respiratory syndrome coro ···················· ···················· ···················· ···················· ········A··········· ····················
5763 MW206066.1 |Severe acute respiratory syndrome coro ···················· ···················· ···················· ···················· ···················· ····················
5764 MW206067.1 |Severe acute respiratory syndrome coro ···················· ···················· ···················· ···················· ···················· ····················
5765 MW206068.1 |Severe acute respiratory syndrome coro ···················· ···················· ···················· ···················· ···················· ····················
5766 MW206069.1 |Severe acute respiratory syndrome coro ···················· ···················· ···················· ···················· ···················· ····················
5767 MW206070.1 |Severe acute respiratory syndrome coro ···················· ···················· ···················· ···················· ···················· ····················
5768 MW206071.1 |Severe acute respiratory syndrome coro ···················· ···················· ···················· ···················· ···················· ····················
5769 MW206072.1 |Severe acute respiratory syndrome coro ···················· ···················· ···················· ···················· ···················· ····················
5770 MW206073.1 |Severe acute respiratory syndrome coro ···················· ···················· ···················· ···················· ···················· ····················
5771 MW206074.1 |Severe acute respiratory syndrome coro ···················· ···················· ···················· ···················· ···················· ····················
5772 MW206075.1 |Severe acute respiratory syndrome coro ···················· ···················· ···················· ···················· ···················· ····················
5773 MW206076.1 |Severe acute respiratory syndrome coro ···················· ···················· ···················· ···················· ···················· ····················
5774 MW206077.1 |Severe acute respiratory syndrome coro ···················· ···················· ···················· ···················· ···················· ····················
5775 MW206078.1 |Severe acute respiratory syndrome coro ···················· ···················· ···················· ···················· ···················· ····················
5776 MW206079.1 |Severe acute respiratory syndrome coro ···················· ·A·················· ···················· ···················· ···················· ····················
5777 MW206080.1 |Severe acute respiratory syndrome coro ···················· ···················· ···················· ···················· ···················· ····················
5778 MW206081.1 |Severe acute respiratory syndrome coro ···················· ···················· ···················· ···················· ···················· ····················
5779 MW206082.1 |Severe acute respiratory syndrome coro ···················· ···················· ···················· ···················· ···················· ····················
5780 MW206083.1 |Severe acute respiratory syndrome coro ···················· ·A·················· ···················· ···················· ···················· ····················
5781 MW206084.1 |Severe acute respiratory syndrome coro ···················· ···················· ···················· ···················· ···················· ····················
5782 MW206085.1 |Severe acute respiratory syndrome coro ···················· ···················· ···················· ···················· ···················· ····················
5783 MW206086.1 |Severe acute respiratory syndrome coro ···················· ···················· ···················· ···················· ···················· ····················
5784 MW206087.1 |Severe acute respiratory syndrome coro ···················· ···················· ···················· ···················· ···················· ····················
5785 MW206088.1 |Severe acute respiratory syndrome coro ···················· ···················· ···················· ···················· ···················· ····················
5786 MW206089.1 |Severe acute respiratory syndrome coro ···················· ···················· ···················· ···················· ···················· ····················
5787 MW206090.1 |Severe acute respiratory syndrome coro ···················· ···················· ···················· ···················· ···················· ····················
5788 MW206091.1 |Severe acute respiratory syndrome coro ···················· ···················· ···················· ···················· ···················· ····················
5789 MW206092.1 |Severe acute respiratory syndrome coro ···················· ···················· ···················· ···················· ···················· ····················
5790 MW206093.1 |Severe acute respiratory syndrome coro ···················· ···················· ···················· ···················· ···················· ····················
5791 MW206094.1 |Severe acute respiratory syndrome coro ···················· ···················· ···················· ···················· ··········G········· ····················
5792 MW206095.1 |Severe acute respiratory syndrome coro ···················· ···················· ···················· ···················· ···················· ····················
5793 MW206096.1 |Severe acute respiratory syndrome coro ···················· ···················· ···················· ···················· ···················· ····················
5794 MW206097.1 |Severe acute respiratory syndrome coro ···················· ···················· ···················· ···················· ···················· ····················
5795 MW206098.1 |Severe acute respiratory syndrome coro ···················· ···················· ···················· ···················· ···················· ····················
5796 MW206099.1 |Severe acute respiratory syndrome coro ···················· ···················· ···················· ···················· ···················· ····················
5797 MW206100.1 |Severe acute respiratory syndrome coro ···················· ···················· ···················· ···················· ···················· ····················
5798 MW206101.1 |Severe acute respiratory syndrome coro ···················· ···················· ···················· ···················· ···················· ····················
5799 MW206102.1 |Severe acute respiratory syndrome coro ···················· ···················· ···················· ···················· ···················· ····················
5800 MW206103.1 |Severe acute respiratory syndrome coro ···················· ···················· ···················· ···················· ···················· ····················
5801 MW206104.1 |Severe acute respiratory syndrome coro ···················· ···················· ···················· ···················· ···················· ····················
5802 MW206105.1 |Severe acute respiratory syndrome coro ···················· ···················· ···················· ···················· ···················· ····················
```

| | | | | | | | | |
|---|---|---|---|---|---|---|---|---|
| 5803 | MW206106.1 | \|Severe acute respiratory syndrome coro | ···················· | ···················· | ···················· | ···················· | ···················· | ···················· |
| 5804 | MW206107.1 | \|Severe acute respiratory syndrome coro | ···················· | ···················· | ···················· | ···················· | ···················· | ···················· |
| 5805 | MW206108.1 | \|Severe acute respiratory syndrome coro | ···················· | ···················· | ···················· | ···················· | ···················· | ···················· |
| 5806 | MW206109.1 | \|Severe acute respiratory syndrome coro | ···················· | ···················· | ···················· | ···················· | ···················· | ···················· |
| 5807 | MW206110.1 | \|Severe acute respiratory syndrome coro | ···················· | ···················· | ···················· | ···················· | ···················· | ···················· |
| 5808 | MW206111.1 | \|Severe acute respiratory syndrome coro | ···················· | ···················· | ···················· | ···················· | ···················· | ···················· |
| 5809 | MW206112.1 | \|Severe acute respiratory syndrome coro | ···················· | ···················· | ···················· | ···················· | ···················· | ···················· |
| 5810 | MW206113.1 | \|Severe acute respiratory syndrome coro | ···················· | ···················· | ···················· | ···················· | ···················· | ···················· |
| 5811 | MW206114.1 | \|Severe acute respiratory syndrome coro | ···················· | ···················· | ···················· | ···················· | ···················· | ···················· |
| 5812 | MW206115.1 | \|Severe acute respiratory syndrome coro | ···················· | ···················· | ···················· | ···················· | ···················· | ···················· |
| 5813 | MW206116.1 | \|Severe acute respiratory syndrome coro | ···················· | ···················· | ···················· | ···················· | ···················· | ···················· |
| 5814 | MW206117.1 | \|Severe acute respiratory syndrome coro | ···················· | ···················· | ···················· | ···················· | ···················· | ···················· |
| 5815 | MW206118.1 | \|Severe acute respiratory syndrome coro | ···················· | ···················· | ···················· | ···················· | ···················· | ···················· |
| 5816 | MW206119.1 | \|Severe acute respiratory syndrome coro | ···················· | ···················· | ···················· | ···················· | ···················· | ···················· |
| 5817 | MW206120.1 | \|Severe acute respiratory syndrome coro | ···················· | ···················· | ···················· | ···················· | ···················· | ···················· |
| 5818 | MW206121.1 | \|Severe acute respiratory syndrome coro | ···················· | ···················· | ···················· | ···················· | ···················· | ···················· |
| 5819 | MW206122.1 | \|Severe acute respiratory syndrome coro | ···················· | ···················· | ···················· | ···················· | ···················· | ···················· |
| 5820 | MW206123.1 | \|Severe acute respiratory syndrome coro | ···················· | ···················· | ···················· | ···················· | ···················· | ···················· |
| 5821 | MW206124.1 | \|Severe acute respiratory syndrome coro | ···················· | ···················· | ···················· | ···················· | ···················· | ···················· |
| 5822 | MW206125.1 | \|Severe acute respiratory syndrome coro | ···················· | ···················· | ···················· | ···················· | ···················· | ···················· |
| 5823 | MW206126.1 | \|Severe acute respiratory syndrome coro | ···················· | ···················· | ···················· | ···················· | ···················· | ···················· |
| 5824 | MW206127.1 | \|Severe acute respiratory syndrome coro | ···················· | ···················· | ···················· | ···················· | ···················· | ···················· |
| 5825 | MW206128.1 | \|Severe acute respiratory syndrome coro | ···················· | ···················· | ···················· | ···················· | ···················· | ···················· |
| 5826 | MW206129.1 | \|Severe acute respiratory syndrome coro | ···················· | ···················· | ···················· | ···················· | ···················· | ···················· |
| 5827 | MW206130.1 | \|Severe acute respiratory syndrome coro | ···················· | ···················· | ···················· | ···················· | ·········A·········· | ···················· |
| 5828 | MW206131.1 | \|Severe acute respiratory syndrome coro | ···················· | ···················· | ···················· | ···················· | ···················· | ···················· |
| 5829 | MW206132.1 | \|Severe acute respiratory syndrome coro | ···················· | ···················· | ···················· | ···················· | ···················· | ···················· |
| 5830 | MW206133.1 | \|Severe acute respiratory syndrome coro | ···················· | ···················· | ···················· | ···················· | ···················· | ···················· |
| 5831 | MW206134.1 | \|Severe acute respiratory syndrome coro | ···················· | ···················· | ···················· | ···················· | ···················· | ···················· |
| 5832 | MW206135.1 | \|Severe acute respiratory syndrome coro | ···················· | ···················· | ···················· | ···················· | ···················· | ···················· |
| 5833 | MW206136.1 | \|Severe acute respiratory syndrome coro | ···················· | ·A·················· | ···················· | ···················· | ···················· | ···················· |
| 5834 | MW206137.1 | \|Severe acute respiratory syndrome coro | ···················· | ···················· | ···················· | ···················· | ···················· | ···················· |
| 5835 | MW206138.1 | \|Severe acute respiratory syndrome coro | ···················· | ···················· | ···················· | ···················· | ···················· | ···················· |
| 5836 | MW206139.1 | \|Severe acute respiratory syndrome coro | ···················· | ···················· | ···················· | ···················· | ···················· | ···················· |
| 5837 | MW206140.1 | \|Severe acute respiratory syndrome coro | ···················· | ···················· | ···················· | ···················· | ···················· | ···················· |
| 5838 | MW206141.1 | \|Severe acute respiratory syndrome coro | ···················· | ···················· | ···················· | ···················· | ···················· | ···················· |
| 5839 | MW206142.1 | \|Severe acute respiratory syndrome coro | ···················· | ···················· | ···················· | ···················· | ···················· | ···················· |
| 5840 | MW206143.1 | \|Severe acute respiratory syndrome coro | ···················· | ···················· | ···················· | ···················· | ···················· | ···················· |
| 5841 | MW206144.1 | \|Severe acute respiratory syndrome coro | ···················· | ···················· | ···················· | ···················· | ···················· | ···················· |
| 5842 | MW206145.1 | \|Severe acute respiratory syndrome coro | ···················· | ···················· | ···················· | ···················· | ···················· | ···················· |
| 5843 | MW206146.1 | \|Severe acute respiratory syndrome coro | ···················· | ···················· | A··················· | ···················· | ···················· | ···················· |
| 5844 | MW206147.1 | \|Severe acute respiratory syndrome coro | ···················· | ···················· | ···················· | ···················· | ·········A·········· | ···················· |
| 5845 | MW206148.1 | \|Severe acute respiratory syndrome coro | ···················· | ···················· | ···················· | ···················· | ·········A·········· | ···················· |
| 5846 | MW206149.1 | \|Severe acute respiratory syndrome coro | ···················· | ···················· | ···················· | ···················· | ···················· | ···················· |
| 5847 | MW206150.1 | \|Severe acute respiratory syndrome coro | ···················· | ···················· | ···················· | ···················· | ···················· | ···················· |
| 5848 | MW206151.1 | \|Severe acute respiratory syndrome coro | ···················· | ···················· | ···················· | ···················· | ···················· | ···················· |
| 5849 | MW206152.1 | \|Severe acute respiratory syndrome coro | ···················· | ···················· | ···················· | ···················· | ···················· | ···················· |
| 5850 | MW206153.1 | \|Severe acute respiratory syndrome coro | ···················· | ···················· | ···················· | ···················· | ···················· | ···················· |
| 5851 | MW206154.1 | \|Severe acute respiratory syndrome coro | ···················· | ···················· | ···················· | ···················· | ···················· | ···················· |
| 5852 | MW206155.1 | \|Severe acute respiratory syndrome coro | ···················· | ···················· | ···················· | ···················· | ···················· | ···················· |
| 5853 | MW206156.1 | \|Severe acute respiratory syndrome coro | ···················· | ···················· | ···················· | ···················· | ···················· | ···················· |
| 5854 | MW206157.1 | \|Severe acute respiratory syndrome coro | ···················· | ···················· | ···················· | ···················· | ···················· | ···················· |
| 5855 | MW206158.1 | \|Severe acute respiratory syndrome coro | ···················· | ···················· | ···················· | ···················· | ···················· | ···················· |
| 5856 | MW206159.1 | \|Severe acute respiratory syndrome coro | ···················· | ···················· | ···················· | ···················· | ···················· | ···················· |
| 5857 | MW206160.1 | \|Severe acute respiratory syndrome coro | ···················· | ···················· | ···················· | ···················· | ···················· | ···················· |
| 5858 | MW206161.1 | \|Severe acute respiratory syndrome coro | ···················· | ···················· | ···················· | ···················· | ···················· | ···················· |
| 5859 | MW206162.1 | \|Severe acute respiratory syndrome coro | ···················· | ···················· | ···················· | ···················· | ···················· | ···················· |
| 5860 | MW206163.1 | \|Severe acute respiratory syndrome coro | ···················· | ·A·················· | ···················· | ···················· | ···················· | ···················· |
| 5861 | MW206164.1 | \|Severe acute respiratory syndrome coro | ···················· | ·A·················· | ···················· | ···················· | ···················· | ···················· |
| 5862 | MW206165.1 | \|Severe acute respiratory syndrome coro | ···················· | ···················· | ···················· | ···················· | ···················· | ···················· |
| 5863 | MW206166.1 | \|Severe acute respiratory syndrome coro | ···················· | ·A·················· | ···················· | ···················· | ···················· | ···················· |
| 5864 | MW206167.1 | \|Severe acute respiratory syndrome coro | ···················· | ···················· | ···················· | ···················· | ···················· | ···················· |
| 5865 | MW206168.1 | \|Severe acute respiratory syndrome coro | ···················· | ···················· | ···················· | ···················· | ···················· | ···················· |
| 5866 | MW206169.1 | \|Severe acute respiratory syndrome coro | ···················· | ···················· | ···················· | ···················· | ·········A·········· | ···················· |
| 5867 | MW206170.1 | \|Severe acute respiratory syndrome coro | ···················· | ···················· | ···················· | ···················· | ···················· | ···················· |
| 5868 | MW206171.1 | \|Severe acute respiratory syndrome coro | ···················· | ···················· | ···················· | ···················· | ···················· | ···················· |
| 5869 | MW206172.1 | \|Severe acute respiratory syndrome coro | ···················· | ···················· | ···················· | ···················· | ···················· | ···················· |
| 5870 | MW206173.1 | \|Severe acute respiratory syndrome coro | ···················· | ···················· | ···················· | ···················· | ···················· | ···················· |
| 5871 | MW206174.1 | \|Severe acute respiratory syndrome coro | ···················· | ···················· | ···················· | ···················· | ···················· | ···················· |
| 5872 | MW206175.1 | \|Severe acute respiratory syndrome coro | ···················· | ···················· | ···················· | ···················· | ···················· | ···················· |
| 5873 | MW206176.1 | \|Severe acute respiratory syndrome coro | ···················· | ···················· | ···················· | ···················· | ···················· | ···················· |
| 5874 | MW206177.1 | \|Severe acute respiratory syndrome coro | ···················· | ···················· | ···················· | ···················· | ···················· | ···················· |
| 5875 | MW206178.1 | \|Severe acute respiratory syndrome coro | ···················· | ···················· | ···················· | ···················· | ···················· | ···················· |
| 5876 | MW206179.1 | \|Severe acute respiratory syndrome coro | ···················· | ···················· | ···················· | ···················· | ···················· | ···················· |
| 5877 | MW206180.1 | \|Severe acute respiratory syndrome coro | ···················· | ···················· | ···················· | ···················· | ···················· | ···················· |
| 5878 | MW206181.1 | \|Severe acute respiratory syndrome coro | ···················· | ···················· | ···················· | ···················· | ···················· | ···················· |
| 5879 | MW206182.1 | \|Severe acute respiratory syndrome coro | ···················· | ···················· | ···················· | ···················· | ···················· | ···················· |
| 5880 | MW206183.1 | \|Severe acute respiratory syndrome coro | ···················· | ···················· | ···················· | ···················· | ···················· | ···················· |

```
5881 MW206184.1 |Severe acute respiratory syndrome coro   ....................  ....................  ....................  ....................  ....................  ....................
5882 MW206185.1 |Severe acute respiratory syndrome coro   ....................  ....................  ....................  ....................  .........A..........  ....................
5883 MW206186.1 |Severe acute respiratory syndrome coro   ....................  ....................  ....................  ....................  ....................  ....................
5884 MW206187.1 |Severe acute respiratory syndrome coro   ....................  ....................  ....................  ....................  ....................  ....................
5885 MW206188.1 |Severe acute respiratory syndrome coro   ....................  ....................  ....................  ....................  ....................  ....................
5886 MW206189.1 |Severe acute respiratory syndrome coro   ....................  ....................  ....................  ....................  ....................  ....................
5887 MW206190.1 |Severe acute respiratory syndrome coro   ....................  ....................  ....................  ....................  ....................  ....................
5888 MW206191.1 |Severe acute respiratory syndrome coro   ....................  ....................  ....................  ....................  ...C................  ....................
5889 MW206192.1 |Severe acute respiratory syndrome coro   ....................  ....................  ....................  ....................  ....................  ....................
5890 MW206193.1 |Severe acute respiratory syndrome coro   ....................  ....................  ....................  ....................  ....................  ....................
5891 MW206194.1 |Severe acute respiratory syndrome coro   ....................  ....................  ....................  ....................  ....................  ....................
5892 MW206195.1 |Severe acute respiratory syndrome coro   ....................  ....................  ....................  ....................  ....................  ....................
5893 MW206196.1 |Severe acute respiratory syndrome coro   ....................  ....................  ....................  ....................  ....................  ....................
5894 MW206197.1 |Severe acute respiratory syndrome coro   ....................  ....................  ....................  ....................  ....................  ....................
5895 MW206198.1 |Severe acute respiratory syndrome coro   ....................  ....................  ....................  ....................  ....................  ....................
5896 MW206199.1 |Severe acute respiratory syndrome coro   ....................  ....................  ....................  ....................  ....................  ....................
5897 MW206200.1 |Severe acute respiratory syndrome coro   ....................  ....................  ....................  ....................  ....................  ....................
5898 MW206201.1 |Severe acute respiratory syndrome coro   ....................  ....................  ....................  ....................  ....................  ....................
5899 MW206202.1 |Severe acute respiratory syndrome coro   ....................  ....................  ....................  ....................  ....................  ....................
5900 MW206203.1 |Severe acute respiratory syndrome coro   ....................  ....................  ....................  ....................  .........A..........  ....................
5901 MW206204.1 |Severe acute respiratory syndrome coro   ....................  ....................  ....................  ....................  ....................  ....................
5902 MW206205.1 |Severe acute respiratory syndrome coro   ....................  ....................  ....................  ....................  ....................  ....................
5903 MW206206.1 |Severe acute respiratory syndrome coro   ....................  ....................  ....................  ....................  ....................  ....................
5904 MW206207.1 |Severe acute respiratory syndrome coro   ....................  ....................  ....................  ....................  ....................  ....................
5905 MW206208.1 |Severe acute respiratory syndrome coro   ....................  ....................  ....................  ....................  ....................  ....................
5906 MW206209.1 |Severe acute respiratory syndrome coro   ....................  ....................  ....................  ....................  ....................  ....................
5907 MW206210.1 |Severe acute respiratory syndrome coro   ....................  ....................  ....................  ....................  ....................  ....................
5908 MW206211.1 |Severe acute respiratory syndrome coro   ....................  ....................  ....................  ....................  ....................  ....................
5909 MW206212.1 |Severe acute respiratory syndrome coro   ....................  ....................  ....................  ....................  ....................  ....................
5910 MW206213.1 |Severe acute respiratory syndrome coro   ....................  ....................  ....................  ....................  ....................  ....................
5911 MW206214.1 |Severe acute respiratory syndrome coro   ....................  ....................  ....................  ....................  ....................  ....................
5912 MW206215.1 |Severe acute respiratory syndrome coro   ....................  ....................  ....................  ....................  ....................  ....................
5913 MW206216.1 |Severe acute respiratory syndrome coro   ....................  ....................  ....................  ....................  ....................  ....................
5914 MW206217.1 |Severe acute respiratory syndrome coro   ....................  ....................  ....................  ....................  ....................  ....................
5915 MW206218.1 |Severe acute respiratory syndrome coro   ....................  ....................  ....................  ....................  ....................  ....................
5916 MW206219.1 |Severe acute respiratory syndrome coro   ....................  ....................  ....................  ....................  ....................  ....................
5917 MW206220.1 |Severe acute respiratory syndrome coro   ....................  ....................  ....................  ....................  ....................  ....................
5918 MW206221.1 |Severe acute respiratory syndrome coro   ....................  ....................  ....................  ....................  ....................  ....................
5919 MW206222.1 |Severe acute respiratory syndrome coro   ....................  ....................  ....................  ....................  ....................  ....................
5920 MW206223.1 |Severe acute respiratory syndrome coro   ....................  ....................  ....................  ....................  ....................  ....................
5921 MW206224.1 |Severe acute respiratory syndrome coro   ....................  ....................  ....................  ....................  ....................  ....................
5922 MW206225.1 |Severe acute respiratory syndrome coro   ....................  ....................  ....................  ....................  ....................  ....................
5923 MW206226.1 |Severe acute respiratory syndrome coro   ....................  ....................  ....................  ....................  ....................  ....................
5924 MW206227.1 |Severe acute respiratory syndrome coro   ....................  ....................  ....................  ....................  ....................  ....................
5925 MW206228.1 |Severe acute respiratory syndrome coro   ....................  ....................  ....................  ....................  ....................  ....................
5926 MW206229.1 |Severe acute respiratory syndrome coro   ....................  ....................  ....................  ....................  ....................  ....................
5927 MW206230.1 |Severe acute respiratory syndrome coro   ....................  .A..................  ....................  ....................  ....................  ....................
5928 MW206231.1 |Severe acute respiratory syndrome coro   ....................  ....................  ....................  ....................  ....................  ....................
5929 MW206232.1 |Severe acute respiratory syndrome coro   ....................  ....................  ....................  ....................  ....................  ....................
5930 MW206233.1 |Severe acute respiratory syndrome coro   ....................  ....................  ....................  ....................  ....................  ....................
5931 MW206234.1 |Severe acute respiratory syndrome coro   ....................  ....................  ....................  ....................  ....................  ....................
5932 MW206235.1 |Severe acute respiratory syndrome coro   ....................  ....................  ....................  ....................  .........A..........  ....................
5933 MW206236.1 |Severe acute respiratory syndrome coro   ....................  ....................  ....................  ....................  ....................  ....................
5934 MW206237.1 |Severe acute respiratory syndrome coro   ....................  ....................  ....................  ....................  ....................  ....................
5935 MW206238.1 |Severe acute respiratory syndrome coro   ....................  ....................  ....................  ....................  .........A..........  ....................
5936 MW206239.1 |Severe acute respiratory syndrome coro   ....................  ....................  ....................  ....................  ....................  ....................
5937 MW206240.1 |Severe acute respiratory syndrome coro   ....................  ....................  ....................  ....................  ....................  ....................
5938 MW206241.1 |Severe acute respiratory syndrome coro   ....................  ....................  ....................  ....................  ....................  ....................
5939 MW206242.1 |Severe acute respiratory syndrome coro   ....................  ....................  ....................  ....................  ....................  ....................
5940 MW206243.1 |Severe acute respiratory syndrome coro   ....................  ....................  ....................  ....................  ....................  ....................
5941 MW206244.1 |Severe acute respiratory syndrome coro   ....................  ....................  ....................  ....................  ....................  ....................
5942 MW206245.1 |Severe acute respiratory syndrome coro   ....................  ....................  ....................  ....................  ....................  ....................
5943 MW206246.1 |Severe acute respiratory syndrome coro   ....................  ....................  ....................  ....................  ....................  ....................
5944 MW206247.1 |Severe acute respiratory syndrome coro   ....................  ....................  ....................  ....................  ....................  ....................
5945 MW206248.1 |Severe acute respiratory syndrome coro   ....................  ....................  ....................  ....................  ....................  ....................
5946 MW206249.1 |Severe acute respiratory syndrome coro   ....................  ....................  ....................  ....................  ....................  ....................
5947 MW206250.1 |Severe acute respiratory syndrome coro   ....................  ....................  ....................  ....................  ....................  ....................
5948 MW206251.1 |Severe acute respiratory syndrome coro   ....................  ....................  ....................  ....................  ....................  ....................
5949 MW206252.1 |Severe acute respiratory syndrome coro   ....................  ....................  ....................  ....................  ....................  ....................
5950 MW206253.1 |Severe acute respiratory syndrome coro   ....................  ....................  ....................  ....................  ....................  ....................
5951 MW206254.1 |Severe acute respiratory syndrome coro   ....................  ....................  ....................  ....................  ....................  ....................
5952 MW206255.1 |Severe acute respiratory syndrome coro   ....................  ....................  ....................  ....................  ....................  ....................
5953 MW206256.1 |Severe acute respiratory syndrome coro   ....................  ....................  ....................  ....................  ....................  ....................
5954 MW206257.1 |Severe acute respiratory syndrome coro   ....................  ....................  ....................  ....................  ....................  ....................
5955 MW206258.1 |Severe acute respiratory syndrome coro   ....................  ....................  ....................  ....................  ....................  ....................
5956 MW206259.1 |Severe acute respiratory syndrome coro   ....................  ....................  ....................  ....................  .........A..........  ....................
5957 MW206260.1 |Severe acute respiratory syndrome coro   ....................  ....................  ....................  ....................  ....................  ....................
5958 MW206261.1 |Severe acute respiratory syndrome coro   ....................  ....................  ....................  ....................  ....................  ....................
```

5959 MW206262.1 |Severe acute respiratory syndrome coro ···················· ···················· ···················· ···················· ···················· ····················
5960 MW206263.1 |Severe acute respiratory syndrome coro ···················· ···················· ···················· ···················· ···················· ····················
5961 MW206264.1 |Severe acute respiratory syndrome coro ···················· ···················· ···················· ···················· ···················· ····················
5962 MW206265.1 |Severe acute respiratory syndrome coro ···················· ···················· ···················· ···················· ···················· ····················
5963 MW206266.1 |Severe acute respiratory syndrome coro ···················· ···················· ···················· ···················· ···················· ····················
5964 MW206267.1 |Severe acute respiratory syndrome coro ···················· ···················· ···················· ···················· ···················· ····················
5965 MW206268.1 |Severe acute respiratory syndrome coro ···················· ···················· ···················· ···················· ···················· ····················
5966 MW206269.1 |Severe acute respiratory syndrome coro ···················· ···················· ···················· ···················· ···················· ····················
5967 MW206270.1 |Severe acute respiratory syndrome coro ···················· ···················· ···················· ···················· ···················· ····················
5968 MW206271.1 |Severe acute respiratory syndrome coro ···················· ···················· ···················· ···················· ···················· ····················
5969 MW206272.1 |Severe acute respiratory syndrome coro ···················· ···················· ···················· ···················· ···················· ····················
5970 MW206273.1 |Severe acute respiratory syndrome coro ···················· ···················· ···················· ···················· ···················· ····················
5971 MW206274.1 |Severe acute respiratory syndrome coro ···················· ···················· ···················· ···················· ···················· ····················
5972 MW206275.1 |Severe acute respiratory syndrome coro ···················· ···················· ···················· ···················· ···················· ····················
5973 MW206279.1 |Severe acute respiratory syndrome coro ···················· ···················· ···················· ···················· ···················· ····················
5974 MW206281.1 |Severe acute respiratory syndrome coro ···················· ···················· ···················· ···················· ···················· ····················
5975 MW206282.1 |Severe acute respiratory syndrome coro ···················· ···················· ···················· ···················· ···················· ····················
5976 MW206283.1 |Severe acute respiratory syndrome coro ···················· ···················· ···················· ···················· ···················· ····················
5977 MW206284.1 |Severe acute respiratory syndrome coro ···················· ···················· ···················· ···················· ···················· ····················
5978 MW206285.1 |Severe acute respiratory syndrome coro ···················· ···················· ···················· ···················· ···················· ····················
5979 MW206286.1 |Severe acute respiratory syndrome coro ···················· ···················· ···················· ···················· ···················· ····················
5980 MW206288.1 |Severe acute respiratory syndrome coro ···················· ·A·················· ···················· ···················· ···················· ····················
5981 MW206289.1 |Severe acute respiratory syndrome coro ···················· ···················· ···················· ···················· ···················· ····················
5982 MW206290.1 |Severe acute respiratory syndrome coro ···················· ·A·················· ···················· ···················· ···················· ····················
5983 MW206291.1 |Severe acute respiratory syndrome coro ···················· ···················· ···················· ···················· ···················· ····················
5984 MW206292.1 |Severe acute respiratory syndrome coro ···················· ···················· ···················· ···················· ···················· ····················
5985 MW206293.1 |Severe acute respiratory syndrome coro ···················· ···················· ···················· ···················· ···················· ····················
5986 MW206294.1 |Severe acute respiratory syndrome coro ···················· ···················· ···················· ···················· ···················· ····················
5987 MW206295.1 |Severe acute respiratory syndrome coro ···················· ···················· ···················· ···················· ···················· ····················
5988 MW206296.1 |Severe acute respiratory syndrome coro ···················· ···················· ···················· ···················· ···················· ····················
5989 MW206297.1 |Severe acute respiratory syndrome coro ···················· ···················· ···················· ···················· ···················· ····················
5990 MW206298.1 |Severe acute respiratory syndrome coro ···················· ···················· ···················· ···················· ···················· ····················
5991 MW206299.1 |Severe acute respiratory syndrome coro ···················· ···················· ···················· ···················· ···················· ····················
5992 MW206300.1 |Severe acute respiratory syndrome coro ···················· ···················· ···················· ···················· ···················· ····················
5993 MW206301.1 |Severe acute respiratory syndrome coro ···················· ···················· ···················· ···················· ···················· ····················
5994 MW206302.1 |Severe acute respiratory syndrome coro ···················· ···················· ···················· ···················· ···················· ····················
5995 MW206303.1 |Severe acute respiratory syndrome coro ···················· ···················· ···················· ···················· ···················· ····················
5996 MW206304.1 |Severe acute respiratory syndrome coro ···················· ···················· ···················· ···················· ···················· ····················
5997 MW206305.1 |Severe acute respiratory syndrome coro ···················· ···················· ···················· ···················· ···················· ····················
5998 MW206306.1 |Severe acute respiratory syndrome coro ···················· ···················· ···················· ···················· ···················· ····················
5999 MW206307.1 |Severe acute respiratory syndrome coro ···················· ···················· ···················· ···················· ···················· ····················
6000 MW206308.1 |Severe acute respiratory syndrome coro ···················· ·A·················· ···················· ···················· ···················· ····················
6001 MW206309.1 |Severe acute respiratory syndrome coro ···················· ·A·················· ···················· ···················· ···················· ····················
6002 MW206310.1 |Severe acute respiratory syndrome coro ···················· ···················· ···················· ···················· ···················· ····················
6003 MW206311.1 |Severe acute respiratory syndrome coro ···················· ···················· ···················· ···················· ···················· ····················
6004 MW206312.1 |Severe acute respiratory syndrome coro ···················· ·A·················· ···················· ···················· ···················· ····················
6005 MW206313.1 |Severe acute respiratory syndrome coro ···················· ···················· ···················· ···················· ···················· ····················
6006 MW206314.1 |Severe acute respiratory syndrome coro ···················· ···················· ···················· ···················· ···················· ····················
6007 MW206315.1 |Severe acute respiratory syndrome coro ···················· ···················· ···················· ···················· ···················· ····················
6008 MW206316.1 |Severe acute respiratory syndrome coro ···················· ···················· ···················· ···················· ···················· ····················
6009 MW206317.1 |Severe acute respiratory syndrome coro ···················· ·A·················· ···················· ···················· ···················· ····················
6010 MW206318.1 |Severe acute respiratory syndrome coro ···················· ···················· ···················· ···················· ···················· ····················
6011 MW206319.1 |Severe acute respiratory syndrome coro ···················· ···················· ···················· ···················· ···················· ····················
6012 MW206320.1 |Severe acute respiratory syndrome coro ···················· ···················· ···················· ···················· ···················· ····················
6013 MW206321.1 |Severe acute respiratory syndrome coro ···················· ···················· ···················· ···················· ···················· ····················
6014 MW206322.1 |Severe acute respiratory syndrome coro ···················· ·A·················· ···················· ···················· ···················· ····················
6015 MW206323.1 |Severe acute respiratory syndrome coro ···················· ···················· ···················· ···················· ···················· ····················
6016 MW206324.1 |Severe acute respiratory syndrome coro ···················· ···················· ···················· ···················· ···················· ····················
6017 MW206325.1 |Severe acute respiratory syndrome coro ···················· ···················· ···················· ···················· ···················· ····················
6018 MW206326.1 |Severe acute respiratory syndrome coro ···················· ···················· ···················· ···················· ·········A·········· ····················
6019 MW206327.1 |Severe acute respiratory syndrome coro ···················· ···················· ···················· ···················· ···················· ····················
6020 MW206328.1 |Severe acute respiratory syndrome coro ···················· ···················· ···················· ···················· ···················· ····················
6021 MW206329.1 |Severe acute respiratory syndrome coro ···················· ···················· ···················· ···················· ···················· ····················
6022 MW206330.1 |Severe acute respiratory syndrome coro ···················· ···················· ···················· ···················· ···················· ····················
6023 MW206331.1 |Severe acute respiratory syndrome coro ···················· ···················· ···················· ···················· ···················· ····················
6024 MW206332.1 |Severe acute respiratory syndrome coro ···················· ···················· ···················· ···················· ···················· ····················
6025 MW206333.1 |Severe acute respiratory syndrome coro ···················· ···················· ···················· ···················· ···················· ····················
6026 MW206334.1 |Severe acute respiratory syndrome coro ···················· ···················· ···················· ···················· ···················· ····················
6027 MW206335.1 |Severe acute respiratory syndrome coro ···················· ···················· ···················· ···················· ···················· ····················
6028 MW206336.1 |Severe acute respiratory syndrome coro ···················· ···················· ···················· ···················· ···················· ····················
6029 MW206337.1 |Severe acute respiratory syndrome coro ···················· ···················· ···················· ···················· ···················· ····················
6030 MW206338.1 |Severe acute respiratory syndrome coro ···················· ···················· ···················· ···················· ···················· ····················
6031 MW206339.1 |Severe acute respiratory syndrome coro ···················· ·A·················· ···················· ···················· ···················· ····················
6032 MW206341.1 |Severe acute respiratory syndrome coro ···················· ···················· ···················· ···················· ···················· ····················
6033 MW206342.1 |Severe acute respiratory syndrome coro ···················· ···················· ···················· ···················· ···················· ····················
6034 MW206343.1 |Severe acute respiratory syndrome coro ···················· ···················· ···················· ···················· ···················· ····················
6035 MW206344.1 |Severe acute respiratory syndrome coro ···················· ···················· ···················· ···················· ···················· ····················
6036 MW206345.1 |Severe acute respiratory syndrome coro ···················· ···················· ···················· ···················· ···················· ····················

6037 MW206346.1 |Severe acute respiratory syndrome coro ···················· ·A·················· ···················· ···················· ···················· ····················
6038 MW206347.1 |Severe acute respiratory syndrome coro ···················· ···················· ···················· ···················· ·········T·········· ····················
6039 MW206348.1 |Severe acute respiratory syndrome coro ···················· ···················· ···················· ···················· ···················· ····················
6040 MW206349.1 |Severe acute respiratory syndrome coro ···················· ·A·················· ···················· ···················· ···················· ····················
6041 MW206350.1 |Severe acute respiratory syndrome coro ···················· ···················· ···················· ···················· ···················· ····················
6042 MW206351.1 |Severe acute respiratory syndrome coro ···················· ···················· ···················· ···················· ···················· ····················
6043 MW206352.1 |Severe acute respiratory syndrome coro ···················· ···················· ···················· ···················· ···················· ····················
6044 MW206353.1 |Severe acute respiratory syndrome coro ···················· ···················· ···················· ···················· ···················· ····················
6045 MW206354.1 |Severe acute respiratory syndrome coro ···················· ··G················· ···················· ···················· ···················· ····················
6046 MW206355.1 |Severe acute respiratory syndrome coro ···················· ···················· ···················· ···················· ···················· ····················
6047 MW206356.1 |Severe acute respiratory syndrome coro ···················· ···················· ···················· ···················· ···················· ····················
6048 MW206357.1 |Severe acute respiratory syndrome coro ···················· ···················· ···················· ···················· ···················· ····················
6049 MW206358.1 |Severe acute respiratory syndrome coro ···················· ···················· ···················· ···················· ···················· ····················
6050 MW206359.1 |Severe acute respiratory syndrome coro ···················· ···················· ···················· ···················· ···················· ····················
6051 MW206360.1 |Severe acute respiratory syndrome coro ···················· ···················· ···················· ···················· ···················· ····················
6052 MW206361.1 |Severe acute respiratory syndrome coro ···················· ···················· ···················· ···················· ···················· ····················
6053 MW206362.1 |Severe acute respiratory syndrome coro ···················· ···················· ···················· ···················· ···················· ····················
6054 MW206363.1 |Severe acute respiratory syndrome coro ···················· ···················· ···················· ···················· ···················· ····················
6055 MW206398.1 |Severe acute respiratory syndrome coro ···················· ·A·················· ···················· ···················· ···················· ····················
6058 MW206409.1 |Severe acute respiratory syndrome coro ···················· ·············A······ ···················· ···················· ···················· ····················
6059 MW206414.1 |Severe acute respiratory syndrome coro ···················· ···················· ···················· ···················· ···················· ····················
6060 MW206415.1 |Severe acute respiratory syndrome coro ···················· ···················· ···················· ···················· ···················· ····················
6061 MW206417.1 |Severe acute respiratory syndrome coro ···················· ···················· ···················· ···················· ···················· ····················
6062 MW206418.1 |Severe acute respiratory syndrome coro ···················· ···················· ···················· ···················· ···················· ····················
6063 MW206428.1 |Severe acute respiratory syndrome coro ···················· ···················· ···················· ···················· ···················· ····················
6064 MW206431.1 |Severe acute respiratory syndrome coro ···················· ···················· ···················· ···················· ···················· ····················
6065 MW206433.1 |Severe acute respiratory syndrome coro ···················· ···················· ···················· ···················· ···················· ····················
6066 MW206434.1 |Severe acute respiratory syndrome coro ···················· ···················· ···················· ···················· ···················· ····················
6067 MW206440.1 |Severe acute respiratory syndrome coro ···················· ·A·················· ···················· ···················· ···················· ····················
6068 MW206448.1 |Severe acute respiratory syndrome coro ···················· ·············A······ ···················· ···················· ···················· ····················
6069 MW206449.1 |Severe acute respiratory syndrome coro ···················· ···················· ···················· ···················· ···················· ····················
6070 MW206450.1 |Severe acute respiratory syndrome coro ···················· ·A·················· ···················· ···················· ···················· ····················
6072 MW206455.1 |Severe acute respiratory syndrome coro ···················· ···················· ···················· ···················· ···················· ····················
6075 MW206459.1 |Severe acute respiratory syndrome coro ···················· ···················· ···················· ···················· ···················· ····················
6076 MW206460.1 |Severe acute respiratory syndrome coro ···················· ···················· ···················· ···················· ···················· ····················
6077 MW206479.1 |Severe acute respiratory syndrome coro ···················· ···················· ···················· ···················· ···················· ····················
6078 MW206480.1 |Severe acute respiratory syndrome coro ···················· ···················· ···················· ···················· ···················· ····················
6079 MW206481.1 |Severe acute respiratory syndrome coro ···················· ···················· ···················· ···················· ···················· ····················
6080 MW206483.1 |Severe acute respiratory syndrome coro ···················· ···················· ···················· ···················· ···················· ····················
6081 MW206485.1 |Severe acute respiratory syndrome coro ···················· ···················· ···················· ···················· ···················· ····················
6082 MW206486.1 |Severe acute respiratory syndrome coro ···················· ···················· ···················· ···················· ···················· ····················
6084 MW206490.1 |Severe acute respiratory syndrome coro ···················· ···················· ···················· ···················· ···················· ····················
6085 MW206492.1 |Severe acute respiratory syndrome coro ···················· ···················· ···················· ···················· ···················· ····················
6086 MW206493.1 |Severe acute respiratory syndrome coro ···················· ···················· ···················· ···················· ···················· ····················
6087 MW206496.1 |Severe acute respiratory syndrome coro ···················· ···················· ···················· ···················· ···················· ····················
6088 MW206501.1 |Severe acute respiratory syndrome coro ···················· ···················· ···················· ···················· ···················· ····················
6089 MW206502.1 |Severe acute respiratory syndrome coro ···················· ···················· ···················· ···················· ···················· ····················
6090 MW206506.1 |Severe acute respiratory syndrome coro ···················· ···················· ···················· ···················· ···················· ····················
6091 MW206508.1 |Severe acute respiratory syndrome coro ···················· ···················· ···················· ···················· ···················· ····················
6092 MW206509.1 |Severe acute respiratory syndrome coro ···················· ···················· ···················· ···················· ···················· ····················
6093 MW206510.1 |Severe acute respiratory syndrome coro ···················· ···················· ···················· ···················· ···················· ····················
6094 MW206511.1 |Severe acute respiratory syndrome coro ···················· ···················· ···················· ···················· ···················· ····················
6095 MW206512.1 |Severe acute respiratory syndrome coro ···················· ···················· ···················· ···················· ···················· ····················
6096 MW206513.1 |Severe acute respiratory syndrome coro ···················· ···················· ···················· ···················· ···················· ····················
6097 MW206514.1 |Severe acute respiratory syndrome coro ···················· ···················· ···················· ···················· ···················· ····················
6099 MW206516.1 |Severe acute respiratory syndrome coro ···················· ···················· ···················· ···················· ···················· ····················
6100 MW206517.1 |Severe acute respiratory syndrome coro ···················· ···················· ···················· ···················· ···················· ····················
6101 MW206518.1 |Severe acute respiratory syndrome coro ···················· ···················· ···················· ···················· ···················· ····················
6102 MW206519.1 |Severe acute respiratory syndrome coro ···················· ···················· ···················· ···················· ···················· ····················
6103 MW206521.1 |Severe acute respiratory syndrome coro ···················· ···················· ···················· ···················· ···················· ····················
6104 MW206522.1 |Severe acute respiratory syndrome coro ···················· ···················· ···················· ···················· ···················· ····················
6105 MW206523.1 |Severe acute respiratory syndrome coro ···················· ···················· ···················· ···················· ···················· ····················
6106 MW206524.1 |Severe acute respiratory syndrome coro ···················· ···················· ···················· ···················· ···················· ····················
6107 MW206525.1 |Severe acute respiratory syndrome coro ···················· ···················· ···················· ···················· ···················· ····················
6108 MW206528.1 |Severe acute respiratory syndrome coro ···················· ···················· ···················· ···················· ···················· ····················
6109 MW206529.1 |Severe acute respiratory syndrome coro ···················· ···················· ···················· ···················· ···················· ····················
6110 MW206530.1 |Severe acute respiratory syndrome coro ···················· ···················· ···················· ···················· ···················· ····················
6111 MW206531.1 |Severe acute respiratory syndrome coro ···················· ···················· ···················· ···················· ···················· ····················
6112 MW206532.1 |Severe acute respiratory syndrome coro ···················· ···················· ···················· ···················· ···················· ····················
6113 MW206533.1 |Severe acute respiratory syndrome coro ···················· ···················· ···················· ···················· ···················· ····················
6114 MW206534.1 |Severe acute respiratory syndrome coro ···················· ···················· ···················· ···················· ···················· ····················
6115 MW206535.1 |Severe acute respiratory syndrome coro ···················· ···················· ···················· ···················· ···················· ····················
6116 MW206536.1 |Severe acute respiratory syndrome coro ···················· ···················· ···················· ···················· ···················· ····················
6117 MW206539.1 |Severe acute respiratory syndrome coro ···················· ···················· ···················· ···················· ···················· ····················
6118 MW206540.1 |Severe acute respiratory syndrome coro ···················· ···················· ···················· ···················· ···················· ····················
6119 MW206541.1 |Severe acute respiratory syndrome coro ···················· ···················· ···················· ···················· ···················· ····················
6120 MW206542.1 |Severe acute respiratory syndrome coro ···················· ···················· ···················· ···················· ···················· ····················
6121 MW206547.1 |Severe acute respiratory syndrome coro ···················· ···················· ···················· ···················· ···················· ····················

| | | | | | | | | | |
|---|---|---|---|---|---|---|---|---|---|
| 6123 | MW206554.1 | \|Severe acute respiratory syndrome coro | ···················· | ···················· | ···················· | ···················· | ···················· | ···················· | |
| 6124 | MW206555.1 | \|Severe acute respiratory syndrome coro | ···················· | ···················· | ···················· | ···················· | ···················· | ···················· | |
| 6126 | MW206561.1 | \|Severe acute respiratory syndrome coro | ···················· | ···················· | ···················· | ···················· | ···················· | ···················· | |
| 6127 | MW206568.1 | \|Severe acute respiratory syndrome coro | ···················· | ···················· | ···················· | ···················· | ···················· | ···················· | |
| 6128 | MW206573.1 | \|Severe acute respiratory syndrome coro | ···················· | ···················· | ···················· | ···················· | ···················· | ···················· | |
| 6129 | MW206582.1 | \|Severe acute respiratory syndrome coro | ···················· | ···················· | ···················· | ···················· | ···················· | ···················· | |
| 6130 | MW206583.1 | \|Severe acute respiratory syndrome coro | ···················· | ···················· | ···················· | ···················· | ···················· | ···················· | |
| 6131 | MW206585.1 | \|Severe acute respiratory syndrome coro | ···················· | ···················· | ···················· | ···················· | ···················· | ···················· | |
| 6132 | MW206587.1 | \|Severe acute respiratory syndrome coro | ···················· | ···················· | ···················· | ···················· | ···················· | ···················· | |
| 6133 | MW206588.1 | \|Severe acute respiratory syndrome coro | ···················· | ···················· | ···················· | ···················· | ···················· | ···················· | |
| 6134 | MW206589.1 | \|Severe acute respiratory syndrome coro | ···················· | ···················· | ···················· | ···················· | ·NNN················ | ···················· | 1 |
| 6135 | MW206590.1 | \|Severe acute respiratory syndrome coro | ···················· | ···················· | ···················· | ···················· | ···················· | ···················· | |
| 6136 | MW206593.1 | \|Severe acute respiratory syndrome coro | ···················· | ···················· | ···················· | ···················· | ···················· | ···················· | |
| 6137 | MW206594.1 | \|Severe acute respiratory syndrome coro | ···················· | ···················· | ···················· | ···················· | ···················· | ···················· | |
| 6138 | MW206595.1 | \|Severe acute respiratory syndrome coro | ···················· | ···················· | ···················· | ···················· | ···················· | ···················· | |
| 6139 | MW206596.1 | \|Severe acute respiratory syndrome coro | ···················· | ···················· | ···················· | ···················· | ···················· | ···················· | |
| 6140 | MW206597.1 | \|Severe acute respiratory syndrome coro | ···················· | ···················· | ···················· | ···················· | ·NN················· | ···················· | |
| 6141 | MW206600.1 | \|Severe acute respiratory syndrome coro | ···················· | ···················· | ···················· | ···················· | ···················· | ···················· | |
| 6142 | MW206601.1 | \|Severe acute respiratory syndrome coro | ···················· | ···················· | ···················· | ···················· | ···················· | ···················· | |
| 6143 | MW206604.1 | \|Severe acute respiratory syndrome coro | ···················· | ···················· | ···················· | ···················· | ···················· | ···················· | |
| 6144 | MW206605.1 | \|Severe acute respiratory syndrome coro | ···················· | ···················· | ···················· | ···················· | ···················· | ···················· | |
| 6145 | MW206606.1 | \|Severe acute respiratory syndrome coro | ···················· | ·A·················· | ···················· | ···················· | ···················· | ···················· | |
| 6146 | MW206607.1 | \|Severe acute respiratory syndrome coro | ···················· | ···················· | ···················· | ···················· | ···················· | ···················· | |
| 6147 | MW206608.1 | \|Severe acute respiratory syndrome coro | ···················· | ···················· | ···················· | ···················· | ···················· | ···················· | |
| 6149 | MW206610.1 | \|Severe acute respiratory syndrome coro | ···················· | ···················· | ···················· | ···················· | ···················· | ···················· | |
| 6150 | MW206611.1 | \|Severe acute respiratory syndrome coro | ···················· | ···················· | ···················· | ···················· | ···················· | ···················· | |
| 6151 | MW206615.1 | \|Severe acute respiratory syndrome coro | ···················· | ···················· | ···················· | ···················· | ···················· | ···················· | |
| 6152 | MW206616.1 | \|Severe acute respiratory syndrome coro | ···················· | ···················· | ···················· | ···················· | ···················· | ···················· | |
| 6153 | MW206617.1 | \|Severe acute respiratory syndrome coro | ···················· | ···················· | ···················· | ···················· | ···················· | ···················· | |
| 6154 | MW206619.1 | \|Severe acute respiratory syndrome coro | ···················· | ···················· | ···················· | ···················· | ···················· | ···················· | |
| 6155 | MW206620.1 | \|Severe acute respiratory syndrome coro | ···················· | ···················· | ···················· | ···················· | ···················· | ···················· | |
| 6156 | MW206624.1 | \|Severe acute respiratory syndrome coro | ···················· | ···················· | ···················· | ···················· | ···················· | ···················· | |
| 6157 | MW206625.1 | \|Severe acute respiratory syndrome coro | ···················· | ···················· | ···················· | ···················· | ···················· | ···················· | |
| 6158 | MW206627.1 | \|Severe acute respiratory syndrome coro | ···················· | ···················· | ···················· | ···················· | ···················· | ···················· | |
| 6159 | MW206628.1 | \|Severe acute respiratory syndrome coro | ···················· | ···················· | ···················· | ···················· | ···················· | ···················· | |
| 6160 | MW206635.1 | \|Severe acute respiratory syndrome coro | ···················· | ···················· | ···················· | ···················· | ···················· | ···················· | |
| 6161 | MW208007.1 | \|Severe acute respiratory syndrome coro | ···················· | ···················· | ···················· | ···················· | ···················· | ···················· | |
| 6162 | MW208008.1 | \|Severe acute respiratory syndrome coro | ···················· | ···················· | ···················· | ···················· | ···················· | ···················· | |
| 6163 | MW208009.1 | \|Severe acute respiratory syndrome coro | ···················· | ···················· | ···················· | ···················· | ···················· | ···················· | |
| 6164 | MW208010.1 | \|Severe acute respiratory syndrome coro | ···················· | ···················· | ···················· | ···················· | ···················· | ···················· | |
| 6165 | MW208012.1 | \|Severe acute respiratory syndrome coro | ···················· | ······A············· | ···················· | ···················· | ···················· | ···················· | |
| 6166 | MW208013.1 | \|Severe acute respiratory syndrome coro | ···················· | ·A·················· | ···················· | ···················· | ···················· | ···················· | |
| 6167 | MW208014.1 | \|Severe acute respiratory syndrome coro | ···················· | ···················· | ···················· | ···················· | ···················· | ···················· | |
| 6168 | MW208015.1 | \|Severe acute respiratory syndrome coro | ···················· | ·A·················· | ···················· | ···················· | ···················· | ···················· | |
| 6169 | MW208016.1 | \|Severe acute respiratory syndrome coro | ···················· | ···················· | ···················· | ···················· | ···················· | ···················· | |
| 6170 | MW208017.1 | \|Severe acute respiratory syndrome coro | ···················· | ·A·················· | ···················· | ···················· | ···················· | ···················· | |
| 6171 | MW208018.1 | \|Severe acute respiratory syndrome coro | ···················· | ···················· | ···················· | ···················· | ···················· | ···················· | |
| 6172 | MW208019.1 | \|Severe acute respiratory syndrome coro | ···················· | ······A············· | ···················· | ···················· | ···················· | ···················· | |
| 6173 | MW208020.1 | \|Severe acute respiratory syndrome coro | ···················· | ···················· | ···················· | ···················· | ···················· | ···················· | |
| 6174 | MW208021.1 | \|Severe acute respiratory syndrome coro | ···················· | ···················· | ···················· | ···················· | ···················· | ···················· | |
| 6175 | MW208022.1 | \|Severe acute respiratory syndrome coro | ···················· | ···················· | ···················· | ···················· | ···················· | ···················· | |
| 6176 | MW208023.1 | \|Severe acute respiratory syndrome coro | ···················· | ···················· | ···················· | ···················· | ···················· | ···················· | |
| 6177 | MW208024.1 | \|Severe acute respiratory syndrome coro | ···················· | ···················· | ···················· | ···················· | ···················· | ···················· | |
| 6178 | MW208025.1 | \|Severe acute respiratory syndrome coro | ···················· | ···················· | ···················· | ···················· | ···················· | ···················· | |
| 6179 | MW208026.1 | \|Severe acute respiratory syndrome coro | ···················· | ···················· | ···················· | ···················· | ···················· | ···················· | |
| 6180 | MW208027.1 | \|Severe acute respiratory syndrome coro | ········T··········· | ·A·················· | ···················· | ···················· | ···················· | ···················· | |
| 6181 | MW208028.1 | \|Severe acute respiratory syndrome coro | ···················· | ···················· | ···················· | ···················· | ···················· | ···················· | |
| 6182 | MW208029.1 | \|Severe acute respiratory syndrome coro | ···················· | ·A·················· | ···················· | ···················· | ···················· | ···················· | |
| 6183 | MW208030.1 | \|Severe acute respiratory syndrome coro | ···················· | ···················· | ···················· | ···················· | ···················· | ···················· | |
| 6184 | MW208031.1 | \|Severe acute respiratory syndrome coro | ···················· | ···················· | ···················· | ···················· | ···················· | ···················· | |
| 6185 | MW208032.1 | \|Severe acute respiratory syndrome coro | ···················· | ····A··············· | ···················· | ···················· | ···················· | ···················· | |
| 6186 | MW208033.1 | \|Severe acute respiratory syndrome coro | ···················· | ···················· | ···················· | ···················· | ···················· | ···················· | |
| 6187 | MW208034.1 | \|Severe acute respiratory syndrome coro | ···················· | ···················· | ···················· | ···················· | ···················· | ···················· | |
| 6188 | MW208036.1 | \|Severe acute respiratory syndrome coro | ···················· | ···················· | ···················· | ···················· | ···················· | ···················· | |
| 6189 | MW208037.1 | \|Severe acute respiratory syndrome coro | ···················· | ···················· | ···················· | ···················· | ···················· | ···················· | |
| 6190 | MW208039.1 | \|Severe acute respiratory syndrome coro | ···················· | ·A·················· | ···················· | ···················· | ···················· | ···················· | |
| 6191 | MW210930.1 | \|Severe acute respiratory syndrome coro | ···················· | ·A·················· | ···················· | ···················· | ···················· | ···················· | |
| 6192 | MW210931.1 | \|Severe acute respiratory syndrome coro | ···················· | ·A·················· | ···················· | ···················· | ···················· | ···················· | |
| 6193 | MW210932.1 | \|Severe acute respiratory syndrome coro | ···················· | ·A·················· | ···················· | ···················· | ···················· | ···················· | |
| 6194 | MW210933.1 | \|Severe acute respiratory syndrome coro | ···················· | ·A·················· | ···················· | ···················· | ···················· | ···················· | |
| 6195 | MW210934.1 | \|Severe acute respiratory syndrome coro | ···················· | ·A·················· | ···················· | ···················· | ···················· | ···················· | |
| 6196 | MW210935.1 | \|Severe acute respiratory syndrome coro | ···················· | ···················· | ···················· | ···················· | ···················· | ···················· | |
| 6197 | MW210936.1 | \|Severe acute respiratory syndrome coro | ···················· | ···················· | ···················· | ···················· | ···················· | ···················· | |
| 6198 | MW210937.1 | \|Severe acute respiratory syndrome coro | ···················· | ···················· | ···················· | ···················· | ···················· | ···················· | |
| 6199 | MW210938.1 | \|Severe acute respiratory syndrome coro | ···················· | ···················· | ···················· | ···················· | ···················· | ···················· | |
| 6200 | MW210939.1 | \|Severe acute respiratory syndrome coro | ···················· | ···················· | ···················· | ···················· | ···················· | ···················· | |
| 6201 | MW210940.1 | \|Severe acute respiratory syndrome coro | ···················· | ···················· | ···················· | ···················· | ···················· | ···················· | |
| 6202 | MW210941.1 | \|Severe acute respiratory syndrome coro | ···················· | ···················· | ···················· | ···················· | ···················· | ···················· | |

| | | | | | | | | | |
|---|---|---|---|---|---|---|---|---|---|
| 6203 | MW210942.1 | \|Severe acute respiratory syndrome coro | .................... | .................... | .................... | .................... | .................... | .................... | |
| 6204 | MW210960.1 | \|Severe acute respiratory syndrome coro | .................... | .A.................. | .................... | .................... | .................... | .................... | |
| 6205 | MW210965.1 | \|Severe acute respiratory syndrome coro | .................... | .................... | .................... | .................... | .................... | .................... | |
| 6206 | MW210975.1 | \|Severe acute respiratory syndrome coro | .................... | .................... | .................... | .................... | .................... | .................... | |
| 6207 | MW210976.1 | \|Severe acute respiratory syndrome coro | .................... | .................... | .................... | .................... | .................... | .................... | |
| 6208 | MW210977.1 | \|Severe acute respiratory syndrome coro | .................... | .................... | .................... | .................... | .................... | .................... | |
| 6209 | MW210978.1 | \|Severe acute respiratory syndrome coro | .................... | .................... | .................... | .................... | .................... | .................... | |
| 6210 | MW210979.1 | \|Severe acute respiratory syndrome coro | .................... | .................... | .................... | .................... | .................... | .................... | |
| 6211 | MW210980.1 | \|Severe acute respiratory syndrome coro | .................... | .................... | .................... | .................... | .................... | .................... | |
| 6212 | MW210982.1 | \|Severe acute respiratory syndrome coro | .................... | .................... | .................... | .................... | .................... | .................... | |
| 6213 | MW210983.1 | \|Severe acute respiratory syndrome coro | .................... | .................... | .................... | .................... | .................... | .................... | |
| 6214 | MW210984.1 | \|Severe acute respiratory syndrome coro | .................... | .................... | .................... | .................... | .................... | .................... | |
| 6215 | MW210985.1 | \|Severe acute respiratory syndrome coro | .................... | .................... | .................... | .................... | .................... | .................... | |
| 6216 | MW210986.1 | \|Severe acute respiratory syndrome coro | .................... | .................... | .................... | .................... | .................... | .................... | |
| 6217 | MW210987.1 | \|Severe acute respiratory syndrome coro | .................... | .................... | .................... | .................... | .................... | .................... | |
| 6218 | MW210988.1 | \|Severe acute respiratory syndrome coro | .................... | .................... | .................... | .................... | .................... | .................... | |
| 6219 | MW210989.1 | \|Severe acute respiratory syndrome coro | .................... | .................... | .................... | .................... | .................... | .................... | |
| 6220 | MW210990.1 | \|Severe acute respiratory syndrome coro | .................... | .................... | .................... | .................... | .................... | .................... | |
| 6221 | MW210991.1 | \|Severe acute respiratory syndrome coro | .................... | .................... | .................... | .................... | .................... | .................... | |
| 6222 | MW210992.1 | \|Severe acute respiratory syndrome coro | .................... | .................... | .................... | .................... | .................... | .................... | |
| 6223 | MW210993.1 | \|Severe acute respiratory syndrome coro | .................... | .................... | .................... | .................... | .................... | .................... | |
| 6224 | MW210994.1 | \|Severe acute respiratory syndrome coro | .................... | .................... | .................... | .................... | .................... | .................... | |
| 6225 | MW210995.1 | \|Severe acute respiratory syndrome coro | .................... | .................... | .................... | .................... | .................... | .................... | |
| 6226 | MW210996.1 | \|Severe acute respiratory syndrome coro | .................... | .................... | .................... | .................... | .................... | .................... | |
| 6227 | MW210997.1 | \|Severe acute respiratory syndrome coro | .................... | .................... | .................... | .................... | .................... | .................... | |
| 6228 | MW210998.1 | \|Severe acute respiratory syndrome coro | .................... | .A.................. | .................... | .................... | .................... | .................... | |
| 6229 | MW210999.1 | \|Severe acute respiratory syndrome coro | .................... | .A.................. | .................... | .................... | .................... | .................... | |
| 6230 | MW211001.1 | \|Severe acute respiratory syndrome coro | .................... | .A.................. | .................... | .................... | .................... | .................... | |
| 6231 | MW211002.1 | \|Severe acute respiratory syndrome coro | .................... | .................... | .................... | .................... | .................... | .................... | |
| 6232 | MW211003.1 | \|Severe acute respiratory syndrome coro | .................... | .................... | .................... | .................... | .................... | .................... | |
| 6233 | MW211004.1 | \|Severe acute respiratory syndrome coro | .................... | .................... | .................... | .................... | .................... | .................... | |
| 6234 | MW211005.1 | \|Severe acute respiratory syndrome coro | ..........T......... | .................... | .................... | .................... | .................... | .................... | |
| 6235 | MW211006.1 | \|Severe acute respiratory syndrome coro | ..........T......... | .................... | .................... | .................... | .................... | .................... | |
| 6236 | MW211008.1 | \|Severe acute respiratory syndrome coro | .................... | .................... | .................... | .................... | .................... | .................... | |
| 6237 | MW211009.1 | \|Severe acute respiratory syndrome coro | .................... | .................... | .................... | .................... | .................... | .................... | |
| 6238 | MW211010.1 | \|Severe acute respiratory syndrome coro | .................... | .................... | .................... | .................... | .................... | .................... | |
| 6239 | MW199143.1 | \|Severe acute respiratory syndrome coro | .................... | .................... | .................... | .................... | .................... | .................... | |
| 6240 | MW199149.1 | \|Severe acute respiratory syndrome coro | .................... | .................... | .................... | .................... | .................... | .................... | |
| 6241 | MW199150.1 | \|Severe acute respiratory syndrome coro | .................... | .................... | .................... | .................... | .................... | .................... | |
| 6242 | MW199186.1 | \|Severe acute respiratory syndrome coro | .................... | .................... | .................... | .................... | .................... | .................... | |
| 6243 | MW201576.1 | \|Severe acute respiratory syndrome coro | .................... | .A.................. | .................... | .................... | .................... | .................... | |
| 6244 | MW201577.1 | \|Severe acute respiratory syndrome coro | .................... | .................... | .................... | .................... | .................... | .................... | |
| 6245 | MW201578.1 | \|Severe acute respiratory syndrome coro | .................... | .................... | .................... | .................... | .................... | .................... | |
| 6246 | MW202150.1 | \|Severe acute respiratory syndrome coro | .................... | .............A...... | .................... | .................... | .................... | .................... | |
| 6247 | MW202151.1 | \|Severe acute respiratory syndrome coro | .................... | .................... | .................... | .................... | .................... | .................... | |
| 6248 | MW202152.1 | \|Severe acute respiratory syndrome coro | .................... | .............A...... | .................... | .................... | .................... | .................... | |
| 6249 | MW202153.1 | \|Severe acute respiratory syndrome coro | .................... | .............A...... | .................... | .................... | .................... | .................... | |
| 6250 | MW202154.1 | \|Severe acute respiratory syndrome coro | .................... | .............A...... | .................... | .................... | .................... | .................... | |
| 6251 | MW202155.1 | \|Severe acute respiratory syndrome coro | .................... | .............A...... | .................... | .................... | .................... | .................... | |
| 6252 | MW202156.1 | \|Severe acute respiratory syndrome coro | .................... | .............A...... | .................... | .................... | .................... | .................... | |
| 6253 | MW202157.1 | \|Severe acute respiratory syndrome coro | .................... | .............A...... | .................... | .................... | .................... | .................... | |
| 6254 | MW202158.1 | \|Severe acute respiratory syndrome coro | .................... | .............A...... | .................... | .................... | .................... | .................... | |
| 6255 | MW202159.1 | \|Severe acute respiratory syndrome coro | .................... | .............A...... | .................... | .................... | .................... | .................... | |
| 6256 | MW202160.1 | \|Severe acute respiratory syndrome coro | .................... | .............A...... | .................... | .................... | .................... | .................... | |
| 6257 | MW202161.1 | \|Severe acute respiratory syndrome coro | .................... | .............A...... | .................... | .................... | .................... | .................... | |
| 6258 | MW202162.1 | \|Severe acute respiratory syndrome coro | .................... | .A.................. | .................... | .................... | .................... | .................... | |
| 6259 | MW202163.1 | \|Severe acute respiratory syndrome coro | .................... | .............A...... | .................... | .................... | .NNN................ | .................... | 1 |
| 6260 | MW202164.1 | \|Severe acute respiratory syndrome coro | .................... | .............A...... | .................... | .................... | .................... | .................... | |
| 6261 | MW202165.1 | \|Severe acute respiratory syndrome coro | .................... | .................... | .................... | .................... | .................... | .................... | |
| 6262 | MW202166.1 | \|Severe acute respiratory syndrome coro | .................... | .............A...... | .................... | .................... | .................... | .................... | |
| 6263 | MW202167.1 | \|Severe acute respiratory syndrome coro | .................... | .............A...... | .................... | .................... | .................... | .................... | |
| 6264 | MW202168.1 | \|Severe acute respiratory syndrome coro | .................... | .................... | .................... | .................... | .................... | .................... | |
| 6265 | MW202169.1 | \|Severe acute respiratory syndrome coro | .................... | .................... | .................... | .................... | .................... | .................... | |
| 6266 | MW202170.1 | \|Severe acute respiratory syndrome coro | .................... | .A.................. | .................... | .................... | .................... | .................... | |
| 6267 | MW202171.1 | \|Severe acute respiratory syndrome coro | .................... | .................... | .................... | .................... | .................... | .................... | |
| 6268 | MW202172.1 | \|Severe acute respiratory syndrome coro | .................... | .................... | .................... | .................... | .................... | .................... | |
| 6269 | MW202173.1 | \|Severe acute respiratory syndrome coro | .................... | .A.................. | .................... | .................... | .................... | .................... | |
| 6270 | MW202174.1 | \|Severe acute respiratory syndrome coro | .................... | ......A......A...... | .................... | .................... | .................... | .................... | |
| 6271 | MW202175.1 | \|Severe acute respiratory syndrome coro | .................... | .................... | .................... | .................... | .................... | .................... | |
| 6272 | MW202176.1 | \|Severe acute respiratory syndrome coro | .................... | .................... | .................... | .................... | .................... | .................... | |
| 6273 | MW202177.1 | \|Severe acute respiratory syndrome coro | .................... | .................... | .................... | .................... | .................... | .................... | |
| 6274 | MW202178.1 | \|Severe acute respiratory syndrome coro | .................... | .................... | .................... | .................... | .................... | .................... | |
| 6275 | MW202179.1 | \|Severe acute respiratory syndrome coro | .................... | .................... | .................... | .................... | .................... | .................... | |
| 6276 | MW202180.1 | \|Severe acute respiratory syndrome coro | .................... | .................... | .................... | .................... | .................... | .................... | |
| 6277 | MW202181.1 | \|Severe acute respiratory syndrome coro | .................... | .................... | .................... | .................... | .................... | .................... | |
| 6278 | MW202182.1 | \|Severe acute respiratory syndrome coro | .................... | .................... | .................... | .................... | .................... | .................... | |
| 6279 | MW202183.1 | \|Severe acute respiratory syndrome coro | .................... | .................... | .................... | .................... | .................... | .................... | |
| 6280 | MW202184.1 | \|Severe acute respiratory syndrome coro | .................... | .................... | .................... | .................... | .................... | .................... | |

| | | | | | | | | | |
|---|---|---|---|---|---|---|---|---|---|
| 6281 | MW202185.1 | \|Severe acute respiratory syndrome coro | ···················· | ···················· | ···················· | ···················· | ···················· | ···················· | |
| 6282 | MW202186.1 | \|Severe acute respiratory syndrome coro | ···················· | ···················· | ···················· | ···················· | ···················· | ···················· | |
| 6283 | MW202187.1 | \|Severe acute respiratory syndrome coro | ···················· | ···················· | ···················· | ···················· | ···················· | ···················· | |
| 6284 | MW202188.1 | \|Severe acute respiratory syndrome coro | ···················· | ···················· | ···················· | ···················· | ··NN················ | ···················· | |
| 6285 | MW202189.1 | \|Severe acute respiratory syndrome coro | ···················· | ···················· | ···················· | ···················· | ···················· | ···················· | |
| 6286 | MW202190.1 | \|Severe acute respiratory syndrome coro | ···················· | ···················· | ···················· | ···················· | ···················· | ···················· | |
| 6287 | MW202191.1 | \|Severe acute respiratory syndrome coro | ···················· | ···················· | ···················· | ···················· | ···················· | ···················· | |
| 6288 | MW202192.1 | \|Severe acute respiratory syndrome coro | ···················· | ···················· | ···················· | ···················· | ···················· | ···················· | |
| 6289 | MW202193.1 | \|Severe acute respiratory syndrome coro | ···················· | ···················· | ···················· | ···················· | ···················· | ···················· | |
| 6290 | MW202194.1 | \|Severe acute respiratory syndrome coro | ···················· | ···················· | ···················· | ···················· | ···················· | ···················· | |
| 6291 | MW202195.1 | \|Severe acute respiratory syndrome coro | ···················· | ···················· | ···················· | ···················· | ···················· | ···················· | |
| 6292 | MW202196.1 | \|Severe acute respiratory syndrome coro | ···················· | ···················· | ···················· | ···················· | ···················· | ···················· | |
| 6293 | MW202197.1 | \|Severe acute respiratory syndrome coro | ···················· | ···················· | ···················· | ···················· | ···················· | ···················· | |
| 6294 | MW202198.1 | \|Severe acute respiratory syndrome coro | ···················· | ···················· | ···················· | ···················· | ···················· | ···················· | |
| 6295 | MW202199.1 | \|Severe acute respiratory syndrome coro | ···················· | ·············A······ | ···················· | ···················· | ···················· | ···················· | |
| 6296 | MW202200.1 | \|Severe acute respiratory syndrome coro | ···················· | ·············A······ | ···················· | ···················· | ···················· | ···················· | |
| 6297 | MW202201.1 | \|Severe acute respiratory syndrome coro | ···················· | ·············A······ | ···················· | ···················· | ···················· | ···················· | |
| 6298 | MW202202.1 | \|Severe acute respiratory syndrome coro | ···················· | ·············A······ | ···················· | ···················· | ···················· | ···················· | |
| 6299 | MW202203.1 | \|Severe acute respiratory syndrome coro | ···················· | ·············A······ | ···················· | ···················· | ···················· | ···················· | |
| 6300 | MW202204.1 | \|Severe acute respiratory syndrome coro | ···················· | ·············A······ | ···················· | ···················· | ···················· | ···················· | |
| 6301 | MW202205.1 | \|Severe acute respiratory syndrome coro | ···················· | ·············A······ | ···················· | ···················· | ···················· | ···················· | |
| 6302 | MW202206.1 | \|Severe acute respiratory syndrome coro | ···················· | ·············A······ | ···················· | ···················· | ···················· | ···················· | |
| 6303 | MW202207.1 | \|Severe acute respiratory syndrome coro | ···················· | ·············A······ | ···················· | ···················· | ···················· | ···················· | |
| 6304 | MW202208.1 | \|Severe acute respiratory syndrome coro | ···················· | ·············A······ | ···················· | ···················· | ···················· | ···················· | |
| 6305 | MW202209.1 | \|Severe acute respiratory syndrome coro | ···················· | ·············A······ | ···················· | ···················· | ···················· | ···················· | |
| 6306 | MW202210.1 | \|Severe acute respiratory syndrome coro | ···················· | ···················· | ···················· | ···················· | ···················· | ···················· | |
| 6307 | MW202211.1 | \|Severe acute respiratory syndrome coro | ···················· | ···················· | ···················· | ···················· | ···················· | ···················· | |
| 6308 | MW202212.1 | \|Severe acute respiratory syndrome coro | ···················· | ···················· | ···················· | ···················· | ···················· | ···················· | |
| 6309 | MW202213.1 | \|Severe acute respiratory syndrome coro | ···················· | ···················· | ···················· | ···················· | ···················· | ···················· | |
| 6310 | MW202214.1 | \|Severe acute respiratory syndrome coro | ···················· | ···················· | ···················· | ···················· | ···················· | ···················· | |
| 6311 | MW202215.1 | \|Severe acute respiratory syndrome coro | ···················· | ···················· | ···················· | ···················· | ···················· | ···················· | |
| 6312 | MW202216.1 | \|Severe acute respiratory syndrome coro | ···················· | ·············A······ | ···················· | ···················· | ···················· | ···················· | |
| 6313 | MW202217.1 | \|Severe acute respiratory syndrome coro | ···················· | ···················· | ···················· | ···················· | ···················· | ···················· | |
| 6314 | MW202218.1 | \|Severe acute respiratory syndrome coro | ···················· | ···················· | ···················· | ···················· | ···················· | ···················· | |
| 6315 | MW202219.1 | \|Severe acute respiratory syndrome coro | ···················· | ·A·················· | ···················· | ···················· | ···················· | ···················· | |
| 6316 | MW202220.1 | \|Severe acute respiratory syndrome coro | ···················· | ·············A······ | ···················· | ···················· | ···················· | ···················· | |
| 6317 | MW202221.1 | \|Severe acute respiratory syndrome coro | ···················· | ·············A······ | ···················· | ···················· | ···················· | ···················· | |
| 6318 | MW202222.1 | \|Severe acute respiratory syndrome coro | ···················· | ·············A······ | ···················· | ···················· | ···················· | ···················· | |
| 6319 | MW202223.1 | \|Severe acute respiratory syndrome coro | ···················· | ·············A······ | ···················· | ···················· | ···················· | ···················· | |
| 6320 | MW202224.1 | \|Severe acute respiratory syndrome coro | ···················· | ·············A······ | ···················· | ···················· | ···················· | ···················· | |
| 6321 | MW202225.1 | \|Severe acute respiratory syndrome coro | ···················· | ·············A······ | ···················· | ···················· | NNN················· | ···················· | 1 |
| 6322 | MW202226.1 | \|Severe acute respiratory syndrome coro | ···················· | ·············A······ | ···················· | ···················· | ···················· | ···················· | |
| 6323 | MW202227.1 | \|Severe acute respiratory syndrome coro | ···················· | ·············A······ | ···················· | ···················· | ···················· | ···················· | |
| 6324 | MW193407.1 | \|Severe acute respiratory syndrome coro | ···················· | ···················· | ···················· | ···················· | ···················· | ···················· | |
| 6325 | MW193408.1 | \|Severe acute respiratory syndrome coro | ···················· | ···················· | ···················· | ···················· | ···················· | ···················· | |
| 6326 | MW193889.1 | \|Severe acute respiratory syndrome coro | ···················· | ···················· | ···················· | ···················· | ···················· | ···················· | |
| 6327 | MW193967.2 | \|Severe acute respiratory syndrome coro | ···················· | ···················· | ···················· | ···················· | ···················· | ···················· | |
| 6328 | MW194121.1 | \|Severe acute respiratory syndrome coro | ···················· | ····A··············· | ···················· | ···················· | ···················· | ···················· | |
| 6329 | MW195531.1 | \|Severe acute respiratory syndrome coro | ···················· | ···················· | ···················· | ···················· | ···················· | ···················· | |
| 6330 | MW195532.1 | \|Severe acute respiratory syndrome coro | ···················· | ···················· | ···················· | ···················· | ···················· | ···················· | |
| 6331 | MW195533.1 | \|Severe acute respiratory syndrome coro | ···················· | ·A·················· | ···················· | ···················· | ···················· | ···················· | |
| 6332 | MW195534.1 | \|Severe acute respiratory syndrome coro | ···················· | ···················· | ···················· | ···················· | ···················· | ···················· | |
| 6333 | MW195535.1 | \|Severe acute respiratory syndrome coro | ···················· | ···················· | ···················· | ···················· | ···················· | ···················· | |
| 6334 | MW195536.1 | \|Severe acute respiratory syndrome coro | ···················· | ···················· | ···················· | ···················· | ···················· | ···················· | |
| 6335 | MW195537.1 | \|Severe acute respiratory syndrome coro | ···················· | ···················· | ···················· | ···················· | ···················· | ···················· | |
| 6336 | MW195538.1 | \|Severe acute respiratory syndrome coro | ···················· | ···················· | ···················· | ···················· | ···················· | ···················· | |
| 6337 | MW195539.1 | \|Severe acute respiratory syndrome coro | ···················· | ···················· | ···················· | ···················· | ···················· | ···················· | |
| 6338 | MW195540.1 | \|Severe acute respiratory syndrome coro | ···················· | ···················· | ···················· | ···················· | ···················· | ···················· | |
| 6339 | MW195541.1 | \|Severe acute respiratory syndrome coro | ···················· | ···················· | ···················· | ···················· | ···················· | ···················· | |
| 6340 | MW195542.1 | \|Severe acute respiratory syndrome coro | ···················· | ···················· | ···················· | ···················· | ···················· | ···················· | |
| 6341 | MW195543.1 | \|Severe acute respiratory syndrome coro | ···················· | ···················· | ···················· | ···················· | ···················· | ···················· | |
| 6342 | MW195544.1 | \|Severe acute respiratory syndrome coro | ···················· | ·A·················· | ···················· | ···················· | ···················· | ···················· | |
| 6343 | MW195545.1 | \|Severe acute respiratory syndrome coro | ···················· | ·A·················· | ···················· | ···················· | ···················· | ···················· | |
| 6344 | MW195546.1 | \|Severe acute respiratory syndrome coro | ···················· | ·A·················· | ···················· | ···················· | ···················· | ···················· | |
| 6345 | MW197470.1 | \|Severe acute respiratory syndrome coro | ···················· | ···················· | ···················· | ···················· | ···················· | ···················· | |
| 6346 | MW197471.1 | \|Severe acute respiratory syndrome coro | ···················· | ···················· | ···················· | ···················· | ···················· | ···················· | |
| 6347 | MW197472.1 | \|Severe acute respiratory syndrome coro | ···················· | ···················· | ···················· | ···················· | ···················· | ···················· | |
| 6348 | MW197473.1 | \|Severe acute respiratory syndrome coro | ···················· | ···················· | ···················· | ···················· | ···················· | ···················· | |
| 6349 | MW197474.1 | \|Severe acute respiratory syndrome coro | ···················· | ···················· | ···················· | ···················· | ···················· | ···················· | |
| 6350 | MW197475.1 | \|Severe acute respiratory syndrome coro | ···················· | ···················· | ···················· | ···················· | ···················· | ···················· | |
| 6351 | MW197476.1 | \|Severe acute respiratory syndrome coro | ···················· | ···················· | ···················· | ···················· | ···················· | ···················· | |
| 6352 | MW197477.1 | \|Severe acute respiratory syndrome coro | ···················· | ···················· | ···················· | ···················· | ···················· | ···················· | |
| 6353 | MW197479.1 | \|Severe acute respiratory syndrome coro | ···················· | ···················· | ···················· | ···················· | ···················· | ···················· | |
| 6354 | MW197480.1 | \|Severe acute respiratory syndrome coro | ···················· | ···················· | ···················· | ···················· | ···················· | ···················· | |
| 6355 | MW197481.1 | \|Severe acute respiratory syndrome coro | ···················· | ···················· | ···················· | ···················· | ···················· | ···················· | |
| 6356 | MW197482.1 | \|Severe acute respiratory syndrome coro | ···················· | ···················· | ···················· | ···················· | ···················· | ···················· | |
| 6357 | MW197483.1 | \|Severe acute respiratory syndrome coro | ···················· | ·············A······ | ···················· | ···················· | ···················· | ···················· | |
| 6358 | MW197485.1 | \|Severe acute respiratory syndrome coro | ···················· | ···················· | ···················· | ···················· | ···················· | ···················· | |

| | | | | | | | | |
|---|---|---|---|---|---|---|---|---|
| 6359 | MW197486.1 | \|Severe acute respiratory syndrome coro | ···················· | ·A·················· | ···················· | ···················· | ···················· | ···················· |
| 6360 | MW197487.1 | \|Severe acute respiratory syndrome coro | ···················· | ···················· | ···················· | ···················· | ···················· | ···················· |
| 6361 | MW197488.1 | \|Severe acute respiratory syndrome coro | ···················· | ···················· | ···················· | ···················· | ···················· | ···················· |
| 6362 | MW197489.1 | \|Severe acute respiratory syndrome coro | ···················· | ·A·················· | ···················· | ···················· | ···················· | ···················· |
| 6363 | MW197490.1 | \|Severe acute respiratory syndrome coro | ···················· | ···················· | ···················· | ···················· | ···················· | ···················· |
| 6364 | MW197491.1 | \|Severe acute respiratory syndrome coro | ···················· | ···················· | ···················· | ···················· | ···················· | ···················· |
| 6365 | MW197492.1 | \|Severe acute respiratory syndrome coro | ···················· | ···················· | ···················· | ···················· | ···················· | ···················· |
| 6366 | MW197493.1 | \|Severe acute respiratory syndrome coro | ···················· | ···················· | ···················· | ···················· | ···················· | ···················· |
| 6367 | MW197494.1 | \|Severe acute respiratory syndrome coro | ···················· | ···················· | ···················· | ···················· | ···················· | ···················· |
| 6368 | MW197496.1 | \|Severe acute respiratory syndrome coro | ···················· | ·A·················· | ···················· | ···················· | ···················· | ···················· |
| 6369 | MW197497.1 | \|Severe acute respiratory syndrome coro | ···················· | ·A·················· | ···················· | ···················· | ···················· | ···················· |
| 6370 | MW197498.1 | \|Severe acute respiratory syndrome coro | ···················· | ···················· | ···················· | ···················· | ···················· | ···················· |
| 6371 | MW059036.1 | \|Severe acute respiratory syndrome coro | ···················· | ···················· | ···················· | ···················· | ···················· | ···················· |
| 6372 | MW181704.1 | \|Severe acute respiratory syndrome coro | ···················· | ···················· | ···················· | ···················· | ···················· | ···················· |
| 6373 | MW181705.1 | \|Severe acute respiratory syndrome coro | ···················· | ···················· | ···················· | ···················· | ···················· | ···················· |
| 6374 | MW181706.1 | \|Severe acute respiratory syndrome coro | ···················· | ···················· | ···················· | ···················· | ···················· | ···················· |
| 6375 | MW181707.1 | \|Severe acute respiratory syndrome coro | ···················· | ···················· | ···················· | ···················· | ···················· | ···················· |
| 6376 | MW181708.1 | \|Severe acute respiratory syndrome coro | ···················· | ···················· | ···················· | ···················· | ···················· | ···················· |
| 6377 | MW181709.1 | \|Severe acute respiratory syndrome coro | ···················· | ···················· | ···················· | ···················· | ···················· | ···················· |
| 6378 | MW181710.1 | \|Severe acute respiratory syndrome coro | ···················· | ···················· | ···················· | ···················· | ···················· | ···················· |
| 6379 | MW181711.1 | \|Severe acute respiratory syndrome coro | ···················· | ···················· | ···················· | ···················· | ···················· | ···················· |
| 6380 | MW181712.1 | \|Severe acute respiratory syndrome coro | ···················· | ···················· | ···················· | ···················· | ···················· | ···················· |
| 6381 | MW181713.1 | \|Severe acute respiratory syndrome coro | ···················· | ···················· | ···················· | ···················· | ···················· | ···················· |
| 6382 | MW181714.1 | \|Severe acute respiratory syndrome coro | ···················· | ···················· | ···················· | ···················· | ···················· | ···················· |
| 6383 | MW181716.1 | \|Severe acute respiratory syndrome coro | ···················· | ···················· | ···················· | ···················· | ···················· | ···················· |
| 6384 | MW181718.1 | \|Severe acute respiratory syndrome coro | ···················· | ···················· | ···················· | ···················· | ···················· | ···················· |
| 6385 | MW181719.1 | \|Severe acute respiratory syndrome coro | ···················· | ···················· | ···················· | ···················· | ···················· | ···················· |
| 6386 | MW181720.1 | \|Severe acute respiratory syndrome coro | ···················· | ···················· | ···················· | ···················· | ···················· | ···················· |
| 6387 | MW181721.1 | \|Severe acute respiratory syndrome coro | ···················· | ···················· | ···················· | ···················· | ···················· | ···················· |
| 6388 | MW181722.1 | \|Severe acute respiratory syndrome coro | ···················· | ···················· | ···················· | ···················· | ···················· | ···················· |
| 6389 | MW181723.1 | \|Severe acute respiratory syndrome coro | ···················· | ···················· | ···················· | ···················· | ···················· | ···················· |
| 6390 | MW181724.1 | \|Severe acute respiratory syndrome coro | ···················· | ···················· | ···················· | ···················· | ···················· | ···················· |
| 6391 | MW181725.1 | \|Severe acute respiratory syndrome coro | ···················· | ···················· | ···················· | ···················· | ···················· | ···················· |
| 6392 | MW181726.1 | \|Severe acute respiratory syndrome coro | ···················· | ···················· | ···················· | ···················· | ···················· | ···················· |
| 6393 | MW181727.1 | \|Severe acute respiratory syndrome coro | ···················· | ···················· | ···················· | ···················· | ···················· | ···················· |
| 6394 | MW181728.1 | \|Severe acute respiratory syndrome coro | ···················· | ···················· | ···················· | ···················· | ···················· | ···················· |
| 6395 | MW181729.1 | \|Severe acute respiratory syndrome coro | ···················· | ···················· | ···················· | ···················· | ···················· | ···················· |
| 6396 | MW181730.1 | \|Severe acute respiratory syndrome coro | ···················· | ···················· | ···················· | ···················· | ···················· | ···················· |
| 6397 | MW181731.1 | \|Severe acute respiratory syndrome coro | ···················· | ···················· | ···················· | ···················· | ···················· | ···················· |
| 6398 | MW181733.1 | \|Severe acute respiratory syndrome coro | ···················· | ···················· | ···················· | ···················· | ···················· | ···················· |
| 6399 | MW181737.1 | \|Severe acute respiratory syndrome coro | ···················· | ···················· | ···················· | ···················· | ···················· | ···················· |
| 6400 | MW181740.1 | \|Severe acute respiratory syndrome coro | ···················· | ·A·················· | ···················· | ···················· | ···················· | ··A················· |
| 6401 | MW181741.1 | \|Severe acute respiratory syndrome coro | ···················· | ·A·················· | ···················· | ···················· | ···················· | ··A················· |
| 6402 | MW181742.1 | \|Severe acute respiratory syndrome coro | ···················· | ·A·················· | ···················· | ···················· | ···················· | ··A················· |
| 6403 | MW181743.1 | \|Severe acute respiratory syndrome coro | ···················· | ·A·················· | ···················· | ···················· | ···················· | ··A················· |
| 6404 | MW181744.1 | \|Severe acute respiratory syndrome coro | ···················· | ···················· | ···················· | ···················· | ···················· | ···················· |
| 6405 | MW181745.1 | \|Severe acute respiratory syndrome coro | ···················· | ···················· | ···················· | ···················· | ···················· | ···················· |
| 6406 | MW181746.1 | \|Severe acute respiratory syndrome coro | ···················· | ···················· | ···················· | ···················· | ···················· | ···················· |
| 6407 | MW181747.1 | \|Severe acute respiratory syndrome coro | ···················· | ···················· | ···················· | ···················· | ···················· | ···················· |
| 6408 | MW181748.1 | \|Severe acute respiratory syndrome coro | ···················· | ···················· | ···················· | ···················· | ···················· | ···················· |
| 6409 | MW181749.1 | \|Severe acute respiratory syndrome coro | ···················· | ···················· | ···················· | ···················· | ···················· | ···················· |
| 6410 | MW181751.1 | \|Severe acute respiratory syndrome coro | ···················· | ···················· | ···················· | ···················· | ···················· | ···················· |
| 6411 | MW181752.1 | \|Severe acute respiratory syndrome coro | ···················· | ···················· | ···················· | ···················· | ···················· | ···················· |
| 6412 | MW181753.1 | \|Severe acute respiratory syndrome coro | ···················· | ·A·················· | ···················· | ···················· | ···················· | ··A················· |
| 6413 | MW181754.1 | \|Severe acute respiratory syndrome coro | ···················· | ·A·················· | ···················· | ···················· | ···················· | ··A················· |
| 6414 | MW181758.1 | \|Severe acute respiratory syndrome coro | ···················· | ·A·················· | ···················· | ···················· | ···················· | ···················· |
| 6415 | MW181759.1 | \|Severe acute respiratory syndrome coro | ···················· | ···················· | ···················· | ···················· | ···················· | ···················· |
| 6416 | MW181761.1 | \|Severe acute respiratory syndrome coro | ···················· | ·A·················· | ···················· | ···················· | ···················· | ··A················· |
| 6417 | MW181764.1 | \|Severe acute respiratory syndrome coro | ···················· | ···················· | ···················· | ···················· | ···················· | ···················· |
| 6418 | MT412338.1 | \|Severe acute respiratory syndrome coro | ···················· | ···················· | ···················· | ···················· | ···················· | ···················· |
| 6419 | MT412339.1 | \|Severe acute respiratory syndrome coro | ···················· | ···················· | ···················· | ···················· | ···················· | ···················· |
| 6420 | MT412340.1 | \|Severe acute respiratory syndrome coro | ···················· | ···················· | ···················· | ···················· | ···················· | ···················· |
| 6421 | MT581410.1 | \|Severe acute respiratory syndrome coro | ···················· | ···················· | ···················· | ···················· | ···················· | ···················· |
| 6422 | MT581411.1 | \|Severe acute respiratory syndrome coro | ···················· | ···················· | ···················· | ···················· | ···················· | ···················· |
| 6423 | MT581412.1 | \|Severe acute respiratory syndrome coro | ···················· | ···················· | ···················· | ···················· | ···················· | ···················· |
| 6424 | MT581413.1 | \|Severe acute respiratory syndrome coro | ···················· | ···················· | ···················· | ···················· | ···················· | ···················· |
| 6425 | MT581414.1 | \|Severe acute respiratory syndrome coro | ···················· | ···················· | ···················· | ···················· | ···················· | ···················· |
| 6426 | MT581415.1 | \|Severe acute respiratory syndrome coro | ···················· | ···················· | ···················· | ···················· | ···················· | ···················· |
| 6427 | MT581416.1 | \|Severe acute respiratory syndrome coro | ···················· | ···················· | ···················· | ···················· | ···················· | ···················· |
| 6428 | MT581417.1 | \|Severe acute respiratory syndrome coro | ···················· | ···················· | ···················· | ···················· | ···················· | ···················· |
| 6429 | MT581418.1 | \|Severe acute respiratory syndrome coro | ···················· | ···················· | ···················· | ···················· | ···················· | ···················· |
| 6430 | MT581419.1 | \|Severe acute respiratory syndrome coro | ···················· | ···················· | ···················· | ···················· | ···················· | ···················· |
| 6431 | MT581420.1 | \|Severe acute respiratory syndrome coro | ···················· | ···················· | ···················· | ···················· | ···················· | ···················· |
| 6432 | MT581421.1 | \|Severe acute respiratory syndrome coro | ···················· | ···················· | ···················· | ···················· | ···················· | ···················· |
| 6433 | MT581422.1 | \|Severe acute respiratory syndrome coro | ···················· | ···················· | ···················· | ···················· | ···················· | ···················· |
| 6434 | MT581423.1 | \|Severe acute respiratory syndrome coro | ···················· | ···················· | ···················· | ···················· | ···················· | ···················· |
| 6435 | MT581426.1 | \|Severe acute respiratory syndrome coro | ···················· | ···················· | ···················· | ···················· | ···················· | ···················· |
| 6436 | MT581427.1 | \|Severe acute respiratory syndrome coro | ···················· | ···················· | ···················· | ···················· | ···················· | ···················· |

```
6437 MT581428.1 |Severe acute respiratory syndrome coro    ....................  ....................  ....................  ....................  ....................  ....................
6438 MT581429.1 |Severe acute respiratory syndrome coro    ....................  ....................  ....................  ....................  ....................  ....................
6439 MT581430.1 |Severe acute respiratory syndrome coro    ....................  ....................  ....................  ....................  ....................  ....................
6440 MT581431.1 |Severe acute respiratory syndrome coro    ....................  ....................  ....................  ....................  ....................  ....................
6441 MT581432.1 |Severe acute respiratory syndrome coro    ....................  ....................  ....................  ....................  ....................  ....................
6442 MT581434.1 |Severe acute respiratory syndrome coro    ....................  ....................  ....................  ....................  ....................  ....................
6443 MT581435.1 |Severe acute respiratory syndrome coro    ....................  ....................  ....................  ....................  ....................  ....................
6444 MT581436.1 |Severe acute respiratory syndrome coro    ....................  ....................  ....................  ....................  ....................  ....................
6445 MT607972.1 |Severe acute respiratory syndrome coro    ....................  ....................  ....................  ....................  ....................  ....................
6446 MT607973.1 |Severe acute respiratory syndrome coro    ....................  ....................  ....................  ....................  ....................  ....................
6447 MT607974.1 |Severe acute respiratory syndrome coro    ....................  ....................  ....................  ....................  ....................  ....................
6448 MT607975.1 |Severe acute respiratory syndrome coro    ....................  ....................  ....................  ....................  ....................  ....................
6449 MT607976.1 |Severe acute respiratory syndrome coro    ....................  ....................  ....................  ....................  ....................  ....................
6450 MT676412.1 |Severe acute respiratory syndrome coro    ....................  ....................  ....................  ....................  ....................  ....................
6451 MT676413.1 |Severe acute respiratory syndrome coro    ....................  ....................  ....................  ....................  ....................  ....................
6452 MT676414.1 |Severe acute respiratory syndrome coro    ....................  ....................  ....................  ....................  ....................  ....................
6453 MT676415.1 |Severe acute respiratory syndrome coro    ....................  ....................  ....................  ....................  ....................  ...............A....
6454 MT676416.1 |Severe acute respiratory syndrome coro    ....................  ....................  ....................  ....................  ....................  ....................
6455 MT676417.1 |Severe acute respiratory syndrome coro    ....................  ....................  ....................  ....................  ....................  ....................
6456 MT676418.1 |Severe acute respiratory syndrome coro    ....................  ....................  ....................  ....................  ....................  ....................
6457 MT676419.1 |Severe acute respiratory syndrome coro    ....................  ....................  ....................  ....................  ....................  ....................
6458 MT676420.1 |Severe acute respiratory syndrome coro    ....................  ....................  ....................  ....................  ....................  ....................
6459 MT676421.1 |Severe acute respiratory syndrome coro    ....................  ....................  ....................  ....................  ....................  ....................
6460 LR897977.1 |Severe acute respiratory syndrome coro    ......-.............  ....................  ....................  ....................  ....................  .......-............
6461 LR897981.1 |Severe acute respiratory syndrome coro    ......-.............  ....................  ....................  ....................  ....................  .......-............
6462 LR897983.1 |Severe acute respiratory syndrome coro    ......-.............  ....................  ....................  ....................  ....................  .......-............
6463 LR897985.1 |Severe acute respiratory syndrome coro    ......-.............  ....................  ....................  ....................  ....................  .......-............
6464 LR897986.1 |Severe acute respiratory syndrome coro    ......-.............  ....................  ....................  ....................  ....................  .......-............
6465 LR897987.1 |Severe acute respiratory syndrome coro    ......-.............  ....................  ....................  ....................  ....................  .......-............
6466 LR897989.1 |Severe acute respiratory syndrome coro    ......-.............  ....................  ....................  ....................  ....................  .......-............
6467 LR897990.1 |Severe acute respiratory syndrome coro    ......-.............  ....................  ....................  ....................  ....................  .......-............
6468 LR897991.1 |Severe acute respiratory syndrome coro    ......-.............  ....................  ....................  ....................  ....................  .......-............
6469 LR897992.1 |Severe acute respiratory syndrome coro    ......-.............  ....................  ....................  ....................  ....................  .......-............
6470 LR897993.1 |Severe acute respiratory syndrome coro    ......-.............  ....................  ....................  ....................  ....................  .......-............
6471 LR897994.1 |Severe acute respiratory syndrome coro    ......-.............  ....................  ....................  ....................  ....................  .......-............
6472 LR897995.1 |Severe acute respiratory syndrome coro    ......-.............  ....................  ....................  ....................  ....................  .......-............
6473 LR897996.1 |Severe acute respiratory syndrome coro    ......-.............  ....................  ....................  ....................  ....................  .......-............
6474 LR897997.1 |Severe acute respiratory syndrome coro    ......-.............  ....................  ....................  ....................  ....................  .......-............
6475 LR898015.1 |Severe acute respiratory syndrome coro    ......-.............  ....................  ....................  ....................  ....................  .......-............
6476 LR898016.1 |Severe acute respiratory syndrome coro    ......-.............  ....................  ....................  ....................  ....................  .......-............
6477 LR898017.1 |Severe acute respiratory syndrome coro    ......-.............  ....................  ....................  ....................  ....................  .......-............
6478 LR898018.1 |Severe acute respiratory syndrome coro    ......-.............  ....................  ....................  ....................  ....................  .......-............
6479 LR898019.1 |Severe acute respiratory syndrome coro    ......-.............  ....................  ....................  ....................  ....................  .......-............
6480 LR898020.1 |Severe acute respiratory syndrome coro    ......-.............  ....................  ....................  ....................  ....................  .......-............
6481 LR898021.1 |Severe acute respiratory syndrome coro    ......-.............  ....................  ....................  ....................  ....................  .......-............
6482 LR898022.1 |Severe acute respiratory syndrome coro    ......-.............  ....................  ....................  ....................  ....................  .......-............
6483 LR898023.1 |Severe acute respiratory syndrome coro    ......-.............  ....................  ....................  ....................  ....................  .......-............
6484 LR898037.1 |Severe acute respiratory syndrome coro    ......-.............  ....................  ....................  ....................  ....................  .......-............
6485 LR898038.1 |Severe acute respiratory syndrome coro    ......-.............  ....................  ....................  ....................  ....................  .......-............
6486 LR898039.1 |Severe acute respiratory syndrome coro    ......-.............  ....................  ....................  ....................  ....................  .......-............
6487 LR898040.1 |Severe acute respiratory syndrome coro    ......-.............  ....................  ....................  ....................  ....................  .......-............
6488 LR898041.1 |Severe acute respiratory syndrome coro    ......-.............  ....................  ....................  ....................  ....................  .......-............
6489 LR898042.1 |Severe acute respiratory syndrome coro    ......-.............  ....................  ....................  ....................  ....................  .......-............
6490 LR898043.1 |Severe acute respiratory syndrome coro    ......-.............  ....................  ....................  ....................  ....................  .......-............
6491 LR898044.1 |Severe acute respiratory syndrome coro    ......-.............  ....................  ....................  ....................  ....................  .......-............
6492 LR898045.1 |Severe acute respiratory syndrome coro    ......-.............  ....................  ....................  ....................  ....................  .......-............
6493 LR898046.1 |Severe acute respiratory syndrome coro    ......-.............  ....................  ....................  ....................  ....................  .......-............
6494 LR898047.1 |Severe acute respiratory syndrome coro    ......-.............  ....................  ....................  ....................  ....................  .......-............
6495 MW190183.1 |Severe acute respiratory syndrome coro    ....................  ....................  ....................  ....................  ....................  ....................
6496 MW190194.1 |Severe acute respiratory syndrome coro    ....................  ....................  ....................  ....................  ....................  ....................
6497 MW190195.1 |Severe acute respiratory syndrome coro    ....................  ....................  ....................  ....................  ....................  ....................
6498 MW190196.1 |Severe acute respiratory syndrome coro    ....................  ....................  ....................  ....................  ....................  ....................
6499 MW190197.1 |Severe acute respiratory syndrome coro    ....................  ....................  ....................  ....................  ....................  ....................
6500 MW190198.1 |Severe acute respiratory syndrome coro    ....................  ....................  ....................  ....................  ....................  ....................
6501 MW190199.1 |Severe acute respiratory syndrome coro    ....................  ....................  ....................  ....................  ....................  ....................
6502 MW190200.1 |Severe acute respiratory syndrome coro    ....................  ....................  ....................  ....................  ....................  ....................
6503 MW190201.1 |Severe acute respiratory syndrome coro    ....................  ....................  ....................  ....................  ....................  ....................
6504 MW190202.1 |Severe acute respiratory syndrome coro    ....................  ....................  ....................  ....................  ....................  ....................
6505 MW190203.1 |Severe acute respiratory syndrome coro    ....................  ....................  ....................  ....................  ....................  ....................
6506 MW190204.1 |Severe acute respiratory syndrome coro    ....................  ....................  ....................  ....................  ....................  ....................
6507 MW190205.1 |Severe acute respiratory syndrome coro    ....................  ....................  ....................  ....................  ....................  ....................
6508 MW190206.1 |Severe acute respiratory syndrome coro    ....................  ....................  ....................  ....................  ....................  ....................
6509 MW190207.1 |Severe acute respiratory syndrome coro    ....................  ....................  ....................  ....................  ....................  ....................
6510 MW190208.1 |Severe acute respiratory syndrome coro    ....................  ....................  ....................  ....................  ....................  ....................
6511 MW190209.1 |Severe acute respiratory syndrome coro    ....................  ....................  ....................  ....................  ....................  ....................
6512 MW190210.1 |Severe acute respiratory syndrome coro    ....................  ....................  ....................  ....................  ....................  ....................
6513 MW190211.1 |Severe acute respiratory syndrome coro    ....................  ....................  ....................  ....................  ....................  ....................
6514 MW190212.1 |Severe acute respiratory syndrome coro    ....................  ....................  ....................  ....................  ....................  ....................
```

6515 MW190213.1 |Severe acute respiratory syndrome coro ···················· ···················· ···················· ···················· ···················· ····················
6516 MW190214.1 |Severe acute respiratory syndrome coro ···················· ···················· ···················· ···················· ···················· ····················
6517 MW190215.1 |Severe acute respiratory syndrome coro ···················· ···················· ···················· ···················· ···················· ····················
6518 MW190216.1 |Severe acute respiratory syndrome coro ···················· ···················· ···················· ···················· ···················· ····················
6519 MW190217.1 |Severe acute respiratory syndrome coro ···················· ···················· ···················· ···················· ···················· ····················
6520 MW190218.1 |Severe acute respiratory syndrome coro ···················· ···················· ···················· ···················· ···················· ····················
6521 MW190219.1 |Severe acute respiratory syndrome coro ···················· ···················· ···················· ···················· ···················· ····················
6522 MW190220.1 |Severe acute respiratory syndrome coro ···················· ···················· ···················· ···················· ···················· ····················
6523 MW190221.1 |Severe acute respiratory syndrome coro ···················· ···················· ···················· ···················· ···················· ····················
6524 MW190223.1 |Severe acute respiratory syndrome coro ···················· ···················· ···················· ···················· ···················· ····················
6525 MW190224.1 |Severe acute respiratory syndrome coro ···················· ···················· ···················· ···················· ···················· ····················
6526 MW190225.1 |Severe acute respiratory syndrome coro ···················· ···················· ···················· ···················· ···················· ····················
6527 MW190226.1 |Severe acute respiratory syndrome coro ···················· ···················· ···················· ···················· ···················· ····················
6528 MW190227.1 |Severe acute respiratory syndrome coro ···················· ···················· ···················· ···················· ···················· ····················
6529 MW190228.1 |Severe acute respiratory syndrome coro ···················· ····A··············· ···················· ···················· ···················· ····················
6530 MW190229.1 |Severe acute respiratory syndrome coro ···················· ···················· ···················· ···················· ···················· ····················
6531 MW190230.1 |Severe acute respiratory syndrome coro ···················· ···················· ···················· ···················· ···················· ····················
6532 MW190231.1 |Severe acute respiratory syndrome coro ···················· ···················· ···················· ···················· ···················· ····················
6533 MW190232.1 |Severe acute respiratory syndrome coro ···················· ···················· ···················· ···················· ···················· ····················
6534 MW190233.1 |Severe acute respiratory syndrome coro ···················· ···················· ···················· ···················· ···················· ····················
6535 MW190234.1 |Severe acute respiratory syndrome coro ···················· ···················· ···················· ···················· ···················· ····················
6536 MW190235.1 |Severe acute respiratory syndrome coro ···················· ···················· ···················· ···················· ···················· ····················
6537 MW190236.1 |Severe acute respiratory syndrome coro ···················· ···················· ···················· ···················· ···················· ····················
6538 MW190237.1 |Severe acute respiratory syndrome coro ···················· ···················· ···················· ···················· ···················· ····················
6539 MW190238.1 |Severe acute respiratory syndrome coro ···················· ···················· ···················· ···················· ···················· ····················
6540 MW190239.1 |Severe acute respiratory syndrome coro ···················· ···················· ···················· ···················· ···················· ····················
6541 MW190240.1 |Severe acute respiratory syndrome coro ···················· ···················· ···················· ···················· ···················· ····················
6542 MW190241.1 |Severe acute respiratory syndrome coro ···················· ···················· ···················· ···················· ···················· ····················
6543 MW190242.1 |Severe acute respiratory syndrome coro ···················· ···················· ···················· ···················· ···················· ····················
6544 MW190243.1 |Severe acute respiratory syndrome coro ···················· ···················· ···················· ···················· ···················· ····················
6545 MW190244.1 |Severe acute respiratory syndrome coro ···················· ···················· ···················· ···················· ···················· ····················
6546 MW190245.1 |Severe acute respiratory syndrome coro ···················· ···················· ···················· ···················· ···················· ····················
6547 MW190246.1 |Severe acute respiratory syndrome coro ···················· ···················· ···················· ···················· ···················· ····················
6548 MW190247.1 |Severe acute respiratory syndrome coro ···················· ···················· ···················· ···················· ···················· ····················
6549 MW190248.1 |Severe acute respiratory syndrome coro ···················· ···················· ···················· ···················· ···················· ····················
6550 MW190249.1 |Severe acute respiratory syndrome coro ···················· ···················· ···················· ···················· ···················· ····················
6551 MW190250.1 |Severe acute respiratory syndrome coro ···················· ···················· ···················· ···················· ···················· ····················
6552 MW190251.1 |Severe acute respiratory syndrome coro ···················· ···················· ···················· ···················· ···················· ····················
6553 MW190252.1 |Severe acute respiratory syndrome coro ···················· ···················· ···················· ···················· ···················· ····················
6554 MW190253.1 |Severe acute respiratory syndrome coro ···················· ···················· ···················· ···················· ···················· ····················
6555 MW190254.1 |Severe acute respiratory syndrome coro ···················· ···················· ···················· ···················· ···················· ····················
6556 MW190255.1 |Severe acute respiratory syndrome coro ···················· ···················· ···················· ···················· ···················· ····················
6557 MW190256.1 |Severe acute respiratory syndrome coro ···T················ ···················· ···················· ···················· ···················· ····················
6558 MW190257.1 |Severe acute respiratory syndrome coro ···················· ···················· ···················· ···················· ···················· ····················
6559 MW190258.1 |Severe acute respiratory syndrome coro ···················· ···················· ···················· ···················· ···················· ····················
6560 MW190259.1 |Severe acute respiratory syndrome coro ···················· ···················· ···················· ···················· ···················· ····················
6561 MW190260.1 |Severe acute respiratory syndrome coro ···················· ···················· ···················· ···················· ···················· ····················
6562 MW190261.1 |Severe acute respiratory syndrome coro ···················· ···················· ···················· ···················· ···················· ····················
6563 MW190262.1 |Severe acute respiratory syndrome coro ···················· ···················· ···················· ···················· ···················· ····················
6564 MW190263.1 |Severe acute respiratory syndrome coro ···················· ···················· ···················· ···················· ···················· ····················
6565 MW190264.1 |Severe acute respiratory syndrome coro ···················· ···················· ···················· ···················· ···················· ····················
6566 MW190265.1 |Severe acute respiratory syndrome coro ···················· ···················· ···················· ···················· ···················· ····················
6567 MW190266.1 |Severe acute respiratory syndrome coro ···················· ···················· ···················· ···················· ···················· ····················
6568 MW190267.1 |Severe acute respiratory syndrome coro ···················· ···················· ···················· ···················· ···················· ····················
6569 MW190268.1 |Severe acute respiratory syndrome coro ···················· ···················· ···················· ···················· ···················· ····················
6570 MW190269.1 |Severe acute respiratory syndrome coro ···················· ···················· ···················· ···················· ···················· ····················
6571 MW190270.1 |Severe acute respiratory syndrome coro ···················· ···················· ···················· ···················· ···················· ····················
6572 MW190271.1 |Severe acute respiratory syndrome coro ···················· ···················· ···················· ···················· ···················· ····················
6573 MW190272.1 |Severe acute respiratory syndrome coro ···················· ···················· ···················· ···················· ···················· ····················
6574 MW190273.1 |Severe acute respiratory syndrome coro ···················· ···················· ···················· ···················· ···················· ····················
6575 MW190274.1 |Severe acute respiratory syndrome coro ···················· ···················· ···················· ···················· ···················· ····················
6576 MW190275.1 |Severe acute respiratory syndrome coro ···················· ···················· ···················· ···················· ···················· ····················
6577 MW190276.1 |Severe acute respiratory syndrome coro ···················· ···················· ···················· ···················· ···················· ····················
6578 MW190277.1 |Severe acute respiratory syndrome coro ···················· ···················· ···················· ···················· ···················· ····················
6579 MW190278.1 |Severe acute respiratory syndrome coro ···················· ···················· ···················· ···················· ···················· ····················
6580 MW190279.1 |Severe acute respiratory syndrome coro ···················· ···················· ···················· ···················· ···················· ····················
6581 MW190280.1 |Severe acute respiratory syndrome coro ···················· ···················· ···················· ···················· ···················· ····················
6582 MW190281.1 |Severe acute respiratory syndrome coro ···················· ···················· ···················· ···················· ···················· ····················
6583 MW190282.1 |Severe acute respiratory syndrome coro ···················· ···················· ···················· ···················· ···················· ····················
6584 MW190283.1 |Severe acute respiratory syndrome coro ···················· ···················· ···················· ···················· ···················· ····················
6585 MW190284.1 |Severe acute respiratory syndrome coro ···················· ···················· ···················· ···················· ···················· ····················
6586 MW190285.1 |Severe acute respiratory syndrome coro ···················· ···················· ···················· ···················· ···················· ····················
6587 MW190286.1 |Severe acute respiratory syndrome coro ···················· ···················· ···················· ···················· ···················· ····················
6588 MW190287.1 |Severe acute respiratory syndrome coro ···················· ···················· ···················· ···················· ···················· ····················
6589 MW190288.1 |Severe acute respiratory syndrome coro ···················· ···················· ···················· ···················· ···················· ····················
6590 MW190289.1 |Severe acute respiratory syndrome coro ···················· ···················· ···················· ···················· ···················· ····················
6591 MW190290.1 |Severe acute respiratory syndrome coro ···················· ···················· ···················· ···················· ···················· ····················
6592 MW190291.1 |Severe acute respiratory syndrome coro ···················· ···················· ···················· ···················· ···················· ····················

```
6593 MW190292.1 |Severe acute respiratory syndrome coro  ....................  ....................  ....................  ....................  ....................  ....................
6594 MW190293.1 |Severe acute respiratory syndrome coro  ....................  ....................  ....................  ....................  ....................  ....................
6595 MW190294.1 |Severe acute respiratory syndrome coro  ....................  ....................  ....................  ....................  ....................  ....................
6596 MW190295.1 |Severe acute respiratory syndrome coro  ....................  ....................  ....................  ....................  ....................  ....................
6597 MW190296.1 |Severe acute respiratory syndrome coro  ....................  ....................  ....................  ....................  ....................  ....................
6598 MW190297.1 |Severe acute respiratory syndrome coro  ....................  ....................  ....................  ....................  ....................  ....................
6599 MW190298.1 |Severe acute respiratory syndrome coro  ....................  ....................  ....................  ....................  ....................  ....................
6600 MW190299.1 |Severe acute respiratory syndrome coro  ....................  ....................  ....................  ....................  ....................  ....................
6601 MW190300.1 |Severe acute respiratory syndrome coro  ....................  ....................  ....................  ....................  ....................  ....................
6602 MW190301.1 |Severe acute respiratory syndrome coro  ....................  ....................  ....................  ....................  ....................  ....................
6603 MW190302.1 |Severe acute respiratory syndrome coro  ....................  ....................  ....................  ....................  ....................  ....................
6604 MW190303.1 |Severe acute respiratory syndrome coro  ....................  ....................  ....................  ....................  ....................  ....................
6605 MW190304.1 |Severe acute respiratory syndrome coro  ....................  ....................  ....................  ....................  ....................  ....................
6606 MW190305.1 |Severe acute respiratory syndrome coro  ....................  ....................  ....................  ....................  ....................  ....................
6607 MW190306.1 |Severe acute respiratory syndrome coro  ....................  ....................  ....................  ....................  ....................  ....................
6608 MW190307.1 |Severe acute respiratory syndrome coro  ....................  ....................  ....................  ....................  ....................  ....................
6609 MW190308.1 |Severe acute respiratory syndrome coro  ....................  ....................  ....................  ....................  ....................  ....................
6610 MW190309.1 |Severe acute respiratory syndrome coro  ....................  ....................  ....................  ....................  ....................  ....................
6611 MW190310.1 |Severe acute respiratory syndrome coro  ....................  ....................  ....................  ....................  ....................  ....................
6612 MW190311.1 |Severe acute respiratory syndrome coro  ....................  ....................  ....................  ....................  ....................  ....................
6613 MW190312.1 |Severe acute respiratory syndrome coro  ....................  ....................  ....................  ....................  ....................  ....................
6614 MW190313.1 |Severe acute respiratory syndrome coro  ....................  ....................  ....................  ....................  ....................  ....................
6615 MW190314.1 |Severe acute respiratory syndrome coro  ....................  ....................  ....................  ....................  ....................  ....................
6616 MW190315.1 |Severe acute respiratory syndrome coro  ....................  ....................  ....................  ....................  ....................  ....................
6617 MW190316.1 |Severe acute respiratory syndrome coro  ....................  ....................  ....................  ....................  ....................  ....................
6618 MW190317.1 |Severe acute respiratory syndrome coro  ....................  ....................  ....................  ....................  ....................  ....................
6619 MW190318.1 |Severe acute respiratory syndrome coro  ....................  ....................  ....................  ....................  ....................  ....................
6620 MW190319.1 |Severe acute respiratory syndrome coro  ....................  ....................  ....................  ....................  ....................  ....................
6621 MW190320.1 |Severe acute respiratory syndrome coro  ....................  ....................  ....................  ....................  ....................  ....................
6622 MW190321.1 |Severe acute respiratory syndrome coro  ....................  ....................  ....................  ....................  ....................  ....................
6623 MW190322.1 |Severe acute respiratory syndrome coro  ....................  ....................  ....................  ....................  ....................  ....................
6624 MW190323.1 |Severe acute respiratory syndrome coro  ....................  ....................  ....................  ....................  ....................  ....................
6625 MW190324.1 |Severe acute respiratory syndrome coro  ....................  ....................  ....................  ....................  ....................  ....................
6626 MW190325.1 |Severe acute respiratory syndrome coro  ....................  ....................  ....................  ....................  ....................  ....................
6627 MW190326.1 |Severe acute respiratory syndrome coro  ....................  ....................  ....................  ....................  ....................  ....................
6628 MW190327.1 |Severe acute respiratory syndrome coro  ....................  ....................  ....................  ....................  ....................  ....................
6629 MW190328.1 |Severe acute respiratory syndrome coro  ....................  ....................  ....................  ....................  ....................  ....................
6630 MW190329.1 |Severe acute respiratory syndrome coro  ....................  ....................  ....................  ....................  ....................  ....................
6631 MW190330.1 |Severe acute respiratory syndrome coro  ....................  ....................  ....................  ....................  ....................  ....................
6632 MW190331.1 |Severe acute respiratory syndrome coro  ....................  ....................  ....................  ....................  ....................  ....................
6633 MW190332.1 |Severe acute respiratory syndrome coro  ....................  ....................  ....................  ....................  ....................  ....................
6634 MW190333.1 |Severe acute respiratory syndrome coro  ....................  ....................  ....................  ....................  ....................  ....................
6635 MW190334.1 |Severe acute respiratory syndrome coro  ....................  ....................  ....................  ....................  ....................  ....................
6636 MW190335.1 |Severe acute respiratory syndrome coro  ....................  ....................  ....................  ....................  ....................  ....................
6637 MW190336.1 |Severe acute respiratory syndrome coro  ....................  ....................  ....................  ....................  ....................  ....................
6638 MW190337.1 |Severe acute respiratory syndrome coro  ....................  ....................  ....................  ....................  ....................  ....................
6639 MW190338.1 |Severe acute respiratory syndrome coro  ....................  ....................  ....................  ....................  ....................  ....................
6640 MW190339.1 |Severe acute respiratory syndrome coro  ....................  ....................  ....................  ....................  ....................  ....................
6641 MW190340.1 |Severe acute respiratory syndrome coro  ....................  ....................  ....................  ....................  ....................  ....................
6642 MW190341.1 |Severe acute respiratory syndrome coro  ....................  ....................  ....................  ....................  ....................  ....................
6643 MW190342.1 |Severe acute respiratory syndrome coro  ....................  ....................  ....................  ....................  ....................  ....................
6644 MW190343.1 |Severe acute respiratory syndrome coro  ....................  ....................  ....................  ....................  ....................  ....................
6645 MW190344.1 |Severe acute respiratory syndrome coro  ....................  ....................  ....................  ....................  ....................  ....................
6646 MW190345.1 |Severe acute respiratory syndrome coro  ....................  ....................  ....................  ....................  ....................  ....................
6647 MW190346.1 |Severe acute respiratory syndrome coro  ....................  ....................  ....................  ....................  ....................  ....................
6648 MW190347.1 |Severe acute respiratory syndrome coro  ....................  ....................  ....................  ....................  ....................  ....................
6649 MW190348.1 |Severe acute respiratory syndrome coro  ....................  ....................  ....................  ....................  ....................  ....................
6650 MW190349.1 |Severe acute respiratory syndrome coro  ....................  ....................  ....................  ....................  ....................  ....................
6651 MW190350.1 |Severe acute respiratory syndrome coro  ....................  ....................  ....................  ....................  ....................  ....................
6652 MW190351.1 |Severe acute respiratory syndrome coro  ....................  ....................  ....................  ....................  ....................  ....................
6653 MW190352.1 |Severe acute respiratory syndrome coro  ....................  ....................  ....................  ....................  ....................  ....................
6654 MW190353.1 |Severe acute respiratory syndrome coro  ....................  ....................  ....................  ....................  ....................  ....................
6655 MW190354.1 |Severe acute respiratory syndrome coro  ....................  ....................  ....................  ....................  ....................  ....................
6656 MW190355.1 |Severe acute respiratory syndrome coro  ....................  ....................  ....................  ....................  ....................  ....................
6657 MW190356.1 |Severe acute respiratory syndrome coro  ....................  ....................  ....................  ....................  ....................  ....................
6658 MW190357.1 |Severe acute respiratory syndrome coro  ....................  ....................  ....................  ....................  ....................  ....................
6659 MW190358.1 |Severe acute respiratory syndrome coro  ....................  ....................  ....................  ....................  ....................  ....................
6660 MW190359.1 |Severe acute respiratory syndrome coro  ....................  ....................  ....................  ....................  ....................  ....................
6661 MW190360.1 |Severe acute respiratory syndrome coro  ....................  ....................  ....................  ....................  ....................  ....................
6662 MW190361.1 |Severe acute respiratory syndrome coro  ....................  ....................  ....................  ....................  ....................  ....................
6663 MW190362.1 |Severe acute respiratory syndrome coro  ....................  ....................  ....................  ....................  ....................  ....................
6664 MW190363.1 |Severe acute respiratory syndrome coro  ....................  ....................  ....................  ....................  ....................  ....................
6665 MW190364.1 |Severe acute respiratory syndrome coro  ....................  ....................  ....................  ....................  ....................  ....................
6666 MW190365.1 |Severe acute respiratory syndrome coro  ....................  ....................  ....................  ....................  ....................  ....................
6667 MW190366.1 |Severe acute respiratory syndrome coro  ....................  ....................  ....................  ....................  ....................  ....................
6668 MW190367.1 |Severe acute respiratory syndrome coro  ....................  ....................  ....................  ....................  ....................  ....................
6669 MW190368.1 |Severe acute respiratory syndrome coro  ....................  ....................  ....................  ....................  ....................  ....................
6670 MW190369.1 |Severe acute respiratory syndrome coro  ....................  ....................  ....................  ....................  ....................  ....................
```

```
6671 MW190370.1 |Severe acute respiratory syndrome coro    ....................  ....................  ....................  ....................  ....................  ....................
6672 MW190372.1 |Severe acute respiratory syndrome coro    ....................  ....................  ....................  ....................  ....................  ....................
6673 MW190373.1 |Severe acute respiratory syndrome coro    ....................  ....................  ....................  ....................  ....................  ....................
6674 MW190374.1 |Severe acute respiratory syndrome coro    ....................  ....................  ....................  ....................  ....................  ....................
6675 MW190375.1 |Severe acute respiratory syndrome coro    ....................  ....................  ....................  ....................  ....................  ....................
6676 MW190376.1 |Severe acute respiratory syndrome coro    ....................  ....................  ....................  ....................  ....................  ....................
6677 MW190377.1 |Severe acute respiratory syndrome coro    ....................  ....................  ....................  ....................  ....................  ....................
6678 MW190378.1 |Severe acute respiratory syndrome coro    ....................  ....................  ....................  ....................  ....................  ....................
6679 MW190379.1 |Severe acute respiratory syndrome coro    ....................  ....................  ....................  ....................  ....................  ....................
6680 MW190380.1 |Severe acute respiratory syndrome coro    ....................  ....................  ....................  ....................  ....................  ....................
6681 MW190381.1 |Severe acute respiratory syndrome coro    ....................  ....................  ....................  ....................  ....................  ....................
6682 MW190382.1 |Severe acute respiratory syndrome coro    ....................  ....................  ....................  ....................  ....................  ....................
6683 MW190383.1 |Severe acute respiratory syndrome coro    ....................  ....................  ....................  ....................  ....................  ....................
6684 MW190384.1 |Severe acute respiratory syndrome coro    ....................  ....................  ....................  ....................  ....................  ....................
6685 MW190385.1 |Severe acute respiratory syndrome coro    ....................  ....................  ....................  ....................  ....................  ....................
6686 MW190386.1 |Severe acute respiratory syndrome coro    ....................  ....................  ....................  ....................  ....................  ....................
6687 MW190387.1 |Severe acute respiratory syndrome coro    ....................  ....................  ....................  ....................  ....................  ....................
6688 MW190388.1 |Severe acute respiratory syndrome coro    ....................  ....................  ....................  ....................  ....................  ....................
6689 MW190389.1 |Severe acute respiratory syndrome coro    ....................  ....................  ....................  ....................  ....................  ....................
6690 MW190390.1 |Severe acute respiratory syndrome coro    ....................  ....................  ....................  ....................  ....................  ....................
6691 MW190391.1 |Severe acute respiratory syndrome coro    ....................  ....................  ....................  ....................  ....................  ....................
6692 MW190392.1 |Severe acute respiratory syndrome coro    ....................  ....................  ....................  ....................  ....................  ....................
6693 MW190393.1 |Severe acute respiratory syndrome coro    ....................  ....................  ....................  ....................  ....................  ....................
6694 MW190394.1 |Severe acute respiratory syndrome coro    ....................  ....................  ....................  ....................  ....................  ....................
6695 MW190395.1 |Severe acute respiratory syndrome coro    ....................  ....................  ....................  ....................  ....................  ....................
6696 MW190396.1 |Severe acute respiratory syndrome coro    ....................  ....................  ....................  ....................  ....................  ....................
6697 MW190397.1 |Severe acute respiratory syndrome coro    ....................  ....................  ....................  ....................  ....................  ....................
6698 MW190398.1 |Severe acute respiratory syndrome coro    ....................  ....................  ....................  ....................  ....................  ....................
6699 MW190399.1 |Severe acute respiratory syndrome coro    ....................  ....................  ....................  ....................  ....................  ....................
6700 MW190400.1 |Severe acute respiratory syndrome coro    ....................  ....................  ....................  ....................  ....................  ....................
6701 MW190401.1 |Severe acute respiratory syndrome coro    ....................  ....................  ....................  ....................  ....................  ....................
6702 MW190402.1 |Severe acute respiratory syndrome coro    ....................  ....................  ....................  ....................  ....................  ....................
6703 MW190403.1 |Severe acute respiratory syndrome coro    ....................  ....................  ....................  ....................  ....................  ....................
6704 MW190404.1 |Severe acute respiratory syndrome coro    ....................  ....................  ....................  ....................  ....................  ....................
6705 MW190405.1 |Severe acute respiratory syndrome coro    ....................  ....................  ....................  ....................  ....................  ....................
6706 MW190406.1 |Severe acute respiratory syndrome coro    ....................  ....................  ....................  ....................  ....................  ....................
6707 MW190407.1 |Severe acute respiratory syndrome coro    ....................  ....................  ....................  ....................  ....................  ....................
6708 MW190408.1 |Severe acute respiratory syndrome coro    ....................  ....................  ....................  ....................  ....................  ....................
6709 MW190409.1 |Severe acute respiratory syndrome coro    ....................  ....................  ....................  ....................  ....................  ....................
6710 MW190410.1 |Severe acute respiratory syndrome coro    ....................  ....................  ....................  ....................  ....................  ....................
6711 MW190411.1 |Severe acute respiratory syndrome coro    ....................  ....................  ....................  ....................  ....................  ....................
6712 MW190412.1 |Severe acute respiratory syndrome coro    ....................  ....................  ....................  ....................  ....................  ....................
6713 MW190413.1 |Severe acute respiratory syndrome coro    ....................  ....................  ....................  ....................  ....................  ....................
6714 MW190414.1 |Severe acute respiratory syndrome coro    ....................  ....................  ....................  ....................  ....................  ....................
6715 MW190415.1 |Severe acute respiratory syndrome coro    ....................  ....................  ....................  ....................  ....................  ....................
6716 MW190416.1 |Severe acute respiratory syndrome coro    ....................  ....................  ....................  ....................  ....................  ....................
6717 MW190417.1 |Severe acute respiratory syndrome coro    ....................  ....................  ....................  ....................  ....................  ....................
6718 MW190418.1 |Severe acute respiratory syndrome coro    ....................  ....................  ....................  ....................  ....................  ....................
6719 MW190419.1 |Severe acute respiratory syndrome coro    ....................  ....................  ....................  ....................  ....................  ....................
6720 MW190420.1 |Severe acute respiratory syndrome coro    ....................  ....................  ....................  ....................  ....................  ....................
6721 MW190421.1 |Severe acute respiratory syndrome coro    ....................  ....................  ....................  ....................  ....................  ....................
6722 MW190422.1 |Severe acute respiratory syndrome coro    ....................  ....................  ....................  ....................  ....................  ....................
6723 MW190423.1 |Severe acute respiratory syndrome coro    ....................  ....................  ....................  ....................  ....................  ....................
6724 MW190424.1 |Severe acute respiratory syndrome coro    ....................  ....................  ....................  ....................  ....................  ....................
6725 MW190425.1 |Severe acute respiratory syndrome coro    ....................  ....................  ....................  ....................  ....................  ....................
6726 MW190426.1 |Severe acute respiratory syndrome coro    ....................  ....................  ....................  ....................  ....................  ....................
6727 MW190427.1 |Severe acute respiratory syndrome coro    ....................  ....................  ....................  ....................  ....................  ....................
6728 MW190428.1 |Severe acute respiratory syndrome coro    ....................  ....................  ....................  ....................  ....................  ....................
6729 MW190429.1 |Severe acute respiratory syndrome coro    ....................  ....................  ....................  ....................  ....................  ....................
6730 MW190430.1 |Severe acute respiratory syndrome coro    ....................  ....................  ....................  ....................  ....................  ....................
6731 MW190431.1 |Severe acute respiratory syndrome coro    ....................  ....................  ....................  ....................  ....................  ....................
6732 MW190432.1 |Severe acute respiratory syndrome coro    ....................  ....................  ....................  ....................  ....................  ....................
6733 MW190433.1 |Severe acute respiratory syndrome coro    ....................  ....................  ....................  ....................  ....................  ....................
6734 MW190434.1 |Severe acute respiratory syndrome coro    ....................  ....................  ....................  ....................  ....................  ....................
6735 MW190435.1 |Severe acute respiratory syndrome coro    ....................  ....................  ....................  ....................  ....................  ....................
6736 MW190436.1 |Severe acute respiratory syndrome coro    ....................  ....................  ....................  ....................  ....................  ....................
6737 MW190437.1 |Severe acute respiratory syndrome coro    ....................  ....................  ....................  ....................  ....................  ....................
6738 MW190438.1 |Severe acute respiratory syndrome coro    ....................  ....................  ....................  ....................  ....................  ....................
6739 MW190439.1 |Severe acute respiratory syndrome coro    ....................  ....................  ....................  ....................  ....................  ....................
6740 MW190440.1 |Severe acute respiratory syndrome coro    ....................  ....................  ....................  ....................  ....................  ....................
6741 MW190441.1 |Severe acute respiratory syndrome coro    ....................  ....................  ....................  ....................  ....................  ....................
6742 MW190442.1 |Severe acute respiratory syndrome coro    ....................  ....................  ....................  ....................  ....................  ....................
6743 MW190443.1 |Severe acute respiratory syndrome coro    ....................  ....................  ....................  ....................  ....................  ....................
6744 MW190444.1 |Severe acute respiratory syndrome coro    ....................  ....................  ....................  ....................  ....................  ....................
6745 MW190445.1 |Severe acute respiratory syndrome coro    ....................  ....................  ....................  ....................  ....................  ....................
6746 MW190446.1 |Severe acute respiratory syndrome coro    ....................  ....................  ....................  ....................  ....................  ....................
6747 MW190447.1 |Severe acute respiratory syndrome coro    ....................  ....................  ....................  ....................  ....................  ....................
6748 MW190448.1 |Severe acute respiratory syndrome coro    ....................  ....................  ....................  ....................  ....................  ....................
```

| | | | | | | | | |
|---|---|---|---|---|---|---|---|---|
| 6749 | MW190449.1 | \|Severe acute respiratory syndrome coro | ···················· | ···················· | ···················· | ···················· | ···················· | ···················· |
| 6750 | MW190450.1 | \|Severe acute respiratory syndrome coro | ···················· | ···················· | ···················· | ···················· | ···················· | ···················· |
| 6751 | MW190451.1 | \|Severe acute respiratory syndrome coro | ···················· | ···················· | ···················· | ···················· | ···················· | ···················· |
| 6752 | MW190452.1 | \|Severe acute respiratory syndrome coro | ···················· | ···················· | ···················· | ···················· | ···················· | ···················· |
| 6753 | MW190453.1 | \|Severe acute respiratory syndrome coro | ···················· | ···················· | ···················· | ···················· | ···················· | ···················· |
| 6754 | MW190454.1 | \|Severe acute respiratory syndrome coro | ···················· | ···················· | ···················· | ···················· | ···················· | ···················· |
| 6755 | MW190455.1 | \|Severe acute respiratory syndrome coro | ···················· | ···················· | ···················· | ···················· | ···················· | ···················· |
| 6756 | MW190456.1 | \|Severe acute respiratory syndrome coro | ···················· | ···················· | ···················· | ···················· | ···················· | ···················· |
| 6757 | MW190457.1 | \|Severe acute respiratory syndrome coro | ···················· | ···················· | ···················· | ···················· | ···················· | ···················· |
| 6758 | MW190458.1 | \|Severe acute respiratory syndrome coro | ···················· | ···················· | ···················· | ···················· | ···················· | ···················· |
| 6759 | MW190459.1 | \|Severe acute respiratory syndrome coro | ···················· | ···················· | ···················· | ···················· | ···················· | ···················· |
| 6760 | MW190460.1 | \|Severe acute respiratory syndrome coro | ···················· | ···················· | ···················· | ···················· | ···················· | ···················· |
| 6761 | MW190461.1 | \|Severe acute respiratory syndrome coro | ···················· | ···················· | ···················· | ···················· | ···················· | ···················· |
| 6762 | MW190462.1 | \|Severe acute respiratory syndrome coro | ···················· | ···················· | ···················· | ···················· | ···················· | ···················· |
| 6763 | MW190463.1 | \|Severe acute respiratory syndrome coro | ···················· | ···················· | ···················· | ···················· | ···················· | ···················· |
| 6764 | MW190464.1 | \|Severe acute respiratory syndrome coro | ···················· | ···················· | ···················· | ···················· | ···················· | ···················· |
| 6765 | MW190465.1 | \|Severe acute respiratory syndrome coro | ···················· | ···················· | ···················· | ···················· | ···················· | ···················· |
| 6766 | MW190466.1 | \|Severe acute respiratory syndrome coro | ···················· | ···················· | ···················· | ···················· | ···················· | ···················· |
| 6767 | MW190467.1 | \|Severe acute respiratory syndrome coro | ···················· | ···················· | ···················· | ···················· | ···················· | ···················· |
| 6768 | MW190468.1 | \|Severe acute respiratory syndrome coro | ···················· | ···················· | ···················· | ···················· | ···················· | ···················· |
| 6769 | MW190469.1 | \|Severe acute respiratory syndrome coro | ···················· | ···················· | ···················· | ···················· | ···················· | ···················· |
| 6770 | MW190470.1 | \|Severe acute respiratory syndrome coro | ···················· | ···················· | ···················· | ···················· | ···················· | ···················· |
| 6771 | MW190471.1 | \|Severe acute respiratory syndrome coro | ···················· | ···················· | ···················· | ···················· | ···················· | ···················· |
| 6772 | MW190472.1 | \|Severe acute respiratory syndrome coro | ···················· | ···················· | ···················· | ···················· | ···················· | ···················· |
| 6773 | MW190473.1 | \|Severe acute respiratory syndrome coro | ···················· | ···················· | ···················· | ···················· | ···················· | ···················· |
| 6774 | MW190474.1 | \|Severe acute respiratory syndrome coro | ···················· | ···················· | ···················· | ···················· | ···················· | ···················· |
| 6775 | MW190475.1 | \|Severe acute respiratory syndrome coro | ···················· | ···················· | ···················· | ···················· | ···················· | ···················· |
| 6776 | MW190476.1 | \|Severe acute respiratory syndrome coro | ···················· | ···················· | ···················· | ···················· | ···················· | ···················· |
| 6777 | MW190477.1 | \|Severe acute respiratory syndrome coro | ···················· | ···················· | ···················· | ···················· | ···················· | ···················· |
| 6778 | MW190478.1 | \|Severe acute respiratory syndrome coro | ···················· | ···················· | ···················· | ···················· | ···················· | ···················· |
| 6779 | MW190479.1 | \|Severe acute respiratory syndrome coro | ···················· | ···················· | ···················· | ···················· | ···················· | ···················· |
| 6780 | MW190480.1 | \|Severe acute respiratory syndrome coro | ···················· | ···················· | ···················· | ···················· | ···················· | ···················· |
| 6781 | MW190481.1 | \|Severe acute respiratory syndrome coro | ···················· | ···················· | ···················· | ···················· | ···················· | ···················· |
| 6782 | MW190482.1 | \|Severe acute respiratory syndrome coro | ···················· | ···················· | ···················· | ···················· | ···················· | ···················· |
| 6783 | MW190483.1 | \|Severe acute respiratory syndrome coro | ···················· | ···················· | ···················· | ···················· | ···················· | ···················· |
| 6784 | MW190484.1 | \|Severe acute respiratory syndrome coro | ···················· | ···················· | ···················· | ···················· | ···················· | ···················· |
| 6785 | MW190485.1 | \|Severe acute respiratory syndrome coro | ···················· | ···················· | ···················· | ···················· | ···················· | ···················· |
| 6786 | MW190486.1 | \|Severe acute respiratory syndrome coro | ···················· | ···················· | ···················· | ···················· | ···················· | ···················· |
| 6787 | MW190487.1 | \|Severe acute respiratory syndrome coro | ···················· | ···················· | ···················· | ···················· | ···················· | ···················· |
| 6788 | MW190488.1 | \|Severe acute respiratory syndrome coro | ···················· | ···················· | ···················· | ···················· | ···················· | ···················· |
| 6789 | MW190489.1 | \|Severe acute respiratory syndrome coro | ···················· | ···················· | ···················· | ···················· | ···················· | ···················· |
| 6790 | MW190490.1 | \|Severe acute respiratory syndrome coro | ···················· | ···················· | ···················· | ···················· | ···················· | ···················· |
| 6791 | MW190491.1 | \|Severe acute respiratory syndrome coro | ···················· | ···················· | ···················· | ···················· | ···················· | ···················· |
| 6792 | MW190492.1 | \|Severe acute respiratory syndrome coro | ···················· | ·········A·········· | ···················· | ···················· | ···················· | ···················· |
| 6793 | MW190493.1 | \|Severe acute respiratory syndrome coro | ···················· | ···················· | ···················· | ···················· | ···················· | ···················· |
| 6794 | MW190495.1 | \|Severe acute respiratory syndrome coro | ···················· | ···················· | ···················· | ···················· | ···················· | ···················· |
| 6795 | MW190496.1 | \|Severe acute respiratory syndrome coro | ···················· | ···················· | ···················· | ···················· | ···················· | ···················· |
| 6796 | MW190497.1 | \|Severe acute respiratory syndrome coro | ···················· | ···················· | ···················· | ···················· | ···················· | ···················· |
| 6797 | MW190498.1 | \|Severe acute respiratory syndrome coro | ···················· | ···················· | ···················· | ···················· | ···················· | ···················· |
| 6798 | MW190499.1 | \|Severe acute respiratory syndrome coro | ···················· | ···················· | ···················· | ···················· | ···················· | ···················· |
| 6799 | MW190500.1 | \|Severe acute respiratory syndrome coro | ···················· | ···················· | ···················· | ···················· | ···················· | ···················· |
| 6800 | MW190501.1 | \|Severe acute respiratory syndrome coro | ···················· | ···················· | ···················· | ···················· | ···················· | ···················· |
| 6801 | MW190502.1 | \|Severe acute respiratory syndrome coro | ···················· | ···················· | ···················· | ···················· | ···················· | ···················· |
| 6802 | MW190503.1 | \|Severe acute respiratory syndrome coro | ···················· | ···················· | ···················· | ···················· | ···················· | ···················· |
| 6803 | MW190504.1 | \|Severe acute respiratory syndrome coro | ···················· | ···················· | ···················· | ···················· | ···················· | ···················· |
| 6804 | MW190505.1 | \|Severe acute respiratory syndrome coro | ···················· | ···················· | ···················· | ···················· | ···················· | ···················· |
| 6805 | MW190506.1 | \|Severe acute respiratory syndrome coro | ···················· | ···················· | ···················· | ···················· | ···················· | ···················· |
| 6806 | MW190507.1 | \|Severe acute respiratory syndrome coro | ···················· | ···················· | ···················· | ···················· | ···················· | ···················· |
| 6807 | MW190508.1 | \|Severe acute respiratory syndrome coro | ···················· | ···················· | ···················· | ···················· | ···················· | ···················· |
| 6808 | MW190509.1 | \|Severe acute respiratory syndrome coro | ···················· | ···················· | ···················· | ···················· | ···················· | ···················· |
| 6809 | MW190510.1 | \|Severe acute respiratory syndrome coro | ···················· | ······A············· | ···················· | ···················· | ···················· | ···················· |
| 6810 | MW190511.1 | \|Severe acute respiratory syndrome coro | ···················· | ···················· | ···················· | ···················· | ···················· | ···················· |
| 6811 | MW190512.1 | \|Severe acute respiratory syndrome coro | ···················· | ···················· | ···················· | ···················· | ···················· | ···················· |
| 6812 | MW190513.1 | \|Severe acute respiratory syndrome coro | ···················· | ···················· | ···················· | ···················· | ···················· | ···················· |
| 6813 | MW190514.1 | \|Severe acute respiratory syndrome coro | ···················· | ···················· | ···················· | ···················· | ···················· | ···················· |
| 6814 | MW190516.1 | \|Severe acute respiratory syndrome coro | ···················· | ···················· | ···················· | ···················· | ···················· | ···················· |
| 6815 | MW190517.1 | \|Severe acute respiratory syndrome coro | ···················· | ···················· | ···················· | ···················· | ···················· | ···················· |
| 6816 | MW190518.1 | \|Severe acute respiratory syndrome coro | ···················· | ···················· | ···················· | ···················· | ···················· | ···················· |
| 6817 | MW190519.1 | \|Severe acute respiratory syndrome coro | ···················· | ···················· | ···················· | ···················· | ···················· | ···················· |
| 6818 | MW190520.1 | \|Severe acute respiratory syndrome coro | ···················· | ···················· | ···················· | ···················· | ···················· | ···················· |
| 6819 | MW190521.1 | \|Severe acute respiratory syndrome coro | ···················· | ···················· | ···················· | ···················· | ···················· | ···················· |
| 6820 | MW190522.1 | \|Severe acute respiratory syndrome coro | ···················· | ···················· | ···················· | ···················· | ···················· | ···················· |
| 6821 | MW190523.1 | \|Severe acute respiratory syndrome coro | ···················· | ···················· | ···················· | ···················· | ···················· | ···················· |
| 6822 | MW190524.1 | \|Severe acute respiratory syndrome coro | ···················· | ···················· | ···················· | ···················· | ···················· | ···················· |
| 6823 | MW190525.1 | \|Severe acute respiratory syndrome coro | ···················· | ···················· | ···················· | ···················· | ···················· | ···················· |
| 6824 | MW190526.1 | \|Severe acute respiratory syndrome coro | ···················· | ···················· | ···················· | ···················· | ···················· | ···················· |
| 6825 | MW190527.1 | \|Severe acute respiratory syndrome coro | ···················· | ···················· | ···················· | ···················· | ···················· | ···················· |
| 6826 | MW190528.1 | \|Severe acute respiratory syndrome coro | ···················· | ···················· | ···················· | ···················· | ···················· | ···················· |

| | | | | | | | | |
|---|---|---|---|---|---|---|---|---|
| 6827 | MW190529.1 | Severe acute respiratory syndrome coro | .................... | .................... | .................... | .................... | .................... | .................... |
| 6828 | MW190530.1 | Severe acute respiratory syndrome coro | .................... | .................... | .................... | .................... | .................... | .................... |
| 6829 | MW190531.1 | Severe acute respiratory syndrome coro | .................... | .................... | .................... | .................... | .................... | .................... |
| 6830 | MW190532.1 | Severe acute respiratory syndrome coro | .................... | .................... | .................... | .................... | .................... | .................... |
| 6831 | MW190533.1 | Severe acute respiratory syndrome coro | .................... | .................... | .................... | .................... | .................... | .................... |
| 6832 | MW190534.1 | Severe acute respiratory syndrome coro | .................... | .................... | .................... | .................... | .................... | .................... |
| 6833 | MW190535.1 | Severe acute respiratory syndrome coro | .................... | .................... | .................... | .................... | .................... | .................... |
| 6834 | MW190536.1 | Severe acute respiratory syndrome coro | .................... | .................... | .................... | .................... | .................... | .................... |
| 6835 | MW190537.1 | Severe acute respiratory syndrome coro | .................... | .................... | .................... | .................... | .................... | .................... |
| 6836 | MW190538.1 | Severe acute respiratory syndrome coro | .................... | .................... | .................... | .................... | .................... | .................... |
| 6837 | MW190539.1 | Severe acute respiratory syndrome coro | .................... | .................... | .................... | .................... | .................... | .................... |
| 6838 | MW190540.1 | Severe acute respiratory syndrome coro | .................... | .................... | .................... | .................... | .................... | .................... |
| 6839 | MW190541.1 | Severe acute respiratory syndrome coro | .................... | .................... | .................... | .................... | .................... | .................... |
| 6840 | MW190542.1 | Severe acute respiratory syndrome coro | .................... | .................... | .................... | .................... | .................... | .................... |
| 6841 | MW190543.1 | Severe acute respiratory syndrome coro | .................... | .................... | .................... | .................... | .................... | .................... |
| 6842 | MW190545.1 | Severe acute respiratory syndrome coro | .................... | .................... | .................... | .................... | .................... | .................... |
| 6843 | MW190546.1 | Severe acute respiratory syndrome coro | .................... | .................... | .................... | .................... | .................... | .................... |
| 6844 | MW190547.1 | Severe acute respiratory syndrome coro | .................... | .................... | .................... | .................... | .................... | .................... |
| 6845 | MW190548.1 | Severe acute respiratory syndrome coro | .................... | .................... | .................... | .................... | .................... | .................... |
| 6846 | MW190549.1 | Severe acute respiratory syndrome coro | .................... | .................... | .................... | .................... | .................... | .................... |
| 6847 | MW190550.1 | Severe acute respiratory syndrome coro | .................... | .................... | .................... | .................... | .................... | .................... |
| 6848 | MW190551.1 | Severe acute respiratory syndrome coro | .................... | .................... | .................... | .................... | .................... | .................... |
| 6849 | MW190552.1 | Severe acute respiratory syndrome coro | .................... | .................... | .................... | .................... | .................... | .................... |
| 6850 | MW190553.1 | Severe acute respiratory syndrome coro | .................... | .................... | .................... | .................... | .................... | .................... |
| 6851 | MW190554.1 | Severe acute respiratory syndrome coro | .................... | .................... | .................... | .................... | .................... | .................... |
| 6852 | MW190556.1 | Severe acute respiratory syndrome coro | .................... | .................... | .................... | .................... | .................... | .................... |
| 6853 | MW190557.1 | Severe acute respiratory syndrome coro | .................... | .................... | .................... | .................... | .................... | .................... |
| 6854 | MW190558.1 | Severe acute respiratory syndrome coro | .................... | .................... | .................... | .................... | .................... | .................... |
| 6855 | MW190559.1 | Severe acute respiratory syndrome coro | .................... | .................... | .................... | .................... | .................... | .................... |
| 6856 | MW190560.1 | Severe acute respiratory syndrome coro | .................... | .................... | .................... | .................... | .................... | .................... |
| 6857 | MW190561.1 | Severe acute respiratory syndrome coro | .................... | .................... | .................... | .................... | .................... | .................... |
| 6858 | MW190562.1 | Severe acute respiratory syndrome coro | .................... | .................... | .................... | .................... | .................... | .................... |
| 6859 | MW190563.1 | Severe acute respiratory syndrome coro | .................... | .................... | .................... | .................... | .................... | .................... |
| 6860 | MW190564.1 | Severe acute respiratory syndrome coro | .................... | .................... | .................... | .................... | .................... | .................... |
| 6861 | MW190565.1 | Severe acute respiratory syndrome coro | .................... | .................... | .................... | .................... | .................... | .................... |
| 6862 | MW190566.1 | Severe acute respiratory syndrome coro | .................... | .................... | .................... | .................... | .................... | .................... |
| 6863 | MW190567.1 | Severe acute respiratory syndrome coro | .................... | .................... | .................... | .................... | .................... | .................... |
| 6864 | MW190568.1 | Severe acute respiratory syndrome coro | .................... | .................... | .................... | .................... | .................... | .................... |
| 6865 | MW190569.1 | Severe acute respiratory syndrome coro | .................... | .................... | .................... | .................... | .................... | .................... |
| 6866 | MW190570.1 | Severe acute respiratory syndrome coro | .................... | .................... | .................... | .................... | .................... | .................... |
| 6867 | MW190571.1 | Severe acute respiratory syndrome coro | .................... | .................... | .................... | .................... | .................... | .................... |
| 6868 | MW190572.1 | Severe acute respiratory syndrome coro | .................... | .................... | .................... | .................... | .................... | .................... |
| 6869 | MW190573.1 | Severe acute respiratory syndrome coro | .................... | .................... | .................... | .................... | .................... | .................... |
| 6870 | MW190574.1 | Severe acute respiratory syndrome coro | .................... | .................... | .................... | .................... | .................... | .................... |
| 6871 | MW190575.1 | Severe acute respiratory syndrome coro | .................... | .................... | .................... | .................... | .................... | .................... |
| 6872 | MW190576.1 | Severe acute respiratory syndrome coro | .................... | .................... | .................... | .................... | .................... | .................... |
| 6873 | MW190577.1 | Severe acute respiratory syndrome coro | .................... | .................... | .................... | .................... | .................... | .................... |
| 6874 | MW190578.1 | Severe acute respiratory syndrome coro | .................... | .................... | .................... | .................... | .................... | .................... |
| 6875 | MW190579.1 | Severe acute respiratory syndrome coro | .................... | .................... | .................... | .................... | .................... | .................... |
| 6876 | MW190580.1 | Severe acute respiratory syndrome coro | .................... | .................... | .................... | .................... | .................... | .................... |
| 6877 | MW190581.1 | Severe acute respiratory syndrome coro | .................... | .................... | .................... | .................... | .................... | .................... |
| 6878 | MW190582.1 | Severe acute respiratory syndrome coro | .................... | .................... | .................... | .................... | .................... | .................... |
| 6879 | MW190583.1 | Severe acute respiratory syndrome coro | .................... | .................... | .................... | .................... | .................... | .................... |
| 6880 | MW190584.1 | Severe acute respiratory syndrome coro | .................... | .................... | .................... | .................... | .................... | .................... |
| 6881 | MW190585.1 | Severe acute respiratory syndrome coro | .................... | .................... | .................... | .................... | .................... | .................... |
| 6882 | MW190586.1 | Severe acute respiratory syndrome coro | .................... | .................... | .................... | .................... | .................... | .................... |
| 6883 | MW190587.1 | Severe acute respiratory syndrome coro | .................... | .................... | .................... | .................... | .................... | .................... |
| 6884 | MW190588.1 | Severe acute respiratory syndrome coro | .................... | .................... | .................... | .................... | .................... | .................... |
| 6885 | MW190589.1 | Severe acute respiratory syndrome coro | .................... | .................... | .................... | .................... | .................... | .................... |
| 6886 | MW190590.1 | Severe acute respiratory syndrome coro | .................... | .................... | .................... | .................... | .................... | .................... |
| 6887 | MW190591.1 | Severe acute respiratory syndrome coro | .................... | .................... | .................... | .................... | .................... | .................... |
| 6888 | MW190592.1 | Severe acute respiratory syndrome coro | .................... | .................... | .................... | .................... | .................... | .................... |
| 6889 | MW190593.1 | Severe acute respiratory syndrome coro | .................... | .................... | .................... | .................... | .................... | .................... |
| 6890 | MW190594.1 | Severe acute respiratory syndrome coro | .................... | .................... | .................... | .................... | .................... | .................... |
| 6891 | MW190595.1 | Severe acute respiratory syndrome coro | .................... | .................... | .................... | .................... | .................... | .................... |
| 6892 | MW190596.1 | Severe acute respiratory syndrome coro | .................... | .................... | .................... | .................... | .................... | .................... |
| 6893 | MW190597.1 | Severe acute respiratory syndrome coro | .................... | .................... | .................... | .................... | .................... | .................... |
| 6894 | MW190598.1 | Severe acute respiratory syndrome coro | .................... | .................... | .................... | .................... | .................... | .................... |
| 6895 | MW190599.1 | Severe acute respiratory syndrome coro | .................... | .................... | .................... | .................... | .................... | .................... |
| 6896 | MW190600.1 | Severe acute respiratory syndrome coro | .................... | .................... | .................... | .................... | .................... | .................... |
| 6897 | MW190601.1 | Severe acute respiratory syndrome coro | .................... | .................... | .................... | .................... | .................... | .................... |
| 6898 | MW190602.1 | Severe acute respiratory syndrome coro | .................... | .................... | .................... | .................... | .................... | .................... |
| 6899 | MW190603.1 | Severe acute respiratory syndrome coro | .................... | .................... | .................... | .................... | .................... | .................... |
| 6900 | MW190604.1 | Severe acute respiratory syndrome coro | .................... | .................... | .................... | .................... | .................... | .................... |
| 6901 | MW190605.1 | Severe acute respiratory syndrome coro | .................... | .................... | .................... | .................... | .................... | .................... |
| 6902 | MW190606.1 | Severe acute respiratory syndrome coro | .................... | .................... | .................... | .................... | .................... | .................... |
| 6903 | MW190607.1 | Severe acute respiratory syndrome coro | .................... | .................... | .................... | .................... | .................... | .................... |
| 6904 | MW190608.1 | Severe acute respiratory syndrome coro | .................... | .................... | .................... | .................... | .................... | .................... |

```
6905 MW190609.1 |Severe acute respiratory syndrome coro    ....................  ....................  ....................  ....................  ....................  ....................
6906 MW190610.1 |Severe acute respiratory syndrome coro    ....................  ....................  ....................  ....................  ....................  ....................
6907 MW190611.1 |Severe acute respiratory syndrome coro    ....................  ....................  ....................  ....................  ....................  ....................
6908 MW190612.1 |Severe acute respiratory syndrome coro    ....................  ....................  ....................  ....................  ....................  ....................
6909 MW190613.1 |Severe acute respiratory syndrome coro    ....................  ....................  ....................  ....................  ....................  ....................
6910 MW190614.1 |Severe acute respiratory syndrome coro    ....................  ....................  ....................  ....................  ....................  ....................
6911 MW190615.1 |Severe acute respiratory syndrome coro    ....................  ....................  ....................  ....................  ....................  ....................
6912 MW190616.1 |Severe acute respiratory syndrome coro    ....................  ....................  ....................  ....................  ....................  ....................
6913 MW190617.1 |Severe acute respiratory syndrome coro    ....................  ....................  ....................  ....................  ....................  ....................
6914 MW190618.1 |Severe acute respiratory syndrome coro    ....................  ....................  ....................  ....................  ....................  ....................
6915 MW190619.1 |Severe acute respiratory syndrome coro    ....................  ....................  ....................  ....................  ....................  ....................
6916 MW190620.1 |Severe acute respiratory syndrome coro    ....................  ....................  ....................  ....................  ....................  ....................
6917 MW190621.1 |Severe acute respiratory syndrome coro    ....................  ....................  ....................  ....................  ....................  ....................
6918 MW190622.1 |Severe acute respiratory syndrome coro    ....................  ....................  ....................  ....................  ....................  ....................
6919 MW190623.1 |Severe acute respiratory syndrome coro    ....................  ....................  ....................  ....................  ....................  ....................
6920 MW190624.1 |Severe acute respiratory syndrome coro    ....................  ....................  ....................  ....................  ....................  ....................
6921 MW190625.1 |Severe acute respiratory syndrome coro    ....................  ....................  ....................  ....................  ....................  ....................
6922 MW190626.1 |Severe acute respiratory syndrome coro    ....................  ....................  ....................  ....................  ....................  ....................
6923 MW190627.1 |Severe acute respiratory syndrome coro    ....................  ....................  ....................  ....................  ....................  ....................
6924 MW190628.1 |Severe acute respiratory syndrome coro    ....................  ....................  ....................  ....................  ....................  ....................
6925 MW190629.1 |Severe acute respiratory syndrome coro    ....................  ....................  ....................  ....................  ....................  ....................
6926 MW190631.1 |Severe acute respiratory syndrome coro    ....................  ....................  ....................  ....................  ....................  ....................
6927 MW190632.1 |Severe acute respiratory syndrome coro    ....................  ....................  ....................  ....................  ....................  ....................
6928 MW190633.1 |Severe acute respiratory syndrome coro    ....................  ....................  ....................  ....................  ....................  ....................
6929 MW190634.1 |Severe acute respiratory syndrome coro    ....................  ....................  ....................  ....................  ....................  ....................
6930 MW190635.1 |Severe acute respiratory syndrome coro    ....................  ....................  ....................  ....................  ....................  ....................
6931 MW190636.1 |Severe acute respiratory syndrome coro    ....................  ....................  ....................  ....................  ....................  ....................
6932 MW190637.1 |Severe acute respiratory syndrome coro    ....................  ....................  ....................  ....................  ....................  ....................
6933 MW190638.1 |Severe acute respiratory syndrome coro    ....................  ....................  ....................  ....................  ....................  ....................
6934 MW190639.1 |Severe acute respiratory syndrome coro    ....................  ....................  ....................  ....................  ....................  ....................
6935 MW190640.1 |Severe acute respiratory syndrome coro    ....................  ....................  ....................  ....................  ....................  ....................
6936 MW190641.1 |Severe acute respiratory syndrome coro    ....................  ....................  ....................  ....................  ....................  ....................
6937 MW190642.1 |Severe acute respiratory syndrome coro    ....................  ....................  ....................  ....................  ....................  ....................
6938 MW190643.1 |Severe acute respiratory syndrome coro    ....................  ....................  ....................  ....................  ....................  ....................
6939 MW190644.1 |Severe acute respiratory syndrome coro    ....................  ....................  ....................  ....................  ....................  ....................
6940 MW190646.1 |Severe acute respiratory syndrome coro    ....................  ....................  ....................  ....................  ....................  ....................
6941 MW190647.1 |Severe acute respiratory syndrome coro    ....................  ....................  ....................  ....................  ....................  ....................
6942 MW190648.1 |Severe acute respiratory syndrome coro    ....................  ....................  ....................  ....................  ....................  ....................
6943 MW190649.1 |Severe acute respiratory syndrome coro    ....................  ....................  ....................  ....................  ....................  ....................
6944 MW190650.1 |Severe acute respiratory syndrome coro    ....................  ....................  ....................  ....................  ....................  ....................
6945 MW190651.1 |Severe acute respiratory syndrome coro    ....................  ....................  ....................  ....................  ....................  ....................
6946 MW190652.1 |Severe acute respiratory syndrome coro    ....................  ....................  ....................  ....................  ....................  ....................
6947 MW190653.1 |Severe acute respiratory syndrome coro    ....................  ....................  ....................  ....................  ....................  ....................
6948 MW190654.1 |Severe acute respiratory syndrome coro    ....................  ....................  ....................  ....................  ....................  ....................
6949 MW190655.1 |Severe acute respiratory syndrome coro    ....................  ....................  ....................  ....................  ....................  ....................
6950 MW190656.1 |Severe acute respiratory syndrome coro    ....................  ....................  ....................  ....................  ....................  ....................
6951 MW190658.1 |Severe acute respiratory syndrome coro    ....................  ....................  ....................  ....................  ....................  ....................
6952 MW190659.1 |Severe acute respiratory syndrome coro    ....................  ....................  ....................  ....................  ....................  ....................
6953 MW190660.1 |Severe acute respiratory syndrome coro    ....................  ....................  ....................  ....................  ....................  ....................
6954 MW190661.1 |Severe acute respiratory syndrome coro    ....................  ....................  ....................  ....................  ....................  ....................
6955 MW190662.1 |Severe acute respiratory syndrome coro    ....................  ....................  ....................  ....................  ....................  ....................
6956 MW190663.1 |Severe acute respiratory syndrome coro    ....................  ....................  ....................  ....................  ....................  ....................
6957 MW190665.1 |Severe acute respiratory syndrome coro    ....................  ....................  ....................  ....................  ....................  ....................
6958 MW190666.1 |Severe acute respiratory syndrome coro    ....................  ....................  ....................  ....................  ....................  ....................
6959 MW190667.1 |Severe acute respiratory syndrome coro    ....................  ....................  ....................  ....................  ....................  ....................
6960 MW190668.1 |Severe acute respiratory syndrome coro    ....................  ....................  ....................  ....................  ....................  ....................
6961 MW190669.1 |Severe acute respiratory syndrome coro    ....................  ....................  ....................  ....................  ....................  ....................
6962 MW190670.1 |Severe acute respiratory syndrome coro    ....................  ....................  ....................  ....................  ....................  ....................
6963 MW190671.1 |Severe acute respiratory syndrome coro    ....................  ....................  ....................  ....................  ....................  ....................
6964 MW190672.1 |Severe acute respiratory syndrome coro    ....................  ....................  ....................  ....................  ....................  ....................
6965 MW190673.1 |Severe acute respiratory syndrome coro    ....................  ....................  ....................  ....................  ....................  ....................
6966 MW190674.1 |Severe acute respiratory syndrome coro    ....................  ....................  ....................  ....................  ....................  ....................
6967 MW190675.1 |Severe acute respiratory syndrome coro    ....................  ....................  ....................  ....................  ....................  ....................
6968 MW190676.1 |Severe acute respiratory syndrome coro    ....................  ....................  ....................  ....................  ....................  ....................
6969 MW190677.1 |Severe acute respiratory syndrome coro    ....................  ....................  ....................  ....................  ....................  ....................
6970 MW190678.1 |Severe acute respiratory syndrome coro    ....................  ....................  ....................  ....................  ....................  ....................
6971 MW190679.1 |Severe acute respiratory syndrome coro    ....................  ....................  ....................  ....................  ....................  ....................
6972 MW190680.1 |Severe acute respiratory syndrome coro    ....................  ....................  ....................  ....................  ....................  ....................
6973 MW190681.1 |Severe acute respiratory syndrome coro    ....................  ....................  ....................  ....................  ....................  ....................
6974 MW190682.1 |Severe acute respiratory syndrome coro    ....................  ....................  ....................  ....................  ....................  ....................
6975 MW190684.1 |Severe acute respiratory syndrome coro    ....................  ....................  ....................  ....................  ....................  ....................
6976 MW190685.1 |Severe acute respiratory syndrome coro    ....................  ....................  ....................  ....................  ....................  ....................
6977 MW190686.1 |Severe acute respiratory syndrome coro    ....................  ....................  ....................  ....................  ....................  ....................
6978 MW190687.1 |Severe acute respiratory syndrome coro    ....................  ....................  ....................  ....................  ....................  ....................
6979 MW190688.1 |Severe acute respiratory syndrome coro    ....................  ....................  ....................  ....................  ....................  ....................
6980 MW190690.1 |Severe acute respiratory syndrome coro    ....................  ....................  ....................  ....................  ....................  ....................
6981 MW190691.1 |Severe acute respiratory syndrome coro    ....................  ....................  ....................  ....................  ....................  ....................
6982 MW190692.1 |Severe acute respiratory syndrome coro    ....................  ....................  ....................  ....................  ....................  ....................
```

```
6983 MW190693.1 |Severe acute respiratory syndrome coro    ....................  ....................  ....................  ....................  ....................  ....................
6984 MW190694.1 |Severe acute respiratory syndrome coro    ....................  ....................  ....................  ....................  ....................  ....................
6985 MW190695.1 |Severe acute respiratory syndrome coro    ....................  ....................  ....................  ....................  ....................  ....................
6986 MW190696.1 |Severe acute respiratory syndrome coro    ....................  ....................  ....................  ....................  ....................  ....................
6987 MW190697.1 |Severe acute respiratory syndrome coro    ....................  ....................  ....................  ....................  ....................  ....................
6988 MW190698.1 |Severe acute respiratory syndrome coro    ....................  ....................  ....................  ....................  ....................  ....................
6989 MW190699.1 |Severe acute respiratory syndrome coro    ....................  ....................  ....................  ....................  ....................  ....................
6990 MW190700.1 |Severe acute respiratory syndrome coro    ....................  ....................  ....................  ....................  ....................  ....................
6991 MW190701.1 |Severe acute respiratory syndrome coro    ....................  ....................  ....................  ....................  ....................  ....................
6992 MW190702.1 |Severe acute respiratory syndrome coro    ....................  ....................  ....................  ....................  ....................  ....................
6993 MW190703.1 |Severe acute respiratory syndrome coro    ....................  ....................  ....................  ....................  ....................  ....................
6994 MW190704.1 |Severe acute respiratory syndrome coro    ....................  ....................  ....................  ....................  ....................  ....................
6995 MW190705.1 |Severe acute respiratory syndrome coro    ....................  ....................  ....................  ....................  ....................  ....................
6996 MW190706.1 |Severe acute respiratory syndrome coro    ....................  ....................  ....................  ....................  ....................  ....................
6997 MW190707.1 |Severe acute respiratory syndrome coro    ....................  ....................  ....................  ....................  ....................  ....................
6998 MW190708.1 |Severe acute respiratory syndrome coro    ....................  ....................  ....................  ....................  ....................  ....................
6999 MW190709.1 |Severe acute respiratory syndrome coro    ....................  ....................  ....................  ....................  ....................  ....................
7000 MW190710.1 |Severe acute respiratory syndrome coro    ....................  ....................  ....................  ....................  ....................  ....................
7001 MW190711.1 |Severe acute respiratory syndrome coro    ....................  ....................  ....................  ....................  ....................  ....................
7002 MW190712.1 |Severe acute respiratory syndrome coro    ....................  ....................  ....................  ....................  ....................  ....................
7003 MW190713.1 |Severe acute respiratory syndrome coro    ....................  ....................  ....................  ....................  ....................  ....................
7004 MW190714.1 |Severe acute respiratory syndrome coro    ....................  ....................  ....................  ....................  ....................  ....................
7005 MW190715.1 |Severe acute respiratory syndrome coro    ....................  ....................  ....................  ....................  ....................  ....................
7006 MW190716.1 |Severe acute respiratory syndrome coro    ....................  ....................  ....................  ....................  ....................  ....................
7007 MW190717.1 |Severe acute respiratory syndrome coro    ....................  ....................  ....................  ....................  ....................  ....................
7008 MW190718.1 |Severe acute respiratory syndrome coro    ....................  ....................  ....................  ....................  ....................  ....................
7009 MW190719.1 |Severe acute respiratory syndrome coro    ....................  ....................  ....................  ....................  ....................  ....................
7010 MW190720.1 |Severe acute respiratory syndrome coro    ....................  ....................  ....................  ....................  ....................  ....................
7011 MW190721.1 |Severe acute respiratory syndrome coro    ....................  ....................  ....................  ....................  ....................  ....................
7012 MW190722.1 |Severe acute respiratory syndrome coro    ....................  ....................  ....................  ....................  ....................  ....................
7013 MW190723.1 |Severe acute respiratory syndrome coro    ....................  ....................  ....................  ....................  ....................  ....................
7014 MW190724.1 |Severe acute respiratory syndrome coro    ....................  ....................  ....................  ....................  ....................  ....................
7015 MW190725.1 |Severe acute respiratory syndrome coro    ....................  ....................  ....................  ....................  ....................  ....................
7016 MW190726.1 |Severe acute respiratory syndrome coro    ....................  ....................  ....................  ....................  ....................  ....................
7017 MW190727.1 |Severe acute respiratory syndrome coro    ....................  ....................  ....................  ....................  ....................  ....................
7018 MW190729.1 |Severe acute respiratory syndrome coro    ....................  ....................  ....................  ....................  ....................  ....................
7019 MW190730.1 |Severe acute respiratory syndrome coro    ....................  ....................  ....................  ....................  ....................  ....................
7020 MW190731.1 |Severe acute respiratory syndrome coro    ....................  ....................  ....................  ....................  ....................  ....................
7021 MW190732.1 |Severe acute respiratory syndrome coro    ....................  ....................  ....................  ....................  ....................  ....................
7022 MW190733.1 |Severe acute respiratory syndrome coro    ....................  ....................  ....................  ....................  ....................  ....................
7023 MW190734.1 |Severe acute respiratory syndrome coro    ....................  ....................  ....................  ....................  ....................  ....................
7024 MW190735.1 |Severe acute respiratory syndrome coro    ....................  ....................  ....................  ....................  ....................  ....................
7025 MW190736.1 |Severe acute respiratory syndrome coro    ....................  ....................  ....................  ....................  ....................  ....................
7026 MW190737.1 |Severe acute respiratory syndrome coro    ....................  ....................  ....................  ....................  ....................  ....................
7027 MW190738.1 |Severe acute respiratory syndrome coro    ....................  ....................  ....................  ....................  ....................  ....................
7028 MW190739.1 |Severe acute respiratory syndrome coro    ....................  ....................  ....................  ....................  ....................  ....................
7029 MW190740.1 |Severe acute respiratory syndrome coro    ....................  ....................  ....................  ....................  ....................  ....................
7030 MW190741.1 |Severe acute respiratory syndrome coro    ....................  ....................  ....................  ....................  ....................  ....................
7031 MW190742.1 |Severe acute respiratory syndrome coro    ....................  ....................  ....................  ....................  ....................  ....................
7032 MW190743.1 |Severe acute respiratory syndrome coro    ....................  ....................  ....................  ....................  ....................  ....................
7033 MW190744.1 |Severe acute respiratory syndrome coro    ....................  ....................  ....................  ....................  ....................  ....................
7034 MW190745.1 |Severe acute respiratory syndrome coro    ....................  ....................  ....................  ....................  ....................  ....................
7035 MW190746.1 |Severe acute respiratory syndrome coro    ....................  ....................  ....................  ....................  ....................  ....................
7036 MW190747.1 |Severe acute respiratory syndrome coro    ....................  ....................  ....................  ....................  ....................  ....................
7037 MW190748.1 |Severe acute respiratory syndrome coro    ....................  ....................  ....................  ....................  ....................  ....................
7038 MW190749.1 |Severe acute respiratory syndrome coro    ....................  ....................  ....................  ....................  ....................  ....................
7039 MW190750.1 |Severe acute respiratory syndrome coro    ....................  ....................  ....................  ....................  ....................  ....................
7040 MW190751.1 |Severe acute respiratory syndrome coro    ....................  ....................  ....................  ....................  ....................  ....................
7041 MW190752.1 |Severe acute respiratory syndrome coro    ....................  ....................  ....................  ....................  ....................  ....................
7042 MW190753.1 |Severe acute respiratory syndrome coro    ....................  ....................  ....................  ....................  ....................  ....................
7043 MW190754.1 |Severe acute respiratory syndrome coro    ....................  ....................  ....................  ....................  ....................  ....................
7044 MW190755.1 |Severe acute respiratory syndrome coro    ....................  ....................  ....................  ....................  ....................  ....................
7045 MW190756.1 |Severe acute respiratory syndrome coro    ....................  ....................  ....................  ....................  ....................  ....................
7046 MW190757.1 |Severe acute respiratory syndrome coro    ....................  ....................  ....................  ....................  ....................  ....................
7047 MW190758.1 |Severe acute respiratory syndrome coro    ....................  ....................  ....................  ....................  ....................  ....................
7048 MW190759.1 |Severe acute respiratory syndrome coro    ....................  ....................  ....................  ....................  ....................  ....................
7049 MW190760.1 |Severe acute respiratory syndrome coro    ....................  ....................  ....................  ....................  ....................  ....................
7050 MW190761.1 |Severe acute respiratory syndrome coro    ....................  ....................  ....................  ....................  ....................  ....................
7051 MW190762.1 |Severe acute respiratory syndrome coro    ....................  ....................  ....................  ....................  ....................  ....................
7052 MW190763.1 |Severe acute respiratory syndrome coro    ....................  ....................  ....................  ....................  ....................  ....................
7053 MW190764.1 |Severe acute respiratory syndrome coro    ....................  ....................  ....................  ....................  ....................  ....................
7054 MW190765.1 |Severe acute respiratory syndrome coro    ....................  ....................  ....................  ....................  ....................  ....................
7055 MW190766.1 |Severe acute respiratory syndrome coro    ....................  ....................  ....................  ....................  ....................  ....................
7056 MW190767.1 |Severe acute respiratory syndrome coro    ....................  ....................  ....................  ....................  ....................  ....................
7057 MW190768.1 |Severe acute respiratory syndrome coro    ....................  ....................  ....................  ....................  ....................  ....................
7058 MW190769.1 |Severe acute respiratory syndrome coro    ....................  ....................  ....................  ....................  ....................  ....................
7059 MW190770.1 |Severe acute respiratory syndrome coro    ....................  ....................  ....................  ....................  ....................  ....................
7060 MW190771.1 |Severe acute respiratory syndrome coro    ....................  ....................  ....................  ....................  ....................  ....................
```

| | | | | | | | |
|---|---|---|---|---|---|---|---|
| 7061 | MW190772.1 \|Severe acute respiratory syndrome coro | .................... | .................... | ..................... | .................... | .................... | .................... |
| 7062 | MW190773.1 \|Severe acute respiratory syndrome coro | .................... | .................... | ..................... | .................... | .................... | .................... |
| 7063 | MW190774.1 \|Severe acute respiratory syndrome coro | .................... | .................... | ..................... | .................... | .................... | .................... |
| 7064 | MW190775.1 \|Severe acute respiratory syndrome coro | .................... | .................... | ..................... | .................... | .................... | .................... |
| 7065 | MW190776.1 \|Severe acute respiratory syndrome coro | .................... | .................... | ..................... | .................... | .................... | .................... |
| 7066 | MW190777.1 \|Severe acute respiratory syndrome coro | .................... | .................... | ..................... | .................... | .................... | .................... |
| 7067 | MW190778.1 \|Severe acute respiratory syndrome coro | .................... | .................... | ..................... | .................... | .................... | .................... |
| 7068 | MW190779.1 \|Severe acute respiratory syndrome coro | .................... | .................... | ..................... | .................... | .................... | .................... |
| 7069 | MW190780.1 \|Severe acute respiratory syndrome coro | .................... | .................... | ..................... | .................... | .................... | .................... |
| 7070 | MW190781.1 \|Severe acute respiratory syndrome coro | .................... | .................... | ..................... | .................... | .................... | .................... |
| 7071 | MW190782.1 \|Severe acute respiratory syndrome coro | .................... | .................... | ..................... | .................... | .................... | .................... |
| 7072 | MW190783.1 \|Severe acute respiratory syndrome coro | .................... | .................... | ..................... | .................... | .................... | .................... |
| 7073 | MW190784.1 \|Severe acute respiratory syndrome coro | .................... | .................... | ..................... | .................... | .................... | .................... |
| 7074 | MW190785.1 \|Severe acute respiratory syndrome coro | .................... | .................... | ..................... | .................... | .................... | .................... |
| 7075 | MW190786.1 \|Severe acute respiratory syndrome coro | .................... | .................... | ..................... | .................... | .................... | .................... |
| 7076 | MW190787.1 \|Severe acute respiratory syndrome coro | .................... | .................... | ..................... | .................... | .................... | .................... |
| 7077 | MW190788.1 \|Severe acute respiratory syndrome coro | .................... | .................... | ..................... | .................... | .................... | .................... |
| 7078 | MW190789.1 \|Severe acute respiratory syndrome coro | .................... | .................... | ..................... | .................... | .................... | .................... |
| 7079 | MW190790.1 \|Severe acute respiratory syndrome coro | .................... | .................... | ..................... | .................... | .................... | .................... |
| 7080 | MW190791.1 \|Severe acute respiratory syndrome coro | .................... | .................... | ..................... | .................... | .................... | .................... |
| 7081 | MW190792.1 \|Severe acute respiratory syndrome coro | .................... | .................... | ..................... | .................... | .................... | .................... |
| 7082 | MW190794.1 \|Severe acute respiratory syndrome coro | .................... | .................... | ..................... | .................... | .................... | .................... |
| 7083 | MW190795.1 \|Severe acute respiratory syndrome coro | .................... | .................... | ..................... | .................... | .................... | .................... |
| 7084 | MW190796.1 \|Severe acute respiratory syndrome coro | .................... | .................... | ..................... | .................... | .................... | .................... |
| 7085 | MW190798.1 \|Severe acute respiratory syndrome coro | .................... | .................... | ..................... | .................... | .................... | .................... |
| 7086 | MW190799.1 \|Severe acute respiratory syndrome coro | .................... | .................... | ..................... | .................... | .................... | .................... |
| 7087 | MW190800.1 \|Severe acute respiratory syndrome coro | .................... | .................... | ..................... | .................... | .................... | .................... |
| 7088 | MW190801.1 \|Severe acute respiratory syndrome coro | .................... | .................... | ..................... | .................... | .................... | .................... |
| 7089 | MW190803.1 \|Severe acute respiratory syndrome coro | .................... | .................... | ..................... | .................... | .................... | .................... |
| 7090 | MW190804.1 \|Severe acute respiratory syndrome coro | .................... | .................... | ..................... | .................... | .................... | .................... |
| 7091 | MW190805.1 \|Severe acute respiratory syndrome coro | .................... | .................... | ..................... | .................... | .................... | .................... |
| 7092 | MW190806.1 \|Severe acute respiratory syndrome coro | .................... | .................... | ..................... | .................... | .................... | .................... |
| 7093 | MW190807.1 \|Severe acute respiratory syndrome coro | .................... | .................... | ..................... | .................... | .................... | .................... |
| 7094 | MW190808.1 \|Severe acute respiratory syndrome coro | .................... | .................... | ..................... | .................... | .................... | .................... |
| 7095 | MW190809.1 \|Severe acute respiratory syndrome coro | .................... | .................... | ..................... | .................... | .................... | .................... |
| 7096 | MW190810.1 \|Severe acute respiratory syndrome coro | .................... | .................... | ..................... | .................... | .................... | .................... |
| 7097 | MW190811.1 \|Severe acute respiratory syndrome coro | .................... | .................... | ..................... | .................... | .................... | .................... |
| 7098 | MW190812.1 \|Severe acute respiratory syndrome coro | .................... | .................... | ..................... | .................... | .................... | .................... |
| 7099 | MW190813.1 \|Severe acute respiratory syndrome coro | .................... | .................... | ..................... | .................... | .................... | .................... |
| 7100 | MW190814.1 \|Severe acute respiratory syndrome coro | .................... | .................... | ..................... | .................... | .................... | .................... |
| 7101 | MW190815.1 \|Severe acute respiratory syndrome coro | .................... | .................... | ..................... | .................... | .................... | .................... |
| 7102 | MW190816.1 \|Severe acute respiratory syndrome coro | .................... | .................... | ..................... | .................... | .................... | .................... |
| 7103 | MW190817.1 \|Severe acute respiratory syndrome coro | .................... | .................... | ..................... | .................... | .................... | .................... |
| 7104 | MW190818.1 \|Severe acute respiratory syndrome coro | .................... | .................... | ..................... | .................... | .................... | .................... |
| 7105 | MW190819.1 \|Severe acute respiratory syndrome coro | .................... | .................... | ..................... | .................... | .................... | .................... |
| 7106 | MW190820.1 \|Severe acute respiratory syndrome coro | .................... | .................... | ..................... | .................... | .................... | .................... |
| 7107 | MW190822.1 \|Severe acute respiratory syndrome coro | .................... | .................... | ..................... | .................... | .................... | .................... |
| 7108 | MW190823.1 \|Severe acute respiratory syndrome coro | .................... | .................... | ..................... | .................... | .................... | .................... |
| 7109 | MW190824.1 \|Severe acute respiratory syndrome coro | .................... | .................... | ..................... | .................... | .................... | .................... |
| 7110 | MW190825.1 \|Severe acute respiratory syndrome coro | .................... | .................... | ..................... | .................... | .................... | .................... |
| 7111 | MW190826.1 \|Severe acute respiratory syndrome coro | .................... | .................... | ..................... | .................... | .................... | .................... |
| 7112 | MW190827.1 \|Severe acute respiratory syndrome coro | .................... | .................... | ..................... | .................... | .................... | .................... |
| 7113 | MW190828.1 \|Severe acute respiratory syndrome coro | .................... | .................... | ..................... | .................... | .................... | .................... |
| 7114 | MW190829.1 \|Severe acute respiratory syndrome coro | .................... | .................... | ..................... | .................... | .................... | .................... |
| 7115 | MW190830.1 \|Severe acute respiratory syndrome coro | .................... | .................... | ..................... | .................... | .................... | .................... |
| 7116 | MW190831.1 \|Severe acute respiratory syndrome coro | .................... | .................... | ..................... | .................... | .................... | .................... |
| 7117 | MW190832.1 \|Severe acute respiratory syndrome coro | .................... | .................... | ..................... | .................... | .................... | .................... |
| 7118 | MW190833.1 \|Severe acute respiratory syndrome coro | .................... | .................... | ..................... | .................... | .................... | .................... |
| 7119 | MW190834.1 \|Severe acute respiratory syndrome coro | .................... | .................... | ..................... | .................... | .................... | .................... |
| 7120 | MW190835.1 \|Severe acute respiratory syndrome coro | .................... | .................... | ..................... | .................... | .................... | .................... |
| 7121 | MW190837.1 \|Severe acute respiratory syndrome coro | .................... | .................... | ..................... | .................... | .................... | .................... |
| 7122 | MW190838.1 \|Severe acute respiratory syndrome coro | .................... | .................... | ..................... | .................... | .................... | .................... |
| 7123 | MW190839.1 \|Severe acute respiratory syndrome coro | .................... | .................... | ..................... | .................... | .................... | .................... |
| 7124 | MW190840.1 \|Severe acute respiratory syndrome coro | .................... | .................... | ..................... | .................... | .................... | .................... |
| 7125 | MW190841.1 \|Severe acute respiratory syndrome coro | .................... | .................... | ..................... | .................... | .................... | .................... |
| 7126 | MW190842.1 \|Severe acute respiratory syndrome coro | .................... | .................... | ..................... | .................... | .................... | .................... |
| 7127 | MW190843.1 \|Severe acute respiratory syndrome coro | .................... | .................... | ..................... | .................... | .................... | .................... |
| 7128 | MW190844.1 \|Severe acute respiratory syndrome coro | .................... | .................... | ..................... | .................... | .................... | .................... |
| 7129 | MW190845.1 \|Severe acute respiratory syndrome coro | .................... | .................... | ..................... | .................... | .................... | .................... |
| 7130 | MW190846.1 \|Severe acute respiratory syndrome coro | .................... | .................... | ..................... | .................... | .................... | .................... |
| 7131 | MW190847.1 \|Severe acute respiratory syndrome coro | .................... | .................... | ..................... | .................... | .................... | .................... |
| 7132 | MW190848.1 \|Severe acute respiratory syndrome coro | .................... | .................... | ..................... | .................... | .................... | .................... |
| 7133 | MW190849.1 \|Severe acute respiratory syndrome coro | .................... | .................... | ..................... | .................... | .................... | .................... |
| 7134 | MW190850.1 \|Severe acute respiratory syndrome coro | .................... | .................... | ..................... | .................... | .................... | .................... |
| 7135 | MW190851.1 \|Severe acute respiratory syndrome coro | .................... | .................... | ..................... | .................... | .................... | .................... |
| 7136 | MW190852.1 \|Severe acute respiratory syndrome coro | .................... | .................... | ..................... | .................... | .................... | .................... |
| 7137 | MW190853.1 \|Severe acute respiratory syndrome coro | .................... | .................... | ..................... | .................... | .................... | .................... |
| 7138 | MW190854.1 \|Severe acute respiratory syndrome coro | .................... | .................... | ..................... | .................... | .................... | .................... |

7139 MW190855.1 |Severe acute respiratory syndrome coro .................... .................... .................... .................... .................... ....................
7140 MW190856.1 |Severe acute respiratory syndrome coro .................... .................... .................... .................... .................... ....................
7141 MW190857.1 |Severe acute respiratory syndrome coro .................... .................... .................... .................... .................... ....................
7142 MW190858.1 |Severe acute respiratory syndrome coro .................... .................... .................... .................... .................... ....................
7143 MW190859.1 |Severe acute respiratory syndrome coro .................... .................... .................... .................... .................... ....................
7144 MW190860.1 |Severe acute respiratory syndrome coro .................... .................... .................... .................... .................... ....................
7145 MW190861.1 |Severe acute respiratory syndrome coro .................... .................... .................... .................... .................... ....................
7146 MW190862.1 |Severe acute respiratory syndrome coro .................... .................... .................... .................... .................... ....................
7147 MW190863.1 |Severe acute respiratory syndrome coro .................... .................... .................... .................... .................... ....................
7148 MW190864.1 |Severe acute respiratory syndrome coro .................... .................... .................... .................... .................... ....................
7149 MW190865.1 |Severe acute respiratory syndrome coro .................... .................... .................... .................... .................... ....................
7150 MW190866.1 |Severe acute respiratory syndrome coro .................... .................... .................... .................... .................... ....................
7151 MW190867.1 |Severe acute respiratory syndrome coro .................... .................... .................... .................... .................... ....................
7152 MW190868.1 |Severe acute respiratory syndrome coro .................... .................... .................... .................... .................... ....................
7153 MW190869.1 |Severe acute respiratory syndrome coro .................... .................... .................... .................... .................... ....................
7154 MW190870.1 |Severe acute respiratory syndrome coro .................... .................... .................... .................... .................... ....................
7155 MW190871.1 |Severe acute respiratory syndrome coro .................... .................... .................... .................... .................... ....................
7156 MW190872.1 |Severe acute respiratory syndrome coro .................... .................... .................... .................... .................... ....................
7157 MW190873.1 |Severe acute respiratory syndrome coro .................... .................... .................... .................... .................... ....................
7158 MW190874.1 |Severe acute respiratory syndrome coro .................... .................... .................... .................... .................... ....................
7159 MW190875.1 |Severe acute respiratory syndrome coro .................... .................... .................... .................... .................... ....................
7160 MW190876.1 |Severe acute respiratory syndrome coro .................... .................... .................... .................... .................... ....................
7161 MW190877.1 |Severe acute respiratory syndrome coro .................... .................... .................... .................... .................... ....................
7162 MW190878.1 |Severe acute respiratory syndrome coro .................... .................... .................... .................... .................... ....................
7163 MW190880.1 |Severe acute respiratory syndrome coro .................... .................... .................... .................... .................... ....................
7164 MW190881.1 |Severe acute respiratory syndrome coro .................... .................... .................... .................... .................... ....................
7165 MW190882.1 |Severe acute respiratory syndrome coro .................... .................... .................... .................... .................... ....................
7166 MW190883.1 |Severe acute respiratory syndrome coro .................... .................... .................... .................... .................... ....................
7167 MW190884.1 |Severe acute respiratory syndrome coro .................... .................... .................... .................... .................... ....................
7168 MW190885.1 |Severe acute respiratory syndrome coro .................... .................... .................... .................... .................... ....................
7169 MW190887.1 |Severe acute respiratory syndrome coro .................... .................... .................... .................... .................... ....................
7170 MW190888.1 |Severe acute respiratory syndrome coro .................... .................... .................... .................... .................... ....................
7171 MW190889.1 |Severe acute respiratory syndrome coro .................... .................... .................... .................... .................... ....................
7172 MW190890.1 |Severe acute respiratory syndrome coro .................... .................... .................... .................... .................... ....................
7173 MW190891.1 |Severe acute respiratory syndrome coro .................... .................... .................... .................... .................... ....................
7174 MW190892.1 |Severe acute respiratory syndrome coro .................... .................... .................... .................... .................... ....................
7175 MW190893.1 |Severe acute respiratory syndrome coro .................... .................... .................... .................... .................... ....................
7176 MW190894.1 |Severe acute respiratory syndrome coro .................... .................... .................... .................... .................... ....................
7177 MW190895.1 |Severe acute respiratory syndrome coro .................... .................... .................... .................... .................... ....................
7178 MW190896.1 |Severe acute respiratory syndrome coro .................... .................... .................... .................... .................... ....................
7179 MW190897.1 |Severe acute respiratory syndrome coro .................... .................... .................... .................... .................... ....................
7180 MW190898.1 |Severe acute respiratory syndrome coro .................... .................... .................... .................... .................... ....................
7181 MW190899.1 |Severe acute respiratory syndrome coro .................... .................... .................... .................... .................... ....................
7182 MW190900.1 |Severe acute respiratory syndrome coro .................... .................... .................... .................... .................... ....................
7183 MW190901.1 |Severe acute respiratory syndrome coro .................... .................... .................... .................... .................... ....................
7184 MW190902.1 |Severe acute respiratory syndrome coro .................... .................... .................... .................... .................... ....................
7185 MW190903.1 |Severe acute respiratory syndrome coro .................... .................... .................... .................... .................... ....................
7186 MW190904.1 |Severe acute respiratory syndrome coro .................... .................... .................... .................... .................... ....................
7187 MW190905.1 |Severe acute respiratory syndrome coro .................... .................... .................... .................... .................... ....................
7188 MW190906.1 |Severe acute respiratory syndrome coro .................... .................... .................... .................... .................... ....................
7189 MW190907.1 |Severe acute respiratory syndrome coro .................... .................... .................... .................... .................... ....................
7190 MW190908.1 |Severe acute respiratory syndrome coro .................... .................... .................... .................... .................... ....................
7191 MW190909.1 |Severe acute respiratory syndrome coro .................... .................... .................... .................... .................... ....................
7192 MW190910.1 |Severe acute respiratory syndrome coro .................... .................... .................... .................... .................... ....................
7193 MW190911.1 |Severe acute respiratory syndrome coro .................... .................... .................... .................... .................... ....................
7194 MW190912.1 |Severe acute respiratory syndrome coro .................... .................... .................... .................... .................... ....................
7195 MW190913.1 |Severe acute respiratory syndrome coro .................... .................... .................... .................... .................... ....................
7196 MW190914.1 |Severe acute respiratory syndrome coro .................... .................... .................... .................... .................... ....................
7197 MW190915.1 |Severe acute respiratory syndrome coro .................... .................... .................... .................... .................... ....................
7198 MW190916.1 |Severe acute respiratory syndrome coro .................... .................... .................... .................... .................... ....................
7199 MW190917.1 |Severe acute respiratory syndrome coro .................... .................... .................... .................... .................... ....................
7200 MW190918.1 |Severe acute respiratory syndrome coro .................... .................... .................... .................... .................... ....................
7201 MW190919.1 |Severe acute respiratory syndrome coro .................... .................... .................... .................... .................... ....................
7202 MW190920.1 |Severe acute respiratory syndrome coro .................... .................... .................... .................... .................... ....................
7203 MW190921.1 |Severe acute respiratory syndrome coro .................... .................... .................... .................... .................... ....................
7204 MW190922.1 |Severe acute respiratory syndrome coro .................... .................... .................... .................... .................... ....................
7205 MW190923.1 |Severe acute respiratory syndrome coro .................... .................... .................... .................... .................... ....................
7206 MW190924.1 |Severe acute respiratory syndrome coro .................... .................... .................... .................... .................... ....................
7207 MW190925.1 |Severe acute respiratory syndrome coro .................... .................... .................... .................... .................... ....................
7208 MW190926.1 |Severe acute respiratory syndrome coro .................... .................... .................... .................... .................... ....................
7209 MW190927.1 |Severe acute respiratory syndrome coro .................... .................... .................... .................... .................... ....................
7210 MW190928.1 |Severe acute respiratory syndrome coro .................... .................... .................... .................... .................... ....................
7211 MW190929.1 |Severe acute respiratory syndrome coro .................... .................... .................... .................... .................... ....................
7212 MW190930.1 |Severe acute respiratory syndrome coro .................... .................... .................... .................... .................... ....................
7213 MW190931.1 |Severe acute respiratory syndrome coro .................... .................... .................... .................... .................... ....................
7214 MW190932.1 |Severe acute respiratory syndrome coro .................... .................... .................... .................... .................... ....................
7215 MW190933.1 |Severe acute respiratory syndrome coro .................... .................... .................... .................... .................... ....................
7216 MW190934.1 |Severe acute respiratory syndrome coro .................... .................... .................... .................... .................... ....................

| | | | | | | | |
|---|---|---|---|---|---|---|---|
| 7217 | MW190935.1 \|Severe acute respiratory syndrome coro | ···················· | ···················· | ···················· | ···················· | ···················· | ···················· |
| 7218 | MW190936.1 \|Severe acute respiratory syndrome coro | ···················· | ···················· | ···················· | ···················· | ···················· | ···················· |
| 7219 | MW190937.1 \|Severe acute respiratory syndrome coro | ···················· | ···················· | ···················· | ···················· | ···················· | ···················· |
| 7220 | MW190938.1 \|Severe acute respiratory syndrome coro | ···················· | ···················· | ···················· | ···················· | ···················· | ···················· |
| 7221 | MW190940.1 \|Severe acute respiratory syndrome coro | ···················· | ···················· | ···················· | ···················· | ···················· | ···················· |
| 7222 | MW190941.1 \|Severe acute respiratory syndrome coro | ···················· | ···················· | ···················· | ···················· | ···················· | ···················· |
| 7223 | MW190942.1 \|Severe acute respiratory syndrome coro | ···················· | ···················· | ···················· | ···················· | ···················· | ···················· |
| 7224 | MW190943.1 \|Severe acute respiratory syndrome coro | ···················· | ···················· | ···················· | ···················· | ···················· | ···················· |
| 7225 | MW190944.1 \|Severe acute respiratory syndrome coro | ···················· | ···················· | ···················· | ···················· | ···················· | ···················· |
| 7226 | MW190945.1 \|Severe acute respiratory syndrome coro | ···················· | ···················· | ···················· | ···················· | ···················· | ···················· |
| 7227 | MW190946.1 \|Severe acute respiratory syndrome coro | ···················· | ···················· | ···················· | ···················· | ···················· | ···················· |
| 7228 | MW190947.1 \|Severe acute respiratory syndrome coro | ···················· | ···················· | ···················· | ···················· | ···················· | ···················· |
| 7229 | MW190948.1 \|Severe acute respiratory syndrome coro | ···················· | ···················· | ···················· | ···················· | ···················· | ···················· |
| 7230 | MW190949.1 \|Severe acute respiratory syndrome coro | ···················· | ···················· | ···················· | ···················· | ···················· | ···················· |
| 7231 | MW190950.1 \|Severe acute respiratory syndrome coro | ···················· | ···················· | ···················· | ···················· | ···················· | ···················· |
| 7232 | MW190951.1 \|Severe acute respiratory syndrome coro | ···················· | ···················· | ···················· | ···················· | ···················· | ···················· |
| 7233 | MW190952.1 \|Severe acute respiratory syndrome coro | ···················· | ···················· | ···················· | ···················· | ···················· | ···················· |
| 7234 | MW190953.1 \|Severe acute respiratory syndrome coro | ···················· | ···················· | ···················· | ···················· | ···················· | ···················· |
| 7235 | MW190954.1 \|Severe acute respiratory syndrome coro | ···················· | ···················· | ···················· | ···················· | ···················· | ···················· |
| 7236 | MW190955.1 \|Severe acute respiratory syndrome coro | ···················· | ···················· | ···················· | ···················· | ···················· | ···················· |
| 7237 | MW190956.1 \|Severe acute respiratory syndrome coro | ···················· | ···················· | ···················· | ···················· | ···················· | ···················· |
| 7238 | MW190957.1 \|Severe acute respiratory syndrome coro | ···················· | ···················· | ···················· | ···················· | ···················· | ···················· |
| 7239 | MW190958.1 \|Severe acute respiratory syndrome coro | ···················· | ···················· | ···················· | ···················· | ···················· | ···················· |
| 7240 | MW190959.1 \|Severe acute respiratory syndrome coro | ···················· | ···················· | ···················· | ···················· | ···················· | ···················· |
| 7241 | MW190960.1 \|Severe acute respiratory syndrome coro | ···················· | ···················· | ···················· | ···················· | ···················· | ···················· |
| 7242 | MW190961.1 \|Severe acute respiratory syndrome coro | ···················· | ···················· | ···················· | ···················· | ···················· | ···················· |
| 7243 | MW190962.1 \|Severe acute respiratory syndrome coro | ···················· | ···················· | ···················· | ···················· | ···················· | ···················· |
| 7244 | MW190963.1 \|Severe acute respiratory syndrome coro | ···················· | ···················· | ···················· | ···················· | ···················· | ···················· |
| 7245 | MW190964.1 \|Severe acute respiratory syndrome coro | ···················· | ···················· | ···················· | ···················· | ···················· | ···················· |
| 7246 | MW190965.1 \|Severe acute respiratory syndrome coro | ···················· | ···················· | ···················· | ···················· | ···················· | ···················· |
| 7247 | MW190966.1 \|Severe acute respiratory syndrome coro | ···················· | ···················· | ···················· | ···················· | ···················· | ···················· |
| 7248 | MW190967.1 \|Severe acute respiratory syndrome coro | ···················· | ···················· | ···················· | ···················· | ···················· | ···················· |
| 7249 | MW190968.1 \|Severe acute respiratory syndrome coro | ···················· | ···················· | ···················· | ···················· | ···················· | ···················· |
| 7250 | MW190969.1 \|Severe acute respiratory syndrome coro | ···················· | ···················· | ···················· | ···················· | ···················· | ···················· |
| 7251 | MW190970.1 \|Severe acute respiratory syndrome coro | ···················· | ···················· | ···················· | ···················· | ···················· | ···················· |
| 7252 | MW190971.1 \|Severe acute respiratory syndrome coro | ···················· | ···················· | ···················· | ···················· | ···················· | ···················· |
| 7253 | MW190972.1 \|Severe acute respiratory syndrome coro | ···················· | ···················· | ···················· | ···················· | ···················· | ···················· |
| 7254 | MW190973.1 \|Severe acute respiratory syndrome coro | ···················· | ···················· | ···················· | ···················· | ···················· | ···················· |
| 7255 | MW190974.1 \|Severe acute respiratory syndrome coro | ···················· | ···················· | ···················· | ···················· | ···················· | ···················· |
| 7256 | MW190975.1 \|Severe acute respiratory syndrome coro | ···················· | ···················· | ···················· | ···················· | ···················· | ···················· |
| 7257 | MW190976.1 \|Severe acute respiratory syndrome coro | ···················· | ···················· | ···················· | ···················· | ···················· | ···················· |
| 7258 | MW190977.1 \|Severe acute respiratory syndrome coro | ···················· | ···················· | ···················· | ···················· | ···················· | ···················· |
| 7259 | MW190978.1 \|Severe acute respiratory syndrome coro | ···················· | ···················· | ···················· | ···················· | ···················· | ···················· |
| 7260 | MW190979.1 \|Severe acute respiratory syndrome coro | ···················· | ···················· | ···················· | ···················· | ···················· | ···················· |
| 7261 | MW190980.1 \|Severe acute respiratory syndrome coro | ···················· | ···················· | ···················· | ···················· | ···················· | ···················· |
| 7262 | MW190981.1 \|Severe acute respiratory syndrome coro | ···················· | ···················· | ···················· | ···················· | ···················· | ···················· |
| 7263 | MW190982.1 \|Severe acute respiratory syndrome coro | ···················· | ···················· | ···················· | ···················· | ···················· | ···················· |
| 7264 | MW190983.1 \|Severe acute respiratory syndrome coro | ···················· | ···················· | ···················· | ···················· | ···················· | ···················· |
| 7265 | MW190984.1 \|Severe acute respiratory syndrome coro | ···················· | ···················· | ···················· | ···················· | ···················· | ···················· |
| 7266 | MW190985.1 \|Severe acute respiratory syndrome coro | ···················· | ···················· | ···················· | ···················· | ···················· | ···················· |
| 7267 | MW190986.1 \|Severe acute respiratory syndrome coro | ···················· | ···················· | ···················· | ···················· | ···················· | ···················· |
| 7268 | MW190987.1 \|Severe acute respiratory syndrome coro | ···················· | ···················· | ···················· | ···················· | ···················· | ···················· |
| 7269 | MW190988.1 \|Severe acute respiratory syndrome coro | ···················· | ···················· | ···················· | ···················· | ···················· | ···················· |
| 7270 | MW190989.1 \|Severe acute respiratory syndrome coro | ···················· | ···················· | ···················· | ···················· | ···················· | ···················· |
| 7271 | MW190990.1 \|Severe acute respiratory syndrome coro | ···················· | ···················· | ···················· | ···················· | ···················· | ···················· |
| 7272 | MW190991.1 \|Severe acute respiratory syndrome coro | ···················· | ···················· | ···················· | ···················· | ···················· | ···················· |
| 7273 | MW190992.1 \|Severe acute respiratory syndrome coro | ···················· | ···················· | ···················· | ···················· | ···················· | ···················· |
| 7274 | MW190993.1 \|Severe acute respiratory syndrome coro | ···················· | ···················· | ···················· | ···················· | ···················· | ···················· |
| 7275 | MW190994.1 \|Severe acute respiratory syndrome coro | ···················· | ···················· | ···················· | ···················· | ···················· | ···················· |
| 7276 | MW190995.1 \|Severe acute respiratory syndrome coro | ···················· | ···················· | ···················· | ···················· | ···················· | ···················· |
| 7277 | MW190996.1 \|Severe acute respiratory syndrome coro | ···················· | ···················· | ···················· | ···················· | ···················· | ···················· |
| 7278 | MW190999.1 \|Severe acute respiratory syndrome coro | ···················· | ···················· | ···················· | ···················· | ···················· | ···················· |
| 7279 | MW191000.1 \|Severe acute respiratory syndrome coro | ···················· | ···················· | ···················· | ···················· | ···················· | ···················· |
| 7280 | MW191001.1 \|Severe acute respiratory syndrome coro | ···················· | ···················· | ···················· | ···················· | ···················· | ···················· |
| 7281 | MW191002.1 \|Severe acute respiratory syndrome coro | ···················· | ···················· | ···················· | ···················· | ···················· | ···················· |
| 7282 | MW191003.1 \|Severe acute respiratory syndrome coro | ···················· | ···················· | ···················· | ···················· | ···················· | ···················· |
| 7283 | MW191004.1 \|Severe acute respiratory syndrome coro | ···················· | ···················· | ···················· | ···················· | ···················· | ···················· |
| 7284 | MW191005.1 \|Severe acute respiratory syndrome coro | ···················· | ···················· | ···················· | ···················· | ···················· | ···················· |
| 7285 | MW191006.1 \|Severe acute respiratory syndrome coro | ···················· | ···················· | ···················· | ···················· | ···················· | ···················· |
| 7286 | MW191007.1 \|Severe acute respiratory syndrome coro | ···················· | ···················· | ···················· | ···················· | ···················· | ···················· |
| 7287 | MW191008.1 \|Severe acute respiratory syndrome coro | ···················· | ···················· | ···················· | ···················· | ······A············· | ···················· |
| 7288 | MW191009.1 \|Severe acute respiratory syndrome coro | ···················· | ···················· | ···················· | ···················· | ···················· | ···················· |
| 7289 | MW191010.1 \|Severe acute respiratory syndrome coro | ···················· | ···················· | ···················· | ···················· | ···················· | ···················· |
| 7290 | MW191011.1 \|Severe acute respiratory syndrome coro | ···················· | ···················· | ···················· | ···················· | ···················· | ···················· |
| 7291 | MW191012.1 \|Severe acute respiratory syndrome coro | ···················· | ···················· | ···················· | ···················· | ···················· | ···················· |
| 7292 | MW191013.1 \|Severe acute respiratory syndrome coro | ···················· | ···················· | ···················· | ···················· | ···················· | ···················· |
| 7293 | MW191014.1 \|Severe acute respiratory syndrome coro | ···················· | ···················· | ···················· | ···················· | ···················· | ···················· |
| 7294 | MW191015.1 \|Severe acute respiratory syndrome coro | ···················· | ···················· | ···················· | ···················· | ···················· | ···················· |

```
7295 MW191016.1 |Severe acute respiratory syndrome coro    ....................  ....................  .....................  ....................  ....................  ....................
7296 MW191017.1 |Severe acute respiratory syndrome coro    ....................  ....................  .....................  ....................  ....................  ....................
7297 MW191018.1 |Severe acute respiratory syndrome coro    ....................  ....................  .....................  ....................  ....................  ....................
7298 MW191019.1 |Severe acute respiratory syndrome coro    ....................  ....................  .....................  ....................  ....................  ....................
7299 MW191020.1 |Severe acute respiratory syndrome coro    ....................  ....................  .....................  ....................  ....................  ....................
7300 MW191021.1 |Severe acute respiratory syndrome coro    ....................  ....................  .....................  ....................  ....................  ....................
7301 MW191022.1 |Severe acute respiratory syndrome coro    ....................  ....................  .....................  ....................  ....................  ....................
7302 MW191023.1 |Severe acute respiratory syndrome coro    ....................  ....................  .....................  ....................  ....................  ....................
7303 MW191025.1 |Severe acute respiratory syndrome coro    ....................  ....................  .....................  ....................  ....................  ....................
7304 MW191026.1 |Severe acute respiratory syndrome coro    ....................  ....................  .....................  ....................  ....................  ....................
7305 MW191027.1 |Severe acute respiratory syndrome coro    ....................  ....................  .....................  ....................  ....................  ....................
7306 MW191028.1 |Severe acute respiratory syndrome coro    ....................  ....................  .....................  ....................  ....................  ....................
7307 MW191029.1 |Severe acute respiratory syndrome coro    ....................  ....................  .....................  ....................  ....................  ....................
7308 MW191030.1 |Severe acute respiratory syndrome coro    ....................  ....................  .....................  ....................  ....................  ....................
7309 MW191032.1 |Severe acute respiratory syndrome coro    ....................  ....................  .....................  ....................  ....................  ....................
7310 MW191033.1 |Severe acute respiratory syndrome coro    ....................  ....................  .....................  ....................  ....................  ....................
7311 MW191034.1 |Severe acute respiratory syndrome coro    ....................  ....................  .....................  ....................  ....................  ....................
7312 MW191035.1 |Severe acute respiratory syndrome coro    ....................  ....................  .....................  ....................  ....................  ....................
7313 MW191036.1 |Severe acute respiratory syndrome coro    ....................  ....................  .....................  ....................  ....................  ....................
7314 MW191037.1 |Severe acute respiratory syndrome coro    ....................  ....................  .....................  ....................  ....................  ....................
7315 MW191038.1 |Severe acute respiratory syndrome coro    ....................  ....................  .....................  ....................  ....................  ....................
7316 MW191039.1 |Severe acute respiratory syndrome coro    ....................  ....................  .....................  ....................  ....................  ....................
7317 MW191040.1 |Severe acute respiratory syndrome coro    ....................  ....................  .....................  ....................  ....................  ....................
7318 MW191041.1 |Severe acute respiratory syndrome coro    ....................  ....................  .....................  ....................  ....................  ....................
7319 MW191042.1 |Severe acute respiratory syndrome coro    ....................  ....................  .....................  ....................  ....................  ....................
7320 MW191043.1 |Severe acute respiratory syndrome coro    ....................  ....................  .....................  ....................  ....................  ....................
7321 MW191044.1 |Severe acute respiratory syndrome coro    ....................  ....................  .....................  ....................  ....................  ....................
7322 MW191045.1 |Severe acute respiratory syndrome coro    ....................  ....................  .....................  ....................  ....................  ....................
7323 MW191046.1 |Severe acute respiratory syndrome coro    ....................  ....................  .....................  ....................  ....................  ....................
7324 MW191047.1 |Severe acute respiratory syndrome coro    ....................  ....................  .....................  ....................  ....................  ....................
7325 MW191048.1 |Severe acute respiratory syndrome coro    ....................  ....................  .....................  ....................  ....................  ....................
7326 MW191049.1 |Severe acute respiratory syndrome coro    ....................  ....................  .....................  ....................  ....................  ....................
7327 MW191050.1 |Severe acute respiratory syndrome coro    ....................  ....................  .....................  ....................  ....................  ....................
7328 MW191051.1 |Severe acute respiratory syndrome coro    ....................  ....................  .....................  ....................  ....................  ....................
7329 MW191052.1 |Severe acute respiratory syndrome coro    ....................  ....................  .....................  ....................  ....................  ....................
7330 MW191053.1 |Severe acute respiratory syndrome coro    ....................  ....................  .....................  ....................  ....................  ....................
7331 MW191054.1 |Severe acute respiratory syndrome coro    ....................  ....................  .....................  ....................  ....................  ....................
7332 MW191055.1 |Severe acute respiratory syndrome coro    ....................  ....................  .....................  ....................  ....................  ....................
7333 MW191056.1 |Severe acute respiratory syndrome coro    ....................  ....................  .....................  ....................  ....................  ....................
7334 MW191057.1 |Severe acute respiratory syndrome coro    ....................  ....................  .....................  ....................  ....................  ....................
7335 MW191058.1 |Severe acute respiratory syndrome coro    ....................  ....................  .....................  ....................  ....................  ....................
7336 MW191059.1 |Severe acute respiratory syndrome coro    ....................  ....................  .....................  ....................  ....................  ....................
7337 MW191060.1 |Severe acute respiratory syndrome coro    ....................  ....................  .....................  ....................  ....................  ....................
7338 MW191061.1 |Severe acute respiratory syndrome coro    ....................  ....................  .....................  ....................  ....................  ....................
7339 MW191062.1 |Severe acute respiratory syndrome coro    ....................  ....................  .....................  ....................  ....................  ....................
7340 MW191063.1 |Severe acute respiratory syndrome coro    ....................  ....................  .....................  ....................  ....................  ....................
7341 MW191065.1 |Severe acute respiratory syndrome coro    ....................  ....................  .....................  ....................  ....................  ....................
7342 MW191066.1 |Severe acute respiratory syndrome coro    ....................  ....................  .....................  ....................  ....................  ....................
7343 MW191067.1 |Severe acute respiratory syndrome coro    ....................  ....................  .....................  ....................  ....................  ....................
7344 MW191068.1 |Severe acute respiratory syndrome coro    ....................  ....................  .....................  ....................  ....................  ....................
7345 MW191069.1 |Severe acute respiratory syndrome coro    ....................  ....................  .....................  ....................  ....................  ....................
7346 MW191070.1 |Severe acute respiratory syndrome coro    ....................  ....................  .....................  ....................  ....................  ....................
7347 MW191071.1 |Severe acute respiratory syndrome coro    ....................  ....................  .....................  ....................  ....................  ....................
7348 MW191072.1 |Severe acute respiratory syndrome coro    ....................  ....................  .....................  ....................  ....................  ....................
7349 MW191073.1 |Severe acute respiratory syndrome coro    ....................  ....................  .....................  ....................  ....................  ....................
7350 MW191074.1 |Severe acute respiratory syndrome coro    ....................  ....................  .....................  ....................  ....................  ....................
7351 MW191075.1 |Severe acute respiratory syndrome coro    ....................  ....................  .....................  ....................  ....................  ....................
7352 MW191076.1 |Severe acute respiratory syndrome coro    ....................  ....................  .....................  ....................  ....................  ....................
7353 MW191077.1 |Severe acute respiratory syndrome coro    ....................  ....................  .....................  ....................  ....................  ....................
7354 MW191078.1 |Severe acute respiratory syndrome coro    ....................  ....................  .....................  ....................  ....................  ....................
7355 MW191079.1 |Severe acute respiratory syndrome coro    ....................  ....................  .....................  ....................  ....................  ....................
7356 MW191080.1 |Severe acute respiratory syndrome coro    ....................  ....................  .....................  ....................  ....................  ....................
7357 MW191081.1 |Severe acute respiratory syndrome coro    ....................  ....................  .....................  ....................  ....................  ....................
7358 MW191082.1 |Severe acute respiratory syndrome coro    ....................  ....................  .....................  ....................  ....................  ....................
7359 MW191083.1 |Severe acute respiratory syndrome coro    ....................  ....................  .....................  ....................  ....................  ....................
7360 MW191084.1 |Severe acute respiratory syndrome coro    ....................  ....................  .....................  ....................  ....................  ....................
7361 MW191085.1 |Severe acute respiratory syndrome coro    ....................  ....................  .....................  ....................  ....................  ....................
7362 MW191086.1 |Severe acute respiratory syndrome coro    ....................  ....................  .....................  ....................  ....................  ....................
7363 MW191087.1 |Severe acute respiratory syndrome coro    ....................  ....................  .....................  ....................  ....................  ....................
7364 MW191088.1 |Severe acute respiratory syndrome coro    ....................  ....................  .....................  ....................  ....................  ....................
7365 MW191089.1 |Severe acute respiratory syndrome coro    ....................  ....................  .....................  ....................  ....................  ....................
7366 MW191090.1 |Severe acute respiratory syndrome coro    ....................  ....................  .....................  ....................  ....................  ....................
7367 MW191091.1 |Severe acute respiratory syndrome coro    ....................  ....................  .....................  ....................  ....................  ....................
7368 MW191092.1 |Severe acute respiratory syndrome coro    ....................  ....................  .....................  ....................  ....................  ....................
7369 MW191093.1 |Severe acute respiratory syndrome coro    ....................  ....................  .....................  ....................  ....................  ....................
7370 MW191094.1 |Severe acute respiratory syndrome coro    ....................  ....................  .....................  ....................  ....................  ....................
7371 MW191095.1 |Severe acute respiratory syndrome coro    ....................  ....................  .....................  ....................  ....................  ....................
7372 MW191096.1 |Severe acute respiratory syndrome coro    ....................  ....................  .....................  ....................  ....................  ....................
```

| | | | | | | | | |
|---|---|---|---|---|---|---|---|---|
| 7373 | MW191097.1 | \|Severe acute respiratory syndrome coro | ···················· | ···················· | ···················· | ···················· | ···················· | ···················· |
| 7374 | MW191098.1 | \|Severe acute respiratory syndrome coro | ···················· | ···················· | ···················· | ···················· | ···················· | ···················· |
| 7375 | MW191099.1 | \|Severe acute respiratory syndrome coro | ···················· | ···················· | ···················· | ···················· | ···················· | ···················· |
| 7376 | MW191101.1 | \|Severe acute respiratory syndrome coro | ···················· | ···················· | ···················· | ···················· | ···················· | ···················· |
| 7377 | MW191102.1 | \|Severe acute respiratory syndrome coro | ···················· | ···················· | ···················· | ···················· | ···················· | ···················· |
| 7378 | MW191103.1 | \|Severe acute respiratory syndrome coro | ···················· | ···················· | ···················· | ···················· | ···················· | ···················· |
| 7379 | MW191104.1 | \|Severe acute respiratory syndrome coro | ···················· | ···················· | ···················· | ···················· | ···················· | ···················· |
| 7380 | MW191105.1 | \|Severe acute respiratory syndrome coro | ···················· | ···················· | ···················· | ···················· | ···················· | ···················· |
| 7381 | MW191106.1 | \|Severe acute respiratory syndrome coro | ···················· | ···················· | ···················· | ···················· | ···················· | ···················· |
| 7382 | MW191107.1 | \|Severe acute respiratory syndrome coro | ···················· | ···················· | ···················· | ···················· | ···················· | ···················· |
| 7383 | MW191108.1 | \|Severe acute respiratory syndrome coro | ···················· | ···················· | ···················· | ···················· | ···················· | ···················· |
| 7384 | MW191109.1 | \|Severe acute respiratory syndrome coro | ···················· | ···················· | ···················· | ···················· | ···················· | ···················· |
| 7385 | MW191110.1 | \|Severe acute respiratory syndrome coro | ···················· | ···················· | ···················· | ···················· | ···················· | ···················· |
| 7386 | MW191111.1 | \|Severe acute respiratory syndrome coro | ···················· | ···················· | ···················· | ···················· | ···················· | ···················· |
| 7387 | MW191112.1 | \|Severe acute respiratory syndrome coro | ···················· | ···················· | ···················· | ···················· | ···················· | ···················· |
| 7388 | MW191113.1 | \|Severe acute respiratory syndrome coro | ···················· | ···················· | ···················· | ···················· | ···················· | ···················· |
| 7389 | MW191114.1 | \|Severe acute respiratory syndrome coro | ···················· | ···················· | ···················· | ···················· | ···················· | ···················· |
| 7390 | MW191115.1 | \|Severe acute respiratory syndrome coro | ···················· | ···················· | ···················· | ···················· | ···················· | ···················· |
| 7391 | MW191116.1 | \|Severe acute respiratory syndrome coro | ···················· | ···················· | ···················· | ···················· | ···················· | ···················· |
| 7392 | MW191117.1 | \|Severe acute respiratory syndrome coro | ···················· | ···················· | ···················· | ···················· | ···················· | ···················· |
| 7393 | MW191118.1 | \|Severe acute respiratory syndrome coro | ···················· | ···················· | ···················· | ···················· | ···················· | ···················· |
| 7394 | MW191119.1 | \|Severe acute respiratory syndrome coro | ···················· | ···················· | ···················· | ···················· | ···················· | ···················· |
| 7395 | MW191120.1 | \|Severe acute respiratory syndrome coro | ···················· | ···················· | ···················· | ···················· | ···················· | ···················· |
| 7396 | MW191121.1 | \|Severe acute respiratory syndrome coro | ···················· | ···················· | ···················· | ···················· | ···················· | ···················· |
| 7397 | MW191122.1 | \|Severe acute respiratory syndrome coro | ···················· | ···················· | ···················· | ···················· | ···················· | ···················· |
| 7398 | MW191123.1 | \|Severe acute respiratory syndrome coro | ···················· | ···················· | ···················· | ···················· | ···················· | ···················· |
| 7399 | MW191124.1 | \|Severe acute respiratory syndrome coro | ···················· | ···················· | ···················· | ···················· | ···················· | ···················· |
| 7400 | MW191125.1 | \|Severe acute respiratory syndrome coro | ···················· | ···················· | ···················· | ···················· | ···················· | ···················· |
| 7401 | MW191126.1 | \|Severe acute respiratory syndrome coro | ···················· | ···················· | ···················· | ···················· | ···················· | ···················· |
| 7402 | MW191127.1 | \|Severe acute respiratory syndrome coro | ···················· | ···················· | ···················· | ···················· | ···················· | ···················· |
| 7403 | MW191128.1 | \|Severe acute respiratory syndrome coro | ···················· | ···················· | ···················· | ···················· | ···················· | ···················· |
| 7404 | MW191129.1 | \|Severe acute respiratory syndrome coro | ···················· | ···················· | ···················· | ···················· | ···················· | ···················· |
| 7405 | MW191130.1 | \|Severe acute respiratory syndrome coro | ···················· | ···················· | ···················· | ···················· | ···················· | ···················· |
| 7406 | MW191132.1 | \|Severe acute respiratory syndrome coro | ···················· | ···················· | ···················· | ···················· | ···················· | ···················· |
| 7407 | MW191134.1 | \|Severe acute respiratory syndrome coro | ···················· | ···················· | ···················· | ···················· | ···················· | ···················· |
| 7408 | MW191135.1 | \|Severe acute respiratory syndrome coro | ···················· | ···················· | ···················· | ···················· | ···················· | ···················· |
| 7409 | MW191136.1 | \|Severe acute respiratory syndrome coro | ···················· | ···················· | ···················· | ···················· | ···················· | ···················· |
| 7410 | MW191137.1 | \|Severe acute respiratory syndrome coro | ···················· | ···················· | ···················· | ···················· | ···················· | ···················· |
| 7411 | MW191138.1 | \|Severe acute respiratory syndrome coro | ···················· | ···················· | ···················· | ···················· | ···················· | ···················· |
| 7412 | MW191139.1 | \|Severe acute respiratory syndrome coro | ···················· | ···················· | ···················· | ···················· | ···················· | ···················· |
| 7413 | MW191140.1 | \|Severe acute respiratory syndrome coro | ···················· | ···················· | ···················· | ···················· | ···················· | ···················· |
| 7414 | MW191141.1 | \|Severe acute respiratory syndrome coro | ···················· | ···················· | ···················· | ···················· | ···················· | ···················· |
| 7415 | MW191142.1 | \|Severe acute respiratory syndrome coro | ···················· | ···················· | ···················· | ···················· | ···················· | ···················· |
| 7416 | MW191143.1 | \|Severe acute respiratory syndrome coro | ···················· | ···················· | ···················· | ···················· | ···················· | ···················· |
| 7417 | MW191144.1 | \|Severe acute respiratory syndrome coro | ···················· | ···················· | ···················· | ···················· | ···················· | ···················· |
| 7418 | MW191145.1 | \|Severe acute respiratory syndrome coro | ···················· | ···················· | ···················· | ···················· | ···················· | ···················· |
| 7419 | MW191146.1 | \|Severe acute respiratory syndrome coro | ···················· | ···················· | ···················· | ···················· | ···················· | ···················· |
| 7420 | MW191147.1 | \|Severe acute respiratory syndrome coro | ···················· | ···················· | ···················· | ···················· | ···················· | ···················· |
| 7421 | MW191148.1 | \|Severe acute respiratory syndrome coro | ···················· | ···················· | ···················· | ···················· | ···················· | ···················· |
| 7422 | MW191149.1 | \|Severe acute respiratory syndrome coro | ···················· | ···················· | ···················· | ···················· | ···················· | ···················· |
| 7423 | MW191150.1 | \|Severe acute respiratory syndrome coro | ···················· | ···················· | ···················· | ···················· | ···················· | ···················· |
| 7424 | MW191151.1 | \|Severe acute respiratory syndrome coro | ···················· | ···················· | ···················· | ···················· | ···················· | ···················· |
| 7425 | MW191152.1 | \|Severe acute respiratory syndrome coro | ···················· | ···················· | ···················· | ···················· | ···················· | ···················· |
| 7426 | MW191153.1 | \|Severe acute respiratory syndrome coro | ···················· | ···················· | ···················· | ···················· | ···················· | ···················· |
| 7427 | MW191154.1 | \|Severe acute respiratory syndrome coro | ···················· | ···················· | ···················· | ···················· | ···················· | ···················· |
| 7428 | MW191155.1 | \|Severe acute respiratory syndrome coro | ···················· | ···················· | ···················· | ···················· | ···················· | ···················· |
| 7429 | MW191156.1 | \|Severe acute respiratory syndrome coro | ···················· | ···················· | ···················· | ···················· | ···················· | ···················· |
| 7430 | MW191157.1 | \|Severe acute respiratory syndrome coro | ···················· | ···················· | ···················· | ···················· | ···················· | ···················· |
| 7431 | MW191158.1 | \|Severe acute respiratory syndrome coro | ···················· | ···················· | ···················· | ···················· | ···················· | ···················· |
| 7432 | MW191159.1 | \|Severe acute respiratory syndrome coro | ···················· | ···················· | ···················· | ···················· | ···················· | ···················· |
| 7433 | MW191160.1 | \|Severe acute respiratory syndrome coro | ···················· | ···················· | ···················· | ···················· | ···················· | ···················· |
| 7434 | MW191161.1 | \|Severe acute respiratory syndrome coro | ···················· | ···················· | ···················· | ···················· | ···················· | ···················· |
| 7435 | MW191163.1 | \|Severe acute respiratory syndrome coro | ···················· | ···················· | ···················· | ···················· | ···················· | ···················· |
| 7436 | MW191164.1 | \|Severe acute respiratory syndrome coro | ···················· | ···················· | ···················· | ···················· | ···················· | ···················· |
| 7437 | MW191165.1 | \|Severe acute respiratory syndrome coro | ···················· | ···················· | ···················· | ···················· | ···················· | ···················· |
| 7438 | MW191166.1 | \|Severe acute respiratory syndrome coro | ···················· | ···················· | ···················· | ···················· | ···················· | ···················· |
| 7439 | MW191167.1 | \|Severe acute respiratory syndrome coro | ···················· | ···················· | ···················· | ···················· | ···················· | ···················· |
| 7440 | MW191168.1 | \|Severe acute respiratory syndrome coro | ···················· | ···················· | ···················· | ···················· | ···················· | ···················· |
| 7441 | MW191169.1 | \|Severe acute respiratory syndrome coro | ···················· | ···················· | ···················· | ···················· | ···················· | ···················· |
| 7442 | MW191170.1 | \|Severe acute respiratory syndrome coro | ···················· | ···················· | ···················· | ···················· | ···················· | ···················· |
| 7443 | MW191171.1 | \|Severe acute respiratory syndrome coro | ···················· | ···················· | ···················· | ···················· | ···················· | ···················· |
| 7444 | MW191172.1 | \|Severe acute respiratory syndrome coro | ···················· | ·············A······ | ···················· | ···················· | ···················· | ···················· |
| 7445 | MW191173.1 | \|Severe acute respiratory syndrome coro | ···················· | ···················· | ···················· | ···················· | ···················· | ···················· |
| 7446 | MW191174.1 | \|Severe acute respiratory syndrome coro | ···················· | ···················· | ···················· | ···················· | ···················· | ···················· |
| 7447 | MW191175.1 | \|Severe acute respiratory syndrome coro | ···················· | ···················· | ···················· | ···················· | ···················· | ···················· |
| 7448 | MW191176.1 | \|Severe acute respiratory syndrome coro | ···················· | ···················· | ···················· | ···················· | ···················· | ···················· |
| 7449 | MW191177.1 | \|Severe acute respiratory syndrome coro | ···················· | ···················· | ···················· | ···················· | ···················· | ···················· |
| 7450 | MW191178.1 | \|Severe acute respiratory syndrome coro | ···················· | ···················· | ···················· | ···················· | ···················· | ···················· |

| | | | | | | | | |
|---|---|---|---|---|---|---|---|---|
| 7451 | MW191179.1 | Severe acute respiratory syndrome coro | .................... | .................... | .................... | .................... | .................... | .................... |
| 7452 | MW191180.1 | Severe acute respiratory syndrome coro | .................... | .................... | .................... | .................... | .................... | .................... |
| 7453 | MW191181.1 | Severe acute respiratory syndrome coro | .................... | .................... | .................... | .................... | .................... | .................... |
| 7454 | MW191182.1 | Severe acute respiratory syndrome coro | .................... | .................... | .................... | .................... | .................... | .................... |
| 7455 | MW191183.1 | Severe acute respiratory syndrome coro | .................... | .................... | .................... | .................... | .................... | .................... |
| 7456 | MW191184.1 | Severe acute respiratory syndrome coro | .................... | .................... | .................... | .................... | .................... | .................... |
| 7457 | MW191185.1 | Severe acute respiratory syndrome coro | .................... | .................... | .................... | .................... | .................... | .................... |
| 7458 | MW191186.1 | Severe acute respiratory syndrome coro | .................... | .................... | .................... | .................... | .................... | .................... |
| 7459 | MW191187.1 | Severe acute respiratory syndrome coro | .................... | .................... | .................... | .................... | .................... | .................... |
| 7460 | MW191188.1 | Severe acute respiratory syndrome coro | .................... | .................... | .................... | .................... | .................... | .................... |
| 7461 | MW191189.1 | Severe acute respiratory syndrome coro | .................... | .................... | .................... | .................... | .................... | .................... |
| 7462 | MW191190.1 | Severe acute respiratory syndrome coro | .................... | .................... | .................... | .................... | .................... | .................... |
| 7463 | MW191191.1 | Severe acute respiratory syndrome coro | .................... | .................... | .................... | .................... | .................... | .................... |
| 7464 | MW191192.1 | Severe acute respiratory syndrome coro | .................... | .................... | .................... | .................... | .................... | .................... |
| 7465 | MW191193.1 | Severe acute respiratory syndrome coro | .................... | .................... | .................... | .................... | .................... | .................... |
| 7466 | MW191194.1 | Severe acute respiratory syndrome coro | .................... | .................... | .................... | .................... | .................... | .................... |
| 7467 | MW191195.1 | Severe acute respiratory syndrome coro | .................... | .................... | .................... | .................... | .................... | .................... |
| 7468 | MW191196.1 | Severe acute respiratory syndrome coro | .................... | .................... | .................... | .................... | .................... | .................... |
| 7469 | MW191197.1 | Severe acute respiratory syndrome coro | .................... | .................... | .................... | .................... | .................... | .................... |
| 7470 | MW191198.1 | Severe acute respiratory syndrome coro | .................... | .................... | .................... | .................... | .................... | .................... |
| 7471 | MW191199.1 | Severe acute respiratory syndrome coro | .................... | .................... | .................... | .................... | .................... | .................... |
| 7472 | MW191200.1 | Severe acute respiratory syndrome coro | .................... | .................... | .................... | .................... | .................... | .................... |
| 7473 | MW191201.1 | Severe acute respiratory syndrome coro | .................... | .................... | .................... | .................... | .................... | .................... |
| 7474 | MW191202.1 | Severe acute respiratory syndrome coro | .................... | .................... | .................... | .................... | .................... | .................... |
| 7475 | MW191203.1 | Severe acute respiratory syndrome coro | .................... | .................... | .................... | .................... | .................... | .................... |
| 7476 | MW191204.1 | Severe acute respiratory syndrome coro | .................... | .................... | .................... | .................... | .................... | .................... |
| 7477 | MW191205.1 | Severe acute respiratory syndrome coro | .................... | .................... | .................... | .................... | .................... | .................... |
| 7478 | MW191206.1 | Severe acute respiratory syndrome coro | .................... | .................... | .................... | .................... | .................... | .................... |
| 7479 | MW191207.1 | Severe acute respiratory syndrome coro | .................... | .................... | .................... | .................... | .................... | .................... |
| 7480 | MW191208.1 | Severe acute respiratory syndrome coro | .................... | .................... | .................... | .................... | .................... | .................... |
| 7481 | MW191209.1 | Severe acute respiratory syndrome coro | .................... | .................... | .................... | .................... | .................... | .................... |
| 7482 | MW191210.1 | Severe acute respiratory syndrome coro | .................... | .................... | .................... | .................... | .................... | .................... |
| 7483 | MW191211.1 | Severe acute respiratory syndrome coro | .................... | .................... | .................... | .................... | .................... | .................... |
| 7484 | MW191212.1 | Severe acute respiratory syndrome coro | .................... | .................... | .................... | .................... | .................... | .................... |
| 7485 | MW191213.1 | Severe acute respiratory syndrome coro | .................... | .................... | .................... | .................... | .................... | .................... |
| 7486 | MW191214.1 | Severe acute respiratory syndrome coro | .................... | .................... | .................... | .................... | .................... | .................... |
| 7487 | MW191215.1 | Severe acute respiratory syndrome coro | .................... | .................... | .................... | .................... | .................... | .................... |
| 7488 | MW191216.1 | Severe acute respiratory syndrome coro | .................... | .................... | .................... | .................... | .................... | .................... |
| 7489 | MW191217.1 | Severe acute respiratory syndrome coro | .................... | .................... | .................... | .................... | .................... | .................... |
| 7490 | MW191218.1 | Severe acute respiratory syndrome coro | .................... | .................... | .................... | .................... | .................... | .................... |
| 7491 | MW191219.1 | Severe acute respiratory syndrome coro | .................... | .................... | .................... | .................... | .................... | .................... |
| 7492 | MW191220.1 | Severe acute respiratory syndrome coro | .................... | .................... | .................... | .................... | .................... | .................... |
| 7493 | MW191221.1 | Severe acute respiratory syndrome coro | .................... | .................... | .................... | .................... | .................... | .................... |
| 7494 | MW191222.1 | Severe acute respiratory syndrome coro | .................... | .................... | .................... | .................... | .................... | .................... |
| 7495 | MW191223.1 | Severe acute respiratory syndrome coro | .................... | .................... | .................... | .................... | .................... | .................... |
| 7496 | MW191224.1 | Severe acute respiratory syndrome coro | .................... | .................... | .................... | .................... | .................... | .................... |
| 7497 | MW191225.1 | Severe acute respiratory syndrome coro | .................... | .................... | .................... | .................... | .................... | .................... |
| 7498 | MW191226.1 | Severe acute respiratory syndrome coro | .................... | .................... | .................... | .................... | .................... | .................... |
| 7499 | MW191227.1 | Severe acute respiratory syndrome coro | .................... | .................... | .................... | .................... | .................... | .................... |
| 7500 | MW191228.1 | Severe acute respiratory syndrome coro | .................... | .................... | .................... | .................... | .................... | .................... |
| 7501 | MW191229.1 | Severe acute respiratory syndrome coro | .................... | .................... | .................... | .................... | .................... | .................... |
| 7502 | MW191230.1 | Severe acute respiratory syndrome coro | .................... | .................... | .................... | .................... | .................... | .................... |
| 7503 | MW191231.1 | Severe acute respiratory syndrome coro | .................... | .................... | .................... | .................... | .................... | .................... |
| 7504 | MW191232.1 | Severe acute respiratory syndrome coro | .................... | .................... | .................... | .................... | .................... | .................... |
| 7505 | MW191233.1 | Severe acute respiratory syndrome coro | .................... | .................... | .................... | .................... | .................... | .................... |
| 7506 | MW191234.1 | Severe acute respiratory syndrome coro | .................... | .................... | .................... | .................... | .................... | .................... |
| 7507 | MW191235.1 | Severe acute respiratory syndrome coro | .................... | .................... | .................... | .................... | .................... | .................... |
| 7508 | MW191236.1 | Severe acute respiratory syndrome coro | .................... | .................... | .................... | .................... | .................... | .................... |
| 7509 | MW191237.1 | Severe acute respiratory syndrome coro | .................... | .................... | .................... | .................... | .................... | .................... |
| 7510 | MW191238.1 | Severe acute respiratory syndrome coro | .................... | .................... | .................... | .................... | .................... | .................... |
| 7511 | MW191239.1 | Severe acute respiratory syndrome coro | .................... | .................... | .................... | .................... | .................... | .................... |
| 7512 | MW191240.1 | Severe acute respiratory syndrome coro | .................... | .................... | .................... | .................... | .................... | .................... |
| 7513 | MW191241.1 | Severe acute respiratory syndrome coro | .................... | .................... | .................... | .................... | .................... | .................... |
| 7514 | MW191242.1 | Severe acute respiratory syndrome coro | .................... | .................... | .................... | .................... | .................... | .................... |
| 7515 | MW191243.1 | Severe acute respiratory syndrome coro | .................... | .................... | .................... | .................... | .................... | .................... |
| 7516 | MW191244.1 | Severe acute respiratory syndrome coro | .................... | .................... | .................... | .................... | .................... | .................... |
| 7517 | MW191245.1 | Severe acute respiratory syndrome coro | .................... | .................... | .................... | .................... | .................... | .................... |
| 7518 | MW191246.1 | Severe acute respiratory syndrome coro | .................... | .................... | .................... | .................... | .................... | .................... |
| 7519 | MW191247.1 | Severe acute respiratory syndrome coro | .................... | .................... | .................... | .................... | .................... | .................... |
| 7520 | MW191248.1 | Severe acute respiratory syndrome coro | .................... | .................... | .................... | .................... | .................... | .................... |
| 7521 | MW191249.1 | Severe acute respiratory syndrome coro | .................... | .................... | .................... | .................... | .................... | .................... |
| 7522 | MW191250.1 | Severe acute respiratory syndrome coro | .................... | .................... | .................... | .................... | .................... | .................... |
| 7523 | MW191251.1 | Severe acute respiratory syndrome coro | .................... | .................... | .................... | .................... | .................... | .................... |
| 7524 | MW191252.1 | Severe acute respiratory syndrome coro | .................... | .................... | .................... | .................... | .................... | .................... |
| 7525 | MW191253.1 | Severe acute respiratory syndrome coro | .................... | .................... | .................... | .................... | .................... | .................... |
| 7526 | MW191255.1 | Severe acute respiratory syndrome coro | .................... | .................... | .................... | .................... | .................... | .................... |
| 7527 | MW191256.1 | Severe acute respiratory syndrome coro | .................... | .................... | .................... | .................... | .................... | .................... |
| 7528 | MW191257.1 | Severe acute respiratory syndrome coro | .................... | .................... | .................... | .................... | .................... | .................... |

```
7529 MW191258.1 |Severe acute respiratory syndrome coro    ....................  ....................  ....................  ....................  ....................  ....................
7530 MW191259.1 |Severe acute respiratory syndrome coro    ....................  ....................  ....................  ....................  ....................  ....................
7531 MW191260.1 |Severe acute respiratory syndrome coro    ....................  ....................  ....................  ....................  ....................  ....................
7532 MW191261.1 |Severe acute respiratory syndrome coro    ....................  ....................  ....................  ....................  ....................  ....................
7533 MW191262.1 |Severe acute respiratory syndrome coro    ....................  ....................  ....................  ....................  ....................  ....................
7534 MW191263.1 |Severe acute respiratory syndrome coro    ....................  ....................  ....................  ....................  ....................  ....................
7535 MW191264.1 |Severe acute respiratory syndrome coro    ....................  ....................  ....................  ....................  ....................  ....................
7536 MW191265.1 |Severe acute respiratory syndrome coro    ....................  ....................  ....................  ....................  ....................  ....................
7537 MW191266.1 |Severe acute respiratory syndrome coro    ....................  ....................  ....................  ....................  ....................  ....................
7538 MW191267.1 |Severe acute respiratory syndrome coro    ....................  ....................  ....................  ....................  ....................  ....................
7539 MW191268.1 |Severe acute respiratory syndrome coro    ....................  ....................  ....................  ....................  ....................  ....................
7540 MW191269.1 |Severe acute respiratory syndrome coro    ....................  ....................  ....................  ....................  ....................  ....................
7541 MW191270.1 |Severe acute respiratory syndrome coro    ....................  ....................  ....................  ....................  ....................  ....................
7542 MW191271.1 |Severe acute respiratory syndrome coro    ....................  ....................  ....................  ....................  ....................  ....................
7543 MW191272.1 |Severe acute respiratory syndrome coro    ....................  ....................  ....................  ....................  ....................  ....................
7544 MW191273.1 |Severe acute respiratory syndrome coro    ....................  ....................  ....................  ....................  ....................  ....................
7545 MW191274.1 |Severe acute respiratory syndrome coro    ....................  ....................  ....................  ....................  ....................  ....................
7546 MW191275.1 |Severe acute respiratory syndrome coro    ....................  ....................  ....................  ....................  ....................  ....................
7547 MW191276.1 |Severe acute respiratory syndrome coro    ....................  ....................  ....................  ....................  ....................  ....................
7548 MW191277.1 |Severe acute respiratory syndrome coro    ....................  ....................  ....................  ....................  ....................  ....................
7549 MW191278.1 |Severe acute respiratory syndrome coro    ....................  ....................  ....................  ....................  ....................  ....................
7550 MW191279.1 |Severe acute respiratory syndrome coro    ....................  ....................  ....................  ....................  ....................  ....................
7551 MW191280.1 |Severe acute respiratory syndrome coro    ....................  ....................  ....................  ....................  ....................  ....................
7552 MW191281.1 |Severe acute respiratory syndrome coro    ....................  ....................  ....................  ....................  ....................  ....................
7553 MW191282.1 |Severe acute respiratory syndrome coro    ....................  ....................  ....................  ....................  ....................  ....................
7554 MW191283.1 |Severe acute respiratory syndrome coro    ....................  ....................  ....................  ....................  ....................  ....................
7555 MW191284.1 |Severe acute respiratory syndrome coro    ....................  ....................  ....................  ....................  ....................  ....................
7556 MW191285.1 |Severe acute respiratory syndrome coro    ....................  ....................  ....................  ....................  ....................  ....................
7557 MW191286.1 |Severe acute respiratory syndrome coro    ....................  ....................  ....................  ....................  ....................  ....................
7558 MW191287.1 |Severe acute respiratory syndrome coro    ....................  ....................  ....................  ....................  ....................  ....................
7559 MW191288.1 |Severe acute respiratory syndrome coro    ....................  ....................  ....................  ....................  ....................  ....................
7560 MW191289.1 |Severe acute respiratory syndrome coro    ....................  ....................  ....................  ....................  ....................  ....................
7561 MW191290.1 |Severe acute respiratory syndrome coro    ....................  ....................  ....................  ....................  ....................  ....................
7562 MW191292.1 |Severe acute respiratory syndrome coro    ....................  ....................  ....................  ....................  ....................  ....................
7563 MW191293.1 |Severe acute respiratory syndrome coro    ....................  ....................  ....................  ....................  ....................  ....................
7564 MW191294.1 |Severe acute respiratory syndrome coro    ....................  ....................  ....................  ....................  ....................  ....................
7565 MW191295.1 |Severe acute respiratory syndrome coro    ....................  ....................  ....................  ....................  ....................  ....................
7566 MW191296.1 |Severe acute respiratory syndrome coro    ....................  ....................  ....................  ....................  ....................  ....................
7567 MW191297.1 |Severe acute respiratory syndrome coro    ....................  ....................  ....................  ....................  ....................  ....................
7568 MW191298.1 |Severe acute respiratory syndrome coro    ....................  ....................  ....................  ....................  ....................  ....................
7569 MW191299.1 |Severe acute respiratory syndrome coro    ....................  ....................  ....................  ....................  ....................  ....................
7570 MW191300.1 |Severe acute respiratory syndrome coro    ....................  ....................  ....................  ....................  ....................  ....................
7571 MW191301.1 |Severe acute respiratory syndrome coro    ....................  ....................  ....................  ....................  ....................  ....................
7572 MW191302.1 |Severe acute respiratory syndrome coro    ....................  ....................  ....................  ....................  ....................  ....................
7573 MW191303.1 |Severe acute respiratory syndrome coro    ....................  ....................  ....................  ....................  ....................  ....................
7574 MW191304.1 |Severe acute respiratory syndrome coro    ....................  ....................  ....................  ....................  ....................  ....................
7575 MW191305.1 |Severe acute respiratory syndrome coro    ....................  ....................  ....................  ....................  ....................  ....................
7576 MW191306.1 |Severe acute respiratory syndrome coro    ....................  ....................  ....................  ....................  ....................  ....................
7577 MW191307.1 |Severe acute respiratory syndrome coro    ....................  ....................  ....................  ....................  ....................  ....................
7578 MW191308.1 |Severe acute respiratory syndrome coro    ....................  ....................  ....................  ....................  ....................  ....................
7579 MW191309.1 |Severe acute respiratory syndrome coro    ....................  ....................  ....................  ....................  ....................  ....................
7580 MW191310.1 |Severe acute respiratory syndrome coro    ....................  ....................  ....................  ....................  ....................  ....................
7581 MW191311.1 |Severe acute respiratory syndrome coro    ....................  ....................  ....................  ....................  ....................  ....................
7582 MW191312.1 |Severe acute respiratory syndrome coro    ....................  ....................  ....................  ....................  ....................  ....................
7583 MW191313.1 |Severe acute respiratory syndrome coro    ....................  ....................  ....................  ....................  ....................  ....................
7584 MW191314.1 |Severe acute respiratory syndrome coro    ....................  ....................  ....................  ....................  ....................  ....................
7585 MW191315.1 |Severe acute respiratory syndrome coro    ....................  ....................  ....................  ....................  ....................  ....................
7586 MW191316.1 |Severe acute respiratory syndrome coro    ....................  ....................  ....................  ....................  ....................  ....................
7587 MW191317.1 |Severe acute respiratory syndrome coro    ....................  ....................  ....................  ....................  ....................  ....................
7588 MW191318.1 |Severe acute respiratory syndrome coro    ....................  ....................  ....................  ....................  ....................  ....................
7589 MW191319.1 |Severe acute respiratory syndrome coro    ....................  ....................  ....................  ....................  ....................  ....................
7590 MW191320.1 |Severe acute respiratory syndrome coro    ....................  ....................  ....................  ....................  ....................  ....................
7591 MW191321.1 |Severe acute respiratory syndrome coro    ....................  ....................  ....................  ....................  ....................  ....................
7592 MW191322.1 |Severe acute respiratory syndrome coro    ....................  ....................  ....................  ....................  ....................  ....................
7593 MW191323.1 |Severe acute respiratory syndrome coro    ....................  ....................  ....................  ....................  ....................  ....................
7594 MW191324.1 |Severe acute respiratory syndrome coro    ....................  ....................  ....................  ....................  ....................  ....................
7595 MW191325.1 |Severe acute respiratory syndrome coro    ....................  ....................  ....................  ....................  ....................  ....................
7596 MW191326.1 |Severe acute respiratory syndrome coro    ....................  ....................  ....................  ....................  ....................  ....................
7597 MW191327.1 |Severe acute respiratory syndrome coro    ....................  ....................  ....................  ....................  ....................  ....................
7598 MW191328.1 |Severe acute respiratory syndrome coro    ....................  ....................  ....................  ....................  ....................  ....................
7599 MW191329.1 |Severe acute respiratory syndrome coro    ....................  ....................  ....................  ....................  ....................  ....................
7600 MW191330.1 |Severe acute respiratory syndrome coro    ....................  ....................  ....................  ....................  ....................  ....................
7601 MW191331.1 |Severe acute respiratory syndrome coro    ....................  ....................  ....................  ....................  ....................  ....................
7602 MW191332.1 |Severe acute respiratory syndrome coro    ....................  ....................  ....................  ....................  ....................  ....................
7603 MW191333.1 |Severe acute respiratory syndrome coro    ....................  ....................  ....................  ....................  ....................  ....................
7604 MW191334.1 |Severe acute respiratory syndrome coro    ....................  ....................  ....................  ....................  ....................  ....................
7605 MW191335.1 |Severe acute respiratory syndrome coro    ....................  ....................  ....................  ....................  ....................  ....................
7606 MW191336.1 |Severe acute respiratory syndrome coro    ....................  ....................  ....................  ....................  ....................  ....................
```

```
7607 MW191337.1 |Severe acute respiratory syndrome coro    .................... .................... .................... .................... .................... ....................
7608 MW191338.1 |Severe acute respiratory syndrome coro    .................... .................... .................... .................... .................... ....................
7609 MW191339.1 |Severe acute respiratory syndrome coro    .................... .................... .................... .................... .................... ....................
7610 MW191340.1 |Severe acute respiratory syndrome coro    .................... .................... .................... .................... .................... ....................
7611 MW191341.1 |Severe acute respiratory syndrome coro    .................... .................... .................... .................... .................... ....................
7612 MW191342.1 |Severe acute respiratory syndrome coro    .................... .................... .................... .................... .................... ....................
7613 MW191343.1 |Severe acute respiratory syndrome coro    .................... .................... .................... .................... .................... ....................
7614 MW191344.1 |Severe acute respiratory syndrome coro    .................... .................... .................... .................... .................... ....................
7615 MW191345.1 |Severe acute respiratory syndrome coro    .................... .................... .................... .................... .................... ....................
7616 MW191346.1 |Severe acute respiratory syndrome coro    .................... .................... .................... .................... .................... ....................
7617 MW191347.1 |Severe acute respiratory syndrome coro    .................... .................... .................... .................... .................... ....................
7618 MW191348.1 |Severe acute respiratory syndrome coro    .................... .................... .................... .................... .................... ....................
7619 MW191349.1 |Severe acute respiratory syndrome coro    .................... .................... .................... .................... .................... ....................
7620 MW191350.1 |Severe acute respiratory syndrome coro    .................... .................... .................... .................... .................... ....................
7621 MW191351.1 |Severe acute respiratory syndrome coro    .................... .................... .................... .................... .................... ....................
7622 MW191352.1 |Severe acute respiratory syndrome coro    .................... .................... .................... .................... .................... ....................
7623 MW191353.1 |Severe acute respiratory syndrome coro    .................... .................... .................... .................... .................... ....................
7624 MW191354.1 |Severe acute respiratory syndrome coro    .................... .................... .................... .................... .................... ....................
7625 MW191355.1 |Severe acute respiratory syndrome coro    .................... .................... .................... .................... .................... ....................
7626 MW191356.1 |Severe acute respiratory syndrome coro    .................... .................... .................... .................... .................... ....................
7627 MW191357.1 |Severe acute respiratory syndrome coro    .................... .................... .................... .................... .................... ....................
7628 MW191358.1 |Severe acute respiratory syndrome coro    .................... .................... .................... .................... .................... ....................
7629 MW191359.1 |Severe acute respiratory syndrome coro    .................... .................... .................... .................... .................... ....................
7630 MW191360.1 |Severe acute respiratory syndrome coro    .................... .................... .................... .................... .................... ....................
7631 MW191361.1 |Severe acute respiratory syndrome coro    .................... .................... .................... .................... .................... ....................
7632 MW191362.1 |Severe acute respiratory syndrome coro    .................... .................... .................... .................... .................... ....................
7633 MW191363.1 |Severe acute respiratory syndrome coro    .................... .................... .................... .................... .................... ....................
7634 MW191364.1 |Severe acute respiratory syndrome coro    .................... .................... .................... .................... .................... ....................
7635 MW191365.1 |Severe acute respiratory syndrome coro    .................... .................... .................... .................... .................... ....................
7636 MW191366.1 |Severe acute respiratory syndrome coro    .................... .................... .................... .................... .................... ....................
7637 MW191367.1 |Severe acute respiratory syndrome coro    .................... .................... .................... .................... .................... ....................
7638 MW191368.1 |Severe acute respiratory syndrome coro    .................... .................... .................... .................... .................... ....................
7639 MW191369.1 |Severe acute respiratory syndrome coro    .................... .................... .................... .................... .................... ....................
7640 MW191370.1 |Severe acute respiratory syndrome coro    .................... .................... .................... .................... .................... ....................
7641 MW191371.1 |Severe acute respiratory syndrome coro    .................... .................... .................... .................... .................... ....................
7642 MW191372.1 |Severe acute respiratory syndrome coro    .................... .................... .................... .................... .................... ....................
7643 MW191373.1 |Severe acute respiratory syndrome coro    .................... .................... .................... .................... .................... ....................
7644 MW191374.1 |Severe acute respiratory syndrome coro    .................... .................... .................... .................... .................... ....................
7645 MW191375.1 |Severe acute respiratory syndrome coro    .................... .................... .................... .................... .................... ....................
7646 MW191376.1 |Severe acute respiratory syndrome coro    .................... .................... .................... .................... .................... ....................
7647 MW191377.1 |Severe acute respiratory syndrome coro    .................... .................... .................... .................... .................... ....................
7648 MW191378.1 |Severe acute respiratory syndrome coro    .................... .................... .................... .................... .................... ....................
7649 MW191379.1 |Severe acute respiratory syndrome coro    .................... .................... .................... .................... .................... ....................
7650 MW191380.1 |Severe acute respiratory syndrome coro    .................... .................... .................... .................... .................... ....................
7651 MW191381.1 |Severe acute respiratory syndrome coro    .................... .................... .................... .................... .................... ....................
7652 MW191382.1 |Severe acute respiratory syndrome coro    .................... .................... .................... .................... .................... ....................
7653 MW191383.1 |Severe acute respiratory syndrome coro    .................... .................... .................... .................... .................... ....................
7654 MW191384.1 |Severe acute respiratory syndrome coro    .................... .................... .................... .................... .................... ....................
7655 MW191385.1 |Severe acute respiratory syndrome coro    .................... .................... .................... .................... .................... ....................
7656 MW191386.1 |Severe acute respiratory syndrome coro    .................... .................... .................... .................... .................... ....................
7657 MW191387.1 |Severe acute respiratory syndrome coro    .................... .................... .................... .................... .................... ....................
7658 MW191388.1 |Severe acute respiratory syndrome coro    .................... .................... .................... .................... .................... ....................
7659 MW191389.1 |Severe acute respiratory syndrome coro    .................... .................... .................... .................... .................... ....................
7660 MW191390.1 |Severe acute respiratory syndrome coro    .................... .................... .................... .................... .................... ....................
7661 MW191391.1 |Severe acute respiratory syndrome coro    .................... .................... .................... .................... .................... ....................
7662 MW191392.1 |Severe acute respiratory syndrome coro    .................... .................... .................... .................... .................... ....................
7663 MW191393.1 |Severe acute respiratory syndrome coro    .................... .................... .................... .................... .................... ....................
7664 MW191394.1 |Severe acute respiratory syndrome coro    .................... .................... .................... .................... .................... ....................
7665 MW191395.1 |Severe acute respiratory syndrome coro    .................... .................... .................... .................... .................... ....................
7666 MW191396.1 |Severe acute respiratory syndrome coro    .................... .................... .................... .................... .................... ....................
7667 MW191397.1 |Severe acute respiratory syndrome coro    .................... .................... .................... .................... .................... ....................
7668 MW191398.1 |Severe acute respiratory syndrome coro    .................... .................... .................... .................... .................... ....................
7669 MW191399.1 |Severe acute respiratory syndrome coro    .................... .................... .................... .................... .................... ....................
7670 MW191400.1 |Severe acute respiratory syndrome coro    .................... .................... .................... .................... .................... ....................
7671 MW191401.1 |Severe acute respiratory syndrome coro    .................... .................... .................... .................... .................... ....................
7672 MW191402.1 |Severe acute respiratory syndrome coro    .................... .................... .................... .................... .................... ....................
7673 MW191403.1 |Severe acute respiratory syndrome coro    .................... .................... .................... .................... .................... ....................
7674 MW191404.1 |Severe acute respiratory syndrome coro    .................... .................... .................... .................... .................... ....................
7675 MW191405.1 |Severe acute respiratory syndrome coro    .................... .................... .................... .................... .................... ....................
7676 MW191406.1 |Severe acute respiratory syndrome coro    .................... .................... .................... .................... .................... ....................
7677 MW191407.1 |Severe acute respiratory syndrome coro    .................... .................... .................... .................... .................... ....................
7678 MW191408.1 |Severe acute respiratory syndrome coro    .................... .................... .................... .................... .................... ....................
7679 MW191409.1 |Severe acute respiratory syndrome coro    .................... .................... .................... .................... .................... ....................
7680 MW191410.1 |Severe acute respiratory syndrome coro    .................... .................... .................... .................... .................... ....................
7681 MW191411.1 |Severe acute respiratory syndrome coro    .................... .................... .................... .................... .................... ....................
7682 MW191412.1 |Severe acute respiratory syndrome coro    .................... .................... .................... .................... .................... ....................
7683 MW191413.1 |Severe acute respiratory syndrome coro    .................... .................... .................... .................... .................... ....................
7684 MW191414.1 |Severe acute respiratory syndrome coro    .................... .................... .................... .................... .................... ....................
```

| | | | | | | | | |
|---|---|---|---|---|---|---|---|---|
| 7685 | MW191415.1 | Severe acute respiratory syndrome coro | .................... | .................... | .................... | .................... | .................... | .................... |
| 7686 | MW191416.1 | Severe acute respiratory syndrome coro | .................... | .................... | .................... | .................... | .................... | .................... |
| 7687 | MW191417.1 | Severe acute respiratory syndrome coro | .................... | .................... | .................... | .................... | .................... | .................... |
| 7688 | MW191418.1 | Severe acute respiratory syndrome coro | .................... | .................... | .................... | .................... | .................... | .................... |
| 7689 | MW191419.1 | Severe acute respiratory syndrome coro | .................... | .................... | .................... | .................... | .................... | .................... |
| 7690 | MW191420.1 | Severe acute respiratory syndrome coro | .................... | .................... | .................... | .................... | .................... | .................... |
| 7691 | MW191421.1 | Severe acute respiratory syndrome coro | .................... | .................... | .................... | .................... | .................... | .................... |
| 7692 | MW191422.1 | Severe acute respiratory syndrome coro | .................... | .................... | .................... | .................... | .................... | .................... |
| 7693 | MW191423.1 | Severe acute respiratory syndrome coro | .................... | .................... | .................... | .................... | .................... | .................... |
| 7694 | MW191424.1 | Severe acute respiratory syndrome coro | .................... | .................... | .................... | .................... | .................... | .................... |
| 7695 | MW191425.1 | Severe acute respiratory syndrome coro | .................... | .................... | .................... | .................... | .................... | .................... |
| 7696 | MW191426.1 | Severe acute respiratory syndrome coro | .................... | .................... | .................... | .................... | .................... | .................... |
| 7697 | MW191427.1 | Severe acute respiratory syndrome coro | .................... | .................... | .................... | .................... | .................... | .................... |
| 7698 | MW191428.1 | Severe acute respiratory syndrome coro | .................... | .................... | .................... | .................... | .................... | .................... |
| 7699 | MW191429.1 | Severe acute respiratory syndrome coro | .................... | .................... | .................... | .................... | .................... | .................... |
| 7700 | MW191430.1 | Severe acute respiratory syndrome coro | .................... | .................... | .................... | .................... | .................... | .................... |
| 7701 | MW191431.1 | Severe acute respiratory syndrome coro | .................... | .................... | .................... | .................... | .................... | .................... |
| 7702 | MW191432.1 | Severe acute respiratory syndrome coro | .................... | .A.................. | .................... | .................... | .................... | .................... |
| 7703 | MW191433.1 | Severe acute respiratory syndrome coro | .................... | .................... | .................... | .................... | .................... | .................... |
| 7704 | MW191434.1 | Severe acute respiratory syndrome coro | .................... | .................... | .................... | .................... | .................... | .................... |
| 7705 | MW191435.1 | Severe acute respiratory syndrome coro | .................... | .................... | .................... | .................... | .................... | .................... |
| 7706 | MW191436.1 | Severe acute respiratory syndrome coro | .................... | .................... | .................... | .................... | .................... | .................... |
| 7707 | MW191437.1 | Severe acute respiratory syndrome coro | .................... | .................... | .................... | .................... | .................... | .................... |
| 7708 | MW191438.1 | Severe acute respiratory syndrome coro | .................... | .................... | .................... | .................... | .................... | .................... |
| 7709 | MW191439.1 | Severe acute respiratory syndrome coro | .................... | .................... | .................... | .................... | .................... | .................... |
| 7710 | MW191440.1 | Severe acute respiratory syndrome coro | .................... | .................... | .................... | .................... | .................... | .................... |
| 7711 | MW191441.1 | Severe acute respiratory syndrome coro | .................... | .................... | .................... | .................... | .................... | .................... |
| 7712 | MW191442.1 | Severe acute respiratory syndrome coro | .................... | .................... | .................... | .................... | .................... | .................... |
| 7713 | MW191443.1 | Severe acute respiratory syndrome coro | .................... | .................... | .................... | .................... | .................... | .................... |
| 7714 | MW191444.1 | Severe acute respiratory syndrome coro | .................... | .................... | .................... | .................... | .................... | .................... |
| 7715 | MW191445.1 | Severe acute respiratory syndrome coro | .................... | .................... | .................... | .................... | .................... | .................... |
| 7716 | MW191446.1 | Severe acute respiratory syndrome coro | .................... | .................... | .................... | .................... | .................... | .................... |
| 7717 | MW191447.1 | Severe acute respiratory syndrome coro | .................... | .................... | .................... | .................... | .................... | .................... |
| 7718 | MW191448.1 | Severe acute respiratory syndrome coro | .................... | .................... | .................... | .................... | .................... | .................... |
| 7719 | MW191449.1 | Severe acute respiratory syndrome coro | .................... | .................... | .................... | .................... | .................... | .................... |
| 7720 | MW191450.1 | Severe acute respiratory syndrome coro | .................... | .................... | .................... | .................... | .................... | .................... |
| 7721 | MW191452.1 | Severe acute respiratory syndrome coro | .................... | .................... | .................... | .................... | .................... | .................... |
| 7722 | MW191453.1 | Severe acute respiratory syndrome coro | .................... | .................... | .................... | .................... | .................... | .................... |
| 7723 | MW191454.1 | Severe acute respiratory syndrome coro | .................... | .................... | .................... | .................... | .................... | .................... |
| 7724 | MW191455.1 | Severe acute respiratory syndrome coro | .................... | .................... | .................... | .................... | .................... | .................... |
| 7725 | MW191456.1 | Severe acute respiratory syndrome coro | .................... | .................... | .................... | .................... | .................... | .................... |
| 7726 | MW191457.1 | Severe acute respiratory syndrome coro | .................... | .................... | .................... | .................... | .................... | .................... |
| 7727 | MW191458.1 | Severe acute respiratory syndrome coro | .................... | .................... | .................... | .................... | .................... | .................... |
| 7728 | MW191459.1 | Severe acute respiratory syndrome coro | .................... | .................... | .................... | .................... | .................... | .................... |
| 7729 | MW191460.1 | Severe acute respiratory syndrome coro | .................... | .................... | .................... | .................... | .................... | .................... |
| 7730 | MW191461.1 | Severe acute respiratory syndrome coro | .................... | .................... | .................... | .................... | .................... | .................... |
| 7731 | MW191462.1 | Severe acute respiratory syndrome coro | .................... | .................... | .................... | .................... | .................... | .................... |
| 7732 | MW191463.1 | Severe acute respiratory syndrome coro | .................... | .................... | .................... | .................... | .................... | .................... |
| 7733 | MW191464.1 | Severe acute respiratory syndrome coro | .................... | .................... | .................... | .................... | .................... | .................... |
| 7734 | MW191465.1 | Severe acute respiratory syndrome coro | .................... | .................... | .................... | .................... | .................... | .................... |
| 7735 | MW191466.1 | Severe acute respiratory syndrome coro | .................... | .................... | .................... | .................... | .................... | .................... |
| 7736 | MW191467.1 | Severe acute respiratory syndrome coro | .................... | .................... | .................... | .................... | .................... | .................... |
| 7737 | MW191468.1 | Severe acute respiratory syndrome coro | .................... | .................... | .................... | .................... | .................... | .................... |
| 7738 | MW191469.1 | Severe acute respiratory syndrome coro | .................... | .................... | .................... | .................... | .................... | .................... |
| 7739 | MW191470.1 | Severe acute respiratory syndrome coro | .................... | .................... | .................... | .................... | .................... | .................... |
| 7740 | MW191471.1 | Severe acute respiratory syndrome coro | .................... | .................... | .................... | .................... | .................... | .................... |
| 7741 | MW191472.1 | Severe acute respiratory syndrome coro | .................... | .................... | .................... | .................... | .................... | .................... |
| 7742 | MW191473.1 | Severe acute respiratory syndrome coro | .................... | .................... | .................... | .................... | .................... | .................... |
| 7743 | MW191474.1 | Severe acute respiratory syndrome coro | .................... | .................... | .................... | .................... | .................... | .................... |
| 7744 | MW191475.1 | Severe acute respiratory syndrome coro | .................... | .................... | .................... | .................... | .................... | .................... |
| 7745 | MW191476.1 | Severe acute respiratory syndrome coro | .................... | .................... | .................... | .................... | .................... | .................... |
| 7746 | MW191477.1 | Severe acute respiratory syndrome coro | .................... | .................... | .................... | .................... | .................... | .................... |
| 7747 | MW191478.1 | Severe acute respiratory syndrome coro | .................... | .................... | .................... | .................... | .................... | .................... |
| 7748 | MW191479.1 | Severe acute respiratory syndrome coro | .................... | .................... | .................... | .................... | .................... | .................... |
| 7749 | MW191480.1 | Severe acute respiratory syndrome coro | .................... | .................... | .................... | .................... | .................... | .................... |
| 7750 | MW191481.1 | Severe acute respiratory syndrome coro | .................... | .................... | .................... | .................... | .................... | .................... |
| 7751 | MW191482.1 | Severe acute respiratory syndrome coro | .................... | .................... | .................... | .................... | .................... | .................... |
| 7752 | MW191483.1 | Severe acute respiratory syndrome coro | .................... | .................... | .................... | .................... | .................... | .................... |
| 7753 | MW191484.1 | Severe acute respiratory syndrome coro | .................... | .................... | .................... | .................... | .................... | .................... |
| 7754 | MW191485.1 | Severe acute respiratory syndrome coro | .................... | .................... | .................... | .................... | .................... | .................... |
| 7755 | MW191486.1 | Severe acute respiratory syndrome coro | .................... | .................... | .................... | .................... | .................... | .................... |
| 7756 | MW191487.1 | Severe acute respiratory syndrome coro | .................... | .................... | .................... | .................... | .................... | .................... |
| 7757 | MW191488.1 | Severe acute respiratory syndrome coro | .................... | .................... | .................... | .................... | .................... | .................... |
| 7758 | MW191489.1 | Severe acute respiratory syndrome coro | .................... | .................... | .................... | .................... | .................... | .................... |
| 7759 | MW191490.1 | Severe acute respiratory syndrome coro | .................... | .................... | .................... | .................... | .................... | .................... |
| 7760 | MW191491.1 | Severe acute respiratory syndrome coro | .................... | .................... | .................... | .................... | .................... | .................... |
| 7761 | MW191492.1 | Severe acute respiratory syndrome coro | .................... | .................... | .................... | .................... | .................... | .................... |
| 7762 | MW191501.1 | Severe acute respiratory syndrome coro | .................... | .................... | .................... | .................... | .................... | .................... |

| | | | | | | | | |
|---|---|---|---|---|---|---|---|---|
| 7763 | MW191509.1 | \|Severe acute respiratory syndrome coro | ···················· | ···················· | ····················· | ···················· | ···················· | ···················· |
| 7764 | MW191510.1 | \|Severe acute respiratory syndrome coro | ···················· | ···················· | ····················· | ···················· | ···················· | ···················· |
| 7765 | MW191511.1 | \|Severe acute respiratory syndrome coro | ···················· | ···················· | ····················· | ···················· | ···················· | ···················· |
| 7766 | MW191512.1 | \|Severe acute respiratory syndrome coro | ···················· | ···················· | ····················· | ···················· | ···················· | ···················· |
| 7768 | MW192064.1 | \|Severe acute respiratory syndrome coro | ···················· | ···················· | ····················· | ···················· | ···················· | ···················· |
| 7769 | MW192065.1 | \|Severe acute respiratory syndrome coro | ···················· | ···················· | ····················· | ···················· | ···················· | ···················· |
| 7770 | MW192066.1 | \|Severe acute respiratory syndrome coro | ···················· | ···················· | ····················· | ···················· | ···················· | ···················· |
| 7771 | MW192067.1 | \|Severe acute respiratory syndrome coro | ···················· | ·············A······ | ····················· | ···················· | ···················· | ···················· |
| 7772 | MW192068.1 | \|Severe acute respiratory syndrome coro | ···················· | ···················· | ····················· | ···················· | ···················· | ···················· |
| 7773 | MW192069.1 | \|Severe acute respiratory syndrome coro | ···················· | ···················· | ····················· | ···················· | ···················· | ···················· |
| 7774 | MW192070.1 | \|Severe acute respiratory syndrome coro | ···················· | ···················· | ····················· | ···················· | ···················· | ···················· |
| 7775 | MW192071.1 | \|Severe acute respiratory syndrome coro | ···················· | ···················· | ····················· | ···················· | ···················· | ···················· |
| 7776 | MW192072.1 | \|Severe acute respiratory syndrome coro | ···················· | ···················· | ····················· | ···················· | ···················· | ···················· |
| 7777 | MW192073.1 | \|Severe acute respiratory syndrome coro | ···················· | ···················· | ····················· | ···················· | ···················· | ···················· |
| 7778 | MW192074.1 | \|Severe acute respiratory syndrome coro | ···················· | ···················· | ····················· | ···················· | ···················· | ···················· |
| 7779 | MW192075.1 | \|Severe acute respiratory syndrome coro | ···················· | ···················· | ····················· | ···················· | ···················· | ···················· |
| 7780 | MW192076.1 | \|Severe acute respiratory syndrome coro | ···················· | ···················· | ····················· | ···················· | ···················· | ···················· |
| 7781 | MW192077.1 | \|Severe acute respiratory syndrome coro | ···················· | ···················· | ····················· | ···················· | ··N················· | ···················· |
| 7782 | MW192078.1 | \|Severe acute respiratory syndrome coro | ···················· | ·············A······ | ····················· | ···················· | ···················· | ···················· |
| 7783 | MW192079.1 | \|Severe acute respiratory syndrome coro | ···················· | ···················· | ····················· | ···················· | ···················· | ···················· |
| 7784 | MW192080.1 | \|Severe acute respiratory syndrome coro | ···················· | ···················· | ····················· | ···················· | ···················· | ···················· |
| 7785 | MW192081.1 | \|Severe acute respiratory syndrome coro | ···················· | ···················· | ····················· | ···················· | ···················· | ···················· |
| 7786 | MW192082.1 | \|Severe acute respiratory syndrome coro | ···················· | ···················· | ····················· | ···················· | ···················· | ···················· |
| 7787 | MW192083.1 | \|Severe acute respiratory syndrome coro | ···················· | ·············A······ | ····················· | ···················· | ···················· | ···················· |
| 7788 | MW192084.1 | \|Severe acute respiratory syndrome coro | ···················· | ···················· | ····················· | ···················· | ···················· | ···················· |
| 7789 | MW192085.1 | \|Severe acute respiratory syndrome coro | ···················· | ···················· | ····················· | ···················· | ···················· | ···················· |
| 7790 | MW192086.1 | \|Severe acute respiratory syndrome coro | ···················· | ···················· | ····················· | ···················· | ···················· | ···················· |
| 7791 | MW192087.1 | \|Severe acute respiratory syndrome coro | ···················· | ·A·················· | ····················· | ···················· | ···················· | ···················· |
| 7792 | MW192088.1 | \|Severe acute respiratory syndrome coro | ···················· | ···················· | ····················· | ···················· | ···················· | ···················· |
| 7793 | MW192089.1 | \|Severe acute respiratory syndrome coro | ···················· | ·A·················· | ····················· | ···················· | ···················· | ···················· |
| 7794 | MW192090.1 | \|Severe acute respiratory syndrome coro | ···················· | ···················· | ····················· | ···················· | ···················· | ···················· |
| 7795 | MW192091.1 | \|Severe acute respiratory syndrome coro | ···················· | ·A·················· | ····················· | ···················· | ···················· | ···················· |
| 7796 | MW192092.1 | \|Severe acute respiratory syndrome coro | ···················· | ···················· | ····················· | ···················· | ···················· | ···················· |
| 7797 | MW192093.1 | \|Severe acute respiratory syndrome coro | ···················· | ·A·················· | ····················· | ···················· | ···················· | ···················· |
| 7798 | MW192094.1 | \|Severe acute respiratory syndrome coro | ···················· | ·A·················· | ····················· | ···················· | ···················· | ···················· |
| 7799 | MW192095.1 | \|Severe acute respiratory syndrome coro | ···················· | ·A·················· | ····················· | ···················· | ···················· | ···················· |
| 7800 | MW192096.1 | \|Severe acute respiratory syndrome coro | ···················· | ·A·················· | ····················· | ···················· | ···················· | ···················· |
| 7801 | MW192097.1 | \|Severe acute respiratory syndrome coro | ···················· | ·A·················· | ····················· | ···················· | ···················· | ···················· |
| 7802 | MW192098.1 | \|Severe acute respiratory syndrome coro | ···················· | ·A·················· | ····················· | ···················· | ···················· | ···················· |
| 7803 | MW192099.1 | \|Severe acute respiratory syndrome coro | ···················· | ···················· | ····················· | ···················· | ···················· | ···················· |
| 7804 | MW192100.1 | \|Severe acute respiratory syndrome coro | ···················· | ···················· | ····················· | ···················· | ···················· | ···················· |
| 7805 | MW192101.1 | \|Severe acute respiratory syndrome coro | ···················· | ···················· | ····················· | ···················· | ···················· | ···················· |
| 7806 | MW192102.1 | \|Severe acute respiratory syndrome coro | ···················· | ·A·················· | ····················· | ···················· | ···················· | ···················· |
| 7807 | MW192103.1 | \|Severe acute respiratory syndrome coro | ···················· | ·A·················· | ····················· | ···················· | ···················· | ···················· |
| 7808 | MW192104.1 | \|Severe acute respiratory syndrome coro | ···················· | ·A·················· | ····················· | ···················· | ···················· | ···················· |
| 7809 | MW192105.1 | \|Severe acute respiratory syndrome coro | ···················· | ·············A······ | ····················· | ···················· | ···················· | ···················· |
| 7810 | MW192106.1 | \|Severe acute respiratory syndrome coro | ···················· | ·············A······ | ····················· | ···················· | ···················· | ···················· |
| 7811 | MW192107.1 | \|Severe acute respiratory syndrome coro | ···················· | ···················· | ····················· | ···················· | ···················· | ···················· |
| 7812 | MW192108.1 | \|Severe acute respiratory syndrome coro | ···················· | ···················· | ····················· | ···················· | ···················· | ···················· |
| 7813 | MW192109.1 | \|Severe acute respiratory syndrome coro | ···················· | ···················· | ····················· | ···················· | ···················· | ···················· |
| 7814 | MW192110.1 | \|Severe acute respiratory syndrome coro | ···················· | ···················· | ····················· | ···················· | ···················· | ···················· |
| 7815 | MW192111.1 | \|Severe acute respiratory syndrome coro | ···················· | ···················· | ····················· | ···················· | ···················· | ···················· |
| 7816 | MW192112.1 | \|Severe acute respiratory syndrome coro | ···················· | ···················· | ····················· | ···················· | ···················· | ···················· |
| 7817 | MW192113.1 | \|Severe acute respiratory syndrome coro | ···················· | ·A·················· | ····················· | ···················· | ···················· | ···················· |
| 7818 | MW192114.1 | \|Severe acute respiratory syndrome coro | ···················· | ·A·················· | ····················· | ···················· | ···················· | ···················· |
| 7819 | MW192115.1 | \|Severe acute respiratory syndrome coro | ···················· | ···················· | ····················· | ···················· | ···················· | ···················· |
| 7820 | MW192116.1 | \|Severe acute respiratory syndrome coro | ···················· | ···················· | ····················· | ···················· | ···················· | ···················· |
| 7821 | MW192117.1 | \|Severe acute respiratory syndrome coro | ···················· | ···················· | ····················· | ···················· | ···················· | ···················· |
| 7822 | MW192118.1 | \|Severe acute respiratory syndrome coro | ···················· | ···················· | ····················· | ···················· | ···················· | ···················· |
| 7823 | MW192119.1 | \|Severe acute respiratory syndrome coro | ···················· | ·A·················· | ····················· | ···················· | ···················· | ···················· |
| 7824 | MW192120.1 | \|Severe acute respiratory syndrome coro | ···················· | ·A·················· | ····················· | ···················· | ···················· | ···················· |
| 7825 | MW192121.1 | \|Severe acute respiratory syndrome coro | ···················· | ·A·················· | ····················· | ···················· | ···················· | ···················· |
| 7826 | MW192122.1 | \|Severe acute respiratory syndrome coro | ···················· | ···················· | ····················· | ···················· | ···················· | ···················· |
| 7827 | MW192123.1 | \|Severe acute respiratory syndrome coro | ···················· | ·A·················· | ····················· | ···················· | ···················· | ···················· |
| 7828 | MW192124.1 | \|Severe acute respiratory syndrome coro | ···················· | ·A·················· | ····················· | ···················· | ···················· | ···················· |
| 7829 | MW192125.1 | \|Severe acute respiratory syndrome coro | ···················· | ···················· | ····················· | ···················· | ···················· | ···················· |
| 7830 | MW192126.1 | \|Severe acute respiratory syndrome coro | ···················· | ···················· | ····················· | ···················· | ··NN················ | ···················· |
| 7831 | MW192127.1 | \|Severe acute respiratory syndrome coro | ···················· | ···················· | ····················· | ···················· | ···················· | ···················· |
| 7832 | MW192766.1 | \|Severe acute respiratory syndrome coro | ···················· | ···················· | ····················· | ···················· | ···················· | ···················· |
| 7833 | MW192771.1 | \|Severe acute respiratory syndrome coro | ···················· | ···················· | ····················· | ···················· | ···················· | ···················· |
| 7834 | MW192772.1 | \|Severe acute respiratory syndrome coro | ···················· | ···················· | ····················· | ···················· | ···················· | ···················· |
| 7835 | MW186669.1 | \|Severe acute respiratory syndrome coro | ···················· | ···················· | ····················· | ···················· | ···················· | ···················· |
| 7836 | MW181434.1 | \|Severe acute respiratory syndrome coro | ···················· | ···················· | ····················· | ···················· | ···················· | ···················· |
| 7837 | MW181442.1 | \|Severe acute respiratory syndrome coro | ···················· | ···················· | ····················· | ···················· | ···················· | ···················· |
| 7838 | MW181459.1 | \|Severe acute respiratory syndrome coro | ···················· | ···················· | ····················· | ···················· | ···················· | ···················· |
| 7839 | MW181466.1 | \|Severe acute respiratory syndrome coro | ···················· | ···················· | ····················· | ···················· | ···················· | ···················· |
| 7840 | MW181473.1 | \|Severe acute respiratory syndrome coro | ···················· | ···················· | ····················· | ···················· | ···················· | ···················· |
| 7841 | MW181474.1 | \|Severe acute respiratory syndrome coro | ···················· | ···················· | ····················· | ···················· | ···················· | ···················· |

| | | | | | | | | |
|---|---|---|---|---|---|---|---|---|
| 7842 | MW181500.1 | \|Severe acute respiratory syndrome coro | .................... | .................... | .................... | .................... | .................... | .................... |
| 7843 | MW181505.1 | \|Severe acute respiratory syndrome coro | .................... | .................... | .................... | .................... | .................... | .................... |
| 7844 | MW181518.1 | \|Severe acute respiratory syndrome coro | .................... | .................... | .................... | .................... | .................... | .................... |
| 7845 | MW181520.1 | \|Severe acute respiratory syndrome coro | .................... | .................... | .................... | .................... | .................... | .................... |
| 7846 | MW181526.1 | \|Severe acute respiratory syndrome coro | .................... | .................... | .................... | .................... | .................... | .................... |
| 7847 | MW181529.1 | \|Severe acute respiratory syndrome coro | .................... | .................... | .................... | .................... | .................... | .................... |
| 7848 | MW181537.1 | \|Severe acute respiratory syndrome coro | .................... | .................... | .................... | .................... | .................... | .................... |
| 7849 | MW181549.1 | \|Severe acute respiratory syndrome coro | .................... | .................... | .................... | .................... | .................... | .................... |
| 7850 | MW181562.1 | \|Severe acute respiratory syndrome coro | .................... | .................... | .................... | .................... | .................... | .................... |
| 7851 | MW181565.1 | \|Severe acute respiratory syndrome coro | .................... | .A.................. | .................... | .................... | .................... | .................... |
| 7852 | MW181566.1 | \|Severe acute respiratory syndrome coro | .................... | .................... | .................... | .................... | .................... | .................... |
| 7853 | MW181574.1 | \|Severe acute respiratory syndrome coro | .................... | .................... | .................... | .................... | .................... | .................... |
| 7854 | MW181575.1 | \|Severe acute respiratory syndrome coro | .................... | .............A...... | .................... | .................... | .................... | .................... |
| 7855 | MW181576.1 | \|Severe acute respiratory syndrome coro | .................... | .A.................. | .................... | .................... | .................... | .................... |
| 7856 | MW181577.1 | \|Severe acute respiratory syndrome coro | .................... | .................... | .................... | .................... | .................... | .................... |
| 7857 | MW181578.1 | \|Severe acute respiratory syndrome coro | .................... | .................... | .................... | .................... | .................... | .................... |
| 7858 | MW181579.1 | \|Severe acute respiratory syndrome coro | .................... | .................... | .................... | .................... | .................... | .................... |
| 7859 | MW181580.1 | \|Severe acute respiratory syndrome coro | .................... | .................... | .................... | .................... | .................... | .................... |
| 7860 | MW181581.1 | \|Severe acute respiratory syndrome coro | .................... | .................... | .................... | .................... | .................... | .................... |
| 7861 | MW181582.1 | \|Severe acute respiratory syndrome coro | .................... | .................... | .................... | .................... | .................... | .................... |
| 7862 | MW181583.1 | \|Severe acute respiratory syndrome coro | .................... | .................... | .................... | .................... | .................... | .................... |
| 7863 | MW181584.1 | \|Severe acute respiratory syndrome coro | .................... | .................... | .................... | .................... | .................... | .................... |
| 7864 | MW181585.1 | \|Severe acute respiratory syndrome coro | .................... | .................... | .................... | .................... | .................... | .................... |
| 7865 | MW181586.1 | \|Severe acute respiratory syndrome coro | .................... | .................... | .................... | .................... | .................... | .................... |
| 7866 | MW181587.1 | \|Severe acute respiratory syndrome coro | .................... | ............C....... | .................... | .................... | .................... | .................... |
| 7867 | MW181588.1 | \|Severe acute respiratory syndrome coro | .................... | .................... | .................... | .................... | .................... | .................... |
| 7868 | MW181589.1 | \|Severe acute respiratory syndrome coro | .................... | .................... | .................... | .................... | .................... | .................... |
| 7869 | MW181590.1 | \|Severe acute respiratory syndrome coro | .................... | .................... | .................... | .................... | .................... | .................... |
| 7870 | MW181591.1 | \|Severe acute respiratory syndrome coro | .................... | .................... | .................... | .................... | .................... | .................... |
| 7871 | MW181592.1 | \|Severe acute respiratory syndrome coro | .................... | .................... | .................... | .................... | .................... | .................... |
| 7872 | MW181593.1 | \|Severe acute respiratory syndrome coro | .................... | .A.................. | .................... | .................... | .................... | .................... |
| 7873 | MW181621.1 | \|Severe acute respiratory syndrome coro | .................... | .................... | .................... | .................... | .................... | .................... |
| 7874 | MW181771.1 | \|Severe acute respiratory syndrome coro | .................... | .................... | .................... | .................... | .................... | .................... |
| 7875 | MW181772.1 | \|Severe acute respiratory syndrome coro | .................... | .................... | .................... | .................... | .................... | .................... |
| 7876 | MW181773.1 | \|Severe acute respiratory syndrome coro | .................... | .................... | .................... | .................... | .................... | .................... |
| 7877 | MW181797.1 | \|Severe acute respiratory syndrome coro | .................... | .................... | .................... | .................... | .................... | .................... |
| 7878 | MW181803.1 | \|Severe acute respiratory syndrome coro | .................... | .................... | .................... | .................... | .................... | .................... |
| 7879 | MW181804.1 | \|Severe acute respiratory syndrome coro | .................... | .................... | .................... | .................... | .................... | .................... |
| 7880 | MW181805.1 | \|Severe acute respiratory syndrome coro | .................... | .................... | .................... | .................... | .................... | .................... |
| 7881 | MW181806.1 | \|Severe acute respiratory syndrome coro | .................... | .................... | .................... | .................... | .................... | .................... |
| 7882 | MW181807.1 | \|Severe acute respiratory syndrome coro | .................... | .................... | .................... | .................... | .................... | .................... |
| 7883 | MW181811.1 | \|Severe acute respiratory syndrome coro | .................... | .................... | .................... | .................... | .................... | .................... |
| 7884 | MW181814.1 | \|Severe acute respiratory syndrome coro | .................... | .................... | .................... | .................... | .................... | .................... |
| 7885 | MW181828.1 | \|Severe acute respiratory syndrome coro | .................... | .................... | .................... | .................... | .................... | .................... |
| 7886 | MW181829.1 | \|Severe acute respiratory syndrome coro | .................... | .................... | .................... | .................... | .................... | .................... |
| 7887 | MW181831.2 | \|Severe acute respiratory syndrome coro | .................... | .................... | .................... | .................... | .................... | .................... |
| 7888 | MW181832.1 | \|Severe acute respiratory syndrome coro | .................... | .................... | .................... | .................... | .................... | .................... |
| 7889 | MW181833.1 | \|Severe acute respiratory syndrome coro | .................... | .................... | .................... | .................... | .................... | .................... |
| 7890 | MW181834.1 | \|Severe acute respiratory syndrome coro | .................... | .................... | .................... | .................... | .................... | .................... |
| 7891 | MW181835.1 | \|Severe acute respiratory syndrome coro | .................... | .................... | .................... | .................... | .................... | .................... |
| 7892 | MW181837.1 | \|Severe acute respiratory syndrome coro | .................... | .................... | .................... | .................... | .................... | .................... |
| 7893 | MW181838.2 | \|Severe acute respiratory syndrome coro | .................... | .................... | .................... | .................... | .................... | .................... |
| 7894 | MW181839.1 | \|Severe acute respiratory syndrome coro | .................... | .................... | .................... | .................... | .................... | .................... |
| 7895 | MW181840.1 | \|Severe acute respiratory syndrome coro | .................... | .................... | .................... | .................... | .................... | .................... |
| 7896 | MW181843.1 | \|Severe acute respiratory syndrome coro | .................... | .................... | .................... | .................... | .................... | .................... |
| 7897 | MW181844.1 | \|Severe acute respiratory syndrome coro | .................... | .................... | .................... | .................... | .................... | .................... |
| 7898 | MW181845.1 | \|Severe acute respiratory syndrome coro | .................... | .................... | .................... | .................... | .................... | .................... |
| 7899 | MW181846.1 | \|Severe acute respiratory syndrome coro | .................... | .................... | .................... | .................... | .................... | .................... |
| 7900 | MW181847.1 | \|Severe acute respiratory syndrome coro | .................... | .................... | .................... | .................... | .................... | .................... |
| 7901 | MW181848.1 | \|Severe acute respiratory syndrome coro | .................... | .................... | .................... | .................... | .................... | .................... |
| 7902 | MW183931.1 | \|Severe acute respiratory syndrome coro | .................... | .................... | .................... | .................... | .................... | .................... |
| 7903 | MW183932.1 | \|Severe acute respiratory syndrome coro | .................... | .................... | .................... | .................... | .................... | .................... |
| 7904 | MW183934.1 | \|Severe acute respiratory syndrome coro | .................... | .................... | .................... | .................... | .................... | .................... |
| 7905 | MW183935.1 | \|Severe acute respiratory syndrome coro | .................... | .................... | .................... | .................... | .................... | .................... |
| 7906 | MW183936.1 | \|Severe acute respiratory syndrome coro | .................... | .................... | .................... | .................... | .................... | .................... |
| 7907 | MW183938.1 | \|Severe acute respiratory syndrome coro | .................... | .................... | .................... | .................... | .................... | .................... |
| 7908 | MW183939.1 | \|Severe acute respiratory syndrome coro | .................... | .................... | .................... | .................... | .................... | .................... |
| 7909 | MW183941.1 | \|Severe acute respiratory syndrome coro | .................... | .................... | .................... | .................... | .................... | .................... |
| 7910 | MW183942.1 | \|Severe acute respiratory syndrome coro | .................... | .................... | .................... | .................... | .................... | .................... |
| 7911 | MW183943.1 | \|Severe acute respiratory syndrome coro | .................... | .................... | .................... | .................... | .................... | .................... |
| 7912 | MW183944.1 | \|Severe acute respiratory syndrome coro | .................... | .................... | .................... | .................... | .................... | .................... |
| 7913 | MW183945.1 | \|Severe acute respiratory syndrome coro | .................... | .................... | .................... | .................... | .................... | .................... |
| 7914 | MW183946.1 | \|Severe acute respiratory syndrome coro | .................... | .................... | .................... | .................... | .................... | .................... |
| 7915 | MW183947.1 | \|Severe acute respiratory syndrome coro | .................... | .................... | .................... | .................... | .................... | .................... |
| 7917 | MW183951.1 | \|Severe acute respiratory syndrome coro | .................... | .................... | .................... | .................... | .................... | .................... |
| 7918 | MW183952.1 | \|Severe acute respiratory syndrome coro | .................... | .................... | .................... | .................... | .................... | .................... |
| 7919 | MW183953.1 | \|Severe acute respiratory syndrome coro | .................... | .................... | .................... | .................... | .................... | .................... |
| 7920 | MW183956.1 | \|Severe acute respiratory syndrome coro | .................... | .................... | .................... | .................... | .................... | .................... |

| | | | | | | | | | |
|---|---|---|---|---|---|---|---|---|---|
| 7921 | MW183957.1 | Severe acute respiratory syndrome coro | ···················· | ···················· | ···················· | ···················· | ···················· | ···················· | |
| 7922 | MW183959.1 | Severe acute respiratory syndrome coro | ···················· | ···················· | ···················· | ···················· | ···················· | ···················· | |
| 7923 | MW183960.1 | Severe acute respiratory syndrome coro | ···················· | ···················· | ···················· | ···················· | ·NNN················ | ···················· | 1 |
| 7924 | MW183961.1 | Severe acute respiratory syndrome coro | ···················· | ···················· | ···················· | ···················· | ···················· | ···················· | |
| 7925 | MW183962.1 | Severe acute respiratory syndrome coro | ···················· | ···················· | ···················· | ···················· | ···················· | ···················· | |
| 7926 | MW183963.1 | Severe acute respiratory syndrome coro | ···················· | ···················· | ···················· | ···················· | ···················· | ···················· | |
| 7927 | MW183964.1 | Severe acute respiratory syndrome coro | ···················· | ···················· | ···················· | ···················· | ···················· | ···················· | |
| 7928 | MW183965.1 | Severe acute respiratory syndrome coro | ···················· | ···················· | ···················· | ···················· | ···················· | ···················· | |
| 7929 | MW183966.1 | Severe acute respiratory syndrome coro | ···················· | ···················· | ···················· | ···················· | ···················· | ···················· | |
| 7931 | MW183970.1 | Severe acute respiratory syndrome coro | ···················· | ···················· | ···················· | ···················· | ···················· | ···················· | |
| 7932 | MW183971.1 | Severe acute respiratory syndrome coro | ···················· | ···················· | ···················· | ···················· | ···················· | ···················· | |
| 7933 | MW183972.1 | Severe acute respiratory syndrome coro | ···················· | ···················· | ···················· | ···················· | ···················· | ···················· | |
| 7934 | MW183975.1 | Severe acute respiratory syndrome coro | ···················· | ···················· | ···················· | ···················· | ···················· | ···················· | |
| 7935 | MW183976.1 | Severe acute respiratory syndrome coro | ···················· | ···················· | ···················· | ···················· | ···················· | ···················· | |
| 7936 | MW183977.1 | Severe acute respiratory syndrome coro | ···················· | ···················· | ···················· | ···················· | ···················· | ···················· | |
| 7937 | MW183978.1 | Severe acute respiratory syndrome coro | ···················· | ···················· | ···················· | ···················· | ···················· | ···················· | |
| 7938 | MW183980.1 | Severe acute respiratory syndrome coro | ···················· | ···················· | ···················· | ···················· | ···················· | ···················· | |
| 7939 | MW183981.1 | Severe acute respiratory syndrome coro | ···················· | ···················· | ···················· | ···················· | ···················· | ···················· | |
| 7940 | MW183982.1 | Severe acute respiratory syndrome coro | ···················· | ···················· | ···················· | ···················· | ···················· | ···················· | |
| 7941 | MW183983.1 | Severe acute respiratory syndrome coro | ···················· | ···················· | ···················· | ···················· | ···················· | ···················· | |
| 7942 | MW183984.1 | Severe acute respiratory syndrome coro | ···················· | ···················· | ···················· | ···················· | ···················· | ···················· | |
| 7943 | MW183985.1 | Severe acute respiratory syndrome coro | ···················· | ···················· | ···················· | ···················· | ···················· | ···················· | |
| 7944 | MW183986.1 | Severe acute respiratory syndrome coro | ···················· | ···················· | ···················· | ···················· | ···················· | ···················· | |
| 7945 | MW183987.1 | Severe acute respiratory syndrome coro | ···················· | ···················· | ···················· | ···················· | ···················· | ···················· | |
| 7946 | MW183988.1 | Severe acute respiratory syndrome coro | ···················· | ···················· | ···················· | ···················· | ···················· | ···················· | |
| 7947 | MW183989.1 | Severe acute respiratory syndrome coro | ···················· | ···················· | ···················· | ···················· | ···················· | ···················· | |
| 7948 | MW183990.1 | Severe acute respiratory syndrome coro | ···················· | ···················· | ···················· | ···················· | ···················· | ···················· | |
| 7950 | MW183994.1 | Severe acute respiratory syndrome coro | ···················· | ···················· | ···················· | ···················· | ···················· | ···················· | |
| 7951 | MW183995.1 | Severe acute respiratory syndrome coro | ···················· | ···················· | ···················· | ···················· | ···················· | ···················· | |
| 7952 | MW183996.1 | Severe acute respiratory syndrome coro | ···················· | ···················· | ···················· | ···················· | ···················· | ···················· | |
| 7953 | MW183997.1 | Severe acute respiratory syndrome coro | ···················· | ···················· | ···················· | ···················· | ···················· | ···················· | |
| 7954 | MW183998.1 | Severe acute respiratory syndrome coro | ···················· | ···················· | ···················· | ···················· | ···················· | ···················· | |
| 7955 | MW183999.1 | Severe acute respiratory syndrome coro | ···················· | ···················· | ···················· | ···················· | ···················· | ···················· | |
| 7956 | MW184000.1 | Severe acute respiratory syndrome coro | ···················· | ···················· | ···················· | ···················· | ···················· | ···················· | |
| 7957 | MW184001.1 | Severe acute respiratory syndrome coro | ···················· | ···················· | ···················· | ···················· | ···················· | ···················· | |
| 7958 | MW184002.1 | Severe acute respiratory syndrome coro | ···················· | ···················· | ···················· | ···················· | ···················· | ···················· | |
| 7959 | MW184003.1 | Severe acute respiratory syndrome coro | ···················· | ···················· | ···················· | ···················· | ···················· | ···················· | |
| 7960 | MW184004.1 | Severe acute respiratory syndrome coro | ···················· | ···················· | ···················· | ···················· | ···················· | ···················· | |
| 7962 | MW184007.1 | Severe acute respiratory syndrome coro | ···················· | ···················· | ···················· | ···················· | ···················· | ···················· | |
| 7963 | MW184009.1 | Severe acute respiratory syndrome coro | ···················· | ···················· | ···················· | ···················· | ···················· | ···················· | |
| 7964 | MW184010.1 | Severe acute respiratory syndrome coro | ···················· | ···················· | ···················· | ···················· | ···················· | ···················· | |
| 7965 | MW184013.1 | Severe acute respiratory syndrome coro | ···················· | ···················· | ···················· | ···················· | ···················· | ···················· | |
| 7966 | MW184014.1 | Severe acute respiratory syndrome coro | ···················· | ···················· | ···················· | ···················· | ···················· | ···················· | |
| 7967 | MW184015.1 | Severe acute respiratory syndrome coro | ···················· | ···················· | ···················· | ···················· | ···················· | ···················· | |
| 7968 | MW184016.1 | Severe acute respiratory syndrome coro | ···················· | ···················· | ···················· | ···················· | ···················· | ···················· | |
| 7969 | MW184019.1 | Severe acute respiratory syndrome coro | ···················· | ···················· | ···················· | ···················· | ···················· | ···················· | |
| 7970 | MW184020.1 | Severe acute respiratory syndrome coro | ···················· | ···················· | ···················· | ···················· | ···················· | ···················· | |
| 7971 | MW184021.1 | Severe acute respiratory syndrome coro | ···················· | ···················· | ···················· | ···················· | ···················· | ···················· | |
| 7972 | MW184022.1 | Severe acute respiratory syndrome coro | ···················· | ···················· | ···················· | ···················· | ···················· | ···················· | |
| 7973 | MW184023.1 | Severe acute respiratory syndrome coro | ···················· | ···················· | ···················· | ···················· | ···················· | ···················· | |
| 7974 | MW184025.1 | Severe acute respiratory syndrome coro | ···················· | ···················· | ···················· | ···················· | ···················· | ···················· | |
| 7975 | MW184026.1 | Severe acute respiratory syndrome coro | ···················· | ···················· | ···················· | ···················· | ···················· | ···················· | |
| 7976 | MW184027.1 | Severe acute respiratory syndrome coro | ···················· | ···················· | ···················· | ···················· | ···················· | ···················· | |
| 7977 | MW184029.1 | Severe acute respiratory syndrome coro | ···················· | ···················· | ···················· | ···················· | ···················· | ···················· | |
| 7978 | MW184030.1 | Severe acute respiratory syndrome coro | ···················· | ···················· | ···················· | ···················· | ···················· | ···················· | |
| 7979 | MW184032.1 | Severe acute respiratory syndrome coro | ···················· | ·····A·············· | ···················· | ···················· | ···················· | ···················· | |
| 7980 | MW184033.1 | Severe acute respiratory syndrome coro | ···················· | ···················· | ···················· | ···················· | ···················· | ···················· | |
| 7981 | MW184035.1 | Severe acute respiratory syndrome coro | ···················· | ···················· | ···················· | ···················· | ···················· | ···················· | |
| 7982 | MW184036.1 | Severe acute respiratory syndrome coro | ···················· | ···················· | ···················· | ···················· | ···················· | ···················· | |
| 7983 | MW184037.1 | Severe acute respiratory syndrome coro | ···················· | ···················· | ···················· | ···················· | ···················· | ···················· | |
| 7984 | MW184038.1 | Severe acute respiratory syndrome coro | ···················· | ···················· | ···················· | ···················· | ···················· | ···················· | |
| 7985 | MW184041.1 | Severe acute respiratory syndrome coro | ···················· | ···················· | ···················· | ···················· | ···················· | ···················· | |
| 7986 | MW184042.1 | Severe acute respiratory syndrome coro | ···················· | ···················· | ···················· | ···················· | ···················· | ···················· | |
| 7987 | MW184044.1 | Severe acute respiratory syndrome coro | ···················· | ···················· | ···················· | ···················· | ···················· | ···················· | |
| 7988 | MW184045.1 | Severe acute respiratory syndrome coro | ···················· | ···················· | ···················· | ···················· | ···················· | ···················· | |
| 7989 | MW184046.1 | Severe acute respiratory syndrome coro | ···················· | ···················· | ···················· | ···················· | ···················· | ···················· | |
| 7990 | MW184048.1 | Severe acute respiratory syndrome coro | ···················· | ···················· | ···················· | ···················· | ···················· | ···················· | |
| 7991 | MW184049.1 | Severe acute respiratory syndrome coro | ···················· | ···················· | ···················· | ···················· | ···················· | ···················· | |
| 7992 | MW184050.1 | Severe acute respiratory syndrome coro | ···················· | ···················· | ···················· | ···················· | ···················· | ···················· | |
| 7993 | MW184051.1 | Severe acute respiratory syndrome coro | ···················· | ···················· | ···················· | ···················· | ···················· | ···················· | |
| 7994 | MW184052.1 | Severe acute respiratory syndrome coro | ···················· | ···················· | ···················· | ···················· | ···················· | ···················· | |
| 7995 | MW184053.1 | Severe acute respiratory syndrome coro | ···················· | ···················· | ···················· | ···················· | ···················· | ···················· | |
| 7996 | MW184054.1 | Severe acute respiratory syndrome coro | ···················· | ···················· | ···················· | ···················· | ···················· | ···················· | |
| 7997 | MW184056.1 | Severe acute respiratory syndrome coro | ···················· | ···················· | ···················· | ···················· | ···················· | ···················· | |
| 7998 | MW184057.1 | Severe acute respiratory syndrome coro | ···················· | ···················· | ···················· | ···················· | ···················· | ···················· | |
| 7999 | MW184058.1 | Severe acute respiratory syndrome coro | ···················· | ···················· | ···················· | ···················· | ···················· | ···················· | |
| 8000 | MW184060.1 | Severe acute respiratory syndrome coro | ···················· | ···················· | ···················· | ···················· | ···················· | ···················· | |
| 8001 | MW184061.1 | Severe acute respiratory syndrome coro | ···················· | ···················· | ···················· | ···················· | ···················· | ···················· | |

8002 MW184062.1 |Severe acute respiratory syndrome coro .................... .................... .................... .................... .................... ....................
8003 MW184063.1 |Severe acute respiratory syndrome coro .................... .................... .................... .................... .................... ....................
8004 MW184064.1 |Severe acute respiratory syndrome coro .................... .................... .................... .................... .................... ....................
8005 MW184066.1 |Severe acute respiratory syndrome coro .................... .................... .................... .................... .................... ....................
8006 MW184067.1 |Severe acute respiratory syndrome coro .................... .................... .................... .................... .................... ....................
8007 MW184068.1 |Severe acute respiratory syndrome coro .................... .................... .................... .................... .................... ....................
8008 MW184069.1 |Severe acute respiratory syndrome coro .................... .................... .................... .................... .................... ....................
8009 MW184070.1 |Severe acute respiratory syndrome coro .................... .................... .................... .................... .................... ....................
8010 MW184073.1 |Severe acute respiratory syndrome coro .................... .................... .................... .................... .................... ....................
8011 MW184074.1 |Severe acute respiratory syndrome coro .................... .................... .................... .................... .................... ....................
8012 MW184075.1 |Severe acute respiratory syndrome coro .................... .................... .................... .................... .................... ....................
8013 MW184077.1 |Severe acute respiratory syndrome coro .................... .................... .................... .................... .................... ....................
8014 MW184079.1 |Severe acute respiratory syndrome coro .................... .................... .................... .................... .................... ....................
8015 MW184082.1 |Severe acute respiratory syndrome coro .................... .................... .................... .................... .................... ....................
8016 MW184083.1 |Severe acute respiratory syndrome coro .................... .................... .................... .................... .................... ....................
8017 MW184084.1 |Severe acute respiratory syndrome coro .................... .................... .................... .................... .................... ....................
8018 MW184085.1 |Severe acute respiratory syndrome coro .................... .................... .................... .................... .................... ....................
8019 MW184087.1 |Severe acute respiratory syndrome coro .................... .................... .................... .................... .................... ....................
8020 MW184088.1 |Severe acute respiratory syndrome coro .................... .................... .................... .................... .................... ....................
8021 MW184089.1 |Severe acute respiratory syndrome coro .................... .................... .................... .................... .................... ....................
8022 MW184091.1 |Severe acute respiratory syndrome coro .................... .................... .................... .................... .................... ....................
8023 MW184092.1 |Severe acute respiratory syndrome coro .................... .................... .................... .................... .................... ....................
8024 MW184093.1 |Severe acute respiratory syndrome coro .................... .................... .................... .................... .................... ....................
8025 MW184094.1 |Severe acute respiratory syndrome coro .................... .................... .................... .................... .................... ....................
8026 MW184095.1 |Severe acute respiratory syndrome coro .................... .................... .................... .................... .................... ....................
8027 MW184097.1 |Severe acute respiratory syndrome coro .................... .................... .................... .................... .................... ....................
8028 MW184098.1 |Severe acute respiratory syndrome coro .................... .................... .................... .................... .................... ....................
8029 MW184099.1 |Severe acute respiratory syndrome coro .................... .................... .................... .................... .................... ....................
8030 MW184100.1 |Severe acute respiratory syndrome coro .................... .................... .................... .................... .................... ....................
8031 MW184101.1 |Severe acute respiratory syndrome coro .................... .................... .................... .................... .................... ....................
8032 MW184102.1 |Severe acute respiratory syndrome coro .................... .................... .................... .................... .................... ....................
8033 MW184103.1 |Severe acute respiratory syndrome coro .................... .................... .................... .................... .................... ....................
8034 MW184105.1 |Severe acute respiratory syndrome coro .................... .................... .................... .................... .................... ....................
8035 MW184107.1 |Severe acute respiratory syndrome coro .................... .................... .................... .................... .................... ....................
8036 MW184108.1 |Severe acute respiratory syndrome coro .................... .................... .................... .................... .................... ....................
8037 MW184109.1 |Severe acute respiratory syndrome coro .................... .................... .................... .................... .................... ....................
8038 MW184110.1 |Severe acute respiratory syndrome coro .................... .................... .................... .................... .................... ....................
8039 MW184111.1 |Severe acute respiratory syndrome coro .................... ......A............. .................... .................... .................... ....................
8040 MW184112.1 |Severe acute respiratory syndrome coro .................... .................... .................... .................... .................... ....................
8041 MW184113.1 |Severe acute respiratory syndrome coro .................... .................... .................... .................... .................... ....................
8042 MW184115.1 |Severe acute respiratory syndrome coro .................... .................... .................... .................... .................... ....................
8043 MW184116.1 |Severe acute respiratory syndrome coro .................... .................... .................... .................... .................... ....................
8044 MW184117.1 |Severe acute respiratory syndrome coro .................... .................... .................... .................... .................... ....................
8045 MW184121.1 |Severe acute respiratory syndrome coro .................... .................... .................... .................... .................... ....................
8046 MW184122.1 |Severe acute respiratory syndrome coro .................... .................... .................... .................... .................... ....................
8047 MW184123.1 |Severe acute respiratory syndrome coro .................... .................... .................... .................... .................... ....................
8048 MW184124.1 |Severe acute respiratory syndrome coro .................... .................... .................... .................... .................... ....................
8049 MW184125.1 |Severe acute respiratory syndrome coro .................... .................... .................... .................... .................... ....................
8050 MW184126.1 |Severe acute respiratory syndrome coro .................... .................... .................... .................... .................... ....................
8051 MW184127.1 |Severe acute respiratory syndrome coro .................... .................... .................... .................... .................... ....................
8052 MW184128.1 |Severe acute respiratory syndrome coro .................... .................... .................... .................... .................... ....................
8053 MW184129.1 |Severe acute respiratory syndrome coro .................... .................... .................... .................... .................... ....................
8054 MW184130.1 |Severe acute respiratory syndrome coro .................... .................... .................... .................... .................... ....................
8055 MW184131.1 |Severe acute respiratory syndrome coro .................... .................... .................... .................... .................... ....................
8056 MW184132.1 |Severe acute respiratory syndrome coro .................... .................... .................... .................... .................... ....................
8057 MW184134.1 |Severe acute respiratory syndrome coro .................... .................... .................... .................... .................... ....................
8058 MW184136.1 |Severe acute respiratory syndrome coro .................... .................... .................... .................... .................... ....................
8059 MW184137.1 |Severe acute respiratory syndrome coro .................... .................... .................... .................... .................... ....................
8060 MW184138.1 |Severe acute respiratory syndrome coro .................... .................... .................... .................... .................... ....................
8061 MW184139.1 |Severe acute respiratory syndrome coro .................... .................... .................... .................... .................... ....................
8062 MW184140.1 |Severe acute respiratory syndrome coro .................... .................... .................... .................... .................... ....................
8063 MW184141.1 |Severe acute respiratory syndrome coro .................... .................... .................... .................... .................... ....................
8064 MW184142.1 |Severe acute respiratory syndrome coro .................... .................... .................... .................... .................... ....................
8065 MW184143.1 |Severe acute respiratory syndrome coro .................... .................... .................... .................... .................... ....................
8066 MW184144.1 |Severe acute respiratory syndrome coro .................... .................... .................... .................... .................... ....................
8067 MW184148.1 |Severe acute respiratory syndrome coro .................... .................... .................... .................... .................... ....................
8068 MW184149.1 |Severe acute respiratory syndrome coro .................... .................... .................... .................... .................... ....................
8069 MW184150.1 |Severe acute respiratory syndrome coro .................... .................... .................... .................... .................... ....................
8070 MW184151.1 |Severe acute respiratory syndrome coro .................... .................... .................... .................... .................... ....................
8071 MW184152.1 |Severe acute respiratory syndrome coro .................... .................... .................... .................... .................... ....................
8072 MW184153.1 |Severe acute respiratory syndrome coro .................... .................... .................... .................... .................... ....................
8073 MW184154.1 |Severe acute respiratory syndrome coro .................... .................... .................... .................... .................... ....................
8074 MW184155.1 |Severe acute respiratory syndrome coro .................... .................... .................... .................... .................... ....................
8075 MW184156.1 |Severe acute respiratory syndrome coro .................... .................... .................... .................... .................... ....................
8076 MW184157.1 |Severe acute respiratory syndrome coro .................... .................... .................... .................... .................... ....................
8077 MW184158.1 |Severe acute respiratory syndrome coro .................... .................... .................... .................... .................... ....................
8078 MW184159.1 |Severe acute respiratory syndrome coro .................... .................... .................... .................... .................... ....................
8079 MW184160.1 |Severe acute respiratory syndrome coro .................... .................... .................... .................... .................... ....................

| | | | | | | | | |
|---|---|---|---|---|---|---|---|---|
| 8080 | MW184161.1 | Severe acute respiratory syndrome coro | .................... | .................... | ..................... | .................... | .................... | .................... |
| 8081 | MW184162.1 | Severe acute respiratory syndrome coro | .................... | .................... | ..................... | .................... | .................... | .................... |
| 8082 | MW184163.1 | Severe acute respiratory syndrome coro | .................... | .................... | ..................... | .................... | .................... | .................... |
| 8083 | MW184164.1 | Severe acute respiratory syndrome coro | .................... | .................... | ..................... | .................... | .................... | .................... |
| 8084 | MW184166.1 | Severe acute respiratory syndrome coro | .................... | .................... | ..................... | .................... | .................... | .................... |
| 8085 | MW184168.1 | Severe acute respiratory syndrome coro | .................... | .................... | ..................... | .................... | .................... | .................... |
| 8086 | MW184169.1 | Severe acute respiratory syndrome coro | .................... | .................... | ..................... | .................... | .................... | .................... |
| 8087 | MW184170.1 | Severe acute respiratory syndrome coro | .................... | .................... | ..................... | .................... | .................... | .................... |
| 8088 | MW184171.1 | Severe acute respiratory syndrome coro | .................... | .................... | ..................... | .................... | .................... | .................... |
| 8089 | MW184172.1 | Severe acute respiratory syndrome coro | .................... | .................... | ..................... | .................... | .................... | .................... |
| 8090 | MW184174.1 | Severe acute respiratory syndrome coro | .................... | .................... | ..................... | .................... | .................... | .................... |
| 8091 | MW184175.1 | Severe acute respiratory syndrome coro | .................... | .................... | ..................... | .................... | .................... | .................... |
| 8092 | MW184176.1 | Severe acute respiratory syndrome coro | .................... | .................... | ..................... | .................... | .................... | .................... |
| 8093 | MW184177.1 | Severe acute respiratory syndrome coro | .................... | .................... | ..................... | .................... | .................... | .................... |
| 8094 | MW184178.1 | Severe acute respiratory syndrome coro | .................... | .................... | ..................... | .................... | .................... | .................... |
| 8095 | MW184180.1 | Severe acute respiratory syndrome coro | .................... | .................... | ..................... | .................... | .................... | .................... |
| 8096 | MW184182.1 | Severe acute respiratory syndrome coro | .................... | .................... | ..................... | .................... | .................... | .................... |
| 8097 | MW184183.1 | Severe acute respiratory syndrome coro | .................... | .................... | ..................... | .................... | .................... | .................... |
| 8098 | MW184184.1 | Severe acute respiratory syndrome coro | .................... | .................... | ..................... | .................... | .................... | .................... |
| 8099 | MW184185.1 | Severe acute respiratory syndrome coro | .................... | .................... | ..................... | .................... | .................... | .................... |
| 8100 | MW184186.1 | Severe acute respiratory syndrome coro | .................... | .................... | ..................... | .................... | .................... | .................... |
| 8101 | MW184188.1 | Severe acute respiratory syndrome coro | .................... | .................... | ..................... | .................... | .................... | .................... |
| 8102 | MW184190.1 | Severe acute respiratory syndrome coro | .................... | .................... | ..................... | .................... | .................... | .................... |
| 8103 | MW184191.1 | Severe acute respiratory syndrome coro | .................... | .................... | ..................... | .................... | .................... | .................... |
| 8104 | MW184194.1 | Severe acute respiratory syndrome coro | .................... | .................... | ..................... | .................... | .................... | .................... |
| 8105 | MW184195.1 | Severe acute respiratory syndrome coro | .................... | .................... | ..................... | .................... | .................... | .................... |
| 8106 | MW184196.1 | Severe acute respiratory syndrome coro | .................... | .................... | ..................... | .................... | .................... | .................... |
| 8107 | MW184197.1 | Severe acute respiratory syndrome coro | .................... | .................... | ..................... | .................... | .................... | .................... |
| 8108 | MW184198.1 | Severe acute respiratory syndrome coro | .................... | .................... | ..................... | .................... | .................... | .................... |
| 8109 | MW184199.1 | Severe acute respiratory syndrome coro | .................... | .................... | ..................... | .................... | .................... | .................... |
| 8110 | MW184200.1 | Severe acute respiratory syndrome coro | .................... | .................... | ..................... | .................... | .................... | .................... |
| 8111 | MW184201.1 | Severe acute respiratory syndrome coro | .................... | .................... | ..................... | .................... | .................... | .................... |
| 8112 | MW184202.1 | Severe acute respiratory syndrome coro | .................... | .................... | ..................... | .................... | .................... | .................... |
| 8113 | MW184203.1 | Severe acute respiratory syndrome coro | .................... | .................... | ..................... | .................... | .................... | .................... |
| 8114 | MW184205.1 | Severe acute respiratory syndrome coro | .................... | .................... | ..................... | .................... | .................... | .................... |
| 8115 | MW184206.1 | Severe acute respiratory syndrome coro | .................... | .................... | ..................... | .................... | .................... | .................... |
| 8116 | MW184207.1 | Severe acute respiratory syndrome coro | .................... | .................... | ..................... | .................... | .................... | .................... |
| 8117 | MW184208.1 | Severe acute respiratory syndrome coro | .................... | .................... | ..................... | .................... | .................... | .................... |
| 8118 | MW184209.1 | Severe acute respiratory syndrome coro | .................... | .................... | ..................... | .................... | .................... | .................... |
| 8119 | MW184212.1 | Severe acute respiratory syndrome coro | .................... | .................... | ..................... | .................... | .................... | .................... |
| 8120 | MW184213.1 | Severe acute respiratory syndrome coro | .................... | .................... | ..................... | .................... | .................... | .................... |
| 8121 | MW184214.1 | Severe acute respiratory syndrome coro | .................... | .................... | ..................... | .................... | .................... | .................... |
| 8122 | MW184215.1 | Severe acute respiratory syndrome coro | .................... | .................... | ..................... | .................... | .................... | .................... |
| 8123 | MW184216.1 | Severe acute respiratory syndrome coro | .................... | .................... | ..................... | .................... | .................... | .................... |
| 8124 | MW184217.1 | Severe acute respiratory syndrome coro | .................... | .................... | ..................... | .................... | .................... | .................... |
| 8125 | MW184218.1 | Severe acute respiratory syndrome coro | .................... | .................... | ..................... | .................... | .................... | .................... |
| 8126 | MW184219.1 | Severe acute respiratory syndrome coro | .................... | .................... | ..................... | .................... | .................... | .................... |
| 8127 | MW184220.1 | Severe acute respiratory syndrome coro | .................... | .................... | ..................... | .................... | .................... | .................... |
| 8128 | MW184222.1 | Severe acute respiratory syndrome coro | .................... | .................... | ..................... | .................... | .................... | .................... |
| 8129 | MW184223.1 | Severe acute respiratory syndrome coro | .................... | .................... | ..................... | .................... | .................... | .................... |
| 8130 | MW184226.1 | Severe acute respiratory syndrome coro | .................... | .................... | ..................... | .................... | .................... | .................... |
| 8131 | MW184228.1 | Severe acute respiratory syndrome coro | .................... | .................... | ..................... | .................... | .................... | .................... |
| 8132 | MW184230.1 | Severe acute respiratory syndrome coro | .................... | .................... | ..................... | .................... | .................... | .................... |
| 8133 | MW184231.1 | Severe acute respiratory syndrome coro | .................... | .................... | ..................... | .................... | .................... | .................... |
| 8134 | MW184232.1 | Severe acute respiratory syndrome coro | .................... | .................... | ..................... | .................... | .................... | .................... |
| 8135 | MW184233.1 | Severe acute respiratory syndrome coro | .................... | .................... | ..................... | .................... | .................... | .................... |
| 8136 | MW184234.1 | Severe acute respiratory syndrome coro | .................... | .................... | ..................... | .................... | .................... | .................... |
| 8137 | MW184235.1 | Severe acute respiratory syndrome coro | .................... | .................... | ..................... | .................... | .................... | .................... |
| 8138 | MW184236.1 | Severe acute respiratory syndrome coro | .................... | .................... | ..................... | .................... | .................... | .................... |
| 8139 | MW184238.1 | Severe acute respiratory syndrome coro | .................... | .................... | ..................... | .................... | .................... | .................... |
| 8140 | MW184239.1 | Severe acute respiratory syndrome coro | .................... | .................... | ..................... | .................... | .................... | .................... |
| 8141 | MW184241.1 | Severe acute respiratory syndrome coro | .................... | .................... | ..................... | .................... | .................... | .................... |
| 8142 | MW184243.1 | Severe acute respiratory syndrome coro | .................... | .................... | ..................... | .................... | .................... | .................... |
| 8143 | MW184244.1 | Severe acute respiratory syndrome coro | .................... | .................... | ..................... | .................... | .................... | .................... |
| 8144 | MW184245.1 | Severe acute respiratory syndrome coro | .................... | .................... | ..................... | .................... | .................... | .................... |
| 8145 | MW184247.1 | Severe acute respiratory syndrome coro | .................... | .................... | ..................... | .................... | .................... | .................... |
| 8146 | MW184248.1 | Severe acute respiratory syndrome coro | .................... | .................... | ..................... | .................... | .................... | .................... |
| 8147 | MW184249.1 | Severe acute respiratory syndrome coro | .................... | .................... | ..................... | .................... | .................... | .................... |
| 8148 | MW184250.1 | Severe acute respiratory syndrome coro | .................... | .................... | ..................... | .................... | .................... | .................... |
| 8149 | MW184252.1 | Severe acute respiratory syndrome coro | .................... | .................... | ..................... | .................... | .................... | .................... |
| 8150 | MW184254.1 | Severe acute respiratory syndrome coro | .................... | .................... | ..................... | .................... | .................... | .................... |
| 8151 | MW184255.1 | Severe acute respiratory syndrome coro | .................... | .................... | ..................... | .................... | .................... | .................... |
| 8152 | MW184256.1 | Severe acute respiratory syndrome coro | .................... | .................... | ..................... | .................... | .................... | .................... |
| 8153 | MW184257.1 | Severe acute respiratory syndrome coro | .................... | .................... | ..................... | .................... | .................... | .................... |
| 8154 | MW184258.1 | Severe acute respiratory syndrome coro | .................... | .................... | ..................... | .................... | .................... | .................... |
| 8155 | MW184259.1 | Severe acute respiratory syndrome coro | .................... | .................... | ..................... | .................... | .................... | .................... |
| 8156 | MW184260.1 | Severe acute respiratory syndrome coro | .................... | .................... | ..................... | .................... | .................... | .................... |
| 8157 | MW184261.1 | Severe acute respiratory syndrome coro | .................... | .................... | ..................... | .................... | .................... | .................... |

8158 MW184263.1 |Severe acute respiratory syndrome coro ···················· ···················· ···················· ···················· ···················· ····················
8159 MW184264.1 |Severe acute respiratory syndrome coro ···················· ···················· ···················· ···················· ···················· ····················
8160 MW184266.1 |Severe acute respiratory syndrome coro ···················· ···················· ···················· ···················· ···················· ····················
8162 MW184268.1 |Severe acute respiratory syndrome coro ···················· ···················· ···················· ···················· ···················· ····················
8163 MW184269.1 |Severe acute respiratory syndrome coro ···················· ···················· ···················· ···················· ···················· ····················
8164 MW184270.1 |Severe acute respiratory syndrome coro ···················· ···················· ···················· ···················· ···················· ····················
8165 MW184271.1 |Severe acute respiratory syndrome coro ···················· ···················· ···················· ···················· ···················· ····················
8166 MW184272.1 |Severe acute respiratory syndrome coro ···················· ···················· ···················· ···················· ···················· ····················
8167 MW184273.1 |Severe acute respiratory syndrome coro ···················· ···················· ···················· ···················· ···················· ····················
8168 MW184274.1 |Severe acute respiratory syndrome coro ···················· ···················· ···················· ···················· ···················· ····················
8169 MW184275.1 |Severe acute respiratory syndrome coro ···················· ···················· ···················· ···················· ···················· ····················
8170 MW184276.1 |Severe acute respiratory syndrome coro ···················· ···················· ···················· ···················· ···················· ····················
8171 MW184277.1 |Severe acute respiratory syndrome coro ···················· ···················· ···················· ···················· ···················· ····················
8172 MW184278.1 |Severe acute respiratory syndrome coro ···················· ···················· ···················· ···················· ···················· ····················
8173 MW184279.1 |Severe acute respiratory syndrome coro ···················· ···················· ···················· ···················· ···················· ····················
8174 MW184280.1 |Severe acute respiratory syndrome coro ···················· ···················· ···················· ···················· ···················· ····················
8175 MW184281.1 |Severe acute respiratory syndrome coro ···················· ···················· ···················· ···················· ···················· ····················
8176 MW184282.1 |Severe acute respiratory syndrome coro ···················· ···················· ···················· ···················· ···················· ····················
8177 MW184283.1 |Severe acute respiratory syndrome coro ···················· ···················· ···················· ···················· ···················· ····················
8179 MW184286.1 |Severe acute respiratory syndrome coro ···················· ···················· ···················· ···················· ···················· ····················
8180 MW184288.1 |Severe acute respiratory syndrome coro ···················· ···················· ···················· ···················· ···················· ····················
8181 MW184289.1 |Severe acute respiratory syndrome coro ···················· ···················· ···················· ···················· ···················· ····················
8182 MW184290.1 |Severe acute respiratory syndrome coro ···················· ···················· ···················· ···················· ···················· ····················
8183 MW184291.1 |Severe acute respiratory syndrome coro ···················· ···················· ···················· ···················· ···················· ····················
8184 MW184293.1 |Severe acute respiratory syndrome coro ···················· ···················· ···················· ···················· ···················· ····················
8185 MW184294.1 |Severe acute respiratory syndrome coro ···················· ···················· ···················· ···················· ···················· ····················
8186 MW184295.1 |Severe acute respiratory syndrome coro ···················· ···················· ···················· ···················· ···················· ····················
8187 MW184297.1 |Severe acute respiratory syndrome coro ···················· ···················· ···················· ···················· ···················· ····················
8188 MW184298.1 |Severe acute respiratory syndrome coro ···················· ···················· ···················· ···················· ···················· ····················
8189 MW184299.1 |Severe acute respiratory syndrome coro ···················· ···················· ···················· ···················· ···················· ····················
8190 MW184300.1 |Severe acute respiratory syndrome coro ···················· ···················· ···················· ···················· ···················· ····················
8191 MW184301.1 |Severe acute respiratory syndrome coro ···················· ···················· ···················· ···················· ···················· ····················
8192 MW184302.1 |Severe acute respiratory syndrome coro ···················· ···················· ···················· ···················· ···················· ····················
8193 MW184305.1 |Severe acute respiratory syndrome coro ···················· ···················· ···················· ···················· ···················· ····················
8194 MW184306.1 |Severe acute respiratory syndrome coro ···················· ···················· ···················· ···················· ···················· ····················
8195 MW184307.1 |Severe acute respiratory syndrome coro ···················· ···················· ···················· ···················· ···················· ····················
8196 MW184308.1 |Severe acute respiratory syndrome coro ···················· ···················· ···················· ···················· ···················· ····················
8197 MW184310.1 |Severe acute respiratory syndrome coro ···················· ···················· ···················· ···················· ···················· ····················
8198 MW184311.1 |Severe acute respiratory syndrome coro ···················· ···················· ···················· ···················· ···················· ····················
8199 MW184314.1 |Severe acute respiratory syndrome coro ···················· ···················· ···················· ···················· ···················· ····················
8200 MW184315.1 |Severe acute respiratory syndrome coro ···················· ···················· ···················· ···················· ···················· ····················
8201 MW184317.1 |Severe acute respiratory syndrome coro ···················· ···················· ···················· ···················· ···················· ····················
8202 MW184318.1 |Severe acute respiratory syndrome coro ···················· ···················· ···················· ···················· ···················· ····················
8203 MW184320.1 |Severe acute respiratory syndrome coro ···················· ···················· ···················· ···················· ···················· ····················
8204 MW184321.1 |Severe acute respiratory syndrome coro ···················· ···················· ···················· ···················· ···················· ····················
8205 MW184322.1 |Severe acute respiratory syndrome coro ···················· ···················· ···················· ···················· ···················· ····················
8206 MW184323.1 |Severe acute respiratory syndrome coro ···················· ···················· ···················· ···················· ···················· ····················
8207 MW184325.1 |Severe acute respiratory syndrome coro ···················· ···················· ···················· ···················· ···················· ····················
8208 MW184326.1 |Severe acute respiratory syndrome coro ···················· ···················· ···················· ···················· ···················· ····················
8209 MW184327.1 |Severe acute respiratory syndrome coro ···················· ···················· ···················· ···················· ···················· ····················
8210 MW184328.1 |Severe acute respiratory syndrome coro ···················· ···················· ···C················ ···················· ···················· ····················
8211 MW184329.1 |Severe acute respiratory syndrome coro ···················· ···················· ···················· ···················· ···················· ····················
8212 MW184330.1 |Severe acute respiratory syndrome coro ···················· ···················· ···················· ···················· ···················· ····················
8213 MW184332.1 |Severe acute respiratory syndrome coro ···················· ···················· ···················· ···················· ···················· ····················
8214 MW184333.1 |Severe acute respiratory syndrome coro ···················· ···················· ···················· ···················· ···················· ····················
8215 MW184335.1 |Severe acute respiratory syndrome coro ···················· ···················· ···················· ···················· ···················· ····················
8216 MW184336.1 |Severe acute respiratory syndrome coro ···················· ···················· ···················· ···················· ···················· ····················
8217 MW184337.1 |Severe acute respiratory syndrome coro ···················· ···················· ···················· ···················· ···················· ····················
8218 MW184339.1 |Severe acute respiratory syndrome coro ···················· ···················· ···················· ···················· ···················· ····················
8219 MW184340.1 |Severe acute respiratory syndrome coro ···················· ···················· ···················· ···················· ···················· ····················
8220 MW184341.1 |Severe acute respiratory syndrome coro ···················· ···················· ···················· ···················· ···················· ····················
8221 MW184343.1 |Severe acute respiratory syndrome coro ···················· ···················· ···················· ···················· ···················· ····················
8222 MW184344.1 |Severe acute respiratory syndrome coro ···················· ···················· ···················· ···················· ···················· ····················
8223 MW184345.1 |Severe acute respiratory syndrome coro ···················· ···················· ···················· ···················· ···················· ····················
8224 MW184346.1 |Severe acute respiratory syndrome coro ···················· ···················· ···················· ···················· ···················· ····················
8225 MW184347.1 |Severe acute respiratory syndrome coro ···················· ···················· ···················· ···················· ···················· ····················
8226 MW184349.1 |Severe acute respiratory syndrome coro ···················· ···················· ···················· ···················· ···················· ····················
8227 MW184350.1 |Severe acute respiratory syndrome coro ···················· ···················· ···················· ···················· ···················· ····················
8228 MW184351.1 |Severe acute respiratory syndrome coro ···················· ···················· ···················· ···················· ···················· ····················
8229 MW184352.1 |Severe acute respiratory syndrome coro ···················· ···················· ···················· ···················· ···················· ····················
8230 MW184353.1 |Severe acute respiratory syndrome coro ···················· ···················· ···················· ···················· ···················· ····················
8231 MW184354.1 |Severe acute respiratory syndrome coro ···················· ···················· ···················· ···················· ···················· ····················
8232 MW184356.1 |Severe acute respiratory syndrome coro ···················· ···················· ···················· ···················· ···················· ····················
8233 MW184357.1 |Severe acute respiratory syndrome coro ···················· ···················· ···················· ···················· ···················· ····················
8234 MW184358.1 |Severe acute respiratory syndrome coro ···················· ···················· ···················· ···················· ···················· ····················
8235 MW184359.1 |Severe acute respiratory syndrome coro ···················· ···················· ···················· ···················· ···················· ····················
8236 MW184360.1 |Severe acute respiratory syndrome coro ···················· ···················· ···················· ···················· ···················· ····················
8237 MW184361.1 |Severe acute respiratory syndrome coro ···················· ···················· ···················· ···················· ···················· ····················

8238 MW184363.1 |Severe acute respiratory syndrome coro .................... .................... .................... .................... .................... ....................
8239 MW184365.1 |Severe acute respiratory syndrome coro .................... .................... .................... .................... .................... ....................
8240 MW184366.1 |Severe acute respiratory syndrome coro .................... .................... .................... .................... .................... ....................
8241 MW184367.1 |Severe acute respiratory syndrome coro .................... .................... .................... .................... .................... ....................
8242 MW184368.1 |Severe acute respiratory syndrome coro .................... .................... .................... .................... .................... ....................
8243 MW184370.1 |Severe acute respiratory syndrome coro .................... .................... .................... .................... .................... ....................
8244 MW184371.1 |Severe acute respiratory syndrome coro .................... .................... .................... .................... .................... ....................
8245 MW184372.1 |Severe acute respiratory syndrome coro .................... .................... .................... .................... .................... ....................
8246 MW184373.1 |Severe acute respiratory syndrome coro .................... .................... .................... .................... .................... ....................
8247 MW184374.1 |Severe acute respiratory syndrome coro .................... .................... .................... .................... .................... ....................
8248 MW184376.1 |Severe acute respiratory syndrome coro .................... .................... .................... .................... .................... ....................
8249 MW184377.1 |Severe acute respiratory syndrome coro .................... ......A............. .................... .................... .................... ....................
8250 MW184378.1 |Severe acute respiratory syndrome coro .................... .................... .................... .................... .................... ....................
8251 MW184379.1 |Severe acute respiratory syndrome coro .................... .................... .................... .................... .................... ....................
8252 MW184380.1 |Severe acute respiratory syndrome coro .................... .................... .................... .................... .................... ....................
8253 MW184381.1 |Severe acute respiratory syndrome coro .................... .................... .................... .................... .................... ....................
8254 MW184382.1 |Severe acute respiratory syndrome coro .................... .................... .................... .................... .................... ....................
8256 MW184384.1 |Severe acute respiratory syndrome coro .................... .................... .................... .................... .................... ....................
8257 MW184385.1 |Severe acute respiratory syndrome coro .................... .................... .................... .................... .................... ....................
8258 MW184387.1 |Severe acute respiratory syndrome coro .................... .................... .................... .................... .................... ....................
8259 MW184388.1 |Severe acute respiratory syndrome coro .................... .................... .................... .................... .................... ....................
8260 MW184389.1 |Severe acute respiratory syndrome coro .................... .................... .................... .................... .................... ....................
8261 MW184390.1 |Severe acute respiratory syndrome coro .................... .................... .................... .................... .................... ....................
8262 MW184392.1 |Severe acute respiratory syndrome coro .................... .................... .................... .................... .................... ....................
8263 MW184393.1 |Severe acute respiratory syndrome coro .................... .................... .................... .................... .................... ....................
8264 MW184394.1 |Severe acute respiratory syndrome coro .................... .................... .................... .................... .................... ....................
8265 MW184396.1 |Severe acute respiratory syndrome coro .................... .................... .................... .................... .................... ....................
8267 MW184398.1 |Severe acute respiratory syndrome coro .................... .................... .................... .................... .................... ....................
8268 MW184400.1 |Severe acute respiratory syndrome coro .................... .................... .................... .................... .................... ....................
8269 MW184401.1 |Severe acute respiratory syndrome coro .................... .................... .................... .................... .................... ....................
8270 MW184402.1 |Severe acute respiratory syndrome coro .................... .................... .................... .................... .................... ....................
8272 MW184404.1 |Severe acute respiratory syndrome coro .................... .................... .................... .................... .................... ....................
8273 MW184405.1 |Severe acute respiratory syndrome coro .................... .................... .................... .................... .................... ....................
8274 MW184406.1 |Severe acute respiratory syndrome coro .................... .................... .................... .................... .................... ....................
8275 MW184407.1 |Severe acute respiratory syndrome coro .................... .................... .................... .................... .................... ....................
8276 MW184408.1 |Severe acute respiratory syndrome coro .................... .................... .................... .................... .................... ....................
8277 MW184409.1 |Severe acute respiratory syndrome coro .................... .................... .................... .................... .................... ....................
8278 MW184410.1 |Severe acute respiratory syndrome coro .................... .................... .................... .................... .................... ....................
8279 MW184412.1 |Severe acute respiratory syndrome coro .................... .................... .................... .................... .................... ....................
8280 MW184413.1 |Severe acute respiratory syndrome coro .................... .................... .................... .................... .................... ....................
8281 MW184414.1 |Severe acute respiratory syndrome coro .................... .................... .................... .................... .................... ....................
8282 MW184415.1 |Severe acute respiratory syndrome coro .................... .................... .................... .................... .................... ....................
8283 MW184416.1 |Severe acute respiratory syndrome coro .................... .................... .................... .................... .................... ....................
8284 MW184418.1 |Severe acute respiratory syndrome coro .................... .................... .................... .................... .................... ....................
8285 MW184419.1 |Severe acute respiratory syndrome coro .................... .................... .................... .................... .................... ....................
8286 MW184420.1 |Severe acute respiratory syndrome coro .................... .................... .................... .................... .................... ....................
8287 MW184421.1 |Severe acute respiratory syndrome coro .................... .................... .................... .................... .................... ....................
8288 MW184422.1 |Severe acute respiratory syndrome coro .................... .................... .................... .................... .................... ....................
8289 MW184424.1 |Severe acute respiratory syndrome coro .................... .................... .................... .................... .................... ....................
8290 MW184425.1 |Severe acute respiratory syndrome coro .................... .................... .................... .................... .................... ....................
8291 MW184426.1 |Severe acute respiratory syndrome coro .................... .................... .................... .................... .................... ....................
8292 MW184428.1 |Severe acute respiratory syndrome coro .................... .................... .................... .................... .................... ....................
8293 MW184429.1 |Severe acute respiratory syndrome coro .................... .................... .................... .................... .................... ....................
8294 MW184430.1 |Severe acute respiratory syndrome coro .................... .................... .................... .................... .................... ....................
8295 MW184431.1 |Severe acute respiratory syndrome coro .................... .................... .................... .................... .................... ....................
8296 MW184432.1 |Severe acute respiratory syndrome coro .................... .................... .................... .................... .................... ....................
8297 MW184433.1 |Severe acute respiratory syndrome coro .................... .................... .................... .................... .................... ....................
8298 MW184434.1 |Severe acute respiratory syndrome coro .................... .................... .................... .................... .................... ....................
8299 MW184435.1 |Severe acute respiratory syndrome coro .................... .................... .................... .................... .................... ....................
8300 MW184437.1 |Severe acute respiratory syndrome coro .................... .................... .................... .................... .................... ....................
8301 MW184438.1 |Severe acute respiratory syndrome coro .................... .................... .................... .................... .................... ....................
8302 MW184439.1 |Severe acute respiratory syndrome coro .................... .................... .................... .................... .................... ....................
8304 MW184442.1 |Severe acute respiratory syndrome coro .................... .................... .................... .................... .................... ....................
8305 MW184444.1 |Severe acute respiratory syndrome coro .................... .................... .................... .................... .................... ....................
8306 MW184446.1 |Severe acute respiratory syndrome coro .................... .................... .................... .................... .................... ....................
8307 MW184447.1 |Severe acute respiratory syndrome coro .................... .................... .................... .................... .................... ....................
8308 MW184451.1 |Severe acute respiratory syndrome coro .................... .................... .................... .................... .................... ....................
8309 MW184452.1 |Severe acute respiratory syndrome coro .................... .................... .................... .................... .................... ....................
8310 MW184455.1 |Severe acute respiratory syndrome coro .................... .................... .................... .................... .................... ....................
8311 MW184459.1 |Severe acute respiratory syndrome coro .................... .................... .................... .................... .................... ....................
8313 MW184461.1 |Severe acute respiratory syndrome coro .................... .................... .................... .................... .................... ....................
8314 MW184462.1 |Severe acute respiratory syndrome coro .................... .................... .................... .................... .................... ....................
8315 MW184463.1 |Severe acute respiratory syndrome coro .................... .................... .................... .................... .................... ....................
8316 MW184467.1 |Severe acute respiratory syndrome coro .................... .................... .................... .................... .................... ....................
8317 MW184468.1 |Severe acute respiratory syndrome coro .................... .................... .................... .................... .................... ....................
8318 MW184470.1 |Severe acute respiratory syndrome coro .................... .................... .................... .................... .................... ....................
8319 MW184471.1 |Severe acute respiratory syndrome coro .................... .................... .................... .................... .................... ....................
8320 MW184473.1 |Severe acute respiratory syndrome coro .................... .................... .................... .................... .................... ....................

| | | | | | | | | |
|---|---|---|---|---|---|---|---|---|
| 8321 | MW184474.1 | \|Severe acute respiratory syndrome coro | ···················· | ···················· | ····················· | ···················· | ···················· | ···················· |
| 8322 | MW184475.1 | \|Severe acute respiratory syndrome coro | ···················· | ···················· | ····················· | ···················· | ···················· | ···················· |
| 8323 | MW184476.1 | \|Severe acute respiratory syndrome coro | ···················· | ···················· | ····················· | ···················· | ···················· | ···················· |
| 8324 | MW184479.1 | \|Severe acute respiratory syndrome coro | ···················· | ···················· | ····················· | ···················· | ···················· | ···················· |
| 8325 | MW184480.1 | \|Severe acute respiratory syndrome coro | ···················· | ···················· | ····················· | ···················· | ···················· | ···················· |
| 8326 | MW184481.1 | \|Severe acute respiratory syndrome coro | ···················· | ···················· | ····················· | ···················· | ···················· | ···················· |
| 8327 | MW184482.1 | \|Severe acute respiratory syndrome coro | ···················· | ···················· | ····················· | ···················· | ···················· | ···················· |
| 8328 | MW184483.1 | \|Severe acute respiratory syndrome coro | ···················· | ···················· | ····················· | ···················· | ···················· | ···················· |
| 8329 | MW184484.1 | \|Severe acute respiratory syndrome coro | ···················· | ···················· | ····················· | ···················· | ···················· | ···················· |
| 8330 | MW184485.1 | \|Severe acute respiratory syndrome coro | ···················· | ···················· | ····················· | ···················· | ···················· | ···················· |
| 8332 | MW184487.1 | \|Severe acute respiratory syndrome coro | ···················· | ···················· | ····················· | ···················· | ···················· | ···················· |
| 8333 | MW184489.1 | \|Severe acute respiratory syndrome coro | ···················· | ···················· | ····················· | ···················· | ···················· | ···················· |
| 8334 | MW184491.1 | \|Severe acute respiratory syndrome coro | ···················· | ···················· | ····················· | ···················· | ···················· | ···················· |
| 8335 | MW184493.1 | \|Severe acute respiratory syndrome coro | ···················· | ···················· | ····················· | ···················· | ···················· | ···················· |
| 8336 | MW184495.1 | \|Severe acute respiratory syndrome coro | ···················· | ···················· | ····················· | ···················· | ···················· | ···················· |
| 8337 | MW184496.1 | \|Severe acute respiratory syndrome coro | ···················· | ···················· | ····················· | ···················· | ···················· | ···················· |
| 8338 | MW184497.1 | \|Severe acute respiratory syndrome coro | ···················· | ···················· | ····················· | ···················· | ···················· | ···················· |
| 8339 | MW184498.1 | \|Severe acute respiratory syndrome coro | ···················· | ···················· | ····················· | ···················· | ···················· | ···················· |
| 8340 | MW184500.1 | \|Severe acute respiratory syndrome coro | ···················· | ···················· | ····················· | ···················· | ···················· | ···················· |
| 8341 | MW184503.1 | \|Severe acute respiratory syndrome coro | ···················· | ···················· | ····················· | ···················· | ···················· | ···················· |
| 8342 | MW184504.1 | \|Severe acute respiratory syndrome coro | ···················· | ···················· | ····················· | ···················· | ···················· | ···················· |
| 8343 | MW184505.1 | \|Severe acute respiratory syndrome coro | ···················· | ···················· | ····················· | ···················· | ···················· | ···················· |
| 8344 | MW184509.1 | \|Severe acute respiratory syndrome coro | ···················· | ···················· | ····················· | ···················· | ···················· | ···················· |
| 8345 | MW184510.1 | \|Severe acute respiratory syndrome coro | ···················· | ···················· | ····················· | ···················· | ···················· | ···················· |
| 8346 | MW184511.1 | \|Severe acute respiratory syndrome coro | ···················· | ···················· | ····················· | ···················· | ···················· | ···················· |
| 8347 | MW184513.1 | \|Severe acute respiratory syndrome coro | ···················· | ···················· | ····················· | ···················· | ···················· | ···················· |
| 8348 | MW184514.1 | \|Severe acute respiratory syndrome coro | ···················· | ···················· | ····················· | ···················· | ···················· | ···················· |
| 8349 | MW184518.1 | \|Severe acute respiratory syndrome coro | ···················· | ···················· | ····················· | ···················· | ···················· | ···················· |
| 8350 | MW184519.1 | \|Severe acute respiratory syndrome coro | ···················· | ···················· | ····················· | ···················· | ···················· | ···················· |
| 8351 | MW184522.1 | \|Severe acute respiratory syndrome coro | ···················· | ···················· | ····················· | ···················· | ···················· | ···················· |
| 8352 | MW184523.1 | \|Severe acute respiratory syndrome coro | ···················· | ···················· | ····················· | ···················· | ···················· | ···················· |
| 8353 | MW184524.1 | \|Severe acute respiratory syndrome coro | ···················· | ···················· | ····················· | ···················· | ···················· | ···················· |
| 8354 | MW184525.1 | \|Severe acute respiratory syndrome coro | ···················· | ···················· | ····················· | ···················· | ···················· | ···················· |
| 8355 | MW184526.1 | \|Severe acute respiratory syndrome coro | ···················· | ···················· | ····················· | ···················· | ···················· | ···················· |
| 8356 | MW184527.1 | \|Severe acute respiratory syndrome coro | ···················· | ···················· | ····················· | ···················· | ···················· | ···················· |
| 8357 | MW184528.1 | \|Severe acute respiratory syndrome coro | ···················· | ···················· | ····················· | ···················· | ···················· | ···················· |
| 8358 | MW184529.1 | \|Severe acute respiratory syndrome coro | ···················· | ···················· | ····················· | ···················· | ···················· | ···················· |
| 8359 | MW184530.1 | \|Severe acute respiratory syndrome coro | ···················· | ···················· | ····················· | ···················· | ···················· | ···················· |
| 8360 | MW184531.1 | \|Severe acute respiratory syndrome coro | ···················· | ···················· | ····················· | ···················· | ···················· | ···················· |
| 8361 | MW184532.1 | \|Severe acute respiratory syndrome coro | ···················· | ···················· | ····················· | ···················· | ···················· | ···················· |
| 8362 | MW184534.1 | \|Severe acute respiratory syndrome coro | ···················· | ···················· | ····················· | ···················· | ···················· | ···················· |
| 8363 | MW184535.1 | \|Severe acute respiratory syndrome coro | ···················· | ···················· | ····················· | ···················· | ···················· | ···················· |
| 8364 | MW184536.1 | \|Severe acute respiratory syndrome coro | ···················· | ···················· | ····················· | ···················· | ···················· | ···················· |
| 8365 | MW184537.1 | \|Severe acute respiratory syndrome coro | ···················· | ···················· | ····················· | ···················· | ···················· | ···················· |
| 8366 | MW184538.1 | \|Severe acute respiratory syndrome coro | ···················· | ···················· | ····················· | ···················· | ···················· | ···················· |
| 8367 | MW184539.1 | \|Severe acute respiratory syndrome coro | ···················· | ···················· | ····················· | ···················· | ···················· | ···················· |
| 8368 | MW184540.1 | \|Severe acute respiratory syndrome coro | ···················· | ···················· | ····················· | ···················· | ···················· | ···················· |
| 8369 | MW184541.1 | \|Severe acute respiratory syndrome coro | ···················· | ·····A·············· | ····················· | ···················· | ···················· | ···················· |
| 8370 | MW184542.1 | \|Severe acute respiratory syndrome coro | ···················· | ···················· | ····················· | ···················· | ···················· | ···················· |
| 8371 | MW184543.1 | \|Severe acute respiratory syndrome coro | ···················· | ···················· | ····················· | ···················· | ···················· | ···················· |
| 8372 | MW184544.1 | \|Severe acute respiratory syndrome coro | ···················· | ···················· | ····················· | ···················· | ···················· | ···················· |
| 8373 | MW184545.1 | \|Severe acute respiratory syndrome coro | ···················· | ···················· | ····················· | ···················· | ···················· | ···················· |
| 8374 | MW184547.1 | \|Severe acute respiratory syndrome coro | ···················· | ···················· | ····················· | ···················· | ···················· | ···················· |
| 8375 | MW184548.1 | \|Severe acute respiratory syndrome coro | ···················· | ···················· | ····················· | ···················· | ···················· | ···················· |
| 8376 | MW184550.1 | \|Severe acute respiratory syndrome coro | ···················· | ···················· | ····················· | ···················· | ···················· | ···················· |
| 8377 | MW184551.1 | \|Severe acute respiratory syndrome coro | ···················· | ···················· | ····················· | ···················· | ···················· | ···················· |
| 8378 | MW184552.1 | \|Severe acute respiratory syndrome coro | ···················· | ···················· | ····················· | ···················· | ···················· | ···················· |
| 8379 | MW184553.1 | \|Severe acute respiratory syndrome coro | ···················· | ···················· | ····················· | ···················· | ···················· | ···················· |
| 8380 | MW184555.1 | \|Severe acute respiratory syndrome coro | ···················· | ···················· | ····················· | ···················· | ···················· | ···················· |
| 8381 | MW184557.1 | \|Severe acute respiratory syndrome coro | ···················· | ···················· | ····················· | ···················· | ···················· | ···················· |
| 8382 | MW184558.1 | \|Severe acute respiratory syndrome coro | ···················· | ···················· | ····················· | ···················· | ···················· | ···················· |
| 8384 | MW184561.1 | \|Severe acute respiratory syndrome coro | ···················· | ···················· | ····················· | ···················· | ···················· | ···················· |
| 8385 | MW184562.1 | \|Severe acute respiratory syndrome coro | ···················· | ···················· | ····················· | ···················· | ···················· | ···················· |
| 8386 | MW184563.1 | \|Severe acute respiratory syndrome coro | ···················· | ···················· | ····················· | ···················· | ···················· | ···················· |
| 8387 | MW184564.1 | \|Severe acute respiratory syndrome coro | ···················· | ···················· | ····················· | ···················· | ···················· | ···················· |
| 8388 | MW184565.1 | \|Severe acute respiratory syndrome coro | ···················· | ···················· | ····················· | ···················· | ···················· | ···················· |
| 8389 | MW184566.1 | \|Severe acute respiratory syndrome coro | ···················· | ···················· | ····················· | ···················· | ···················· | ···················· |
| 8390 | MW184567.1 | \|Severe acute respiratory syndrome coro | ···················· | ···················· | ····················· | ···················· | ···················· | ···················· |
| 8391 | MW184568.1 | \|Severe acute respiratory syndrome coro | ···················· | ···················· | ····················· | ···················· | ···················· | ···················· |
| 8392 | MW184569.1 | \|Severe acute respiratory syndrome coro | ···················· | ···················· | ····················· | ···················· | ···················· | ···················· |
| 8393 | MW184570.1 | \|Severe acute respiratory syndrome coro | ···················· | ···················· | ····················· | ···················· | ···················· | ···················· |
| 8394 | MW184571.1 | \|Severe acute respiratory syndrome coro | ···················· | ···················· | ····················· | ···················· | ···················· | ···················· |
| 8395 | MW184572.1 | \|Severe acute respiratory syndrome coro | ···················· | ···················· | ····················· | ···················· | ···················· | ···················· |
| 8396 | MW184574.1 | \|Severe acute respiratory syndrome coro | ···················· | ···················· | ····················· | ···················· | ···················· | ···················· |
| 8397 | MW184575.1 | \|Severe acute respiratory syndrome coro | ···················· | ···················· | ····················· | ···················· | ···················· | ···················· |
| 8398 | MW184576.1 | \|Severe acute respiratory syndrome coro | ···················· | ···················· | ····················· | ···················· | ···················· | ···················· |
| 8399 | MW184577.1 | \|Severe acute respiratory syndrome coro | ···················· | ···················· | ····················· | ···················· | ···················· | ···················· |
| 8400 | MW184578.1 | \|Severe acute respiratory syndrome coro | ···················· | ···················· | ····················· | ···················· | ···················· | ···················· |

| | | | | | | | | |
|---|---|---|---|---|---|---|---|---|
| 8401 | MW184579.1 | \|Severe acute respiratory syndrome coro | ···················· | ···················· | ···················· | ···················· | ···················· | ···················· |
| 8402 | MW184582.1 | \|Severe acute respiratory syndrome coro | ···················· | ···················· | ···················· | ···················· | ···················· | ···················· |
| 8403 | MW184583.1 | \|Severe acute respiratory syndrome coro | ···················· | ···················· | ···················· | ···················· | ···················· | ···················· |
| 8404 | MW184584.1 | \|Severe acute respiratory syndrome coro | ···················· | ···················· | ···················· | ···················· | ···················· | ···················· |
| 8405 | MW184585.1 | \|Severe acute respiratory syndrome coro | ···················· | ···················· | ···················· | ···················· | ···················· | ···················· |
| 8407 | MW184589.1 | \|Severe acute respiratory syndrome coro | ···················· | ···················· | ···················· | ···················· | ···················· | ···················· |
| 8408 | MW184590.1 | \|Severe acute respiratory syndrome coro | ···················· | ···················· | ···················· | ···················· | ···················· | ···················· |
| 8409 | MW184591.1 | \|Severe acute respiratory syndrome coro | ···················· | ···················· | ···················· | ···················· | ···················· | ···················· |
| 8410 | MW184592.1 | \|Severe acute respiratory syndrome coro | ···················· | ···················· | ···················· | ···················· | ···················· | ···················· |
| 8411 | MW184594.1 | \|Severe acute respiratory syndrome coro | ···················· | ···················· | ···················· | ···················· | ···················· | ···················· |
| 8412 | MW184595.1 | \|Severe acute respiratory syndrome coro | ···················· | ···················· | ···················· | ···················· | ···················· | ···················· |
| 8413 | MW184598.1 | \|Severe acute respiratory syndrome coro | ···················· | ···················· | ···················· | ···················· | ···················· | ···················· |
| 8414 | MW184599.1 | \|Severe acute respiratory syndrome coro | ···················· | ···················· | ···················· | ···················· | ···················· | ···················· |
| 8415 | MW184601.1 | \|Severe acute respiratory syndrome coro | ···················· | ···················· | ···················· | ···················· | ···················· | ···················· |
| 8416 | MW184603.1 | \|Severe acute respiratory syndrome coro | ···················· | ···················· | ···················· | ···················· | ···················· | ···················· |
| 8417 | MW184604.1 | \|Severe acute respiratory syndrome coro | ···················· | ···················· | ···················· | ···················· | ···················· | ···················· |
| 8418 | MW184605.1 | \|Severe acute respiratory syndrome coro | ···················· | ···················· | ···················· | ···················· | ···················· | ···················· |
| 8419 | MW184606.1 | \|Severe acute respiratory syndrome coro | ···················· | ···················· | ···················· | ···················· | ···················· | ···················· |
| 8421 | MW184609.1 | \|Severe acute respiratory syndrome coro | ···················· | ···················· | ···················· | ···················· | ···················· | ···················· |
| 8422 | MW184610.1 | \|Severe acute respiratory syndrome coro | ···················· | ···················· | ···················· | ···················· | ···················· | ···················· |
| 8423 | MW184611.1 | \|Severe acute respiratory syndrome coro | ···················· | ···················· | ···················· | ···················· | ···················· | ···················· |
| 8424 | MW184612.1 | \|Severe acute respiratory syndrome coro | ···················· | ···················· | ···················· | ···················· | ···················· | ···················· |
| 8426 | MW184614.1 | \|Severe acute respiratory syndrome coro | ···················· | ···················· | ···················· | ···················· | ···················· | ···················· |
| 8427 | MW184616.1 | \|Severe acute respiratory syndrome coro | ···················· | ···················· | ···················· | ···················· | ···················· | ···················· |
| 8428 | MW184617.1 | \|Severe acute respiratory syndrome coro | ···················· | ···················· | ···················· | ···················· | ···················· | ···················· |
| 8429 | MW184618.1 | \|Severe acute respiratory syndrome coro | ···················· | ···················· | ···················· | ···················· | ···················· | ···················· |
| 8430 | MW184621.1 | \|Severe acute respiratory syndrome coro | ···················· | ···················· | ···················· | ···················· | ···················· | ···················· |
| 8431 | MW184622.1 | \|Severe acute respiratory syndrome coro | ···················· | ···················· | ···················· | ···················· | ···················· | ···················· |
| 8432 | MW184623.1 | \|Severe acute respiratory syndrome coro | ···················· | ···················· | ···················· | ···················· | ···················· | ···················· |
| 8433 | MW184624.1 | \|Severe acute respiratory syndrome coro | ···················· | ···················· | ···················· | ···················· | ···················· | ···················· |
| 8434 | MW184625.1 | \|Severe acute respiratory syndrome coro | ···················· | ···················· | ···················· | ···················· | ···················· | ···················· |
| 8435 | MW184626.1 | \|Severe acute respiratory syndrome coro | ···················· | ···················· | ···················· | ···················· | ···················· | ···················· |
| 8436 | MW184627.1 | \|Severe acute respiratory syndrome coro | ···················· | ···················· | ···················· | ···················· | ···················· | ···················· |
| 8437 | MW184629.1 | \|Severe acute respiratory syndrome coro | ···················· | ···················· | ···················· | ···················· | ···················· | ···················· |
| 8439 | MW184632.1 | \|Severe acute respiratory syndrome coro | ···················· | ···················· | ···················· | ···················· | ···················· | ···················· |
| 8440 | MW184633.1 | \|Severe acute respiratory syndrome coro | ···················· | ···················· | ···················· | ···················· | ···················· | ···················· |
| 8441 | MW184634.1 | \|Severe acute respiratory syndrome coro | ···················· | ···················· | ···················· | ···················· | ···················· | ···················· |
| 8442 | MW184635.1 | \|Severe acute respiratory syndrome coro | ···················· | ···················· | ···················· | ···················· | ···················· | ···················· |
| 8443 | MW184636.1 | \|Severe acute respiratory syndrome coro | ···················· | ···················· | ···················· | ···················· | ···················· | ···················· |
| 8444 | MW184637.1 | \|Severe acute respiratory syndrome coro | ···················· | ···················· | ···················· | ···················· | ···················· | ···················· |
| 8445 | MW184638.1 | \|Severe acute respiratory syndrome coro | ···················· | ···················· | ···················· | ···················· | ···················· | ···················· |
| 8446 | MW184639.1 | \|Severe acute respiratory syndrome coro | ···················· | ···················· | ···················· | ···················· | ···················· | ···················· |
| 8447 | MW184640.1 | \|Severe acute respiratory syndrome coro | ···················· | ···················· | ···················· | ···················· | ···················· | ···················· |
| 8448 | MW184641.1 | \|Severe acute respiratory syndrome coro | ···················· | ···················· | ···················· | ···················· | ···················· | ···················· |
| 8449 | MW184642.1 | \|Severe acute respiratory syndrome coro | ···················· | ···················· | ···················· | ···················· | ···················· | ···················· |
| 8450 | MW184643.1 | \|Severe acute respiratory syndrome coro | ···················· | ···················· | ···················· | ···················· | ···················· | ···················· |
| 8452 | MW184646.1 | \|Severe acute respiratory syndrome coro | ···················· | ···················· | ···················· | ···················· | ···················· | ···················· |
| 8453 | MW184647.1 | \|Severe acute respiratory syndrome coro | ···················· | ···················· | ···················· | ···················· | ···················· | ···················· |
| 8454 | MW184648.1 | \|Severe acute respiratory syndrome coro | ···················· | ···················· | ···················· | ···················· | ···················· | ···················· |
| 8456 | MW184650.1 | \|Severe acute respiratory syndrome coro | ···················· | ···················· | ···················· | ···················· | ···················· | ···················· |
| 8457 | MW184652.1 | \|Severe acute respiratory syndrome coro | ···················· | ···················· | ···················· | ···················· | ···················· | ···················· |
| 8458 | MW184653.1 | \|Severe acute respiratory syndrome coro | ···················· | ···················· | ···················· | ···················· | ···················· | ···················· |
| 8459 | MW184654.1 | \|Severe acute respiratory syndrome coro | ···················· | ···················· | ···················· | ···················· | ···················· | ···················· |
| 8460 | MW184655.1 | \|Severe acute respiratory syndrome coro | ···················· | ···················· | ···················· | ···················· | ···················· | ···················· |
| 8461 | MW184657.1 | \|Severe acute respiratory syndrome coro | ···················· | ···················· | ···················· | ···················· | ···················· | ···················· |
| 8462 | MW184658.1 | \|Severe acute respiratory syndrome coro | ···················· | ···················· | ···················· | ···················· | ···················· | ···················· |
| 8463 | MW184659.1 | \|Severe acute respiratory syndrome coro | ···················· | ···················· | ···················· | ···················· | ···················· | ···················· |
| 8464 | MW184661.1 | \|Severe acute respiratory syndrome coro | ···················· | ·····A·············· | ···················· | ···················· | ···················· | ···················· |
| 8465 | MW184662.1 | \|Severe acute respiratory syndrome coro | ···················· | ···················· | ···················· | ···················· | ···················· | ···················· |
| 8466 | MW184663.1 | \|Severe acute respiratory syndrome coro | ···················· | ···················· | ···················· | ···················· | ···················· | ···················· |
| 8467 | MW184664.1 | \|Severe acute respiratory syndrome coro | ···················· | ···················· | ···················· | ···················· | ···················· | ···················· |
| 8468 | MW184665.1 | \|Severe acute respiratory syndrome coro | ···················· | ···················· | ···················· | ···················· | ···················· | ···················· |
| 8469 | MW184666.1 | \|Severe acute respiratory syndrome coro | ···················· | ···················· | ···················· | ···················· | ···················· | ···················· |
| 8470 | MW184668.1 | \|Severe acute respiratory syndrome coro | ···················· | ···················· | ···················· | ···················· | ···················· | ···················· |
| 8471 | MW184670.1 | \|Severe acute respiratory syndrome coro | ···················· | ···················· | ···················· | ···················· | ···················· | ···················· |
| 8472 | MW184671.1 | \|Severe acute respiratory syndrome coro | ···················· | ···················· | ···················· | ···················· | ···················· | ···················· |
| 8473 | MW184673.1 | \|Severe acute respiratory syndrome coro | ···················· | ···················· | ···················· | ···················· | ···················· | ···················· |
| 8474 | MW184674.1 | \|Severe acute respiratory syndrome coro | ···················· | ···················· | ···················· | ···················· | ···················· | ···················· |
| 8475 | MW184675.1 | \|Severe acute respiratory syndrome coro | ···················· | ···················· | ···················· | ···················· | ···················· | ···················· |
| 8476 | MW184676.1 | \|Severe acute respiratory syndrome coro | ···················· | ···················· | ···················· | ···················· | ···················· | ···················· |
| 8477 | MW184677.1 | \|Severe acute respiratory syndrome coro | ···················· | ···················· | ···················· | ···················· | ···················· | ···················· |
| 8478 | MW184678.1 | \|Severe acute respiratory syndrome coro | ···················· | ···················· | ···················· | ···················· | ···················· | ···················· |
| 8479 | MW184679.1 | \|Severe acute respiratory syndrome coro | ···················· | ···················· | ···················· | ···················· | ···················· | ···················· |
| 8480 | MW184680.1 | \|Severe acute respiratory syndrome coro | ···················· | ···················· | ···················· | ···················· | ···················· | ···················· |
| 8481 | MW184683.1 | \|Severe acute respiratory syndrome coro | ···················· | ···················· | ···················· | ···················· | ···················· | ···················· |
| 8482 | MW184689.1 | \|Severe acute respiratory syndrome coro | ···················· | ···················· | ···················· | ···················· | ···················· | ···················· |
| 8483 | MW184690.1 | \|Severe acute respiratory syndrome coro | ···················· | ···················· | ···················· | ···················· | ···················· | ···················· |
| 8484 | MW184691.1 | \|Severe acute respiratory syndrome coro | ···················· | ···················· | ···················· | ···················· | ···················· | ···················· |

```
8485 MW184692.1 |Severe acute respiratory syndrome coro    ····················  ····················  ·····················  ····················  ····················  ····················
8486 MW184694.1 |Severe acute respiratory syndrome coro    ····················  ····················  ·····················  ····················  ····················  ····················
8487 MW184696.1 |Severe acute respiratory syndrome coro    ····················  ····················  ·····················  ····················  ····················  ····················
8488 MW184697.1 |Severe acute respiratory syndrome coro    ····················  ····················  ·····················  ····················  ····················  ····················
8489 MW184699.1 |Severe acute respiratory syndrome coro    ····················  ····················  ·····················  ····················  ····················  ····················
8490 MW184700.1 |Severe acute respiratory syndrome coro    ····················  ····················  ·····················  ····················  ····················  ····················
8491 MW184702.1 |Severe acute respiratory syndrome coro    ····················  ····················  ·····················  ····················  ····················  ····················
8492 MW184703.1 |Severe acute respiratory syndrome coro    ····················  ····················  ·····················  ····················  ····················  ····················
8493 MW184704.1 |Severe acute respiratory syndrome coro    ····················  ····················  ·····················  ····················  ····················  ····················
8494 MW184705.1 |Severe acute respiratory syndrome coro    ····················  ····················  ·····················  ····················  ····················  ····················
8495 MW184706.1 |Severe acute respiratory syndrome coro    ····················  ····················  ·····················  ····················  ····················  ····················
8496 MW184707.1 |Severe acute respiratory syndrome coro    ····················  ····················  ·····················  ····················  ····················  ····················
8497 MW184709.1 |Severe acute respiratory syndrome coro    ····················  ····················  ·····················  ····················  ····················  ····················
8498 MW184710.1 |Severe acute respiratory syndrome coro    ····················  ····················  ·····················  ····················  ····················  ····················
8499 MW184711.1 |Severe acute respiratory syndrome coro    ····················  ····················  ·····················  ····················  ····················  ····················
8500 MW184712.1 |Severe acute respiratory syndrome coro    ····················  ····················  ·····················  ····················  ····················  ····················
8501 MW184713.1 |Severe acute respiratory syndrome coro    ····················  ····················  ·····················  ····················  ····················  ····················
8502 MW184714.1 |Severe acute respiratory syndrome coro    ····················  ····················  ·····················  ····················  ····················  ····················
8503 MW184716.1 |Severe acute respiratory syndrome coro    ····················  ····················  ·····················  ····················  ····················  ····················
8504 MW184718.1 |Severe acute respiratory syndrome coro    ····················  ····················  ·····················  ····················  ····················  ····················
8505 MW184719.1 |Severe acute respiratory syndrome coro    ····················  ····················  ·····················  ····················  ····················  ····················
8506 MW184720.1 |Severe acute respiratory syndrome coro    ····················  ····················  ·····················  ····················  ····················  ····················
8507 MW184723.1 |Severe acute respiratory syndrome coro    ····················  ····················  ·····················  ····················  ····················  ····················
8508 MW184725.1 |Severe acute respiratory syndrome coro    ····················  ····················  ·····················  ····················  ····················  ····················
8509 MW184727.1 |Severe acute respiratory syndrome coro    ····················  ······A·············  ·····················  ····················  ····················  ····················
8510 MW184729.1 |Severe acute respiratory syndrome coro    ····················  ····················  ·····················  ····················  ····················  ····················
8511 MW184731.1 |Severe acute respiratory syndrome coro    ····················  ····················  ·····················  ····················  ····················  ····················
8512 MW184732.1 |Severe acute respiratory syndrome coro    ····················  ····················  ·····················  ····················  ····················  ····················
8513 MW184733.1 |Severe acute respiratory syndrome coro    ····················  ····················  ·····················  ····················  ····················  ····················
8514 MW184735.1 |Severe acute respiratory syndrome coro    ····················  ····················  ·····················  ····················  ····················  ····················
8515 MW184737.1 |Severe acute respiratory syndrome coro    ····················  ····················  ·····················  ····················  ····················  ····················
8516 MW184739.1 |Severe acute respiratory syndrome coro    ····················  ····················  ·····················  ····················  ····················  ····················
8517 MW184740.1 |Severe acute respiratory syndrome coro    ····················  ····················  ·····················  ····················  ····················  ····················
8518 MW184741.1 |Severe acute respiratory syndrome coro    ····················  ····················  ·····················  ····················  ····················  ····················
8519 MW184743.1 |Severe acute respiratory syndrome coro    ····················  ····················  ·····················  ····················  ····················  ····················
8520 MW184745.1 |Severe acute respiratory syndrome coro    ····················  ····················  ·····················  ····················  ····················  ····················
8521 MW184747.1 |Severe acute respiratory syndrome coro    ····················  ····················  ·····················  ····················  ····················  ····················
8522 MW184748.1 |Severe acute respiratory syndrome coro    ····················  ····················  ·····················  ····················  ····················  ····················
8523 MW184749.1 |Severe acute respiratory syndrome coro    ····················  ····················  ·····················  ····················  ····················  ····················
8524 MW184750.1 |Severe acute respiratory syndrome coro    ····················  ····················  ·····················  ····················  ····················  ····················
8525 MW184751.1 |Severe acute respiratory syndrome coro    ····················  ····················  ···C·················  ····················  ····················  ····················
8526 MW184753.1 |Severe acute respiratory syndrome coro    ····················  ····················  ·····················  ····················  ····················  ····················
8527 MW184754.1 |Severe acute respiratory syndrome coro    ····················  ····················  ·····················  ····················  ····················  ····················
8528 MW184755.1 |Severe acute respiratory syndrome coro    ····················  ····················  ·····················  ····················  ····················  ····················
8529 MW184756.1 |Severe acute respiratory syndrome coro    ····················  ····················  ·····················  ····················  ····················  ····················
8530 MW184757.1 |Severe acute respiratory syndrome coro    ····················  ····················  ·····················  ····················  ····················  ····················
8531 MW184759.1 |Severe acute respiratory syndrome coro    ····················  ····················  ·····················  ····················  ····················  ····················
8532 MW184760.1 |Severe acute respiratory syndrome coro    ····················  ····················  ·····················  ····················  ····················  ····················
8533 MW184761.1 |Severe acute respiratory syndrome coro    ····················  ····················  ·····················  ····················  ····················  ····················
8534 MW184762.1 |Severe acute respiratory syndrome coro    ····················  ····················  ·····················  ····················  ····················  ····················
8535 MW184763.1 |Severe acute respiratory syndrome coro    ····················  ····················  ·····················  ····················  ····················  ····················
8536 MW184766.1 |Severe acute respiratory syndrome coro    ····················  ····················  ·····················  ····················  ····················  ····················
8537 MW184767.1 |Severe acute respiratory syndrome coro    ····················  ····················  ·····················  ····················  ····················  ····················
8538 MW184768.1 |Severe acute respiratory syndrome coro    ····················  ····················  ·····················  ····················  ····················  ····················
8539 MW184770.1 |Severe acute respiratory syndrome coro    ····················  ····················  ·····················  ····················  ····················  ····················
8540 MW184771.1 |Severe acute respiratory syndrome coro    ····················  ····················  ·····················  ····················  ····················  ····················
8541 MW184772.1 |Severe acute respiratory syndrome coro    ····················  ····················  ·····················  ····················  ····················  ····················
8542 MW184775.1 |Severe acute respiratory syndrome coro    ····················  ····················  ·····················  ····················  ····················  ····················
8543 MW184776.1 |Severe acute respiratory syndrome coro    ····················  ····················  ·····················  ····················  ····················  ····················
8544 MW184777.1 |Severe acute respiratory syndrome coro    ····················  ····················  ·····················  ····················  ····················  ····················
8545 MW184780.1 |Severe acute respiratory syndrome coro    ····················  ····················  ·····················  ····················  ····················  ····················
8546 MW184781.1 |Severe acute respiratory syndrome coro    ····················  ····················  ·····················  ····················  ····················  ····················
8547 MW184782.1 |Severe acute respiratory syndrome coro    ····················  ····················  ·····················  ····················  ····················  ····················
8548 MW184783.1 |Severe acute respiratory syndrome coro    ····················  ····················  ·····················  ····················  ····················  ····················
8549 MW184784.1 |Severe acute respiratory syndrome coro    ····················  ····················  ·····················  ····················  ····················  ····················
8550 MW184785.1 |Severe acute respiratory syndrome coro    ····················  ····················  ·····················  ····················  ····················  ····················
8551 MW184786.1 |Severe acute respiratory syndrome coro    ····················  ····················  ·····················  ····················  ····················  ····················
8552 MW184787.1 |Severe acute respiratory syndrome coro    ····················  ····················  ·····················  ····················  ····················  ····················
8553 MW184788.1 |Severe acute respiratory syndrome coro    ····················  ····················  ·····················  ····················  ····················  ····················
8554 MW184790.1 |Severe acute respiratory syndrome coro    ····················  ····················  ·····················  ····················  ····················  ····················
8555 MW184791.1 |Severe acute respiratory syndrome coro    ····················  ····················  ·····················  ····················  ····················  ····················
8556 MW184792.1 |Severe acute respiratory syndrome coro    ····················  ····················  ·····················  ····················  ····················  ····················
8557 MW184793.1 |Severe acute respiratory syndrome coro    ····················  ····················  ·····················  ····················  ····················  ····················
8558 MW184794.1 |Severe acute respiratory syndrome coro    ····················  ····················  ·····················  ····················  ····················  ····················
8559 MW184795.1 |Severe acute respiratory syndrome coro    ····················  ····················  ·····················  ····················  ····················  ····················
8560 MW184796.1 |Severe acute respiratory syndrome coro    ····················  ····················  ·····················  ····················  ····················  ····················
8561 MW184797.1 |Severe acute respiratory syndrome coro    ····················  ····················  ·····················  ····················  ····················  ····················
8562 MW184798.1 |Severe acute respiratory syndrome coro    ····················  ····················  ·····················  ····················  ····················  ····················
```

| | | | | | | | | |
|---|---|---|---|---|---|---|---|---|
| 8563 | MW184799.1 | \|Severe acute respiratory syndrome coro | .................... | .................... | .................... | .................... | .................... | .................... |
| 8564 | MW184801.1 | \|Severe acute respiratory syndrome coro | .................... | .................... | .................... | .................... | .................... | .................... |
| 8565 | MW184802.1 | \|Severe acute respiratory syndrome coro | .................... | .................... | .................... | .................... | .................... | .................... |
| 8566 | MW184803.1 | \|Severe acute respiratory syndrome coro | .................... | .................... | .................... | .................... | .................... | .................... |
| 8567 | MW184804.1 | \|Severe acute respiratory syndrome coro | .................... | .................... | .................... | .................... | .................... | .................... |
| 8568 | MW184805.1 | \|Severe acute respiratory syndrome coro | .................... | .................... | .................... | .................... | .................... | .................... |
| 8569 | MW184806.1 | \|Severe acute respiratory syndrome coro | .................... | .................... | .................... | .................... | .................... | .................... |
| 8570 | MW184811.1 | \|Severe acute respiratory syndrome coro | .................... | .................... | .................... | .................... | .................... | .................... |
| 8571 | MW184813.1 | \|Severe acute respiratory syndrome coro | .................... | .................... | .................... | .................... | .................... | .................... |
| 8573 | MW184815.1 | \|Severe acute respiratory syndrome coro | .................... | .................... | .................... | .................... | .................... | .................... |
| 8575 | MW184817.1 | \|Severe acute respiratory syndrome coro | .................... | .................... | .................... | .................... | .................... | .................... |
| 8576 | MW184820.1 | \|Severe acute respiratory syndrome coro | .................... | .................... | .................... | .................... | .................... | .................... |
| 8577 | MW184821.1 | \|Severe acute respiratory syndrome coro | .................... | .................... | .................... | .................... | .................... | .................... |
| 8578 | MW184822.1 | \|Severe acute respiratory syndrome coro | .................... | .................... | .................... | .................... | .................... | .................... |
| 8579 | MW184823.1 | \|Severe acute respiratory syndrome coro | .................... | .................... | .................... | .................... | .................... | .................... |
| 8580 | MW184825.1 | \|Severe acute respiratory syndrome coro | .................... | .................... | .................... | .................... | .................... | .................... |
| 8581 | MW184827.1 | \|Severe acute respiratory syndrome coro | .................... | .................... | .................... | .................... | .................... | .................... |
| 8582 | MW184828.1 | \|Severe acute respiratory syndrome coro | .................... | .................... | .................... | .................... | .................... | .................... |
| 8583 | MW184829.1 | \|Severe acute respiratory syndrome coro | .................... | .................... | .................... | .................... | .................... | .................... |
| 8584 | MW184830.1 | \|Severe acute respiratory syndrome coro | .................... | .................... | .................... | .................... | .................... | .................... |
| 8585 | MW184831.1 | \|Severe acute respiratory syndrome coro | .................... | .................... | .................... | .................... | .................... | .................... |
| 8586 | MW184832.1 | \|Severe acute respiratory syndrome coro | .................... | .................... | .................... | .................... | .................... | .................... |
| 8587 | MW184833.1 | \|Severe acute respiratory syndrome coro | .................... | .................... | .................... | .................... | .................... | .................... |
| 8588 | MW184834.1 | \|Severe acute respiratory syndrome coro | .................... | .................... | .................... | .................... | .................... | .................... |
| 8589 | MW184835.1 | \|Severe acute respiratory syndrome coro | .................... | .................... | .................... | .................... | .................... | .................... |
| 8590 | MW184836.1 | \|Severe acute respiratory syndrome coro | .................... | .................... | .................... | .................... | .................... | .................... |
| 8591 | MW184837.1 | \|Severe acute respiratory syndrome coro | .................... | .................... | .................... | .................... | .................... | .................... |
| 8592 | MW184838.1 | \|Severe acute respiratory syndrome coro | .................... | .................... | .................... | .................... | .................... | .................... |
| 8593 | MW184839.1 | \|Severe acute respiratory syndrome coro | .................... | .................... | .................... | .................... | .................... | .................... |
| 8594 | MW184840.1 | \|Severe acute respiratory syndrome coro | .................... | .................... | .................... | .................... | .................... | .................... |
| 8595 | MW184841.1 | \|Severe acute respiratory syndrome coro | .................... | .................... | .................... | .................... | .................... | .................... |
| 8596 | MW184842.1 | \|Severe acute respiratory syndrome coro | .................... | .................... | .................... | .................... | .................... | .................... |
| 8597 | MW184844.1 | \|Severe acute respiratory syndrome coro | .................... | .................... | .................... | .................... | .................... | .................... |
| 8598 | MW184845.1 | \|Severe acute respiratory syndrome coro | .................... | ......A............. | .................... | .................... | .................... | .................... |
| 8600 | MW184849.1 | \|Severe acute respiratory syndrome coro | .................... | .................... | .................... | .................... | .................... | .................... |
| 8601 | MW184851.1 | \|Severe acute respiratory syndrome coro | .................... | .................... | .................... | .................... | .................... | .................... |
| 8602 | MW184852.1 | \|Severe acute respiratory syndrome coro | .................... | .................... | .................... | .................... | .................... | .................... |
| 8603 | MW184853.1 | \|Severe acute respiratory syndrome coro | .................... | .................... | .................... | .................... | .................... | .................... |
| 8604 | MW184854.1 | \|Severe acute respiratory syndrome coro | .................... | .................... | .................... | .................... | .................... | .................... |
| 8605 | MW184855.1 | \|Severe acute respiratory syndrome coro | .................... | .................... | .................... | .................... | .................... | .................... |
| 8606 | MW184856.1 | \|Severe acute respiratory syndrome coro | .................... | .................... | .................... | .................... | .................... | .................... |
| 8607 | MW184858.1 | \|Severe acute respiratory syndrome coro | .................... | .................... | .................... | .................... | .................... | .................... |
| 8608 | MW184859.1 | \|Severe acute respiratory syndrome coro | .................... | .................... | .................... | .................... | .................... | .................... |
| 8609 | MW184860.1 | \|Severe acute respiratory syndrome coro | .................... | .................... | .................... | .................... | .................... | .................... |
| 8610 | MW184861.1 | \|Severe acute respiratory syndrome coro | .................... | .................... | .................... | .................... | .................... | .................... |
| 8611 | MW184862.1 | \|Severe acute respiratory syndrome coro | .................... | .................... | .................... | .................... | .................... | .................... |
| 8612 | MW184864.1 | \|Severe acute respiratory syndrome coro | .................... | .................... | .................... | .................... | .................... | .................... |
| 8613 | MW184865.1 | \|Severe acute respiratory syndrome coro | .................... | .................... | .................... | .................... | .................... | .................... |
| 8614 | MW184866.1 | \|Severe acute respiratory syndrome coro | .................... | .................... | .................... | .................... | .................... | .................... |
| 8615 | MW184867.1 | \|Severe acute respiratory syndrome coro | .................... | .................... | .................... | .................... | .................... | .................... |
| 8616 | MW184869.1 | \|Severe acute respiratory syndrome coro | .................... | .................... | .................... | .................... | .................... | .................... |
| 8617 | MW184870.1 | \|Severe acute respiratory syndrome coro | .................... | .................... | .................... | .................... | .................... | .................... |
| 8618 | MW184871.1 | \|Severe acute respiratory syndrome coro | .................... | .................... | .................... | .................... | .................... | .................... |
| 8619 | MW184872.1 | \|Severe acute respiratory syndrome coro | .................... | .................... | .................... | .................... | .................... | .................... |
| 8620 | MW184874.1 | \|Severe acute respiratory syndrome coro | .................... | .................... | .................... | .................... | .................... | .................... |
| 8621 | MW184875.1 | \|Severe acute respiratory syndrome coro | .................... | .................... | .................... | .................... | .................... | .................... |
| 8622 | MW184876.1 | \|Severe acute respiratory syndrome coro | .................... | .................... | .................... | .................... | .........R.......... | .................... |
| 8623 | MW184877.1 | \|Severe acute respiratory syndrome coro | .................... | .................... | .................... | .................... | .................... | .................... |
| 8624 | MW184879.1 | \|Severe acute respiratory syndrome coro | .................... | ......A............. | .................... | .................... | .................... | .................... |
| 8625 | MW184882.1 | \|Severe acute respiratory syndrome coro | .................... | .................... | .................... | .................... | .................... | .................... |
| 8626 | MW184883.1 | \|Severe acute respiratory syndrome coro | .................... | .................... | .................... | .................... | .................... | .................... |
| 8627 | MW184884.1 | \|Severe acute respiratory syndrome coro | .................... | .................... | .................... | .................... | .................... | .................... |
| 8628 | MW184885.1 | \|Severe acute respiratory syndrome coro | .................... | .................... | .................... | .................... | .................... | .................... |
| 8629 | MW184886.1 | \|Severe acute respiratory syndrome coro | .................... | .................... | .................... | .................... | .................... | .................... |
| 8630 | MW184887.1 | \|Severe acute respiratory syndrome coro | .................... | .................... | .................... | .................... | .................... | .................... |
| 8631 | MW184888.1 | \|Severe acute respiratory syndrome coro | .................... | .................... | .................... | .................... | .................... | .................... |
| 8633 | MW184891.1 | \|Severe acute respiratory syndrome coro | .................... | .................... | .................... | .................... | .................... | .................... |
| 8634 | MW184892.1 | \|Severe acute respiratory syndrome coro | .................... | .................... | .................... | .................... | .................... | .................... |
| 8635 | MW184893.1 | \|Severe acute respiratory syndrome coro | .................... | .................... | .................... | .................... | .................... | .................... |
| 8636 | MW184894.1 | \|Severe acute respiratory syndrome coro | .................... | .................... | .................... | .................... | .................... | .................... |
| 8637 | MW184895.1 | \|Severe acute respiratory syndrome coro | .................... | .................... | .................... | .................... | .................... | .................... |
| 8638 | MW184896.1 | \|Severe acute respiratory syndrome coro | .................... | .................... | .................... | .................... | .................... | .................... |
| 8639 | MW184897.1 | \|Severe acute respiratory syndrome coro | .................... | .................... | .................... | .................... | .................... | .................... |
| 8640 | MW184899.1 | \|Severe acute respiratory syndrome coro | .................... | .................... | .................... | .................... | .................... | .................... |
| 8641 | MW184900.1 | \|Severe acute respiratory syndrome coro | .................... | .................... | .................... | .................... | .................... | .................... |
| 8642 | MW184901.1 | \|Severe acute respiratory syndrome coro | .................... | .................... | .................... | .................... | .................... | .................... |
| 8643 | MW184903.1 | \|Severe acute respiratory syndrome coro | .................... | .................... | .................... | .................... | .................... | .................... |
| 8644 | MW184904.1 | \|Severe acute respiratory syndrome coro | .................... | .................... | .................... | .................... | .................... | .................... |

```
8645 MW184906.1 |Severe acute respiratory syndrome coro    .................... .................... .................... .................... .................... ....................
8646 MW184907.1 |Severe acute respiratory syndrome coro    .................... .................... .................... .................... .................... ....................
8647 MW184908.1 |Severe acute respiratory syndrome coro    .................... .................... .................... .................... .................... ....................
8648 MW184909.1 |Severe acute respiratory syndrome coro    .................... .................... .................... .................... .................... ....................
8649 MW184911.1 |Severe acute respiratory syndrome coro    .................... .................... .................... .................... .................... ....................
8650 MW184914.1 |Severe acute respiratory syndrome coro    .................... .................... .................... .................... .................... ....................
8651 MW184915.1 |Severe acute respiratory syndrome coro    .................... .................... .................... .................... .................... ....................
8652 MW184916.1 |Severe acute respiratory syndrome coro    .................... .................... .................... .................... .................... ....................
8653 MW184917.1 |Severe acute respiratory syndrome coro    .................... .................... .................... .................... .................... ....................
8654 MW184918.1 |Severe acute respiratory syndrome coro    .................... .................... .................... .................... .................... ....................
8655 MW184919.1 |Severe acute respiratory syndrome coro    .................... .................... .................... .................... .................... ....................
8656 MW184922.1 |Severe acute respiratory syndrome coro    .................... .................... .................... .................... .................... ....................
8657 MW184923.1 |Severe acute respiratory syndrome coro    .................... .................... .................... .................... .................... ....................
8658 MW184924.1 |Severe acute respiratory syndrome coro    .................... .................... .................... .................... .................... ....................
8659 MW184925.1 |Severe acute respiratory syndrome coro    .................... .................... .................... .................... .................... ....................
8660 MW184926.1 |Severe acute respiratory syndrome coro    .................... .................... .................... .................... .................... ....................
8661 MW184927.1 |Severe acute respiratory syndrome coro    .................... .................... .................... .................... .................... ....................
8662 MW184928.1 |Severe acute respiratory syndrome coro    .................... .................... .................... .................... .................... ....................
8663 MW184929.1 |Severe acute respiratory syndrome coro    .................... .................... .................... .................... .................... ....................
8664 MW184930.1 |Severe acute respiratory syndrome coro    .................... .................... .................... .................... .................... ....................
8665 MW184931.1 |Severe acute respiratory syndrome coro    .................... .................... .................... .................... .................... ....................
8666 MW184933.1 |Severe acute respiratory syndrome coro    .................... .................... .................... .................... .................... ....................
8667 MW184934.1 |Severe acute respiratory syndrome coro    .................... .................... .................... .................... .................... ....................
8669 MW184938.1 |Severe acute respiratory syndrome coro    .................... .................... .................... .................... .................... ....................
8670 MW184939.1 |Severe acute respiratory syndrome coro    .................... .................... .................... .................... .................... ....................
8671 MW184940.1 |Severe acute respiratory syndrome coro    .................... .................... .................... .................... .................... ....................
8672 MW184941.1 |Severe acute respiratory syndrome coro    .................... .................... .................... .................... .................... ....................
8673 MW184944.1 |Severe acute respiratory syndrome coro    .................... .................... .................... .................... .................... ....................
8674 MW184945.1 |Severe acute respiratory syndrome coro    .................... .................... .................... .................... .................... ....................
8675 MW184947.1 |Severe acute respiratory syndrome coro    .................... .................... .................... .................... .................... ....................
8676 MW184948.1 |Severe acute respiratory syndrome coro    .................... .................... .................... .................... .................... ....................
8677 MW184949.1 |Severe acute respiratory syndrome coro    .................... .................... .................... .................... .................... ....................
8678 MW184950.1 |Severe acute respiratory syndrome coro    .................... .................... .................... .................... .................... ....................
8679 MW184951.1 |Severe acute respiratory syndrome coro    .................... .................... .................... .................... .................... ....................
8680 MW184952.1 |Severe acute respiratory syndrome coro    .................... .................... .................... .................... .................... ....................
8681 MW184953.1 |Severe acute respiratory syndrome coro    .................... .................... .................... .................... .................... ....................
8682 MW184954.1 |Severe acute respiratory syndrome coro    .................... .................... .................... .................... .................... ....................
8683 MW184956.1 |Severe acute respiratory syndrome coro    .................... .................... .................... .................... .................... ....................
8684 MW184960.1 |Severe acute respiratory syndrome coro    .................... .................... .................... .................... .................... ....................
8685 MW184961.1 |Severe acute respiratory syndrome coro    .................... .................... .................... .................... .................... ....................
8686 MW184966.1 |Severe acute respiratory syndrome coro    .................... .................... .................... .................... .................... ....................
8687 MW184968.1 |Severe acute respiratory syndrome coro    .................... .................... .................... .................... .................... ....................
8688 MW184969.1 |Severe acute respiratory syndrome coro    .................... .................... .................... .................... .................... ....................
8689 MW184970.1 |Severe acute respiratory syndrome coro    .................... .................... .................... .................... .................... ....................
8690 MW184972.1 |Severe acute respiratory syndrome coro    .................... .................... .................... .................... .................... ....................
8691 MW184973.1 |Severe acute respiratory syndrome coro    .................... .................... .................... .................... .................... ....................
8692 MW184974.1 |Severe acute respiratory syndrome coro    .................... .................... .................... .................... .................... ....................
8693 MW184975.1 |Severe acute respiratory syndrome coro    .................... .................... .................... .................... .................... ....................
8694 MW184976.1 |Severe acute respiratory syndrome coro    .................... .................... .................... .................... .................... ....................
8695 MW184977.1 |Severe acute respiratory syndrome coro    .................... .................... .................... .................... .................... ....................
8696 MW184978.1 |Severe acute respiratory syndrome coro    .................... .................... .................... .................... .................... ....................
8697 MW184979.1 |Severe acute respiratory syndrome coro    .................... .................... .................... .................... .................... ....................
8698 MW184980.1 |Severe acute respiratory syndrome coro    .................... .................... .................... .................... .................... ....................
8699 MW184981.1 |Severe acute respiratory syndrome coro    .................... .................... .................... .................... .................... ....................
8700 MW184982.1 |Severe acute respiratory syndrome coro    .................... .................... .................... .................... .................... ....................
8701 MW184983.1 |Severe acute respiratory syndrome coro    .................... .................... .................... .................... .................... ....................
8702 MW184984.1 |Severe acute respiratory syndrome coro    .................... .................... .................... .................... .................... ....................
8703 MW184987.1 |Severe acute respiratory syndrome coro    .................... .................... .................... .................... .................... ....................
8704 MW184988.1 |Severe acute respiratory syndrome coro    .................... .................... .................... .................... .................... ....................
8705 MW184989.1 |Severe acute respiratory syndrome coro    .................... .................... .................... .................... .................... ....................
8706 MW184990.1 |Severe acute respiratory syndrome coro    .................... .................... .................... .................... .................... ....................
8707 MW184991.1 |Severe acute respiratory syndrome coro    .................... .................... .................... .................... .................... ....................
8708 MW184992.1 |Severe acute respiratory syndrome coro    .................... .................... .................... .................... .................... ....................
8709 MW184993.1 |Severe acute respiratory syndrome coro    .................... .................... .................... .................... .................... ....................
8710 MW184994.1 |Severe acute respiratory syndrome coro    .................... .................... .................... .................... .................... ....................
8711 MW184995.1 |Severe acute respiratory syndrome coro    .................... .................... .................... .................... .................... ....................
8712 MW184996.1 |Severe acute respiratory syndrome coro    .................... .................... .................... .................... .................... ....................
8713 MW184997.1 |Severe acute respiratory syndrome coro    .................... .................... .................... .................... .................... ....................
8714 MW184999.1 |Severe acute respiratory syndrome coro    .................... .................... .................... .................... .................... ....................
8715 MW185001.1 |Severe acute respiratory syndrome coro    .................... .................... .................... .................... .................... ....................
8716 MW185003.1 |Severe acute respiratory syndrome coro    .................... .................... .................... .................... .................... ....................
8717 MW185004.1 |Severe acute respiratory syndrome coro    .................... .................... .................... .................... .................... ....................
8718 MW185007.1 |Severe acute respiratory syndrome coro    .................... .................... .................... .................... .................... ....................
8719 MW185008.1 |Severe acute respiratory syndrome coro    .................... .................... .................... .................... .................... ....................
8720 MW185009.1 |Severe acute respiratory syndrome coro    .................... .................... .................... .................... .................... ....................
8721 MW185010.1 |Severe acute respiratory syndrome coro    .................... .................... .................... .................... .................... ....................
8722 MW185012.1 |Severe acute respiratory syndrome coro    .................... .................... .................... .................... .................... ....................
8723 MW185013.1 |Severe acute respiratory syndrome coro    .................... .................... .................... .................... .................... ....................
```

8724 MW185014.1 |Severe acute respiratory syndrome coro .................... .................... .................... .................... .................... ....................
8725 MW185015.1 |Severe acute respiratory syndrome coro .................... .................... .................... .................... .................... ....................
8726 MW185016.1 |Severe acute respiratory syndrome coro .................... .................... .................... .................... .................... ....................
8727 MW185017.1 |Severe acute respiratory syndrome coro .................... .................... .................... .................... .................... ....................
8728 MW185018.1 |Severe acute respiratory syndrome coro .................... .................... .................... .................... .................... ....................
8729 MW185020.1 |Severe acute respiratory syndrome coro .................... .................... .................... .................... .................... ....................
8730 MW185023.1 |Severe acute respiratory syndrome coro .................... .................... .................... .................... .................... ....................
8731 MW185024.1 |Severe acute respiratory syndrome coro .................... .................... .................... .................... .................... ....................
8732 MW185026.1 |Severe acute respiratory syndrome coro .................... .................... .................... .................... .................... ....................
8733 MW185027.1 |Severe acute respiratory syndrome coro .................... .................... .................... .................... .................... ....................
8734 MW185028.1 |Severe acute respiratory syndrome coro .................... .................... .................... .................... .................... ....................
8735 MW185029.1 |Severe acute respiratory syndrome coro .................... .................... .................... .................... .................... ....................
8736 MW185030.1 |Severe acute respiratory syndrome coro .................... ......A............. .................... .................... .................... ....................
8737 MW185031.1 |Severe acute respiratory syndrome coro .................... .................... .................... .................... .................... ....................
8738 MW185032.1 |Severe acute respiratory syndrome coro .................... .................... .................... .................... .................... ....................
8739 MW185034.1 |Severe acute respiratory syndrome coro .................... .................... .................... .................... .................... ....................
8740 MW185035.1 |Severe acute respiratory syndrome coro .................... .................... .................... .................... .................... ....................
8741 MW185037.1 |Severe acute respiratory syndrome coro .................... .................... .................... .................... .................... ....................
8742 MW185038.1 |Severe acute respiratory syndrome coro .................... .................... .................... .................... .................... ....................
8743 MW185040.1 |Severe acute respiratory syndrome coro .................... .................... .................... .................... .................... ....................
8744 MW185042.1 |Severe acute respiratory syndrome coro .................... .................... .................... .................... .................... ....................
8745 MW185044.1 |Severe acute respiratory syndrome coro .................... .................... .................... .................... .................... ....................
8746 MW185045.1 |Severe acute respiratory syndrome coro .................... .................... .................... .................... .................... ....................
8747 MW185046.1 |Severe acute respiratory syndrome coro .................... .................... .................... .................... .................... ....................
8748 MW185047.1 |Severe acute respiratory syndrome coro .................... .................... .................... .................... .................... ....................
8749 MW185048.1 |Severe acute respiratory syndrome coro .................... .................... .................... .................... .................... ....................
8750 MW185049.1 |Severe acute respiratory syndrome coro .................... .................... .................... .................... .................... ....................
8752 MW185052.1 |Severe acute respiratory syndrome coro .................... .................... .................... .................... .................... ....................
8753 MW185053.1 |Severe acute respiratory syndrome coro .................... .................... .................... .................... .................... ....................
8754 MW185055.1 |Severe acute respiratory syndrome coro .................... .................... .................... .................... .................... ....................
8755 MW185057.1 |Severe acute respiratory syndrome coro .................... .................... .................... .................... .................... ....................
8756 MW185058.1 |Severe acute respiratory syndrome coro .................... .................... .................... .................... .................... ....................
8757 MW185059.1 |Severe acute respiratory syndrome coro .................... .................... .................... .................... .................... ....................
8758 MW185061.1 |Severe acute respiratory syndrome coro .................... .................... .................... .................... .................... ....................
8759 MW185062.1 |Severe acute respiratory syndrome coro .................... .................... .................... .................... .................... ....................
8760 MW185064.1 |Severe acute respiratory syndrome coro .................... .................... .................... .................... .................... ....................
8761 MW185065.1 |Severe acute respiratory syndrome coro .................... .................... .................... .................... .................... ....................
8762 MW185066.1 |Severe acute respiratory syndrome coro .................... .................... .................... .................... .................... ....................
8763 MW185067.1 |Severe acute respiratory syndrome coro .................... .................... .................... .................... .................... ....................
8764 MW185069.1 |Severe acute respiratory syndrome coro .................... .................... .................... .................... .................... ....................
8765 MW185070.1 |Severe acute respiratory syndrome coro .................... .................... .................... .................... .................... ....................
8766 MW185071.1 |Severe acute respiratory syndrome coro .................... .................... .................... .................... .................... ....................
8767 MW185072.1 |Severe acute respiratory syndrome coro .................... .................... .................... .................... .................... ....................
8768 MW185073.1 |Severe acute respiratory syndrome coro .................... .................... .................... .................... .................... ....................
8769 MW185074.1 |Severe acute respiratory syndrome coro .................... .................... .................... .................... .................... ....................
8770 MW185075.1 |Severe acute respiratory syndrome coro .................... .................... .................... .................... .................... ....................
8771 MW185076.1 |Severe acute respiratory syndrome coro .................... .................... .................... .................... .................... ....................
8772 MW185077.1 |Severe acute respiratory syndrome coro .................... .................... .................... .................... .................... ....................
8773 MW185078.1 |Severe acute respiratory syndrome coro .................... .................... .................... .................... .................... ....................
8774 MW185080.1 |Severe acute respiratory syndrome coro .................... .................... .................... .................... .................... ....................
8775 MW185081.1 |Severe acute respiratory syndrome coro .................... .................... .................... .................... .................... ....................
8777 MW185086.1 |Severe acute respiratory syndrome coro .................... .................... .................... .................... .................... ....................
8778 MW185087.1 |Severe acute respiratory syndrome coro .................... .................... .................... .................... .................... ....................
8779 MW185088.1 |Severe acute respiratory syndrome coro .................... .................... .................... .................... .................... ....................
8780 MW185089.1 |Severe acute respiratory syndrome coro .................... .................... .................... .................... .................... ....................
8781 MW185090.1 |Severe acute respiratory syndrome coro .................... .................... .................... .................... .................... ....................
8782 MW185091.1 |Severe acute respiratory syndrome coro .................... .................... .................... .................... .................... ....................
8783 MW185093.1 |Severe acute respiratory syndrome coro .................... .................... .................... .................... .................... ....................
8784 MW185094.1 |Severe acute respiratory syndrome coro .................... .................... .................... .................... .................... ....................
8785 MW185096.1 |Severe acute respiratory syndrome coro .................... .................... .................... .................... .................... ....................
8786 MW185098.1 |Severe acute respiratory syndrome coro .................... .................... .................... .................... .................... ....................
8787 MW185099.1 |Severe acute respiratory syndrome coro .................... .................... .................... .................... .................... ....................
8788 MW185101.1 |Severe acute respiratory syndrome coro .................... .................... .................... .................... .................... ....................
8789 MW185102.1 |Severe acute respiratory syndrome coro .................... .................... .................... .................... .................... ....................
8792 MW185105.1 |Severe acute respiratory syndrome coro .................... .................... .................... .................... .................... ....................
8793 MW185106.1 |Severe acute respiratory syndrome coro .................... .................... .................... .................... .................... ....................
8794 MW185107.1 |Severe acute respiratory syndrome coro .................... .................... .................... .................... .................... ....................
8795 MW185108.1 |Severe acute respiratory syndrome coro .................... .................... .................... .................... .................... ....................
8796 MW185109.1 |Severe acute respiratory syndrome coro .................... .................... .................... .................... .................... ....................
8797 MW185111.1 |Severe acute respiratory syndrome coro .................... .................... .................... .................... .................... ....................
8798 MW185113.1 |Severe acute respiratory syndrome coro .................... .................... .................... .................... .................... ....................
8799 MW185114.1 |Severe acute respiratory syndrome coro .................... .................... .................... .................... .................... ....................
8800 MW185116.1 |Severe acute respiratory syndrome coro .................... .................... .................... .................... .................... ....................
8801 MW185118.1 |Severe acute respiratory syndrome coro .................... .................... .................... .................... .................... ....................
8802 MW185119.1 |Severe acute respiratory syndrome coro .................... .................... .................... .................... .................... ....................
8803 MW185120.1 |Severe acute respiratory syndrome coro .................... .................... .................... .................... .................... ....................
8804 MW185121.1 |Severe acute respiratory syndrome coro .................... .................... .................... .................... .................... ....................
8805 MW185122.1 |Severe acute respiratory syndrome coro .................... .................... .................... .................... .................... ....................

| | | | | | | | |
|---|---|---|---|---|---|---|---|
| 8806 MW185123.1 | Severe acute respiratory syndrome coro | .................... | .................... | ..................... | .................... | .................... | .................... |
| 8808 MW185126.1 | Severe acute respiratory syndrome coro | .................... | .................... | ..................... | .................... | .................... | .................... |
| 8809 MW185128.1 | Severe acute respiratory syndrome coro | .................... | .................... | ..................... | .................... | .................... | .................... |
| 8810 MW185129.1 | Severe acute respiratory syndrome coro | .................... | .................... | ..................... | .................... | .................... | .................... |
| 8811 MW185130.1 | Severe acute respiratory syndrome coro | .................... | .................... | ..................... | .................... | .................... | .................... |
| 8812 MW185131.1 | Severe acute respiratory syndrome coro | .................... | .................... | ..................... | .................... | .................... | .................... |
| 8813 MW185132.1 | Severe acute respiratory syndrome coro | .................... | .................... | ..................... | .................... | .................... | .................... |
| 8814 MW185133.1 | Severe acute respiratory syndrome coro | .................... | .................... | ..................... | .................... | .................... | .................... |
| 8815 MW185135.1 | Severe acute respiratory syndrome coro | .................... | .................... | ..................... | .................... | .................... | .................... |
| 8816 MW185136.1 | Severe acute respiratory syndrome coro | .................... | .................... | ..................... | .................... | .................... | .................... |
| 8817 MW185137.1 | Severe acute respiratory syndrome coro | .................... | .................... | ..................... | .................... | .................... | .................... |
| 8818 MW185138.1 | Severe acute respiratory syndrome coro | .................... | .................... | ..................... | .................... | .................... | .................... |
| 8819 MW185140.1 | Severe acute respiratory syndrome coro | .................... | .................... | ..................... | .................... | .................... | .................... |
| 8820 MW185141.1 | Severe acute respiratory syndrome coro | .................... | .................... | ..................... | .................... | .................... | .................... |
| 8821 MW185142.1 | Severe acute respiratory syndrome coro | .................... | .................... | ..................... | .................... | .................... | .................... |
| 8822 MW185143.1 | Severe acute respiratory syndrome coro | .................... | .................... | ..................... | .................... | .................... | .................... |
| 8823 MW185144.1 | Severe acute respiratory syndrome coro | .................... | .................... | ..................... | .................... | .................... | .................... |
| 8824 MW185145.1 | Severe acute respiratory syndrome coro | .................... | .................... | ..................... | .................... | .................... | .................... |
| 8825 MW185146.1 | Severe acute respiratory syndrome coro | .................... | .................... | ..................... | .................... | .................... | .................... |
| 8826 MW185148.1 | Severe acute respiratory syndrome coro | .................... | .................... | ..................... | .................... | .................... | .................... |
| 8827 MW185150.1 | Severe acute respiratory syndrome coro | .................... | .................... | ..................... | .................... | .................... | .................... |
| 8828 MW185151.1 | Severe acute respiratory syndrome coro | .................... | .................... | ..................... | .................... | .................... | .................... |
| 8829 MW185152.1 | Severe acute respiratory syndrome coro | .................... | .................... | ..................... | .................... | .................... | .................... |
| 8830 MW185153.1 | Severe acute respiratory syndrome coro | .................... | .................... | ..................... | .................... | .................... | .................... |
| 8831 MW185155.1 | Severe acute respiratory syndrome coro | .................... | .................... | ..................... | .................... | .................... | .................... |
| 8832 MW185156.1 | Severe acute respiratory syndrome coro | .................... | .................... | ..................... | .................... | .................... | .................... |
| 8833 MW185158.1 | Severe acute respiratory syndrome coro | .................... | .................... | ..................... | .................... | .................... | .................... |
| 8834 MW185159.1 | Severe acute respiratory syndrome coro | .................... | .................... | ..................... | .................... | .................... | .................... |
| 8835 MW185161.1 | Severe acute respiratory syndrome coro | .................... | .................... | ..................... | .................... | .................... | .................... |
| 8836 MW185162.1 | Severe acute respiratory syndrome coro | .................... | .................... | ..................... | .................... | .................... | .................... |
| 8837 MW185164.1 | Severe acute respiratory syndrome coro | .................... | .................... | ..................... | .................... | .................... | .................... |
| 8838 MW185167.1 | Severe acute respiratory syndrome coro | .................... | .................... | ..................... | .................... | .................... | .................... |
| 8839 MW185168.1 | Severe acute respiratory syndrome coro | .................... | .................... | ..................... | .................... | .................... | .................... |
| 8840 MW185169.1 | Severe acute respiratory syndrome coro | .................... | .................... | ..................... | .................... | .................... | .................... |
| 8841 MW185171.1 | Severe acute respiratory syndrome coro | .................... | .................... | ..................... | .................... | .................... | .................... |
| 8842 MW185173.1 | Severe acute respiratory syndrome coro | .................... | .................... | ..................... | .................... | .................... | .................... |
| 8843 MW185174.1 | Severe acute respiratory syndrome coro | .................... | .................... | ..................... | .................... | .................... | .................... |
| 8844 MW185175.1 | Severe acute respiratory syndrome coro | .................... | .................... | ..................... | .................... | .................... | .................... |
| 8845 MW185176.1 | Severe acute respiratory syndrome coro | .................... | .................... | ..................... | .................... | .................... | .................... |
| 8846 MW185178.1 | Severe acute respiratory syndrome coro | .................... | .................... | ..................... | .................... | .................... | .................... |
| 8847 MW185179.1 | Severe acute respiratory syndrome coro | .................... | .................... | ..................... | .................... | .................... | .................... |
| 8848 MW185180.1 | Severe acute respiratory syndrome coro | .................... | .................... | ..................... | .................... | .................... | .................... |
| 8849 MW185181.1 | Severe acute respiratory syndrome coro | .................... | .................... | ..................... | .................... | .................... | .................... |
| 8850 MW185182.1 | Severe acute respiratory syndrome coro | .................... | .................... | ..................... | .................... | .................... | .................... |
| 8851 MW185184.1 | Severe acute respiratory syndrome coro | .................... | .................... | ..................... | .................... | .................... | .................... |
| 8852 MW185185.1 | Severe acute respiratory syndrome coro | .................... | .................... | ..................... | .................... | .................... | .................... |
| 8853 MW185186.1 | Severe acute respiratory syndrome coro | .................... | .................... | ..................... | .................... | .................... | .................... |
| 8855 MW185189.1 | Severe acute respiratory syndrome coro | .................... | .................... | ..................... | .................... | .................... | .................... |
| 8856 MW185191.1 | Severe acute respiratory syndrome coro | .................... | .................... | ..................... | .................... | .................... | .................... |
| 8857 MW185192.1 | Severe acute respiratory syndrome coro | .................... | .................... | ..................... | .................... | .................... | .................... |
| 8858 MW185193.1 | Severe acute respiratory syndrome coro | .................... | .................... | ..................... | .................... | .................... | .................... |
| 8859 MW185194.1 | Severe acute respiratory syndrome coro | .................... | .................... | ..................... | .................... | .................... | .................... |
| 8860 MW185196.1 | Severe acute respiratory syndrome coro | .................... | .................... | ..................... | .................... | .................... | .................... |
| 8862 MW185198.1 | Severe acute respiratory syndrome coro | .................... | .................... | ..................... | .................... | .................... | .................... |
| 8863 MW185199.1 | Severe acute respiratory syndrome coro | .................... | .................... | ..................... | .................... | .................... | .................... |
| 8865 MW185201.1 | Severe acute respiratory syndrome coro | .................... | .................... | ..................... | .................... | .................... | .................... |
| 8866 MW185202.1 | Severe acute respiratory syndrome coro | .................... | .................... | ..................... | .................... | .................... | .................... |
| 8867 MW185203.1 | Severe acute respiratory syndrome coro | .................... | .................... | ..................... | .................... | .................... | .................... |
| 8868 MW185207.1 | Severe acute respiratory syndrome coro | .................... | .................... | ..................... | .................... | .................... | .................... |
| 8869 MW185208.1 | Severe acute respiratory syndrome coro | .................... | .................... | ..................... | .................... | .................... | .................... |
| 8870 MW185210.1 | Severe acute respiratory syndrome coro | .................... | .................... | ..................... | .................... | .................... | .................... |
| 8871 MW185212.1 | Severe acute respiratory syndrome coro | .................... | .................... | ..................... | .................... | .................... | .................... |
| 8872 MW185213.1 | Severe acute respiratory syndrome coro | .................... | .................... | ..................... | .................... | .................... | .................... |
| 8873 MW185215.1 | Severe acute respiratory syndrome coro | .................... | .................... | ..................... | .................... | .................... | .................... |
| 8874 MW185216.1 | Severe acute respiratory syndrome coro | .................... | .................... | ..................... | .................... | .................... | .................... |
| 8875 MW185217.1 | Severe acute respiratory syndrome coro | .................... | .................... | ..................... | .................... | .................... | .................... |
| 8876 MW185218.1 | Severe acute respiratory syndrome coro | .................... | .................... | ..................... | .................... | .................... | .................... |
| 8877 MW185219.1 | Severe acute respiratory syndrome coro | .................... | .................... | ..................... | .................... | .................... | .................... |
| 8878 MW185221.1 | Severe acute respiratory syndrome coro | .................... | .................... | ..................... | .................... | .................... | .................... |
| 8879 MW185222.1 | Severe acute respiratory syndrome coro | .................... | .................... | ..................... | .................... | .................... | .................... |
| 8880 MW185223.1 | Severe acute respiratory syndrome coro | .................... | .................... | ..................... | .................... | .................... | .................... |
| 8881 MW185224.1 | Severe acute respiratory syndrome coro | .................... | .................... | ..................... | .................... | .................... | .................... |
| 8882 MW185226.1 | Severe acute respiratory syndrome coro | .................... | .................... | ..................... | .................... | .................... | .................... |
| 8883 MW185227.1 | Severe acute respiratory syndrome coro | .................... | .................... | ..................... | .................... | .................... | .................... |
| 8884 MW185228.1 | Severe acute respiratory syndrome coro | .................... | .................... | ..................... | .................... | .................... | .................... |
| 8885 MW185231.1 | Severe acute respiratory syndrome coro | .................... | .................... | ..................... | .................... | .................... | .................... |
| 8887 MW185233.1 | Severe acute respiratory syndrome coro | .................... | .................... | ..................... | .................... | .................... | .................... |
| 8888 MW185234.1 | Severe acute respiratory syndrome coro | .................... | .................... | ..................... | .................... | .................... | .................... |

| | | | | | | | | | |
|---|---|---|---|---|---|---|---|---|---|
| 8889 | MW185237.1 | \|Severe acute respiratory syndrome coro | ···················· | ···················· | ···················· | ···················· | ···················· | ···················· | |
| 8890 | MW185238.1 | \|Severe acute respiratory syndrome coro | ···················· | ···················· | ···················· | ···················· | ···················· | ···················· | |
| 8891 | MW185239.1 | \|Severe acute respiratory syndrome coro | ···················· | ···················· | ···················· | ···················· | ···················· | ···················· | |
| 8892 | MW185242.1 | \|Severe acute respiratory syndrome coro | ···················· | ···················· | ···················· | ···················· | ···················· | ···················· | |
| 8893 | MW185245.1 | \|Severe acute respiratory syndrome coro | ···················· | ···················· | ···················· | ···················· | ···················· | ···················· | |
| 8894 | MW185247.1 | \|Severe acute respiratory syndrome coro | ···················· | ···················· | ···················· | ···················· | ···················· | ···················· | |
| 8895 | MW185248.1 | \|Severe acute respiratory syndrome coro | ···················· | ···················· | ···················· | ···················· | ···················· | ···················· | |
| 8896 | MW185249.1 | \|Severe acute respiratory syndrome coro | ···················· | ···················· | ···················· | ···················· | ···················· | ···················· | |
| 8897 | MW185250.1 | \|Severe acute respiratory syndrome coro | ···················· | ···················· | ···················· | ···················· | ···················· | ···················· | |
| 8898 | MW185251.1 | \|Severe acute respiratory syndrome coro | ···················· | ···················· | ···················· | ···················· | ···················· | ···················· | |
| 8899 | MW185252.1 | \|Severe acute respiratory syndrome coro | ···················· | ···················· | ···················· | ···················· | ···················· | ···················· | |
| 8900 | MW185253.1 | \|Severe acute respiratory syndrome coro | ···················· | ···················· | ···················· | ···················· | ···················· | ···················· | |
| 8901 | MW185256.1 | \|Severe acute respiratory syndrome coro | ···················· | ···················· | ···················· | ···················· | ···················· | ···················· | |
| 8902 | MW185257.1 | \|Severe acute respiratory syndrome coro | ···················· | ···················· | ···················· | ···················· | ···················· | ···················· | |
| 8903 | MW185258.1 | \|Severe acute respiratory syndrome coro | ···················· | ···················· | ···················· | ···················· | ···················· | ···················· | |
| 8904 | MW185259.1 | \|Severe acute respiratory syndrome coro | ···················· | ···················· | ···················· | ···················· | ···················· | ···················· | |
| 8905 | MW185261.1 | \|Severe acute respiratory syndrome coro | ···················· | ···················· | ···················· | ···················· | ···················· | ···················· | |
| 8906 | MW185262.1 | \|Severe acute respiratory syndrome coro | ···················· | ···················· | ···················· | ···················· | ···················· | ···················· | |
| 8907 | MW185263.1 | \|Severe acute respiratory syndrome coro | ···················· | ···················· | ···················· | ···················· | ···················· | ···················· | |
| 8908 | MW185264.1 | \|Severe acute respiratory syndrome coro | ···················· | ···················· | ···················· | ···················· | ···················· | ···················· | |
| 8909 | MW185265.1 | \|Severe acute respiratory syndrome coro | ···················· | ···················· | ···················· | ···················· | ···················· | ···················· | |
| 8910 | MW185266.1 | \|Severe acute respiratory syndrome coro | ···················· | ···················· | ···················· | ···················· | ···················· | ···················· | |
| 8911 | MW185267.1 | \|Severe acute respiratory syndrome coro | ···················· | ···················· | ···················· | ···················· | ······R············· | ···················· | |
| 8912 | MW185268.1 | \|Severe acute respiratory syndrome coro | ···················· | ···················· | ···················· | ···················· | ···················· | ···················· | |
| 8913 | MW185269.1 | \|Severe acute respiratory syndrome coro | ···················· | ···················· | ···················· | ···················· | ···················· | ···················· | |
| 8914 | MW185271.1 | \|Severe acute respiratory syndrome coro | ···················· | ···················· | ···················· | ···················· | ···················· | ···················· | |
| 8915 | MW185272.1 | \|Severe acute respiratory syndrome coro | ···················· | ···················· | ···················· | ···················· | ···················· | ···················· | |
| 8916 | MW185273.1 | \|Severe acute respiratory syndrome coro | ···················· | ···················· | ···················· | ···················· | ···················· | ···················· | |
| 8917 | MW185274.1 | \|Severe acute respiratory syndrome coro | ···················· | ···················· | ···················· | ···················· | ···················· | ···················· | |
| 8918 | MW185275.1 | \|Severe acute respiratory syndrome coro | ···················· | ···················· | ···················· | ···················· | ···················· | ···················· | |
| 8919 | MW185277.1 | \|Severe acute respiratory syndrome coro | ···················· | ···················· | ···················· | ···················· | ···················· | ···················· | |
| 8920 | MW185278.1 | \|Severe acute respiratory syndrome coro | ···················· | ···················· | ···················· | ···················· | ···················· | ···················· | |
| 8921 | MW185279.1 | \|Severe acute respiratory syndrome coro | ···················· | ···················· | ···················· | ···················· | ···················· | ···················· | |
| 8922 | MW185280.1 | \|Severe acute respiratory syndrome coro | ···················· | ···················· | ···················· | ···················· | ···················· | ···················· | |
| 8923 | MW185282.1 | \|Severe acute respiratory syndrome coro | ···················· | ···················· | ···················· | ···················· | ···················· | ···················· | |
| 8924 | MW185283.1 | \|Severe acute respiratory syndrome coro | ···················· | ···················· | ···················· | ···················· | ···················· | ···················· | |
| 8925 | MW185285.1 | \|Severe acute respiratory syndrome coro | ···················· | ···················· | ···················· | ···················· | ···················· | ···················· | |
| 8927 | MW185287.1 | \|Severe acute respiratory syndrome coro | ···················· | ···················· | ···················· | ···················· | ···················· | ···················· | |
| 8928 | MW185288.1 | \|Severe acute respiratory syndrome coro | ···················· | ···················· | ···················· | ···················· | ···················· | ···················· | |
| 8929 | MW185289.1 | \|Severe acute respiratory syndrome coro | ···················· | ···················· | ···················· | ···················· | ···················· | ···················· | |
| 8930 | MW185290.1 | \|Severe acute respiratory syndrome coro | ···················· | ···················· | ···················· | ···················· | ···················· | ···················· | |
| 8931 | MW185291.1 | \|Severe acute respiratory syndrome coro | ···················· | ···················· | ···················· | ···················· | ···················· | ···················· | |
| 8932 | MW185293.1 | \|Severe acute respiratory syndrome coro | ···················· | ···················· | ···················· | ···················· | ···················· | ···················· | |
| 8933 | MW185295.1 | \|Severe acute respiratory syndrome coro | ···················· | ···················· | ···················· | ···················· | ···················· | ···················· | |
| 8934 | MW185296.1 | \|Severe acute respiratory syndrome coro | ···················· | ···················· | ···················· | ···················· | ···················· | ···················· | |
| 8935 | MW185297.1 | \|Severe acute respiratory syndrome coro | ···················· | ···················· | ···················· | ···················· | ···················· | ···················· | |
| 8936 | MW185300.1 | \|Severe acute respiratory syndrome coro | ···················· | ···················· | ···················· | ···················· | ···················· | ···················· | |
| 8937 | MW185301.1 | \|Severe acute respiratory syndrome coro | ···················· | ···················· | ···················· | ···················· | ···················· | ···················· | |
| 8938 | MW185303.1 | \|Severe acute respiratory syndrome coro | ···················· | ···················· | ···················· | ···················· | ···················· | ···················· | |
| 8939 | MW185306.1 | \|Severe acute respiratory syndrome coro | ···················· | ···················· | ···················· | ···················· | ···················· | ···················· | |
| 8940 | MW185308.1 | \|Severe acute respiratory syndrome coro | ···················· | ···················· | ···················· | ···················· | ···················· | ···················· | |
| 8941 | MW185309.1 | \|Severe acute respiratory syndrome coro | ···················· | ···················· | ···················· | ···················· | ···················· | ···················· | |
| 8942 | MW185310.1 | \|Severe acute respiratory syndrome coro | ···················· | ···················· | ···················· | ···················· | ···················· | ···················· | |
| 8943 | MW185311.1 | \|Severe acute respiratory syndrome coro | ···················· | ···················· | ···················· | ···················· | ···················· | ···················· | |
| 8944 | MW185312.1 | \|Severe acute respiratory syndrome coro | ···················· | ···················· | ···················· | ···················· | ···················· | ···················· | |
| 8945 | MW185313.1 | \|Severe acute respiratory syndrome coro | ···················· | ···················· | ···················· | ···················· | ···················· | ···················· | |
| 8946 | MW185314.1 | \|Severe acute respiratory syndrome coro | ···················· | ···················· | ···················· | ···················· | ···················· | ···················· | |
| 8947 | MW185316.1 | \|Severe acute respiratory syndrome coro | ···················· | ···················· | ···················· | ···················· | ···················· | ···················· | |
| 8948 | MW185318.1 | \|Severe acute respiratory syndrome coro | ···················· | ···················· | ···················· | ···················· | ···················· | ···················· | |
| 8949 | MW185320.1 | \|Severe acute respiratory syndrome coro | ···················· | ···················· | ···················· | ···················· | ···················· | ···················· | |
| 8950 | MW185321.1 | \|Severe acute respiratory syndrome coro | ···················· | ···················· | ···················· | ···················· | ···················· | ···················· | |
| 8951 | MW185322.1 | \|Severe acute respiratory syndrome coro | ···················· | ···················· | ···················· | ···················· | ···················· | ···················· | |
| 8952 | MW185323.1 | \|Severe acute respiratory syndrome coro | ···················· | ···················· | ···················· | ···················· | ···················· | ···················· | |
| 8954 | MW185327.1 | \|Severe acute respiratory syndrome coro | ···················· | ···················· | ···················· | ···················· | ···················· | ···················· | |
| 8955 | MW185329.1 | \|Severe acute respiratory syndrome coro | ···················· | ···················· | ···················· | ···················· | ···················· | ···················· | |
| 8956 | MW185330.1 | \|Severe acute respiratory syndrome coro | ···················· | ···················· | ···················· | ···················· | ···················· | ···················· | |
| 8957 | MW185334.1 | \|Severe acute respiratory syndrome coro | ···················· | ···················· | ···················· | ···················· | ···················· | ···················· | |
| 8958 | MW185335.1 | \|Severe acute respiratory syndrome coro | ···················· | ···················· | ···················· | ···················· | ···················· | ···················· | |
| 8959 | MW185336.1 | \|Severe acute respiratory syndrome coro | ···················· | ···················· | ···················· | ···················· | ···················· | ···················· | |
| 8960 | MW185339.1 | \|Severe acute respiratory syndrome coro | ···················· | ···················· | ···················· | ···················· | ···················· | ···················· | |
| 8961 | MW185343.1 | \|Severe acute respiratory syndrome coro | ···················· | ···················· | ···················· | ···················· | ···················· | ···················· | |
| 8962 | MW185344.1 | \|Severe acute respiratory syndrome coro | ···················· | ···················· | ···················· | ···················· | ···················· | ···················· | |
| 8963 | MW185345.1 | \|Severe acute respiratory syndrome coro | ···················· | ···················· | ···················· | ···················· | ···················· | ···················· | |
| 8964 | MW185346.1 | \|Severe acute respiratory syndrome coro | ···················· | ···················· | ···················· | ···················· | ···················· | ···················· | |
| 8965 | MW185347.1 | \|Severe acute respiratory syndrome coro | ···················· | ···················· | ···················· | ···················· | ···················· | NNN················· | 1 |
| 8966 | MW185348.1 | \|Severe acute respiratory syndrome coro | ···················· | ···················· | ···················· | ···················· | ···················· | ···················· | |
| 8967 | MW185349.1 | \|Severe acute respiratory syndrome coro | ···················· | ···················· | ···················· | ···················· | NNN················· | ···················· | 1 |
| 8968 | MW185350.1 | \|Severe acute respiratory syndrome coro | ···················· | ···················· | ···················· | ···················· | ···················· | ···················· | |

```
8969 MW185351.1 |Severe acute respiratory syndrome coro   .................... .................... .................... .................... .................... ....................
8970 MW185352.1 |Severe acute respiratory syndrome coro   .................... .................... .................... .................... .................... ....................
8971 MW185354.1 |Severe acute respiratory syndrome coro   .................... .................... .................... .................... .................... ....................
8972 MW185356.1 |Severe acute respiratory syndrome coro   .................... .................... .................... .................... .................... ....................
8973 MW185357.1 |Severe acute respiratory syndrome coro   .................... .................... .................... .................... .................... ....................
8974 MW185359.1 |Severe acute respiratory syndrome coro   .................... .................... .................... .................... .................... ....................
8975 MW185360.1 |Severe acute respiratory syndrome coro   .................... .................... .................... .................... .................... ....................
8976 MW185362.1 |Severe acute respiratory syndrome coro   .................... .................... .................... .................... .................... ....................
8977 MW185363.1 |Severe acute respiratory syndrome coro   .................... .................... .................... .................... .................... ....................
8978 MW185364.1 |Severe acute respiratory syndrome coro   .................... .................... .................... .................... .................... ....................
8979 MW185365.1 |Severe acute respiratory syndrome coro   .................... .................... .................... .................... .................... ....................
8981 MW185367.1 |Severe acute respiratory syndrome coro   .................... .................... .................... .................... .................... ....................
8982 MW185368.1 |Severe acute respiratory syndrome coro   .................... .................... .................... .................... .................... ....................
8983 MW185369.1 |Severe acute respiratory syndrome coro   .................... .................... .................... .................... .................... ....................
8984 MW185370.1 |Severe acute respiratory syndrome coro   .................... .................... .................... .................... .................... ....................
8985 MW185371.1 |Severe acute respiratory syndrome coro   .................... .................... .................... .................... .................... ....................
8986 MW185372.1 |Severe acute respiratory syndrome coro   .................... .................... .................... .................... .................... ....................
8987 MW185373.1 |Severe acute respiratory syndrome coro   .................... .................... .................... .................... .................... ....................
8988 MW185374.1 |Severe acute respiratory syndrome coro   .................... .................... .................... .................... .................... ....................
8989 MW185375.1 |Severe acute respiratory syndrome coro   .................... .................... .................... .................... .................... ....................
8990 MW185376.1 |Severe acute respiratory syndrome coro   .................... .................... .................... .................... .................... ....................
8991 MW185377.1 |Severe acute respiratory syndrome coro   .................... .................... .................... .................... .................... ....................
8992 MW185379.1 |Severe acute respiratory syndrome coro   .................... .................... .................... .................... .................... ....................
8993 MW185380.1 |Severe acute respiratory syndrome coro   .................... .................... .................... .................... .................... ....................
8994 MW185381.1 |Severe acute respiratory syndrome coro   .................... .................... .................... .................... .................... ....................
8995 MW185382.1 |Severe acute respiratory syndrome coro   .................... .................... .................... .................... .................... ....................
8996 MW185386.1 |Severe acute respiratory syndrome coro   .................... .................... .................... .................... .................... ....................
8997 MW185389.1 |Severe acute respiratory syndrome coro   .................... .................... .................... .................... .................... ....................
8998 MW185391.1 |Severe acute respiratory syndrome coro   .................... .................... .................... .................... .................... ....................
8999 MW185392.1 |Severe acute respiratory syndrome coro   .................... .................... .................... .................... .................... ....................
9000 MW185393.1 |Severe acute respiratory syndrome coro   .................... .................... .................... .................... .................... ....................
9001 MW185394.1 |Severe acute respiratory syndrome coro   .................... .................... .................... .................... .................... ....................
9002 MW185395.1 |Severe acute respiratory syndrome coro   .................... .................... .................... .................... .................... ....................
9003 MW185396.1 |Severe acute respiratory syndrome coro   .................... .................... .................... .................... .................... ....................
9004 MW185397.1 |Severe acute respiratory syndrome coro   .................... .................... .................... .................... .................... ....................
9005 MW185398.1 |Severe acute respiratory syndrome coro   .................... .................... .................... .................... .................... ....................
9006 MW185399.1 |Severe acute respiratory syndrome coro   .................... .................... .................... .................... .................... ....................
9007 MW185400.1 |Severe acute respiratory syndrome coro   .................... .................... .................... .................... .................... ....................
9008 MW185401.1 |Severe acute respiratory syndrome coro   .................... .................... .................... .................... .................... ....................
9009 MW185402.1 |Severe acute respiratory syndrome coro   .................... .................... .................... .................... .................... ....................
9010 MW185403.1 |Severe acute respiratory syndrome coro   .................... .................... .................... .................... .................... ....................
9011 MW185406.1 |Severe acute respiratory syndrome coro   .................... .................... .................... .................... .................... ....................
9012 MW185408.1 |Severe acute respiratory syndrome coro   .................... .................... .................... .................... .................... ....................
9013 MW185409.1 |Severe acute respiratory syndrome coro   .................... .................... .................... .................... .................... ....................
9014 MW185410.1 |Severe acute respiratory syndrome coro   .................... .................... .................... .................... .................... ....................
9015 MW185411.1 |Severe acute respiratory syndrome coro   .................... .................... .................... .................... .................... ....................
9016 MW185414.1 |Severe acute respiratory syndrome coro   .................... .................... .................... .................... .................... ....................
9017 MW185415.1 |Severe acute respiratory syndrome coro   .................... .................... .................... .................... .................... ....................
9018 MW185416.1 |Severe acute respiratory syndrome coro   .................... .................... .................... .................... .................... ....................
9019 MW185417.1 |Severe acute respiratory syndrome coro   .................... .................... .................... .................... .................... ....................
9020 MW185418.1 |Severe acute respiratory syndrome coro   .................... .................... .................... .................... .................... ....................
9021 MW185419.1 |Severe acute respiratory syndrome coro   .................... .................... .................... .................... .................... ....................
9022 MW185421.1 |Severe acute respiratory syndrome coro   .................... .................... .................... .................... .................... ....................
9024 MW185423.1 |Severe acute respiratory syndrome coro   .................... .................... .................... .................... .................... ....................
9025 MW185424.1 |Severe acute respiratory syndrome coro   .................... .................... .................... .................... .................... ....................
9026 MW185427.1 |Severe acute respiratory syndrome coro   .................... .................... .................... .................... .................... ....................
9027 MW185428.1 |Severe acute respiratory syndrome coro   .................... .................... .................... .................... .................... ....................
9028 MW185432.1 |Severe acute respiratory syndrome coro   .................... .................... .................... .................... .................... ....................
9029 MW185433.1 |Severe acute respiratory syndrome coro   .................... .................... .................... .................... .................... ....................
9030 MW185434.1 |Severe acute respiratory syndrome coro   .................... .................... .................... .................... .................... ....................
9031 MW185435.1 |Severe acute respiratory syndrome coro   .................... .................... .................... .................... .................... ....................
9032 MW185436.1 |Severe acute respiratory syndrome coro   .................... .................... .................... .................... .................... ....................
9033 MW185437.1 |Severe acute respiratory syndrome coro   .................... .................... .................... .................... .................... ....................
9034 MW185440.1 |Severe acute respiratory syndrome coro   .................... .................... .................... .................... .................... ....................
9035 MW185441.1 |Severe acute respiratory syndrome coro   .................... .................... .................... .................... .................... ....................
9036 MW185442.1 |Severe acute respiratory syndrome coro   .................... .................... .................... .................... .................... ....................
9037 MW185443.1 |Severe acute respiratory syndrome coro   .................... .................... .................... .................... .................... ....................
9038 MW185444.1 |Severe acute respiratory syndrome coro   .................... .................... .................... .................... .................... ....................
9039 MW185445.1 |Severe acute respiratory syndrome coro   .................... .................... .................... .................... .................... ....................
9040 MW185446.1 |Severe acute respiratory syndrome coro   .................... .................... .................... .................... .................... ....................
9041 MW185448.1 |Severe acute respiratory syndrome coro   .................... .................... .................... .................... .................... ....................
9042 MW185450.1 |Severe acute respiratory syndrome coro   .................... .................... .................... .................... .................... ....................
9043 MW185451.1 |Severe acute respiratory syndrome coro   .................... .................... .................... .................... .................... ....................
9044 MW185453.1 |Severe acute respiratory syndrome coro   .................... .................... .................... .................... .................... ....................
9045 MW185454.1 |Severe acute respiratory syndrome coro   .................... .................... .................... .................... .................... ....................
9046 MW185458.1 |Severe acute respiratory syndrome coro   .................... .................... .................... .................... .................... ....................
9047 MW185459.1 |Severe acute respiratory syndrome coro   .................... .................... .................... .................... .................... ....................
9048 MW185462.1 |Severe acute respiratory syndrome coro   .................... .................... .................... .................... .................... ....................
```

```
9049 MW185463.1 |Severe acute respiratory syndrome coro    ....................  ....................  .....................  ....................  ....................  ....................
9050 MW185466.1 |Severe acute respiratory syndrome coro    ....................  ....................  .....................  ....................  ....................  ....................
9051 MW185467.1 |Severe acute respiratory syndrome coro    ....................  ....................  .....................  ....................  ....................  ....................
9052 MW185468.1 |Severe acute respiratory syndrome coro    ....................  ....................  .....................  ....................  ....................  ....................
9053 MW185469.1 |Severe acute respiratory syndrome coro    ....................  ....................  .....................  ....................  ....................  ....................
9054 MW185470.1 |Severe acute respiratory syndrome coro    ....................  ....................  .....................  ....................  ....................  ....................
9055 MW185471.1 |Severe acute respiratory syndrome coro    ....................  ....................  .....................  ....................  ....................  ....................
9056 MW185472.1 |Severe acute respiratory syndrome coro    ....................  ....................  .....................  ....................  ....................  ....................
9057 MW185477.1 |Severe acute respiratory syndrome coro    ....................  ....................  .....................  ....................  ....................  ....................
9058 MW185478.1 |Severe acute respiratory syndrome coro    ....................  ....................  .....................  ....................  ....................  ....................
9059 MW185479.1 |Severe acute respiratory syndrome coro    ....................  ....................  .....................  ....................  ....................  ....................
9060 MW185480.1 |Severe acute respiratory syndrome coro    ....................  ....................  .....................  ....................  ....................  ....................
9061 MW185482.1 |Severe acute respiratory syndrome coro    ....................  ....................  .....................  ....................  ....................  ....................
9062 MW185484.1 |Severe acute respiratory syndrome coro    ....................  ....................  .....................  ....................  ....................  ....................
9063 MW185488.1 |Severe acute respiratory syndrome coro    ....................  ....................  .....................  ....................  ....................  ....................
9064 MW185489.1 |Severe acute respiratory syndrome coro    ....................  ....................  .....................  ....................  ....................  ....................
9065 MW185490.1 |Severe acute respiratory syndrome coro    ....................  ....................  .....................  ....................  ....................  ....................
9066 MW185491.1 |Severe acute respiratory syndrome coro    ....................  ....................  .....................  ....................  ....................  ....................
9067 MW185492.1 |Severe acute respiratory syndrome coro    ....................  ....................  .....................  ....................  ....................  ....................
9068 MW185493.1 |Severe acute respiratory syndrome coro    ....................  ....................  .....................  ....................  ....................  ....................
9069 MW185494.1 |Severe acute respiratory syndrome coro    ....................  ....................  .....................  ....................  ....................  ....................
9070 MW185496.1 |Severe acute respiratory syndrome coro    ....................  ....................  .....................  ....................  ....................  ....................
9071 MW185497.1 |Severe acute respiratory syndrome coro    ....................  ....................  .....................  ....................  ....................  ....................
9072 MW185498.1 |Severe acute respiratory syndrome coro    ....................  ....................  .....................  ....................  ....................  ....................
9073 MW185499.1 |Severe acute respiratory syndrome coro    ....................  ....................  .....................  ....................  ....................  ....................
9074 MW185500.1 |Severe acute respiratory syndrome coro    ....................  ....................  .....................  ....................  ....................  ....................
9075 MW185501.1 |Severe acute respiratory syndrome coro    ....................  ....................  .....................  ....................  ....................  ....................
9076 MW185503.1 |Severe acute respiratory syndrome coro    ....................  ....................  .....................  ....................  ....................  ....................
9077 MW185505.1 |Severe acute respiratory syndrome coro    ....................  ....................  .....................  ....................  ....................  ....................
9078 MW185506.1 |Severe acute respiratory syndrome coro    ....................  ....................  .....................  ....................  ....................  ....................
9079 MW185507.1 |Severe acute respiratory syndrome coro    ....................  ....................  .....................  ....................  ....................  ....................
9080 MW185508.1 |Severe acute respiratory syndrome coro    ....................  ....................  .....................  ....................  ....................  ....................
9082 MW185510.1 |Severe acute respiratory syndrome coro    ....................  ....................  .....................  ....................  ....................  ....................
9083 MW185513.1 |Severe acute respiratory syndrome coro    ....................  ....................  .....................  ....................  ....................  ....................
9084 MW185514.1 |Severe acute respiratory syndrome coro    ....................  ....................  .....................  ....................  ....................  ....................
9085 MW185515.1 |Severe acute respiratory syndrome coro    ....................  ....................  .....................  ....................  ....................  ....................
9086 MW185516.1 |Severe acute respiratory syndrome coro    ....................  ....................  .....................  ....................  ....................  ....................
9087 MW185517.1 |Severe acute respiratory syndrome coro    ....................  ....................  .....................  ....................  ....................  ....................
9088 MW185518.1 |Severe acute respiratory syndrome coro    ....................  ....................  .....................  ....................  ....................  ....................
9089 MW185520.1 |Severe acute respiratory syndrome coro    ....................  ....................  .....................  ....................  ....................  ....................
9090 MW185521.1 |Severe acute respiratory syndrome coro    ....................  ....................  .....................  ....................  ....................  ....................
9091 MW185523.1 |Severe acute respiratory syndrome coro    ....................  ....................  .....................  ....................  ....................  ....................
9092 MW185525.1 |Severe acute respiratory syndrome coro    ....................  ....................  .....................  ....................  ....................  ....................
9093 MW185526.1 |Severe acute respiratory syndrome coro    ....................  ....................  .....................  ....................  ....................  ....................
9094 MW185527.1 |Severe acute respiratory syndrome coro    ....................  ....................  .....................  ....................  ....................  ....................
9095 MW185528.1 |Severe acute respiratory syndrome coro    ....................  ....................  .....................  ....................  ....................  ....................
9096 MW185531.1 |Severe acute respiratory syndrome coro    ....................  ....................  .....................  ....................  ....................  ....................
9097 MW185532.1 |Severe acute respiratory syndrome coro    ....................  ....................  .....................  ....................  ....................  ....................
9098 MW185534.1 |Severe acute respiratory syndrome coro    ....................  ....................  .....................  ....................  ....................  ....................
9099 MW185535.1 |Severe acute respiratory syndrome coro    ....................  ....................  .....................  ....................  ....................  ....................
9100 MW185538.1 |Severe acute respiratory syndrome coro    ....................  ....................  .....................  ....................  ....................  ....................
9101 MW185539.1 |Severe acute respiratory syndrome coro    ....................  ....................  .....................  ....................  ....................  ....................
9102 MW185540.1 |Severe acute respiratory syndrome coro    ....................  ....................  .....................  ....................  ....................  ....................
9103 MW185541.1 |Severe acute respiratory syndrome coro    ....................  ....................  .....................  ....................  ....................  ....................
9104 MW185542.1 |Severe acute respiratory syndrome coro    ....................  ....................  .....................  ....................  ....................  ....................
9105 MW185543.1 |Severe acute respiratory syndrome coro    ....................  ....................  .....................  ....................  ....................  ....................
9106 MW185546.1 |Severe acute respiratory syndrome coro    ....................  ....................  .....................  ....................  ....................  ....................
9107 MW185547.1 |Severe acute respiratory syndrome coro    ....................  ....................  .....................  ....................  ....................  ....................
9108 MW185548.1 |Severe acute respiratory syndrome coro    ....................  ....................  .....................  ....................  ....................  ....................
9109 MW185549.1 |Severe acute respiratory syndrome coro    ....................  ....................  .....................  ....................  ....................  ....................
9110 MW185550.1 |Severe acute respiratory syndrome coro    ....................  ....................  .....................  ....................  ....................  ....................
9111 MW185552.1 |Severe acute respiratory syndrome coro    ....................  ....................  .....................  ....................  ....................  ....................
9112 MW185553.1 |Severe acute respiratory syndrome coro    ....................  ....................  .....................  ....................  ....................  ....................
9113 MW185554.1 |Severe acute respiratory syndrome coro    ....................  ....................  .....................  ....................  ....................  ....................
9114 MW185556.1 |Severe acute respiratory syndrome coro    ....................  ....................  .....................  ....................  ....................  ....................
9115 MW185557.1 |Severe acute respiratory syndrome coro    ....................  ....................  .....................  ....................  ....................  ....................
9116 MW185558.1 |Severe acute respiratory syndrome coro    ....................  ....................  .....................  ....................  ....................  ....................
9117 MW185559.1 |Severe acute respiratory syndrome coro    ....................  ....................  .....................  ....................  ....................  ....................
9118 MW185560.1 |Severe acute respiratory syndrome coro    ....................  ....................  .....................  ....................  ....................  ....................
9119 MW185562.1 |Severe acute respiratory syndrome coro    ....................  ....................  .....................  ....................  ....................  ....................
9120 MW185563.1 |Severe acute respiratory syndrome coro    ....................  ....................  .....................  ....................  ....................  ....................
9121 MW185565.1 |Severe acute respiratory syndrome coro    ....................  ....................  .....................  ....................  ....................  ....................
9122 MW185568.1 |Severe acute respiratory syndrome coro    ....................  ....................  .....................  ....................  ....................  ....................
9123 MW185569.1 |Severe acute respiratory syndrome coro    ....................  ....................  .....................  ....................  ....................  ....................
9124 MW185570.1 |Severe acute respiratory syndrome coro    ....................  ....................  .....................  ....................  ....................  ....................
9125 MW185572.1 |Severe acute respiratory syndrome coro    ....................  ....................  .....................  ....................  ....................  ....................
9126 MW185573.1 |Severe acute respiratory syndrome coro    ....................  ....................  .....................  ....................  ....................  ....................
9127 MW185574.1 |Severe acute respiratory syndrome coro    ....................  ....................  .....................  ....................  ....................  ....................
```

| | | | | | | | | | |
|---|---|---|---|---|---|---|---|---|---|
| 9128 | MW185575.1 | \|Severe acute respiratory syndrome coro | ···················· | ···················· | ···················· | ···················· | ···················· | ···················· | |
| 9129 | MW185576.1 | \|Severe acute respiratory syndrome coro | ···················· | ···················· | ···················· | ···················· | ···················· | ···················· | |
| 9130 | MW185578.1 | \|Severe acute respiratory syndrome coro | ···················· | ···················· | ···················· | ···················· | ···················· | ···················· | |
| 9131 | MW185579.1 | \|Severe acute respiratory syndrome coro | ···················· | ···················· | ···················· | ···················· | ···················· | ···················· | |
| 9132 | MW185580.1 | \|Severe acute respiratory syndrome coro | ···················· | ···················· | ···················· | ···················· | ···················· | ···················· | |
| 9133 | MW185581.1 | \|Severe acute respiratory syndrome coro | ···················· | ···················· | ···················· | ···················· | ···················· | ···················· | |
| 9134 | MW185582.1 | \|Severe acute respiratory syndrome coro | ···················· | ···················· | ···················· | ···················· | ···················· | ···················· | |
| 9135 | MW185584.1 | \|Severe acute respiratory syndrome coro | ···················· | ···················· | ···················· | ···················· | ···················· | ···················· | |
| 9136 | MW185585.1 | \|Severe acute respiratory syndrome coro | ···················· | ···················· | ···················· | ···················· | ···················· | ···················· | |
| 9137 | MW185586.1 | \|Severe acute respiratory syndrome coro | ···················· | ···················· | ···················· | ···················· | ···················· | ···················· | |
| 9138 | MW185587.1 | \|Severe acute respiratory syndrome coro | ···················· | ···················· | ···················· | ···················· | ···················· | ···················· | |
| 9139 | MW185588.1 | \|Severe acute respiratory syndrome coro | ···················· | ···················· | ···················· | ···················· | ···················· | ···················· | |
| 9140 | MW185589.1 | \|Severe acute respiratory syndrome coro | ···················· | ···················· | ···················· | ···················· | ···················· | ···················· | |
| 9141 | MW185590.1 | \|Severe acute respiratory syndrome coro | ···················· | ···················· | ···················· | ···················· | ···················· | ···················· | |
| 9142 | MW185593.1 | \|Severe acute respiratory syndrome coro | ···················· | ···················· | ···················· | ···················· | ···················· | ···················· | |
| 9143 | MW185595.1 | \|Severe acute respiratory syndrome coro | ···················· | ···················· | ···················· | ···················· | ···················· | ···················· | |
| 9144 | MW185596.1 | \|Severe acute respiratory syndrome coro | ···················· | ···················· | ···················· | ···················· | ···················· | ···················· | |
| 9145 | MW185597.1 | \|Severe acute respiratory syndrome coro | ···················· | ···················· | ···················· | ···················· | ···················· | ···················· | |
| 9146 | MW185598.1 | \|Severe acute respiratory syndrome coro | ···················· | ···················· | ···················· | ···················· | ···················· | ···················· | |
| 9147 | MW185599.1 | \|Severe acute respiratory syndrome coro | ···················· | ···················· | ···················· | ···················· | ···················· | ···················· | |
| 9148 | MW185600.1 | \|Severe acute respiratory syndrome coro | ···················· | ···················· | ···················· | ···················· | ···················· | ···················· | |
| 9149 | MW185603.1 | \|Severe acute respiratory syndrome coro | ···················· | ···················· | ···················· | ···················· | ···················· | ···················· | |
| 9150 | MW185606.1 | \|Severe acute respiratory syndrome coro | ···················· | ···················· | ···················· | ···················· | ···················· | ···················· | |
| 9151 | MW185607.1 | \|Severe acute respiratory syndrome coro | ···················· | ···················· | ···················· | ···················· | ···················· | ···················· | |
| 9152 | MW185608.1 | \|Severe acute respiratory syndrome coro | ···················· | ···················· | ···················· | ···················· | ···················· | ···················· | |
| 9153 | MW185609.1 | \|Severe acute respiratory syndrome coro | ···················· | ···················· | ···················· | ···················· | ···················· | ···················· | |
| 9154 | MW185611.1 | \|Severe acute respiratory syndrome coro | ···················· | ···················· | ···················· | ···················· | ···················· | ···················· | |
| 9155 | MW185614.1 | \|Severe acute respiratory syndrome coro | ···················· | ···················· | ···················· | ···················· | ···················· | ···················· | |
| 9156 | MW185615.1 | \|Severe acute respiratory syndrome coro | ···················· | ···················· | ···················· | ···················· | ···················· | ···················· | |
| 9157 | MW185617.1 | \|Severe acute respiratory syndrome coro | ···················· | ···················· | ···················· | ···················· | ···················· | ···················· | |
| 9158 | MW185618.1 | \|Severe acute respiratory syndrome coro | ···················· | ···················· | ···················· | ···················· | ···················· | ···················· | |
| 9159 | MW185619.1 | \|Severe acute respiratory syndrome coro | ···················· | ···················· | ···················· | ···················· | ···················· | ···················· | |
| 9160 | MW185620.1 | \|Severe acute respiratory syndrome coro | ···················· | ···················· | ···················· | ···················· | ···················· | ···················· | |
| 9161 | MW185621.1 | \|Severe acute respiratory syndrome coro | ···················· | ···················· | ···················· | ···················· | ···················· | ···················· | |
| 9162 | MW185624.1 | \|Severe acute respiratory syndrome coro | ···················· | ···················· | ···················· | ···················· | ···················· | ···················· | |
| 9163 | MW185625.1 | \|Severe acute respiratory syndrome coro | ···················· | ···················· | ···················· | ···················· | ···················· | ···················· | |
| 9164 | MW185627.1 | \|Severe acute respiratory syndrome coro | ···················· | ···················· | ···················· | ···················· | ···················· | ···················· | |
| 9165 | MW185628.1 | \|Severe acute respiratory syndrome coro | ···················· | ···················· | ···················· | ···················· | ···················· | ···················· | |
| 9166 | MW185629.1 | \|Severe acute respiratory syndrome coro | ···················· | ···················· | ···················· | ···················· | ···················· | ···················· | |
| 9167 | MW185630.1 | \|Severe acute respiratory syndrome coro | ···················· | ···················· | ···················· | ···················· | ···················· | ···················· | |
| 9168 | MW185632.1 | \|Severe acute respiratory syndrome coro | ···················· | ······A············· | ···················· | ···················· | ···················· | ···················· | |
| 9169 | MW185636.1 | \|Severe acute respiratory syndrome coro | ···················· | ···················· | ···················· | ···················· | ···················· | ···················· | |
| 9170 | MW185638.1 | \|Severe acute respiratory syndrome coro | ···················· | ···················· | ···················· | ···················· | ···················· | ···················· | |
| 9171 | MW185640.1 | \|Severe acute respiratory syndrome coro | ···················· | ···················· | ···················· | ···················· | ···················· | ···················· | |
| 9172 | MW185641.1 | \|Severe acute respiratory syndrome coro | ···················· | ···················· | ···················· | ···················· | ···················· | ···················· | |
| 9173 | MW185642.1 | \|Severe acute respiratory syndrome coro | ···················· | ···················· | ···················· | ···················· | ···················· | ···················· | |
| 9174 | MW185643.1 | \|Severe acute respiratory syndrome coro | ···················· | ···················· | ···················· | ···················· | ···················· | ···················· | |
| 9175 | MW185645.1 | \|Severe acute respiratory syndrome coro | ···················· | ···················· | ···················· | ···················· | ···················· | ···················· | |
| 9176 | MW185646.1 | \|Severe acute respiratory syndrome coro | ···················· | ···················· | ···················· | ···················· | ···················· | ···················· | |
| 9177 | MW185647.1 | \|Severe acute respiratory syndrome coro | ···················· | ···················· | ···················· | ···················· | ···················· | ···················· | |
| 9178 | MW185648.1 | \|Severe acute respiratory syndrome coro | ···················· | ···················· | ···················· | ···················· | ···················· | ···················· | |
| 9179 | MW185649.1 | \|Severe acute respiratory syndrome coro | ···················· | ···················· | ···················· | ···················· | ···················· | ···················· | |
| 9180 | MW185650.1 | \|Severe acute respiratory syndrome coro | ···················· | ···················· | ···················· | ···················· | ···················· | ···················· | |
| 9181 | MW185652.1 | \|Severe acute respiratory syndrome coro | ···················· | ···················· | ···················· | ···················· | ···················· | ···················· | |
| 9182 | MW185653.1 | \|Severe acute respiratory syndrome coro | ···················· | ···················· | ···················· | ···················· | ···················· | ···················· | |
| 9183 | MW185654.1 | \|Severe acute respiratory syndrome coro | ···················· | ···················· | ···················· | ···················· | ···················· | ···················· | |
| 9184 | MW185655.1 | \|Severe acute respiratory syndrome coro | ···················· | ···················· | ···················· | ···················· | ···················· | ···················· | |
| 9185 | MW185657.1 | \|Severe acute respiratory syndrome coro | ···················· | ···················· | ···················· | ···················· | ···················· | ···················· | |
| 9186 | MW185658.1 | \|Severe acute respiratory syndrome coro | ···················· | ···················· | ···················· | ···················· | ···················· | ···················· | |
| 9187 | MW185659.1 | \|Severe acute respiratory syndrome coro | ···················· | ···················· | ···················· | ···················· | ···················· | ···················· | |
| 9188 | MW185660.1 | \|Severe acute respiratory syndrome coro | ···················· | ···················· | ···················· | ···················· | ···················· | ···················· | |
| 9189 | MW185661.1 | \|Severe acute respiratory syndrome coro | ···················· | ···················· | ···················· | ···················· | ···················· | ···················· | |
| 9190 | MW185662.1 | \|Severe acute respiratory syndrome coro | ···················· | ···················· | ···················· | ···················· | ···················· | ···················· | |
| 9191 | MW185663.1 | \|Severe acute respiratory syndrome coro | ···················· | ···················· | ···················· | ···················· | ···················· | ···················· | |
| 9192 | MW185664.1 | \|Severe acute respiratory syndrome coro | ···················· | ···················· | ···················· | ···················· | ···················· | ···················· | |
| 9193 | MW185665.1 | \|Severe acute respiratory syndrome coro | ···················· | ···················· | ···················· | ···················· | ···················· | ···················· | |
| 9194 | MW185666.1 | \|Severe acute respiratory syndrome coro | ···················· | ···················· | ···················· | ···················· | ··················NN | ···················· | 1 |
| 9195 | MW185667.1 | \|Severe acute respiratory syndrome coro | ···················· | ···················· | ···················· | ···················· | ···················· | ···················· | |
| 9196 | MW185669.1 | \|Severe acute respiratory syndrome coro | ···················· | ···················· | ···················· | ···················· | ···················· | ···················· | |
| 9197 | MW185670.1 | \|Severe acute respiratory syndrome coro | ···················· | ···················· | ···················· | ···················· | ···················· | ···················· | |
| 9198 | MW185671.1 | \|Severe acute respiratory syndrome coro | ···················· | ···················· | ···················· | ···················· | ···················· | ···················· | |
| 9199 | MW185672.1 | \|Severe acute respiratory syndrome coro | ···················· | ···················· | ···················· | ···················· | ···················· | ···················· | |
| 9200 | MW185674.1 | \|Severe acute respiratory syndrome coro | ···················· | ···················· | ···················· | ···················· | ···················· | ···················· | |
| 9201 | MW185675.1 | \|Severe acute respiratory syndrome coro | ···················· | ···················· | ···················· | ···················· | ···················· | ···················· | |
| 9202 | MW185676.1 | \|Severe acute respiratory syndrome coro | ···················· | ···················· | ···················· | ···················· | ···················· | ···················· | |
| 9203 | MW185677.1 | \|Severe acute respiratory syndrome coro | ···················· | ···················· | ···················· | ···················· | ···················· | ···················· | |
| 9204 | MW185678.1 | \|Severe acute respiratory syndrome coro | ···················· | ···················· | ···················· | ···················· | ···················· | ···················· | |
| 9205 | MW185679.1 | \|Severe acute respiratory syndrome coro | ···················· | ···················· | ···················· | ···················· | ···················· | ···················· | |

9206 MW185680.1 |Severe acute respiratory syndrome coro ···················· ···················· ···················· ···················· ···················· ····················
9207 MW185681.1 |Severe acute respiratory syndrome coro ···················· ···················· ···················· ···················· ···················· ····················
9208 MW185682.1 |Severe acute respiratory syndrome coro ···················· ···················· ···················· ···················· ···················· ····················
9209 MW185683.1 |Severe acute respiratory syndrome coro ···················· ···················· ···················· ···················· ···················· ····················
9210 MW185685.1 |Severe acute respiratory syndrome coro ···················· ···················· ···················· ···················· ···················· ····················
9211 MW185686.1 |Severe acute respiratory syndrome coro ···················· ···················· ···················· ···················· ···················· ····················
9212 MW185688.1 |Severe acute respiratory syndrome coro ···················· ···················· ···················· ···················· ···················· ····················
9213 MW185691.1 |Severe acute respiratory syndrome coro ···················· ···················· ···················· ···················· ···················· ····················
9214 MW185692.1 |Severe acute respiratory syndrome coro ···················· ···················· ···················· ···················· ···················· ····················
9215 MW185694.1 |Severe acute respiratory syndrome coro ···················· ···················· ···················· ···················· ···················· ····················
9216 MW185695.1 |Severe acute respiratory syndrome coro ···················· ···················· ···················· ···················· ···················· ····················
9217 MW185696.1 |Severe acute respiratory syndrome coro ···················· ···················· ···················· ···················· ···················· ····················
9218 MW185697.1 |Severe acute respiratory syndrome coro ···················· ···················· ···················· ···················· ···················· ····················
9219 MW185698.1 |Severe acute respiratory syndrome coro ···················· ···················· ···················· ···················· ···················· ····················
9220 MW185699.1 |Severe acute respiratory syndrome coro ···················· ···················· ···················· ···················· ···················· ····················
9221 MW185700.1 |Severe acute respiratory syndrome coro ···················· ···················· ···················· ···················· ···················· ····················
9222 MW185701.1 |Severe acute respiratory syndrome coro ···················· ···················· ···················· ···················· ···················· ····················
9223 MW185703.1 |Severe acute respiratory syndrome coro ···················· ···················· ···················· ···················· ···················· ····················
9224 MW185704.1 |Severe acute respiratory syndrome coro ···················· ···················· ···················· ···················· ···················· ····················
9225 MW185705.1 |Severe acute respiratory syndrome coro ···················· ···················· ···················· ···················· ···················· ····················
9226 MW185706.1 |Severe acute respiratory syndrome coro ···················· ···················· ···················· ···················· ···················· ····················
9227 MW185707.1 |Severe acute respiratory syndrome coro ···················· ···················· ···················· ···················· ···················· ····················
9228 MW185708.1 |Severe acute respiratory syndrome coro ···················· ···················· ···················· ···················· ···················· ····················
9229 MW185709.1 |Severe acute respiratory syndrome coro ···················· ···················· ···················· ···················· ···················· ····················
9230 MW185710.1 |Severe acute respiratory syndrome coro ···················· ······A············· ···················· ···················· ···················· ····················
9231 MW185711.1 |Severe acute respiratory syndrome coro ···················· ···················· ···················· ···················· ···················· ····················
9232 MW185713.1 |Severe acute respiratory syndrome coro ···················· ···················· ···················· ···················· ···················· ····················
9233 MW185714.1 |Severe acute respiratory syndrome coro ···················· ···················· ···················· ···················· ···················· ····················
9234 MW185715.1 |Severe acute respiratory syndrome coro ···················· ···················· ···················· ···················· ···················· ····················
9235 MW185716.1 |Severe acute respiratory syndrome coro ···················· ···················· ···················· ···················· ···················· ····················
9236 MW185717.1 |Severe acute respiratory syndrome coro ···················· ···················· ···················· ···················· ···················· ····················
9237 MW185718.1 |Severe acute respiratory syndrome coro ···················· ···················· ···················· ···················· ···················· ····················
9238 MW185721.1 |Severe acute respiratory syndrome coro ···················· ···················· ···················· ···················· ···················· ····················
9239 MW185722.1 |Severe acute respiratory syndrome coro ···················· ···················· ···················· ···················· ···················· ····················
9240 MW185723.1 |Severe acute respiratory syndrome coro ···················· ···················· ···················· ···················· ···················· ····················
9241 MW185725.1 |Severe acute respiratory syndrome coro ···················· ···················· ···················· ···················· ···················· ····················
9242 MW185726.1 |Severe acute respiratory syndrome coro ···················· ···················· ···················· ···················· ···················· ····················
9243 MW185729.1 |Severe acute respiratory syndrome coro ···················· ···················· ···················· ···················· ···················· ····················
9244 MW185730.1 |Severe acute respiratory syndrome coro ···················· ···················· ···················· ···················· ···················· ····················
9245 MW185731.1 |Severe acute respiratory syndrome coro ···················· ···················· ···················· ···················· ···················· ····················
9246 MW185732.1 |Severe acute respiratory syndrome coro ···················· ···················· ···················· ···················· ···················· ····················
9247 MW185733.1 |Severe acute respiratory syndrome coro ···················· ···················· ···················· ···················· ···················· ····················
9248 MW185735.1 |Severe acute respiratory syndrome coro ···················· ···················· ···················· ···················· ···················· ····················
9249 MW185737.1 |Severe acute respiratory syndrome coro ···················· ···················· ···················· ···················· ···················· ····················
9250 MW185738.1 |Severe acute respiratory syndrome coro ···················· ···················· ···················· ···················· ···················· ····················
9251 MW185739.1 |Severe acute respiratory syndrome coro ···················· ···················· ···················· ···················· ···················· ····················
9252 MW185740.1 |Severe acute respiratory syndrome coro ···················· ···················· ···················· ···················· ···················· ····················
9253 MW185741.1 |Severe acute respiratory syndrome coro ···················· ···················· ···················· ···················· ···················· ····················
9254 MW185742.1 |Severe acute respiratory syndrome coro ···················· ···················· ···················· ···················· ···················· ····················
9255 MW185743.1 |Severe acute respiratory syndrome coro ···················· ···················· ···················· ···················· ···················· ····················
9256 MW185746.1 |Severe acute respiratory syndrome coro ···················· ···················· ···················· ···················· ···················· ····················
9257 MW185747.1 |Severe acute respiratory syndrome coro ···················· ···················· ···················· ···················· ···················· ····················
9258 MW185748.1 |Severe acute respiratory syndrome coro ···················· ···················· ···················· ···················· ···················· ····················
9259 MW185749.1 |Severe acute respiratory syndrome coro ···················· ···················· ···················· ···················· ···················· ····················
9260 MW185751.1 |Severe acute respiratory syndrome coro ···················· ···················· ···················· ···················· ···················· ····················
9261 MW185752.1 |Severe acute respiratory syndrome coro ···················· ···················· ···················· ···················· ···················· ····················
9262 MW185753.1 |Severe acute respiratory syndrome coro ···················· ···················· ···················· ···················· ···················· ····················
9263 MW185755.1 |Severe acute respiratory syndrome coro ···················· ···················· ···················· ···················· ···················· ····················
9264 MW185757.1 |Severe acute respiratory syndrome coro ···················· ···················· ···················· ···················· ···················· ····················
9265 MW185758.1 |Severe acute respiratory syndrome coro ···················· ···················· ···················· ···················· ···················· ····················
9266 MW185759.1 |Severe acute respiratory syndrome coro ···················· ···················· ···················· ···················· ···················· ····················
9267 MW185760.1 |Severe acute respiratory syndrome coro ···················· ···················· ···················· ···················· ···················· ····················
9268 MW185761.1 |Severe acute respiratory syndrome coro ···················· ···················· ···················· ···················· ···················· ····················
9269 MW185763.1 |Severe acute respiratory syndrome coro ···················· ···················· ···················· ···················· ···················· ····················
9270 MW185764.1 |Severe acute respiratory syndrome coro ···················· ···················· ···················· ···················· ···················· ····················
9271 MW185765.1 |Severe acute respiratory syndrome coro ···················· ···················· ···················· ···················· ···················· ····················
9272 MW185766.1 |Severe acute respiratory syndrome coro ···················· ···················· ···················· ···················· ···················· ····················
9273 MW185767.1 |Severe acute respiratory syndrome coro ···················· ···················· ···················· ···················· ···················· ····················
9274 MW185768.1 |Severe acute respiratory syndrome coro ···················· ···················· ···················· ···················· ···················· ····················
9275 MW185769.1 |Severe acute respiratory syndrome coro ···················· ···················· ···················· ···················· ···················· ····················
9276 MW185770.1 |Severe acute respiratory syndrome coro ···················· ···················· ···················· ···················· ···················· ····················
9277 MW185771.1 |Severe acute respiratory syndrome coro ···················· ···················· ···················· ···················· ···················· ····················
9278 MW185772.1 |Severe acute respiratory syndrome coro ···················· ···················· ···················· ···················· ···················· ····················
9279 MW185773.1 |Severe acute respiratory syndrome coro ···················· ···················· ···················· ···················· ···················· ····················
9281 MW185775.1 |Severe acute respiratory syndrome coro ···················· ···················· ···················· ···················· ···················· ····················
9282 MW185776.1 |Severe acute respiratory syndrome coro ···················· ···················· ···················· ···················· ···················· ····················
9283 MW185777.1 |Severe acute respiratory syndrome coro ···················· ···················· ···················· ···················· ···················· ····················
9284 MW185778.1 |Severe acute respiratory syndrome coro ···················· ···················· ···················· ···················· ···················· ····················

```
9285 MW185779.1 |Severe acute respiratory syndrome coro    ....................  ....................  ....................  ....................  ....................  ....................
9286 MW173245.1 |Severe acute respiratory syndrome coro    ....................  ....................  ....................  ....................  ....................  ....................
9287 MW173250.1 |Severe acute respiratory syndrome coro    ....................  .A..................  ....................  ....................  ....................  ....................
9288 MW173252.1 |Severe acute respiratory syndrome coro    ....................  ....................  ....................  ....................  ....................  ....................
9289 MW173253.1 |Severe acute respiratory syndrome coro    ....................  ....................  ....................  ....................  ....................  ....................
9290 MW173254.1 |Severe acute respiratory syndrome coro    ....................  ....................  ....................  ....................  ....................  ....................
9291 MW173298.1 |Severe acute respiratory syndrome coro    ....................  .A..................  ....................  ....................  ....................  ..A.................
9292 MW173401.1 |Severe acute respiratory syndrome coro    ....................  ....................  ....................  ....................  ....................  ....................
9293 MW173402.1 |Severe acute respiratory syndrome coro    ....................  .............A......  ....................  ....................  ....................  ....................
9294 MW173404.1 |Severe acute respiratory syndrome coro    ....................  .A..................  ....................  ....................  ....................  ....................
9295 MW173406.1 |Severe acute respiratory syndrome coro    ....................  ....................  ....................  ....................  ....................  ....................
9296 MW173407.1 |Severe acute respiratory syndrome coro    ....................  ....................  ....................  ....................  ....................  ....................
9297 MW173408.1 |Severe acute respiratory syndrome coro    ....................  ....................  ....................  ....................  ....................  ....................
9298 MW173409.1 |Severe acute respiratory syndrome coro    ....................  ....................  ....................  ....................  ....................  ....................
9299 MW173410.1 |Severe acute respiratory syndrome coro    ....................  ....................  ....................  ....................  ....................  ....................
9300 MW173412.1 |Severe acute respiratory syndrome coro    ....................  ....................  ....................  ....................  ....................  ....................
9301 MW173414.1 |Severe acute respiratory syndrome coro    ....................  ....................  ....................  ....................  ....................  ....................
9302 MW173417.1 |Severe acute respiratory syndrome coro    ....................  ....................  ....................  ....................  ....................  ....................
9303 MW173419.1 |Severe acute respiratory syndrome coro    ....................  .A..................  ....................  ....................  ....................  ....................
9304 MW173423.1 |Severe acute respiratory syndrome coro    ....................  .A..................  ....................  ....................  ....................  ....................
9305 MW173426.1 |Severe acute respiratory syndrome coro    ....................  .A..................  ....................  ....................  ....................  ....................
9306 MW173427.1 |Severe acute respiratory syndrome coro    ....................  ....................  ....................  ....................  ....................  ....................
9307 MW173433.1 |Severe acute respiratory syndrome coro    ....................  ....................  ....................  ....................  ....................  ....................
9308 MW173436.1 |Severe acute respiratory syndrome coro    ....................  ....................  ....................  ....................  ....................  ....................
9309 MW173440.1 |Severe acute respiratory syndrome coro    ....................  ....................  ....................  ....................  ....................  ....................
9310 MW173441.1 |Severe acute respiratory syndrome coro    ....................  ....................  ....................  ....................  ....................  ....................
9311 MW173442.1 |Severe acute respiratory syndrome coro    ....................  ....................  ....................  ....................  ....................  ....................
9312 MW173443.1 |Severe acute respiratory syndrome coro    ....................  ....................  ....................  ....................  ....................  ....................
9313 MW173444.1 |Severe acute respiratory syndrome coro    ....................  ....................  ....................  ....................  ....................  ....................
9315 MW173448.1 |Severe acute respiratory syndrome coro    ....................  ....................  ....................  ....................  ....................  ....................
9316 MW173452.1 |Severe acute respiratory syndrome coro    ....................  ....................  ....................  ....................  ....................  ....................
9317 MW173453.1 |Severe acute respiratory syndrome coro    ....................  ....................  ....................  ....................  ....................  ....................
9318 MW173454.1 |Severe acute respiratory syndrome coro    ....................  ....................  ....................  ....................  ....................  ....................
9319 MW173459.1 |Severe acute respiratory syndrome coro    ....................  ....................  ....................  ....................  ....................  ....................
9320 MW173460.1 |Severe acute respiratory syndrome coro    ....................  .A..................  ....................  ....................  ....................  ..A.................
9321 MW173463.1 |Severe acute respiratory syndrome coro    ....................  .A..................  ....................  ....................  ....................  ..A.................
9322 MW180935.1 |Severe acute respiratory syndrome coro    ....................  ....................  ....................  ....................  ....................  ....................
9323 MW180936.1 |Severe acute respiratory syndrome coro    ....................  ....................  ....................  ....................  ....................  ....................
9324 MW165854.1 |Severe acute respiratory syndrome coro    ....................  .A..................  ....................  ....................  ....................  ..A.................
9325 MW165857.1 |Severe acute respiratory syndrome coro    ....................  .A..................  ....................  ....................  ....................  ..A.................
9326 MW165861.1 |Severe acute respiratory syndrome coro    ....................  .A..................  ....................  ....................  ....................  ..A.................
9327 MW165862.1 |Severe acute respiratory syndrome coro    ....................  .A..................  ....................  ....................  ....................  ....................
9328 MW165865.1 |Severe acute respiratory syndrome coro    ....................  .A..................  ....................  ....................  ....................  ..A.................
9329 MW165869.1 |Severe acute respiratory syndrome coro    ....................  ....................  ....................  ....................  ....................  ....................
9330 MW165872.1 |Severe acute respiratory syndrome coro    ....................  ....................  ....................  ....................  ....................  ....................
9331 MW165873.1 |Severe acute respiratory syndrome coro    ....................  .A..................  ....................  ....................  ....................  ..A.................
9332 MW165876.1 |Severe acute respiratory syndrome coro    ....................  .A..................  ....................  ....................  ....................  ....................
9333 MW165885.1 |Severe acute respiratory syndrome coro    ....................  .A..................  ....................  ....................  ....................  ..A.................
9334 MW166053.1 |Severe acute respiratory syndrome coro    ....................  ....................  ....................  ....................  ....................  ....................
9335 MW166054.1 |Severe acute respiratory syndrome coro    ....................  ....................  ....................  ....................  ....................  ....................
9336 MW166055.1 |Severe acute respiratory syndrome coro    ....................  ....................  ....................  ....................  ....................  ....................
9337 MW166056.1 |Severe acute respiratory syndrome coro    ....................  ....................  ....................  ....................  ....................  ....................
9338 MW166064.1 |Severe acute respiratory syndrome coro    ....................  ....................  ....................  ....................  ....................  ....................
9339 MW166065.1 |Severe acute respiratory syndrome coro    ....................  ....................  ....................  ....................  ....................  ....................
9340 MW166066.1 |Severe acute respiratory syndrome coro    ....................  ....................  ....................  ....................  ....................  ....................
9342 MW166069.1 |Severe acute respiratory syndrome coro    ....................  ....................  ....................  ....................  ....................  ....................
9343 MW166070.1 |Severe acute respiratory syndrome coro    ....................  ....................  ....................  ....................  ....................  ....................
9345 MW166072.1 |Severe acute respiratory syndrome coro    ....................  ....................  ....................  ....................  ....................  ....................
9346 MW166073.1 |Severe acute respiratory syndrome coro    ....................  ....................  ....................  ....................  ....................  ....................
9347 MW166075.1 |Severe acute respiratory syndrome coro    ....................  ....................  ....................  ....................  ....................  ....................
9348 MW166077.1 |Severe acute respiratory syndrome coro    ....................  ....................  ....................  ....................  ....................  ....................
9349 MW166078.1 |Severe acute respiratory syndrome coro    ....................  ....................  ....................  ....................  ....................  ....................
9350 MW166079.1 |Severe acute respiratory syndrome coro    ....................  ....................  ....................  ....................  ....................  ....................
9351 MW166080.1 |Severe acute respiratory syndrome coro    ....................  ....................  ....................  ....................  ....................  ....................
9352 MW166081.1 |Severe acute respiratory syndrome coro    ....................  ....................  ....................  ....................  ....................  ....................
9353 MW166082.1 |Severe acute respiratory syndrome coro    ....................  ....................  ....................  ....................  ....................  ....................
9354 MW166083.1 |Severe acute respiratory syndrome coro    ....................  ....................  ....................  ....................  ....................  ....................
9355 MW166084.1 |Severe acute respiratory syndrome coro    ....................  ....................  ....................  ....................  ....................  ....................
9356 MW166086.1 |Severe acute respiratory syndrome coro    ....................  ....................  ....................  ....................  ....................  ....................
9357 MW166087.1 |Severe acute respiratory syndrome coro    ....................  ....................  ....................  ....................  ....................  ....................
9358 MW166088.1 |Severe acute respiratory syndrome coro    ....................  ....................  ....................  ....................  ....................  ....................
9359 MW166092.1 |Severe acute respiratory syndrome coro    ....................  ....................  ....................  ....................  ....................  ....................
9361 MW166096.1 |Severe acute respiratory syndrome coro    ....................  ....................  ....................  ....................  ....................  ....................
9362 MW166097.1 |Severe acute respiratory syndrome coro    ....................  ....................  ....................  ....................  ....................  ....................
9363 MW166098.1 |Severe acute respiratory syndrome coro    ....................  ....................  ....................  ....................  ....................  ....................
9364 MW166108.1 |Severe acute respiratory syndrome coro    ....................  ....................  ....................  ....................  ....................  ....................
9365 MW166110.1 |Severe acute respiratory syndrome coro    ....................  ....................  ....................  ....................  ....................  ....................
9366 MW166113.1 |Severe acute respiratory syndrome coro    ....................  ....................  ....................  ....................  ....................  ....................
```

9367 MW166116.1 |Severe acute respiratory syndrome coro ···················· ···················· ···················· ···················· ···················· ····················
9368 MW166117.1 |Severe acute respiratory syndrome coro ···················· ···················· ···················· ···················· ···················· ····················
9369 MW166118.1 |Severe acute respiratory syndrome coro ···················· ···················· ···················· ···················· ···················· ····················
9370 MW166119.1 |Severe acute respiratory syndrome coro ···················· ···················· ···················· ···················· ···················· ····················
9371 MW166120.1 |Severe acute respiratory syndrome coro ··············G····· ···················· ···················· ···················· ···················· ····················
9372 MW166122.1 |Severe acute respiratory syndrome coro ···················· ···················· ···················· ···················· ···················· ····················
9373 MW166123.1 |Severe acute respiratory syndrome coro ···················· ·········A·········· ···················· ···················· ···················· ····················
9374 MW166125.1 |Severe acute respiratory syndrome coro ···················· ···················· ···················· ···················· ···················· ····················
9375 MW166126.1 |Severe acute respiratory syndrome coro ···················· ···················· ···················· ···················· ···················· ····················
9376 MW166127.1 |Severe acute respiratory syndrome coro ···················· ···················· ···················· ···················· ···················· ····················
9377 MW166129.1 |Severe acute respiratory syndrome coro ···················· ···················· ···················· ···················· ···················· ····················
9378 MW166130.1 |Severe acute respiratory syndrome coro ···················· ···················· ···················· ···················· ···················· ····················
9379 MW166131.1 |Severe acute respiratory syndrome coro ···················· ···················· ···················· ···················· ···················· ····················
9380 MW166133.1 |Severe acute respiratory syndrome coro ···················· ···················· ···················· ···················· ···················· ····················
9381 MW166136.1 |Severe acute respiratory syndrome coro ···················· ···················· ···················· ···················· ···················· ····················
9382 MW166138.1 |Severe acute respiratory syndrome coro ···················· ···················· ···················· ···················· ···················· ····················
9383 MW166139.1 |Severe acute respiratory syndrome coro ···················· ···················· ···················· ···················· ···················· ····················
9384 MW166141.1 |Severe acute respiratory syndrome coro ···················· ···················· ···················· ···················· ···················· ····················
9385 MW166142.1 |Severe acute respiratory syndrome coro ···················· ···················· ···················· ···················· ···················· ····················
9386 MW166144.1 |Severe acute respiratory syndrome coro ···················· ···················· ···················· ···················· ···················· ····················
9387 MW166145.1 |Severe acute respiratory syndrome coro ···················· ···················· ···················· ···················· ···················· ····················
9388 MW166149.1 |Severe acute respiratory syndrome coro ···················· ···················· ···················· ···················· ···················· ····················
9389 MW166150.1 |Severe acute respiratory syndrome coro ···················· ···················· ···················· ···················· ···················· ····················
9390 MW166151.1 |Severe acute respiratory syndrome coro ···················· ···················· ···················· ···················· ···················· ····················
9391 MW166152.1 |Severe acute respiratory syndrome coro ···················· ···················· ···················· ···················· ···················· ····················
9392 MW166154.1 |Severe acute respiratory syndrome coro ···················· ···················· ···················· ···················· ···················· ····················
9393 MW166165.1 |Severe acute respiratory syndrome coro ···················· ···················· ···················· ···················· ···················· ····················
9394 MW166167.1 |Severe acute respiratory syndrome coro ···················· ···················· ···················· ···················· ···················· ····················
9395 MW166214.1 |Severe acute respiratory syndrome coro ···················· ·A·················· ···················· ···················· ···················· ···A················
9396 MW166221.1 |Severe acute respiratory syndrome coro ···················· ·A·················· ···················· ···················· ···················· ···A················
9397 MW166349.1 |Severe acute respiratory syndrome coro ···················· ·A·················· ···················· ···················· ···················· ···A················
9398 MW166864.1 |Severe acute respiratory syndrome coro ···················· ···················· ···················· ···················· ···················· ····················
9399 MW167078.1 |Severe acute respiratory syndrome coro ···················· ·A·················· ···················· ···················· ···················· ···A················
9400 MW167112.1 |Severe acute respiratory syndrome coro ···················· ·A·················· ···················· ···················· ···················· ···A················
9401 MW172414.1 |Severe acute respiratory syndrome coro ···················· ···················· ···················· ···················· ···················· ····················
9402 MW172415.1 |Severe acute respiratory syndrome coro ···················· ···················· ···················· ···················· ···················· ····················
9403 MW172507.1 |Severe acute respiratory syndrome coro ···················· ···················· ···················· ···················· ···················· ····················
9404 MW172508.1 |Severe acute respiratory syndrome coro ···················· ···················· ···················· ···················· ···················· ····················
9405 MW172509.1 |Severe acute respiratory syndrome coro ···················· ···················· ···················· ···················· ···················· ····················
9406 MW172510.1 |Severe acute respiratory syndrome coro ···················· ···················· ···················· ···················· ···················· ····················
9407 MW172513.1 |Severe acute respiratory syndrome coro ···················· ···················· ···················· ···················· ···················· ····················
9408 MW172514.1 |Severe acute respiratory syndrome coro ···················· ···················· ···················· ···················· ···················· ····················
9409 MW172515.1 |Severe acute respiratory syndrome coro ···················· ···················· ···················· ···················· ···················· ····················
9410 MW172517.1 |Severe acute respiratory syndrome coro ···················· ···················· ···················· ···················· ···················· ····················
9411 MW172518.1 |Severe acute respiratory syndrome coro ···················· ···················· ···················· ···················· ···················· ····················
9412 MW172520.1 |Severe acute respiratory syndrome coro ···················· ···················· ···················· ···················· ···················· ····················
9413 MW172558.1 |Severe acute respiratory syndrome coro ···················· ···················· ···················· ···················· ···················· ····················
9414 MW172563.1 |Severe acute respiratory syndrome coro ···················· ···················· ···················· ···················· ···················· ····················
9415 MW172570.1 |Severe acute respiratory syndrome coro ···················· ···················· ···················· ···················· ···················· ····················
9416 MW172571.1 |Severe acute respiratory syndrome coro ···················· ···················· ···················· ···················· ···················· ····················
9417 MW172572.1 |Severe acute respiratory syndrome coro ···················· ···················· ···················· ···················· ···················· ····················
9418 MW172574.1 |Severe acute respiratory syndrome coro ···················· ···················· ···················· ···················· ···················· ····················
9419 MW172576.1 |Severe acute respiratory syndrome coro ···················· ···················· ···················· ···················· ···················· ····················
9420 MW172579.1 |Severe acute respiratory syndrome coro ···················· ···················· ···················· ···················· ···················· ····················
9421 MW172581.1 |Severe acute respiratory syndrome coro ···················· ···················· ···················· ···················· ···················· ····················
9422 MW172721.1 |Severe acute respiratory syndrome coro ···················· ···················· ···················· ···················· ···················· ····················
9423 MW172727.1 |Severe acute respiratory syndrome coro ···················· ···················· ···················· ···················· ···················· ····················
9424 MW172730.1 |Severe acute respiratory syndrome coro ···················· ···················· ···················· ···················· ···················· ····················
9425 MW172733.1 |Severe acute respiratory syndrome coro ···················· ···················· ···················· ···················· ···················· ····················
9426 MW172737.1 |Severe acute respiratory syndrome coro ···················· ···················· ···················· ···················· ···················· ····················
9427 MW172742.1 |Severe acute respiratory syndrome coro ···················· ···················· ···················· ···················· ···················· ····················
9428 MW172756.1 |Severe acute respiratory syndrome coro ···················· ···················· ···················· ···················· ···················· ····················
9429 MW172762.1 |Severe acute respiratory syndrome coro ···················· ···················· ···················· ···················· ···················· ····················
9430 MW172766.1 |Severe acute respiratory syndrome coro ···················· ···················· ···················· ···················· ···················· ····················
9431 MW172768.1 |Severe acute respiratory syndrome coro ···················· ···················· ···················· ···················· ···················· ····················
9432 MW172771.1 |Severe acute respiratory syndrome coro ···················· ···················· ···················· ···················· ···················· ····················
9433 MW172813.1 |Severe acute respiratory syndrome coro ···················· ···················· ···················· ···················· ···················· ····················
9434 MW173146.1 |Severe acute respiratory syndrome coro ···················· ···················· ···················· ···················· ···················· ····················
9435 MW173147.1 |Severe acute respiratory syndrome coro ···················· ···················· ···················· ···················· ···················· ····················
9436 MW173148.1 |Severe acute respiratory syndrome coro ···················· ·A·················· ···················· ···················· ···················· ····················
9437 MW173149.1 |Severe acute respiratory syndrome coro ···················· ···················· ···················· ···················· ···················· ····················
9438 MW173150.1 |Severe acute respiratory syndrome coro ···················· ···················· ···················· ···················· ···················· ····················
9439 MW173151.1 |Severe acute respiratory syndrome coro ···················· ···················· ···················· ···················· ···················· ····················
9440 MW173152.1 |Severe acute respiratory syndrome coro ···················· ···················· ···················· ···················· ···················· ····················
9441 MW173153.1 |Severe acute respiratory syndrome coro ···················· ···················· ···················· ···················· ···················· ····················
9442 MW173154.1 |Severe acute respiratory syndrome coro ···················· ···················· ···················· ···················· ···················· ····················
9443 MW173155.1 |Severe acute respiratory syndrome coro ···················· ···················· ···················· ···················· ···················· ····················
9444 MW173156.1 |Severe acute respiratory syndrome coro ···················· ···················· ···················· ···················· ···················· ····················

9445 MW173157.1 |Severe acute respiratory syndrome coro .................... .................... ..................... .................... .................... ....................
9446 MW173158.1 |Severe acute respiratory syndrome coro .................... .................... ..................... .................... .................... ....................
9447 MW173159.1 |Severe acute respiratory syndrome coro .................... .................... ..................... .................... .................... ....................
9448 MW173160.1 |Severe acute respiratory syndrome coro .................... .................... ..................... .................... .................... ....................
9449 MW173161.1 |Severe acute respiratory syndrome coro .................... .................... ..................... .................... .................... ....................
9450 MW173162.1 |Severe acute respiratory syndrome coro .................... .................... ..................... .................... .................... ....................
9451 MW173163.1 |Severe acute respiratory syndrome coro .................... .................... ..................... .................... .................... ....................
9452 MW173164.1 |Severe acute respiratory syndrome coro .................... .A.................. ..................... .................... .................... ....................
9453 MW173165.1 |Severe acute respiratory syndrome coro .................... .................... ..................... .................... .................... ....................
9454 MW173166.1 |Severe acute respiratory syndrome coro .................... .................... ..................... .................... .................... ....................
9455 MW173167.1 |Severe acute respiratory syndrome coro .................... .................... ..................... .................... .................... ....................
9456 MW173168.1 |Severe acute respiratory syndrome coro .................... .................... ..................... .................... .................... ....................
9457 MW173169.1 |Severe acute respiratory syndrome coro .................... .................... ..................... .................... .................... ....................
9458 MW173170.1 |Severe acute respiratory syndrome coro .................... .................... ..................... .................... .................... ....................
9459 MW173171.1 |Severe acute respiratory syndrome coro .................... .................... ..................... .................... .................... ....................
9460 MW173212.1 |Severe acute respiratory syndrome coro .................... .................... ..................... .................... .................... ....................
9461 MW173213.1 |Severe acute respiratory syndrome coro .................... .................... ..................... .................... .................... ....................
9462 MW161259.1 |Severe acute respiratory syndrome coro .................... .................... ..................... .................... .................... ....................
9463 MW153038.1 |Severe acute respiratory syndrome coro .................... .................... ..................... .................... .................... ....................
9464 MW153039.1 |Severe acute respiratory syndrome coro .................... .................... ..................... .................... .................... ....................
9467 MW153047.1 |Severe acute respiratory syndrome coro .................... .................... ..................... .................... .................... ....................
9469 MW153051.1 |Severe acute respiratory syndrome coro .................... .................... ..................... .................... .................... ....................
9473 MW153063.1 |Severe acute respiratory syndrome coro .................... .................... ..................... .................... .................... ....................
9474 MW153072.1 |Severe acute respiratory syndrome coro .................... .................... ..................... .................... .................... ....................
9475 MW153074.1 |Severe acute respiratory syndrome coro .................... .................... ..................... .................... .................... ....................
9476 MW153075.1 |Severe acute respiratory syndrome coro .................... .................... ..................... .................... .................... ....................
9478 MW153078.1 |Severe acute respiratory syndrome coro .................... .................... ..................... .................... .................... ....................
9479 MW153079.1 |Severe acute respiratory syndrome coro .................... .................... ..................... .................... .................... ....................
9481 MW153081.1 |Severe acute respiratory syndrome coro .................... .................... ..................... .................... .................... ....................
9482 MW153082.1 |Severe acute respiratory syndrome coro .................... .................... ..................... .................... .................... ....................
9483 MW153083.1 |Severe acute respiratory syndrome coro .................... .................... ..................... .................... .................... ....................
9484 MW153084.1 |Severe acute respiratory syndrome coro .................... .................... ..................... .................... .................... ....................
9485 MW153085.1 |Severe acute respiratory syndrome coro .................... .................... ..................... .................... .................... ....................
9486 MW153086.1 |Severe acute respiratory syndrome coro .................... .................... ..................... .................... .................... ....................
9487 MW153087.1 |Severe acute respiratory syndrome coro .................... .................... ..................... .................... .................... ....................
9488 MW153088.1 |Severe acute respiratory syndrome coro .................... .................... ..................... .................... .................... ....................
9489 MW153089.1 |Severe acute respiratory syndrome coro .................... .................... ..................... .................... .................... ....................
9490 MW153090.1 |Severe acute respiratory syndrome coro .................... .................... ..................... .................... .................... ....................
9491 MW153091.1 |Severe acute respiratory syndrome coro .................... .................... ..................... .................... .................... ....................
9492 MW153094.1 |Severe acute respiratory syndrome coro .................... .................... ..................... .................... .................... ....................
9493 MW153095.1 |Severe acute respiratory syndrome coro .................... .................... ..................... .................... .................... ....................
9494 MW153096.1 |Severe acute respiratory syndrome coro .................... .................... ..................... .................... .................... ....................
9495 MW153097.1 |Severe acute respiratory syndrome coro .................... .................... ..................... .................... .................... ....................
9496 MW153098.1 |Severe acute respiratory syndrome coro .................... .................... ..................... .................... .................... ....................
9497 MW153099.1 |Severe acute respiratory syndrome coro .................... .................... ..................... .................... .................... ....................
9498 MW153100.1 |Severe acute respiratory syndrome coro .................... .................... ..................... .................... .................... ....................
9499 MW153101.1 |Severe acute respiratory syndrome coro .................... .................... ..................... .................... .................... ....................
9500 MW153102.1 |Severe acute respiratory syndrome coro .................... .................... ..................... .................... .................... ....................
9501 MW153103.1 |Severe acute respiratory syndrome coro .................... .................... ..................... .................... .................... ....................
9502 MW153104.1 |Severe acute respiratory syndrome coro .................... .................... ..................... .................... .................... ....................
9503 MW153105.1 |Severe acute respiratory syndrome coro .................... .................... ..................... .................... .................... ....................
9504 MW153106.1 |Severe acute respiratory syndrome coro .................... .................... ..................... .................... .................... ....................
9505 MW153107.1 |Severe acute respiratory syndrome coro .................... .................... ..................... .................... .................... ....................
9506 MW153108.1 |Severe acute respiratory syndrome coro .................... .................... ..................... .................... .................... ....................
9507 MW153109.1 |Severe acute respiratory syndrome coro .................... .................... ..................... .................... .................... ....................
9508 MW153110.1 |Severe acute respiratory syndrome coro .................... .................... ..................... .................... .................... ....................
9509 MW153111.1 |Severe acute respiratory syndrome coro .................... .................... ..................... .................... .................... ....................
9510 MW153112.1 |Severe acute respiratory syndrome coro .................... .................... ..................... .................... .................... ....................
9511 MW153113.1 |Severe acute respiratory syndrome coro .................... .................... ..................... .................... .................... ....................
9512 MW153114.1 |Severe acute respiratory syndrome coro .................... .................... ..................... .................... .................... ....................
9513 MW153115.1 |Severe acute respiratory syndrome coro .................... .................... ..................... .................... .................... ....................
9514 MW153116.1 |Severe acute respiratory syndrome coro .................... .................... ..................... .................... .................... ....................
9515 MW153117.1 |Severe acute respiratory syndrome coro .................... .................... ..................... .................... .................... ....................
9516 MW153118.1 |Severe acute respiratory syndrome coro .................... .................... ..................... .................... .................... ....................
9517 MW153119.1 |Severe acute respiratory syndrome coro .................... .................... ..................... .................... .................... ....................
9518 MW153120.1 |Severe acute respiratory syndrome coro .................... .................... ..................... .................... .................... ....................
9519 MW153121.1 |Severe acute respiratory syndrome coro .................... .................... ..................... .................... .................... ....................
9520 MW153122.1 |Severe acute respiratory syndrome coro .................... .................... ..................... .................... .................... ....................
9521 MW153123.1 |Severe acute respiratory syndrome coro .................... .................... ..................... .................... .................... ....................
9522 MW153126.1 |Severe acute respiratory syndrome coro .................... .................... ..................... .................... .................... ....................
9523 MW153127.1 |Severe acute respiratory syndrome coro .................... .................... ..................... .................... .................... ....................
9524 MW153128.1 |Severe acute respiratory syndrome coro .................... .................... ..................... .................... .................... ....................
9525 MW153129.1 |Severe acute respiratory syndrome coro .................... .................... ..................... .................... .................... ....................
9526 MW153130.1 |Severe acute respiratory syndrome coro .................... .................... ..................... .................... .................... ....................
9527 MW153131.1 |Severe acute respiratory syndrome coro .................... .................... ..................... .................... .................... ....................
9528 MW153132.1 |Severe acute respiratory syndrome coro .................... .................... ..................... .................... .................... ....................
9529 MW153133.1 |Severe acute respiratory syndrome coro .................... .................... ..................... .................... .................... ....................
9530 MW153134.1 |Severe acute respiratory syndrome coro .................... .................... ..................... .................... .................... ....................

```
9531 MW153135.1 |Severe acute respiratory syndrome coro    ....................  ....................  ....................  ....................  ....................  ....................
9532 MW153136.1 |Severe acute respiratory syndrome coro    ....................  ....................  ....................  ....................  ....................  ....................
9533 MW153137.1 |Severe acute respiratory syndrome coro    ....................  ....................  ....................  ....................  ....................  ....................
9534 MW153138.1 |Severe acute respiratory syndrome coro    ....................  ......A.............  ....................  ....................  ....................  ....................
9535 MW153139.1 |Severe acute respiratory syndrome coro    ....................  ....................  ....................  ....................  ....................  ....................
9536 MW153140.1 |Severe acute respiratory syndrome coro    ....................  ....................  ....................  ....................  ....................  ....................
9537 MW153142.1 |Severe acute respiratory syndrome coro    ....................  ....................  ....................  ....................  ....................  ....................
9538 MW153143.1 |Severe acute respiratory syndrome coro    ....................  ....................  ....................  ....................  ....................  ....................
9539 MW153144.1 |Severe acute respiratory syndrome coro    ....................  ....................  ....................  ....................  ....................  ....................
9540 MW153145.1 |Severe acute respiratory syndrome coro    ....................  ....................  ....................  ....................  ....................  ....................
9541 MW153146.1 |Severe acute respiratory syndrome coro    ....................  ....................  ....................  ....................  ....................  ....................
9542 MW153148.1 |Severe acute respiratory syndrome coro    ....................  ....................  ....................  ....................  ....................  ....................
9543 MW153149.1 |Severe acute respiratory syndrome coro    ....................  ....................  ....................  ....................  ....................  ....................
9544 MW153151.1 |Severe acute respiratory syndrome coro    ....................  ....................  ....................  ....................  ....................  ....................
9545 MW153152.1 |Severe acute respiratory syndrome coro    ....................  ....................  ....................  ....................  ....................  ....................
9546 MW153153.1 |Severe acute respiratory syndrome coro    ....................  ....................  ....................  ....................  ....................  ....................
9547 MW153154.1 |Severe acute respiratory syndrome coro    ....................  ....................  ....................  ....................  ....................  ....................
9548 MW153155.1 |Severe acute respiratory syndrome coro    ....................  ....................  ....................  ....................  ....................  ....................
9549 MW153156.1 |Severe acute respiratory syndrome coro    ....................  ....................  ....................  ....................  ....................  ....................
9550 MW153157.1 |Severe acute respiratory syndrome coro    ....................  ....................  ....................  ....................  ....................  ....................
9551 MW153158.1 |Severe acute respiratory syndrome coro    ....................  ....................  ....................  ....................  ....................  ....................
9552 MW153159.1 |Severe acute respiratory syndrome coro    ....................  ....................  ....................  ....................  ....................  ....................
9553 MW153160.1 |Severe acute respiratory syndrome coro    ....................  ....................  ....................  ....................  ....................  ....................
9554 MW153162.1 |Severe acute respiratory syndrome coro    ....................  ....................  ....................  ....................  ....................  ....................
9555 MW153163.1 |Severe acute respiratory syndrome coro    ....................  ....................  ....................  ....................  ....................  ....................
9556 MW153164.1 |Severe acute respiratory syndrome coro    ....................  ....................  ....................  ....................  ....................  ....................
9557 MW153165.1 |Severe acute respiratory syndrome coro    ....................  ....................  ....................  ....................  ....................  ....................
9558 MW153166.1 |Severe acute respiratory syndrome coro    ....................  ....................  ....................  ....................  ....................  ....................
9559 MW153167.1 |Severe acute respiratory syndrome coro    ....................  ....................  ....................  ....................  ....................  ....................
9560 MW153168.1 |Severe acute respiratory syndrome coro    ....................  ....................  ....................  ....................  ....................  ....................
9561 MW153169.1 |Severe acute respiratory syndrome coro    ....................  ....................  ....................  ....................  ....................  ....................
9562 MW153170.1 |Severe acute respiratory syndrome coro    ....................  ....................  ....................  ....................  ....................  ....................
9563 MW153172.1 |Severe acute respiratory syndrome coro    ....................  ....................  ....................  ....................  ....................  ....................
9564 MW153173.1 |Severe acute respiratory syndrome coro    ....................  ....................  ....................  ....................  ....................  ....................
9565 MW153175.1 |Severe acute respiratory syndrome coro    ....................  ....................  ....................  ....................  ....................  ....................
9566 MW153176.1 |Severe acute respiratory syndrome coro    ....................  ....................  ....................  ....................  ....................  ....................
9567 MW153177.1 |Severe acute respiratory syndrome coro    ....................  ....................  ....................  ....................  ....................  ....................
9568 MW153178.1 |Severe acute respiratory syndrome coro    ....................  ....................  ....................  ....................  ....................  ....................
9569 MW153181.1 |Severe acute respiratory syndrome coro    ....................  ....................  ....................  ....................  ....................  ....................
9570 MW153182.1 |Severe acute respiratory syndrome coro    ....................  ....................  ....................  ....................  ....................  ....................
9571 MW153183.1 |Severe acute respiratory syndrome coro    ....................  ....................  ....................  ....................  ....................  ....................
9572 MW153184.1 |Severe acute respiratory syndrome coro    ....................  ....................  ....................  ....................  ....................  ....................
9573 MW153185.1 |Severe acute respiratory syndrome coro    ....................  ....................  ....................  ....................  ....................  ....................
9574 MW153187.1 |Severe acute respiratory syndrome coro    ....................  ....................  ....................  ....................  ....................  ....................
9575 MW153188.1 |Severe acute respiratory syndrome coro    ....................  ....................  ....................  ....................  ....................  ....................
9576 MW153189.1 |Severe acute respiratory syndrome coro    ....................  ....................  ....................  ....................  ....................  ....................
9577 MW153190.1 |Severe acute respiratory syndrome coro    ....................  ....................  ....................  ....................  ....................  ....................
9578 MW153191.1 |Severe acute respiratory syndrome coro    ....................  ....................  ....................  ....................  ....................  ....................
9579 MW153192.1 |Severe acute respiratory syndrome coro    ....................  ....................  ....................  ....................  ....................  ....................
9580 MW153193.1 |Severe acute respiratory syndrome coro    ....................  ....................  ....................  ....................  ....................  ....................
9581 MW153194.1 |Severe acute respiratory syndrome coro    ....................  ....................  ....................  ....................  ....................  ....................
9582 MW153195.1 |Severe acute respiratory syndrome coro    ....................  ....................  ....................  ....................  ....................  ....................
9583 MW153196.1 |Severe acute respiratory syndrome coro    ....................  ....................  ....................  ....................  ....................  ....................
9584 MW153197.1 |Severe acute respiratory syndrome coro    ....................  ....................  ....................  ....................  ....................  ....................
9585 MW153198.1 |Severe acute respiratory syndrome coro    ....................  ....................  ....................  ....................  ....................  ....................
9586 MW153199.1 |Severe acute respiratory syndrome coro    ....................  ....................  ....................  ....................  ....................  ....................
9587 MW153200.1 |Severe acute respiratory syndrome coro    ....................  ....................  ....................  ....................  ....................  ....................
9588 MW153202.1 |Severe acute respiratory syndrome coro    ....................  ....................  ....................  ....................  ....................  ....................
9589 MW153203.1 |Severe acute respiratory syndrome coro    ....................  ....................  ....................  ....................  ....................  ....................
9590 MW153204.1 |Severe acute respiratory syndrome coro    ....................  ....................  ....................  ....................  ....................  ....................
9591 MW153205.1 |Severe acute respiratory syndrome coro    ....................  ....................  ....................  ....................  ....................  ....................
9592 MW153206.1 |Severe acute respiratory syndrome coro    ....................  ....................  ....................  ....................  ....................  ....................
9593 MW153208.1 |Severe acute respiratory syndrome coro    ....................  ....................  ....................  ....................  ....................  ....................
9594 MW153209.1 |Severe acute respiratory syndrome coro    ....................  ....................  ....................  ....................  ....................  ....................
9595 MW153210.1 |Severe acute respiratory syndrome coro    ....................  ....................  ....................  ....................  ....................  ....................
9596 MW153211.1 |Severe acute respiratory syndrome coro    ....................  ....................  ....................  ....................  ....................  ....................
9597 MW153212.1 |Severe acute respiratory syndrome coro    ....................  ....................  ....................  ....................  ....................  ....................
9598 MW153213.1 |Severe acute respiratory syndrome coro    ....................  ....................  ....................  ....................  ....................  ....................
9599 MW153214.1 |Severe acute respiratory syndrome coro    ....................  ....................  ....................  ....................  ....................  ....................
9600 MW153215.1 |Severe acute respiratory syndrome coro    ....................  ....................  ....................  ....................  ....................  ....................
9601 MW153216.1 |Severe acute respiratory syndrome coro    ....................  ......A.............  ....................  ....................  ....................  ....................
9602 MW153217.1 |Severe acute respiratory syndrome coro    ....................  ....................  ....................  ....................  ....................  ....................
9603 MW153219.1 |Severe acute respiratory syndrome coro    ....................  ....................  ....................  ....................  ....................  ....................
9604 MW153220.1 |Severe acute respiratory syndrome coro    ....................  ....................  ....................  ....................  ....................  ....................
9605 MW153223.1 |Severe acute respiratory syndrome coro    ....................  ....................  ....................  ....................  ....................  ....................
9606 MW153226.1 |Severe acute respiratory syndrome coro    ....................  ....................  ....................  ....................  ....................  ....................
9607 MW153227.1 |Severe acute respiratory syndrome coro    ....................  ....................  ....................  ....................  ....................  ....................
9608 MW153228.1 |Severe acute respiratory syndrome coro    ....................  ....................  ....................  ....................  ....................  ....................
```

```
9609 MW153229.1 |Severe acute respiratory syndrome coro    ....................  ....................  .....................  ....................  ....................  ....................
9610 MW153230.1 |Severe acute respiratory syndrome coro    ....................  ....................  .....................  ....................  ....................  ....................
9611 MW153231.1 |Severe acute respiratory syndrome coro    ....................  ....................  .....................  ....................  ....................  ....................
9612 MW153232.1 |Severe acute respiratory syndrome coro    ....................  ....................  .....................  ....................  ....................  ....................
9613 MW153233.1 |Severe acute respiratory syndrome coro    ....................  ....................  .....................  ....................  ....................  ....................
9614 MW153234.1 |Severe acute respiratory syndrome coro    ....................  ....................  .....................  ....................  ....................  ....................
9615 MW153235.1 |Severe acute respiratory syndrome coro    ....................  ....................  .....................  ....................  ....................  ....................
9616 MW153236.1 |Severe acute respiratory syndrome coro    ....................  ....................  .....................  ....................  ....................  ....................
9617 MW153237.1 |Severe acute respiratory syndrome coro    ....................  ....................  .....................  ....................  ....................  ....................
9618 MW153238.1 |Severe acute respiratory syndrome coro    ....................  ....................  .....................  ....................  ....................  ....................
9619 MW153239.1 |Severe acute respiratory syndrome coro    ....................  ....................  .....................  ....................  ....................  ....................
9620 MW153240.1 |Severe acute respiratory syndrome coro    ....................  ....................  .....................  ....................  ....................  ....................
9621 MW153241.1 |Severe acute respiratory syndrome coro    ....................  ....................  .....................  ....................  ....................  ....................
9622 MW153242.1 |Severe acute respiratory syndrome coro    ....................  ....................  .....................  ....................  ....................  ....................
9623 MW153243.1 |Severe acute respiratory syndrome coro    ....................  ....................  .....................  ....................  ....................  ....................
9624 MW153244.1 |Severe acute respiratory syndrome coro    ....................  ....................  .....................  ....................  ....................  ....................
9625 MW153245.1 |Severe acute respiratory syndrome coro    ....................  ....................  .....................  ....................  ....................  ....................
9626 MW153246.1 |Severe acute respiratory syndrome coro    ....................  ....................  .....................  ....................  ....................  ....................
9627 MW153247.1 |Severe acute respiratory syndrome coro    ....................  ....................  .....................  ....................  ....................  ....................
9628 MW153249.1 |Severe acute respiratory syndrome coro    ....................  ....................  .....................  ....................  ....................  ....................
9629 MW153250.1 |Severe acute respiratory syndrome coro    ....................  ....................  .....................  ....................  ....................  ....................
9630 MW153251.1 |Severe acute respiratory syndrome coro    ....................  ....................  .....................  ....................  ....................  ....................
9631 MW153252.1 |Severe acute respiratory syndrome coro    ....................  ....................  .....................  ....................  ....................  ....................
9632 MW153253.1 |Severe acute respiratory syndrome coro    ....................  ....................  .....................  ....................  ....................  ....................
9633 MW153255.1 |Severe acute respiratory syndrome coro    ....................  ....................  .....................  ....................  ....................  ....................
9634 MW153258.1 |Severe acute respiratory syndrome coro    ....................  ....................  .....................  ....................  ....................  ....................
9635 MW153259.1 |Severe acute respiratory syndrome coro    ....................  ....................  .....................  ....................  ....................  ....................
9636 MW153260.1 |Severe acute respiratory syndrome coro    ....................  ....................  .....................  ....................  ....................  ....................
9637 MW153261.1 |Severe acute respiratory syndrome coro    ....................  ....................  .....................  ....................  ....................  ....................
9638 MW153262.1 |Severe acute respiratory syndrome coro    ....................  ....................  .....................  ....................  ....................  ....................
9639 MW153263.1 |Severe acute respiratory syndrome coro    ....................  ....................  .....................  ....................  ....................  ....................
9640 MW153264.1 |Severe acute respiratory syndrome coro    ....................  ....................  .....................  ....................  ....................  ....................
9641 MW153266.1 |Severe acute respiratory syndrome coro    ....................  ....................  .....................  ....................  ....................  ....................
9642 MW153267.1 |Severe acute respiratory syndrome coro    ....................  ....................  .....................  ....................  ....................  ....................
9643 MW153268.1 |Severe acute respiratory syndrome coro    ....................  ....................  .....................  ....................  ....................  ....................
9644 MW153269.1 |Severe acute respiratory syndrome coro    ....................  ....................  .....................  ....................  ....................  ....................
9645 MW153270.1 |Severe acute respiratory syndrome coro    ....................  ....................  .....................  ....................  ....................  ....................
9646 MW153271.1 |Severe acute respiratory syndrome coro    ....................  ....................  .....................  ....................  ....................  ....................
9647 MW153272.1 |Severe acute respiratory syndrome coro    ....................  ....................  .....................  ....................  ....................  ....................
9648 MW153273.1 |Severe acute respiratory syndrome coro    ....................  ....................  .....................  ....................  ....................  ....................
9649 MW153274.1 |Severe acute respiratory syndrome coro    ....................  ....................  .....................  ....................  ....................  ....................
9650 MW153275.1 |Severe acute respiratory syndrome coro    ....................  ....................  .....................  ....................  ....................  ....................
9651 MW153276.1 |Severe acute respiratory syndrome coro    ....................  ....................  .....................  ....................  ....................  ....................
9652 MW153277.1 |Severe acute respiratory syndrome coro    ....................  ....................  .....................  ....................  ....................  ....................
9653 MW153278.1 |Severe acute respiratory syndrome coro    ....................  ....................  .....................  ....................  ....................  ....................
9654 MW153279.1 |Severe acute respiratory syndrome coro    ....................  ....................  .....................  ....................  ....................  ....................
9655 MW153280.1 |Severe acute respiratory syndrome coro    ....................  ....................  .....................  ....................  ....................  ....................
9656 MW153281.1 |Severe acute respiratory syndrome coro    ....................  ....................  .....................  ....................  ....................  ....................
9657 MW153282.1 |Severe acute respiratory syndrome coro    ....................  ....................  .....................  ....................  ....................  ....................
9658 MW153283.1 |Severe acute respiratory syndrome coro    ....................  ....................  .....................  ....................  ....................  ....................
9659 MW153285.1 |Severe acute respiratory syndrome coro    ....................  ....................  .....................  ....................  ....................  ....................
9660 MW153286.1 |Severe acute respiratory syndrome coro    ....................  ....................  .....................  ....................  ....................  ....................
9661 MW153292.1 |Severe acute respiratory syndrome coro    ....................  ....................  .....................  ....................  ....................  ....................
9662 MW153293.1 |Severe acute respiratory syndrome coro    ....................  ....................  .....................  ....................  ....................  ....................
9663 MW153294.1 |Severe acute respiratory syndrome coro    ....................  ....................  .....................  ....................  ....................  ....................
9664 MW153295.1 |Severe acute respiratory syndrome coro    ....................  ....................  .....................  ....................  ....................  ....................
9665 MW153296.1 |Severe acute respiratory syndrome coro    ....................  ....................  .....................  ....................  ....................  ....................
9666 MW153299.1 |Severe acute respiratory syndrome coro    ....................  ....................  .....................  ....................  ....................  ....................
9667 MW153300.1 |Severe acute respiratory syndrome coro    ....................  ....................  .....................  ....................  ....................  ....................
9668 MW153301.1 |Severe acute respiratory syndrome coro    ....................  ....................  .....................  ....................  ....................  ....................
9669 MW153302.1 |Severe acute respiratory syndrome coro    ....................  ....................  .....................  ....................  ....................  ....................
9670 MW153304.1 |Severe acute respiratory syndrome coro    ....................  ....................  .....................  ....................  ....................  ....................
9671 MW153305.1 |Severe acute respiratory syndrome coro    ....................  ....................  .....................  ....................  ....................  ....................
9672 MW153306.1 |Severe acute respiratory syndrome coro    ....................  ....................  .....................  ....................  ....................  ....................
9673 MW153311.1 |Severe acute respiratory syndrome coro    ....................  ....................  .....................  ....................  ....................  ....................
9674 MW153316.1 |Severe acute respiratory syndrome coro    ....................  ....................  .....................  ....................  ....................  ....................
9675 MW153318.1 |Severe acute respiratory syndrome coro    ....................  ....................  .....................  ....................  ....................  ....................
9676 MW153320.1 |Severe acute respiratory syndrome coro    ....................  ....................  .....................  ....................  ....................  ....................
9677 MW153322.1 |Severe acute respiratory syndrome coro    ....................  ....................  .....................  ....................  ....................  ....................
9678 MW153323.1 |Severe acute respiratory syndrome coro    ....................  ....................  .....................  ....................  ....................  ....................
9679 MW153324.1 |Severe acute respiratory syndrome coro    ....................  ....................  .....................  ....................  ....................  ....................
9680 MW153325.1 |Severe acute respiratory syndrome coro    ....................  ....................  .....................  ....................  ....................  ....................
9681 MW153326.1 |Severe acute respiratory syndrome coro    ....................  ....................  .....................  ....................  ....................  ....................
9682 MW153327.1 |Severe acute respiratory syndrome coro    ....................  ....................  .....................  ....................  ....................  ....................
9683 MW153328.1 |Severe acute respiratory syndrome coro    ....................  ....................  .....................  ....................  ....................  ....................
9684 MW153329.1 |Severe acute respiratory syndrome coro    ....................  ....................  .....................  ....................  ....................  ....................
9685 MW153331.1 |Severe acute respiratory syndrome coro    ....................  ....................  .....................  ....................  ....................  ....................
9686 MW153332.1 |Severe acute respiratory syndrome coro    ....................  ....................  .....................  ....................  ....................  ....................
```

```
9687 MW153333.1 |Severe acute respiratory syndrome coro    ····················  ····················  ·····················  ····················  ····················  ····················
9688 MW153334.1 |Severe acute respiratory syndrome coro    ····················  ····················  ·····················  ····················  ····················  ····················
9689 MW153337.1 |Severe acute respiratory syndrome coro    ····················  ····················  ·····················  ····················  ····················  ····················
9690 MW153338.1 |Severe acute respiratory syndrome coro    ····················  ····················  ·····················  ····················  ····················  ····················
9691 MW153339.1 |Severe acute respiratory syndrome coro    ····················  ····················  ·····················  ····················  ····················  ····················
9692 MW153340.1 |Severe acute respiratory syndrome coro    ····················  ····················  ·····················  ····················  ····················  ····················
9693 MW153341.1 |Severe acute respiratory syndrome coro    ····················  ····················  ·····················  ····················  ····················  ····················
9694 MW153342.1 |Severe acute respiratory syndrome coro    ····················  ······A·············  ·····················  ····················  ····················  ····················
9695 MW153343.1 |Severe acute respiratory syndrome coro    ····················  ····················  ·····················  ····················  ····················  ····················
9696 MW153345.1 |Severe acute respiratory syndrome coro    ····················  ····················  ·····················  ····················  ····················  ····················
9697 MW153346.1 |Severe acute respiratory syndrome coro    ····················  ····················  ·····················  ····················  ····················  ····················
9698 MW153347.1 |Severe acute respiratory syndrome coro    ····················  ····················  ·····················  ····················  ····················  ····················
9699 MW153350.1 |Severe acute respiratory syndrome coro    ····················  ····················  ·····················  ····················  ····················  ····················
9700 MW153351.1 |Severe acute respiratory syndrome coro    ····················  ····················  ·····················  ····················  ····················  ····················
9701 MW153352.1 |Severe acute respiratory syndrome coro    ····················  ····················  ·····················  ····················  ····················  ····················
9702 MW153354.1 |Severe acute respiratory syndrome coro    ····················  ····················  ·····················  ····················  ····················  ····················
9703 MW153356.1 |Severe acute respiratory syndrome coro    ····················  ····················  ·····················  ····················  ····················  ····················
9704 MW153357.1 |Severe acute respiratory syndrome coro    ····················  ····················  ·····················  ····················  ····················  ····················
9705 MW153358.1 |Severe acute respiratory syndrome coro    ····················  ····················  ·····················  ····················  ····················  ····················
9706 MW153360.1 |Severe acute respiratory syndrome coro    ····················  ····················  ·····················  ····················  ····················  ····················
9707 MW153361.1 |Severe acute respiratory syndrome coro    ····················  ····················  ·····················  ····················  ····················  ····················
9708 MW153363.1 |Severe acute respiratory syndrome coro    ····················  ····················  ·····················  ····················  ····················  ····················
9709 MW153364.1 |Severe acute respiratory syndrome coro    ····················  ····················  ·····················  ····················  ····················  ····················
9710 MW153365.1 |Severe acute respiratory syndrome coro    ····················  ····················  ·····················  ····················  ····················  ····················
9711 MW153368.1 |Severe acute respiratory syndrome coro    ····················  ····················  ·····················  ····················  ····················  ····················
9712 MW153369.1 |Severe acute respiratory syndrome coro    ····················  ····················  ·····················  ····················  ····················  ····················
9713 MW153371.1 |Severe acute respiratory syndrome coro    ····················  ····················  ·····················  ····················  ····················  ····················
9714 MW153372.1 |Severe acute respiratory syndrome coro    ····················  ····················  ·····················  ····················  ····················  ····················
9715 MW153373.1 |Severe acute respiratory syndrome coro    ····················  ····················  ·····················  ····················  ····················  ····················
9716 MW153374.1 |Severe acute respiratory syndrome coro    ····················  ····················  ·····················  ····················  ····················  ····················
9717 MW153375.1 |Severe acute respiratory syndrome coro    ····················  ····················  ·····················  ····················  ····················  ····················
9718 MW153376.1 |Severe acute respiratory syndrome coro    ····················  ····················  ·····················  ····················  ····················  ····················
9719 MW153377.1 |Severe acute respiratory syndrome coro    ····················  ····················  ·····················  ····················  ····················  ····················
9720 MW153379.1 |Severe acute respiratory syndrome coro    ····················  ····················  ·····················  ····················  ····················  ····················
9721 MW153380.1 |Severe acute respiratory syndrome coro    ····················  ····················  ·····················  ····················  ····················  ····················
9722 MW153381.1 |Severe acute respiratory syndrome coro    ····················  ····················  ·····················  ····················  ····················  ····················
9723 MW153382.1 |Severe acute respiratory syndrome coro    ····················  ····················  ·····················  ····················  ····················  ····················
9724 MW153383.1 |Severe acute respiratory syndrome coro    ····················  ····················  ·····················  ····················  ····················  ····················
9725 MW153384.1 |Severe acute respiratory syndrome coro    ····················  ····················  ·····················  ····················  ····················  ····················
9726 MW153385.1 |Severe acute respiratory syndrome coro    ····················  ····················  ·····················  ····················  ····················  ····················
9727 MW153387.1 |Severe acute respiratory syndrome coro    ····················  ····················  ·····················  ····················  ····················  ····················
9728 MW153389.1 |Severe acute respiratory syndrome coro    ····················  ····················  ·····················  ····················  ····················  ····················
9729 MW153390.1 |Severe acute respiratory syndrome coro    ····················  ····················  ·····················  ····················  ····················  ····················
9730 MW153392.1 |Severe acute respiratory syndrome coro    ····················  ····················  ·····················  ····················  ····················  ····················
9731 MW153394.1 |Severe acute respiratory syndrome coro    ····················  ····················  ·····················  ····················  ····················  ····················
9732 MW153395.1 |Severe acute respiratory syndrome coro    ····················  ····················  ·····················  ····················  ····················  ····················
9733 MW153396.1 |Severe acute respiratory syndrome coro    ····················  ····················  ·····················  ····················  ····················  ····················
9734 MW153397.1 |Severe acute respiratory syndrome coro    ····················  ····················  ·····················  ····················  ····················  ····················
9735 MW153398.1 |Severe acute respiratory syndrome coro    ····················  ····················  ·····················  ····················  ····················  ····················
9736 MW153399.1 |Severe acute respiratory syndrome coro    ····················  ····················  ·····················  ····················  ····················  ····················
9737 MW153400.1 |Severe acute respiratory syndrome coro    ····················  ····················  ·····················  ····················  ····················  ····················
9738 MW153401.1 |Severe acute respiratory syndrome coro    ····················  ····················  ·····················  ····················  ····················  ····················
9739 MW153402.1 |Severe acute respiratory syndrome coro    ····················  ····················  ·····················  ····················  ····················  ····················
9740 MW153403.1 |Severe acute respiratory syndrome coro    ····················  ····················  ·····················  ····················  ····················  ····················
9741 MW153404.1 |Severe acute respiratory syndrome coro    ····················  ····················  ·····················  ····················  ····················  ····················
9742 MW153405.1 |Severe acute respiratory syndrome coro    ····················  ····················  ·····················  ····················  ····················  ····················
9743 MW153406.1 |Severe acute respiratory syndrome coro    ····················  ····················  ·····················  ····················  ····················  ····················
9744 MW153407.1 |Severe acute respiratory syndrome coro    ····················  ····················  ·····················  ····················  ····················  ····················
9745 MW153408.1 |Severe acute respiratory syndrome coro    ····················  ····················  ·····················  ····················  ····················  ····················
9746 MW153409.1 |Severe acute respiratory syndrome coro    ····················  ····················  ·····················  ····················  ····················  ····················
9747 MW153411.1 |Severe acute respiratory syndrome coro    ····················  ····················  ·····················  ····················  ····················  ····················
9748 MW153412.1 |Severe acute respiratory syndrome coro    ····················  ····················  ·····················  ····················  ····················  ····················
9749 MW153413.1 |Severe acute respiratory syndrome coro    ····················  ····················  ·····················  ····················  ····················  ····················
9750 MW153415.1 |Severe acute respiratory syndrome coro    ····················  ····················  ·····················  ····················  ····················  ····················
9751 MW153416.1 |Severe acute respiratory syndrome coro    ····················  ····················  ·····················  ····················  ····················  ····················
9752 MW153417.1 |Severe acute respiratory syndrome coro    ····················  ····················  ·····················  ····················  ····················  ····················
9753 MW153418.1 |Severe acute respiratory syndrome coro    ····················  ····················  ·····················  ····················  ····················  ····················
9754 MW153420.1 |Severe acute respiratory syndrome coro    ····················  ····················  ·····················  ····················  ····················  ····················
9755 MW153421.1 |Severe acute respiratory syndrome coro    ····················  ····················  ·····················  ····················  ····················  ····················
9756 MW153422.1 |Severe acute respiratory syndrome coro    ····················  ····················  ·····················  ····················  ····················  ····················
9757 MW153423.1 |Severe acute respiratory syndrome coro    ····················  ····················  ·····················  ····················  ····················  ····················
9758 MW153424.1 |Severe acute respiratory syndrome coro    ····················  ····················  ·····················  ····················  ····················  ····················
9759 MW153425.1 |Severe acute respiratory syndrome coro    ····················  ····················  ·····················  ····················  ····················  ····················
9760 MW153426.1 |Severe acute respiratory syndrome coro    ····················  ····················  ·····················  ····················  ····················  ····················
9761 MW153427.1 |Severe acute respiratory syndrome coro    ····················  ····················  ·····················  ····················  ····················  ····················
9762 MW153428.1 |Severe acute respiratory syndrome coro    ····················  ····················  ·····················  ····················  ····················  ····················
9763 MW153429.1 |Severe acute respiratory syndrome coro    ····················  ····················  ·····················  ····················  ····················  ····················
9764 MW153430.1 |Severe acute respiratory syndrome coro    ····················  ····················  ·····················  ····················  ····················  ····················
```

| | | | | | | | | |
|---|---|---|---|---|---|---|---|---|
| 9765 | MW153431.1 | \|Severe acute respiratory syndrome coro | .................... | .................... | ..................... | .................... | .................... | .................... |
| 9766 | MW153432.1 | \|Severe acute respiratory syndrome coro | .................... | .................... | ..................... | .................... | .................... | .................... |
| 9767 | MW153433.1 | \|Severe acute respiratory syndrome coro | .................... | .................... | ..................... | .................... | .................... | .................... |
| 9768 | MW153434.1 | \|Severe acute respiratory syndrome coro | .................... | .................... | ..................... | .................... | .................... | .................... |
| 9769 | MW153435.1 | \|Severe acute respiratory syndrome coro | .................... | .................... | ..................... | .................... | .................... | .................... |
| 9770 | MW153436.1 | \|Severe acute respiratory syndrome coro | .................... | .................... | ..................... | .................... | .................... | .................... |
| 9771 | MW153437.1 | \|Severe acute respiratory syndrome coro | .................... | .................... | ..................... | .................... | .................... | .................... |
| 9772 | MW153438.1 | \|Severe acute respiratory syndrome coro | .................... | .................... | ..................... | .................... | .................... | .................... |
| 9773 | MW153439.1 | \|Severe acute respiratory syndrome coro | .................... | .................... | ..................... | .................... | .................... | .................... |
| 9774 | MW153440.1 | \|Severe acute respiratory syndrome coro | .................... | .................... | ..................... | .................... | .................... | .................... |
| 9775 | MW153441.1 | \|Severe acute respiratory syndrome coro | .................... | .................... | ..................... | .................... | .................... | .................... |
| 9776 | MW153444.1 | \|Severe acute respiratory syndrome coro | .................... | .................... | ..................... | .................... | .................... | .................... |
| 9777 | MW153445.1 | \|Severe acute respiratory syndrome coro | .................... | .................... | ..................... | .................... | .................... | .................... |
| 9778 | MW153446.1 | \|Severe acute respiratory syndrome coro | .................... | .................... | ..................... | .................... | .................... | .................... |
| 9779 | MW153447.1 | \|Severe acute respiratory syndrome coro | .................... | .................... | ..................... | .................... | .................... | .................... |
| 9780 | MW153448.1 | \|Severe acute respiratory syndrome coro | .................... | .................... | ..................... | .................... | .................... | .................... |
| 9781 | MW153449.1 | \|Severe acute respiratory syndrome coro | .................... | .................... | ..................... | .................... | .................... | .................... |
| 9782 | MW153451.1 | \|Severe acute respiratory syndrome coro | .................... | .................... | ..................... | .................... | .................... | .................... |
| 9783 | MW153452.1 | \|Severe acute respiratory syndrome coro | .................... | .................... | ..................... | .................... | .................... | .................... |
| 9784 | MW153453.1 | \|Severe acute respiratory syndrome coro | .................... | .................... | ..................... | .................... | .................... | .................... |
| 9785 | MW153454.1 | \|Severe acute respiratory syndrome coro | .................... | .................... | ..................... | .................... | .................... | .................... |
| 9786 | MW153455.1 | \|Severe acute respiratory syndrome coro | .................... | .................... | ..................... | .................... | .................... | .................... |
| 9787 | MW153456.1 | \|Severe acute respiratory syndrome coro | .................... | .................... | ..................... | .................... | .................... | .................... |
| 9788 | MW153457.1 | \|Severe acute respiratory syndrome coro | .................... | .................... | ..................... | .................... | .................... | .................... |
| 9789 | MW153458.1 | \|Severe acute respiratory syndrome coro | .................... | .................... | ..................... | .................... | .................... | .................... |
| 9790 | MW153459.1 | \|Severe acute respiratory syndrome coro | .................... | .................... | ..................... | .................... | .................... | .................... |
| 9791 | MW153460.1 | \|Severe acute respiratory syndrome coro | .................... | .................... | ..................... | .................... | .................... | .................... |
| 9792 | MW153461.1 | \|Severe acute respiratory syndrome coro | .................... | .................... | ..................... | .................... | .................... | .................... |
| 9793 | MW153462.1 | \|Severe acute respiratory syndrome coro | .................... | .................... | ..................... | .................... | .................... | .................... |
| 9794 | MW153463.1 | \|Severe acute respiratory syndrome coro | .................... | .................... | ..................... | .................... | .................... | .................... |
| 9795 | MW153464.1 | \|Severe acute respiratory syndrome coro | .................... | .................... | ..................... | .................... | .................... | .................... |
| 9796 | MW153465.1 | \|Severe acute respiratory syndrome coro | .................... | .................... | ..................... | .................... | .................... | .................... |
| 9797 | MW153466.1 | \|Severe acute respiratory syndrome coro | .................... | .................... | ..................... | .................... | .................... | .................... |
| 9798 | MW153467.1 | \|Severe acute respiratory syndrome coro | .................... | .................... | ..................... | .................... | .................... | .................... |
| 9799 | MW153468.1 | \|Severe acute respiratory syndrome coro | .................... | .................... | ..................... | .................... | .................... | .................... |
| 9800 | MW153469.1 | \|Severe acute respiratory syndrome coro | .................... | .................... | ..................... | .................... | .................... | .................... |
| 9801 | MW153470.1 | \|Severe acute respiratory syndrome coro | .................... | .................... | ..................... | .................... | .................... | .................... |
| 9802 | MW153471.1 | \|Severe acute respiratory syndrome coro | .................... | .................... | ..................... | .................... | .................... | .................... |
| 9803 | MW153472.1 | \|Severe acute respiratory syndrome coro | .................... | .................... | ..................... | .................... | .................... | .................... |
| 9804 | MW153473.1 | \|Severe acute respiratory syndrome coro | .................... | .................... | ..................... | .................... | .................... | .................... |
| 9805 | MW153474.1 | \|Severe acute respiratory syndrome coro | .................... | .................... | ..................... | .................... | .................... | .................... |
| 9806 | MW153475.1 | \|Severe acute respiratory syndrome coro | .................... | .................... | ..................... | .................... | .................... | .................... |
| 9807 | MW153476.1 | \|Severe acute respiratory syndrome coro | .................... | .................... | ..................... | .................... | .................... | .................... |
| 9808 | MW153477.1 | \|Severe acute respiratory syndrome coro | .................... | .................... | ..................... | .................... | .................... | .................... |
| 9809 | MW153478.1 | \|Severe acute respiratory syndrome coro | .................... | ......A............. | ..................... | .................... | .................... | .................... |
| 9810 | MW153479.1 | \|Severe acute respiratory syndrome coro | .................... | ......A............. | ..................... | .................... | .................... | .................... |
| 9811 | MW153480.1 | \|Severe acute respiratory syndrome coro | .................... | .................... | ..................... | .................... | .................... | .................... |
| 9812 | MW153481.1 | \|Severe acute respiratory syndrome coro | .................... | .................... | ..................... | .................... | .................... | .................... |
| 9813 | MW153482.1 | \|Severe acute respiratory syndrome coro | .................... | .................... | ..................... | .................... | .................... | .................... |
| 9814 | MW153483.1 | \|Severe acute respiratory syndrome coro | .................... | .................... | ..................... | .................... | .................... | .................... |
| 9815 | MW153484.1 | \|Severe acute respiratory syndrome coro | .................... | .................... | ..................... | .................... | .................... | .................... |
| 9816 | MW153486.1 | \|Severe acute respiratory syndrome coro | .................... | .................... | ..................... | .................... | .................... | .................... |
| 9817 | MW153487.1 | \|Severe acute respiratory syndrome coro | .................... | .................... | ..................... | .................... | .................... | .................... |
| 9818 | MW153488.1 | \|Severe acute respiratory syndrome coro | .................... | .................... | ..................... | .................... | .................... | .................... |
| 9819 | MW153489.1 | \|Severe acute respiratory syndrome coro | .................... | .................... | ..................... | .................... | .................... | .................... |
| 9820 | MW153490.1 | \|Severe acute respiratory syndrome coro | .................... | .................... | ..................... | .................... | .................... | .................... |
| 9821 | MW153491.1 | \|Severe acute respiratory syndrome coro | .................... | .................... | ..................... | .................... | .................... | .................... |
| 9822 | MW153492.1 | \|Severe acute respiratory syndrome coro | .................... | .................... | ..................... | .................... | .................... | .................... |
| 9823 | MW153493.1 | \|Severe acute respiratory syndrome coro | .................... | .................... | ..................... | .................... | .................... | .................... |
| 9824 | MW153495.1 | \|Severe acute respiratory syndrome coro | .................... | .................... | ..................... | .................... | .................... | .................... |
| 9825 | MW153496.1 | \|Severe acute respiratory syndrome coro | .................... | .................... | ..................... | .................... | .................... | .................... |
| 9826 | MW153497.1 | \|Severe acute respiratory syndrome coro | .................... | .................... | ..................... | .................... | .................... | .................... |
| 9827 | MW153498.1 | \|Severe acute respiratory syndrome coro | .................... | .................... | ..................... | .................... | .................... | .................... |
| 9828 | MW153499.1 | \|Severe acute respiratory syndrome coro | .................... | .................... | ..................... | .................... | .................... | .................... |
| 9829 | MW153500.1 | \|Severe acute respiratory syndrome coro | .................... | .................... | ..................... | .................... | .................... | .................... |
| 9830 | MW153501.1 | \|Severe acute respiratory syndrome coro | .................... | .................... | ..................... | .................... | .................... | .................... |
| 9831 | MW153502.1 | \|Severe acute respiratory syndrome coro | .................... | .................... | ..................... | .................... | .................... | .................... |
| 9832 | MW153503.1 | \|Severe acute respiratory syndrome coro | .................... | .................... | ..................... | .................... | .................... | .................... |
| 9833 | MW153504.1 | \|Severe acute respiratory syndrome coro | .................... | .................... | ..................... | .................... | .................... | .................... |
| 9834 | MW153505.1 | \|Severe acute respiratory syndrome coro | .................... | .................... | ..................... | .................... | .................... | .................... |
| 9835 | MW153506.1 | \|Severe acute respiratory syndrome coro | .................... | .................... | ..................... | .................... | .................... | .................... |
| 9836 | MW153507.1 | \|Severe acute respiratory syndrome coro | .................... | .................... | ..................... | .................... | .................... | .................... |
| 9837 | MW153508.1 | \|Severe acute respiratory syndrome coro | .................... | .................... | ..................... | .................... | .................... | .................... |
| 9838 | MW153509.1 | \|Severe acute respiratory syndrome coro | .................... | .................... | ..................... | .................... | .................... | .................... |
| 9839 | MW153510.1 | \|Severe acute respiratory syndrome coro | .................... | .................... | ..................... | .................... | .................... | .................... |
| 9840 | MW153511.1 | \|Severe acute respiratory syndrome coro | .................... | .................... | ..................... | .................... | .................... | .................... |
| 9841 | MW153512.1 | \|Severe acute respiratory syndrome coro | .................... | .................... | ..................... | .................... | .................... | .................... |
| 9842 | MW153513.1 | \|Severe acute respiratory syndrome coro | .................... | .................... | ..................... | .................... | .................... | .................... |

```
9843 MW153514.1 |Severe acute respiratory syndrome coro   ....................  ....................  ....................  ....................  ....................  ....................
9844 MW153515.1 |Severe acute respiratory syndrome coro   ....................  ....................  ....................  ....................  ....................  ....................
9845 MW153516.1 |Severe acute respiratory syndrome coro   ....................  ....................  ....................  ....................  ....................  ....................
9846 MW153517.1 |Severe acute respiratory syndrome coro   ....................  ....................  ....................  ....................  ....................  ....................
9847 MW153518.1 |Severe acute respiratory syndrome coro   ....................  ....................  ....................  ....................  ....................  ....................
9848 MW153520.1 |Severe acute respiratory syndrome coro   ....................  ....................  ....................  ....................  ....................  ....................
9849 MW153521.1 |Severe acute respiratory syndrome coro   ....................  ....................  ....................  ....................  ....................  ....................
9850 MW153522.1 |Severe acute respiratory syndrome coro   ....................  ....................  ....................  ....................  ....................  ....................
9851 MW153525.1 |Severe acute respiratory syndrome coro   ....................  ....................  ....................  ....................  ....................  ....................
9852 MW153526.1 |Severe acute respiratory syndrome coro   ....................  ....................  ....................  ....................  ....................  ....................
9853 MW153527.1 |Severe acute respiratory syndrome coro   ....................  ....................  ....................  ....................  ....................  ....................
9854 MW153528.1 |Severe acute respiratory syndrome coro   ....................  ....................  ....................  ....................  ....................  ....................
9855 MW153529.1 |Severe acute respiratory syndrome coro   ....................  ....................  ....................  ....................  ....................  ....................
9856 MW153530.1 |Severe acute respiratory syndrome coro   ....................  ....................  ....................  ....................  ....................  ....................
9857 MW153532.1 |Severe acute respiratory syndrome coro   ....................  ....................  ....................  ....................  ....................  ....................
9858 MW153533.1 |Severe acute respiratory syndrome coro   ....................  ....................  ....................  ....................  ....................  ....................
9859 MW153534.1 |Severe acute respiratory syndrome coro   ....................  ....................  ....................  ....................  ....................  ....................
9860 MW153536.1 |Severe acute respiratory syndrome coro   ....................  ....................  ....................  ....................  ....................  ....................
9861 MW153538.1 |Severe acute respiratory syndrome coro   ....................  ....................  ....................  ....................  ....................  ....................
9862 MW153540.1 |Severe acute respiratory syndrome coro   ....................  ....................  ....................  ....................  ....................  ....................
9863 MW153541.1 |Severe acute respiratory syndrome coro   ....................  ....................  ....................  ....................  ....................  ....................
9864 MW153543.1 |Severe acute respiratory syndrome coro   ....................  ....................  ....................  ....................  ....................  ....................
9865 MW153544.1 |Severe acute respiratory syndrome coro   ....................  ....................  ....................  ....................  ....................  ....................
9866 MW153545.1 |Severe acute respiratory syndrome coro   ....................  ....................  ....................  ....................  ....................  ....................
9867 MW153546.1 |Severe acute respiratory syndrome coro   ....................  ....................  ....................  ....................  ....................  ....................
9868 MW153547.1 |Severe acute respiratory syndrome coro   ....................  ....................  ....................  ....................  ....................  ....................
9869 MW153549.1 |Severe acute respiratory syndrome coro   ....................  ....................  ....................  ....................  ....................  ....................
9870 MW153550.1 |Severe acute respiratory syndrome coro   ....................  ....................  ....................  ....................  ....................  ....................
9871 MW153551.1 |Severe acute respiratory syndrome coro   ....................  ....................  ....................  ....................  ....................  ....................
9872 MW153552.1 |Severe acute respiratory syndrome coro   ....................  ....................  ....................  ....................  ....................  ....................
9873 MW153553.1 |Severe acute respiratory syndrome coro   ....................  ....................  ....................  ....................  ....................  ....................
9874 MW153554.1 |Severe acute respiratory syndrome coro   ....................  ....................  ....................  ....................  ....................  ....................
9875 MW153555.1 |Severe acute respiratory syndrome coro   ....................  ....................  ....................  ....................  ....................  ....................
9876 MW153556.1 |Severe acute respiratory syndrome coro   ....................  ....................  ....................  ....................  ....................  ....................
9877 MW153557.1 |Severe acute respiratory syndrome coro   ....................  ....................  ....................  ....................  ....................  ....................
9878 MW153558.1 |Severe acute respiratory syndrome coro   ....................  ....................  ....................  ....................  ....................  ....................
9879 MW153559.1 |Severe acute respiratory syndrome coro   ....................  ....................  ....................  ....................  ....................  ....................
9880 MW153560.1 |Severe acute respiratory syndrome coro   ....................  ....................  ....................  ....................  ....................  ....................
9881 MW153561.1 |Severe acute respiratory syndrome coro   ....................  ....................  ....................  ....................  ....................  ....................
9882 MW153562.1 |Severe acute respiratory syndrome coro   ....................  ....................  ....................  ....................  ....................  ....................
9883 MW153563.1 |Severe acute respiratory syndrome coro   ....................  ....................  ....................  ....................  ....................  ....................
9884 MW153564.1 |Severe acute respiratory syndrome coro   ....................  ....................  ....................  ....................  ....................  ....................
9885 MW153566.1 |Severe acute respiratory syndrome coro   ....................  ....................  ....................  ....................  ....................  ....................
9886 MW153567.1 |Severe acute respiratory syndrome coro   ....................  ....................  ....................  ....................  ....................  ....................
9887 MW153568.1 |Severe acute respiratory syndrome coro   ....................  ....................  ....................  ....................  ....................  ....................
9888 MW153569.1 |Severe acute respiratory syndrome coro   ....................  ....................  ....................  ....................  ....................  ....................
9889 MW153570.1 |Severe acute respiratory syndrome coro   ....................  ....................  ....................  ....................  ....................  ....................
9890 MW153571.1 |Severe acute respiratory syndrome coro   ....................  ....................  ....................  ....................  ....................  ....................
9891 MW153573.1 |Severe acute respiratory syndrome coro   ....................  ....................  ....................  ....................  ....................  ....................
9892 MW153574.1 |Severe acute respiratory syndrome coro   ....................  ....................  ....................  ....................  ....................  ....................
9893 MW153575.1 |Severe acute respiratory syndrome coro   ....................  ....................  ....................  ....................  ....................  ....................
9894 MW153576.1 |Severe acute respiratory syndrome coro   ....................  ....................  ....................  ....................  ....................  ....................
9895 MW153578.1 |Severe acute respiratory syndrome coro   ....................  ....................  ....................  ....................  ....................  ....................
9896 MW153579.1 |Severe acute respiratory syndrome coro   ....................  ....................  ....................  ....................  ....................  ....................
9897 MW153581.1 |Severe acute respiratory syndrome coro   ....................  ....................  ....................  ....................  ....................  ....................
9898 MW153582.1 |Severe acute respiratory syndrome coro   ....................  ....................  ....................  ....................  ....................  ....................
9899 MW153583.1 |Severe acute respiratory syndrome coro   ....................  ....................  ....................  ....................  ....................  ....................
9900 MW153584.1 |Severe acute respiratory syndrome coro   ....................  ....................  ....................  ....................  ....................  ....................
9901 MW153585.1 |Severe acute respiratory syndrome coro   ....................  ....................  ....................  ....................  ....................  ....................
9902 MW153586.1 |Severe acute respiratory syndrome coro   ....................  ....................  ....................  ....................  ....................  ....................
9903 MW153587.1 |Severe acute respiratory syndrome coro   ....................  ....................  ....................  ....................  ....................  ....................
9904 MW153588.1 |Severe acute respiratory syndrome coro   ....................  ....................  ....................  ....................  ....................  ....................
9905 MW153590.1 |Severe acute respiratory syndrome coro   ....................  ....................  ....................  ....................  ....................  ....................
9906 MW153591.1 |Severe acute respiratory syndrome coro   ....................  ....................  ....................  ....................  ....................  ....................
9907 MW153592.1 |Severe acute respiratory syndrome coro   ....................  ....................  ....................  ....................  ....................  ....................
9908 MW153593.1 |Severe acute respiratory syndrome coro   ....................  ....................  ....................  ....................  ....................  ....................
9909 MW153594.1 |Severe acute respiratory syndrome coro   ....................  ....................  ....................  ....................  ....................  ....................
9910 MW153595.1 |Severe acute respiratory syndrome coro   ....................  ....................  ....................  ....................  ....................  ....................
9911 MW153596.1 |Severe acute respiratory syndrome coro   ....................  ....................  ....................  ....................  ....................  ....................
9912 MW153597.1 |Severe acute respiratory syndrome coro   ....................  ....................  ....................  ....................  ....................  ....................
9913 MW153598.1 |Severe acute respiratory syndrome coro   ....................  ....................  ....................  ....................  ....................  ....................
9914 MW153600.1 |Severe acute respiratory syndrome coro   ....................  ....................  ....................  ....................  ....................  ....................
9915 MW153601.1 |Severe acute respiratory syndrome coro   ....................  ....................  ....................  ....................  ....................  ....................
9916 MW153602.1 |Severe acute respiratory syndrome coro   ....................  ....................  ....................  ....................  ....................  ....................
9917 MW153603.1 |Severe acute respiratory syndrome coro   ....................  ....................  ....................  ....................  ....................  ....................
9918 MW153605.1 |Severe acute respiratory syndrome coro   ....................  ....................  ....................  ....................  ....................  ....................
9919 MW153606.1 |Severe acute respiratory syndrome coro   ....................  ....................  ....................  ....................  ....................  ....................
9920 MW153607.1 |Severe acute respiratory syndrome coro   ....................  ....................  ....................  ....................  ....................  ....................
```

| | | | | | | | | |
|---|---|---|---|---|---|---|---|---|
| 9921 | MW153608.1 | \|Severe acute respiratory syndrome coro | .................... | .................... | .................... | .................... | .................... | .................... |
| 9922 | MW153609.1 | \|Severe acute respiratory syndrome coro | .................... | .................... | .................... | .................... | .................... | .................... |
| 9923 | MW153610.1 | \|Severe acute respiratory syndrome coro | .................... | .................... | .................... | .................... | .................... | .................... |
| 9924 | MW153611.1 | \|Severe acute respiratory syndrome coro | .................... | .................... | .................... | .................... | .................... | .................... |
| 9925 | MW153612.1 | \|Severe acute respiratory syndrome coro | .................... | .................... | .................... | .................... | .................... | .................... |
| 9926 | MW153613.1 | \|Severe acute respiratory syndrome coro | .................... | .................... | .................... | .................... | .................... | .................... |
| 9927 | MW153614.1 | \|Severe acute respiratory syndrome coro | .................... | .................... | .................... | .................... | .................... | .................... |
| 9928 | MW153615.1 | \|Severe acute respiratory syndrome coro | .................... | .................... | .................... | .................... | .................... | .................... |
| 9929 | MW153616.1 | \|Severe acute respiratory syndrome coro | .................... | .................... | .................... | .................... | .................... | .................... |
| 9930 | MW153617.1 | \|Severe acute respiratory syndrome coro | .................... | .................... | .................... | .................... | .................... | .................... |
| 9931 | MW153618.1 | \|Severe acute respiratory syndrome coro | .................... | .................... | .................... | .................... | .................... | .................... |
| 9932 | MW153620.1 | \|Severe acute respiratory syndrome coro | .................... | .................... | .................... | .................... | .................... | .................... |
| 9933 | MW153621.1 | \|Severe acute respiratory syndrome coro | .................... | .................... | .................... | .................... | .................... | .................... |
| 9934 | MW153622.1 | \|Severe acute respiratory syndrome coro | .................... | .................... | .................... | .................... | .................... | .................... |
| 9935 | MW153623.1 | \|Severe acute respiratory syndrome coro | .................... | .................... | .................... | .................... | .................... | .................... |
| 9936 | MW153627.1 | \|Severe acute respiratory syndrome coro | .................... | .................... | .................... | .................... | .................... | .................... |
| 9937 | MW153628.1 | \|Severe acute respiratory syndrome coro | .................... | .................... | .................... | .................... | .................... | .................... |
| 9938 | MW153629.1 | \|Severe acute respiratory syndrome coro | .................... | .................... | .................... | .................... | .................... | .................... |
| 9939 | MW153630.1 | \|Severe acute respiratory syndrome coro | .................... | .................... | .................... | .................... | .................... | .................... |
| 9940 | MW153631.1 | \|Severe acute respiratory syndrome coro | .................... | .................... | .................... | .................... | .................... | .................... |
| 9941 | MW153633.1 | \|Severe acute respiratory syndrome coro | .................... | .................... | .................... | .................... | .................... | .................... |
| 9942 | MW153634.1 | \|Severe acute respiratory syndrome coro | .................... | .................... | .................... | .................... | .................... | .................... |
| 9943 | MW153635.1 | \|Severe acute respiratory syndrome coro | .................... | .................... | .................... | .................... | .................... | .................... |
| 9944 | MW153636.1 | \|Severe acute respiratory syndrome coro | .................... | .................... | .................... | .................... | .................... | .................... |
| 9945 | MW153637.1 | \|Severe acute respiratory syndrome coro | .................... | .................... | .................... | .................... | .................... | .................... |
| 9946 | MW153641.1 | \|Severe acute respiratory syndrome coro | .................... | .................... | .................... | .................... | .................... | .................... |
| 9947 | MW153642.1 | \|Severe acute respiratory syndrome coro | .................... | .................... | .................... | .................... | .................... | .................... |
| 9948 | MW153643.1 | \|Severe acute respiratory syndrome coro | .................... | .................... | .................... | .................... | .................... | .................... |
| 9949 | MW153644.1 | \|Severe acute respiratory syndrome coro | .................... | .................... | .................... | .................... | .................... | .................... |
| 9950 | MW153645.1 | \|Severe acute respiratory syndrome coro | .................... | .................... | .................... | .................... | .................... | .................... |
| 9951 | MW153646.1 | \|Severe acute respiratory syndrome coro | .................... | .................... | .................... | .................... | .................... | .................... |
| 9952 | MW153647.1 | \|Severe acute respiratory syndrome coro | .................... | .................... | .................... | .................... | .................... | .................... |
| 9953 | MW153648.1 | \|Severe acute respiratory syndrome coro | .................... | .................... | .................... | .................... | .................... | .................... |
| 9954 | MW153649.1 | \|Severe acute respiratory syndrome coro | .................... | .................... | .................... | .................... | .................... | .................... |
| 9955 | MW153650.1 | \|Severe acute respiratory syndrome coro | .................... | .................... | .................... | .................... | .................... | .................... |
| 9956 | MW153651.1 | \|Severe acute respiratory syndrome coro | .................... | .................... | .................... | .................... | .................... | .................... |
| 9957 | MW153652.1 | \|Severe acute respiratory syndrome coro | .................... | .................... | .................... | .................... | .................... | .................... |
| 9958 | MW153654.1 | \|Severe acute respiratory syndrome coro | .................... | .................... | .................... | .................... | .................... | .................... |
| 9959 | MW153656.1 | \|Severe acute respiratory syndrome coro | .................... | .................... | .................... | .................... | .................... | .................... |
| 9960 | MW153657.1 | \|Severe acute respiratory syndrome coro | .................... | .................... | .................... | .................... | .................... | .................... |
| 9961 | MW153658.1 | \|Severe acute respiratory syndrome coro | .................... | .................... | .................... | .................... | .................... | .................... |
| 9962 | MW153659.1 | \|Severe acute respiratory syndrome coro | .................... | .................... | .................... | .................... | .................... | .................... |
| 9963 | MW153660.1 | \|Severe acute respiratory syndrome coro | .................... | .................... | .................... | .................... | .................... | .................... |
| 9964 | MW153661.1 | \|Severe acute respiratory syndrome coro | .................... | .................... | .................... | .................... | .................... | .................... |
| 9965 | MW153662.1 | \|Severe acute respiratory syndrome coro | .................... | .................... | .................... | .................... | .................... | .................... |
| 9966 | MW153663.1 | \|Severe acute respiratory syndrome coro | .................... | .................... | .................... | .................... | .................... | .................... |
| 9967 | MW153664.1 | \|Severe acute respiratory syndrome coro | .................... | .................... | .................... | .................... | .................... | .................... |
| 9968 | MW153665.1 | \|Severe acute respiratory syndrome coro | .................... | .................... | .................... | .................... | .................... | .................... |
| 9969 | MW153666.1 | \|Severe acute respiratory syndrome coro | .................... | .................... | .................... | .................... | .................... | .................... |
| 9970 | MW153667.1 | \|Severe acute respiratory syndrome coro | .................... | .................... | .................... | .................... | .................... | .................... |
| 9971 | MW153668.1 | \|Severe acute respiratory syndrome coro | .................... | .................... | .................... | .................... | .................... | .................... |
| 9972 | MW153669.1 | \|Severe acute respiratory syndrome coro | .................... | .................... | .................... | .................... | .................... | .................... |
| 9973 | MW153670.1 | \|Severe acute respiratory syndrome coro | .................... | .................... | .................... | .................... | .................... | .................... |
| 9974 | MW153671.1 | \|Severe acute respiratory syndrome coro | .................... | .................... | .................... | .................... | .................... | .................... |
| 9975 | MW153672.1 | \|Severe acute respiratory syndrome coro | .................... | .................... | .................... | .................... | .................... | .................... |
| 9976 | MW153673.1 | \|Severe acute respiratory syndrome coro | .................... | .................... | .................... | .................... | .................... | .................... |
| 9977 | MW153674.1 | \|Severe acute respiratory syndrome coro | .................... | .................... | .................... | .................... | .................... | .................... |
| 9978 | MW153675.1 | \|Severe acute respiratory syndrome coro | .................... | .................... | .................... | .................... | .................... | .................... |
| 9979 | MW153677.1 | \|Severe acute respiratory syndrome coro | .................... | .................... | .................... | .................... | .................... | .................... |
| 9980 | MW153678.1 | \|Severe acute respiratory syndrome coro | .................... | .................... | .................... | .................... | .................... | .................... |
| 9981 | MW153679.1 | \|Severe acute respiratory syndrome coro | .................... | .................... | .................... | .................... | .................... | .................... |
| 9982 | MW153680.1 | \|Severe acute respiratory syndrome coro | .................... | .................... | .................... | .................... | .................... | .................... |
| 9983 | MW153681.1 | \|Severe acute respiratory syndrome coro | .................... | .................... | .................... | .................... | .................... | .................... |
| 9984 | MW153682.1 | \|Severe acute respiratory syndrome coro | .................... | .................... | .................... | .................... | .................... | .................... |
| 9985 | MW153683.1 | \|Severe acute respiratory syndrome coro | .................... | .................... | .................... | .................... | .................... | .................... |
| 9986 | MW153685.1 | \|Severe acute respiratory syndrome coro | .................... | .................... | .................... | .................... | .................... | .................... |
| 9987 | MW153686.1 | \|Severe acute respiratory syndrome coro | .................... | .................... | .................... | .................... | .................... | .................... |
| 9988 | MW153687.1 | \|Severe acute respiratory syndrome coro | .................... | .................... | .................... | .................... | .................... | .................... |
| 9989 | MW153688.1 | \|Severe acute respiratory syndrome coro | .................... | .................... | .................... | .................... | .................... | .................... |
| 9990 | MW153689.1 | \|Severe acute respiratory syndrome coro | .................... | .................... | .................... | .................... | .................... | .................... |
| 9991 | MW153690.1 | \|Severe acute respiratory syndrome coro | ........T........... | .................... | .................... | .................... | .................... | .................... |
| 9992 | MW153691.1 | \|Severe acute respiratory syndrome coro | .................... | .................... | .................... | .................... | .................... | .................... |
| 9993 | MW153692.1 | \|Severe acute respiratory syndrome coro | .................... | .................... | .................... | .................... | .................... | .................... |
| 9994 | MW153693.1 | \|Severe acute respiratory syndrome coro | .................... | .................... | .................... | .................... | .................... | .................... |
| 9995 | MW153694.1 | \|Severe acute respiratory syndrome coro | .................... | .................... | .................... | .................... | .................... | .................... |
| 9996 | MW153695.1 | \|Severe acute respiratory syndrome coro | .................... | .................... | .................... | .................... | .................... | .................... |
| 9997 | MW153696.1 | \|Severe acute respiratory syndrome coro | .................... | .................... | .................... | .................... | .................... | .................... |
| 9998 | MW153697.1 | \|Severe acute respiratory syndrome coro | .................... | .................... | .................... | .................... | .................... | .................... |

| | | | | | | | | |
|---|---|---|---|---|---|---|---|---|
| 9999 | MW153698.1 | \|Severe acute respiratory syndrome coro | ···················· | ···················· | ···················· | ···················· | ···················· | ···················· |
| 10000 | MW153699.1 | \|Severe acute respiratory syndrome coro | ···················· | ···················· | ···················· | ···················· | ···················· | ···················· |
| 10001 | MW153700.1 | \|Severe acute respiratory syndrome coro | ···················· | ···················· | ···················· | ···················· | ···················· | ···················· |
| 10002 | MW153701.1 | \|Severe acute respiratory syndrome coro | ···················· | ···················· | ···················· | ···················· | ···················· | ···················· |
| 10003 | MW153702.1 | \|Severe acute respiratory syndrome coro | ···················· | ···················· | ···················· | ···················· | ···················· | ···················· |
| 10004 | MW153703.1 | \|Severe acute respiratory syndrome coro | ···················· | ···················· | ···················· | ···················· | ···················· | ···················· |
| 10005 | MW153704.1 | \|Severe acute respiratory syndrome coro | ···················· | ···················· | ···················· | ···················· | ···················· | ···················· |
| 10006 | MW153705.1 | \|Severe acute respiratory syndrome coro | ···················· | ···················· | ···················· | ···················· | ···················· | ···················· |
| 10007 | MW153706.1 | \|Severe acute respiratory syndrome coro | ···················· | ···················· | ···················· | ···················· | ···················· | ···················· |
| 10008 | MW153707.1 | \|Severe acute respiratory syndrome coro | ···················· | ···················· | ···················· | ···················· | ···················· | ···················· |
| 10009 | MW153708.1 | \|Severe acute respiratory syndrome coro | ···················· | ···················· | ···················· | ···················· | ···················· | ···················· |
| 10010 | MW153709.1 | \|Severe acute respiratory syndrome coro | ···················· | ···················· | ···················· | ···················· | ···················· | ···················· |
| 10011 | MW153710.1 | \|Severe acute respiratory syndrome coro | ···················· | ···················· | ···················· | ···················· | ···················· | ···················· |
| 10012 | MW153711.1 | \|Severe acute respiratory syndrome coro | ···················· | ···················· | ···················· | ···················· | ···················· | ···················· |
| 10013 | MW153712.1 | \|Severe acute respiratory syndrome coro | ···················· | ···················· | ···················· | ···················· | ···················· | ···················· |
| 10014 | MW153713.1 | \|Severe acute respiratory syndrome coro | ···················· | ······A············· | ···················· | ···················· | ···················· | ···················· |
| 10015 | MW153714.1 | \|Severe acute respiratory syndrome coro | ···················· | ···················· | ···················· | ···················· | ···················· | ···················· |
| 10016 | MW153716.1 | \|Severe acute respiratory syndrome coro | ···················· | ···················· | ···················· | ···················· | ···················· | ···················· |
| 10017 | MW153718.1 | \|Severe acute respiratory syndrome coro | ···················· | ···················· | ···················· | ···················· | ···················· | ···················· |
| 10018 | MW153719.1 | \|Severe acute respiratory syndrome coro | ···················· | ···················· | ···················· | ···················· | ···················· | ···················· |
| 10019 | MW153721.1 | \|Severe acute respiratory syndrome coro | ···················· | ···················· | ···················· | ···················· | ···················· | ···················· |
| 10020 | MW153722.1 | \|Severe acute respiratory syndrome coro | ···················· | ···················· | ···················· | ···················· | ···················· | ···················· |
| 10021 | MW153724.1 | \|Severe acute respiratory syndrome coro | ···················· | ···················· | ···················· | ···················· | ···················· | ···················· |
| 10022 | MW153726.1 | \|Severe acute respiratory syndrome coro | ···················· | ···················· | ···················· | ···················· | ···················· | ···················· |
| 10023 | MW153727.1 | \|Severe acute respiratory syndrome coro | ···················· | ···················· | ···················· | ···················· | ···················· | ···················· |
| 10024 | MW153728.1 | \|Severe acute respiratory syndrome coro | ···················· | ···················· | ···················· | ···················· | ···················· | ···················· |
| 10025 | MW153729.1 | \|Severe acute respiratory syndrome coro | ···················· | ···················· | ···················· | ···················· | ···················· | ···················· |
| 10026 | MW153730.1 | \|Severe acute respiratory syndrome coro | ···················· | ···················· | ···················· | ···················· | ···················· | ···················· |
| 10027 | MW153731.1 | \|Severe acute respiratory syndrome coro | ···················· | ···················· | ···················· | ···················· | ···················· | ···················· |
| 10028 | MW153732.1 | \|Severe acute respiratory syndrome coro | ···················· | ···················· | ···················· | ···················· | ···················· | ···················· |
| 10029 | MW153733.1 | \|Severe acute respiratory syndrome coro | ···················· | ···················· | ···················· | ···················· | ···················· | ···················· |
| 10030 | MW153734.1 | \|Severe acute respiratory syndrome coro | ···················· | ···················· | ···················· | ···················· | ···················· | ···················· |
| 10031 | MW153735.1 | \|Severe acute respiratory syndrome coro | ···················· | ···················· | ···················· | ···················· | ···················· | ···················· |
| 10032 | MW153736.1 | \|Severe acute respiratory syndrome coro | ···················· | ···················· | ···················· | ···················· | ···················· | ···················· |
| 10033 | MW153737.1 | \|Severe acute respiratory syndrome coro | ···················· | ···················· | ···················· | ···················· | ···················· | ···················· |
| 10034 | MW153738.1 | \|Severe acute respiratory syndrome coro | ···················· | ···················· | ···················· | ···················· | ···················· | ···················· |
| 10035 | MW153739.1 | \|Severe acute respiratory syndrome coro | ···················· | ···················· | ···················· | ···················· | ···················· | ···················· |
| 10036 | MW153740.1 | \|Severe acute respiratory syndrome coro | ···················· | ···················· | ···················· | ···················· | ···················· | ···················· |
| 10037 | MW153741.1 | \|Severe acute respiratory syndrome coro | ···················· | ···················· | ···················· | ···················· | ···················· | ···················· |
| 10038 | MW153742.1 | \|Severe acute respiratory syndrome coro | ···················· | ···················· | ···················· | ···················· | ···················· | ···················· |
| 10039 | MW153743.1 | \|Severe acute respiratory syndrome coro | ···················· | ···················· | ···················· | ···················· | ···················· | ···················· |
| 10040 | MW153744.1 | \|Severe acute respiratory syndrome coro | ···················· | ···················· | ···················· | ···················· | ···················· | ···················· |
| 10041 | MW153746.1 | \|Severe acute respiratory syndrome coro | ···················· | ···················· | ···················· | ···················· | ···················· | ···················· |
| 10042 | MW153747.1 | \|Severe acute respiratory syndrome coro | ···················· | ···················· | ···················· | ···················· | ···················· | ···················· |
| 10043 | MW153748.1 | \|Severe acute respiratory syndrome coro | ···················· | ···················· | ···················· | ···················· | ···················· | ···················· |
| 10044 | MW153749.1 | \|Severe acute respiratory syndrome coro | ···················· | ···················· | ···················· | ···················· | ···················· | ···················· |
| 10045 | MW153750.1 | \|Severe acute respiratory syndrome coro | ···················· | ···················· | ···················· | ···················· | ···················· | ···················· |
| 10046 | MW153751.1 | \|Severe acute respiratory syndrome coro | ···················· | ···················· | ···················· | ···················· | ···················· | ···················· |
| 10047 | MW153752.1 | \|Severe acute respiratory syndrome coro | ···················· | ···················· | ···················· | ···················· | ···················· | ···················· |
| 10048 | MW153753.1 | \|Severe acute respiratory syndrome coro | ···················· | ···················· | ···················· | ···················· | ···················· | ···················· |
| 10049 | MW153754.1 | \|Severe acute respiratory syndrome coro | ···················· | ···················· | ···················· | ···················· | ···················· | ···················· |
| 10050 | MW153755.1 | \|Severe acute respiratory syndrome coro | ···················· | ···················· | ···················· | ···················· | ···················· | ···················· |
| 10051 | MW153756.1 | \|Severe acute respiratory syndrome coro | ···················· | ···················· | ···················· | ···················· | ···················· | ···················· |
| 10052 | MW153757.1 | \|Severe acute respiratory syndrome coro | ···················· | ···················· | ···················· | ···················· | ···················· | ···················· |
| 10053 | MW153758.1 | \|Severe acute respiratory syndrome coro | ·····G·············· | ···················· | ···················· | ···················· | ···················· | ···················· |
| 10054 | MW153759.1 | \|Severe acute respiratory syndrome coro | ···················· | ···················· | ···················· | ···················· | ···················· | ···················· |
| 10055 | MW153760.1 | \|Severe acute respiratory syndrome coro | ···················· | ···················· | ···················· | ···················· | ···················· | ···················· |
| 10056 | MW153762.1 | \|Severe acute respiratory syndrome coro | ···················· | ···················· | ···················· | ···················· | ···················· | ···················· |
| 10057 | MW153764.1 | \|Severe acute respiratory syndrome coro | ···················· | ···················· | ···················· | ···················· | ···················· | ···················· |
| 10058 | MW153765.1 | \|Severe acute respiratory syndrome coro | ···················· | ···················· | ···················· | ···················· | ···················· | ···················· |
| 10059 | MW153766.1 | \|Severe acute respiratory syndrome coro | ···················· | ···················· | ···················· | ···················· | ···················· | ···················· |
| 10060 | MW153767.1 | \|Severe acute respiratory syndrome coro | ···················· | ···················· | ···················· | ···················· | ···················· | ···················· |
| 10061 | MW153768.1 | \|Severe acute respiratory syndrome coro | ···················· | ···················· | ···················· | ···················· | ···················· | ···················· |
| 10062 | MW153769.1 | \|Severe acute respiratory syndrome coro | ···················· | ···················· | ···················· | ···················· | ···················· | ···················· |
| 10063 | MW153771.1 | \|Severe acute respiratory syndrome coro | ···················· | ···················· | ···················· | ···················· | ···················· | ···················· |
| 10064 | MW153774.1 | \|Severe acute respiratory syndrome coro | ···················· | ···················· | ···················· | ···················· | ···················· | ···················· |
| 10065 | MW153775.1 | \|Severe acute respiratory syndrome coro | ···················· | ···················· | ···················· | ···················· | ···················· | ···················· |
| 10066 | MW153776.1 | \|Severe acute respiratory syndrome coro | ···················· | ···················· | ···················· | ···················· | ···················· | ···················· |
| 10067 | MW153778.1 | \|Severe acute respiratory syndrome coro | ···················· | ···················· | ···················· | ···················· | ···················· | ···················· |
| 10068 | MW153779.1 | \|Severe acute respiratory syndrome coro | ···················· | ···················· | ···················· | ···················· | ···················· | ···················· |
| 10069 | MW153780.1 | \|Severe acute respiratory syndrome coro | ···················· | ···················· | ···················· | ···················· | ···················· | ···················· |
| 10070 | MW153781.1 | \|Severe acute respiratory syndrome coro | ···················· | ···················· | ···················· | ···················· | ···················· | ···················· |
| 10071 | MW153782.1 | \|Severe acute respiratory syndrome coro | ···················· | ···················· | ···················· | ···················· | ···················· | ···················· |
| 10072 | MW153783.1 | \|Severe acute respiratory syndrome coro | ···················· | ···················· | ···················· | ···················· | ···················· | ···················· |
| 10073 | MW153784.1 | \|Severe acute respiratory syndrome coro | ···················· | ···················· | ···················· | ···················· | ···················· | ···················· |
| 10074 | MW153785.1 | \|Severe acute respiratory syndrome coro | ···················· | ···················· | ···················· | ···················· | ···················· | ···················· |
| 10075 | MW153786.1 | \|Severe acute respiratory syndrome coro | ···················· | ···················· | ···················· | ···················· | ···················· | ···················· |
| 10076 | MW153787.1 | \|Severe acute respiratory syndrome coro | ···················· | ···················· | ···················· | ···················· | ···················· | ···················· |

| | | | | | | | | |
|---|---|---|---|---|---|---|---|---|
| 10077 | MW153788.1 | Severe acute respiratory syndrome coro | .................... | .................... | ..................... | .................... | .................... | .................... |
| 10078 | MW153789.1 | Severe acute respiratory syndrome coro | .................... | .................... | ..................... | .................... | .................... | .................... |
| 10079 | MW153790.1 | Severe acute respiratory syndrome coro | .................... | .................... | ..................... | .................... | .................... | .................... |
| 10080 | MW153791.1 | Severe acute respiratory syndrome coro | .................... | .................... | ..................... | .................... | .................... | .................... |
| 10081 | MW153792.1 | Severe acute respiratory syndrome coro | .................... | .................... | ..................... | .................... | .................... | .................... |
| 10082 | MW153793.1 | Severe acute respiratory syndrome coro | .................... | .................... | ..................... | .................... | .................... | .................... |
| 10083 | MW153794.1 | Severe acute respiratory syndrome coro | .................... | .................... | ..................... | .................... | .................... | .................... |
| 10084 | MW153795.1 | Severe acute respiratory syndrome coro | .................... | .................... | ..................... | .................... | .................... | .................... |
| 10085 | MW153796.1 | Severe acute respiratory syndrome coro | .................... | .................... | ..................... | .................... | .................... | .................... |
| 10086 | MW153797.1 | Severe acute respiratory syndrome coro | .................... | .................... | ..................... | .................... | .................... | .................... |
| 10087 | MW153799.1 | Severe acute respiratory syndrome coro | .................... | .................... | ..................... | .................... | .................... | .................... |
| 10088 | MW153801.1 | Severe acute respiratory syndrome coro | .................... | .................... | ..................... | .................... | .................... | .................... |
| 10089 | MW153802.1 | Severe acute respiratory syndrome coro | .................... | .................... | ..................... | .................... | .................... | .................... |
| 10090 | MW153803.1 | Severe acute respiratory syndrome coro | .................... | .................... | ..................... | .................... | .................... | .................... |
| 10091 | MW153804.1 | Severe acute respiratory syndrome coro | .................... | .................... | ..................... | .................... | .................... | .................... |
| 10092 | MW153805.1 | Severe acute respiratory syndrome coro | .................... | .................... | ..................... | .................... | .................... | .................... |
| 10093 | MW153806.1 | Severe acute respiratory syndrome coro | .................... | .................... | ..................... | .................... | .................... | .................... |
| 10094 | MW153807.1 | Severe acute respiratory syndrome coro | .................... | .................... | ..................... | .................... | .................... | .................... |
| 10095 | MW153809.1 | Severe acute respiratory syndrome coro | .................... | .................... | ..................... | .................... | .................... | .................... |
| 10096 | MW153811.1 | Severe acute respiratory syndrome coro | .................... | .................... | ..................... | .................... | .................... | .................... |
| 10097 | MW153812.1 | Severe acute respiratory syndrome coro | .................... | .................... | ..................... | .................... | .................... | .................... |
| 10098 | MW153813.1 | Severe acute respiratory syndrome coro | .................... | .................... | ..................... | .................... | .................... | .................... |
| 10099 | MW153814.1 | Severe acute respiratory syndrome coro | .................... | .................... | ..................... | .................... | .................... | .................... |
| 10100 | MW153815.1 | Severe acute respiratory syndrome coro | .................... | .................... | ..................... | .................... | .................... | .................... |
| 10101 | MW153816.1 | Severe acute respiratory syndrome coro | .................... | .................... | ..................... | .................... | .................... | .................... |
| 10102 | MW153817.1 | Severe acute respiratory syndrome coro | .................... | .................... | ..................... | .................... | .................... | .................... |
| 10103 | MW153818.1 | Severe acute respiratory syndrome coro | .................... | .................... | ..................... | .................... | .................... | .................... |
| 10104 | MW153819.1 | Severe acute respiratory syndrome coro | .................... | .................... | ..................... | .................... | .................... | .................... |
| 10105 | MW153820.1 | Severe acute respiratory syndrome coro | .................... | .................... | ..................... | .................... | .................... | .................... |
| 10106 | MW153821.1 | Severe acute respiratory syndrome coro | .................... | .................... | ..................... | .................... | .................... | .................... |
| 10107 | MW153822.1 | Severe acute respiratory syndrome coro | .................... | .................... | ..................... | .................... | .................... | .................... |
| 10108 | MW153823.1 | Severe acute respiratory syndrome coro | .................... | .................... | ..................... | .................... | .................... | .................... |
| 10109 | MW153824.1 | Severe acute respiratory syndrome coro | .................... | .................... | ..................... | .................... | .................... | .................... |
| 10110 | MW153825.1 | Severe acute respiratory syndrome coro | .................... | .................... | ..................... | .................... | .................... | .................... |
| 10111 | MW153826.1 | Severe acute respiratory syndrome coro | .................... | .................... | ..................... | .................... | .................... | .................... |
| 10112 | MW153827.1 | Severe acute respiratory syndrome coro | .................... | .................... | ..................... | .................... | .................... | .................... |
| 10113 | MW153828.1 | Severe acute respiratory syndrome coro | .................... | .................... | ..................... | .................... | .................... | .................... |
| 10114 | MW153829.1 | Severe acute respiratory syndrome coro | .................... | .................... | ..................... | .................... | .................... | .................... |
| 10115 | MW153830.1 | Severe acute respiratory syndrome coro | .................... | .................... | ..................... | .................... | .................... | .................... |
| 10116 | MW153831.1 | Severe acute respiratory syndrome coro | .................... | .................... | ..................... | .................... | .................... | .................... |
| 10117 | MW153832.1 | Severe acute respiratory syndrome coro | .................... | .................... | ..................... | .................... | .................... | .................... |
| 10118 | MW153833.1 | Severe acute respiratory syndrome coro | .................... | .................... | ..................... | .................... | .................... | .................... |
| 10119 | MW153834.1 | Severe acute respiratory syndrome coro | .................... | .................... | ..................... | .................... | .................... | .................... |
| 10120 | MW153835.1 | Severe acute respiratory syndrome coro | .................... | .................... | ..................... | .................... | .................... | .................... |
| 10121 | MW153836.1 | Severe acute respiratory syndrome coro | .................... | .................... | ..................... | .................... | .................... | .................... |
| 10122 | MW153837.1 | Severe acute respiratory syndrome coro | .................... | .................... | ..................... | .................... | .................... | .................... |
| 10123 | MW153838.1 | Severe acute respiratory syndrome coro | .................... | .................... | ..................... | .................... | .................... | .................... |
| 10124 | MW153839.1 | Severe acute respiratory syndrome coro | .................... | .................... | ..................... | .................... | .................... | .................... |
| 10125 | MW153840.1 | Severe acute respiratory syndrome coro | .................... | .................... | ..................... | .................... | .................... | .................... |
| 10126 | MW153841.1 | Severe acute respiratory syndrome coro | .................... | .................... | ..................... | .................... | .................... | .................... |
| 10127 | MW153842.1 | Severe acute respiratory syndrome coro | .................... | .................... | ..................... | .................... | .................... | .................... |
| 10128 | MW153843.1 | Severe acute respiratory syndrome coro | .................... | .................... | ..................... | .................... | .................... | .................... |
| 10129 | MW153845.1 | Severe acute respiratory syndrome coro | .................... | .................... | ..................... | .................... | .................... | .................... |
| 10130 | MW153846.1 | Severe acute respiratory syndrome coro | .................... | .................... | ..................... | .................... | .................... | .................... |
| 10131 | MW153847.1 | Severe acute respiratory syndrome coro | .................... | .................... | ..................... | .................... | .................... | .................... |
| 10132 | MW153848.1 | Severe acute respiratory syndrome coro | .................... | .................... | ..................... | .................... | .................... | .................... |
| 10133 | MW153849.1 | Severe acute respiratory syndrome coro | .................... | .................... | ..................... | .................... | .................... | .................... |
| 10134 | MW153850.1 | Severe acute respiratory syndrome coro | .................... | .................... | ..................... | .................... | .................... | .................... |
| 10135 | MW153851.1 | Severe acute respiratory syndrome coro | .................... | .................... | ..................... | .................... | .................... | .................... |
| 10136 | MW153853.1 | Severe acute respiratory syndrome coro | .................... | .................... | ..................... | .................... | .................... | .................... |
| 10137 | MW153854.1 | Severe acute respiratory syndrome coro | .................... | .................... | ..................... | .................... | .................... | .................... |
| 10138 | MW153856.1 | Severe acute respiratory syndrome coro | .................... | .................... | ..................... | .................... | .................... | .................... |
| 10139 | MW153858.1 | Severe acute respiratory syndrome coro | .................... | .................... | ..................... | .................... | .................... | .................... |
| 10140 | MW153859.1 | Severe acute respiratory syndrome coro | .................... | .................... | ..................... | .................... | .................... | .................... |
| 10141 | MW153860.1 | Severe acute respiratory syndrome coro | .................... | .................... | ..................... | .................... | .................... | .................... |
| 10142 | MW153861.1 | Severe acute respiratory syndrome coro | .................... | .................... | ..................... | .................... | .................... | .................... |
| 10143 | MW153862.1 | Severe acute respiratory syndrome coro | .................... | .................... | ..................... | .................... | .................... | .................... |
| 10144 | MW153863.1 | Severe acute respiratory syndrome coro | .................... | .................... | ..................... | .................... | .................... | .................... |
| 10145 | MW153864.1 | Severe acute respiratory syndrome coro | .................... | .................... | ..................... | .................... | .................... | .................... |
| 10146 | MW153865.1 | Severe acute respiratory syndrome coro | .................... | .................... | ..................... | .................... | .................... | .................... |
| 10147 | MW153866.1 | Severe acute respiratory syndrome coro | .................... | .................... | ..................... | .................... | .................... | .................... |
| 10148 | MW153867.1 | Severe acute respiratory syndrome coro | .................... | .................... | ..................... | .................... | .................... | .................... |
| 10149 | MW153868.1 | Severe acute respiratory syndrome coro | .................... | .................... | ..................... | .................... | .................... | .................... |
| 10150 | MW153869.1 | Severe acute respiratory syndrome coro | .................... | .................... | ..................... | .................... | .................... | .................... |
| 10151 | MW153870.1 | Severe acute respiratory syndrome coro | .................... | .................... | ..................... | .................... | .................... | .................... |
| 10152 | MW153871.1 | Severe acute respiratory syndrome coro | .................... | .................... | ..................... | .................... | .................... | .................... |
| 10153 | MW153872.1 | Severe acute respiratory syndrome coro | .................... | .................... | ..................... | .................... | .................... | .................... |
| 10154 | MW153873.1 | Severe acute respiratory syndrome coro | .................... | .................... | ..................... | .................... | .................... | .................... |

```
10155 MW153875.1 |Severe acute respiratory syndrome coro    ....................  ....................  ....................  ....................  ....................  ....................
10156 MW153876.1 |Severe acute respiratory syndrome coro    ....................  ....................  ....................  ....................  ....................  ....................
10157 MW153877.1 |Severe acute respiratory syndrome coro    ....................  ....................  ....................  ....................  ....................  ....................
10158 MW153878.1 |Severe acute respiratory syndrome coro    ....................  ....................  ....................  ....................  ....................  ....................
10159 MW153879.1 |Severe acute respiratory syndrome coro    ....................  ....................  ....................  ....................  ....................  ....................
10160 MW153880.1 |Severe acute respiratory syndrome coro    ....................  ....................  ....................  ....................  ....................  ....................
10161 MW153881.1 |Severe acute respiratory syndrome coro    ....................  ....................  ....................  ....................  ....................  ....................
10162 MW153882.1 |Severe acute respiratory syndrome coro    ....................  ....................  ....................  ....................  ....................  ....................
10163 MW153883.1 |Severe acute respiratory syndrome coro    ....................  ....................  ....................  ....................  ....................  ....................
10164 MW153884.1 |Severe acute respiratory syndrome coro    ....................  ....................  ....................  ....................  ....................  ....................
10165 MW153885.1 |Severe acute respiratory syndrome coro    ....................  ....................  ....................  ....................  ....................  ....................
10166 MW153888.1 |Severe acute respiratory syndrome coro    ....................  ....................  ....................  ....................  ....................  ....................
10167 MW153889.1 |Severe acute respiratory syndrome coro    ....................  ....................  ....................  ....................  ....................  ....................
10168 MW153891.1 |Severe acute respiratory syndrome coro    ....................  ....................  ....................  ....................  ....................  ....................
10169 MW153893.1 |Severe acute respiratory syndrome coro    ....................  ....................  ....................  ....................  ....................  ....................
10170 MW153896.1 |Severe acute respiratory syndrome coro    ....................  ....................  ....................  ....................  ....................  ....................
10171 MW153897.1 |Severe acute respiratory syndrome coro    ....................  ....................  ....................  ....................  ....................  ....................
10172 MW153898.1 |Severe acute respiratory syndrome coro    ....................  ....................  ....................  ....................  ....................  ....................
10173 MW153899.1 |Severe acute respiratory syndrome coro    ....................  ....................  ....................  ....................  ....................  ....................
10174 MW153900.1 |Severe acute respiratory syndrome coro    ....................  ....................  ....................  ....................  ....................  ....................
10175 MW153901.1 |Severe acute respiratory syndrome coro    ....................  ....................  ....................  ....................  ....................  ....................
10176 MW153902.1 |Severe acute respiratory syndrome coro    ....................  ....................  ....................  ....................  ....................  ....................
10177 MW153903.1 |Severe acute respiratory syndrome coro    ....................  ....................  ....................  ....................  ....................  ....................
10178 MW153905.1 |Severe acute respiratory syndrome coro    ....................  ....................  ....................  ....................  ....................  ....................
10179 MW153906.1 |Severe acute respiratory syndrome coro    ....................  ....................  ....................  ....................  ....................  ....................
10180 MW153907.1 |Severe acute respiratory syndrome coro    ....................  ....................  ....................  ....................  ....................  ....................
10181 MW153908.1 |Severe acute respiratory syndrome coro    ....................  ....................  ....................  ....................  ....................  ....................
10182 MW153909.1 |Severe acute respiratory syndrome coro    ....................  ....................  ....................  ....................  ....................  ....................
10183 MW153910.1 |Severe acute respiratory syndrome coro    ....................  ....................  ....................  ....................  ....................  ....................
10184 MW153911.1 |Severe acute respiratory syndrome coro    ....................  ....................  ....................  ....................  ....................  ....................
10185 MW153912.1 |Severe acute respiratory syndrome coro    ....................  ....................  ....................  ....................  ....................  ....................
10186 MW153913.1 |Severe acute respiratory syndrome coro    ....................  ....................  ....................  ....................  ....................  ....................
10187 MW153914.1 |Severe acute respiratory syndrome coro    ....................  ....................  ....................  ....................  ....................  ....................
10188 MW153915.1 |Severe acute respiratory syndrome coro    ....................  ....................  ....................  ....................  ....................  ....................
10189 MW153916.1 |Severe acute respiratory syndrome coro    ....................  ....................  ....................  ....................  ....................  ....................
10190 MW153917.1 |Severe acute respiratory syndrome coro    ....................  ....................  ....................  ....................  ....................  ....................
10191 MW153918.1 |Severe acute respiratory syndrome coro    ....................  ....................  ....................  ....................  ....................  ....................
10192 MW153919.1 |Severe acute respiratory syndrome coro    ....................  ....................  ....................  ....................  ....................  ....................
10193 MW153920.1 |Severe acute respiratory syndrome coro    ....................  ....................  ....................  ....................  ....................  ....................
10194 MW153921.1 |Severe acute respiratory syndrome coro    ....................  ....................  ....................  ....................  ....................  ....................
10195 MW153922.1 |Severe acute respiratory syndrome coro    ....................  ....................  ....................  ....................  ....................  ....................
10196 MW153923.1 |Severe acute respiratory syndrome coro    ....................  ....................  ....................  ....................  ....................  ....................
10197 MW153925.1 |Severe acute respiratory syndrome coro    ....................  ....................  ....................  ....................  ....................  ....................
10198 MW153926.1 |Severe acute respiratory syndrome coro    ....................  ....................  ....................  ....................  ....................  ....................
10199 MW153927.1 |Severe acute respiratory syndrome coro    ....................  ....................  ....................  ....................  ....................  ....................
10200 MW153928.1 |Severe acute respiratory syndrome coro    ....................  ....................  ....................  ....................  ....................  ....................
10201 MW153929.1 |Severe acute respiratory syndrome coro    ....................  ....................  ....................  ....................  ....................  ....................
10202 MW153931.1 |Severe acute respiratory syndrome coro    ....................  ....................  ....................  ....................  ....................  ....................
10203 MW153932.1 |Severe acute respiratory syndrome coro    ....................  ....................  ....................  ....................  ....................  ....................
10204 MW153933.1 |Severe acute respiratory syndrome coro    ....................  ....................  ....................  ....................  ....................  ....................
10205 MW153934.1 |Severe acute respiratory syndrome coro    ....................  ......A.............  ....................  ....................  ....................  ....................
10206 MW153935.1 |Severe acute respiratory syndrome coro    ....................  ......A.............  ....................  ....................  ....................  ....................
10207 MW153936.1 |Severe acute respiratory syndrome coro    ....................  ....................  ....................  ....................  ....................  ....................
10208 MW153937.1 |Severe acute respiratory syndrome coro    ....................  ....................  ....................  ....................  ....................  ....................
10209 MW153938.1 |Severe acute respiratory syndrome coro    ....................  ....................  ....................  ....................  ....................  ....................
10210 MW153939.1 |Severe acute respiratory syndrome coro    ....................  ......A.............  ....................  ....................  ....................  ....................
10211 MW153940.1 |Severe acute respiratory syndrome coro    ....................  ......A.............  ....................  ....................  ....................  ....................
10212 MW153941.1 |Severe acute respiratory syndrome coro    ....................  ....................  ....................  ....................  ....................  ....................
10213 MW153942.1 |Severe acute respiratory syndrome coro    ....................  ....................  ....................  ....................  ....................  ....................
10214 MW153943.1 |Severe acute respiratory syndrome coro    ....................  ....................  ....................  ....................  ....................  ....................
10215 MW153944.1 |Severe acute respiratory syndrome coro    ....................  ....................  ....................  ....................  ....................  ....................
10216 MW153945.1 |Severe acute respiratory syndrome coro    ....................  ......A.............  ....................  ....................  ....................  ....................
10217 MW153946.1 |Severe acute respiratory syndrome coro    ....................  ....................  ....................  ....................  ....................  ....................
10218 MW153947.1 |Severe acute respiratory syndrome coro    ....................  ....................  ....................  ....................  ....................  ....................
10219 MW153948.1 |Severe acute respiratory syndrome coro    ....................  ....................  ....................  ....................  ....................  ....................
10220 MW153949.1 |Severe acute respiratory syndrome coro    ....................  ....................  ....................  ....................  ....................  ....................
10221 MW153950.1 |Severe acute respiratory syndrome coro    ....................  ....................  ....................  ....................  ....................  ....................
10222 MW153951.1 |Severe acute respiratory syndrome coro    ....................  ....................  ....................  ....................  ....................  ....................
10223 MW153952.1 |Severe acute respiratory syndrome coro    ....................  ....................  ....................  ....................  ....................  ....................
10224 MW153953.1 |Severe acute respiratory syndrome coro    ....................  ....................  ....................  ....................  ....................  ....................
10225 MW153954.1 |Severe acute respiratory syndrome coro    ....................  ....................  ....................  ....................  ....................  ....................
10226 MW153955.1 |Severe acute respiratory syndrome coro    ....................  ....................  ....................  ....................  ....................  ....................
10227 MW153956.1 |Severe acute respiratory syndrome coro    ....................  ....................  ....................  ....................  ....................  ....................
10228 MW153957.1 |Severe acute respiratory syndrome coro    ....................  ....................  ....................  ....................  ....................  ....................
10229 MW153958.1 |Severe acute respiratory syndrome coro    ....................  ....................  ....................  ....................  ....................  ....................
10230 MW153959.1 |Severe acute respiratory syndrome coro    ....................  ....................  ....................  ....................  ....................  ....................
10231 MW153960.1 |Severe acute respiratory syndrome coro    ....................  ....................  ....................  ....................  ....................  ....................
10232 MW153961.1 |Severe acute respiratory syndrome coro    ....................  ....................  ....................  ....................  ....................  ....................
```

```
10233 MW153962.1 |Severe acute respiratory syndrome coro    .................... .................... .................... .................... .................... ....................
10234 MW153963.1 |Severe acute respiratory syndrome coro    .................... .................... .................... .................... .................... ....................
10235 MW153964.1 |Severe acute respiratory syndrome coro    .................... .................... .................... .................... .................... ....................
10236 MW153965.1 |Severe acute respiratory syndrome coro    .................... .................... .................... .................... .................... ....................
10237 MW153966.1 |Severe acute respiratory syndrome coro    .................... .................... .................... .................... .................... ....................
10238 MW153967.1 |Severe acute respiratory syndrome coro    .................... .................... .................... .................... .................... ....................
10239 MW153968.1 |Severe acute respiratory syndrome coro    .................... .................... .................... .................... .................... ....................
10240 MW153969.1 |Severe acute respiratory syndrome coro    .................... .................... .................... .................... .................... ....................
10241 MW153970.1 |Severe acute respiratory syndrome coro    .................... .................... .................... .................... .................... ....................
10242 MW153971.1 |Severe acute respiratory syndrome coro    .................... .................... .................... .................... .................... ....................
10243 MW153972.1 |Severe acute respiratory syndrome coro    .................... .................... .................... .................... .................... ....................
10244 MW153973.1 |Severe acute respiratory syndrome coro    .................... .................... .................... .................... .................... ....................
10245 MW153974.1 |Severe acute respiratory syndrome coro    .................... .................... .................... .................... .................... ....................
10246 MW153975.1 |Severe acute respiratory syndrome coro    .................... .................... .................... .................... .................... ....................
10247 MW153976.1 |Severe acute respiratory syndrome coro    .................... .................... .................... .................... .................... ....................
10248 MW153977.1 |Severe acute respiratory syndrome coro    .................... .................... .................... .................... .................... ....................
10249 MW153978.1 |Severe acute respiratory syndrome coro    .................... .................... .................... .................... .................... ....................
10250 MW153979.1 |Severe acute respiratory syndrome coro    .................... .................... .................... .................... .................... ....................
10251 MW153980.1 |Severe acute respiratory syndrome coro    .................... .................... .................... .................... .................... ....................
10252 MW153981.1 |Severe acute respiratory syndrome coro    .................... .................... .................... .................... .................... ....................
10253 MW153982.1 |Severe acute respiratory syndrome coro    .................... .................... .................... .................... .................... ....................
10254 MW153983.1 |Severe acute respiratory syndrome coro    .................... .................... .................... .................... .................... ....................
10255 MW153984.1 |Severe acute respiratory syndrome coro    .................... .................... .................... .................... .................... ....................
10256 MW153985.1 |Severe acute respiratory syndrome coro    .................... .................... .................... .................... .................... ....................
10257 MW153986.1 |Severe acute respiratory syndrome coro    .................... .................... .................... .................... .................... ....................
10258 MW153987.1 |Severe acute respiratory syndrome coro    .................... .................... .................... .................... .................... ....................
10259 MW153988.1 |Severe acute respiratory syndrome coro    .................... .................... .................... .................... .................... ....................
10260 MW153989.1 |Severe acute respiratory syndrome coro    .................... .................... .................... .................... .................... ....................
10261 MW153990.1 |Severe acute respiratory syndrome coro    .................... .................... .................... .................... .................... ....................
10262 MW153992.1 |Severe acute respiratory syndrome coro    .................... .................... .................... .................... .................... ....................
10263 MW153993.1 |Severe acute respiratory syndrome coro    .................... .................... .................... .................... .................... ....................
10264 MW153994.1 |Severe acute respiratory syndrome coro    .................... .................... .................... .................... .................... ....................
10265 MW153995.1 |Severe acute respiratory syndrome coro    .................... .................... .................... .................... .................... ....................
10266 MW153996.1 |Severe acute respiratory syndrome coro    .................... .................... .................... .................... .................... ....................
10267 MW153997.1 |Severe acute respiratory syndrome coro    .................... .................... .................... .................... .................... ....................
10268 MW153998.1 |Severe acute respiratory syndrome coro    .................... .................... .................... .................... .................... ....................
10269 MW153999.1 |Severe acute respiratory syndrome coro    .................... .................... .................... .................... .................... ....................
10270 MW154000.1 |Severe acute respiratory syndrome coro    .................... .................... .................... .................... .................... ....................
10271 MW154001.1 |Severe acute respiratory syndrome coro    .................... .................... .................... .................... .................... ....................
10272 MW154002.1 |Severe acute respiratory syndrome coro    .................... .................... .................... .................... .................... ....................
10273 MW154003.1 |Severe acute respiratory syndrome coro    .................... .................... .................... .................... .................... ....................
10274 MW154005.1 |Severe acute respiratory syndrome coro    .................... .................... .................... .................... .................... ....................
10275 MW154006.1 |Severe acute respiratory syndrome coro    .................... .................... .................... .................... .................... ....................
10276 MW154007.1 |Severe acute respiratory syndrome coro    .................... .................... .................... .................... .................... ....................
10277 MW154008.1 |Severe acute respiratory syndrome coro    .................... .................... .................... .................... .................... ....................
10278 MW154010.1 |Severe acute respiratory syndrome coro    .................... .................... .................... .................... .................... ....................
10279 MW154011.1 |Severe acute respiratory syndrome coro    .................... .................... .................... .................... .................... ....................
10280 MW154012.1 |Severe acute respiratory syndrome coro    .................... .................... .................... .................... .................... ....................
10281 MW154013.1 |Severe acute respiratory syndrome coro    .................... .................... .................... .................... .................... ....................
10282 MW154014.1 |Severe acute respiratory syndrome coro    .................... .................... .................... .................... .................... ....................
10283 MW154015.1 |Severe acute respiratory syndrome coro    .................... .................... .................... .................... .................... ....................
10284 MW154016.1 |Severe acute respiratory syndrome coro    .................... .................... .................... .................... .................... ....................
10285 MW154017.1 |Severe acute respiratory syndrome coro    .................... .................... .................... .................... .................... ....................
10286 MW154018.1 |Severe acute respiratory syndrome coro    .................... .................... .................... .................... .................... ....................
10287 MW154019.1 |Severe acute respiratory syndrome coro    .................... .................... .................... .................... .................... ....................
10288 MW154020.1 |Severe acute respiratory syndrome coro    .................... .................... .................... .................... .................... ....................
10289 MW154021.1 |Severe acute respiratory syndrome coro    .................... .................... .................... .................... .................... ....................
10290 MW154022.1 |Severe acute respiratory syndrome coro    .................... .................... .................... .................... .................... ....................
10291 MW154023.1 |Severe acute respiratory syndrome coro    .................... .................... .................... .................... .................... ....................
10292 MW154024.1 |Severe acute respiratory syndrome coro    .................... .................... .................... .................... .................... ....................
10293 MW154025.1 |Severe acute respiratory syndrome coro    .................... .................... .................... .................... .................... ....................
10294 MW154026.1 |Severe acute respiratory syndrome coro    .................... .................... .................... .................... .................... ....................
10295 MW154027.1 |Severe acute respiratory syndrome coro    .................... .................... .................... .................... .................... ....................
10296 MW154028.1 |Severe acute respiratory syndrome coro    .................... .................... .................... .................... .................... ....................
10297 MW154029.1 |Severe acute respiratory syndrome coro    .................... .................... .................... .................... .................... ....................
10298 MW154030.1 |Severe acute respiratory syndrome coro    .................... .................... .................... .................... .................... ....................
10299 MW154031.1 |Severe acute respiratory syndrome coro    .................... .................... .................... .................... .................... ....................
10300 MW154032.1 |Severe acute respiratory syndrome coro    .................... .................... .................... .................... .................... ....................
10301 MW154033.1 |Severe acute respiratory syndrome coro    .................... .................... .................... .................... .................... ....................
10302 MW154034.1 |Severe acute respiratory syndrome coro    .................... .................... .................... .................... .................... ....................
10303 MW154035.1 |Severe acute respiratory syndrome coro    .................... .................... .................... .................... .................... ....................
10304 MW154036.1 |Severe acute respiratory syndrome coro    .................... .................... .................... .................... .................... ....................
10305 MW154037.1 |Severe acute respiratory syndrome coro    .................... .................... .................... .................... .................... ....................
10306 MW154038.1 |Severe acute respiratory syndrome coro    .................... .................... .................... .................... .................... ....................
10307 MW154039.1 |Severe acute respiratory syndrome coro    .................... .................... .................... .................... .................... ....................
10308 MW154040.1 |Severe acute respiratory syndrome coro    .................... .................... .................... .................... .................... ....................
10309 MW154042.1 |Severe acute respiratory syndrome coro    .................... .................... .................... .................... .................... ....................
10310 MW154043.1 |Severe acute respiratory syndrome coro    .................... .................... .................... .................... .................... ....................
```

```
10311 MW154044.1 |Severe acute respiratory syndrome coro    ....................  ....................  ....................  ....................  ....................  ....................
10312 MW154045.1 |Severe acute respiratory syndrome coro    ....................  ....................  ....................  ....................  ....................  ....................
10313 MW154046.1 |Severe acute respiratory syndrome coro    ....................  ....................  ....................  ....................  ....................  ....................
10314 MW154047.1 |Severe acute respiratory syndrome coro    ....................  ....................  ....................  ....................  ....................  ....................
10315 MW154048.1 |Severe acute respiratory syndrome coro    ....................  ....................  ....................  ....................  ....................  ....................
10316 MW154049.1 |Severe acute respiratory syndrome coro    ....................  ....................  ....................  ....................  ....................  ....................
10317 MW154050.1 |Severe acute respiratory syndrome coro    ....................  ....................  ....................  ....................  ....................  ....................
10318 MW154051.1 |Severe acute respiratory syndrome coro    ....................  ....................  ....................  ....................  ....................  ....................
10319 MW154052.1 |Severe acute respiratory syndrome coro    ....................  ....................  ....................  ....................  ....................  ....................
10320 MW154053.1 |Severe acute respiratory syndrome coro    ....................  ....................  ....................  ....................  ....................  ....................
10321 MW154054.1 |Severe acute respiratory syndrome coro    ....................  ....................  ....................  ....................  ....................  ....................
10322 MW154055.1 |Severe acute respiratory syndrome coro    ....................  ....................  ....................  ....................  ....................  ....................
10323 MW154056.1 |Severe acute respiratory syndrome coro    ....................  ....................  ....................  ....................  ....................  ....................
10324 MW154057.1 |Severe acute respiratory syndrome coro    ....................  ....................  ....................  ....................  ....................  ....................
10325 MW154058.1 |Severe acute respiratory syndrome coro    ....................  ....................  ....................  ....................  ....................  ....................
10326 MW154059.1 |Severe acute respiratory syndrome coro    ....................  ....................  ....................  ....................  ....................  ....................
10327 MW154060.1 |Severe acute respiratory syndrome coro    ....................  ....................  ....................  ....................  ....................  ....................
10328 MW154061.1 |Severe acute respiratory syndrome coro    ....................  ....................  ....................  ....................  ....................  ....................
10329 MW154062.1 |Severe acute respiratory syndrome coro    ....................  ....................  ....................  ....................  ....................  ....................
10330 MW154063.1 |Severe acute respiratory syndrome coro    ....................  ....................  ....................  ....................  ....................  ....................
10331 MW154064.1 |Severe acute respiratory syndrome coro    ....................  ....................  ....................  ....................  ....................  ....................
10332 MW154065.1 |Severe acute respiratory syndrome coro    ....................  ....................  ....................  ....................  ....................  ....................
10333 MW154066.1 |Severe acute respiratory syndrome coro    ....................  ....................  ....................  ....................  ....................  ....................
10334 MW154067.1 |Severe acute respiratory syndrome coro    ....................  ....................  ....................  ....................  ....................  ....................
10335 MW154068.1 |Severe acute respiratory syndrome coro    ....................  ....................  ....................  ....................  ....................  ....................
10336 MW154069.1 |Severe acute respiratory syndrome coro    ....................  ....................  ....................  ....................  ....................  ....................
10337 MW154070.1 |Severe acute respiratory syndrome coro    ....................  ....................  ....................  ....................  ....................  ....................
10338 MW154071.1 |Severe acute respiratory syndrome coro    ....................  ....................  ....................  ....................  ....................  ....................
10339 MW154072.1 |Severe acute respiratory syndrome coro    ....................  ....................  ....................  ....................  ....................  ....................
10340 MW154073.1 |Severe acute respiratory syndrome coro    ....................  ....................  ....................  ....................  ....................  ....................
10341 MW154074.1 |Severe acute respiratory syndrome coro    ....................  ....................  ....................  ....................  ....................  ....................
10342 MW154075.1 |Severe acute respiratory syndrome coro    ....................  ....................  ....................  ....................  ....................  ....................
10343 MW154076.1 |Severe acute respiratory syndrome coro    ....................  ....................  ....................  ....................  ....................  ....................
10344 MW154077.1 |Severe acute respiratory syndrome coro    ....................  ....................  ....................  ....................  ....................  ....................
10345 MW154078.1 |Severe acute respiratory syndrome coro    ....................  ....................  ....................  ....................  ....................  ....................
10346 MW154079.1 |Severe acute respiratory syndrome coro    ....................  ....................  ....................  ....................  ....................  ....................
10347 MW154080.1 |Severe acute respiratory syndrome coro    ....................  ....................  ....................  ....................  ....................  ....................
10348 MW154081.1 |Severe acute respiratory syndrome coro    ....................  ....................  ....................  ....................  ....................  ....................
10349 MW154082.1 |Severe acute respiratory syndrome coro    ....................  ....................  ....................  ....................  ....................  ....................
10350 MW154083.1 |Severe acute respiratory syndrome coro    ....................  ....................  ....................  ....................  ....................  ....................
10351 MW154084.1 |Severe acute respiratory syndrome coro    ....................  ....................  ....................  ....................  ....................  ....................
10352 MW154085.1 |Severe acute respiratory syndrome coro    ....................  ....................  ....................  ....................  ....................  ....................
10353 MW154086.1 |Severe acute respiratory syndrome coro    ....................  ....................  ....................  ....................  ....................  ....................
10354 MW154087.1 |Severe acute respiratory syndrome coro    ....................  ....................  ....................  ....................  ....................  ....................
10355 MW154088.1 |Severe acute respiratory syndrome coro    ....................  ....................  ....................  ....................  ....................  ....................
10356 MW154089.1 |Severe acute respiratory syndrome coro    ....................  ....................  ....................  ....................  ....................  ....................
10357 MW154090.1 |Severe acute respiratory syndrome coro    ....................  ....................  ....................  ....................  ....................  ....................
10358 MW154095.1 |Severe acute respiratory syndrome coro    ....................  ....................  ....................  ....................  ....................  ....................
10359 MW154096.1 |Severe acute respiratory syndrome coro    ....................  ....................  ....................  ....................  ....................  ....................
10360 MW154098.1 |Severe acute respiratory syndrome coro    ....................  ....................  ....................  ....................  ....................  ....................
10361 MW154099.1 |Severe acute respiratory syndrome coro    ....................  ....................  ....................  ....................  ....................  ....................
10362 MW154100.1 |Severe acute respiratory syndrome coro    ....................  ....................  ....................  ....................  ....................  ....................
10363 MW154101.1 |Severe acute respiratory syndrome coro    ....................  ....................  ....................  ....................  ....................  ....................
10364 MW154103.1 |Severe acute respiratory syndrome coro    ....................  ....................  ....................  ....................  ....................  ....................
10365 MW154104.1 |Severe acute respiratory syndrome coro    ....................  ....................  ....................  ....................  ....................  ....................
10366 MW154105.1 |Severe acute respiratory syndrome coro    ....................  ....................  ....................  ....................  ....................  ....................
10367 MW154106.1 |Severe acute respiratory syndrome coro    ....................  ....................  ....................  ....................  ....................  ....................
10368 MW154107.1 |Severe acute respiratory syndrome coro    ....................  ....................  ....................  ....................  ....................  ....................
10369 MW154108.1 |Severe acute respiratory syndrome coro    ....................  ....................  ....................  ....................  ....................  ....................
10370 MW154109.1 |Severe acute respiratory syndrome coro    ....................  ....................  ....................  ....................  ....................  ....................
10371 MW154110.1 |Severe acute respiratory syndrome coro    ....................  ....................  ....................  ....................  ....................  ....................
10372 MW154111.1 |Severe acute respiratory syndrome coro    ....................  ....................  ....................  ....................  ....................  ....................
10373 MW154112.1 |Severe acute respiratory syndrome coro    ....................  ....................  ....................  ....................  ....................  ....................
10374 MW154113.1 |Severe acute respiratory syndrome coro    ....................  ....................  ....................  ....................  ....................  ....................
10375 MW154115.1 |Severe acute respiratory syndrome coro    ....................  ....................  ....................  ....................  ....................  ....................
10376 MW154116.1 |Severe acute respiratory syndrome coro    ....................  ....................  ....................  ....................  ....................  ....................
10377 MW154117.1 |Severe acute respiratory syndrome coro    ....................  ....................  ....................  ....................  ....................  ....................
10378 MW154119.1 |Severe acute respiratory syndrome coro    ....................  ....................  ....................  ....................  ....................  ....................
10379 MW154120.1 |Severe acute respiratory syndrome coro    ....................  ....................  ....................  ....................  ....................  ....................
10380 MW154121.1 |Severe acute respiratory syndrome coro    ....................  ....................  ....................  ....................  ....................  ....................
10381 MW154122.1 |Severe acute respiratory syndrome coro    ....................  ....................  ....................  ....................  ....................  ....................
10382 MW154123.1 |Severe acute respiratory syndrome coro    ....................  ....................  ....................  ....................  ....................  ....................
10383 MW154124.1 |Severe acute respiratory syndrome coro    ....................  ....................  ....................  ....................  ....................  ....................
10384 MW154125.1 |Severe acute respiratory syndrome coro    ....................  ....................  ....................  ....................  ....................  ....................
10385 MW154126.1 |Severe acute respiratory syndrome coro    ....................  ....................  ....................  ....................  ....................  ....................
10386 MW154127.1 |Severe acute respiratory syndrome coro    ....................  ....................  ....................  ....................  ....................  ....................
10388 MW154130.1 |Severe acute respiratory syndrome coro    ....................  ....................  ....................  ....................  ....................  ....................
10389 MW154131.1 |Severe acute respiratory syndrome coro    ....................  ....................  ....................  ....................  ....................  ....................
```

10390 MW154132.1 |Severe acute respiratory syndrome coro .................... .................... .................... .................... .................... ....................
10391 MW154133.1 |Severe acute respiratory syndrome coro .................... .................... .................... .................... .................... ....................
10392 MW154134.1 |Severe acute respiratory syndrome coro .................... .................... .................... .................... .................... ....................
10393 MW154135.1 |Severe acute respiratory syndrome coro .................... .................... .................... .................... .................... ....................
10394 MW154136.1 |Severe acute respiratory syndrome coro .................... .................... .................... .................... .................... ....................
10395 MW154137.1 |Severe acute respiratory syndrome coro .................... .................... .................... .................... .................... ....................
10396 MW154138.1 |Severe acute respiratory syndrome coro .................... .................... .................... .................... .................... ....................
10397 MW154139.1 |Severe acute respiratory syndrome coro .................... .................... .................... .................... .................... ....................
10398 MW154140.1 |Severe acute respiratory syndrome coro .................... .................... .................... .................... .................... ....................
10399 MW154142.1 |Severe acute respiratory syndrome coro .................... .................... .................... .................... .................... ....................
10400 MW154143.1 |Severe acute respiratory syndrome coro .................... .................... .................... .................... .................... ....................
10401 MW154146.1 |Severe acute respiratory syndrome coro .................... .................... .................... .................... .................... ....................
10402 MW154147.1 |Severe acute respiratory syndrome coro .................... .................... .................... .................... .................... ....................
10403 MW154148.1 |Severe acute respiratory syndrome coro .................... .................... .................... .................... .................... ....................
10404 MW154149.1 |Severe acute respiratory syndrome coro .................... .................... .................... .................... .................... ....................
10405 MW154151.1 |Severe acute respiratory syndrome coro .................... .................... .................... .................... .................... ....................
10406 MW154152.1 |Severe acute respiratory syndrome coro .................... .................... .................... .................... .................... ....................
10407 MW154153.1 |Severe acute respiratory syndrome coro .................... .................... .................... .................... .................... ....................
10408 MW154154.1 |Severe acute respiratory syndrome coro .................... .................... .................... .................... .................... ....................
10409 MW154155.1 |Severe acute respiratory syndrome coro .................... .................... .................... .................... .................... ....................
10410 MW154156.1 |Severe acute respiratory syndrome coro .................... .................... .................... .................... .................... ....................
10411 MW154157.1 |Severe acute respiratory syndrome coro .................... .................... .................... .................... .................... ....................
10412 MW154158.1 |Severe acute respiratory syndrome coro .................... .................... .................... .................... .................... ....................
10413 MW154160.1 |Severe acute respiratory syndrome coro .................... .................... .................... .................... .................... ....................
10414 MW154161.1 |Severe acute respiratory syndrome coro .................... .................... .................... .................... .................... ....................
10415 MW154162.1 |Severe acute respiratory syndrome coro .................... .................... .................... .................... .................... ....................
10416 MW154163.1 |Severe acute respiratory syndrome coro .................... .................... .................... .................... .................... ....................
10417 MW154164.1 |Severe acute respiratory syndrome coro .................... .................... .................... .................... .................... ....................
10418 MW154165.1 |Severe acute respiratory syndrome coro .................... .................... .................... .................... .................... ....................
10419 MW154166.1 |Severe acute respiratory syndrome coro .................... .................... .................... .................... .................... ....................
10420 MW154168.1 |Severe acute respiratory syndrome coro .................... .................... .................... .................... .................... ....................
10421 MW154169.1 |Severe acute respiratory syndrome coro .................... .................... .................... .................... .................... ....................
10422 MW154171.1 |Severe acute respiratory syndrome coro .................... .................... .................... .................... .................... ....................
10423 MW154172.1 |Severe acute respiratory syndrome coro .................... .................... .................... .................... .................... ....................
10424 MW154173.1 |Severe acute respiratory syndrome coro .................... .................... .................... .................... .................... ....................
10425 MW154174.1 |Severe acute respiratory syndrome coro .................... .................... .................... .................... .................... ....................
10426 MW154177.1 |Severe acute respiratory syndrome coro .................... .................... .................... .................... .................... ....................
10427 MW154178.1 |Severe acute respiratory syndrome coro .................... .................... .................... .................... .................... ....................
10428 MW154179.1 |Severe acute respiratory syndrome coro .................... .................... .................... .................... .................... ....................
10429 MW154181.1 |Severe acute respiratory syndrome coro .................... .................... .................... .................... .................... ....................
10430 MW154182.1 |Severe acute respiratory syndrome coro .................... .................... .................... .................... .................... ....................
10431 MW154183.1 |Severe acute respiratory syndrome coro .................... .................... .................... .................... .................... ....................
10432 MW154184.1 |Severe acute respiratory syndrome coro .................... .................... .................... .................... .................... ....................
10433 MW154185.1 |Severe acute respiratory syndrome coro .................... .................... .................... .................... .................... ....................
10434 MW154186.1 |Severe acute respiratory syndrome coro .................... .................... .................... .................... .................... ....................
10435 MW154187.1 |Severe acute respiratory syndrome coro .................... .................... .................... .................... .................... ....................
10436 MW154188.1 |Severe acute respiratory syndrome coro .................... .................... .................... .................... .................... ....................
10437 MW154189.1 |Severe acute respiratory syndrome coro .................... .................... .................... .................... .................... ....................
10438 MW154190.1 |Severe acute respiratory syndrome coro .................... .................... .................... .................... .................... ....................
10439 MW154191.1 |Severe acute respiratory syndrome coro .................... .................... .................... .................... .................... ....................
10440 MW154192.1 |Severe acute respiratory syndrome coro .................... .................... .................... .................... .................... ....................
10441 MW154193.1 |Severe acute respiratory syndrome coro .................... .................... .................... .................... .................... ....................
10442 MW154194.1 |Severe acute respiratory syndrome coro .................... .................... .................... .................... .................... ....................
10443 MW154196.1 |Severe acute respiratory syndrome coro .................... .................... .................... .................... .................... ....................
10444 MW154197.1 |Severe acute respiratory syndrome coro .................... .................... .................... .................... .................... ....................
10445 MW154198.1 |Severe acute respiratory syndrome coro .................... .................... .................... .................... .................... ....................
10446 MW154199.1 |Severe acute respiratory syndrome coro .................... .................... .................... .................... .................... ....................
10447 MW154201.1 |Severe acute respiratory syndrome coro .................... .................... .................... .................... .................... ....................
10448 MW154202.1 |Severe acute respiratory syndrome coro .................... .................... .................... .................... .................... ....................
10449 MW154203.1 |Severe acute respiratory syndrome coro .................... .................... .................... .................... .................... ....................
10450 MW154204.1 |Severe acute respiratory syndrome coro .................... .................... .................... .................... .................... ....................
10451 MW154205.1 |Severe acute respiratory syndrome coro .................... ..........A......... .................... .................... .................... ....................
10452 MW154206.1 |Severe acute respiratory syndrome coro .................... .................... .................... .................... .................... ....................
10453 MW154207.1 |Severe acute respiratory syndrome coro .................... .................... .................... .................... .................... ....................
10454 MW154208.1 |Severe acute respiratory syndrome coro .................... .................... .................... .................... .................... ....................
10455 MW154209.1 |Severe acute respiratory syndrome coro .................... .................... .................... .................... .................... ....................
10456 MW154211.1 |Severe acute respiratory syndrome coro .................... .................... .................... .................... .................... ....................
10457 MW154212.1 |Severe acute respiratory syndrome coro .................... .................... .................... .................... .................... ....................
10458 MW154213.1 |Severe acute respiratory syndrome coro .................... .................... .................... .................... .................... ....................
10459 MW154214.1 |Severe acute respiratory syndrome coro .................... .................... .................... .................... .................... ....................
10460 MW154216.1 |Severe acute respiratory syndrome coro .................... .................... .................... .................... .................... ....................
10461 MW154217.1 |Severe acute respiratory syndrome coro .................... .................... .................... .................... .................... ....................
10462 MW154218.1 |Severe acute respiratory syndrome coro .................... .................... .................... .................... .................... ....................
10463 MW154219.1 |Severe acute respiratory syndrome coro .................... .................... .................... .................... .................... ....................
10464 MW154221.1 |Severe acute respiratory syndrome coro .................... .................... .................... .................... .................... ....................
10465 MW154222.1 |Severe acute respiratory syndrome coro .................... .................... .................... .................... .................... ....................
10466 MW154223.1 |Severe acute respiratory syndrome coro .................... .................... .................... .................... .................... ....................
10467 MW154224.1 |Severe acute respiratory syndrome coro .................... .................... .................... .................... .................... ....................

10468 MW154225.1 |Severe acute respiratory syndrome coro .................... .................... .................... .................... .................... ....................
10469 MW154226.1 |Severe acute respiratory syndrome coro .................... .................... .................... .................... .................... ....................
10470 MW154227.1 |Severe acute respiratory syndrome coro .................... .................... .................... .................... .................... ....................
10471 MW154228.1 |Severe acute respiratory syndrome coro .................... .................... .................... .................... .................... ....................
10472 MW154229.1 |Severe acute respiratory syndrome coro .................... .................... .................... .................... .................... ....................
10473 MW154230.1 |Severe acute respiratory syndrome coro .................... .................... .................... .................... .................... ....................
10474 MW154231.1 |Severe acute respiratory syndrome coro .................... .................... .................... .................... .................... ....................
10475 MW154232.1 |Severe acute respiratory syndrome coro .................... .................... .................... .................... .................... ....................
10476 MW154233.1 |Severe acute respiratory syndrome coro .................... .................... .................... .................... .................... ....................
10477 MW154234.1 |Severe acute respiratory syndrome coro .................... .................... .................... .................... .................... ....................
10478 MW154235.1 |Severe acute respiratory syndrome coro .................... .................... .................... .................... .................... ....................
10479 MW154236.1 |Severe acute respiratory syndrome coro .................... .................... .................... .................... .................... ....................
10480 MW154237.1 |Severe acute respiratory syndrome coro .................... .................... .................... .................... .................... ....................
10481 MW154238.1 |Severe acute respiratory syndrome coro .................... .................... .................... .................... .................... ....................
10482 MW154239.1 |Severe acute respiratory syndrome coro .................... .................... .................... .................... .................... ....................
10483 MW154240.1 |Severe acute respiratory syndrome coro .................... .................... .................... .................... .................... ....................
10484 MW154241.1 |Severe acute respiratory syndrome coro .................... .................... .................... .................... .................... ....................
10485 MW154242.1 |Severe acute respiratory syndrome coro .................... .................... .................... .................... .................... ....................
10486 MW154243.1 |Severe acute respiratory syndrome coro .................... .................... .................... .................... .................... ....................
10487 MW154244.1 |Severe acute respiratory syndrome coro .................... .................... .................... .................... .................... ....................
10488 MW154245.1 |Severe acute respiratory syndrome coro .................... .................... .................... .................... .................... ....................
10489 MW154247.1 |Severe acute respiratory syndrome coro .................... ......A............. .................... .................... .................... ....................
10490 MW154248.1 |Severe acute respiratory syndrome coro .................... .................... .................... .................... .................... ....................
10491 MW154250.1 |Severe acute respiratory syndrome coro .................... .................... .................... .................... .................... ....................
10492 MW154251.1 |Severe acute respiratory syndrome coro .................... .................... .................... .................... .................... ....................
10493 MW154252.1 |Severe acute respiratory syndrome coro .................... .................... .................... .................... .................... ....................
10494 MW154253.1 |Severe acute respiratory syndrome coro .................... .................... .................... .................... .................... ....................
10495 MW154254.1 |Severe acute respiratory syndrome coro .................... .................... .................... .................... .................... ....................
10496 MW154255.1 |Severe acute respiratory syndrome coro .................... .................... .................... .................... .................... ....................
10497 MW154256.1 |Severe acute respiratory syndrome coro .................... .................... .................... .................... .................... ....................
10498 MW154258.1 |Severe acute respiratory syndrome coro .................... .................... .................... .................... .................... ....................
10499 MW154259.1 |Severe acute respiratory syndrome coro .................... .................... .................... .................... .................... ....................
10500 MW154260.1 |Severe acute respiratory syndrome coro .................... .................... .................... .................... .................... ....................
10501 MW154261.1 |Severe acute respiratory syndrome coro .................... .................... .................... .................... .................... ....................
10502 MW154262.1 |Severe acute respiratory syndrome coro .................... .................... .................... .................... .................... ....................
10503 MW154264.1 |Severe acute respiratory syndrome coro .................... .................... .................... .................... .................... ....................
10504 MW154265.1 |Severe acute respiratory syndrome coro .................... .................... .................... .................... .................... ....................
10505 MW154266.1 |Severe acute respiratory syndrome coro .................... .................... .................... .................... .................... ....................
10506 MW154267.1 |Severe acute respiratory syndrome coro .................... .................... .................... .................... .................... ....................
10507 MW154268.1 |Severe acute respiratory syndrome coro .................... .................... .................... .................... .................... ....................
10508 MW154269.1 |Severe acute respiratory syndrome coro .................... .................... .................... .................... .................... ....................
10509 MW154270.1 |Severe acute respiratory syndrome coro .................... .................... .................... .................... .................... ....................
10510 MW154271.1 |Severe acute respiratory syndrome coro .................... .................... .................... .................... .................... ....................
10511 MW154272.1 |Severe acute respiratory syndrome coro .................... .................... .................... .................... .................... ....................
10512 MW154273.1 |Severe acute respiratory syndrome coro .................... .................... .................... .................... .................... ....................
10513 MW154274.1 |Severe acute respiratory syndrome coro .................... .................... .................... .................... .................... ....................
10514 MW154275.1 |Severe acute respiratory syndrome coro .................... .................... .................... .................... .................... ....................
10515 MW154278.1 |Severe acute respiratory syndrome coro .................... .................... .................... .................... .................... ....................
10516 MW154279.1 |Severe acute respiratory syndrome coro .................... .................... .................... .................... .................... ....................
10517 MW154280.1 |Severe acute respiratory syndrome coro .................... .................... .................... .................... .................... ....................
10518 MW154281.1 |Severe acute respiratory syndrome coro .................... .................... .................... .................... .................... ....................
10519 MW154282.1 |Severe acute respiratory syndrome coro .................... .................... .................... .................... .................... ....................
10520 MW154283.1 |Severe acute respiratory syndrome coro .................... .................... .................... .................... .................... ....................
10521 MW154285.1 |Severe acute respiratory syndrome coro .................... .................... .................... .................... .................... ....................
10522 MW154287.1 |Severe acute respiratory syndrome coro .................... .................... .................... .................... .................... ....................
10523 MW154289.1 |Severe acute respiratory syndrome coro .................... .................... .................... .................... .................... ....................
10524 MW154290.1 |Severe acute respiratory syndrome coro .................... .................... .................... .................... .................... ....................
10525 MW154291.1 |Severe acute respiratory syndrome coro .................... .................... .................... .................... .................... ....................
10526 MW154292.1 |Severe acute respiratory syndrome coro .................... .................... .................... .................... .................... ....................
10527 MW154293.1 |Severe acute respiratory syndrome coro .................... ..........A......... .................... .................... .................... ....................
10528 MW154295.1 |Severe acute respiratory syndrome coro .................... .................... .................... .................... .................... ....................
10529 MW154296.1 |Severe acute respiratory syndrome coro .................... .................... .................... .................... .................... ....................
10530 MW154297.1 |Severe acute respiratory syndrome coro .................... .................... .................... .................... .................... ....................
10531 MW154298.1 |Severe acute respiratory syndrome coro .................... ......A............. .................... .................... .................... ....................
10532 MW154299.1 |Severe acute respiratory syndrome coro .................... .................... .................... .................... .................... ....................
10533 MW154301.1 |Severe acute respiratory syndrome coro .................... .................... .................... .................... .................... ....................
10534 MW154302.1 |Severe acute respiratory syndrome coro .................... .................... .................... .................... .................... ....................
10535 MW154304.1 |Severe acute respiratory syndrome coro .................... .................... .................... .................... .................... ....................
10536 MW154305.1 |Severe acute respiratory syndrome coro .................... .................... .................... .................... .................... ....................
10537 MW154306.1 |Severe acute respiratory syndrome coro .................... .................... .................... .................... .................... ....................
10538 MW154307.1 |Severe acute respiratory syndrome coro .................... .................... .................... .................... .................... ....................
10539 MW154308.1 |Severe acute respiratory syndrome coro .................... .................... .................... .................... .................... ....................
10540 MW154309.1 |Severe acute respiratory syndrome coro .................... .................... .................... .................... .................... ....................
10541 MW154310.1 |Severe acute respiratory syndrome coro .................... .................... .................... .................... .................... ....................
10542 MW154311.1 |Severe acute respiratory syndrome coro .................... .................... .................... .................... .................... ....................
10543 MW154312.1 |Severe acute respiratory syndrome coro .................... .................... .................... .................... .................... ....................
10544 MW154313.1 |Severe acute respiratory syndrome coro .................... .................... .................... .................... .................... ....................
10545 MW154314.1 |Severe acute respiratory syndrome coro .................... .................... .................... .................... .................... ....................

| | | | | | | | | |
|---|---|---|---|---|---|---|---|---|
| 10546 | MW154315.1 | \|Severe acute respiratory syndrome coro | .................... | .................... | .................... | .................... | .................... | .................... |
| 10547 | MW154316.1 | \|Severe acute respiratory syndrome coro | .................... | .................... | .................... | .................... | .................... | .................... |
| 10548 | MW154317.1 | \|Severe acute respiratory syndrome coro | .................... | .................... | .................... | .................... | .................... | .................... |
| 10549 | MW154318.1 | \|Severe acute respiratory syndrome coro | .................... | .................... | .................... | .................... | .................... | .................... |
| 10550 | MW154319.1 | \|Severe acute respiratory syndrome coro | .................... | .................... | .................... | .................... | .................... | .................... |
| 10551 | MW154320.1 | \|Severe acute respiratory syndrome coro | .................... | .................... | .................... | .................... | .................... | .................... |
| 10552 | MW154321.1 | \|Severe acute respiratory syndrome coro | .................... | .................... | .................... | .................... | .................... | .................... |
| 10553 | MW154323.1 | \|Severe acute respiratory syndrome coro | .................... | .................... | .................... | .................... | .................... | .................... |
| 10554 | MW154324.1 | \|Severe acute respiratory syndrome coro | .................... | .................... | .................... | .................... | .................... | .................... |
| 10555 | MW154325.1 | \|Severe acute respiratory syndrome coro | .................... | .................... | .................... | .................... | .................... | .................... |
| 10556 | MW154326.1 | \|Severe acute respiratory syndrome coro | .................... | .................... | .................... | .................... | .................... | .................... |
| 10557 | MW154327.1 | \|Severe acute respiratory syndrome coro | .................... | .................... | .................... | .................... | .................... | .................... |
| 10558 | MW154329.1 | \|Severe acute respiratory syndrome coro | .................... | .................... | .................... | .................... | .................... | .................... |
| 10559 | MW154330.1 | \|Severe acute respiratory syndrome coro | .................... | .................... | .................... | .................... | .................... | .................... |
| 10560 | MW154331.1 | \|Severe acute respiratory syndrome coro | .................... | .................... | .................... | .................... | .................... | .................... |
| 10561 | MW154332.1 | \|Severe acute respiratory syndrome coro | .................... | .................... | .................... | .................... | .................... | .................... |
| 10562 | MW154333.1 | \|Severe acute respiratory syndrome coro | .................... | .................... | .................... | .................... | .................... | .................... |
| 10563 | MW154334.1 | \|Severe acute respiratory syndrome coro | .................... | .................... | .................... | .................... | .................... | .................... |
| 10564 | MW154335.1 | \|Severe acute respiratory syndrome coro | .................... | .................... | .................... | .................... | .................... | .................... |
| 10565 | MW154336.1 | \|Severe acute respiratory syndrome coro | .................... | .................... | .................... | .................... | .................... | .................... |
| 10566 | MW154338.1 | \|Severe acute respiratory syndrome coro | .................... | .................... | .................... | .................... | .................... | .................... |
| 10567 | MW154339.1 | \|Severe acute respiratory syndrome coro | .................... | .................... | .................... | .................... | .................... | .................... |
| 10568 | MW154340.1 | \|Severe acute respiratory syndrome coro | .................... | .................... | .................... | .................... | .................... | .................... |
| 10569 | MW154341.1 | \|Severe acute respiratory syndrome coro | .................... | .................... | .................... | .................... | .................... | .................... |
| 10570 | MW154342.1 | \|Severe acute respiratory syndrome coro | .................... | .................... | .................... | .................... | .................... | .................... |
| 10571 | MW154343.1 | \|Severe acute respiratory syndrome coro | .................... | .................... | .................... | .................... | .................... | .................... |
| 10572 | MW154345.1 | \|Severe acute respiratory syndrome coro | .................... | .................... | .................... | .................... | .................... | .................... |
| 10573 | MW154349.1 | \|Severe acute respiratory syndrome coro | .................... | .................... | .................... | .................... | .................... | .................... |
| 10574 | MW154350.1 | \|Severe acute respiratory syndrome coro | .................... | .................... | .................... | .................... | .................... | .................... |
| 10575 | MW154351.1 | \|Severe acute respiratory syndrome coro | .................... | .................... | .................... | .................... | .................... | .................... |
| 10576 | MW154352.1 | \|Severe acute respiratory syndrome coro | .................... | .................... | .................... | .................... | .................... | .................... |
| 10577 | MW154353.1 | \|Severe acute respiratory syndrome coro | .................... | .................... | .................... | .................... | .................... | .................... |
| 10578 | MW154355.1 | \|Severe acute respiratory syndrome coro | .................... | .................... | .................... | .................... | .................... | .................... |
| 10579 | MW154356.1 | \|Severe acute respiratory syndrome coro | .................... | .................... | .................... | .................... | .................... | .................... |
| 10580 | MW154357.1 | \|Severe acute respiratory syndrome coro | .................... | .................... | .................... | .................... | .................... | .................... |
| 10581 | MW154358.1 | \|Severe acute respiratory syndrome coro | .................... | .................... | .................... | .................... | .................... | .................... |
| 10582 | MW154359.1 | \|Severe acute respiratory syndrome coro | .................... | .................... | .................... | .................... | .................... | .................... |
| 10583 | MW154360.1 | \|Severe acute respiratory syndrome coro | .................... | .................... | .................... | .................... | .................... | .................... |
| 10584 | MW154362.1 | \|Severe acute respiratory syndrome coro | .................... | .................... | .................... | .................... | .................... | .................... |
| 10585 | MW154363.1 | \|Severe acute respiratory syndrome coro | .................... | .................... | .................... | .................... | .................... | .................... |
| 10586 | MW154364.1 | \|Severe acute respiratory syndrome coro | .................... | .................... | .................... | .................... | .................... | .................... |
| 10587 | MW154365.1 | \|Severe acute respiratory syndrome coro | .................... | .................... | .................... | .................... | .................... | .................... |
| 10588 | MW154366.1 | \|Severe acute respiratory syndrome coro | .................... | .................... | .................... | .................... | .................... | .................... |
| 10589 | MW154367.1 | \|Severe acute respiratory syndrome coro | .................... | .................... | .................... | .................... | .................... | .................... |
| 10590 | MW154368.1 | \|Severe acute respiratory syndrome coro | .................... | .................... | .................... | .................... | .................... | .................... |
| 10591 | MW154369.1 | \|Severe acute respiratory syndrome coro | .................... | .................... | .................... | .................... | .................... | .................... |
| 10592 | MW154370.1 | \|Severe acute respiratory syndrome coro | .................... | .................... | .................... | .................... | .................... | .................... |
| 10593 | MW154371.1 | \|Severe acute respiratory syndrome coro | .................... | .................... | .................... | .................... | .................... | .................... |
| 10594 | MW154372.1 | \|Severe acute respiratory syndrome coro | .................... | .................... | .................... | .................... | .................... | .................... |
| 10595 | MW154373.1 | \|Severe acute respiratory syndrome coro | .................... | .................... | .................... | .................... | .................... | .................... |
| 10596 | MW154374.1 | \|Severe acute respiratory syndrome coro | .................... | .................... | .................... | .................... | .................... | .................... |
| 10597 | MW154376.1 | \|Severe acute respiratory syndrome coro | .................... | ......A............. | .................... | .................... | .................... | .................... |
| 10598 | MW154378.1 | \|Severe acute respiratory syndrome coro | .................... | .................... | .................... | .................... | .................... | .................... |
| 10599 | MW154379.1 | \|Severe acute respiratory syndrome coro | .................... | .................... | .................... | .................... | .................... | .................... |
| 10600 | MW154380.1 | \|Severe acute respiratory syndrome coro | .................... | .................... | .................... | .................... | .................... | .................... |
| 10601 | MW154381.1 | \|Severe acute respiratory syndrome coro | .................... | .................... | .................... | .................... | .................... | .................... |
| 10602 | MW154382.1 | \|Severe acute respiratory syndrome coro | .................... | .................... | .................... | .................... | .................... | .................... |
| 10603 | MW154383.1 | \|Severe acute respiratory syndrome coro | .................... | .................... | .................... | .................... | .................... | .................... |
| 10604 | MW154385.1 | \|Severe acute respiratory syndrome coro | .................... | .................... | .................... | .................... | .................... | .................... |
| 10605 | MW154386.1 | \|Severe acute respiratory syndrome coro | .................... | .................... | .................... | .................... | .................... | .................... |
| 10606 | MW154387.1 | \|Severe acute respiratory syndrome coro | .................... | .................... | .................... | .................... | .................... | .................... |
| 10607 | MW154388.1 | \|Severe acute respiratory syndrome coro | .................... | .................... | .................... | .................... | .................... | .................... |
| 10608 | MW154389.1 | \|Severe acute respiratory syndrome coro | .................... | .................... | .................... | .................... | .................... | .................... |
| 10609 | MW154390.1 | \|Severe acute respiratory syndrome coro | .................... | .................... | .................... | .................... | .................... | .................... |
| 10610 | MW154391.1 | \|Severe acute respiratory syndrome coro | .................... | .................... | .................... | .................... | .................... | .................... |
| 10611 | MW154392.1 | \|Severe acute respiratory syndrome coro | .................... | .................... | .................... | .................... | .................... | .................... |
| 10612 | MW154393.1 | \|Severe acute respiratory syndrome coro | .................... | .................... | .................... | .................... | .................... | .................... |
| 10613 | MW154394.1 | \|Severe acute respiratory syndrome coro | .................... | .................... | .................... | .................... | .................... | .................... |
| 10614 | MW154395.1 | \|Severe acute respiratory syndrome coro | .................... | .................... | .................... | .................... | .................... | .................... |
| 10615 | MW154397.1 | \|Severe acute respiratory syndrome coro | .................... | .................... | .................... | .................... | .................... | .................... |
| 10616 | MW154399.1 | \|Severe acute respiratory syndrome coro | .................... | .................... | .................... | .................... | .................... | .................... |
| 10617 | MW154400.1 | \|Severe acute respiratory syndrome coro | .................... | .................... | .................... | .................... | .................... | .................... |
| 10618 | MW154401.1 | \|Severe acute respiratory syndrome coro | .................... | .................... | .................... | .................... | .................... | .................... |
| 10619 | MW154402.1 | \|Severe acute respiratory syndrome coro | .................... | .................... | .................... | .................... | .................... | .................... |
| 10620 | MW154403.1 | \|Severe acute respiratory syndrome coro | .................... | .................... | .................... | .................... | .................... | .................... |
| 10621 | MW154404.1 | \|Severe acute respiratory syndrome coro | .................... | .................... | .................... | .................... | .................... | .................... |
| 10622 | MW154405.1 | \|Severe acute respiratory syndrome coro | .................... | .................... | .................... | .................... | .................... | .................... |
| 10623 | MW154406.1 | \|Severe acute respiratory syndrome coro | .................... | .................... | .................... | .................... | .................... | .................... |

10624 MW154407.1 |Severe acute respiratory syndrome coro .................... .................... ..................... .................... .................... ....................
10625 MW154408.1 |Severe acute respiratory syndrome coro .................... .................... ..................... .................... .................... ....................
10626 MW154409.1 |Severe acute respiratory syndrome coro .................... .................... ..................... .................... .................... ....................
10627 MW154411.1 |Severe acute respiratory syndrome coro .................... .................... ..................... .................... .................... ....................
10628 MW154412.1 |Severe acute respiratory syndrome coro .................... .................... ..................... .................... .................... ....................
10629 MW154413.1 |Severe acute respiratory syndrome coro .................... .................... ..................... .................... .................... ....................
10630 MW154417.1 |Severe acute respiratory syndrome coro .................... .................... ..................... .................... .................... ....................
10631 MW154419.1 |Severe acute respiratory syndrome coro .................... .................... ..................... .................... .................... ....................
10632 MW154421.1 |Severe acute respiratory syndrome coro .................... .................... ..................... .................... .................... ....................
10633 MW154423.1 |Severe acute respiratory syndrome coro .................... .................... ..................... .................... .................... ....................
10634 MW154424.1 |Severe acute respiratory syndrome coro .................... .................... ..................... .................... .................... ....................
10635 MW154425.1 |Severe acute respiratory syndrome coro .................... .................... ..................... .................... .................... ....................
10636 MW154426.1 |Severe acute respiratory syndrome coro .................... .................... ..................... .................... .................... ....................
10637 MW154428.1 |Severe acute respiratory syndrome coro .................... .................... ..................... .................... .................... ....................
10638 MW154429.1 |Severe acute respiratory syndrome coro .................... .................... ..................... .................... .................... ....................
10639 MW154430.1 |Severe acute respiratory syndrome coro .................... .................... ..................... .................... .................... ....................
10640 MW154431.1 |Severe acute respiratory syndrome coro .................... .................... ..................... .................... .................... ....................
10641 MW154432.1 |Severe acute respiratory syndrome coro .................... .................... ..................... .................... .................... ....................
10642 MW154433.1 |Severe acute respiratory syndrome coro .................... .................... ..................... .................... .................... ....................
10643 MW154434.1 |Severe acute respiratory syndrome coro .................... .................... ..................... .................... .................... ....................
10644 MW154435.1 |Severe acute respiratory syndrome coro .................... .................... ..................... .................... .................... ....................
10645 MW154437.1 |Severe acute respiratory syndrome coro .................... .................... ..................... .................... .................... ....................
10646 MW154438.1 |Severe acute respiratory syndrome coro .................... .................... ..................... .................... .................... ....................
10647 MW154439.1 |Severe acute respiratory syndrome coro .................... .................... ..................... .................... .................... ....................
10648 MW154440.1 |Severe acute respiratory syndrome coro .................... .................... ..................... .................... .................... ....................
10649 MW154441.1 |Severe acute respiratory syndrome coro .................... .................... ..................... .................... .................... ....................
10650 MW154442.1 |Severe acute respiratory syndrome coro .................... .................... ..................... .................... .................... ....................
10651 MW154443.1 |Severe acute respiratory syndrome coro .................... .................... ..................... .................... .................... ....................
10652 MW154444.1 |Severe acute respiratory syndrome coro .................... .................... ..................... .................... .................... ....................
10653 MW154445.1 |Severe acute respiratory syndrome coro .................... .................... ..................... .................... .................... ....................
10654 MW154446.1 |Severe acute respiratory syndrome coro .................... .................... ..................... .................... .................... ....................
10655 MW154447.1 |Severe acute respiratory syndrome coro .................... .................... ..................... .................... .................... ....................
10656 MW154448.1 |Severe acute respiratory syndrome coro .................... .................... ..................... .................... .................... ....................
10657 MW154449.1 |Severe acute respiratory syndrome coro .................... .................... ..................... .................... .................... ....................
10658 MW154450.1 |Severe acute respiratory syndrome coro .................... .................... ..................... .................... .................... ....................
10659 MW154451.1 |Severe acute respiratory syndrome coro .................... .................... ..................... .................... .................... ....................
10660 MW154452.1 |Severe acute respiratory syndrome coro .................... .................... ..................... .................... .................... ....................
10661 MW154453.1 |Severe acute respiratory syndrome coro .................... .................... ..................... .................... .................... ....................
10662 MW154455.1 |Severe acute respiratory syndrome coro .................... .................... ..................... .................... .................... ....................
10663 MW154456.1 |Severe acute respiratory syndrome coro .................... .................... ..................... .................... .................... ....................
10664 MW154457.1 |Severe acute respiratory syndrome coro .................... .................... ..................... .................... .................... ....................
10665 MW154458.1 |Severe acute respiratory syndrome coro .................... .................... ..................... .................... .................... ....................
10666 MW154459.1 |Severe acute respiratory syndrome coro .................... .................... ..................... .................... .................... ....................
10667 MW154460.1 |Severe acute respiratory syndrome coro .................... .................... ..................... .................... .................... ....................
10668 MW154461.1 |Severe acute respiratory syndrome coro .................... .................... ..................... .................... .................... ....................
10669 MW154463.1 |Severe acute respiratory syndrome coro .................... .................... ..................... .................... .................... ....................
10670 MW154464.1 |Severe acute respiratory syndrome coro .................... .................... ..................... .................... .................... ....................
10671 MW154465.1 |Severe acute respiratory syndrome coro .................... .................... ..................... .................... .................... ....................
10672 MW154466.1 |Severe acute respiratory syndrome coro .................... .................... ..................... .................... .................... ....................
10673 MW154468.1 |Severe acute respiratory syndrome coro .................... .................... ..................... .................... .................... ....................
10674 MW154469.1 |Severe acute respiratory syndrome coro .................... .................... ..................... .................... .................... ....................
10675 MW154470.1 |Severe acute respiratory syndrome coro .................... .................... ..................... .................... .................... ....................
10676 MW154471.1 |Severe acute respiratory syndrome coro .................... .................... ..................... .................... .................... ....................
10677 MW154472.1 |Severe acute respiratory syndrome coro .................... .................... ..................... .................... .................... ....................
10678 MW154473.1 |Severe acute respiratory syndrome coro .................... .................... ..................... .................... .................... ....................
10679 MW154474.1 |Severe acute respiratory syndrome coro .................... .................... ..................... .................... .................... ....................
10680 MW154475.1 |Severe acute respiratory syndrome coro .................... .................... ..................... .................... .................... ....................
10681 MW154476.1 |Severe acute respiratory syndrome coro .................... .................... ..................... .................... .................... ....................
10682 MW154477.1 |Severe acute respiratory syndrome coro .................... .................... ..................... .................... .................... ....................
10683 MW154479.1 |Severe acute respiratory syndrome coro .................... .................... ..................... .................... .................... ....................
10684 MW154480.1 |Severe acute respiratory syndrome coro .................... .................... ..................... .................... .................... ....................
10685 MW154481.1 |Severe acute respiratory syndrome coro .................... .................... ..................... .................... .................... ....................
10686 MW154483.1 |Severe acute respiratory syndrome coro .................... .................... ..................... .................... .................... ....................
10687 MW154484.1 |Severe acute respiratory syndrome coro .................... .................... ..................... .................... .................... ....................
10688 MW154485.1 |Severe acute respiratory syndrome coro .................... .................... ..................... .................... .................... ....................
10689 MW154486.1 |Severe acute respiratory syndrome coro .................... .................... ..................... .................... .................... ....................
10690 MW154487.1 |Severe acute respiratory syndrome coro .................... .................... ..................... .................... .................... ....................
10691 MW154488.1 |Severe acute respiratory syndrome coro .................... .................... ..................... .................... .................... ....................
10692 MW154490.1 |Severe acute respiratory syndrome coro .................... .................... ..................... .................... .................... ....................
10693 MW154491.1 |Severe acute respiratory syndrome coro .................... .................... ..................... .................... .................... ....................
10694 MW154492.1 |Severe acute respiratory syndrome coro .................... .................... ..................... .................... .................... ....................
10695 MW154493.1 |Severe acute respiratory syndrome coro .................... .................... ..................... .................... .................... ....................
10696 MW154494.1 |Severe acute respiratory syndrome coro .................... .................... ..................... .................... .................... ....................
10697 MW154495.1 |Severe acute respiratory syndrome coro .................... .................... ..................... .................... .................... ....................
10698 MW154496.1 |Severe acute respiratory syndrome coro .................... .................... ..................... .................... .................... ....................
10699 MW154497.1 |Severe acute respiratory syndrome coro .................... .................... ..................... .................... .................... ....................
10700 MW154498.1 |Severe acute respiratory syndrome coro .................... .................... ..................... .................... .................... ....................
10701 MW154499.1 |Severe acute respiratory syndrome coro .................... .................... ..................... .................... .................... ....................

| | | | | | | | | |
|---|---|---|---|---|---|---|---|---|
| 10702 | MW154500.1 | \|Severe acute respiratory syndrome coro | .................... | .................... | ..................... | .................... | .................... | .................... |
| 10703 | MW154501.1 | \|Severe acute respiratory syndrome coro | .................... | .................... | ..................... | .................... | .................... | .................... |
| 10704 | MW154502.1 | \|Severe acute respiratory syndrome coro | .................... | .................... | ..................... | .................... | .................... | .................... |
| 10705 | MW154503.1 | \|Severe acute respiratory syndrome coro | .................... | .................... | ..................... | .................... | .................... | .................... |
| 10706 | MW154504.1 | \|Severe acute respiratory syndrome coro | .................... | .................... | ..................... | .................... | .................... | .................... |
| 10707 | MW154505.1 | \|Severe acute respiratory syndrome coro | .................... | .................... | ..................... | .................... | .................... | .................... |
| 10708 | MW154506.1 | \|Severe acute respiratory syndrome coro | .................... | .................... | ..................... | .................... | .................... | .................... |
| 10709 | MW154507.1 | \|Severe acute respiratory syndrome coro | .................... | .................... | ..................... | .................... | .................... | .................... |
| 10710 | MW154508.1 | \|Severe acute respiratory syndrome coro | .................... | .................... | ..................... | .................... | .................... | .................... |
| 10711 | MW154509.1 | \|Severe acute respiratory syndrome coro | .................... | .................... | ..................... | .................... | .................... | .................... |
| 10712 | MW154510.1 | \|Severe acute respiratory syndrome coro | .................... | .................... | ..................... | .................... | .................... | .................... |
| 10713 | MW154511.1 | \|Severe acute respiratory syndrome coro | .................... | .................... | ..................... | .................... | .................... | .................... |
| 10714 | MW154512.1 | \|Severe acute respiratory syndrome coro | .................... | .................... | ..................... | .................... | .................... | .................... |
| 10715 | MW154513.1 | \|Severe acute respiratory syndrome coro | .................... | .................... | ..................... | .................... | .................... | .................... |
| 10716 | MW154514.1 | \|Severe acute respiratory syndrome coro | .................... | .................... | ..................... | .................... | .................... | .................... |
| 10717 | MW154515.1 | \|Severe acute respiratory syndrome coro | .................... | .................... | ..................... | .................... | .................... | .................... |
| 10718 | MW154516.1 | \|Severe acute respiratory syndrome coro | .................... | .................... | ..................... | .................... | .................... | .................... |
| 10719 | MW154517.1 | \|Severe acute respiratory syndrome coro | .................... | .................... | ..................... | .................... | .................... | .................... |
| 10720 | MW154518.1 | \|Severe acute respiratory syndrome coro | .................... | .................... | ..................... | .................... | .................... | .................... |
| 10721 | MW154519.1 | \|Severe acute respiratory syndrome coro | .................... | .................... | ..................... | .................... | .................... | .................... |
| 10722 | MW154520.1 | \|Severe acute respiratory syndrome coro | .................... | .................... | ..................... | .................... | .................... | .................... |
| 10723 | MW154521.1 | \|Severe acute respiratory syndrome coro | .................... | .................... | ..................... | .................... | .................... | .................... |
| 10724 | MW154522.1 | \|Severe acute respiratory syndrome coro | .................... | .................... | ..................... | .................... | .................... | .................... |
| 10725 | MW154523.1 | \|Severe acute respiratory syndrome coro | .................... | .................... | ..................... | .................... | .................... | .................... |
| 10726 | MW154524.1 | \|Severe acute respiratory syndrome coro | .................... | .................... | ..................... | .................... | .................... | .................... |
| 10727 | MW154526.1 | \|Severe acute respiratory syndrome coro | .................... | .................... | ..................... | .................... | .................... | .................... |
| 10728 | MW154527.1 | \|Severe acute respiratory syndrome coro | .................... | .................... | ..................... | .................... | .................... | .................... |
| 10729 | MW154528.1 | \|Severe acute respiratory syndrome coro | .................... | .................... | ..................... | .................... | .................... | .................... |
| 10730 | MW154530.1 | \|Severe acute respiratory syndrome coro | .................... | .................... | ..................... | .................... | .................... | .................... |
| 10731 | MW154531.1 | \|Severe acute respiratory syndrome coro | .................... | .................... | ..................... | .................... | .................... | .................... |
| 10732 | MW154532.1 | \|Severe acute respiratory syndrome coro | .................... | .................... | ..................... | .................... | .................... | .................... |
| 10733 | MW154533.1 | \|Severe acute respiratory syndrome coro | .................... | .................... | ..................... | .................... | .................... | .................... |
| 10734 | MW154534.1 | \|Severe acute respiratory syndrome coro | .................... | .................... | ..................... | .................... | .................... | .................... |
| 10735 | MW154536.1 | \|Severe acute respiratory syndrome coro | .................... | .................... | ..................... | .................... | .................... | .................... |
| 10736 | MW154537.1 | \|Severe acute respiratory syndrome coro | .................... | .................... | ..................... | .................... | .................... | .................... |
| 10737 | MW154538.1 | \|Severe acute respiratory syndrome coro | .................... | .................... | ..................... | .................... | .................... | .................... |
| 10738 | MW154539.1 | \|Severe acute respiratory syndrome coro | .................... | .................... | ..................... | .................... | .................... | .................... |
| 10739 | MW154542.1 | \|Severe acute respiratory syndrome coro | .................... | .................... | ..................... | .................... | .................... | .................... |
| 10740 | MW154543.1 | \|Severe acute respiratory syndrome coro | .................... | .................... | ..................... | .................... | .................... | .................... |
| 10741 | MW154546.1 | \|Severe acute respiratory syndrome coro | .................... | .................... | ..................... | .................... | .................... | .................... |
| 10742 | MW154547.1 | \|Severe acute respiratory syndrome coro | .................... | .................... | ..................... | .................... | .................... | .................... |
| 10743 | MW154548.1 | \|Severe acute respiratory syndrome coro | .................... | .................... | ..................... | .................... | .................... | .................... |
| 10744 | MW154549.1 | \|Severe acute respiratory syndrome coro | .................... | .................... | ..................... | .................... | .................... | .................... |
| 10745 | MW154550.1 | \|Severe acute respiratory syndrome coro | .................... | .................... | ..................... | .................... | .................... | .................... |
| 10746 | MW154552.1 | \|Severe acute respiratory syndrome coro | .................... | .................... | ..................... | .................... | .................... | .................... |
| 10747 | MW154553.1 | \|Severe acute respiratory syndrome coro | .................... | .................... | ..................... | .................... | .................... | .................... |
| 10748 | MW154554.1 | \|Severe acute respiratory syndrome coro | .................... | .................... | ..................... | .................... | .................... | .................... |
| 10749 | MW154555.1 | \|Severe acute respiratory syndrome coro | .................... | .................... | ..................... | .................... | .................... | .................... |
| 10750 | MW154556.1 | \|Severe acute respiratory syndrome coro | .................... | .................... | ..................... | .................... | .................... | .................... |
| 10751 | MW154557.1 | \|Severe acute respiratory syndrome coro | .................... | .................... | ..................... | .................... | .................... | .................... |
| 10752 | MW154559.1 | \|Severe acute respiratory syndrome coro | .................... | .................... | ..................... | .................... | .................... | .................... |
| 10753 | MW154560.1 | \|Severe acute respiratory syndrome coro | .................... | .................... | ..................... | .................... | .................... | .................... |
| 10754 | MW154561.1 | \|Severe acute respiratory syndrome coro | .................... | .................... | ..................... | .................... | .................... | .................... |
| 10755 | MW154562.1 | \|Severe acute respiratory syndrome coro | .................... | .................... | ..................... | .................... | .................... | .................... |
| 10756 | MW154564.1 | \|Severe acute respiratory syndrome coro | .................... | .................... | ..................... | .................... | .................... | .................... |
| 10757 | MW154565.1 | \|Severe acute respiratory syndrome coro | .................... | .................... | ..................... | .................... | .................... | .................... |
| 10758 | MW154566.1 | \|Severe acute respiratory syndrome coro | .................... | .................... | ..................... | .................... | .................... | .................... |
| 10759 | MW154567.1 | \|Severe acute respiratory syndrome coro | .................... | .................... | ..................... | .................... | .................... | .................... |
| 10760 | MW154569.1 | \|Severe acute respiratory syndrome coro | .................... | .................... | ..................... | .................... | .................... | .................... |
| 10761 | MW154570.1 | \|Severe acute respiratory syndrome coro | .................... | .................... | ..................... | .................... | .................... | .................... |
| 10762 | MW154571.1 | \|Severe acute respiratory syndrome coro | .................... | ....A............... | ..................... | .................... | .................... | .................... |
| 10763 | MW154572.1 | \|Severe acute respiratory syndrome coro | .................... | .................... | ..................... | .................... | .................... | .................... |
| 10764 | MW154573.1 | \|Severe acute respiratory syndrome coro | .................... | .................... | ..................... | .................... | .................... | .................... |
| 10765 | MW154574.1 | \|Severe acute respiratory syndrome coro | .................... | .................... | ..................... | .................... | .................... | .................... |
| 10766 | MW154576.1 | \|Severe acute respiratory syndrome coro | .................... | .................... | ..................... | .................... | .................... | .................... |
| 10767 | MW154577.1 | \|Severe acute respiratory syndrome coro | .................... | .................... | ..................... | .................... | .................... | .................... |
| 10768 | MW154578.1 | \|Severe acute respiratory syndrome coro | .................... | .................... | ..................... | .................... | .................... | .................... |
| 10769 | MW154579.1 | \|Severe acute respiratory syndrome coro | .................... | .................... | ..................... | .................... | .................... | .................... |
| 10770 | MW154583.1 | \|Severe acute respiratory syndrome coro | .................... | .................... | ..................... | .................... | .................... | .................... |
| 10771 | MW154584.1 | \|Severe acute respiratory syndrome coro | .................... | .................... | ..................... | .................... | .................... | .................... |
| 10772 | MW154585.1 | \|Severe acute respiratory syndrome coro | .................... | .................... | ..................... | .................... | .................... | .................... |
| 10773 | MW154586.1 | \|Severe acute respiratory syndrome coro | .................... | .................... | ..................... | .................... | .................... | .................... |
| 10774 | MW154588.1 | \|Severe acute respiratory syndrome coro | .................... | .................... | ..................... | .................... | .................... | .................... |
| 10775 | MW154589.1 | \|Severe acute respiratory syndrome coro | .................... | .................... | ..................... | .................... | .................... | .................... |
| 10776 | MW154590.1 | \|Severe acute respiratory syndrome coro | .................... | .................... | ..................... | .................... | .................... | .................... |
| 10777 | MW154591.1 | \|Severe acute respiratory syndrome coro | .................... | .................... | ..................... | .................... | .................... | .................... |
| 10778 | MW154592.1 | \|Severe acute respiratory syndrome coro | .................... | .................... | ..................... | .................... | .................... | .................... |
| 10779 | MW154594.1 | \|Severe acute respiratory syndrome coro | .................... | .................... | ..................... | .................... | .................... | .................... |

| | | | | | | | | |
|---|---|---|---|---|---|---|---|---|
| 10780 | MW154597.1 | \|Severe acute respiratory syndrome coro | .................... | .................... | ..................... | .................... | .................... | .................... |
| 10781 | MW154598.1 | \|Severe acute respiratory syndrome coro | .................... | .................... | ..................... | .................... | .................... | .................... |
| 10782 | MW154599.1 | \|Severe acute respiratory syndrome coro | .................... | .................... | ..................... | .................... | .................... | .................... |
| 10783 | MW154600.1 | \|Severe acute respiratory syndrome coro | .................... | .................... | ..................... | .................... | .................... | .................... |
| 10784 | MW154601.1 | \|Severe acute respiratory syndrome coro | .................... | .................... | ..................... | .................... | .................... | .................... |
| 10785 | MW154602.1 | \|Severe acute respiratory syndrome coro | .................... | .................... | ..................... | .................... | .................... | .................... |
| 10786 | MW154604.1 | \|Severe acute respiratory syndrome coro | .................... | .................... | ..................... | .................... | .................... | .................... |
| 10787 | MW154606.1 | \|Severe acute respiratory syndrome coro | .................... | .................... | ..................... | .................... | .................... | .................... |
| 10788 | MW154607.1 | \|Severe acute respiratory syndrome coro | .................... | .................... | ..................... | .................... | .................... | .................... |
| 10789 | MW154609.1 | \|Severe acute respiratory syndrome coro | .................... | .................... | ..................... | .................... | .................... | .................... |
| 10790 | MW154610.1 | \|Severe acute respiratory syndrome coro | .................... | .................... | ..................... | .................... | .................... | .................... |
| 10791 | MW154611.1 | \|Severe acute respiratory syndrome coro | .................... | .................... | ..................... | .................... | .................... | .................... |
| 10792 | MW154612.1 | \|Severe acute respiratory syndrome coro | .................... | .................... | ..................... | .................... | .................... | .................... |
| 10793 | MW154613.1 | \|Severe acute respiratory syndrome coro | .................... | .................... | ..................... | .................... | .................... | .................... |
| 10794 | MW154616.1 | \|Severe acute respiratory syndrome coro | .................... | .................... | ..................... | .................... | .................... | .................... |
| 10795 | MW154617.1 | \|Severe acute respiratory syndrome coro | .................... | .................... | ..................... | .................... | .................... | .................... |
| 10796 | MW154618.1 | \|Severe acute respiratory syndrome coro | .................... | .................... | ..................... | .................... | .................... | .................... |
| 10797 | MW154619.1 | \|Severe acute respiratory syndrome coro | .................... | .................... | ..................... | .................... | .................... | .................... |
| 10798 | MW154620.1 | \|Severe acute respiratory syndrome coro | .................... | .................... | ..................... | .................... | .................... | .................... |
| 10799 | MW154622.1 | \|Severe acute respiratory syndrome coro | .................... | .................... | ..................... | .................... | .................... | .................... |
| 10800 | MW154623.1 | \|Severe acute respiratory syndrome coro | .................... | .................... | ..................... | .................... | .................... | .................... |
| 10801 | MW154624.1 | \|Severe acute respiratory syndrome coro | .................... | .................... | ..................... | .................... | .................... | .................... |
| 10802 | MW154625.1 | \|Severe acute respiratory syndrome coro | .................... | .................... | ..................... | .................... | .................... | .................... |
| 10803 | MW154626.1 | \|Severe acute respiratory syndrome coro | .................... | .................... | ..................... | .................... | .................... | .................... |
| 10804 | MW154627.1 | \|Severe acute respiratory syndrome coro | .................... | .................... | ..................... | .................... | .................... | .................... |
| 10805 | MW154628.1 | \|Severe acute respiratory syndrome coro | .................... | .................... | ..................... | .................... | .................... | .................... |
| 10806 | MW154629.1 | \|Severe acute respiratory syndrome coro | .................... | .................... | ..................... | .................... | .................... | .................... |
| 10807 | MW154632.1 | \|Severe acute respiratory syndrome coro | .................... | .................... | ..................... | .................... | .................... | .................... |
| 10808 | MW154633.1 | \|Severe acute respiratory syndrome coro | .................... | .................... | ..................... | .................... | .................... | .................... |
| 10809 | MW154635.1 | \|Severe acute respiratory syndrome coro | .................... | .................... | ..................... | .................... | .................... | .................... |
| 10810 | MW154636.1 | \|Severe acute respiratory syndrome coro | .................... | .................... | ..................... | .................... | .................... | .................... |
| 10811 | MW154637.1 | \|Severe acute respiratory syndrome coro | .................... | .................... | ..................... | .................... | .................... | .................... |
| 10812 | MW154638.1 | \|Severe acute respiratory syndrome coro | .................... | .................... | ..................... | .................... | .................... | .................... |
| 10813 | MW154640.1 | \|Severe acute respiratory syndrome coro | .................... | .................... | ..................... | .................... | .................... | .................... |
| 10814 | MW154641.1 | \|Severe acute respiratory syndrome coro | .................... | .................... | ..................... | .................... | .................... | .................... |
| 10815 | MW154644.1 | \|Severe acute respiratory syndrome coro | .................... | .................... | ..................... | .................... | .................... | .................... |
| 10816 | MW154645.1 | \|Severe acute respiratory syndrome coro | .................... | .................... | ..................... | .................... | .................... | .................... |
| 10817 | MW154646.1 | \|Severe acute respiratory syndrome coro | .................... | .................... | ..................... | .................... | .................... | .................... |
| 10818 | MW154647.1 | \|Severe acute respiratory syndrome coro | .................... | .................... | ..................... | .................... | .................... | .................... |
| 10819 | MW154648.1 | \|Severe acute respiratory syndrome coro | .................... | .................... | ..................... | .................... | .................... | .................... |
| 10820 | MW154649.1 | \|Severe acute respiratory syndrome coro | .................... | .................... | ..................... | .................... | .................... | .................... |
| 10821 | MW154650.1 | \|Severe acute respiratory syndrome coro | .................... | .................... | ..................... | .................... | .................... | .................... |
| 10822 | MW154651.1 | \|Severe acute respiratory syndrome coro | .................... | .................... | ..................... | .................... | .................... | .................... |
| 10823 | MW154653.1 | \|Severe acute respiratory syndrome coro | .................... | .................... | ..................... | .................... | .................... | .................... |
| 10824 | MW154655.1 | \|Severe acute respiratory syndrome coro | .................... | .................... | ..................... | .................... | .................... | .................... |
| 10825 | MW154656.1 | \|Severe acute respiratory syndrome coro | .................... | .................... | ..................... | .................... | .................... | .................... |
| 10826 | MW154657.1 | \|Severe acute respiratory syndrome coro | .................... | .................... | ..................... | .................... | .................... | .................... |
| 10827 | MW154658.1 | \|Severe acute respiratory syndrome coro | .................... | .................... | ..................... | .................... | .................... | .................... |
| 10828 | MW154659.1 | \|Severe acute respiratory syndrome coro | .................... | .................... | ..................... | .................... | .................... | .................... |
| 10829 | MW154660.1 | \|Severe acute respiratory syndrome coro | .................... | .................... | ..................... | .................... | .................... | .................... |
| 10830 | MW154661.1 | \|Severe acute respiratory syndrome coro | .................... | .................... | ..................... | .................... | .................... | .................... |
| 10831 | MW154662.1 | \|Severe acute respiratory syndrome coro | .................... | .................... | ..................... | .................... | .................... | .................... |
| 10832 | MW154663.1 | \|Severe acute respiratory syndrome coro | .................... | .................... | ..................... | .................... | .................... | .................... |
| 10833 | MW154665.1 | \|Severe acute respiratory syndrome coro | .................... | .................... | ..................... | .................... | .................... | .................... |
| 10834 | MW154666.1 | \|Severe acute respiratory syndrome coro | .................... | .................... | ..................... | .................... | .................... | .................... |
| 10835 | MW154667.1 | \|Severe acute respiratory syndrome coro | .................... | .................... | ..................... | .................... | .................... | .................... |
| 10836 | MW154668.1 | \|Severe acute respiratory syndrome coro | .................... | .................... | ..................... | .................... | .................... | .................... |
| 10837 | MW154669.1 | \|Severe acute respiratory syndrome coro | .................... | .................... | ..................... | .................... | .................... | .................... |
| 10838 | MW154670.1 | \|Severe acute respiratory syndrome coro | .................... | .................... | ..................... | .................... | .................... | .................... |
| 10839 | MW154671.1 | \|Severe acute respiratory syndrome coro | .................... | .................... | ..................... | .................... | .................... | .................... |
| 10840 | MW154672.1 | \|Severe acute respiratory syndrome coro | .................... | .................... | ..................... | .................... | .................... | .................... |
| 10841 | MW154673.1 | \|Severe acute respiratory syndrome coro | .................... | .................... | ..................... | .................... | .................... | .................... |
| 10842 | MW154674.1 | \|Severe acute respiratory syndrome coro | .................... | .................... | ..................... | .................... | .................... | .................... |
| 10843 | MW154675.1 | \|Severe acute respiratory syndrome coro | .................... | .................... | ..................... | .................... | .................... | .................... |
| 10844 | MW154676.1 | \|Severe acute respiratory syndrome coro | .................... | .................... | ..................... | .................... | .................... | .................... |
| 10845 | MW154677.1 | \|Severe acute respiratory syndrome coro | .................... | .................... | ..................... | .................... | .................... | .................... |
| 10846 | MW154680.1 | \|Severe acute respiratory syndrome coro | .................... | .................... | ..................... | .................... | .................... | .................... |
| 10847 | MW154681.1 | \|Severe acute respiratory syndrome coro | .................... | .................... | ..................... | .................... | .................... | .................... |
| 10848 | MW154682.1 | \|Severe acute respiratory syndrome coro | .................... | .................... | ..................... | .................... | .................... | .................... |
| 10849 | MW154683.1 | \|Severe acute respiratory syndrome coro | .................... | .................... | ..................... | .................... | .................... | .................... |
| 10850 | MW154684.1 | \|Severe acute respiratory syndrome coro | .................... | .................... | ..................... | .................... | .................... | .................... |
| 10851 | MW154685.1 | \|Severe acute respiratory syndrome coro | .................... | .................... | ..................... | .................... | .................... | .................... |
| 10852 | MW154686.1 | \|Severe acute respiratory syndrome coro | .................... | .................... | ..................... | .................... | .................... | .................... |
| 10853 | MW154687.1 | \|Severe acute respiratory syndrome coro | .................... | .................... | ..................... | .................... | .................... | .................... |
| 10854 | MW154688.1 | \|Severe acute respiratory syndrome coro | .................... | .................... | ..................... | .................... | .................... | .................... |
| 10855 | MW154689.1 | \|Severe acute respiratory syndrome coro | .................... | .................... | ..................... | .................... | .................... | .................... |
| 10856 | MW154694.1 | \|Severe acute respiratory syndrome coro | .................... | .................... | ..................... | .................... | .................... | .................... |
| 10857 | MW154695.1 | \|Severe acute respiratory syndrome coro | .................... | .................... | ..................... | .................... | .................... | .................... |

```
10858 MW154696.1 |Severe acute respiratory syndrome coro    ....................  ....................  ....................  ....................  ....................  ....................
10859 MW154698.1 |Severe acute respiratory syndrome coro    ....................  ....................  ....................  ....................  ....................  ....................
10860 MW154699.1 |Severe acute respiratory syndrome coro    ....................  ....................  ....................  ....................  ....................  ....................
10861 MW154700.1 |Severe acute respiratory syndrome coro    ....................  ....................  ....................  ....................  ....................  ....................
10862 MW154701.1 |Severe acute respiratory syndrome coro    ....................  ....................  ....................  ....................  ....................  ....................
10863 MW154702.1 |Severe acute respiratory syndrome coro    ....................  ....................  ....................  ....................  ....................  ....................
10864 MW154703.1 |Severe acute respiratory syndrome coro    ....................  ....................  ....................  ....................  ....................  ....................
10865 MW154704.1 |Severe acute respiratory syndrome coro    ....................  ....................  ....................  ....................  ....................  ....................
10866 MW154705.1 |Severe acute respiratory syndrome coro    ....................  ....................  ....................  ....................  ....................  ....................
10867 MW154706.1 |Severe acute respiratory syndrome coro    ....................  ....................  ....................  ....................  ....................  ....................
10868 MW154708.1 |Severe acute respiratory syndrome coro    ....................  ....................  ....................  ....................  ....................  ....................
10869 MW154709.1 |Severe acute respiratory syndrome coro    ....................  ....................  ....................  ....................  ....................  ....................
10870 MW154710.1 |Severe acute respiratory syndrome coro    ....................  ....................  ....................  ....................  ....................  ....................
10871 MW154711.1 |Severe acute respiratory syndrome coro    ....................  ....................  ....................  ....................  ....................  ....................
10872 MW154712.1 |Severe acute respiratory syndrome coro    ....................  ....................  ....................  ....................  ....................  ....................
10873 MW154713.1 |Severe acute respiratory syndrome coro    ....................  ....................  ....................  ....................  ....................  ....................
10874 MW154714.1 |Severe acute respiratory syndrome coro    ....................  ....................  ....................  ....................  ....................  ....................
10875 MW154723.1 |Severe acute respiratory syndrome coro    ....................  ....................  ....................  ....................  ....................  ....................
10876 MW154724.1 |Severe acute respiratory syndrome coro    ....................  ....................  ....................  ....................  ....................  ....................
10877 MW154725.1 |Severe acute respiratory syndrome coro    ....................  ....................  ....................  ....................  ....................  ....................
10878 MW154726.1 |Severe acute respiratory syndrome coro    ....................  ....................  ....................  ....................  ....................  ....................
10879 MW154727.1 |Severe acute respiratory syndrome coro    ....................  ....................  ....................  ....................  ....................  ....................
10880 MW154728.1 |Severe acute respiratory syndrome coro    ....................  ....................  ....................  ....................  ....................  ....................
10881 MW154729.1 |Severe acute respiratory syndrome coro    ....................  ....................  ....................  ....................  ....................  ....................
10882 MW154730.1 |Severe acute respiratory syndrome coro    ....................  ....................  ....................  ....................  ....................  ....................
10883 MW154731.1 |Severe acute respiratory syndrome coro    ....................  ....................  ....................  ....................  ....................  ....................
10884 MW154732.1 |Severe acute respiratory syndrome coro    ....................  ....................  ....................  ....................  ....................  ....................
10885 MW154734.1 |Severe acute respiratory syndrome coro    ....................  ......A.............  ....................  ....................  ....................  ....................
10886 MW154735.1 |Severe acute respiratory syndrome coro    ....................  ....................  ....................  ....................  ....................  ....................
10887 MW154736.1 |Severe acute respiratory syndrome coro    ....................  ....................  ....................  ....................  ....................  ....................
10888 MW154737.1 |Severe acute respiratory syndrome coro    ....................  ....................  ....................  ....................  ....................  ....................
10889 MW154738.1 |Severe acute respiratory syndrome coro    ....................  ....................  ....................  ....................  ....................  ....................
10890 MW154741.1 |Severe acute respiratory syndrome coro    ....................  ....................  ....................  ....................  ....................  ....................
10891 MW154742.1 |Severe acute respiratory syndrome coro    ....................  ....................  ....................  ....................  ....................  ....................
10892 MW154743.1 |Severe acute respiratory syndrome coro    ....................  ....................  ....................  ....................  ....................  ....................
10893 MW154748.1 |Severe acute respiratory syndrome coro    ....................  ....................  ....................  ....................  ....................  ....................
10894 MW154749.1 |Severe acute respiratory syndrome coro    ....................  ....................  ....................  ....................  ....................  ....................
10895 MW154750.1 |Severe acute respiratory syndrome coro    ....................  ....................  ....................  ....................  ....................  ....................
10896 MW154751.1 |Severe acute respiratory syndrome coro    ....................  ....................  ....................  ....................  ....................  ....................
10897 MW154752.1 |Severe acute respiratory syndrome coro    ....................  ....................  ....................  ....................  ....................  ....................
10898 MW154754.1 |Severe acute respiratory syndrome coro    ....................  ....................  ....................  ....................  ....................  ....................
10899 MW154755.1 |Severe acute respiratory syndrome coro    ....................  ....................  ....................  ....................  ....................  ....................
10900 MW154756.1 |Severe acute respiratory syndrome coro    ....................  ....................  ....................  ....................  ....................  ....................
10901 MW154758.1 |Severe acute respiratory syndrome coro    ....................  ....................  ....................  ....................  ....................  ....................
10902 MW154760.1 |Severe acute respiratory syndrome coro    ....................  ....................  ....................  ....................  ....................  ....................
10903 MW154762.1 |Severe acute respiratory syndrome coro    ....................  ....................  ....................  ....................  ....................  ....................
10904 MW154763.1 |Severe acute respiratory syndrome coro    ....................  ....................  ....................  ....................  ....................  ....................
10905 MW154764.1 |Severe acute respiratory syndrome coro    ....................  ....................  ....................  ....................  ....................  ....................
10906 MW154765.1 |Severe acute respiratory syndrome coro    ....................  ....................  ....................  ....................  ....................  ....................
10907 MW154766.1 |Severe acute respiratory syndrome coro    ....................  ....................  ....................  ....................  ....................  ....................
10908 MW154767.1 |Severe acute respiratory syndrome coro    ....................  ....................  ....................  ....................  ....................  ....................
10909 MW154768.1 |Severe acute respiratory syndrome coro    ....................  ....................  ....................  ....................  ....................  ....................
10910 MW154770.1 |Severe acute respiratory syndrome coro    ....................  ....................  ....................  ....................  ....................  ....................
10911 MW154772.1 |Severe acute respiratory syndrome coro    ....................  ....................  ....................  ....................  ....................  ....................
10912 MW154773.1 |Severe acute respiratory syndrome coro    ....................  ....................  ....................  ....................  ....................  ....................
10913 MW154774.1 |Severe acute respiratory syndrome coro    ....................  ....................  ....................  ....................  ....................  ....................
10914 MW154775.1 |Severe acute respiratory syndrome coro    ....................  ....................  ....................  ....................  ....................  ....................
10915 MW154776.1 |Severe acute respiratory syndrome coro    ....................  ....................  ....................  ....................  ....................  ....................
10916 MW154777.1 |Severe acute respiratory syndrome coro    ....................  ....................  ....................  ....................  ....................  ....................
10917 MW154778.1 |Severe acute respiratory syndrome coro    ....................  ....................  ....................  ....................  ....................  ....................
10918 MW154779.1 |Severe acute respiratory syndrome coro    ....................  ....................  ....................  ....................  ....................  ....................
10919 MW154780.1 |Severe acute respiratory syndrome coro    ....................  ....................  ....................  ....................  ....................  ....................
10920 MW154781.1 |Severe acute respiratory syndrome coro    ....................  ....................  ....................  ....................  ....................  ....................
10921 MW154782.1 |Severe acute respiratory syndrome coro    ....................  ....................  ....................  ....................  ....................  ....................
10922 MW154783.1 |Severe acute respiratory syndrome coro    ....................  ....................  ....................  ....................  ....................  ....................
10923 MW154784.1 |Severe acute respiratory syndrome coro    ....................  ....................  ....................  ....................  ....................  ....................
10924 MW154785.1 |Severe acute respiratory syndrome coro    ....................  ....................  ....................  ....................  ....................  ....................
10925 MW154786.1 |Severe acute respiratory syndrome coro    ....................  ....................  ....................  ....................  ....................  ....................
10926 MW154787.1 |Severe acute respiratory syndrome coro    ....................  ....................  ....................  ....................  ....................  ....................
10927 MW154788.1 |Severe acute respiratory syndrome coro    ....................  ....................  ....................  ....................  ....................  ....................
10928 MW154790.1 |Severe acute respiratory syndrome coro    ....................  ....................  ....................  ....................  ....................  ....................
10929 MW154791.1 |Severe acute respiratory syndrome coro    ....................  ....................  ....................  ....................  ....................  ....................
10930 MW154793.1 |Severe acute respiratory syndrome coro    ....................  ....................  ....................  ....................  ....................  ....................
10931 MW154795.1 |Severe acute respiratory syndrome coro    ....................  ....................  ....................  ....................  ....................  ....................
10932 MW154796.1 |Severe acute respiratory syndrome coro    ....................  ....................  ....................  ....................  ....................  ....................
10933 MW154798.1 |Severe acute respiratory syndrome coro    ....................  ....................  ....................  ....................  ....................  ....................
10934 MW154799.1 |Severe acute respiratory syndrome coro    ....................  ....................  ....................  ....................  ....................  ....................
10935 MW154800.1 |Severe acute respiratory syndrome coro    ....................  ....................  ....................  ....................  ....................  ....................
```

```
10936 MW154801.1 |Severe acute respiratory syndrome coro    ....................  ....................  .....................  ....................  ....................  ....................
10937 MW154802.1 |Severe acute respiratory syndrome coro    ....................  ....................  .....................  ....................  ....................  ....................
10938 MW154804.1 |Severe acute respiratory syndrome coro    ....................  ....................  .....................  ....................  ....................  ....................
10939 MW154805.1 |Severe acute respiratory syndrome coro    ....................  ....................  .....................  ....................  ....................  ....................
10940 MW154806.1 |Severe acute respiratory syndrome coro    ....................  ....................  .....................  ....................  ....................  ....................
10941 MW154808.1 |Severe acute respiratory syndrome coro    ....................  ....................  .....................  ....................  ....................  ....................
10942 MW154809.1 |Severe acute respiratory syndrome coro    ....................  ....................  .....................  ....................  ....................  ....................
10943 MW154810.1 |Severe acute respiratory syndrome coro    ....................  ....................  .....................  ....................  ....................  ....................
10944 MW154811.1 |Severe acute respiratory syndrome coro    ....................  ....................  .....................  ....................  ....................  ....................
10945 MW154812.1 |Severe acute respiratory syndrome coro    ....................  ....................  .....................  ....................  ....................  ....................
10946 MW154813.1 |Severe acute respiratory syndrome coro    ....................  ....................  .....................  ....................  ....................  ....................
10947 MW154814.1 |Severe acute respiratory syndrome coro    ....................  ....................  .....................  ....................  ....................  ....................
10948 MW154816.1 |Severe acute respiratory syndrome coro    ....................  ....................  .....................  ....................  ....................  ....................
10949 MW154817.1 |Severe acute respiratory syndrome coro    ....................  ....................  .....................  ....................  ....................  ....................
10950 MW154818.1 |Severe acute respiratory syndrome coro    ....................  ....................  .....................  ....................  ....................  ....................
10951 MW154819.1 |Severe acute respiratory syndrome coro    ....................  ....................  .....................  ....................  ....................  ....................
10952 MW154820.1 |Severe acute respiratory syndrome coro    ....................  ....................  .....................  ....................  ....................  ....................
10953 MW154821.1 |Severe acute respiratory syndrome coro    ....................  ....................  .....................  ....................  ....................  ....................
10954 MW154823.1 |Severe acute respiratory syndrome coro    ....................  ....................  .....................  ....................  ....................  ....................
10955 MW154824.1 |Severe acute respiratory syndrome coro    ....................  ....................  .....................  ....................  ....................  ....................
10956 MW154827.1 |Severe acute respiratory syndrome coro    ....................  ....................  .....................  ....................  ....................  ....................
10957 MW154828.1 |Severe acute respiratory syndrome coro    ....................  ....................  .....................  ....................  ....................  ....................
10958 MW154829.1 |Severe acute respiratory syndrome coro    ....................  ....................  .....................  ....................  ....................  ....................
10959 MW154831.1 |Severe acute respiratory syndrome coro    ....................  ....................  .....................  ....................  ....................  ....................
10960 MW154832.1 |Severe acute respiratory syndrome coro    ....................  ....................  .....................  ....................  ....................  ....................
10961 MW154833.1 |Severe acute respiratory syndrome coro    ....................  ....................  .....................  ....................  ....................  ....................
10962 MW154834.1 |Severe acute respiratory syndrome coro    ....................  ....................  .....................  ....................  ....................  ....................
10963 MW154835.1 |Severe acute respiratory syndrome coro    ....................  ....................  .....................  ....................  ....................  ....................
10964 MW154837.1 |Severe acute respiratory syndrome coro    ....................  ....................  .....................  ....................  ....................  ....................
10965 MW154838.1 |Severe acute respiratory syndrome coro    ....................  ....................  .....................  ....................  ....................  ....................
10966 MW154840.1 |Severe acute respiratory syndrome coro    ....................  ....................  .....................  ....................  ....................  ....................
10967 MW154841.1 |Severe acute respiratory syndrome coro    ....................  ....................  .....................  ....................  ....................  ....................
10968 MW154842.1 |Severe acute respiratory syndrome coro    ....................  ....................  .....................  ....................  ....................  ....................
10969 MW154843.1 |Severe acute respiratory syndrome coro    ....................  ...A................  .....................  ....................  ....................  ....................
10970 MW154844.1 |Severe acute respiratory syndrome coro    ....................  ....................  .....................  ....................  ....................  ....................
10971 MW154845.1 |Severe acute respiratory syndrome coro    ....................  ....................  .....................  ....................  ....................  ....................
10972 MW154846.1 |Severe acute respiratory syndrome coro    ....................  ....................  .....................  ....................  ....................  ....................
10973 MW154847.1 |Severe acute respiratory syndrome coro    ....................  ....................  .....................  ....................  ....................  ....................
10974 MW154849.1 |Severe acute respiratory syndrome coro    ....................  ....................  .....................  ....................  ....................  ....................
10975 MW154851.1 |Severe acute respiratory syndrome coro    ....................  ....................  .....................  ....................  ....................  ....................
10976 MW154852.1 |Severe acute respiratory syndrome coro    ....................  ....................  .....................  ....................  ....................  ....................
10977 MW154854.1 |Severe acute respiratory syndrome coro    ....................  ....................  .....................  ....................  ....................  ....................
10979 MW154856.1 |Severe acute respiratory syndrome coro    ....................  ....................  .....................  ....................  ....................  ....................
10980 MW154857.1 |Severe acute respiratory syndrome coro    ....................  ....................  .....................  ....................  ....................  ....................
10981 MW154858.1 |Severe acute respiratory syndrome coro    ....................  ....................  .....................  ....................  ....................  ....................
10983 MW154860.1 |Severe acute respiratory syndrome coro    ....................  ....................  .....................  ....................  ....................  ....................
10984 MW154863.1 |Severe acute respiratory syndrome coro    ....................  ....................  .....................  ....................  ....................  ....................
10985 MW154864.1 |Severe acute respiratory syndrome coro    ....................  ....................  .....................  ....................  ....................  ....................
10986 MW154865.1 |Severe acute respiratory syndrome coro    ....................  ....................  .....................  ....................  ....................  ....................
10987 MW154866.1 |Severe acute respiratory syndrome coro    ....................  ....................  .....................  ....................  ....................  ....................
10988 MW154867.1 |Severe acute respiratory syndrome coro    ....................  ....................  .....................  ....................  ....................  ....................
10989 MW154869.1 |Severe acute respiratory syndrome coro    ....................  ....................  .....................  ....................  ....................  ....................
10990 MW154870.1 |Severe acute respiratory syndrome coro    ....................  ....................  .....................  ....................  ....................  ....................
10991 MW154871.1 |Severe acute respiratory syndrome coro    ....................  ....................  .....................  ....................  ....................  ....................
10992 MW154872.1 |Severe acute respiratory syndrome coro    ....................  ....................  .....................  ....................  ....................  ....................
10993 MW154874.1 |Severe acute respiratory syndrome coro    ....................  ....................  .....................  ....................  ....................  ....................
10994 MW154875.1 |Severe acute respiratory syndrome coro    ....................  ....................  .....................  ....................  ....................  ....................
10995 MW154876.1 |Severe acute respiratory syndrome coro    ....................  ....................  .....................  ....................  ....................  ....................
10996 MW154877.1 |Severe acute respiratory syndrome coro    ....................  ....................  .....................  ....................  ....................  ....................
10997 MW154878.1 |Severe acute respiratory syndrome coro    ....................  ....................  .....................  ....................  ....................  ....................
10998 MW154879.1 |Severe acute respiratory syndrome coro    ....................  ....................  .....................  ....................  ....................  ....................
10999 MW154881.1 |Severe acute respiratory syndrome coro    ....................  ....................  .....................  ....................  ....................  ....................
11000 MW154883.1 |Severe acute respiratory syndrome coro    ....................  ....................  .....................  ....................  ....................  ....................
11001 MW154884.1 |Severe acute respiratory syndrome coro    ....................  ....................  .....................  ....................  ....................  ....................
11002 MW154885.1 |Severe acute respiratory syndrome coro    ....................  ....................  .....................  ....................  ....................  ....................
11003 MW154887.1 |Severe acute respiratory syndrome coro    ....................  ....................  .....................  ....................  ....................  ....................
11004 MW154888.1 |Severe acute respiratory syndrome coro    ....................  ....................  .....................  ....................  ....................  ....................
11005 MW154889.1 |Severe acute respiratory syndrome coro    ....................  ....................  .....................  ....................  ....................  ....................
11006 MW154891.1 |Severe acute respiratory syndrome coro    ....................  ....................  .....................  ....................  ....................  ....................
11007 MW154892.1 |Severe acute respiratory syndrome coro    ....................  ....................  .....................  ....................  ....................  ....................
11008 MW154893.1 |Severe acute respiratory syndrome coro    ....................  ....................  .....................  ....................  ....................  ....................
11009 MW154894.1 |Severe acute respiratory syndrome coro    ....................  ....................  .....................  ....................  ....................  ....................
11010 MW154895.1 |Severe acute respiratory syndrome coro    ....................  ....................  .....................  ....................  ....................  ....................
11011 MW154897.1 |Severe acute respiratory syndrome coro    ....................  ....................  .....................  ....................  ....................  ....................
11012 MW154898.1 |Severe acute respiratory syndrome coro    ....................  ....................  .....................  ....................  ....................  ....................
11013 MW154900.1 |Severe acute respiratory syndrome coro    ....................  ....................  .....................  ....................  ....................  ....................
11014 MW154901.1 |Severe acute respiratory syndrome coro    ....................  ....................  .....................  ....................  ....................  ....................
11015 MW154903.1 |Severe acute respiratory syndrome coro    ....................  ....................  .....................  ....................  ....................  ....................
```

11016 MW154904.1 |Severe acute respiratory syndrome coro .................... .................... .................... .................... .................... ....................
11017 MW154905.1 |Severe acute respiratory syndrome coro .................... .................... .................... .................... .................... ....................
11018 MW154906.1 |Severe acute respiratory syndrome coro .................... .................... .................... .................... .................... ....................
11019 MW154907.1 |Severe acute respiratory syndrome coro .................... .................... .................... .................... .................... ....................
11020 MW154908.1 |Severe acute respiratory syndrome coro .................... .................... .................... .................... .................... ....................
11021 MW154909.1 |Severe acute respiratory syndrome coro .................... .................... .................... .................... .................... ....................
11022 MW154910.1 |Severe acute respiratory syndrome coro .................... .................... .................... .................... .................... ....................
11023 MW154911.1 |Severe acute respiratory syndrome coro .................... .................... .................... .................... .................... ....................
11024 MW154913.1 |Severe acute respiratory syndrome coro .................... .................... .................... .................... .................... ....................
11025 MW154915.1 |Severe acute respiratory syndrome coro .................... .................... .................... .................... .................... ....................
11026 MW154917.1 |Severe acute respiratory syndrome coro .................... .................... .................... .................... .................... ....................
11027 MW154920.1 |Severe acute respiratory syndrome coro .................... .................... .................... .................... .................... ....................
11028 MW154923.1 |Severe acute respiratory syndrome coro .................... .................... .................... .................... .................... ....................
11029 MW154926.1 |Severe acute respiratory syndrome coro .................... .................... .................... .................... .................... ....................
11030 MW154927.1 |Severe acute respiratory syndrome coro .................... .................... .................... .................... .................... ....................
11031 MW154928.1 |Severe acute respiratory syndrome coro .................... .................... .................... .................... .................... ....................
11032 MW154929.1 |Severe acute respiratory syndrome coro .................... .................... .................... .................... .................... ....................
11033 MW154930.1 |Severe acute respiratory syndrome coro .................... .................... .................... .................... .................... ....................
11034 MW154931.1 |Severe acute respiratory syndrome coro .................... .................... .................... .................... .................... ....................
11035 MW154932.1 |Severe acute respiratory syndrome coro .................... .................... .................... .................... .................... ....................
11036 MW154934.1 |Severe acute respiratory syndrome coro .................... .................... .................... .................... .................... ....................
11037 MW154935.1 |Severe acute respiratory syndrome coro .................... .................... .................... .................... .................... ....................
11039 MW154938.1 |Severe acute respiratory syndrome coro .................... .................... .................... .................... .................... ....................
11040 MW154939.1 |Severe acute respiratory syndrome coro .................... .................... .................... .................... .................... ....................
11041 MW154940.1 |Severe acute respiratory syndrome coro .................... .................... .................... .................... .................... ....................
11042 MW154941.1 |Severe acute respiratory syndrome coro .................... .................... .................... .................... .................... ....................
11043 MW154942.1 |Severe acute respiratory syndrome coro .................... .................... .................... .................... .................... ....................
11044 MW154944.1 |Severe acute respiratory syndrome coro .................... .................... .................... .................... .................... ....................
11045 MW154946.1 |Severe acute respiratory syndrome coro .................... .................... .................... .................... .................... ....................
11046 MW154947.1 |Severe acute respiratory syndrome coro .................... .................... .................... .................... .................... ....................
11047 MW154949.1 |Severe acute respiratory syndrome coro .................... .................... .................... .................... .................... ....................
11048 MW154951.1 |Severe acute respiratory syndrome coro .................... ......A............. .................... .................... .................... ....................
11049 MW154952.1 |Severe acute respiratory syndrome coro .................... .................... .................... .................... .................... ....................
11050 MW154953.1 |Severe acute respiratory syndrome coro .................... .................... .................... .................... .................... ....................
11051 MW154955.1 |Severe acute respiratory syndrome coro .................... .................... .................... .................... .................... ....................
11052 MW154957.1 |Severe acute respiratory syndrome coro .................... .................... .................... .................... .................... ....................
11053 MW154958.1 |Severe acute respiratory syndrome coro .................... .................... .................... .................... .................... ....................
11054 MW154959.1 |Severe acute respiratory syndrome coro .................... .................... .................... .................... .................... ....................
11055 MW154960.1 |Severe acute respiratory syndrome coro .................... .................... .................... .................... .................... ....................
11056 MW154961.1 |Severe acute respiratory syndrome coro .................... .................... .................... .................... .................... ....................
11057 MW154963.1 |Severe acute respiratory syndrome coro .................... .................... ...C................ .................... .................... ....................
11058 MW154964.1 |Severe acute respiratory syndrome coro .................... .................... .................... .................... .................... ....................
11059 MW154965.1 |Severe acute respiratory syndrome coro .................... .................... .................... .................... .................... ....................
11060 MW154966.1 |Severe acute respiratory syndrome coro .................... .................... .................... .................... .................... ....................
11061 MW154967.1 |Severe acute respiratory syndrome coro .................... .................... .................... .................... .................... ....................
11062 MW154968.1 |Severe acute respiratory syndrome coro .................... .................... .................... .................... .................... ....................
11063 MW154969.1 |Severe acute respiratory syndrome coro .................... .................... .................... .................... .................... ....................
11064 MW154970.1 |Severe acute respiratory syndrome coro .................... .................... .................... .................... .................... ....................
11065 MW154971.1 |Severe acute respiratory syndrome coro .................... .................... .................... .................... .................... ....................
11066 MW154972.1 |Severe acute respiratory syndrome coro .................... .................... .................... .................... .................... ....................
11067 MW154973.1 |Severe acute respiratory syndrome coro .................... .................... .................... .................... .................... ....................
11068 MW154975.1 |Severe acute respiratory syndrome coro .................... .................... .................... .................... .................... ....................
11069 MW154976.1 |Severe acute respiratory syndrome coro .................... .................... .................... .................... .................... ....................
11070 MW154977.1 |Severe acute respiratory syndrome coro .................... .................... ...C................ .................... .................... ....................
11071 MW154978.1 |Severe acute respiratory syndrome coro .................... .................... .................... .................... .................... ....................
11072 MW154979.1 |Severe acute respiratory syndrome coro .................... .................... .................... .................... .................... ....................
11073 MW154980.1 |Severe acute respiratory syndrome coro .................... .................... .................... .................... .................... ....................
11074 MW154981.1 |Severe acute respiratory syndrome coro .................... .................... .................... .................... .................... ....................
11075 MW154982.1 |Severe acute respiratory syndrome coro .................... .................... .................... .................... .................... ....................
11076 MW154983.1 |Severe acute respiratory syndrome coro .................... .................... .................... .................... .................... ....................
11077 MW154984.1 |Severe acute respiratory syndrome coro .................... .................... .................... .................... .................... ....................
11078 MW154985.1 |Severe acute respiratory syndrome coro .................... .................... .................... .................... .................... ....................
11079 MW154986.1 |Severe acute respiratory syndrome coro .................... .................... .................... .................... .................... ....................
11080 MW154987.1 |Severe acute respiratory syndrome coro .................... .................... .................... .................... .................... ....................
11081 MW154988.1 |Severe acute respiratory syndrome coro .................... .................... .................... .................... .................... ....................
11082 MW154989.1 |Severe acute respiratory syndrome coro .................... .................... .................... .................... .................... ....................
11083 MW154990.1 |Severe acute respiratory syndrome coro .................... .................... .................... .................... .................... ....................
11084 MW154991.1 |Severe acute respiratory syndrome coro .................... .................... .................... .................... .................... ....................
11085 MW154993.1 |Severe acute respiratory syndrome coro .................... .................... .................... .................... .................... ....................
11086 MW154995.1 |Severe acute respiratory syndrome coro .................... .................... .................... .................... .................... ....................
11087 MW154996.1 |Severe acute respiratory syndrome coro .................... .................... .................... .................... .................... ....................
11088 MW154997.1 |Severe acute respiratory syndrome coro .................... .................... .................... .................... .................... ....................
11089 MW154999.1 |Severe acute respiratory syndrome coro .................... .................... .................... .................... .................... ....................
11090 MW155001.1 |Severe acute respiratory syndrome coro .................... .................... .................... .................... .................... ....................
11091 MW155002.1 |Severe acute respiratory syndrome coro .................... .................... .................... .................... .................... ....................
11092 MW155003.1 |Severe acute respiratory syndrome coro .................... .................... .................... .................... .................... ....................
11093 MW155004.1 |Severe acute respiratory syndrome coro .................... .................... .................... .................... .................... ....................
11094 MW155005.1 |Severe acute respiratory syndrome coro .................... .................... .................... .................... .................... ....................

| | | | | | | | | | |
|---|---|---|---|---|---|---|---|---|---|
| 11095 | MW155006.1 | Severe acute respiratory syndrome coro | .................... | .................... | .................... | .................... | .................... | .................... | |
| 11098 | MW155009.1 | Severe acute respiratory syndrome coro | .................... | .................... | .................... | .................... | .................... | .................... | |
| 11099 | MW155010.1 | Severe acute respiratory syndrome coro | .................... | .................... | .................... | .................... | NNN................. | .................... | 1 |
| 11100 | MW155011.1 | Severe acute respiratory syndrome coro | .................... | .................... | .................... | .................... | .................... | .................... | |
| 11101 | MW155012.1 | Severe acute respiratory syndrome coro | .................... | .................... | .................... | .................... | .................... | .................... | |
| 11102 | MW155014.1 | Severe acute respiratory syndrome coro | .................... | .................... | .................... | .................... | .................... | .................... | |
| 11103 | MW155015.1 | Severe acute respiratory syndrome coro | .................... | .................... | .................... | .................... | .................... | .................... | |
| 11104 | MW155016.1 | Severe acute respiratory syndrome coro | .................... | .................... | .................... | .................... | .................... | .................... | |
| 11105 | MW155017.1 | Severe acute respiratory syndrome coro | .................... | .................... | .................... | .................... | .................... | .................... | |
| 11106 | MW155018.1 | Severe acute respiratory syndrome coro | .................... | .................... | .................... | .................... | .................... | .................... | |
| 11108 | MW155022.1 | Severe acute respiratory syndrome coro | .................... | .................... | .................... | .................... | .................... | .................... | |
| 11110 | MW155029.1 | Severe acute respiratory syndrome coro | .................... | .................... | .................... | .................... | .................... | .................... | |
| 11111 | MW155030.1 | Severe acute respiratory syndrome coro | .................... | .................... | .................... | .................... | .................... | .................... | |
| 11112 | MW155031.1 | Severe acute respiratory syndrome coro | .................... | .................... | .................... | .................... | .................... | .................... | |
| 11113 | MW155032.1 | Severe acute respiratory syndrome coro | .................... | .................... | .................... | .................... | .................... | .................... | |
| 11114 | MW155034.1 | Severe acute respiratory syndrome coro | .................... | .................... | .................... | .................... | .................... | .................... | |
| 11115 | MW155036.1 | Severe acute respiratory syndrome coro | .................... | .................... | .................... | .................... | .................... | .................... | |
| 11116 | MW155037.1 | Severe acute respiratory syndrome coro | .................... | .................... | .................... | .................... | .................... | .................... | |
| 11117 | MW155038.1 | Severe acute respiratory syndrome coro | .................... | .................... | .................... | .................... | .................... | .................... | |
| 11118 | MW155039.1 | Severe acute respiratory syndrome coro | .................... | .................... | .................... | .................... | .................... | .................... | |
| 11119 | MW155040.1 | Severe acute respiratory syndrome coro | .................... | .................... | .................... | .................... | .................... | .................... | |
| 11120 | MW155041.1 | Severe acute respiratory syndrome coro | .................... | .................... | .................... | .................... | .................... | .................... | |
| 11121 | MW155042.1 | Severe acute respiratory syndrome coro | .................... | .................... | .................... | .................... | .................... | .................... | |
| 11122 | MW155043.1 | Severe acute respiratory syndrome coro | .................... | .................... | .................... | .................... | .................... | .................... | |
| 11123 | MW155044.1 | Severe acute respiratory syndrome coro | .................... | .................... | .................... | .................... | .................... | .................... | |
| 11124 | MW155045.1 | Severe acute respiratory syndrome coro | .................... | .................... | .................... | .................... | .................... | .................... | |
| 11125 | MW155046.1 | Severe acute respiratory syndrome coro | .................... | .................... | .................... | .................... | .................... | .................... | |
| 11126 | MW155048.1 | Severe acute respiratory syndrome coro | .................... | .................... | .................... | .................... | .................... | .................... | |
| 11127 | MW155051.1 | Severe acute respiratory syndrome coro | .................... | .................... | .................... | .................... | .................... | .................... | |
| 11128 | MW155053.1 | Severe acute respiratory syndrome coro | .................... | .................... | .................... | .................... | .................... | .................... | |
| 11129 | MW155055.1 | Severe acute respiratory syndrome coro | .................... | .................... | .................... | .................... | .................... | .................... | |
| 11130 | MW155056.1 | Severe acute respiratory syndrome coro | .................... | .................... | .................... | .................... | .................... | .................... | |
| 11131 | MW155058.1 | Severe acute respiratory syndrome coro | .................... | .................... | .................... | .................... | .................... | .................... | |
| 11132 | MW155059.1 | Severe acute respiratory syndrome coro | .................... | .................... | .................... | .................... | .................... | .................... | |
| 11133 | MW155061.1 | Severe acute respiratory syndrome coro | .................... | .................... | .................... | .................... | .................... | .................... | |
| 11134 | MW155062.1 | Severe acute respiratory syndrome coro | .................... | .................... | .................... | .................... | .................... | .................... | |
| 11135 | MW155064.1 | Severe acute respiratory syndrome coro | .................... | .................... | .................... | .................... | .................... | .................... | |
| 11136 | MW155065.1 | Severe acute respiratory syndrome coro | .................... | .................... | .................... | .................... | .................... | .................... | |
| 11137 | MW155066.1 | Severe acute respiratory syndrome coro | .................... | .................... | .................... | .................... | .................... | .................... | |
| 11138 | MW155067.1 | Severe acute respiratory syndrome coro | .................... | .................... | .................... | .................... | .................... | .................... | |
| 11139 | MW155070.1 | Severe acute respiratory syndrome coro | .................... | .................... | .................... | .................... | .................... | .................... | |
| 11140 | MW155071.1 | Severe acute respiratory syndrome coro | .................... | .................... | .................... | .................... | .................... | .................... | |
| 11141 | MW155072.1 | Severe acute respiratory syndrome coro | .................... | .................... | .................... | .................... | .................... | .................... | |
| 11142 | MW155074.1 | Severe acute respiratory syndrome coro | .................... | .................... | .................... | .................... | .................... | .................... | |
| 11143 | MW155075.1 | Severe acute respiratory syndrome coro | .................... | .................... | .................... | .................... | .................... | .................... | |
| 11144 | MW155077.1 | Severe acute respiratory syndrome coro | .................... | .................... | .................... | .................... | .................... | .................... | |
| 11145 | MW155079.1 | Severe acute respiratory syndrome coro | .................... | .................... | .................... | .................... | .................... | .................... | |
| 11146 | MW155080.1 | Severe acute respiratory syndrome coro | .................... | .................... | .................... | .................... | .................... | .................... | |
| 11147 | MW155081.1 | Severe acute respiratory syndrome coro | .................... | .................... | .................... | .................... | .................... | .................... | |
| 11148 | MW155082.1 | Severe acute respiratory syndrome coro | .................... | .................... | .................... | .................... | .................... | .................... | |
| 11149 | MW155083.1 | Severe acute respiratory syndrome coro | .................... | .................... | .................... | .................... | .................... | .................... | |
| 11150 | MW155084.1 | Severe acute respiratory syndrome coro | .................... | .................... | .................... | .................... | .................... | .................... | |
| 11151 | MW155085.1 | Severe acute respiratory syndrome coro | .................... | .................... | .................... | .................... | .................... | .................... | |
| 11152 | MW155088.1 | Severe acute respiratory syndrome coro | .................... | .................... | .................... | .................... | .................... | .................... | |
| 11153 | MW155090.1 | Severe acute respiratory syndrome coro | .................... | .................... | .................... | .................... | .................... | .................... | |
| 11154 | MW155091.1 | Severe acute respiratory syndrome coro | .................... | .................... | .................... | .................... | .................... | .................... | |
| 11155 | MW155093.1 | Severe acute respiratory syndrome coro | .................... | .................... | .................... | .................... | .................... | .................... | |
| 11156 | MW155094.1 | Severe acute respiratory syndrome coro | .................... | .................... | .................... | .................... | .................... | .................... | |
| 11157 | MW155096.1 | Severe acute respiratory syndrome coro | .................... | .................... | .................... | .................... | .................... | .................... | |
| 11158 | MW155097.1 | Severe acute respiratory syndrome coro | .................... | .................... | .................... | .................... | .................... | .................... | |
| 11159 | MW155098.1 | Severe acute respiratory syndrome coro | .................... | .................... | .................... | .................... | .................... | .................... | |
| 11160 | MW155099.1 | Severe acute respiratory syndrome coro | .................... | .................... | .................... | .................... | .................... | .................... | |
| 11161 | MW155102.1 | Severe acute respiratory syndrome coro | .................... | .................... | .................... | .................... | .................... | .................... | |
| 11162 | MW155103.1 | Severe acute respiratory syndrome coro | .................... | .................... | .................... | .................... | .................... | .................... | |
| 11163 | MW155104.1 | Severe acute respiratory syndrome coro | .................... | .................... | .................... | .................... | .................... | .................... | |
| 11164 | MW155106.1 | Severe acute respiratory syndrome coro | .................... | .................... | .................... | .................... | .................... | .................... | |
| 11165 | MW155107.1 | Severe acute respiratory syndrome coro | .................... | .................... | .................... | .................... | .................... | ..................C. | 1 |
| 11166 | MW155109.1 | Severe acute respiratory syndrome coro | .................... | .................... | .................... | .................... | .................... | .................... | |
| 11167 | MW155110.1 | Severe acute respiratory syndrome coro | .................... | .................... | .................... | .................... | .................... | .................... | |
| 11168 | MW155111.1 | Severe acute respiratory syndrome coro | .................... | .................... | .................... | .................... | .................... | .................... | |
| 11169 | MW155112.1 | Severe acute respiratory syndrome coro | .................... | .................... | .................... | .................... | .................... | .................... | |
| 11170 | MW155113.1 | Severe acute respiratory syndrome coro | .................... | .................... | .................... | .................... | .................... | .................... | |
| 11171 | MW155114.1 | Severe acute respiratory syndrome coro | .................... | .................... | .................... | .................... | .................... | .................... | |
| 11172 | MW155115.1 | Severe acute respiratory syndrome coro | .................... | .................... | .................... | .................... | .................... | .................... | |
| 11173 | MW155117.1 | Severe acute respiratory syndrome coro | .................... | .................... | .................... | .................... | .................... | .................... | |
| 11174 | MW155118.1 | Severe acute respiratory syndrome coro | .................... | .................... | .................... | .................... | .................... | .................... | |
| 11175 | MW155120.1 | Severe acute respiratory syndrome coro | .................... | .................... | .................... | .................... | .................... | .................... | |
| 11176 | MW155121.1 | Severe acute respiratory syndrome coro | .................... | .................... | .................... | .................... | .................... | .................... | |

| | | | | | | | | |
|---|---|---|---|---|---|---|---|---|
| 11177 | MW155122.1 | \|Severe acute respiratory syndrome coro | .................... | .................... | .................... | .................... | .................... | .................... |
| 11178 | MW155123.1 | \|Severe acute respiratory syndrome coro | .................... | .................... | .................... | .................... | .................... | .................... |
| 11179 | MW155124.1 | \|Severe acute respiratory syndrome coro | .................... | .................... | .................... | .................... | .................... | .................... |
| 11180 | MW155125.1 | \|Severe acute respiratory syndrome coro | .................... | .................... | .................... | .................... | .................... | .................... |
| 11181 | MW155126.1 | \|Severe acute respiratory syndrome coro | .................... | .................... | .................... | .................... | .................... | .................... |
| 11182 | MW155127.1 | \|Severe acute respiratory syndrome coro | .................... | .................... | .................... | .................... | .................... | .................... |
| 11183 | MW155128.1 | \|Severe acute respiratory syndrome coro | .................... | .................... | .................... | .................... | .................... | .................... |
| 11184 | MW155129.1 | \|Severe acute respiratory syndrome coro | .................... | .................... | .................... | .................... | .................... | .................... |
| 11185 | MW155130.1 | \|Severe acute respiratory syndrome coro | .................... | .................... | .................... | .................... | .................... | .................... |
| 11186 | MW155132.1 | \|Severe acute respiratory syndrome coro | .................... | .................... | .................... | .................... | .................... | .................... |
| 11187 | MW155133.1 | \|Severe acute respiratory syndrome coro | .................... | .................... | .................... | .................... | .................... | .................... |
| 11188 | MW155134.1 | \|Severe acute respiratory syndrome coro | .................... | .................... | .................... | .................... | .................... | .................... |
| 11189 | MW155136.1 | \|Severe acute respiratory syndrome coro | .................... | .................... | .................... | .................... | .................... | .................... |
| 11190 | MW155137.1 | \|Severe acute respiratory syndrome coro | .................... | .................... | .................... | .................... | .................... | .................... |
| 11191 | MW155138.1 | \|Severe acute respiratory syndrome coro | .................... | .................... | .................... | .................... | .................... | .................... |
| 11192 | MW155140.1 | \|Severe acute respiratory syndrome coro | .................... | .................... | .................... | .................... | .................... | .................... |
| 11193 | MW155143.1 | \|Severe acute respiratory syndrome coro | .................... | .................... | .................... | .................... | .................... | .................... |
| 11194 | MW155144.1 | \|Severe acute respiratory syndrome coro | .................... | .................... | .................... | .................... | .................... | .................... |
| 11195 | MW155146.1 | \|Severe acute respiratory syndrome coro | .................... | .................... | .................... | .................... | .................... | .................... |
| 11196 | MW155148.1 | \|Severe acute respiratory syndrome coro | .................... | .................... | .................... | .................... | .................... | .................... |
| 11197 | MW155149.1 | \|Severe acute respiratory syndrome coro | .................... | .................... | .................... | .................... | .................... | .................... |
| 11198 | MW155151.1 | \|Severe acute respiratory syndrome coro | .................... | .................... | .................... | .................... | .................... | .................... |
| 11199 | MW155153.1 | \|Severe acute respiratory syndrome coro | .................... | .................... | .................... | .................... | .................... | .................... |
| 11200 | MW155154.1 | \|Severe acute respiratory syndrome coro | .................... | .................... | .................... | .................... | .................... | .................... |
| 11201 | MW155155.1 | \|Severe acute respiratory syndrome coro | .................... | .................... | .................... | .................... | .................... | .................... |
| 11202 | MW155156.1 | \|Severe acute respiratory syndrome coro | .................... | .................... | .................... | .................... | .................... | .................... |
| 11203 | MW155158.1 | \|Severe acute respiratory syndrome coro | .................... | .................... | .................... | .................... | .................... | .................... |
| 11204 | MW155160.1 | \|Severe acute respiratory syndrome coro | .................... | .................... | .................... | .................... | .................... | .................... |
| 11205 | MW155161.1 | \|Severe acute respiratory syndrome coro | .................... | .................... | .................... | .................... | .................... | .................... |
| 11206 | MW155162.1 | \|Severe acute respiratory syndrome coro | .................... | .................... | .................... | .................... | .................... | .................... |
| 11207 | MW155163.1 | \|Severe acute respiratory syndrome coro | .................... | .................... | .................... | .................... | .................... | .................... |
| 11208 | MW155164.1 | \|Severe acute respiratory syndrome coro | .................... | .................... | .................... | .................... | .................... | .................... |
| 11209 | MW155165.1 | \|Severe acute respiratory syndrome coro | .................... | .................... | .................... | .................... | .................... | .................... |
| 11210 | MW155166.1 | \|Severe acute respiratory syndrome coro | .................... | .................... | .................... | .................... | .................... | .................... |
| 11211 | MW155167.1 | \|Severe acute respiratory syndrome coro | .................... | .................... | .................... | .................... | .................... | .................... |
| 11212 | MW155168.1 | \|Severe acute respiratory syndrome coro | .................... | .................... | .................... | .................... | .................... | .................... |
| 11213 | MW155169.1 | \|Severe acute respiratory syndrome coro | .................... | .................... | .................... | .................... | .................... | .................... |
| 11214 | MW155172.1 | \|Severe acute respiratory syndrome coro | .................... | .................... | .................... | .................... | .................... | .................... |
| 11215 | MW155173.1 | \|Severe acute respiratory syndrome coro | .................... | .................... | .................... | .................... | .................... | .................... |
| 11216 | MW155174.1 | \|Severe acute respiratory syndrome coro | .................... | .................... | .................... | .................... | .................... | .................... |
| 11217 | MW155175.1 | \|Severe acute respiratory syndrome coro | .................... | .................... | .................... | .................... | .................... | .................... |
| 11218 | MW155178.1 | \|Severe acute respiratory syndrome coro | .................... | .................... | .................... | .................... | .................... | .................... |
| 11219 | MW155179.1 | \|Severe acute respiratory syndrome coro | .................... | .................... | .................... | .................... | .................... | .................... |
| 11220 | MW155180.1 | \|Severe acute respiratory syndrome coro | .................... | .................... | .................... | .................... | .................... | .................... |
| 11221 | MW155182.1 | \|Severe acute respiratory syndrome coro | .................... | .................... | .................... | .................... | .................... | .................... |
| 11222 | MW155183.1 | \|Severe acute respiratory syndrome coro | .................... | .................... | .................... | .................... | .................... | .................... |
| 11223 | MW155185.1 | \|Severe acute respiratory syndrome coro | .................... | .................... | .................... | .................... | .................... | .................... |
| 11224 | MW155186.1 | \|Severe acute respiratory syndrome coro | .................... | .................... | .................... | .................... | .................... | .................... |
| 11225 | MW155187.1 | \|Severe acute respiratory syndrome coro | .................... | .................... | .................... | .................... | .................... | .................... |
| 11226 | MW155188.1 | \|Severe acute respiratory syndrome coro | .................... | .................... | .................... | .................... | .................... | .................... |
| 11227 | MW155190.1 | \|Severe acute respiratory syndrome coro | .................... | .................... | .................... | .................... | .................... | .................... |
| 11228 | MW155193.1 | \|Severe acute respiratory syndrome coro | .................... | .................... | .................... | .................... | .................... | .................... |
| 11229 | MW155194.1 | \|Severe acute respiratory syndrome coro | .................... | .................... | .................... | .................... | .................... | .................... |
| 11230 | MW155195.1 | \|Severe acute respiratory syndrome coro | .................... | .................... | .................... | .................... | .................... | .................... |
| 11231 | MW155197.1 | \|Severe acute respiratory syndrome coro | .................... | .................... | .................... | .................... | .................... | .................... |
| 11232 | MW155198.1 | \|Severe acute respiratory syndrome coro | .................... | .................... | .................... | .................... | .................... | .................... |
| 11233 | MW155199.1 | \|Severe acute respiratory syndrome coro | .................... | .................... | .................... | .................... | .................... | .................... |
| 11234 | MW155200.1 | \|Severe acute respiratory syndrome coro | .................... | .................... | .................... | .................... | .................... | .................... |
| 11235 | MW155202.1 | \|Severe acute respiratory syndrome coro | .................... | .................... | .................... | .................... | .................... | .................... |
| 11236 | MW155203.1 | \|Severe acute respiratory syndrome coro | .................... | .................... | .................... | .................... | .................... | .................... |
| 11237 | MW155206.1 | \|Severe acute respiratory syndrome coro | .................... | .................... | .................... | .................... | .................... | .................... |
| 11238 | MW155207.1 | \|Severe acute respiratory syndrome coro | .................... | .................... | .................... | .................... | .................... | .................... |
| 11239 | MW155208.1 | \|Severe acute respiratory syndrome coro | .................... | .................... | .................... | .................... | .................... | .................... |
| 11240 | MW155209.1 | \|Severe acute respiratory syndrome coro | .................... | .................... | .................... | .................... | .................... | .................... |
| 11241 | MW155212.1 | \|Severe acute respiratory syndrome coro | .................... | .................... | .................... | .................... | .................... | .................... |
| 11242 | MW155213.1 | \|Severe acute respiratory syndrome coro | .................... | .................... | .................... | .................... | .................... | .................... |
| 11243 | MW155214.1 | \|Severe acute respiratory syndrome coro | .................... | .................... | .................... | .................... | .................... | .................... |
| 11244 | MW155215.1 | \|Severe acute respiratory syndrome coro | .................... | .................... | .................... | .................... | .................... | .................... |
| 11245 | MW155217.1 | \|Severe acute respiratory syndrome coro | .................... | .................... | .................... | .................... | .................... | .................... |
| 11246 | MW155219.1 | \|Severe acute respiratory syndrome coro | .................... | .................... | .................... | .................... | .................... | .................... |
| 11247 | MW155220.1 | \|Severe acute respiratory syndrome coro | .................... | .................... | .................... | .................... | .................... | .................... |
| 11248 | MW155221.1 | \|Severe acute respiratory syndrome coro | .................... | .................... | .................... | .................... | .................... | .................... |
| 11249 | MW155222.1 | \|Severe acute respiratory syndrome coro | .................... | .................... | .................... | .................... | .................... | .................... |
| 11250 | MW155223.1 | \|Severe acute respiratory syndrome coro | .................... | .................... | .................... | .................... | .................... | .................... |
| 11251 | MW155224.1 | \|Severe acute respiratory syndrome coro | .................... | .................... | .................... | .................... | .................... | .................... |
| 11252 | MW155226.1 | \|Severe acute respiratory syndrome coro | .................... | .................... | .................... | .................... | .................... | .................... |
| 11253 | MW155227.1 | \|Severe acute respiratory syndrome coro | .................... | .................... | .................... | .................... | .................... | .................... |
| 11254 | MW155229.1 | \|Severe acute respiratory syndrome coro | .................... | .................... | .................... | .................... | .................... | .................... |

11255 MW155230.1 |Severe acute respiratory syndrome coro .................... .................... ..................... .................... .................... ....................
11256 MW155231.1 |Severe acute respiratory syndrome coro .................... .................... ..................... .................... .................... ....................
11257 MW155232.1 |Severe acute respiratory syndrome coro .................... .................... ..................... .................... .................... ....................
11258 MW155234.1 |Severe acute respiratory syndrome coro .................... .................... ..................... .................... .................... ....................
11259 MW155235.1 |Severe acute respiratory syndrome coro .................... .................... ..................... .................... .................... ....................
11260 MW155237.1 |Severe acute respiratory syndrome coro .................... .................... ..................... .................... .................... ....................
11261 MW155239.1 |Severe acute respiratory syndrome coro .................... .................... ..................... .................... .................... ....................
11262 MW155240.1 |Severe acute respiratory syndrome coro .................... .................... ..................... .................... .................... ....................
11263 MW155242.1 |Severe acute respiratory syndrome coro .................... .................... ..................... .................... .................... ....................
11264 MW155243.1 |Severe acute respiratory syndrome coro .................... .................... ..................... .................... .................... ....................
11265 MW155244.1 |Severe acute respiratory syndrome coro .................... .................... ..................... .................... .................... ....................
11266 MW155245.1 |Severe acute respiratory syndrome coro .................... .................... ..................... .................... .................... ....................
11267 MW155247.1 |Severe acute respiratory syndrome coro .................... .................... ..................... .................... .................... ....................
11268 MW155248.1 |Severe acute respiratory syndrome coro .................... .................... ..................... .................... .................... ....................
11269 MW155249.1 |Severe acute respiratory syndrome coro .................... .................... ..................... .................... .................... ....................
11270 MW155254.1 |Severe acute respiratory syndrome coro .................... .................... ..................... .................... .................... ....................
11271 MW155255.1 |Severe acute respiratory syndrome coro .................... .................... ..................... .................... .................... ....................
11272 MW155256.1 |Severe acute respiratory syndrome coro .................... .................... ..................... .................... .................... ....................
11273 MW155257.1 |Severe acute respiratory syndrome coro .................... .................... ..................... .................... .................... ....................
11274 MW155259.1 |Severe acute respiratory syndrome coro .................... .................... ..................... .................... .................... ....................
11275 MW155260.1 |Severe acute respiratory syndrome coro .................... .................... ..................... .................... .................... ....................
11276 MW155261.1 |Severe acute respiratory syndrome coro .................... .................... ..................... .................... .................... ....................
11277 MW155262.1 |Severe acute respiratory syndrome coro .................... .................... ..................... .................... .................... ....................
11278 MW155263.1 |Severe acute respiratory syndrome coro .................... .................... ..................... .................... .................... ....................
11279 MW155264.1 |Severe acute respiratory syndrome coro .................... .................... ..................... .................... .................... ....................
11280 MW155266.1 |Severe acute respiratory syndrome coro .................... .................... ..................... .................... .................... ....................
11281 MW155268.1 |Severe acute respiratory syndrome coro .................... .................... ..................... .................... .................... ....................
11282 MW155271.1 |Severe acute respiratory syndrome coro .................... .................... ..................... .................... .................... ....................
11283 MW155272.1 |Severe acute respiratory syndrome coro .................... .................... ..................... .................... .................... ....................
11284 MW155273.1 |Severe acute respiratory syndrome coro .................... .................... ..................... .................... .................... ....................
11285 MW155275.1 |Severe acute respiratory syndrome coro .................... .................... ..................... .................... .................... ....................
11286 MW155276.1 |Severe acute respiratory syndrome coro .................... .................... ..................... .................... .................... ....................
11287 MW155278.1 |Severe acute respiratory syndrome coro .................... .................... ..................... .................... .................... ....................
11288 MW155281.1 |Severe acute respiratory syndrome coro .................... .................... ..................... .................... .................... ....................
11289 MW155282.1 |Severe acute respiratory syndrome coro .................... .................... ..................... .................... .................... ....................
11290 MW155283.1 |Severe acute respiratory syndrome coro .................... .................... ..................... .................... .................... ....................
11291 MW155284.1 |Severe acute respiratory syndrome coro .................... .................... ..................... .................... .................... ....................
11292 MW155285.1 |Severe acute respiratory syndrome coro .................... .................... ..................... .................... .................... ....................
11293 MW155286.1 |Severe acute respiratory syndrome coro .................... .................... ..................... .................... .................... ....................
11294 MW155287.1 |Severe acute respiratory syndrome coro .................... .................... ..................... .................... .................... ....................
11295 MW155288.1 |Severe acute respiratory syndrome coro .................... .................... ..................... .................... .................... ....................
11296 MW155290.1 |Severe acute respiratory syndrome coro .................... .................... ..................... .................... .................... ....................
11297 MW155291.1 |Severe acute respiratory syndrome coro .................... .................... ..................... .................... .................... ....................
11298 MW155292.1 |Severe acute respiratory syndrome coro .................... .................... ..................... .................... .................... ....................
11299 MW155295.1 |Severe acute respiratory syndrome coro .................... .................... ..................... .................... .................... ....................
11300 MW155296.1 |Severe acute respiratory syndrome coro .................... .................... ..................... .................... .................... ....................
11301 MW155297.1 |Severe acute respiratory syndrome coro .................... .................... ..................... .................... .................... ....................
11302 MW155299.1 |Severe acute respiratory syndrome coro .................... .................... ..................... .................... .................... ....................
11303 MW155300.1 |Severe acute respiratory syndrome coro .................... .................... ..................... .................... .................... ....................
11304 MW155302.1 |Severe acute respiratory syndrome coro .................... .................... ..................... .................... .................... ....................
11305 MW155303.1 |Severe acute respiratory syndrome coro .................... .................... ..................... .................... .................... ....................
11306 MW155305.1 |Severe acute respiratory syndrome coro .................... .................... ..................... .................... .................... ....................
11307 MW155306.1 |Severe acute respiratory syndrome coro .................... .................... ..................... .................... .................... ....................
11308 MW155307.1 |Severe acute respiratory syndrome coro .................... .................... ..................... .................... .................... ....................
11309 MW155308.1 |Severe acute respiratory syndrome coro .................... .................... ..................... .................... .................... ....................
11310 MW155311.1 |Severe acute respiratory syndrome coro .................... .................... ..................... .................... .................... ....................
11311 MW155312.1 |Severe acute respiratory syndrome coro .................... .................... ..................... .................... .................... ....................
11312 MW155313.1 |Severe acute respiratory syndrome coro .................... .................... ..................... .................... .................... ....................
11313 MW155314.1 |Severe acute respiratory syndrome coro .................... .................... ..................... .................... .................... ....................
11314 MW155315.1 |Severe acute respiratory syndrome coro .................... .................... ..................... .................... .................... ....................
11315 MW155316.1 |Severe acute respiratory syndrome coro .................... .................... ..................... .................... .................... ....................
11316 MW155317.1 |Severe acute respiratory syndrome coro .................... .................... ..................... .................... .................... ....................
11317 MW155319.1 |Severe acute respiratory syndrome coro .................... .................... ..................... .................... .................... ....................
11318 MW155320.1 |Severe acute respiratory syndrome coro .................... .................... ..................... .................... .................... ....................
11319 MW155321.1 |Severe acute respiratory syndrome coro .................... .................... ..................... .................... .................... ....................
11320 MW155322.1 |Severe acute respiratory syndrome coro .................... .................... ..................... .................... .................... ....................
11321 MW155323.1 |Severe acute respiratory syndrome coro .................... .................... ..................... .................... .................... ....................
11322 MW155324.1 |Severe acute respiratory syndrome coro .................... .................... ..................... .................... .................... ....................
11323 MW155325.1 |Severe acute respiratory syndrome coro .................... .................... ..................... .................... .................... ....................
11324 MW155326.1 |Severe acute respiratory syndrome coro .................... .................... ..................... .................... .................... ....................
11325 MW155327.1 |Severe acute respiratory syndrome coro .................... .................... ..................... .................... .................... ....................
11326 MW155328.1 |Severe acute respiratory syndrome coro .................... .................... ..................... .................... .................... ....................
11327 MW155329.1 |Severe acute respiratory syndrome coro .................... .................... ..................... .................... .................... ....................
11328 MW155330.1 |Severe acute respiratory syndrome coro .................... .................... ..................... .................... .................... ....................
11329 MW155331.1 |Severe acute respiratory syndrome coro .................... .................... ..................... .................... .................... ....................
11330 MW155332.1 |Severe acute respiratory syndrome coro .................... .................... ..................... .................... .................... ....................
11331 MW155333.1 |Severe acute respiratory syndrome coro .................... .................... ..................... .................... .................... ....................
11332 MW155334.1 |Severe acute respiratory syndrome coro .................... .................... ..................... .................... .................... ....................

```
11333 MW155335.1 |Severe acute respiratory syndrome coro    ....................  ....................  .....................  ....................  ....................  ....................
11334 MW155336.1 |Severe acute respiratory syndrome coro    ....................  ....................  .....................  ....................  ....................  ....................
11335 MW155337.1 |Severe acute respiratory syndrome coro    ....................  ....................  .....................  ....................  ....................  ....................
11336 MW155338.1 |Severe acute respiratory syndrome coro    ....................  ....................  .....................  ....................  ....................  ....................
11337 MW155339.1 |Severe acute respiratory syndrome coro    ....................  ....................  .....................  ....................  ....................  ....................
11338 MW155341.1 |Severe acute respiratory syndrome coro    ....................  ....................  .....................  ....................  ....................  ....................
11339 MW155342.1 |Severe acute respiratory syndrome coro    ....................  ....................  .....................  ....................  ....................  ....................
11340 MW155343.1 |Severe acute respiratory syndrome coro    ....................  ....................  .....................  ....................  ....................  ....................
11341 MW155344.1 |Severe acute respiratory syndrome coro    ....................  ....................  .....................  ....................  ....................  ....................
11342 MW155345.1 |Severe acute respiratory syndrome coro    ....................  ....................  .....................  ....................  ....................  ....................
11343 MW155346.1 |Severe acute respiratory syndrome coro    ....................  ....................  .....................  ....................  ....................  ....................
11344 MW155347.1 |Severe acute respiratory syndrome coro    ....................  ....................  .....................  ....................  ....................  ....................
11345 MW155348.1 |Severe acute respiratory syndrome coro    ....................  ....................  .....................  ....................  ....................  ....................
11346 MW155349.1 |Severe acute respiratory syndrome coro    ....................  ....................  .....................  ....................  ....................  ....................
11347 MW155350.1 |Severe acute respiratory syndrome coro    ....................  ....................  .....................  ....................  ....................  ....................
11348 MW155351.1 |Severe acute respiratory syndrome coro    ....................  ....................  .....................  ....................  ....................  ....................
11349 MW155352.1 |Severe acute respiratory syndrome coro    ....................  ....................  .....................  ....................  ....................  ....................
11350 MW155354.1 |Severe acute respiratory syndrome coro    ....................  ....................  .....................  ....................  ....................  ....................
11351 MW155359.1 |Severe acute respiratory syndrome coro    ....................  ....................  .....................  ....................  ....................  ....................
11352 MW155360.1 |Severe acute respiratory syndrome coro    ....................  ....................  .....................  ....................  ....................  ....................
11354 MW155364.1 |Severe acute respiratory syndrome coro    ....................  ....................  .....................  ....................  ....................  ....................
11355 MW155365.1 |Severe acute respiratory syndrome coro    ....................  ....................  .....................  ....................  ....................  ....................
11356 MW155366.1 |Severe acute respiratory syndrome coro    ....................  ....................  .....................  ....................  ....................  ....................
11357 MW155367.1 |Severe acute respiratory syndrome coro    ....................  ....................  .....................  ....................  ....................  ....................
11358 MW155369.1 |Severe acute respiratory syndrome coro    ....................  ....................  .....................  ....................  ....................  ....................
11359 MW155372.1 |Severe acute respiratory syndrome coro    ....................  ....................  .....................  ....................  ....................  ....................
11360 MW155373.1 |Severe acute respiratory syndrome coro    ....................  ....................  .....................  ....................  ....................  ....................
11361 MW155374.1 |Severe acute respiratory syndrome coro    ....................  ....................  .....................  ....................  ....................  ....................
11362 MW155375.1 |Severe acute respiratory syndrome coro    ....................  ....................  .....................  ....................  ....................  ....................
11363 MW155376.1 |Severe acute respiratory syndrome coro    ....................  ....................  .....................  ....................  ....................  ....................
11364 MW155377.1 |Severe acute respiratory syndrome coro    ....................  ....................  .....................  ....................  ....................  ....................
11365 MW155379.1 |Severe acute respiratory syndrome coro    ....................  ....................  .....................  ....................  ....................  ....................
11366 MW155380.1 |Severe acute respiratory syndrome coro    ....................  ....................  .....................  ....................  ....................  ....................
11367 MW155384.1 |Severe acute respiratory syndrome coro    ....................  ....................  .....................  ....................  ....................  ....................
11368 MW155386.1 |Severe acute respiratory syndrome coro    ....................  ....................  .....................  ....................  ....................  ....................
11369 MW155387.1 |Severe acute respiratory syndrome coro    ....................  ....................  .....................  ....................  ....................  ....................
11370 MW155389.1 |Severe acute respiratory syndrome coro    ....................  ....................  .....................  ....................  ....................  ....................
11371 MW155391.1 |Severe acute respiratory syndrome coro    ....................  ....................  .....................  ....................  ....................  ....................
11372 MW155392.1 |Severe acute respiratory syndrome coro    ....................  ....................  .....................  ....................  ....................  ....................
11373 MW155393.1 |Severe acute respiratory syndrome coro    ....................  ....................  .....................  ....................  ....................  ....................
11374 MW155395.1 |Severe acute respiratory syndrome coro    ....................  ....................  .....................  ....................  ....................  ....................
11375 MW155396.1 |Severe acute respiratory syndrome coro    ....................  ....................  .....................  ....................  ....................  ....................
11376 MW155397.1 |Severe acute respiratory syndrome coro    ....................  ....................  .....................  ....................  ....................  ....................
11377 MW155398.1 |Severe acute respiratory syndrome coro    ....................  ....................  .....................  ....................  ....................  ....................
11378 MW155400.1 |Severe acute respiratory syndrome coro    ....................  ....................  .....................  ....................  ....................  ....................
11379 MW155401.1 |Severe acute respiratory syndrome coro    ....................  ....................  .....................  ....................  ....................  ....................
11381 MW155403.1 |Severe acute respiratory syndrome coro    ....................  ....................  .....................  ....................  ....................  ....................
11382 MW155405.1 |Severe acute respiratory syndrome coro    ....................  ....................  .....................  ....................  ....................  ....................
11383 MW155406.1 |Severe acute respiratory syndrome coro    ....................  ....................  .....................  ....................  ....................  ....................
11384 MW155407.1 |Severe acute respiratory syndrome coro    ....................  ....................  .....................  ....................  ....................  ....................
11385 MW155409.1 |Severe acute respiratory syndrome coro    ....................  ....................  .....................  ....................  ....................  ....................
11386 MW155411.1 |Severe acute respiratory syndrome coro    ....................  ....................  .....................  ....................  ....................  ....................
11387 MW155412.1 |Severe acute respiratory syndrome coro    ....................  ....................  .....................  ....................  ....................  ....................
11388 MW155413.1 |Severe acute respiratory syndrome coro    ....................  ....................  .....................  ....................  ....................  ....................
11389 MW155414.1 |Severe acute respiratory syndrome coro    ....................  ....................  .....................  ....................  ....................  ....................
11390 MW155415.1 |Severe acute respiratory syndrome coro    ....................  ....................  .....................  ....................  ....................  ....................
11391 MW155418.1 |Severe acute respiratory syndrome coro    ....................  ....................  .....................  ....................  ....................  ....................
11392 MW155420.1 |Severe acute respiratory syndrome coro    ....................  ....................  .....................  ....................  ....................  ....................
11393 MW155422.1 |Severe acute respiratory syndrome coro    ....................  ....................  .....................  ....................  ....................  ....................
11394 MW155423.1 |Severe acute respiratory syndrome coro    ....................  ....................  .....................  ....................  ....................  ....................
11395 MW155424.1 |Severe acute respiratory syndrome coro    ....................  ....................  .....................  ....................  ....................  ....................
11396 MW155425.1 |Severe acute respiratory syndrome coro    ....................  ....................  .....................  ....................  ....................  ....................
11397 MW155428.1 |Severe acute respiratory syndrome coro    ....................  ....................  .....................  ....................  ....................  ....................
11398 MW155430.1 |Severe acute respiratory syndrome coro    ....................  ....................  .....................  ....................  ....................  ....................
11399 MW155431.1 |Severe acute respiratory syndrome coro    ....................  ....................  .....................  ....................  ....................  ....................
11400 MW155432.1 |Severe acute respiratory syndrome coro    ....................  ....................  .....................  ....................  ....................  ....................
11401 MW155433.1 |Severe acute respiratory syndrome coro    ....................  ....................  .....................  ....................  ....................  ....................
11402 MW155434.1 |Severe acute respiratory syndrome coro    ....................  ....................  .....................  ....................  ....................  ....................
11403 MW155435.1 |Severe acute respiratory syndrome coro    ....................  ....................  .....................  ....................  ....................  ....................
11404 MW155436.1 |Severe acute respiratory syndrome coro    ....................  ....................  .....................  ....................  ....................  ....................
11405 MW155437.1 |Severe acute respiratory syndrome coro    ....................  ....................  .....................  ....................  ....................  ....................
11406 MW155439.1 |Severe acute respiratory syndrome coro    ....................  ....................  .....................  ....................  ....................  ....................
11407 MW155440.1 |Severe acute respiratory syndrome coro    ....................  ....................  .....................  ....................  ....................  ....................
11408 MW155441.1 |Severe acute respiratory syndrome coro    ....................  ....................  .....................  ....................  ....................  ....................
11409 MW155442.1 |Severe acute respiratory syndrome coro    ....................  ....................  .....................  ....................  ....................  ....................
11410 MW155443.1 |Severe acute respiratory syndrome coro    ....................  ....................  .....................  ....................  ....................  ....................
11411 MW155444.1 |Severe acute respiratory syndrome coro    ....................  ....................  .....................  ....................  ....................  ....................
11412 MW155445.1 |Severe acute respiratory syndrome coro    ....................  ....................  .....................  ....................  ....................  ....................
```

```
11413 MW155446.1 |Severe acute respiratory syndrome coro  ....................  ....................  .....................  ....................  ....................  ....................
11414 MW155447.1 |Severe acute respiratory syndrome coro  ....................  ....................  .....................  ....................  ....................  ....................
11415 MW155448.1 |Severe acute respiratory syndrome coro  ....................  ....................  .....................  ....................  ....................  ....................
11416 MW155449.1 |Severe acute respiratory syndrome coro  ....................  ....................  .....................  ....................  ....................  ....................
11417 MW155450.1 |Severe acute respiratory syndrome coro  ....................  ....................  .....................  ....................  ....................  ....................
11418 MW155451.1 |Severe acute respiratory syndrome coro  ....................  ....................  .....................  ....................  ....................  ....................
11419 MW155453.1 |Severe acute respiratory syndrome coro  ....................  ....................  .....................  ....................  ....................  ....................
11420 MW155454.1 |Severe acute respiratory syndrome coro  ....................  ....................  .....................  ....................  ....................  ....................
11421 MW155455.1 |Severe acute respiratory syndrome coro  ....................  ....................  .....................  ....................  ....................  ....................
11422 MW155456.1 |Severe acute respiratory syndrome coro  ....................  ....................  .....................  ....................  ....................  ....................
11423 MW155458.1 |Severe acute respiratory syndrome coro  ....................  ....................  .....................  ....................  ....................  ....................
11424 MW155459.1 |Severe acute respiratory syndrome coro  ....................  ....................  .....................  ....................  ....................  ....................
11425 MW155460.1 |Severe acute respiratory syndrome coro  ....................  ....................  .....................  ....................  ....................  ....................
11426 MW155461.1 |Severe acute respiratory syndrome coro  ....................  ....................  .....................  ....................  ....................  ....................
11427 MW155463.1 |Severe acute respiratory syndrome coro  ....................  ....................  .....................  ....................  ....................  ....................
11428 MW155466.1 |Severe acute respiratory syndrome coro  ....................  ....................  .....................  ....................  ....................  ....................
11429 MW155467.1 |Severe acute respiratory syndrome coro  ....................  ....................  .....................  ....................  ....................  ....................
11430 MW155468.1 |Severe acute respiratory syndrome coro  ....................  ....................  .....................  ....................  ....................  ....................
11431 MW155469.1 |Severe acute respiratory syndrome coro  ....................  ....................  .....................  ....................  ....................  ....................
11432 MW155471.1 |Severe acute respiratory syndrome coro  ....................  ....................  .....................  ....................  ....................  ....................
11433 MW155472.1 |Severe acute respiratory syndrome coro  ....................  ....................  .....................  ....................  ....................  ....................
11434 MW155473.1 |Severe acute respiratory syndrome coro  ....................  ....................  .....................  ....................  ....................  ....................
11435 MW155474.1 |Severe acute respiratory syndrome coro  ....................  ....................  .....................  ....................  ....................  ....................
11436 MW155475.1 |Severe acute respiratory syndrome coro  ....................  ....................  .....................  ....................  ....................  ....................
11437 MW155476.1 |Severe acute respiratory syndrome coro  ....................  ....................  .....................  ....................  ....................  ....................
11438 MW155477.1 |Severe acute respiratory syndrome coro  ....................  ....................  .....................  ....................  ....................  ....................
11439 MW155478.1 |Severe acute respiratory syndrome coro  ....................  ....................  .....................  ....................  ....................  ....................
11440 MW155479.1 |Severe acute respiratory syndrome coro  ....................  ....................  .....................  ....................  ....................  ....................
11441 MW155481.1 |Severe acute respiratory syndrome coro  ....................  ....................  .....................  ....................  ....................  ....................
11442 MW155483.1 |Severe acute respiratory syndrome coro  ....................  ....................  .....................  ....................  ....................  ....................
11443 MW155486.1 |Severe acute respiratory syndrome coro  ....................  ....................  .....................  ....................  ....................  ....................
11444 MW155488.1 |Severe acute respiratory syndrome coro  ....................  ....................  .....................  ....................  ....................  ....................
11445 MW155491.1 |Severe acute respiratory syndrome coro  ....................  ....................  .....................  ....................  ....................  ....................
11446 MW155492.1 |Severe acute respiratory syndrome coro  ....................  ....................  .....................  ....................  ....................  ....................
11447 MW155494.1 |Severe acute respiratory syndrome coro  ....................  ....................  .....................  ....................  ....................  ....................
11448 MW155499.1 |Severe acute respiratory syndrome coro  ....................  ....................  .....................  ....................  ....................  ....................
11449 MW155500.1 |Severe acute respiratory syndrome coro  ....................  ....................  .....................  ....................  ....................  ....................
11450 MW155501.1 |Severe acute respiratory syndrome coro  ....................  ....................  .....................  ....................  ....................  ....................
11451 MW155502.1 |Severe acute respiratory syndrome coro  ....................  ....................  .....................  ....................  ....................  ....................
11452 MW155503.1 |Severe acute respiratory syndrome coro  ....................  ....................  .....................  ....................  ....................  ....................
11453 MW155504.1 |Severe acute respiratory syndrome coro  ....................  ....................  .....................  ....................  ....................  ....................
11454 MW155505.1 |Severe acute respiratory syndrome coro  ....................  ....................  .....................  ....................  ....................  ....................
11455 MW155506.1 |Severe acute respiratory syndrome coro  ....................  ....................  .....................  ....................  ....................  ....................
11456 MW155507.1 |Severe acute respiratory syndrome coro  ....................  ....................  .....................  ....................  ....................  ....................
11457 MW155508.1 |Severe acute respiratory syndrome coro  ....................  ....................  .....................  ....................  ....................  ....................
11458 MW155512.1 |Severe acute respiratory syndrome coro  ....................  ....................  .....................  ....................  ....................  ....................
11459 MW155513.1 |Severe acute respiratory syndrome coro  ....................  ....................  .....................  ....................  ....................  ....................
11460 MW155517.1 |Severe acute respiratory syndrome coro  ....................  ....................  .....................  ....................  ....................  ....................
11461 MW155518.1 |Severe acute respiratory syndrome coro  ....................  ....................  .....................  ....................  ....................  ....................
11462 MW155519.1 |Severe acute respiratory syndrome coro  ....................  ....................  .....................  ....................  ....................  ....................
11463 MW155520.1 |Severe acute respiratory syndrome coro  ....................  ....................  .....................  ....................  ....................  ....................
11464 MW155521.1 |Severe acute respiratory syndrome coro  ....................  ....................  .....................  ....................  ....................  ....................
11465 MW155522.1 |Severe acute respiratory syndrome coro  ....................  ....................  .....................  ....................  ....................  ....................
11466 MW155523.1 |Severe acute respiratory syndrome coro  ....................  ....................  .....................  ....................  ....................  ....................
11467 MW155524.1 |Severe acute respiratory syndrome coro  ....................  ....................  .....................  ....................  ....................  ....................
11468 MW155525.1 |Severe acute respiratory syndrome coro  ....................  ....................  .....................  ....................  ....................  ....................
11469 MW155526.1 |Severe acute respiratory syndrome coro  ....................  ....................  .....................  ....................  ....................  ....................
11470 MW155527.1 |Severe acute respiratory syndrome coro  ....................  ....................  .....................  ....................  ....................  ....................
11471 MW155528.1 |Severe acute respiratory syndrome coro  ....................  ....................  .....................  ....................  ....................  ....................
11472 MW155529.1 |Severe acute respiratory syndrome coro  ....................  ....................  .....................  ....................  ....................  ....................
11473 MW155530.1 |Severe acute respiratory syndrome coro  ....................  ....................  .....................  ....................  ....................  ....................
11474 MW155531.1 |Severe acute respiratory syndrome coro  ....................  ....................  .....................  ....................  ....................  ....................
11475 MW155532.1 |Severe acute respiratory syndrome coro  ....................  ....................  .....................  ....................  ....................  ....................
11476 MW155533.1 |Severe acute respiratory syndrome coro  ....................  ....................  .....................  ....................  ....................  ....................
11477 MW155534.1 |Severe acute respiratory syndrome coro  ....................  ....................  .....................  ....................  ....................  ....................
11478 MW155535.1 |Severe acute respiratory syndrome coro  ....................  ....................  .....................  ....................  ....................  ....................
11479 MW155536.1 |Severe acute respiratory syndrome coro  ....................  ....................  .....................  ....................  ....................  ....................
11480 MW155537.1 |Severe acute respiratory syndrome coro  ....................  ....................  .....................  ....................  ....................  ....................
11481 MW155538.1 |Severe acute respiratory syndrome coro  ....................  ....................  .....................  ....................  ....................  ....................
11482 MW155539.1 |Severe acute respiratory syndrome coro  ....................  ....................  .....................  ....................  ....................  ....................
11483 MW155540.1 |Severe acute respiratory syndrome coro  ....................  ....................  .....................  ....................  ....................  ....................
11484 MW155541.1 |Severe acute respiratory syndrome coro  ....................  ....................  .....................  ....................  ....................  ....................
11485 MW155542.1 |Severe acute respiratory syndrome coro  ....................  ....................  .....................  ....................  ....................  ....................
11486 MW155543.1 |Severe acute respiratory syndrome coro  ....................  ....................  .....................  ....................  ....................  ....................
11487 MW155544.1 |Severe acute respiratory syndrome coro  ....................  ....................  .....................  ....................  ....................  ....................
11488 MW155545.1 |Severe acute respiratory syndrome coro  ....................  ....................  .....................  ....................  ....................  ....................
11489 MW155546.1 |Severe acute respiratory syndrome coro  ....................  ....................  .....................  ....................  ....................  ....................
11490 MW155547.1 |Severe acute respiratory syndrome coro  ....................  ....................  .....................  ....................  ....................  ....................
```

11491 MW155548.1 |Severe acute respiratory syndrome coro .................... .................... .................... .................... .................... ....................
11492 MW155549.1 |Severe acute respiratory syndrome coro .................... .................... .................... .................... .................... ....................
11493 MW155551.1 |Severe acute respiratory syndrome coro .................... .................... .................... .................... .................... ....................
11494 MW155552.1 |Severe acute respiratory syndrome coro .................... .................... .................... .................... .................... ....................
11495 MW155554.1 |Severe acute respiratory syndrome coro .................... .................... .................... .................... .................... ....................
11496 MW155555.1 |Severe acute respiratory syndrome coro .................... .................... .................... .................... .................... ....................
11497 MW155556.1 |Severe acute respiratory syndrome coro .................... .................... .................... .................... .................... ....................
11498 MW155557.1 |Severe acute respiratory syndrome coro .................... .................... .................... .................... .................... ....................
11499 MW155560.1 |Severe acute respiratory syndrome coro .................... .................... .................... .................... .................... ....................
11500 MW155562.1 |Severe acute respiratory syndrome coro .................... .................... .................... .................... .................... ....................
11501 MW155563.1 |Severe acute respiratory syndrome coro .................... .................... .................... .................... .................... ....................
11502 MW155564.1 |Severe acute respiratory syndrome coro .................... .................... .................... .................... .................... ....................
11503 MW155567.1 |Severe acute respiratory syndrome coro .................... .................... .................... .................... .................... ....................
11504 MW155569.1 |Severe acute respiratory syndrome coro .................... .................... .................... .................... .................... ....................
11505 MW155571.1 |Severe acute respiratory syndrome coro .................... .................... .................... .................... .................... ....................
11506 MW155572.1 |Severe acute respiratory syndrome coro .................... .................... .................... .................... .................... ....................
11507 MW155573.1 |Severe acute respiratory syndrome coro .................... .................... .................... .................... .................... ....................
11508 MW155576.1 |Severe acute respiratory syndrome coro .................... .................... .................... .................... .................... ....................
11509 MW155577.1 |Severe acute respiratory syndrome coro .................... .................... .................... .................... .................... ....................
11510 MW155578.1 |Severe acute respiratory syndrome coro .................... .................... .................... .................... .................... ....................
11511 MW155579.1 |Severe acute respiratory syndrome coro .................... .................... .................... .................... .................... ....................
11512 MW155580.1 |Severe acute respiratory syndrome coro .................... .................... .................... .................... .................... ....................
11513 MW155581.1 |Severe acute respiratory syndrome coro .................... .................... .................... .................... .................... ....................
11514 MW155582.1 |Severe acute respiratory syndrome coro .................... .................... .................... .................... .................... ....................
11515 MW155583.1 |Severe acute respiratory syndrome coro .................... .................... .................... .................... .................... ....................
11516 MW155585.1 |Severe acute respiratory syndrome coro .................... .................... .................... .................... .................... ....................
11517 MW155586.1 |Severe acute respiratory syndrome coro .................... .................... .................... .................... .................... ....................
11518 MW155587.1 |Severe acute respiratory syndrome coro .................... .................... .................... .................... .................... ....................
11519 MW155590.1 |Severe acute respiratory syndrome coro .................... .................... .................... .................... .................... ....................
11520 MW155592.1 |Severe acute respiratory syndrome coro .................... .................... .................... .................... .................... ....................
11521 MW155593.1 |Severe acute respiratory syndrome coro .................... .................... .................... .................... .................... ....................
11522 MW155596.1 |Severe acute respiratory syndrome coro .................... .................... .................... .................... .................... ....................
11523 MW155598.1 |Severe acute respiratory syndrome coro .................... .................... .................... .................... .................... ....................
11524 MW155599.1 |Severe acute respiratory syndrome coro .................... .................... .................... .................... .................... ....................
11525 MW155601.1 |Severe acute respiratory syndrome coro .................... .................... .................... .................... .................... ....................
11526 MW155602.1 |Severe acute respiratory syndrome coro .................... .................... .................... .................... .................... ....................
11527 MW155603.1 |Severe acute respiratory syndrome coro .................... .................... .................... .................... .................... ....................
11528 MW155604.1 |Severe acute respiratory syndrome coro .................... .................... .................... .................... .................... ....................
11529 MW155606.1 |Severe acute respiratory syndrome coro .................... .................... .................... .................... .................... ....................
11530 MW155607.1 |Severe acute respiratory syndrome coro .................... .................... .................... .................... .................... ....................
11531 MW155608.1 |Severe acute respiratory syndrome coro .................... .................... .................... .................... .................... ....................
11532 MW155609.1 |Severe acute respiratory syndrome coro .................... .................... .................... .................... .................... ....................
11533 MW155610.1 |Severe acute respiratory syndrome coro .................... .................... .................... .................... .................... ....................
11534 MW155611.1 |Severe acute respiratory syndrome coro .................... .................... .................... .................... .................... ....................
11535 MW155616.1 |Severe acute respiratory syndrome coro .................... .................... .................... .................... .................... ....................
11536 MW155619.1 |Severe acute respiratory syndrome coro .................... .................... .................... .................... .................... ....................
11537 MW155620.1 |Severe acute respiratory syndrome coro .................... .................... .................... .................... .................... ....................
11538 MW155621.1 |Severe acute respiratory syndrome coro .................... .................... .................... .................... .................... ....................
11539 MW155622.1 |Severe acute respiratory syndrome coro .................... .................... .................... .................... .................... ....................
11540 MW155623.1 |Severe acute respiratory syndrome coro .................... .................... .................... .................... .................... ....................
11541 MW155624.1 |Severe acute respiratory syndrome coro .................... .................... .................... .................... .................... ....................
11542 MW155625.1 |Severe acute respiratory syndrome coro .................... .................... .................... .................... .................... ....................
11543 MW155626.1 |Severe acute respiratory syndrome coro .................... .................... .................... .................... .................... ....................
11544 MW155627.1 |Severe acute respiratory syndrome coro .................... .................... .................... .................... .................... ....................
11545 MW155628.1 |Severe acute respiratory syndrome coro .................... .................... .................... .................... .................... ....................
11546 MW155629.1 |Severe acute respiratory syndrome coro .................... .................... .................... .................... .................... ....................
11547 MW155630.1 |Severe acute respiratory syndrome coro .................... .................... .................... .................... .................... ....................
11548 MW155631.1 |Severe acute respiratory syndrome coro .................... .................... .................... .................... .................... ....................
11549 MW155632.1 |Severe acute respiratory syndrome coro .................... .................... .................... .................... .................... ....................
11551 MW155635.1 |Severe acute respiratory syndrome coro .................... .................... .................... .................... .................... ....................
11552 MW155636.1 |Severe acute respiratory syndrome coro .................... .................... .................... .................... .................... ....................
11553 MW155637.1 |Severe acute respiratory syndrome coro .................... .................... .................... .................... .................... ....................
11554 MW155638.1 |Severe acute respiratory syndrome coro .................... .................... .................... .................... .................... ....................
11555 MW155639.1 |Severe acute respiratory syndrome coro .................... .................... .................... .................... .................... ....................
11556 MW155640.1 |Severe acute respiratory syndrome coro .................... .................... .................... .................... .................... ....................
11557 MW155641.1 |Severe acute respiratory syndrome coro .................... .................... .................... .................... .................... ....................
11558 MW155643.1 |Severe acute respiratory syndrome coro .................... .................... .................... .................... .................... ....................
11559 MW155644.1 |Severe acute respiratory syndrome coro .................... .................... .................... .................... .................... ....................
11560 MW155645.1 |Severe acute respiratory syndrome coro .................... .................... .................... .................... .................... ....................
11561 MW155646.1 |Severe acute respiratory syndrome coro .................... .................... .................... .................... .................... ....................
11562 MW155647.1 |Severe acute respiratory syndrome coro .................... .................... .................... .................... .................... ....................
11563 MW155648.1 |Severe acute respiratory syndrome coro .................... .................... .................... .................... ...A................ ....................
11564 MW155649.1 |Severe acute respiratory syndrome coro .................... .................... .................... .................... .................... ....................
11565 MW155650.1 |Severe acute respiratory syndrome coro .................... .................... .................... .................... .................... ....................
11566 MW155651.1 |Severe acute respiratory syndrome coro .................... .................... .................... .................... .................... ....................
11567 MW155652.1 |Severe acute respiratory syndrome coro .................... .................... .................... .................... .................... ....................
11568 MW155653.1 |Severe acute respiratory syndrome coro .................... .................... .................... .................... .................... ....................
11569 MW155654.1 |Severe acute respiratory syndrome coro .................... .................... .................... .................... .................... ....................

```
11570 MW155655.1 |Severe acute respiratory syndrome coro    ....................  ....................  .....................  ....................  ....................  ....................
11571 MW155656.1 |Severe acute respiratory syndrome coro    ....................  ....................  .....................  ....................  ....................  ....................
11572 MW155657.1 |Severe acute respiratory syndrome coro    ....................  ....................  .....................  ....................  ....................  ....................
11574 MW155660.1 |Severe acute respiratory syndrome coro    ....................  ....................  .....................  ....................  ....................  ....................
11575 MW155661.1 |Severe acute respiratory syndrome coro    ....................  ....................  .....................  ....................  ....................  ....................
11576 MW155662.1 |Severe acute respiratory syndrome coro    ....................  ....................  .....................  ....................  ....................  ....................
11577 MW155663.1 |Severe acute respiratory syndrome coro    ....................  ....................  .....................  ....................  ....................  ....................
11578 MW155664.1 |Severe acute respiratory syndrome coro    ....................  ....................  .....................  ....................  ....................  ....................
11579 MW155665.1 |Severe acute respiratory syndrome coro    ....................  ....................  .....................  ....................  ....................  ....................
11580 MW155666.1 |Severe acute respiratory syndrome coro    ....................  ....................  .....................  ....................  ....................  ....................
11581 MW155667.1 |Severe acute respiratory syndrome coro    ....................  ....................  .....................  ....................  ....................  ....................
11582 MW155668.1 |Severe acute respiratory syndrome coro    ....................  ....................  .....................  ....................  ....................  ....................
11583 MW155669.1 |Severe acute respiratory syndrome coro    ....................  ....................  .....................  ....................  ....................  ....................
11584 MW155670.1 |Severe acute respiratory syndrome coro    ....................  ....................  .....................  ....................  ....................  ....................
11585 MW155673.1 |Severe acute respiratory syndrome coro    ....................  ....................  .....................  ....................  ....................  ....................
11586 MW155675.1 |Severe acute respiratory syndrome coro    ....................  ....................  .....................  ....................  ....................  ....................
11587 MW155676.1 |Severe acute respiratory syndrome coro    ....................  ....................  .....................  ....................  ....................  ....................
11588 MW155677.1 |Severe acute respiratory syndrome coro    ....................  ....................  .....................  ....................  ....................  ....................
11589 MW155678.1 |Severe acute respiratory syndrome coro    ....................  ....................  .....................  ....................  ....................  ....................
11590 MW155679.1 |Severe acute respiratory syndrome coro    ....................  ....................  .....................  ....................  ....................  ....................
11591 MW155680.1 |Severe acute respiratory syndrome coro    ....................  ....................  ...........T.........  ....................  ....................  ....................
11592 MW155681.1 |Severe acute respiratory syndrome coro    ....................  ....................  .....................  ....................  ....................  ....................
11593 MW155682.1 |Severe acute respiratory syndrome coro    ....................  ....................  .....................  ....................  ....................  ....................
11594 MW155683.1 |Severe acute respiratory syndrome coro    ....................  ....................  .....................  ....................  ....................  ....................
11595 MW155684.1 |Severe acute respiratory syndrome coro    ....................  ....................  .....................  ....................  ....................  ....................
11596 MW155685.1 |Severe acute respiratory syndrome coro    ....................  ....................  .....................  ....................  ....................  ....................
11597 MW155686.1 |Severe acute respiratory syndrome coro    ....................  ....................  .....................  ....................  ....................  ....................
11598 MW155687.1 |Severe acute respiratory syndrome coro    ....................  ....................  .....................  ....................  ....................  ....................
11599 MW155689.1 |Severe acute respiratory syndrome coro    ....................  ....................  .....................  ....................  ....................  ....................
11600 MW155690.1 |Severe acute respiratory syndrome coro    ....................  ....................  .....................  ....................  ....................  ....................
11601 MW155691.1 |Severe acute respiratory syndrome coro    ....................  ....................  .....................  ....................  ....................  ....................
11602 MW155692.1 |Severe acute respiratory syndrome coro    ....................  ....................  .....................  ....................  ....................  ....................
11603 MW155693.1 |Severe acute respiratory syndrome coro    ....................  ....................  .....................  ....................  ....................  ....................
11604 MW155694.1 |Severe acute respiratory syndrome coro    ....................  ....................  .....................  ....................  ....................  ....................
11605 MW155695.1 |Severe acute respiratory syndrome coro    ....................  ....................  .....................  ....................  ....................  ....................
11606 MW155696.1 |Severe acute respiratory syndrome coro    ....................  ....................  .....................  ....................  ....................  ....................
11607 MW155697.1 |Severe acute respiratory syndrome coro    ....................  ....................  .....................  ....................  ....................  ....................
11608 MW155698.1 |Severe acute respiratory syndrome coro    ....................  ....................  .....................  ....................  ....................  ....................
11609 MW155699.1 |Severe acute respiratory syndrome coro    ....................  ....................  .....................  ....................  ....................  ....................
11610 MW155700.1 |Severe acute respiratory syndrome coro    ....................  ....................  .....................  ....................  ....................  ....................
11611 MW155701.1 |Severe acute respiratory syndrome coro    ....................  ....................  .....................  ....................  ....................  ....................
11612 MW155702.1 |Severe acute respiratory syndrome coro    ....................  ....................  .....................  ....................  ....................  ....................
11613 MW155703.1 |Severe acute respiratory syndrome coro    ....................  ....................  .....................  ....................  ....................  ....................
11614 MW155705.1 |Severe acute respiratory syndrome coro    ....................  ....................  .....................  ....................  ....................  ....................
11615 MW155706.1 |Severe acute respiratory syndrome coro    ....................  ....................  .....................  ....................  ....................  ....................
11616 MW155707.1 |Severe acute respiratory syndrome coro    ....................  ....................  .....................  ....................  ....................  ....................
11617 MW155708.1 |Severe acute respiratory syndrome coro    ....................  ....................  .....................  ....................  ....................  ....................
11618 MW155709.1 |Severe acute respiratory syndrome coro    ....................  ....................  .....................  ....................  ....................  ....................
11619 MW155710.1 |Severe acute respiratory syndrome coro    ....................  ....G...............  .....................  ....................  ....................  ....................
11620 MW155711.1 |Severe acute respiratory syndrome coro    ....................  ....................  .....................  ....................  ....................  ....................
11621 MW155712.1 |Severe acute respiratory syndrome coro    ....................  ....................  .....................  ....................  ....................  ....................
11622 MW155713.1 |Severe acute respiratory syndrome coro    ....................  ....................  .....................  ....................  ....................  ....................
11623 MW155715.1 |Severe acute respiratory syndrome coro    ....................  ....................  .....................  ....................  ....................  ....................
11624 MW155716.1 |Severe acute respiratory syndrome coro    ....................  ....................  .....................  ....................  ....................  ....................
11625 MW155717.1 |Severe acute respiratory syndrome coro    ....................  ....................  .....................  ....................  ....................  ....................
11626 MW155718.1 |Severe acute respiratory syndrome coro    ....................  ....................  .....................  ....................  ....................  ....................
11627 MW155719.1 |Severe acute respiratory syndrome coro    ....................  ....................  .....................  ....................  ....................  ....................
11628 MW155721.1 |Severe acute respiratory syndrome coro    ....................  ....................  .....................  ....................  ....................  ....................
11629 MW155722.1 |Severe acute respiratory syndrome coro    ....................  ....................  .....................  ....................  ....................  ....................
11630 MW155723.1 |Severe acute respiratory syndrome coro    ....................  ....................  .....................  ....................  ....................  ....................
11631 MW155724.1 |Severe acute respiratory syndrome coro    ....................  ....................  .....................  ....................  ....................  ....................
11632 MW155725.1 |Severe acute respiratory syndrome coro    ....................  ....................  .....................  ....................  ....................  ....................
11633 MW155726.1 |Severe acute respiratory syndrome coro    ....................  ....................  .....................  ....................  ....................  ....................
11634 MW155727.1 |Severe acute respiratory syndrome coro    ....................  ....................  .....................  ....................  ....................  ....................
11635 MW155728.1 |Severe acute respiratory syndrome coro    ....................  ....................  .....................  ....................  ....................  ....................
11636 MW155730.1 |Severe acute respiratory syndrome coro    ....................  ....................  .....................  ....................  ....................  ....................
11637 MW155731.1 |Severe acute respiratory syndrome coro    ....................  ....................  .....................  ....................  ....................  ....................
11638 MW155732.1 |Severe acute respiratory syndrome coro    ....................  ....................  .....................  ....................  ....................  ....................
11639 MW155733.1 |Severe acute respiratory syndrome coro    ....................  ....................  .....................  ....................  ....................  ....................
11640 MW155734.1 |Severe acute respiratory syndrome coro    ....................  ....................  .....................  ....................  ....................  ....................
11641 MW155735.1 |Severe acute respiratory syndrome coro    ....................  ....................  .....................  ....................  ....................  ....................
11642 MW155736.1 |Severe acute respiratory syndrome coro    ....................  ....................  .....................  ....................  ....................  ....................
11643 MW155737.1 |Severe acute respiratory syndrome coro    ....................  ....................  .....................  ....................  ....................  ....................
11644 MW155740.1 |Severe acute respiratory syndrome coro    ....................  ....................  .....................  ....................  ....................  ....................
11646 MW155743.1 |Severe acute respiratory syndrome coro    ....................  ....................  .....................  ....................  ....................  ....................
11647 MW155747.1 |Severe acute respiratory syndrome coro    ....................  ....................  .....................  ....................  ....................  ....................
11648 MW155748.1 |Severe acute respiratory syndrome coro    ....................  ....................  .....................  ....................  ....................  ....................
11649 MW155750.1 |Severe acute respiratory syndrome coro    ....................  ....................  .....................  ....................  ....................  ....................
```

| | | | | | | | | |
|---|---|---|---|---|---|---|---|---|
| 11650 | MW155752.1 | \|Severe acute respiratory syndrome coro | .................... | .................... | ..................... | .................... | .................... | .................... |
| 11651 | MW155753.1 | \|Severe acute respiratory syndrome coro | .................... | .................... | ..................... | .................... | .................... | .................... |
| 11652 | MW155755.1 | \|Severe acute respiratory syndrome coro | .................... | .................... | ..................... | .................... | .................... | .................... |
| 11653 | MW155757.1 | \|Severe acute respiratory syndrome coro | .................... | .................... | ..................... | .................... | .................... | .................... |
| 11654 | MW155758.1 | \|Severe acute respiratory syndrome coro | .................... | .................... | ..................... | .................... | .................... | .................... |
| 11656 | MW155760.1 | \|Severe acute respiratory syndrome coro | .................... | .................... | ..................... | .................... | .................... | .................... |
| 11657 | MW155761.1 | \|Severe acute respiratory syndrome coro | .................... | .................... | ..................... | .................... | .................... | .................... |
| 11658 | MW155763.1 | \|Severe acute respiratory syndrome coro | .................... | .................... | ..................... | .................... | .................... | .................... |
| 11659 | MW155764.1 | \|Severe acute respiratory syndrome coro | .................... | .................... | ..................... | .................... | .................... | .................... |
| 11660 | MW155765.1 | \|Severe acute respiratory syndrome coro | .................... | .................... | ..................... | .................... | .................... | .................... |
| 11661 | MW155766.1 | \|Severe acute respiratory syndrome coro | .................... | .................... | ..................... | .................... | .................... | .................... |
| 11662 | MW155767.1 | \|Severe acute respiratory syndrome coro | .................... | .................... | ..................... | .................... | .................... | .................... |
| 11663 | MW155768.1 | \|Severe acute respiratory syndrome coro | .................... | .................... | ..................... | .................... | .................... | .................... |
| 11664 | MW155769.1 | \|Severe acute respiratory syndrome coro | .................... | .................... | ..................... | .................... | .................... | .................... |
| 11665 | MW155770.1 | \|Severe acute respiratory syndrome coro | .................... | .................... | ..................... | .................... | .................... | .................... |
| 11666 | MW155771.1 | \|Severe acute respiratory syndrome coro | .................... | .................... | ..................... | .................... | .................... | .................... |
| 11667 | MW155772.1 | \|Severe acute respiratory syndrome coro | .................... | .................... | ..................... | .................... | .................... | .................... |
| 11668 | MW155774.1 | \|Severe acute respiratory syndrome coro | .................... | .................... | ..................... | .................... | .................... | .................... |
| 11669 | MW155775.1 | \|Severe acute respiratory syndrome coro | .................... | .................... | ..................... | .................... | .................... | .................... |
| 11670 | MW155776.1 | \|Severe acute respiratory syndrome coro | .................... | .................... | ..................... | .................... | .................... | .................... |
| 11671 | MW155777.1 | \|Severe acute respiratory syndrome coro | .................... | .................... | ..................... | .................... | .................... | .................... |
| 11672 | MW155778.1 | \|Severe acute respiratory syndrome coro | .................... | .................... | ..................... | .................... | .................... | .................... |
| 11673 | MW155779.1 | \|Severe acute respiratory syndrome coro | .................... | .................... | ..................... | .................... | .................... | .................... |
| 11674 | MW155780.1 | \|Severe acute respiratory syndrome coro | .................... | .................... | ..................... | .................... | .................... | .................... |
| 11675 | MW155781.1 | \|Severe acute respiratory syndrome coro | .................... | .................... | ..................... | .................... | .................... | .................... |
| 11676 | MW155783.1 | \|Severe acute respiratory syndrome coro | .................... | .................... | ..................... | .................... | .................... | .................... |
| 11677 | MW155784.1 | \|Severe acute respiratory syndrome coro | .................... | .................... | ..................... | .................... | .................... | .................... |
| 11678 | MW155785.1 | \|Severe acute respiratory syndrome coro | .................... | .................... | ..................... | .................... | .................... | .................... |
| 11679 | MW155786.1 | \|Severe acute respiratory syndrome coro | .................... | .................... | ..................... | .................... | .................... | .................... |
| 11680 | MW155788.1 | \|Severe acute respiratory syndrome coro | .................... | ......A............. | ..................... | .................... | .................... | .................... |
| 11681 | MW155790.1 | \|Severe acute respiratory syndrome coro | .................... | ......A............. | ..................... | .................... | .................... | .................... |
| 11682 | MW155791.1 | \|Severe acute respiratory syndrome coro | .................... | .................... | ..................... | .................... | .................... | .................... |
| 11683 | MW155792.1 | \|Severe acute respiratory syndrome coro | .................... | .................... | ..................... | .................... | .................... | .................... |
| 11684 | MW155793.1 | \|Severe acute respiratory syndrome coro | .................... | .................... | ..................... | .................... | .................... | .................... |
| 11685 | MW155794.1 | \|Severe acute respiratory syndrome coro | .................... | .................... | ..................... | .................... | .................... | .................... |
| 11686 | MW155795.1 | \|Severe acute respiratory syndrome coro | .................... | .................... | ..................... | .................... | .................... | .................... |
| 11687 | MW155797.1 | \|Severe acute respiratory syndrome coro | .................... | .................... | ..................... | .................... | .................... | .................... |
| 11688 | MW155798.1 | \|Severe acute respiratory syndrome coro | .................... | .................... | ..................... | .................... | .................... | .................... |
| 11689 | MW155800.1 | \|Severe acute respiratory syndrome coro | .................... | .................... | ..................... | .................... | .................... | .................... |
| 11690 | MW155802.1 | \|Severe acute respiratory syndrome coro | .................... | .................... | ..................... | .................... | .................... | .................... |
| 11691 | MW155804.1 | \|Severe acute respiratory syndrome coro | .................... | .................... | ..................... | .................... | .................... | .................... |
| 11692 | MW155805.1 | \|Severe acute respiratory syndrome coro | .................... | .................... | ..................... | .................... | .................... | .................... |
| 11693 | MW155806.1 | \|Severe acute respiratory syndrome coro | .................... | .................... | ..................... | .................... | .................... | .................... |
| 11694 | MW155807.1 | \|Severe acute respiratory syndrome coro | .................... | .................... | ..................... | .................... | .................... | .................... |
| 11695 | MW155808.1 | \|Severe acute respiratory syndrome coro | .................... | .................... | ..................... | .................... | .................... | .................... |
| 11696 | MW155811.1 | \|Severe acute respiratory syndrome coro | .................... | .................... | ..................... | .................... | .................... | .................... |
| 11697 | MW155812.1 | \|Severe acute respiratory syndrome coro | .................... | .................... | ..................... | .................... | .................... | .................... |
| 11698 | MW155813.1 | \|Severe acute respiratory syndrome coro | .................... | ......A............. | ..................... | .................... | .................... | .................... |
| 11699 | MW155814.1 | \|Severe acute respiratory syndrome coro | .................... | .................... | ..................... | .................... | .................... | .................... |
| 11700 | MW155815.1 | \|Severe acute respiratory syndrome coro | .................... | .................... | ..................... | .................... | .................... | .................... |
| 11701 | MW155816.1 | \|Severe acute respiratory syndrome coro | .................... | .................... | ..................... | .................... | .................... | .................... |
| 11702 | MW155817.1 | \|Severe acute respiratory syndrome coro | .................... | ......A............. | ..................... | .................... | .................... | .................... |
| 11703 | MW155818.1 | \|Severe acute respiratory syndrome coro | .................... | .................... | ..................... | .................... | .................... | .................... |
| 11704 | MW155819.1 | \|Severe acute respiratory syndrome coro | .................... | .................... | ..................... | .................... | .................... | .................... |
| 11705 | MW155821.1 | \|Severe acute respiratory syndrome coro | .................... | .................... | ..................... | .................... | .................... | .................... |
| 11707 | MW155823.1 | \|Severe acute respiratory syndrome coro | .................... | .................... | ..................... | .................... | .................... | .................... |
| 11708 | MW155825.1 | \|Severe acute respiratory syndrome coro | .................... | .................... | ..................... | .................... | .................... | .................... |
| 11709 | MW155826.1 | \|Severe acute respiratory syndrome coro | .................... | .................... | ..................... | .................... | .................... | .................... |
| 11710 | MW155827.1 | \|Severe acute respiratory syndrome coro | .................... | .................... | ..................... | .................... | .................... | .................... |
| 11711 | MW155828.1 | \|Severe acute respiratory syndrome coro | .................... | .................... | ..................... | .................... | .................... | .................... |
| 11712 | MW155830.1 | \|Severe acute respiratory syndrome coro | .................... | .................... | ..................... | .................... | .................... | .................... |
| 11713 | MW155831.1 | \|Severe acute respiratory syndrome coro | .................... | .................... | ..................... | .................... | .................... | .................... |
| 11714 | MW155832.1 | \|Severe acute respiratory syndrome coro | .................... | .................... | ..................... | .................... | .................... | .................... |
| 11715 | MW155833.1 | \|Severe acute respiratory syndrome coro | .................... | .................... | ..................... | .................... | .................... | .................... |
| 11716 | MW155834.1 | \|Severe acute respiratory syndrome coro | .................... | .................... | ..................... | .................... | .................... | .................... |
| 11717 | MW155835.1 | \|Severe acute respiratory syndrome coro | .................... | .................... | ..................... | .................... | .................... | .................... |
| 11718 | MW155836.1 | \|Severe acute respiratory syndrome coro | .................... | .................... | ..................... | .................... | .................... | .................... |
| 11719 | MW155837.1 | \|Severe acute respiratory syndrome coro | .................... | .................... | ..................... | .................... | .................... | .................... |
| 11720 | MW155838.1 | \|Severe acute respiratory syndrome coro | .................... | .................... | ..................... | .................... | .................... | .................... |
| 11721 | MW155839.1 | \|Severe acute respiratory syndrome coro | .................... | .................... | ..................... | .................... | .................... | .................... |
| 11722 | MW155840.1 | \|Severe acute respiratory syndrome coro | .................... | .................... | ..................... | .................... | .................... | .................... |
| 11723 | MW155841.1 | \|Severe acute respiratory syndrome coro | .................... | .................... | ..................... | .................... | .................... | .................... |
| 11724 | MW155842.1 | \|Severe acute respiratory syndrome coro | .................... | .................... | ..................... | .................... | .................... | .................... |
| 11725 | MW155843.1 | \|Severe acute respiratory syndrome coro | .................... | .................... | ..................... | .................... | .................... | .................... |
| 11726 | MW155844.1 | \|Severe acute respiratory syndrome coro | .................... | .................... | ..................... | .................... | .................... | .................... |
| 11727 | MW155846.1 | \|Severe acute respiratory syndrome coro | .................... | .................... | ..................... | .................... | .................... | .................... |
| 11728 | MW155847.1 | \|Severe acute respiratory syndrome coro | .................... | .................... | ..................... | .................... | .................... | .................... |
| 11729 | MW155848.1 | \|Severe acute respiratory syndrome coro | .................... | .................... | ..................... | .................... | .................... | .................... |

| | | | | | | | | |
|---|---|---|---|---|---|---|---|---|
| 11730 | MW155850.1 | \|Severe acute respiratory syndrome coro | .................... | .................... | .................... | .................... | .................... | .................... |
| 11731 | MW155851.1 | \|Severe acute respiratory syndrome coro | .................... | .................... | .................... | .................... | .................... | .................... |
| 11732 | MW155853.1 | \|Severe acute respiratory syndrome coro | .................... | .................... | .................... | .................... | .................... | .................... |
| 11733 | MW155855.1 | \|Severe acute respiratory syndrome coro | .................... | .................... | .................... | .................... | .................... | .................... |
| 11734 | MW155856.1 | \|Severe acute respiratory syndrome coro | .................... | .................... | .................... | .................... | .................... | .................... |
| 11735 | MW155857.1 | \|Severe acute respiratory syndrome coro | .................... | .................... | .................... | .................... | .................... | .................... |
| 11736 | MW155861.1 | \|Severe acute respiratory syndrome coro | .................... | .................... | .................... | .................... | .................... | .................... |
| 11737 | MW155862.1 | \|Severe acute respiratory syndrome coro | .................... | .................... | .................... | .................... | .................... | .................... |
| 11738 | MW155863.1 | \|Severe acute respiratory syndrome coro | .................... | .................... | .................... | .................... | .................... | .................... |
| 11739 | MW155864.1 | \|Severe acute respiratory syndrome coro | .................... | .....A.............. | .................... | .................... | .................... | .................... |
| 11740 | MW155866.1 | \|Severe acute respiratory syndrome coro | .................... | .................... | .................... | .................... | .................... | .................... |
| 11741 | MW155867.1 | \|Severe acute respiratory syndrome coro | .................... | .................... | .................... | .................... | .................... | .................... |
| 11742 | MW155869.1 | \|Severe acute respiratory syndrome coro | .................... | .................... | .................... | .................... | .................... | .................... |
| 11743 | MW155870.1 | \|Severe acute respiratory syndrome coro | .................... | .................... | .................... | .................... | .................... | .................... |
| 11744 | MW155872.1 | \|Severe acute respiratory syndrome coro | .................... | .................... | .................... | .................... | .................... | .................... |
| 11745 | MW155873.1 | \|Severe acute respiratory syndrome coro | .................... | .................... | .................... | .................... | .................... | .................... |
| 11746 | MW155877.1 | \|Severe acute respiratory syndrome coro | .................... | .................... | .................... | .................... | .................... | .................... |
| 11747 | MW155878.1 | \|Severe acute respiratory syndrome coro | .................... | .................... | .................... | .................... | .................... | .................... |
| 11748 | MW155879.1 | \|Severe acute respiratory syndrome coro | .................... | .................... | .................... | .................... | .................... | .................... |
| 11749 | MW155880.1 | \|Severe acute respiratory syndrome coro | .................... | .................... | .................... | .................... | .................... | .................... |
| 11750 | MW155881.1 | \|Severe acute respiratory syndrome coro | .................... | .................... | .................... | .................... | .................... | .................... |
| 11751 | MW155882.1 | \|Severe acute respiratory syndrome coro | .................... | .................... | .................... | .................... | .................... | .................... |
| 11752 | MW155883.1 | \|Severe acute respiratory syndrome coro | .................... | .................... | .................... | .................... | .................... | .................... |
| 11753 | MW155884.1 | \|Severe acute respiratory syndrome coro | .................... | .................... | .................... | .................... | .................... | .................... |
| 11754 | MW155885.1 | \|Severe acute respiratory syndrome coro | .................... | .................... | .................... | .................... | .................... | .................... |
| 11755 | MW155886.1 | \|Severe acute respiratory syndrome coro | .................... | .................... | .................... | .................... | .................... | .................... |
| 11756 | MW155887.1 | \|Severe acute respiratory syndrome coro | .................... | .................... | .................... | .................... | .................... | .................... |
| 11757 | MW155888.1 | \|Severe acute respiratory syndrome coro | .................... | .................... | .................... | .................... | .................... | .................... |
| 11758 | MW155890.1 | \|Severe acute respiratory syndrome coro | .................... | .................... | .................... | .................... | .................... | .................... |
| 11759 | MW155891.1 | \|Severe acute respiratory syndrome coro | .................... | .................... | .................... | .................... | .................... | .................... |
| 11760 | MW155892.1 | \|Severe acute respiratory syndrome coro | .................... | .................... | .................... | .................... | .................... | .................... |
| 11761 | MW155893.1 | \|Severe acute respiratory syndrome coro | .................... | .................... | .................... | .................... | .................... | .................... |
| 11762 | MW155894.1 | \|Severe acute respiratory syndrome coro | .................... | .................... | .................... | .................... | .................... | .................... |
| 11763 | MW155895.1 | \|Severe acute respiratory syndrome coro | .................... | .................... | .................... | .................... | .................... | .................... |
| 11764 | MW155896.1 | \|Severe acute respiratory syndrome coro | .................... | .................... | .................... | .................... | .................... | .................... |
| 11765 | MW155897.1 | \|Severe acute respiratory syndrome coro | .................... | .................... | .................... | .................... | .................... | .................... |
| 11766 | MW155899.1 | \|Severe acute respiratory syndrome coro | .................... | .................... | .................... | .................... | .................... | .................... |
| 11767 | MW155900.1 | \|Severe acute respiratory syndrome coro | .................... | .................... | .................... | .................... | .................... | .................... |
| 11768 | MW155901.1 | \|Severe acute respiratory syndrome coro | .................... | .................... | .................... | .................... | .................... | .................... |
| 11769 | MW155902.1 | \|Severe acute respiratory syndrome coro | .................... | .................... | .................... | .................... | .................... | .................... |
| 11770 | MW155903.1 | \|Severe acute respiratory syndrome coro | .................... | .................... | .................... | .................... | .................... | .................... |
| 11771 | MW155904.1 | \|Severe acute respiratory syndrome coro | .................... | .....A.............. | .................... | .................... | .................... | .................... |
| 11772 | MW155905.1 | \|Severe acute respiratory syndrome coro | .................... | .................... | .................... | .................... | .................... | .................... |
| 11773 | MW155907.1 | \|Severe acute respiratory syndrome coro | .................... | .................... | .................... | .................... | .................... | .................... |
| 11774 | MW155908.1 | \|Severe acute respiratory syndrome coro | .................... | .................... | .................... | .................... | .................... | .................... |
| 11775 | MW155909.1 | \|Severe acute respiratory syndrome coro | .................... | .................... | .................... | .................... | .................... | .................... |
| 11776 | MW155910.1 | \|Severe acute respiratory syndrome coro | .................... | .................... | .................... | .................... | .................... | .................... |
| 11777 | MW155912.1 | \|Severe acute respiratory syndrome coro | .................... | .................... | .................... | .................... | .................... | .................... |
| 11778 | MW155913.1 | \|Severe acute respiratory syndrome coro | .................... | .................... | .................... | .................... | .................... | .................... |
| 11779 | MW155914.1 | \|Severe acute respiratory syndrome coro | .................... | .................... | .................... | .................... | .................... | .................... |
| 11780 | MW155915.1 | \|Severe acute respiratory syndrome coro | .................... | .................... | .................... | .................... | .................... | .................... |
| 11781 | MW155917.1 | \|Severe acute respiratory syndrome coro | .................... | .................... | .................... | .................... | .................... | .................... |
| 11782 | MW155920.1 | \|Severe acute respiratory syndrome coro | .................... | .................... | .................... | .................... | .................... | .................... |
| 11783 | MW155921.1 | \|Severe acute respiratory syndrome coro | .................... | .................... | .................... | .................... | .................... | .................... |
| 11784 | MW155922.1 | \|Severe acute respiratory syndrome coro | .................... | .................... | .................... | .................... | .................... | .................... |
| 11785 | MW155926.1 | \|Severe acute respiratory syndrome coro | .................... | .................... | .................... | .................... | .................... | .................... |
| 11786 | MW155928.1 | \|Severe acute respiratory syndrome coro | .................... | .................... | .................... | .................... | .................... | .................... |
| 11787 | MW155929.1 | \|Severe acute respiratory syndrome coro | .................... | .................... | .................... | .................... | .................... | .................... |
| 11788 | MW155934.1 | \|Severe acute respiratory syndrome coro | .................... | .................... | .................... | .................... | .................... | .................... |
| 11789 | MW155935.1 | \|Severe acute respiratory syndrome coro | .................... | .................... | .................... | .................... | .................... | .................... |
| 11790 | MW155936.1 | \|Severe acute respiratory syndrome coro | .................... | .................... | .................... | .................... | .................... | .................... |
| 11791 | MW155937.1 | \|Severe acute respiratory syndrome coro | .................... | .................... | .................... | .................... | .................... | .................... |
| 11792 | MW155938.1 | \|Severe acute respiratory syndrome coro | .................... | .................... | .................... | .................... | .................... | .................... |
| 11793 | MW155939.1 | \|Severe acute respiratory syndrome coro | .................... | .................... | .................... | .................... | .................... | .................... |
| 11794 | MW155940.1 | \|Severe acute respiratory syndrome coro | .................... | .................... | .................... | .................... | .................... | .................... |
| 11795 | MW155941.1 | \|Severe acute respiratory syndrome coro | .................... | .................... | .................... | .................... | .................... | .................... |
| 11796 | MW155942.1 | \|Severe acute respiratory syndrome coro | .................... | .................... | .................... | .................... | .................... | .................... |
| 11797 | MW155943.1 | \|Severe acute respiratory syndrome coro | .................... | .................... | .................... | .................... | .................... | .................... |
| 11798 | MW155944.1 | \|Severe acute respiratory syndrome coro | .................... | .................... | .................... | .................... | .................... | .................... |
| 11799 | MW155945.1 | \|Severe acute respiratory syndrome coro | .................... | .................... | .................... | .................... | .................... | .................... |
| 11800 | MW155946.1 | \|Severe acute respiratory syndrome coro | .................... | .................... | .................... | .................... | .................... | .................... |
| 11801 | MW155947.1 | \|Severe acute respiratory syndrome coro | .................... | .................... | .................... | .................... | .................... | .................... |
| 11802 | MW155948.1 | \|Severe acute respiratory syndrome coro | .................... | .................... | .................... | .................... | .................... | .................... |
| 11803 | MW155949.1 | \|Severe acute respiratory syndrome coro | .................... | .................... | .................... | .................... | .................... | .................... |
| 11804 | MW155951.1 | \|Severe acute respiratory syndrome coro | .................... | .................... | .................... | .................... | .................... | .................... |
| 11805 | MW155953.1 | \|Severe acute respiratory syndrome coro | .................... | .................... | .................... | .................... | .................... | .................... |
| 11806 | MW155955.1 | \|Severe acute respiratory syndrome coro | .................... | .................... | .................... | .................... | .................... | .................... |
| 11807 | MW155956.1 | \|Severe acute respiratory syndrome coro | .................... | .................... | .................... | .................... | .................... | .................... |

```
11808 MW155957.1 |Severe acute respiratory syndrome coro    ....................  ....................  ....................  ....................  ....................  ....................
11809 MW155958.1 |Severe acute respiratory syndrome coro    ....................  ....................  ....................  ....................  ....................  ....................
11810 MW155959.1 |Severe acute respiratory syndrome coro    ....................  ....................  ....................  ....................  ....................  ....................
11811 MW155962.1 |Severe acute respiratory syndrome coro    ....................  ....................  ....................  ....................  ....................  ....................
11812 MW155963.1 |Severe acute respiratory syndrome coro    ....................  ....................  ....................  ....................  ....................  ....................
11813 MW155966.1 |Severe acute respiratory syndrome coro    ....................  ....................  ....................  ....................  ....................  ....................
11814 MW155968.1 |Severe acute respiratory syndrome coro    ....................  ....................  ....................  ....................  ....................  ....................
11815 MW155969.1 |Severe acute respiratory syndrome coro    ....................  ....................  ....................  ....................  ....................  ....................
11816 MW155970.1 |Severe acute respiratory syndrome coro    ....................  ....................  ....................  ....................  ....................  ....................
11817 MW155971.1 |Severe acute respiratory syndrome coro    ....................  ....................  ....................  ....................  ....................  ....................
11818 MW155973.1 |Severe acute respiratory syndrome coro    ....................  ....................  ....................  ....................  ....................  ....................
11819 MW155974.1 |Severe acute respiratory syndrome coro    ....................  ....................  ....................  ....................  ....................  ....................
11820 MW155975.1 |Severe acute respiratory syndrome coro    ....................  ....................  ....................  ....................  ....................  ....................
11821 MW155977.1 |Severe acute respiratory syndrome coro    ....................  ....................  ....................  ....................  ....................  ....................
11822 MW155978.1 |Severe acute respiratory syndrome coro    ....................  ....................  ....................  ....................  ....................  ....................
11823 MW155980.1 |Severe acute respiratory syndrome coro    ....................  ....................  ....................  ....................  ....................  ....................
11824 MW155981.1 |Severe acute respiratory syndrome coro    ....................  ....................  ....................  ....................  ....................  ....................
11825 MW155982.1 |Severe acute respiratory syndrome coro    ....................  ....................  ....................  ....................  ....................  ....................
11826 MW155983.1 |Severe acute respiratory syndrome coro    ....................  ....................  ....................  ....................  ....................  ....................
11827 MW155984.1 |Severe acute respiratory syndrome coro    ....................  ....................  ....................  ....................  ....................  ....................
11828 MW155986.1 |Severe acute respiratory syndrome coro    ....................  ....................  ....................  ....................  ....................  ....................
11829 MW155987.1 |Severe acute respiratory syndrome coro    ....................  ....................  ....................  ....................  ....................  ....................
11830 MW155988.1 |Severe acute respiratory syndrome coro    ....................  ....................  ....................  ....................  ....................  ....................
11831 MW155989.1 |Severe acute respiratory syndrome coro    ....................  ....................  ....................  ....................  ....................  ....................
11832 MW155990.1 |Severe acute respiratory syndrome coro    ....................  ....................  ....................  ....................  ....................  ....................
11833 MW155991.1 |Severe acute respiratory syndrome coro    ....................  ....................  ....................  ....................  ....................  ....................
11834 MW155992.1 |Severe acute respiratory syndrome coro    ....................  ....................  ....................  ....................  ....................  ....................
11835 MW155994.1 |Severe acute respiratory syndrome coro    ....................  ....................  ....................  ....................  ....................  ....................
11836 MW155995.1 |Severe acute respiratory syndrome coro    ....................  ....................  ....................  ....................  ....................  ....................
11837 MW155998.1 |Severe acute respiratory syndrome coro    ....................  ....................  ....................  ....................  ....................  ....................
11838 MW155999.1 |Severe acute respiratory syndrome coro    ....................  ....................  ....................  ....................  ....................  ....................
11839 MW156000.1 |Severe acute respiratory syndrome coro    ....................  ....................  ....................  ....................  ....................  ....................
11840 MW156001.1 |Severe acute respiratory syndrome coro    ....................  ....................  ....................  ....................  ....................  ....................
11841 MW156002.1 |Severe acute respiratory syndrome coro    ....................  ....................  ....................  ....................  ....................  ....................
11842 MW156003.1 |Severe acute respiratory syndrome coro    ....................  ....................  ....................  ....................  ....................  ....................
11843 MW156004.1 |Severe acute respiratory syndrome coro    ....................  ....................  ....................  ....................  ....................  ....................
11844 MW156006.1 |Severe acute respiratory syndrome coro    ....................  ....................  ....................  ....................  ....................  ....................
11845 MW156007.1 |Severe acute respiratory syndrome coro    ....................  ....................  ....................  ....................  ....................  ....................
11846 MW156009.1 |Severe acute respiratory syndrome coro    ....................  ....................  ....................  ....................  ....................  ....................
11847 MW156010.1 |Severe acute respiratory syndrome coro    ....................  ....................  ....................  ....................  ....................  ....................
11848 MW156011.1 |Severe acute respiratory syndrome coro    ....................  ....................  ....................  ....................  ....................  ....................
11849 MW156012.1 |Severe acute respiratory syndrome coro    ....................  ....................  ....................  ....................  ....................  ....................
11850 MW156016.1 |Severe acute respiratory syndrome coro    ....................  ....................  ....................  ....................  ....................  ....................
11851 MW156018.1 |Severe acute respiratory syndrome coro    ....................  ....................  ....................  ....................  ....................  ....................
11852 MW156019.1 |Severe acute respiratory syndrome coro    ....................  ....................  ....................  ....................  ....................  ....................
11853 MW156020.1 |Severe acute respiratory syndrome coro    ....................  ....................  ....................  ....................  ....................  ....................
11854 MW156021.1 |Severe acute respiratory syndrome coro    ....................  ....................  ....................  ....................  ....................  ....................
11855 MW156022.1 |Severe acute respiratory syndrome coro    ....................  ....................  ....................  ....................  ....................  ....................
11856 MW156025.1 |Severe acute respiratory syndrome coro    ....................  ....................  ....................  ....................  ....................  ....................
11857 MW156027.1 |Severe acute respiratory syndrome coro    ....................  ....................  ....................  ....................  ....................  ....................
11858 MW156028.1 |Severe acute respiratory syndrome coro    ....................  ....................  ....................  ....................  ....................  ....................
11859 MW156029.1 |Severe acute respiratory syndrome coro    ....................  ....................  ....................  ....................  ....................  ....................
11860 MW156030.1 |Severe acute respiratory syndrome coro    ....................  ....................  ....................  ....................  ....................  ....................
11861 MW156031.1 |Severe acute respiratory syndrome coro    ....................  ....................  ....................  ....................  ....................  ....................
11862 MW156032.1 |Severe acute respiratory syndrome coro    ....................  ....................  ....................  ....................  ....................  ....................
11863 MW156033.1 |Severe acute respiratory syndrome coro    ....................  ....................  ....................  ....................  ....................  ....................
11864 MW156034.1 |Severe acute respiratory syndrome coro    ....................  ....................  ....................  ....................  ....................  ....................
11865 MW156036.1 |Severe acute respiratory syndrome coro    ....................  ....................  ....................  ....................  ....................  ....................
11866 MW156039.1 |Severe acute respiratory syndrome coro    ....................  ....................  ....................  ....................  ....................  ....................
11867 MW156040.1 |Severe acute respiratory syndrome coro    ....................  ....................  ....................  ....................  ....................  ....................
11868 MW156041.1 |Severe acute respiratory syndrome coro    ....................  ....................  ....................  ....................  ....................  ....................
11869 MW156042.1 |Severe acute respiratory syndrome coro    ....................  ....................  ....................  ....................  ....................  ....................
11870 MW156043.1 |Severe acute respiratory syndrome coro    ....................  ....................  ....................  ....................  ....................  ....................
11871 MW156045.1 |Severe acute respiratory syndrome coro    ....................  ....................  ....................  ....................  ....................  ....................
11872 MW156047.1 |Severe acute respiratory syndrome coro    ....................  ....................  ....................  ....................  ....................  ....................
11873 MW156048.1 |Severe acute respiratory syndrome coro    ....................  ....................  ....................  ....................  ....................  ....................
11874 MW156049.1 |Severe acute respiratory syndrome coro    ....................  ....................  ....................  ....................  ....................  ....................
11875 MW156050.1 |Severe acute respiratory syndrome coro    ....................  ....................  ....................  ....................  ....................  ....................
11876 MW156051.1 |Severe acute respiratory syndrome coro    ....................  ....................  ....................  ....................  ....................  ....................
11877 MW156052.1 |Severe acute respiratory syndrome coro    ....................  ....................  ....................  ....................  ....................  ....................
11878 MW156054.1 |Severe acute respiratory syndrome coro    ....................  ....................  ....................  ....................  ....................  ....................
11879 MW156055.1 |Severe acute respiratory syndrome coro    ....................  ....................  ....................  ....................  ....................  ....................
11880 MW156057.1 |Severe acute respiratory syndrome coro    ....................  ....................  ....................  ....................  ....................  ....................
11881 MW156058.1 |Severe acute respiratory syndrome coro    ....................  ....................  ....................  ....................  ....................  ....................
11882 MW156059.1 |Severe acute respiratory syndrome coro    ....................  ....................  ....................  ....................  ....................  ....................
11883 MW156060.1 |Severe acute respiratory syndrome coro    ....................  ....................  ....................  ....................  ....................  ....................
11884 MW156061.1 |Severe acute respiratory syndrome coro    ....................  ....................  ....................  ....................  ....................  ....................
11885 MW156063.1 |Severe acute respiratory syndrome coro    ....................  ....................  ....................  ....................  ....................  ....................
```

```
11886 MW156064.1 |Severe acute respiratory syndrome coro    ····················  ····················  ·····················  ····················  ····················  ····················
11887 MW156066.1 |Severe acute respiratory syndrome coro    ····················  ····················  ·····················  ····················  ····················  ····················
11888 MW156069.1 |Severe acute respiratory syndrome coro    ····················  ····················  ·····················  ····················  ····················  ····················
11889 MW156070.1 |Severe acute respiratory syndrome coro    ····················  ····················  ·····················  ····················  ····················  ····················
11890 MW156071.1 |Severe acute respiratory syndrome coro    ····················  ····················  ·····················  ····················  ····················  ····················
11891 MW156073.1 |Severe acute respiratory syndrome coro    ····················  ····················  ·····················  ····················  ····················  ····················
11892 MW156074.1 |Severe acute respiratory syndrome coro    ····················  ····················  ·····················  ····················  ····················  ····················
11893 MW156075.1 |Severe acute respiratory syndrome coro    ····················  ····················  ·····················  ····················  ····················  ····················
11894 MW156076.1 |Severe acute respiratory syndrome coro    ····················  ····················  ·····················  ····················  ····················  ····················
11895 MW156078.1 |Severe acute respiratory syndrome coro    ····················  ····················  ·····················  ····················  ····················  ····················
11896 MW156080.1 |Severe acute respiratory syndrome coro    ····················  ····················  ·····················  ····················  ····················  ····················
11897 MW156081.1 |Severe acute respiratory syndrome coro    ····················  ····················  ·····················  ····················  ····················  ····················
11898 MW156082.1 |Severe acute respiratory syndrome coro    ····················  ····················  ·····················  ····················  ····················  ····················
11899 MW156083.1 |Severe acute respiratory syndrome coro    ····················  ····················  ·····················  ····················  ····················  ····················
11900 MW156084.1 |Severe acute respiratory syndrome coro    ····················  ····················  ·····················  ····················  ····················  ····················
11901 MW156086.1 |Severe acute respiratory syndrome coro    ····················  ····················  ·····················  ····················  ····················  ····················
11902 MW156087.1 |Severe acute respiratory syndrome coro    ····················  ····················  ·····················  ····················  ····················  ····················
11903 MW156088.1 |Severe acute respiratory syndrome coro    ····················  ····················  ·····················  ····················  ····················  ····················
11904 MW156089.1 |Severe acute respiratory syndrome coro    ····················  ····················  ·····················  ····················  ····················  ····················
11905 MW156090.1 |Severe acute respiratory syndrome coro    ····················  ····················  ·····················  ····················  ····················  ····················
11906 MW156091.1 |Severe acute respiratory syndrome coro    ····················  ····················  ·····················  ····················  ····················  ····················
11907 MW156094.1 |Severe acute respiratory syndrome coro    ····················  ····················  ·····················  ····················  ····················  ····················
11908 MW156097.1 |Severe acute respiratory syndrome coro    ····················  ····················  ·····················  ····················  ····················  ····················
11909 MW156099.1 |Severe acute respiratory syndrome coro    ····················  ····················  ·····················  ····················  ····················  ····················
11910 MW156100.1 |Severe acute respiratory syndrome coro    ····················  ····················  ·····················  ····················  ····················  ····················
11911 MW156101.1 |Severe acute respiratory syndrome coro    ····················  ····················  ·····················  ····················  ····················  ····················
11912 MW156103.1 |Severe acute respiratory syndrome coro    ····················  ····················  ·····················  ····················  ····················  ····················
11913 MW156104.1 |Severe acute respiratory syndrome coro    ····················  ····················  ·····················  ····················  ····················  ····················
11914 MW156105.1 |Severe acute respiratory syndrome coro    ····················  ····················  ·····················  ····················  ····················  ····················
11915 MW156108.1 |Severe acute respiratory syndrome coro    ····················  ····················  ·····················  ····················  ····················  ····················
11916 MW156109.1 |Severe acute respiratory syndrome coro    ····················  ····················  ·····················  ····················  ····················  ····················
11917 MW156110.1 |Severe acute respiratory syndrome coro    ····················  ····················  ·····················  ····················  ····················  ····················
11918 MW156112.1 |Severe acute respiratory syndrome coro    ····················  ····················  ·····················  ····················  ····················  ····················
11919 MW156113.1 |Severe acute respiratory syndrome coro    ····················  ····················  ·····················  ····················  ····················  ····················
11920 MW156114.1 |Severe acute respiratory syndrome coro    ····················  ····················  ·····················  ····················  ····················  ····················
11921 MW156115.1 |Severe acute respiratory syndrome coro    ····················  ····················  ·····················  ····················  ····················  ····················
11922 MW156119.1 |Severe acute respiratory syndrome coro    ····················  ····················  ·····················  ····················  ····················  ····················
11923 MW156120.1 |Severe acute respiratory syndrome coro    ····················  ····················  ·····················  ····················  ····················  ····················
11924 MW156121.1 |Severe acute respiratory syndrome coro    ····················  ····················  ·····················  ····················  ····················  ····················
11925 MW156122.1 |Severe acute respiratory syndrome coro    ····················  ····················  ·····················  ····················  ····················  ····················
11926 MW156123.1 |Severe acute respiratory syndrome coro    ····················  ····················  ·····················  ····················  ····················  ····················
11927 MW156124.1 |Severe acute respiratory syndrome coro    ····················  ····················  ·····················  ····················  ····················  ····················
11928 MW156125.1 |Severe acute respiratory syndrome coro    ····················  ····················  ·····················  ····················  ····················  ····················
11929 MW156127.1 |Severe acute respiratory syndrome coro    ····················  ····················  ·····················  ····················  ····················  ····················
11930 MW156128.1 |Severe acute respiratory syndrome coro    ····················  ····················  ·····················  ····················  ····················  ····················
11931 MW156129.1 |Severe acute respiratory syndrome coro    ····················  ····················  ·····················  ····················  ····················  ····················
11932 MW156130.1 |Severe acute respiratory syndrome coro    ····················  ····················  ·····················  ····················  ····················  ····················
11933 MW156132.1 |Severe acute respiratory syndrome coro    ····················  ····················  ·····················  ····················  ····················  ····················
11934 MW156133.1 |Severe acute respiratory syndrome coro    ····················  ····················  ·····················  ····················  ····················  ····················
11935 MW156135.1 |Severe acute respiratory syndrome coro    ····················  ····················  ·····················  ····················  ····················  ····················
11936 MW156136.1 |Severe acute respiratory syndrome coro    ····················  ····················  ···C·················  ····················  ····················  ····················
11937 MW156138.1 |Severe acute respiratory syndrome coro    ····················  ····················  ·····················  ····················  ····················  ····················
11938 MW156139.1 |Severe acute respiratory syndrome coro    ····················  ····················  ·····················  ····················  ····················  ····················
11939 MW156141.1 |Severe acute respiratory syndrome coro    ····················  ····················  ·····················  ····················  ····················  ····················
11940 MW156142.1 |Severe acute respiratory syndrome coro    ····················  ····················  ·····················  ····················  ····················  ····················
11941 MW156143.1 |Severe acute respiratory syndrome coro    ····················  ····················  ·····················  ····················  ····················  ····················
11942 MW156144.1 |Severe acute respiratory syndrome coro    ····················  ····················  ·····················  ····················  ····················  ····················
11943 MW156145.1 |Severe acute respiratory syndrome coro    ····················  ····················  ·····················  ····················  ····················  ····················
11944 MW156146.1 |Severe acute respiratory syndrome coro    ····················  ····················  ·····················  ····················  ····················  ····················
11945 MW156147.1 |Severe acute respiratory syndrome coro    ····················  ····················  ·····················  ····················  ····················  ····················
11946 MW156149.1 |Severe acute respiratory syndrome coro    ····················  ····················  ·····················  ····················  ····················  ····················
11947 MW156150.1 |Severe acute respiratory syndrome coro    ····················  ····················  ·····················  ····················  ····················  ····················
11948 MW156151.1 |Severe acute respiratory syndrome coro    ····················  ····················  ·····················  ····················  ····················  ····················
11949 MW156154.1 |Severe acute respiratory syndrome coro    ····················  ····················  ·····················  ····················  ····················  ····················
11950 MW156155.1 |Severe acute respiratory syndrome coro    ····················  ····················  ·····················  ····················  ····················  ····················
11951 MW156156.1 |Severe acute respiratory syndrome coro    ····················  ····················  ·····················  ····················  ····················  ····················
11952 MW156157.1 |Severe acute respiratory syndrome coro    ····················  ····················  ·····················  ····················  ····················  ····················
11953 MW156158.1 |Severe acute respiratory syndrome coro    ····················  ····················  ·····················  ····················  ····················  ····················
11954 MW156159.1 |Severe acute respiratory syndrome coro    ····················  ····················  ·····················  ····················  ····················  ····················
11955 MW156160.1 |Severe acute respiratory syndrome coro    ····················  ····················  ·····················  ····················  ····················  ····················
11956 MW156161.1 |Severe acute respiratory syndrome coro    ····················  ····················  ·····················  ····················  ····················  ····················
11957 MW156162.1 |Severe acute respiratory syndrome coro    ····················  ····················  ·····················  ····················  ····················  ····················
11958 MW156163.1 |Severe acute respiratory syndrome coro    ····················  ····················  ·····················  ····················  ····················  ····················
11959 MW156164.1 |Severe acute respiratory syndrome coro    ····················  ····················  ·····················  ····················  ····················  ····················
11960 MW156165.1 |Severe acute respiratory syndrome coro    ····················  ····················  ·····················  ····················  ····················  ····················
11961 MW156166.1 |Severe acute respiratory syndrome coro    ····················  ····················  ·····················  ····················  ····················  ····················
11962 MW156168.1 |Severe acute respiratory syndrome coro    ····················  ····················  ·····················  ····················  ····················  ····················
11963 MW156169.1 |Severe acute respiratory syndrome coro    ····················  ····················  ·····················  ····················  ····················  ····················
```

11964 MW156170.1 |Severe acute respiratory syndrome coro .................... .................... ..................... .................... .................... ....................
11965 MW156171.1 |Severe acute respiratory syndrome coro .................... .................... ..................... .................... .................... ....................
11966 MW156172.1 |Severe acute respiratory syndrome coro .................... .................... ..................... .................... .................... ....................
11967 MW156173.1 |Severe acute respiratory syndrome coro .................... .................... ..................... .................... .................... ....................
11968 MW156174.1 |Severe acute respiratory syndrome coro .................... .................... ..................... .................... .................... ....................
11969 MW156175.1 |Severe acute respiratory syndrome coro .................... .................... ..................... .................... .................... ....................
11970 MW156178.1 |Severe acute respiratory syndrome coro .................... .................... ..................... .................... .................... ....................
11971 MW156179.1 |Severe acute respiratory syndrome coro .................... .................... ..................... .................... .................... ....................
11972 MW156180.1 |Severe acute respiratory syndrome coro .................... .................... ..................... .................... .................... ....................
11973 MW156181.1 |Severe acute respiratory syndrome coro .................... .................... ..................... .................... .................... ....................
11974 MW156182.1 |Severe acute respiratory syndrome coro .................... .................... ..................... .................... .................... ....................
11975 MW156183.1 |Severe acute respiratory syndrome coro .................... .................... ..................... .................... .................... ....................
11976 MW156185.1 |Severe acute respiratory syndrome coro .................... .................... ..................... .................... .................... ....................
11977 MW156186.1 |Severe acute respiratory syndrome coro .................... .................... ..................... .................... .................... ....................
11978 MW156187.1 |Severe acute respiratory syndrome coro .................... .................... ..................... .................... .................... ....................
11979 MW156189.1 |Severe acute respiratory syndrome coro .................... .................... ..................... .................... .................... ....................
11980 MW156190.1 |Severe acute respiratory syndrome coro .................... .................... ..................... .................... .................... ....................
11981 MW156191.1 |Severe acute respiratory syndrome coro .................... .................... ..................... .................... .................... ....................
11982 MW156192.1 |Severe acute respiratory syndrome coro .................... .................... ..................... .................... .................... ....................
11983 MW156193.1 |Severe acute respiratory syndrome coro .................... .................... ..................... .................... .................... ....................
11984 MW156194.1 |Severe acute respiratory syndrome coro .................... .................... ..................... .................... .................... ....................
11985 MW156195.1 |Severe acute respiratory syndrome coro .................... .................... ..................... .................... .................... ....................
11986 MW156197.1 |Severe acute respiratory syndrome coro .................... .................... ..................... .................... .................... ....................
11987 MW156198.1 |Severe acute respiratory syndrome coro .................... .................... ..................... .................... .................... ....................
11988 MW156199.1 |Severe acute respiratory syndrome coro .................... .................... ..................... .................... .................... ....................
11989 MW156200.1 |Severe acute respiratory syndrome coro .................... .................... ..................... .................... .................... ....................
11990 MW156201.1 |Severe acute respiratory syndrome coro .................... .................... ..................... .................... .................... ....................
11991 MW156202.1 |Severe acute respiratory syndrome coro .................... .................... ..................... .................... .................... ....................
11992 MW156204.1 |Severe acute respiratory syndrome coro .................... .................... ..................... .................... .................... ....................
11993 MW156205.1 |Severe acute respiratory syndrome coro .................... .................... ..................... .................... .................... ....................
11994 MW156206.1 |Severe acute respiratory syndrome coro .................... .................... ..................... .................... .................... ....................
11995 MW156208.1 |Severe acute respiratory syndrome coro .................... .................... ..................... .................... .................... ....................
11996 MW156209.1 |Severe acute respiratory syndrome coro .................... .................... ..................... .................... .................... ....................
11997 MW156210.1 |Severe acute respiratory syndrome coro .................... .................... ..................... .................... .................... ....................
11998 MW156211.1 |Severe acute respiratory syndrome coro .................... .................... ..................... .................... .................... ....................
11999 MW156212.1 |Severe acute respiratory syndrome coro .................... .................... ..................... .................... .................... ....................
12000 MW156213.1 |Severe acute respiratory syndrome coro .................... .................... ..................... .................... .................... ....................
12001 MW156214.1 |Severe acute respiratory syndrome coro .................... .................... ..................... .................... .................... ....................
12002 MW156215.1 |Severe acute respiratory syndrome coro .................... .................... ..................... .................... .................... ....................
12003 MW156216.1 |Severe acute respiratory syndrome coro .................... .................... ..................... .................... .................... ....................
12004 MW156217.1 |Severe acute respiratory syndrome coro .................... .................... ..................... .................... .................... ....................
12005 MW156218.1 |Severe acute respiratory syndrome coro .................... .................... ..................... .................... .................... ....................
12006 MW156219.1 |Severe acute respiratory syndrome coro .................... .................... ..................... .................... .................... ....................
12007 MW156220.1 |Severe acute respiratory syndrome coro .................... .................... ..................... .................... .................... ....................
12008 MW156221.1 |Severe acute respiratory syndrome coro .................... .................... ..................... .................... .................... ....................
12009 MW156222.1 |Severe acute respiratory syndrome coro .................... .................... ..................... .................... .................... ....................
12010 MW156226.1 |Severe acute respiratory syndrome coro .................... .................... ..................... .................... .................... ....................
12011 MW156229.1 |Severe acute respiratory syndrome coro .................... .................... ..................... .................... .................... ....................
12012 MW156230.1 |Severe acute respiratory syndrome coro .................... .................... ..................... .................... .................... ....................
12013 MW156231.1 |Severe acute respiratory syndrome coro .................... .................... ..................... .................... .................... ....................
12014 MW156232.1 |Severe acute respiratory syndrome coro .................... .................... ..................... .................... .................... ....................
12015 MW156233.1 |Severe acute respiratory syndrome coro .................... .................... ..................... .................... .................... ....................
12016 MW156234.1 |Severe acute respiratory syndrome coro .................... .................... ..................... .................... .................... ....................
12017 MW156236.1 |Severe acute respiratory syndrome coro .................... .................... ..................... .................... .................... ....................
12018 MW156238.1 |Severe acute respiratory syndrome coro .................... .................... ..................... .................... .................... ....................
12019 MW156239.1 |Severe acute respiratory syndrome coro .................... .................... ..................... .................... .................... ....................
12020 MW156240.1 |Severe acute respiratory syndrome coro .................... .................... ..................... .................... .................... ....................
12021 MW156241.1 |Severe acute respiratory syndrome coro .................... .................... ..................... .................... .................... ....................
12022 MW156242.1 |Severe acute respiratory syndrome coro .................... .................... ..................... .................... .................... ....................
12023 MW156243.1 |Severe acute respiratory syndrome coro .................... .................... ..................... .................... .................... ....................
12024 MW156244.1 |Severe acute respiratory syndrome coro .................... .................... ..................... .................... .................... ....................
12025 MW156245.1 |Severe acute respiratory syndrome coro .................... .................... ..................... .................... .................... ....................
12026 MW156246.1 |Severe acute respiratory syndrome coro .................... .................... ..................... .................... .................... ....................
12027 MW156247.1 |Severe acute respiratory syndrome coro .................... .................... ..................... .................... .................... ....................
12028 MW156248.1 |Severe acute respiratory syndrome coro .................... .................... ..................... .................... .................... ....................
12029 MW156249.1 |Severe acute respiratory syndrome coro .................... .................... ..................... .................... .................... ....................
12030 MW156250.1 |Severe acute respiratory syndrome coro .................... .................... ..................... .................... .................... ....................
12031 MW156251.1 |Severe acute respiratory syndrome coro .................... .................... ..................... .................... .................... ....................
12032 MW156252.1 |Severe acute respiratory syndrome coro .................... .................... ..................... .................... .................... ....................
12033 MW156253.1 |Severe acute respiratory syndrome coro .................... .................... ..................... .................... .................... ....................
12034 MW156254.1 |Severe acute respiratory syndrome coro .................... .................... ..................... .................... .................... ....................
12035 MW156255.1 |Severe acute respiratory syndrome coro .................... .................... ..................... .................... .................... ....................
12036 MW156256.1 |Severe acute respiratory syndrome coro .................... .................... ..................... .................... .................... ....................
12037 MW156257.1 |Severe acute respiratory syndrome coro .................... .................... ..................... .................... .................... ....................
12038 MW156258.1 |Severe acute respiratory syndrome coro .................... .................... ..................... .................... .................... ....................
12039 MW156260.1 |Severe acute respiratory syndrome coro .................... .................... ..................... .................... .................... ....................
12040 MW156261.1 |Severe acute respiratory syndrome coro .................... .................... ..................... .................... .................... ....................
12041 MW156262.1 |Severe acute respiratory syndrome coro .................... .................... ..................... .................... .................... ....................

| | | | | | | | |
|---|---|---|---|---|---|---|---|
| 12042 | MW156264.1 \|Severe acute respiratory syndrome coro | .................... | .................... | .................... | .................... | .................... | .................... |
| 12043 | MW156265.1 \|Severe acute respiratory syndrome coro | .................... | .................... | .................... | .................... | .................... | .................... |
| 12044 | MW156266.1 \|Severe acute respiratory syndrome coro | .................... | .................... | .................... | .................... | .................... | .................... |
| 12045 | MW156267.1 \|Severe acute respiratory syndrome coro | .................... | .................... | .................... | .................... | .................... | .................... |
| 12046 | MW156268.1 \|Severe acute respiratory syndrome coro | .................... | .................... | .................... | .................... | .................... | .................... |
| 12047 | MW156269.1 \|Severe acute respiratory syndrome coro | .................... | .................... | .................... | .................... | .................... | .................... |
| 12048 | MW156271.1 \|Severe acute respiratory syndrome coro | .................... | .................... | .................... | .................... | .................... | .................... |
| 12050 | MW156273.1 \|Severe acute respiratory syndrome coro | .................... | .................... | .................... | .................... | .................... | .................... |
| 12051 | MW156274.1 \|Severe acute respiratory syndrome coro | .................... | .................... | .................... | .................... | .................... | .................... |
| 12052 | MW156276.1 \|Severe acute respiratory syndrome coro | .................... | .................... | .................... | .................... | .................... | .................... |
| 12053 | MW156277.1 \|Severe acute respiratory syndrome coro | .................... | .................... | .................... | .................... | .................... | .................... |
| 12055 | MW156279.1 \|Severe acute respiratory syndrome coro | .................... | .................... | .................... | .................... | .................... | .................... |
| 12056 | MW156280.1 \|Severe acute respiratory syndrome coro | .................... | .................... | .................... | .................... | .................... | .................... |
| 12057 | MW156281.1 \|Severe acute respiratory syndrome coro | .................... | .................... | .................... | .................... | .................... | .................... |
| 12058 | MW156282.1 \|Severe acute respiratory syndrome coro | .................... | .................... | .................... | .................... | .................... | .................... |
| 12059 | MW156283.1 \|Severe acute respiratory syndrome coro | .................... | .................... | .................... | .................... | .................... | .................... |
| 12060 | MW156285.1 \|Severe acute respiratory syndrome coro | .................... | .................... | .................... | .................... | .................... | .................... |
| 12061 | MW156286.1 \|Severe acute respiratory syndrome coro | .................... | .................... | .................... | .................... | .................... | .................... |
| 12062 | MW156287.1 \|Severe acute respiratory syndrome coro | .................... | .................... | .................... | .................... | .................... | .................... |
| 12063 | MW156288.1 \|Severe acute respiratory syndrome coro | .................... | .................... | .................... | .................... | .................... | .................... |
| 12064 | MW156289.1 \|Severe acute respiratory syndrome coro | .................... | .................... | .................... | .................... | .................... | .................... |
| 12065 | MW156290.1 \|Severe acute respiratory syndrome coro | .................... | .................... | .................... | .................... | .................... | .................... |
| 12066 | MW156291.1 \|Severe acute respiratory syndrome coro | .................... | .................... | .................... | .................... | .................... | .................... |
| 12067 | MW156292.1 \|Severe acute respiratory syndrome coro | .................... | .................... | .................... | .................... | .................... | .................... |
| 12068 | MW156293.1 \|Severe acute respiratory syndrome coro | .................... | .................... | .................... | .................... | .................... | .................... |
| 12069 | MW156294.1 \|Severe acute respiratory syndrome coro | .................... | .................... | .................... | .................... | .................... | .................... |
| 12070 | MW156295.1 \|Severe acute respiratory syndrome coro | .................... | .................... | .................... | .................... | .................... | .................... |
| 12072 | MW156297.1 \|Severe acute respiratory syndrome coro | .................... | .................... | .................... | .................... | .................... | .................... |
| 12073 | MW156298.1 \|Severe acute respiratory syndrome coro | .................... | .................... | .................... | .................... | .................... | .................... |
| 12074 | MW156299.1 \|Severe acute respiratory syndrome coro | .................... | .................... | .................... | .................... | .................... | .................... |
| 12075 | MW156300.1 \|Severe acute respiratory syndrome coro | .................... | .................... | .................... | .................... | .................... | .................... |
| 12076 | MW156302.1 \|Severe acute respiratory syndrome coro | .................... | .................... | .................... | .................... | .................... | .................... |
| 12077 | MW156303.1 \|Severe acute respiratory syndrome coro | .................... | .................... | .................... | .................... | .................... | .................... |
| 12078 | MW156304.1 \|Severe acute respiratory syndrome coro | .................... | .................... | .................... | .................... | .................... | .................... |
| 12079 | MW156305.1 \|Severe acute respiratory syndrome coro | .................... | .................... | .................... | .................... | .................... | .................... |
| 12080 | MW156306.1 \|Severe acute respiratory syndrome coro | .................... | .................... | .................... | .................... | .................... | .................... |
| 12081 | MW156307.1 \|Severe acute respiratory syndrome coro | .................... | .................... | .................... | .................... | .................... | .................... |
| 12082 | MW156308.1 \|Severe acute respiratory syndrome coro | .................... | .................... | .................... | .................... | .................... | .................... |
| 12083 | MW156309.1 \|Severe acute respiratory syndrome coro | .................... | .................... | .................... | .................... | .................... | .................... |
| 12084 | MW156310.1 \|Severe acute respiratory syndrome coro | .................... | .................... | .................... | .................... | .................... | .................... |
| 12085 | MW156311.1 \|Severe acute respiratory syndrome coro | .................... | .................... | .................... | .................... | .................... | .................... |
| 12086 | MW156312.1 \|Severe acute respiratory syndrome coro | .................... | .................... | .................... | .................... | .................... | .................... |
| 12087 | MW156313.1 \|Severe acute respiratory syndrome coro | .................... | .................... | .................... | .................... | .................... | .................... |
| 12088 | MW156314.1 \|Severe acute respiratory syndrome coro | .................... | .................... | .................... | .................... | .................... | NN.................. |
| 12089 | MW156315.1 \|Severe acute respiratory syndrome coro | .................... | .................... | .................... | .................... | .................... | .................... |
| 12090 | MW156316.1 \|Severe acute respiratory syndrome coro | .................... | .................... | .................... | .................... | .................... | .................... |
| 12091 | MW156317.1 \|Severe acute respiratory syndrome coro | .................... | .................... | .................... | .................... | .................... | .................... |
| 12092 | MW156318.1 \|Severe acute respiratory syndrome coro | .................... | .................... | .................... | .................... | .................... | .................... |
| 12093 | MW156319.1 \|Severe acute respiratory syndrome coro | .................... | .................... | .................... | .................... | .................... | .................... |
| 12094 | MW156320.1 \|Severe acute respiratory syndrome coro | .................... | .................... | .................... | .................... | .................... | .................... |
| 12095 | MW156321.1 \|Severe acute respiratory syndrome coro | .................... | .................... | .................... | .................... | .................... | .................... |
| 12096 | MW156323.1 \|Severe acute respiratory syndrome coro | .................... | .................... | .................... | .................... | .................... | .................... |
| 12097 | MW156325.1 \|Severe acute respiratory syndrome coro | .................... | .................... | .................... | .................... | .................... | .................... |
| 12098 | MW156326.1 \|Severe acute respiratory syndrome coro | .................... | .................... | .................... | .................... | .................... | .................... |
| 12099 | MW156327.1 \|Severe acute respiratory syndrome coro | .................... | .................... | .................... | .................... | .................... | .................... |
| 12100 | MW156330.1 \|Severe acute respiratory syndrome coro | .................... | .................... | .................... | .................... | .................... | .................... |
| 12101 | MW156332.1 \|Severe acute respiratory syndrome coro | .................... | .................... | .................... | .................... | .................... | .................... |
| 12102 | MW156333.1 \|Severe acute respiratory syndrome coro | .................... | .................... | .................... | .................... | .................... | .................... |
| 12103 | MW156335.1 \|Severe acute respiratory syndrome coro | .................... | .................... | .................... | .................... | .................... | .................... |
| 12104 | MW156336.1 \|Severe acute respiratory syndrome coro | .................... | .................... | .................... | .................... | .................... | .................... |
| 12106 | MW156338.1 \|Severe acute respiratory syndrome coro | .................... | .................... | .................... | .................... | .................... | .................... |
| 12107 | MW156339.1 \|Severe acute respiratory syndrome coro | .................... | .................... | .................... | .................... | .................... | .................... |
| 12108 | MW156340.1 \|Severe acute respiratory syndrome coro | .................... | .................... | .................... | .................... | .................... | .................... |
| 12109 | MW156341.1 \|Severe acute respiratory syndrome coro | .................... | .................... | .................... | .................... | .................... | .................... |
| 12110 | MW156342.1 \|Severe acute respiratory syndrome coro | .................... | .................... | .................... | .................... | .................... | .................... |
| 12111 | MW156343.1 \|Severe acute respiratory syndrome coro | .................... | .................... | .................... | .................... | .................... | .................... |
| 12112 | MW156344.1 \|Severe acute respiratory syndrome coro | .................... | .................... | .................... | .................... | .................... | .................... |
| 12113 | MW156345.1 \|Severe acute respiratory syndrome coro | .................... | .................... | .................... | .................... | .................... | .................... |
| 12114 | MW156346.1 \|Severe acute respiratory syndrome coro | .................... | .................... | .................... | .................... | .................... | .................... |
| 12115 | MW156347.1 \|Severe acute respiratory syndrome coro | .................... | .................... | .................... | .................... | .................... | .................... |
| 12116 | MW156348.1 \|Severe acute respiratory syndrome coro | .................... | .................... | .................... | .................... | .................... | .................... |
| 12117 | MW156349.1 \|Severe acute respiratory syndrome coro | .................... | .................... | .................... | .................... | .................... | .................... |
| 12118 | MW156350.1 \|Severe acute respiratory syndrome coro | .................... | .................... | .................... | .................... | .................... | .................... |
| 12119 | MW156351.1 \|Severe acute respiratory syndrome coro | .................... | .................... | .................... | .................... | .................... | .................... |
| 12120 | MW156352.1 \|Severe acute respiratory syndrome coro | .................... | .................... | .................... | .................... | .................... | .................... |
| 12121 | MW156353.1 \|Severe acute respiratory syndrome coro | .................... | .................... | .................... | .................... | .................... | .................... |
| 12122 | MW156354.1 \|Severe acute respiratory syndrome coro | .................... | .................... | .................... | .................... | .................... | .................... |
| 12123 | MW156355.1 \|Severe acute respiratory syndrome coro | .................... | .................... | .................... | .................... | .................... | .................... |

```
12124 MW156357.1 |Severe acute respiratory syndrome coro    .................... .................... .................... .................... .................... ....................
12125 MW156358.1 |Severe acute respiratory syndrome coro    .................... .................... .................... .................... .................... ....................
12126 MW156360.1 |Severe acute respiratory syndrome coro    .................... .................... .................... .................... .................... ....................
12127 MW156361.1 |Severe acute respiratory syndrome coro    .................... .................... .................... .................... .................... ....................
12128 MW156362.1 |Severe acute respiratory syndrome coro    .................... .................... .................... .................... .................... ....................
12129 MW156363.1 |Severe acute respiratory syndrome coro    .................... .................... .................... .................... .................... ....................
12130 MW156364.1 |Severe acute respiratory syndrome coro    .................... .................... .................... .................... .................... ....................
12131 MW156365.1 |Severe acute respiratory syndrome coro    .................... .................... .................... .................... .................... ....................
12132 MW156366.1 |Severe acute respiratory syndrome coro    .................... .................... .................... .................... .................... ....................
12133 MW156367.1 |Severe acute respiratory syndrome coro    .................... .................... .................... .................... .................... ....................
12134 MW156368.1 |Severe acute respiratory syndrome coro    .................... .................... .................... .................... .................... ....................
12135 MW156370.1 |Severe acute respiratory syndrome coro    .................... .................... .................... .................... .................... ....................
12136 MW156371.1 |Severe acute respiratory syndrome coro    .................... .................... .................... .................... .................... ....................
12137 MW156373.1 |Severe acute respiratory syndrome coro    .................... .................... .................... .................... .................... ....................
12138 MW156374.1 |Severe acute respiratory syndrome coro    .................... .................... .................... .................... .................... ....................
12139 MW156375.1 |Severe acute respiratory syndrome coro    .................... .................... .................... .................... .................... ....................
12140 MW156376.1 |Severe acute respiratory syndrome coro    .................... .................... .................... .................... .................... ....................
12141 MW156377.1 |Severe acute respiratory syndrome coro    .................... .................... .................... .................... .................... ....................
12142 MW156378.1 |Severe acute respiratory syndrome coro    .................... .................... .................... .................... .................... ....................
12143 MW156380.1 |Severe acute respiratory syndrome coro    .................... .................... .................... .................... .................... ....................
12144 MW156381.1 |Severe acute respiratory syndrome coro    .................... .................... .................... .................... .................... ....................
12145 MW156382.1 |Severe acute respiratory syndrome coro    .................... .................... .................... .................... .................... ....................
12146 MW156383.1 |Severe acute respiratory syndrome coro    .................... .................... .................... .................... .................... ....................
12147 MW156384.1 |Severe acute respiratory syndrome coro    .................... .................... .................... .................... .................... ....................
12148 MW156385.1 |Severe acute respiratory syndrome coro    .................... .................... .................... .................... .................... ....................
12149 MW156386.1 |Severe acute respiratory syndrome coro    .................... .................... .................... .................... .................... ....................
12150 MW156387.1 |Severe acute respiratory syndrome coro    .................... .................... .................... .................... .................... ....................
12151 MW156388.1 |Severe acute respiratory syndrome coro    .................... .................... .................... .................... .................... ....................
12152 MW156389.1 |Severe acute respiratory syndrome coro    .................... .................... .................... .................... .................... ....................
12153 MW156390.1 |Severe acute respiratory syndrome coro    .................... .................... .................... .................... .................... ....................
12154 MW156391.1 |Severe acute respiratory syndrome coro    .................... .................... .................... .................... .................... ....................
12155 MW156392.1 |Severe acute respiratory syndrome coro    .................... .................... .................... .................... .................... ....................
12156 MW156393.1 |Severe acute respiratory syndrome coro    .................... .................... .................... .................... .................... ....................
12157 MW156394.1 |Severe acute respiratory syndrome coro    .................... .................... .................... .................... .................... ....................
12158 MW156395.1 |Severe acute respiratory syndrome coro    .................... .................... .................... .................... .................... ....................
12159 MW156396.1 |Severe acute respiratory syndrome coro    .................... .................... .................... .................... .................... ....................
12160 MW156397.1 |Severe acute respiratory syndrome coro    .................... .................... .................... .................... .................... ....................
12161 MW156398.1 |Severe acute respiratory syndrome coro    .................... .................... .................... .................... .................... ....................
12162 MW156399.1 |Severe acute respiratory syndrome coro    .................... .................... .................... .................... .................... ....................
12163 MW156400.1 |Severe acute respiratory syndrome coro    .................... .................... .................... .................... .................... ....................
12164 MW156401.1 |Severe acute respiratory syndrome coro    .................... .................... .................... .................... .................... ....................
12165 MW156402.1 |Severe acute respiratory syndrome coro    .................... .................... .................... .................... .................... ....................
12166 MW156406.1 |Severe acute respiratory syndrome coro    .................... .................... .................... .................... .................... ....................
12167 MW156407.1 |Severe acute respiratory syndrome coro    .................... .................... .................... .................... .................... ....................
12168 MW156408.1 |Severe acute respiratory syndrome coro    .................... .................... .................... .................... .................... ....................
12169 MW156409.1 |Severe acute respiratory syndrome coro    .................... .................... .................... .................... .................... ....................
12170 MW156410.1 |Severe acute respiratory syndrome coro    .................... .................... .................... .................... .................... ....................
12171 MW156411.1 |Severe acute respiratory syndrome coro    .................... .................... .................... .................... .................... ....................
12172 MW156412.1 |Severe acute respiratory syndrome coro    .................... .................... .................... .................... .................... ....................
12173 MW156413.1 |Severe acute respiratory syndrome coro    .................... .................... .................... .................... .................... ....................
12174 MW156414.1 |Severe acute respiratory syndrome coro    .................... .................... .................... .................... .................... ....................
12175 MW156416.1 |Severe acute respiratory syndrome coro    .................... .................... .................... .................... .................... ....................
12176 MW156417.1 |Severe acute respiratory syndrome coro    .................... .................... .................... .................... .................... ....................
12177 MW156420.1 |Severe acute respiratory syndrome coro    .................... .................... .................... .................... .................... ....................
12178 MW156422.1 |Severe acute respiratory syndrome coro    .................... .................... .................... .................... .................... ....................
12179 MW156423.1 |Severe acute respiratory syndrome coro    .................... .................... .................... .................... .................... ....................
12180 MW156424.1 |Severe acute respiratory syndrome coro    .................... .................... .................... .................... .................... ....................
12181 MW156425.1 |Severe acute respiratory syndrome coro    .................... .................... .................... .................... .................... ....................
12182 MW156426.1 |Severe acute respiratory syndrome coro    .................... .................... .................... .................... .................... ....................
12183 MW156427.1 |Severe acute respiratory syndrome coro    .................... .................... .................... .................... .................... ....................
12184 MW156428.1 |Severe acute respiratory syndrome coro    .................... .................... .................... .................... .................... ....................
12185 MW156429.1 |Severe acute respiratory syndrome coro    .................... .................... .................... .................... .................... ....................
12186 MW156430.1 |Severe acute respiratory syndrome coro    .................... .................... .................... .................... .................... ....................
12187 MW156431.1 |Severe acute respiratory syndrome coro    .................... .................... .................... .................... .................... ....................
12188 MW156433.1 |Severe acute respiratory syndrome coro    .................... .................... .................... .................... .................... ....................
12189 MW156434.1 |Severe acute respiratory syndrome coro    .................... .................... .................... .................... .................... ....................
12190 MW156435.1 |Severe acute respiratory syndrome coro    .................... .................... .................... .................... .................... ....................
12191 MW156436.1 |Severe acute respiratory syndrome coro    .................... .................... .................... .................... .................... ....................
12192 MW156437.1 |Severe acute respiratory syndrome coro    .................... .................... .................... .................... .................... ....................
12193 MW156438.1 |Severe acute respiratory syndrome coro    .................... .................... .................... .................... .................... ....................
12194 MW156439.1 |Severe acute respiratory syndrome coro    .................... .................... .................... .................... .................... ....................
12195 MW156441.1 |Severe acute respiratory syndrome coro    .................... .................... .................... .................... .................... ....................
12196 MW156442.1 |Severe acute respiratory syndrome coro    .................... .................... .................... .................... .................... ....................
12197 MW156443.1 |Severe acute respiratory syndrome coro    .................... .................... .................... .................... .................... ....................
12198 MW156444.1 |Severe acute respiratory syndrome coro    .................... .................... .................... .................... .................... ....................
12199 MW156445.1 |Severe acute respiratory syndrome coro    .................... .................... .................... .................... .................... ....................
12200 MW156446.1 |Severe acute respiratory syndrome coro    .................... .................... .................... .................... .................... ....................
12201 MW156447.1 |Severe acute respiratory syndrome coro    .................... .................... .................... .................... .................... ....................
```

| | | | | | | | | |
|---|---|---|---|---|---|---|---|---|
| 12202 | MW156449.1 | \|Severe acute respiratory syndrome coro | .................... | .................... | .................... | .................... | .................... | .................... |
| 12203 | MW156450.1 | \|Severe acute respiratory syndrome coro | .................... | .................... | .................... | .................... | .................... | .................... |
| 12204 | MW156452.1 | \|Severe acute respiratory syndrome coro | .................... | .................... | .................... | .................... | .................... | .................... |
| 12205 | MW156453.1 | \|Severe acute respiratory syndrome coro | .................... | .................... | .................... | .................... | .................... | .................... |
| 12206 | MW156454.1 | \|Severe acute respiratory syndrome coro | .................... | .................... | .................... | .................... | .................... | .................... |
| 12207 | MW156455.1 | \|Severe acute respiratory syndrome coro | .................... | .................... | .................... | .................... | .................... | .................... |
| 12208 | MW156456.1 | \|Severe acute respiratory syndrome coro | .................... | .................... | .................... | .................... | .................... | .................... |
| 12209 | MW156457.1 | \|Severe acute respiratory syndrome coro | .................... | .................... | .................... | .................... | .................... | .................... |
| 12210 | MW156458.1 | \|Severe acute respiratory syndrome coro | .................... | .................... | .................... | .................... | .................... | .................... |
| 12211 | MW156459.1 | \|Severe acute respiratory syndrome coro | .................... | .................... | .................... | .................... | .................... | .................... |
| 12212 | MW156460.1 | \|Severe acute respiratory syndrome coro | .................... | .................... | .................... | .................... | .................... | .................... |
| 12213 | MW156461.1 | \|Severe acute respiratory syndrome coro | .................... | .................... | .................... | .................... | .................... | .................... |
| 12214 | MW156462.1 | \|Severe acute respiratory syndrome coro | .................... | .................... | .................... | .................... | .................... | .................... |
| 12215 | MW156463.1 | \|Severe acute respiratory syndrome coro | .................... | .................... | .................... | .................... | .................... | .................... |
| 12216 | MW156464.1 | \|Severe acute respiratory syndrome coro | .................... | .................... | .................... | .................... | .................... | .................... |
| 12217 | MW156465.1 | \|Severe acute respiratory syndrome coro | .................... | .................... | .................... | .................... | .................... | .................... |
| 12218 | MW156466.1 | \|Severe acute respiratory syndrome coro | .................... | .................... | .................... | .................... | .................... | .................... |
| 12219 | MW156467.1 | \|Severe acute respiratory syndrome coro | .................... | .................... | .................... | .................... | .................... | .................... |
| 12220 | MW156468.1 | \|Severe acute respiratory syndrome coro | .................... | .................... | .................... | .................... | .................... | .................... |
| 12221 | MW156469.1 | \|Severe acute respiratory syndrome coro | .................... | .................... | .................... | .................... | .................... | .................... |
| 12222 | MW156470.1 | \|Severe acute respiratory syndrome coro | .................... | .................... | .................... | .................... | .................... | .................... |
| 12223 | MW156471.1 | \|Severe acute respiratory syndrome coro | .................... | .................... | .................... | .................... | .................... | .................... |
| 12224 | MW156472.1 | \|Severe acute respiratory syndrome coro | .................... | .................... | .................... | .................... | .................... | .................... |
| 12225 | MW156474.1 | \|Severe acute respiratory syndrome coro | .................... | .................... | .................... | .................... | .................... | .................... |
| 12226 | MW156475.1 | \|Severe acute respiratory syndrome coro | .................... | .................... | .................... | .................... | .................... | .................... |
| 12227 | MW156477.1 | \|Severe acute respiratory syndrome coro | .................... | .................... | .................... | .................... | .................... | .................... |
| 12228 | MW156478.1 | \|Severe acute respiratory syndrome coro | .................... | .................... | .................... | .................... | .................... | .................... |
| 12229 | MW156479.1 | \|Severe acute respiratory syndrome coro | .................... | .................... | .................... | .................... | .................... | .................... |
| 12230 | MW156480.1 | \|Severe acute respiratory syndrome coro | .................... | .................... | .................... | .................... | .................... | .................... |
| 12231 | MW156481.1 | \|Severe acute respiratory syndrome coro | .................... | .................... | .................... | .................... | .................... | .................... |
| 12232 | MW156482.1 | \|Severe acute respiratory syndrome coro | .................... | .................... | .................... | .................... | .................... | .................... |
| 12233 | MW156483.1 | \|Severe acute respiratory syndrome coro | .................... | .................... | .................... | .................... | .................... | .................... |
| 12234 | MW156484.1 | \|Severe acute respiratory syndrome coro | .................... | .................... | .................... | .................... | .................... | .................... |
| 12235 | MW156485.1 | \|Severe acute respiratory syndrome coro | .................... | .................... | .................... | .................... | .................... | .................... |
| 12236 | MW156486.1 | \|Severe acute respiratory syndrome coro | .................... | .................... | .................... | .................... | .................... | .................... |
| 12237 | MW156488.1 | \|Severe acute respiratory syndrome coro | .................... | .................... | .................... | .................... | .................... | .................... |
| 12238 | MW156490.1 | \|Severe acute respiratory syndrome coro | .................... | .................... | .................... | .................... | .................... | .................... |
| 12239 | MW156491.1 | \|Severe acute respiratory syndrome coro | .................... | .................... | .................... | .................... | .................... | .................... |
| 12240 | MW156492.1 | \|Severe acute respiratory syndrome coro | .................... | .................... | .................... | .................... | .................... | .................... |
| 12241 | MW156493.1 | \|Severe acute respiratory syndrome coro | .................... | .................... | .................... | .................... | .................... | .................... |
| 12242 | MW156495.1 | \|Severe acute respiratory syndrome coro | .................... | .................... | .................... | .................... | .................... | .................... |
| 12243 | MW156496.1 | \|Severe acute respiratory syndrome coro | .................... | .................... | .................... | .................... | .................... | .................... |
| 12244 | MW156497.1 | \|Severe acute respiratory syndrome coro | .................... | .................... | .................... | .................... | .................... | .................... |
| 12245 | MW156498.1 | \|Severe acute respiratory syndrome coro | .................... | .................... | .................... | .................... | .................... | .................... |
| 12246 | MW156499.1 | \|Severe acute respiratory syndrome coro | .................... | .................... | .................... | .................... | .................... | .................... |
| 12247 | MW156500.1 | \|Severe acute respiratory syndrome coro | .................... | .................... | .................... | .................... | .................... | .................... |
| 12248 | MW156501.1 | \|Severe acute respiratory syndrome coro | .................... | .................... | .................... | .................... | .................... | .................... |
| 12249 | MW156502.1 | \|Severe acute respiratory syndrome coro | .................... | .................... | .................... | .................... | .................... | .................... |
| 12250 | MW156503.1 | \|Severe acute respiratory syndrome coro | .................... | .................... | .................... | .................... | .................... | .................... |
| 12251 | MW156505.1 | \|Severe acute respiratory syndrome coro | .................... | .................... | .................... | .................... | .................... | .................... |
| 12252 | MW156506.1 | \|Severe acute respiratory syndrome coro | .................... | .................... | .................... | .................... | .................... | .................... |
| 12253 | MW156507.1 | \|Severe acute respiratory syndrome coro | .................... | .................... | .................... | .................... | .................... | .................... |
| 12254 | MW156508.1 | \|Severe acute respiratory syndrome coro | .................... | .................... | .................... | .................... | .................... | .................... |
| 12255 | MW156509.1 | \|Severe acute respiratory syndrome coro | .................... | ......A............. | .................... | .................... | .................... | .................... |
| 12256 | MW156511.1 | \|Severe acute respiratory syndrome coro | .................... | .................... | .................... | .................... | .................... | .................... |
| 12257 | MW156512.1 | \|Severe acute respiratory syndrome coro | .................... | .................... | .................... | .................... | .................... | .................... |
| 12258 | MW156513.1 | \|Severe acute respiratory syndrome coro | .................... | .................... | .................... | .................... | .................... | .................... |
| 12259 | MW156515.1 | \|Severe acute respiratory syndrome coro | .................... | .................... | .................... | .................... | .................... | .................... |
| 12260 | MW156516.1 | \|Severe acute respiratory syndrome coro | .................... | .................... | .................... | .................... | .................... | .................... |
| 12261 | MW156517.1 | \|Severe acute respiratory syndrome coro | .................... | .................... | .................... | .................... | .................... | .................... |
| 12262 | MW156519.1 | \|Severe acute respiratory syndrome coro | .................... | .................... | .................... | .................... | .................... | .................... |
| 12263 | MW156520.1 | \|Severe acute respiratory syndrome coro | .................... | .................... | .................... | .................... | .................... | .................... |
| 12264 | MW156521.1 | \|Severe acute respiratory syndrome coro | .................... | .................... | .................... | .................... | .................... | .................... |
| 12265 | MW156522.1 | \|Severe acute respiratory syndrome coro | .................... | .................... | .................... | .................... | .................... | .................... |
| 12266 | MW156523.1 | \|Severe acute respiratory syndrome coro | .................... | .................... | .................... | .................... | .................... | .................... |
| 12267 | MW156524.1 | \|Severe acute respiratory syndrome coro | .................... | .................... | .................... | .................... | .................... | .................... |
| 12268 | MW156525.1 | \|Severe acute respiratory syndrome coro | .................... | .................... | .................... | .................... | .................... | .................... |
| 12269 | MW156526.1 | \|Severe acute respiratory syndrome coro | .................... | .................... | .................... | .................... | .................... | .................... |
| 12270 | MW156528.1 | \|Severe acute respiratory syndrome coro | .................... | .................... | .................... | .................... | .................... | .................... |
| 12271 | MW156529.1 | \|Severe acute respiratory syndrome coro | .................... | .................... | .................... | .................... | .................... | .................... |
| 12272 | MW156531.1 | \|Severe acute respiratory syndrome coro | .................... | .................... | .................... | .................... | .................... | .................... |
| 12273 | MW156535.1 | \|Severe acute respiratory syndrome coro | .................... | .................... | .................... | .................... | .................... | .................... |
| 12274 | MW156537.1 | \|Severe acute respiratory syndrome coro | .................... | .................... | .................... | .................... | .................... | .................... |
| 12275 | MW156538.1 | \|Severe acute respiratory syndrome coro | .................... | .................... | .................... | .................... | .................... | .................... |
| 12276 | MW156539.1 | \|Severe acute respiratory syndrome coro | .................... | .................... | .................... | .................... | .................... | .................... |
| 12277 | MW156540.1 | \|Severe acute respiratory syndrome coro | .................... | .................... | .................... | .................... | .................... | .................... |
| 12278 | MW156541.1 | \|Severe acute respiratory syndrome coro | .................... | .................... | .................... | .................... | .................... | .................... |
| 12279 | MW156542.1 | \|Severe acute respiratory syndrome coro | .................... | .................... | .................... | .................... | .................... | .................... |

12280 MW156544.1 |Severe acute respiratory syndrome coro .................... .................... .................... .................... .................... ....................
12281 MW156546.1 |Severe acute respiratory syndrome coro .................... .................... .................... .................... .................... ....................
12282 MW156547.1 |Severe acute respiratory syndrome coro .................... .................... .................... .................... .................... ....................
12283 MW156549.1 |Severe acute respiratory syndrome coro .................... .................... .................... .................... .................... ....................
12284 MW156550.1 |Severe acute respiratory syndrome coro .................... .................... .................... .................... .................... ....................
12285 MW156552.1 |Severe acute respiratory syndrome coro .................... .................... .................... .................... .................... ....................
12286 MW156553.1 |Severe acute respiratory syndrome coro .................... .................... .................... .................... .................... ....................
12287 MW156554.1 |Severe acute respiratory syndrome coro .................... .................... .................... .................... .................... ....................
12288 MW156555.1 |Severe acute respiratory syndrome coro .................... .................... .................... .................... .................... ....................
12289 MW156556.1 |Severe acute respiratory syndrome coro .................... .................... .................... .................... .................... ....................
12291 MW156559.1 |Severe acute respiratory syndrome coro .................... .................... .................... .................... .................... ....................
12292 MW156560.1 |Severe acute respiratory syndrome coro .................... .................... .................... .................... .................... ....................
12293 MW156561.1 |Severe acute respiratory syndrome coro .................... .................... .................... .................... .................... ....................
12294 MW156562.1 |Severe acute respiratory syndrome coro .................... ...G................ .................... .................... .................... ....................
12295 MW156563.1 |Severe acute respiratory syndrome coro .................... .................... .................... .................... .................... ....................
12296 MW156565.1 |Severe acute respiratory syndrome coro .................... .................... .................... .................... .................... ....................
12297 MW156566.1 |Severe acute respiratory syndrome coro .................... .................... .................... .................... .................... ....................
12298 MW156567.1 |Severe acute respiratory syndrome coro .................... .................... .................... .................... .................... ....................
12299 MW156568.1 |Severe acute respiratory syndrome coro .................... .................... .................... .................... .................... ....................
12300 MW156569.1 |Severe acute respiratory syndrome coro .................... .................... .................... .................... .................... ....................
12301 MW156570.1 |Severe acute respiratory syndrome coro .................... .................... .................... .................... .................... ....................
12302 MW156571.1 |Severe acute respiratory syndrome coro .................... .................... .................... .................... .................... ....................
12303 MW156572.1 |Severe acute respiratory syndrome coro .................... .................... .................... .................... .................... ....................
12304 MW156573.1 |Severe acute respiratory syndrome coro .................... .................... .................... .................... .................... ....................
12305 MW156574.1 |Severe acute respiratory syndrome coro .................... .................... .................... .................... .................... ....................
12306 MW156575.1 |Severe acute respiratory syndrome coro .................... .................... .................... .................... .................... ....................
12307 MW156576.1 |Severe acute respiratory syndrome coro .................... .................... .................... .................... .................... ....................
12308 MW156577.1 |Severe acute respiratory syndrome coro .................... .................... .................... .................... .................... ....................
12309 MW156578.1 |Severe acute respiratory syndrome coro .................... .................... .................... .................... .................... ....................
12310 MW156579.1 |Severe acute respiratory syndrome coro .................... .................... .................... .................... .................... ....................
12311 MW156580.1 |Severe acute respiratory syndrome coro .................... .................... .................... .................... .................... ....................
12312 MW156581.1 |Severe acute respiratory syndrome coro .................... .................... .................... .................... .................... ....................
12313 MW156582.1 |Severe acute respiratory syndrome coro .................... .................... .................... .................... .................... ....................
12314 MW156584.1 |Severe acute respiratory syndrome coro .................... .................... .................... .................... .................... ....................
12315 MW156585.1 |Severe acute respiratory syndrome coro .................... .................... .................... .................... .................... ....................
12316 MW156586.1 |Severe acute respiratory syndrome coro .................... .................... .................... .................... .................... ....................
12317 MW156587.1 |Severe acute respiratory syndrome coro .................... .................... .................... .................... .................... ....................
12318 MW156588.1 |Severe acute respiratory syndrome coro .................... .................... .................... .................... .................... ....................
12319 MW156589.1 |Severe acute respiratory syndrome coro .................... .................... .................... .................... .................... ....................
12320 MW156590.1 |Severe acute respiratory syndrome coro .................... .................... .................... .................... .................... ....................
12321 MW156591.1 |Severe acute respiratory syndrome coro .................... .................... .................... .................... .................... ....................
12322 MW156592.1 |Severe acute respiratory syndrome coro .................... .................... .................... .................... .................... ....................
12323 MW156593.1 |Severe acute respiratory syndrome coro .................... .................... .................... .................... .................... ....................
12324 MW156594.1 |Severe acute respiratory syndrome coro .................... .................... .................... .................... .................... ....................
12325 MW156595.1 |Severe acute respiratory syndrome coro .................... .................... .................... .................... .................... ....................
12327 MW156598.1 |Severe acute respiratory syndrome coro .................... .................... .................... .................... .................... ....................
12328 MW156600.1 |Severe acute respiratory syndrome coro .................... .................... .................... .................... .................... ....................
12329 MW156602.1 |Severe acute respiratory syndrome coro .................... .................... .................... .................... .................... ....................
12330 MW156603.1 |Severe acute respiratory syndrome coro .................... .................... .................... .................... .................... ....................
12331 MW156604.1 |Severe acute respiratory syndrome coro .................... .................... .................... .................... .................... ....................
12332 MW156605.1 |Severe acute respiratory syndrome coro .................... .................... .................... .................... .................... ....................
12333 MW156607.1 |Severe acute respiratory syndrome coro .................... .................... .................... .................... .................... ....................
12334 MW156608.1 |Severe acute respiratory syndrome coro .................... .................... .................... .................... .................... ....................
12335 MW156609.1 |Severe acute respiratory syndrome coro .................... .................... .................... .................... .................... ....................
12336 MW156610.1 |Severe acute respiratory syndrome coro .................... .................... .................... .................... .................... ....................
12337 MW156611.1 |Severe acute respiratory syndrome coro .................... .................... .................... .................... .................... ....................
12338 MW156612.1 |Severe acute respiratory syndrome coro .................... .................... .................... .................... .................... ....................
12339 MW156613.1 |Severe acute respiratory syndrome coro .................... .................... .................... .................... .................... ....................
12340 MW156615.1 |Severe acute respiratory syndrome coro .................... .................... .................... .................... .................... ....................
12341 MW156616.1 |Severe acute respiratory syndrome coro .................... .................... .................... .................... .................... ....................
12342 MW156617.1 |Severe acute respiratory syndrome coro .................... .................... .................... .................... .................... ....................
12343 MW156618.1 |Severe acute respiratory syndrome coro .................... .................... .................... .................... .................... ....................
12344 MW156620.1 |Severe acute respiratory syndrome coro .................... .................... .................... .................... .................... ....................
12345 MW156621.1 |Severe acute respiratory syndrome coro .................... .................... .................... .................... .................... ....................
12346 MW156622.1 |Severe acute respiratory syndrome coro .................... .................... .................... .................... .................... ....................
12347 MW156623.1 |Severe acute respiratory syndrome coro .................... .................... .................... .................... .................... ....................
12348 MW156624.1 |Severe acute respiratory syndrome coro .................... .................... .................... .................... .................... ....................
12349 MW156625.1 |Severe acute respiratory syndrome coro .................... .................... .................... .................... .................... ....................
12350 MW156626.1 |Severe acute respiratory syndrome coro .................... .................... .................... .................... .................... ....................
12351 MW156627.1 |Severe acute respiratory syndrome coro .................... .................... .................... .................... .................... ....................
12352 MW156628.1 |Severe acute respiratory syndrome coro .................... .................... .................... .................... .................... ....................
12353 MW156629.1 |Severe acute respiratory syndrome coro .................... .................... .................... .................... .................... ....................
12354 MW156630.1 |Severe acute respiratory syndrome coro .................... .................... .................... .................... .................... ....................
12355 MW156632.1 |Severe acute respiratory syndrome coro .................... .................... .................... .................... .................... ....................
12356 MW156634.1 |Severe acute respiratory syndrome coro .................... .................... .................... .................... .................... ....................
12357 MW156635.1 |Severe acute respiratory syndrome coro .................... .................... .................... .................... .................... ....................
12358 MW156636.1 |Severe acute respiratory syndrome coro .................... .................... .................... .................... .................... ....................
12359 MW156637.1 |Severe acute respiratory syndrome coro .................... .................... .................... .................... .................... ....................

```
12360 MW156638.1 |Severe acute respiratory syndrome coro    ....................  ....................  ....................  ....................  ....................  ....................
12361 MW156639.1 |Severe acute respiratory syndrome coro    ....................  ....................  ....................  ....................  ....................  ....................
12362 MW156640.1 |Severe acute respiratory syndrome coro    ....................  ....................  ....................  ....................  ....................  ....................
12363 MW156641.1 |Severe acute respiratory syndrome coro    ....................  ....................  ....................  ....................  ....................  ....................
12364 MW156643.1 |Severe acute respiratory syndrome coro    ....................  ....................  ....................  ....................  ....................  ....................
12365 MW156644.1 |Severe acute respiratory syndrome coro    ....................  ....................  ....................  ....................  ....................  ....................
12366 MW156645.1 |Severe acute respiratory syndrome coro    ....................  ....................  ....................  ....................  ....................  ....................
12367 MW156647.1 |Severe acute respiratory syndrome coro    ....................  ....................  ....................  ....................  ....................  ....................
12368 MW156648.1 |Severe acute respiratory syndrome coro    ....................  ....................  ....................  ....................  ....................  ....................
12369 MW156649.1 |Severe acute respiratory syndrome coro    ....................  ....................  ....................  ....................  ....................  ....................
12370 MW156650.1 |Severe acute respiratory syndrome coro    ....................  ....................  ....................  ....................  ....................  ....................
12371 MW156652.1 |Severe acute respiratory syndrome coro    ....................  ....................  ....................  ....................  ....................  ....................
12372 MW156653.1 |Severe acute respiratory syndrome coro    ....................  ....................  ....................  ....................  ....................  ....................
12373 MW156654.1 |Severe acute respiratory syndrome coro    ....................  ....................  ....................  ....................  ....................  ....................
12374 MW156655.1 |Severe acute respiratory syndrome coro    ....................  ....................  ....................  ....................  ....................  ....................
12375 MW156656.1 |Severe acute respiratory syndrome coro    ....................  ....................  ....................  ....................  ....................  ....................
12376 MW156657.1 |Severe acute respiratory syndrome coro    ....................  ....................  ....................  ....................  ....................  ....................
12377 MW156658.1 |Severe acute respiratory syndrome coro    ....................  ....................  ....................  ....................  ....................  ....................
12378 MW156659.1 |Severe acute respiratory syndrome coro    ....................  ....................  ....................  ....................  ....................  ....................
12381 MW156663.1 |Severe acute respiratory syndrome coro    ....................  ....................  ....................  ....................  ....................  ....................
12382 MW156664.1 |Severe acute respiratory syndrome coro    ....................  ....................  ....................  ....................  ....................  ....................
12383 MW156665.1 |Severe acute respiratory syndrome coro    ....................  ....................  ....................  ....................  ....................  ....................
12384 MW156666.1 |Severe acute respiratory syndrome coro    ....................  ....................  ....................  ....................  ....................  ....................
12385 MW156667.1 |Severe acute respiratory syndrome coro    ....................  ....................  ....................  ....................  ....................  ....................
12387 MW156669.1 |Severe acute respiratory syndrome coro    ....................  ....................  ....................  ....................  ....................  ....................
12388 MW156673.1 |Severe acute respiratory syndrome coro    ....................  ....................  ....................  ....................  ....................  ....................
12389 MW156674.1 |Severe acute respiratory syndrome coro    ....................  ....................  ....................  ....................  ....................  ....................
12390 MW156675.1 |Severe acute respiratory syndrome coro    ....................  ....................  ....................  ....................  ....................  ....................
12391 MW156676.1 |Severe acute respiratory syndrome coro    ....................  ....................  ....................  ....................  ....................  ....................
12393 MW156678.1 |Severe acute respiratory syndrome coro    ....................  ....................  ....................  ....................  ....................  ....................
12394 MW156679.1 |Severe acute respiratory syndrome coro    ....................  ....................  ....................  ....................  ....................  ....................
12395 MW156680.1 |Severe acute respiratory syndrome coro    ....................  ....................  ....................  ....................  ....................  ....................
12396 MW156681.1 |Severe acute respiratory syndrome coro    ....................  ....................  ....................  ....................  ....................  ....................
12397 MW156684.1 |Severe acute respiratory syndrome coro    ....................  ....................  ....................  ....................  ....................  ....................
12398 MW156685.1 |Severe acute respiratory syndrome coro    ....................  ....................  ....................  ....................  ....................  ....................
12399 MW156686.1 |Severe acute respiratory syndrome coro    ....................  ....................  ....................  ....................  ....................  ....................
12400 MW156687.1 |Severe acute respiratory syndrome coro    ....................  ....................  ....................  ....................  ....................  ....................
12401 MW156688.1 |Severe acute respiratory syndrome coro    ....................  ....................  ....................  ....................  ....................  ....................
12402 MW156689.1 |Severe acute respiratory syndrome coro    ....................  ....................  ....................  ....................  ....................  ....................
12403 MW156690.1 |Severe acute respiratory syndrome coro    ....................  ....................  ....................  ....................  ....................  ....................
12404 MW156691.1 |Severe acute respiratory syndrome coro    ....................  ....................  ....................  ....................  ....................  ....................
12405 MW156692.1 |Severe acute respiratory syndrome coro    ....................  ....................  ....................  ....................  ....................  ....................
12406 MW156693.1 |Severe acute respiratory syndrome coro    ....................  ....................  ....................  ....................  ....................  ....................
12407 MW156694.1 |Severe acute respiratory syndrome coro    ....................  ....................  ....................  ....................  ....................  ....................
12408 MW156695.1 |Severe acute respiratory syndrome coro    ....................  ....................  ....................  ....................  ....................  ....................
12409 MW156696.1 |Severe acute respiratory syndrome coro    ....................  ....................  ....................  ....................  ....................  ....................
12410 MW156697.1 |Severe acute respiratory syndrome coro    ....................  ....................  ....................  ....................  ....................  ....................
12411 MW156698.1 |Severe acute respiratory syndrome coro    ....................  ....................  ....................  ....................  ....................  ....................
12412 MW156700.1 |Severe acute respiratory syndrome coro    ....................  ....................  ....................  ....................  ....................  ....................
12413 MW156701.1 |Severe acute respiratory syndrome coro    ....................  ....................  ....................  ....................  ....................  ....................
12414 MW156703.1 |Severe acute respiratory syndrome coro    ....................  ....................  ....................  ....................  ....................  ....................
12415 MW156704.1 |Severe acute respiratory syndrome coro    ....................  ....................  ....................  ....................  ....................  ....................
12416 MW156705.1 |Severe acute respiratory syndrome coro    ....................  ....................  ....................  ....................  ....................  ....................
12417 MW156706.1 |Severe acute respiratory syndrome coro    ....................  ....................  ....................  ....................  ....................  ....................
12418 MW156707.1 |Severe acute respiratory syndrome coro    ....................  ....................  ....................  ....................  ....................  ....................
12419 MW156708.1 |Severe acute respiratory syndrome coro    ....................  ....................  ....................  ....................  ....................  ....................
12420 MW156711.1 |Severe acute respiratory syndrome coro    ....................  ....................  ....................  ....................  ....................  ....................
12421 MW156712.1 |Severe acute respiratory syndrome coro    ....................  ....................  ....................  ....................  ....................  ....................
12422 MW156714.1 |Severe acute respiratory syndrome coro    ....................  ....................  ....................  ....................  ....................  ....................
12423 MW156715.1 |Severe acute respiratory syndrome coro    ....................  ....................  ....................  ....................  ....................  ....................
12424 MW156716.1 |Severe acute respiratory syndrome coro    ....................  ....................  ....................  ....................  ....................  ....................
12425 MW156717.1 |Severe acute respiratory syndrome coro    ....................  ....................  ....................  ....................  ....................  ....................
12426 MW156719.1 |Severe acute respiratory syndrome coro    ....................  ....................  ....................  ....................  ....................  ....................
12427 MW156721.1 |Severe acute respiratory syndrome coro    ....................  ....................  ....................  ....................  ....................  ....................
12428 MW156722.1 |Severe acute respiratory syndrome coro    ....................  ....................  ....................  ....................  ....................  ....................
12429 MW156723.1 |Severe acute respiratory syndrome coro    ....................  ....................  ....................  ....................  ....................  ....................
12430 MW156725.1 |Severe acute respiratory syndrome coro    ....................  ....................  ....................  ....................  ....................  ....................
12431 MW156726.1 |Severe acute respiratory syndrome coro    ....................  ....................  ....................  ....................  ....................  ....................
12432 MW156729.1 |Severe acute respiratory syndrome coro    ....................  ....................  ....................  ....................  ....................  ....................
12433 MW156730.1 |Severe acute respiratory syndrome coro    ....................  ....................  ....................  ....................  ....................  ....................
12434 MW156732.1 |Severe acute respiratory syndrome coro    ....................  ....................  ....................  ....................  ....................  ....................
12435 MW156733.1 |Severe acute respiratory syndrome coro    ....................  ....................  ....................  ....................  ....................  ....................
12436 MW156734.1 |Severe acute respiratory syndrome coro    ....................  ....................  ....................  ....................  ....................  ....................
12437 MW156735.1 |Severe acute respiratory syndrome coro    ....................  ....................  ....................  ....................  ....................  ....................
12438 MW156736.1 |Severe acute respiratory syndrome coro    ....................  ....................  ....................  ....................  ....................  ....................
12439 MW156737.1 |Severe acute respiratory syndrome coro    ....................  ....................  ....................  ....................  ....................  ....................
12440 MW156740.1 |Severe acute respiratory syndrome coro    ....................  ....................  ....................  ....................  ....................  ....................
12441 MW156741.1 |Severe acute respiratory syndrome coro    ....................  ....................  ....................  ....................  ....................  ....................
```

```
12442 MW156743.1 |Severe acute respiratory syndrome coro    ....................  ....................  ....................  ....................  ....................  ....................
12443 MW156744.1 |Severe acute respiratory syndrome coro    ....................  ....................  ....................  ....................  ....................  ....................
12444 MW156747.1 |Severe acute respiratory syndrome coro    ....................  ....................  ....................  ....................  ....................  ....................
12445 MW156748.1 |Severe acute respiratory syndrome coro    ....................  ....................  ....................  ....................  ....................  ....................
12446 MW156750.1 |Severe acute respiratory syndrome coro    ....................  ....................  ....................  ....................  ....................  ....................
12447 MW156752.1 |Severe acute respiratory syndrome coro    ....................  ....................  ....................  ....................  ....................  ....................
12448 MW156753.1 |Severe acute respiratory syndrome coro    ....................  ....................  ....................  ....................  ....................  ....................
12449 MW156755.1 |Severe acute respiratory syndrome coro    ....................  ....................  ....................  ....................  ....................  ....................
12450 MW156756.1 |Severe acute respiratory syndrome coro    ....................  ....................  ....................  ....................  ....................  ....................
12451 MW156758.1 |Severe acute respiratory syndrome coro    ....................  ....................  ....................  ....................  ....................  ....................
12452 MW156759.1 |Severe acute respiratory syndrome coro    ....................  ....................  ....................  ....................  ....................  ....................
12453 MW156762.1 |Severe acute respiratory syndrome coro    ....................  ....................  ....................  ....................  ....................  ....................
12454 MW156767.1 |Severe acute respiratory syndrome coro    ....................  ....................  ....................  ....................  ....................  ....................
12455 MW156768.1 |Severe acute respiratory syndrome coro    ....................  ....................  ....................  ....................  ....................  ....................
12456 MW156769.1 |Severe acute respiratory syndrome coro    ....................  ....................  ....................  ....................  ....................  ....................
12457 MW156770.1 |Severe acute respiratory syndrome coro    ....................  ....................  ....................  ....................  ....................  ....................
12458 MW156772.1 |Severe acute respiratory syndrome coro    ....................  ....................  ....................  ....................  ....................  ....................
12459 MW156773.1 |Severe acute respiratory syndrome coro    ....................  ....................  ....................  ....................  ....................  ....................
12460 MW156775.1 |Severe acute respiratory syndrome coro    ....................  ....................  ....................  ....................  ....................  ....................
12461 MW156776.1 |Severe acute respiratory syndrome coro    ....................  ....................  ....................  ....................  ....................  ....................
12462 MW156777.1 |Severe acute respiratory syndrome coro    ....................  ....................  ....................  ....................  ....................  ....................
12463 MW156778.1 |Severe acute respiratory syndrome coro    ....................  ....................  ....................  ....................  ....................  ....................
12464 MW156779.1 |Severe acute respiratory syndrome coro    ....................  ....................  ....................  ....................  ....................  ....................
12465 MW156781.1 |Severe acute respiratory syndrome coro    ....................  ....................  ....................  ....................  ....................  ....................
12466 MW156782.1 |Severe acute respiratory syndrome coro    ....................  ....................  ....................  ....................  ....................  ....................
12467 MW156784.1 |Severe acute respiratory syndrome coro    ....................  ....................  ....................  ....................  ....................  ....................
12468 MW156786.1 |Severe acute respiratory syndrome coro    ....................  ....................  ....................  ....................  ....................  ....................
12469 MW156787.1 |Severe acute respiratory syndrome coro    ....................  ....................  ....................  ....................  ....................  ....................
12470 MW156788.1 |Severe acute respiratory syndrome coro    ....................  ....................  ....................  ....................  ....................  ....................
12471 MW156789.1 |Severe acute respiratory syndrome coro    ....................  ....................  ....................  ....................  ....................  ....................
12472 MW156791.1 |Severe acute respiratory syndrome coro    ....................  ....................  ....................  ....................  ....................  ....................
12473 MW156792.1 |Severe acute respiratory syndrome coro    ....................  ....................  ....................  ....................  ....................  ....................
12474 MW156793.1 |Severe acute respiratory syndrome coro    ....................  ....................  ....................  ....................  ....................  ....................
12475 MW156794.1 |Severe acute respiratory syndrome coro    ....................  ....................  ....................  ....................  ....................  ....................
12476 MW156795.1 |Severe acute respiratory syndrome coro    ....................  ....................  ....................  ....................  ....................  ....................
12477 MW156797.1 |Severe acute respiratory syndrome coro    ....................  ....................  ....................  ....................  ....................  ....................
12478 MW156798.1 |Severe acute respiratory syndrome coro    ....................  ....................  ....................  ....................  ....................  ....................
12479 MW156800.1 |Severe acute respiratory syndrome coro    ....................  ....................  ....................  ....................  ....................  ....................
12480 MW156802.1 |Severe acute respiratory syndrome coro    ....................  ....................  ....................  ....................  ....................  ....................
12481 MW156805.1 |Severe acute respiratory syndrome coro    ....................  ....................  ....................  ....................  ....................  ....................
12482 MW156806.1 |Severe acute respiratory syndrome coro    ....................  ....................  ....................  ....................  ....................  ....................
12483 MW156807.1 |Severe acute respiratory syndrome coro    ....................  ....................  ....................  ....................  ....................  ....................
12484 MW156809.1 |Severe acute respiratory syndrome coro    ....................  ....................  ....................  ....................  ....................  ....................
12485 MW156810.1 |Severe acute respiratory syndrome coro    ....................  ....................  ....................  ....................  ....................  ....................
12486 MW156812.1 |Severe acute respiratory syndrome coro    ....................  ....................  ....................  ....................  ....................  ....................
12487 MW156817.1 |Severe acute respiratory syndrome coro    ....................  ....................  ....................  ....................  ....................  ....................
12488 MW156818.1 |Severe acute respiratory syndrome coro    ....................  ....................  ....................  ....................  ....................  ....................
12489 MW156822.1 |Severe acute respiratory syndrome coro    ....................  ....................  ....................  ....................  ....................  ....................
12490 MW156823.1 |Severe acute respiratory syndrome coro    ....................  ....................  ....................  ....................  ....................  ....................
12491 MW156824.1 |Severe acute respiratory syndrome coro    ....................  ....................  ....................  ....................  ....................  ....................
12492 MW156825.1 |Severe acute respiratory syndrome coro    ....................  ....................  ....................  ....................  ....................  ....................
12493 MW156826.1 |Severe acute respiratory syndrome coro    ....................  ....................  ....................  ....................  ....................  ....................
12494 MW156827.1 |Severe acute respiratory syndrome coro    ....................  ....................  ....................  ....................  ....................  ....................
12495 MW156829.1 |Severe acute respiratory syndrome coro    ....................  ....................  ....................  ....................  ....................  ....................
12496 MW156830.1 |Severe acute respiratory syndrome coro    ....................  ....................  ....................  ....................  ....................  ....................
12497 MW156831.1 |Severe acute respiratory syndrome coro    ....................  ....................  ....................  ....................  ....................  ....................
12498 MW156832.1 |Severe acute respiratory syndrome coro    ....................  ....................  ....................  ....................  ....................  ....................
12499 MW156833.1 |Severe acute respiratory syndrome coro    ....................  ....................  ....................  ....................  ....................  ....................
12500 MW156834.1 |Severe acute respiratory syndrome coro    ....................  ....................  ....................  ....................  ....................  ....................
12501 MW156835.1 |Severe acute respiratory syndrome coro    ....................  ....................  ....................  ....................  ....................  ....................
12502 MW156836.1 |Severe acute respiratory syndrome coro    ....................  ....................  ....................  ....................  ....................  ....................
12503 MW156837.1 |Severe acute respiratory syndrome coro    ....................  ....................  ....................  ....................  ....................  ....................
12505 MW156839.1 |Severe acute respiratory syndrome coro    ....................  ....................  ....................  ....................  ....................  ....................
12506 MW156842.1 |Severe acute respiratory syndrome coro    ....................  ....................  ....................  ....................  ....................  ....................
12507 MW156843.1 |Severe acute respiratory syndrome coro    ....................  ....................  ....................  ....................  ....................  ....................
12508 MW156844.1 |Severe acute respiratory syndrome coro    ....................  ....................  ....................  ....................  ....................  ....................
12509 MW156846.1 |Severe acute respiratory syndrome coro    ....................  ....................  ....................  ....................  ....................  ....................
12510 MW156848.1 |Severe acute respiratory syndrome coro    ....................  ....................  ....................  ....................  ....................  ....................
12511 MW156849.1 |Severe acute respiratory syndrome coro    ....................  ....................  ....................  ....................  ....................  ....................
12512 MW156850.1 |Severe acute respiratory syndrome coro    ....................  ....................  ....................  ....................  ....................  ....................
12513 MW156851.1 |Severe acute respiratory syndrome coro    ....................  ....................  ....................  ....................  ....................  ....................
12514 MW156853.1 |Severe acute respiratory syndrome coro    ....................  ....................  ....................  ....................  ....................  ....................
12515 MW156855.1 |Severe acute respiratory syndrome coro    ....................  ....................  ....................  ....................  ....................  ....................
12516 MW156856.1 |Severe acute respiratory syndrome coro    ....................  ....................  ....................  ....................  ....................  ....................
12517 MW156857.1 |Severe acute respiratory syndrome coro    ....................  ....................  ....................  ....................  ....................  ....................
12518 MW156858.1 |Severe acute respiratory syndrome coro    ....................  ....................  ....................  ....................  ....................  ....................
12519 MW156860.1 |Severe acute respiratory syndrome coro    ....................  ....................  ....................  ....................  ....................  ....................
12520 MW156861.1 |Severe acute respiratory syndrome coro    ....................  ....................  ....................  ....................  ....................  ....................
```

| | | | | | | | | |
|---|---|---|---|---|---|---|---|---|
| 12521 | MW156862.1 | \|Severe acute respiratory syndrome coro | ···················· | ···················· | ···················· | ···················· | ···················· | ···················· |
| 12522 | MW156863.1 | \|Severe acute respiratory syndrome coro | ···················· | ···················· | ···················· | ···················· | ···················· | ···················· |
| 12523 | MW156865.1 | \|Severe acute respiratory syndrome coro | ···················· | ···················· | ···················· | ···················· | N··················· | ···················· |
| 12524 | MW156866.1 | \|Severe acute respiratory syndrome coro | ···················· | ···················· | ···················· | ···················· | ···················· | ···················· |
| 12525 | MW156868.1 | \|Severe acute respiratory syndrome coro | ···················· | ···················· | ···················· | ···················· | ···················· | ···················· |
| 12526 | MW156871.1 | \|Severe acute respiratory syndrome coro | ···················· | ···················· | ···················· | ···················· | ···················· | ···················· |
| 12527 | MW156872.1 | \|Severe acute respiratory syndrome coro | ···················· | ···················· | ···················· | ···················· | ···················· | ···················· |
| 12528 | MW156873.1 | \|Severe acute respiratory syndrome coro | ···················· | ···················· | ···················· | ···················· | ···················· | ···················· |
| 12529 | MW156874.1 | \|Severe acute respiratory syndrome coro | ···················· | ···················· | ···················· | ···················· | ···················· | ···················· |
| 12530 | MW156875.1 | \|Severe acute respiratory syndrome coro | ···················· | ···················· | ···················· | ···················· | ···················· | ···················· |
| 12531 | MW156876.1 | \|Severe acute respiratory syndrome coro | ···················· | ···················· | ···················· | ···················· | ···················· | ···················· |
| 12532 | MW156877.1 | \|Severe acute respiratory syndrome coro | ···················· | ···················· | ···················· | ···················· | ···················· | ···················· |
| 12533 | MW156878.1 | \|Severe acute respiratory syndrome coro | ···················· | ···················· | ···················· | ···················· | ···················· | ···················· |
| 12534 | MW156880.1 | \|Severe acute respiratory syndrome coro | ···················· | ···················· | ···················· | ···················· | ···················· | ···················· |
| 12535 | MW156883.1 | \|Severe acute respiratory syndrome coro | ···················· | ···················· | ···················· | ···················· | ···················· | ···················· |
| 12536 | MW156884.1 | \|Severe acute respiratory syndrome coro | ···················· | ···················· | ···················· | ···················· | ···················· | ···················· |
| 12537 | MW156885.1 | \|Severe acute respiratory syndrome coro | ···················· | ···················· | ···················· | ···················· | ···················· | ···················· |
| 12538 | MW156886.1 | \|Severe acute respiratory syndrome coro | ···················· | ···················· | ···················· | ···················· | ···················· | ···················· |
| 12539 | MW156887.1 | \|Severe acute respiratory syndrome coro | ···················· | ···················· | ···················· | ···················· | ···················· | ···················· |
| 12540 | MW156888.1 | \|Severe acute respiratory syndrome coro | ···················· | ···················· | ···················· | ···················· | ···················· | ···················· |
| 12541 | MW156889.1 | \|Severe acute respiratory syndrome coro | ···················· | ···················· | ···················· | ···················· | ···················· | ···················· |
| 12542 | MW156890.1 | \|Severe acute respiratory syndrome coro | ···················· | ···················· | ···················· | ···················· | ···················· | ···················· |
| 12543 | MW156891.1 | \|Severe acute respiratory syndrome coro | ···················· | ···················· | ···················· | ···················· | ···················· | ···················· |
| 12545 | MW156894.1 | \|Severe acute respiratory syndrome coro | ···················· | ···················· | ···················· | ···················· | ···················· | ···················· |
| 12546 | MW156895.1 | \|Severe acute respiratory syndrome coro | ···················· | ···················· | ···················· | ···················· | ···················· | ···················· |
| 12547 | MW156897.1 | \|Severe acute respiratory syndrome coro | ···················· | ···················· | ···················· | ···················· | ···················· | ···················· |
| 12548 | MW156898.1 | \|Severe acute respiratory syndrome coro | ···················· | ···················· | ···················· | ···················· | ···················· | ···················· |
| 12549 | MW156899.1 | \|Severe acute respiratory syndrome coro | ···················· | ···················· | ···················· | ···················· | ···················· | ···················· |
| 12550 | MW156901.1 | \|Severe acute respiratory syndrome coro | ···················· | ···················· | ···················· | ···················· | ···················· | ···················· |
| 12551 | MW156902.1 | \|Severe acute respiratory syndrome coro | ···················· | ···················· | ···················· | ···················· | ···················· | ···················· |
| 12552 | MW156904.1 | \|Severe acute respiratory syndrome coro | ···················· | ···················· | ···················· | ···················· | ···················· | ···················· |
| 12553 | MW156905.1 | \|Severe acute respiratory syndrome coro | ···················· | ···················· | ···················· | ···················· | ···················· | ···················· |
| 12554 | MW156906.1 | \|Severe acute respiratory syndrome coro | ···················· | ···················· | ···················· | ···················· | ···················· | ···················· |
| 12555 | MW156907.1 | \|Severe acute respiratory syndrome coro | ···················· | ···················· | ···················· | ···················· | ···················· | ···················· |
| 12556 | MW156908.1 | \|Severe acute respiratory syndrome coro | ···················· | ···················· | ···················· | ···················· | ···················· | ···················· |
| 12557 | MW156909.1 | \|Severe acute respiratory syndrome coro | ···················· | ···················· | ···················· | ···················· | ···················· | ···················· |
| 12558 | MW156910.1 | \|Severe acute respiratory syndrome coro | ···················· | ···················· | ···················· | ···················· | ···················· | ···················· |
| 12559 | MW156911.1 | \|Severe acute respiratory syndrome coro | ···················· | ···················· | ···················· | ···················· | ···················· | ···················· |
| 12560 | MW156912.1 | \|Severe acute respiratory syndrome coro | ···················· | ···················· | ···················· | ···················· | ···················· | ···················· |
| 12561 | MW156913.1 | \|Severe acute respiratory syndrome coro | ···················· | ···················· | ···················· | ···················· | ···················· | ···················· |
| 12562 | MW156915.1 | \|Severe acute respiratory syndrome coro | ···················· | ···················· | ···················· | ···················· | ···················· | ···················· |
| 12563 | MW156916.1 | \|Severe acute respiratory syndrome coro | ···················· | ···················· | ···················· | ···················· | ···················· | ···················· |
| 12564 | MW156917.1 | \|Severe acute respiratory syndrome coro | ···················· | ···················· | ···················· | ···················· | ···················· | ···················· |
| 12565 | MW156918.1 | \|Severe acute respiratory syndrome coro | ···················· | ···················· | ···················· | ···················· | ···················· | ···················· |
| 12566 | MW156920.1 | \|Severe acute respiratory syndrome coro | ···················· | ···················· | ···················· | ···················· | ···················· | ···················· |
| 12567 | MW156922.1 | \|Severe acute respiratory syndrome coro | ···················· | ···················· | ···················· | ···················· | ···················· | ···················· |
| 12568 | MW156923.1 | \|Severe acute respiratory syndrome coro | ···················· | ···················· | ···················· | ···················· | ···················· | ···················· |
| 12569 | MW156925.1 | \|Severe acute respiratory syndrome coro | ···················· | ···················· | ···················· | ···················· | ···················· | ···················· |
| 12570 | MW156926.1 | \|Severe acute respiratory syndrome coro | ···················· | ···················· | ···················· | ···················· | ···················· | ···················· |
| 12571 | MW156927.1 | \|Severe acute respiratory syndrome coro | ···················· | ···················· | ···················· | ···················· | ···················· | ···················· |
| 12572 | MW156930.1 | \|Severe acute respiratory syndrome coro | ···················· | ···················· | ···················· | ···················· | ···················· | ···················· |
| 12573 | MW156931.1 | \|Severe acute respiratory syndrome coro | ···················· | ···················· | ···················· | ···················· | ···················· | ···················· |
| 12574 | MW156934.1 | \|Severe acute respiratory syndrome coro | ···················· | ···················· | ···················· | ···················· | ···················· | ···················· |
| 12575 | MW156935.1 | \|Severe acute respiratory syndrome coro | ···················· | ···················· | ···················· | ···················· | ···················· | ···················· |
| 12576 | MW156936.1 | \|Severe acute respiratory syndrome coro | ···················· | ···················· | ···················· | ···················· | ···················· | ···················· |
| 12577 | MW156937.1 | \|Severe acute respiratory syndrome coro | ···················· | ···················· | ···················· | ···················· | ···················· | ···················· |
| 12578 | MW156938.1 | \|Severe acute respiratory syndrome coro | ···················· | ···················· | ···················· | ···················· | ···················· | ···················· |
| 12579 | MW156940.1 | \|Severe acute respiratory syndrome coro | ···················· | ···················· | ···················· | ···················· | ···················· | ···················· |
| 12580 | MW156941.1 | \|Severe acute respiratory syndrome coro | ···················· | ···················· | ···················· | ···················· | ···················· | ···················· |
| 12581 | MW156942.1 | \|Severe acute respiratory syndrome coro | ···················· | ···················· | ···················· | ···················· | ···················· | ···················· |
| 12582 | MW156943.1 | \|Severe acute respiratory syndrome coro | ···················· | ···················· | ···················· | ···················· | ···················· | ···················· |
| 12583 | MW156944.1 | \|Severe acute respiratory syndrome coro | ···················· | ···················· | ···················· | ···················· | ···················· | ···················· |
| 12584 | MW156946.1 | \|Severe acute respiratory syndrome coro | ···················· | ···················· | ···················· | ···················· | ···················· | ···················· |
| 12585 | MW156947.1 | \|Severe acute respiratory syndrome coro | ···················· | ···················· | ···················· | ···················· | ···················· | ···················· |
| 12586 | MW156948.1 | \|Severe acute respiratory syndrome coro | ···················· | ···················· | ···················· | ···················· | ···················· | ···················· |
| 12587 | MW156950.1 | \|Severe acute respiratory syndrome coro | ···················· | ···················· | ···················· | ···················· | ···················· | ···················· |
| 12588 | MW156954.1 | \|Severe acute respiratory syndrome coro | ···················· | ···················· | ···················· | ···················· | ···················· | ···················· |
| 12589 | MW156956.1 | \|Severe acute respiratory syndrome coro | ···················· | ···················· | ···················· | ···················· | ···················· | ···················· |
| 12590 | MW156957.1 | \|Severe acute respiratory syndrome coro | ···················· | ···················· | ···················· | ···················· | ···················· | ···················· |
| 12591 | MW156960.1 | \|Severe acute respiratory syndrome coro | ···················· | ···················· | ···················· | ···················· | ···················· | ···················· |
| 12592 | MW156963.1 | \|Severe acute respiratory syndrome coro | ···················· | ···················· | ···················· | ···················· | ···················· | ···················· |
| 12593 | MW156964.1 | \|Severe acute respiratory syndrome coro | ···················· | ···················· | ···················· | ···················· | ···················· | ···················· |
| 12594 | MW156965.1 | \|Severe acute respiratory syndrome coro | ···················· | ···················· | ···················· | ···················· | ···················· | ···················· |
| 12595 | MW156966.1 | \|Severe acute respiratory syndrome coro | ···················· | ···················· | ···················· | ···················· | ···················· | ···················· |
| 12596 | MW156969.1 | \|Severe acute respiratory syndrome coro | ···················· | ···················· | ···················· | ···················· | ···················· | ···················· |
| 12597 | MW156970.1 | \|Severe acute respiratory syndrome coro | ···················· | ···················· | ···················· | ···················· | ···················· | ···················· |
| 12598 | MW156972.1 | \|Severe acute respiratory syndrome coro | ···················· | ···················· | ···················· | ···················· | ···················· | ···················· |
| 12599 | MW156973.1 | \|Severe acute respiratory syndrome coro | ···················· | ···················· | ···················· | ···················· | ···················· | ···················· |

12600 MW156974.1 |Severe acute respiratory syndrome coro .................... .................... ..................... .................... .................... ....................
12601 MW156975.1 |Severe acute respiratory syndrome coro .................... .................... ..................... .................... .................... ....................
12602 MW156978.1 |Severe acute respiratory syndrome coro .................... .................... ..................... .................... .................... ....................
12603 MW156980.1 |Severe acute respiratory syndrome coro .................... .................... ..................... .................... .................... ....................
12604 MW156982.1 |Severe acute respiratory syndrome coro .................... .................... ..................... .................... .................... ....................
12605 MW156983.1 |Severe acute respiratory syndrome coro .................... .................... ..................... .................... .................... ....................
12606 MW156984.1 |Severe acute respiratory syndrome coro .................... .................... ..................... .................... .................... ....................
12607 MW156986.1 |Severe acute respiratory syndrome coro .................... .................... ..................... .................... .................... ....................
12608 MW156987.1 |Severe acute respiratory syndrome coro .................... .................... ..................... .................... .................... ....................
12609 MW156989.1 |Severe acute respiratory syndrome coro .................... .................... ..................... .................... .................... ....................
12610 MW156990.1 |Severe acute respiratory syndrome coro .................... .................... ..................... .................... .................... ....................
12611 MW156991.1 |Severe acute respiratory syndrome coro .................... .................... ..................... .................... .................... ....................
12612 MW156992.1 |Severe acute respiratory syndrome coro .................... .................... ..................... .................... .................... ....................
12613 MW156995.1 |Severe acute respiratory syndrome coro .................... .................... ..................... .................... .................... ....................
12614 MW156996.1 |Severe acute respiratory syndrome coro .................... .................... ..................... .................... .................... ....................
12615 MW156997.1 |Severe acute respiratory syndrome coro .................... .................... ..................... .................... .................... ....................
12616 MW156998.1 |Severe acute respiratory syndrome coro .................... .................... ..................... .................... .................... ....................
12617 MW156999.1 |Severe acute respiratory syndrome coro .................... .................... ..................... .................... .................... ....................
12618 MW157000.1 |Severe acute respiratory syndrome coro .................... .................... ..................... .................... .................... ....................
12619 MW157005.1 |Severe acute respiratory syndrome coro .................... .................... ..................... .................... .................... ....................
12620 MW157006.1 |Severe acute respiratory syndrome coro .................... .................... ..................... .................... .................... ....................
12621 MW157007.1 |Severe acute respiratory syndrome coro .................... .................... ..................... .................... .................... ....................
12622 MW157009.1 |Severe acute respiratory syndrome coro .................... .................... ..................... .................... .................... ....................
12623 MW157010.1 |Severe acute respiratory syndrome coro .................... .................... ..................... .................... .................... ....................
12624 MW157011.1 |Severe acute respiratory syndrome coro .................... .................... ..................... .................... .................... ....................
12625 MW157015.1 |Severe acute respiratory syndrome coro .................... .................... ..................... .................... .................... ....................
12626 MW157016.1 |Severe acute respiratory syndrome coro .................... .................... ..................... .................... .................... ....................
12627 MW157017.1 |Severe acute respiratory syndrome coro .................... .................... ..................... .................... .................... ....................
12628 MW157018.1 |Severe acute respiratory syndrome coro .................... .................... ..................... .................... .................... ....................
12629 MW157020.1 |Severe acute respiratory syndrome coro .................... .................... ..................... .................... .................... ....................
12630 MW157021.1 |Severe acute respiratory syndrome coro .................... .................... ..................... .................... .................... ....................
12631 MW157022.1 |Severe acute respiratory syndrome coro .................... .................... ..................... .................... .................... ....................
12632 MW157023.1 |Severe acute respiratory syndrome coro .................... .................... ..................... .................... .................... ....................
12633 MW157025.1 |Severe acute respiratory syndrome coro .................... .................... ..................... .................... .................... ....................
12634 MW157027.1 |Severe acute respiratory syndrome coro .................... .................... ..................... .................... .................... ....................
12635 MW157029.1 |Severe acute respiratory syndrome coro .................... .................... ..................... .................... .................... ....................
12636 MW157033.1 |Severe acute respiratory syndrome coro .................... .................... ..................... .................... .................... ....................
12637 MW157034.1 |Severe acute respiratory syndrome coro .................... .................... ..................... .................... .................... ....................
12638 MW157037.1 |Severe acute respiratory syndrome coro .................... .................... ..................... .................... .................... ....................
12639 MW157039.1 |Severe acute respiratory syndrome coro .................... .................... ..................... .................... .................... ....................
12640 MW157041.1 |Severe acute respiratory syndrome coro .................... .................... ..................... .................... .................... ....................
12641 MW157042.1 |Severe acute respiratory syndrome coro .................... .................... ..................... .................... .................... ....................
12642 MW157043.1 |Severe acute respiratory syndrome coro .................... .................... ..................... .................... .................... ....................
12643 MW157044.1 |Severe acute respiratory syndrome coro .................... .................... ..................... .................... .................... ....................
12644 MW157045.1 |Severe acute respiratory syndrome coro .................... .................... ..................... .................... .................... ....................
12645 MW157046.1 |Severe acute respiratory syndrome coro .................... .................... ..................... .................... .................... ....................
12646 MW157047.1 |Severe acute respiratory syndrome coro .................... .................... ..................... .................... .................... ....................
12647 MW157048.1 |Severe acute respiratory syndrome coro .................... .................... ..................... .................... .................... ....................
12648 MW157049.1 |Severe acute respiratory syndrome coro .................... .................... ..................... .................... .................... ....................
12649 MW157050.1 |Severe acute respiratory syndrome coro .................... .................... ..................... .................... .................... ....................
12650 MW157051.1 |Severe acute respiratory syndrome coro .................... .................... ..................... .................... .................... ....................
12651 MW157054.1 |Severe acute respiratory syndrome coro .................... .................... ..................... .................... .................... ....................
12652 MW157055.1 |Severe acute respiratory syndrome coro .................... .................... ..................... .................... .................... ....................
12653 MW157056.1 |Severe acute respiratory syndrome coro .................... .................... ..................... .................... .................... ....................
12654 MW157057.1 |Severe acute respiratory syndrome coro .................... .................... ..................... .................... .................... ....................
12655 MW157059.1 |Severe acute respiratory syndrome coro .................... .................... ..................... .................... .................... ....................
12656 MW157061.1 |Severe acute respiratory syndrome coro .................... .................... ..................... .................... .................... ....................
12657 MW157062.1 |Severe acute respiratory syndrome coro .................... .................... ..................... .................... .................... ....................
12658 MW157063.1 |Severe acute respiratory syndrome coro .................... .................... ..................... .................... .................... ....................
12659 MW157064.1 |Severe acute respiratory syndrome coro .................... .................... ..................... .................... .................... ....................
12660 MW157065.1 |Severe acute respiratory syndrome coro .................... .................... ..................... .................... .................... ....................
12661 MW157066.1 |Severe acute respiratory syndrome coro .................... .................... ..................... .................... .................... ....................
12662 MW157067.1 |Severe acute respiratory syndrome coro .................... .................... ..................... .................... .................... ....................
12663 MW157068.1 |Severe acute respiratory syndrome coro .................... ....T............... ..................... .................... .................... ....................
12665 MW157071.1 |Severe acute respiratory syndrome coro .................... .................... ..................... .................... .................... ....................
12666 MW157072.1 |Severe acute respiratory syndrome coro .................... .................... ..................... .................... .................... ....................
12667 MW157075.1 |Severe acute respiratory syndrome coro .................... .................... ..................... .................... .................... ....................
12668 MW157076.1 |Severe acute respiratory syndrome coro .................... .................... ..................... .................... .................... ....................
12669 MW157078.1 |Severe acute respiratory syndrome coro .................... .................... ..................... .................... .................... ....................
12671 MW157080.1 |Severe acute respiratory syndrome coro .................... .................... ..................... .................... .................... ....................
12672 MW157081.1 |Severe acute respiratory syndrome coro .................... .................... ..................... .................... .................... ....................
12673 MW157082.1 |Severe acute respiratory syndrome coro .................... .................... ..................... .................... .................... ....................
12674 MW157085.1 |Severe acute respiratory syndrome coro .................... .................... ..................... .................... .................... ....................
12676 MW157089.1 |Severe acute respiratory syndrome coro .................... .................... ..................... .................... .................... ....................
12677 MW157090.1 |Severe acute respiratory syndrome coro .................... .................... ..................... .................... .................... ....................
12678 MW157091.1 |Severe acute respiratory syndrome coro .................... .................... ..................... .................... .................... ....................
12679 MW157094.1 |Severe acute respiratory syndrome coro .................... .................... ..................... .................... .................... ....................
12680 MW157096.1 |Severe acute respiratory syndrome coro .................... .................... ..................... .................... .................... ....................

| | | | | | | | | |
|---|---|---|---|---|---|---|---|---|
| 12681 | MW157097.1 | Severe acute respiratory syndrome coro | .................... | .................... | ..................... | .................... | .................... | .................... |
| 12682 | MW157101.1 | Severe acute respiratory syndrome coro | .................... | .................... | ..................... | .................... | .................... | .................... |
| 12683 | MW157103.1 | Severe acute respiratory syndrome coro | .................... | .................... | ..................... | .................... | .................... | .................... |
| 12684 | MW157104.1 | Severe acute respiratory syndrome coro | .................... | .................... | ..................... | .................... | .................... | .................... |
| 12685 | MW157105.1 | Severe acute respiratory syndrome coro | .................... | .................... | ..................... | .................... | .................... | .................... |
| 12686 | MW157106.1 | Severe acute respiratory syndrome coro | .................... | .................... | ..................... | .................... | .................... | .................... |
| 12687 | MW157107.1 | Severe acute respiratory syndrome coro | .................... | .................... | ..................... | .................... | .................... | .................... |
| 12688 | MW157108.1 | Severe acute respiratory syndrome coro | .................... | .................... | ..................... | .................... | .................... | .................... |
| 12689 | MW157109.1 | Severe acute respiratory syndrome coro | .................... | .................... | ..................... | .................... | .................... | .................... |
| 12690 | MW157110.1 | Severe acute respiratory syndrome coro | .................... | .................... | ..................... | .................... | .................... | .................... |
| 12691 | MW157112.1 | Severe acute respiratory syndrome coro | .................... | .................... | ..................... | .................... | .................... | .................... |
| 12692 | MW157113.1 | Severe acute respiratory syndrome coro | .................... | .................... | ..................... | .................... | .................... | .................... |
| 12693 | MW157114.1 | Severe acute respiratory syndrome coro | .................... | .................... | ..................... | .................... | .................... | .................... |
| 12694 | MW157115.1 | Severe acute respiratory syndrome coro | .................... | .................... | ..................... | .................... | .................... | .................... |
| 12695 | MW157116.1 | Severe acute respiratory syndrome coro | .................... | .................... | ..................... | .................... | .................... | .................... |
| 12696 | MW157119.1 | Severe acute respiratory syndrome coro | .................... | .................... | ..................... | .................... | .................... | .................... |
| 12697 | MW157120.1 | Severe acute respiratory syndrome coro | .................... | .................... | ..................... | .................... | .................... | .................... |
| 12698 | MW157122.1 | Severe acute respiratory syndrome coro | .................... | .................... | ..................... | .................... | .................... | .................... |
| 12699 | MW157124.1 | Severe acute respiratory syndrome coro | .................... | .................... | ..................... | .................... | .................... | .................... |
| 12700 | MW157125.1 | Severe acute respiratory syndrome coro | .................... | .................... | ..................... | .................... | .................... | .................... |
| 12701 | MW157126.1 | Severe acute respiratory syndrome coro | .................... | .................... | ..................... | .................... | .................... | .................... |
| 12702 | MW157127.1 | Severe acute respiratory syndrome coro | .................... | .................... | ..................... | .................... | .................... | .................... |
| 12703 | MW157128.1 | Severe acute respiratory syndrome coro | .................... | .................... | ..................... | .................... | .................... | .................... |
| 12704 | MW157129.1 | Severe acute respiratory syndrome coro | .................... | .................... | ..................... | .................... | .................... | .................... |
| 12705 | MW157133.1 | Severe acute respiratory syndrome coro | .................... | .................... | ..................... | .................... | .................... | .................... |
| 12706 | MW157134.1 | Severe acute respiratory syndrome coro | .................... | .................... | ..................... | .................... | .................... | .................... |
| 12707 | MW157137.1 | Severe acute respiratory syndrome coro | .................... | .................... | ..................... | .................... | .................... | .................... |
| 12708 | MW157141.1 | Severe acute respiratory syndrome coro | .................... | .................... | ..................... | .................... | .................... | .................... |
| 12709 | MW157142.1 | Severe acute respiratory syndrome coro | .................... | .................... | ..................... | .................... | .................... | .................... |
| 12710 | MW157143.1 | Severe acute respiratory syndrome coro | .................... | .................... | ..................... | .................... | .................... | .................... |
| 12711 | MW157144.1 | Severe acute respiratory syndrome coro | .................... | .................... | ..................... | .................... | .................... | .................... |
| 12712 | MW157147.1 | Severe acute respiratory syndrome coro | .................... | .................... | ..................... | .................... | .................... | .................... |
| 12713 | MW157148.1 | Severe acute respiratory syndrome coro | .................... | .................... | ..................... | .................... | .................... | .................... |
| 12714 | MW157149.1 | Severe acute respiratory syndrome coro | .................... | .................... | ..................... | .................... | .................... | .................... |
| 12715 | MW157151.1 | Severe acute respiratory syndrome coro | .................... | .................... | ..................... | .................... | .................... | .................... |
| 12716 | MW157152.1 | Severe acute respiratory syndrome coro | .................... | .................... | ..................... | .................... | .................... | .................... |
| 12717 | MW157154.1 | Severe acute respiratory syndrome coro | .................... | .................... | ..................... | .................... | .................... | .................... |
| 12718 | MW157156.1 | Severe acute respiratory syndrome coro | .................... | .................... | ..................... | .................... | .................... | .................... |
| 12719 | MW157157.1 | Severe acute respiratory syndrome coro | .................... | .................... | ..................... | .................... | .................... | .................... |
| 12720 | MW157159.1 | Severe acute respiratory syndrome coro | .................... | .................... | ..................... | .................... | .................... | .................... |
| 12721 | MW157160.1 | Severe acute respiratory syndrome coro | .................... | .................... | ..................... | .................... | .................... | .................... |
| 12722 | MW157161.1 | Severe acute respiratory syndrome coro | .................... | .................... | ..................... | .................... | .................... | .................... |
| 12723 | MW157165.1 | Severe acute respiratory syndrome coro | .................... | .................... | ..................... | .................... | .................... | .................... |
| 12724 | MW157166.1 | Severe acute respiratory syndrome coro | .................... | .................... | ..................... | .................... | .................... | .................... |
| 12725 | MW157170.1 | Severe acute respiratory syndrome coro | .................... | .................... | ..................... | .................... | .................... | .................... |
| 12726 | MW157171.1 | Severe acute respiratory syndrome coro | .................... | .................... | ..................... | .................... | .................... | .................... |
| 12727 | MW157174.1 | Severe acute respiratory syndrome coro | .................... | .................... | ..................... | .................... | .................... | .................... |
| 12729 | MW157180.1 | Severe acute respiratory syndrome coro | .................... | .................... | ..................... | .................... | .................... | .................... |
| 12730 | MW157183.1 | Severe acute respiratory syndrome coro | .................... | .................... | ..................... | .................... | .................... | .................... |
| 12731 | MW157184.1 | Severe acute respiratory syndrome coro | .................... | .................... | ..................... | .................... | .................... | .................... |
| 12732 | MW157185.1 | Severe acute respiratory syndrome coro | .................... | .................... | ..................... | .................... | .................... | .................... |
| 12733 | MW157188.1 | Severe acute respiratory syndrome coro | .................... | .................... | ..................... | .................... | .................... | .................... |
| 12734 | MW157192.1 | Severe acute respiratory syndrome coro | .................... | .................... | ..................... | .................... | .................... | .................... |
| 12735 | MW157194.1 | Severe acute respiratory syndrome coro | .................... | .................... | ..................... | .................... | .................... | .................... |
| 12736 | MW157195.1 | Severe acute respiratory syndrome coro | .................... | .................... | ..................... | .................... | .................... | .................... |
| 12737 | MW157196.1 | Severe acute respiratory syndrome coro | .................... | .................... | ..................... | .................... | .................... | .................... |
| 12738 | MW157197.1 | Severe acute respiratory syndrome coro | .................... | .................... | ..................... | .................... | .................... | .................... |
| 12739 | MW157199.1 | Severe acute respiratory syndrome coro | .................... | .................... | ..................... | .................... | .................... | .................... |
| 12740 | MW157201.1 | Severe acute respiratory syndrome coro | .................... | .................... | ..................... | .................... | .................... | .................... |
| 12741 | MW157202.1 | Severe acute respiratory syndrome coro | .................... | .................... | ..................... | .................... | .................... | .................... |
| 12742 | MW157205.1 | Severe acute respiratory syndrome coro | .................... | .................... | ..................... | .................... | .................... | .................... |
| 12743 | MW157207.1 | Severe acute respiratory syndrome coro | .................... | .................... | ..................... | .................... | .................... | .................... |
| 12744 | MW157209.1 | Severe acute respiratory syndrome coro | .................... | .................... | ..................... | .................... | .................... | .................... |
| 12745 | MW157212.1 | Severe acute respiratory syndrome coro | .................... | .................... | ..................... | .................... | .................... | .................... |
| 12746 | MW157213.1 | Severe acute respiratory syndrome coro | .................... | .................... | ..................... | .................... | .................... | .................... |
| 12747 | MW157214.1 | Severe acute respiratory syndrome coro | .................... | .................... | ..................... | .................... | .................... | .................... |
| 12748 | MW157218.1 | Severe acute respiratory syndrome coro | .................... | .................... | ..................... | .................... | .................... | .................... |
| 12749 | MW157222.1 | Severe acute respiratory syndrome coro | .................... | .................... | ..................... | .................... | .................... | .................... |
| 12750 | MW157223.1 | Severe acute respiratory syndrome coro | .................... | .................... | ..................... | .................... | .................... | .................... |
| 12751 | MW157227.1 | Severe acute respiratory syndrome coro | .................... | .................... | ..................... | .................... | .................... | .................... |
| 12752 | MW157228.1 | Severe acute respiratory syndrome coro | .................... | .................... | ..................... | .................... | .................... | .................... |
| 12753 | MW157229.1 | Severe acute respiratory syndrome coro | .................... | .................... | ..................... | .................... | .................... | .................... |
| 12754 | MW157230.1 | Severe acute respiratory syndrome coro | .................... | .................... | ..................... | .................... | .................... | .................... |
| 12755 | MW157237.1 | Severe acute respiratory syndrome coro | .................... | .................... | ..................... | .................... | .................... | .................... |
| 12756 | MW157238.1 | Severe acute respiratory syndrome coro | .................... | .................... | ..................... | .................... | .................... | .................... |
| 12757 | MW157239.1 | Severe acute respiratory syndrome coro | .................... | .................... | ..................... | .................... | .................... | .................... |
| 12758 | MW157241.1 | Severe acute respiratory syndrome coro | .................... | .................... | ..................... | .................... | .................... | .................... |
| 12759 | MW157242.1 | Severe acute respiratory syndrome coro | .................... | .................... | ..................... | .................... | .................... | .................... |

| | | | | | | | | |
|---|---|---|---|---|---|---|---|---|
| 12760 | MW157243.1 | \|Severe acute respiratory syndrome coro | .................... | .................... | .................... | .................... | .................... | .................... |
| 12761 | MW157244.1 | \|Severe acute respiratory syndrome coro | .................... | .A.................. | .................... | .................... | .................... | .................... |
| 12762 | MW157245.1 | \|Severe acute respiratory syndrome coro | .................... | .................... | .................... | .................... | .................... | .................... |
| 12763 | MW157246.1 | \|Severe acute respiratory syndrome coro | .................... | .................... | .................... | .................... | .................... | .................... |
| 12764 | MW157247.1 | \|Severe acute respiratory syndrome coro | .................... | .................... | .................... | .................... | .................... | .................... |
| 12765 | MW157249.1 | \|Severe acute respiratory syndrome coro | .................... | .................... | .................... | .................... | .................... | .................... |
| 12766 | MW157250.1 | \|Severe acute respiratory syndrome coro | .................... | .A.................. | .................... | .................... | .................... | .................... |
| 12767 | MW157251.1 | \|Severe acute respiratory syndrome coro | .................... | .................... | .................... | .................... | .................... | .................... |
| 12768 | MW133981.1 | \|Severe acute respiratory syndrome coro | .................... | .................... | .................... | .................... | .................... | .................... |
| 12769 | MW133998.1 | \|Severe acute respiratory syndrome coro | .................... | .................... | .................... | .................... | .................... | .................... |
| 12770 | MW134005.1 | \|Severe acute respiratory syndrome coro | .................... | .................... | .................... | .................... | .................... | .................... |
| 12771 | MW134006.1 | \|Severe acute respiratory syndrome coro | .................... | .................... | .................... | .................... | .................... | .................... |
| 12772 | MW134008.1 | \|Severe acute respiratory syndrome coro | .................... | .................... | .................... | .................... | .................... | .................... |
| 12773 | MW134014.1 | \|Severe acute respiratory syndrome coro | .................... | .................... | .................... | .................... | .................... | .................... |
| 12774 | MW134016.1 | \|Severe acute respiratory syndrome coro | .................... | .A.................. | .................... | .................... | .................... | .................... |
| 12775 | MW134078.1 | \|Severe acute respiratory syndrome coro | .................... | .................... | .................... | .................... | .................... | .................... |
| 12776 | MW134088.1 | \|Severe acute respiratory syndrome coro | .................... | .................... | .................... | .................... | .................... | .................... |
| 12777 | MW134092.1 | \|Severe acute respiratory syndrome coro | .................... | .................... | .................... | .................... | .................... | .................... |
| 12778 | MW134096.1 | \|Severe acute respiratory syndrome coro | .................... | .................... | .................... | .................... | .................... | .................... |
| 12779 | MW134097.1 | \|Severe acute respiratory syndrome coro | .................... | .................... | .................... | .................... | .................... | .................... |
| 12780 | MW134100.1 | \|Severe acute respiratory syndrome coro | .................... | .................... | .................... | .................... | .................... | .................... |
| 12781 | MW134101.1 | \|Severe acute respiratory syndrome coro | .................... | .A.................. | .................... | .................... | .................... | .................... |
| 12782 | MW134104.1 | \|Severe acute respiratory syndrome coro | .................... | .A.................. | .................... | .................... | .................... | .................... |
| 12783 | MW134105.1 | \|Severe acute respiratory syndrome coro | .................... | .................... | .................... | .................... | .................... | .................... |
| 12784 | MW134106.1 | \|Severe acute respiratory syndrome coro | .................... | .................... | ...........T........ | .................... | .................... | .................... |
| 12785 | MW134107.1 | \|Severe acute respiratory syndrome coro | .................... | .A.................. | .................... | .................... | .................... | .................... |
| 12786 | MW134109.1 | \|Severe acute respiratory syndrome coro | .................... | .................... | .................... | .................... | .................... | .................... |
| 12787 | MW134110.1 | \|Severe acute respiratory syndrome coro | .................... | .................... | .................... | .................... | .................... | .................... |
| 12788 | MW134111.1 | \|Severe acute respiratory syndrome coro | .................... | .................... | .................... | .................... | .................... | .................... |
| 12789 | MW134113.1 | \|Severe acute respiratory syndrome coro | .................... | .A.................. | .................... | .................... | .................... | .................... |
| 12790 | MW134115.1 | \|Severe acute respiratory syndrome coro | .................... | .................... | .................... | .................... | .................... | .................... |
| 12791 | MW134117.1 | \|Severe acute respiratory syndrome coro | .................... | .................... | .................... | .................... | .................... | .................... |
| 12792 | MW134119.1 | \|Severe acute respiratory syndrome coro | .................... | .................... | .................... | .................... | .................... | .................... |
| 12793 | MW134120.1 | \|Severe acute respiratory syndrome coro | .................... | .A.................. | .................... | .................... | .................... | .................... |
| 12794 | MW134121.1 | \|Severe acute respiratory syndrome coro | .................... | .A.................. | .................... | .................... | .................... | .................... |
| 12795 | MW134122.1 | \|Severe acute respiratory syndrome coro | .................... | .A.................. | .................... | .................... | .................... | .................... |
| 12796 | MW134124.1 | \|Severe acute respiratory syndrome coro | .................... | .A.................. | .................... | .................... | .................... | .................... |
| 12797 | MW134126.1 | \|Severe acute respiratory syndrome coro | .................... | .................... | .................... | .................... | .................... | .................... |
| 12798 | MW134128.1 | \|Severe acute respiratory syndrome coro | .................... | .................... | .................... | .................... | .................... | .................... |
| 12799 | MW134130.1 | \|Severe acute respiratory syndrome coro | .................... | .................... | .................... | .................... | .................... | .................... |
| 12800 | MW134131.1 | \|Severe acute respiratory syndrome coro | .................... | .A.................. | .................... | .................... | .................... | .................... |
| 12801 | MW134134.1 | \|Severe acute respiratory syndrome coro | .................... | .................... | .................... | .................... | .................... | .................... |
| 12802 | MW134135.1 | \|Severe acute respiratory syndrome coro | .................... | .................... | .................... | .................... | .................... | .................... |
| 12803 | MW134138.1 | \|Severe acute respiratory syndrome coro | .................... | .................... | .................... | .................... | .................... | .................... |
| 12804 | MW134139.1 | \|Severe acute respiratory syndrome coro | .................... | .A.................. | .................... | .................... | .................... | .................... |
| 12805 | MW134140.1 | \|Severe acute respiratory syndrome coro | .................... | .................... | .................... | .................... | .................... | .................... |
| 12806 | MW134141.1 | \|Severe acute respiratory syndrome coro | .................... | .................... | .................... | .................... | .................... | .................... |
| 12807 | MW134153.1 | \|Severe acute respiratory syndrome coro | .................... | .................... | .................... | .................... | .................... | .................... |
| 12808 | MW134154.1 | \|Severe acute respiratory syndrome coro | .................... | .A.................. | .................... | .................... | .................... | .................... |
| 12809 | MW134155.1 | \|Severe acute respiratory syndrome coro | .................... | .................... | .................... | .................... | .................... | .................... |
| 12810 | MW134156.1 | \|Severe acute respiratory syndrome coro | .................... | .................... | .................... | .................... | .................... | .................... |
| 12811 | MW134158.1 | \|Severe acute respiratory syndrome coro | .................... | .................... | .................... | .................... | .................... | .................... |
| 12812 | MW134159.1 | \|Severe acute respiratory syndrome coro | .................... | .................... | .................... | .................... | .................... | .................... |
| 12813 | MW134162.1 | \|Severe acute respiratory syndrome coro | .................... | .................... | .................... | .................... | .................... | .................... |
| 12814 | MW134163.1 | \|Severe acute respiratory syndrome coro | .................... | .................... | .................... | .................... | .................... | .................... |
| 12815 | MW134164.1 | \|Severe acute respiratory syndrome coro | .................... | .................... | .................... | .................... | .................... | .................... |
| 12816 | MW134165.1 | \|Severe acute respiratory syndrome coro | .................... | .A.................. | .................... | .................... | .................... | .................... |
| 12817 | MW134166.1 | \|Severe acute respiratory syndrome coro | .................... | .A.................. | .................... | .................... | .................... | .................... |
| 12818 | MW134167.1 | \|Severe acute respiratory syndrome coro | .................... | .................... | .................... | .................... | .................... | .................... |
| 12819 | MW134168.1 | \|Severe acute respiratory syndrome coro | .................... | .................... | .................... | .................... | .................... | .................... |
| 12820 | MW134169.1 | \|Severe acute respiratory syndrome coro | .................... | .................... | .................... | .................... | .................... | .................... |
| 12821 | MW134170.1 | \|Severe acute respiratory syndrome coro | .................... | .................... | .................... | .................... | .................... | .................... |
| 12822 | MW134171.1 | \|Severe acute respiratory syndrome coro | .................... | .................... | .................... | .................... | .................... | .................... |
| 12823 | MW134172.1 | \|Severe acute respiratory syndrome coro | .................... | .................... | .................... | .................... | .................... | .................... |
| 12824 | MW134173.1 | \|Severe acute respiratory syndrome coro | .................... | .................... | .................... | .................... | .................... | .................... |
| 12825 | MW134174.1 | \|Severe acute respiratory syndrome coro | .................... | .................... | .................... | .................... | .................... | .................... |
| 12826 | MW134175.1 | \|Severe acute respiratory syndrome coro | .................... | .................... | .................... | .................... | .................... | .................... |
| 12827 | MW134176.1 | \|Severe acute respiratory syndrome coro | .................... | .................... | .................... | .................... | .................... | .................... |
| 12828 | MW134177.1 | \|Severe acute respiratory syndrome coro | .................... | .A.................. | .................... | .................... | .................... | .................... |
| 12829 | MW134190.1 | \|Severe acute respiratory syndrome coro | .................... | .................... | .................... | .................... | .................... | .................... |
| 12830 | MW134203.1 | \|Severe acute respiratory syndrome coro | .................... | .................... | .................... | .................... | .................... | .................... |
| 12831 | MW134213.1 | \|Severe acute respiratory syndrome coro | .................... | .................... | .................... | .................... | .................... | .................... |
| 12832 | MW134221.1 | \|Severe acute respiratory syndrome coro | .................... | .A.................. | .................... | .................... | .................... | .................... |
| 12833 | MW134222.1 | \|Severe acute respiratory syndrome coro | .................... | .................... | .................... | .................... | .................... | .................... |
| 12834 | MW134225.1 | \|Severe acute respiratory syndrome coro | .................... | .................... | .................... | .................... | .................... | .................... |
| 12835 | MW134226.1 | \|Severe acute respiratory syndrome coro | .................... | .................... | .................... | .................... | .................... | .................... |
| 12836 | MW134227.1 | \|Severe acute respiratory syndrome coro | .................... | .................... | .................... | .................... | .................... | .................... |
| 12837 | MW134232.1 | \|Severe acute respiratory syndrome coro | .................... | .................... | .................... | .................... | .................... | .................... |

```
12838 MW134234.1 |Severe acute respiratory syndrome coro    ····················  ····················  ····················  ····················  ····················  ····················
12840 MW134244.1 |Severe acute respiratory syndrome coro    ····················  ····················  ····················  ····················  ····················  ····················
12841 MW134247.1 |Severe acute respiratory syndrome coro    ····················  ····················  ····················  ····················  ····················  ····················
12842 MW134251.1 |Severe acute respiratory syndrome coro    ····················  ····················  ····················  ····················  ····················  ····················
12843 MW134252.1 |Severe acute respiratory syndrome coro    ····················  ·A··················  ····················  ····················  ····················  ····················
12844 MW134253.1 |Severe acute respiratory syndrome coro    ····················  ····················  ····················  ····················  ····················  ····················
12845 MW134263.1 |Severe acute respiratory syndrome coro    ····················  ····················  ····················  ····················  ····················  ····················
12846 MW134273.1 |Severe acute respiratory syndrome coro    ····················  ·A··················  ····················  ····················  ····················  ····················
12847 MW134274.1 |Severe acute respiratory syndrome coro    ····················  ····················  ····················  ····················  ····················  ····················
12848 MW134279.1 |Severe acute respiratory syndrome coro    ····················  ·A··················  ····················  ····················  ····················  ····················
12849 MW134283.1 |Severe acute respiratory syndrome coro    ····················  ····················  ····················  ····················  ····················  ····················
12850 MW134289.1 |Severe acute respiratory syndrome coro    ····················  ····················  ····················  ····················  ····················  ····················
12851 MW134293.1 |Severe acute respiratory syndrome coro    ····················  ····················  ····················  ····················  ····················  ····················
12852 MW134294.1 |Severe acute respiratory syndrome coro    ····················  ·A··················  ····················  ····················  ····················  ····················
12853 MW134298.1 |Severe acute respiratory syndrome coro    ····················  ····················  ····················  ····················  ····················  ····················
12854 MW134299.1 |Severe acute respiratory syndrome coro    ····················  ····················  ····················  ····················  ····················  ····················
12855 MW134301.1 |Severe acute respiratory syndrome coro    ····················  ·A··················  ····················  ····················  ····················  ····················
12856 MW134305.1 |Severe acute respiratory syndrome coro    ····················  ····················  ····················  ····················  ····················  ····················
12857 MW134308.1 |Severe acute respiratory syndrome coro    ····················  ····················  ····················  ····················  ····················  ····················
12858 MW134312.1 |Severe acute respiratory syndrome coro    ····················  ····················  ····················  ····················  ····················  ····················
12859 MW134313.1 |Severe acute respiratory syndrome coro    ····················  ·A··················  ····················  ····················  ····················  ····················
12860 MW134315.1 |Severe acute respiratory syndrome coro    ····················  ····················  ····················  ····················  ····················  ····················
12861 MW134316.1 |Severe acute respiratory syndrome coro    ····················  ····················  ····················  ····················  ····················  ····················
12862 MW134317.1 |Severe acute respiratory syndrome coro    ····················  ·A··················  ····················  ····················  ····················  ····················
12863 MW134318.1 |Severe acute respiratory syndrome coro    ····················  ····················  ····················  ····················  ····················  ····················
12864 MW134322.1 |Severe acute respiratory syndrome coro    ····················  ····················  ····················  ····················  ····················  ····················
12865 MW134323.1 |Severe acute respiratory syndrome coro    ····················  ····················  ····················  ····················  ····················  ····················
12866 MW134324.1 |Severe acute respiratory syndrome coro    ····················  ····················  ····················  ····················  ····················  ····················
12867 MW134331.1 |Severe acute respiratory syndrome coro    ····················  ·A··················  ····················  ····················  ····················  ····················
12868 MW134332.1 |Severe acute respiratory syndrome coro    ····················  ·A··················  ····················  ····················  ····················  ···A················
12869 MW134333.1 |Severe acute respiratory syndrome coro    ····················  ·A··················  ····················  ····················  ····················  ····················
12870 MW134334.1 |Severe acute respiratory syndrome coro    ····················  ·A··················  ····················  ····················  ····················  ····················
12871 MW134336.1 |Severe acute respiratory syndrome coro    ····················  ·A··················  ····················  ····················  ····················  ····················
12872 MW134337.1 |Severe acute respiratory syndrome coro    ····················  ·A··················  ····················  ····················  ····················  ····················
12873 MW134339.1 |Severe acute respiratory syndrome coro    ····················  ·A··················  ····················  ····················  ····················  ····················
12874 MW134340.1 |Severe acute respiratory syndrome coro    ····················  ·A··················  ····················  ····················  ····················  ····················
12875 MW134342.1 |Severe acute respiratory syndrome coro    ····················  ·A··················  ····················  ····················  ····················  ····················
12876 MW134343.1 |Severe acute respiratory syndrome coro    ····················  ····················  ····················  ····················  ····················  ····················
12877 MW134344.1 |Severe acute respiratory syndrome coro    ····················  ····················  ····················  ····················  ····················  ····················
12878 MW134346.1 |Severe acute respiratory syndrome coro    ····················  ····················  ····················  ····················  ····················  ····················
12879 MW134347.1 |Severe acute respiratory syndrome coro    ····················  ····················  ····················  ····················  ····················  ····················
12880 MW134352.1 |Severe acute respiratory syndrome coro    ····················  ····················  ····················  ····················  ····················  ····················
12881 MW134353.1 |Severe acute respiratory syndrome coro    ····················  ·A··················  ····················  ····················  ····················  ····················
12882 MW134355.1 |Severe acute respiratory syndrome coro    ····················  ····················  ····················  ····················  ····················  ····················
12883 MW134357.1 |Severe acute respiratory syndrome coro    ····················  ····················  ····················  ····················  ····················  ····················
12884 MW134358.1 |Severe acute respiratory syndrome coro    ····················  ····················  ····················  ····················  ····················  ····················
12885 MW134361.1 |Severe acute respiratory syndrome coro    ····················  ·A··················  ····················  ····················  ····················  ····················
12886 MW134364.1 |Severe acute respiratory syndrome coro    ····················  ·A··················  ····················  ····················  ····················  ····················
12887 MW134365.1 |Severe acute respiratory syndrome coro    ····················  ·A··················  ····················  ····················  ····················  ····················
12888 MW134366.1 |Severe acute respiratory syndrome coro    ····················  ·A··················  ····················  ····················  ····················  ····················
12889 MW134367.1 |Severe acute respiratory syndrome coro    ····················  ·A··················  ····················  ····················  ····················  ····················
12890 MW134368.1 |Severe acute respiratory syndrome coro    ····················  ·A··················  ····················  ····················  ····················  ····················
12891 MW134371.1 |Severe acute respiratory syndrome coro    ····················  ·A··················  ····················  ····················  ····················  ····················
12892 MW134372.1 |Severe acute respiratory syndrome coro    ····················  ·A··················  ····················  ····················  ····················  ····················
12893 MW134374.1 |Severe acute respiratory syndrome coro    ····················  ····················  ····················  ····················  ····················  ····················
12894 MW134380.1 |Severe acute respiratory syndrome coro    ····················  ·A··················  ····················  ····················  ····················  ····················
12895 MW134381.1 |Severe acute respiratory syndrome coro    ····················  ····················  ····················  ····················  ····················  ····················
12896 MW134382.1 |Severe acute respiratory syndrome coro    ····················  ····················  ····················  ····················  ····················  ····················
12897 MW134384.1 |Severe acute respiratory syndrome coro    ····················  ·A··················  ····················  ····················  ····················  ····················
12898 MW134385.1 |Severe acute respiratory syndrome coro    ····················  ····················  ····················  ····················  ····················  ····················
12899 MW134386.1 |Severe acute respiratory syndrome coro    ····················  ·A··················  ····················  ····················  ····················  ····················
12900 MW134387.1 |Severe acute respiratory syndrome coro    ····················  ····················  ····················  ····················  ····················  ····················
12901 MW134388.1 |Severe acute respiratory syndrome coro    ····················  ·A··················  ····················  ····················  ····················  ····················
12902 MW134390.1 |Severe acute respiratory syndrome coro    ····················  ·A··················  ····················  ····················  ····················  ····················
12903 MW134396.1 |Severe acute respiratory syndrome coro    ····················  ·A··················  ····················  ····················  ····················  ····················
12904 MW134401.1 |Severe acute respiratory syndrome coro    ····················  ····················  ····················  ····················  ····················  ····················
12905 MW134402.1 |Severe acute respiratory syndrome coro    ····················  ····················  ····················  ····················  ····················  ····················
12906 MW134403.1 |Severe acute respiratory syndrome coro    ····················  ····················  ····················  ····················  ····················  ····················
12907 MW134404.1 |Severe acute respiratory syndrome coro    ····················  ····················  ····················  ····················  ····················  ····················
12908 MW134406.1 |Severe acute respiratory syndrome coro    ····················  ····················  ····················  ····················  ····················  ····················
12909 MW134407.1 |Severe acute respiratory syndrome coro    ····················  ·A··················  ····················  ····················  ····················  ····················
12910 MW134411.1 |Severe acute respiratory syndrome coro    ····················  ·A··················  ····················  ····················  ····················  ····················
12911 MW134414.1 |Severe acute respiratory syndrome coro    ····················  ·A··················  ····················  ····················  ····················  ····················
12912 MW134417.1 |Severe acute respiratory syndrome coro    ····················  ·A··················  ····················  ····················  ····················  ····················
12913 MW134418.1 |Severe acute respiratory syndrome coro    ····················  ·A··················  ····················  ····················  ····················  ····················
12914 MW134419.1 |Severe acute respiratory syndrome coro    ····················  ·A··················  ····················  ····················  ····················  ····················
12915 MW134420.1 |Severe acute respiratory syndrome coro    ····················  ····················  ····················  ····················  ····················  ····················
12916 MW134421.1 |Severe acute respiratory syndrome coro    ····················  ·A··················  ····················  ····················  ····················  ····················
```

```
12917 MW134422.1 |Severe acute respiratory syndrome coro   ····················  ····················  ····················  ····················  ····················  ····················
12918 MW134423.1 |Severe acute respiratory syndrome coro   ····················  ·A··················  ····················  ····················  ····················  ····················
12919 MW134424.1 |Severe acute respiratory syndrome coro   ····················  ·A··················  ····················  ····················  ····················  ····················
12920 MW134426.1 |Severe acute respiratory syndrome coro   ····················  ····················  ····················  ····················  ····················  ····················
12921 MW134427.1 |Severe acute respiratory syndrome coro   ····················  ·A··················  ····················  ····················  ····················  ····················
12922 MW134428.1 |Severe acute respiratory syndrome coro   ····················  ·A··················  ····················  ····················  ····················  ····················
12923 MW134429.1 |Severe acute respiratory syndrome coro   ····················  ····················  ····················  ····················  ····················  ····················
12924 MW134430.1 |Severe acute respiratory syndrome coro   ····················  ····················  ····················  ····················  ····················  ····················
12925 MW134431.1 |Severe acute respiratory syndrome coro   ····················  ····················  ····················  ····················  ····················  ····················
12926 MW134432.1 |Severe acute respiratory syndrome coro   ····················  ·A··················  ····················  ····················  ····················  ····················
12927 MW134434.1 |Severe acute respiratory syndrome coro   ····················  ·A··················  ····················  ····················  ····················  ····················
12928 MW134435.1 |Severe acute respiratory syndrome coro   ····················  ····················  ····················  ····················  ····················  ····················
12929 MW134436.1 |Severe acute respiratory syndrome coro   ····················  ····················  ····················  ····················  ····················  ····················
12930 MW134437.1 |Severe acute respiratory syndrome coro   ····················  ·A···A··············  ····················  ····················  ····················  ····················
12931 MW134438.1 |Severe acute respiratory syndrome coro   ····················  ·A··················  ····················  ····················  ····················  ····················
12932 MW134439.1 |Severe acute respiratory syndrome coro   ····················  ·A··················  ····················  ····················  ····················  ····················
12933 MW134441.1 |Severe acute respiratory syndrome coro   ····················  ·A··················  ····················  ····················  ····················  ····················
12934 MW134443.1 |Severe acute respiratory syndrome coro   ····················  ·A··················  ····················  ····················  ····················  ····················
12935 MW134444.1 |Severe acute respiratory syndrome coro   ····················  ·A··················  ····················  ····················  ····················  ····················
12936 MW134445.1 |Severe acute respiratory syndrome coro   ····················  ·A··················  ····················  ····················  ····················  ····················
12937 MW134446.1 |Severe acute respiratory syndrome coro   ····················  ····················  ····················  ····················  ····················  ····················
12938 MW134447.1 |Severe acute respiratory syndrome coro   ····················  ·A··················  ····················  ····················  ····················  ····················
12939 MW134448.1 |Severe acute respiratory syndrome coro   ····················  ····················  ····················  ····················  ····················  ····················
12940 MW134449.1 |Severe acute respiratory syndrome coro   ····················  ·A··················  ····················  ····················  ····················  ····················
12941 MW134451.1 |Severe acute respiratory syndrome coro   ····················  ····················  ····················  ····················  ····················  ····················
12942 MW134452.1 |Severe acute respiratory syndrome coro   ····················  ·A··················  ····················  ····················  ····················  ····················
12943 MW134453.1 |Severe acute respiratory syndrome coro   ····················  ····················  ····················  ····················  ····················  ····················
12944 MW134454.1 |Severe acute respiratory syndrome coro   ····················  ····················  ····················  ····················  ····················  ····················
12945 MW134558.1 |Severe acute respiratory syndrome coro   ····················  ····················  ····················  ····················  ····················  ····················
12946 MW134559.1 |Severe acute respiratory syndrome coro   ····················  ····················  ····················  ····················  ····················  ····················
12947 MW134560.1 |Severe acute respiratory syndrome coro   ····················  ····················  ····················  ····················  ····················  ····················
12948 MW134561.1 |Severe acute respiratory syndrome coro   ····················  ····················  ····················  ····················  ····················  ····················
12949 MW134562.1 |Severe acute respiratory syndrome coro   ····················  ····················  ····················  ····················  ····················  ····················
12950 MW134563.1 |Severe acute respiratory syndrome coro   ····················  ····················  ····················  ····················  ····················  ····················
12951 MW134564.1 |Severe acute respiratory syndrome coro   ····················  ····················  ····················  ····················  ····················  ····················
12952 MW134565.1 |Severe acute respiratory syndrome coro   ····················  ····················  ····················  ····················  ····················  ····················
12953 MW134566.1 |Severe acute respiratory syndrome coro   ····················  ····················  ····················  ····················  ····················  ····················
12954 MW134567.1 |Severe acute respiratory syndrome coro   ····················  NN··················  ····················  ····················  ····················  ····················
12955 MW136816.1 |Severe acute respiratory syndrome coro   ····················  ····················  ····················  ····················  ····················  ····················
12956 MW136817.1 |Severe acute respiratory syndrome coro   ····················  ····················  ····················  ····················  ····················  ····················
12957 MW136818.1 |Severe acute respiratory syndrome coro   ····················  ····················  ····················  ····················  ····················  ····················
12958 MW136819.1 |Severe acute respiratory syndrome coro   ····················  ····················  ····················  ····················  ·NN·················  ····················
12959 MW136820.1 |Severe acute respiratory syndrome coro   ····················  ····················  ····················  ····················  ····················  ····················
12960 MW136821.1 |Severe acute respiratory syndrome coro   ····················  ····················  ····················  ····················  ····················  ····················
12961 MW136822.1 |Severe acute respiratory syndrome coro   ····················  ····················  ····················  ····················  ····················  ····················
12962 MW136823.1 |Severe acute respiratory syndrome coro   ····················  ····················  ····················  ····················  ····················  ····················
12963 MW136824.1 |Severe acute respiratory syndrome coro   ····················  ····················  ····················  ····················  ····················  ····················
12964 MW136825.1 |Severe acute respiratory syndrome coro   ····················  ····················  ····················  ····················  ····················  ····················
12965 MW136826.1 |Severe acute respiratory syndrome coro   ····················  ····················  ····················  ····················  ····················  ····················
12966 MW136827.1 |Severe acute respiratory syndrome coro   ····················  ····················  ····················  ····················  ····················  ····················
12967 MW136828.1 |Severe acute respiratory syndrome coro   ····················  ····················  ····················  ····················  ····················  ····················
12968 MW136829.1 |Severe acute respiratory syndrome coro   ····················  ····················  ····················  ····················  ····················  ····················
12969 MW136830.1 |Severe acute respiratory syndrome coro   ····················  ····················  ····················  ····················  ····················  ····················
12970 MW136831.1 |Severe acute respiratory syndrome coro   ····················  ····················  ····················  ····················  ····················  ····················
12971 MW136832.1 |Severe acute respiratory syndrome coro   ····················  ····················  ····················  ····················  ····················  ····················
12972 MW136833.1 |Severe acute respiratory syndrome coro   ····················  ····················  ····················  ····················  ····················  ····················
12973 MW136834.1 |Severe acute respiratory syndrome coro   ····················  ····················  ····················  ····················  ····················  ····················
12974 MW136835.1 |Severe acute respiratory syndrome coro   ····················  ····················  ····················  ····················  ····················  ····················
12975 MW136836.1 |Severe acute respiratory syndrome coro   ····················  ····················  ····················  ····················  ····W···············  ····················
12976 MW136837.1 |Severe acute respiratory syndrome coro   ····················  ····················  ····················  ····················  ····················  ····················
12977 MW136838.1 |Severe acute respiratory syndrome coro   ····················  ····················  ····················  ····················  ····················  ····················
12978 MW136839.1 |Severe acute respiratory syndrome coro   ····················  ····················  ····················  ····················  ····················  ····················
12979 MW136840.1 |Severe acute respiratory syndrome coro   ····················  ····················  ····················  ····················  ····················  ····················
12980 MW136841.1 |Severe acute respiratory syndrome coro   ····················  ····················  ····················  ····················  ····················  ····················
12981 MW136842.1 |Severe acute respiratory syndrome coro   ····················  ····················  ····················  ····················  ····················  ····················
12982 MW136843.1 |Severe acute respiratory syndrome coro   ····················  ····················  ····················  ····················  ····················  ····················
12983 MW136844.1 |Severe acute respiratory syndrome coro   ····················  ····················  ····················  ····················  ····················  ····················
12984 MW136845.1 |Severe acute respiratory syndrome coro   ····················  ····················  ····················  ····················  ····················  ····················
12985 MW136846.1 |Severe acute respiratory syndrome coro   ····················  ····················  ····················  ····················  ····················  ····················
12986 MW136847.1 |Severe acute respiratory syndrome coro   ····················  ····················  ····················  ····················  ····················  ····················
12987 MW136848.1 |Severe acute respiratory syndrome coro   ····················  ····················  ····················  ····················  ····················  ····················
12988 MW136849.1 |Severe acute respiratory syndrome coro   ····················  ····················  ····················  ····················  ····················  ····················
12989 MW136850.1 |Severe acute respiratory syndrome coro   ····················  ···G················  ····················  ····················  ····················  ····················
12990 MW136851.1 |Severe acute respiratory syndrome coro   ····················  ···G················  ····················  ····················  ····················  ····················
12991 MW136852.1 |Severe acute respiratory syndrome coro   ····················  ····················  ····················  ····················  ····················  ····················
12992 MW136853.1 |Severe acute respiratory syndrome coro   ····················  ····················  ····················  ····················  ····················  ····················
12993 MW136854.1 |Severe acute respiratory syndrome coro   ····················  ····················  ····················  ····················  ····················  ····················
12994 MW136855.1 |Severe acute respiratory syndrome coro   ····················  ····················  ····················  ····················  ····N···············  ····················
```

```
12995 MW136856.1 |Severe acute respiratory syndrome coro    ....................  ....................  ....................  ....................  ....................  ....................
12996 MW136857.1 |Severe acute respiratory syndrome coro    ....................  ....................  ....................  ....................  ....................  ....................
12997 MW136858.1 |Severe acute respiratory syndrome coro    ....................  ....G...............  ....................  ....................  ....................  ....................
12998 MW136859.1 |Severe acute respiratory syndrome coro    ....................  ....................  ....................  ....................  ....................  ....................
12999 MW136860.1 |Severe acute respiratory syndrome coro    ....................  ....................  ....................  ....................  ....................  ....................
13000 MW136861.1 |Severe acute respiratory syndrome coro    ....................  ....................  ....................  ....................  ....................  ....................
13001 MW136862.1 |Severe acute respiratory syndrome coro    ....................  ....................  ....................  ....................  ....................  ....................
13002 MW136863.1 |Severe acute respiratory syndrome coro    ....................  ....................  ....................  ....................  ....................  ....................
13003 MW136864.1 |Severe acute respiratory syndrome coro    ....................  ....................  ....................  ....................  ....................  ....................
13004 MW136865.1 |Severe acute respiratory syndrome coro    ....................  ....................  ....................  ....................  ....................  ....................
13005 MW136866.1 |Severe acute respiratory syndrome coro    ....................  ....................  ....................  ....................  ....................  ....................
13006 MW136867.1 |Severe acute respiratory syndrome coro    ....................  ....................  ....................  ....................  ....................  ....................
13007 MW136868.1 |Severe acute respiratory syndrome coro    ....................  ....................  ....................  ....................  ....................  ....................
13008 MW136869.1 |Severe acute respiratory syndrome coro    ....................  ....................  ....................  ....................  ....................  ....................
13009 MW136870.1 |Severe acute respiratory syndrome coro    ....................  ....................  ....................  ....................  ....................  ....................
13010 MW136871.1 |Severe acute respiratory syndrome coro    ....................  ....................  ....................  ....................  ....................  ....................
13011 MW136872.1 |Severe acute respiratory syndrome coro    ....................  ....................  ....................  ....................  ....................  ....................
13012 MW136873.1 |Severe acute respiratory syndrome coro    ....................  ....................  ....................  ....................  ....................  ....................
13013 MW136874.1 |Severe acute respiratory syndrome coro    ....................  ....................  ....................  ....................  ....................  ....................
13014 MW136875.1 |Severe acute respiratory syndrome coro    ....................  ....................  ....................  ....................  ....................  ....................
13015 MW136876.1 |Severe acute respiratory syndrome coro    ....................  ....................  ....................  ....................  ....................  ....................
13016 MW136877.1 |Severe acute respiratory syndrome coro    ....................  ....................  ....................  ....................  ....................  ....................
13017 MW136878.1 |Severe acute respiratory syndrome coro    ....................  ....................  ....................  ....................  ....................  ....................
13018 MW136879.1 |Severe acute respiratory syndrome coro    ....................  ....................  ....................  ....................  ....................  ....................
13019 MW136880.1 |Severe acute respiratory syndrome coro    ....................  ....................  ....................  ....................  ....................  ....................
13020 MW136881.1 |Severe acute respiratory syndrome coro    ....................  ....................  ....................  ....................  ....................  ....................
13021 MW136882.1 |Severe acute respiratory syndrome coro    ....................  ....................  ....................  ....................  ....................  ....................
13022 MW136883.1 |Severe acute respiratory syndrome coro    ....................  ....................  ....................  ....................  ....................  ....................
13023 MW136884.1 |Severe acute respiratory syndrome coro    ....................  ....................  ....................  ....................  ....................  ....................
13024 MW136885.1 |Severe acute respiratory syndrome coro    ....................  ....................  ....................  ....................  ....................  ....................
13025 MW136886.1 |Severe acute respiratory syndrome coro    ....................  ....G.....A.........  ....................  ....................  ....................  ....................
13026 MT810119.1 |Severe acute respiratory syndrome coro    ....................  ....................  ....................  ....................  ....................  ....................
13027 MT952602.1 |Mutant Severe acute respiratory syndro    ....................  ....................  ....................  ....................  ....................  ....................
13028 MW127503.1 |Severe acute respiratory syndrome coro    ....................  .A..................  ....................  ....................  ....................  ....................
13029 MW127504.1 |Severe acute respiratory syndrome coro    ....................  .A..................  ....................  ....................  ....................  ....................
13030 MW127505.1 |Severe acute respiratory syndrome coro    ....................  .A..................  ....................  ....................  ....................  ....................
13031 MW127515.1 |Severe acute respiratory syndrome coro    ....................  ....................  ....................  ....................  ....................  ....................
13032 MW127517.1 |Severe acute respiratory syndrome coro    ....................  ....................  ....................  ....................  ....................  ....................
13033 MW127518.1 |Severe acute respiratory syndrome coro    ....................  ....................  ....................  ....................  ....................  ....................
13034 MW127519.1 |Severe acute respiratory syndrome coro    ....................  ....................  ....................  ....................  ....................  ....................
13035 MW127520.1 |Severe acute respiratory syndrome coro    ....................  ....................  ....................  ....................  ....................  ....................
13036 MW127521.1 |Severe acute respiratory syndrome coro    ....................  ....................  ....................  ....................  ....................  ....................
13037 MW127522.1 |Severe acute respiratory syndrome coro    ....................  ....................  ....................  ....................  ....................  ....................
13038 MW127523.1 |Severe acute respiratory syndrome coro    ....................  ....................  ....................  ....................  ....................  ....................
13039 MW127524.1 |Severe acute respiratory syndrome coro    ....................  ....................  ....................  ....................  ....................  ....................
13040 MW127525.1 |Severe acute respiratory syndrome coro    ....................  ....................  ....................  ....................  ....................  ....................
13041 MW127526.1 |Severe acute respiratory syndrome coro    ....................  ....................  ....................  ....................  ....................  ....................
13042 MW127527.1 |Severe acute respiratory syndrome coro    ....................  ....................  ....................  ....................  ....................  ....................
13043 MW127549.1 |Severe acute respiratory syndrome coro    ....................  ....................  ....................  ....................  ....................  ....................
13044 MW130859.1 |Severe acute respiratory syndrome coro    ....................  ....................  ....................  ....................  ....................  ....................
13045 MW130860.1 |Severe acute respiratory syndrome coro    ....................  ....................  ....................  ....................  ....................  ....................
13046 MW130861.1 |Severe acute respiratory syndrome coro    ....................  ....................  ....................  ....................  ....................  ....................
13047 MW130862.1 |Severe acute respiratory syndrome coro    ....................  ....................  ....................  ....................  ....................  ....................
13048 MW130863.1 |Severe acute respiratory syndrome coro    ....................  ....................  ....................  ....................  ....................  ....................
13049 MW130864.1 |Severe acute respiratory syndrome coro    ....................  ....................  ....................  ....................  ....................  ....................
13050 MW130865.1 |Severe acute respiratory syndrome coro    ....................  ....................  ....................  ....................  ....................  ....................
13051 MW130866.1 |Severe acute respiratory syndrome coro    ....................  ....................  ....................  ....................  ....................  ....................
13052 MW130867.1 |Severe acute respiratory syndrome coro    ....................  ....................  ....................  ....................  ....................  ....................
13053 MW130868.1 |Severe acute respiratory syndrome coro    ....................  ....................  ....................  ....................  ....................  ....................
13054 MW130869.1 |Severe acute respiratory syndrome coro    ....................  ....................  ....................  ....................  ....................  ....................
13055 MW130870.1 |Severe acute respiratory syndrome coro    ....................  ....................  ....................  ....................  ....................  ....................
13056 MW130871.1 |Severe acute respiratory syndrome coro    ....................  ....................  ....................  ....................  ....................  ....................
13057 MW130872.1 |Severe acute respiratory syndrome coro    ....................  ....................  ....................  ....................  ....................  ....................
13058 MW130873.1 |Severe acute respiratory syndrome coro    ....................  ....................  ....................  ....................  ....................  ....................
13059 MW130874.1 |Severe acute respiratory syndrome coro    ....................  ....................  ....................  ....................  ....................  ....................
13060 MW130875.1 |Severe acute respiratory syndrome coro    ....................  ....................  ....................  ....................  ....................  ....................
13061 MW130876.1 |Severe acute respiratory syndrome coro    ....................  ....................  ....................  ....................  ....................  ....................
13062 MW130877.1 |Severe acute respiratory syndrome coro    ....................  ....................  ....................  ....................  ....................  ....................
13063 MW130878.1 |Severe acute respiratory syndrome coro    ....................  ....................  ....................  ....................  ....................  ....................
13064 MW130879.1 |Severe acute respiratory syndrome coro    ....................  ....................  ....................  ....................  ....................  ....................
13065 MW130880.1 |Severe acute respiratory syndrome coro    ....................  ....................  ....................  ....................  ....................  ....................
13066 MW130881.1 |Severe acute respiratory syndrome coro    ....................  ....................  ....................  ....................  ....................  ....................
13067 LC581364.1 |Severe acute respiratory syndrome coro    ....................  ....................  ....................  ....................  ....................  ....................
13068 LC581365.1 |Severe acute respiratory syndrome coro    ....................  ....................  ....................  ....................  ....................  ....................
13069 LC581366.1 |Severe acute respiratory syndrome coro    ....................  ....................  ....................  ....................  ....................  ....................
13070 LC581367.1 |Severe acute respiratory syndrome coro    ....................  ....................  ....................  ....................  ....................  ....................
13071 LC581368.1 |Severe acute respiratory syndrome coro    ....................  ....................  ....................  ....................  ....................  ....................
13072 LC581369.1 |Severe acute respiratory syndrome coro    ....................  ....................  ....................  ....................  ....................  ....................
```

13073 LC581370.1 |Severe acute respiratory syndrome coro ···················· ···················· ···················· ···················· ···················· ····················
13074 LC581371.1 |Severe acute respiratory syndrome coro ···················· ···················· ···················· ···················· ···················· ····················
13075 LC581372.1 |Severe acute respiratory syndrome coro ···················· ···················· ···················· ···················· ···················· ····················
13076 LC581373.1 |Severe acute respiratory syndrome coro ···················· ···················· ···················· ···················· ···················· ····················
13077 LC581374.1 |Severe acute respiratory syndrome coro ···················· ···················· ···················· ···················· ···················· ····················
13078 LC581375.1 |Severe acute respiratory syndrome coro ···················· ···················· ···················· ···················· ···················· ····················
13079 LC581376.1 |Severe acute respiratory syndrome coro ···················· ···················· ···················· ···················· ···················· ····················
13080 MW093453.1 |Severe acute respiratory syndrome coro ···················· ···················· ···················· ···················· ···················· ····················
13081 MW093454.1 |Severe acute respiratory syndrome coro ···················· ···················· ···················· ···················· ···················· ····················
13082 MW093458.1 |Severe acute respiratory syndrome coro ···················· ···················· ···················· ···················· ···················· ····················
13083 MW093467.1 |Severe acute respiratory syndrome coro ···················· ···················· ···················· ···················· ···················· ····················
13084 MW093471.1 |Severe acute respiratory syndrome coro ···················· ···················· ···················· ···················· ···················· ····················
13085 MW093477.1 |Severe acute respiratory syndrome coro ···················· ···················· ···················· ···················· ···················· ····················
13086 MW093478.1 |Severe acute respiratory syndrome coro ···················· ···················· ···················· ···················· ···················· ····················
13087 MW093479.1 |Severe acute respiratory syndrome coro ···················· ···················· ···················· ···················· ···················· ····················
13088 MW093484.1 |Severe acute respiratory syndrome coro ···················· ···················· ···················· ···················· ···················· ····················
13089 MW093485.1 |Severe acute respiratory syndrome coro ···················· ···················· ···················· ···················· ···················· ····················
13090 MW093486.1 |Severe acute respiratory syndrome coro ···················· ···················· ···················· ···················· ···················· ····················
13091 MW093490.1 |Severe acute respiratory syndrome coro ···················· ···················· ···················· ···················· ···················· ····················
13092 MW093532.1 |Severe acute respiratory syndrome coro ···················· ···················· ···················· ···················· ···················· ····················
13093 LR883967.1 |Severe acute respiratory syndrome coro ···················· ···················· ···················· ···················· ···················· ····················
13094 LR883968.1 |Severe acute respiratory syndrome coro ···················· ···················· ···················· ···················· ···················· ····················
13095 LR883969.1 |Severe acute respiratory syndrome coro ···················· ···················· ···················· ···················· ···················· ····················
13096 LR883970.1 |Severe acute respiratory syndrome coro ···················· ···················· ···················· ···················· ···················· ····················
13097 LR883971.1 |Severe acute respiratory syndrome coro ···················· ···················· ···················· ···················· ···················· ····················
13098 LR883972.1 |Severe acute respiratory syndrome coro ···················· ···················· ···················· ···················· ···················· ····················
13099 LR883973.1 |Severe acute respiratory syndrome coro ···················· ···················· ···················· ···················· ···················· ····················
13100 LR883974.1 |Severe acute respiratory syndrome coro ···················· ···················· ···················· ···················· ···················· ····················
13101 LR883975.1 |Severe acute respiratory syndrome coro ···················· ···················· ···················· ···················· ···················· ····················
13102 LR883976.1 |Severe acute respiratory syndrome coro ···················· ···················· ···················· ···················· ···················· ····················
13103 LR883977.1 |Severe acute respiratory syndrome coro ···················· ···················· ···················· ···················· ···················· ····················
13104 LR883978.1 |Severe acute respiratory syndrome coro ···················· ···················· ···················· ···················· ···················· ····················
13105 LR883979.1 |Severe acute respiratory syndrome coro ···················· ···················· ···················· ···················· ···················· ····················
13106 LR883980.1 |Severe acute respiratory syndrome coro ···················· ···················· ···················· ···················· ···················· ····················
13107 LR883981.1 |Severe acute respiratory syndrome coro ···················· ···················· ···················· ···················· ···················· ····················
13108 LR883982.1 |Severe acute respiratory syndrome coro ···················· ···················· ···················· ···················· ···················· ····················
13109 LR883983.1 |Severe acute respiratory syndrome coro ···················· ···················· ···················· ···················· ···················· ····················
13110 LR883984.1 |Severe acute respiratory syndrome coro ···················· ···················· ···················· ···················· ···················· ····················
13111 LR883985.1 |Severe acute respiratory syndrome coro ···················· ···················· ···················· ···················· ···················· ····················
13112 LR883986.1 |Severe acute respiratory syndrome coro ···················· ···················· ···················· ···················· ···················· ····················
13113 LR883987.1 |Severe acute respiratory syndrome coro ···················· ···················· ···················· ···················· ···················· ····················
13114 LR883988.1 |Severe acute respiratory syndrome coro ···················· ···················· ···················· ···················· ···················· ····················
13115 LR883989.1 |Severe acute respiratory syndrome coro ···················· ···················· ···················· ···················· ···················· ····················
13116 LR883990.1 |Severe acute respiratory syndrome coro ···················· ···················· ···················· ···················· ···················· ····················
13117 LR883991.1 |Severe acute respiratory syndrome coro ···················· ···················· ···················· ···················· ···················· ····················
13118 LR883992.1 |Severe acute respiratory syndrome coro ···················· ···················· ···················· ···················· ···················· ····················
13119 LR883993.1 |Severe acute respiratory syndrome coro ···················· ···················· ···················· ···················· ···················· ····················
13120 LR883994.1 |Severe acute respiratory syndrome coro ···················· ···················· ···················· ···················· ···················· ····················
13121 LR883995.1 |Severe acute respiratory syndrome coro ···················· ···················· ···················· ···················· ···················· ····················
13122 LR883996.1 |Severe acute respiratory syndrome coro ···················· ···················· ···················· ···················· ···················· ····················
13123 LR883997.1 |Severe acute respiratory syndrome coro ···················· ···················· ···················· ···················· ···················· ····················
13124 LR883998.1 |Severe acute respiratory syndrome coro ···················· ···················· ···················· ···················· ···················· ····················
13125 LR883999.1 |Severe acute respiratory syndrome coro ···················· ···················· ···················· ···················· ···················· ····················
13126 LR884000.1 |Severe acute respiratory syndrome coro ···················· ···················· ···················· ···················· ···················· ····················
13127 LR884001.1 |Severe acute respiratory syndrome coro ···················· ···················· ···················· ···················· ···················· ····················
13128 LR884002.1 |Severe acute respiratory syndrome coro ···················· ···················· ···················· ···················· ···················· ····················
13129 LR884003.1 |Severe acute respiratory syndrome coro ···················· ···················· ···················· ···················· ···················· ····················
13130 LR884004.1 |Severe acute respiratory syndrome coro ···················· ···················· ···················· ···················· ···················· ····················
13131 LR884005.1 |Severe acute respiratory syndrome coro ···················· ·A·················· ···················· ···················· ···················· ····················
13132 LR884006.1 |Severe acute respiratory syndrome coro ···················· ···················· ···················· ···················· ···················· ····················
13133 LR884007.1 |Severe acute respiratory syndrome coro ···················· ···················· ···················· ···················· ···················· ····················
13134 LR884008.1 |Severe acute respiratory syndrome coro ···················· ···················· ···················· ···················· ···················· ····················
13135 LR884009.1 |Severe acute respiratory syndrome coro ···················· ···················· ···················· ···················· ···················· ····················
13136 LR884010.1 |Severe acute respiratory syndrome coro ···················· ···················· ···················· ···················· ···················· ····················
13137 LR884011.1 |Severe acute respiratory syndrome coro ···················· ···················· ···················· ···················· ···················· ····················
13138 LR884012.1 |Severe acute respiratory syndrome coro ···················· ···················· ···················· ···················· ···················· ····················
13139 LR884013.1 |Severe acute respiratory syndrome coro ···················· ···················· ···················· ···················· ···················· ····················
13140 LR884014.1 |Severe acute respiratory syndrome coro ···················· ···················· ···················· ···················· ···················· ····················
13141 LR884015.1 |Severe acute respiratory syndrome coro ···················· ···················· ···················· ···················· ···················· ····················
13142 LR884016.1 |Severe acute respiratory syndrome coro ···················· ···················· ···················· ···················· ···················· ····················
13143 LR884017.1 |Severe acute respiratory syndrome coro ···················· ···················· ···················· ···················· ···················· ····················
13144 LR884018.1 |Severe acute respiratory syndrome coro ···················· ···················· ···················· ···················· ···················· ····················
13145 LR884019.1 |Severe acute respiratory syndrome coro ···················· ···················· ···················· ···················· ···················· ····················
13146 LR884020.1 |Severe acute respiratory syndrome coro ···················· ···················· ···················· ···················· ···················· ····················
13147 LR884021.1 |Severe acute respiratory syndrome coro ···················· ·A·················· ···················· ···················· ···················· ····················
13148 LR884022.1 |Severe acute respiratory syndrome coro ···················· ···················· ···················· ···················· ···················· ····················
13149 LR884023.1 |Severe acute respiratory syndrome coro ···················· ···················· ···················· ···················· ···················· ····················
13150 LR884024.1 |Severe acute respiratory syndrome coro ···················· ···················· ···················· ···················· ···················· ····················

13151 LR884025.1 |Severe acute respiratory syndrome coro .................... .................... ..................... .................... .................... ....................
13152 LR884026.1 |Severe acute respiratory syndrome coro .................... .................... ..................... .................... .................... ....................
13153 LR884027.1 |Severe acute respiratory syndrome coro .................... .................... ..................... .................... .................... ....................
13154 LR884028.1 |Severe acute respiratory syndrome coro .................... .................... ..................... .................... .................... ....................
13155 LR884029.1 |Severe acute respiratory syndrome coro .................... .................... ..................... .................... .................... ....................
13156 LR884030.1 |Severe acute respiratory syndrome coro .................... .................... ..................... .................... .................... ....................
13157 LR884031.1 |Severe acute respiratory syndrome coro .................... .................... ..................... .................... .................... ....................
13158 LR884032.1 |Severe acute respiratory syndrome coro .................... .................... ..................... .................... .................... ....................
13159 LR884033.1 |Severe acute respiratory syndrome coro .................... .................... ..................... .................... .................... ....................
13160 LR884034.1 |Severe acute respiratory syndrome coro .................... .................... ..................... .................... .................... ....................
13161 LR884035.1 |Severe acute respiratory syndrome coro .................... .................... ..................... .................... .................... ....................
13162 LR884036.1 |Severe acute respiratory syndrome coro .................... .................... ..................... .................... .................... ....................
13163 LR884037.1 |Severe acute respiratory syndrome coro .................... .................... ..................... .................... .................... ..A.................
13164 LR884038.1 |Severe acute respiratory syndrome coro .................... .................... ..................... .................... .................... ....................
13165 LR884039.1 |Severe acute respiratory syndrome coro .................... .................... ..................... .................... .................... ....................
13166 LR884040.1 |Severe acute respiratory syndrome coro .................... .................... ..................... .................... .................... ....................
13167 LR884041.1 |Severe acute respiratory syndrome coro .................... .................... ..................... .................... .................... ....................
13168 LR884042.1 |Severe acute respiratory syndrome coro .................... .................... ..................... .................... .................... ....................
13169 LR884043.1 |Severe acute respiratory syndrome coro .................... .................... ..................... .................... .................... ....................
13170 LR884044.1 |Severe acute respiratory syndrome coro .................... .A.................. ..................... .................... .................... ....................
13171 LR884045.1 |Severe acute respiratory syndrome coro .................... .................... ..................... .................... .................... ....................
13172 LR884046.1 |Severe acute respiratory syndrome coro .................... .................... ..................... .................... .................... ....................
13173 LR884047.1 |Severe acute respiratory syndrome coro .................... .................... ..................... .................... .................... ....................
13174 LR884048.1 |Severe acute respiratory syndrome coro .................... .................... ..................... .................... .................... ....................
13175 LR884049.1 |Severe acute respiratory syndrome coro .................... .................... ..................... .................... .................... ....................
13176 LR884050.1 |Severe acute respiratory syndrome coro .................... .................... ..................... .................... .................... ....................
13177 LR884051.1 |Severe acute respiratory syndrome coro .................... .................... ..................... .................... .................... ....................
13178 LR884052.1 |Severe acute respiratory syndrome coro .................... .................... ..................... .................... .................... ....................
13179 LR884053.1 |Severe acute respiratory syndrome coro .................... .................... ..................... .................... .................... ....................
13180 LR884054.1 |Severe acute respiratory syndrome coro .................... .................... ..................... .................... .................... ....................
13181 LR884055.1 |Severe acute respiratory syndrome coro .................... .................... ..................... .................... .................... ....................
13182 LR884056.1 |Severe acute respiratory syndrome coro .................... .................... ..................... .................... .................... ....................
13183 LR884057.1 |Severe acute respiratory syndrome coro .................... .................... ..................... .................... .................... ....................
13184 LR884058.1 |Severe acute respiratory syndrome coro .................... .................... ..................... .................... .................... ....................
13185 LR884059.1 |Severe acute respiratory syndrome coro .................... .................... ..................... .................... .................... ....................
13186 LR884060.1 |Severe acute respiratory syndrome coro .................... .................... ..................... .................... .................... ....................
13187 LR884061.1 |Severe acute respiratory syndrome coro .................... .................... ..................... .................... .................... ....................
13188 LR884062.1 |Severe acute respiratory syndrome coro .................... .................... ..................... .................... .................... ....................
13189 LR884063.1 |Severe acute respiratory syndrome coro .................... .................... ..................... .................... .................... ....................
13190 LR884064.1 |Severe acute respiratory syndrome coro .................... .................... ..................... .................... .................... ....................
13191 LR884065.1 |Severe acute respiratory syndrome coro .................... .................... ..................... .................... .................... ....................
13192 LR884066.1 |Severe acute respiratory syndrome coro .................... .................... ..................... .................... .................... ....................
13193 LR884067.1 |Severe acute respiratory syndrome coro .................... .................... ..................... .................... .................... ....................
13194 LR884068.1 |Severe acute respiratory syndrome coro .................... .................... ..................... .................... .................... ....................
13195 LR884069.1 |Severe acute respiratory syndrome coro .................... .................... ..................... .................... .................... ....................
13196 LR884070.1 |Severe acute respiratory syndrome coro .................... .................... ..................... .................... .................... ....................
13197 LR884071.1 |Severe acute respiratory syndrome coro .................... .................... ..................... .................... .................... ....................
13198 LR884072.1 |Severe acute respiratory syndrome coro .................... .................... ..................... .................... .................... ....................
13199 LR884073.1 |Severe acute respiratory syndrome coro .................... .................... ..................... .................... .................... ....................
13200 LR884074.1 |Severe acute respiratory syndrome coro .................... .................... ..................... .................... .................... ....................
13201 LR884075.1 |Severe acute respiratory syndrome coro .................... .................... ..................... .................... .................... ....................
13202 LR884076.1 |Severe acute respiratory syndrome coro .................... .................... ..................... .................... .................... ....................
13203 LR884077.1 |Severe acute respiratory syndrome coro .................... .................... ..................... .................... .................... ....................
13204 LR884078.1 |Severe acute respiratory syndrome coro .................... .................... ..................... .................... .................... ....................
13205 LR884079.1 |Severe acute respiratory syndrome coro .................... .................... ..................... .................... .................... ....................
13206 LR884080.1 |Severe acute respiratory syndrome coro .................... .................... ..................... .................... .................... ....................
13207 LR884081.1 |Severe acute respiratory syndrome coro .................... .................... ..................... .................... .................... ....................
13208 LR884082.1 |Severe acute respiratory syndrome coro .................... .................... ..................... .................... .................... ....................
13209 LR884083.1 |Severe acute respiratory syndrome coro .................... .................... ..................... .................... .................... ....................
13210 LR884084.1 |Severe acute respiratory syndrome coro .................... .................... ..................... .................... .................... ....................
13211 LR884085.1 |Severe acute respiratory syndrome coro .................... .................... ..................... .................... .................... ....................
13212 LR884086.1 |Severe acute respiratory syndrome coro .................... .................... ..................... .................... .................... ....................
13213 LR884087.1 |Severe acute respiratory syndrome coro .................... .................... ..................... .................... .................... ....................
13214 LR884088.1 |Severe acute respiratory syndrome coro .................... .................... ..................... .................... .................... ....................
13215 LR884089.1 |Severe acute respiratory syndrome coro .................... .................... ..................... .................... .................... ....................
13216 LR884090.1 |Severe acute respiratory syndrome coro .................... .................... ..................... .................... .................... ....................
13217 LR884091.1 |Severe acute respiratory syndrome coro .................... .................... ..................... .................... .................... ....................
13218 LR884092.1 |Severe acute respiratory syndrome coro .................... .................... ..................... .................... .................... ....................
13219 LR884093.1 |Severe acute respiratory syndrome coro .................... .................... ..................... .................... .................... ....................
13220 LR884094.1 |Severe acute respiratory syndrome coro .................... .................... ..................... .................... .................... ....................
13221 LR884095.1 |Severe acute respiratory syndrome coro .................... .................... ..................... .................... .................... ....................
13222 LR884096.1 |Severe acute respiratory syndrome coro .................... .................... ..................... .................... .................... ....................
13223 LR884097.1 |Severe acute respiratory syndrome coro .................... .................... ..................... .................... .................... ....................
13224 LR884098.1 |Severe acute respiratory syndrome coro .................... .................... ..................... .................... .................... ....................
13225 LR884099.1 |Severe acute respiratory syndrome coro .................... .................... ..................... .................... .................... ....................
13226 LR884100.1 |Severe acute respiratory syndrome coro .................... .................... ..................... .................... .................... ....................
13227 LR884101.1 |Severe acute respiratory syndrome coro .................... .................... ..................... .................... .................... ....................
13228 LR884102.1 |Severe acute respiratory syndrome coro .................... .................... ..................... .................... .................... ....................

13229 LR884103.1 |Severe acute respiratory syndrome coro ···················· ···················· ····················· ···················· ···················· ····················
13230 LR884104.1 |Severe acute respiratory syndrome coro ···················· ···················· ····················· ···················· ···················· ····················
13231 LR884105.1 |Severe acute respiratory syndrome coro ···················· ···················· ····················· ···················· ···················· ····················
13232 LR884106.1 |Severe acute respiratory syndrome coro ···················· ···················· ···········T········· ···················· ···················· ····················
13233 LR884107.1 |Severe acute respiratory syndrome coro ···················· ···················· ····················· ···················· ···················· ····················
13234 LR884108.1 |Severe acute respiratory syndrome coro ···················· ···················· ····················· ···················· ···················· ····················
13235 LR884109.1 |Severe acute respiratory syndrome coro ···················· ···················· ····················· ···················· ···················· ····················
13236 LR884110.1 |Severe acute respiratory syndrome coro ···················· ···················· ····················· ···················· ···················· ····················
13237 LR884111.1 |Severe acute respiratory syndrome coro ···················· ···················· ····················· ···················· ···················· ····················
13238 LR884112.1 |Severe acute respiratory syndrome coro ···················· ···················· ····················· ···················· ···················· ····················
13239 LR884113.1 |Severe acute respiratory syndrome coro ···················· ···················· ····················· ···················· ···················· ····················
13240 LR884114.1 |Severe acute respiratory syndrome coro ···················· ···················· ····················· ···················· ···················· ····················
13241 LR884115.1 |Severe acute respiratory syndrome coro ···················· ···················· ····················· ···················· ···················· ····················
13242 LR884116.1 |Severe acute respiratory syndrome coro ···················· ···················· ····················· ···················· ···················· ····················
13243 LR884117.1 |Severe acute respiratory syndrome coro ···················· ···················· ····················· ···················· ···················· ····················
13244 LR884118.1 |Severe acute respiratory syndrome coro ···················· ···················· ····················· ···················· ···················· ····················
13245 LR884119.1 |Severe acute respiratory syndrome coro ···················· ···················· ····················· ···················· ···················· ····················
13246 LR884120.1 |Severe acute respiratory syndrome coro ···················· ···················· ····················· ···················· ···················· ····················
13247 LR884121.1 |Severe acute respiratory syndrome coro ···················· ···················· ····················· ···················· ···················· ····················
13248 LR884122.1 |Severe acute respiratory syndrome coro ···················· ···················· ····················· ···················· ···················· ····················
13249 LR884123.1 |Severe acute respiratory syndrome coro ···················· ···················· ····················· ···················· ···················· ····················
13250 LR884124.1 |Severe acute respiratory syndrome coro ···················· ···················· ····················· ···················· ···················· ····················
13251 LR884125.1 |Severe acute respiratory syndrome coro ···················· ···················· ····················· ···················· ···················· ····················
13252 LR884126.1 |Severe acute respiratory syndrome coro ···················· ···················· ····················· ···················· ···················· ····················
13253 LR884127.1 |Severe acute respiratory syndrome coro ···················· ···················· ····················· ···················· ···················· ····················
13254 LR884128.1 |Severe acute respiratory syndrome coro ···················· ···················· ····················· ···················· ···················· ····················
13255 LR884129.1 |Severe acute respiratory syndrome coro ···················· ···················· ····················· ···················· ···················· ····················
13256 LR884130.1 |Severe acute respiratory syndrome coro ···················· ···················· ····················· ···················· ···················· ····················
13257 LR884131.1 |Severe acute respiratory syndrome coro ···················· ···················· ····················· ···················· ···················· ····················
13258 LR884132.1 |Severe acute respiratory syndrome coro ···················· ···················· ····················· ···················· ···················· ····················
13259 LR884133.1 |Severe acute respiratory syndrome coro ···················· ···················· ····················· ···················· ···················· ····················
13260 LR884134.1 |Severe acute respiratory syndrome coro ···················· ·A·················· ····················· ···················· ···················· ····················
13261 LR884135.1 |Severe acute respiratory syndrome coro ···················· ···················· ····················· ···················· ···················· ····················
13262 LR884136.1 |Severe acute respiratory syndrome coro ···················· ···················· ····················· ···················· ···················· ····················
13263 LR884137.1 |Severe acute respiratory syndrome coro ···················· ···················· ····················· ···················· ···················· ····················
13264 LR884138.1 |Severe acute respiratory syndrome coro ···················· ···················· ····················· ···················· ···················· ····················
13265 LR884139.1 |Severe acute respiratory syndrome coro ···················· ···················· ····················· ···················· ···················· ····················
13266 LR884140.1 |Severe acute respiratory syndrome coro ···················· ···················· ····················· ···················· ···················· ····················
13267 LR884141.1 |Severe acute respiratory syndrome coro ···················· ···················· ····················· ···················· ···················· ····················
13268 LR884142.1 |Severe acute respiratory syndrome coro ···················· ···················· ····················· ···················· ···················· ····················
13269 LR884143.1 |Severe acute respiratory syndrome coro ···················· ···················· ····················· ···················· ···················· ····················
13270 LR884144.1 |Severe acute respiratory syndrome coro ···················· ·A·················· ····················· ···················· ···················· ····················
13271 LR884145.1 |Severe acute respiratory syndrome coro ···················· ···················· ····················· ···················· ···················· ····················
13272 LR884146.1 |Severe acute respiratory syndrome coro ···················· ···················· ····················· ···················· ···················· ····················
13273 LR884147.1 |Severe acute respiratory syndrome coro ···················· ···················· ····················· ···················· ···················· ····················
13274 LR884148.1 |Severe acute respiratory syndrome coro ···················· ···················· ····················· ···················· ···················· ····················
13275 LR884149.1 |Severe acute respiratory syndrome coro ···················· ···················· ····················· ···················· ···················· ····················
13276 LR884150.1 |Severe acute respiratory syndrome coro ···················· ···················· ····················· ···················· ···················· ····················
13277 LR884151.1 |Severe acute respiratory syndrome coro ···················· ···················· ····················· ···················· ···················· ····················
13278 LR884152.1 |Severe acute respiratory syndrome coro ···················· ···················· ····················· ···················· ···················· ····················
13279 LR884153.1 |Severe acute respiratory syndrome coro ···················· ···················· ····················· ···················· ···················· ····················
13280 LR884154.1 |Severe acute respiratory syndrome coro ···················· ···················· ····················· ···················· ···················· ····················
13281 LR884155.1 |Severe acute respiratory syndrome coro ···················· ···················· ····················· ···················· ···················· ····················
13282 LR884156.1 |Severe acute respiratory syndrome coro ···················· ···················· ····················· ···················· ···················· ····················
13283 LR884157.1 |Severe acute respiratory syndrome coro ···················· ···················· ····················· ···················· ···················· ····················
13284 LR884158.1 |Severe acute respiratory syndrome coro ···················· ···················· ····················· ···················· ···················· ····················
13285 LR884159.1 |Severe acute respiratory syndrome coro ···················· ···················· ····················· ···················· ···················· ····················
13286 LR884160.1 |Severe acute respiratory syndrome coro ···················· ···················· ····················· ···················· ···················· ····················
13287 LR884161.1 |Severe acute respiratory syndrome coro ···················· ···················· ····················· ···················· ···················· ····················
13288 LR884162.1 |Severe acute respiratory syndrome coro ···················· ···················· ····················· ···················· ···················· ····················
13289 LR884163.1 |Severe acute respiratory syndrome coro ···················· ···················· ····················· ···················· ···················· ····················
13290 LR884164.1 |Severe acute respiratory syndrome coro ···················· ···················· ····················· ···················· ···················· ····················
13291 LR884165.1 |Severe acute respiratory syndrome coro ···················· ···················· ····················· ···················· ···················· ····················
13292 LR884166.1 |Severe acute respiratory syndrome coro ···················· ···················· ····················· ···················· ···················· ····················
13293 LR884167.1 |Severe acute respiratory syndrome coro ···················· ···················· ····················· ···················· ···················· ····················
13294 LR884168.1 |Severe acute respiratory syndrome coro ···················· ···················· ····················· ···················· ···················· ····················
13295 LR884169.1 |Severe acute respiratory syndrome coro ···················· ···················· ····················· ···················· ···················· ····················
13296 LR884170.1 |Severe acute respiratory syndrome coro ···················· ···················· ····················· ···················· ···················· ····················
13297 LR884171.1 |Severe acute respiratory syndrome coro ···················· ···················· ····················· ···················· ···················· ····················
13298 LR884172.1 |Severe acute respiratory syndrome coro ···················· ···················· ····················· ···················· ···················· ····················
13299 LR884173.1 |Severe acute respiratory syndrome coro ···················· ···················· ····················· ···················· ···················· ····················
13300 LR884174.1 |Severe acute respiratory syndrome coro ···················· ···A················ ····················· ···················· ···················· ····················
13301 LR884175.1 |Severe acute respiratory syndrome coro ···················· ···················· ····················· ···················· ···················· ····················
13302 LR884176.1 |Severe acute respiratory syndrome coro ···················· ···················· ····················· ···················· ···················· ····················
13303 LR884177.1 |Severe acute respiratory syndrome coro ···················· ···················· ····················· ···················· ···················· ····················
13304 LR884178.1 |Severe acute respiratory syndrome coro ···················· ···················· ····················· ···················· ···················· ····················
13305 LR884179.1 |Severe acute respiratory syndrome coro ···················· ···················· ····················· ···················· ···················· ····················
13306 LR884180.1 |Severe acute respiratory syndrome coro ···················· ···················· ····················· ···················· ···················· ····················

```
13307 LR884181.1 |Severe acute respiratory syndrome coro    ....................  ....................  ....................  ....................  ....................  ....................
13308 LR884182.1 |Severe acute respiratory syndrome coro    ....................  ....................  ....................  ....................  ....................  ....................
13309 LR884183.1 |Severe acute respiratory syndrome coro    ....................  ....................  ....................  ....................  ....................  ....................
13310 LR884184.1 |Severe acute respiratory syndrome coro    ....................  ....................  ....................  ....................  ....................  ....................
13311 LR884185.1 |Severe acute respiratory syndrome coro    ....................  ....................  ....................  ....................  ....................  ....................
13312 LR884186.1 |Severe acute respiratory syndrome coro    ....................  ....................  ....................  ....................  ....................  ....................
13313 LR884187.1 |Severe acute respiratory syndrome coro    ....................  ....................  ....................  ....................  ....................  ....................
13314 LR884188.1 |Severe acute respiratory syndrome coro    ....................  ....................  ....................  ....................  ....................  ....................
13315 LR884189.1 |Severe acute respiratory syndrome coro    ....................  ....................  ....................  ....................  ....................  ....................
13316 LR884190.1 |Severe acute respiratory syndrome coro    ....................  ....................  ....................  ....................  ....................  ....................
13317 MW084406.1 |Severe acute respiratory syndrome coro    ....................  ....................  ....................  ....................  ....................  ....................
13318 MW084414.1 |Severe acute respiratory syndrome coro    ....................  ....................  ....................  ....................  ....................  ....................
13319 MW084419.1 |Severe acute respiratory syndrome coro    ....................  ....................  ....................  ....................  ....................  ....................
13320 MW084426.1 |Severe acute respiratory syndrome coro    ....................  ....................  ....................  ....................  ....................  ....................
13321 MW084440.1 |Severe acute respiratory syndrome coro    ....................  ....................  ....................  ....................  ....................  ....................
13322 MW084441.1 |Severe acute respiratory syndrome coro    ....................  ....................  ....................  ....................  ....................  ....................
13323 MW084469.1 |Severe acute respiratory syndrome coro    ....................  ....................  ....................  ....................  ....................  ....................
13324 MW084505.1 |Severe acute respiratory syndrome coro    ....................  ....................  ....................  ....................  ....................  ....................
13325 MW084509.1 |Severe acute respiratory syndrome coro    ....................  ....................  ....................  ....................  ....................  ....................
13326 MW084516.1 |Severe acute respiratory syndrome coro    ....................  ....................  ....................  ....................  ....................  ....................
13327 MW084517.1 |Severe acute respiratory syndrome coro    ....................  ....................  ....................  ....................  ....................  ....................
13328 MW084518.1 |Severe acute respiratory syndrome coro    ....................  ....................  ....................  ....................  ....................  ....................
13329 MW084520.1 |Severe acute respiratory syndrome coro    ....................  ....................  ....................  ....................  ....................  ....................
13330 MW084522.1 |Severe acute respiratory syndrome coro    ....................  ....................  ....................  ....................  ....................  ....................
13331 MW084525.1 |Severe acute respiratory syndrome coro    ....................  ....................  ....................  ....................  ....................  ....................
13332 MW084527.1 |Severe acute respiratory syndrome coro    ....................  ....................  ....................  ....................  ....................  ....................
13333 MW084536.1 |Severe acute respiratory syndrome coro    ....................  ....................  ....................  ....................  ....................  ....................
13334 MW084553.1 |Severe acute respiratory syndrome coro    ....................  ....................  ....................  ....................  ....................  ....................
13335 MW084559.1 |Severe acute respiratory syndrome coro    ....................  ....................  ....................  ....................  ....................  ....................
13336 MW084561.1 |Severe acute respiratory syndrome coro    ....................  ....................  ....................  ....................  ....................  ....................
13337 MW084567.1 |Severe acute respiratory syndrome coro    ....................  ....................  ....................  ....................  ....................  ....................
13338 MW084577.1 |Severe acute respiratory syndrome coro    ....................  ....................  ....................  ....................  ....................  ....................
13339 MW084583.1 |Severe acute respiratory syndrome coro    ....................  ....................  ....................  ....................  ....................  ....................
13340 MW084585.1 |Severe acute respiratory syndrome coro    ....................  ....................  ....................  ....................  ....................  ....................
13341 MW084596.1 |Severe acute respiratory syndrome coro    ....................  ....................  ....................  ....................  ....................  ....................
13342 MW084599.1 |Severe acute respiratory syndrome coro    ....................  ....................  ....................  ....................  ....................  ....................
13343 MW084600.1 |Severe acute respiratory syndrome coro    ....................  ....................  ....................  ....................  ....................  ....................
13344 MW084601.1 |Severe acute respiratory syndrome coro    ....................  ....................  ....................  ....................  ....................  ....................
13345 MW084605.1 |Severe acute respiratory syndrome coro    ....................  ....................  ....................  ....................  ....................  ....................
13346 MW086594.1 |Severe acute respiratory syndrome coro    ....................  ....................  ....................  ....................  ....................  ....................
13347 MW086595.1 |Severe acute respiratory syndrome coro    ....................  ....................  ....................  ....................  ....................  ....................
13348 MW086596.1 |Severe acute respiratory syndrome coro    ....................  ....................  ....................  ....................  ....................  ....................
13349 MW086597.1 |Severe acute respiratory syndrome coro    ....................  ....................  ....................  ....................  ....................  ....................
13350 MW086598.1 |Severe acute respiratory syndrome coro    ....................  ....................  ....................  ....................  ....................  ....................
13351 MW086893.1 |Severe acute respiratory syndrome coro    ....................  ...G................  ....................  ....................  ....................  ....................
13352 MW086894.1 |Severe acute respiratory syndrome coro    ....................  ....................  ....................  ....................  ....................  ....................
13353 MW086895.1 |Severe acute respiratory syndrome coro    ....................  ....................  ....................  ....................  ....................  ....................
13354 MW086896.1 |Severe acute respiratory syndrome coro    ....................  ....................  ....................  ....................  ....................  ....................
13355 MW086897.1 |Severe acute respiratory syndrome coro    ....................  ....................  ....................  ....................  ....................  ....................
13356 MW086898.1 |Severe acute respiratory syndrome coro    ....................  ....................  ....................  ....................  ....................  ....................
13357 MW086899.1 |Severe acute respiratory syndrome coro    ....................  ....................  ....................  ....................  ....................  ....................
13358 MW086900.1 |Severe acute respiratory syndrome coro    ....................  ....................  ....................  ....................  ....................  ....................
13359 MW086901.1 |Severe acute respiratory syndrome coro    ....................  ....................  ....................  ....................  ....................  ....................
13360 MW086902.1 |Severe acute respiratory syndrome coro    ....................  ....................  ....................  ....................  ....................  ....................
13361 MW086903.1 |Severe acute respiratory syndrome coro    ....................  ....................  ....................  ....................  ....................  ....................
13362 MW086904.1 |Severe acute respiratory syndrome coro    ....................  ....................  ....................  ....................  ....................  ....................
13363 MW086905.1 |Severe acute respiratory syndrome coro    ....................  ....................  ....................  ....................  ....................  ....................
13364 MW086906.1 |Severe acute respiratory syndrome coro    ....................  ....................  ....................  ....................  ....................  ....................
13365 MW086907.1 |Severe acute respiratory syndrome coro    ....................  ....................  ....................  ....................  ....................  ....................
13366 MW086908.1 |Severe acute respiratory syndrome coro    ....................  ....................  ....................  ....................  ....................  ....................
13367 MW086909.1 |Severe acute respiratory syndrome coro    ....................  ....................  ....................  ....................  ....................  ....................
13368 MW086910.1 |Severe acute respiratory syndrome coro    ....................  ....................  ....................  ....................  ....................  ....................
13369 MW086911.1 |Severe acute respiratory syndrome coro    ....................  ....................  ....................  ....................  ....................  ....................
13370 MW086912.1 |Severe acute respiratory syndrome coro    ....................  ....................  ....................  ....................  ....................  ....................
13371 MW086913.1 |Severe acute respiratory syndrome coro    ....................  ....................  ....................  ....................  ....................  ....................
13372 MW086914.1 |Severe acute respiratory syndrome coro    ....................  ....................  ....................  ....................  ....................  ....................
13373 MW086915.1 |Severe acute respiratory syndrome coro    ....................  ....................  ....................  ....................  ....................  ....................
13374 MW086916.1 |Severe acute respiratory syndrome coro    ....................  ....................  ....................  ....................  ....................  ....................
13375 MW086917.1 |Severe acute respiratory syndrome coro    ....................  ....................  ....................  ....................  ....................  ....................
13376 MW086918.1 |Severe acute respiratory syndrome coro    ....................  ....................  ....................  ....................  ....................  ....................
13377 MW086919.1 |Severe acute respiratory syndrome coro    ....................  ....................  ....................  ....................  ....................  ....................
13378 MW086920.1 |Severe acute respiratory syndrome coro    ....................  ....................  ....................  ....................  ....................  ....................
13379 MW086921.1 |Severe acute respiratory syndrome coro    ....................  ....................  ....................  ....................  ....................  ....................
13380 MW077466.1 |Severe acute respiratory syndrome coro    ....................  .A..................  ....................  ....................  ....................  ....................
13381 MW077467.1 |Severe acute respiratory syndrome coro    ....................  .A..................  ....................  ....................  ....................  ....................
13382 MW077468.1 |Severe acute respiratory syndrome coro    ....................  .A..................  ....................  ....................  ....................  ....................
13383 MW077469.1 |Severe acute respiratory syndrome coro    ....................  .A..................  ....................  ....................  ....................  ....................
13384 MW077470.1 |Severe acute respiratory syndrome coro    ....................  .A..................  ....................  ....................  ....................  ....................
```

13385 MW077471.1 |Severe acute respiratory syndrome coro ···················· ···················· ···················· ···················· ···················· ····················
13386 MW077472.1 |Severe acute respiratory syndrome coro ···················· ·A·················· ···················· ···················· ···················· ····················
13387 MW077473.1 |Severe acute respiratory syndrome coro ···················· ·A·················· ···················· ···················· ···················· ····················
13388 MW077474.1 |Severe acute respiratory syndrome coro ···················· ···················· ···················· ···················· ···················· ····················
13389 MW077475.1 |Severe acute respiratory syndrome coro ···················· ···················· ···················· ···················· ···················· ····················
13390 MW077476.1 |Severe acute respiratory syndrome coro ···················· ············A······· ···················· ···················· ···················· ····················
13391 MW077477.1 |Severe acute respiratory syndrome coro ···················· ···················· ···················· ···················· ···················· ····················
13392 MW077478.1 |Severe acute respiratory syndrome coro ···················· ···················· ···················· ···················· ···················· ····················
13393 MW077479.1 |Severe acute respiratory syndrome coro ···················· ···················· ···················· ···················· ···················· ····················
13394 MW077480.1 |Severe acute respiratory syndrome coro ···················· ···················· ···················· ···················· ···················· ····················
13395 MW077481.1 |Severe acute respiratory syndrome coro ···················· ···················· ···················· ···················· ···················· ····················
13396 MW077482.1 |Severe acute respiratory syndrome coro ···················· ·A·················· ···················· ···················· ···················· ····················
13397 MW077483.1 |Severe acute respiratory syndrome coro ···················· ···················· ···················· ···················· ···················· ····················
13398 MW077485.1 |Severe acute respiratory syndrome coro ···················· ···················· ···················· ···················· ···················· ····················
13399 MW077486.1 |Severe acute respiratory syndrome coro ···················· ············A······· ···················· ···················· ···················· ····················
13400 MW077487.1 |Severe acute respiratory syndrome coro ···················· ···················· ···················· ···················· ···················· ····················
13401 MW077489.1 |Severe acute respiratory syndrome coro ···················· ············A······· ···················· ···················· ···················· ····················
13402 MW077491.1 |Severe acute respiratory syndrome coro ···················· ···················· ···················· ···················· ···················· ····················
13403 MW077492.1 |Severe acute respiratory syndrome coro ···················· ···················· ···················· ···················· ···················· ····················
13404 MW077493.1 |Severe acute respiratory syndrome coro ···················· ···················· ···················· ···················· ···················· ····················
13405 MW077494.1 |Severe acute respiratory syndrome coro ···················· ···················· ···················· ···················· ···················· ····················
13406 MW077495.1 |Severe acute respiratory syndrome coro ···················· ·A·················· ···················· ···················· ···················· ····················
13407 MW077496.1 |Severe acute respiratory syndrome coro ···················· ···················· ···················· ···················· ···················· ····················
13408 MW077497.1 |Severe acute respiratory syndrome coro ···················· ·A·················· ···················· ···················· ···················· ····················
13409 MW077498.1 |Severe acute respiratory syndrome coro ···················· ·A·················· ···················· ···················· ···················· ····················
13410 MW077500.1 |Severe acute respiratory syndrome coro ···················· ·A·················· ···················· ···················· ···················· ····················
13411 MW077501.1 |Severe acute respiratory syndrome coro ···················· ·A·················· ···················· ···················· ···················· ····················
13412 MW077502.1 |Severe acute respiratory syndrome coro ···················· ·A·················· ···················· ···················· ···················· ····················
13413 MW079418.1 |Severe acute respiratory syndrome coro ···················· ·A·················· ···················· ···················· ···················· ····················
13414 MW079419.1 |Severe acute respiratory syndrome coro ···················· ·A·················· ···················· ···················· ···················· ····················
13415 MW079421.1 |Severe acute respiratory syndrome coro ···················· ·A·················· ···················· ···················· ···················· ····················
13416 MW079422.1 |Severe acute respiratory syndrome coro ···················· ·A·················· ···················· ···················· ···················· ····················
13417 MW079428.1 |Severe acute respiratory syndrome coro ···················· ···················· ···················· ···················· ···················· ···A················
13418 MW079429.1 |Severe acute respiratory syndrome coro ···················· ···················· ···················· ···················· ···················· ····················
13419 MW079819.1 |Severe acute respiratory syndrome coro ···················· ···················· ···················· ···················· ···················· ····················
13420 MW079820.1 |Severe acute respiratory syndrome coro ···················· ·A·················· ···················· ···················· ···················· ····················
13421 MW079821.1 |Severe acute respiratory syndrome coro ···················· ·A·················· ···················· ···················· ···················· ····················
13422 MW079822.1 |Severe acute respiratory syndrome coro ···················· ·A·················· ···················· ···················· ···················· ····················
13423 MW079823.1 |Severe acute respiratory syndrome coro ···················· ·A··G··············· ···················· ···················· ···················· ····················
13424 MW079824.1 |Severe acute respiratory syndrome coro ···················· ···················· ···················· ···················· ···················· ····················
13425 MW079825.1 |Severe acute respiratory syndrome coro ···················· ···················· ···················· ···················· ···················· ····················
13426 MW079826.1 |Severe acute respiratory syndrome coro ···················· ···················· ···················· ···················· ···················· ····················
13427 MW079827.1 |Severe acute respiratory syndrome coro ···················· ···················· ···················· ···················· ···················· ····················
13428 MW079828.1 |Severe acute respiratory syndrome coro ···················· ···················· ···················· ···················· ···················· ····················
13429 MW079829.1 |Severe acute respiratory syndrome coro ···················· ···················· ···················· ···················· ···················· ····················
13430 MW079830.1 |Severe acute respiratory syndrome coro ···················· ···················· ···················· ···················· ···················· ····················
13431 MW079831.1 |Severe acute respiratory syndrome coro ···················· ···················· ···················· ···················· ···················· ····················
13432 MW079832.1 |Severe acute respiratory syndrome coro ···················· ···················· ···················· ···················· ···················· ····················
13433 MW079833.1 |Severe acute respiratory syndrome coro ···················· ···················· ···················· ···················· ···················· ····················
13434 MW079834.1 |Severe acute respiratory syndrome coro ···················· ···················· ···················· ···················· ···················· ····················
13435 MW079835.1 |Severe acute respiratory syndrome coro ···················· ···················· ···················· ···················· ···················· ····················
13436 MW079836.1 |Severe acute respiratory syndrome coro ···················· ···················· ···················· ···················· ···················· ····················
13437 MW079837.1 |Severe acute respiratory syndrome coro ···················· ···················· ···················· ···················· ···················· ····················
13438 MW079838.1 |Severe acute respiratory syndrome coro ···················· ···················· ···················· ···················· ···················· ····················
13439 MW079839.1 |Severe acute respiratory syndrome coro ···················· ···················· ···················· ···················· ···················· ····················
13440 MW079840.1 |Severe acute respiratory syndrome coro ···················· ···················· ···················· ···················· ···················· ····················
13441 MW079841.1 |Severe acute respiratory syndrome coro ···················· ···················· ···················· ···················· ···················· ····················
13442 MW079842.1 |Severe acute respiratory syndrome coro ···················· ···················· ···················· ···················· ···················· ····················
13443 MT998442.1 |Severe acute respiratory syndrome coro ···················· ·A·················· ···················· ···················· ···················· ····················
13444 MT998443.1 |Severe acute respiratory syndrome coro ···················· ···················· ···················· ···················· ···················· ····················
13445 MT998444.1 |Severe acute respiratory syndrome coro ···················· ···················· ···················· ···················· ···················· ····················
13446 MW075732.1 |Severe acute respiratory syndrome coro ···················· ···················· ···················· ···················· ···················· ····················
13447 MW075733.1 |Severe acute respiratory syndrome coro ···················· ···················· ···················· ···················· ···················· ····················
13448 MW075735.1 |Severe acute respiratory syndrome coro ···················· ···················· ···················· ···················· ···················· ····················
13449 MW075737.1 |Severe acute respiratory syndrome coro ···················· ···················· ···················· ···················· ···················· ····················
13450 MW075738.1 |Severe acute respiratory syndrome coro ···················· ···················· ···················· ···················· ···················· ····················
13451 MW075739.1 |Severe acute respiratory syndrome coro ···················· ···················· ···················· ···················· ···················· ····················
13452 MW075740.1 |Severe acute respiratory syndrome coro ···················· ···················· ···················· ···················· ···················· ····················
13453 MW075741.1 |Severe acute respiratory syndrome coro ···················· ···················· ···················· ···················· ···················· ····················
13454 MW075742.1 |Severe acute respiratory syndrome coro ···················· ···················· ···················· ···················· ···················· ····················
13455 MW075744.1 |Severe acute respiratory syndrome coro ···················· ···················· ···················· ···················· ···················· ····················
13456 MW075745.1 |Severe acute respiratory syndrome coro ···················· ·A·················· ···················· ···················· ···················· ····················
13457 MW075747.1 |Severe acute respiratory syndrome coro ···················· ·A·················· ···················· ···················· ···················· ····················
13458 MW075748.1 |Severe acute respiratory syndrome coro ···················· ···················· ···················· ···················· ···················· ····················
13459 MW075749.1 |Severe acute respiratory syndrome coro ···················· ···················· ···················· ···················· ···················· ····················
13460 MW075751.1 |Severe acute respiratory syndrome coro ···················· ···················· ···················· ···················· ···················· ····················
13461 MW075752.1 |Severe acute respiratory syndrome coro ···················· ···················· ···················· ···················· ···················· ····················
13462 MW075753.1 |Severe acute respiratory syndrome coro ···················· ···················· ···················· ···················· ···················· ····················

| | | | | | | | | |
|---|---|---|---|---|---|---|---|---|
| 13463 | MW075754.1 | Severe acute respiratory syndrome coro | ···················· | ···················· | ···················· | ···················· | ···················· | ···················· |
| 13464 | MW075755.1 | Severe acute respiratory syndrome coro | ···················· | ···················· | ···················· | ···················· | ···················· | ···················· |
| 13465 | MW075756.1 | Severe acute respiratory syndrome coro | ···················· | ···················· | ···················· | ···················· | ···················· | ···················· |
| 13466 | MW075757.1 | Severe acute respiratory syndrome coro | ···················· | ···················· | ···················· | ···················· | ···················· | ···················· |
| 13467 | MW075758.1 | Severe acute respiratory syndrome coro | ···················· | ···················· | ···················· | ···················· | ···················· | ···················· |
| 13468 | MW075760.1 | Severe acute respiratory syndrome coro | ···················· | ···················· | ···················· | ···················· | ···················· | ···················· |
| 13469 | MW075761.1 | Severe acute respiratory syndrome coro | ···················· | ···················· | ···················· | ···················· | ···················· | ···················· |
| 13470 | MW075762.1 | Severe acute respiratory syndrome coro | ···················· | ···················· | ···················· | ···················· | ···················· | ···················· |
| 13471 | MW075763.1 | Severe acute respiratory syndrome coro | ···················· | ···················· | ···················· | ···················· | ···················· | ···················· |
| 13472 | MW075764.1 | Severe acute respiratory syndrome coro | ···················· | ···················· | ···················· | ···················· | ···················· | ···················· |
| 13473 | MW075765.1 | Severe acute respiratory syndrome coro | ···················· | ·A·················· | ···················· | ···················· | ···················· | ···················· |
| 13474 | MW075766.1 | Severe acute respiratory syndrome coro | ···················· | ···················· | ···················· | ···················· | ···················· | ···················· |
| 13475 | MW075767.1 | Severe acute respiratory syndrome coro | ···················· | ···················· | ···················· | ···················· | ···················· | ···················· |
| 13476 | MW075768.1 | Severe acute respiratory syndrome coro | ···················· | ·A·················· | ···················· | ···················· | ···················· | ···················· |
| 13477 | MW075777.1 | Severe acute respiratory syndrome coro | ···················· | ···················· | ···················· | ···················· | ···················· | ···················· |
| 13478 | MW075778.1 | Severe acute respiratory syndrome coro | ···················· | ···················· | ···················· | ···················· | ···················· | ···················· |
| 13479 | MW075779.1 | Severe acute respiratory syndrome coro | ···················· | ···················· | ···················· | ···················· | ···················· | ···················· |
| 13480 | MW075781.1 | Severe acute respiratory syndrome coro | ···················· | ·A·················· | ···················· | ···················· | ···················· | ···················· |
| 13481 | MW075782.1 | Severe acute respiratory syndrome coro | ···················· | ···················· | ···················· | ···················· | ···················· | ···················· |
| 13482 | MW075783.1 | Severe acute respiratory syndrome coro | ···················· | ···················· | ···················· | ···················· | ···················· | ···················· |
| 13483 | MW075784.1 | Severe acute respiratory syndrome coro | ···················· | ···A················ | ···················· | ···················· | ···················· | ···················· |
| 13484 | MW075785.1 | Severe acute respiratory syndrome coro | ···················· | ···················· | ···················· | ···················· | ···················· | ···················· |
| 13485 | MW075786.1 | Severe acute respiratory syndrome coro | ···················· | ·A·················· | ···················· | ···················· | ···················· | ···················· |
| 13486 | MW075787.1 | Severe acute respiratory syndrome coro | ···················· | ···················· | ···················· | ···················· | ···················· | ···················· |
| 13487 | MW075788.1 | Severe acute respiratory syndrome coro | ···················· | ···················· | ···················· | ···················· | ···················· | ···················· |
| 13488 | MW075789.1 | Severe acute respiratory syndrome coro | ···················· | ···A················ | ···················· | ···················· | ···················· | ···················· |
| 13489 | MW075790.1 | Severe acute respiratory syndrome coro | ···················· | ···················· | ···················· | ···················· | ···················· | ···················· |
| 13490 | MW075791.1 | Severe acute respiratory syndrome coro | ···················· | ···················· | ···················· | ···················· | ···················· | ···················· |
| 13491 | MW075793.1 | Severe acute respiratory syndrome coro | ···················· | ·············A······ | ···················· | ···················· | ···················· | ···················· |
| 13492 | MW075794.1 | Severe acute respiratory syndrome coro | ···················· | ···················· | ···················· | ···················· | ···················· | ···················· |
| 13493 | MW075795.1 | Severe acute respiratory syndrome coro | ···················· | ···················· | ···················· | ···················· | ···················· | ···················· |
| 13494 | MW075796.1 | Severe acute respiratory syndrome coro | ···················· | ···················· | ···················· | ···················· | ···················· | ···················· |
| 13495 | MW075797.1 | Severe acute respiratory syndrome coro | ···················· | ···················· | ···················· | ···················· | ···················· | ···················· |
| 13496 | MW075798.1 | Severe acute respiratory syndrome coro | ···················· | ···················· | ···················· | ···················· | ···················· | ···················· |
| 13497 | MW075799.1 | Severe acute respiratory syndrome coro | ···················· | ···················· | ···················· | ···················· | ···················· | ···················· |
| 13498 | MW075801.1 | Severe acute respiratory syndrome coro | ···················· | ···················· | ···················· | ···················· | ···················· | ···················· |
| 13499 | MW075802.1 | Severe acute respiratory syndrome coro | ···················· | ···A················ | ···················· | ···················· | ···················· | ···················· |
| 13500 | MW075803.1 | Severe acute respiratory syndrome coro | ···················· | ···················· | ···················· | ···················· | ···················· | ···················· |
| 13501 | MW075804.1 | Severe acute respiratory syndrome coro | ···················· | ···················· | ···················· | ···················· | ···················· | ···················· |
| 13502 | MW075808.1 | Severe acute respiratory syndrome coro | ···················· | ···A················ | ···················· | ···················· | ···················· | ···················· |
| 13503 | MW064314.1 | Severe acute respiratory syndrome coro | ···················· | ···················· | ···················· | ···················· | ···················· | ···················· |
| 13504 | MW064315.1 | Severe acute respiratory syndrome coro | ···················· | ···················· | ···················· | ···················· | ···················· | ···················· |
| 13505 | MW064316.1 | Severe acute respiratory syndrome coro | ···················· | ···················· | ···················· | ···················· | ···················· | ···················· |
| 13506 | MW064317.1 | Severe acute respiratory syndrome coro | ···················· | ···················· | ···················· | ···················· | ···················· | ···················· |
| 13507 | MW064318.1 | Severe acute respiratory syndrome coro | ···················· | ···················· | ···················· | ···················· | ···················· | ···················· |
| 13508 | MW064319.1 | Severe acute respiratory syndrome coro | ···················· | ···················· | ···················· | ···················· | ···················· | ···················· |
| 13509 | MW064320.1 | Severe acute respiratory syndrome coro | ···················· | ···················· | ···················· | ···················· | ···················· | ···················· |
| 13510 | MW064321.1 | Severe acute respiratory syndrome coro | ···················· | ···················· | ···················· | ···················· | ···················· | ···················· |
| 13511 | MW064322.1 | Severe acute respiratory syndrome coro | ···················· | ···················· | ···················· | ···················· | ···················· | ···················· |
| 13512 | MW064323.1 | Severe acute respiratory syndrome coro | ···················· | ···················· | ···················· | ···················· | ···················· | ···················· |
| 13513 | MW064324.1 | Severe acute respiratory syndrome coro | ···················· | ···················· | ···················· | ···················· | ···················· | ···················· |
| 13514 | MW064325.1 | Severe acute respiratory syndrome coro | ···················· | ···················· | ···················· | ···················· | ···················· | ···················· |
| 13515 | MW064326.1 | Severe acute respiratory syndrome coro | ···················· | ···················· | ···················· | ···················· | ···················· | ···················· |
| 13516 | MW064327.1 | Severe acute respiratory syndrome coro | ···················· | ···················· | ···················· | ···················· | ···················· | ···················· |
| 13517 | MW064328.1 | Severe acute respiratory syndrome coro | ···················· | ···················· | ···················· | ···················· | ···················· | ···················· |
| 13518 | MW064329.1 | Severe acute respiratory syndrome coro | ···················· | ···················· | ···················· | ···················· | ···················· | ···················· |
| 13519 | MW064331.1 | Severe acute respiratory syndrome coro | ···················· | ···················· | ···················· | ···················· | ···················· | ···················· |
| 13520 | MW064333.1 | Severe acute respiratory syndrome coro | ···················· | ···················· | ···················· | ···················· | ···················· | ···················· |
| 13521 | MW064334.1 | Severe acute respiratory syndrome coro | ···················· | ·············A······ | ···················· | ···················· | ···················· | ···················· |
| 13522 | MW064335.1 | Severe acute respiratory syndrome coro | ···················· | ···················· | ···················· | ···················· | ···················· | ···················· |
| 13523 | MW064336.1 | Severe acute respiratory syndrome coro | ···················· | ···················· | ···················· | ···················· | ···················· | ···················· |
| 13524 | MW064337.1 | Severe acute respiratory syndrome coro | ···················· | ···················· | ···················· | ···················· | ···················· | ···················· |
| 13525 | MW064338.1 | Severe acute respiratory syndrome coro | ···················· | ···················· | ···················· | ···················· | ···················· | ···················· |
| 13526 | MW064339.1 | Severe acute respiratory syndrome coro | ···················· | ···················· | ···················· | ···················· | ···················· | ···················· |
| 13527 | MW064340.1 | Severe acute respiratory syndrome coro | ···················· | ···················· | ···················· | ···················· | ···················· | ···················· |
| 13528 | MW064341.1 | Severe acute respiratory syndrome coro | ···················· | ···················· | ···················· | ···················· | ···················· | ···················· |
| 13529 | MW064342.1 | Severe acute respiratory syndrome coro | ···················· | ···················· | ···················· | ···················· | ···················· | ···················· |
| 13530 | MW064343.1 | Severe acute respiratory syndrome coro | ···················· | ···················· | ···················· | ···················· | ···················· | ···················· |
| 13531 | MW064344.1 | Severe acute respiratory syndrome coro | ···················· | ···················· | ···················· | ···················· | ···················· | ···················· |
| 13532 | MW064345.1 | Severe acute respiratory syndrome coro | ···················· | ···················· | ···················· | ···················· | ···················· | ···················· |
| 13533 | MW064346.1 | Severe acute respiratory syndrome coro | ···················· | ···················· | ···················· | ···················· | ···················· | ···················· |
| 13534 | MW064347.1 | Severe acute respiratory syndrome coro | ···················· | ···················· | ···················· | ···················· | ···················· | ···················· |
| 13535 | MW064348.1 | Severe acute respiratory syndrome coro | ···················· | ···················· | ···················· | ···················· | ···················· | ···················· |
| 13536 | MW064349.1 | Severe acute respiratory syndrome coro | ···················· | ···················· | ···················· | ···················· | ···················· | ···················· |
| 13537 | MW064350.1 | Severe acute respiratory syndrome coro | ···················· | ···················· | ···················· | ···················· | ···················· | ···················· |
| 13538 | MW064351.1 | Severe acute respiratory syndrome coro | ···················· | ···················· | ···················· | ···················· | ···················· | ···················· |
| 13539 | MW064352.1 | Severe acute respiratory syndrome coro | ···················· | ···················· | ···················· | ···················· | ···················· | ···················· |
| 13540 | MW064353.1 | Severe acute respiratory syndrome coro | ···················· | ···················· | ···················· | ···················· | ···················· | ···················· |

| | | | | | | | | |
|---|---|---|---|---|---|---|---|---|
| 13541 | MW064354.1 | \|Severe acute respiratory syndrome coro | .................... | .................... | ..................... | .................... | .................... | .................... |
| 13542 | MW064355.1 | \|Severe acute respiratory syndrome coro | .................... | .................... | ..................... | .................... | .................... | .................... |
| 13543 | MW064356.1 | \|Severe acute respiratory syndrome coro | .................... | .................... | ..................... | .................... | .................... | .................... |
| 13544 | MW064357.1 | \|Severe acute respiratory syndrome coro | .................... | .................... | ..................... | .................... | .................... | .................... |
| 13545 | MW064358.1 | \|Severe acute respiratory syndrome coro | .................... | .................... | ..................... | .................... | .................... | .................... |
| 13546 | MW064359.1 | \|Severe acute respiratory syndrome coro | .................... | .................... | ..................... | .................... | .................... | .................... |
| 13547 | MW064360.1 | \|Severe acute respiratory syndrome coro | .................... | .................... | ..................... | .................... | .................... | .................... |
| 13548 | MW064361.1 | \|Severe acute respiratory syndrome coro | .................... | .................... | ..................... | .................... | .................... | .................... |
| 13549 | MW064362.1 | \|Severe acute respiratory syndrome coro | .................... | .................... | ..................... | .................... | .................... | .................... |
| 13550 | MW064363.1 | \|Severe acute respiratory syndrome coro | .................... | .................... | ..................... | .................... | .................... | .................... |
| 13551 | MW064364.1 | \|Severe acute respiratory syndrome coro | .................... | .................... | ..................... | .................... | .................... | .................... |
| 13552 | MW064365.1 | \|Severe acute respiratory syndrome coro | .................... | .................... | ..................... | .................... | .................... | .................... |
| 13553 | MW064366.1 | \|Severe acute respiratory syndrome coro | .................... | .................... | ..................... | .................... | .................... | .................... |
| 13554 | MW064367.1 | \|Severe acute respiratory syndrome coro | .................... | .................... | ..................... | .................... | .................... | .................... |
| 13555 | MW064368.1 | \|Severe acute respiratory syndrome coro | .................... | .................... | ..................... | .................... | .................... | .................... |
| 13556 | MW064369.1 | \|Severe acute respiratory syndrome coro | .................... | .................... | ..................... | .................... | .................... | .................... |
| 13557 | MW064370.1 | \|Severe acute respiratory syndrome coro | .................... | .................... | ..................... | .................... | .................... | .................... |
| 13558 | MW064371.1 | \|Severe acute respiratory syndrome coro | .................... | .................... | ..................... | .................... | .................... | .................... |
| 13559 | MW064372.1 | \|Severe acute respiratory syndrome coro | .................... | .................... | ..................... | .................... | .................... | .................... |
| 13560 | MW064373.1 | \|Severe acute respiratory syndrome coro | .................... | .................... | ..................... | .................... | .................... | .................... |
| 13561 | MW064374.1 | \|Severe acute respiratory syndrome coro | .................... | .................... | ..................... | .................... | .................... | .................... |
| 13562 | MW064375.1 | \|Severe acute respiratory syndrome coro | .................... | .................... | ..................... | .................... | .................... | .................... |
| 13563 | MW064376.1 | \|Severe acute respiratory syndrome coro | .................... | .................... | ..................... | .................... | .................... | .................... |
| 13564 | MW064377.1 | \|Severe acute respiratory syndrome coro | .................... | .................... | ..................... | .................... | .................... | .................... |
| 13565 | MW064378.1 | \|Severe acute respiratory syndrome coro | .................... | .................... | ..................... | .................... | .................... | .................... |
| 13566 | MW064379.1 | \|Severe acute respiratory syndrome coro | .................... | .................... | ..................... | .................... | .................... | .................... |
| 13567 | MW064380.1 | \|Severe acute respiratory syndrome coro | .................... | .................... | ..................... | .................... | .................... | .................... |
| 13568 | MW064381.1 | \|Severe acute respiratory syndrome coro | .................... | .................... | ..................... | .................... | .................... | .................... |
| 13569 | MW064382.1 | \|Severe acute respiratory syndrome coro | .................... | .................... | ..................... | .................... | .................... | .................... |
| 13570 | MW064383.1 | \|Severe acute respiratory syndrome coro | .................... | .................... | ..................... | .................... | .................... | .................... |
| 13571 | MW064385.1 | \|Severe acute respiratory syndrome coro | .................... | .................... | ..................... | .................... | .................... | .................... |
| 13572 | MW064386.1 | \|Severe acute respiratory syndrome coro | .................... | .................... | ..................... | .................... | .................... | .................... |
| 13573 | MW064387.1 | \|Severe acute respiratory syndrome coro | .................... | .................... | ..................... | .................... | .................... | .................... |
| 13574 | MW064388.1 | \|Severe acute respiratory syndrome coro | .................... | .................... | ..................... | .................... | .................... | .................... |
| 13575 | MW064389.1 | \|Severe acute respiratory syndrome coro | .................... | .................... | ..................... | .................... | .................... | .................... |
| 13576 | MW064390.1 | \|Severe acute respiratory syndrome coro | .................... | .................... | ..................... | .................... | .................... | .................... |
| 13577 | MW064391.1 | \|Severe acute respiratory syndrome coro | .................... | .................... | ..................... | .................... | .................... | .................... |
| 13578 | MW064392.1 | \|Severe acute respiratory syndrome coro | .................... | .................... | ..................... | .................... | .................... | .................... |
| 13579 | MW064393.1 | \|Severe acute respiratory syndrome coro | .................... | .................... | ..................... | .................... | .................... | .................... |
| 13580 | MW064394.1 | \|Severe acute respiratory syndrome coro | .................... | .................... | ..................... | .................... | .................... | .................... |
| 13581 | MW064395.1 | \|Severe acute respiratory syndrome coro | .................... | .................... | ..................... | .................... | .................... | .................... |
| 13582 | MW064396.1 | \|Severe acute respiratory syndrome coro | .................... | .................... | ..................... | .................... | .................... | .................... |
| 13583 | MW064397.1 | \|Severe acute respiratory syndrome coro | .................... | .................... | ..................... | .................... | .................... | .................... |
| 13584 | MW064398.1 | \|Severe acute respiratory syndrome coro | .................... | .................... | ..................... | .................... | .................... | .................... |
| 13585 | MW064399.1 | \|Severe acute respiratory syndrome coro | .................... | .................... | ..................... | .................... | .................... | .................... |
| 13586 | MW064400.1 | \|Severe acute respiratory syndrome coro | .................... | .................... | ..................... | .................... | .................... | .................... |
| 13587 | MW064401.1 | \|Severe acute respiratory syndrome coro | .................... | .................... | ..................... | .................... | .................... | .................... |
| 13588 | MW064402.1 | \|Severe acute respiratory syndrome coro | .................... | .................... | ..................... | .................... | .................... | .................... |
| 13589 | MW064403.1 | \|Severe acute respiratory syndrome coro | .................... | .................... | ..................... | .................... | .................... | .................... |
| 13590 | MW064404.1 | \|Severe acute respiratory syndrome coro | ...T................ | .................... | ..................... | .................... | .................... | .................... |
| 13591 | MW064405.1 | \|Severe acute respiratory syndrome coro | .................... | .................... | ..................... | .................... | .................... | .................... |
| 13592 | MW064406.1 | \|Severe acute respiratory syndrome coro | .................... | .................... | ..................... | .................... | .................... | .................... |
| 13593 | MW064407.1 | \|Severe acute respiratory syndrome coro | .................... | .................... | ..................... | .................... | .................... | .................... |
| 13594 | MW064408.1 | \|Severe acute respiratory syndrome coro | .................... | .................... | ..................... | .................... | .................... | .................... |
| 13595 | MW064409.1 | \|Severe acute respiratory syndrome coro | .................... | .................... | ..................... | .................... | .................... | .................... |
| 13596 | MW064410.1 | \|Severe acute respiratory syndrome coro | .................... | .................... | ..................... | .................... | .................... | .................... |
| 13597 | MW064411.1 | \|Severe acute respiratory syndrome coro | .................... | .................... | ..................... | .................... | .................... | .................... |
| 13598 | MW064412.1 | \|Severe acute respiratory syndrome coro | .................... | .................... | ..................... | .................... | .................... | .................... |
| 13599 | MW064413.1 | \|Severe acute respiratory syndrome coro | .................... | .................... | ..................... | .................... | .................... | .................... |
| 13600 | MW064414.1 | \|Severe acute respiratory syndrome coro | .................... | .................... | ..................... | .................... | .................... | .................... |
| 13601 | MW064415.1 | \|Severe acute respiratory syndrome coro | .................... | .................... | ..................... | .................... | .................... | .................... |
| 13602 | MW064416.1 | \|Severe acute respiratory syndrome coro | .................... | .................... | ..................... | .................... | .................... | .................... |
| 13603 | MW064417.1 | \|Severe acute respiratory syndrome coro | .................... | .................... | ..................... | .................... | .................... | .................... |
| 13604 | MW064418.1 | \|Severe acute respiratory syndrome coro | .................... | .................... | ..................... | .................... | .................... | .................... |
| 13605 | MW064419.1 | \|Severe acute respiratory syndrome coro | .................... | .................... | ..................... | .................... | .................... | .................... |
| 13606 | MW064420.1 | \|Severe acute respiratory syndrome coro | .................... | .................... | ..................... | .................... | .................... | .................... |
| 13607 | MW064421.1 | \|Severe acute respiratory syndrome coro | .................... | .................... | ..................... | .................... | .................... | .................... |
| 13608 | MW064422.1 | \|Severe acute respiratory syndrome coro | .................... | .................... | ..................... | .................... | .................... | .................... |
| 13609 | MW064423.1 | \|Severe acute respiratory syndrome coro | .................... | .................... | ..................... | .................... | .................... | .................... |
| 13610 | MW064424.1 | \|Severe acute respiratory syndrome coro | .................... | .................... | ..................... | .................... | .................... | .................... |
| 13611 | MW064425.1 | \|Severe acute respiratory syndrome coro | .................... | .................... | ..................... | .................... | .................... | .................... |
| 13612 | MW064426.1 | \|Severe acute respiratory syndrome coro | .................... | .................... | ..................... | .................... | .................... | .................... |
| 13613 | MW064427.1 | \|Severe acute respiratory syndrome coro | .................... | .................... | ..................... | .................... | .................... | .................... |
| 13614 | MW064428.1 | \|Severe acute respiratory syndrome coro | .................... | .................... | ..................... | .................... | .................... | .................... |
| 13615 | MW064429.1 | \|Severe acute respiratory syndrome coro | .................... | .................... | ..................... | .................... | .................... | .................... |
| 13616 | MW064430.1 | \|Severe acute respiratory syndrome coro | .................... | .................... | ..................... | .................... | .................... | .................... |
| 13617 | MW064432.1 | \|Severe acute respiratory syndrome coro | .................... | .................... | ..................... | .................... | .................... | .................... |
| 13618 | MW064433.1 | \|Severe acute respiratory syndrome coro | .................... | .................... | ..................... | .................... | .................... | .................... |

```
13619 MW064434.1 |Severe acute respiratory syndrome coro  ....................  ....................  ....................  ....................  ....................  ....................
13620 MW064435.1 |Severe acute respiratory syndrome coro  ....................  ....................  ....................  ....................  ....................  ....................
13621 MW064436.1 |Severe acute respiratory syndrome coro  ....................  ....................  ....................  ....................  ....................  ....................
13622 MW064437.1 |Severe acute respiratory syndrome coro  ....................  ....................  ....................  ....................  ....................  ....................
13623 MW064438.1 |Severe acute respiratory syndrome coro  ....................  ....................  ....................  ....................  ....................  ....................
13624 MW064439.1 |Severe acute respiratory syndrome coro  ....................  ....................  ....................  ....................  ....................  ....................
13625 MW064440.1 |Severe acute respiratory syndrome coro  ....................  ....................  ....................  ....................  ....................  ....................
13626 MW064441.1 |Severe acute respiratory syndrome coro  ....................  ....................  ....................  ....................  ....................  ....................
13627 MW064442.1 |Severe acute respiratory syndrome coro  ....................  ....................  ....................  ....................  ....................  ....................
13628 MW064443.1 |Severe acute respiratory syndrome coro  ....................  ....................  ....................  ....................  ....................  ....................
13629 MW064444.1 |Severe acute respiratory syndrome coro  ....................  ....................  ....................  ....................  ....................  ....................
13630 MW064445.1 |Severe acute respiratory syndrome coro  ....................  ....................  ....................  ....................  ....................  ....................
13631 MW064446.1 |Severe acute respiratory syndrome coro  ....................  ....................  ....................  ....................  ....................  ....................
13632 MW064447.1 |Severe acute respiratory syndrome coro  ....................  ....................  ....................  ....................  ....................  ....................
13633 MW064448.1 |Severe acute respiratory syndrome coro  ....................  ....................  ....................  ....................  ....................  ....................
13634 MW064449.1 |Severe acute respiratory syndrome coro  ....................  ....................  ....................  ....................  ....................  ....................
13635 MW064450.1 |Severe acute respiratory syndrome coro  ....................  ....................  ....................  ....................  ....................  ....................
13636 MW064451.1 |Severe acute respiratory syndrome coro  ....................  ....................  ....................  ....................  ....................  ....................
13637 MW064452.1 |Severe acute respiratory syndrome coro  ....................  ....................  ....................  ....................  ....................  ....................
13638 MW064453.1 |Severe acute respiratory syndrome coro  ....................  ....................  ....................  ....................  ....................  ....................
13639 MW064454.1 |Severe acute respiratory syndrome coro  ....................  ....................  ....................  ....................  ....................  ....................
13640 MW064455.1 |Severe acute respiratory syndrome coro  ....................  ....................  ....................  ....................  ....................  ....................
13641 MW064456.1 |Severe acute respiratory syndrome coro  ....................  ....................  ....................  ....................  ....................  ....................
13642 MW064457.1 |Severe acute respiratory syndrome coro  ....................  ....................  ....................  ....................  ....................  ....................
13643 MW064458.1 |Severe acute respiratory syndrome coro  ....................  ....................  ....................  ....................  ....................  ....................
13644 MW064459.1 |Severe acute respiratory syndrome coro  ....................  ....................  ....................  ....................  ....................  ....................
13645 MW064460.1 |Severe acute respiratory syndrome coro  ....................  ....................  ....................  ....................  ....................  ....................
13646 MW064461.1 |Severe acute respiratory syndrome coro  ....................  ....................  ....................  ....................  ....................  ....................
13647 MW064462.1 |Severe acute respiratory syndrome coro  ....................  ....................  ....................  ....................  ....................  ....................
13648 MW064463.1 |Severe acute respiratory syndrome coro  ....................  ....................  ....................  ....................  ....................  ....................
13649 MW064464.1 |Severe acute respiratory syndrome coro  ....................  ....................  ....................  ....................  ....................  ....................
13650 MW064465.1 |Severe acute respiratory syndrome coro  ....................  ....................  ....................  ....................  ....................  ....................
13651 MW064466.1 |Severe acute respiratory syndrome coro  ....................  ....................  ....................  ....................  ....................  ....................
13652 MW064467.1 |Severe acute respiratory syndrome coro  ....................  ....................  ....................  ....................  ....................  ....................
13653 MW064468.1 |Severe acute respiratory syndrome coro  ....................  ....................  ....................  ....................  ....................  ....................
13654 MW064469.1 |Severe acute respiratory syndrome coro  ....................  ....................  ....................  ....................  ....................  ....................
13655 MW064470.1 |Severe acute respiratory syndrome coro  ....................  ....................  ....................  ....................  ....................  ....................
13656 MW064472.1 |Severe acute respiratory syndrome coro  ....................  ....................  ....................  ....................  ....................  ....................
13657 MW064473.1 |Severe acute respiratory syndrome coro  ....................  ....................  ....................  ....................  ....................  ....................
13658 MW064474.1 |Severe acute respiratory syndrome coro  ....................  ....................  ....................  ....................  ....................  ....................
13659 MW064475.1 |Severe acute respiratory syndrome coro  ....................  ....................  ....................  ....................  ....................  ....................
13660 MW064476.1 |Severe acute respiratory syndrome coro  ....................  ....................  ....................  ....................  ....................  ....................
13661 MW064477.1 |Severe acute respiratory syndrome coro  ....................  ....................  ....................  ....................  ....................  ....................
13662 MW064478.1 |Severe acute respiratory syndrome coro  ....................  ....................  ....................  ....................  ....................  ....................
13663 MW064479.1 |Severe acute respiratory syndrome coro  ....................  ....................  ....................  ....................  ....................  ....................
13664 MW064480.1 |Severe acute respiratory syndrome coro  ....................  ....................  ....................  ....................  ....................  ....................
13665 MW064481.1 |Severe acute respiratory syndrome coro  ....................  ....................  ....................  ....................  ....................  ....................
13666 MW064482.1 |Severe acute respiratory syndrome coro  ....................  ....................  ....................  ....................  ....................  ....................
13667 MW064483.1 |Severe acute respiratory syndrome coro  ....................  ....................  ....................  ....................  ....................  ....................
13668 MW064484.1 |Severe acute respiratory syndrome coro  ....................  ....................  ....................  ....................  ....................  ....................
13669 MW064485.1 |Severe acute respiratory syndrome coro  ....................  ....................  ....................  ....................  ....................  ....................
13670 MW064486.1 |Severe acute respiratory syndrome coro  ....................  ....................  ....................  ....................  ....................  ....................
13671 MW064487.1 |Severe acute respiratory syndrome coro  ....................  ....................  ....................  ....................  ....................  ....................
13672 MW064488.1 |Severe acute respiratory syndrome coro  ....................  ....................  ....................  ....................  ....................  ....................
13673 MW064489.1 |Severe acute respiratory syndrome coro  ....................  ....................  ....................  ....................  ....................  ....................
13674 MW064490.1 |Severe acute respiratory syndrome coro  ....................  ....................  ....................  ....................  ....................  ....................
13675 MW064491.1 |Severe acute respiratory syndrome coro  ....................  ....................  ....................  ....................  ....................  ....................
13676 MW064492.1 |Severe acute respiratory syndrome coro  ....................  ....................  ....................  ....................  ....................  ....................
13677 MW064493.1 |Severe acute respiratory syndrome coro  ....................  ....................  ....................  ....................  ....................  ....................
13678 MW064494.1 |Severe acute respiratory syndrome coro  ....................  ....................  ....................  ....................  ....................  ....................
13679 MW064496.1 |Severe acute respiratory syndrome coro  ....................  ....................  ....................  ....................  ....................  ....................
13680 MW064497.1 |Severe acute respiratory syndrome coro  ....................  ....................  ....................  ....................  ....................  ....................
13681 MW064498.1 |Severe acute respiratory syndrome coro  ....................  ....................  ....................  ....................  ....................  ....................
13682 MW064499.1 |Severe acute respiratory syndrome coro  ....................  ....................  ....................  ....................  ....................  ....................
13683 MW064500.1 |Severe acute respiratory syndrome coro  ....................  ....................  ....................  ....................  ....................  ....................
13684 MW064501.1 |Severe acute respiratory syndrome coro  ....................  ....................  ....................  ....................  ....................  ....................
13685 MW064502.1 |Severe acute respiratory syndrome coro  ....................  ....................  ....................  ....................  ....................  ....................
13686 MW064503.1 |Severe acute respiratory syndrome coro  ....................  ....................  ....................  ....................  ....................  ....................
13687 MW064504.1 |Severe acute respiratory syndrome coro  ....................  ....................  ....................  ....................  ....................  ....................
13688 MW064505.1 |Severe acute respiratory syndrome coro  ....................  ....................  ....................  ....................  ....................  ....................
13689 MW064506.1 |Severe acute respiratory syndrome coro  ....................  ....................  ....................  ....................  ....................  ....................
13690 MW064507.1 |Severe acute respiratory syndrome coro  ....................  ....................  ....................  ....................  ....................  ....................
13691 MW064508.1 |Severe acute respiratory syndrome coro  ....................  ....................  ....................  ....................  ....................  ....................
13692 MW064509.1 |Severe acute respiratory syndrome coro  ....................  ....................  ....................  ....................  ....................  ....................
13693 MW064510.1 |Severe acute respiratory syndrome coro  ....................  ....................  ....................  ....................  ....................  ....................
13694 MW064511.1 |Severe acute respiratory syndrome coro  ....................  ....................  ....................  ....................  ....................  ....................
13695 MW064512.1 |Severe acute respiratory syndrome coro  ....................  ....................  ....................  ....................  ....................  ....................
13696 MW064513.1 |Severe acute respiratory syndrome coro  ....................  ....................  ....................  ....................  ....................  ....................
```

```
13697 MW064514.1 |Severe acute respiratory syndrome coro    ....................  ....................  .....................  ....................  ....................  ....................
13698 MW064515.1 |Severe acute respiratory syndrome coro    ....................  ....................  .....................  ....................  ....................  ....................
13699 MW064516.1 |Severe acute respiratory syndrome coro    ....................  ....................  .....................  ....................  ....................  ....................
13700 MW064517.1 |Severe acute respiratory syndrome coro    ....................  ....................  .....................  ....................  ....................  ....................
13701 MW064518.1 |Severe acute respiratory syndrome coro    ....................  ....................  .....................  ....................  ....................  ....................
13702 MW064519.1 |Severe acute respiratory syndrome coro    ....................  ....................  .....................  ....................  ....................  ....................
13703 MW064520.1 |Severe acute respiratory syndrome coro    ....................  ....................  .....................  ....................  ....................  ....................
13704 MW064521.1 |Severe acute respiratory syndrome coro    ....................  ....................  .....................  ....................  ....................  ....................
13705 MW064522.1 |Severe acute respiratory syndrome coro    ....................  ....................  .....................  ....................  ....................  ....................
13706 MW064523.1 |Severe acute respiratory syndrome coro    ....................  ....................  .....................  ....................  ....................  ....................
13707 MW064524.1 |Severe acute respiratory syndrome coro    ....................  ....................  .....................  ....................  ....................  ....................
13708 MW064525.1 |Severe acute respiratory syndrome coro    ....................  ....................  .....................  ....................  ....................  ....................
13709 MW064526.1 |Severe acute respiratory syndrome coro    ....................  ....................  .....................  ....................  ....................  ....................
13710 MW064527.1 |Severe acute respiratory syndrome coro    ....................  ....................  .....................  ....................  ....................  ....................
13711 MW064528.1 |Severe acute respiratory syndrome coro    ....................  ....................  .....................  ....................  ....................  ....................
13712 MW064529.1 |Severe acute respiratory syndrome coro    ....................  ....................  .....................  ....................  ....................  ....................
13713 MW064530.1 |Severe acute respiratory syndrome coro    ....................  ....................  .....................  ....................  ....................  ....................
13714 MW064531.1 |Severe acute respiratory syndrome coro    ....................  ....................  .....................  ....................  ....................  ....................
13715 MW064532.1 |Severe acute respiratory syndrome coro    ....................  ....................  .....................  ....................  ....................  ....................
13716 MW064533.1 |Severe acute respiratory syndrome coro    ....................  ....................  .....................  ....................  ....................  ....................
13717 MW064534.1 |Severe acute respiratory syndrome coro    ....................  ....................  .....................  ....................  ....................  ....................
13718 MW064535.1 |Severe acute respiratory syndrome coro    ....................  ....................  .....................  ....................  ....................  ....................
13719 MW064537.1 |Severe acute respiratory syndrome coro    ....................  ....................  .....................  ....................  ....................  ....................
13720 MW064538.1 |Severe acute respiratory syndrome coro    ....................  ....................  .....................  ....................  ....................  ....................
13721 MW064539.1 |Severe acute respiratory syndrome coro    ....................  ....................  .....................  ....................  ....................  ....................
13722 MW064541.1 |Severe acute respiratory syndrome coro    ....................  ....................  .....................  ....................  ....................  ....................
13723 MW064542.1 |Severe acute respiratory syndrome coro    ....................  ....................  .....................  ....................  ....................  ....................
13724 MW064543.1 |Severe acute respiratory syndrome coro    ....................  ....................  .....................  ....................  ....................  ....................
13725 MW064544.1 |Severe acute respiratory syndrome coro    ....................  ....................  .....................  ....................  ....................  ....................
13726 MW064545.1 |Severe acute respiratory syndrome coro    ....................  ....................  .....................  ....................  ....................  ....................
13727 MW064546.1 |Severe acute respiratory syndrome coro    ....................  ....................  .....................  ....................  ....................  ....................
13728 MW064547.1 |Severe acute respiratory syndrome coro    ....................  ....................  .....................  ....................  ....................  ....................
13729 MW064548.1 |Severe acute respiratory syndrome coro    ....................  ....................  .....................  ....................  ....................  ....................
13730 MW064549.1 |Severe acute respiratory syndrome coro    ....................  ....................  .....................  ....................  ....................  ....................
13731 MW064551.1 |Severe acute respiratory syndrome coro    ....................  ....................  .....................  ....................  ....................  ....................
13732 MW064553.1 |Severe acute respiratory syndrome coro    ....................  ....................  .....................  ....................  ....................  ....................
13733 MW064554.1 |Severe acute respiratory syndrome coro    ....................  ....................  .....................  ....................  ....................  ....................
13734 MW064555.1 |Severe acute respiratory syndrome coro    ....................  ....................  .....................  ....................  ....................  ....................
13735 MW064556.1 |Severe acute respiratory syndrome coro    ....................  ....................  .....................  ....................  ....................  ....................
13736 MW064557.1 |Severe acute respiratory syndrome coro    ....................  ....................  .....................  ....................  ....................  ....................
13737 MW064558.1 |Severe acute respiratory syndrome coro    ....................  ....................  .....................  ....................  ....................  ....................
13738 MW064560.1 |Severe acute respiratory syndrome coro    ....................  ....................  .....................  ....................  ....................  ....................
13739 MW064561.1 |Severe acute respiratory syndrome coro    ....................  ....................  .....................  ....................  ....................  ....................
13740 MW064562.1 |Severe acute respiratory syndrome coro    ....................  ....................  .....................  ....................  ....................  ....................
13741 MW064563.1 |Severe acute respiratory syndrome coro    ....................  ....................  .....................  ....................  ....................  ....................
13742 MW064564.1 |Severe acute respiratory syndrome coro    ....................  ....................  .....................  ....................  ....................  ....................
13743 MW064565.1 |Severe acute respiratory syndrome coro    ....................  ....................  .....................  ....................  ....................  ....................
13744 MW064566.1 |Severe acute respiratory syndrome coro    ....................  ....................  .....................  ....................  ....................  ....................
13745 MW064567.1 |Severe acute respiratory syndrome coro    ....................  ....................  .....................  ....................  ....................  ....................
13746 MW064568.1 |Severe acute respiratory syndrome coro    ....................  ....................  .....................  ....................  ....................  ....................
13747 MW064569.1 |Severe acute respiratory syndrome coro    ....................  ....................  .....................  ....................  ....................  ....................
13748 MW064570.1 |Severe acute respiratory syndrome coro    ....................  ....................  .....................  ....................  ....................  ....................
13749 MW064571.1 |Severe acute respiratory syndrome coro    ....................  ....................  .....................  ....................  ....................  ....................
13750 MW064572.1 |Severe acute respiratory syndrome coro    ....................  ....................  .....................  ....................  ....................  ....................
13751 MW064573.1 |Severe acute respiratory syndrome coro    ....................  ....................  .....................  ....................  ....................  ....................
13752 MW064574.1 |Severe acute respiratory syndrome coro    ....................  ....................  .....................  ....................  ....................  ....................
13753 MW064575.1 |Severe acute respiratory syndrome coro    ....................  ....................  .....................  ....................  ....................  ....................
13754 MW064576.1 |Severe acute respiratory syndrome coro    ....................  ....................  .....................  ....................  ....................  ....................
13755 MW064577.1 |Severe acute respiratory syndrome coro    ....................  ....................  .....................  ....................  ....................  ....................
13756 MW064578.1 |Severe acute respiratory syndrome coro    ....................  ....................  .....................  ....................  ....................  ....................
13757 MW064579.1 |Severe acute respiratory syndrome coro    ....................  ....................  .....................  ....................  ....................  ....................
13758 MW064581.1 |Severe acute respiratory syndrome coro    ....................  ....................  .....................  ....................  ....................  ....................
13759 MW064582.1 |Severe acute respiratory syndrome coro    ....................  ....................  .....................  ....................  ....................  ....................
13760 MW064583.1 |Severe acute respiratory syndrome coro    ....................  ....................  .....................  ....................  ....................  ....................
13761 MW064584.1 |Severe acute respiratory syndrome coro    ....................  ....................  .....................  ....................  ....................  ....................
13762 MW064585.1 |Severe acute respiratory syndrome coro    ....................  ....................  .....................  ....................  ....................  ....................
13763 MW064586.1 |Severe acute respiratory syndrome coro    ....................  ....................  .....................  ....................  ....................  ....................
13764 MW064587.1 |Severe acute respiratory syndrome coro    ....................  ....................  .....................  ....................  ....................  ....................
13765 MW064588.1 |Severe acute respiratory syndrome coro    ....................  ....................  .....................  ....................  ....................  ....................
13766 MW064589.1 |Severe acute respiratory syndrome coro    ....................  ....................  .....................  ....................  ....................  ....................
13767 MW064590.1 |Severe acute respiratory syndrome coro    ....................  ....................  .....................  ....................  ....................  ....................
13768 MW064591.1 |Severe acute respiratory syndrome coro    ....................  ....................  .....................  ....................  ....................  ....................
13769 MW064592.1 |Severe acute respiratory syndrome coro    ....................  ....................  .....................  ....................  ....................  ....................
13770 MW064593.1 |Severe acute respiratory syndrome coro    ....................  ....................  .....................  ....................  ....................  ....................
13771 MW064594.1 |Severe acute respiratory syndrome coro    ....................  ....................  .....................  ....................  ....................  ....................
13772 MW064597.1 |Severe acute respiratory syndrome coro    ....................  ....................  .....................  ....................  ....................  ....................
13773 MW064598.1 |Severe acute respiratory syndrome coro    ....................  ....................  .....................  ....................  ....................  ....................
13774 MW064599.1 |Severe acute respiratory syndrome coro    ....................  ....................  .....................  ....................  ....................  ....................
```

13775 MW064600.1 |Severe acute respiratory syndrome coro .................... .................... ..................... .................... .................... ....................
13776 MW064601.1 |Severe acute respiratory syndrome coro .................... .................... ..................... .................... .................... ....................
13777 MW064602.1 |Severe acute respiratory syndrome coro .................... .................... ..................... .................... .................... ....................
13778 MW064603.1 |Severe acute respiratory syndrome coro .................... .................... ..................... .................... .................... ....................
13779 MW064604.1 |Severe acute respiratory syndrome coro .................... .................... ..................... .................... .................... ....................
13780 MW064605.1 |Severe acute respiratory syndrome coro .................... .................... ..................... .................... .................... ....................
13781 MW064606.1 |Severe acute respiratory syndrome coro .................... .................... ..................... .................... .................... ....................
13782 MW064607.1 |Severe acute respiratory syndrome coro .................... .................... ..................... .................... .................... ....................
13783 MW064608.1 |Severe acute respiratory syndrome coro .................... .................... ..................... .................... .................... ....................
13784 MW064609.1 |Severe acute respiratory syndrome coro .................... .................... ..................... .................... .................... ....................
13785 MW064610.1 |Severe acute respiratory syndrome coro .................... .................... ..................... .................... .................... ....................
13786 MW064611.1 |Severe acute respiratory syndrome coro .................... .................... ..................... .................... .................... ....................
13787 MW064612.1 |Severe acute respiratory syndrome coro .................... .................... ..................... .................... .................... ....................
13788 MW064613.1 |Severe acute respiratory syndrome coro .................... .................... ..................... .................... .................... ....................
13789 MW064614.1 |Severe acute respiratory syndrome coro .................... .................... ..................... .................... .................... ....................
13790 MW064615.1 |Severe acute respiratory syndrome coro .................... .................... ..................... .................... .................... ....................
13791 MW064616.1 |Severe acute respiratory syndrome coro .................... .................... ..................... .................... .................... ....................
13792 MW064617.1 |Severe acute respiratory syndrome coro .................... .................... ..................... .................... .................... ....................
13793 MW064618.1 |Severe acute respiratory syndrome coro .................... .................... ..................... .................... .................... ....................
13794 MW064619.1 |Severe acute respiratory syndrome coro .................... .................... ..................... .................... .................... ....................
13795 MW064620.1 |Severe acute respiratory syndrome coro .................... .................... ..................... .................... .................... ....................
13796 MW064621.1 |Severe acute respiratory syndrome coro .................... .................... ..................... .................... .................... ....................
13797 MW064622.1 |Severe acute respiratory syndrome coro .................... .................... ..................... .................... .................... ....................
13798 MW064623.1 |Severe acute respiratory syndrome coro .................... .................... ..................... .................... .................... ....................
13799 MW064624.1 |Severe acute respiratory syndrome coro .................... .................... ..................... .................... .................... ....................
13800 MW064625.1 |Severe acute respiratory syndrome coro .................... .................... ..................... .................... .................... ....................
13801 MW064626.1 |Severe acute respiratory syndrome coro .................... .................... ..................... .................... .................... ....................
13802 MW064627.1 |Severe acute respiratory syndrome coro .................... .................... ..................... .................... .................... ....................
13803 MW064628.1 |Severe acute respiratory syndrome coro .................... .................... ..................... .................... .................... ....................
13804 MW064629.1 |Severe acute respiratory syndrome coro .................... .................... ..................... .................... .................... ....................
13805 MW064630.1 |Severe acute respiratory syndrome coro .................... .................... ..................... .................... .................... ....................
13806 MW064631.1 |Severe acute respiratory syndrome coro .................... .................... ..................... .................... .................... ....................
13807 MW064632.1 |Severe acute respiratory syndrome coro .................... .................... ..................... .................... .................... ....................
13808 MW064633.1 |Severe acute respiratory syndrome coro .................... .................... ..................... .................... .................... ....................
13809 MW064634.1 |Severe acute respiratory syndrome coro .................... .................... ..................... .................... .................... ....................
13810 MW064635.1 |Severe acute respiratory syndrome coro .................... .................... ..................... .................... .................... ....................
13811 MW064636.1 |Severe acute respiratory syndrome coro .................... .................... ..................... .................... .................... ....................
13812 MW064637.1 |Severe acute respiratory syndrome coro .................... .................... ..................... .................... .................... ....................
13813 MW064638.1 |Severe acute respiratory syndrome coro .................... .................... ..................... .................... .................... ....................
13814 MW064639.1 |Severe acute respiratory syndrome coro .................... .................... ..................... .................... .................... ....................
13815 MW064640.1 |Severe acute respiratory syndrome coro .................... .................... ..................... .................... .................... ....................
13816 MW064641.1 |Severe acute respiratory syndrome coro .................... .................... ..................... .................... .................... ....................
13817 MW064642.1 |Severe acute respiratory syndrome coro .................... .................... ..................... .................... .................... ....................
13818 MW064643.1 |Severe acute respiratory syndrome coro .................... .................... ..................... .................... .................... ....................
13819 MW064644.1 |Severe acute respiratory syndrome coro .................... .................... ............T........ .................... .................... ....................
13820 MW064645.1 |Severe acute respiratory syndrome coro .................... .................... ..................... .................... .................... ....................
13821 MW064646.1 |Severe acute respiratory syndrome coro .................... .................... ..................... .................... .................... ....................
13822 MW064647.1 |Severe acute respiratory syndrome coro .................... .................... ..................... .................... .................... ....................
13823 MW064648.1 |Severe acute respiratory syndrome coro .................... .................... ..................... .................... .................... ....................
13824 MW064649.1 |Severe acute respiratory syndrome coro .................... .................... ..................... .................... .................... ....................
13825 MW064650.1 |Severe acute respiratory syndrome coro .................... .................... ..................... .................... .................... ....................
13826 MW064651.1 |Severe acute respiratory syndrome coro .................... .................... ..................... .................... .................... ....................
13827 MW064652.1 |Severe acute respiratory syndrome coro .................... .................... ..................... .................... .................... ....................
13828 MW064653.1 |Severe acute respiratory syndrome coro .................... .................... ..................... .................... .................... ....................
13829 MW064654.1 |Severe acute respiratory syndrome coro .................... .................... ..................... .................... .................... ....................
13830 MW064655.1 |Severe acute respiratory syndrome coro .................... .................... ..................... .................... .................... ....................
13831 MW064656.1 |Severe acute respiratory syndrome coro .................... .................... ..................... .................... .................... ....................
13832 MW064657.1 |Severe acute respiratory syndrome coro .................... .................... ..................... .................... .................... ....................
13833 MW064658.1 |Severe acute respiratory syndrome coro .................... .................... ..................... .................... .................... ....................
13834 MW064659.1 |Severe acute respiratory syndrome coro .................... .................... ..................... .................... .................... ....................
13835 MW064660.1 |Severe acute respiratory syndrome coro .................... .................... ..................... .................... .................... ....................
13836 MW064661.1 |Severe acute respiratory syndrome coro .................... .................... ..................... .................... .................... ....................
13837 MW064662.1 |Severe acute respiratory syndrome coro .................... .................... ..................... .................... .................... ....................
13838 MW064663.1 |Severe acute respiratory syndrome coro .................... .................... ..................... .................... .................... ....................
13839 MW064664.1 |Severe acute respiratory syndrome coro .................... .................... ..................... .................... .................... ....................
13840 MW064665.1 |Severe acute respiratory syndrome coro .................... .................... ..................... .................... .................... ....................
13841 MW064666.1 |Severe acute respiratory syndrome coro .................... .................... ..................... .................... .................... ....................
13842 MW064667.1 |Severe acute respiratory syndrome coro .................... .................... ..................... .................... .................... ....................
13843 MW064668.1 |Severe acute respiratory syndrome coro .................... .................... ..................... .................... .................... ....................
13844 MW064669.1 |Severe acute respiratory syndrome coro .................... .................... ..................... .................... .................... ....................
13845 MW064670.1 |Severe acute respiratory syndrome coro .................... .................... ..................... .................... .................... ....................
13846 MW064671.1 |Severe acute respiratory syndrome coro .................... .................... ..................... .................... .................... ....................
13847 MW064672.1 |Severe acute respiratory syndrome coro .................... .................... ..................... .................... .................... ....................
13848 MW064673.1 |Severe acute respiratory syndrome coro .................... .................... ..................... .................... .................... ....................
13849 MW064674.1 |Severe acute respiratory syndrome coro .................... .................... ..................... .................... .................... ....................
13850 MW064675.1 |Severe acute respiratory syndrome coro .................... .................... ..................... .................... .................... ....................
13851 MW064676.1 |Severe acute respiratory syndrome coro .................... .................... ..................... .................... .................... ....................
13852 MW064677.1 |Severe acute respiratory syndrome coro .................... .................... ..................... .................... .................... ....................

| | | | | | | | | |
|---|---|---|---|---|---|---|---|---|
| 13853 | MW064678.1 | \|Severe acute respiratory syndrome coro | .................... | .................... | .................... | .................... | .................... | .................... |
| 13854 | MW064679.1 | \|Severe acute respiratory syndrome coro | .................... | .................... | .................... | .................... | .................... | .................... |
| 13855 | MW064680.1 | \|Severe acute respiratory syndrome coro | .................... | .................... | .................... | .................... | .................... | .................... |
| 13856 | MW064681.1 | \|Severe acute respiratory syndrome coro | .................... | .................... | .................... | .................... | .................... | .................... |
| 13857 | MW064683.1 | \|Severe acute respiratory syndrome coro | .................... | .................... | .................... | .................... | .................... | .................... |
| 13858 | MW064684.1 | \|Severe acute respiratory syndrome coro | .................... | .................... | .................... | .................... | .................... | .................... |
| 13859 | MW064685.1 | \|Severe acute respiratory syndrome coro | .................... | .................... | .................... | .................... | .................... | .................... |
| 13860 | MW064687.1 | \|Severe acute respiratory syndrome coro | .................... | .................... | .................... | .................... | .................... | .................... |
| 13861 | MW064688.1 | \|Severe acute respiratory syndrome coro | .................... | .................... | .................... | .................... | .................... | .................... |
| 13862 | MW064689.1 | \|Severe acute respiratory syndrome coro | .................... | .................... | .................... | .................... | .................... | .................... |
| 13863 | MW064690.1 | \|Severe acute respiratory syndrome coro | .................... | .................... | .................... | .................... | .................... | .................... |
| 13864 | MW064691.1 | \|Severe acute respiratory syndrome coro | .................... | .................... | .................... | .................... | .................... | .................... |
| 13865 | MW064692.1 | \|Severe acute respiratory syndrome coro | .................... | .................... | .................... | .................... | .................... | .................... |
| 13866 | MW064694.1 | \|Severe acute respiratory syndrome coro | .................... | .................... | .................... | .................... | .................... | .................... |
| 13867 | MW064695.1 | \|Severe acute respiratory syndrome coro | .................... | .................... | .................... | .................... | .................... | .................... |
| 13868 | MW064696.1 | \|Severe acute respiratory syndrome coro | .................... | .................... | .................... | .................... | .................... | .................... |
| 13869 | MW064697.1 | \|Severe acute respiratory syndrome coro | .................... | .................... | .................... | .................... | .................... | .................... |
| 13870 | MW064699.1 | \|Severe acute respiratory syndrome coro | .................... | .................... | .................... | .................... | .................... | .................... |
| 13871 | MW064701.1 | \|Severe acute respiratory syndrome coro | .................... | .................... | .................... | .................... | .................... | .................... |
| 13872 | MW064702.1 | \|Severe acute respiratory syndrome coro | .................... | .................... | .................... | .................... | .................... | .................... |
| 13873 | MW064703.1 | \|Severe acute respiratory syndrome coro | .................... | .................... | .................... | .................... | .................... | .................... |
| 13874 | MW064704.1 | \|Severe acute respiratory syndrome coro | .................... | .................... | .................... | .................... | .................... | .................... |
| 13875 | MW064705.1 | \|Severe acute respiratory syndrome coro | .................... | .................... | .................... | .................... | .................... | .................... |
| 13876 | MW064707.1 | \|Severe acute respiratory syndrome coro | .................... | .................... | .................... | .................... | .................... | .................... |
| 13877 | MW064708.1 | \|Severe acute respiratory syndrome coro | .................... | .................... | .................... | .................... | .................... | .................... |
| 13878 | MW064709.1 | \|Severe acute respiratory syndrome coro | .................... | .................... | .................... | .................... | .................... | .................... |
| 13879 | MW064710.1 | \|Severe acute respiratory syndrome coro | .................... | .................... | .................... | .................... | .................... | .................... |
| 13880 | MW064711.1 | \|Severe acute respiratory syndrome coro | .................... | .................... | .................... | .................... | .................... | .................... |
| 13881 | MW064712.1 | \|Severe acute respiratory syndrome coro | .................... | .................... | .................... | .................... | .................... | .................... |
| 13882 | MW064713.1 | \|Severe acute respiratory syndrome coro | .................... | .................... | .................... | .................... | .................... | .................... |
| 13883 | MW064714.1 | \|Severe acute respiratory syndrome coro | .................... | .................... | .................... | .................... | .................... | .................... |
| 13884 | MW064715.1 | \|Severe acute respiratory syndrome coro | .................... | .................... | .................... | .................... | .................... | .................... |
| 13885 | MW064716.1 | \|Severe acute respiratory syndrome coro | .................... | .................... | .................... | .................... | .................... | .................... |
| 13886 | MW064717.1 | \|Severe acute respiratory syndrome coro | .................... | .................... | .................... | .................... | .................... | .................... |
| 13887 | MW064719.1 | \|Severe acute respiratory syndrome coro | .................... | .................... | .................... | .................... | .................... | .................... |
| 13888 | MW064721.1 | \|Severe acute respiratory syndrome coro | .................... | .................... | .................... | .................... | .................... | .................... |
| 13889 | MW064722.1 | \|Severe acute respiratory syndrome coro | .................... | .................... | .................... | .................... | .................... | .................... |
| 13890 | MW064723.1 | \|Severe acute respiratory syndrome coro | .................... | .................... | .................... | .................... | .................... | .................... |
| 13891 | MW064724.1 | \|Severe acute respiratory syndrome coro | .................... | .................... | .................... | .................... | .................... | .................... |
| 13892 | MW064725.1 | \|Severe acute respiratory syndrome coro | .................... | .................... | .................... | .................... | .................... | .................... |
| 13893 | MW064726.1 | \|Severe acute respiratory syndrome coro | .................... | .................... | .................... | .................... | .................... | .................... |
| 13894 | MW064727.1 | \|Severe acute respiratory syndrome coro | .................... | .................... | .................... | .................... | .................... | .................... |
| 13895 | MW064728.1 | \|Severe acute respiratory syndrome coro | .................... | .................... | .................... | .................... | .................... | .................... |
| 13896 | MW064729.1 | \|Severe acute respiratory syndrome coro | .................... | .................... | .................... | .................... | .................... | .................... |
| 13897 | MW064730.1 | \|Severe acute respiratory syndrome coro | .................... | .................... | .................... | .................... | .................... | .................... |
| 13898 | MW064732.1 | \|Severe acute respiratory syndrome coro | .................... | .................... | .................... | .................... | .................... | .................... |
| 13899 | MW064733.1 | \|Severe acute respiratory syndrome coro | .................... | .................... | .................... | .................... | .................... | .................... |
| 13900 | MW064734.1 | \|Severe acute respiratory syndrome coro | .................... | .................... | .................... | .................... | .................... | .................... |
| 13901 | MW064735.1 | \|Severe acute respiratory syndrome coro | .................... | .................... | .................... | .................... | .................... | .................... |
| 13902 | MW064736.1 | \|Severe acute respiratory syndrome coro | .................... | .................... | .................... | .................... | .................... | .................... |
| 13903 | MW064737.1 | \|Severe acute respiratory syndrome coro | .................... | .................... | .................... | .................... | .................... | .................... |
| 13904 | MW064738.1 | \|Severe acute respiratory syndrome coro | .................... | .................... | .................... | .................... | .................... | .................... |
| 13905 | MW064739.1 | \|Severe acute respiratory syndrome coro | .................... | .................... | .................... | .................... | .................... | .................... |
| 13906 | MW064740.1 | \|Severe acute respiratory syndrome coro | .................... | .................... | .................... | .................... | .................... | .................... |
| 13907 | MW064741.1 | \|Severe acute respiratory syndrome coro | .................... | .................... | .................... | .................... | .................... | .................... |
| 13908 | MW064742.1 | \|Severe acute respiratory syndrome coro | .................... | .................... | .................... | .................... | .................... | .................... |
| 13909 | MW064743.1 | \|Severe acute respiratory syndrome coro | .................... | .................... | .................... | .................... | .................... | .................... |
| 13910 | MW064744.1 | \|Severe acute respiratory syndrome coro | .................... | .................... | .................... | .................... | .................... | .................... |
| 13911 | MW064745.1 | \|Severe acute respiratory syndrome coro | .................... | .................... | .................... | .................... | .................... | .................... |
| 13912 | MW064746.1 | \|Severe acute respiratory syndrome coro | .................... | .................... | .................... | .................... | .................... | .................... |
| 13913 | MW064747.1 | \|Severe acute respiratory syndrome coro | .................... | .................... | .................... | .................... | .................... | .................... |
| 13914 | MW064748.1 | \|Severe acute respiratory syndrome coro | .................... | .................... | .................... | .................... | .................... | .................... |
| 13915 | MW064749.1 | \|Severe acute respiratory syndrome coro | .................... | .................... | .................... | .................... | .................... | .................... |
| 13916 | MW064750.1 | \|Severe acute respiratory syndrome coro | .................... | .................... | .................... | .................... | .................... | .................... |
| 13917 | MW064751.1 | \|Severe acute respiratory syndrome coro | .................... | .................... | .................... | .................... | .................... | .................... |
| 13918 | MW064752.1 | \|Severe acute respiratory syndrome coro | .................... | .................... | .................... | .................... | .................... | .................... |
| 13919 | MW064753.1 | \|Severe acute respiratory syndrome coro | .................... | .................... | .................... | .................... | .................... | .................... |
| 13920 | MW064754.1 | \|Severe acute respiratory syndrome coro | .................... | .................... | .................... | .................... | .................... | .................... |
| 13921 | MW064755.1 | \|Severe acute respiratory syndrome coro | .................... | .................... | .................... | .................... | .................... | .................... |
| 13922 | MW064756.1 | \|Severe acute respiratory syndrome coro | .................... | .................... | .................... | .................... | .................... | .................... |
| 13923 | MW064757.1 | \|Severe acute respiratory syndrome coro | .................... | .................... | .................... | .................... | .................... | .................... |
| 13924 | MW064758.1 | \|Severe acute respiratory syndrome coro | .................... | .................... | .................... | .................... | .................... | .................... |
| 13925 | MW064759.1 | \|Severe acute respiratory syndrome coro | .................... | .................... | .................... | .................... | .................... | .................... |
| 13926 | MW064760.1 | \|Severe acute respiratory syndrome coro | .................... | .................... | .................... | .................... | .................... | .................... |
| 13927 | MW064761.1 | \|Severe acute respiratory syndrome coro | .................... | .................... | .................... | .................... | .................... | .................... |
| 13928 | MW064762.1 | \|Severe acute respiratory syndrome coro | .................... | .................... | .................... | .................... | .................... | .................... |
| 13929 | MW064763.1 | \|Severe acute respiratory syndrome coro | .................... | .................... | .................... | .................... | .................... | .................... |
| 13930 | MW064764.1 | \|Severe acute respiratory syndrome coro | .................... | .................... | .................... | .................... | .................... | .................... |

| | | | | | | | | | |
|---|---|---|---|---|---|---|---|---|---|
| 13931 | MW064765.1 | \|Severe acute respiratory syndrome coro | .................... | .................... | .................... | .................... | .................... | .................... | |
| 13932 | MW064766.1 | \|Severe acute respiratory syndrome coro | .................... | .................... | .................... | .................... | .................... | .................... | |
| 13933 | MW064767.1 | \|Severe acute respiratory syndrome coro | .................... | .................... | .................... | .................... | .................... | .................... | |
| 13934 | MW064769.1 | \|Severe acute respiratory syndrome coro | .................... | .................... | .................... | .................... | .................... | .................... | |
| 13935 | MW064770.1 | \|Severe acute respiratory syndrome coro | .................... | .................... | .................... | .................... | .................... | .................... | |
| 13936 | MW064771.1 | \|Severe acute respiratory syndrome coro | .................... | .................... | .................... | .................... | .................... | .................... | |
| 13937 | MW064772.1 | \|Severe acute respiratory syndrome coro | .................... | .................... | .................... | .................... | .................... | .................... | |
| 13938 | MW064774.1 | \|Severe acute respiratory syndrome coro | .................... | .................... | .................... | .................... | .................... | .................... | |
| 13939 | MW064775.1 | \|Severe acute respiratory syndrome coro | .................... | .................... | .................... | .................... | .................... | .................... | |
| 13940 | MW064776.1 | \|Severe acute respiratory syndrome coro | .................... | .................... | .................... | .................... | .................... | .................... | |
| 13941 | MW064777.1 | \|Severe acute respiratory syndrome coro | .................... | .................... | .................... | .................... | .................... | .................... | |
| 13942 | MW064778.1 | \|Severe acute respiratory syndrome coro | .................... | .................... | .................... | .................... | .................... | .................... | |
| 13943 | MW064779.1 | \|Severe acute respiratory syndrome coro | .................... | .................... | .................... | .................... | .................... | .................... | |
| 13944 | MW064780.1 | \|Severe acute respiratory syndrome coro | .................... | .................... | .................... | .................... | ...A................ | .................... | |
| 13945 | MW064782.1 | \|Severe acute respiratory syndrome coro | .................... | .................... | .................... | .................... | .................... | .................... | |
| 13946 | MW064784.1 | \|Severe acute respiratory syndrome coro | .................... | .................... | .................... | .................... | .................... | .................... | |
| 13947 | MW064785.1 | \|Severe acute respiratory syndrome coro | .................... | .................... | .................... | .................... | .................... | .................... | |
| 13948 | MW064786.1 | \|Severe acute respiratory syndrome coro | .................... | .................... | .................... | .................... | .................... | .................... | |
| 13949 | MW064787.1 | \|Severe acute respiratory syndrome coro | .................... | .................... | .................... | .................... | .................... | .................... | |
| 13950 | MW064788.1 | \|Severe acute respiratory syndrome coro | .................... | .................... | .................... | .................... | .................... | .................... | |
| 13951 | MW064789.1 | \|Severe acute respiratory syndrome coro | .................... | .................... | .................... | .................... | .................... | .................... | |
| 13952 | MW064790.1 | \|Severe acute respiratory syndrome coro | .................... | .................... | .................... | .................... | .................... | .................... | |
| 13953 | MW064791.1 | \|Severe acute respiratory syndrome coro | .................... | .................... | .................... | .................... | .................... | .................... | |
| 13954 | MW064792.1 | \|Severe acute respiratory syndrome coro | .................... | .................... | .................... | .................... | .................... | .................... | |
| 13955 | MW064793.1 | \|Severe acute respiratory syndrome coro | .................... | .................... | .................... | .................... | .................... | .................... | |
| 13956 | MW064794.1 | \|Severe acute respiratory syndrome coro | .................... | .................... | .................... | .................... | .................... | .................... | |
| 13957 | MW064795.1 | \|Severe acute respiratory syndrome coro | .................... | .................... | .................... | .................... | .................... | .................... | |
| 13958 | MW064796.1 | \|Severe acute respiratory syndrome coro | .................... | .................... | .................... | .................... | .................... | .................... | |
| 13959 | MW064797.1 | \|Severe acute respiratory syndrome coro | .................... | .................... | .................... | .................... | .................... | .................... | |
| 13960 | MW064798.1 | \|Severe acute respiratory syndrome coro | .................... | .................... | .................... | .................... | .................... | .................... | |
| 13961 | MW064799.1 | \|Severe acute respiratory syndrome coro | .................... | .................... | .................... | .................... | .................... | .................... | |
| 13962 | MW064800.1 | \|Severe acute respiratory syndrome coro | .................... | .................... | .................... | .................... | .................... | .................... | |
| 13963 | MW064801.1 | \|Severe acute respiratory syndrome coro | .................... | .................... | .................... | .................... | .................... | .................... | |
| 13964 | MW064802.1 | \|Severe acute respiratory syndrome coro | .................... | .................... | .................... | .................... | .................... | .................... | |
| 13965 | MW064803.1 | \|Severe acute respiratory syndrome coro | .................... | .................... | .................... | .................... | .................... | .................... | |
| 13966 | MW064804.1 | \|Severe acute respiratory syndrome coro | .................... | .................... | .................... | .................... | .................... | .................... | |
| 13967 | MW064805.1 | \|Severe acute respiratory syndrome coro | .................... | .................... | .................... | .................... | .................... | .................... | |
| 13968 | MW064806.1 | \|Severe acute respiratory syndrome coro | .................... | .................... | .................... | .................... | .................... | .................... | |
| 13969 | MW064807.1 | \|Severe acute respiratory syndrome coro | .................... | .................... | .................... | .................... | .................... | .................... | |
| 13970 | MW064808.1 | \|Severe acute respiratory syndrome coro | .................... | ...................A | .................... | .................... | .................... | .................... | 1 |
| 13971 | MW064809.1 | \|Severe acute respiratory syndrome coro | .................... | .................... | .................... | .................... | .................... | .................... | |
| 13972 | MW064810.1 | \|Severe acute respiratory syndrome coro | .................... | .................... | .................... | .................... | .................... | .................... | |
| 13973 | MW064812.1 | \|Severe acute respiratory syndrome coro | .................... | .................... | .................... | .................... | .................... | .................... | |
| 13974 | MW064813.1 | \|Severe acute respiratory syndrome coro | .................... | .................... | .................... | .................... | .................... | .................... | |
| 13975 | MW064814.1 | \|Severe acute respiratory syndrome coro | .................... | .................... | .................... | .................... | .................... | .................... | |
| 13976 | MW064815.1 | \|Severe acute respiratory syndrome coro | .................... | .................... | .................... | .................... | .................... | .................... | |
| 13977 | MW064816.1 | \|Severe acute respiratory syndrome coro | .................... | ...................A | .................... | .................... | .................... | .................... | 1 |
| 13978 | MW064817.1 | \|Severe acute respiratory syndrome coro | .................... | .................... | .................... | .................... | .................... | .................... | |
| 13979 | MW064818.1 | \|Severe acute respiratory syndrome coro | .................... | .................... | .................... | .................... | .................... | .................... | |
| 13980 | MW064819.1 | \|Severe acute respiratory syndrome coro | .................... | .................... | .................... | .................... | .................... | .................... | |
| 13981 | MW064820.1 | \|Severe acute respiratory syndrome coro | .................... | .................... | .................... | .................... | .................... | .................... | |
| 13982 | MW064821.1 | \|Severe acute respiratory syndrome coro | .................... | .................... | .................... | .................... | .................... | .................... | |
| 13983 | MW064823.1 | \|Severe acute respiratory syndrome coro | .................... | .................... | .................... | .................... | .................... | .................... | |
| 13984 | MW064824.1 | \|Severe acute respiratory syndrome coro | .................... | .................... | .................... | .................... | .................... | .................... | |
| 13985 | MW064825.1 | \|Severe acute respiratory syndrome coro | .................... | .................... | .................... | .................... | .................... | .................... | |
| 13986 | MW064826.1 | \|Severe acute respiratory syndrome coro | .................... | .................... | .................... | .................... | .................... | .................... | |
| 13987 | MW064827.1 | \|Severe acute respiratory syndrome coro | .................... | .................... | .................... | .................... | .................... | .................... | |
| 13988 | MW064828.1 | \|Severe acute respiratory syndrome coro | .................... | .................... | .................... | .................... | .................... | .................... | |
| 13989 | MW064829.1 | \|Severe acute respiratory syndrome coro | .................... | .................... | .................... | .................... | .................... | .................... | |
| 13990 | MW064830.1 | \|Severe acute respiratory syndrome coro | .................... | .................... | .................... | .................... | .................... | .................... | |
| 13991 | MW064831.1 | \|Severe acute respiratory syndrome coro | .................... | .................... | .................... | .................... | .................... | .................... | |
| 13992 | MW064832.1 | \|Severe acute respiratory syndrome coro | .................... | .................... | .................... | .................... | .................... | .................... | |
| 13993 | MW064834.1 | \|Severe acute respiratory syndrome coro | .................... | .................... | .................... | .................... | .................... | .................... | |
| 13994 | MW064835.1 | \|Severe acute respiratory syndrome coro | .................... | .................... | .................... | .................... | .................... | .................... | |
| 13995 | MW064837.1 | \|Severe acute respiratory syndrome coro | .................... | .................... | .................... | .................... | .................... | .................... | |
| 13996 | MW064838.1 | \|Severe acute respiratory syndrome coro | .................... | .................... | .................... | .................... | .................... | .................... | |
| 13997 | MW064839.1 | \|Severe acute respiratory syndrome coro | .................... | .................... | .................... | .................... | .................... | .................... | |
| 13998 | MW064840.1 | \|Severe acute respiratory syndrome coro | .................... | .................... | .................... | .................... | .................... | .................... | |
| 13999 | MW064841.1 | \|Severe acute respiratory syndrome coro | .................... | .................... | .................... | .................... | .................... | .................... | |
| 14000 | MW064842.1 | \|Severe acute respiratory syndrome coro | .................... | .................... | .................... | .................... | .................... | .................... | |
| 14001 | MW064843.1 | \|Severe acute respiratory syndrome coro | .................... | .................... | .................... | .................... | .................... | .................... | |
| 14002 | MW064844.1 | \|Severe acute respiratory syndrome coro | .................... | .................... | .................... | .................... | .................... | .................... | |
| 14003 | MW064845.1 | \|Severe acute respiratory syndrome coro | .................... | .................... | .................... | .................... | .................... | .................... | |
| 14004 | MW064846.1 | \|Severe acute respiratory syndrome coro | .................... | .................... | .................... | .................... | .................... | .................... | |
| 14005 | MW064847.1 | \|Severe acute respiratory syndrome coro | .................... | .................... | .................... | .................... | .................... | .................... | |
| 14006 | MW064848.1 | \|Severe acute respiratory syndrome coro | .................... | .................... | .................... | .................... | .................... | .................... | |
| 14007 | MW064849.1 | \|Severe acute respiratory syndrome coro | .................... | .................... | .................... | .................... | .................... | .................... | |
| 14008 | MW064850.1 | \|Severe acute respiratory syndrome coro | .................... | .................... | .................... | .................... | .................... | .................... | |

```
14009 MW064851.1 |Severe acute respiratory syndrome coro    ....................  ....................  .....................  ....................  ....................  ....................
14010 MW064852.1 |Severe acute respiratory syndrome coro    ....................  ....................  .....................  ....................  ....................  ....................
14011 MW064853.1 |Severe acute respiratory syndrome coro    ....................  ....................  .....................  ....................  ....................  ....................
14012 MW064854.1 |Severe acute respiratory syndrome coro    ....................  ....................  .....................  ....................  ....................  ....................
14013 MW064856.1 |Severe acute respiratory syndrome coro    ....................  ....................  .....................  ....................  ....................  ....................
14014 MW064857.1 |Severe acute respiratory syndrome coro    ....................  ....................  .....................  ....................  ....................  ....................
14015 MW064858.1 |Severe acute respiratory syndrome coro    ....................  ....................  .....................  ....................  ....................  ....................
14016 MW064859.1 |Severe acute respiratory syndrome coro    ....................  ....................  .....................  ....................  ....................  ....................
14017 MW064860.1 |Severe acute respiratory syndrome coro    ....................  ....................  .....................  ....................  ....................  ....................
14018 MW064862.1 |Severe acute respiratory syndrome coro    ....................  ....................  .....................  ....................  ....................  ....................
14019 MW064863.1 |Severe acute respiratory syndrome coro    ....................  ....................  .....................  ....................  ....................  ....................
14020 MW064864.1 |Severe acute respiratory syndrome coro    ....................  ....................  .....................  ....................  ....................  ....................
14021 MW064865.1 |Severe acute respiratory syndrome coro    ....................  ....................  .....................  ....................  ....................  ....................
14022 MW064866.1 |Severe acute respiratory syndrome coro    ....................  ....................  .....................  ....................  ....................  ....................
14023 MW064867.1 |Severe acute respiratory syndrome coro    ....................  ....................  .....................  ....................  ....................  ....................
14024 MW064868.1 |Severe acute respiratory syndrome coro    ....................  ....................  .....................  ....................  ....................  ....................
14025 MW064869.1 |Severe acute respiratory syndrome coro    ....................  ....................  .....................  ....................  ....................  ....................
14026 MW064870.1 |Severe acute respiratory syndrome coro    ....................  ....................  .....................  ....................  ....................  ....................
14027 MW064871.1 |Severe acute respiratory syndrome coro    ....................  ....................  .....................  ....................  ....................  ....................
14028 MW064872.1 |Severe acute respiratory syndrome coro    ....................  ....................  .....................  ....................  ....................  ....................
14029 MW064873.1 |Severe acute respiratory syndrome coro    ....................  ....................  .....................  ....................  ....................  ....................
14030 MW064874.1 |Severe acute respiratory syndrome coro    ....................  ....................  .....................  ....................  ....................  ....................
14031 MW064875.1 |Severe acute respiratory syndrome coro    ....................  ....................  .....................  ....................  ....................  ....................
14032 MW064876.1 |Severe acute respiratory syndrome coro    ....................  ....................  .....................  ....................  ....................  ....................
14033 MW064877.1 |Severe acute respiratory syndrome coro    ....................  ....................  .....................  ....................  ....................  ....................
14034 MW064878.1 |Severe acute respiratory syndrome coro    ....................  ....................  .....................  ....................  ....................  ....................
14035 MW064879.1 |Severe acute respiratory syndrome coro    ....................  ....................  .....................  ....................  ....................  ....................
14036 MW064880.1 |Severe acute respiratory syndrome coro    ....................  ....................  .....................  ....................  ....................  ....................
14037 MW064881.1 |Severe acute respiratory syndrome coro    ....................  ....................  .....................  ....................  ....................  ....................
14038 MW064882.1 |Severe acute respiratory syndrome coro    ....................  ....................  .....................  ....................  ....................  ....................
14039 MW064883.1 |Severe acute respiratory syndrome coro    ....................  ....................  .....................  ....................  ....................  ....................
14040 MW064884.1 |Severe acute respiratory syndrome coro    ....................  ....................  .....................  ....................  ....................  ....................
14041 MW064885.1 |Severe acute respiratory syndrome coro    ....................  ....................  .....................  ....................  ....................  ....................
14042 MW064886.1 |Severe acute respiratory syndrome coro    ....................  ....................  .....................  ....................  ....................  ....................
14043 MW064888.1 |Severe acute respiratory syndrome coro    ....................  ....................  .....................  ....................  ....................  ....................
14044 MW064889.1 |Severe acute respiratory syndrome coro    ....................  ....................  .....................  ....................  ....................  ....................
14045 MW064890.1 |Severe acute respiratory syndrome coro    ....................  ....................  .....................  ....................  ....................  ....................
14046 MW064891.1 |Severe acute respiratory syndrome coro    ....................  ....................  .....................  ....................  ....................  ....................
14047 MW064892.1 |Severe acute respiratory syndrome coro    ....................  ....................  .....................  ....................  ....................  ....................
14048 MW064893.1 |Severe acute respiratory syndrome coro    ....................  ....................  .....................  ....................  ....................  ....................
14049 MW064894.1 |Severe acute respiratory syndrome coro    ....................  ....................  .....................  ....................  ....................  ....................
14050 MW064895.1 |Severe acute respiratory syndrome coro    ....................  ....................  .....................  ....................  ....................  ....................
14051 MW064896.1 |Severe acute respiratory syndrome coro    ....................  ....................  .....................  ....................  ....................  ....................
14052 MW064897.1 |Severe acute respiratory syndrome coro    ....................  ....................  .....................  ....................  ....................  ....................
14053 MW064898.1 |Severe acute respiratory syndrome coro    ....................  ....................  .....................  ....................  ....................  ....................
14054 MW064899.1 |Severe acute respiratory syndrome coro    ....................  ....................  .....................  ....................  ....................  ....................
14055 MW064900.1 |Severe acute respiratory syndrome coro    ....................  ....................  .....................  ....................  ....................  ....................
14056 MW064901.1 |Severe acute respiratory syndrome coro    ....................  ....................  .....................  ....................  ....................  ....................
14057 MW064902.1 |Severe acute respiratory syndrome coro    ....................  ....................  .....................  ....................  ....................  ....................
14058 MW064903.1 |Severe acute respiratory syndrome coro    ....................  ....................  .....................  ....................  ....................  ....................
14059 MW064904.1 |Severe acute respiratory syndrome coro    ....................  ....................  .....................  ....................  ....................  ....................
14060 MW064905.1 |Severe acute respiratory syndrome coro    ....................  ....................  .....................  ....................  ....................  ....................
14061 MW064906.1 |Severe acute respiratory syndrome coro    ....................  ....................  .....................  ....................  ....................  ....................
14062 MW064907.1 |Severe acute respiratory syndrome coro    ....................  ....................  .....................  ....................  ....................  ....................
14063 MW064908.1 |Severe acute respiratory syndrome coro    ....................  ....................  .....................  ....................  ....................  ....................
14064 MW064909.1 |Severe acute respiratory syndrome coro    ....................  ....................  .....................  ....................  ....................  ....................
14065 MW064910.1 |Severe acute respiratory syndrome coro    ....................  ....................  .....................  ....................  ....................  ....................
14066 MW064911.1 |Severe acute respiratory syndrome coro    ....................  ....................  .....................  ....................  ....................  ....................
14067 MW064912.1 |Severe acute respiratory syndrome coro    ....................  ....................  .....................  ....................  ....................  ....................
14068 MW064913.1 |Severe acute respiratory syndrome coro    ....................  ....................  .....................  ....................  ....................  ....................
14069 MW064914.1 |Severe acute respiratory syndrome coro    ....................  ....................  .....................  ....................  ....................  ....................
14070 MW064915.1 |Severe acute respiratory syndrome coro    ....................  ....................  .....................  ....................  ....................  ....................
14071 MW064916.1 |Severe acute respiratory syndrome coro    ....................  ....................  .....................  ....................  ....................  ....................
14072 MW064917.1 |Severe acute respiratory syndrome coro    ....................  ....................  .....................  ....................  ....................  ....................
14073 MW064918.1 |Severe acute respiratory syndrome coro    ....................  ....................  .....................  ....................  ....................  ....................
14074 MW064919.1 |Severe acute respiratory syndrome coro    ....................  ....................  .....................  ....................  ....................  ....................
14075 MW064920.1 |Severe acute respiratory syndrome coro    ....................  ....................  .....................  ....................  ....................  ....................
14076 MW064921.1 |Severe acute respiratory syndrome coro    ....................  ....................  .....................  ....................  ....................  ....................
14077 MW064922.1 |Severe acute respiratory syndrome coro    ....................  ....................  .....................  ....................  ....................  ....................
14078 MW064923.1 |Severe acute respiratory syndrome coro    ....................  ....................  .....................  ....................  ....................  ....................
14079 MW064924.1 |Severe acute respiratory syndrome coro    ....................  ....................  .....................  ....................  ....................  ....................
14080 MW064925.1 |Severe acute respiratory syndrome coro    ....................  ....................  .....................  ....................  ....................  ....................
14081 MW064926.1 |Severe acute respiratory syndrome coro    ....................  ....................  .....................  ....................  ....................  ....................
14082 MW064927.1 |Severe acute respiratory syndrome coro    ....................  ....................  .....................  ....................  ....................  ....................
14083 MW064928.1 |Severe acute respiratory syndrome coro    ....................  ....................  .....................  ....................  ....................  ....................
14084 MW064929.1 |Severe acute respiratory syndrome coro    ....................  ....................  .....................  ....................  ....................  ....................
14085 MW064930.1 |Severe acute respiratory syndrome coro    ....................  ....................  .....................  ....................  ....................  ....................
14086 MW064931.1 |Severe acute respiratory syndrome coro    ....................  ....................  .....................  ....................  ....................  ....................
```

```
14087 MW064932.1 |Severe acute respiratory syndrome coro    .................... .................... .................... .................... .................... ....................
14088 MW064933.1 |Severe acute respiratory syndrome coro    .................... .................... .................... .................... .................... ....................
14089 MW064934.1 |Severe acute respiratory syndrome coro    .................... .................... .................... .................... .................... ....................
14090 MW064935.1 |Severe acute respiratory syndrome coro    .................... .................... .................... .................... .................... ....................
14091 MW064936.1 |Severe acute respiratory syndrome coro    .................... .................... .................... .................... .................... ....................
14092 MW064937.1 |Severe acute respiratory syndrome coro    .................... .................... .................... .................... .................... ....................
14093 MW064938.1 |Severe acute respiratory syndrome coro    .................... .................... .................... .................... .................... ....................
14094 MW064940.1 |Severe acute respiratory syndrome coro    .................... .................... .................... .................... .................... ....................
14095 MW064941.1 |Severe acute respiratory syndrome coro    .................... .................... .................... .................... .................... ....................
14096 MW064942.1 |Severe acute respiratory syndrome coro    .................... .................... .................... .................... .................... ....................
14097 MW064943.1 |Severe acute respiratory syndrome coro    .................... .................... .................... .................... .................... ....................
14098 MW064944.1 |Severe acute respiratory syndrome coro    .................... .................... .................... .................... .................... ....................
14099 MW064945.1 |Severe acute respiratory syndrome coro    .................... .................... .................... .................... .................... ....................
14100 MW064946.1 |Severe acute respiratory syndrome coro    .................... .................... .................... .................... .................... ....................
14101 MW064947.1 |Severe acute respiratory syndrome coro    .................... .................... .................... .................... .................... ....................
14102 MW064948.1 |Severe acute respiratory syndrome coro    .................... .................... .................... .................... .................... ....................
14103 MW064949.1 |Severe acute respiratory syndrome coro    .................... .................... .................... .................... .................... ....................
14104 MW064950.1 |Severe acute respiratory syndrome coro    .................... .................... .................... .................... .................... ....................
14105 MW064951.1 |Severe acute respiratory syndrome coro    .................... .................... .................... .................... .................... ....................
14106 MW064952.1 |Severe acute respiratory syndrome coro    .................... .................... .................... .................... .................... ....................
14107 MW064953.1 |Severe acute respiratory syndrome coro    .................... .................... .................... .................... .................... ....................
14108 MW064954.1 |Severe acute respiratory syndrome coro    .................... .................... .................... .................... .................... ....................
14109 MW064955.1 |Severe acute respiratory syndrome coro    .................... .................... .................... .................... .................... ....................
14110 MW064956.1 |Severe acute respiratory syndrome coro    .................... .................... .................... .................... .................... ....................
14111 MW064957.1 |Severe acute respiratory syndrome coro    .................... .................... .................... .................... .................... ....................
14112 MW064958.1 |Severe acute respiratory syndrome coro    .................... .................... .................... .................... .................... ....................
14113 MW064959.1 |Severe acute respiratory syndrome coro    .................... .................... .................... .................... .................... ....................
14114 MW064960.1 |Severe acute respiratory syndrome coro    .................... .................... .................... .................... .................... ....................
14115 MW064961.1 |Severe acute respiratory syndrome coro    .................... .................... .................... .................... .................... ....................
14116 MW064962.1 |Severe acute respiratory syndrome coro    .................... .................... .................... .................... .................... ....................
14117 MW064963.1 |Severe acute respiratory syndrome coro    .................... .................... .................... .................... .................... ....................
14118 MW064964.1 |Severe acute respiratory syndrome coro    .................... .................... .................... .................... .................... ....................
14119 MW064965.1 |Severe acute respiratory syndrome coro    .................... .................... .................... .................... .................... ....................
14120 MW064966.1 |Severe acute respiratory syndrome coro    .................... .................... .................... .................... .................... ....................
14121 MW064967.1 |Severe acute respiratory syndrome coro    .................... .................... .................... .................... .................... ....................
14122 MW064968.1 |Severe acute respiratory syndrome coro    .................... .................... .................... .................... .................... ....................
14123 MW064969.1 |Severe acute respiratory syndrome coro    .................... .................... .................... .................... .................... ....................
14124 MW064970.1 |Severe acute respiratory syndrome coro    .................... .................... .................... .................... .................... ....................
14125 MW064971.1 |Severe acute respiratory syndrome coro    .................... .................... .................... .................... .................... ....................
14126 MW064972.1 |Severe acute respiratory syndrome coro    .................... .................... .................... .................... .................... ....................
14127 MW064973.1 |Severe acute respiratory syndrome coro    .................... .................... .................... .................... .................... ....................
14128 MW064974.1 |Severe acute respiratory syndrome coro    .................... .................... .................... .................... .................... ....................
14129 MW064975.1 |Severe acute respiratory syndrome coro    .................... .................... .................... .................... .................... ....................
14130 MW064976.1 |Severe acute respiratory syndrome coro    .................... .................... .................... .................... .................... ....................
14131 MW064977.1 |Severe acute respiratory syndrome coro    .................... .................... .................... .................... .................... ....................
14132 MW064978.1 |Severe acute respiratory syndrome coro    .................... .................... .................... .................... .................... ....................
14133 MW064979.1 |Severe acute respiratory syndrome coro    .................... .................... .................... .................... .................... ....................
14134 MW064980.1 |Severe acute respiratory syndrome coro    .................... .................... .................... .................... .................... ....................
14135 MW064981.1 |Severe acute respiratory syndrome coro    .................... .................... .................... .................... .................... ....................
14136 MW064982.1 |Severe acute respiratory syndrome coro    .................... .................... .................... .................... .................... ....................
14137 MW064984.1 |Severe acute respiratory syndrome coro    .................... .................... .................... .................... .................... ....................
14138 MW064985.1 |Severe acute respiratory syndrome coro    .................... .................... .................... .................... .................... ....................
14139 MW064986.1 |Severe acute respiratory syndrome coro    .................... .................... .................... .................... .................... ....................
14140 MW064987.1 |Severe acute respiratory syndrome coro    .................... .................... .................... .................... .................... ....................
14141 MW064989.1 |Severe acute respiratory syndrome coro    .................... .................... .................... .................... .................... ....................
14142 MW064990.1 |Severe acute respiratory syndrome coro    .................... .................... .................... .................... .................... ....................
14143 MW064992.1 |Severe acute respiratory syndrome coro    .................... .................... .................... .................... .................... ....................
14144 MW064993.1 |Severe acute respiratory syndrome coro    .................... .................... .................... .................... .................... ....................
14145 MW064994.1 |Severe acute respiratory syndrome coro    .................... .................... .................... .................... .................... ....................
14146 MW064995.1 |Severe acute respiratory syndrome coro    .................... .................... .................... .................... .................... ....................
14147 MW064996.1 |Severe acute respiratory syndrome coro    .................... .................... .................... .................... .................... ....................
14148 MW064997.1 |Severe acute respiratory syndrome coro    .................... .................... .................... .................... .................... ....................
14149 MW064998.1 |Severe acute respiratory syndrome coro    .................... .................... .................... .................... .................... ....................
14150 MW064999.1 |Severe acute respiratory syndrome coro    .................... .................... .................... .................... .................... ....................
14151 MW065000.1 |Severe acute respiratory syndrome coro    .................... .................... .................... .................... .................... ....................
14152 MW065001.1 |Severe acute respiratory syndrome coro    .................... .................... .................... .................... .................... ....................
14153 MW065002.1 |Severe acute respiratory syndrome coro    .................... .................... .................... .................... .................... ....................
14154 MW065003.1 |Severe acute respiratory syndrome coro    .................... .................... .................... .................... .................... ....................
14155 MW065004.1 |Severe acute respiratory syndrome coro    .................... .................... .................... .................... .................... ....................
14156 MW065005.1 |Severe acute respiratory syndrome coro    .................... .................... .................... .................... .................... ....................
14157 MW065006.1 |Severe acute respiratory syndrome coro    .................... .................... .................... .................... .................... ....................
14158 MW065007.1 |Severe acute respiratory syndrome coro    .................... .................... .................... .................... .................... ....................
14159 MW065008.1 |Severe acute respiratory syndrome coro    .................... .................... .................... .................... .................... ....................
14160 MW065009.1 |Severe acute respiratory syndrome coro    .................... .................... .................... .................... .................... ....................
14161 MW065010.1 |Severe acute respiratory syndrome coro    .................... .................... .................... .................... .................... ....................
14162 MW065011.1 |Severe acute respiratory syndrome coro    .................... .................... .................... .................... .................... ....................
14163 MW065012.1 |Severe acute respiratory syndrome coro    .................... .................... .................... .................... .................... ....................
14164 MW065013.1 |Severe acute respiratory syndrome coro    .................... .................... .................... .................... .................... ....................
```

```
14165 MW065014.1 |Severe acute respiratory syndrome coro    ....................  ....................  .....................  ....................  ....................  ....................
14166 MW065015.1 |Severe acute respiratory syndrome coro    ....................  ....................  .....................  ....................  ....................  ....................
14167 MW065016.1 |Severe acute respiratory syndrome coro    ....................  ....................  .....................  ....................  ....................  ....................
14168 MW065017.1 |Severe acute respiratory syndrome coro    ....................  ....................  .....................  ....................  ....................  ....................
14169 MW065018.1 |Severe acute respiratory syndrome coro    ....................  ....................  .....................  ....................  ....................  ....................
14170 MW065019.1 |Severe acute respiratory syndrome coro    ....................  ....................  .....................  ....................  ....................  ....................
14171 MW065020.1 |Severe acute respiratory syndrome coro    ....................  ....................  .....................  ....................  ....................  ....................
14172 MW065021.1 |Severe acute respiratory syndrome coro    ....................  ....................  .....................  ....................  ....................  ....................
14173 MW065022.1 |Severe acute respiratory syndrome coro    ....................  ....................  .....................  ....................  ....................  ....................
14174 MW065023.1 |Severe acute respiratory syndrome coro    ....................  ....................  .....................  ....................  ....................  ....................
14175 MW065024.1 |Severe acute respiratory syndrome coro    ....................  ....................  .....................  ....................  ....................  ....................
14176 MW065025.1 |Severe acute respiratory syndrome coro    ....................  ....................  .....................  ....................  ....................  ....................
14177 MW065026.1 |Severe acute respiratory syndrome coro    ....................  ....................  .....................  ....................  ....................  ....................
14178 MW065027.1 |Severe acute respiratory syndrome coro    ....................  ....................  .....................  ....................  ....................  ....................
14179 MW065028.1 |Severe acute respiratory syndrome coro    ....................  ....................  .....................  ....................  ....................  ....................
14180 MW065029.1 |Severe acute respiratory syndrome coro    ....................  ....................  .....................  ....................  ....................  ....................
14181 MW065030.1 |Severe acute respiratory syndrome coro    ....................  ....................  .....................  ....................  ....................  ....................
14182 MW065031.1 |Severe acute respiratory syndrome coro    ....................  ....................  .....................  ....................  ....................  ....................
14183 MW065032.1 |Severe acute respiratory syndrome coro    ....................  ....................  .....................  ....................  ....................  ....................
14184 MW065033.1 |Severe acute respiratory syndrome coro    ....................  ....................  .....................  ....................  ....................  ....................
14185 MW065034.1 |Severe acute respiratory syndrome coro    ....................  ....................  .....................  ....................  ....................  ....................
14186 MW065035.1 |Severe acute respiratory syndrome coro    ....................  ....................  .....................  ....................  ....................  ....................
14187 MW065036.1 |Severe acute respiratory syndrome coro    ....................  ....................  .....................  ....................  ....................  ....................
14188 MW065037.1 |Severe acute respiratory syndrome coro    ....................  ....................  .....................  ....................  ....................  ....................
14189 MW065038.1 |Severe acute respiratory syndrome coro    ....................  ....................  .....................  ....................  ....................  ....................
14190 MW065039.1 |Severe acute respiratory syndrome coro    ....................  ....................  .....................  ....................  ....................  ....................
14191 MW065040.1 |Severe acute respiratory syndrome coro    ....................  ....................  .....................  ....................  ....................  ....................
14192 MW065041.1 |Severe acute respiratory syndrome coro    ....................  ....................  .....................  ....................  ....................  ....................
14193 MW065042.1 |Severe acute respiratory syndrome coro    ....................  ....................  .....................  ....................  ....................  ....................
14194 MW065043.1 |Severe acute respiratory syndrome coro    ....................  ....................  .....................  ....................  ....................  ....................
14195 MW065044.1 |Severe acute respiratory syndrome coro    ....................  ....................  .....................  ....................  ....................  ....................
14196 MW065046.1 |Severe acute respiratory syndrome coro    ....................  ....................  .....................  ....................  ....................  ....................
14197 MW065047.1 |Severe acute respiratory syndrome coro    ....................  ....................  .....................  ....................  ....................  ....................
14198 MW065048.1 |Severe acute respiratory syndrome coro    ....................  ....................  .....................  ....................  ....................  ....................
14199 MW065049.1 |Severe acute respiratory syndrome coro    ....................  ....................  .....................  ....................  ....................  ....................
14200 MW065050.1 |Severe acute respiratory syndrome coro    ....................  ....................  .....................  ....................  ....................  ....................
14201 MW065051.1 |Severe acute respiratory syndrome coro    ....................  ....................  .....................  ....................  ....................  ....................
14202 MW065052.1 |Severe acute respiratory syndrome coro    ....................  ....................  .....................  ....................  ....................  ....................
14203 MW065053.1 |Severe acute respiratory syndrome coro    ....................  ....................  .....................  ....................  ....................  ....................
14204 MW065054.1 |Severe acute respiratory syndrome coro    ....................  ....................  .....................  ....................  ....................  ....................
14205 MW065055.1 |Severe acute respiratory syndrome coro    ....................  ....................  .....................  ....................  ....................  ....................
14206 MW065056.1 |Severe acute respiratory syndrome coro    ....................  ....................  .....................  ....................  ....................  ....................
14207 MW065057.1 |Severe acute respiratory syndrome coro    ....................  ....................  .....................  ....................  ....................  ....................
14208 MW065058.1 |Severe acute respiratory syndrome coro    ....................  ....................  .....................  ....................  ....................  ....................
14209 MW065059.1 |Severe acute respiratory syndrome coro    ....................  ....................  .....................  ....................  ....................  ....................
14210 MW065060.1 |Severe acute respiratory syndrome coro    ....................  ....................  .....................  ....................  ....................  ....................
14211 MW065061.1 |Severe acute respiratory syndrome coro    ....................  ....................  .....................  ....................  ....................  ....................
14212 MW065062.1 |Severe acute respiratory syndrome coro    ....................  ....................  .....................  ....................  ....................  ....................
14213 MW065063.1 |Severe acute respiratory syndrome coro    ....................  ....................  .....................  ....................  ....................  ....................
14214 MW065064.1 |Severe acute respiratory syndrome coro    ....................  ....................  .....................  ....................  ....................  ....................
14215 MW065065.1 |Severe acute respiratory syndrome coro    ....................  ....................  .....................  ....................  ....................  ....................
14216 MW065066.1 |Severe acute respiratory syndrome coro    ....................  ....................  .....................  ....................  ....................  ....................
14217 MW065067.1 |Severe acute respiratory syndrome coro    ....................  ....................  .....................  ....................  ....................  ....................
14218 MW065068.1 |Severe acute respiratory syndrome coro    ....................  ....................  .....................  ....................  ....................  ....................
14219 MW065069.1 |Severe acute respiratory syndrome coro    ....................  ....................  .....................  ....................  ....................  ....................
14220 MW065070.1 |Severe acute respiratory syndrome coro    ....................  ....................  .....................  ....................  ....................  ....................
14221 MW065071.1 |Severe acute respiratory syndrome coro    ....................  ....................  .....................  ....................  ....................  ....................
14222 MW065072.1 |Severe acute respiratory syndrome coro    ....................  ....................  .....................  ....................  ....................  ....................
14223 MW065073.1 |Severe acute respiratory syndrome coro    ....................  ....................  .....................  ....................  ....................  ....................
14224 MW065074.1 |Severe acute respiratory syndrome coro    ....................  ....................  .....................  ....................  ....................  ....................
14225 MW065076.1 |Severe acute respiratory syndrome coro    ....................  ....................  .....................  ....................  ....................  ....................
14226 MW065077.1 |Severe acute respiratory syndrome coro    ....................  ....................  .....................  ....................  ....................  ....................
14227 MW065078.1 |Severe acute respiratory syndrome coro    ....................  ....................  .....................  ....................  ....................  ....................
14228 MW065079.1 |Severe acute respiratory syndrome coro    ....................  ....................  .....................  ....................  ....................  ....................
14229 MW065080.1 |Severe acute respiratory syndrome coro    ....................  ....................  .....................  ....................  ....................  ....................
14230 MW065081.1 |Severe acute respiratory syndrome coro    ....................  ....................  .....................  ....................  ....................  ....................
14231 MW065082.1 |Severe acute respiratory syndrome coro    ....................  ....................  .....................  ....................  ....................  ....................
14232 MW065083.1 |Severe acute respiratory syndrome coro    ....................  ....................  .....................  ....................  ....................  ....................
14233 MW065084.1 |Severe acute respiratory syndrome coro    ....................  ....................  .....................  ....................  ....................  ....................
14234 MW065085.1 |Severe acute respiratory syndrome coro    ....................  ....................  .....................  ....................  ....................  ....................
14235 MW065086.1 |Severe acute respiratory syndrome coro    ....................  ....................  .....................  ....................  ....................  ....................
14236 MW065087.1 |Severe acute respiratory syndrome coro    ....................  ....................  .....................  ....................  ....................  ....................
14237 MW065088.1 |Severe acute respiratory syndrome coro    ....................  ....................  .....................  ....................  ....................  ....................
14238 MW065089.1 |Severe acute respiratory syndrome coro    ....................  ....................  .....................  ....................  ....................  ....................
14239 MW065090.1 |Severe acute respiratory syndrome coro    ....................  ....................  .....................  ....................  ....................  ....................
14240 MW065091.1 |Severe acute respiratory syndrome coro    ....................  ....................  .....................  ....................  ....................  ....................
14241 MW065092.1 |Severe acute respiratory syndrome coro    ....................  ....................  .....................  ....................  ....................  ....................
14242 MW065093.1 |Severe acute respiratory syndrome coro    ....................  ....................  .....................  ....................  ....................  ....................
```

14243 MW065094.1 |Severe acute respiratory syndrome coro .................... .................... .................... .................... .................... ....................
14244 MW065095.1 |Severe acute respiratory syndrome coro .................... .................... .................... .................... .................... ....................
14245 MW065096.1 |Severe acute respiratory syndrome coro .................... .................... .................... .................... .................... ....................
14246 MW065097.1 |Severe acute respiratory syndrome coro .................... .................... .................... .................... .................... ....................
14247 MW065098.1 |Severe acute respiratory syndrome coro .................... .................... .................... .................... .................... ....................
14248 MW065099.1 |Severe acute respiratory syndrome coro .................... .................... .................... .................... .................... ....................
14249 MW065100.1 |Severe acute respiratory syndrome coro .................... .................... .................... .................... .................... ....................
14250 MW065101.1 |Severe acute respiratory syndrome coro .................... .................... .................... .................... .................... ....................
14251 MW065103.1 |Severe acute respiratory syndrome coro .................... .................... .................... .................... .................... ....................
14252 MW065104.1 |Severe acute respiratory syndrome coro .................... .................... .................... .................... .................... ....................
14253 MW065106.1 |Severe acute respiratory syndrome coro .................... .................... .................... .................... .................... ....................
14254 MW065107.1 |Severe acute respiratory syndrome coro .................... .................... .................... .................... .................... ....................
14255 MW065108.1 |Severe acute respiratory syndrome coro .................... .................... .................... .................... .................... ....................
14256 MW065109.1 |Severe acute respiratory syndrome coro .................... .................... .................... .................... .................... ....................
14257 MW065110.1 |Severe acute respiratory syndrome coro .................... .................... .................... .................... .................... ....................
14258 MW065111.1 |Severe acute respiratory syndrome coro .................... .................... .................... .................... .................... ....................
14259 MW065112.1 |Severe acute respiratory syndrome coro .................... .................... .................... .................... .................... ....................
14260 MW065113.1 |Severe acute respiratory syndrome coro .................... .................... .................... .................... .................... ....................
14261 MW065114.1 |Severe acute respiratory syndrome coro .................... .................... .................... .................... .................... ....................
14262 MW065115.1 |Severe acute respiratory syndrome coro .................... .................... .................... .................... .................... ....................
14263 MW065116.1 |Severe acute respiratory syndrome coro .................... .................... .................... .................... .................... ....................
14264 MW065117.1 |Severe acute respiratory syndrome coro .................... .................... .................... .................... .................... ....................
14265 MW065118.1 |Severe acute respiratory syndrome coro .................... .................... .................... .................... .................... ....................
14266 MW065119.1 |Severe acute respiratory syndrome coro .................... .................... .................... .................... .................... ....................
14267 MW065120.1 |Severe acute respiratory syndrome coro .................... .................... .................... .................... .................... ....................
14268 MW065121.1 |Severe acute respiratory syndrome coro .................... .................... .................... .................... .................... ....................
14269 MW065122.1 |Severe acute respiratory syndrome coro .................... .................... .................... .................... .................... ....................
14270 MW065123.1 |Severe acute respiratory syndrome coro .................... .................... .................... .................... .................... ....................
14271 MW065124.1 |Severe acute respiratory syndrome coro .................... .................... .................... .................... .................... ....................
14272 MW065125.1 |Severe acute respiratory syndrome coro .................... .................... .................... .................... .................... ....................
14273 MW065126.1 |Severe acute respiratory syndrome coro .................... .................... .................... .................... .................... ....................
14274 MW065127.1 |Severe acute respiratory syndrome coro .................... .................... .................... .................... .................... ....................
14275 MW065128.1 |Severe acute respiratory syndrome coro .................... .................... .................... .................... .................... ....................
14276 MW065129.1 |Severe acute respiratory syndrome coro .................... .................... .................... .................... .................... ....................
14277 MW065130.1 |Severe acute respiratory syndrome coro .................... .................... .................... .................... .................... ....................
14278 MW065132.1 |Severe acute respiratory syndrome coro .................... .................... .................... .................... .................... ....................
14279 MW065133.1 |Severe acute respiratory syndrome coro .................... .................... .................... .................... .................... ....................
14280 MW065134.1 |Severe acute respiratory syndrome coro .................... .................... .................... .................... .................... ....................
14281 MW065135.1 |Severe acute respiratory syndrome coro .................... .................... .................... .................... .................... ....................
14282 MW065136.1 |Severe acute respiratory syndrome coro .................... .................... .................... .................... .................... ....................
14283 MW065137.1 |Severe acute respiratory syndrome coro .................... .................... .................... .................... .................... ....................
14284 MW065138.1 |Severe acute respiratory syndrome coro .................... .................... .................... .................... .................... ....................
14285 MW065139.1 |Severe acute respiratory syndrome coro .................... .................... .................... .................... .................... ....................
14286 MW065140.1 |Severe acute respiratory syndrome coro .................... .................... .................... .................... .................... ....................
14287 MW065141.1 |Severe acute respiratory syndrome coro .................... .................... .................... .................... .................... ....................
14288 MW065142.1 |Severe acute respiratory syndrome coro .................... .................... .................... .................... .................... ....................
14289 MW065143.1 |Severe acute respiratory syndrome coro .................... .................... .................... .................... .................... ....................
14290 MW065144.1 |Severe acute respiratory syndrome coro .................... .................... .................... .................... .................... ....................
14291 MW065146.1 |Severe acute respiratory syndrome coro .................... .................... .................... .................... .................... ....................
14292 MW065147.1 |Severe acute respiratory syndrome coro .................... .................... .................... .................... .................... ....................
14293 MW065148.1 |Severe acute respiratory syndrome coro .................... .................... .................... .................... .................... ....................
14294 MW065149.1 |Severe acute respiratory syndrome coro .................... .................... .................... .................... .................... ....................
14295 MW065150.1 |Severe acute respiratory syndrome coro .................... .................... .................... .................... .................... ....................
14296 MW065151.1 |Severe acute respiratory syndrome coro .................... .................... .................... .................... .................... ....................
14297 MW065152.1 |Severe acute respiratory syndrome coro .................... .................... .................... .................... .................... ....................
14298 MW065153.1 |Severe acute respiratory syndrome coro .................... .................... .................... .................... .................... ....................
14299 MW065154.1 |Severe acute respiratory syndrome coro .................... .................... .................... .................... .................... ....................
14300 MW065155.1 |Severe acute respiratory syndrome coro .................... .................... .................... .................... .................... ....................
14301 MW065156.1 |Severe acute respiratory syndrome coro .................... .................... .................... .................... .................... ....................
14302 MW065158.1 |Severe acute respiratory syndrome coro .................... .................... .................... .................... .................... ....................
14303 MW065160.1 |Severe acute respiratory syndrome coro .................... .................... .................... .................... .................... ....................
14304 MW065161.1 |Severe acute respiratory syndrome coro .................... .................... .................... .................... .................... ....................
14305 MW065162.1 |Severe acute respiratory syndrome coro .................... .................... .................... .................... .................... ....................
14306 MW065163.1 |Severe acute respiratory syndrome coro .................... .................... .................... .................... .................... ....................
14307 MW065164.1 |Severe acute respiratory syndrome coro .................... .................... .................... .................... .................... ....................
14308 MW065165.1 |Severe acute respiratory syndrome coro .................... .................... .................... .................... .................... ....................
14309 MW065166.1 |Severe acute respiratory syndrome coro .................... .................... .................... .................... .................... ....................
14310 MW065167.1 |Severe acute respiratory syndrome coro .................... .................... .................... .................... .................... ....................
14311 MW065168.1 |Severe acute respiratory syndrome coro .................... .................... .................... .................... .................... ....................
14312 MW065169.1 |Severe acute respiratory syndrome coro .................... .................... .................... .................... .................... ....................
14313 MW065170.1 |Severe acute respiratory syndrome coro .................... .................... .................... .................... .................... ....................
14314 MW065171.1 |Severe acute respiratory syndrome coro .................... .................... .................... .................... .................... ....................
14315 MW065172.1 |Severe acute respiratory syndrome coro .................... .................... .................... .................... .................... ....................
14316 MW065173.1 |Severe acute respiratory syndrome coro .................... .................... .................... .................... .................... ....................
14317 MW065174.1 |Severe acute respiratory syndrome coro .................... .................... .................... .................... .................... ....................
14318 MW065175.1 |Severe acute respiratory syndrome coro .................... .................... .................... .................... .................... ....................
14319 MW065176.1 |Severe acute respiratory syndrome coro .................... .................... .................... .................... .................... ....................
14320 MW065177.1 |Severe acute respiratory syndrome coro .................... .................... .................... .................... .................... ....................

| | | | | | | | | |
|---|---|---|---|---|---|---|---|---|
| 14321 | MW065178.1 | |Severe acute respiratory syndrome coro | .................... | .................... | .................... | .................... | .................... | .................... |
| 14322 | MW065179.1 | |Severe acute respiratory syndrome coro | .................... | .................... | .................... | .................... | .................... | .................... |
| 14323 | MW065180.1 | |Severe acute respiratory syndrome coro | .................... | .................... | .................... | .................... | .................... | .................... |
| 14324 | MW065181.1 | |Severe acute respiratory syndrome coro | .................... | .................... | .................... | .................... | .................... | .................... |
| 14325 | MW065182.1 | |Severe acute respiratory syndrome coro | .................... | .................... | .................... | .................... | .................... | .................... |
| 14326 | MW065183.1 | |Severe acute respiratory syndrome coro | .................... | .................... | .................... | .................... | .................... | .................... |
| 14327 | MW065184.1 | |Severe acute respiratory syndrome coro | .................... | .................... | .................... | .................... | .................... | .................... |
| 14328 | MW065185.1 | |Severe acute respiratory syndrome coro | .................... | .................... | .................... | .................... | .................... | .................... |
| 14329 | MW065186.1 | |Severe acute respiratory syndrome coro | .................... | .................... | .................... | .................... | .................... | .................... |
| 14330 | MW065187.1 | |Severe acute respiratory syndrome coro | .................... | .................... | .................... | .................... | .................... | .................... |
| 14331 | MW065188.1 | |Severe acute respiratory syndrome coro | .................... | .................... | .................... | .................... | .................... | .................... |
| 14332 | MW065189.1 | |Severe acute respiratory syndrome coro | .................... | .................... | .................... | .................... | .................... | .................... |
| 14333 | MW065190.1 | |Severe acute respiratory syndrome coro | .................... | .................... | .................... | .................... | .................... | .................... |
| 14334 | MW065191.1 | |Severe acute respiratory syndrome coro | .................... | .................... | .................... | .................... | .................... | .................... |
| 14335 | MW065192.1 | |Severe acute respiratory syndrome coro | .................... | .................... | .................... | .................... | .................... | .................... |
| 14336 | MW065193.1 | |Severe acute respiratory syndrome coro | .................... | .................... | .................... | .................... | .................... | .................... |
| 14337 | MW065194.1 | |Severe acute respiratory syndrome coro | .................... | .................... | .................... | .................... | .................... | .................... |
| 14338 | MW065195.1 | |Severe acute respiratory syndrome coro | .................... | .................... | .................... | .................... | .................... | .................... |
| 14339 | MW065196.1 | |Severe acute respiratory syndrome coro | .................... | .................... | .................... | .................... | .................... | .................... |
| 14340 | MW065197.1 | |Severe acute respiratory syndrome coro | .................... | .................... | .................... | .................... | .................... | .................... |
| 14341 | MW065198.1 | |Severe acute respiratory syndrome coro | .................... | .................... | .................... | .................... | .................... | .................... |
| 14342 | MW065200.1 | |Severe acute respiratory syndrome coro | .................... | .................... | .................... | .................... | .................... | .................... |
| 14343 | MW065201.1 | |Severe acute respiratory syndrome coro | .................... | .................... | .................... | .................... | .................... | .................... |
| 14344 | MW065203.1 | |Severe acute respiratory syndrome coro | .................... | .................... | .................... | .................... | .................... | .................... |
| 14345 | MW065204.1 | |Severe acute respiratory syndrome coro | .................... | .................... | .................... | .................... | .................... | .................... |
| 14346 | MW065205.1 | |Severe acute respiratory syndrome coro | .................... | .................... | .................... | .................... | .................... | .................... |
| 14347 | MW065206.1 | |Severe acute respiratory syndrome coro | .................... | .................... | .................... | .................... | .................... | .................... |
| 14348 | MW065207.1 | |Severe acute respiratory syndrome coro | .................... | .................... | .................... | .................... | .................... | .................... |
| 14349 | MW065208.1 | |Severe acute respiratory syndrome coro | .................... | .................... | .................... | .................... | .................... | .................... |
| 14350 | MW065210.1 | |Severe acute respiratory syndrome coro | .................... | .................... | .................... | .................... | .................... | .................... |
| 14351 | MW065211.1 | |Severe acute respiratory syndrome coro | .................... | .................... | .................... | .................... | .................... | .................... |
| 14352 | MW065213.1 | |Severe acute respiratory syndrome coro | .................... | .................... | .................... | .................... | .................... | .................... |
| 14353 | MW065214.1 | |Severe acute respiratory syndrome coro | .................... | .................... | .................... | .................... | .................... | .................... |
| 14354 | MW065215.1 | |Severe acute respiratory syndrome coro | .................... | .................... | .................... | .................... | .................... | .................... |
| 14355 | MW065216.1 | |Severe acute respiratory syndrome coro | .................... | .................... | .................... | .................... | .................... | .................... |
| 14356 | MW065217.1 | |Severe acute respiratory syndrome coro | .................... | .................... | .................... | .................... | .................... | .................... |
| 14357 | MW065218.1 | |Severe acute respiratory syndrome coro | .................... | .................... | .................... | .................... | .................... | .................... |
| 14358 | MW065219.1 | |Severe acute respiratory syndrome coro | .................... | .................... | .................... | .................... | .................... | .................... |
| 14359 | MW065220.1 | |Severe acute respiratory syndrome coro | .................... | .................... | .................... | .................... | .................... | .................... |
| 14360 | MW065221.1 | |Severe acute respiratory syndrome coro | .................... | .................... | .................... | .................... | .................... | .................... |
| 14361 | MW065222.1 | |Severe acute respiratory syndrome coro | .................... | .................... | .................... | .................... | .................... | .................... |
| 14362 | MW065223.1 | |Severe acute respiratory syndrome coro | .................... | ..........A......... | .................... | .................... | .................... | .................... |
| 14363 | MW065224.1 | |Severe acute respiratory syndrome coro | .................... | .................... | .................... | .................... | .................... | .................... |
| 14364 | MW065226.1 | |Severe acute respiratory syndrome coro | .................... | .................... | .................... | .................... | .................... | .................... |
| 14365 | MW065227.1 | |Severe acute respiratory syndrome coro | .................... | .................... | .................... | .................... | .................... | .................... |
| 14366 | MW065228.1 | |Severe acute respiratory syndrome coro | .................... | .................... | .................... | .................... | .................... | .................... |
| 14367 | MW065229.1 | |Severe acute respiratory syndrome coro | .................... | .................... | .................... | .................... | .................... | .................... |
| 14368 | MW065230.1 | |Severe acute respiratory syndrome coro | .................... | .................... | .................... | .................... | .................... | .................... |
| 14369 | MW065231.1 | |Severe acute respiratory syndrome coro | .................... | .................... | .................... | .................... | .................... | .................... |
| 14370 | MW065232.1 | |Severe acute respiratory syndrome coro | .................... | .................... | .................... | .................... | .................... | .................... |
| 14371 | MW065233.1 | |Severe acute respiratory syndrome coro | .................... | .................... | .................... | .................... | .................... | .................... |
| 14372 | MW065234.1 | |Severe acute respiratory syndrome coro | .................... | .................... | .................... | .................... | .................... | .................... |
| 14373 | MW065235.1 | |Severe acute respiratory syndrome coro | .................... | .................... | .................... | .................... | .................... | .................... |
| 14374 | MW065236.1 | |Severe acute respiratory syndrome coro | .................... | .................... | .................... | .................... | .................... | .................... |
| 14375 | MW065237.1 | |Severe acute respiratory syndrome coro | .................... | .................... | .................... | .................... | .................... | .................... |
| 14376 | MW065238.1 | |Severe acute respiratory syndrome coro | .................... | .................... | .................... | .................... | .................... | .................... |
| 14377 | MW065239.1 | |Severe acute respiratory syndrome coro | .................... | .................... | .................... | .................... | .................... | .................... |
| 14378 | MW065240.1 | |Severe acute respiratory syndrome coro | .................... | .................... | .................... | .................... | .................... | .................... |
| 14379 | MW065242.1 | |Severe acute respiratory syndrome coro | .................... | .................... | .................... | .................... | .................... | .................... |
| 14380 | MW065243.1 | |Severe acute respiratory syndrome coro | .................... | .................... | .................... | .................... | .................... | .................... |
| 14381 | MW065244.1 | |Severe acute respiratory syndrome coro | .................... | .................... | .................... | .................... | .................... | .................... |
| 14382 | MW065245.1 | |Severe acute respiratory syndrome coro | .................... | .................... | .................... | .................... | .................... | .................... |
| 14383 | MW065247.1 | |Severe acute respiratory syndrome coro | .................... | .................... | .................... | .................... | .................... | .................... |
| 14384 | MW065248.1 | |Severe acute respiratory syndrome coro | .................... | .................... | .................... | .................... | .................... | .................... |
| 14385 | MW065249.1 | |Severe acute respiratory syndrome coro | .................... | .................... | .................... | .................... | .................... | .................... |
| 14386 | MW065250.1 | |Severe acute respiratory syndrome coro | .................... | .................... | .................... | .................... | .................... | .................... |
| 14387 | MW065251.1 | |Severe acute respiratory syndrome coro | .................... | .................... | .................... | .................... | .................... | .................... |
| 14388 | MW065252.1 | |Severe acute respiratory syndrome coro | .................... | .................... | .................... | .................... | .................... | .................... |
| 14389 | MW065253.1 | |Severe acute respiratory syndrome coro | .................... | .................... | .................... | .................... | .................... | .................... |
| 14390 | MW065254.1 | |Severe acute respiratory syndrome coro | .................... | .................... | .................... | .................... | .................... | .................... |
| 14391 | MW065255.1 | |Severe acute respiratory syndrome coro | .................... | .................... | .................... | .................... | .................... | .................... |
| 14392 | MW065256.1 | |Severe acute respiratory syndrome coro | .................... | .................... | .................... | .................... | .................... | .................... |
| 14393 | MW065257.1 | |Severe acute respiratory syndrome coro | .................... | .................... | .................... | .................... | .................... | .................... |
| 14394 | MW065258.1 | |Severe acute respiratory syndrome coro | .................... | .................... | .................... | .................... | .................... | .................... |
| 14395 | MW065259.1 | |Severe acute respiratory syndrome coro | .................... | .................... | .................... | .................... | .................... | .................... |
| 14396 | MW065260.1 | |Severe acute respiratory syndrome coro | .................... | .................... | .................... | .................... | .................... | .................... |
| 14397 | MW065261.1 | |Severe acute respiratory syndrome coro | .................... | .................... | .................... | .................... | .................... | .................... |
| 14398 | MW065262.1 | |Severe acute respiratory syndrome coro | .................... | .................... | .................... | .................... | .................... | .................... |

| | | | | | | | | | |
|---|---|---|---|---|---|---|---|---|---|
| 14399 | MW065263.1 | \|Severe acute respiratory syndrome coro | .................... | .................... | .................... | .................... | .................... | .................... | |
| 14400 | MW065264.1 | \|Severe acute respiratory syndrome coro | .................... | .................... | .................... | .................... | .................... | .................... | |
| 14401 | MW065265.1 | \|Severe acute respiratory syndrome coro | .................... | .................... | .................... | .................... | .................... | .................... | |
| 14402 | MW065266.1 | \|Severe acute respiratory syndrome coro | .................... | .................... | .................... | .................... | .................... | .................... | |
| 14403 | MW065267.1 | \|Severe acute respiratory syndrome coro | .................... | .................... | .................... | .................... | .................... | .................... | |
| 14404 | MW065268.1 | \|Severe acute respiratory syndrome coro | .................... | .................... | .................... | .................... | .................... | .................... | |
| 14405 | MW065269.1 | \|Severe acute respiratory syndrome coro | .................... | .................... | .................... | .................... | .................... | .................... | |
| 14406 | MW065270.1 | \|Severe acute respiratory syndrome coro | .................... | .................... | .................... | .................... | .................... | .................... | |
| 14407 | MW065271.1 | \|Severe acute respiratory syndrome coro | .................... | .................... | .................... | .................... | .................... | .................... | |
| 14408 | MW065272.1 | \|Severe acute respiratory syndrome coro | .................... | .................... | .................... | .................... | .................... | .................... | |
| 14409 | MW065273.1 | \|Severe acute respiratory syndrome coro | .................... | .................... | .................... | .................... | .................... | .................... | |
| 14410 | MW065275.1 | \|Severe acute respiratory syndrome coro | .................... | .................... | .................... | .................... | .................... | .................... | |
| 14411 | MW065276.1 | \|Severe acute respiratory syndrome coro | .................... | .................... | .................... | .................... | .................... | .................... | |
| 14412 | MW065277.1 | \|Severe acute respiratory syndrome coro | .................... | .................... | .................... | .................... | .................... | .................... | |
| 14413 | MW065278.1 | \|Severe acute respiratory syndrome coro | .................... | .................... | .................... | .................... | .................... | .................... | |
| 14414 | MW065279.1 | \|Severe acute respiratory syndrome coro | .................... | .................... | .................... | .................... | .................... | .................... | |
| 14415 | MW065280.1 | \|Severe acute respiratory syndrome coro | .................... | .................... | .................... | .................... | .................... | .................... | |
| 14416 | MW065281.1 | \|Severe acute respiratory syndrome coro | .................... | .................... | .................... | .................... | .................... | .................... | |
| 14417 | MW065282.1 | \|Severe acute respiratory syndrome coro | .................... | .................... | .................... | .................... | .................... | .................... | |
| 14418 | MW065283.1 | \|Severe acute respiratory syndrome coro | .................... | .................... | .................... | .................... | .................... | .................... | |
| 14419 | MW065284.1 | \|Severe acute respiratory syndrome coro | .................... | .................... | .................... | .................... | .................... | .................... | |
| 14420 | MW065285.1 | \|Severe acute respiratory syndrome coro | .................... | .................... | .................... | .................... | .................... | .................... | |
| 14421 | MW065286.1 | \|Severe acute respiratory syndrome coro | .................... | .................... | .................... | .................... | .................... | .................... | |
| 14422 | MW065287.1 | \|Severe acute respiratory syndrome coro | .................... | .................... | .................... | .................... | .................... | .................... | |
| 14423 | MW065288.1 | \|Severe acute respiratory syndrome coro | .................... | .................... | .................... | .................... | .................... | .................... | |
| 14424 | MW065289.1 | \|Severe acute respiratory syndrome coro | .................... | .................... | .................... | .................... | .................... | .................... | |
| 14425 | MW065290.1 | \|Severe acute respiratory syndrome coro | .................... | .................... | .................... | .................... | .................... | .................... | |
| 14426 | MW065291.1 | \|Severe acute respiratory syndrome coro | .................... | .................... | .................... | .................... | .................... | .................... | |
| 14427 | MW065292.1 | \|Severe acute respiratory syndrome coro | .................... | .................... | .................... | .................... | .................... | .................... | |
| 14428 | MW065293.1 | \|Severe acute respiratory syndrome coro | .................... | .................... | .................... | .................... | .................... | .................... | |
| 14429 | MW065294.1 | \|Severe acute respiratory syndrome coro | .................... | .................... | .................... | .................... | .................... | .................... | |
| 14430 | MW065295.1 | \|Severe acute respiratory syndrome coro | .................... | .................... | .................... | .................... | .................... | .................... | |
| 14431 | MW065296.1 | \|Severe acute respiratory syndrome coro | .................... | .................... | .................... | .................... | .................... | .................... | |
| 14432 | MW065297.1 | \|Severe acute respiratory syndrome coro | .................... | ...................A | .................... | .................... | .................... | .................... | 1 |
| 14433 | MW065298.1 | \|Severe acute respiratory syndrome coro | .................... | .................... | .................... | .................... | .................... | .................... | |
| 14434 | MW065299.1 | \|Severe acute respiratory syndrome coro | .................... | .................... | .................... | .................... | .................... | .................... | |
| 14435 | MW065300.1 | \|Severe acute respiratory syndrome coro | .................... | .................... | .................... | .................... | .................... | .................... | |
| 14436 | MW065301.1 | \|Severe acute respiratory syndrome coro | .................... | .................... | .................... | .................... | .................... | .................... | |
| 14437 | MW065302.1 | \|Severe acute respiratory syndrome coro | .................... | .................... | .................... | .................... | .................... | .................... | |
| 14438 | MW065303.1 | \|Severe acute respiratory syndrome coro | .................... | .................... | .................... | .................... | .................... | .................... | |
| 14439 | MW065304.1 | \|Severe acute respiratory syndrome coro | .................... | .................... | .................... | .................... | .................... | .................... | |
| 14440 | MW065305.1 | \|Severe acute respiratory syndrome coro | .................... | .................... | .................... | .................... | .................... | .................... | |
| 14441 | MW065306.1 | \|Severe acute respiratory syndrome coro | .................... | .................... | .................... | .................... | .................... | .................... | |
| 14442 | MW065307.1 | \|Severe acute respiratory syndrome coro | .................... | .................... | .................... | .................... | .................... | .................... | |
| 14443 | MW065308.1 | \|Severe acute respiratory syndrome coro | .................... | .................... | .................... | .................... | .................... | .................... | |
| 14444 | MW065309.1 | \|Severe acute respiratory syndrome coro | .................... | .................... | .................... | .................... | .................... | .................... | |
| 14445 | MW065310.1 | \|Severe acute respiratory syndrome coro | .................... | .................... | .................... | .................... | .................... | .................... | |
| 14446 | MW065312.1 | \|Severe acute respiratory syndrome coro | .................... | .................... | .................... | .................... | .................... | .................... | |
| 14447 | MW065313.1 | \|Severe acute respiratory syndrome coro | .................... | .................... | .................... | .................... | .................... | .................... | |
| 14448 | MW065314.1 | \|Severe acute respiratory syndrome coro | .................... | .................... | .................... | .................... | .................... | .................... | |
| 14449 | MW065315.1 | \|Severe acute respiratory syndrome coro | .................... | .................... | .................... | .................... | .................... | .................... | |
| 14450 | MW065316.1 | \|Severe acute respiratory syndrome coro | .................... | .................... | .................... | .................... | .................... | .................... | |
| 14451 | MW065317.1 | \|Severe acute respiratory syndrome coro | .................... | .................... | .................... | .................... | .................... | .................... | |
| 14452 | MW065318.1 | \|Severe acute respiratory syndrome coro | .................... | .................... | .................... | .................... | .................... | .................... | |
| 14453 | MW065319.1 | \|Severe acute respiratory syndrome coro | .................... | .................... | .................... | .................... | .................... | .................... | |
| 14454 | MW065320.1 | \|Severe acute respiratory syndrome coro | .................... | .................... | .................... | .................... | .................... | .................... | |
| 14455 | MW065321.1 | \|Severe acute respiratory syndrome coro | .................... | .................... | .................... | .................... | .................... | .................... | |
| 14456 | MW065322.1 | \|Severe acute respiratory syndrome coro | .................... | .................... | .................... | .................... | .................... | .................... | |
| 14457 | MW065323.1 | \|Severe acute respiratory syndrome coro | .................... | .................... | .................... | .................... | .................... | .................... | |
| 14458 | MW065324.1 | \|Severe acute respiratory syndrome coro | .................... | .................... | .................... | .................... | .................... | .................... | |
| 14459 | MW065325.1 | \|Severe acute respiratory syndrome coro | .................... | .................... | .................... | .................... | .................... | .................... | |
| 14460 | MW065326.1 | \|Severe acute respiratory syndrome coro | .................... | .................... | .................... | .................... | .................... | .................... | |
| 14461 | MW065327.1 | \|Severe acute respiratory syndrome coro | .................... | .................... | .................... | .................... | .................... | .................... | |
| 14462 | MW065329.1 | \|Severe acute respiratory syndrome coro | .................... | .................... | .................... | .................... | .................... | .................... | |
| 14463 | MW065330.1 | \|Severe acute respiratory syndrome coro | .................... | .................... | .................... | .................... | .................... | .................... | |
| 14464 | MW065331.1 | \|Severe acute respiratory syndrome coro | .................... | .................... | .................... | .................... | .................... | .................... | |
| 14465 | MW065332.1 | \|Severe acute respiratory syndrome coro | .................... | .................... | .................... | .................... | .................... | .................... | |
| 14466 | MW065333.1 | \|Severe acute respiratory syndrome coro | .................... | .................... | .................... | .................... | .................... | .................... | |
| 14467 | MW065334.1 | \|Severe acute respiratory syndrome coro | .................... | .................... | .................... | .................... | .................... | .................... | |
| 14468 | MW065335.1 | \|Severe acute respiratory syndrome coro | .................... | .................... | .................... | .................... | .................... | .................... | |
| 14469 | MW065336.1 | \|Severe acute respiratory syndrome coro | .................... | .................... | .................... | .................... | .................... | .................... | |
| 14470 | MW065337.1 | \|Severe acute respiratory syndrome coro | .................... | .................... | .................... | .................... | .................... | .................... | |
| 14471 | MW065338.1 | \|Severe acute respiratory syndrome coro | .................... | .................... | .................... | .................... | .................... | .................... | |
| 14472 | MW065339.1 | \|Severe acute respiratory syndrome coro | .................... | .................... | .................... | .................... | .................... | .................... | |
| 14473 | MW065340.1 | \|Severe acute respiratory syndrome coro | .................... | .................... | .................... | .................... | .................... | .................... | |
| 14474 | MW065341.1 | \|Severe acute respiratory syndrome coro | .................... | .................... | .................... | .................... | .................... | .................... | |
| 14475 | MW065342.1 | \|Severe acute respiratory syndrome coro | .................... | .................... | .................... | .................... | .................... | .................... | |
| 14476 | MW065343.1 | \|Severe acute respiratory syndrome coro | .................... | .................... | .................... | .................... | .................... | .................... | |

```
14477 MW065344.1 |Severe acute respiratory syndrome coro    ....................  ....................  .....................  ....................  ....................  ....................
14478 MW065345.1 |Severe acute respiratory syndrome coro    ....................  ....................  .....................  ....................  ....................  ....................
14479 MW065346.1 |Severe acute respiratory syndrome coro    ....................  ....................  .....................  ....................  ....................  ....................
14480 MW065347.1 |Severe acute respiratory syndrome coro    ....................  ....................  .....................  ....................  ....................  ....................
14481 MW065348.1 |Severe acute respiratory syndrome coro    ....................  ....................  .....................  ....................  ....................  ....................
14482 MW065349.1 |Severe acute respiratory syndrome coro    ....................  ....................  .....................  ....................  ....................  ....................
14483 MW065350.1 |Severe acute respiratory syndrome coro    ....................  ....................  .....................  ....................  ....................  ....................
14484 MW065351.1 |Severe acute respiratory syndrome coro    ....................  ....................  .....................  ....................  ....................  ....................
14485 MW065352.1 |Severe acute respiratory syndrome coro    ....................  ....................  .....................  ....................  ....................  ....................
14486 MW065353.1 |Severe acute respiratory syndrome coro    ....................  ....................  .....................  ....................  ....................  ....................
14487 MW065354.1 |Severe acute respiratory syndrome coro    ....................  ....................  .....................  ....................  ....................  ....................
14488 MW065355.1 |Severe acute respiratory syndrome coro    ....................  ....................  .....................  ....................  ....................  ....................
14489 MW065357.1 |Severe acute respiratory syndrome coro    ....................  ....................  .....................  ....................  ....................  ....................
14490 MW065358.1 |Severe acute respiratory syndrome coro    ....................  ....................  .....................  ....................  ....................  ....................
14491 MW065359.1 |Severe acute respiratory syndrome coro    ....................  ....................  .....................  ....................  ....................  ....................
14492 MW065360.1 |Severe acute respiratory syndrome coro    ....................  ....................  .....................  ....................  ....................  ....................
14493 MW065361.1 |Severe acute respiratory syndrome coro    ....................  ....................  .....................  ....................  ....................  ....................
14494 MW065362.1 |Severe acute respiratory syndrome coro    ....................  ....................  .....................  ....................  ....................  ....................
14495 MW065363.1 |Severe acute respiratory syndrome coro    ....................  ....................  .....................  ....................  ....................  ....................
14496 MW065364.1 |Severe acute respiratory syndrome coro    ....................  ....................  .....................  ....................  ....................  ....................
14497 MW065365.1 |Severe acute respiratory syndrome coro    ....................  ....................  .....................  ....................  ....................  ....................
14498 MW065366.1 |Severe acute respiratory syndrome coro    ....................  ....................  .....................  ....................  ....................  ....................
14499 MW065367.1 |Severe acute respiratory syndrome coro    ....................  ....................  .....................  ....................  ....................  ....................
14500 MW065368.1 |Severe acute respiratory syndrome coro    ....................  ....................  .....................  ....................  ....................  ....................
14501 MW065369.1 |Severe acute respiratory syndrome coro    ....................  ....................  .....................  ....................  ....................  ....................
14502 MW065370.1 |Severe acute respiratory syndrome coro    ....................  ....................  .....................  ....................  ....................  ....................
14503 MW065371.1 |Severe acute respiratory syndrome coro    ....................  ....................  .....................  ....................  ....................  ....................
14504 MW065372.1 |Severe acute respiratory syndrome coro    ....................  ....................  .....................  ....................  ....................  ....................
14505 MW065373.1 |Severe acute respiratory syndrome coro    ....................  ....................  .....................  ....................  ....................  ....................
14506 MW065374.1 |Severe acute respiratory syndrome coro    ....................  ....................  .....................  ....................  ....................  ....................
14507 MW065375.1 |Severe acute respiratory syndrome coro    ....................  ....................  .....................  ....................  ....................  ....................
14508 MW065376.1 |Severe acute respiratory syndrome coro    ....................  ....................  .....................  ....................  ....................  ....................
14509 MW065377.1 |Severe acute respiratory syndrome coro    ....................  ....................  .....................  ....................  ....................  ....................
14510 MW065378.1 |Severe acute respiratory syndrome coro    ....................  ....................  .....................  ....................  ....................  ....................
14511 MW065379.1 |Severe acute respiratory syndrome coro    ....................  ....................  .....................  ....................  ....................  ....................
14512 MW065380.1 |Severe acute respiratory syndrome coro    ....................  ....................  .....................  ....................  ....................  ....................
14513 MW065381.1 |Severe acute respiratory syndrome coro    ....................  ....................  .....................  ....................  ....................  ....................
14514 MW065382.1 |Severe acute respiratory syndrome coro    ....................  ....................  .....................  ....................  ....................  ....................
14515 MW065383.1 |Severe acute respiratory syndrome coro    ....................  ....................  .....................  ....................  ....................  ....................
14516 MW065384.1 |Severe acute respiratory syndrome coro    ....................  ....................  .....................  ....................  ....................  ....................
14517 MW065385.1 |Severe acute respiratory syndrome coro    ....................  ....................  .....................  ....................  ....................  ....................
14518 MW065386.1 |Severe acute respiratory syndrome coro    ....................  ....................  .....................  ....................  ....................  ....................
14519 MW065387.1 |Severe acute respiratory syndrome coro    ....................  ....................  .....................  ....................  ....................  ....................
14520 MW065388.1 |Severe acute respiratory syndrome coro    ....................  ....................  .....................  ....................  ....................  ....................
14521 MW065389.1 |Severe acute respiratory syndrome coro    ....................  ....................  .....................  ....................  ....................  ....................
14522 MW065390.1 |Severe acute respiratory syndrome coro    ....................  ....................  .....................  ....................  ....................  ....................
14523 MW065391.1 |Severe acute respiratory syndrome coro    ....................  ....................  .....................  ....................  ....................  ....................
14524 MW065392.1 |Severe acute respiratory syndrome coro    ....................  ....................  .....................  ....................  ....................  ....................
14525 MW065393.1 |Severe acute respiratory syndrome coro    ....................  ....................  .....................  ....................  ....................  ....................
14526 MW065394.1 |Severe acute respiratory syndrome coro    ....................  ....................  .....................  ....................  ....................  ....................
14527 MW065395.1 |Severe acute respiratory syndrome coro    ....................  ....................  .....................  ....................  ....................  ....................
14528 MW065396.1 |Severe acute respiratory syndrome coro    ....................  ....................  .....................  ....................  ....................  ....................
14529 MW065397.1 |Severe acute respiratory syndrome coro    ....................  ....................  .....................  ....................  ....................  ....................
14530 MW065398.1 |Severe acute respiratory syndrome coro    ....................  ....................  .....................  ....................  ....................  ....................
14531 MW065399.1 |Severe acute respiratory syndrome coro    ....................  ....................  .....................  ....................  ....................  ....................
14532 MW065400.1 |Severe acute respiratory syndrome coro    ....................  ....................  .....................  ....................  ....................  ....................
14533 MW065401.1 |Severe acute respiratory syndrome coro    ....................  ....................  .....................  ....................  ....................  ....................
14534 MW065402.1 |Severe acute respiratory syndrome coro    ....................  ....................  .....................  ....................  ....................  ....................
14535 MW065403.1 |Severe acute respiratory syndrome coro    ....................  ....................  .....................  ....................  ....................  ....................
14536 MW065404.1 |Severe acute respiratory syndrome coro    ....................  ....................  .....................  ....................  ....................  ....................
14537 MW065405.1 |Severe acute respiratory syndrome coro    ....................  ....................  .....................  ....................  ....................  ....................
14538 MW065406.1 |Severe acute respiratory syndrome coro    ....................  ....................  .....................  ....................  ....................  ....................
14539 MW065407.1 |Severe acute respiratory syndrome coro    ....................  ....................  .....................  ....................  ....................  ....................
14540 MW065408.1 |Severe acute respiratory syndrome coro    ....................  ....................  .....................  ....................  ....................  ....................
14541 MW065409.1 |Severe acute respiratory syndrome coro    ....................  ....................  .....................  ....................  ....................  ....................
14542 MW065410.1 |Severe acute respiratory syndrome coro    ....................  ....................  .....................  ....................  ....................  ....................
14543 MW065411.1 |Severe acute respiratory syndrome coro    ....................  ....................  .....................  ....................  ....................  ....................
14544 MW065412.1 |Severe acute respiratory syndrome coro    ....................  ....................  .....................  ....................  ....................  ....................
14545 MW065413.1 |Severe acute respiratory syndrome coro    ....................  ....................  .....................  ....................  ....................  ....................
14546 MW065414.1 |Severe acute respiratory syndrome coro    ....................  ....................  .....................  ....................  ....................  ....................
14547 MW065415.1 |Severe acute respiratory syndrome coro    ....................  ....................  .....................  ....................  ....................  ....................
14548 MW065416.1 |Severe acute respiratory syndrome coro    ....................  ....................  .....................  ....................  ....................  ....................
14549 MW065417.1 |Severe acute respiratory syndrome coro    ....................  ....................  .....................  ....................  ....................  ....................
14550 MW065418.1 |Severe acute respiratory syndrome coro    ....................  ....................  .....................  ....................  ....................  ....................
14551 MW065419.1 |Severe acute respiratory syndrome coro    ....................  ....................  .....................  ....................  ....................  ....................
14552 MW065420.1 |Severe acute respiratory syndrome coro    ....................  ....................  .....................  ....................  ....................  ....................
14553 MW065421.1 |Severe acute respiratory syndrome coro    ....................  ....................  .....................  ....................  ....................  ....................
14554 MW065422.1 |Severe acute respiratory syndrome coro    ....................  ....................  .....................  ....................  ....................  ....................
```

14555 MW065424.1 |Severe acute respiratory syndrome coro ···················· ···················· ····················· ···················· ···················· ····················
14556 MW065425.1 |Severe acute respiratory syndrome coro ···················· ···················· ····················· ···················· ···················· ····················
14557 MW065426.1 |Severe acute respiratory syndrome coro ···················· ···················· ····················· ···················· ···················· ····················
14558 MW065427.1 |Severe acute respiratory syndrome coro ···················· ···················· ····················· ···················· ···················· ····················
14559 MW065428.1 |Severe acute respiratory syndrome coro ···················· ···················· ····················· ···················· ···················· ····················
14560 MW065429.1 |Severe acute respiratory syndrome coro ···················· ···················· ····················· ···················· ···················· ····················
14561 MW065430.1 |Severe acute respiratory syndrome coro ···················· ···················· ····················· ···················· ···················· ····················
14562 MW065431.1 |Severe acute respiratory syndrome coro ···················· ···················· ····················· ···················· ···················· ····················
14563 MW065432.1 |Severe acute respiratory syndrome coro ···················· ···················· ····················· ···················· ···················· ····················
14564 MW065433.1 |Severe acute respiratory syndrome coro ···················· ···················· ····················· ···················· ···················· ····················
14565 MW065434.1 |Severe acute respiratory syndrome coro ···················· ···················· ····················· ···················· ···················· ····················
14566 MW065435.1 |Severe acute respiratory syndrome coro ···················· ···················· ····················· ···················· ···················· ····················
14567 MW065436.1 |Severe acute respiratory syndrome coro ···················· ···················· ····················· ···················· ···················· ····················
14568 MW065437.1 |Severe acute respiratory syndrome coro ···················· ···················· ····················· ···················· ···················· ····················
14569 MW065438.1 |Severe acute respiratory syndrome coro ···················· ···················· ····················· ···················· ···················· ····················
14570 MW065439.1 |Severe acute respiratory syndrome coro ···················· ···················· ····················· ···················· ···················· ····················
14571 MW065440.1 |Severe acute respiratory syndrome coro ···················· ···················· ····················· ···················· ···················· ····················
14572 MW065441.1 |Severe acute respiratory syndrome coro ···················· ···················· ····················· ···················· ···················· ····················
14573 MW065442.1 |Severe acute respiratory syndrome coro ···················· ···················· ····················· ···················· ···················· ····················
14574 MW065443.1 |Severe acute respiratory syndrome coro ···················· ···················· ····················· ···················· ···················· ····················
14575 MW065444.1 |Severe acute respiratory syndrome coro ···················· ···················· ····················· ···················· ···················· ····················
14576 MW065445.1 |Severe acute respiratory syndrome coro ···················· ···················· ····················· ···················· ···················· ····················
14577 MW065446.1 |Severe acute respiratory syndrome coro ···················· ···················· ····················· ···················· ···················· ····················
14578 MW065447.1 |Severe acute respiratory syndrome coro ···················· ···················· ····················· ···················· ···················· ····················
14579 MW065448.1 |Severe acute respiratory syndrome coro ···················· ···················· ····················· ···················· ···················· ····················
14580 MW065449.1 |Severe acute respiratory syndrome coro ···················· ···················· ····················· ···················· ···················· ····················
14581 MW065450.1 |Severe acute respiratory syndrome coro ···················· ···················· ····················· ···················· ···················· ····················
14582 MW065451.1 |Severe acute respiratory syndrome coro ···················· ···················· ····················· ···················· ···················· ····················
14583 MW065452.1 |Severe acute respiratory syndrome coro ···················· ···················· ····················· ···················· ···················· ····················
14584 MW065453.1 |Severe acute respiratory syndrome coro ···················· ···················· ····················· ···················· ···················· ····················
14585 MW065454.1 |Severe acute respiratory syndrome coro ···················· ···················· ····················· ···················· ···················· ····················
14586 MW065455.1 |Severe acute respiratory syndrome coro ···················· ···················· ····················· ···················· ···················· ····················
14587 MW065456.1 |Severe acute respiratory syndrome coro ···················· ···················· ····················· ···················· ···················· ····················
14588 MW065457.1 |Severe acute respiratory syndrome coro ···················· ···················· ····················· ···················· ···················· ····················
14589 MW065458.1 |Severe acute respiratory syndrome coro ···················· ···················· ····················· ···················· ···················· ····················
14590 MW065459.1 |Severe acute respiratory syndrome coro ···················· ···················· ····················· ···················· ···················· ····················
14591 MW065460.1 |Severe acute respiratory syndrome coro ···················· ···················· ····················· ···················· ···················· ····················
14592 MW065461.1 |Severe acute respiratory syndrome coro ···················· ···················· ····················· ···················· ···················· ····················
14593 MW065462.1 |Severe acute respiratory syndrome coro ···················· ···················· ····················· ···················· ···················· ····················
14594 MW065463.1 |Severe acute respiratory syndrome coro ···················· ···················· ····················· ···················· ···················· ····················
14595 MW065464.1 |Severe acute respiratory syndrome coro ···················· ···················· ····················· ···················· ···················· ····················
14596 MW065465.1 |Severe acute respiratory syndrome coro ···················· ···················· ····················· ···················· ···················· ····················
14597 MW065466.1 |Severe acute respiratory syndrome coro ···················· ···················· ····················· ···················· ···················· ····················
14598 MW065467.1 |Severe acute respiratory syndrome coro ···················· ···················· ····················· ···················· ···················· ····················
14599 MW065468.1 |Severe acute respiratory syndrome coro ···················· ···················· ····················· ···················· ···················· ····················
14600 MW065469.1 |Severe acute respiratory syndrome coro ···················· ···················· ····················· ···················· ···················· ····················
14601 MW065470.1 |Severe acute respiratory syndrome coro ···················· ···················· ····················· ···················· ···················· ····················
14602 MW065471.1 |Severe acute respiratory syndrome coro ···················· ···················· ····················· ···················· ···················· ····················
14603 MW065472.1 |Severe acute respiratory syndrome coro ···················· ···················· ····················· ···················· ···················· ····················
14604 MW065473.1 |Severe acute respiratory syndrome coro ···················· ···················· ····················· ···················· ···················· ····················
14605 MW065474.1 |Severe acute respiratory syndrome coro ···················· ···················· ····················· ···················· ···················· ····················
14606 MW065475.1 |Severe acute respiratory syndrome coro ···················· ···················· ····················· ···················· ···················· ····················
14607 MW065476.1 |Severe acute respiratory syndrome coro ···················· ···················· ····················· ···················· ···················· ····················
14608 MW065477.1 |Severe acute respiratory syndrome coro ···················· ···················· ····················· ···················· ···················· ····················
14609 MW065478.1 |Severe acute respiratory syndrome coro ···················· ···················· ····················· ···················· ···················· ····················
14610 MW065479.1 |Severe acute respiratory syndrome coro ···················· ···················· ····················· ···················· ···················· ····················
14611 MW065480.1 |Severe acute respiratory syndrome coro ···················· ···················· ····················· ···················· ···················· ····················
14612 MW065481.1 |Severe acute respiratory syndrome coro ···················· ···················· ····················· ···················· ···················· ····················
14613 MW065482.1 |Severe acute respiratory syndrome coro ···················· ···················· ····················· ···················· ···················· ····················
14614 MW065483.1 |Severe acute respiratory syndrome coro ···················· ···················· ····················· ···················· ···················· ····················
14615 MW067675.1 |Severe acute respiratory syndrome coro ···················· ···················· ····················· ···················· ···················· ····················
14616 MW067676.1 |Severe acute respiratory syndrome coro ···················· ···················· ····················· ···················· ···················· ····················
14617 MW067677.1 |Severe acute respiratory syndrome coro ···················· ···················· ····················· ···················· ···················· ····················
14618 MW067678.1 |Severe acute respiratory syndrome coro ···················· ···················· ····················· ···················· ···················· ····················
14619 MW067679.1 |Severe acute respiratory syndrome coro ···················· ···················· ····················· ···················· ···················· ····················
14620 MW067680.1 |Severe acute respiratory syndrome coro ···················· ···················· ····················· ···················· ···················· ····················
14621 MW067681.1 |Severe acute respiratory syndrome coro ···················· ···················· ····················· ···················· ···················· ····················
14622 MW067682.1 |Severe acute respiratory syndrome coro ···················· ···················· ····················· ···················· ···················· ····················
14623 MW067684.1 |Severe acute respiratory syndrome coro ···················· ···················· ····················· ···················· ···················· ····················
14624 MW067685.1 |Severe acute respiratory syndrome coro ···················· ···················· ····················· ···················· ···················· ····················
14625 MW067686.1 |Severe acute respiratory syndrome coro ···················· ···················· ····················· ···················· ···················· ····················
14626 MW067687.1 |Severe acute respiratory syndrome coro ···················· ···················· ····················· ···················· ···················· ····················
14627 MW067688.1 |Severe acute respiratory syndrome coro ···················· ···················· ····················· ···················· ···················· ····················
14628 MW067689.1 |Severe acute respiratory syndrome coro ···················· ···················· ····················· ···················· ···················· ····················
14629 MW067690.1 |Severe acute respiratory syndrome coro ···················· ···················· ····················· ···················· ···················· ····················
14630 MW067691.1 |Severe acute respiratory syndrome coro ···················· ···················· ····················· ···················· ···················· ····················
14631 MW067692.1 |Severe acute respiratory syndrome coro ···················· ···················· ····················· ···················· ···················· ····················
14632 MW067693.1 |Severe acute respiratory syndrome coro ···················· ···················· ····················· ···················· ···················· ····················

| | | | | | | | |
|---|---|---|---|---|---|---|---|
| 14633 MW067694.1 | Severe acute respiratory syndrome coro | .................... | .................... | .................... | .................... | .................... | .................... |
| 14635 MW067696.1 | Severe acute respiratory syndrome coro | .................... | .................... | .................... | .................... | .................... | .................... |
| 14636 MW067697.1 | Severe acute respiratory syndrome coro | .................... | .................... | .................... | .................... | .................... | .................... |
| 14637 MW067698.1 | Severe acute respiratory syndrome coro | .................... | .................... | .................... | .................... | .................... | .................... |
| 14638 MW067699.1 | Severe acute respiratory syndrome coro | .................... | .................... | .................... | .................... | .................... | .................... |
| 14639 MW067700.1 | Severe acute respiratory syndrome coro | .................... | .................... | .................... | .................... | .................... | .................... |
| 14640 MW067701.1 | Severe acute respiratory syndrome coro | .................... | .................... | .................... | .................... | .................... | .................... |
| 14641 MW067702.1 | Severe acute respiratory syndrome coro | .................... | .................... | .................... | .................... | .................... | .................... |
| 14642 MW067703.1 | Severe acute respiratory syndrome coro | .................... | .................... | .................... | .................... | .................... | .................... |
| 14643 MW067704.1 | Severe acute respiratory syndrome coro | .................... | .................... | .................... | .................... | .................... | .................... |
| 14644 MW067705.1 | Severe acute respiratory syndrome coro | .................... | .................... | .................... | .................... | .................... | .................... |
| 14645 MW067706.1 | Severe acute respiratory syndrome coro | .................... | .................... | .................... | .................... | .................... | .................... |
| 14646 MW067707.1 | Severe acute respiratory syndrome coro | .................... | .................... | .................... | .................... | .................... | .................... |
| 14647 MW067708.1 | Severe acute respiratory syndrome coro | .................... | .................... | .................... | .................... | .................... | .................... |
| 14648 MW067709.1 | Severe acute respiratory syndrome coro | .................... | .................... | .................... | .................... | .................... | .................... |
| 14649 MW067710.1 | Severe acute respiratory syndrome coro | .................... | .................... | .................... | .................... | .................... | .................... |
| 14650 MW067711.1 | Severe acute respiratory syndrome coro | .................... | .................... | .................... | .................... | .................... | .................... |
| 14651 MW067712.1 | Severe acute respiratory syndrome coro | .................... | .................... | .................... | .................... | .................... | .................... |
| 14652 MW067713.1 | Severe acute respiratory syndrome coro | .................... | .................... | .................... | .................... | .................... | .................... |
| 14653 MW067714.1 | Severe acute respiratory syndrome coro | .................... | .................... | .................... | .................... | .................... | .................... |
| 14654 MW067715.1 | Severe acute respiratory syndrome coro | .................... | .................... | .................... | .................... | .................... | .................... |
| 14655 MW067716.1 | Severe acute respiratory syndrome coro | .................... | .................... | .................... | .................... | .................... | .................... |
| 14656 MW067717.1 | Severe acute respiratory syndrome coro | .................... | .................... | .................... | .................... | .................... | .................... |
| 14657 MW067718.1 | Severe acute respiratory syndrome coro | .................... | .................... | .................... | .................... | .................... | .................... |
| 14658 MW067719.1 | Severe acute respiratory syndrome coro | .................... | .................... | .................... | .................... | .................... | .................... |
| 14659 MW067720.1 | Severe acute respiratory syndrome coro | .................... | .................... | .................... | .................... | .................... | .................... |
| 14660 MW067721.1 | Severe acute respiratory syndrome coro | .................... | .................... | .................... | .................... | .................... | .................... |
| 14661 MW067722.1 | Severe acute respiratory syndrome coro | .................... | .................... | .................... | .................... | .................... | .................... |
| 14662 MW067723.1 | Severe acute respiratory syndrome coro | .................... | .................... | .................... | .................... | .................... | .................... |
| 14663 MW067724.1 | Severe acute respiratory syndrome coro | .................... | .................... | .................... | .................... | .................... | .................... |
| 14664 MW067725.1 | Severe acute respiratory syndrome coro | .................... | .................... | .................... | .................... | .................... | .................... |
| 14665 MW067726.1 | Severe acute respiratory syndrome coro | .................... | .................... | .................... | .................... | .................... | .................... |
| 14666 MW067727.1 | Severe acute respiratory syndrome coro | .................... | .................... | .................... | .................... | .................... | .................... |
| 14667 MW067728.1 | Severe acute respiratory syndrome coro | .................... | .................... | .................... | .................... | .................... | .................... |
| 14668 MW067729.1 | Severe acute respiratory syndrome coro | .................... | .................... | .................... | .................... | .................... | .................... |
| 14669 MW067730.1 | Severe acute respiratory syndrome coro | .................... | .................... | .................... | .................... | .................... | .................... |
| 14670 MW067731.1 | Severe acute respiratory syndrome coro | .................... | .................... | .................... | .................... | .................... | .................... |
| 14671 MW067732.1 | Severe acute respiratory syndrome coro | .................... | .................... | .................... | .................... | .................... | .................... |
| 14672 MW067733.1 | Severe acute respiratory syndrome coro | .................... | .A.................. | .................... | .................... | .................... | .................... |
| 14673 MW067734.1 | Severe acute respiratory syndrome coro | .................... | .................... | .................... | .................... | .................... | .........A.......... |
| 14674 MW067735.1 | Severe acute respiratory syndrome coro | .................... | .................... | .................... | .................... | .................... | .................... |
| 14675 MW067736.1 | Severe acute respiratory syndrome coro | .................... | .................... | .................... | .................... | .................... | .................... |
| 14676 MW067737.1 | Severe acute respiratory syndrome coro | .................... | .................... | .................... | .................... | .................... | .................... |
| 14677 MW067738.1 | Severe acute respiratory syndrome coro | .................... | .................... | .................... | .................... | .................... | .................... |
| 14678 MW067739.1 | Severe acute respiratory syndrome coro | .................... | .A.................. | .................... | .................... | .................... | .................... |
| 14679 MW067740.1 | Severe acute respiratory syndrome coro | .................... | .A.................. | .................... | .................... | .................... | .................... |
| 14680 MW067741.1 | Severe acute respiratory syndrome coro | .................... | .................... | .................... | .................... | .................... | .................... |
| 14681 MW067742.1 | Severe acute respiratory syndrome coro | .................... | .................... | .................... | .................... | .................... | .................... |
| 14682 MW067743.1 | Severe acute respiratory syndrome coro | .................... | .................... | .................... | .................... | .................... | .................... |
| 14683 MW067744.1 | Severe acute respiratory syndrome coro | .................... | .................... | .................... | .................... | .................... | .................... |
| 14684 MW067745.1 | Severe acute respiratory syndrome coro | .................... | .................... | .................... | .................... | .................... | .........A.......... |
| 14685 MW067746.1 | Severe acute respiratory syndrome coro | .................... | .................... | .................... | .................... | .................... | .................... |
| 14686 MW067747.1 | Severe acute respiratory syndrome coro | .................... | .................... | .................... | .................... | .................... | .................... |
| 14687 MW067748.1 | Severe acute respiratory syndrome coro | .................... | .................... | .................... | .................... | .................... | .................... |
| 14688 MW067749.1 | Severe acute respiratory syndrome coro | .................... | .A.................. | .................... | .................... | .................... | .................... |
| 14689 MW067751.1 | Severe acute respiratory syndrome coro | .................... | .................... | .................... | .................... | .................... | .................... |
| 14690 MW067752.1 | Severe acute respiratory syndrome coro | .................... | .A.................. | .................... | .................... | .................... | .................... |
| 14691 MW067753.1 | Severe acute respiratory syndrome coro | .................... | .................... | .................... | .................... | .................... | .................... |
| 14692 MW067757.1 | Severe acute respiratory syndrome coro | .................... | .................... | .................... | .................... | .................... | .................... |
| 14693 MW067758.1 | Severe acute respiratory syndrome coro | .................... | .................... | .................... | .................... | .................... | .................... |
| 14694 MW067759.1 | Severe acute respiratory syndrome coro | .................... | .................... | .................... | .................... | .................... | .................... |
| 14695 MW067760.1 | Severe acute respiratory syndrome coro | .................... | .................... | .................... | .................... | .................... | .................... |
| 14696 MW067761.1 | Severe acute respiratory syndrome coro | .................... | .................... | .................... | .................... | .................... | .................... |
| 14697 MW067762.1 | Severe acute respiratory syndrome coro | .................... | .................... | .................... | .................... | .................... | .................... |
| 14698 MW067763.1 | Severe acute respiratory syndrome coro | .................... | .................... | .................... | .................... | .................... | .................... |
| 14699 MW067764.1 | Severe acute respiratory syndrome coro | .................... | .................... | .................... | .................... | .................... | .................... |
| 14700 MW067765.1 | Severe acute respiratory syndrome coro | .................... | .................... | .................... | .................... | .................... | .........A.......... |
| 14701 MW067766.1 | Severe acute respiratory syndrome coro | .................... | .................... | .................... | .................... | .................... | .................... |
| 14702 MW067768.1 | Severe acute respiratory syndrome coro | .................... | .................... | .................... | .................... | .................... | .................... |
| 14703 MW067769.1 | Severe acute respiratory syndrome coro | .................... | .................... | .................... | .................... | .................... | .................... |
| 14704 MW067770.1 | Severe acute respiratory syndrome coro | .................... | .................... | .................... | .................... | .................... | .................... |
| 14705 MW067771.1 | Severe acute respiratory syndrome coro | .................... | .................... | .................... | .................... | .................... | .................... |
| 14706 MW067773.1 | Severe acute respiratory syndrome coro | .................... | .................... | .................... | .................... | .................... | .................... |
| 14707 MW067774.1 | Severe acute respiratory syndrome coro | .................... | .................... | .................... | .................... | .................... | .................... |
| 14708 MW067775.1 | Severe acute respiratory syndrome coro | .................... | .A.................. | .................... | .................... | .................... | .................... |
| 14709 MW067777.1 | Severe acute respiratory syndrome coro | .................... | .................... | .................... | .................... | .................... | .................... |
| 14710 MW067778.1 | Severe acute respiratory syndrome coro | .................... | .................... | .................... | .................... | .................... | .................... |
| 14711 MW067779.1 | Severe acute respiratory syndrome coro | .................... | .................... | .................... | .................... | .................... | .................... |

| | | | | | | | | |
|---|---|---|---|---|---|---|---|---|
| 14712 | MW067780.1 | \|Severe acute respiratory syndrome coro | .................... | .................... | .................... | .................... | .................... | .................... |
| 14713 | MW067781.1 | \|Severe acute respiratory syndrome coro | .................... | .................... | .................... | .................... | .................... | .................... |
| 14714 | MW067782.1 | \|Severe acute respiratory syndrome coro | .................... | .................... | .................... | .................... | .................... | .................... |
| 14715 | MW067783.1 | \|Severe acute respiratory syndrome coro | .................... | .................... | .................... | .................... | .................... | .................... |
| 14716 | MW067784.1 | \|Severe acute respiratory syndrome coro | .................... | .................... | .................... | .................... | .................... | .................... |
| 14717 | MW067785.1 | \|Severe acute respiratory syndrome coro | .................... | .................... | .................... | .................... | .................... | .................... |
| 14718 | MW067786.1 | \|Severe acute respiratory syndrome coro | .................... | .................... | .................... | .................... | .................... | .................... |
| 14719 | MW067787.1 | \|Severe acute respiratory syndrome coro | .................... | .................... | .................... | .................... | .................... | .................... |
| 14720 | MW067788.1 | \|Severe acute respiratory syndrome coro | .................... | .................... | .................... | .................... | .................... | .................... |
| 14721 | MW067789.1 | \|Severe acute respiratory syndrome coro | .................... | .................... | .................... | .................... | .................... | .................... |
| 14722 | MW067790.1 | \|Severe acute respiratory syndrome coro | .................... | .................... | .................... | .................... | .................... | .................... |
| 14723 | MW067791.1 | \|Severe acute respiratory syndrome coro | .................... | .................... | .................... | .................... | .................... | .................... |
| 14724 | MW067792.1 | \|Severe acute respiratory syndrome coro | .................... | .................... | .................... | .................... | .................... | .................... |
| 14725 | MW067793.1 | \|Severe acute respiratory syndrome coro | .................... | .................... | .................... | .................... | .................... | .................... |
| 14726 | MW067794.1 | \|Severe acute respiratory syndrome coro | .................... | .................... | .................... | .................... | .................... | .................... |
| 14727 | MW067795.1 | \|Severe acute respiratory syndrome coro | .................... | .................... | .................... | .................... | .................... | .................... |
| 14728 | MW067796.1 | \|Severe acute respiratory syndrome coro | .................... | .................... | .................... | .................... | .................... | .................... |
| 14729 | MW067797.1 | \|Severe acute respiratory syndrome coro | .................... | .................... | .................... | .................... | .................... | .................... |
| 14730 | MW067798.1 | \|Severe acute respiratory syndrome coro | .................... | .................... | .................... | .................... | .................... | .................... |
| 14731 | MW067799.1 | \|Severe acute respiratory syndrome coro | .................... | .................... | .................... | .................... | .................... | .................... |
| 14732 | MW067800.1 | \|Severe acute respiratory syndrome coro | .................... | .................... | .................... | .................... | .................... | .................... |
| 14733 | MW067802.1 | \|Severe acute respiratory syndrome coro | .................... | .................... | .................... | .................... | .................... | .................... |
| 14734 | MW067803.1 | \|Severe acute respiratory syndrome coro | .................... | .................... | .................... | .................... | .................... | .................... |
| 14735 | MW067804.1 | \|Severe acute respiratory syndrome coro | .................... | .................... | .................... | .................... | .................... | .................... |
| 14736 | MW067805.1 | \|Severe acute respiratory syndrome coro | .................... | .................... | .................... | .................... | .................... | .................... |
| 14737 | MW067806.1 | \|Severe acute respiratory syndrome coro | .................... | .................... | .................... | .................... | .................... | .................... |
| 14738 | MW067809.1 | \|Severe acute respiratory syndrome coro | .................... | .................... | .................... | .................... | .................... | .................... |
| 14739 | MW067810.1 | \|Severe acute respiratory syndrome coro | .................... | .................... | .................... | .................... | .................... | .................... |
| 14740 | MW067811.1 | \|Severe acute respiratory syndrome coro | .................... | .................... | .................... | .................... | .................... | .................... |
| 14741 | MW067812.1 | \|Severe acute respiratory syndrome coro | .................... | .................... | .................... | .................... | .................... | .................... |
| 14742 | MW067813.1 | \|Severe acute respiratory syndrome coro | .................... | .................... | .................... | .................... | .................... | .................... |
| 14743 | MW067814.1 | \|Severe acute respiratory syndrome coro | .................... | .................... | .................... | .................... | .................... | .................... |
| 14744 | MW067815.1 | \|Severe acute respiratory syndrome coro | .................... | .................... | .................... | .................... | .................... | .................... |
| 14745 | MW067816.1 | \|Severe acute respiratory syndrome coro | .................... | .................... | .................... | .................... | .................... | .................... |
| 14746 | MW067817.1 | \|Severe acute respiratory syndrome coro | .................... | .................... | .................... | .................... | .................... | .................... |
| 14747 | MW067818.1 | \|Severe acute respiratory syndrome coro | .................... | .................... | .................... | .................... | .................... | .................... |
| 14748 | MW067820.1 | \|Severe acute respiratory syndrome coro | .................... | .................... | .................... | .................... | .................... | .................... |
| 14749 | MW067821.1 | \|Severe acute respiratory syndrome coro | .................... | .................... | .................... | .................... | .................... | .................... |
| 14750 | MW067822.1 | \|Severe acute respiratory syndrome coro | .................... | .................... | .................... | .................... | .................... | .................... |
| 14751 | MW067823.1 | \|Severe acute respiratory syndrome coro | .................... | .................... | .................... | .................... | .................... | .................... |
| 14752 | MW067825.1 | \|Severe acute respiratory syndrome coro | .................... | .................... | .................... | .................... | .................... | .................... |
| 14753 | MW067826.1 | \|Severe acute respiratory syndrome coro | .................... | .................... | .................... | .................... | .................... | .................... |
| 14754 | MW067827.1 | \|Severe acute respiratory syndrome coro | .................... | .................... | .................... | .................... | .................... | .................... |
| 14755 | MW070033.1 | \|Severe acute respiratory syndrome coro | .................... | .................... | .................... | .................... | .................... | .................... |
| 14756 | MW070035.1 | \|Severe acute respiratory syndrome coro | .................... | .................... | .................... | .................... | .................... | .................... |
| 14757 | MW070036.1 | \|Severe acute respiratory syndrome coro | .................... | .................... | .................... | .................... | .................... | .................... |
| 14758 | MW070037.1 | \|Severe acute respiratory syndrome coro | .................... | .................... | .................... | .................... | .................... | .................... |
| 14759 | MW070038.1 | \|Severe acute respiratory syndrome coro | .................... | .................... | .................... | .................... | .................... | .................... |
| 14760 | MW070039.1 | \|Severe acute respiratory syndrome coro | .................... | .................... | .................... | .................... | .................... | .................... |
| 14761 | MW070040.1 | \|Severe acute respiratory syndrome coro | .................... | .................... | .................... | .................... | .................... | .................... |
| 14762 | MW070041.1 | \|Severe acute respiratory syndrome coro | .................... | .................... | .................... | .................... | .................... | .................... |
| 14763 | MW070042.1 | \|Severe acute respiratory syndrome coro | .................... | .................... | .................... | .................... | .................... | .................... |
| 14764 | MW070043.1 | \|Severe acute respiratory syndrome coro | .................... | .................... | .................... | .................... | .................... | .................... |
| 14765 | MW070044.1 | \|Severe acute respiratory syndrome coro | .................... | .................... | .................... | .................... | .................... | .................... |
| 14766 | MW070045.1 | \|Severe acute respiratory syndrome coro | .................... | .................... | .................... | .................... | .................... | .................... |
| 14767 | MW070046.1 | \|Severe acute respiratory syndrome coro | .................... | .................... | .................... | .................... | .................... | .................... |
| 14768 | MW070048.1 | \|Severe acute respiratory syndrome coro | .................... | .................... | .................... | .................... | .................... | .................... |
| 14769 | MW070049.1 | \|Severe acute respiratory syndrome coro | .................... | .................... | .................... | .................... | .................... | .................... |
| 14770 | MW070050.1 | \|Severe acute respiratory syndrome coro | .................... | .................... | .................... | .................... | .................... | .................... |
| 14771 | MW070051.1 | \|Severe acute respiratory syndrome coro | .................... | .................... | .................... | .................... | .................... | .................... |
| 14772 | MW070054.1 | \|Severe acute respiratory syndrome coro | .................... | .................... | .................... | .................... | ......T............. | .................... |
| 14773 | MW070057.1 | \|Severe acute respiratory syndrome coro | .................... | .................... | .................... | .................... | .................... | .................... |
| 14774 | MW070059.1 | \|Severe acute respiratory syndrome coro | .................... | .................... | .................... | .................... | .................... | .................... |
| 14775 | MW070060.1 | \|Severe acute respiratory syndrome coro | .................... | .................... | .................... | .................... | .................... | .................... |
| 14776 | MW070061.1 | \|Severe acute respiratory syndrome coro | .................... | .................... | .................... | .................... | .................... | .................... |
| 14777 | MW070062.1 | \|Severe acute respiratory syndrome coro | .................... | .................... | .................... | .................... | .................... | .................... |
| 14778 | MW070063.1 | \|Severe acute respiratory syndrome coro | .................... | .................... | .................... | .................... | .................... | .................... |
| 14779 | MW070064.1 | \|Severe acute respiratory syndrome coro | .................... | .................... | .................... | .................... | .................... | .................... |
| 14780 | MW070065.1 | \|Severe acute respiratory syndrome coro | .................... | .................... | .................... | .................... | .................... | .................... |
| 14781 | MW070066.1 | \|Severe acute respiratory syndrome coro | .................... | .................... | .................... | .................... | .................... | .................... |
| 14782 | MW070067.1 | \|Severe acute respiratory syndrome coro | .................... | .................... | .................... | .................... | .................... | .................... |
| 14783 | MW070068.1 | \|Severe acute respiratory syndrome coro | .................... | .................... | .................... | .................... | .................... | .................... |
| 14784 | MW070069.1 | \|Severe acute respiratory syndrome coro | .................... | .................... | .................... | .................... | .................... | .................... |
| 14785 | MW070070.1 | \|Severe acute respiratory syndrome coro | .................... | .................... | .................... | .................... | .................... | .................... |
| 14786 | MW070071.1 | \|Severe acute respiratory syndrome coro | .................... | .................... | .................... | .................... | .................... | .................... |
| 14787 | MW070072.1 | \|Severe acute respiratory syndrome coro | .................... | .................... | .................... | .................... | .................... | .................... |
| 14788 | MW070073.1 | \|Severe acute respiratory syndrome coro | .................... | .................... | .................... | .................... | .................... | .................... |
| 14789 | MW070074.1 | \|Severe acute respiratory syndrome coro | .................... | .................... | .................... | .................... | .................... | .................... |

```
14790 MW070075.1 |Severe acute respiratory syndrome coro    ....................  ....................  .....................  ....................  ....................  ....................
14791 MW070076.1 |Severe acute respiratory syndrome coro    ....................  ....................  .....................  ....................  ....................  ....................
14792 MW070077.1 |Severe acute respiratory syndrome coro    ....................  ....................  .....................  ....................  ....................  ....................
14793 MW070078.1 |Severe acute respiratory syndrome coro    ....................  ....................  .....................  ....................  ....................  ....................
14794 MW070079.1 |Severe acute respiratory syndrome coro    ....................  ....................  .....................  ....................  ....................  ....................
14795 MW070080.1 |Severe acute respiratory syndrome coro    ....................  ....................  .....................  ....................  ....................  ....................
14796 MW070081.1 |Severe acute respiratory syndrome coro    ....................  ...A................  .....................  ....................  ....................  ....................
14797 MW070082.1 |Severe acute respiratory syndrome coro    ....................  ....................  .....................  ....................  ....................  ....................
14798 MW070084.1 |Severe acute respiratory syndrome coro    ....................  ....................  .....................  ....................  ....................  ....................
14799 MW070085.1 |Severe acute respiratory syndrome coro    ....................  ....................  .....................  ....................  ....................  ....................
14800 MW070086.1 |Severe acute respiratory syndrome coro    ....................  ....................  .....................  ....................  ....................  ....................
14801 MW070087.1 |Severe acute respiratory syndrome coro    ....................  ....................  .....................  ....................  ....................  ....................
14802 MW070088.1 |Severe acute respiratory syndrome coro    ....................  ....................  .....................  ....................  ....................  ....................
14803 MW070089.1 |Severe acute respiratory syndrome coro    ....................  ....................  .....................  ....................  ....................  ....................
14804 MW070090.1 |Severe acute respiratory syndrome coro    ....................  ....................  .....................  ....................  ....................  ....................
14805 MW070091.1 |Severe acute respiratory syndrome coro    ....................  .A..................  .....................  ....................  ....................  ....................
14806 MW070092.1 |Severe acute respiratory syndrome coro    ....................  .A..................  .....................  ....................  ....................  ....................
14807 MW070093.1 |Severe acute respiratory syndrome coro    ....................  .A..................  .....................  ....................  ....................  ....................
14808 MW070096.1 |Severe acute respiratory syndrome coro    ....................  ....................  .....................  ....................  ....................  ....................
14809 MW070097.1 |Severe acute respiratory syndrome coro    ....................  ....................  .....................  ....................  ....................  ....................
14810 MW070098.1 |Severe acute respiratory syndrome coro    ....................  ....................  .....................  ....................  ....................  ....................
14811 MW070099.1 |Severe acute respiratory syndrome coro    ....................  ....................  .....................  ....................  ....................  ....................
14812 MW070101.1 |Severe acute respiratory syndrome coro    ....................  ....................  .....................  ....................  ....................  ....................
14813 MW070102.1 |Severe acute respiratory syndrome coro    ....................  ....................  .....................  ....................  ....................  ....................
14814 MW070103.1 |Severe acute respiratory syndrome coro    ....................  ....................  .....................  ....................  ....................  ....................
14815 MW070105.1 |Severe acute respiratory syndrome coro    ....................  ....................  .....................  ....................  ....................  ....................
14816 MW070106.1 |Severe acute respiratory syndrome coro    ....................  ....................  .....................  ....................  ....................  ....................
14817 MW070107.1 |Severe acute respiratory syndrome coro    ....................  ....................  .....................  ....................  ....................  ....................
14818 MW070108.1 |Severe acute respiratory syndrome coro    ....................  ....................  .....................  ....................  ....................  ....................
14819 MW070109.1 |Severe acute respiratory syndrome coro    ....................  ....................  .....................  ....................  ....................  ....................
14820 MW070110.1 |Severe acute respiratory syndrome coro    ....................  ....................  .....................  ....................  ....................  ....................
14821 MW070111.1 |Severe acute respiratory syndrome coro    ....................  ...A................  .....................  ....................  ....................  ....................
14822 MW070112.1 |Severe acute respiratory syndrome coro    ....................  ....................  .....................  ....................  ....................  ....................
14823 MW070113.1 |Severe acute respiratory syndrome coro    ....................  .A..................  .....................  ....................  ....................  ....................
14824 MW056032.1 |Severe acute respiratory syndrome coro    ....................  ....................  .....................  ....................  ....................  ....................
14825 MW056033.1 |Severe acute respiratory syndrome coro    ....................  ....................  .....................  ....................  ....................  ....................
14826 MW056034.1 |Severe acute respiratory syndrome coro    ....................  ....................  .....................  ....................  ....................  ....................
14827 MW056078.1 |Severe acute respiratory syndrome coro    ....................  ....................  .....................  ....................  ....................  ....................
14828 MW056086.1 |Severe acute respiratory syndrome coro    ....................  ...T................  .....................  ....................  N...................  ....................
14829 MW056092.1 |Severe acute respiratory syndrome coro    ....................  ....................  .........T...........  ....................  ....................  ....................
14830 MW056093.1 |Severe acute respiratory syndrome coro    ....................  ....................  .....................  ....................  ....................  ....................
14831 MW056097.1 |Severe acute respiratory syndrome coro    ....................  ....................  .....................  ....................  ....................  ....................
14832 MW056101.1 |Severe acute respiratory syndrome coro    ....................  ....................  .....................  ....................  ....................  ....................
14833 MW056102.1 |Severe acute respiratory syndrome coro    ....................  .............A......  .....................  ....................  ....................  ....................
14834 MW056104.1 |Severe acute respiratory syndrome coro    ....................  ....................  .....................  ....................  ....................  ....................
14835 MW056105.1 |Severe acute respiratory syndrome coro    ....................  ....................  .....................  ....................  ....................  ....................
14836 MW056107.1 |Severe acute respiratory syndrome coro    ....................  ....................  .....................  ....................  ....................  ....................
14837 MW056109.1 |Severe acute respiratory syndrome coro    ....................  ....................  .....................  ....................  ....................  ....................
14838 MW056110.1 |Severe acute respiratory syndrome coro    ....................  ....................  .....................  ....................  ....................  ....................
14839 MW056111.1 |Severe acute respiratory syndrome coro    ....................  ....................  .....................  ....................  ....................  ....................
14841 MW056114.1 |Severe acute respiratory syndrome coro    ....................  ....................  .....................  ....................  ....................  ....................
14842 MW056115.1 |Severe acute respiratory syndrome coro    ....................  ....................  .....................  ....................  ....................  ....................
14844 MW056117.1 |Severe acute respiratory syndrome coro    ....................  ....................  .....................  ....................  ....................  ....................
14845 MW056118.1 |Severe acute respiratory syndrome coro    ....................  ....................  .....................  ....................  ....................  ....................
14846 MW056122.1 |Severe acute respiratory syndrome coro    ....................  ....................  .....................  ....................  ....................  ....................
14847 MW056124.1 |Severe acute respiratory syndrome coro    ....................  ....................  .....................  ....................  ....................  ....................
14848 MW056125.1 |Severe acute respiratory syndrome coro    ....................  ....................  .....................  ....................  ....................  ....................
14849 MW056126.1 |Severe acute respiratory syndrome coro    ....................  ....................  .....................  ....................  ....................  ....................
14850 MW056127.1 |Severe acute respiratory syndrome coro    ....................  .............A......  .....................  ....................  ....................  ....................
14851 MW056128.1 |Severe acute respiratory syndrome coro    ....................  ....................  .....................  ....................  ....................  ....................
14852 MW056131.1 |Severe acute respiratory syndrome coro    ....................  ....................  .....................  ....................  ....................  ....................
14854 MW056136.1 |Severe acute respiratory syndrome coro    ....................  ....................  .....................  ....................  ....................  ....................
14855 MW056137.1 |Severe acute respiratory syndrome coro    ....................  .............A......  .....................  ....................  ....................  ....................
14856 MW056138.1 |Severe acute respiratory syndrome coro    ....................  ....................  .....................  ....................  ....................  ....................
14857 MW056141.1 |Severe acute respiratory syndrome coro    ....................  ....................  .....................  ....................  ....................  ....................
14858 MW056146.1 |Severe acute respiratory syndrome coro    ....................  ....................  .....................  ....................  ....................  ....................
14859 MW056147.1 |Severe acute respiratory syndrome coro    ....................  ....................  .....................  ....................  ....................  ....................
14860 MW056149.1 |Severe acute respiratory syndrome coro    ....................  ....................  .....................  ....................  ....................  ....................
14861 MW056151.1 |Severe acute respiratory syndrome coro    ....................  ....................  .....................  ....................  ....................  ....................
14862 MW056152.1 |Severe acute respiratory syndrome coro    ....................  ....................  .....................  ....................  ....................  ....................
14863 MW056155.1 |Severe acute respiratory syndrome coro    ....................  .............A......  .....................  ....................  ....................  ....................
14864 MW056156.1 |Severe acute respiratory syndrome coro    ....................  ....................  .....................  ....................  ....................  ....................
14865 MW056159.1 |Severe acute respiratory syndrome coro    ....................  ....................  .....................  ....................  ....................  ....................
14866 MW056161.1 |Severe acute respiratory syndrome coro    ....................  ....................  .....................  ....................  ....................  ....................
14867 MW056166.1 |Severe acute respiratory syndrome coro    ....................  ....................  .....................  ....................  ....................  ....................
14868 MW056167.1 |Severe acute respiratory syndrome coro    ....................  ....................  .....................  ....................  ....................  ....................
14869 MW056168.1 |Severe acute respiratory syndrome coro    ....................  ...A................  .....................  ....................  ....................  ....................
14870 MW056169.1 |Severe acute respiratory syndrome coro    ....................  ....................  .....................  ....................  ....................  ....................
```

14871 MW056170.1 |Severe acute respiratory syndrome coro ···················· ···················· ···················· ···················· ···················· ····················
14873 MW056172.1 |Severe acute respiratory syndrome coro ···················· ···················· ···················· ···················· ···················· ····················
14874 MW056173.1 |Severe acute respiratory syndrome coro ···················· ···················· ···················· ···················· ···················· ····················
14875 MW056174.1 |Severe acute respiratory syndrome coro ···················· ···················· ···················· ···················· ···················· ····················
14876 MW056175.1 |Severe acute respiratory syndrome coro ···················· ···················· ···················· ···················· ···················· ····················
14877 MW056176.1 |Severe acute respiratory syndrome coro ···················· ···················· ···················· ···················· ···················· ····················
14878 MW056177.1 |Severe acute respiratory syndrome coro ···················· ·A·················· ···················· ···················· ···················· ····················
14879 MW052620.1 |Severe acute respiratory syndrome coro ···················· ···················· ···················· ···················· ···················· ····················
14880 MW052624.1 |Severe acute respiratory syndrome coro ···················· ···················· ···················· ···················· ···················· ····················
14881 MW052625.1 |Severe acute respiratory syndrome coro ···················· ···················· ···················· ···················· ···················· ····················
14882 MW052629.1 |Severe acute respiratory syndrome coro ···················· ·A·················· ···················· ···················· ···················· ····················
14883 MW052631.1 |Severe acute respiratory syndrome coro ···················· ···················· ···················· ···················· ···················· ····················
14884 MW052632.1 |Severe acute respiratory syndrome coro ···················· ···················· ···················· ···················· ···················· ····················
14885 MW052633.1 |Severe acute respiratory syndrome coro ···················· ···················· ···················· ···················· ···················· ····················
14886 MW052634.1 |Severe acute respiratory syndrome coro ···················· ···G················ ···················· ···················· ···················· ····················
14887 MW052635.1 |Severe acute respiratory syndrome coro ···················· ···G················ ···················· ···················· ···················· ····················
14888 MW052636.1 |Severe acute respiratory syndrome coro ···················· ···················· ···················· ···················· ···················· ····················
14889 MW052637.1 |Severe acute respiratory syndrome coro ···················· ···················· ···················· ···················· ···················· ····················
14890 MW052638.1 |Severe acute respiratory syndrome coro ···················· ···················· ···················· ···················· ···················· ····················
14891 MW052639.1 |Severe acute respiratory syndrome coro ···················· ···················· ···················· ···················· ···················· ····················
14892 MW052640.1 |Severe acute respiratory syndrome coro ···················· ···················· ···················· ···················· ···················· ····················
14893 MW052641.1 |Severe acute respiratory syndrome coro ···················· ···················· ···················· ···················· ···················· ····················
14894 MW052642.1 |Severe acute respiratory syndrome coro ···················· ···················· ···················· ···················· ···················· ····················
14895 MW052643.1 |Severe acute respiratory syndrome coro ···················· ···················· ···················· ···················· ···················· ····················
14896 MW052644.1 |Severe acute respiratory syndrome coro ···················· ···················· ···················· ···················· ···················· ····················
14897 MW052645.1 |Severe acute respiratory syndrome coro ···················· ···················· ···················· ···················· ···················· ····················
14898 MW052646.1 |Severe acute respiratory syndrome coro ···················· ···················· ···················· ···················· ···················· ····················
14899 MW052647.1 |Severe acute respiratory syndrome coro ···················· ···················· ···················· ···················· ···················· ····················
14900 MW052648.1 |Severe acute respiratory syndrome coro ···················· ···················· ···················· ···················· ···················· ····················
14901 MW052649.1 |Severe acute respiratory syndrome coro ···················· ···G················ ···················· ···················· ···················· ····················
14902 MW052650.1 |Severe acute respiratory syndrome coro ···················· ···················· ···················· ···················· ···················· ····················
14903 MW052651.1 |Severe acute respiratory syndrome coro ···················· ···················· ···················· ···················· ···················· ····················
14904 MW052652.1 |Severe acute respiratory syndrome coro ···················· ···················· ···················· ···················· ···················· ····················
14905 MW052653.1 |Severe acute respiratory syndrome coro ···················· ···················· ···················· ···················· ···················· ····················
14906 MW052654.1 |Severe acute respiratory syndrome coro ···················· ···················· ···················· ···················· ···················· ····················
14907 MW052655.1 |Severe acute respiratory syndrome coro ···················· ···················· ···················· ···················· ···················· ····················
14908 MW052656.1 |Severe acute respiratory syndrome coro ···················· ···················· ···················· ···················· ···················· ····················
14909 MW052657.1 |Severe acute respiratory syndrome coro ···················· ···················· ···················· ···················· ···················· ····················
14910 MW052658.1 |Severe acute respiratory syndrome coro ···················· ···················· ···················· ···················· ···················· ····················
14911 MW052659.1 |Severe acute respiratory syndrome coro ···················· ···················· ···················· ···················· ···················· ····················
14912 MW052660.1 |Severe acute respiratory syndrome coro ···················· ···················· ···················· ···················· ···················· ····················
14913 MW052661.1 |Severe acute respiratory syndrome coro ···················· ···················· ···················· ···················· ···················· ····················
14914 MW052662.1 |Severe acute respiratory syndrome coro ···················· ···················· ···················· ···················· ···················· ····················
14915 MW052663.1 |Severe acute respiratory syndrome coro ···················· ···················· ···················· ···················· ···················· ····················
14916 MW052664.1 |Severe acute respiratory syndrome coro ···················· ···················· ···················· ···················· ···················· ····················
14917 MW052665.1 |Severe acute respiratory syndrome coro ···················· ···················· ···················· ···················· ···················· ····················
14918 MW052666.1 |Severe acute respiratory syndrome coro ···················· ···················· ···················· ···················· ···················· ····················
14919 MW052667.1 |Severe acute respiratory syndrome coro ···················· ···················· ···················· ···················· ···················· ····················
14920 MW052668.1 |Severe acute respiratory syndrome coro ···················· ···················· ···················· ···················· ···················· ····················
14921 MW052669.1 |Severe acute respiratory syndrome coro ···················· ···················· ···················· ···················· ···················· ····················
14922 MW052670.1 |Severe acute respiratory syndrome coro ···················· ···················· ···················· ···················· ···················· ····················
14923 MW052671.1 |Severe acute respiratory syndrome coro ···················· ···················· ···················· ···················· ···················· ····················
14924 MW052672.1 |Severe acute respiratory syndrome coro ···················· ···················· ···················· ···················· ···················· ····················
14925 MW052673.1 |Severe acute respiratory syndrome coro ···················· ···················· ···················· ···················· ···················· ····················
14926 MW052674.1 |Severe acute respiratory syndrome coro ···················· ···················· ···················· ···················· ···················· ····················
14927 MW052675.1 |Severe acute respiratory syndrome coro ···················· ···················· ···················· ···················· ···················· ····················
14928 MW052676.1 |Severe acute respiratory syndrome coro ···················· ···················· ···················· ···················· ···················· ····················
14929 MW052677.1 |Severe acute respiratory syndrome coro ···················· ···················· ···················· ···················· ···················· ····················
14930 MW052678.1 |Severe acute respiratory syndrome coro ···················· ···G················ ···················· ···················· ···················· ····················
14931 MW052679.1 |Severe acute respiratory syndrome coro ···················· ···················· ···················· ···················· ···················· ····················
14932 MW052680.1 |Severe acute respiratory syndrome coro ········T··········· ···················· ···················· ···················· ···················· ····················
14933 MW052681.1 |Severe acute respiratory syndrome coro ···················· ···················· ···················· ···················· ···················· ····················
14934 MW052682.1 |Severe acute respiratory syndrome coro ···················· ···················· ···················· ···················· ···················· ····················
14935 MW052683.1 |Severe acute respiratory syndrome coro ···················· ···················· ···················· ···················· ···················· ····················
14936 MW052684.1 |Severe acute respiratory syndrome coro ···················· ···················· ···················· ···················· ···················· ····················
14937 MW052685.1 |Severe acute respiratory syndrome coro ···················· ···················· ···················· ···················· ···················· ····················
14938 MW052686.1 |Severe acute respiratory syndrome coro ···················· ···················· ···················· ···················· ···················· ····················
14939 MW052687.1 |Severe acute respiratory syndrome coro ···················· ···················· ···················· ···················· ···················· ····················
14940 MW052688.1 |Severe acute respiratory syndrome coro ···················· ···················· ···················· ···················· ···················· ····················
14941 MW052689.1 |Severe acute respiratory syndrome coro ···················· ···················· ···················· ···················· ···················· ····················
14942 MW052690.1 |Severe acute respiratory syndrome coro ···················· ···················· ···················· ···················· ···················· ····················
14943 MW052691.1 |Severe acute respiratory syndrome coro ···················· ···················· ···················· ···················· ···················· ····················
14944 MW052692.1 |Severe acute respiratory syndrome coro ···················· ·A·················· ···················· ···················· ···················· ····················
14945 MW052693.1 |Severe acute respiratory syndrome coro ···················· ·A·················· ···················· ···················· ···················· ····················
14946 MW053297.1 |Severe acute respiratory syndrome coro ···················· ···A················ ···················· ···················· ···················· ····················
14947 MW053298.1 |Severe acute respiratory syndrome coro ···················· ···A················ ···················· ···················· ···················· ····················
14948 MW053299.1 |Severe acute respiratory syndrome coro ···················· ···A················ ···················· ···················· ···················· ····················
14949 MW053300.1 |Severe acute respiratory syndrome coro ···················· ···A················ ···················· ···················· ···················· ····················

| | | | | | | | | |
|---|---|---|---|---|---|---|---|---|
| 14950 | MW053301.1 | \|Severe acute respiratory syndrome coro | ···················· | ····A··············· | ···················· | ···················· | ···················· | ···················· |
| 14951 | MW053302.1 | \|Severe acute respiratory syndrome coro | ···················· | ····A··············· | ···················· | ···················· | ···················· | ···················· |
| 14952 | MW053303.1 | \|Severe acute respiratory syndrome coro | ···················· | ····A··············· | ···················· | ···················· | ···················· | ···················· |
| 14953 | MW053304.1 | \|Severe acute respiratory syndrome coro | ···················· | ····A··············· | ···················· | ···················· | ···················· | ···················· |
| 14954 | MW053305.1 | \|Severe acute respiratory syndrome coro | ···················· | ····A··············· | ···················· | ···················· | ···················· | ···················· |
| 14955 | MW053306.1 | \|Severe acute respiratory syndrome coro | ···················· | ····A··············· | ···················· | ···················· | ···················· | ···················· |
| 14956 | MW053307.1 | \|Severe acute respiratory syndrome coro | ···················· | ····A··············· | ···················· | ···················· | ···················· | ···················· |
| 14957 | MW053308.1 | \|Severe acute respiratory syndrome coro | ···················· | ····A··············· | ···················· | ···················· | ···················· | ···················· |
| 14958 | MW053309.1 | \|Severe acute respiratory syndrome coro | ···················· | ····A··············· | ···················· | ···················· | ···················· | ···················· |
| 14959 | MW053310.1 | \|Severe acute respiratory syndrome coro | ···················· | ····A··············· | ···················· | ···················· | ···················· | ···················· |
| 14960 | MW053311.1 | \|Severe acute respiratory syndrome coro | ···················· | ····A··············· | ···················· | ···················· | ···················· | ···················· |
| 14961 | MW053312.1 | \|Severe acute respiratory syndrome coro | ···················· | ····A··············· | ···················· | ···················· | ···················· | ···················· |
| 14962 | MW053313.1 | \|Severe acute respiratory syndrome coro | ···················· | ····A··············· | ···················· | ···················· | ···················· | ···················· |
| 14963 | MW053314.1 | \|Severe acute respiratory syndrome coro | ···················· | ····A··············· | ···················· | ···················· | ···················· | ···················· |
| 14964 | MW053315.1 | \|Severe acute respiratory syndrome coro | ···················· | ····A··············· | ···················· | ···················· | ···················· | ···················· |
| 14965 | MW053316.1 | \|Severe acute respiratory syndrome coro | ···················· | ····A··············· | ···················· | ···················· | ···················· | ···················· |
| 14966 | MW053317.1 | \|Severe acute respiratory syndrome coro | ···················· | ····A··············· | ···················· | ···················· | ···················· | ···················· |
| 14967 | MW053318.1 | \|Severe acute respiratory syndrome coro | ···················· | ····A··············· | ···················· | ···················· | ···················· | ···················· |
| 14968 | MW053319.1 | \|Severe acute respiratory syndrome coro | ···················· | ····A··············· | ···················· | ···················· | ···················· | ···················· |
| 14969 | MW053708.1 | \|Severe acute respiratory syndrome coro | ···················· | ···················· | ···················· | ···················· | ···················· | ···················· |
| 14970 | MW053750.1 | \|Severe acute respiratory syndrome coro | ···················· | ···················· | ···················· | ···················· | ···················· | ···················· |
| 14971 | MW053767.1 | \|Severe acute respiratory syndrome coro | ···················· | ···················· | ···················· | ···················· | ···················· | ···················· |
| 14972 | MW053869.1 | \|Severe acute respiratory syndrome coro | ···················· | ···················· | ···················· | ···················· | ···················· | ···················· |
| 14973 | MW053874.1 | \|Severe acute respiratory syndrome coro | ···················· | ···················· | ···················· | ···················· | ···················· | ···················· |
| 14974 | MW053875.1 | \|Severe acute respiratory syndrome coro | ···················· | ···················· | ···················· | ···················· | ···················· | ···················· |
| 14975 | MW053876.1 | \|Severe acute respiratory syndrome coro | ···················· | ···················· | ···················· | ···················· | ···················· | ···················· |
| 14976 | MW053883.1 | \|Severe acute respiratory syndrome coro | ···················· | ···················· | ···················· | ···················· | ···················· | ···················· |
| 14977 | MW053885.1 | \|Severe acute respiratory syndrome coro | ···················· | ···················· | ···················· | ···················· | ···················· | ···················· |
| 14978 | MW053921.1 | \|Severe acute respiratory syndrome coro | ···················· | ···················· | ···················· | ···················· | ···················· | ···················· |
| 14979 | MW053925.1 | \|Severe acute respiratory syndrome coro | ···················· | ···················· | ···················· | ···················· | ···················· | ···················· |
| 14980 | MW053929.1 | \|Severe acute respiratory syndrome coro | ···················· | ···················· | ···················· | ···················· | ···················· | ···················· |
| 14981 | MW053935.1 | \|Severe acute respiratory syndrome coro | ···················· | ···················· | ···················· | ···················· | ···················· | ···················· |
| 14982 | MW053938.1 | \|Severe acute respiratory syndrome coro | ···················· | ···················· | ···················· | ···················· | ···················· | ···················· |
| 14983 | MW053950.1 | \|Severe acute respiratory syndrome coro | ···················· | ···················· | ···················· | ···················· | ···················· | ···················· |
| 14984 | MW053957.1 | \|Severe acute respiratory syndrome coro | ···················· | ···················· | ···················· | ···················· | ···················· | ···················· |
| 14985 | MW053961.1 | \|Severe acute respiratory syndrome coro | ···················· | ···················· | ···················· | ···················· | ···················· | ···················· |
| 14986 | MW053976.1 | \|Severe acute respiratory syndrome coro | ···················· | ···················· | ···················· | ···················· | ···················· | ···················· |
| 14987 | MW053980.1 | \|Severe acute respiratory syndrome coro | ···················· | ···················· | ···················· | ···················· | ···················· | ···················· |
| 14988 | MW053981.1 | \|Severe acute respiratory syndrome coro | ···················· | ···················· | ···················· | ···················· | ···················· | ···················· |
| 14989 | MW054091.1 | \|Severe acute respiratory syndrome coro | ···················· | ···················· | ···················· | ···················· | ···················· | ···················· |
| 14990 | MW054103.1 | \|Severe acute respiratory syndrome coro | ···················· | ···················· | ···················· | ···················· | ···················· | ···················· |
| 14991 | MW054108.1 | \|Severe acute respiratory syndrome coro | ···················· | ···················· | ···················· | ···················· | ···················· | ···················· |
| 14992 | MW054109.1 | \|Severe acute respiratory syndrome coro | ···················· | ···················· | ···················· | ···················· | ···················· | ···················· |
| 14993 | MW054137.1 | \|Severe acute respiratory syndrome coro | ···················· | ···················· | ···················· | ···················· | ···················· | ···················· |
| 14994 | MW054154.1 | \|Severe acute respiratory syndrome coro | ···················· | ·A·················· | ···················· | ···················· | ···················· | ···················· |
| 14995 | MW048997.1 | \|Severe acute respiratory syndrome coro | ···················· | ···················· | ···················· | ···················· | ···················· | ···················· |
| 14996 | MW048998.1 | \|Severe acute respiratory syndrome coro | ···················· | ···················· | ···················· | ···················· | ···················· | ···················· |
| 14997 | MW048999.1 | \|Severe acute respiratory syndrome coro | ···················· | ···················· | ···················· | ···················· | ···················· | ···················· |
| 14998 | MW049000.1 | \|Severe acute respiratory syndrome coro | ···················· | ···················· | ···················· | ···················· | ···················· | ···················· |
| 14999 | MW049001.1 | \|Severe acute respiratory syndrome coro | ···················· | ···················· | ···················· | ···················· | ···················· | ···················· |
| 15000 | MW049002.1 | \|Severe acute respiratory syndrome coro | ···················· | ···················· | ···················· | ···················· | ···················· | ···················· |
| 15001 | MW049003.1 | \|Severe acute respiratory syndrome coro | ···················· | ···················· | ···················· | ···················· | ···················· | ···················· |
| 15002 | MW049005.1 | \|Severe acute respiratory syndrome coro | ···················· | ···················· | ···················· | ···················· | ···················· | ···················· |
| 15003 | MW049006.1 | \|Severe acute respiratory syndrome coro | ···················· | ···················· | ···················· | ···················· | ···················· | ···················· |
| 15004 | MW049007.1 | \|Severe acute respiratory syndrome coro | ···················· | ···················· | ···················· | ···················· | ···················· | ···················· |
| 15005 | MW049008.1 | \|Severe acute respiratory syndrome coro | ···················· | ···················· | ···················· | ···················· | ···················· | ···················· |
| 15006 | MW049009.1 | \|Severe acute respiratory syndrome coro | ···················· | ···················· | ···················· | ···················· | ···················· | ···················· |
| 15007 | MW049010.1 | \|Severe acute respiratory syndrome coro | ···················· | ···················· | ···················· | ···················· | ···················· | ···················· |
| 15008 | MW049011.1 | \|Severe acute respiratory syndrome coro | ···················· | ···················· | ···················· | ···················· | ···················· | ···················· |
| 15009 | MW049012.1 | \|Severe acute respiratory syndrome coro | ···················· | ···················· | ···················· | ···················· | ···················· | ···················· |
| 15010 | MW049013.1 | \|Severe acute respiratory syndrome coro | ···················· | ···················· | ···················· | ···················· | ···················· | ···················· |
| 15011 | MW049014.1 | \|Severe acute respiratory syndrome coro | ···················· | ···················· | ···················· | ···················· | ···················· | ···················· |
| 15012 | MW049015.1 | \|Severe acute respiratory syndrome coro | ···················· | ···················· | ···················· | ···················· | ···················· | ···················· |
| 15013 | MW049016.1 | \|Severe acute respiratory syndrome coro | ···················· | ···················· | ···················· | ···················· | ···················· | ···················· |
| 15014 | MW049017.1 | \|Severe acute respiratory syndrome coro | ···················· | ···················· | ···················· | ···················· | ···················· | ···················· |
| 15015 | MW049018.1 | \|Severe acute respiratory syndrome coro | ···················· | ···················· | ···················· | ···················· | ···················· | ···················· |
| 15016 | MW049019.1 | \|Severe acute respiratory syndrome coro | ···················· | ···················· | ···················· | ···················· | ···················· | ···················· |
| 15017 | MW049020.1 | \|Severe acute respiratory syndrome coro | ···················· | ···················· | ···················· | ···················· | ···················· | ···················· |
| 15018 | MW049021.1 | \|Severe acute respiratory syndrome coro | ···················· | ·A·················· | ···················· | ···················· | ···················· | ···················· |
| 15019 | MW049022.1 | \|Severe acute respiratory syndrome coro | ···················· | ···················· | ···················· | ···················· | ···················· | ···················· |
| 15020 | MW049023.1 | \|Severe acute respiratory syndrome coro | ···················· | ···················· | ···················· | ···················· | ···················· | ···················· |
| 15021 | MT704310.1 | \|Severe acute respiratory syndrome coro | ···················· | ···················· | ···················· | ···················· | ···················· | ···················· |
| 15022 | MT704311.1 | \|Severe acute respiratory syndrome coro | ···················· | ···················· | ···················· | ···················· | ···················· | ···················· |
| 15023 | MT704312.1 | \|Severe acute respiratory syndrome coro | ···················· | ···················· | ···················· | ···················· | ···················· | ···················· |
| 15024 | MT704313.1 | \|Severe acute respiratory syndrome coro | ···················· | ···················· | ···················· | ···················· | ···················· | ···················· |
| 15025 | MT704314.1 | \|Severe acute respiratory syndrome coro | ···················· | ···················· | ···················· | ···················· | ···C················ | ···················· |
| 15026 | MT704315.1 | \|Severe acute respiratory syndrome coro | ···················· | ···················· | ···················· | ···················· | ···················· | ···················· |
| 15027 | MT704316.1 | \|Severe acute respiratory syndrome coro | ···················· | ···················· | ···················· | ···················· | ···················· | ···················· |

```
15028 MT704317.1 |Severe acute respiratory syndrome coro    ....................  ....................  .....................  ....................  ....................  ....................
15029 MT747978.1 |Severe acute respiratory syndrome coro    ....................  ....................  .....................  ....................  ....................  ....................
15030 MW047016.1 |Severe acute respiratory syndrome coro    ....................  ....................  .....................  ....................  ....................  ....................
15031 MW047017.1 |Severe acute respiratory syndrome coro    ....................  ....................  .....................  ....................  ....................  ....................
15032 MW047018.1 |Severe acute respiratory syndrome coro    ....................  ....................  .....................  ....................  ....................  ....................
15033 MW047085.1 |Severe acute respiratory syndrome coro    ....................  ....................  .....................  ....................  ....................  ....................
15034 MW047307.1 |Severe acute respiratory syndrome coro    ....................  ....................  .....................  ....................  ....................  ....................
15035 MW047308.1 |Severe acute respiratory syndrome coro    ....................  ....................  .....................  ....................  ....................  ....................
15036 MW047309.1 |Severe acute respiratory syndrome coro    ....................  ....................  .....................  ....................  ....................  ....................
15037 MW047310.1 |Severe acute respiratory syndrome coro    ....................  ....................  .....................  ....................  ....................  ....................
15038 MW047311.1 |Severe acute respiratory syndrome coro    ....................  ....................  .....................  ....................  ....................  ....................
15039 MW047312.1 |Severe acute respiratory syndrome coro    ....................  ....................  .....................  ....................  ....................  ....................
15040 MW047313.1 |Severe acute respiratory syndrome coro    ....................  ....................  .....................  ....................  ....................  ....................
15041 MW047314.1 |Severe acute respiratory syndrome coro    ....................  ....................  .....................  ....................  ....................  ....................
15042 MW047315.1 |Severe acute respiratory syndrome coro    ....................  ....................  .....................  ....................  ....................  ....................
15043 MW047316.1 |Severe acute respiratory syndrome coro    ....................  ....................  .....................  ....................  ....................  ....................
15044 MW047317.1 |Severe acute respiratory syndrome coro    ....................  ....................  .....................  ....................  ....................  ....................
15045 MW047318.1 |Severe acute respiratory syndrome coro    ....................  ....................  .....................  ....................  ....................  ....................
15046 MW041156.1 |Severe acute respiratory syndrome coro    ....................  ....................  .....................  ....................  ....................  ....................
15047 MW042923.1 |Severe acute respiratory syndrome coro    ....................  .A..................  .....................  ....................  ....................  ....................
15048 MW042925.1 |Severe acute respiratory syndrome coro    ....................  ....................  .....................  ....................  ....................  ....................
15049 MW040606.1 |Severe acute respiratory syndrome coro    ....................  ....................  .....................  ....................  ....................  ....................
15051 MW040621.1 |Severe acute respiratory syndrome coro    ....................  ....................  .....................  ....................  ....................  ....................
15052 MW040622.1 |Severe acute respiratory syndrome coro    ....................  ....................  .....................  ....................  ....................  ....................
15053 MW040623.1 |Severe acute respiratory syndrome coro    ....................  ....................  .....................  ....................  ....................  ....................
15054 MW040624.1 |Severe acute respiratory syndrome coro    ....................  ....................  .....................  ....................  ....................  ....................
15055 MW040625.1 |Severe acute respiratory syndrome coro    ....................  ....................  .....................  ....................  ....................  ....................
15056 MW040627.1 |Severe acute respiratory syndrome coro    ....................  ....................  .....................  ....................  ....................  ....................
15057 MW040628.1 |Severe acute respiratory syndrome coro    ....................  ....................  .....................  ....................  ....................  ....................
15058 MW040629.1 |Severe acute respiratory syndrome coro    ....................  ....................  .....................  ....................  ....................  ....................
15059 MW040630.1 |Severe acute respiratory syndrome coro    ....................  ....................  .....................  ....................  ....................  ....................
15060 MW040631.1 |Severe acute respiratory syndrome coro    ....................  ....................  .....................  ....................  ....................  ....................
15061 MW040632.1 |Severe acute respiratory syndrome coro    ....................  ....................  .....................  ....................  ....................  ....................
15062 MW040633.1 |Severe acute respiratory syndrome coro    ....................  ....................  .....................  ....................  ....................  ....................
15063 MW040634.1 |Severe acute respiratory syndrome coro    ....................  ....................  .....................  ....................  ....................  ....................
15064 MW040635.1 |Severe acute respiratory syndrome coro    ....................  ....................  .....................  ....................  ....................  ....................
15065 MW040636.1 |Severe acute respiratory syndrome coro    ....................  ....................  .....................  ....................  ....................  ....................
15066 MW040638.1 |Severe acute respiratory syndrome coro    ....................  ....................  .....................  ....................  ....................  ....................
15068 MW040640.1 |Severe acute respiratory syndrome coro    ....................  .A..................  .....................  ....................  ....................  ....................
15069 MW040641.1 |Severe acute respiratory syndrome coro    ....................  ....................  .....................  ....................  ....................  ....................
15070 MW040642.1 |Severe acute respiratory syndrome coro    ....................  ....................  .....................  ....................  ....................  ....................
15071 MW040643.1 |Severe acute respiratory syndrome coro    ....................  ....................  .....................  ....................  ....................  ....................
15072 MW040644.1 |Severe acute respiratory syndrome coro    ....................  ....................  .....................  ....................  ....................  ....................
15073 MW040645.1 |Severe acute respiratory syndrome coro    ....................  ....................  .....................  ....................  ....................  ....................
15074 MW040646.1 |Severe acute respiratory syndrome coro    ....................  ....................  .....................  ....................  ....................  ....................
15075 MW040647.1 |Severe acute respiratory syndrome coro    ....................  ....................  .....................  ....................  ....................  ....................
15076 MW040648.1 |Severe acute respiratory syndrome coro    ....................  ....................  .....................  ....................  ....................  ....................
15078 MW040650.1 |Severe acute respiratory syndrome coro    ....................  ....................  .....................  ....................  ....................  ....................
15079 MW040651.1 |Severe acute respiratory syndrome coro    ....................  ....................  .....................  ....................  ....................  ....................
15080 MW040652.1 |Severe acute respiratory syndrome coro    ....................  ....................  .....................  ....................  ....................  ....................
15081 MW040653.1 |Severe acute respiratory syndrome coro    ....................  .A..................  .....................  ....................  ....................  ....................
15082 MW040654.1 |Severe acute respiratory syndrome coro    ....................  ....................  .....................  ....................  ....................  ....................
15083 MW040655.1 |Severe acute respiratory syndrome coro    ....................  ....................  .....................  ....................  ....................  ....................
15084 MW040659.1 |Severe acute respiratory syndrome coro    ....................  ....................  .....................  ....................  ....................  ....................
15085 MW040660.1 |Severe acute respiratory syndrome coro    ....................  ....................  .....................  ....................  ....................  ....................
15086 MW040664.1 |Severe acute respiratory syndrome coro    ....................  ....................  .....................  ....................  ....................  ....................
15087 MW040665.1 |Severe acute respiratory syndrome coro    ....................  ....................  .....................  ....................  ....................  ....................
15088 MW040666.1 |Severe acute respiratory syndrome coro    ....................  ....................  .....................  ....................  ....................  ....................
15089 MW040667.1 |Severe acute respiratory syndrome coro    ....................  ....................  .....................  ....................  ....................  ....................
15090 MW040668.1 |Severe acute respiratory syndrome coro    ....................  ....................  .....................  ....................  ....................  ....................
15092 MW040673.1 |Severe acute respiratory syndrome coro    ....................  ....................  .....................  ....................  ....................  ....................
15093 MW040675.1 |Severe acute respiratory syndrome coro    ....................  ....................  .....................  ....................  ....................  ....................
15094 MW040678.1 |Severe acute respiratory syndrome coro    ....................  ....................  .....................  ....................  ....................  ....................
15095 MW040679.1 |Severe acute respiratory syndrome coro    ....................  ....................  .....................  ....................  ....................  ....................
15096 MW040681.1 |Severe acute respiratory syndrome coro    ....................  ....................  .....................  ....................  ....................  ....................
15097 MW040683.1 |Severe acute respiratory syndrome coro    ....................  ....................  .....................  ....................  ....................  ....................
15098 MW040686.1 |Severe acute respiratory syndrome coro    ....................  ....................  .....................  ....................  ....................  ....................
15099 MW040687.1 |Severe acute respiratory syndrome coro    ....................  ....................  .....................  ....................  ....................  ....................
15100 MW040690.1 |Severe acute respiratory syndrome coro    ....................  ....................  .....................  ....................  ....................  ....................
15101 MW040693.1 |Severe acute respiratory syndrome coro    ....................  ....................  .....................  ....................  ....................  ....................
15102 MW040694.1 |Severe acute respiratory syndrome coro    ....................  ....................  .....................  ....................  ....................  ....................
15103 MW040695.1 |Severe acute respiratory syndrome coro    ....................  ....................  .....................  ....................  ....................  ....................
15104 MW040696.1 |Severe acute respiratory syndrome coro    ....................  ....................  .....................  ....................  ....................  ....................
15105 MW040697.1 |Severe acute respiratory syndrome coro    ....................  ....................  .....................  ....................  ....................  ....................
15106 MT594401.1 |Severe acute respiratory syndrome coro    ....................  ....................  .....................  ....................  ....................  ....................
15107 MT594402.1 |Severe acute respiratory syndrome coro    ....................  ....................  .....................  ....................  ....................  ....................
15108 MW039234.1 |Severe acute respiratory syndrome coro    ....................  ....................  .....................  ....................  ....................  ....................
15109 MW039235.1 |Severe acute respiratory syndrome coro    ....................  ....................  .....................  ....................  ....................  ....................
```

| | | | | | | | | | |
|---|---|---|---|---|---|---|---|---|---|
| 15110 | MW039450.1 | Severe acute respiratory syndrome coro | ···················· | ···················· | ···················· | ···················· | ···················· | ···················· | |
| 15111 | MW035370.1 | Severe acute respiratory syndrome coro | ···················· | ···················· | ···················· | ···················· | ···················· | ···················· | |
| 15112 | MW035371.1 | Severe acute respiratory syndrome coro | ···················· | ···················· | ···················· | ···················· | ···················· | ···················· | |
| 15113 | MW035373.1 | Severe acute respiratory syndrome coro | ···················· | ···················· | ···················· | ···················· | ···················· | ···················· | |
| 15114 | MW035374.1 | Severe acute respiratory syndrome coro | ···················· | ···················· | ···················· | ···················· | ···················· | ···················· | |
| 15115 | MW035376.1 | Severe acute respiratory syndrome coro | ···················· | ···················· | ···················· | ···················· | ···················· | ···················· | |
| 15116 | MW035377.1 | Severe acute respiratory syndrome coro | ···················· | ···················· | ···················· | ···················· | ···················· | ···················· | |
| 15117 | MW035378.1 | Severe acute respiratory syndrome coro | ···················· | ···················· | ···················· | ···················· | ···················· | ···················· | |
| 15118 | MW035379.1 | Severe acute respiratory syndrome coro | ···················· | ···················· | ···················· | ···················· | ···················· | ···················· | |
| 15119 | MW035380.1 | Severe acute respiratory syndrome coro | ···················· | ···················· | ···················· | ···················· | ···················· | ···················· | |
| 15120 | MW035381.1 | Severe acute respiratory syndrome coro | ···················· | ···················· | ···················· | ···················· | ···················· | ···················· | |
| 15121 | MW035382.1 | Severe acute respiratory syndrome coro | ···················· | ···················· | ···················· | ···················· | ···················· | ···················· | |
| 15122 | MW035384.1 | Severe acute respiratory syndrome coro | ···················· | ···················· | ···················· | ···················· | ···················· | ···················· | |
| 15123 | MW035385.1 | Severe acute respiratory syndrome coro | ···················· | ···················· | ···················· | ···················· | ···················· | ···················· | |
| 15124 | MW035386.1 | Severe acute respiratory syndrome coro | ···················· | ·············A······ | ···················· | ···················· | ···················· | ···················· | |
| 15125 | MW035388.1 | Severe acute respiratory syndrome coro | ···················· | ···················· | ···················· | ···················· | ···················· | ···················· | |
| 15126 | MW035389.1 | Severe acute respiratory syndrome coro | ···················· | ···················· | ···················· | ···················· | ···················· | ···················· | |
| 15127 | MW035391.1 | Severe acute respiratory syndrome coro | ···················· | ···················· | ···················· | ···················· | ···················· | ···················· | |
| 15128 | MW035392.1 | Severe acute respiratory syndrome coro | ···················· | ···················· | ···················· | ···················· | ···················· | ···················· | |
| 15129 | MW035393.1 | Severe acute respiratory syndrome coro | ···················· | ···················· | ···················· | ···················· | ···················· | ···················· | |
| 15130 | MW035394.1 | Severe acute respiratory syndrome coro | ···················· | ···················· | ···················· | ···················· | ···················· | ···················· | |
| 15131 | MW035395.1 | Severe acute respiratory syndrome coro | ···················· | ···················· | ···················· | ···················· | ···················· | ···················· | |
| 15132 | MW035396.1 | Severe acute respiratory syndrome coro | ···················· | ·A·················· | ···················· | ···················· | ···················· | ···················· | |
| 15133 | MW035397.1 | Severe acute respiratory syndrome coro | ···················· | ···················· | ···················· | ···················· | ···················· | ···················· | |
| 15134 | MW035399.1 | Severe acute respiratory syndrome coro | ···················· | ···················· | ···················· | ···················· | ···················· | ···················· | |
| 15135 | MW035400.1 | Severe acute respiratory syndrome coro | ···················· | ···················· | ···················· | ···················· | ···················· | ···················· | |
| 15136 | MW035401.1 | Severe acute respiratory syndrome coro | ···················· | ···················· | ···················· | ···················· | ···················· | ···················· | |
| 15137 | MW035402.1 | Severe acute respiratory syndrome coro | ···················· | ···················· | ···················· | ···················· | ···················· | ···················· | |
| 15138 | MW035403.1 | Severe acute respiratory syndrome coro | ···················· | ···················· | ···················· | ···················· | ···················· | ···················· | |
| 15139 | MW035404.1 | Severe acute respiratory syndrome coro | ···················· | ···················· | ···················· | ···················· | ···················· | ···················· | |
| 15140 | MW035405.1 | Severe acute respiratory syndrome coro | ···················· | ···················· | ···················· | ···················· | ···················· | ···················· | |
| 15141 | MW035406.1 | Severe acute respiratory syndrome coro | ···················· | ·A·················· | ···················· | ···················· | ···················· | ···················· | |
| 15142 | MW035407.1 | Severe acute respiratory syndrome coro | ···················· | ···················· | ···················· | ···················· | ···················· | ···················· | |
| 15143 | MW035408.1 | Severe acute respiratory syndrome coro | ···················· | ···················· | ···················· | ···················· | ···················· | ···················· | |
| 15144 | MW035409.1 | Severe acute respiratory syndrome coro | ···················· | ···················· | ···················· | ···················· | ···················· | ···················· | |
| 15145 | MW035410.1 | Severe acute respiratory syndrome coro | ···················· | ···················· | ···················· | ···················· | ···················· | ···················· | |
| 15146 | MW035411.1 | Severe acute respiratory syndrome coro | ···················· | ···················· | ···················· | ···················· | ···················· | ···················· | |
| 15147 | MW035412.1 | Severe acute respiratory syndrome coro | ···················· | ···················· | ···················· | ···················· | ···················· | ···················· | |
| 15148 | MW035413.1 | Severe acute respiratory syndrome coro | ···················· | ···················· | ···················· | ···················· | ···················· | ···················· | |
| 15149 | MW035414.1 | Severe acute respiratory syndrome coro | ···················· | ···················· | ···················· | ···················· | ···················· | ···················· | |
| 15150 | MW035415.1 | Severe acute respiratory syndrome coro | ···················· | ···················· | ···················· | ···················· | ···················· | ···················· | |
| 15151 | MW035416.1 | Severe acute respiratory syndrome coro | ···················· | ···················· | ···················· | ···················· | ···················· | ···················· | |
| 15152 | MW035418.1 | Severe acute respiratory syndrome coro | ···················· | ··················C· | ···················· | ···················· | ···················· | ···················· | 1 |
| 15153 | MW035419.1 | Severe acute respiratory syndrome coro | ···················· | ···················· | ···················· | ···················· | ···················· | ···················· | |
| 15154 | MW035420.1 | Severe acute respiratory syndrome coro | ···················· | ···················· | ···················· | ···················· | ···················· | ···················· | |
| 15155 | MW035422.1 | Severe acute respiratory syndrome coro | ···················· | ···················· | ···················· | ···················· | ···················· | ···················· | |
| 15156 | MW035423.1 | Severe acute respiratory syndrome coro | ···················· | ···················· | ···················· | ···················· | ···················· | ···················· | |
| 15157 | MW035425.1 | Severe acute respiratory syndrome coro | ···················· | ···················· | ···················· | ···················· | ···················· | ···················· | |
| 15158 | MW035426.1 | Severe acute respiratory syndrome coro | ···················· | ···················· | ···················· | ···················· | ···················· | ···················· | |
| 15159 | MW035427.1 | Severe acute respiratory syndrome coro | ···················· | ···················· | ···················· | ···················· | ···················· | ···················· | |
| 15160 | MW035428.1 | Severe acute respiratory syndrome coro | ···················· | ·A·················· | ···················· | ···················· | ···················· | ···················· | |
| 15161 | MW035429.1 | Severe acute respiratory syndrome coro | ···················· | ···················· | ···················· | ···················· | ···················· | ···················· | |
| 15162 | MW035430.1 | Severe acute respiratory syndrome coro | ···················· | ···················· | ···················· | ···················· | ···················· | ···················· | |
| 15163 | MW035431.1 | Severe acute respiratory syndrome coro | ···················· | ·A·················· | ···················· | ···················· | ···················· | ···················· | |
| 15164 | MW035432.1 | Severe acute respiratory syndrome coro | ···················· | ···················· | ···················· | ···················· | ···················· | ···················· | |
| 15165 | MW035433.1 | Severe acute respiratory syndrome coro | ···················· | ···················· | ···················· | ···················· | ···················· | ···················· | |
| 15166 | MW035434.1 | Severe acute respiratory syndrome coro | ···················· | ···················· | ···················· | ···················· | ···················· | ···················· | |
| 15167 | MW035435.1 | Severe acute respiratory syndrome coro | ···················· | ·A·················· | ···················· | ···················· | ···················· | ···················· | |
| 15168 | MW035436.1 | Severe acute respiratory syndrome coro | ···················· | ···················· | ···················· | ···················· | ···················· | ···················· | |
| 15169 | MW035437.1 | Severe acute respiratory syndrome coro | ···················· | ···················· | ···················· | ···················· | ···················· | ···················· | |
| 15170 | MW035438.1 | Severe acute respiratory syndrome coro | ···················· | ···················· | ···················· | ···················· | ···················· | ···················· | |
| 15171 | MW035439.1 | Severe acute respiratory syndrome coro | ···················· | ···················· | ···················· | ···················· | ···················· | ···················· | |
| 15172 | MW035440.1 | Severe acute respiratory syndrome coro | ···················· | ···················· | ···················· | ···················· | ···················· | ···················· | |
| 15173 | MW035441.1 | Severe acute respiratory syndrome coro | ···················· | ···················· | ···················· | ···················· | ···················· | ···················· | |
| 15174 | MW035442.1 | Severe acute respiratory syndrome coro | ···················· | ···················· | ···················· | ···················· | ···················· | ···················· | |
| 15175 | MW035443.1 | Severe acute respiratory syndrome coro | ···················· | ···················· | ···················· | ···················· | ···················· | ···················· | |
| 15176 | MW035444.1 | Severe acute respiratory syndrome coro | ···················· | ···················· | ···················· | ···················· | ···················· | ···················· | |
| 15177 | MW035445.1 | Severe acute respiratory syndrome coro | ···················· | ···················· | ···················· | ···················· | ···················· | ···················· | |
| 15178 | MW035446.1 | Severe acute respiratory syndrome coro | ···················· | ···················· | ···················· | ···················· | ···················· | ···················· | |
| 15179 | MW035447.1 | Severe acute respiratory syndrome coro | ···················· | ···················· | ···················· | ···················· | ···················· | ···················· | |
| 15180 | MW035448.1 | Severe acute respiratory syndrome coro | ···················· | ···················· | ···················· | ···················· | ···················· | ···················· | |
| 15181 | MW035449.1 | Severe acute respiratory syndrome coro | ···················· | ·A·················· | ···················· | ···················· | ···················· | ···················· | |
| 15182 | MW035450.1 | Severe acute respiratory syndrome coro | ···················· | ···················· | ···················· | ···················· | ···················· | ···················· | |
| 15183 | MW035451.1 | Severe acute respiratory syndrome coro | ···················· | ···················· | ···················· | ···················· | ···················· | ···················· | |
| 15184 | MW035452.1 | Severe acute respiratory syndrome coro | ···················· | ···················· | ···················· | ···················· | ···················· | ···················· | |
| 15185 | MW035453.1 | Severe acute respiratory syndrome coro | ···················· | ·A·················· | ···················· | ···················· | ···················· | ···················· | |
| 15186 | MW035454.1 | Severe acute respiratory syndrome coro | ···················· | ···················· | ···················· | ···················· | ···················· | ···················· | |
| 15187 | MW035455.1 | Severe acute respiratory syndrome coro | ···················· | ···················· | ···················· | ···················· | ···················· | ···················· | |

15188 MW035456.1 |Severe acute respiratory syndrome coro .................... .................... .................... .................... .................... ....................
15189 MW035458.1 |Severe acute respiratory syndrome coro .................... .A.................. .................... .................... .................... ....................
15190 MW035459.1 |Severe acute respiratory syndrome coro .................... .................... .................... .................... .................... ....................
15191 MW035460.1 |Severe acute respiratory syndrome coro .................... .................... .................... .................... .................... ....................
15192 MW035461.1 |Severe acute respiratory syndrome coro .................... .................... .................... .................... .................... ....................
15193 MW035462.1 |Severe acute respiratory syndrome coro .................... .................... .................... .................... .................... ....................
15194 MW035463.1 |Severe acute respiratory syndrome coro .................... .................... .................... .................... .................... ....................
15195 MW035464.1 |Severe acute respiratory syndrome coro .................... .................... .................... .................... .................... ....................
15196 MW035465.1 |Severe acute respiratory syndrome coro .................... ..C................. .................... .................... .................... ....................
15197 MW035466.1 |Severe acute respiratory syndrome coro .................... .................... .................... .................... .................... ....................
15198 MW035468.1 |Severe acute respiratory syndrome coro .................... .................... .................... .................... .................... ....................
15199 MW035469.1 |Severe acute respiratory syndrome coro .................... .................... .................... .................... .................... ....................
15200 MW035471.1 |Severe acute respiratory syndrome coro .................... .................... .................... .................... .................... ....................
15201 MW035472.1 |Severe acute respiratory syndrome coro .................... .................... .................... .................... .................... ....................
15202 MW035473.1 |Severe acute respiratory syndrome coro .................... .A.................. .................... .................... .................... ....................
15203 MW035474.1 |Severe acute respiratory syndrome coro .................... .................... .................... .................... .................... ....................
15204 MW035475.1 |Severe acute respiratory syndrome coro .................... .................... .................... .................... .................... ....................
15205 MW035476.1 |Severe acute respiratory syndrome coro .................... .................... .................... .................... .................... ....................
15206 MW035484.1 |Severe acute respiratory syndrome coro .................... .................... .................... .................... .................... ....................
15207 MW035495.1 |Severe acute respiratory syndrome coro .................... .................... .................... .................... .................... ....................
15208 MW035496.1 |Severe acute respiratory syndrome coro .................... .................... .................... .................... .................... ....................
15209 MW035497.1 |Severe acute respiratory syndrome coro .................... .................... .................... .................... .................... ....................
15210 MW035498.1 |Severe acute respiratory syndrome coro .................... .................... .................... .................... .................... ....................
15211 MW035499.1 |Severe acute respiratory syndrome coro .................... ..C................. .................... .................... .................... ....................
15212 MW035500.1 |Severe acute respiratory syndrome coro .................... .................... .................... .................... .................... ....................
15213 MW035501.1 |Severe acute respiratory syndrome coro .................... .................... .................... .................... .................... ....................
15214 MW035502.1 |Severe acute respiratory syndrome coro .................... .................... .................... .................... .................... ....................
15215 MW035503.1 |Severe acute respiratory syndrome coro .................... .............A...... .................... .................... .................... ....................
15216 MW035504.1 |Severe acute respiratory syndrome coro .................... .................... .................... .................... .................... ....................
15217 MW035505.1 |Severe acute respiratory syndrome coro .................... .................... .................... .................... .................... ....................
15218 MW035506.1 |Severe acute respiratory syndrome coro .................... .................... .................... .................... .................... ....................
15219 MW035507.1 |Severe acute respiratory syndrome coro .................... .................... .................... .................... .................... ....................
15220 MW035509.1 |Severe acute respiratory syndrome coro .................... .................... .................... .................... .................... ....................
15221 MW035511.1 |Severe acute respiratory syndrome coro .................... .................... .................... .................... .................... ....................
15222 MW035512.1 |Severe acute respiratory syndrome coro .................... .................... .................... .................... .................... ....................
15223 MW035514.1 |Severe acute respiratory syndrome coro .................... .................... .................... .................... .................... ....................
15224 MW035515.1 |Severe acute respiratory syndrome coro .................... .................... .................... .................... .................... ....................
15225 MW035516.1 |Severe acute respiratory syndrome coro .................... .................... .................... .................... .................... ....................
15226 MW035517.1 |Severe acute respiratory syndrome coro .................... .................... .................... .................... .................... ....................
15227 MW035518.1 |Severe acute respiratory syndrome coro .................... .................... .................... .................... .................... ....................
15228 MW035519.1 |Severe acute respiratory syndrome coro .................... .................... .................... .................... .................... ....................
15229 MW035520.1 |Severe acute respiratory syndrome coro .................... .................... .................... .................... .................... ....................
15230 MW035521.1 |Severe acute respiratory syndrome coro .................... .................... .................... .................... .................... ....................
15231 MW035522.1 |Severe acute respiratory syndrome coro .................... .................... .................... .................... .................... ....................
15232 MW035524.1 |Severe acute respiratory syndrome coro .................... .................... .................... .................... .................... ....................
15233 MW035525.1 |Severe acute respiratory syndrome coro .................... .................... .................... .................... .................... ....................
15234 MW035526.1 |Severe acute respiratory syndrome coro .................... .................... .................... .................... .................... ....................
15235 MW035527.1 |Severe acute respiratory syndrome coro .................... .................... .................... .................... .................... ....................
15236 MW035528.1 |Severe acute respiratory syndrome coro .................... .................... .................... .................... .................... ....................
15237 MW035529.1 |Severe acute respiratory syndrome coro .................... .................... .................... .................... .................... ....................
15238 MW035530.1 |Severe acute respiratory syndrome coro .................... .A.................. .................... .................... .................... ....................
15239 MW035531.1 |Severe acute respiratory syndrome coro .................... .................... .................... .................... .................... ....................
15240 MW035532.1 |Severe acute respiratory syndrome coro .................... .................... .................... .................... .................... ....................
15241 MW035533.1 |Severe acute respiratory syndrome coro .................... .................... .................... .................... .................... ....................
15242 MW035534.1 |Severe acute respiratory syndrome coro .................... .................... .................... .................... .................... ....................
15243 MW035535.1 |Severe acute respiratory syndrome coro .................... .................... .................... .................... .................... ....................
15244 MW035536.1 |Severe acute respiratory syndrome coro .................... .................... .................... .................... .................... ....................
15245 MW035537.1 |Severe acute respiratory syndrome coro .................... .................... .................... .................... .................... ....................
15246 MW035538.1 |Severe acute respiratory syndrome coro .................... .................... .................... .................... .................... ....................
15247 MW035539.1 |Severe acute respiratory syndrome coro .................... .................... .................... .................... .................... ....................
15248 MW035540.1 |Severe acute respiratory syndrome coro .................... .................... .................... .................... .................... ....................
15249 MW035541.1 |Severe acute respiratory syndrome coro .................... .................... .................... .................... .................... ....................
15250 MW035542.1 |Severe acute respiratory syndrome coro .................... .................... .................... .................... .................... ....................
15251 MW035543.1 |Severe acute respiratory syndrome coro .................... .................... .................... .................... .................... ....................
15252 MW035545.1 |Severe acute respiratory syndrome coro .................... .................... .................... .................... .................... ....................
15253 MW035546.1 |Severe acute respiratory syndrome coro .................... .................... .................... .................... .................... ....................
15254 MW035548.1 |Severe acute respiratory syndrome coro .................... .................... .................... .................... .................... ....................
15255 MW035549.1 |Severe acute respiratory syndrome coro .................... .................... .................... .................... .................... ....................
15256 MW035550.1 |Severe acute respiratory syndrome coro .................... .................... .................... .................... .................... ....................
15257 MW035551.1 |Severe acute respiratory syndrome coro .................... .................... .................... .................... .................... ....................
15258 MW035552.1 |Severe acute respiratory syndrome coro .................... .................... .................... .................... .................... ....................
15259 MW035553.1 |Severe acute respiratory syndrome coro .................... .................... .................... .................... .................... ....................
15260 MW035554.1 |Severe acute respiratory syndrome coro .................... .................... .................... .................... .................... ....................
15261 MW035555.1 |Severe acute respiratory syndrome coro .................... .................... .................... .................... .................... ....................
15262 MW035556.1 |Severe acute respiratory syndrome coro .................... .................... .................... .................... .................... ....................
15263 MW035557.1 |Severe acute respiratory syndrome coro .................... .................... .................... .................... .................... ....................
15264 MW035558.1 |Severe acute respiratory syndrome coro .................... .................... .................... .................... .................... ....................
15265 MW035559.1 |Severe acute respiratory syndrome coro .................... .................... .................... .................... .................... ....................

| | | | | | | | | |
|---|---|---|---|---|---|---|---|---|
| 15266 | MW035560.1 | \|Severe acute respiratory syndrome coro | ···················· | ···················· | ···················· | ···················· | ···················· | ···················· |
| 15267 | MW035561.1 | \|Severe acute respiratory syndrome coro | ···················· | ···················· | ···················· | ···················· | ···················· | ···················· |
| 15268 | MW035562.1 | \|Severe acute respiratory syndrome coro | ···················· | ···················· | ···················· | ···················· | ···················· | ···················· |
| 15269 | MW035564.1 | \|Severe acute respiratory syndrome coro | ···················· | ···················· | ···················· | ···················· | ···················· | ···················· |
| 15270 | MW035565.1 | \|Severe acute respiratory syndrome coro | ···················· | ···················· | ···················· | ···················· | ···················· | ···················· |
| 15271 | MW035566.1 | \|Severe acute respiratory syndrome coro | ···················· | ···················· | ···················· | ···················· | ···················· | ···················· |
| 15272 | MW035567.1 | \|Severe acute respiratory syndrome coro | ···················· | ···················· | ···················· | ···················· | ···················· | ···················· |
| 15273 | MW035568.1 | \|Severe acute respiratory syndrome coro | ···················· | ···················· | ···················· | ···················· | ···················· | ···················· |
| 15274 | MW035569.1 | \|Severe acute respiratory syndrome coro | ···················· | ···················· | ···················· | ···················· | ···················· | ···················· |
| 15275 | MW035570.1 | \|Severe acute respiratory syndrome coro | ···················· | ···················· | ···················· | ···················· | ···················· | ···················· |
| 15276 | MW035571.1 | \|Severe acute respiratory syndrome coro | ···················· | ···················· | ···················· | ···················· | ···················· | ···················· |
| 15277 | MW035572.1 | \|Severe acute respiratory syndrome coro | ···················· | ···················· | ···················· | ···················· | ···················· | ···················· |
| 15278 | MW035573.1 | \|Severe acute respiratory syndrome coro | ···················· | ···················· | ···················· | ···················· | ···················· | ···················· |
| 15279 | MW035574.1 | \|Severe acute respiratory syndrome coro | ···················· | ···················· | ···················· | ···················· | ···················· | ···················· |
| 15280 | MW035575.1 | \|Severe acute respiratory syndrome coro | ···················· | ···················· | ···················· | ···················· | ···················· | ···················· |
| 15281 | MW035576.1 | \|Severe acute respiratory syndrome coro | ···················· | ···················· | ···················· | ···················· | ···················· | ···················· |
| 15282 | MW035577.1 | \|Severe acute respiratory syndrome coro | ···················· | ···················· | ···················· | ···················· | ···················· | ···················· |
| 15283 | MW035578.1 | \|Severe acute respiratory syndrome coro | ···················· | ···················· | ···················· | ···················· | ···················· | ···················· |
| 15284 | MW035579.1 | \|Severe acute respiratory syndrome coro | ···················· | ···················· | ···················· | ···················· | ···················· | ···················· |
| 15285 | MW035918.1 | \|Severe acute respiratory syndrome coro | ···················· | ···················· | ···················· | ···················· | ···················· | ···················· |
| 15286 | MW035919.1 | \|Severe acute respiratory syndrome coro | ···················· | ···················· | ···················· | ···················· | ···················· | ···················· |
| 15287 | MW035920.1 | \|Severe acute respiratory syndrome coro | ···················· | ···················· | ···················· | ···················· | ···················· | ···················· |
| 15288 | MW035921.1 | \|Severe acute respiratory syndrome coro | ··············Y····· | ···················· | ···················· | ···················· | ···················· | ···················· |
| 15289 | MW035922.1 | \|Severe acute respiratory syndrome coro | ···················· | ···················· | ···················· | ···················· | ···················· | ···················· |
| 15290 | MW035923.1 | \|Severe acute respiratory syndrome coro | ···················· | ···················· | ···················· | ···················· | ···················· | ···················· |
| 15291 | MW035924.1 | \|Severe acute respiratory syndrome coro | ···················· | ···················· | ···················· | ···················· | ···················· | ···················· |
| 15292 | MW035925.1 | \|Severe acute respiratory syndrome coro | ···················· | ···················· | ···················· | ···················· | ···················· | ···················· |
| 15293 | MW035926.1 | \|Severe acute respiratory syndrome coro | ···················· | ···················· | ···················· | ···················· | ···················· | ···················· |
| 15294 | MW035927.1 | \|Severe acute respiratory syndrome coro | ···················· | ···················· | ···················· | ···················· | ···················· | ···················· |
| 15295 | MW035928.1 | \|Severe acute respiratory syndrome coro | ···················· | ···················· | ···················· | ···················· | ···················· | ···················· |
| 15296 | MW035929.1 | \|Severe acute respiratory syndrome coro | ···················· | ···················· | ···················· | ···················· | ···················· | ···················· |
| 15297 | MW035930.1 | \|Severe acute respiratory syndrome coro | ···················· | ···················· | ···················· | ···················· | ···················· | ···················· |
| 15298 | MW035931.1 | \|Severe acute respiratory syndrome coro | ···················· | ···················· | ···················· | ···················· | ···················· | ···················· |
| 15299 | MW035932.1 | \|Severe acute respiratory syndrome coro | ···················· | ···················· | ···················· | ···················· | ···················· | ···················· |
| 15300 | MW035933.1 | \|Severe acute respiratory syndrome coro | ···················· | ···················· | ···················· | ···················· | ···················· | ···················· |
| 15301 | MW035934.1 | \|Severe acute respiratory syndrome coro | ···················· | ···················· | ···················· | ···················· | ···················· | ···················· |
| 15302 | MW035935.1 | \|Severe acute respiratory syndrome coro | ···················· | ···················· | ···················· | ···················· | ···················· | ···················· |
| 15303 | MW035936.1 | \|Severe acute respiratory syndrome coro | ··············Y····· | ·A·················· | ···················· | ···················· | ···················· | ···················· |
| 15304 | MW035937.1 | \|Severe acute respiratory syndrome coro | ···················· | ···················· | ···················· | ···················· | ···················· | ···················· |
| 15305 | MW035938.1 | \|Severe acute respiratory syndrome coro | ···················· | ···················· | ···················· | ···················· | ···················· | ···················· |
| 15306 | MW035939.1 | \|Severe acute respiratory syndrome coro | ···················· | ···················· | ···················· | ···················· | ···················· | ···················· |
| 15307 | MW035940.1 | \|Severe acute respiratory syndrome coro | ···················· | ···················· | ···················· | ···················· | ···················· | ···················· |
| 15308 | MW035941.1 | \|Severe acute respiratory syndrome coro | ···················· | ···················· | ···················· | ···················· | ···················· | ···················· |
| 15309 | MW035942.1 | \|Severe acute respiratory syndrome coro | ···················· | ···················· | ···················· | ···················· | ···················· | ···················· |
| 15310 | MW035943.1 | \|Severe acute respiratory syndrome coro | ···················· | ···················· | ···················· | ···················· | ···················· | ···················· |
| 15311 | MW035944.1 | \|Severe acute respiratory syndrome coro | ···················· | ···················· | ···················· | ···················· | ···················· | ···················· |
| 15312 | MW035945.1 | \|Severe acute respiratory syndrome coro | ···················· | ···················· | ···················· | ···················· | ···················· | ···················· |
| 15313 | MW035946.1 | \|Severe acute respiratory syndrome coro | ···················· | ···················· | ···················· | ···················· | ···················· | ···················· |
| 15314 | MW035947.1 | \|Severe acute respiratory syndrome coro | ···················· | ···················· | ···················· | ···················· | ···················· | ···················· |
| 15315 | MW035948.1 | \|Severe acute respiratory syndrome coro | ···················· | ···················· | ···················· | ···················· | ···················· | ···················· |
| 15316 | MW035949.1 | \|Severe acute respiratory syndrome coro | ···················· | ···················· | ···················· | ···················· | ···················· | ···················· |
| 15317 | MW035950.1 | \|Severe acute respiratory syndrome coro | ···················· | ···················· | ···················· | ···················· | ···················· | ···················· |
| 15318 | MW035951.1 | \|Severe acute respiratory syndrome coro | ···················· | ···················· | ···················· | ···················· | ···················· | ···················· |
| 15319 | MW035952.1 | \|Severe acute respiratory syndrome coro | ···················· | ···················· | ···················· | ···················· | ···················· | ···················· |
| 15320 | MW035953.1 | \|Severe acute respiratory syndrome coro | ···················· | ···················· | ···················· | ···················· | ···················· | ···················· |
| 15321 | MW035954.1 | \|Severe acute respiratory syndrome coro | ···················· | ···················· | ···················· | ···················· | ···················· | ···················· |
| 15322 | MW035955.1 | \|Severe acute respiratory syndrome coro | ···················· | ···················· | ···················· | ···················· | ···················· | ···················· |
| 15323 | MW035956.1 | \|Severe acute respiratory syndrome coro | ········T··········· | ···················· | ···················· | ···················· | ···················· | ···················· |
| 15324 | MW035957.1 | \|Severe acute respiratory syndrome coro | ···················· | ···················· | ···················· | ···················· | ···················· | ···················· |
| 15325 | MW035958.1 | \|Severe acute respiratory syndrome coro | ···················· | ···················· | ···················· | ···················· | ···················· | ···················· |
| 15326 | MW035959.1 | \|Severe acute respiratory syndrome coro | ···················· | ···················· | ···················· | ···················· | ···················· | ···················· |
| 15327 | MW035960.1 | \|Severe acute respiratory syndrome coro | ···················· | ···················· | ···················· | ···················· | ···················· | ···················· |
| 15328 | MW035962.1 | \|Severe acute respiratory syndrome coro | ···················· | ·A·················· | ···················· | ···················· | ···················· | ···················· |
| 15329 | MW035963.1 | \|Severe acute respiratory syndrome coro | ···················· | ···················· | ···················· | ···················· | ···················· | ···················· |
| 15330 | MW035964.1 | \|Severe acute respiratory syndrome coro | ···················· | ···················· | ···················· | ···················· | ···················· | ···················· |
| 15331 | MW035965.1 | \|Severe acute respiratory syndrome coro | ···················· | ···················· | ···················· | ···················· | ···················· | ···················· |
| 15332 | MW035966.1 | \|Severe acute respiratory syndrome coro | ···················· | ···················· | ···················· | ···················· | ···················· | ···················· |
| 15333 | MW035967.1 | \|Severe acute respiratory syndrome coro | ···················· | ···················· | ···················· | ···················· | ···················· | ···················· |
| 15334 | MW035968.1 | \|Severe acute respiratory syndrome coro | ···················· | ···················· | ···················· | ···················· | ···················· | ···················· |
| 15335 | MW035970.1 | \|Severe acute respiratory syndrome coro | ···················· | ···················· | ···················· | ···················· | ···················· | ···················· |
| 15336 | MW035971.1 | \|Severe acute respiratory syndrome coro | ···················· | ···················· | ···················· | ···················· | ···················· | ···················· |
| 15337 | MW035972.1 | \|Severe acute respiratory syndrome coro | ···················· | ···················· | ···················· | ···················· | ···················· | ···················· |
| 15338 | MW035973.1 | \|Severe acute respiratory syndrome coro | ···················· | ···················· | ···················· | ···················· | ···················· | ···················· |
| 15339 | MW035974.1 | \|Severe acute respiratory syndrome coro | ···················· | ···················· | ···················· | ···················· | ···················· | ···················· |
| 15340 | MW035976.1 | \|Severe acute respiratory syndrome coro | ···················· | ···················· | ···················· | ···················· | ···················· | ···················· |
| 15341 | MW035984.1 | \|Severe acute respiratory syndrome coro | ···················· | ···················· | ···················· | ···················· | ···················· | ···················· |
| 15342 | MW035987.1 | \|Severe acute respiratory syndrome coro | ···················· | ···················· | ···················· | ···················· | ···················· | ···················· |
| 15343 | MW035988.1 | \|Severe acute respiratory syndrome coro | ···················· | ···················· | ···················· | ···················· | ···················· | ···················· |

| | | | | | | | |
|---|---|---|---|---|---|---|---|
| 15344 MW035990.1 | Severe acute respiratory syndrome coro | ···················· | ···················· | ···················· | ···················· | ···················· | ···················· |
| 15346 MW035997.1 | Severe acute respiratory syndrome coro | ···················· | ···················· | ···················· | ···················· | ···················· | ···················· |
| 15347 MW035999.1 | Severe acute respiratory syndrome coro | ···················· | ···················· | ···················· | ···················· | ···················· | ···················· |
| 15348 MW036000.1 | Severe acute respiratory syndrome coro | ···················· | ···················· | ···················· | ···················· | ···················· | ···················· |
| 15349 MW036003.1 | Severe acute respiratory syndrome coro | ···················· | ···················· | ···················· | ···················· | ···················· | ···················· |
| 15350 MW036006.1 | Severe acute respiratory syndrome coro | ···················· | ···················· | ···················· | ···················· | ···················· | ···················· |
| 15351 MW036007.1 | Severe acute respiratory syndrome coro | ···················· | ·A·················· | ···················· | ···················· | ···················· | ···················· |
| 15352 MW036008.1 | Severe acute respiratory syndrome coro | ···················· | ···················· | ···················· | ···················· | ···················· | ···················· |
| 15353 MW036010.1 | Severe acute respiratory syndrome coro | ···················· | ···················· | ···················· | ···················· | ···················· | ···················· |
| 15354 MW036011.1 | Severe acute respiratory syndrome coro | ···················· | ···················· | ···················· | ···················· | ···················· | ···················· |
| 15355 MW036013.1 | Severe acute respiratory syndrome coro | ···················· | ···················· | ···················· | ···················· | ···················· | ···················· |
| 15356 MW036014.1 | Severe acute respiratory syndrome coro | ···················· | ···················· | ···················· | ···················· | ···················· | ···················· |
| 15357 MW036016.1 | Severe acute respiratory syndrome coro | ···················· | ···················· | ···················· | ···················· | ···················· | ···················· |
| 15358 MW036017.1 | Severe acute respiratory syndrome coro | ···················· | ···················· | ···················· | ···················· | ···················· | ···················· |
| 15359 MW036018.1 | Severe acute respiratory syndrome coro | ···················· | ···················· | ···················· | ···················· | ···················· | ···················· |
| 15360 MW036019.1 | Severe acute respiratory syndrome coro | ···················· | ···················· | ···················· | ···················· | ···················· | ···················· |
| 15361 MW036020.1 | Severe acute respiratory syndrome coro | ···················· | ···················· | ···················· | ···················· | ···················· | ···················· |
| 15362 MW036021.1 | Severe acute respiratory syndrome coro | ···················· | ···················· | ···················· | ···················· | ···················· | ···················· |
| 15363 MW036022.1 | Severe acute respiratory syndrome coro | ···················· | ···················· | ···················· | ···················· | ···················· | ···················· |
| 15364 MW036023.1 | Severe acute respiratory syndrome coro | ···················· | ···················· | ···················· | ···················· | ···················· | ···················· |
| 15365 MW036024.1 | Severe acute respiratory syndrome coro | ···················· | ···················· | ···················· | ···················· | ···················· | ···················· |
| 15366 MW036026.1 | Severe acute respiratory syndrome coro | ···················· | ···················· | ···················· | ···················· | ···················· | ···················· |
| 15367 MW036027.1 | Severe acute respiratory syndrome coro | ···················· | ·A·················· | ···················· | ···················· | ···················· | ···················· |
| 15368 MW036028.1 | Severe acute respiratory syndrome coro | ···················· | ···················· | ···················· | ···················· | ···················· | ···················· |
| 15369 MW036029.1 | Severe acute respiratory syndrome coro | ···················· | ···················· | ···················· | ···················· | ···················· | ···················· |
| 15370 MW036031.1 | Severe acute respiratory syndrome coro | ···················· | ···················· | ···················· | ···················· | ···················· | ···················· |
| 15371 MW036032.1 | Severe acute respiratory syndrome coro | ···················· | ···················· | ···················· | ···················· | ···················· | ···················· |
| 15372 MW036034.1 | Severe acute respiratory syndrome coro | ···················· | ·A·················· | ···················· | ···················· | ···················· | ···················· |
| 15373 MW036035.1 | Severe acute respiratory syndrome coro | ···················· | ···················· | ···················· | ···················· | ···················· | ···················· |
| 15374 MW036038.1 | Severe acute respiratory syndrome coro | ···················· | ···················· | ···················· | ···················· | ···················· | ···················· |
| 15375 MW036039.1 | Severe acute respiratory syndrome coro | ···················· | ···················· | ···················· | ···················· | ···················· | ···················· |
| 15376 MW036041.1 | Severe acute respiratory syndrome coro | ···················· | ···················· | ···················· | ···················· | ···················· | ···················· |
| 15377 MW036042.1 | Severe acute respiratory syndrome coro | ···················· | ···················· | ···················· | ···················· | ···················· | ···················· |
| 15378 MW036043.1 | Severe acute respiratory syndrome coro | ···················· | ···················· | ···················· | ···················· | ···················· | ···················· |
| 15379 MW036044.1 | Severe acute respiratory syndrome coro | ···················· | ·A·················· | ···················· | ···················· | ···················· | ···················· |
| 15380 MW036046.1 | Severe acute respiratory syndrome coro | ···················· | ···················· | ···················· | ···················· | ···················· | ···················· |
| 15381 MW036047.1 | Severe acute respiratory syndrome coro | ···················· | ···················· | ···················· | ···················· | ···················· | ···················· |
| 15382 MW036049.1 | Severe acute respiratory syndrome coro | ···················· | ·A·················· | ···················· | ···················· | ···················· | ···················· |
| 15383 MW036050.1 | Severe acute respiratory syndrome coro | ···················· | ···················· | ···················· | ···················· | ···················· | ···················· |
| 15384 MW036051.1 | Severe acute respiratory syndrome coro | ···················· | ···················· | ···················· | ···················· | ···················· | ···················· |
| 15385 MW036053.1 | Severe acute respiratory syndrome coro | ···················· | ···················· | ···················· | ···················· | ···················· | ···················· |
| 15386 MW036054.1 | Severe acute respiratory syndrome coro | ···················· | ···················· | ···················· | ···················· | ···················· | ···················· |
| 15387 MW036055.1 | Severe acute respiratory syndrome coro | ···················· | ···················· | ···················· | ···················· | ···················· | ···················· |
| 15388 MW036056.1 | Severe acute respiratory syndrome coro | ···················· | ···················· | ···················· | ···················· | ···················· | ···················· |
| 15389 MW036057.1 | Severe acute respiratory syndrome coro | ···················· | ···················· | ···················· | ···················· | ···················· | ···················· |
| 15390 MW036060.1 | Severe acute respiratory syndrome coro | ···················· | ·A·················· | ···················· | ···················· | ···················· | ···················· |
| 15391 MW036062.1 | Severe acute respiratory syndrome coro | ···················· | ···················· | ···················· | ···················· | ···················· | ···················· |
| 15392 MW036063.1 | Severe acute respiratory syndrome coro | ···················· | ·A·················· | ···················· | ···················· | ···················· | ···················· |
| 15393 MW036067.1 | Severe acute respiratory syndrome coro | ···················· | ···················· | ···················· | ···················· | ···················· | ···················· |
| 15394 MW036068.1 | Severe acute respiratory syndrome coro | ···················· | ···················· | ···················· | ···················· | ···················· | ···················· |
| 15395 MW036069.1 | Severe acute respiratory syndrome coro | ···················· | ·A·················· | ···················· | ···················· | ···················· | ···················· |
| 15396 MW036070.1 | Severe acute respiratory syndrome coro | ···················· | ···················· | ···················· | ···················· | ···················· | ···················· |
| 15397 MW036071.1 | Severe acute respiratory syndrome coro | ···················· | ···················· | ···················· | ···················· | ···················· | ···················· |
| 15398 MW036072.1 | Severe acute respiratory syndrome coro | ···················· | ·A·················· | ···················· | ···················· | ···················· | ···················· |
| 15399 MW036073.1 | Severe acute respiratory syndrome coro | ···················· | ···················· | ···················· | ···················· | ···················· | ···················· |
| 15400 MW036074.1 | Severe acute respiratory syndrome coro | ···················· | ·A·················· | ···················· | ···················· | ···················· | ···················· |
| 15401 MW036080.1 | Severe acute respiratory syndrome coro | ···················· | ···················· | ···················· | ···················· | ···················· | ···················· |
| 15402 MW036081.1 | Severe acute respiratory syndrome coro | ···················· | ···················· | ···················· | ···················· | ···················· | ···················· |
| 15403 MW036083.1 | Severe acute respiratory syndrome coro | ···················· | ·A·················· | ···················· | ···················· | ···················· | ···················· |
| 15404 MW036084.1 | Severe acute respiratory syndrome coro | ···················· | ···················· | ···················· | ···················· | ···················· | ···················· |
| 15405 MW036086.1 | Severe acute respiratory syndrome coro | ···················· | ···················· | ···················· | ···················· | ···················· | ···················· |
| 15406 MW030193.1 | Severe acute respiratory syndrome coro | ···················· | ···················· | ···················· | ···················· | ···················· | ···················· |
| 15407 MW030194.1 | Severe acute respiratory syndrome coro | ···················· | ···················· | ···················· | ···················· | ···················· | ···················· |
| 15408 MW030195.1 | Severe acute respiratory syndrome coro | ···················· | ···················· | ···················· | ···················· | ···················· | ···················· |
| 15409 MW030196.1 | Severe acute respiratory syndrome coro | ···················· | ···················· | ···················· | ···················· | ···················· | ···················· |
| 15410 MW030197.1 | Severe acute respiratory syndrome coro | ···················· | ···················· | ···················· | ···················· | ···················· | ···················· |
| 15411 MW030198.1 | Severe acute respiratory syndrome coro | ···················· | ···················· | ···················· | ···················· | ···················· | ···················· |
| 15412 MW030199.1 | Severe acute respiratory syndrome coro | ···················· | ···················· | ···················· | ···················· | ···················· | ···················· |
| 15413 MW030200.1 | Severe acute respiratory syndrome coro | ···················· | ···················· | ···················· | ···················· | ···················· | ···················· |
| 15414 MW030201.1 | Severe acute respiratory syndrome coro | ···················· | ···················· | ···················· | ···················· | ···················· | ···················· |
| 15415 MW030202.1 | Severe acute respiratory syndrome coro | ···················· | ···················· | ···················· | ···················· | ···················· | ···················· |
| 15416 MW030203.1 | Severe acute respiratory syndrome coro | ···················· | ···················· | ···················· | ···················· | ···················· | ···················· |
| 15417 MW030204.1 | Severe acute respiratory syndrome coro | ···················· | ···················· | ···················· | ···················· | ···················· | ···················· |
| 15418 MW030205.1 | Severe acute respiratory syndrome coro | ···················· | ···················· | ···················· | ···················· | ···················· | ···················· |
| 15419 MW030206.1 | Severe acute respiratory syndrome coro | ···················· | ···················· | ···················· | ···················· | ···················· | ···················· |
| 15420 MW030207.1 | Severe acute respiratory syndrome coro | ···················· | ···················· | ···················· | ···················· | ···················· | ···················· |
| 15421 MW030208.1 | Severe acute respiratory syndrome coro | ···················· | ···················· | ···················· | ···················· | ···················· | ···················· |
| 15422 MW030209.1 | Severe acute respiratory syndrome coro | ···················· | ···················· | ···················· | ···················· | ···················· | ···················· |

```
15423 MW030210.1 |Severe acute respiratory syndrome coro .................... .................... .................... .................... .................... ....................
15424 MW030211.1 |Severe acute respiratory syndrome coro .................... .................... .................... .................... .................... ....................
15425 MW030212.1 |Severe acute respiratory syndrome coro .................... .................... .................... .................... .................... ....................
15426 MW030213.1 |Severe acute respiratory syndrome coro .................... .................... .................... .................... .................... ....................
15427 MW030214.1 |Severe acute respiratory syndrome coro .................... .................... .................... .................... .................... ....................
15428 MW030215.1 |Severe acute respiratory syndrome coro .................... .................... .................... .................... .................... ....................
15429 MW030216.1 |Severe acute respiratory syndrome coro .................... .................... .................... .................... .................... ....................
15430 MW030217.1 |Severe acute respiratory syndrome coro .................... .................... .................... .................... .................... ....................
15431 MW030218.1 |Severe acute respiratory syndrome coro .................... .................... .................... .................... .................... ....................
15432 MW030219.1 |Severe acute respiratory syndrome coro .................... .................... .................... .................... .................... ....................
15433 MW030220.1 |Severe acute respiratory syndrome coro .................... .................... .................... .................... .................... ....................
15434 MW030221.1 |Severe acute respiratory syndrome coro .................... .................... .................... .................... .................... ....................
15435 MW030222.1 |Severe acute respiratory syndrome coro .................... .................... .................... .................... .................... ....................
15436 MW030223.1 |Severe acute respiratory syndrome coro .................... .................... .................... .................... .................... ....................
15437 MW030224.1 |Severe acute respiratory syndrome coro .................... .................... .................... .................... .................... ....................
15438 MW030225.1 |Severe acute respiratory syndrome coro .................... .................... .................... .................... .................... ....................
15439 MW030226.1 |Severe acute respiratory syndrome coro .................... .................... .................... .................... .................... ....................
15440 MW030227.1 |Severe acute respiratory syndrome coro .................... .................... .................... .................... .................... ....................
15441 MW030228.1 |Severe acute respiratory syndrome coro .................... .................... .................... .................... .................... ....................
15442 MW030229.1 |Severe acute respiratory syndrome coro .................... .................... .................... .................... .................... ....................
15443 MW030230.1 |Severe acute respiratory syndrome coro .................... .................... .................... .................... .................... ....................
15444 MW030231.1 |Severe acute respiratory syndrome coro .................... .................... .................... .................... .................... ....................
15445 MW030232.1 |Severe acute respiratory syndrome coro .................... .................... .................... .................... .................... ....................
15446 MW030233.1 |Severe acute respiratory syndrome coro .................... .................... .................... .................... .................... ....................
15447 MW030234.1 |Severe acute respiratory syndrome coro .................... .................... .................... .................... .................... ....................
15448 MW030235.1 |Severe acute respiratory syndrome coro .................... .................... .................... .................... .................... ....................
15449 MW030236.1 |Severe acute respiratory syndrome coro .................... .................... .................... .................... .................... ....................
15450 MW030237.1 |Severe acute respiratory syndrome coro .................... .................... .................... .................... .................... ....................
15451 MW030238.1 |Severe acute respiratory syndrome coro .................... .................... .................... .................... .................... ....................
15452 MW030239.1 |Severe acute respiratory syndrome coro .................... .................... .................... .................... .................... ....................
15453 MW030240.1 |Severe acute respiratory syndrome coro .................... .................... .................... .................... .................... ....................
15454 MW030241.1 |Severe acute respiratory syndrome coro .................... .................... .................... .................... .................... ....................
15455 MW030242.1 |Severe acute respiratory syndrome coro .................... .................... .................... .................... .................... ....................
15456 MW030243.1 |Severe acute respiratory syndrome coro .................... .................... .................... .................... .................... ....................
15457 MW030244.1 |Severe acute respiratory syndrome coro .................... .................... .................... .................... .................... ....................
15458 MW030245.1 |Severe acute respiratory syndrome coro .................... .................... .................... .................... .................... ....................
15459 MW030246.1 |Severe acute respiratory syndrome coro .................... .................... .................... .................... .................... ....................
15460 MW030247.1 |Severe acute respiratory syndrome coro .................... .................... .................... .................... .................... ....................
15461 MW030248.1 |Severe acute respiratory syndrome coro .................... .................... .................... .................... .................... ....................
15462 MW030249.1 |Severe acute respiratory syndrome coro .................... .................... .................... .................... .................... ....................
15463 MW030250.1 |Severe acute respiratory syndrome coro .................... .................... .................... .................... .................... ....................
15464 MW030251.1 |Severe acute respiratory syndrome coro .................... .................... .................... .................... .................... ....................
15465 MW030252.1 |Severe acute respiratory syndrome coro .................... .................... .................... .................... .................... ....................
15466 MW030253.1 |Severe acute respiratory syndrome coro .................... .................... .................... .................... .................... ....................
15467 MW030254.1 |Severe acute respiratory syndrome coro .................... .................... .................... .................... .................... ....................
15468 MW030255.1 |Severe acute respiratory syndrome coro .................... .................... .................... .................... .................... ....................
15469 MW030256.1 |Severe acute respiratory syndrome coro .................... .................... .................... .................... .................... ....................
15470 MW030257.1 |Severe acute respiratory syndrome coro .................... .................... .................... .................... .................... ....................
15471 MW030258.1 |Severe acute respiratory syndrome coro .................... .................... .................... .................... .................... ....................
15472 MW030259.1 |Severe acute respiratory syndrome coro .................... .................... .................... .................... .................... ....................
15473 MW030260.1 |Severe acute respiratory syndrome coro .................... .................... .................... .................... .................... ....................
15474 MW030261.1 |Severe acute respiratory syndrome coro .................... .................... .................... .................... .................... ....................
15475 MW030262.1 |Severe acute respiratory syndrome coro .................... .................... .................... .................... .................... ....................
15476 MW030263.1 |Severe acute respiratory syndrome coro .................... .................... .................... .................... .................... ....................
15477 MW030264.1 |Severe acute respiratory syndrome coro .................... .................... .................... .................... .................... ....................
15478 MW030265.1 |Severe acute respiratory syndrome coro .................... .................... .................... .................... .................... ....................
15479 MW030266.1 |Severe acute respiratory syndrome coro .................... .................... .................... .................... .................... ....................
15480 MW030267.1 |Severe acute respiratory syndrome coro .................... .................... .................... .................... .................... ....................
15481 MW030268.1 |Severe acute respiratory syndrome coro .................... .................... .................... .................... .................... ....................
15482 MW030269.1 |Severe acute respiratory syndrome coro .................... .................... .................... .................... .................... ....................
15483 MW030270.1 |Severe acute respiratory syndrome coro .................... .................... .................... .................... .................... ...................N 1
15484 MW030271.1 |Severe acute respiratory syndrome coro .................... .................... .................... .................... .................... ....................
15485 MW030272.1 |Severe acute respiratory syndrome coro .................... .................... .................... .................... .................... ....................
15486 MW030273.1 |Severe acute respiratory syndrome coro .................... .................... .................... .................... .................... ....................
15487 MW030274.1 |Severe acute respiratory syndrome coro .................... .................... .................... .................... .................... ....................
15488 MW030275.1 |Severe acute respiratory syndrome coro .................... .................... .................... .................... .................... ....................
15489 MW030276.1 |Severe acute respiratory syndrome coro .................... .................... .................... .................... .................... ....................
15490 MW030277.1 |Severe acute respiratory syndrome coro .................... .................... .................... .................... .................... ....................
15491 MW030278.1 |Severe acute respiratory syndrome coro .................... .................... .................... .................... .................... ....................
15492 MW030985.1 |Severe acute respiratory syndrome coro .................... .................... .................... .................... .................... ....................
15493 MW030988.1 |Severe acute respiratory syndrome coro .................... .................... .................... .................... .................... ....................
15494 MW030989.1 |Severe acute respiratory syndrome coro .................... .............A...... .................... .................... .................... ....................
15495 MW030993.1 |Severe acute respiratory syndrome coro .................... .................... .................... .................... .................... ....................
15496 MW030994.1 |Severe acute respiratory syndrome coro .................... .................... .................... .................... .................... ....................
15497 MW030996.1 |Severe acute respiratory syndrome coro .................... .................... .................... .................... .................... ....................
15498 MW030998.1 |Severe acute respiratory syndrome coro .................... .................... .................... .................... .................... ....................
15499 MW031000.1 |Severe acute respiratory syndrome coro .................... .................... .................... .................... .................... ....................
15500 MW031001.1 |Severe acute respiratory syndrome coro .................... .................... .................... .................... .................... ....................
```

```
15501 MW031003.1 |Severe acute respiratory syndrome coro   ....................  ....................  ....................  ....................  ....................  ....................
15502 MW031004.1 |Severe acute respiratory syndrome coro   ....................  ....................  ....................  ....................  ....................  ....................
15503 MW031009.1 |Severe acute respiratory syndrome coro   ....................  ....................  ....................  ....................  ....................  ....................
15504 MW031010.1 |Severe acute respiratory syndrome coro   ....................  ....................  ....................  ....................  ....................  ....................
15505 MW031011.1 |Severe acute respiratory syndrome coro   ....................  ....................  ....................  ....................  ....................  ....................
15506 MW031014.1 |Severe acute respiratory syndrome coro   ....................  ....................  ....................  ....................  ....................  ....................
15507 MW031015.1 |Severe acute respiratory syndrome coro   ....................  ....................  ....................  ....................  ....................  ....................
15508 MW031016.1 |Severe acute respiratory syndrome coro   ....................  .A..................  ....................  ....................  ....................  ....................
15509 MW031017.1 |Severe acute respiratory syndrome coro   ....................  ....................  ....................  ....................  ....................  ....................
15510 MW031018.1 |Severe acute respiratory syndrome coro   ....................  ....................  ....................  ....................  ....................  ....................
15511 MW031021.1 |Severe acute respiratory syndrome coro   ....................  ....................  ....................  ....................  ....................  ....................
15512 MW031022.1 |Severe acute respiratory syndrome coro   ....................  ....................  ....................  ....................  ....................  ....................
15513 MW031023.1 |Severe acute respiratory syndrome coro   ....................  ....................  ....................  ....................  ....................  ....................
15514 MW031025.1 |Severe acute respiratory syndrome coro   ....................  .A..................  ....................  ....................  ....................  ....................
15515 MW031027.1 |Severe acute respiratory syndrome coro   ....................  ....................  ....................  ....................  ....................  ....................
15516 MW031028.1 |Severe acute respiratory syndrome coro   ....................  ....................  ....................  ....................  ....................  ....................
15517 MW031029.1 |Severe acute respiratory syndrome coro   ....................  ....................  ....................  ....................  ....................  ....................
15518 MW031030.1 |Severe acute respiratory syndrome coro   ....................  ....................  ....................  ....................  ....................  ....................
15519 MW031033.1 |Severe acute respiratory syndrome coro   ....................  ....................  ....................  ....................  ....................  ....................
15520 MW031034.1 |Severe acute respiratory syndrome coro   ....................  ....................  ....................  ....................  ....................  ....................
15521 MW031035.1 |Severe acute respiratory syndrome coro   ....................  ....................  ....................  ....................  ....................  ....................
15522 MW031037.1 |Severe acute respiratory syndrome coro   ....................  ....................  ....................  ....................  ....................  ....................
15523 MW031038.1 |Severe acute respiratory syndrome coro   ....................  ....................  ....................  ....................  ....................  ....................
15524 MW031039.1 |Severe acute respiratory syndrome coro   ....................  ....................  ....................  ....................  ....................  ....................
15525 MW031040.1 |Severe acute respiratory syndrome coro   ....................  ....................  ....................  ....................  ....................  ....................
15526 MW031043.1 |Severe acute respiratory syndrome coro   ....................  ....................  ....................  ....................  ....................  ....................
15527 MW031044.1 |Severe acute respiratory syndrome coro   ....................  ....................  ....................  ....................  ....................  ....................
15528 MW031045.1 |Severe acute respiratory syndrome coro   ....................  ....................  ....................  ....................  ....................  ....................
15529 MW031046.1 |Severe acute respiratory syndrome coro   ....................  ....................  ....................  ....................  ....................  ....................
15530 MW031047.1 |Severe acute respiratory syndrome coro   ....................  ....................  ....................  ....................  ....................  ....................
15531 MW031048.1 |Severe acute respiratory syndrome coro   ....................  ....................  ....................  ....................  ....................  ....................
15532 MW031049.1 |Severe acute respiratory syndrome coro   ....................  ....................  ....................  ....................  ....................  ....................
15533 MW031050.1 |Severe acute respiratory syndrome coro   ....................  ....................  ....................  ....................  ....................  ....................
15534 MW031051.1 |Severe acute respiratory syndrome coro   ....................  ....................  ....................  ....................  ....................  ....................
15535 MW031052.1 |Severe acute respiratory syndrome coro   ....................  ....................  ....................  ....................  ....................  ....................
15536 MW031053.1 |Severe acute respiratory syndrome coro   ....................  ....................  ....................  ....................  ....................  ....................
15537 MW031054.1 |Severe acute respiratory syndrome coro   ....................  ....................  ....................  ....................  ....................  ....................
15538 MW031055.1 |Severe acute respiratory syndrome coro   ....................  .A..................  ....................  ....................  ....................  ....................
15539 MW031056.1 |Severe acute respiratory syndrome coro   ....................  ....................  ....................  ....................  ....................  ....................
15540 MW031058.1 |Severe acute respiratory syndrome coro   ....................  ....................  ....................  ....................  ....................  ....................
15541 MW031059.1 |Severe acute respiratory syndrome coro   ....................  ....................  ....................  ....................  ....................  ....................
15542 MW031060.1 |Severe acute respiratory syndrome coro   ....................  ....................  ....................  ....................  ....................  ....................
15543 MW031061.1 |Severe acute respiratory syndrome coro   ....................  ....................  ....................  ....................  ....................  ....................
15544 MW031063.1 |Severe acute respiratory syndrome coro   ....................  ....................  ....................  ....................  ....................  ....................
15545 MW031064.1 |Severe acute respiratory syndrome coro   ....................  ....................  ....................  ....................  ....................  ....................
15546 MW031065.1 |Severe acute respiratory syndrome coro   ....................  ....................  ....................  ....................  ....................  ....................
15547 MW031066.1 |Severe acute respiratory syndrome coro   ....................  ....................  ....................  ....................  ....................  ....................
15548 MW031067.1 |Severe acute respiratory syndrome coro   ....................  ....................  ....................  ....................  ....................  ....................
15549 MW031068.1 |Severe acute respiratory syndrome coro   ....................  ....................  ....................  ....................  ....................  ....................
15550 MW031069.1 |Severe acute respiratory syndrome coro   ....................  ....................  ....................  ....................  ....................  ....................
15551 MW031070.1 |Severe acute respiratory syndrome coro   ....................  ....................  ....................  ....................  ....................  ....................
15552 MW031071.1 |Severe acute respiratory syndrome coro   ....................  ....................  ....................  ....................  ....................  ....................
15553 MW031072.1 |Severe acute respiratory syndrome coro   ....................  ....................  ....................  ....................  ....................  ....................
15554 MW031073.1 |Severe acute respiratory syndrome coro   ....................  ....................  ....................  ....................  ....................  ....................
15555 MW031074.1 |Severe acute respiratory syndrome coro   ....................  ....................  ....................  ....................  ....................  ....................
15556 MW031077.1 |Severe acute respiratory syndrome coro   ....................  ....................  ....................  ....................  ...TT...............  ....................
15557 MW031078.1 |Severe acute respiratory syndrome coro   ....................  ....................  ....................  ....................  ....................  ....................
15558 MW031079.1 |Severe acute respiratory syndrome coro   ....................  ....................  ....................  ....................  ....................  ....................
15559 MW031080.1 |Severe acute respiratory syndrome coro   ....................  ....................  ....................  ....................  ....................  ....................
15560 MW031082.1 |Severe acute respiratory syndrome coro   ....................  ....................  ....................  ....................  ....................  ....................
15561 MW031084.1 |Severe acute respiratory syndrome coro   ....................  .A..................  ....................  ....................  ....................  ....................
15562 MW031086.1 |Severe acute respiratory syndrome coro   ....................  ....................  ....................  ....................  ....................  ....................
15563 MW031088.1 |Severe acute respiratory syndrome coro   ....................  ....................  ....................  ....................  ....................  ....................
15564 MW031089.1 |Severe acute respiratory syndrome coro   ....................  ....................  ....................  ....................  ....................  ....................
15565 MW031090.1 |Severe acute respiratory syndrome coro   ....................  ....................  ....................  ....................  ....................  ....................
15566 MW031091.1 |Severe acute respiratory syndrome coro   ....................  ....................  ....................  ....................  ....................  ....................
15567 MW031092.1 |Severe acute respiratory syndrome coro   ....................  ....................  ....................  ....................  ....................  ....................
15568 MW031093.1 |Severe acute respiratory syndrome coro   ....................  ....................  ....................  ....................  ....................  ....................
15569 MW031799.1 |Severe acute respiratory syndrome coro   ....................  ....................  ....................  ....................  ....................  ....................
15570 MW031800.1 |Severe acute respiratory syndrome coro   ....................  ....................  ....................  ....................  ....................  ....................
15571 MW031801.1 |Severe acute respiratory syndrome coro   ....................  ....................  ....................  ....................  ....................  ....................
15572 MW031802.1 |Severe acute respiratory syndrome coro   ....................  ....................  ....................  ....................  ....................  ....................
15573 MW031803.1 |Severe acute respiratory syndrome coro   ....................  ....................  ....................  ....................  ....................  ....................
15574 MT577009.1 |Severe acute respiratory syndrome coro   ....................  ....................  ....................  ....................  ....................  ....................
15575 MT577010.1 |Severe acute respiratory syndrome coro   ....................  ....................  ....................  ....................  ....................  ....................
15576 MT577011.1 |Severe acute respiratory syndrome coro   ....................  ....................  ....................  ....................  ....................  ....................
15577 MT577012.1 |Severe acute respiratory syndrome coro   ....................  ....................  ....................  ....................  ....................  ....................
15578 MT577013.1 |Severe acute respiratory syndrome coro   ....................  ....................  ....................  ....................  ....................  ....................
```

```
15579 MT577014.1 |Severe acute respiratory syndrome coro    ····················  ····················  ····················  ····················  ····················  ····················
15580 MT577015.1 |Severe acute respiratory syndrome coro    ····················  ····················  ····················  ····················  ····················  ····················
15581 MW020061.1 |Severe acute respiratory syndrome coro    ····················  ····················  ····················  ····················  ····················  ····················
15582 MW020062.1 |Severe acute respiratory syndrome coro    ····················  ····················  ····················  ····················  ····················  ····················
15583 MW020063.1 |Severe acute respiratory syndrome coro    ····················  ····················  ····················  ····················  ····················  ····················
15584 MW020064.1 |Severe acute respiratory syndrome coro    ····················  ····················  ····················  ····················  ····················  ····················
15585 MW020065.1 |Severe acute respiratory syndrome coro    ····················  ····················  ····················  ····················  ····················  ····················
15586 MW020066.1 |Severe acute respiratory syndrome coro    ····················  ····················  ····················  ····················  ····················  ····················
15587 MW020067.1 |Severe acute respiratory syndrome coro    ····················  ····················  ····················  ····················  ····················  ····················
15588 MW020068.1 |Severe acute respiratory syndrome coro    ····················  ····················  ····················  ····················  ····················  ····················
15589 MW020069.1 |Severe acute respiratory syndrome coro    ····················  ····················  ····················  ····················  ····················  ····················
15590 MW020070.1 |Severe acute respiratory syndrome coro    ····················  ····················  ····················  ····················  ····················  ····················
15591 MW020071.1 |Severe acute respiratory syndrome coro    ····················  ····················  ····················  ····················  ····················  ····················
15592 MW020072.1 |Severe acute respiratory syndrome coro    ····················  ····················  ····················  ····················  ····················  ····················
15593 MW020073.1 |Severe acute respiratory syndrome coro    ····················  ····················  ····················  ····················  ····················  ····················
15594 MW020074.1 |Severe acute respiratory syndrome coro    ····················  ····················  ····················  ····················  ····················  ····················
15595 MW020075.1 |Severe acute respiratory syndrome coro    ····················  ····················  ····················  ····················  ····················  ····················
15596 MW020076.1 |Severe acute respiratory syndrome coro    ····················  ····················  ····················  ····················  ····················  ····················
15597 MW020077.1 |Severe acute respiratory syndrome coro    ····················  ····················  ····················  ····················  ····················  ····················
15598 MW020078.1 |Severe acute respiratory syndrome coro    ····················  ····················  ····················  ····················  ····················  ····················
15599 MW020079.1 |Severe acute respiratory syndrome coro    ····················  ·············A······  ····················  ····················  ····················  ····················
15600 MW020080.1 |Severe acute respiratory syndrome coro    ····················  ····················  ····················  ····················  ····················  ····················
15601 MW020081.1 |Severe acute respiratory syndrome coro    ····················  ····················  ····················  ····················  ····················  ····················
15602 MW020082.1 |Severe acute respiratory syndrome coro    ········T···········  ····················  ····················  ····················  ····················  ····················
15603 MW020083.1 |Severe acute respiratory syndrome coro    ····················  ····················  ····················  ····················  ····················  ····················
15604 MW020084.1 |Severe acute respiratory syndrome coro    ····················  ·A··················  ····················  ····················  ····················  ····················
15605 MW020085.1 |Severe acute respiratory syndrome coro    ····················  ····················  ····················  ····················  ····················  ····················
15606 MW020086.1 |Severe acute respiratory syndrome coro    ····················  ····················  ····················  ····················  ····················  ····················
15607 MW021454.1 |Severe acute respiratory syndrome coro    ····················  ····················  ····················  ····················  ····················  ····················
15608 MW021455.1 |Severe acute respiratory syndrome coro    ····················  ····················  ····················  ····················  ····················  ····················
15609 MW021456.1 |Severe acute respiratory syndrome coro    ····················  ····················  ····················  ····················  ····················  ····················
15610 MW021457.1 |Severe acute respiratory syndrome coro    ····················  ····················  ····················  ····················  ····················  ····················
15611 MW021458.1 |Severe acute respiratory syndrome coro    ····················  ·········A··········  ····················  ····················  ····················  ····················
15612 MW021459.1 |Severe acute respiratory syndrome coro    ····················  ····················  ····················  ····················  ····················  ····················
15613 MW021460.1 |Severe acute respiratory syndrome coro    ····················  ····················  ····················  ····················  ····················  ····················
15614 MW021461.1 |Severe acute respiratory syndrome coro    ····················  ·A··················  ····················  ····················  ····················  ····················
15615 MW021463.1 |Severe acute respiratory syndrome coro    ····················  ····················  ····················  ····················  ····················  ····················
15616 MW021464.1 |Severe acute respiratory syndrome coro    ····················  ····················  ····················  ····················  ····················  ····················
15617 MW021465.1 |Severe acute respiratory syndrome coro    ····················  ·A··················  ····················  ····················  ····················  ····················
15618 MW021466.1 |Severe acute respiratory syndrome coro    ····················  ····················  ····················  ····················  ····················  ····················
15619 MW021468.1 |Severe acute respiratory syndrome coro    ····················  ····················  ····················  ····················  ····················  ····················
15620 MW021469.1 |Severe acute respiratory syndrome coro    ····················  ····················  ····················  ····················  ····················  ····················
15621 MW021470.1 |Severe acute respiratory syndrome coro    ····················  ····················  ····················  ····················  ····················  ····················
15622 MW021512.1 |Severe acute respiratory syndrome coro    ····················  ····················  ····················  ····················  ····················  ····················
15623 MW021513.1 |Severe acute respiratory syndrome coro    ····················  ····················  ····················  ····················  ····················  ····················
15624 MW021514.1 |Severe acute respiratory syndrome coro    ····················  ····················  ····················  ····················  ····················  ····················
15625 MW023388.1 |Severe acute respiratory syndrome coro    ····················  ····················  ····················  ····················  ····················  ····················
15626 MW023389.1 |Severe acute respiratory syndrome coro    ····················  ····················  ····················  ····················  ····················  ····················
15627 MW023390.1 |Severe acute respiratory syndrome coro    ····················  ····················  ····················  ····················  ····················  ····················
15628 MW023391.1 |Severe acute respiratory syndrome coro    ····················  ····················  ····················  ····················  ····················  ····················
15629 MW023392.1 |Severe acute respiratory syndrome coro    ····················  ····················  ····················  ····················  ····················  ····················
15630 MW023393.1 |Severe acute respiratory syndrome coro    ····················  ····················  ····················  ····················  ····················  ····················
15631 MW023394.1 |Severe acute respiratory syndrome coro    ····················  ····················  ····················  ····················  ····················  ····················
15632 MW023404.1 |Severe acute respiratory syndrome coro    ····················  ····················  ····················  ····················  ····················  ····················
15633 MW023409.1 |Severe acute respiratory syndrome coro    ····················  ····················  ····················  ····················  ····················  ····················
15634 MW023415.1 |Severe acute respiratory syndrome coro    ····················  ····················  ····················  ····················  ····················  ····················
15635 MW023416.1 |Severe acute respiratory syndrome coro    ····················  ····················  ····················  ····················  ····················  ····················
15636 MW023418.1 |Severe acute respiratory syndrome coro    ····················  ····················  ····················  ····················  ····················  ····················
15637 MW023424.1 |Severe acute respiratory syndrome coro    ····················  ····················  ····················  ····················  ····················  ····················
15638 MW023428.1 |Severe acute respiratory syndrome coro    ····················  ····················  ····················  ····················  ····················  ····················
15639 MW023429.1 |Severe acute respiratory syndrome coro    ····················  ····················  ····················  ····················  ····················  ····················
15641 MW023436.1 |Severe acute respiratory syndrome coro    ····················  ····················  ····················  ····················  ····················  ····················
15642 MW023437.1 |Severe acute respiratory syndrome coro    ····················  ····················  ····················  ····················  ····················  ····················
15643 MW023439.1 |Severe acute respiratory syndrome coro    ····················  ····················  ····················  ····················  ····················  ····················
15644 MW023440.1 |Severe acute respiratory syndrome coro    ····················  ····················  ····················  ····················  ····················  ····················
15645 MW023442.1 |Severe acute respiratory syndrome coro    ····················  ····················  ····················  ····················  ····················  ····················
15646 MW023443.1 |Severe acute respiratory syndrome coro    ····················  ····A···············  ····················  ····················  ····················  ····················
15647 MW023446.1 |Severe acute respiratory syndrome coro    ····················  ····················  ····················  ····················  ····················  ····················
15648 MW023447.1 |Severe acute respiratory syndrome coro    ····················  ····················  ····················  ····················  ····················  ····················
15649 MW023448.1 |Severe acute respiratory syndrome coro    ····················  ····················  ····················  ····················  ····················  ····················
15650 MW023449.1 |Severe acute respiratory syndrome coro    ····················  ····················  ····················  ····················  ····················  ····················
15651 MW023450.1 |Severe acute respiratory syndrome coro    ····················  ····················  ····················  ····················  ····················  ····················
15652 MW023452.1 |Severe acute respiratory syndrome coro    ····················  ····················  ····················  ····················  ····················  ····················
15653 MW023453.1 |Severe acute respiratory syndrome coro    ····················  ····················  ····················  ····················  ····················  ····················
15654 MW023456.1 |Severe acute respiratory syndrome coro    ····················  ····················  ····················  ····················  ····················  ····················
15655 MW023457.1 |Severe acute respiratory syndrome coro    ····················  ····················  ····················  ····················  ····················  ····················
15657 MW023460.1 |Severe acute respiratory syndrome coro    ····················  ····················  ····················  ····················  ····················  ····················
15658 MW023461.1 |Severe acute respiratory syndrome coro    ····················  ····················  ····················  ····················  ····················  ····················
```

```
15659 MW023462.1 |Severe acute respiratory syndrome coro .................... .................... .................... .................... .................... ....................
15660 MW023466.1 |Severe acute respiratory syndrome coro .................... .............A...... .................... .................... .................... ....................
15661 MW023470.1 |Severe acute respiratory syndrome coro .................... .................... .................... .................... .................... ....................
15662 MW023471.1 |Severe acute respiratory syndrome coro .................... .................... .................... .................... .................... ....................
15663 MW023474.1 |Severe acute respiratory syndrome coro .................... .................... .................... .................... .................... ....................
15664 MW023476.1 |Severe acute respiratory syndrome coro .................... .................... .................... .................... .................... ....................
15665 MW023478.1 |Severe acute respiratory syndrome coro .................... .................... .................... .................... .................... ....................
15667 MW023482.1 |Severe acute respiratory syndrome coro .................... .................... .................... .................... .................... ....................
15669 MW023486.1 |Severe acute respiratory syndrome coro .................... .................... .................... .................... .................... ....................
15670 MW023487.1 |Severe acute respiratory syndrome coro .................... .................... .................... .................... .................... ....................
15671 MW023494.1 |Severe acute respiratory syndrome coro .................... .................... .................... .................... .................... ....................
15672 MW023495.1 |Severe acute respiratory syndrome coro .................... .................... .................... .................... .................... ....................
15673 MW023498.1 |Severe acute respiratory syndrome coro .................... .................... .................... .................... .................... ....................
15674 MW023500.1 |Severe acute respiratory syndrome coro .................... .................... .................... .................... .................... ....................
15675 MW023501.1 |Severe acute respiratory syndrome coro .................... .................... .................... .................... .................... ....................
15676 MW023502.1 |Severe acute respiratory syndrome coro .................... .................... .................... .................... .................... ....................
15677 MW023511.1 |Severe acute respiratory syndrome coro .................... .................... .................... .................... .................... ....................
15678 MW023516.1 |Severe acute respiratory syndrome coro .................... .................... .................... .................... .................... ....................
15679 MW023517.1 |Severe acute respiratory syndrome coro .................... .................... .................... .................... .................... ....................
15680 MW023522.1 |Severe acute respiratory syndrome coro .................... .................... .................... .................... .................... ....................
15682 MW023525.1 |Severe acute respiratory syndrome coro .................... .................... .................... .................... .................... ....................
15684 MW023542.1 |Severe acute respiratory syndrome coro .................... .................... .................... .................... .................... ....................
15685 MW023547.1 |Severe acute respiratory syndrome coro .................... .................... .................... .................... .................... ....................
15686 MW023550.1 |Severe acute respiratory syndrome coro .................... .................... .................... .................... .................... ....................
15687 MW023552.1 |Severe acute respiratory syndrome coro .................... .................... .................... .................... .................... ....................
15691 MW023564.1 |Severe acute respiratory syndrome coro .................... .................... .................... .................... .................... ....................
15692 MW023570.1 |Severe acute respiratory syndrome coro ..........T......... .................... .................... .................... .................... ....................
15693 MW023576.1 |Severe acute respiratory syndrome coro .................... .................... .................... .................... .................... ....................
15694 MW023579.1 |Severe acute respiratory syndrome coro .................... .................... .................... .................... .................... ....................
15695 MW023580.1 |Severe acute respiratory syndrome coro .................... .................... .................... .................... .................... ....................
15696 MW023584.1 |Severe acute respiratory syndrome coro .................... .................... .................... .................... .................... ....................
15697 MW023592.1 |Severe acute respiratory syndrome coro .................... .................... .................... .................... .................... ....................
15698 MW023935.1 |Severe acute respiratory syndrome coro .................... .................... .................... .................... .................... ....................
15699 MW018448.1 |Severe acute respiratory syndrome coro .................... .................... .................... .................... .................... ....................
15700 MW012266.1 |Severe acute respiratory syndrome coro .................... .................... .................... .................... .................... ....................
15701 MW012267.1 |Severe acute respiratory syndrome coro .................... .................... .................... .................... .................... ....................
15702 MW012268.1 |Severe acute respiratory syndrome coro .................... .................... .................... .................... .................... ....................
15703 MW012269.1 |Severe acute respiratory syndrome coro .................... .................... .................... .................... .................... ....................
15704 MW012270.1 |Severe acute respiratory syndrome coro .................... .................... .................... .................... .................... ....................
15705 MW012271.1 |Severe acute respiratory syndrome coro .................... .................... .................... .................... .................... ....................
15706 MW012272.1 |Severe acute respiratory syndrome coro .................... .................... .................... .................... .................... ....................
15707 MW012273.1 |Severe acute respiratory syndrome coro .................... .................... .................... .................... .................... ....................
15708 MW012274.1 |Severe acute respiratory syndrome coro .................... .................... .................... .................... .................... ....................
15709 MW012275.1 |Severe acute respiratory syndrome coro .................... .................... .................... .................... .................... ....................
15710 MW012276.1 |Severe acute respiratory syndrome coro .................... .................... .................... .................... .................... ....................
15711 MW012277.1 |Severe acute respiratory syndrome coro .................... .................... .................... .................... .................... ....................
15712 MW012278.1 |Severe acute respiratory syndrome coro .................... .................... .................... .................... .................... ....................
15713 MW012279.1 |Severe acute respiratory syndrome coro .................... .................... .................... .................... .................... ....................
15714 MW014808.1 |Severe acute respiratory syndrome coro .................... .................... .................... .................... .................... ....................
15715 MW008562.1 |Severe acute respiratory syndrome coro .................... .................... .................... .................... .................... ....................
15716 MW008565.1 |Severe acute respiratory syndrome coro .................... .................... .................... .................... .................... ....................
15717 MW008566.1 |Severe acute respiratory syndrome coro .................... .................... .................... .................... .................... ....................
15718 MW008567.1 |Severe acute respiratory syndrome coro .................... .................... .................... .................... .................... ....................
15719 MW008568.1 |Severe acute respiratory syndrome coro .................... .................... .................... .................... .................... ....................
15720 MW008569.1 |Severe acute respiratory syndrome coro .................... .................... .................... .................... .................... ....................
15721 MW008570.1 |Severe acute respiratory syndrome coro .................... .................... .................... .................... .................... ....................
15722 MW008571.1 |Severe acute respiratory syndrome coro .................... .................... .................... .................... .................... ....................
15723 MW008573.1 |Severe acute respiratory syndrome coro .................... .................... .................... .................... .................... ....................
15724 MW008574.1 |Severe acute respiratory syndrome coro .................... .................... .................... .................... .................... ....................
15725 MW008575.1 |Severe acute respiratory syndrome coro .................... .................... .................... .................... .................... ....................
15726 MW008576.1 |Severe acute respiratory syndrome coro .................... .................... .................... .................... .................... ....................
15727 MW008577.1 |Severe acute respiratory syndrome coro .................... .................... .................... .................... .................... ....................
15728 MW008578.1 |Severe acute respiratory syndrome coro .................... .................... .................... .................... .................... ....................
15729 MW008579.1 |Severe acute respiratory syndrome coro .................... .................... .................... .................... .................... ....................
15730 MW008580.1 |Severe acute respiratory syndrome coro .................... .................... .................... .................... .................... ....................
15731 MW008581.1 |Severe acute respiratory syndrome coro .................... .................... .................... .................... .................... ....................
15732 MW008582.1 |Severe acute respiratory syndrome coro .................... .................... .................... .................... .................... ....................
15733 MW008583.1 |Severe acute respiratory syndrome coro .................... .................... .................... .................... .................... ....................
15734 MW008584.1 |Severe acute respiratory syndrome coro .................... .................... .................... .................... .................... ....................
15735 MW008585.1 |Severe acute respiratory syndrome coro .................... .................... .................... .................... .................... ....................
15736 MW008587.1 |Severe acute respiratory syndrome coro .................... .................... .................... .................... .................... ....................
15737 MW008590.1 |Severe acute respiratory syndrome coro .................... .................... .................... .................... .................... ....................
15738 MW009055.1 |Severe acute respiratory syndrome coro .................... .................... .................... .................... .................... ....................
15739 MW009107.1 |Severe acute respiratory syndrome coro .................... .................... .................... .................... .................... ....................
15740 MW009255.1 |Severe acute respiratory syndrome coro .................... .................... .................... .................... .................... ....................
15741 MW010028.1 |Severe acute respiratory syndrome coro .................... .................... .................... .................... .................... ....................
15742 MW010029.1 |Severe acute respiratory syndrome coro .................... .................... .................... .................... .................... ....................
15743 MW010235.1 |Severe acute respiratory syndrome coro .................... .................... .................... .................... .................... ....................
```

15744 MW010236.1 |Severe acute respiratory syndrome coro ···················· ···················· ···················· ···················· ···················· ····················
15745 MW010245.1 |Severe acute respiratory syndrome coro ···················· ···················· ···················· ···················· ···················· ····················
15746 MW010246.1 |Severe acute respiratory syndrome coro ···················· ···················· ···················· ···················· ···················· ····················
15747 MW010247.1 |Severe acute respiratory syndrome coro ···················· ···················· ···················· ···················· ···················· ····················
15748 MW010248.1 |Severe acute respiratory syndrome coro ···················· ···················· ···················· ···················· ···················· ····················
15749 MW010249.1 |Severe acute respiratory syndrome coro ···················· ···················· ···················· ···················· ···················· ····················
15750 MW010250.1 |Severe acute respiratory syndrome coro ···················· ···················· ···················· ···················· ···················· ····················
15751 MW010251.1 |Severe acute respiratory syndrome coro ···················· ···················· ···················· ···················· ···················· ····················
15752 MW010252.1 |Severe acute respiratory syndrome coro ···················· ···················· ···················· ···················· ···················· ····················
15753 MW010253.1 |Severe acute respiratory syndrome coro ···················· ···················· ···················· ···················· ···················· ····················
15754 MW010254.1 |Severe acute respiratory syndrome coro ···················· ···················· ···················· ···················· ···················· ····················
15755 MW010255.1 |Severe acute respiratory syndrome coro ···················· ···················· ···················· ···················· ···················· ····················
15756 MW011762.1 |Severe acute respiratory syndrome coro ···················· ···················· ···················· ···················· ···················· ····················
15757 MW011763.1 |Severe acute respiratory syndrome coro ···················· ···················· ···················· ···················· ···················· ····················
15758 MW011764.1 |Severe acute respiratory syndrome coro ···················· ···················· ···················· ···················· ···················· ····················
15759 MW011765.1 |Severe acute respiratory syndrome coro ···················· ···················· ···················· ···················· ···················· ····················
15760 MW011766.1 |Severe acute respiratory syndrome coro ···················· ···················· ···················· ···················· ···················· ····················
15761 MW011767.1 |Severe acute respiratory syndrome coro ···················· ···················· ···················· ···················· ···················· ····················
15762 MW011768.1 |Severe acute respiratory syndrome coro ···················· ···················· ···················· ···················· ···················· ····················
15763 MW000354.1 |Severe acute respiratory syndrome coro ···················· ···················· ···················· ···················· ···················· ····················
15764 MW000355.1 |Severe acute respiratory syndrome coro ···················· ············C······· ···················· ···················· ···················· ····················
15765 MW000356.1 |Severe acute respiratory syndrome coro ···················· ············C······· ···················· ···················· ···················· ····················
15766 MW000357.1 |Severe acute respiratory syndrome coro ···················· ···················· ···················· ···················· ···················· ····················
15767 MW000358.1 |Severe acute respiratory syndrome coro ···················· ···················· ···················· ···················· ···················· ····················
15768 MW000359.1 |Severe acute respiratory syndrome coro ···················· ···················· ···················· ···················· ···················· ····················
15769 MW000360.1 |Severe acute respiratory syndrome coro ···················· ···················· ···················· ···················· ···················· ····················
15770 MW000361.1 |Severe acute respiratory syndrome coro ···················· ···················· ···················· ···················· ···················· ····················
15771 MW000363.1 |Severe acute respiratory syndrome coro ···················· ···················· ···················· ···················· ···················· ····················
15772 MW000364.1 |Severe acute respiratory syndrome coro ···················· ···················· ···················· ···················· ···················· ····················
15773 MW000365.1 |Severe acute respiratory syndrome coro ···················· ···················· ···················· ···················· ···················· ····················
15774 MW000366.1 |Severe acute respiratory syndrome coro ···················· ···················· ···················· ···················· ···················· ····················
15775 MW000369.1 |Severe acute respiratory syndrome coro ···················· ···················· ···················· ···················· ···················· ····················
15776 MW000371.1 |Severe acute respiratory syndrome coro ···················· ···················· ···················· ···················· ···················· ····················
15777 MW001232.1 |Severe acute respiratory syndrome coro ···················· ···················· ···················· ···················· ···················· ····················
15778 MW001233.1 |Severe acute respiratory syndrome coro ···················· ···················· ···················· ···················· ···················· ····················
15779 MW001234.1 |Severe acute respiratory syndrome coro ···················· ···················· ···················· ···················· ···················· ····················
15780 MW001235.1 |Severe acute respiratory syndrome coro ···················· ···················· ···················· ···················· ···················· ····················
15781 MW001236.1 |Severe acute respiratory syndrome coro ···················· ···················· ···················· ···················· ···················· ····················
15782 MW001237.1 |Severe acute respiratory syndrome coro ···················· ···················· ···················· ···················· ···················· ····················
15783 MW001238.1 |Severe acute respiratory syndrome coro ···················· ···················· ···················· ···················· ···················· ····················
15784 MW001239.1 |Severe acute respiratory syndrome coro ···················· ···················· ···················· ···················· ···················· ····················
15785 MW001240.1 |Severe acute respiratory syndrome coro ···················· ···················· ···················· ···················· ···················· ····················
15786 MW001241.1 |Severe acute respiratory syndrome coro ···················· ···················· ···················· ···················· ···················· ····················
15787 MW001242.1 |Severe acute respiratory syndrome coro ···················· ···················· ···················· ···················· ···················· ····················
15788 MW001243.1 |Severe acute respiratory syndrome coro ···················· ···················· ···················· ···················· ···················· ····················
15789 MW001244.1 |Severe acute respiratory syndrome coro ···················· ···················· ···················· ···················· ···················· ····················
15790 MW001245.1 |Severe acute respiratory syndrome coro ···················· ···················· ···················· ···················· ···················· ····················
15791 MW001246.1 |Severe acute respiratory syndrome coro ···················· ···················· ···················· ···················· ···················· ····················
15792 MW001247.1 |Severe acute respiratory syndrome coro ···················· ···················· ···················· ···················· ···················· ····················
15793 MW001248.1 |Severe acute respiratory syndrome coro ···················· ···················· ···················· ···················· ···················· ····················
15794 MW001249.1 |Severe acute respiratory syndrome coro ···················· ···················· ···················· ···················· ···················· ····················
15795 MW001250.1 |Severe acute respiratory syndrome coro ···················· ···················· ···················· ···················· ···················· ····················
15796 MW001251.1 |Severe acute respiratory syndrome coro ···················· ···················· ···················· ···················· ···················· ····················
15797 MW001252.1 |Severe acute respiratory syndrome coro ···················· ···················· ···················· ···················· ···················· ····················
15798 MW001253.1 |Severe acute respiratory syndrome coro ···················· ···················· ···················· ···················· ···················· ····················
15799 MW001254.1 |Severe acute respiratory syndrome coro ···················· ···················· ···················· ···················· ···················· ····················
15800 MW001255.1 |Severe acute respiratory syndrome coro ···················· ···················· ···················· ···················· ···················· ····················
15801 MW001256.1 |Severe acute respiratory syndrome coro ···················· ···················· ···················· ···················· ···················· ····················
15802 MW001257.1 |Severe acute respiratory syndrome coro ···················· ···················· ···················· ···················· ···················· ····················
15803 MW001258.1 |Severe acute respiratory syndrome coro ···················· ···················· ···················· ···················· ···················· ····················
15804 MW001259.1 |Severe acute respiratory syndrome coro ···················· ···················· ···················· ···················· ···················· ····················
15805 MW001260.1 |Severe acute respiratory syndrome coro ···················· ···················· ···················· ···················· ···················· ····················
15806 MW001261.1 |Severe acute respiratory syndrome coro ···················· ···················· ···················· ···················· ···················· ····················
15807 MW001262.1 |Severe acute respiratory syndrome coro ···················· ···················· ···················· ···················· ···················· ····················
15808 MW001263.1 |Severe acute respiratory syndrome coro ···················· ···················· ···················· ···················· ···················· ····················
15809 MW001264.1 |Severe acute respiratory syndrome coro ···················· ···················· ···················· ···················· ···················· ····················
15810 MW001265.1 |Severe acute respiratory syndrome coro ···················· ···················· ···················· ···················· ···················· ····················
15811 MW001266.1 |Severe acute respiratory syndrome coro ···················· ···················· ···················· ···················· ···················· ····················
15812 MW001267.1 |Severe acute respiratory syndrome coro ···················· ···················· ···················· ···················· ···················· ····················
15813 MW001268.1 |Severe acute respiratory syndrome coro ···················· ···················· ···················· ···················· ···················· ····················
15814 MW001269.1 |Severe acute respiratory syndrome coro ···················· ···················· ···················· ···················· ···················· ····················
15815 MW001270.1 |Severe acute respiratory syndrome coro ···················· ···················· ···················· ···················· ···················· ····················
15816 MW001271.1 |Severe acute respiratory syndrome coro ···················· ···················· ···················· ···················· ···················· ····················
15817 MW001272.1 |Severe acute respiratory syndrome coro ···················· ···················· ···················· ···················· ···················· ····················
15818 MW001273.1 |Severe acute respiratory syndrome coro ···················· ···················· ···················· ···················· ···················· ····················
15819 MW001274.1 |Severe acute respiratory syndrome coro ···················· ···················· ···················· ···················· ···················· ····················
15820 MW001275.1 |Severe acute respiratory syndrome coro ···················· ···················· ···················· ···················· ···················· ····················
15821 MW001276.1 |Severe acute respiratory syndrome coro ···················· ···················· ···················· ···················· ···················· ····················

15822 MW001277.1 |Severe acute respiratory syndrome coro ···················· ···················· ····················· ···················· ···················· ····················
15823 MW001278.1 |Severe acute respiratory syndrome coro ···················· ···················· ····················· ···················· ···················· ····················
15824 MW001279.1 |Severe acute respiratory syndrome coro ···················· ···················· ····················· ···················· ···················· ····················
15825 MW001280.1 |Severe acute respiratory syndrome coro ···················· ···················· ····················· ···················· ···················· ····················
15826 MW001281.1 |Severe acute respiratory syndrome coro ···················· ···················· ····················· ···················· ···················· ····················
15827 MW001282.1 |Severe acute respiratory syndrome coro ···················· ···················· ····················· ···················· ···················· ····················
15828 MW001283.1 |Severe acute respiratory syndrome coro ···················· ···················· ····················· ···················· ···················· ····················
15829 MW001284.1 |Severe acute respiratory syndrome coro ···················· ···················· ····················· ···················· ···················· ····················
15830 MW001285.1 |Severe acute respiratory syndrome coro ···················· ···················· ····················· ···················· ···················· ····················
15831 MW001286.1 |Severe acute respiratory syndrome coro ···················· ·A·················· ····················· ···················· ···················· ····················
15832 MW001287.1 |Severe acute respiratory syndrome coro ···················· ···················· ····················· ···················· ···················· ····················
15833 MW001288.1 |Severe acute respiratory syndrome coro ···················· ···················· ····················· ···················· ···················· ····················
15834 MW001667.1 |Severe acute respiratory syndrome coro ···················· ···················· ····················· ···················· ···················· ····················
15835 MW001668.1 |Severe acute respiratory syndrome coro ···················· ···················· ····················· ···················· ···················· ····················
15836 MW001669.1 |Severe acute respiratory syndrome coro ···················· ···················· ····················· ···················· ···················· ····················
15837 MW001670.1 |Severe acute respiratory syndrome coro ···················· ···················· ····················· ···················· ···················· ····················
15838 MW001671.1 |Severe acute respiratory syndrome coro ···················· ···················· ····················· ···················· ···················· ····················
15839 MW001672.1 |Severe acute respiratory syndrome coro ···················· ···················· ····················· ···················· ···················· ····················
15840 MW001673.1 |Severe acute respiratory syndrome coro ···················· ·A·················· ····················· ···················· ···················· ····················
15841 MW001674.1 |Severe acute respiratory syndrome coro ···················· ·A·················· ····················· ···················· ··C················· ····················
15842 MW001675.1 |Severe acute respiratory syndrome coro ···················· ·A·················· ····················· ···················· ···················· ····················
15843 MW001692.1 |Severe acute respiratory syndrome coro ···················· ·A·················· ····················· ···················· ···················· ····················
15844 MW001693.1 |Severe acute respiratory syndrome coro ···················· ·A·················· ····················· ···················· ···················· ····················
15845 MW001880.1 |Severe acute respiratory syndrome coro ···················· ···················· ····················· ···················· ···················· ····················
15846 MW001881.1 |Severe acute respiratory syndrome coro ···················· ···················· ····················· ···················· ···················· ····················
15847 MW001882.1 |Severe acute respiratory syndrome coro ···················· ···················· ····················· ···················· ···················· ····················
15848 MW001883.1 |Severe acute respiratory syndrome coro ···················· ···················· ····················· ···················· ···················· ····················
15849 MW001884.1 |Severe acute respiratory syndrome coro ···················· ···················· ····················· ···················· ···················· ····················
15850 MW001885.1 |Severe acute respiratory syndrome coro ···················· ···················· ····················· ···················· ···················· ····················
15851 MW001886.1 |Severe acute respiratory syndrome coro ···················· ···················· ····················· ···················· ···················· ····················
15852 MW001887.1 |Severe acute respiratory syndrome coro ···················· ·A·················· ····················· ···················· ···················· ····················
15853 MW001888.1 |Severe acute respiratory syndrome coro ···················· ·A·················· ····················· ···················· ···················· ····················
15854 MW001889.1 |Severe acute respiratory syndrome coro ···················· ···················· ····················· ···················· ···················· ····················
15855 MW001890.1 |Severe acute respiratory syndrome coro ···················· ·A·················· ····················· ···················· ···················· ····················
15856 MW001891.1 |Severe acute respiratory syndrome coro ···················· ···················· ····················· ···················· ···················· ····················
15857 MW001892.1 |Severe acute respiratory syndrome coro ···················· ···················· ····················· ···················· ···················· ····················
15858 MW001893.1 |Severe acute respiratory syndrome coro ···················· ·A·················· ····················· ···················· ···················· ····················
15859 MW001894.1 |Severe acute respiratory syndrome coro ···················· ·A·················· ····················· ···················· ···················· ····················
15860 MW001895.1 |Severe acute respiratory syndrome coro ···················· ···················· ····················· ···················· ···················· ····················
15861 MW001896.1 |Severe acute respiratory syndrome coro ···················· ·A·················· ····················· ···················· ···················· ····················
15862 MW001897.1 |Severe acute respiratory syndrome coro ···················· ···················· ····················· ···················· ···················· ····················
15863 MW001898.1 |Severe acute respiratory syndrome coro ···················· ·A·················· ····················· ···················· ···················· ····················
15864 MW001899.1 |Severe acute respiratory syndrome coro ···················· ·A·················· ····················· ···················· ···················· ····················
15865 MW001900.1 |Severe acute respiratory syndrome coro ···················· ·A·················· ····················· ···················· ···················· ····················
15866 MW001901.1 |Severe acute respiratory syndrome coro ···················· ·A·················· ····················· ···················· ···················· ····················
15867 MW001902.1 |Severe acute respiratory syndrome coro ···················· ·A·················· ····················· ···················· ···················· ····················
15868 MW001903.1 |Severe acute respiratory syndrome coro ···················· ·A·················· ····················· ···················· ···················· ····················
15869 MW001904.1 |Severe acute respiratory syndrome coro ···················· ·A·················· ····················· ···················· ···················· ····················
15870 MW001905.1 |Severe acute respiratory syndrome coro ···················· ·A·················· ····················· ···················· ···················· ····················
15871 MW001906.1 |Severe acute respiratory syndrome coro ···················· ···················· ····················· ···················· ···················· ····················
15872 MW001907.1 |Severe acute respiratory syndrome coro ···················· ·A·················· ····················· ···················· ···················· ····················
15873 MW001908.1 |Severe acute respiratory syndrome coro ···················· ···················· ····················· ···················· ···················· ····················
15874 MW001909.1 |Severe acute respiratory syndrome coro ···················· ···················· ····················· ···················· ···················· ····················
15875 MW001910.1 |Severe acute respiratory syndrome coro ···················· ···················· ····················· ···················· ···················· ····················
15876 MW001911.1 |Severe acute respiratory syndrome coro ···················· ···················· ····················· ···················· ···················· ····················
15877 MW001912.1 |Severe acute respiratory syndrome coro ···················· ···················· ····················· ···················· ···················· ····················
15878 MW001913.1 |Severe acute respiratory syndrome coro ···················· ···················· ····················· ···················· ···················· ····················
15879 MW001914.1 |Severe acute respiratory syndrome coro ···················· ·A·················· ····················· ···················· ···················· ····················
15880 MW001915.1 |Severe acute respiratory syndrome coro ···················· ·A·················· ····················· ···················· ···················· ····················
15881 MW001916.1 |Severe acute respiratory syndrome coro ···················· ·A·················· ····················· ···················· ···················· ····················
15882 MW001917.1 |Severe acute respiratory syndrome coro ···················· ·A·················· ····················· ···················· ···················· ····················
15883 MW001918.1 |Severe acute respiratory syndrome coro ···················· ·A·················· ····················· ···················· ···················· ····················
15884 MW001919.1 |Severe acute respiratory syndrome coro ···················· ·A·················· ····················· ···················· ···················· ····················
15885 MW001920.1 |Severe acute respiratory syndrome coro ···················· ·A·················· ····················· ···················· ···················· ····················
15886 MW001921.1 |Severe acute respiratory syndrome coro ···················· ·A·················· ····················· ···················· ···················· ····················
15887 MW001922.1 |Severe acute respiratory syndrome coro ···················· ·A·················· ····················· ···················· ···················· ····················
15888 MW001923.1 |Severe acute respiratory syndrome coro ···················· ·A·················· ····················· ···················· ···················· ····················
15889 MW001924.1 |Severe acute respiratory syndrome coro ···················· ·A·················· ····················· ···················· ···················· ····················
15890 MW001925.1 |Severe acute respiratory syndrome coro ···················· ·A·················· ····················· ···················· ···················· ····················
15891 MW001926.1 |Severe acute respiratory syndrome coro ···················· ·A·················· ····················· ···················· ···················· ····················
15892 MW001927.1 |Severe acute respiratory syndrome coro ···················· ·A·················· ····················· ···················· ···················· ····················
15893 MW001928.1 |Severe acute respiratory syndrome coro ···················· ·A·················· ····················· ···················· ···················· ····················
15894 MW001929.1 |Severe acute respiratory syndrome coro ···················· ·A·················· ····················· ···················· ···················· ····················
15895 MW001930.1 |Severe acute respiratory syndrome coro ···················· ·A·················· ····················· ···················· ···················· ····················
15896 MW001931.1 |Severe acute respiratory syndrome coro ···················· ·A·················· ····················· ···················· ···················· ····················
15897 MW002806.1 |Severe acute respiratory syndrome coro ···················· ···················· ····················· ···················· ···················· ····················
15898 MW002807.1 |Severe acute respiratory syndrome coro ···················· ···················· ····················· ···················· ···················· ····················
15899 MW002808.1 |Severe acute respiratory syndrome coro ···················· ···················· ····················· ···················· ···················· ····················

15900 MW002809.1 |Severe acute respiratory syndrome coro ···················· ···················· ····················· ···················· ···················· ····················
15901 MW002811.1 |Severe acute respiratory syndrome coro ···················· ···················· ····················· ···················· ···················· ····················
15902 MW002812.1 |Severe acute respiratory syndrome coro ···················· ···················· ····················· ···················· ···················· ····················
15903 MW002813.1 |Severe acute respiratory syndrome coro ···················· ···················· ····················· ···················· ···················· ····················
15904 MW002815.1 |Severe acute respiratory syndrome coro ···················· ·A·················· ····················· ···················· ···················· ····················
15905 MW002816.1 |Severe acute respiratory syndrome coro ···················· ·A·················· ····················· ···················· ···················· ····················
15906 MW002817.1 |Severe acute respiratory syndrome coro ···················· ·A·················· ····················· ···················· ···················· ····················
15907 MW002818.1 |Severe acute respiratory syndrome coro ···················· ·A·················· ····················· ···················· ···················· ····················
15908 MW002822.1 |Severe acute respiratory syndrome coro ···················· ·········A·········· ····················· ···················· ···················· ····················
15909 MW002823.1 |Severe acute respiratory syndrome coro ···················· ···················· ····················· ···················· ···················· ····················
15910 MW002825.1 |Severe acute respiratory syndrome coro ···················· ···················· ····················· ···················· ···················· ····················
15911 MW002826.1 |Severe acute respiratory syndrome coro ···················· ···················· ····················· ···················· ···················· ····················
15912 MW002827.1 |Severe acute respiratory syndrome coro ···················· ···················· ····················· ···················· ···················· ····················
15913 MW002828.1 |Severe acute respiratory syndrome coro ···················· ···················· ····················· ···················· ···················· ····················
15914 MW002829.1 |Severe acute respiratory syndrome coro ···················· ···················· ····················· ···················· ···················· ····················
15915 MW002831.1 |Severe acute respiratory syndrome coro ···················· ···················· ····················· ···················· ···················· ····················
15916 MW002833.1 |Severe acute respiratory syndrome coro ···················· ···················· ····················· ···················· ···················· ····················
15917 MW002834.1 |Severe acute respiratory syndrome coro ···················· ···················· ····················· ···················· ···················· ····················
15918 MW002836.1 |Severe acute respiratory syndrome coro ···················· ···················· ····················· ···················· ···················· ····················
15919 MW002837.1 |Severe acute respiratory syndrome coro ···················· ···················· ····················· ···················· ···················· ····················
15920 MW004168.1 |Severe acute respiratory syndrome coro ···················· ···················· ····················· ···················· ···················· ····················
15921 MW006555.1 |Severe acute respiratory syndrome coro ···················· ···················· ····················· ···················· ···················· ····················
15922 MW006556.1 |Severe acute respiratory syndrome coro ···················· ···················· ····················· ···················· ···················· ····················
15923 MW006557.1 |Severe acute respiratory syndrome coro ···················· ···················· ····················· ···················· ···················· ····················
15924 MW006559.1 |Severe acute respiratory syndrome coro ···················· ···················· ····················· ···················· ···················· ····················
15925 MW006560.1 |Severe acute respiratory syndrome coro ···················· ···················· ····················· ···················· ···················· ····················
15926 MW006561.1 |Severe acute respiratory syndrome coro ···················· ···················· ····················· ···················· ···················· ····················
15927 MW006563.1 |Severe acute respiratory syndrome coro ···················· ···················· ····················· ···················· ···················· ····················
15928 MW006564.1 |Severe acute respiratory syndrome coro ···················· ···················· ····················· ···················· ···················· ····················
15929 MW006565.1 |Severe acute respiratory syndrome coro ···················· ···················· ····················· ···················· ···················· ····················
15930 MW006567.1 |Severe acute respiratory syndrome coro ···················· ···················· ····················· ···················· ···················· ····················
15931 MW006569.1 |Severe acute respiratory syndrome coro ···················· ···················· ····················· ···················· ···················· ····················
15932 MW006571.1 |Severe acute respiratory syndrome coro ···················· ···················· ····················· ···················· ···················· ····················
15933 MW006573.1 |Severe acute respiratory syndrome coro ···················· ···················· ····················· ···················· ···················· ····················
15934 MW006574.1 |Severe acute respiratory syndrome coro ···················· ···················· ····················· ···················· ···················· ····················
15935 MW006575.1 |Severe acute respiratory syndrome coro ···················· ···················· ····················· ···················· ···················· ····················
15936 MW006576.1 |Severe acute respiratory syndrome coro ···················· ···················· ····················· ···················· ···················· ····················
15937 MW006577.1 |Severe acute respiratory syndrome coro ···················· ···················· ····················· ···················· ···················· ····················
15938 MW006578.1 |Severe acute respiratory syndrome coro ···················· ···················· ····················· ···················· ···················· ····················
15939 MW006579.1 |Severe acute respiratory syndrome coro ···················· ···················· ····················· ···················· ···················· ····················
15940 MW006583.1 |Severe acute respiratory syndrome coro ···················· ···················· ····················· ···················· ···················· ····················
15941 MW006585.1 |Severe acute respiratory syndrome coro ···················· ···················· ····················· ···················· ···················· ····················
15942 MW006586.1 |Severe acute respiratory syndrome coro ···················· ···················· ····················· ···················· ···················· ····················
15943 MW006590.1 |Severe acute respiratory syndrome coro ···················· ···················· ····················· ···················· ···················· ····················
15944 MW006591.1 |Severe acute respiratory syndrome coro ···················· ····T··············· ····················· ···················· ···················· ····················
15945 MW006592.1 |Severe acute respiratory syndrome coro ···················· ···················· ····················· ···················· ···················· ····················
15946 MW006593.1 |Severe acute respiratory syndrome coro ···················· ···················· ····················· ···················· ···················· ····················
15947 MW006594.1 |Severe acute respiratory syndrome coro ···················· ···················· ····················· ···················· ···················· ····················
15948 MW006595.1 |Severe acute respiratory syndrome coro ···················· ···················· ····················· ···················· ···················· ····················
15949 MW006596.1 |Severe acute respiratory syndrome coro ···················· ···················· ····················· ···················· ···················· ····················
15950 MW006598.1 |Severe acute respiratory syndrome coro ···················· ···················· ····················· ···················· ···················· ····················
15951 MW006599.1 |Severe acute respiratory syndrome coro ···················· ···················· ····················· ···················· ···················· ····················
15952 MW006600.1 |Severe acute respiratory syndrome coro ···················· ···················· ····················· ···················· ···················· ····················
15953 MW006601.1 |Severe acute respiratory syndrome coro ···················· ···················· ····················· ···················· ···················· ····················
15954 MW006603.1 |Severe acute respiratory syndrome coro ···················· ···················· ····················· ···················· ···················· ····················
15955 MW006604.1 |Severe acute respiratory syndrome coro ···················· ···················· ····················· ···················· ···················· ····················
15956 MW006605.1 |Severe acute respiratory syndrome coro ···················· ···················· ····················· ···················· ···················· ····················
15957 MW006606.1 |Severe acute respiratory syndrome coro ···················· ·A·················· ····················· ···················· ···················· ····················
15958 MW006607.1 |Severe acute respiratory syndrome coro ···················· ·A·················· ····················· ···················· ···················· ····················
15959 MT997203.1 |Severe acute respiratory syndrome coro ···················· ···················· ····················· ···················· ···················· ····················
15960 MT997204.1 |Severe acute respiratory syndrome coro ···················· ···················· ····················· ···················· ···················· ····················
15961 MT997205.1 |Severe acute respiratory syndrome coro ···················· ···················· ····················· ···················· ···················· ····················
15962 MT997206.1 |Severe acute respiratory syndrome coro ···················· ···················· ···········T········· ···················· ···················· ····················
15963 MT997208.1 |Severe acute respiratory syndrome coro ···················· ···················· ···········T········· ···················· ···················· ····················
15964 MT997209.1 |Severe acute respiratory syndrome coro ···················· ·A·················· ····················· ···················· ···················· ····················
15965 MT997210.1 |Severe acute respiratory syndrome coro ···················· ···················· ····················· ···················· ···················· ····················
15966 MT997212.1 |Severe acute respiratory syndrome coro ···················· ···················· ····················· ···················· ···················· ····················
15967 MT997213.1 |Severe acute respiratory syndrome coro ···················· ···················· ····················· ···················· ···················· ····················
15968 MT997214.1 |Severe acute respiratory syndrome coro ···················· ···················· ····················· ···················· ···················· ····················
15969 MT997215.1 |Severe acute respiratory syndrome coro ···················· ···················· ····················· ···················· ···················· ····················
15970 MT997216.1 |Severe acute respiratory syndrome coro ···················· ···················· ····················· ···················· ···················· ····················
15971 MT997218.1 |Severe acute respiratory syndrome coro ···················· ···················· ····················· ···················· ···················· ····················
15972 MT997219.1 |Severe acute respiratory syndrome coro ···················· ···················· ····················· ···················· ···················· ····················
15973 MT997220.1 |Severe acute respiratory syndrome coro ···················· ···················· ····················· ···················· ···················· ····················
15974 MT997221.1 |Severe acute respiratory syndrome coro ···················· ···················· ····················· ···················· ···················· ····················
15975 MT997222.1 |Severe acute respiratory syndrome coro ···················· ···················· ····················· ···················· ···················· ····················
15976 MT997223.1 |Severe acute respiratory syndrome coro ···················· ···················· ····················· ···················· ···················· ····················
15977 MT997224.1 |Severe acute respiratory syndrome coro ···················· ·A·················· ····················· ···················· ···················· ····················

15978 MT997225.1 |Severe acute respiratory syndrome coro ···················· ···················· ···················· ···················· ···················· ····················
15979 MT997226.1 |Severe acute respiratory syndrome coro ···················· ·A·················· ···················· ···················· ···················· ···A················
15980 MT997227.1 |Severe acute respiratory syndrome coro ···················· ·A·················· ···················· ···················· ···················· ····················
15981 MT997228.1 |Severe acute respiratory syndrome coro ···················· ···················· ···················· ···················· ···················· ····················
15982 MT997229.1 |Severe acute respiratory syndrome coro ···················· ···················· ···················· ···················· ···················· ····················
15983 MT997230.1 |Severe acute respiratory syndrome coro ···················· ···················· ···················· ···················· ···················· ····················
15984 MT997232.1 |Severe acute respiratory syndrome coro ···················· ···················· ···················· ···················· ···················· ····················
15985 MT997233.1 |Severe acute respiratory syndrome coro ···················· ·A·················· ···················· ···················· ···················· ····················
15986 MT997235.1 |Severe acute respiratory syndrome coro ···················· ·A·················· ···················· ···················· ···················· ···A················
15987 MT997236.1 |Severe acute respiratory syndrome coro ···················· ···A················ ···················· ···················· ···················· ····················
15988 MT997237.1 |Severe acute respiratory syndrome coro ···················· ·A·················· ···················· ···················· ···················· ····················
15989 MT997240.1 |Severe acute respiratory syndrome coro ···················· ·A·················· ···················· ···················· ···················· ····················
15990 MT997241.1 |Severe acute respiratory syndrome coro ···················· ···················· ···················· ···················· ···················· ····················
15991 MT997242.1 |Severe acute respiratory syndrome coro ···················· ·A·················· ···················· ···················· ···················· ····················
15992 MT997243.1 |Severe acute respiratory syndrome coro ···················· ·A·················· ···················· ···················· ···················· ····················
15993 MT997685.1 |Severe acute respiratory syndrome coro ···················· ···················· ···················· ···················· ···················· ····················
15994 MT997686.1 |Severe acute respiratory syndrome coro ···················· ···················· ···················· ···················· ···················· ····················
15995 MT997687.1 |Severe acute respiratory syndrome coro ···················· ···················· ···················· ···················· ···················· ····················
15996 MT997688.1 |Severe acute respiratory syndrome coro ···················· ···················· ···················· ···················· ···················· ····················
15997 MT997689.1 |Severe acute respiratory syndrome coro ···················· ·A·················· ···················· ···················· ···················· ····················
15998 MT997690.1 |Severe acute respiratory syndrome coro ···················· ·A·················· ···················· ···················· ···················· ····················
15999 MT997691.1 |Severe acute respiratory syndrome coro ···················· ·A·················· ···················· ···················· ···················· ····················
16000 MT997692.1 |Severe acute respiratory syndrome coro ···················· ···················· ···················· ···················· ···················· ····················
16001 MT997693.1 |Severe acute respiratory syndrome coro ···················· ···················· ···················· ···················· ···················· ····················
16002 MT997694.1 |Severe acute respiratory syndrome coro ···················· ···················· ···················· ···················· ···················· ····················
16003 MT997695.1 |Severe acute respiratory syndrome coro ···················· ···················· ···················· ···················· ···················· ····················
16004 MT997696.1 |Severe acute respiratory syndrome coro ···················· ···················· ···················· ···················· ···················· ····················
16005 MT997697.1 |Severe acute respiratory syndrome coro ···················· ···················· ···················· ···················· ···················· ····················
16006 MT997698.1 |Severe acute respiratory syndrome coro ···················· ···················· ···················· ···················· ···················· ····················
16007 MT997699.1 |Severe acute respiratory syndrome coro ···················· ···················· ···················· ···················· ···················· ····················
16008 MT997700.1 |Severe acute respiratory syndrome coro ···················· ···················· ···················· ···················· ···················· ····················
16009 MT997701.1 |Severe acute respiratory syndrome coro ···················· ···················· ···················· ···················· ···················· ····················
16010 MT997702.1 |Severe acute respiratory syndrome coro ···················· ···················· ···················· ···················· ···················· ····················
16011 MT997703.1 |Severe acute respiratory syndrome coro ···················· ···················· ···················· ···················· ···················· ····················
16012 MT997704.1 |Severe acute respiratory syndrome coro ···················· ···················· ···················· ···················· ···················· ····················
16013 MT997705.1 |Severe acute respiratory syndrome coro ···················· ···················· ···················· ···················· ···················· ····················
16014 MT997706.1 |Severe acute respiratory syndrome coro ···················· ···················· ···················· ···················· ···················· ····················
16015 MT997707.1 |Severe acute respiratory syndrome coro ···················· ···················· ···················· ···················· ···················· ····················
16016 MT997708.1 |Severe acute respiratory syndrome coro ···················· ···················· ···················· ···················· ···················· ····················
16017 MT997709.1 |Severe acute respiratory syndrome coro ···················· ···················· ···················· ···················· ···················· ····················
16018 MT997710.1 |Severe acute respiratory syndrome coro ···················· ···················· ···················· ···················· ···················· ····················
16019 MT997711.1 |Severe acute respiratory syndrome coro ···················· ···················· ···················· ···················· ···················· ····················
16020 MT997712.1 |Severe acute respiratory syndrome coro ···················· ···················· ···················· ···················· ···················· ····················
16021 MT997713.1 |Severe acute respiratory syndrome coro ···················· ···················· ···················· ···················· ···················· ····················
16022 MT997714.1 |Severe acute respiratory syndrome coro ···················· ···················· ···················· ···················· ···················· ····················
16023 MT997715.1 |Severe acute respiratory syndrome coro ···················· ···················· ···················· ···················· ···················· ····················
16024 MT997716.1 |Severe acute respiratory syndrome coro ···················· ···················· ···················· ···················· ···················· ····················
16025 MT997717.1 |Severe acute respiratory syndrome coro ···················· ···················· ···················· ···················· ···················· ····················
16026 MT997718.1 |Severe acute respiratory syndrome coro ···················· ···················· ···················· ···················· ···················· ····················
16027 MT997719.1 |Severe acute respiratory syndrome coro ···················· ···················· ···················· ···················· ···················· ····················
16028 MT997720.1 |Severe acute respiratory syndrome coro ···················· ···················· ···················· ···················· ···················· ····················
16029 MT997721.1 |Severe acute respiratory syndrome coro ···················· ···················· ···················· ···················· ···················· ····················
16030 MT997722.1 |Severe acute respiratory syndrome coro ···················· ···················· ···················· ···················· ···················· ····················
16031 MT997723.1 |Severe acute respiratory syndrome coro ···················· ···················· ···················· ···················· ···················· ····················
16032 MT997724.1 |Severe acute respiratory syndrome coro ···················· ···················· ···················· ···················· ···················· ····················
16033 MT997725.1 |Severe acute respiratory syndrome coro ···················· ···················· ···················· ···················· ···················· ····················
16034 MT997726.1 |Severe acute respiratory syndrome coro ···················· ···················· ···················· ···················· ···················· ····················
16035 MT997727.1 |Severe acute respiratory syndrome coro ···················· ···················· ···················· ···················· ···················· ····················
16036 MT997728.1 |Severe acute respiratory syndrome coro ···················· ···················· ···················· ···················· ···················· ····················
16037 MT997729.1 |Severe acute respiratory syndrome coro ···················· ···················· ···················· ···················· ···················· ····················
16038 MT997730.1 |Severe acute respiratory syndrome coro ···················· ···················· ···················· ···················· ···················· ····················
16039 MT997731.1 |Severe acute respiratory syndrome coro ···················· ···················· ···················· ···················· ···················· ····················
16040 MT997732.1 |Severe acute respiratory syndrome coro ···················· ···················· ···················· ···················· ···················· ····················
16041 MT997733.1 |Severe acute respiratory syndrome coro ···················· ···················· ···················· ···················· ···················· ····················
16042 MT997734.1 |Severe acute respiratory syndrome coro ···················· ···················· ···················· ···················· ···················· ····················
16043 MT997735.1 |Severe acute respiratory syndrome coro ···················· ···················· ···················· ···················· ···················· ····················
16044 MT997736.1 |Severe acute respiratory syndrome coro ···················· ···················· ···················· ···················· ··NN················ ····················
16045 MT997737.1 |Severe acute respiratory syndrome coro ···················· ···················· ···················· ···················· ···················· ····················
16046 MT997738.1 |Severe acute respiratory syndrome coro ···················· ···················· ···················· ···················· ···················· ····················
16047 MT997739.1 |Severe acute respiratory syndrome coro ···················· ···················· ···················· ···················· ···················· ····················
16048 MT997740.1 |Severe acute respiratory syndrome coro ···················· ···················· ···················· ···················· ···················· ····················
16049 MT997741.1 |Severe acute respiratory syndrome coro ···················· ···················· ···················· ···················· ···················· ····················
16050 MT997742.1 |Severe acute respiratory syndrome coro ···················· ···················· ···················· ···················· ···················· ····················
16051 MT997743.1 |Severe acute respiratory syndrome coro ···················· ···················· ···················· ···················· ···················· ····················
16052 MT997744.1 |Severe acute respiratory syndrome coro ···················· ···················· ···················· ···················· ···················· ····················
16053 MT997745.1 |Severe acute respiratory syndrome coro ···················· ···················· ···················· ···················· ···················· ····················
16054 MT997746.1 |Severe acute respiratory syndrome coro ···················· ···················· ···················· ···················· ···················· ····················
16055 MT997747.1 |Severe acute respiratory syndrome coro ···················· ···················· ···················· ···················· ···················· ····················

16056 MT997762.1 |Severe acute respiratory syndrome coro .................... .................... .................... .................... .................... ....................
16057 MT997763.1 |Severe acute respiratory syndrome coro .................... .................... .................... .................... .................... ....................
16058 MT997764.1 |Severe acute respiratory syndrome coro .................... .................... .................... .................... .................... ....................
16059 MT997765.1 |Severe acute respiratory syndrome coro .................... .................... .................... .................... .................... ....................
16060 MT997766.1 |Severe acute respiratory syndrome coro .................... .................... .................... .................... .................... ....................
16061 MT997767.1 |Severe acute respiratory syndrome coro .................... .................... .................... .................... .................... ....................
16062 MT997768.1 |Severe acute respiratory syndrome coro .................... .................... .................... .................... .................... ....................
16063 MT997769.1 |Severe acute respiratory syndrome coro .................... .................... .................... .................... .................... ....................
16064 MT997770.1 |Severe acute respiratory syndrome coro .................... .................... .................... .................... .................... ....................
16065 MT997771.1 |Severe acute respiratory syndrome coro .................... .................... .................... .................... .................... ....................
16066 MT997949.1 |Severe acute respiratory syndrome coro .................... .................... .................... .................... .................... ....................
16067 MT997950.1 |Severe acute respiratory syndrome coro .................... .................... .................... .................... .................... ....................
16068 MT997951.1 |Severe acute respiratory syndrome coro .................... .................... .................... .................... .................... ....................
16069 MT997952.1 |Severe acute respiratory syndrome coro .................... .................... .................... .................... .................... ....................
16070 MT997953.1 |Severe acute respiratory syndrome coro .................... .................... .................... .................... .................... ....................
16071 MT997954.1 |Severe acute respiratory syndrome coro .................... .................... .................... .................... .................... ....................
16072 MT997955.1 |Severe acute respiratory syndrome coro .................... .................... .................... .................... .................... ....................
16073 MT997956.1 |Severe acute respiratory syndrome coro .................... .................... .................... .................... .................... ....................
16074 MT997957.1 |Severe acute respiratory syndrome coro .................... .................... .................... .................... .................... ....................
16075 MT997958.1 |Severe acute respiratory syndrome coro .................... .................... .................... .................... .................... ....................
16076 MT997959.1 |Severe acute respiratory syndrome coro .................... .................... .................... .................... .................... ....................
16077 MT997960.1 |Severe acute respiratory syndrome coro .................... .................... .................... .................... .................... ....................
16078 MT997961.1 |Severe acute respiratory syndrome coro .................... .................... .................... .................... .................... ....................
16079 MT997962.1 |Severe acute respiratory syndrome coro .................... .................... .................... .................... .................... ....................
16080 MT997963.1 |Severe acute respiratory syndrome coro .................... .................... .................... .................... .................... ....................
16081 MT997964.1 |Severe acute respiratory syndrome coro .................... .................... .................... .................... .................... ....................
16082 MT997965.1 |Severe acute respiratory syndrome coro .................... .................... .................... .................... .................... ....................
16083 MT997966.1 |Severe acute respiratory syndrome coro .................... .................... .................... .................... .................... ....................
16084 MT997967.1 |Severe acute respiratory syndrome coro .................... .................... .................... .................... .................... ....................
16085 MT997968.1 |Severe acute respiratory syndrome coro .................... .................... .................... .................... .................... ....................
16086 MT997969.1 |Severe acute respiratory syndrome coro .................... .................... .................... .................... .................... ....................
16087 MT997970.1 |Severe acute respiratory syndrome coro .................... .................... .................... .................... .................... ....................
16088 MT997971.1 |Severe acute respiratory syndrome coro .................... .................... .................... .................... .................... ....................
16089 MT997972.1 |Severe acute respiratory syndrome coro .................... .................... .................... .................... .................... ....................
16090 MT997973.1 |Severe acute respiratory syndrome coro .................... .................... .................... .................... .................... ....................
16091 MT997974.1 |Severe acute respiratory syndrome coro .................... .................... .................... .................... .................... ....................
16092 MT997975.1 |Severe acute respiratory syndrome coro .................... .................... .................... .................... .................... ....................
16093 MT997976.1 |Severe acute respiratory syndrome coro .................... .................... .................... .................... .................... ....................
16094 MT997977.1 |Severe acute respiratory syndrome coro .................... .................... .................... .................... .................... ....................
16095 MT997978.1 |Severe acute respiratory syndrome coro .................... .................... .................... .................... .................... ....................
16096 MT997979.1 |Severe acute respiratory syndrome coro .................... .................... .................... .................... .................... ....................
16097 MT997980.1 |Severe acute respiratory syndrome coro .................... .................... .................... .................... .................... ....................
16098 MT997981.1 |Severe acute respiratory syndrome coro .................... .................... .................... .................... .................... ....................
16099 MT997982.1 |Severe acute respiratory syndrome coro .................... .................... .................... .................... .................... ....................
16100 MT997983.1 |Severe acute respiratory syndrome coro .................... .................... .................... .................... .................... ....................
16101 MT997984.1 |Severe acute respiratory syndrome coro .................... .................... .................... .................... .................... ....................
16102 MT997985.1 |Severe acute respiratory syndrome coro .................... .................... .................... .................... .................... ....................
16103 MT997986.1 |Severe acute respiratory syndrome coro .................... .................... .................... .................... .................... ....................
16104 MT997987.1 |Severe acute respiratory syndrome coro .................... .................... .................... .................... .................... ....................
16105 MT997988.1 |Severe acute respiratory syndrome coro .................... .................... .................... .................... .................... ....................
16106 MT997989.1 |Severe acute respiratory syndrome coro .................... .................... .................... .................... .................... ....................
16107 MT997990.1 |Severe acute respiratory syndrome coro .................... .................... .................... .................... .................... ....................
16108 MT997991.1 |Severe acute respiratory syndrome coro .................... .................... .................... .................... .................... ....................
16109 MT997992.1 |Severe acute respiratory syndrome coro .................... .................... .................... .................... .................... ....................
16110 MT997993.1 |Severe acute respiratory syndrome coro .................... .................... .................... .................... .................... ....................
16111 MT997994.1 |Severe acute respiratory syndrome coro .................... .................... .................... .................... .................... ....................
16112 MT994849.1 |Severe acute respiratory syndrome coro .................... .................... .................... .................... .................... ....................
16113 MT994881.1 |Severe acute respiratory syndrome coro .................... .................... .................... .................... .................... ....................
16114 MT994884.1 |Severe acute respiratory syndrome coro .................... .................... .................... .................... .................... ....................
16115 MT994901.1 |Severe acute respiratory syndrome coro .................... .................... .................... .................... .................... ....................
16116 MT994902.1 |Severe acute respiratory syndrome coro .................... .................... .................... .................... .................... ....................
16117 MT994903.1 |Severe acute respiratory syndrome coro .................... .................... .................... .................... .................... ....................
16118 MT994904.1 |Severe acute respiratory syndrome coro .................... .................... .................... .................... .................... ....................
16119 MT994905.1 |Severe acute respiratory syndrome coro .................... .................... .................... .................... .................... ....................
16120 MT994906.1 |Severe acute respiratory syndrome coro .................... .................... .................... .................... .................... ....................
16121 MT994907.1 |Severe acute respiratory syndrome coro .................... .................... .................... .................... .................... ....................
16122 MT994908.1 |Severe acute respiratory syndrome coro .................... .................... .................... .................... .................... ....................
16123 MT994909.1 |Severe acute respiratory syndrome coro .................... .................... .................... .................... .................... ....................
16124 MT994910.1 |Severe acute respiratory syndrome coro .................... .................... .................... .................... .................... ....................
16125 MT994911.1 |Severe acute respiratory syndrome coro .................... .................... .................... .................... .................... ....................
16126 MT994912.1 |Severe acute respiratory syndrome coro .................... .................... .................... .................... .................... ....................
16127 MT994913.1 |Severe acute respiratory syndrome coro .................... .................... .................... .................... .................... ....................
16128 MT994914.1 |Severe acute respiratory syndrome coro .................... .................... .................... .................... .................... ....................
16129 MT994915.1 |Severe acute respiratory syndrome coro .................... .................... .................... .................... .................... ....................
16130 MT994916.1 |Severe acute respiratory syndrome coro .................... .................... .................... .................... .................... ....................
16131 MT994917.1 |Severe acute respiratory syndrome coro .................... .................... .................... .................... .................... ....................
16132 MT994918.1 |Severe acute respiratory syndrome coro .................... .................... .................... .................... .................... ....................
16133 MT994919.1 |Severe acute respiratory syndrome coro .................... .................... .................... .................... .................... ....................

```
16134 MT994920.1 |Severe acute respiratory syndrome coro    ····················  ····················  ·····················  ····················  ····················  ····················
16135 MT994921.1 |Severe acute respiratory syndrome coro    ····················  ····················  ·····················  ····················  ····················  ····················
16136 MT994922.1 |Severe acute respiratory syndrome coro    ····················  ····················  ·····················  ····················  ····················  ····················
16137 MT994923.1 |Severe acute respiratory syndrome coro    ····················  ····················  ·····················  ····················  ····················  ····················
16138 MT994924.1 |Severe acute respiratory syndrome coro    ····················  ····················  ·····················  ····················  ····················  ····················
16139 MT994925.1 |Severe acute respiratory syndrome coro    ····················  ····················  ·····················  ····················  ····················  ····················
16140 MT994926.1 |Severe acute respiratory syndrome coro    ····················  ····················  ·····················  ····················  ····················  ····················
16141 MT994927.1 |Severe acute respiratory syndrome coro    ····················  ····················  ·····················  ····················  ····················  ····················
16142 MT994928.1 |Severe acute respiratory syndrome coro    ····················  ····················  ·····················  ····················  ····················  ····················
16143 MT994929.1 |Severe acute respiratory syndrome coro    ····················  ····················  ·····················  ····················  ····················  ····················
16144 MT994930.1 |Severe acute respiratory syndrome coro    ····················  ····················  ·····················  ····················  ····················  ····················
16145 MT994931.1 |Severe acute respiratory syndrome coro    ····················  ····················  ·····················  ····················  ····················  ····················
16146 MT994932.1 |Severe acute respiratory syndrome coro    ····················  ····················  ·····················  ····················  ····················  ····················
16147 MT994933.1 |Severe acute respiratory syndrome coro    ····················  ····················  ·····················  ····················  ····················  ····················
16148 MT994934.1 |Severe acute respiratory syndrome coro    ····················  ····················  ·····················  ····················  ····················  ····················
16149 MT994935.1 |Severe acute respiratory syndrome coro    ····················  ····················  ·····················  ····················  ····················  ····················
16150 MT994936.1 |Severe acute respiratory syndrome coro    ····················  ····················  ·····················  ····················  ····················  ····················
16151 MT994937.1 |Severe acute respiratory syndrome coro    ····················  ····················  ·····················  ····················  ····················  ····················
16152 MT994938.1 |Severe acute respiratory syndrome coro    ····················  ····················  ·····················  ····················  ····················  ····················
16153 MT994939.1 |Severe acute respiratory syndrome coro    ····················  ····················  ·····················  ····················  ····················  ····················
16154 MT994940.1 |Severe acute respiratory syndrome coro    ····················  ····················  ·····················  ····················  ····················  ····················
16155 MT994941.1 |Severe acute respiratory syndrome coro    ····················  ····················  ·····················  ····················  ····················  ····················
16156 MT994942.1 |Severe acute respiratory syndrome coro    ····················  ····················  ·····················  ····················  ····················  ····················
16157 MT994943.1 |Severe acute respiratory syndrome coro    ····················  ····················  ·····················  ····················  ····················  ····················
16158 MT994944.1 |Severe acute respiratory syndrome coro    ····················  ····················  ·····················  ····················  ····················  ····················
16159 MT994945.1 |Severe acute respiratory syndrome coro    ····················  ····················  ·····················  ····················  ····················  ····················
16160 MT994946.1 |Severe acute respiratory syndrome coro    ····················  ····················  ·····················  ····················  ····················  ····················
16161 MT994947.1 |Severe acute respiratory syndrome coro    ····················  ····················  ·····················  ····················  ····················  ····················
16162 MT994948.1 |Severe acute respiratory syndrome coro    ····················  ····················  ·····················  ····················  ····················  ····················
16163 MT994949.1 |Severe acute respiratory syndrome coro    ····················  ····················  ·····················  ····················  ····················  ····················
16164 MT994950.1 |Severe acute respiratory syndrome coro    ····················  ····················  ·····················  ····················  ····················  ····················
16165 MT994951.1 |Severe acute respiratory syndrome coro    ····················  ····················  ·····················  ····················  ····················  ····················
16166 MT994952.1 |Severe acute respiratory syndrome coro    ····················  ····················  ·····················  ····················  ····················  ····················
16167 MT994953.1 |Severe acute respiratory syndrome coro    ····················  ····················  ·····················  ····················  ····················  ····················
16168 MT994954.1 |Severe acute respiratory syndrome coro    ····················  ····················  ·····················  ····················  ····················  ····················
16169 MT994955.1 |Severe acute respiratory syndrome coro    ····················  ····················  ·····················  ····················  ····················  ····················
16170 MT994956.1 |Severe acute respiratory syndrome coro    ····················  ····················  ·····················  ····················  ····················  ····················
16171 MT994957.1 |Severe acute respiratory syndrome coro    ····················  ····················  ·····················  ····················  ····················  ····················
16172 MT994958.1 |Severe acute respiratory syndrome coro    ····················  ····················  ·····················  ····················  ····················  ····················
16173 MT994959.1 |Severe acute respiratory syndrome coro    ····················  ····················  ·····················  ····················  ····················  ····················
16174 MT994960.1 |Severe acute respiratory syndrome coro    ····················  ····················  ·····················  ····················  ····················  ····················
16175 MT994961.1 |Severe acute respiratory syndrome coro    ····················  ····················  ·····················  ····················  ····················  ····················
16176 MT994962.1 |Severe acute respiratory syndrome coro    ····················  ····················  ·····················  ····················  ····················  ····················
16177 MT994963.1 |Severe acute respiratory syndrome coro    ····················  ····················  ·····················  ····················  ····················  ····················
16178 MT994964.1 |Severe acute respiratory syndrome coro    ····················  ····················  ·····················  ····················  ····················  ····················
16179 MT994965.1 |Severe acute respiratory syndrome coro    ····················  ····················  ·····················  ····················  ····················  ····················
16180 MT994966.1 |Severe acute respiratory syndrome coro    ····················  ····················  ·····················  ····················  ····················  ····················
16181 MT994967.1 |Severe acute respiratory syndrome coro    ····················  ····················  ·····················  ····················  ····················  ····················
16182 MT994968.1 |Severe acute respiratory syndrome coro    ····················  ····················  ·····················  ····················  ····················  ····················
16183 MT994969.1 |Severe acute respiratory syndrome coro    ····················  ····················  ·····················  ····················  ····················  ····················
16184 MT994970.1 |Severe acute respiratory syndrome coro    ····················  ····················  ·····················  ····················  ····················  ····················
16185 MT994971.1 |Severe acute respiratory syndrome coro    ····················  ····················  ·····················  ····················  ····················  ····················
16186 MT994972.1 |Severe acute respiratory syndrome coro    ····················  ····················  ·····················  ····················  ····················  ····················
16187 MT994973.1 |Severe acute respiratory syndrome coro    ····················  ····················  ·····················  ····················  ····················  ····················
16188 MT994974.1 |Severe acute respiratory syndrome coro    ····················  ····················  ·····················  ····················  ····················  ····················
16189 MT994975.1 |Severe acute respiratory syndrome coro    ····················  ····················  ·····················  ····················  ····················  ····················
16190 MT994976.1 |Severe acute respiratory syndrome coro    ····················  ····················  ·····················  ····················  ····················  ····················
16191 MT994977.1 |Severe acute respiratory syndrome coro    ····················  ····················  ·····················  ····················  ····················  ····················
16192 MT994978.1 |Severe acute respiratory syndrome coro    ····················  ····················  ·····················  ····················  ····················  ····················
16193 MT994979.1 |Severe acute respiratory syndrome coro    ····················  ····················  ·····················  ····················  ····················  ····················
16194 MT994980.1 |Severe acute respiratory syndrome coro    ····················  ····················  ·····················  ····················  ····················  ····················
16195 MT994981.1 |Severe acute respiratory syndrome coro    ····················  ····················  ·····················  ····················  ····················  ····················
16196 MT994982.1 |Severe acute respiratory syndrome coro    ····················  ····················  ·····················  ····················  ····················  ····················
16197 MT994983.1 |Severe acute respiratory syndrome coro    ····················  ····················  ·····················  ····················  ····················  ····················
16198 MT994984.1 |Severe acute respiratory syndrome coro    ····················  ····················  ·····················  ····················  ····················  ····················
16199 MT994985.1 |Severe acute respiratory syndrome coro    ····················  ····················  ·····················  ····················  ····················  ····················
16200 MT994986.1 |Severe acute respiratory syndrome coro    ····················  ····················  ·····················  ····················  ····················  ····················
16201 MT994987.1 |Severe acute respiratory syndrome coro    ····················  ····················  ·····················  ····················  ····················  ····················
16202 MT994988.1 |Severe acute respiratory syndrome coro    ····················  ····················  ·····················  ····················  ····················  ····················
16203 MT994989.1 |Severe acute respiratory syndrome coro    ····················  ····················  ·····················  ····················  ····················  ····················
16204 MT994632.1 |Severe acute respiratory syndrome coro    ····················  ····················  ·····················  ····················  ····················  ····················
16205 MT992723.1 |Severe acute respiratory syndrome coro    ····················  ····················  ·····················  ····················  ····················  ····················
16206 MT992724.1 |Severe acute respiratory syndrome coro    ····················  ·A··················  ·····················  ····················  ····················  ····················
16207 MT992725.1 |Severe acute respiratory syndrome coro    ····················  ·A··················  ·····················  ····················  ····················  ····················
16208 MT992726.1 |Severe acute respiratory syndrome coro    ····················  ····················  ·····················  ····················  ····················  ····················
16209 MT992727.1 |Severe acute respiratory syndrome coro    ····················  ····················  ·····················  ····················  ····················  ····················
16210 MT992728.1 |Severe acute respiratory syndrome coro    ····················  ····················  ·····················  ····················  ····················  ····················
16211 MT992729.1 |Severe acute respiratory syndrome coro    ····················  ····················  ·····················  ····················  ····················  ····················
```

```
16212 MT992730.1 |Severe acute respiratory syndrome coro    ····················  ····················  ····················  ····················  ····················  ····················
16213 MT992731.1 |Severe acute respiratory syndrome coro    ····················  ····················  ····················  ····················  ····················  ·········A··········
16214 MT992732.1 |Severe acute respiratory syndrome coro    ····················  ····················  ····················  ····················  ····················  ····················
16215 MT992733.1 |Severe acute respiratory syndrome coro    ····················  ····················  ····················  ····················  ····················  ····················
16216 MT992734.1 |Severe acute respiratory syndrome coro    ····················  ····················  ····················  ····················  ····················  ····················
16217 MT992735.1 |Severe acute respiratory syndrome coro    ····················  ····················  ····················  ····················  ····················  ····················
16218 MT992736.1 |Severe acute respiratory syndrome coro    ····················  ····················  ····················  ····················  ····················  ····················
16219 MT992737.1 |Severe acute respiratory syndrome coro    ····················  ····················  ····················  ····················  ····················  ····················
16220 MT992738.1 |Severe acute respiratory syndrome coro    ····················  ····················  ····················  ····················  ····················  ····················
16221 MT992739.1 |Severe acute respiratory syndrome coro    ····················  ····················  ····················  ····················  ····················  ····················
16222 MT992740.1 |Severe acute respiratory syndrome coro    ····················  ····················  ····················  ····················  ····················  ····················
16223 MT992741.1 |Severe acute respiratory syndrome coro    ····················  ····················  ····················  ····················  ····················  ····················
16224 MT992742.1 |Severe acute respiratory syndrome coro    ····················  ····················  ····················  ····················  ····················  ····················
16225 MT992743.1 |Severe acute respiratory syndrome coro    ····················  ····················  ····················  ····················  ····················  ····················
16226 MT992744.1 |Severe acute respiratory syndrome coro    ····················  ····················  ····················  ····················  ····················  ····················
16227 MT992745.1 |Severe acute respiratory syndrome coro    ····················  ····················  ····················  ····················  ····················  ····················
16228 MT992746.1 |Severe acute respiratory syndrome coro    ····················  ····················  ····················  ····················  ····················  ····················
16229 MT992747.1 |Severe acute respiratory syndrome coro    ····················  ····················  ····················  ····················  ····················  ····················
16230 MT992748.1 |Severe acute respiratory syndrome coro    ····················  ····················  ····················  ····················  ····················  ····················
16231 MT992749.1 |Severe acute respiratory syndrome coro    ····················  ····················  ····················  ····················  ····················  ····················
16232 MT992750.1 |Severe acute respiratory syndrome coro    ····················  ····················  ····················  ····················  ····················  ····················
16233 MT992751.1 |Severe acute respiratory syndrome coro    ····················  ····················  ····················  ····················  ····················  ····················
16234 MT993864.1 |Severe acute respiratory syndrome coro    ····················  ····················  ····················  ····················  ····················  ····················
16235 MT993866.1 |Severe acute respiratory syndrome coro    ····················  ····················  ····················  ····················  ····················  ····················
16236 MT993868.1 |Severe acute respiratory syndrome coro    ····················  ····················  ····················  ····················  ····················  ····················
16237 MT993870.1 |Severe acute respiratory syndrome coro    ····················  ····················  ····················  ····················  ····················  ····················
16238 MT993871.1 |Severe acute respiratory syndrome coro    ····················  ····················  ····················  ····················  ····················  ····················
16239 MT993872.1 |Severe acute respiratory syndrome coro    ····················  ····················  ····················  ····················  ····················  ····················
16240 MT993873.1 |Severe acute respiratory syndrome coro    ····················  ····················  ····················  ····················  ····················  ····················
16241 MT993874.1 |Severe acute respiratory syndrome coro    ····················  ····················  ····················  ····················  ····················  ····················
16242 MT993875.1 |Severe acute respiratory syndrome coro    ····················  ····················  ····················  ····················  ····················  ····················
16243 MT993876.1 |Severe acute respiratory syndrome coro    ····················  ····················  ····················  ····················  ····················  ····················
16244 MT993877.1 |Severe acute respiratory syndrome coro    ····················  ····················  ····················  ····················  ····················  ····················
16245 MT993878.1 |Severe acute respiratory syndrome coro    ····················  ····················  ····················  ····················  ····················  ····················
16246 MT993879.1 |Severe acute respiratory syndrome coro    ····················  ····················  ····················  ····················  ····················  ····················
16247 MT993880.1 |Severe acute respiratory syndrome coro    ····················  ····················  ····················  ····················  ····················  ····················
16248 MT993881.1 |Severe acute respiratory syndrome coro    ····················  ····················  ····················  ····················  ····················  ····················
16249 MT993882.1 |Severe acute respiratory syndrome coro    ····················  ····················  ····················  ····················  ····················  ····················
16250 MT993883.1 |Severe acute respiratory syndrome coro    ····················  ····················  ····················  ····················  ····················  ····················
16251 MT993884.1 |Severe acute respiratory syndrome coro    ····················  ····················  ····················  ····················  ····················  ····················
16252 MT993885.1 |Severe acute respiratory syndrome coro    ····················  ····················  ····················  ····················  ····················  ····················
16253 MT993886.1 |Severe acute respiratory syndrome coro    ····················  ····················  ····················  ····················  ····················  ····················
16254 MT993887.1 |Severe acute respiratory syndrome coro    ····················  ····················  ····················  ····················  ····················  ····················
16255 MT994286.1 |Severe acute respiratory syndrome coro    ····················  ····················  ····················  ····················  ····················  ····················
16256 MT994393.1 |Severe acute respiratory syndrome coro    ····················  ····················  ····················  ····················  ····················  ····················
16257 MT994394.1 |Severe acute respiratory syndrome coro    ····················  ·A··················  ····················  ····················  ····················  ····················
16258 MT994395.1 |Severe acute respiratory syndrome coro    ····················  ····················  ····················  ····················  ····················  ····················
16259 MT990449.1 |Severe acute respiratory syndrome coro    ····················  ····················  ····················  ····················  ····················  ····················
16260 MT990450.1 |Severe acute respiratory syndrome coro    ····················  ····················  ····················  ····················  ····················  ····················
16261 MT981086.1 |Severe acute respiratory syndrome coro    ····················  ····················  ····················  ····················  ····················  ····················
16262 MT981087.1 |Severe acute respiratory syndrome coro    ····················  ····················  ····················  ····················  ····················  ····················
16263 MT981088.1 |Severe acute respiratory syndrome coro    ····················  ····················  ····················  ····················  ····················  ····················
16264 MT981089.1 |Severe acute respiratory syndrome coro    ····················  ····················  ····················  ····················  ····················  ····················
16265 MT981090.1 |Severe acute respiratory syndrome coro    ····················  ····················  ····················  ····················  ····················  ····················
16266 MT981091.1 |Severe acute respiratory syndrome coro    ····················  ····················  ····················  ····················  ····················  ····················
16267 MT981092.1 |Severe acute respiratory syndrome coro    ····················  ····················  ····················  ····················  ····················  ····················
16268 MT981093.1 |Severe acute respiratory syndrome coro    ····················  ·A··················  ····················  ····················  ····················  ····················
16269 MT981094.1 |Severe acute respiratory syndrome coro    ····················  ·A··················  ····················  ····················  ····················  ····················
16270 MT981095.1 |Severe acute respiratory syndrome coro    ····················  ····················  ····················  ····················  ····················  ····················
16271 MT981096.1 |Severe acute respiratory syndrome coro    ····················  ····················  ····················  ····················  ····················  ····················
16272 MT981097.1 |Severe acute respiratory syndrome coro    ····················  ·A··················  ····················  ····················  ····················  ····················
16273 MT981098.1 |Severe acute respiratory syndrome coro    ····················  ····················  ····················  ····················  ····················  ····················
16274 MT981099.1 |Severe acute respiratory syndrome coro    ····················  ····················  ····················  ····················  ····················  ····················
16275 MT981100.1 |Severe acute respiratory syndrome coro    ····················  ····················  ····················  ····················  ····················  ····················
16276 MT981101.1 |Severe acute respiratory syndrome coro    ····················  ····················  ····················  ····················  ····················  ····················
16277 MT981102.1 |Severe acute respiratory syndrome coro    ····················  ····················  ····················  ····················  ····················  ····················
16278 MT981103.1 |Severe acute respiratory syndrome coro    ····················  ····················  ····················  ····················  ····················  ····················
16279 MT981104.1 |Severe acute respiratory syndrome coro    ····················  ····················  ····················  ····················  ····················  ····················
16280 MT981105.1 |Severe acute respiratory syndrome coro    ····················  ····················  ····················  ····················  ····················  ····················
16281 MT981106.1 |Severe acute respiratory syndrome coro    ····················  ····················  ····················  ····················  ····················  ····················
16282 MT981107.1 |Severe acute respiratory syndrome coro    ····················  ····················  ····················  ····················  ····················  ····················
16283 MT981108.1 |Severe acute respiratory syndrome coro    ····················  ····················  ····················  ····················  ····················  ····················
16284 MT981418.1 |Severe acute respiratory syndrome coro    ····················  ····················  ····················  ····················  ····················  ····················
16285 MT981419.1 |Severe acute respiratory syndrome coro    ····················  ····················  ····················  ····················  ····················  ····················
16286 MT981420.1 |Severe acute respiratory syndrome coro    ····················  ····················  ····················  ····················  ····················  ····················
16288 MT981422.1 |Severe acute respiratory syndrome coro    ····················  ····················  ····················  ····················  ····················  ····················
16289 MT981423.1 |Severe acute respiratory syndrome coro    ····················  ····················  ····················  ····················  ····················  ····················
16290 MT981424.1 |Severe acute respiratory syndrome coro    ····················  ····················  ····················  ····················  ····················  ····················
```

| | | | | | | | |
|---|---|---|---|---|---|---|---|
| 16291 | MT981425.1 \|Severe acute respiratory syndrome coro | ···················· | ···················· | ···················· | ···················· | ···················· | ···················· |
| 16292 | MT981426.1 \|Severe acute respiratory syndrome coro | ···················· | ···················· | ···················· | ···················· | ···················· | ···················· |
| 16293 | MT981427.1 \|Severe acute respiratory syndrome coro | ···················· | ···················· | ···················· | ···················· | ···················· | ···················· |
| 16294 | MT981428.1 \|Severe acute respiratory syndrome coro | ···················· | ···················· | ···················· | ···················· | ···················· | ···················· |
| 16295 | MT981429.1 \|Severe acute respiratory syndrome coro | ···················· | ·········A·········· | ···················· | ···················· | ···················· | ···················· |
| 16296 | MT981430.1 \|Severe acute respiratory syndrome coro | ···················· | ···················· | ···················· | ···················· | ···················· | ···················· |
| 16297 | MT981431.1 \|Severe acute respiratory syndrome coro | ···················· | ···················· | ···················· | ···················· | ···················· | ···················· |
| 16298 | MT981432.1 \|Severe acute respiratory syndrome coro | ···················· | ···················· | ···················· | ···················· | ···················· | ···················· |
| 16299 | MT981433.1 \|Severe acute respiratory syndrome coro | ···················· | ···················· | ···················· | ···················· | ···················· | ···················· |
| 16300 | MT981434.1 \|Severe acute respiratory syndrome coro | ···················· | ···················· | ···················· | ···················· | ···················· | ···················· |
| 16301 | MT981435.1 \|Severe acute respiratory syndrome coro | ···················· | ···················· | ···················· | ···················· | ···················· | ···················· |
| 16302 | MT981436.1 \|Severe acute respiratory syndrome coro | ···················· | ···················· | ···················· | ···················· | ···················· | ···················· |
| 16303 | MT981437.1 \|Severe acute respiratory syndrome coro | ···················· | ···················· | ···················· | ···················· | ···················· | ···················· |
| 16304 | MT981438.1 \|Severe acute respiratory syndrome coro | ···················· | ···················· | ···················· | ···················· | ···················· | ···················· |
| 16305 | MT981439.1 \|Severe acute respiratory syndrome coro | ···················· | ···················· | ···················· | ···················· | ···················· | ···················· |
| 16306 | MT981440.1 \|Severe acute respiratory syndrome coro | ···················· | ···················· | ···················· | ···················· | ···················· | ···················· |
| 16307 | MT981441.1 \|Severe acute respiratory syndrome coro | ···················· | ···················· | ···················· | ···················· | ···················· | ···················· |
| 16308 | MT981449.1 \|Severe acute respiratory syndrome coro | ···················· | ···················· | ···················· | ···················· | ···················· | ···················· |
| 16309 | MT981450.1 \|Severe acute respiratory syndrome coro | ···················· | ···················· | ···················· | ···················· | ···················· | ···················· |
| 16310 | MT981451.1 \|Severe acute respiratory syndrome coro | ···················· | ···················· | ···················· | ···················· | ···················· | ···················· |
| 16311 | MT981452.1 \|Severe acute respiratory syndrome coro | ···················· | ···················· | ···················· | ···················· | ···················· | ···················· |
| 16312 | MT981453.1 \|Severe acute respiratory syndrome coro | ···················· | ···················· | ···················· | ···················· | ···················· | ···················· |
| 16313 | MT981454.1 \|Severe acute respiratory syndrome coro | ···················· | ····A·A············· | ···················· | ···················· | ···················· | ···················· |
| 16314 | MT981455.1 \|Severe acute respiratory syndrome coro | ···················· | ···················· | ···················· | ···················· | ···················· | ···················· |
| 16315 | MT981456.1 \|Severe acute respiratory syndrome coro | ···················· | ···················· | ···················· | ···················· | ···················· | ···················· |
| 16316 | MT981457.1 \|Severe acute respiratory syndrome coro | ···················· | ···················· | ···················· | ···················· | ···················· | ···················· |
| 16317 | MT981459.1 \|Severe acute respiratory syndrome coro | ···················· | ···················· | ···················· | ···················· | ···················· | ···················· |
| 16318 | MT981460.1 \|Severe acute respiratory syndrome coro | ···················· | ···················· | ···················· | ···················· | ···················· | ···················· |
| 16319 | MT981461.1 \|Severe acute respiratory syndrome coro | ···················· | ···················· | ···················· | ···················· | ···················· | ···················· |
| 16320 | MT981462.1 \|Severe acute respiratory syndrome coro | ···················· | ···················· | ···················· | ···················· | ···················· | ···················· |
| 16321 | MT981463.1 \|Severe acute respiratory syndrome coro | ···················· | ···················· | ···················· | ···················· | ···················· | ···················· |
| 16322 | MT981464.1 \|Severe acute respiratory syndrome coro | ···················· | ···················· | ···················· | ···················· | ···················· | ···················· |
| 16323 | MT981465.1 \|Severe acute respiratory syndrome coro | ········T··········· | ···················· | ···················· | ···················· | ···················· | ···················· |
| 16324 | MT981466.1 \|Severe acute respiratory syndrome coro | ···················· | ···················· | ···················· | ···················· | ···················· | ···················· |
| 16325 | MT981467.1 \|Severe acute respiratory syndrome coro | ···················· | ···················· | ···················· | ···················· | ···················· | ···················· |
| 16326 | MT981483.1 \|Severe acute respiratory syndrome coro | ···················· | ···················· | ···················· | ···················· | ···················· | ···················· |
| 16327 | MT981485.1 \|Severe acute respiratory syndrome coro | ···················· | ···················· | ···················· | ···················· | ···················· | ···················· |
| 16328 | MT981487.1 \|Severe acute respiratory syndrome coro | ···················· | ·········A·········· | ···················· | ···················· | ···················· | ···················· |
| 16329 | MT981488.1 \|Severe acute respiratory syndrome coro | ···················· | ···················· | ···················· | ···················· | ···················· | ···················· |
| 16330 | MT981489.1 \|Severe acute respiratory syndrome coro | ···················· | ···················· | ···················· | ···················· | ···················· | ···················· |
| 16331 | MT981490.1 \|Severe acute respiratory syndrome coro | ···················· | ···················· | ···················· | ···················· | ···················· | ···················· |
| 16332 | MT844088.1 \|Mutant Severe acute respiratory syndro | ···················· | ···················· | ···················· | ···················· | ···················· | ···················· |
| 16333 | MT844089.1 \|Mutant Severe acute respiratory syndro | ···················· | ···················· | ···················· | ···················· | ···················· | ···················· |
| 16334 | MT974069.1 \|Severe acute respiratory syndrome coro | ···················· | ···················· | ···················· | ···················· | ···················· | ···················· |
| 16335 | MT974071.1 \|Severe acute respiratory syndrome coro | ···················· | ···················· | ···················· | ···················· | ···················· | ···················· |
| 16336 | MT965767.1 \|Severe acute respiratory syndrome coro | ···················· | ···················· | ···················· | ···················· | ···················· | ···················· |
| 16337 | MT965769.1 \|Severe acute respiratory syndrome coro | ···················· | ·A·················· | ···················· | ···················· | ···················· | ···A················ |
| 16338 | MT966076.1 \|Severe acute respiratory syndrome coro | ···················· | ···················· | ···················· | ···················· | ···················· | ···················· |
| 16339 | MT966077.1 \|Severe acute respiratory syndrome coro | ···················· | ···················· | ···················· | ···················· | ···················· | ···················· |
| 16342 | MT966083.1 \|Severe acute respiratory syndrome coro | ···················· | ···················· | ···················· | ···················· | ···················· | ···················· |
| 16343 | MT966084.1 \|Severe acute respiratory syndrome coro | ···················· | ···················· | ···················· | ···················· | ···················· | ···················· |
| 16344 | MT966086.1 \|Severe acute respiratory syndrome coro | ···················· | ···················· | ···················· | ···················· | ···················· | ···················· |
| 16345 | MT966087.1 \|Severe acute respiratory syndrome coro | ···················· | ···················· | ···················· | ···················· | ···················· | ···················· |
| 16346 | MT966089.1 \|Severe acute respiratory syndrome coro | ···················· | ···················· | ···················· | ···················· | ···················· | ···················· |
| 16347 | MT966091.1 \|Severe acute respiratory syndrome coro | ···················· | ···················· | ···················· | ···················· | ···················· | ···················· |
| 16348 | MT966094.1 \|Severe acute respiratory syndrome coro | ···················· | ···················· | ···················· | ···················· | ···················· | ···················· |
| 16349 | MT966095.1 \|Severe acute respiratory syndrome coro | ···················· | ···················· | ···················· | ···················· | ···················· | ···················· |
| 16350 | MT966096.1 \|Severe acute respiratory syndrome coro | ···················· | ···················· | ···················· | ···················· | ···················· | ···················· |
| 16351 | MT966097.1 \|Severe acute respiratory syndrome coro | ···················· | ···················· | ···················· | ···················· | ···················· | ···················· |
| 16352 | MT966099.1 \|Severe acute respiratory syndrome coro | ···················· | ···················· | ···················· | ···················· | ···················· | ···················· |
| 16354 | MT966101.1 \|Severe acute respiratory syndrome coro | ···················· | ···················· | ···················· | ···················· | ···················· | ···················· |
| 16355 | MT966102.1 \|Severe acute respiratory syndrome coro | ···················· | ···················· | ···················· | ···················· | ···················· | ···················· |
| 16356 | MT966104.1 \|Severe acute respiratory syndrome coro | ···················· | ···················· | ···················· | ···················· | ···················· | ···················· |
| 16357 | MT966105.1 \|Severe acute respiratory syndrome coro | ···················· | ···················· | ···················· | ···················· | ···················· | ···················· |
| 16358 | MT966106.1 \|Severe acute respiratory syndrome coro | ···················· | ···················· | ···················· | ···················· | ···················· | ···················· |
| 16361 | MT966110.1 \|Severe acute respiratory syndrome coro | ···················· | ···················· | ···················· | ···················· | ···················· | ···················· |
| 16362 | MT966113.1 \|Severe acute respiratory syndrome coro | ···················· | ···················· | ···················· | ···················· | ···················· | ···················· |
| 16363 | MT966126.1 \|Severe acute respiratory syndrome coro | ···················· | ···················· | ···················· | ···················· | ···················· | ···················· |
| 16364 | MT966129.1 \|Severe acute respiratory syndrome coro | ···················· | ···················· | ···················· | ···················· | ···················· | ···················· |
| 16365 | MT966133.1 \|Severe acute respiratory syndrome coro | ···················· | ···················· | ···················· | ···················· | ···················· | ···············A···· |
| 16366 | MT966138.1 \|Severe acute respiratory syndrome coro | ···················· | ···················· | ···················· | ···················· | ···················· | ···················· |
| 16368 | MT966141.1 \|Severe acute respiratory syndrome coro | ···················· | ···················· | ···················· | ···················· | ···················· | ···················· |
| 16369 | MT966142.1 \|Severe acute respiratory syndrome coro | ···················· | ···················· | ···················· | ···················· | ···················· | ···················· |
| 16370 | MT966144.1 \|Severe acute respiratory syndrome coro | ···················· | ···················· | ···················· | ···················· | ···················· | ···················· |
| 16371 | MT966145.1 \|Severe acute respiratory syndrome coro | ···················· | ···················· | ···················· | ···················· | ···················· | ···················· |
| 16372 | MT966146.1 \|Severe acute respiratory syndrome coro | ···················· | ···················· | ···················· | ···················· | ···················· | ···················· |
| 16373 | MT966148.1 \|Severe acute respiratory syndrome coro | ···················· | ···················· | ···················· | ···················· | ···················· | ···················· |
| 16374 | MT966149.1 \|Severe acute respiratory syndrome coro | ···················· | ········A··········· | ···················· | ···················· | ···················· | ···················· |

| | | | | | | | | | |
|---|---|---|---|---|---|---|---|---|---|
| 16375 | MT966154.1 | \|Severe acute respiratory syndrome coro | .................... | .................... | .................... | .................... | NNN................. | .................... | 1 |
| 16376 | MT966155.1 | \|Severe acute respiratory syndrome coro | .................... | .................... | .................... | .................... | .................... | .................... | |
| 16377 | MT966156.1 | \|Severe acute respiratory syndrome coro | .................... | .............A...... | .................... | .................... | .................... | .................... | |
| 16378 | MT966157.1 | \|Severe acute respiratory syndrome coro | .................... | .................... | .................... | .................... | .................... | .................... | |
| 16380 | MT966159.1 | \|Severe acute respiratory syndrome coro | .................... | .................... | .................... | .................... | .................... | .................... | |
| 16382 | MT966161.1 | \|Severe acute respiratory syndrome coro | .................... | .................... | .................... | .................... | .................... | .................... | |
| 16383 | MT966162.1 | \|Severe acute respiratory syndrome coro | .................... | .................... | .................... | .................... | .................... | .................... | |
| 16384 | MT966163.1 | \|Severe acute respiratory syndrome coro | .................... | .................... | .................... | .................... | .................... | .................... | |
| 16385 | MT966164.1 | \|Severe acute respiratory syndrome coro | .................... | .................... | .................... | .................... | .................... | .................... | |
| 16387 | MT966167.1 | \|Severe acute respiratory syndrome coro | .................... | .................... | .................... | .................... | .................... | .................... | |
| 16388 | MT966169.1 | \|Severe acute respiratory syndrome coro | .................... | .................... | .................... | .................... | .................... | .................... | |
| 16389 | MT966171.1 | \|Severe acute respiratory syndrome coro | .................... | .................... | .................... | .................... | .................... | .................... | |
| 16390 | MT966174.1 | \|Severe acute respiratory syndrome coro | .................... | .................... | .................... | .................... | .................... | .................... | |
| 16391 | MT966176.1 | \|Severe acute respiratory syndrome coro | .................... | .................... | .................... | .................... | .................... | .................... | |
| 16392 | MT966177.1 | \|Severe acute respiratory syndrome coro | .................... | .................... | .................... | .................... | NNN................. | .................... | 1 |
| 16393 | MT966178.1 | \|Severe acute respiratory syndrome coro | .................... | .................... | .................... | .................... | .................... | .................... | |
| 16394 | MT966179.1 | \|Severe acute respiratory syndrome coro | .................... | .................... | .................... | .................... | .................... | .................... | |
| 16395 | MT966180.1 | \|Severe acute respiratory syndrome coro | .................... | .................... | .................... | .................... | .................... | .................... | |
| 16396 | MT966181.1 | \|Severe acute respiratory syndrome coro | .................... | .................... | .................... | .................... | .................... | .................... | |
| 16397 | MT966183.1 | \|Severe acute respiratory syndrome coro | .................... | .................... | .................... | .................... | .................... | .................... | |
| 16400 | MT966191.1 | \|Severe acute respiratory syndrome coro | .................... | .................... | .................... | .................... | .................... | .................... | |
| 16401 | MT966192.1 | \|Severe acute respiratory syndrome coro | .................... | .................... | .................... | .................... | .................... | .................... | |
| 16402 | MT966193.1 | \|Severe acute respiratory syndrome coro | .................... | .................... | .................... | .................... | .................... | .................... | |
| 16403 | MT966195.1 | \|Severe acute respiratory syndrome coro | .................... | .................... | .................... | .................... | .................... | .................... | |
| 16404 | MT966197.1 | \|Severe acute respiratory syndrome coro | .................... | .................... | .................... | .................... | .................... | .................... | |
| 16405 | MT966199.1 | \|Severe acute respiratory syndrome coro | .................... | .................... | .................... | .................... | .................... | .................... | |
| 16406 | MT966201.1 | \|Severe acute respiratory syndrome coro | .................... | .................... | .................... | .................... | .................... | .................... | |
| 16407 | MT966204.1 | \|Severe acute respiratory syndrome coro | .................... | .................... | .................... | .................... | .................... | .................... | |
| 16408 | MT966205.1 | \|Severe acute respiratory syndrome coro | .................... | .................... | .................... | .................... | .................... | .................... | |
| 16409 | MT966206.1 | \|Severe acute respiratory syndrome coro | .................... | .................... | .................... | .................... | .................... | .................... | |
| 16410 | MT966209.1 | \|Severe acute respiratory syndrome coro | .................... | .................... | .................... | .................... | .................... | .................... | |
| 16411 | MT966210.1 | \|Severe acute respiratory syndrome coro | .................... | .................... | .................... | .................... | .................... | .................... | |
| 16412 | MT966211.1 | \|Severe acute respiratory syndrome coro | .................... | .................... | .................... | .................... | .................... | .................... | |
| 16413 | MT966212.1 | \|Severe acute respiratory syndrome coro | .................... | .................... | .................... | .................... | .................... | .................... | |
| 16414 | MT966213.1 | \|Severe acute respiratory syndrome coro | .................... | .................... | .................... | .................... | .................... | .................... | |
| 16415 | MT966214.1 | \|Severe acute respiratory syndrome coro | .................... | .................... | .................... | .................... | .................... | .................... | |
| 16416 | MT966216.1 | \|Severe acute respiratory syndrome coro | .................... | .................... | .................... | .................... | .................... | .................... | |
| 16417 | MT966217.1 | \|Severe acute respiratory syndrome coro | .................... | .................... | .................... | .................... | .................... | .................... | |
| 16418 | MT966218.1 | \|Severe acute respiratory syndrome coro | .................... | .................... | .................... | .................... | .................... | .................... | |
| 16419 | MT966219.1 | \|Severe acute respiratory syndrome coro | .................... | .................... | .................... | .................... | .................... | .................... | |
| 16420 | MT966223.1 | \|Severe acute respiratory syndrome coro | .................... | .................... | .................... | .................... | .................... | .................... | |
| 16421 | MT966224.1 | \|Severe acute respiratory syndrome coro | .................... | .................... | .................... | .................... | .................... | .................... | |
| 16422 | MT966225.1 | \|Severe acute respiratory syndrome coro | .................... | .................... | .................... | .................... | .................... | .................... | |
| 16423 | MT966226.1 | \|Severe acute respiratory syndrome coro | .................... | .................... | .................... | .................... | .................... | .................... | |
| 16424 | MT966227.1 | \|Severe acute respiratory syndrome coro | .................... | .................... | .................... | .................... | .................... | .................... | |
| 16425 | MT966228.1 | \|Severe acute respiratory syndrome coro | .................... | .................... | .................... | .................... | .................... | .................... | |
| 16427 | MT966230.1 | \|Severe acute respiratory syndrome coro | .................... | .................... | .................... | .................... | .................... | .................... | |
| 16428 | MT966231.1 | \|Severe acute respiratory syndrome coro | .................... | .................... | .................... | .................... | .................... | .................... | |
| 16429 | MT966233.1 | \|Severe acute respiratory syndrome coro | .................... | .................... | .................... | .................... | .................... | .................... | |
| 16430 | MT966234.1 | \|Severe acute respiratory syndrome coro | .................... | .............A...... | .................... | .................... | .................... | .................... | |
| 16432 | MT966238.1 | \|Severe acute respiratory syndrome coro | .................... | .................... | .................... | .................... | .................... | .................... | |
| 16433 | MT966239.1 | \|Severe acute respiratory syndrome coro | .................... | .................... | .................... | .................... | .................... | .................... | |
| 16434 | MT966241.1 | \|Severe acute respiratory syndrome coro | .................... | .................... | .................... | .................... | .................... | .................... | |
| 16435 | MT966243.1 | \|Severe acute respiratory syndrome coro | .................... | .................... | .................... | .................... | .................... | .................... | |
| 16436 | MT966244.1 | \|Severe acute respiratory syndrome coro | .................... | .................... | .................... | .................... | .................... | .................... | |
| 16437 | MT966245.1 | \|Severe acute respiratory syndrome coro | .................... | .................... | .................... | .................... | .................... | .................... | |
| 16438 | MT966246.1 | \|Severe acute respiratory syndrome coro | .................... | .................... | .................... | .................... | .................... | .................... | |
| 16439 | MT966247.1 | \|Severe acute respiratory syndrome coro | .................... | .................... | .................... | .................... | .................... | .................... | |
| 16440 | MT966248.1 | \|Severe acute respiratory syndrome coro | .................... | .................... | .................... | .................... | .................... | .................... | |
| 16441 | MT966249.1 | \|Severe acute respiratory syndrome coro | .................... | .................... | .................... | .................... | .................... | .................... | |
| 16442 | MT966251.1 | \|Severe acute respiratory syndrome coro | .................... | .................... | .................... | .................... | .................... | .................... | |
| 16443 | MT966252.1 | \|Severe acute respiratory syndrome coro | .................... | .................... | .................... | .................... | .................... | .................... | |
| 16445 | MT966254.1 | \|Severe acute respiratory syndrome coro | .................... | .................... | .................... | .................... | .................... | .................... | |
| 16446 | MT966255.1 | \|Severe acute respiratory syndrome coro | .................... | .................... | .................... | .................... | .................... | .................... | |
| 16447 | MT966256.1 | \|Severe acute respiratory syndrome coro | .................... | .................... | .................... | .................... | .................... | .................... | |
| 16448 | MT966257.1 | \|Severe acute respiratory syndrome coro | .................... | .................... | .................... | .................... | .................... | .................... | |
| 16449 | MT966262.1 | \|Severe acute respiratory syndrome coro | .................... | .................... | .................... | .................... | .................... | .................... | |
| 16450 | MT967899.1 | \|Severe acute respiratory syndrome coro | .................... | .................... | .................... | .................... | .................... | .................... | |
| 16451 | MT967900.1 | \|Severe acute respiratory syndrome coro | .................... | .................... | .................... | .................... | .................... | .................... | |
| 16452 | MT967901.1 | \|Severe acute respiratory syndrome coro | .................... | .................... | .................... | .................... | .................... | .................... | |
| 16453 | MT967902.1 | \|Severe acute respiratory syndrome coro | .................... | .................... | .................... | .................... | .................... | .................... | |
| 16454 | MT967903.1 | \|Severe acute respiratory syndrome coro | .................... | .................... | .................... | .................... | .................... | .................... | |
| 16455 | MT967904.1 | \|Severe acute respiratory syndrome coro | .................... | .................... | .................... | .................... | .................... | .................... | |
| 16456 | MT967905.1 | \|Severe acute respiratory syndrome coro | .................... | .................... | .................... | .................... | .................... | .................... | |
| 16457 | MT967906.1 | \|Severe acute respiratory syndrome coro | .................... | .................... | .................... | .................... | .................... | .................... | |
| 16458 | MT967907.1 | \|Severe acute respiratory syndrome coro | .................... | .................... | .................... | .................... | .................... | .................... | |
| 16459 | MT967908.1 | \|Severe acute respiratory syndrome coro | .................... | .................... | .................... | .................... | .................... | .................... | |
| 16460 | MT967909.1 | \|Severe acute respiratory syndrome coro | .................... | .................... | .................... | .................... | .................... | .................... | |

16461 MT967910.1 |Severe acute respiratory syndrome coro ···················· ···················· ····················· ···················· ···················· ····················
16462 MT967918.1 |Severe acute respiratory syndrome coro ···················· ···················· ····················· ···················· ···················· ····················
16463 MT967919.1 |Severe acute respiratory syndrome coro ···················· ···················· ····················· ···················· ···················· ····················
16464 MT967920.1 |Severe acute respiratory syndrome coro ···················· ···················· ····················· ···················· ···················· ····················
16465 MT967921.1 |Severe acute respiratory syndrome coro ···················· ···················· ····················· ···················· ···················· ····················
16466 MT967922.1 |Severe acute respiratory syndrome coro ···················· ···················· ····················· ···················· ···················· ····················
16467 MT967923.1 |Severe acute respiratory syndrome coro ···················· ···················· ····················· ···················· ···················· ····················
16468 MT967924.1 |Severe acute respiratory syndrome coro ···················· ···················· ····················· ···················· ···················· ····················
16469 MT967925.1 |Severe acute respiratory syndrome coro ···················· ···················· ····················· ···················· ···················· ····················
16470 MT967926.1 |Severe acute respiratory syndrome coro ···················· ···················· ····················· ···················· ···················· ····················
16471 MT967927.1 |Severe acute respiratory syndrome coro ···················· ···················· ····················· ···················· ···················· ····················
16472 MT967928.1 |Severe acute respiratory syndrome coro ···················· ···················· ····················· ···················· ···················· ····················
16473 MT967929.1 |Severe acute respiratory syndrome coro ···················· ···················· ····················· ···················· ···················· ····················
16474 MT967930.1 |Severe acute respiratory syndrome coro ···················· ···················· ····················· ···················· ···················· ····················
16475 MT967931.1 |Severe acute respiratory syndrome coro ···················· ···················· ····················· ···················· ···················· ····················
16476 MT967932.1 |Severe acute respiratory syndrome coro ···················· ···················· ····················· ···················· ···················· ····················
16477 MT967933.1 |Severe acute respiratory syndrome coro ···················· ···················· ····················· ···················· ···················· ····················
16478 MT967934.1 |Severe acute respiratory syndrome coro ···················· ···················· ····················· ···················· ···················· ····················
16479 MT967935.1 |Severe acute respiratory syndrome coro ···················· ···················· ····················· ···················· ···················· ····················
16480 MT967936.1 |Severe acute respiratory syndrome coro ···················· ···················· ····················· ···················· ···················· ····················
16481 MT967937.1 |Severe acute respiratory syndrome coro ···················· ···················· ····················· ···················· ···················· ····················
16482 MT967938.1 |Severe acute respiratory syndrome coro ···················· ···················· ····················· ···················· ···················· ····················
16483 MT967939.1 |Severe acute respiratory syndrome coro ···················· ···················· ····················· ···················· ···················· ····················
16484 MT967940.1 |Severe acute respiratory syndrome coro ···················· ···················· ····················· ···················· ···················· ····················
16485 MT967941.1 |Severe acute respiratory syndrome coro ···················· ···················· ····················· ···················· ···················· ····················
16486 MT967942.1 |Severe acute respiratory syndrome coro ···················· ···················· ····················· ···················· ···················· ····················
16487 MT967943.1 |Severe acute respiratory syndrome coro ···················· ···················· ····················· ···················· ···················· ····················
16488 MT967944.1 |Severe acute respiratory syndrome coro ···················· ···················· ····················· ···················· ···················· ····················
16489 MT967945.1 |Severe acute respiratory syndrome coro ···················· ···················· ····················· ···················· ···················· ····················
16490 MT967946.1 |Severe acute respiratory syndrome coro ···················· ···················· ····················· ···················· ···················· ····················
16491 MT967947.1 |Severe acute respiratory syndrome coro ···················· ···················· ····················· ···················· ···················· ····················
16492 MT967948.1 |Severe acute respiratory syndrome coro ···················· ···················· ····················· ···················· ···················· ····················
16493 MT967949.1 |Severe acute respiratory syndrome coro ···················· ···················· ····················· ···················· ···················· ····················
16494 MT967950.1 |Severe acute respiratory syndrome coro ···················· ···················· ····················· ···················· ···················· ····················
16495 MT967951.1 |Severe acute respiratory syndrome coro ···················· ·A·················· ····················· ···················· ···················· ····················
16496 MT967952.1 |Severe acute respiratory syndrome coro ···················· ···················· ····················· ···················· ···················· ····················
16497 MT967953.1 |Severe acute respiratory syndrome coro ···················· ···················· ····················· ···················· ···················· ····················
16498 MT967954.1 |Severe acute respiratory syndrome coro ···················· ···················· ····················· ···················· ···················· ····················
16499 MT967955.1 |Severe acute respiratory syndrome coro ···················· ···················· ····················· ···················· ···················· ····················
16500 MT967956.1 |Severe acute respiratory syndrome coro ···················· ···················· ····················· ···················· ···················· ····················
16501 MT967957.1 |Severe acute respiratory syndrome coro ···················· ···················· ····················· ···················· ···················· ····················
16502 MT967958.1 |Severe acute respiratory syndrome coro ···················· ···················· ····················· ···················· ···················· ····················
16503 MT967959.1 |Severe acute respiratory syndrome coro ···················· ···················· ····················· ···················· ···················· ····················
16504 MT967960.1 |Severe acute respiratory syndrome coro ···················· ···················· ····················· ···················· ···················· ····················
16505 MT967961.1 |Severe acute respiratory syndrome coro ···················· ···················· ····················· ···················· ···················· ····················
16506 MT967962.1 |Severe acute respiratory syndrome coro ···················· ·A·················· ····················· ···················· ···················· ····················
16507 MT967963.1 |Severe acute respiratory syndrome coro ···················· ···················· ····················· ···················· ···················· ····················
16508 MT967964.1 |Severe acute respiratory syndrome coro ···················· ···················· ····················· ···················· ···················· ····················
16509 MT967965.1 |Severe acute respiratory syndrome coro ···················· ···················· ····················· ···················· ···················· ····················
16510 MT967966.1 |Severe acute respiratory syndrome coro ···················· ···················· ····················· ···················· ···················· ····················
16511 MT967967.1 |Severe acute respiratory syndrome coro ···················· ···················· ····················· ···················· ···················· ····················
16512 MT967968.1 |Severe acute respiratory syndrome coro ···················· ···················· ····················· ···················· ···················· ····················
16513 MT967969.1 |Severe acute respiratory syndrome coro ···················· ···················· ····················· ···················· ···················· ····················
16514 MT967970.1 |Severe acute respiratory syndrome coro ···················· ···················· ····················· ···················· ···················· ····················
16515 MT967971.1 |Severe acute respiratory syndrome coro ···················· ···················· ····················· ···················· ···················· ····················
16516 MT967972.1 |Severe acute respiratory syndrome coro ···················· ···················· ····················· ···················· ···················· ····················
16517 MT967973.1 |Severe acute respiratory syndrome coro ···················· ···················· ····················· ···················· ···················· ····················
16518 MT967974.1 |Severe acute respiratory syndrome coro ···················· ···················· ····················· ···················· ···················· ····················
16519 MT967975.1 |Severe acute respiratory syndrome coro ···················· ···················· ····················· ···················· ···················· ····················
16520 MT967976.1 |Severe acute respiratory syndrome coro ···················· ···················· ···A················· ···················· ···················· ····················
16521 MT967977.1 |Severe acute respiratory syndrome coro ···················· ···················· ···A················· ···················· ···················· ····················
16522 MT967978.1 |Severe acute respiratory syndrome coro ···················· ···················· ····················· ···················· ···················· ····················
16523 MT967979.1 |Severe acute respiratory syndrome coro ···················· ···················· ····················· ···················· ···················· ····················
16524 MT967980.1 |Severe acute respiratory syndrome coro ···················· ···················· ····················· ···················· ···················· ····················
16525 MT967981.1 |Severe acute respiratory syndrome coro ···················· ···················· ····················· ···················· ···················· ····················
16526 MT967982.1 |Severe acute respiratory syndrome coro ···················· ···················· ····················· ···················· ···················· ····················
16527 MT967983.1 |Severe acute respiratory syndrome coro ···················· ···················· ····················· ···················· ···················· ····················
16528 MT967984.1 |Severe acute respiratory syndrome coro ···················· ···················· ····················· ···················· ···················· ····················
16529 MT968051.1 |Severe acute respiratory syndrome coro ···················· ···················· ····················· ···················· ···················· ····················
16530 MT968052.1 |Severe acute respiratory syndrome coro ···················· ···················· ····················· ···················· ···················· ····················
16531 MT968053.1 |Severe acute respiratory syndrome coro ···················· ···················· ····················· ···················· ···················· ····················
16532 MT968054.1 |Severe acute respiratory syndrome coro ···················· ···················· ····················· ···················· ···················· ····················
16533 MT968055.1 |Severe acute respiratory syndrome coro ···················· ···················· ····················· ···················· ···················· ····················
16534 MT968057.1 |Severe acute respiratory syndrome coro ···················· ···················· ····················· ···················· ···················· ····················
16535 MT968058.1 |Severe acute respiratory syndrome coro ···················· ···················· ····················· ···················· ···················· ····················
16536 MT968059.1 |Severe acute respiratory syndrome coro ···················· ···················· ····················· ···················· ···················· ····················
16537 MT968060.1 |Severe acute respiratory syndrome coro ···················· ···················· ····················· ···················· ···················· ····················
16538 MT968061.1 |Severe acute respiratory syndrome coro ···················· ···················· ····················· ···················· ···················· ····················

```
16539 MT968062.1 |Severe acute respiratory syndrome coro    ....................  ....................  .....................  ....................  ....................  ....................
16540 MT968063.1 |Severe acute respiratory syndrome coro    ....................  ....................  .....................  ....................  ....................  ....................
16541 MT968064.1 |Severe acute respiratory syndrome coro    ....................  ....................  .....................  ....................  ....................  ....................
16542 MT968065.1 |Severe acute respiratory syndrome coro    ....................  ....................  .....................  ....................  ....................  ....................
16543 MT968066.1 |Severe acute respiratory syndrome coro    ....................  ....................  .....................  ....................  ....................  ....................
16544 MT968067.1 |Severe acute respiratory syndrome coro    ....................  ....................  .....................  ....................  ....................  ....................
16545 MT968068.1 |Severe acute respiratory syndrome coro    ....................  ....................  .....................  ....................  ....................  ....................
16546 MT968069.1 |Severe acute respiratory syndrome coro    ....................  ....................  .....................  ....................  ....................  ....................
16547 MT968070.1 |Severe acute respiratory syndrome coro    ....................  ....................  .....................  ....................  ....................  ....................
16548 MT968071.1 |Severe acute respiratory syndrome coro    ....................  ....................  .....................  ....................  ....................  ....................
16549 MT968072.1 |Severe acute respiratory syndrome coro    ....................  ....................  .....................  ....................  ....................  ....................
16550 MT968073.1 |Severe acute respiratory syndrome coro    ....................  ....................  .....................  ....................  ....................  ....................
16551 MT968074.1 |Severe acute respiratory syndrome coro    ....................  ....................  .....................  ....................  ....................  ....................
16552 MT968075.1 |Severe acute respiratory syndrome coro    ....................  ....................  .....................  ....................  ....................  ....................
16553 MT968076.1 |Severe acute respiratory syndrome coro    ....................  ....................  .....................  ....................  ....................  ....................
16554 MT968077.1 |Severe acute respiratory syndrome coro    ....................  ....................  .....................  ....................  ....................  ....................
16555 MT969352.1 |Severe acute respiratory syndrome coro    ....................  ....................  .....................  ....................  ....................  ....................
16556 MT969354.1 |Severe acute respiratory syndrome coro    ....................  ....................  .....................  ....................  ....................  ....................
16558 MT969357.1 |Severe acute respiratory syndrome coro    ....................  ....................  .....................  ....................  ....................  ....................
16559 MT969358.1 |Severe acute respiratory syndrome coro    ....................  ....................  .....................  ....................  ....................  ....................
16560 MT969359.1 |Severe acute respiratory syndrome coro    ....................  ....................  .....................  ....................  ....................  ....................
16561 MT969361.1 |Severe acute respiratory syndrome coro    ....................  ....................  .....................  ....................  ....................  ....................
16562 MT969363.1 |Severe acute respiratory syndrome coro    ....................  ....................  .....................  ....................  ....................  ....................
16563 MT969364.1 |Severe acute respiratory syndrome coro    ....................  ....................  .....................  ....................  ....................  ....................
16564 MT969366.1 |Severe acute respiratory syndrome coro    ....................  ....................  .....................  ....................  ....................  ....................
16565 MT969367.1 |Severe acute respiratory syndrome coro    ....................  ....................  .....................  ....................  ....................  ....................
16566 MT969368.1 |Severe acute respiratory syndrome coro    ....................  ....................  .....................  ....................  ....................  ....................
16567 MT969369.1 |Severe acute respiratory syndrome coro    ....................  ....................  .....................  ....................  ....................  ....................
16568 MT969370.1 |Severe acute respiratory syndrome coro    ....................  ....................  .....................  ....................  ....................  ....................
16569 MT969371.1 |Severe acute respiratory syndrome coro    ....................  ....................  .....................  ....................  ....................  ....................
16570 MT969372.1 |Severe acute respiratory syndrome coro    ....................  ....................  .....................  ....................  ....................  ....................
16571 MT969373.1 |Severe acute respiratory syndrome coro    ....................  ....................  .....................  ....................  ....................  ....................
16572 MT969374.1 |Severe acute respiratory syndrome coro    ....................  ....................  .....................  ....................  ....................  ....................
16573 MT969375.1 |Severe acute respiratory syndrome coro    ....................  ....................  .....................  ....................  ....................  ....................
16574 MT969376.1 |Severe acute respiratory syndrome coro    ....................  ....................  .....................  ....................  ....................  ....................
16575 MT969377.1 |Severe acute respiratory syndrome coro    ....................  ....................  .....................  ....................  ....................  ....................
16577 MT969379.1 |Severe acute respiratory syndrome coro    ....................  ....................  .....................  ....................  ....................  ....................
16578 MT969380.1 |Severe acute respiratory syndrome coro    ....................  ....................  .....................  ....................  ....................  ....................
16579 MT969381.1 |Severe acute respiratory syndrome coro    ....................  ....................  .....................  ....................  ....................  ....................
16580 MT969382.1 |Severe acute respiratory syndrome coro    ....................  ....................  .....................  ....................  ....................  ....................
16581 MT969383.1 |Severe acute respiratory syndrome coro    ....................  ....................  .....................  ....................  ....................  ....................
16582 MT969384.1 |Severe acute respiratory syndrome coro    ....................  ....................  .....................  ....................  ....................  ....................
16583 MT969385.1 |Severe acute respiratory syndrome coro    ....................  ....................  .....................  ....................  ....................  ....................
16584 MT969386.1 |Severe acute respiratory syndrome coro    ....................  ....................  .....................  ....................  ....................  ....................
16585 MT969387.1 |Severe acute respiratory syndrome coro    ....................  ....................  .....................  ....................  ....................  ....................
16586 MT969388.1 |Severe acute respiratory syndrome coro    ....................  ....................  .....................  ....................  ....................  ....................
16587 MT969389.1 |Severe acute respiratory syndrome coro    ....................  ....................  .....................  ....................  ....................  ....................
16588 MT969390.1 |Severe acute respiratory syndrome coro    ....................  ....................  .....................  ....................  ....................  ....................
16589 MT969391.1 |Severe acute respiratory syndrome coro    ....................  ....................  .....................  ....................  ....................  ....................
16590 MT969393.1 |Severe acute respiratory syndrome coro    ....................  ....................  .....................  ....................  ....................  ....................
16591 MT969395.1 |Severe acute respiratory syndrome coro    ....................  ....................  .....................  ....................  ....................  ....................
16592 MT969397.1 |Severe acute respiratory syndrome coro    ....................  ....................  .....................  ....................  ....................  ....................
16593 MT969398.1 |Severe acute respiratory syndrome coro    ....................  ....................  .....................  ....................  ....................  ....................
16594 MT969399.1 |Severe acute respiratory syndrome coro    ....................  ....................  .....................  ....................  ....................  ....................
16595 MT969400.1 |Severe acute respiratory syndrome coro    ....................  ....................  .....................  ....................  ....................  ....................
16596 MT969401.1 |Severe acute respiratory syndrome coro    ....................  ....................  .....................  ....................  ....................  ....................
16597 MT969402.1 |Severe acute respiratory syndrome coro    ....................  ....................  .....................  ....................  ....................  ....................
16598 MT969403.1 |Severe acute respiratory syndrome coro    ....................  ....................  .....................  ....................  ....................  ....................
16599 MT969404.1 |Severe acute respiratory syndrome coro    ....................  ....................  .....................  ....................  ....................  ....................
16601 MT969408.1 |Severe acute respiratory syndrome coro    ....................  ....................  .....................  ....................  ....................  ....................
16602 MT969409.1 |Severe acute respiratory syndrome coro    ....................  ....................  .....................  ....................  ....................  ....................
16604 MT969411.1 |Severe acute respiratory syndrome coro    ....................  ....................  .....................  ....................  ....................  ....................
16605 MT969412.1 |Severe acute respiratory syndrome coro    ....................  ....................  .....................  ....................  ....................  ....................
16607 MT969414.1 |Severe acute respiratory syndrome coro    ....................  ....................  .....................  ....................  ....................  ....................
16608 MT969415.1 |Severe acute respiratory syndrome coro    ....................  ....................  .....................  ....................  ....................  ....................
16610 MT969417.1 |Severe acute respiratory syndrome coro    ....................  ....................  .....................  ....................  ....................  ....................
16611 MT969418.1 |Severe acute respiratory syndrome coro    ....................  ....................  .....................  ....................  ....................  ....................
16612 MT969419.1 |Severe acute respiratory syndrome coro    ....................  ....................  .....................  ....................  ....................  ....................
16613 MT969421.1 |Severe acute respiratory syndrome coro    ....................  ....................  .....................  ....................  ....................  ....................
16614 MT969422.1 |Severe acute respiratory syndrome coro    ....................  ....................  .....................  ....................  ....................  ....................
16615 MT969423.1 |Severe acute respiratory syndrome coro    ....................  ....................  .....................  ....................  ....................  ....................
16616 MT969424.1 |Severe acute respiratory syndrome coro    ....................  ....................  .....................  ....................  ....................  ....................
16617 MT969425.1 |Severe acute respiratory syndrome coro    ....................  ....................  .....................  ....................  ....................  ....................
16618 MT969426.1 |Severe acute respiratory syndrome coro    ....................  ....................  .....................  ....................  ....................  ....................
16619 MT969427.1 |Severe acute respiratory syndrome coro    ....................  ....................  .....................  ....................  ....................  ....................
16620 MT969428.1 |Severe acute respiratory syndrome coro    ....................  ....................  .....................  ....................  ....................  ....................
16621 MT969429.1 |Severe acute respiratory syndrome coro    ....................  ....................  .....................  ....................  ....................  ....................
16622 MT969430.1 |Severe acute respiratory syndrome coro    ....................  ....................  .....................  ....................  ....................  ....................
```

| | | | | | | | | | |
|---|---|---|---|---|---|---|---|---|---|
| 16623 | MT969431.1 | \|Severe acute respiratory syndrome coro | .................... | .................... | .................... | .................... | .................... | .................... | |
| 16624 | MT969432.1 | \|Severe acute respiratory syndrome coro | .................... | .................... | .................... | .................... | .................... | .................... | |
| 16625 | MT969433.1 | \|Severe acute respiratory syndrome coro | .................... | .................... | .................... | .................... | .................... | .................... | |
| 16627 | MT969436.1 | \|Severe acute respiratory syndrome coro | .................... | .................... | .................... | .................... | NN.................. | .................... | |
| 16628 | MT969437.1 | \|Severe acute respiratory syndrome coro | .................... | .................... | .................... | .................... | N................... | .................... | |
| 16629 | MT969438.1 | \|Severe acute respiratory syndrome coro | .................... | .................... | .................... | .................... | .................... | .................... | |
| 16630 | MT969439.1 | \|Severe acute respiratory syndrome coro | .................... | .................... | .................... | .................... | .................... | .................... | |
| 16631 | MT969440.1 | \|Severe acute respiratory syndrome coro | .................... | .................... | .................... | .................... | NNN................. | .................... | 1 |
| 16632 | MT969441.1 | \|Severe acute respiratory syndrome coro | .................... | .................... | .................... | .................... | .................... | .................... | |
| 16633 | MT969443.1 | \|Severe acute respiratory syndrome coro | .................... | .................... | .................... | .................... | .................... | .................... | |
| 16634 | MT969445.1 | \|Severe acute respiratory syndrome coro | .................... | .................... | .................... | .................... | .................... | .................... | |
| 16636 | MT969447.1 | \|Severe acute respiratory syndrome coro | .................... | .................... | .................... | .................... | .................... | .................... | |
| 16637 | MT969448.1 | \|Severe acute respiratory syndrome coro | .................... | .................... | .................... | .................... | .................... | .................... | |
| 16638 | MT969449.1 | \|Severe acute respiratory syndrome coro | .................... | .................... | .................... | .................... | .................... | .................... | |
| 16640 | MT969451.1 | \|Severe acute respiratory syndrome coro | .................... | .................... | .................... | .................... | .................... | .................... | |
| 16641 | MT969452.1 | \|Severe acute respiratory syndrome coro | .................... | .................... | .................... | .................... | .................... | .................... | |
| 16646 | MT969457.1 | \|Severe acute respiratory syndrome coro | .................... | .................... | .................... | .................... | ..NN................ | .................... | |
| 16647 | MT969458.1 | \|Severe acute respiratory syndrome coro | .................... | .................... | .................... | .................... | NNN................. | .................... | 1 |
| 16648 | MT969459.1 | \|Severe acute respiratory syndrome coro | .................... | .................... | .................... | .................... | .................... | .................... | |
| 16649 | MT969460.1 | \|Severe acute respiratory syndrome coro | .................... | .................... | .................... | .................... | .NN................. | .................... | |
| 16651 | MT969463.1 | \|Severe acute respiratory syndrome coro | .................... | .................... | .................... | .................... | .................... | .................... | |
| 16652 | MT969464.1 | \|Severe acute respiratory syndrome coro | .................... | .................... | .................... | .................... | .................... | .................... | |
| 16653 | MT969465.1 | \|Severe acute respiratory syndrome coro | .................... | .................... | .................... | .................... | .................... | .................... | |
| 16654 | MT969466.1 | \|Severe acute respiratory syndrome coro | .................... | .................... | .................... | .................... | .................... | .................... | |
| 16655 | MT969467.1 | \|Severe acute respiratory syndrome coro | .................... | .................... | .................... | .................... | .................... | .................... | |
| 16656 | MT969468.1 | \|Severe acute respiratory syndrome coro | .................... | .................... | .................... | .................... | .................... | .................... | |
| 16657 | MT969469.1 | \|Severe acute respiratory syndrome coro | .................... | .................... | .................... | .................... | .................... | .................... | |
| 16658 | MT969470.1 | \|Severe acute respiratory syndrome coro | .................... | .................... | .................... | .................... | .................... | .................... | |
| 16659 | MT969473.1 | \|Severe acute respiratory syndrome coro | .................... | .................... | .................... | .................... | .................... | .................... | |
| 16660 | MT969475.1 | \|Severe acute respiratory syndrome coro | .................... | .................... | .................... | .................... | .................... | .................... | |
| 16661 | MT969476.1 | \|Severe acute respiratory syndrome coro | .................... | .................... | .................... | .................... | .................... | .................... | |
| 16662 | MT969477.1 | \|Severe acute respiratory syndrome coro | .................... | .................... | .................... | .................... | .................... | .................... | |
| 16663 | MT969478.1 | \|Severe acute respiratory syndrome coro | .................... | .................... | .................... | .................... | .................... | .................... | |
| 16664 | MT969479.1 | \|Severe acute respiratory syndrome coro | .................... | .................... | .................... | .................... | .................... | .................... | |
| 16665 | MT969480.1 | \|Severe acute respiratory syndrome coro | .................... | .................... | .................... | .................... | .................... | .................... | |
| 16667 | MT969482.1 | \|Severe acute respiratory syndrome coro | .................... | .................... | .................... | .................... | .................... | .................... | |
| 16668 | MT969483.1 | \|Severe acute respiratory syndrome coro | .................... | .................... | .................... | .................... | .................... | .................... | |
| 16669 | MT969484.1 | \|Severe acute respiratory syndrome coro | .................... | .................... | .................... | .................... | .................... | .................... | |
| 16670 | MT969485.1 | \|Severe acute respiratory syndrome coro | .................... | .................... | .................... | .................... | .................... | .................... | |
| 16671 | MT969488.1 | \|Severe acute respiratory syndrome coro | .................... | .................... | .................... | .................... | .................... | .................... | |
| 16672 | MT969489.1 | \|Severe acute respiratory syndrome coro | .................... | .................... | .................... | .................... | .................... | .................... | |
| 16673 | MT969490.1 | \|Severe acute respiratory syndrome coro | .................... | .................... | .................... | .................... | .................... | .................... | |
| 16674 | MT969491.1 | \|Severe acute respiratory syndrome coro | .................... | .................... | .................... | .................... | .................... | .................... | |
| 16675 | MT969492.1 | \|Severe acute respiratory syndrome coro | .................... | .................... | .................... | .................... | .................... | .................... | |
| 16676 | MT969493.1 | \|Severe acute respiratory syndrome coro | .................... | .................... | .................... | .................... | .................... | .................... | |
| 16677 | MT969494.1 | \|Severe acute respiratory syndrome coro | .................... | .................... | .................... | .................... | .................... | .................... | |
| 16678 | MT969495.1 | \|Severe acute respiratory syndrome coro | .................... | .................... | .................... | .................... | .................... | .................... | |
| 16679 | MT969496.1 | \|Severe acute respiratory syndrome coro | .................... | .................... | .................... | .................... | .................... | .................... | |
| 16680 | MT969497.1 | \|Severe acute respiratory syndrome coro | .................... | .................... | .................... | .................... | .................... | .................... | |
| 16682 | MT969499.1 | \|Severe acute respiratory syndrome coro | .................... | .................... | .................... | .................... | .................... | .................... | |
| 16683 | MT969500.1 | \|Severe acute respiratory syndrome coro | .................... | .................... | .................... | .................... | .................... | .................... | |
| 16684 | MT969501.1 | \|Severe acute respiratory syndrome coro | .................... | .................... | .................... | .................... | .................... | .................... | |
| 16685 | MT969502.1 | \|Severe acute respiratory syndrome coro | .................... | .................... | .................... | .................... | .................... | .................... | |
| 16686 | MT969503.1 | \|Severe acute respiratory syndrome coro | .................... | .................... | .................... | .................... | ..N................. | .................... | |
| 16687 | MT969504.1 | \|Severe acute respiratory syndrome coro | .................... | .................... | .................... | .................... | .................... | .................... | |
| 16688 | MT969505.1 | \|Severe acute respiratory syndrome coro | .................... | .................... | .................... | .................... | .................... | .................... | |
| 16689 | MT969506.1 | \|Severe acute respiratory syndrome coro | .................... | .................... | .................... | .................... | .................... | .................... | |
| 16690 | MT969507.1 | \|Severe acute respiratory syndrome coro | .................... | .................... | .................... | .................... | .................... | .................... | |
| 16691 | MT969508.1 | \|Severe acute respiratory syndrome coro | .................... | .................... | .................... | .................... | .................... | .................... | |
| 16692 | MT969509.1 | \|Severe acute respiratory syndrome coro | .................... | .................... | .................... | .................... | .................... | .................... | |
| 16693 | MT969510.1 | \|Severe acute respiratory syndrome coro | .................... | .................... | .................... | .................... | .................... | .................... | |
| 16694 | MT969511.1 | \|Severe acute respiratory syndrome coro | .................... | .................... | .................... | .................... | .................... | .................... | |
| 16695 | MT969512.1 | \|Severe acute respiratory syndrome coro | .................... | .................... | .................... | .................... | .................... | .................... | |
| 16697 | MT969514.1 | \|Severe acute respiratory syndrome coro | .................... | .................... | .................... | .................... | .................... | .................... | |
| 16698 | MT969515.1 | \|Severe acute respiratory syndrome coro | .................... | .................... | .................... | .................... | .................... | .................... | |
| 16699 | MT969516.1 | \|Severe acute respiratory syndrome coro | .................... | .................... | .................... | .................... | .................... | .................... | |
| 16700 | MT969517.1 | \|Severe acute respiratory syndrome coro | .................... | .................... | .................... | .................... | .................... | .................... | |
| 16702 | MT969519.1 | \|Severe acute respiratory syndrome coro | .................... | .................... | .................... | .................... | .................... | .................... | |
| 16703 | MT969520.1 | \|Severe acute respiratory syndrome coro | .................... | .................... | .................... | .................... | .................... | .................... | |
| 16704 | MT969521.1 | \|Severe acute respiratory syndrome coro | .................... | .................... | .................... | .................... | .................... | .................... | |
| 16705 | MT969522.1 | \|Severe acute respiratory syndrome coro | .................... | .................... | .................... | .................... | .................... | .................... | |
| 16706 | MT969523.1 | \|Severe acute respiratory syndrome coro | .................... | .................... | .................... | .................... | .................... | .................... | |
| 16707 | MT969524.1 | \|Severe acute respiratory syndrome coro | .................... | .................... | .................... | .................... | .................... | .................... | |
| 16708 | MT969525.1 | \|Severe acute respiratory syndrome coro | .................... | .................... | .................... | .................... | .................... | .................... | |
| 16709 | MT969526.1 | \|Severe acute respiratory syndrome coro | .................... | .................... | .................... | .................... | .................... | .................... | |
| 16710 | MT969528.1 | \|Severe acute respiratory syndrome coro | .................... | .................... | .................... | .................... | .................... | .................... | |
| 16711 | MT969530.1 | \|Severe acute respiratory syndrome coro | .................... | .................... | .................... | .................... | .................... | .................... | |
| 16712 | MT969531.1 | \|Severe acute respiratory syndrome coro | .................... | .................... | .................... | .................... | .................... | .................... | |

| | | | | | | | | | |
|---|---|---|---|---|---|---|---|---|---|
| 16713 | MT969532.1 | \|Severe acute respiratory syndrome coro | .................... | .................... | .................... | .................... | .................... | .................... | |
| 16714 | MT969533.1 | \|Severe acute respiratory syndrome coro | .................... | .................... | .................... | .................... | .................... | .................... | |
| 16716 | MT969536.1 | \|Severe acute respiratory syndrome coro | .................... | .................... | .................... | .................... | .................... | .................... | |
| 16717 | MT969537.1 | \|Severe acute respiratory syndrome coro | .................... | .................... | .................... | .................... | .................... | .................... | |
| 16718 | MT969538.1 | \|Severe acute respiratory syndrome coro | .................... | .................... | .................... | .................... | NNN................. | .................... | 1 |
| 16719 | MT969539.1 | \|Severe acute respiratory syndrome coro | .................... | .................... | .................... | .................... | .................... | .................... | |
| 16720 | MT969540.1 | \|Severe acute respiratory syndrome coro | .................... | .................... | .................... | .................... | .................... | .................... | |
| 16721 | MT969541.1 | \|Severe acute respiratory syndrome coro | .................... | .................... | .................... | .................... | .................... | .................... | |
| 16722 | MT969542.1 | \|Severe acute respiratory syndrome coro | .................... | .................... | .................... | .................... | .................... | .................... | |
| 16724 | MT969544.1 | \|Severe acute respiratory syndrome coro | .................... | .................... | .................... | .................... | .................... | .................... | |
| 16725 | MT969545.1 | \|Severe acute respiratory syndrome coro | .................... | .................... | .................... | .................... | .................... | .................... | |
| 16726 | MT969546.1 | \|Severe acute respiratory syndrome coro | .................... | .................... | .................... | .................... | .................... | .................... | |
| 16727 | MT969547.1 | \|Severe acute respiratory syndrome coro | .................... | .................... | .................... | .................... | .................... | .................... | |
| 16728 | MT969548.1 | \|Severe acute respiratory syndrome coro | .................... | .................... | .................... | .................... | .................... | .................... | |
| 16730 | MT969550.1 | \|Severe acute respiratory syndrome coro | .................... | .................... | .................... | .................... | .................... | .................... | |
| 16731 | MT969551.1 | \|Severe acute respiratory syndrome coro | .................... | .................... | .................... | .................... | .................... | .................... | |
| 16732 | MT969552.1 | \|Severe acute respiratory syndrome coro | .................... | .................... | .................... | .................... | .................... | .................... | |
| 16733 | MT969553.1 | \|Severe acute respiratory syndrome coro | .................... | .................... | .................... | .................... | .................... | .................... | |
| 16734 | MT969554.1 | \|Severe acute respiratory syndrome coro | .................... | .................... | .................... | .................... | .................... | .................... | |
| 16735 | MT969555.1 | \|Severe acute respiratory syndrome coro | .................... | .................... | .................... | .................... | .................... | .................... | |
| 16736 | MT969556.1 | \|Severe acute respiratory syndrome coro | .................... | .................... | .................... | .................... | .................... | .................... | |
| 16737 | MT969557.1 | \|Severe acute respiratory syndrome coro | .................... | .................... | .................... | .................... | .................... | .................... | |
| 16738 | MT969558.1 | \|Severe acute respiratory syndrome coro | .................... | .................... | .................... | .................... | .................... | .................... | |
| 16739 | MT969559.1 | \|Severe acute respiratory syndrome coro | .................... | .................... | .................... | .................... | .................... | .................... | |
| 16740 | MT969560.1 | \|Severe acute respiratory syndrome coro | .................... | .................... | .................... | .................... | .................... | .................... | |
| 16741 | MT969561.1 | \|Severe acute respiratory syndrome coro | .................... | .................... | .................... | .................... | .................... | .................... | |
| 16742 | MT969562.1 | \|Severe acute respiratory syndrome coro | .................... | .................... | .................... | .................... | .................... | .................... | |
| 16743 | MT969563.1 | \|Severe acute respiratory syndrome coro | .................... | .................... | .................... | .................... | .................... | .................... | |
| 16744 | MT969564.1 | \|Severe acute respiratory syndrome coro | .................... | .................... | .................... | .................... | .................... | .................... | |
| 16745 | MT969565.1 | \|Severe acute respiratory syndrome coro | .................... | .................... | .................... | .................... | .................... | .................... | |
| 16746 | MT969566.1 | \|Severe acute respiratory syndrome coro | .................... | .................... | .................... | .................... | .................... | .................... | |
| 16747 | MT969567.1 | \|Severe acute respiratory syndrome coro | .................... | .................... | .................... | .................... | .................... | .................... | |
| 16748 | MT969568.1 | \|Severe acute respiratory syndrome coro | .................... | .................... | .................... | .................... | .................... | .................... | |
| 16749 | MT969569.1 | \|Severe acute respiratory syndrome coro | .................... | .................... | .................... | .................... | .................... | .................... | |
| 16750 | MT969570.1 | \|Severe acute respiratory syndrome coro | .................... | .................... | .................... | .................... | .................... | .................... | |
| 16751 | MT969571.1 | \|Severe acute respiratory syndrome coro | .................... | .................... | .................... | .................... | .................... | .................... | |
| 16752 | MT969572.1 | \|Severe acute respiratory syndrome coro | .................... | .................... | .................... | .................... | .................... | .................... | |
| 16753 | MT969573.1 | \|Severe acute respiratory syndrome coro | .................... | .................... | .................... | .................... | .................... | .................... | |
| 16754 | MT969574.1 | \|Severe acute respiratory syndrome coro | .................... | .................... | .................... | .................... | .................... | .................... | |
| 16755 | MT969575.1 | \|Severe acute respiratory syndrome coro | .................... | .................... | .................... | .................... | .................... | .................... | |
| 16756 | MT969576.1 | \|Severe acute respiratory syndrome coro | .................... | .................... | .................... | .................... | .................... | .................... | |
| 16757 | MT969577.1 | \|Severe acute respiratory syndrome coro | .................... | .................... | .................... | .................... | .................... | .................... | |
| 16758 | MT969578.1 | \|Severe acute respiratory syndrome coro | .................... | .................... | .................... | .................... | .................... | .................... | |
| 16759 | MT969579.1 | \|Severe acute respiratory syndrome coro | .................... | .................... | .................... | .................... | .................... | .................... | |
| 16760 | MT969580.1 | \|Severe acute respiratory syndrome coro | .................... | .................... | .................... | .................... | .................... | .................... | |
| 16761 | MT969581.1 | \|Severe acute respiratory syndrome coro | .................... | .................... | .................... | .................... | .................... | .................... | |
| 16762 | MT969582.1 | \|Severe acute respiratory syndrome coro | .................... | .................... | .................... | .................... | .................... | .................... | |
| 16763 | MT969583.1 | \|Severe acute respiratory syndrome coro | .................... | .................... | .................... | .................... | .................... | .................... | |
| 16764 | MT969584.1 | \|Severe acute respiratory syndrome coro | .................... | .................... | .................... | .................... | .................... | .................... | |
| 16765 | MT969585.1 | \|Severe acute respiratory syndrome coro | .................... | .................... | .................... | .................... | .................... | .................... | |
| 16766 | MT969586.1 | \|Severe acute respiratory syndrome coro | .................... | .................... | .................... | .................... | .................... | .................... | |
| 16767 | MT969587.1 | \|Severe acute respiratory syndrome coro | .................... | .................... | .................... | .................... | .................... | .................... | |
| 16768 | MT969588.1 | \|Severe acute respiratory syndrome coro | .................... | .................... | .................... | .................... | .................... | .................... | |
| 16769 | MT969589.1 | \|Severe acute respiratory syndrome coro | .................... | .................... | .................... | .................... | .................... | .................... | |
| 16770 | MT969590.1 | \|Severe acute respiratory syndrome coro | .................... | .................... | .................... | .................... | .................... | .................... | |
| 16771 | MT969591.1 | \|Severe acute respiratory syndrome coro | .................... | .................... | .................... | .................... | .................... | .................... | |
| 16772 | MT969592.1 | \|Severe acute respiratory syndrome coro | .................... | .................... | .................... | .................... | .................... | .................... | |
| 16773 | MT969593.1 | \|Severe acute respiratory syndrome coro | .................... | .................... | .................... | .................... | .................... | .................... | |
| 16774 | MT969594.1 | \|Severe acute respiratory syndrome coro | .................... | .................... | .................... | .................... | .................... | .................... | |
| 16775 | MT969595.1 | \|Severe acute respiratory syndrome coro | .................... | .................... | .................... | .................... | .................... | .................... | |
| 16776 | MT969596.1 | \|Severe acute respiratory syndrome coro | .................... | .................... | .................... | .................... | .................... | .................... | |
| 16777 | MT969597.1 | \|Severe acute respiratory syndrome coro | .................... | .................... | .................... | .................... | .................... | .................... | |
| 16778 | MT969598.1 | \|Severe acute respiratory syndrome coro | .................... | .................... | .................... | .................... | .................... | .................... | |
| 16779 | MT969599.1 | \|Severe acute respiratory syndrome coro | .................... | .................... | .................... | .................... | .................... | .................... | |
| 16780 | MT969600.1 | \|Severe acute respiratory syndrome coro | .................... | .................... | .................... | .................... | .................... | .................... | |
| 16781 | MT969601.1 | \|Severe acute respiratory syndrome coro | .................... | .................... | .................... | .................... | .................... | .................... | |
| 16782 | MT969602.1 | \|Severe acute respiratory syndrome coro | .................... | .................... | .................... | .................... | .................... | .................... | |
| 16783 | MT969603.1 | \|Severe acute respiratory syndrome coro | .................... | .................... | .................... | .................... | .................... | .................... | |
| 16784 | MT969604.1 | \|Severe acute respiratory syndrome coro | .................... | .................... | .................... | .................... | .................... | .................... | |
| 16785 | MT969605.1 | \|Severe acute respiratory syndrome coro | .................... | .................... | .................... | .................... | .................... | .................... | |
| 16786 | MT969606.1 | \|Severe acute respiratory syndrome coro | .................... | .................... | .................... | .................... | .................... | .................... | |
| 16787 | MT969607.1 | \|Severe acute respiratory syndrome coro | .................... | .................... | .................... | .................... | .................... | .................... | |
| 16788 | MT969608.1 | \|Severe acute respiratory syndrome coro | .................... | .................... | .................... | .................... | .................... | .................... | |
| 16789 | MT969609.1 | \|Severe acute respiratory syndrome coro | .................... | .................... | .................... | .................... | .................... | .................... | |
| 16790 | MT969610.1 | \|Severe acute respiratory syndrome coro | .................... | .................... | .................... | .................... | .................... | .................... | |
| 16791 | MT969611.1 | \|Severe acute respiratory syndrome coro | .................... | .................... | .................... | .................... | .................... | .................... | |
| 16792 | MT969612.1 | \|Severe acute respiratory syndrome coro | .................... | .................... | .................... | .................... | .................... | .................... | |
| 16793 | MT969613.1 | \|Severe acute respiratory syndrome coro | .................... | .................... | .................... | .................... | .NNN................ | .................... | 1 |

| | | | | | | | | | |
|---|---|---|---|---|---|---|---|---|---|
| 16794 | MT969614.1 | \|Severe acute respiratory syndrome coro | ···················· | ···················· | ···················· | ···················· | ···················· | ···················· | |
| 16795 | MT969615.1 | \|Severe acute respiratory syndrome coro | ···················· | ···················· | ···················· | ···················· | ···················· | ···················· | |
| 16796 | MT969616.1 | \|Severe acute respiratory syndrome coro | ···················· | ···················· | ···················· | ···················· | ···················· | ···················· | |
| 16797 | MT969617.1 | \|Severe acute respiratory syndrome coro | ···················· | ···················· | ···················· | ···················· | ···················· | ···················· | |
| 16798 | MT969618.1 | \|Severe acute respiratory syndrome coro | ···················· | ···················· | ···················· | ···················· | ···················· | ···················· | |
| 16799 | MT969619.1 | \|Severe acute respiratory syndrome coro | ···················· | ···················· | ···················· | ···················· | ···················· | ···················· | |
| 16800 | MT969620.1 | \|Severe acute respiratory syndrome coro | ···················· | ···················· | ···················· | ···················· | ···················· | ···················· | |
| 16801 | MT969621.1 | \|Severe acute respiratory syndrome coro | ···················· | ···················· | ···················· | ···················· | ···················· | ···················· | |
| 16802 | MT969622.1 | \|Severe acute respiratory syndrome coro | ···················· | ···················· | ···················· | ···················· | ···················· | ···················· | |
| 16803 | MT969623.1 | \|Severe acute respiratory syndrome coro | ···················· | ···················· | ···················· | ···················· | ···················· | ···················· | |
| 16805 | MT969625.1 | \|Severe acute respiratory syndrome coro | ···················· | ···················· | ···················· | ···················· | ···················· | ···················· | |
| 16806 | MT969626.1 | \|Severe acute respiratory syndrome coro | ···················· | ···················· | ···················· | ···················· | ···················· | ···················· | |
| 16807 | MT969627.1 | \|Severe acute respiratory syndrome coro | ···················· | ···················· | ···················· | ···················· | ···················· | ···················· | |
| 16808 | MT969629.1 | \|Severe acute respiratory syndrome coro | ···················· | ···················· | ···················· | ···················· | ···················· | ···················· | |
| 16811 | MT969632.1 | \|Severe acute respiratory syndrome coro | ···················· | ···················· | ···················· | ···················· | ···················· | ···················· | |
| 16812 | MT969635.1 | \|Severe acute respiratory syndrome coro | ···················· | ···················· | ···················· | ···················· | ···················· | ···················· | |
| 16813 | MT969636.1 | \|Severe acute respiratory syndrome coro | ···················· | ···················· | ···················· | ···················· | ···················· | ···················· | |
| 16815 | MT969638.1 | \|Severe acute respiratory syndrome coro | ···················· | ···················· | ···················· | ···················· | ···················· | ···················· | |
| 16816 | MT969640.1 | \|Severe acute respiratory syndrome coro | ···················· | ···················· | ···················· | ···················· | ···················· | ···················· | |
| 16817 | MT969641.1 | \|Severe acute respiratory syndrome coro | ···················· | ···················· | ···················· | ···················· | ···················· | ···················· | |
| 16818 | MT969642.1 | \|Severe acute respiratory syndrome coro | ···················· | ···················· | ···················· | ···················· | ···················· | ···················· | |
| 16819 | MT969643.1 | \|Severe acute respiratory syndrome coro | ···················· | ···················· | ···················· | ···················· | ···················· | ···················· | |
| 16820 | MT969644.1 | \|Severe acute respiratory syndrome coro | ···················· | ···················· | ···················· | ···················· | ···················· | ···················· | |
| 16822 | MT969646.1 | \|Severe acute respiratory syndrome coro | ···················· | ···················· | ···················· | ···················· | NNN················· | ···················· | 1 |
| 16823 | MT969647.1 | \|Severe acute respiratory syndrome coro | ···················· | ···················· | ···················· | ···················· | ···················· | ···················· | |
| 16824 | MT969648.1 | \|Severe acute respiratory syndrome coro | ···················· | ···················· | ···················· | ···················· | ···················· | ···················· | |
| 16825 | MT969649.1 | \|Severe acute respiratory syndrome coro | ···················· | ···················· | ···················· | ···················· | ···················· | ···················· | |
| 16826 | MT969650.1 | \|Severe acute respiratory syndrome coro | ···················· | ···················· | ···················· | ···················· | ···················· | ···················· | |
| 16827 | MT969651.1 | \|Severe acute respiratory syndrome coro | ···················· | ···················· | ···················· | ···················· | ···················· | ···················· | |
| 16828 | MT969652.1 | \|Severe acute respiratory syndrome coro | ···················· | ···················· | ···················· | ···················· | ···················· | ···················· | |
| 16829 | MT969653.1 | \|Severe acute respiratory syndrome coro | ···················· | ···················· | ···················· | ···················· | ···················· | ···················· | |
| 16830 | MT969654.1 | \|Severe acute respiratory syndrome coro | ···················· | ···················· | ···················· | ···················· | ···················· | ···················· | |
| 16831 | MT969655.1 | \|Severe acute respiratory syndrome coro | ···················· | ···················· | ···················· | ···················· | ···················· | ···················· | |
| 16832 | MT969656.1 | \|Severe acute respiratory syndrome coro | ···················· | ···················· | ···················· | ···················· | ···················· | ···················· | |
| 16833 | MT969657.1 | \|Severe acute respiratory syndrome coro | ···················· | ···················· | ···················· | ···················· | ···················· | ···················· | |
| 16834 | MT969658.1 | \|Severe acute respiratory syndrome coro | ···················· | ···················· | ···················· | ···················· | ···················· | ···················· | |
| 16835 | MT969659.1 | \|Severe acute respiratory syndrome coro | ···················· | ···················· | ···················· | ···················· | ···················· | ···················· | |
| 16836 | MT969660.1 | \|Severe acute respiratory syndrome coro | ···················· | ···················· | ···················· | ···················· | ···················· | ···················· | |
| 16837 | MT969661.1 | \|Severe acute respiratory syndrome coro | ···················· | ···················· | ···················· | ···················· | ···················· | ···················· | |
| 16838 | MT969662.1 | \|Severe acute respiratory syndrome coro | ···················· | ···················· | ···················· | ···················· | ···················· | ···················· | |
| 16839 | MT969663.1 | \|Severe acute respiratory syndrome coro | ···················· | ···················· | ···················· | ···················· | ···················· | ···················· | |
| 16840 | MT969664.1 | \|Severe acute respiratory syndrome coro | ···················· | ···················· | ···················· | ···················· | ···················· | ···················· | |
| 16841 | MT969665.1 | \|Severe acute respiratory syndrome coro | ···················· | ···················· | ···················· | ···················· | ···················· | ···················· | |
| 16842 | MT969666.1 | \|Severe acute respiratory syndrome coro | ···················· | ···················· | ···················· | ···················· | ···················· | ···················· | |
| 16843 | MT969667.1 | \|Severe acute respiratory syndrome coro | ···················· | ···················· | ···················· | ···················· | ···················· | ···················· | |
| 16844 | MT969668.1 | \|Severe acute respiratory syndrome coro | ···················· | ···················· | ···················· | ···················· | ···················· | ···················· | |
| 16845 | MT969669.1 | \|Severe acute respiratory syndrome coro | ···················· | ···················· | ···················· | ···················· | ···················· | ···················· | |
| 16847 | MT969671.1 | \|Severe acute respiratory syndrome coro | ···················· | ···················· | ···················· | ···················· | NNN················· | ···················· | 1 |
| 16848 | MT969672.1 | \|Severe acute respiratory syndrome coro | ···················· | ···················· | ···················· | ···················· | ···················· | ···················· | |
| 16849 | MT969673.1 | \|Severe acute respiratory syndrome coro | ···················· | ···················· | ···················· | ···················· | ···················· | ···················· | |
| 16850 | MT969674.1 | \|Severe acute respiratory syndrome coro | ···················· | ···················· | ···················· | ···················· | ···················· | ···················· | |
| 16851 | MT969675.1 | \|Severe acute respiratory syndrome coro | ···················· | ···················· | ···················· | ···················· | ···················· | ···················· | |
| 16852 | MT969676.1 | \|Severe acute respiratory syndrome coro | ···················· | ···················· | ···················· | ···················· | ···················· | ···················· | |
| 16853 | MT969677.1 | \|Severe acute respiratory syndrome coro | ···················· | ···················· | ···················· | ···················· | ···················· | ···················· | |
| 16855 | MT969679.1 | \|Severe acute respiratory syndrome coro | ···················· | ···················· | ···················· | ···················· | ···················· | ···················· | |
| 16857 | MT969681.1 | \|Severe acute respiratory syndrome coro | ···················· | ···················· | ···················· | ···················· | ···················· | ···················· | |
| 16859 | MT969683.1 | \|Severe acute respiratory syndrome coro | ···················· | ···················· | ···················· | ···················· | ···················· | ···················· | |
| 16860 | MT969685.1 | \|Severe acute respiratory syndrome coro | ···················· | ···················· | ···················· | ···················· | ···················· | ···················· | |
| 16861 | MT969686.1 | \|Severe acute respiratory syndrome coro | ···················· | ···················· | ···················· | ···················· | ···················· | ···················· | |
| 16862 | MT969687.1 | \|Severe acute respiratory syndrome coro | ···················· | ···················· | ···················· | ···················· | ···················· | ···················· | |
| 16863 | MT969688.1 | \|Severe acute respiratory syndrome coro | ···················· | ···················· | ···················· | ···················· | ···················· | ···················· | |
| 16864 | MT969689.1 | \|Severe acute respiratory syndrome coro | ···················· | ···················· | ···················· | ···················· | ···················· | ···················· | |
| 16865 | MT969690.1 | \|Severe acute respiratory syndrome coro | ···················· | ···················· | ···················· | ···················· | ···················· | ···················· | |
| 16866 | MT969691.1 | \|Severe acute respiratory syndrome coro | ···················· | ···················· | ···················· | ···················· | ···················· | ···················· | |
| 16868 | MT969693.1 | \|Severe acute respiratory syndrome coro | ···················· | ···················· | ···················· | ···················· | ···················· | ···················· | |
| 16869 | MT969694.1 | \|Severe acute respiratory syndrome coro | ···················· | ···················· | ···················· | ···················· | ···················· | ···················· | |
| 16870 | MT969695.1 | \|Severe acute respiratory syndrome coro | ···················· | ···················· | ···················· | ···················· | ···················· | ···················· | |
| 16871 | MT969696.1 | \|Severe acute respiratory syndrome coro | ···················· | ···················· | ···················· | ···················· | ···················· | ···················· | |
| 16872 | MT969697.1 | \|Severe acute respiratory syndrome coro | ···················· | ···················· | ···················· | ···················· | ···················· | ···················· | |
| 16873 | MT969698.1 | \|Severe acute respiratory syndrome coro | ···················· | ···················· | ···················· | ···················· | ···················· | ···················· | |
| 16874 | MT969699.1 | \|Severe acute respiratory syndrome coro | ···················· | ···················· | ···················· | ···················· | ···················· | ···················· | |
| 16875 | MT969700.1 | \|Severe acute respiratory syndrome coro | ···················· | ···················· | ···················· | ···················· | ···················· | ···················· | |
| 16876 | MT969701.1 | \|Severe acute respiratory syndrome coro | ···················· | ···················· | ···················· | ···················· | ···················· | ···················· | |
| 16877 | MT969702.1 | \|Severe acute respiratory syndrome coro | ···················· | ···················· | ···················· | ···················· | ···················· | ···················· | |
| 16878 | MT969703.1 | \|Severe acute respiratory syndrome coro | ···················· | ···················· | ···················· | ···················· | ···················· | ···················· | |
| 16879 | MT969704.1 | \|Severe acute respiratory syndrome coro | ···················· | ···················· | ···················· | ···················· | ···················· | ···················· | |
| 16880 | MT969705.1 | \|Severe acute respiratory syndrome coro | ···················· | ···················· | ···················· | ···················· | ···················· | ···················· | |
| 16881 | MT969706.1 | \|Severe acute respiratory syndrome coro | ···················· | ···················· | ···················· | ···················· | ···················· | ···················· | |

| | | | | | | | | |
|---|---|---|---|---|---|---|---|---|
| 16883 | MT969708.1 | \|Severe acute respiratory syndrome coro | .................... | .................... | ..................... | .................... | .................... | .................... |
| 16884 | MT969709.1 | \|Severe acute respiratory syndrome coro | .................... | .................... | ..................... | .................... | .................... | .................... |
| 16885 | MT969710.1 | \|Severe acute respiratory syndrome coro | .................... | .................... | ..................... | .................... | .................... | .................... |
| 16886 | MT969711.1 | \|Severe acute respiratory syndrome coro | .................... | .................... | ..................... | .................... | .................... | .................... |
| 16887 | MT969712.1 | \|Severe acute respiratory syndrome coro | .................... | .................... | ..................... | .................... | .................... | .................... |
| 16888 | MT969713.1 | \|Severe acute respiratory syndrome coro | .................... | .................... | ..................... | .................... | .................... | .................... |
| 16889 | MT969714.1 | \|Severe acute respiratory syndrome coro | .................... | .................... | ..................... | .................... | .................... | .................... |
| 16890 | MT969715.1 | \|Severe acute respiratory syndrome coro | .................... | .................... | ..................... | .................... | .................... | .................... |
| 16891 | MT969716.1 | \|Severe acute respiratory syndrome coro | .................... | .................... | ..................... | .................... | .................... | .................... |
| 16892 | MT969717.1 | \|Severe acute respiratory syndrome coro | .................... | .................... | ..................... | .................... | .................... | .................... |
| 16893 | MT969718.1 | \|Severe acute respiratory syndrome coro | .................... | .................... | ..................... | .................... | .................... | .................... |
| 16894 | MT969719.1 | \|Severe acute respiratory syndrome coro | .................... | .................... | ..................... | .................... | .................... | .................... |
| 16895 | MT969720.1 | \|Severe acute respiratory syndrome coro | .................... | .................... | ..................... | .................... | .................... | .................... |
| 16896 | MT969721.1 | \|Severe acute respiratory syndrome coro | .................... | .................... | ..................... | .................... | .................... | .................... |
| 16897 | MT969723.1 | \|Severe acute respiratory syndrome coro | .................... | .................... | ..................... | .................... | .................... | .................... |
| 16899 | MT969725.1 | \|Severe acute respiratory syndrome coro | .................... | .................... | ..................... | .................... | .................... | .................... |
| 16900 | MT969726.1 | \|Severe acute respiratory syndrome coro | .................... | .................... | ..................... | .................... | .................... | .................... |
| 16901 | MT969727.1 | \|Severe acute respiratory syndrome coro | .................... | .................... | ..................... | .................... | .................... | .................... |
| 16902 | MT969728.1 | \|Severe acute respiratory syndrome coro | .................... | .................... | ..................... | .................... | .................... | .................... |
| 16903 | MT969729.1 | \|Severe acute respiratory syndrome coro | .................... | .................... | ..................... | .................... | .................... | .................... |
| 16904 | MT969730.1 | \|Severe acute respiratory syndrome coro | .................... | .................... | ..................... | .................... | .................... | .................... |
| 16905 | MT969731.1 | \|Severe acute respiratory syndrome coro | .................... | .................... | ..................... | .................... | .................... | .................... |
| 16906 | MT969732.1 | \|Severe acute respiratory syndrome coro | .................... | .................... | ..................... | .................... | .................... | .................... |
| 16907 | MT969733.1 | \|Severe acute respiratory syndrome coro | .................... | .................... | ..................... | .................... | .................... | .................... |
| 16908 | MT969734.1 | \|Severe acute respiratory syndrome coro | .................... | .................... | ..................... | .................... | .................... | .................... |
| 16909 | MT969736.1 | \|Severe acute respiratory syndrome coro | .................... | .................... | ..................... | .................... | .................... | .................... |
| 16910 | MT969737.1 | \|Severe acute respiratory syndrome coro | .................... | .................... | ..................... | .................... | .................... | .................... |
| 16911 | MT969738.1 | \|Severe acute respiratory syndrome coro | .................... | .................... | ..................... | .................... | .................... | .................... |
| 16912 | MT969739.1 | \|Severe acute respiratory syndrome coro | .................... | .................... | ..................... | .................... | .................... | .................... |
| 16913 | MT969740.1 | \|Severe acute respiratory syndrome coro | .................... | .................... | ..................... | .................... | .................... | .................... |
| 16914 | MT969741.1 | \|Severe acute respiratory syndrome coro | .................... | .................... | ..................... | .................... | .................... | .................... |
| 16916 | MT969743.1 | \|Severe acute respiratory syndrome coro | .................... | .................... | ..................... | .................... | .................... | .................... |
| 16917 | MT969744.1 | \|Severe acute respiratory syndrome coro | .................... | .................... | ..................... | .................... | .................... | .................... |
| 16918 | MT969745.1 | \|Severe acute respiratory syndrome coro | .................... | .................... | ..................... | .................... | .................... | .................... |
| 16919 | MT969746.1 | \|Severe acute respiratory syndrome coro | .................... | .................... | ..................... | .................... | .................... | .................... |
| 16920 | MT969747.1 | \|Severe acute respiratory syndrome coro | .................... | .................... | ..................... | .................... | .................... | .................... |
| 16921 | MT969748.1 | \|Severe acute respiratory syndrome coro | .................... | .................... | ..................... | .................... | .................... | .................... |
| 16922 | MT969749.1 | \|Severe acute respiratory syndrome coro | .................... | .................... | ..................... | .................... | .................... | .................... |
| 16923 | MT969750.1 | \|Severe acute respiratory syndrome coro | .................... | .................... | ..................... | .................... | .................... | .................... |
| 16924 | MT969751.1 | \|Severe acute respiratory syndrome coro | .................... | .................... | ..................... | .................... | .................... | .................... |
| 16925 | MT969752.1 | \|Severe acute respiratory syndrome coro | .................... | .................... | ..................... | .................... | .................... | .................... |
| 16926 | MT969753.1 | \|Severe acute respiratory syndrome coro | .................... | .................... | ..................... | .................... | .................... | .................... |
| 16927 | MT969754.1 | \|Severe acute respiratory syndrome coro | .................... | .................... | ..................... | .................... | .................... | .................... |
| 16929 | MT969756.1 | \|Severe acute respiratory syndrome coro | .................... | .................... | ..................... | .................... | .................... | .................... |
| 16930 | MT969757.1 | \|Severe acute respiratory syndrome coro | .................... | .................... | ..................... | .................... | .................... | .................... |
| 16931 | MT969758.1 | \|Severe acute respiratory syndrome coro | .................... | .................... | ..................... | .................... | .................... | .................... |
| 16932 | MT969759.1 | \|Severe acute respiratory syndrome coro | .................... | .................... | ..................... | .................... | .................... | .................... |
| 16933 | MT969760.1 | \|Severe acute respiratory syndrome coro | .................... | .................... | ..................... | .................... | .................... | .................... |
| 16934 | MT969761.1 | \|Severe acute respiratory syndrome coro | .................... | .................... | ..................... | .................... | .................... | .................... |
| 16935 | MT969762.1 | \|Severe acute respiratory syndrome coro | .................... | .................... | ..................... | .................... | .................... | .................... |
| 16936 | MT969763.1 | \|Severe acute respiratory syndrome coro | .................... | .................... | ..................... | .................... | .................... | .................... |
| 16937 | MT969764.1 | \|Severe acute respiratory syndrome coro | .................... | .................... | ..................... | .................... | .................... | .................... |
| 16938 | MT969765.1 | \|Severe acute respiratory syndrome coro | .................... | .................... | ..................... | .................... | .................... | .................... |
| 16939 | MT969766.1 | \|Severe acute respiratory syndrome coro | .................... | .................... | ..................... | .................... | .................... | .................... |
| 16940 | MT969767.1 | \|Severe acute respiratory syndrome coro | .................... | .................... | ..................... | .................... | .................... | .................... |
| 16941 | MT969768.1 | \|Severe acute respiratory syndrome coro | .................... | .................... | ..................... | .................... | .................... | .................... |
| 16942 | MT969769.1 | \|Severe acute respiratory syndrome coro | .................... | .................... | ..................... | .................... | .................... | .................... |
| 16943 | MT969770.1 | \|Severe acute respiratory syndrome coro | .................... | .................... | ..................... | .................... | .................... | .................... |
| 16944 | MT969771.1 | \|Severe acute respiratory syndrome coro | .................... | .................... | ..................... | .................... | .................... | .................... |
| 16945 | MT969772.1 | \|Severe acute respiratory syndrome coro | .................... | .................... | ..................... | .................... | .................... | .................... |
| 16947 | MT969774.1 | \|Severe acute respiratory syndrome coro | .................... | .................... | ..................... | .................... | .................... | .................... |
| 16948 | MT969775.1 | \|Severe acute respiratory syndrome coro | .................... | .................... | ..................... | .................... | .................... | .................... |
| 16949 | MT969776.1 | \|Severe acute respiratory syndrome coro | .................... | .................... | ..................... | .................... | .................... | .................... |
| 16950 | MT969777.1 | \|Severe acute respiratory syndrome coro | .................... | .................... | ..................... | .................... | .................... | .................... |
| 16951 | MT969778.1 | \|Severe acute respiratory syndrome coro | .................... | .................... | ..................... | .................... | .................... | .................... |
| 16952 | MT969779.1 | \|Severe acute respiratory syndrome coro | .................... | .................... | ..................... | .................... | .................... | .................... |
| 16953 | MT969780.1 | \|Severe acute respiratory syndrome coro | .................... | .................... | ..................... | .................... | .................... | .................... |
| 16954 | MT969781.1 | \|Severe acute respiratory syndrome coro | .................... | .................... | ..................... | .................... | .................... | .................... |
| 16955 | MT969782.1 | \|Severe acute respiratory syndrome coro | .................... | .................... | ..................... | .................... | .................... | .................... |
| 16957 | MT969784.1 | \|Severe acute respiratory syndrome coro | .................... | .................... | ..................... | .................... | .................... | .................... |
| 16958 | MT969785.1 | \|Severe acute respiratory syndrome coro | .................... | .................... | ..................... | .................... | .................... | .................... |
| 16959 | MT969786.1 | \|Severe acute respiratory syndrome coro | .................... | .................... | ..................... | .................... | .................... | .................... |
| 16960 | MT969787.1 | \|Severe acute respiratory syndrome coro | .................... | .................... | ..................... | .................... | .................... | .................... |
| 16961 | MT969788.1 | \|Severe acute respiratory syndrome coro | .................... | .................... | ..................... | .................... | .................... | .................... |
| 16962 | MT969789.1 | \|Severe acute respiratory syndrome coro | .................... | .................... | ..................... | .................... | .................... | .................... |
| 16963 | MT969790.1 | \|Severe acute respiratory syndrome coro | .................... | .................... | ..................... | .................... | .................... | .................... |
| 16964 | MT969791.1 | \|Severe acute respiratory syndrome coro | .................... | .................... | ..................... | .................... | .................... | .................... |
| 16965 | MT969792.1 | \|Severe acute respiratory syndrome coro | .................... | .................... | ..................... | .................... | .................... | .................... |

| | | | | | | | | | |
|---|---|---|---|---|---|---|---|---|---|
| 16966 | MT969793.1 | \|Severe acute respiratory syndrome coro | ···················· | ···················· | ···················· | ···················· | ···················· | ···················· | |
| 16967 | MT969794.1 | \|Severe acute respiratory syndrome coro | ···················· | ···················· | ···················· | ···················· | ···················· | ···················· | |
| 16968 | MT969795.1 | \|Severe acute respiratory syndrome coro | ···················· | ···················· | ···················· | ···················· | ···················· | ···················· | |
| 16969 | MT969796.1 | \|Severe acute respiratory syndrome coro | ···················· | ···················· | ···················· | ···················· | ···················· | ···················· | |
| 16970 | MT969797.1 | \|Severe acute respiratory syndrome coro | ···················· | ···················· | ···················· | ···················· | ···················· | ···················· | |
| 16971 | MT969799.1 | \|Severe acute respiratory syndrome coro | ···················· | ···················· | ···················· | ···················· | ···················· | ···················· | |
| 16972 | MT969800.1 | \|Severe acute respiratory syndrome coro | ···················· | ···················· | ···················· | ···················· | ···················· | ···················· | |
| 16973 | MT969801.1 | \|Severe acute respiratory syndrome coro | ···················· | ···················· | ···················· | ···················· | ···················· | ···················· | |
| 16974 | MT969802.1 | \|Severe acute respiratory syndrome coro | ···················· | ···················· | ···················· | ···················· | ···················· | ···················· | |
| 16975 | MT969803.1 | \|Severe acute respiratory syndrome coro | ···················· | ···················· | ···················· | ···················· | ···················· | ···················· | |
| 16976 | MT969804.1 | \|Severe acute respiratory syndrome coro | ···················· | ···················· | ···················· | ···················· | ···················· | ···················· | |
| 16977 | MT969805.1 | \|Severe acute respiratory syndrome coro | ···················· | ···················· | ···················· | ···················· | ···················· | ···················· | |
| 16978 | MT969806.1 | \|Severe acute respiratory syndrome coro | ···················· | ···················· | ···················· | ···················· | ···················· | ···················· | |
| 16979 | MT969807.1 | \|Severe acute respiratory syndrome coro | ···················· | ···················· | ···················· | ···················· | ···················· | ···················· | |
| 16980 | MT969808.1 | \|Severe acute respiratory syndrome coro | ···················· | ···················· | ···················· | ···················· | ···················· | ···················· | |
| 16981 | MT969809.1 | \|Severe acute respiratory syndrome coro | ···················· | ···················· | ···················· | ···················· | ···················· | ···················· | |
| 16982 | MT969810.1 | \|Severe acute respiratory syndrome coro | ···················· | ···················· | ···················· | ···················· | ···················· | ···················· | |
| 16983 | MT969811.1 | \|Severe acute respiratory syndrome coro | ···················· | ···················· | ···················· | ···················· | ···················· | ···················· | |
| 16984 | MT969812.1 | \|Severe acute respiratory syndrome coro | ···················· | ···················· | ···················· | ···················· | ···················· | ···················· | |
| 16985 | MT969813.1 | \|Severe acute respiratory syndrome coro | ···················· | ···················· | ···················· | ···················· | ···················· | ···················· | |
| 16986 | MT969814.1 | \|Severe acute respiratory syndrome coro | ···················· | ···················· | ···················· | ···················· | ···················· | ···················· | |
| 16987 | MT969815.1 | \|Severe acute respiratory syndrome coro | ···················· | ···················· | ···················· | ···················· | ···················· | ···················· | |
| 16988 | MT969816.1 | \|Severe acute respiratory syndrome coro | ···················· | ···················· | ···················· | ···················· | ···················· | ···················· | |
| 16989 | MT969817.1 | \|Severe acute respiratory syndrome coro | ···················· | ···················· | ···················· | ···················· | ···················· | ···················· | |
| 16990 | MT969818.1 | \|Severe acute respiratory syndrome coro | ···················· | ···················· | ···················· | ···················· | ···················· | ···················· | |
| 16991 | MT969819.1 | \|Severe acute respiratory syndrome coro | ···················· | ···················· | ···················· | ···················· | ···················· | ···················· | |
| 16992 | MT969820.1 | \|Severe acute respiratory syndrome coro | ···················· | ···················· | ···················· | ···················· | ···················· | ···················· | |
| 16993 | MT969821.1 | \|Severe acute respiratory syndrome coro | ···················· | ···················· | ···················· | ···················· | ···················· | ···················· | |
| 16994 | MT969822.1 | \|Severe acute respiratory syndrome coro | ···················· | ···················· | ···················· | ···················· | ···················· | ···················· | |
| 16995 | MT969823.1 | \|Severe acute respiratory syndrome coro | ···················· | ···················· | ···················· | ···················· | ···················· | ···················· | |
| 16996 | MT969824.1 | \|Severe acute respiratory syndrome coro | ···················· | ···················· | ···················· | ···················· | ···················· | ···················· | |
| 16997 | MT969825.1 | \|Severe acute respiratory syndrome coro | ···················· | ···················· | ···················· | ···················· | ···················· | ···················· | |
| 16998 | MT969826.1 | \|Severe acute respiratory syndrome coro | ···················· | ···················· | ···················· | ···················· | ···················· | ···················· | |
| 16999 | MT969827.1 | \|Severe acute respiratory syndrome coro | ···················· | ···················· | ···················· | ···················· | ···················· | ···················· | |
| 17000 | MT969828.1 | \|Severe acute respiratory syndrome coro | ···················· | ···················· | ···················· | ···················· | ···················· | ···················· | |
| 17001 | MT969829.1 | \|Severe acute respiratory syndrome coro | ···················· | ···················· | ···················· | ···················· | ···················· | ···················· | |
| 17002 | MT969830.1 | \|Severe acute respiratory syndrome coro | ···················· | ···················· | ···················· | ···················· | ···················· | ···················· | |
| 17003 | MT969831.1 | \|Severe acute respiratory syndrome coro | ···················· | ···················· | ···················· | ···················· | ···················· | ···················· | |
| 17004 | MT969832.1 | \|Severe acute respiratory syndrome coro | ···················· | ···················· | ···················· | ···················· | ···················· | ···················· | |
| 17006 | MT969834.1 | \|Severe acute respiratory syndrome coro | ···················· | ···················· | ···················· | ···················· | ···················· | ···················· | |
| 17007 | MT969835.1 | \|Severe acute respiratory syndrome coro | ···················· | ···················· | ···················· | ···················· | ···················· | ···················· | |
| 17008 | MT969836.1 | \|Severe acute respiratory syndrome coro | ···················· | ···················· | ···················· | ···················· | ···················· | ···················· | |
| 17009 | MT969837.1 | \|Severe acute respiratory syndrome coro | ···················· | ···················· | ···················· | ···················· | ···················· | ···················· | |
| 17010 | MT969838.1 | \|Severe acute respiratory syndrome coro | ···················· | ···················· | ···················· | ···················· | ···················· | ···················· | |
| 17011 | MT969839.1 | \|Severe acute respiratory syndrome coro | ···················· | ···················· | ···················· | ···················· | ···················· | ···················· | |
| 17013 | MT969841.1 | \|Severe acute respiratory syndrome coro | ···················· | ···················· | ···················· | ···················· | ···················· | ···················· | |
| 17014 | MT969842.1 | \|Severe acute respiratory syndrome coro | ···················· | ···················· | ···················· | ···················· | ···················· | ···················· | |
| 17015 | MT969844.1 | \|Severe acute respiratory syndrome coro | ···················· | ···················· | ···················· | ···················· | ···················· | ···················· | |
| 17016 | MT969845.1 | \|Severe acute respiratory syndrome coro | ···················· | ···················· | ···················· | ···················· | ···················· | ···················· | |
| 17017 | MT969847.1 | \|Severe acute respiratory syndrome coro | ···················· | ···················· | ···················· | ···················· | ·NN················· | ···················· | |
| 17018 | MT969848.1 | \|Severe acute respiratory syndrome coro | ···················· | ···················· | ···················· | ···················· | ···················· | ···················· | |
| 17019 | MT969849.1 | \|Severe acute respiratory syndrome coro | ···················· | ···················· | ···················· | ···················· | ···················· | ···················· | |
| 17020 | MT969850.1 | \|Severe acute respiratory syndrome coro | ···················· | ···················· | ···················· | ···················· | ···················· | ···················· | |
| 17021 | MT969851.1 | \|Severe acute respiratory syndrome coro | ···················· | ···················· | ···················· | ···················· | ···················· | ···················· | |
| 17022 | MT969852.1 | \|Severe acute respiratory syndrome coro | ···················· | ···················· | ···················· | ···················· | ···················· | ···················· | |
| 17023 | MT969853.1 | \|Severe acute respiratory syndrome coro | ···················· | ···················· | ···················· | ···················· | ···················· | ···················· | |
| 17024 | MT969854.1 | \|Severe acute respiratory syndrome coro | ···················· | ···················· | ···················· | ···················· | ···················· | ···················· | |
| 17025 | MT969855.1 | \|Severe acute respiratory syndrome coro | ···················· | ···················· | ···················· | ···················· | ···················· | ···················· | |
| 17026 | MT969856.1 | \|Severe acute respiratory syndrome coro | ···················· | ···················· | ···················· | ···················· | ···················· | ···················· | |
| 17027 | MT969857.1 | \|Severe acute respiratory syndrome coro | ···················· | ···················· | ···················· | ···················· | NNN················· | ···················· | 1 |
| 17028 | MT969858.1 | \|Severe acute respiratory syndrome coro | ···················· | ···················· | ···················· | ···················· | ···················· | ···················· | |
| 17030 | MT969861.1 | \|Severe acute respiratory syndrome coro | ···················· | ···················· | ···················· | ···················· | ···················· | ···················· | |
| 17031 | MT969862.1 | \|Severe acute respiratory syndrome coro | ···················· | ···················· | ···················· | ···················· | ···················· | ···················· | |
| 17032 | MT969863.1 | \|Severe acute respiratory syndrome coro | ···················· | ···················· | ···················· | ···················· | ···················· | ···················· | |
| 17033 | MT969864.1 | \|Severe acute respiratory syndrome coro | ···················· | ···················· | ···················· | ···················· | ···················· | ···················· | |
| 17035 | MT969867.1 | \|Severe acute respiratory syndrome coro | ···················· | ···················· | ···················· | ···················· | ···················· | ···················· | |
| 17037 | MT969870.1 | \|Severe acute respiratory syndrome coro | ···················· | ···················· | ···················· | ···················· | ···················· | ···················· | |
| 17038 | MT969871.1 | \|Severe acute respiratory syndrome coro | ···················· | ···················· | ···················· | ···················· | ···················· | ···················· | |
| 17039 | MT969872.1 | \|Severe acute respiratory syndrome coro | ···················· | ···················· | ···················· | ···················· | ···················· | ···················· | |
| 17040 | MT969874.1 | \|Severe acute respiratory syndrome coro | ···················· | ···················· | ···················· | ···················· | ···················· | ···················· | |
| 17041 | MT969875.1 | \|Severe acute respiratory syndrome coro | ···················· | ···················· | ···················· | ···················· | ···················· | ···················· | |
| 17042 | MT969876.1 | \|Severe acute respiratory syndrome coro | ···················· | ···················· | ···················· | ···················· | ···················· | ···················· | |
| 17043 | MT969877.1 | \|Severe acute respiratory syndrome coro | ···················· | ···················· | ···················· | ···················· | ···················· | ···················· | |
| 17044 | MT969878.1 | \|Severe acute respiratory syndrome coro | ···················· | ···················· | ···················· | ···················· | ···················· | ···················· | |
| 17045 | MT969879.1 | \|Severe acute respiratory syndrome coro | ···················· | ···················· | ···················· | ···················· | ···················· | ···················· | |
| 17046 | MT969880.1 | \|Severe acute respiratory syndrome coro | ···················· | ···················· | ···················· | ···················· | ···················· | ···················· | |
| 17047 | MT969881.1 | \|Severe acute respiratory syndrome coro | ···················· | ···················· | ···················· | ···················· | ···················· | ···················· | |
| 17048 | MT969882.1 | \|Severe acute respiratory syndrome coro | ···················· | ···················· | ···················· | ···················· | ···················· | ···················· | |

17049 MT969883.1 |Severe acute respiratory syndrome coro .................... .................... ..................... .................... .................... ....................
17050 MT969884.1 |Severe acute respiratory syndrome coro .................... .................... ..................... .................... .................... ....................
17051 MT969885.1 |Severe acute respiratory syndrome coro .................... .................... ..................... .................... .................... ....................
17052 MT969886.1 |Severe acute respiratory syndrome coro .................... .................... ..................... .................... .................... ....................
17053 MT969887.1 |Severe acute respiratory syndrome coro .................... .................... ..................... .................... .................... ....................
17054 MT969888.1 |Severe acute respiratory syndrome coro .................... .................... ..................... .................... .................... ....................
17055 MT969889.1 |Severe acute respiratory syndrome coro .................... .................... ..................... .................... .................... ....................
17056 MT969890.1 |Severe acute respiratory syndrome coro .................... .................... ..................... .................... .................... ....................
17057 MT969891.1 |Severe acute respiratory syndrome coro .................... .................... ..................... .................... .................... ....................
17058 MT969892.1 |Severe acute respiratory syndrome coro .................... .................... ..................... .................... .................... ....................
17059 MT969893.1 |Severe acute respiratory syndrome coro .................... .................... ..................... .................... .................... ....................
17060 MT969894.1 |Severe acute respiratory syndrome coro .................... .................... ..................... .................... .................... ....................
17061 MT969895.1 |Severe acute respiratory syndrome coro .................... .................... ..................... .................... .................... ....................
17062 MT969896.1 |Severe acute respiratory syndrome coro .................... .................... ..................... .................... .................... ....................
17063 MT969897.1 |Severe acute respiratory syndrome coro .................... .................... ..................... .................... .................... ....................
17064 MT969898.1 |Severe acute respiratory syndrome coro .................... .................... ..................... .................... .................... ....................
17065 MT969899.1 |Severe acute respiratory syndrome coro .................... .................... ..................... .................... .................... ....................
17066 MT969900.1 |Severe acute respiratory syndrome coro .................... .................... ..................... .................... .................... ....................
17067 MT969901.1 |Severe acute respiratory syndrome coro .................... .................... ..................... .................... .................... ....................
17068 MT969902.1 |Severe acute respiratory syndrome coro .................... .................... ..................... .................... .................... ....................
17069 MT969903.1 |Severe acute respiratory syndrome coro .................... .................... ..................... .................... .................... ....................
17070 MT969905.1 |Severe acute respiratory syndrome coro .................... .................... ..................... .................... .................... ....................
17071 MT969906.1 |Severe acute respiratory syndrome coro .................... .................... ..................... .................... .................... ....................
17072 MT969907.1 |Severe acute respiratory syndrome coro .................... .................... ..................... .................... .................... ....................
17073 MT969908.1 |Severe acute respiratory syndrome coro .................... .................... ..................... .................... .................... ....................
17074 MT969909.1 |Severe acute respiratory syndrome coro .................... .................... ..................... .................... .................... ....................
17075 MT969910.1 |Severe acute respiratory syndrome coro .................... .................... ..................... .................... .................... ....................
17076 MT969911.1 |Severe acute respiratory syndrome coro .................... .................... ..................... .................... .................... ....................
17077 MT969912.1 |Severe acute respiratory syndrome coro .................... .................... ..................... .................... .................... ....................
17078 MT969913.1 |Severe acute respiratory syndrome coro .................... .................... ..................... .................... .................... ....................
17079 MT969914.1 |Severe acute respiratory syndrome coro .................... .................... ..................... .................... .................... ....................
17080 MT969915.1 |Severe acute respiratory syndrome coro .................... .................... ..................... .................... .................... ....................
17081 MT969916.1 |Severe acute respiratory syndrome coro .................... .................... ..................... .................... .................... ....................
17082 MT969917.1 |Severe acute respiratory syndrome coro .................... .................... ..................... .................... .................... ....................
17083 MT969918.1 |Severe acute respiratory syndrome coro .................... .................... ..................... .................... .................... ....................
17084 MT969919.1 |Severe acute respiratory syndrome coro .................... .................... ..................... .................... .................... ....................
17085 MT969920.1 |Severe acute respiratory syndrome coro .................... .................... ..................... .................... .................... ....................
17086 MT969921.1 |Severe acute respiratory syndrome coro .................... .................... ..................... .................... .................... ....................
17087 MT969922.1 |Severe acute respiratory syndrome coro .................... .................... ..................... .................... .................... ....................
17088 MT969923.1 |Severe acute respiratory syndrome coro .................... .................... ..................... .................... .................... ....................
17089 MT969924.1 |Severe acute respiratory syndrome coro .................... .................... ..................... .................... .................... ....................
17090 MT969925.1 |Severe acute respiratory syndrome coro .................... .................... ..................... .................... .................... ....................
17091 MT969926.1 |Severe acute respiratory syndrome coro .................... .................... ..................... .................... .................... ....................
17092 MT969927.1 |Severe acute respiratory syndrome coro .................... .................... ..................... .................... .................... ....................
17093 MT969928.1 |Severe acute respiratory syndrome coro .................... .................... ..................... .................... .................... ....................
17094 MT969929.1 |Severe acute respiratory syndrome coro .................... .................... ..................... .................... .................... ....................
17095 MT969930.1 |Severe acute respiratory syndrome coro .................... .................... ..................... .................... .................... ....................
17096 MT969931.1 |Severe acute respiratory syndrome coro .................... .................... ..................... .................... .................... ....................
17097 MT969932.1 |Severe acute respiratory syndrome coro .................... .................... ..................... .................... .................... ....................
17098 MT969933.1 |Severe acute respiratory syndrome coro .................... .................... ..................... .................... .................... ....................
17099 MT969934.1 |Severe acute respiratory syndrome coro .................... .................... ..................... .................... .................... ....................
17100 MT969935.1 |Severe acute respiratory syndrome coro .................... .................... ..................... .................... .................... ....................
17101 MT969936.1 |Severe acute respiratory syndrome coro .................... .................... ..................... .................... .................... ....................
17102 MT969937.1 |Severe acute respiratory syndrome coro .................... .................... ..................... .................... .................... ....................
17103 MT969938.1 |Severe acute respiratory syndrome coro .................... .................... ..................... .................... .................... ....................
17104 MT969939.1 |Severe acute respiratory syndrome coro .................... .................... ..................... .................... .................... ....................
17105 MT969940.1 |Severe acute respiratory syndrome coro .................... .................... ..................... .................... .................... ....................
17106 MT969941.1 |Severe acute respiratory syndrome coro .................... .................... ..................... .................... .................... ....................
17107 MT969942.1 |Severe acute respiratory syndrome coro .................... .................... ..................... .................... .................... ....................
17108 MT969943.1 |Severe acute respiratory syndrome coro .................... .................... ..................... .................... .................... ....................
17109 MT969944.1 |Severe acute respiratory syndrome coro .................... .................... ..................... .................... .................... ....................
17110 MT969945.1 |Severe acute respiratory syndrome coro .................... .................... ..................... .................... .................... ....................
17112 MT969947.1 |Severe acute respiratory syndrome coro .................... .................... ..................... .................... .................... ....................
17113 MT969948.1 |Severe acute respiratory syndrome coro .................... .................... ..................... .................... .................... ....................
17114 MT969949.1 |Severe acute respiratory syndrome coro .................... .................... ..................... .................... .................... ....................
17115 MT969950.1 |Severe acute respiratory syndrome coro .................... .................... ..................... .................... .................... ....................
17116 MT969951.1 |Severe acute respiratory syndrome coro .................... .................... ..................... .................... .................... ....................
17117 MT969952.1 |Severe acute respiratory syndrome coro .................... .................... ..................... .................... .................... ....................
17118 MT969953.1 |Severe acute respiratory syndrome coro .................... .................... ..................... .................... .................... ....................
17119 MT969954.1 |Severe acute respiratory syndrome coro .................... .................... ..................... .................... .................... ....................
17120 MT969955.1 |Severe acute respiratory syndrome coro .................... .................... ..................... .................... .................... ....................
17121 MT969956.1 |Severe acute respiratory syndrome coro .................... .................... ..................... .................... .................... ....................
17122 MT969957.1 |Severe acute respiratory syndrome coro .................... .................... ..................... .................... .................... ....................
17123 MT969958.1 |Severe acute respiratory syndrome coro .................... .................... ..................... .................... .................... ....................
17124 MT969959.1 |Severe acute respiratory syndrome coro .................... .................... ..................... .................... .................... ....................
17125 MT969960.1 |Severe acute respiratory syndrome coro .................... .................... ..................... .................... .................... ....................
17126 MT969961.1 |Severe acute respiratory syndrome coro .................... .................... ..................... .................... .................... ....................
17127 MT969962.1 |Severe acute respiratory syndrome coro .................... .................... ..................... .................... .................... ....................

| | | | | | | | | | |
|---|---|---|---|---|---|---|---|---|---|
| 17128 | MT969963.1 | Severe acute respiratory syndrome coro | ···················· | ···················· | ···················· | ···················· | ···················· | ···················· | |
| 17129 | MT969964.1 | Severe acute respiratory syndrome coro | ···················· | ···················· | ···················· | ···················· | ···················· | ···················· | |
| 17130 | MT969965.1 | Severe acute respiratory syndrome coro | ···················· | ···················· | ···················· | ···················· | ···················· | ···················· | |
| 17131 | MT969966.1 | Severe acute respiratory syndrome coro | ···················· | ···················· | ···················· | ···················· | ···················· | ···················· | |
| 17132 | MT969967.1 | Severe acute respiratory syndrome coro | ···················· | ···················· | ···················· | ···················· | ···················· | ···················· | |
| 17133 | MT969968.1 | Severe acute respiratory syndrome coro | ···················· | ···················· | ···················· | ···················· | ···················· | ···················· | |
| 17134 | MT969969.1 | Severe acute respiratory syndrome coro | ···················· | ···················· | ···················· | ···················· | ···················· | ···················· | |
| 17135 | MT969970.1 | Severe acute respiratory syndrome coro | ···················· | ···················· | ···················· | ···················· | ···················· | ···················· | |
| 17136 | MT969971.1 | Severe acute respiratory syndrome coro | ···················· | ···················· | ···················· | ···················· | ···················· | ···················· | |
| 17137 | MT969972.1 | Severe acute respiratory syndrome coro | ···················· | ···················· | ···················· | ···················· | ···················· | ···················· | |
| 17138 | MT969973.1 | Severe acute respiratory syndrome coro | ···················· | ···················· | ···················· | ···················· | ···················· | ···················· | |
| 17139 | MT969974.1 | Severe acute respiratory syndrome coro | ···················· | ···················· | ···················· | ···················· | ···················· | ···················· | |
| 17140 | MT969975.1 | Severe acute respiratory syndrome coro | ···················· | ···················· | ···················· | ···················· | ···················· | ···················· | |
| 17141 | MT969976.1 | Severe acute respiratory syndrome coro | ···················· | ···················· | ···················· | ···················· | ···················· | ···················· | |
| 17142 | MT969977.1 | Severe acute respiratory syndrome coro | ···················· | ···················· | ···················· | ···················· | ···················· | ···················· | |
| 17143 | MT969978.1 | Severe acute respiratory syndrome coro | ···················· | ···················· | ···················· | ···················· | ···················· | ···················· | |
| 17144 | MT969979.1 | Severe acute respiratory syndrome coro | ···················· | ···················· | ···················· | ···················· | ···················· | ···················· | |
| 17145 | MT969981.1 | Severe acute respiratory syndrome coro | ···················· | ···················· | ···················· | ···················· | ···················· | ···················· | |
| 17146 | MT969982.1 | Severe acute respiratory syndrome coro | ···················· | ···················· | ···················· | ···················· | ···················· | ···················· | |
| 17147 | MT969983.1 | Severe acute respiratory syndrome coro | ···················· | ···················· | ···················· | ···················· | ···················· | ···················· | |
| 17148 | MT969984.1 | Severe acute respiratory syndrome coro | ···················· | ···················· | ···················· | ···················· | ···················· | ···················· | |
| 17149 | MT969985.1 | Severe acute respiratory syndrome coro | ···················· | ···················· | ···················· | ···················· | ···················· | ···················· | |
| 17150 | MT969986.1 | Severe acute respiratory syndrome coro | ···················· | ···················· | ···················· | ···················· | ···················· | ···················· | |
| 17151 | MT969987.1 | Severe acute respiratory syndrome coro | ···················· | ···················· | ···················· | ···················· | ···················· | ···················· | |
| 17152 | MT969988.1 | Severe acute respiratory syndrome coro | ···················· | ···················· | ···················· | ···················· | ···················· | ···················· | |
| 17153 | MT969989.1 | Severe acute respiratory syndrome coro | ···················· | ···················· | ···················· | ···················· | ···················· | ···················· | |
| 17154 | MT969990.1 | Severe acute respiratory syndrome coro | ···················· | ···················· | ···················· | ···················· | ···················· | ···················· | |
| 17155 | MT969991.1 | Severe acute respiratory syndrome coro | ···················· | ···················· | ···················· | ···················· | ···················· | ···················· | |
| 17156 | MT969992.1 | Severe acute respiratory syndrome coro | ···················· | ···················· | ···················· | ···················· | ···················· | ···················· | |
| 17157 | MT969993.1 | Severe acute respiratory syndrome coro | ···················· | ···················· | ···················· | ···················· | ···················· | ···················· | |
| 17158 | MT969994.1 | Severe acute respiratory syndrome coro | ···················· | ···················· | ···················· | ···················· | ···················· | ···················· | |
| 17159 | MT969995.1 | Severe acute respiratory syndrome coro | ···················· | ···················· | ···················· | ···················· | ···················· | ···················· | |
| 17160 | MT969996.1 | Severe acute respiratory syndrome coro | ···················· | ···················· | ···················· | ···················· | ···················· | ···················· | |
| 17161 | MT969997.1 | Severe acute respiratory syndrome coro | ···················· | ···················· | ···················· | ···················· | ···················· | ···················· | |
| 17162 | MT969998.1 | Severe acute respiratory syndrome coro | ···················· | ···················· | ···················· | ···················· | ···················· | ···················· | |
| 17163 | MT969999.1 | Severe acute respiratory syndrome coro | ···················· | ···················· | ···················· | ···················· | ···················· | ···················· | |
| 17164 | MT970000.1 | Severe acute respiratory syndrome coro | ···················· | ···················· | ···················· | ···················· | ···················· | ···················· | |
| 17165 | MT970001.1 | Severe acute respiratory syndrome coro | ···················· | ···················· | ···················· | ···················· | NNN················· | ···················· | 1 |
| 17166 | MT970002.1 | Severe acute respiratory syndrome coro | ···················· | ···················· | ···················· | ···················· | ···················· | ···················· | |
| 17167 | MT970003.1 | Severe acute respiratory syndrome coro | ···················· | ···················· | ···················· | ···················· | ···················· | ···················· | |
| 17168 | MT970004.1 | Severe acute respiratory syndrome coro | ···················· | ···················· | ···················· | ···················· | ···················· | ···················· | |
| 17169 | MT970005.1 | Severe acute respiratory syndrome coro | ···················· | ···················· | ···················· | ···················· | ···················· | ···················· | |
| 17170 | MT970006.1 | Severe acute respiratory syndrome coro | ···················· | ···················· | ···················· | ···················· | ···················· | ···················· | |
| 17171 | MT970007.1 | Severe acute respiratory syndrome coro | ···················· | ···················· | ···················· | ···················· | ···················· | ···················· | |
| 17172 | MT970008.1 | Severe acute respiratory syndrome coro | ···················· | ···················· | ···················· | ···················· | ···················· | ···················· | |
| 17173 | MT970009.1 | Severe acute respiratory syndrome coro | ···················· | ···················· | ···················· | ···················· | ···················· | ···················· | |
| 17174 | MT970010.1 | Severe acute respiratory syndrome coro | ···················· | ···················· | ···················· | ···················· | ···················· | ···················· | |
| 17175 | MT970011.1 | Severe acute respiratory syndrome coro | ···················· | ···················· | ···················· | ···················· | ···················· | ···················· | |
| 17176 | MT970012.1 | Severe acute respiratory syndrome coro | ···················· | ···················· | ···················· | ···················· | ···················· | ···················· | |
| 17177 | MT970013.1 | Severe acute respiratory syndrome coro | ···················· | ···················· | ···················· | ···················· | ···················· | ···················· | |
| 17178 | MT970056.1 | Severe acute respiratory syndrome coro | ···················· | ···················· | ···················· | ···················· | ···················· | ···················· | |
| 17179 | MT970057.1 | Severe acute respiratory syndrome coro | ···················· | ···················· | ···················· | ···················· | ···················· | ···················· | |
| 17180 | MT970058.1 | Severe acute respiratory syndrome coro | ···················· | ···················· | ···················· | ···················· | ···················· | ···················· | |
| 17181 | MT970059.1 | Severe acute respiratory syndrome coro | ···················· | ···················· | ···················· | ···················· | ···················· | ···················· | |
| 17182 | MT970060.1 | Severe acute respiratory syndrome coro | ···················· | ···················· | ···················· | ···················· | ···················· | ···················· | |
| 17183 | MT970061.1 | Severe acute respiratory syndrome coro | ···················· | ···················· | ···················· | ···················· | ···················· | ···················· | |
| 17184 | MT970062.1 | Severe acute respiratory syndrome coro | ···················· | ···················· | ···················· | ···················· | ···················· | ···················· | |
| 17185 | MT970063.1 | Severe acute respiratory syndrome coro | ···················· | ···················· | ···················· | ···················· | ···················· | ···················· | |
| 17186 | MT970064.1 | Severe acute respiratory syndrome coro | ···················· | ···················· | ···················· | ···················· | ···················· | ···················· | |
| 17187 | MT970078.1 | Severe acute respiratory syndrome coro | ···················· | ···················· | ···················· | ···················· | ···················· | ···················· | |
| 17191 | MT970086.1 | Severe acute respiratory syndrome coro | ···················· | ···················· | ···················· | ···················· | ···················· | ···················· | |
| 17192 | MT970088.1 | Severe acute respiratory syndrome coro | ···················· | ···················· | ···················· | ···················· | ···················· | ···················· | |
| 17193 | MT970089.1 | Severe acute respiratory syndrome coro | ···················· | ···················· | ···················· | ···················· | NNN················· | ···················· | 1 |
| 17194 | MT970091.1 | Severe acute respiratory syndrome coro | ···················· | ···················· | ···················· | ···················· | ···················· | ···················· | |
| 17195 | MT970112.1 | Severe acute respiratory syndrome coro | ···················· | ···················· | ···················· | ···················· | ···················· | ···················· | |
| 17196 | MT970149.1 | Severe acute respiratory syndrome coro | ···················· | ···················· | ···················· | ···················· | ···················· | ···················· | |
| 17197 | MT970150.1 | Severe acute respiratory syndrome coro | ···················· | ···················· | ···················· | ···················· | ···················· | ···················· | |
| 17198 | MT970151.1 | Severe acute respiratory syndrome coro | ···················· | ···················· | ···················· | ···················· | ···················· | ···················· | |
| 17199 | MT970152.1 | Severe acute respiratory syndrome coro | ···················· | ···················· | ···················· | ···················· | ···················· | ···················· | |
| 17200 | MT970153.1 | Severe acute respiratory syndrome coro | ···················· | ···················· | ···················· | ···················· | ···················· | ···················· | |
| 17201 | MT970154.1 | Severe acute respiratory syndrome coro | ···················· | ···················· | ···················· | ···················· | ···················· | ···················· | |
| 17202 | MT970155.1 | Severe acute respiratory syndrome coro | ···················· | ···················· | ···················· | ···················· | ···················· | ···················· | |
| 17203 | MT970158.1 | Severe acute respiratory syndrome coro | ···················· | ···················· | ···················· | ···················· | ···················· | ···················· | |
| 17204 | MT970159.1 | Severe acute respiratory syndrome coro | ···················· | ···················· | ···················· | ···················· | ···················· | ···················· | |
| 17205 | MT970160.1 | Severe acute respiratory syndrome coro | ···················· | ···················· | ···················· | ···················· | ···················· | ···················· | |
| 17206 | MT970161.1 | Severe acute respiratory syndrome coro | ···················· | ···················· | ···················· | ···················· | ···················· | ···················· | |
| 17207 | MT970162.1 | Severe acute respiratory syndrome coro | ···················· | ···················· | ···················· | ···················· | ···················· | ···················· | |
| 17208 | MT970163.1 | Severe acute respiratory syndrome coro | ···················· | ···················· | ···················· | ···················· | ···················· | ···················· | |

| | | | | | | | | | |
|---|---|---|---|---|---|---|---|---|---|
| 17209 | MT970164.1 | \|Severe acute respiratory syndrome coro | .................... | .................... | .................... | .................... | .................... | .................... | |
| 17210 | MT970165.1 | \|Severe acute respiratory syndrome coro | .................... | .................... | .................... | .................... | .................... | .................... | |
| 17211 | MT970166.1 | \|Severe acute respiratory syndrome coro | .................... | .................... | .................... | .................... | .................... | .................... | |
| 17212 | MT970167.1 | \|Severe acute respiratory syndrome coro | .................... | .................... | .................... | .................... | .................... | .................... | |
| 17213 | MT970168.1 | \|Severe acute respiratory syndrome coro | .................... | .................... | .................... | .................... | .................... | .................... | |
| 17214 | MT970169.1 | \|Severe acute respiratory syndrome coro | .................... | .................... | .................... | .................... | .................... | .................... | |
| 17215 | MT970170.1 | \|Severe acute respiratory syndrome coro | .................... | .................... | .................... | .................... | .................... | .................... | |
| 17216 | MT970171.1 | \|Severe acute respiratory syndrome coro | .................... | .................... | .................... | .................... | .................... | .................... | |
| 17217 | MT970172.1 | \|Severe acute respiratory syndrome coro | .................... | .................... | .................... | .................... | .................... | .................... | |
| 17219 | MT970174.1 | \|Severe acute respiratory syndrome coro | .................... | .................... | .................... | .................... | .................... | .................... | |
| 17220 | MT970175.1 | \|Severe acute respiratory syndrome coro | .................... | .................... | .................... | .................... | .................... | .................... | |
| 17221 | MT970176.1 | \|Severe acute respiratory syndrome coro | .................... | .................... | .................... | .................... | .................... | .................... | |
| 17222 | MT970177.1 | \|Severe acute respiratory syndrome coro | .................... | .................... | .................... | .................... | .................... | .................... | |
| 17223 | MT970178.1 | \|Severe acute respiratory syndrome coro | .................... | .................... | .................... | .................... | .................... | .................... | |
| 17224 | MT970179.1 | \|Severe acute respiratory syndrome coro | .................... | .................... | .................... | .................... | .................... | .................... | |
| 17225 | MT970180.1 | \|Severe acute respiratory syndrome coro | .................... | .................... | .................... | .................... | .................... | .................... | |
| 17226 | MT970181.1 | \|Severe acute respiratory syndrome coro | .................... | .................... | .................... | .................... | .................... | .................... | |
| 17227 | MT970182.1 | \|Severe acute respiratory syndrome coro | .................... | .................... | .................... | .................... | NNN................. | .................... | 1 |
| 17228 | MT970183.1 | \|Severe acute respiratory syndrome coro | .................... | .................... | .................... | .................... | .................... | .................... | |
| 17229 | MT970184.1 | \|Severe acute respiratory syndrome coro | .................... | .................... | .................... | .................... | .................... | .................... | |
| 17230 | MT970185.1 | \|Severe acute respiratory syndrome coro | .................... | .................... | .................... | .................... | .................... | .................... | |
| 17231 | MT970186.1 | \|Severe acute respiratory syndrome coro | .................... | .................... | .................... | .................... | .................... | .................... | |
| 17232 | MT970187.1 | \|Severe acute respiratory syndrome coro | .................... | .................... | .................... | .................... | .................... | .................... | |
| 17233 | MT970188.1 | \|Severe acute respiratory syndrome coro | .................... | .................... | .................... | .................... | .................... | .................... | |
| 17234 | MT970189.1 | \|Severe acute respiratory syndrome coro | .................... | .................... | .................... | .................... | .................... | .................... | |
| 17235 | MT970190.1 | \|Severe acute respiratory syndrome coro | .................... | .................... | .................... | .................... | .................... | .................... | |
| 17236 | MT970191.1 | \|Severe acute respiratory syndrome coro | .................... | .................... | .................... | .................... | .................... | .................... | |
| 17237 | MT970192.1 | \|Severe acute respiratory syndrome coro | .................... | .................... | .................... | .................... | .................... | .................... | |
| 17238 | MT970193.1 | \|Severe acute respiratory syndrome coro | .................... | .................... | .................... | .................... | .................... | .................... | |
| 17239 | MT970194.1 | \|Severe acute respiratory syndrome coro | .................... | .................... | .................... | .................... | .................... | .................... | |
| 17240 | MT970195.1 | \|Severe acute respiratory syndrome coro | .................... | .................... | .................... | .................... | .................... | .................... | |
| 17241 | MT970196.1 | \|Severe acute respiratory syndrome coro | .................... | .................... | .................... | .................... | .................... | .................... | |
| 17242 | MT970197.1 | \|Severe acute respiratory syndrome coro | .................... | .................... | .................... | .................... | .................... | .................... | |
| 17243 | MT970198.1 | \|Severe acute respiratory syndrome coro | .................... | .................... | .................... | .................... | .................... | .................... | |
| 17244 | MT970199.1 | \|Severe acute respiratory syndrome coro | .................... | .................... | .................... | .................... | .................... | .................... | |
| 17245 | MT970200.1 | \|Severe acute respiratory syndrome coro | .................... | .................... | .................... | .................... | .................... | .................... | |
| 17246 | MT970201.1 | \|Severe acute respiratory syndrome coro | .................... | .................... | .................... | .................... | .................... | .................... | |
| 17247 | MT970202.1 | \|Severe acute respiratory syndrome coro | .................... | .................... | .................... | .................... | .................... | .................... | |
| 17248 | MT970203.1 | \|Severe acute respiratory syndrome coro | .................... | .................... | .................... | .................... | .................... | .................... | |
| 17249 | MT970204.1 | \|Severe acute respiratory syndrome coro | .................... | .................... | .................... | .................... | .................... | .................... | |
| 17250 | MT970205.1 | \|Severe acute respiratory syndrome coro | .................... | .................... | .................... | .................... | .................... | .................... | |
| 17251 | MT970206.1 | \|Severe acute respiratory syndrome coro | .................... | .................... | .................... | .................... | .................... | .................... | |
| 17252 | MT970207.1 | \|Severe acute respiratory syndrome coro | .................... | .................... | .................... | .................... | .................... | .................... | |
| 17253 | MT970208.1 | \|Severe acute respiratory syndrome coro | .................... | .................... | .................... | .................... | .................... | .................... | |
| 17254 | MT970209.1 | \|Severe acute respiratory syndrome coro | .................... | .................... | .................... | .................... | .................... | .................... | |
| 17255 | MT970210.1 | \|Severe acute respiratory syndrome coro | .................... | .................... | .................... | .................... | .................... | .................... | |
| 17256 | MT970211.1 | \|Severe acute respiratory syndrome coro | .................... | .................... | .................... | .................... | .................... | .................... | |
| 17257 | MT970212.1 | \|Severe acute respiratory syndrome coro | .................... | .................... | .................... | .................... | .................... | .................... | |
| 17258 | MT970213.1 | \|Severe acute respiratory syndrome coro | .................... | .................... | .................... | .................... | .................... | .................... | |
| 17259 | MT970214.1 | \|Severe acute respiratory syndrome coro | .................... | .................... | .................... | .................... | .................... | .................... | |
| 17260 | MT970215.1 | \|Severe acute respiratory syndrome coro | .................... | .................... | .................... | .................... | .................... | .................... | |
| 17261 | MT970216.1 | \|Severe acute respiratory syndrome coro | .................... | .................... | .................... | .................... | .................... | .................... | |
| 17262 | MT970217.1 | \|Severe acute respiratory syndrome coro | .................... | .................... | .................... | .................... | .................... | .................... | |
| 17263 | MT970218.1 | \|Severe acute respiratory syndrome coro | .................... | .................... | .................... | .................... | .................... | .................... | |
| 17264 | MT970219.1 | \|Severe acute respiratory syndrome coro | .................... | .................... | .................... | .................... | .................... | .................... | |
| 17265 | MT970220.1 | \|Severe acute respiratory syndrome coro | .................... | .................... | .................... | .................... | .................... | .................... | |
| 17266 | MT970221.1 | \|Severe acute respiratory syndrome coro | .................... | .................... | .................... | .................... | .................... | .................... | |
| 17267 | MT970222.1 | \|Severe acute respiratory syndrome coro | .................... | .................... | .................... | .................... | .................... | .................... | |
| 17268 | MT970223.1 | \|Severe acute respiratory syndrome coro | .................... | .................... | .................... | .................... | .................... | .................... | |
| 17269 | MT970224.1 | \|Severe acute respiratory syndrome coro | .................... | .................... | .................... | .................... | .................... | .................... | |
| 17271 | MT970226.1 | \|Severe acute respiratory syndrome coro | .................... | .................... | .................... | .................... | .................... | .................... | |
| 17272 | MT970227.1 | \|Severe acute respiratory syndrome coro | .................... | .................... | .................... | .................... | .................... | .................... | |
| 17273 | MT970228.1 | \|Severe acute respiratory syndrome coro | .................... | .................... | .................... | .................... | .................... | .................... | |
| 17274 | MT970229.1 | \|Severe acute respiratory syndrome coro | .................... | .................... | .................... | .................... | .................... | .................... | |
| 17275 | MT970230.1 | \|Severe acute respiratory syndrome coro | .................... | .................... | .................... | .................... | .................... | .................... | |
| 17276 | MT970231.1 | \|Severe acute respiratory syndrome coro | .................... | .................... | .................... | .................... | .................... | .................... | |
| 17277 | MT970232.1 | \|Severe acute respiratory syndrome coro | .................... | .................... | .................... | .................... | .................... | .................... | |
| 17278 | MT970233.1 | \|Severe acute respiratory syndrome coro | .................... | .................... | .................... | .................... | .................... | .................... | |
| 17279 | MT970234.1 | \|Severe acute respiratory syndrome coro | .................... | .................... | .................... | .................... | .................... | .................... | |
| 17280 | MT970235.1 | \|Severe acute respiratory syndrome coro | .................... | .................... | .................... | .................... | .................... | .................... | |
| 17281 | MT970236.1 | \|Severe acute respiratory syndrome coro | .................... | .................... | .................... | .................... | .................... | .................... | |
| 17282 | MT970237.1 | \|Severe acute respiratory syndrome coro | .................... | .................... | .................... | .................... | .................... | .................... | |
| 17283 | MT970238.1 | \|Severe acute respiratory syndrome coro | .................... | .................... | .................... | .................... | .................... | .................... | |
| 17284 | MT970239.1 | \|Severe acute respiratory syndrome coro | .................... | .................... | .................... | .................... | .................... | .................... | |
| 17285 | MT970240.1 | \|Severe acute respiratory syndrome coro | .................... | .................... | .................... | .................... | .................... | .................... | |
| 17286 | MT970241.1 | \|Severe acute respiratory syndrome coro | .................... | .................... | .................... | .................... | .................... | .................... | |
| 17287 | MT970242.1 | \|Severe acute respiratory syndrome coro | .................... | .................... | .................... | .................... | .................... | .................... | |
| 17288 | MT970243.1 | \|Severe acute respiratory syndrome coro | .................... | .................... | .................... | .................... | .................... | .................... | |

| | | | | | | | | | |
|---|---|---|---|---|---|---|---|---|---|
| 17289 | MT970244.1 | \|Severe acute respiratory syndrome coro | ···················· | ···················· | ···················· | ···················· | ···················· | ···················· | |
| 17290 | MT970245.1 | \|Severe acute respiratory syndrome coro | ···················· | ···················· | ···················· | ···················· | ···················· | ···················· | |
| 17291 | MT970246.1 | \|Severe acute respiratory syndrome coro | ···················· | ···················· | ···················· | ···················· | ···················· | ···················· | |
| 17292 | MT970247.1 | \|Severe acute respiratory syndrome coro | ···················· | ···················· | ···················· | ···················· | ···················· | ···················· | |
| 17293 | MT970248.1 | \|Severe acute respiratory syndrome coro | ···················· | ···················· | ···················· | ···················· | ···················· | ···················· | |
| 17294 | MT970249.1 | \|Severe acute respiratory syndrome coro | ···················· | ···················· | ···················· | ···················· | ···················· | ···················· | |
| 17295 | MT970250.1 | \|Severe acute respiratory syndrome coro | ···················· | ···················· | ···················· | ···················· | ···················· | ···················· | |
| 17296 | MT970251.1 | \|Severe acute respiratory syndrome coro | ···················· | ···················· | ···················· | ···················· | ···················· | ···················· | |
| 17297 | MT970252.1 | \|Severe acute respiratory syndrome coro | ···················· | ···················· | ···················· | ···················· | ···················· | ···················· | |
| 17298 | MT970253.1 | \|Severe acute respiratory syndrome coro | ···················· | ···················· | ···················· | ···················· | ···················· | ···················· | |
| 17299 | MT970254.1 | \|Severe acute respiratory syndrome coro | ···················· | ···················· | ···················· | ···················· | ···················· | ···················· | |
| 17300 | MT970255.1 | \|Severe acute respiratory syndrome coro | ···················· | ···················· | ···················· | ···················· | ···················· | ···················· | |
| 17301 | MT970256.1 | \|Severe acute respiratory syndrome coro | ···················· | ···················· | ···················· | ···················· | ···················· | ···················· | |
| 17302 | MT970257.1 | \|Severe acute respiratory syndrome coro | ···················· | ···················· | ···················· | ···················· | ·NNN················ | ···················· | 1 |
| 17303 | MT970258.1 | \|Severe acute respiratory syndrome coro | ···················· | ···················· | ···················· | ···················· | ···················· | ···················· | |
| 17304 | MT970259.1 | \|Severe acute respiratory syndrome coro | ···················· | ···················· | ···················· | ···················· | ···················· | ···················· | |
| 17305 | MT970260.1 | \|Severe acute respiratory syndrome coro | ···················· | ···················· | ···················· | ···················· | ···················· | ···················· | |
| 17306 | MT970261.1 | \|Severe acute respiratory syndrome coro | ···················· | ···················· | ···················· | ···················· | ···················· | ···················· | |
| 17307 | MT970262.1 | \|Severe acute respiratory syndrome coro | ···················· | ···················· | ···················· | ···················· | ···················· | ···················· | |
| 17308 | MT970263.1 | \|Severe acute respiratory syndrome coro | ···················· | ···················· | ···················· | ···················· | ···················· | ···················· | |
| 17309 | MT970264.1 | \|Severe acute respiratory syndrome coro | ···················· | ···················· | ···················· | ···················· | ···················· | ···················· | |
| 17310 | MT970265.1 | \|Severe acute respiratory syndrome coro | ···················· | ···················· | ···················· | ···················· | ···················· | ···················· | |
| 17311 | MT970267.1 | \|Severe acute respiratory syndrome coro | ···················· | ···················· | ···················· | ···················· | ···················· | ···················· | |
| 17312 | MT970268.1 | \|Severe acute respiratory syndrome coro | ···················· | ···················· | ···················· | ···················· | ···················· | ···················· | |
| 17313 | MT970269.1 | \|Severe acute respiratory syndrome coro | ···················· | ···················· | ···················· | ···················· | ···················· | ···················· | |
| 17314 | MT970270.1 | \|Severe acute respiratory syndrome coro | ···················· | ···················· | ···················· | ···················· | ···················· | ···················· | |
| 17315 | MT970271.1 | \|Severe acute respiratory syndrome coro | ···················· | ···················· | ···················· | ···················· | ···················· | ···················· | |
| 17316 | MT970272.1 | \|Severe acute respiratory syndrome coro | ···················· | ···················· | ···················· | ···················· | ···················· | ···················· | |
| 17317 | MT970273.1 | \|Severe acute respiratory syndrome coro | ···················· | ···················· | ···················· | ···················· | ···················· | ···················· | |
| 17318 | MT970274.1 | \|Severe acute respiratory syndrome coro | ···················· | ···················· | ···················· | ···················· | ···················· | ···················· | |
| 17320 | MT970276.1 | \|Severe acute respiratory syndrome coro | ···················· | ···················· | ···················· | ···················· | ···················· | ···················· | |
| 17321 | MT970277.1 | \|Severe acute respiratory syndrome coro | ···················· | ···················· | ···················· | ···················· | ···················· | ···················· | |
| 17322 | MT970278.1 | \|Severe acute respiratory syndrome coro | ···················· | ···················· | ···················· | ···················· | ···················· | ···················· | |
| 17323 | MT970279.1 | \|Severe acute respiratory syndrome coro | ···················· | ···················· | ···················· | ···················· | ···················· | ···················· | |
| 17324 | MT970280.1 | \|Severe acute respiratory syndrome coro | ···················· | ···················· | ···················· | ···················· | ···················· | ···················· | |
| 17325 | MT970281.1 | \|Severe acute respiratory syndrome coro | ···················· | ···················· | ···················· | ···················· | ···················· | ···················· | |
| 17326 | MT970282.1 | \|Severe acute respiratory syndrome coro | ···················· | ···················· | ···················· | ···················· | ···················· | ···················· | |
| 17327 | MT970283.1 | \|Severe acute respiratory syndrome coro | ···················· | ···················· | ···················· | ···················· | NNN················· | ···················· | 1 |
| 17328 | MT970284.1 | \|Severe acute respiratory syndrome coro | ···················· | ···················· | ···················· | ···················· | ···················· | ···················· | |
| 17329 | MT970285.1 | \|Severe acute respiratory syndrome coro | ···················· | ···················· | ···················· | ···················· | ···················· | ···················· | |
| 17331 | MT970287.1 | \|Severe acute respiratory syndrome coro | ···················· | ···················· | ···················· | ···················· | ···················· | ···················· | |
| 17332 | MT970288.1 | \|Severe acute respiratory syndrome coro | ···················· | ···················· | ···················· | ···················· | ···················· | ···················· | |
| 17333 | MT970289.1 | \|Severe acute respiratory syndrome coro | ···················· | ···················· | ···················· | ···················· | ···················· | ···················· | |
| 17334 | MT970290.1 | \|Severe acute respiratory syndrome coro | ···················· | ···················· | ···················· | ···················· | ···················· | ···················· | |
| 17335 | MT970291.1 | \|Severe acute respiratory syndrome coro | ···················· | ···················· | ···················· | ···················· | ···················· | ···················· | |
| 17336 | MT970293.1 | \|Severe acute respiratory syndrome coro | ···················· | ···················· | ···················· | ···················· | ···················· | ···················· | |
| 17337 | MT970294.1 | \|Severe acute respiratory syndrome coro | ···················· | ···················· | ···················· | ···················· | ···················· | ···················· | |
| 17338 | MT970295.1 | \|Severe acute respiratory syndrome coro | ···················· | ···················· | ···················· | ···················· | ···················· | ···················· | |
| 17339 | MT970296.1 | \|Severe acute respiratory syndrome coro | ···················· | ···················· | ···················· | ···················· | ···················· | ···················· | |
| 17340 | MT970297.1 | \|Severe acute respiratory syndrome coro | ···················· | ···················· | ···················· | ···················· | ···················· | ···················· | |
| 17341 | MT970298.1 | \|Severe acute respiratory syndrome coro | ···················· | ···················· | ···················· | ···················· | ···················· | ···················· | |
| 17342 | MT970299.1 | \|Severe acute respiratory syndrome coro | ···················· | ···················· | ···················· | ···················· | ···················· | ···················· | |
| 17343 | MT970300.1 | \|Severe acute respiratory syndrome coro | ···················· | ···················· | ···················· | ···················· | ···················· | ···················· | |
| 17344 | MT970301.1 | \|Severe acute respiratory syndrome coro | ···················· | ···················· | ···················· | ···················· | ···················· | ···················· | |
| 17345 | MT970302.1 | \|Severe acute respiratory syndrome coro | ···················· | ···················· | ···················· | ···················· | ···················· | ···················· | |
| 17346 | MT970303.1 | \|Severe acute respiratory syndrome coro | ···················· | ···················· | ···················· | ···················· | ···················· | ···················· | |
| 17347 | MT970304.1 | \|Severe acute respiratory syndrome coro | ···················· | ···················· | ···················· | ···················· | ···················· | ···················· | |
| 17348 | MT970305.1 | \|Severe acute respiratory syndrome coro | ···················· | ···················· | ···················· | ···················· | ···················· | ···················· | |
| 17349 | MT970306.1 | \|Severe acute respiratory syndrome coro | ···················· | ···················· | ···················· | ···················· | ···················· | ···················· | |
| 17350 | MT970307.1 | \|Severe acute respiratory syndrome coro | ···················· | ···················· | ···················· | ···················· | ···················· | ···················· | |
| 17351 | MT970308.1 | \|Severe acute respiratory syndrome coro | ···················· | ···················· | ···················· | ···················· | ···················· | ···················· | |
| 17352 | MT970309.1 | \|Severe acute respiratory syndrome coro | ···················· | ···················· | ···················· | ···················· | ···················· | ···················· | |
| 17353 | MT970310.1 | \|Severe acute respiratory syndrome coro | ···················· | ···················· | ···················· | ···················· | ···················· | ···················· | |
| 17354 | MT970311.1 | \|Severe acute respiratory syndrome coro | ···················· | ···················· | ···················· | ···················· | ···················· | ···················· | |
| 17355 | MT970312.1 | \|Severe acute respiratory syndrome coro | ···················· | ···················· | ···················· | ···················· | ···················· | ···················· | |
| 17356 | MT970313.1 | \|Severe acute respiratory syndrome coro | ···················· | ···················· | ···················· | ···················· | ···················· | ···················· | |
| 17357 | MT970314.1 | \|Severe acute respiratory syndrome coro | ···················· | ···················· | ···················· | ···················· | ···················· | ···················· | |
| 17358 | MT970315.1 | \|Severe acute respiratory syndrome coro | ···················· | ···················· | ···················· | ···················· | ···················· | ···················· | |
| 17359 | MT970317.1 | \|Severe acute respiratory syndrome coro | ···················· | ···················· | ···················· | ···················· | ···················· | ···················· | |
| 17360 | MT970318.1 | \|Severe acute respiratory syndrome coro | ···················· | ···················· | ···················· | ···················· | ···················· | ···················· | |
| 17361 | MT970321.1 | \|Severe acute respiratory syndrome coro | ···················· | ···················· | ···················· | ···················· | ···················· | ···················· | |
| 17362 | MT970322.1 | \|Severe acute respiratory syndrome coro | ···················· | ···················· | ···················· | ···················· | ···················· | ···················· | |
| 17363 | MT970323.1 | \|Severe acute respiratory syndrome coro | ···················· | ···················· | ···················· | ···················· | ···················· | ···················· | |
| 17364 | MT970328.1 | \|Severe acute respiratory syndrome coro | ···················· | ···················· | ···················· | ···················· | ···················· | ···················· | |
| 17365 | MT970330.1 | \|Severe acute respiratory syndrome coro | ···················· | ···················· | ···················· | ···················· | ···················· | ···················· | |
| 17366 | MT970331.1 | \|Severe acute respiratory syndrome coro | ···················· | ···················· | ···················· | ···················· | ···················· | ···················· | |
| 17367 | MT970332.1 | \|Severe acute respiratory syndrome coro | ···················· | ···················· | ···················· | ···················· | ···················· | ···················· | |
| 17368 | MT970333.1 | \|Severe acute respiratory syndrome coro | ···················· | ···················· | ···················· | ···················· | ···················· | ···················· | |

```
17369 MT970334.1 |Severe acute respiratory syndrome coro    ....................  ....................  ....................  ....................  ....................  ....................
17370 MT970335.1 |Severe acute respiratory syndrome coro    ....................  ....................  ....................  ....................  ....................  ....................
17371 MT970336.1 |Severe acute respiratory syndrome coro    ....................  ....................  ....................  ....................  ....................  ....................
17372 MT970337.1 |Severe acute respiratory syndrome coro    ....................  ....................  ....................  ....................  ....................  ....................
17373 MT970338.1 |Severe acute respiratory syndrome coro    ....................  ....................  ....................  ....................  ....................  ....................
17375 MT970340.1 |Severe acute respiratory syndrome coro    ....................  ....................  ....................  ....................  ....................  ....................
17376 MT970341.1 |Severe acute respiratory syndrome coro    ....................  ....................  ....................  ....................  ....................  ....................
17377 MT970342.1 |Severe acute respiratory syndrome coro    ....................  ....................  ....................  ....................  ....................  ....................
17378 MT970343.1 |Severe acute respiratory syndrome coro    ....................  ....................  ....................  ....................  ....................  ....................
17379 MT970344.1 |Severe acute respiratory syndrome coro    ....................  ....................  ....................  ....................  ....................  ....................
17381 MT970346.1 |Severe acute respiratory syndrome coro    ....................  ....................  ....................  ....................  ....................  ....................
17382 MT970347.1 |Severe acute respiratory syndrome coro    ....................  ....................  ....................  ....................  ....................  ....................
17383 MT970348.1 |Severe acute respiratory syndrome coro    ....................  ....................  ....................  ....................  ....................  ....................
17384 MT970349.1 |Severe acute respiratory syndrome coro    ....................  ....................  ....................  ....................  ....................  ....................
17385 MT970350.1 |Severe acute respiratory syndrome coro    ....................  ....................  ....................  ....................  ....................  ....................
17386 MT970355.1 |Severe acute respiratory syndrome coro    ....................  ....................  ....................  ....................  ....................  ....................
17387 MT970356.1 |Severe acute respiratory syndrome coro    ....................  ....................  ....................  ....................  ....................  ....................
17388 MT970357.1 |Severe acute respiratory syndrome coro    ....................  ....................  ....................  ....................  ....................  ....................
17389 MT970359.1 |Severe acute respiratory syndrome coro    ....................  ....................  ....................  ....................  ....................  ....................
17390 MT970360.1 |Severe acute respiratory syndrome coro    ....................  ....................  ....................  ....................  ....................  ....................
17391 MT970362.1 |Severe acute respiratory syndrome coro    ....................  ....................  ....................  ....................  ....................  ....................
17392 MT970367.1 |Severe acute respiratory syndrome coro    ....................  ....................  ....................  ....................  ....................  ....................
17393 MT970370.1 |Severe acute respiratory syndrome coro    ....................  ....................  ....................  ....................  ....................  ....................
17394 MT970371.1 |Severe acute respiratory syndrome coro    ....................  ....................  ....................  ....................  ....................  ....................
17395 MT970372.1 |Severe acute respiratory syndrome coro    ....................  ....................  ....................  ....................  ....................  ....................
17396 MT970373.1 |Severe acute respiratory syndrome coro    ....................  ....................  ....................  ....................  ....................  ....................
17397 MT970374.1 |Severe acute respiratory syndrome coro    ....................  ....................  ....................  ....................  ....................  ....................
17398 MT970376.1 |Severe acute respiratory syndrome coro    ....................  ....................  ....................  ....................  ....................  ....................
17399 MT970379.1 |Severe acute respiratory syndrome coro    ....................  ....................  ....................  ....................  ....................  ....................
17400 MT970380.1 |Severe acute respiratory syndrome coro    ....................  ....................  ....................  ....................  ....................  ....................
17401 MT970382.1 |Severe acute respiratory syndrome coro    ....................  ....................  ....................  ....................  ....................  ....................
17402 MT970387.1 |Severe acute respiratory syndrome coro    ....................  ....................  ....................  ....................  ....................  ....................
17403 MT970388.1 |Severe acute respiratory syndrome coro    ....................  ....................  ....................  ....................  ....................  ....................
17404 MT970390.1 |Severe acute respiratory syndrome coro    ....................  ....................  ....................  ....................  ....................  ....................
17405 MT970393.1 |Severe acute respiratory syndrome coro    ....................  ....................  ....................  ....................  ....................  ....................
17406 MT970396.1 |Severe acute respiratory syndrome coro    ....................  ....................  ....................  ....................  ....................  ....................
17407 MT970403.1 |Severe acute respiratory syndrome coro    ....................  ....................  ....................  ....................  ....................  ....................
17408 MT970406.1 |Severe acute respiratory syndrome coro    ....................  ....................  ....................  ....................  ....................  ....................
17409 MT970408.1 |Severe acute respiratory syndrome coro    ....................  ....................  ....................  ....................  ....................  ....................
17410 MT970410.1 |Severe acute respiratory syndrome coro    ....................  ....................  ....................  ....................  ....................  ....................
17411 MT970411.1 |Severe acute respiratory syndrome coro    ....................  ....................  ....................  ....................  ....................  ....................
17412 MT970412.1 |Severe acute respiratory syndrome coro    ....................  ....................  ....................  ....................  ....................  ....................
17413 MT970414.1 |Severe acute respiratory syndrome coro    ....................  ....................  ....................  ....................  ....................  ....................
17414 MT970416.1 |Severe acute respiratory syndrome coro    ....................  ....................  ....................  ....................  ....................  ....................
17415 MT970419.1 |Severe acute respiratory syndrome coro    ....................  ....................  ....................  ....................  ....................  ....................
17416 MT970420.1 |Severe acute respiratory syndrome coro    ....................  ....................  ....................  ....................  ....................  ....................
17417 MT970422.1 |Severe acute respiratory syndrome coro    ....................  ....................  ....................  ....................  ....................  ....................
17418 MT970423.1 |Severe acute respiratory syndrome coro    ....................  ....................  ....................  ....................  ....................  ....................
17419 MT970439.1 |Severe acute respiratory syndrome coro    ....................  ....................  ....................  ....................  ....................  ....................
17420 MT970440.1 |Severe acute respiratory syndrome coro    ....................  ....................  ....................  ....................  ....................  ....................
17421 MT970441.1 |Severe acute respiratory syndrome coro    ....................  ....................  ....................  ....................  ....................  ....................
17422 MT970442.1 |Severe acute respiratory syndrome coro    ....................  ....................  ....................  ....................  ....................  ....................
17423 MT970443.1 |Severe acute respiratory syndrome coro    ....................  ....................  ....................  ....................  ....................  ....................
17424 MT970444.1 |Severe acute respiratory syndrome coro    ....................  ....................  ....................  ....................  ....................  ....................
17425 MT970445.1 |Severe acute respiratory syndrome coro    ....................  ....................  ....................  ....................  ....................  ....................
17426 MT970446.1 |Severe acute respiratory syndrome coro    ....................  ....................  ....................  ....................  ....................  ....................
17427 MT970447.1 |Severe acute respiratory syndrome coro    ....................  ....................  ....................  ....................  ....................  ....................
17428 MT970448.1 |Severe acute respiratory syndrome coro    ....................  ....................  ....................  ....................  ....................  ....................
17429 MT970449.1 |Severe acute respiratory syndrome coro    ....................  ....................  ....................  ....................  ....................  ....................
17430 MT970450.1 |Severe acute respiratory syndrome coro    ....................  ....................  ....................  ....................  ....................  ....................
17431 MT970451.1 |Severe acute respiratory syndrome coro    ....................  ....................  ....................  ....................  ....................  ....................
17432 MT970452.1 |Severe acute respiratory syndrome coro    ....................  ....................  ....................  ....................  ....................  ....................
17433 MT970453.1 |Severe acute respiratory syndrome coro    ....................  ....................  ....................  ....................  ....................  ....................
17434 MT970454.1 |Severe acute respiratory syndrome coro    ....................  ....................  ....................  ....................  ....................  ....................
17435 MT970455.1 |Severe acute respiratory syndrome coro    ....................  ....................  ....................  ....................  ....................  ....................
17436 MT970456.1 |Severe acute respiratory syndrome coro    ....................  ....................  ....................  ....................  ....................  ....................
17437 MT970457.1 |Severe acute respiratory syndrome coro    ....................  ....................  ....................  ....................  ....................  ....................
17438 MT970458.1 |Severe acute respiratory syndrome coro    ....................  ....................  ....................  ....................  ....................  ....................
17439 MT970459.1 |Severe acute respiratory syndrome coro    ....................  ....................  ....................  ....................  ....................  ....................
17440 MT970460.1 |Severe acute respiratory syndrome coro    ....................  ....................  ....................  ....................  ....................  ....................
17441 MT970461.1 |Severe acute respiratory syndrome coro    ....................  ....................  ....................  ....................  ....................  ....................
17442 MT970462.1 |Severe acute respiratory syndrome coro    ....................  ....................  ....................  ....................  ....................  ....................
17443 MT970463.1 |Severe acute respiratory syndrome coro    ....................  ....................  ....................  ....................  ....................  ....................
17444 MT970464.1 |Severe acute respiratory syndrome coro    ....................  ....................  ....................  ....................  ....................  ....................
17445 MT970465.1 |Severe acute respiratory syndrome coro    ....................  ....................  ....................  ....................  ....................  ....................
17446 MT970466.1 |Severe acute respiratory syndrome coro    ....................  ....................  ....................  ....................  ....................  ....................
17447 MT970467.1 |Severe acute respiratory syndrome coro    ....................  ....................  ....................  ....................  ....................  ....................
17448 MT970468.1 |Severe acute respiratory syndrome coro    ....................  ....................  ....................  ....................  ....................  ....................
```

17449 MT970469.1 |Severe acute respiratory syndrome coro .................... .................... .................... .................... .................... ....................
17450 MT970470.1 |Severe acute respiratory syndrome coro .................... .................... .................... .................... .................... ....................
17451 MT970471.1 |Severe acute respiratory syndrome coro .................... .................... .................... .................... .................... ....................
17452 MT970472.1 |Severe acute respiratory syndrome coro .................... .................... .................... .................... .................... ....................
17453 MT970473.1 |Severe acute respiratory syndrome coro .................... .................... .................... .................... .................... ....................
17454 MT970474.1 |Severe acute respiratory syndrome coro .................... .................... .................... .................... .................... ....................
17455 MT970475.1 |Severe acute respiratory syndrome coro .................... .................... .................... .................... .................... ....................
17456 MT970476.1 |Severe acute respiratory syndrome coro .................... .................... .................... .................... .................... ....................
17457 MT970477.1 |Severe acute respiratory syndrome coro .................... .................... .................... .................... .................... ....................
17458 MT970478.1 |Severe acute respiratory syndrome coro .................... .................... .................... .................... .................... ....................
17459 MT970479.1 |Severe acute respiratory syndrome coro .................... .................... .................... .................... .................... ....................
17460 MT970480.1 |Severe acute respiratory syndrome coro .................... .................... .................... .................... .................... ....................
17461 MT970481.1 |Severe acute respiratory syndrome coro .................... .................... .................... .................... .................... ....................
17462 MT970482.1 |Severe acute respiratory syndrome coro .................... .................... .................... .................... .................... ....................
17463 MT970483.1 |Severe acute respiratory syndrome coro .................... .................... .................... .................... .................... ....................
17464 MT970484.1 |Severe acute respiratory syndrome coro .................... .................... .................... .................... .................... ....................
17465 MT970485.1 |Severe acute respiratory syndrome coro .................... .................... .................... .................... .................... ....................
17466 MT970486.1 |Severe acute respiratory syndrome coro .................... .................... .................... .................... .................... ....................
17467 MT970487.1 |Severe acute respiratory syndrome coro .................... .................... .................... .................... .................... ....................
17468 MT970488.1 |Severe acute respiratory syndrome coro .................... .................... .................... .................... .................... ....................
17469 MT970489.1 |Severe acute respiratory syndrome coro .................... .................... .................... .................... .................... ....................
17470 MT970490.1 |Severe acute respiratory syndrome coro .................... .................... .................... .................... .................... ....................
17471 MT970491.1 |Severe acute respiratory syndrome coro .................... .................... .................... .................... .................... ....................
17472 MT970492.1 |Severe acute respiratory syndrome coro .................... .................... .................... .................... .................... ....................
17473 MT970493.1 |Severe acute respiratory syndrome coro .................... .................... .................... .................... .................... ....................
17474 MT970494.1 |Severe acute respiratory syndrome coro .................... .................... .................... .................... .................... ....................
17475 MT970495.1 |Severe acute respiratory syndrome coro .................... .................... .................... .................... .................... ....................
17476 MT970496.1 |Severe acute respiratory syndrome coro .................... .................... .................... .................... .................... ....................
17477 MT970497.1 |Severe acute respiratory syndrome coro .................... .................... .................... .................... .................... ....................
17478 MT970498.1 |Severe acute respiratory syndrome coro .................... .................... .................... .................... .................... ....................
17479 MT970499.1 |Severe acute respiratory syndrome coro .................... .................... .................... .................... .................... ....................
17480 MT970500.1 |Severe acute respiratory syndrome coro .................... .................... .................... .................... .................... ....................
17481 MT970501.1 |Severe acute respiratory syndrome coro .................... .................... .................... .................... .................... ....................
17482 MT970502.1 |Severe acute respiratory syndrome coro .................... .................... .................... .................... .................... ....................
17483 MT970503.1 |Severe acute respiratory syndrome coro .................... .................... .................... .................... .................... ....................
17484 MT970504.1 |Severe acute respiratory syndrome coro .................... .................... .................... .................... .................... ....................
17485 MT970505.1 |Severe acute respiratory syndrome coro .................... .................... .................... .................... .................... ....................
17486 MT970506.1 |Severe acute respiratory syndrome coro .................... .................... .................... .................... .................... ....................
17487 MT970507.1 |Severe acute respiratory syndrome coro .................... .................... .................... .................... .................... ....................
17488 MT970508.1 |Severe acute respiratory syndrome coro .................... .................... .................... .................... .................... ....................
17489 MT970509.1 |Severe acute respiratory syndrome coro .................... .................... .................... .................... .................... ....................
17490 MT970510.1 |Severe acute respiratory syndrome coro .................... .................... .................... .................... .................... ....................
17491 MT970511.1 |Severe acute respiratory syndrome coro .................... .................... .................... .................... .................... ....................
17492 MT970512.1 |Severe acute respiratory syndrome coro .................... .................... .................... .................... .................... ....................
17493 MT970513.1 |Severe acute respiratory syndrome coro .................... .................... .................... .................... .................... ....................
17494 MT970514.1 |Severe acute respiratory syndrome coro .................... .................... .................... .................... .................... ....................
17495 MT970515.1 |Severe acute respiratory syndrome coro .................... .................... .................... .................... .................... ....................
17496 MT970516.1 |Severe acute respiratory syndrome coro .................... .................... .................... .................... .................... ....................
17497 MT970517.1 |Severe acute respiratory syndrome coro .................... .................... .................... .................... .................... ....................
17498 MT970518.1 |Severe acute respiratory syndrome coro .................... .................... .................... .................... .................... ....................
17499 MT970519.1 |Severe acute respiratory syndrome coro .................... .................... .................... .................... .................... ....................
17500 MT970520.1 |Severe acute respiratory syndrome coro .................... .................... .................... .................... .................... ....................
17501 MT970521.1 |Severe acute respiratory syndrome coro .................... .................... .................... .................... .................... ....................
17502 MT970522.1 |Severe acute respiratory syndrome coro .................... .................... .................... .................... .................... ....................
17503 MT970523.1 |Severe acute respiratory syndrome coro .................... .................... .................... .................... .................... ....................
17504 MT970524.1 |Severe acute respiratory syndrome coro .................... .................... .................... .................... .................... ....................
17505 MT970525.1 |Severe acute respiratory syndrome coro .................... .................... .................... .................... .................... ....................
17506 MT970526.1 |Severe acute respiratory syndrome coro .................... .................... .................... .................... .................... ....................
17507 MT970527.1 |Severe acute respiratory syndrome coro .................... .................... .................... .................... .................... ....................
17508 MT970528.1 |Severe acute respiratory syndrome coro .................... .................... .................... .................... .................... ....................
17509 MT970529.1 |Severe acute respiratory syndrome coro .................... .................... .................... .................... .................... ....................
17510 MT970530.1 |Severe acute respiratory syndrome coro .................... .................... .................... .................... .................... ....................
17511 MT970531.1 |Severe acute respiratory syndrome coro .................... .................... .................... .................... .................... ....................
17512 MT970532.1 |Severe acute respiratory syndrome coro .................... .................... .................... .................... .................... ....................
17513 MT970533.1 |Severe acute respiratory syndrome coro .................... .................... .................... .................... .................... ....................
17514 MT970534.1 |Severe acute respiratory syndrome coro .................... .................... .................... .................... .................... ....................
17515 MT970535.1 |Severe acute respiratory syndrome coro .................... .................... .................... .................... .................... ....................
17516 MT970536.1 |Severe acute respiratory syndrome coro .................... .................... .................... .................... .................... ....................
17517 MT970537.1 |Severe acute respiratory syndrome coro .................... .................... .................... .................... .................... ....................
17518 MT970538.1 |Severe acute respiratory syndrome coro .................... .................... .................... .................... .................... ....................
17519 MT970539.1 |Severe acute respiratory syndrome coro .................... .................... .................... .................... .................... ....................
17520 MT970540.1 |Severe acute respiratory syndrome coro .................... .................... .................... .................... .................... ....................
17521 MT970541.1 |Severe acute respiratory syndrome coro .................... .................... .................... .................... .................... ....................
17522 MT970542.1 |Severe acute respiratory syndrome coro .................... .................... .................... .................... .................... ....................
17523 MT970543.1 |Severe acute respiratory syndrome coro .................... .................... .................... .................... .................... ....................
17524 MT970544.1 |Severe acute respiratory syndrome coro .................... .................... .................... .................... .................... ....................
17525 MT970545.1 |Severe acute respiratory syndrome coro .................... .................... .................... .................... .................... ....................
17526 MT970546.1 |Severe acute respiratory syndrome coro .................... .................... .................... .................... .................... ....................

17527 MT970547.1 |Severe acute respiratory syndrome coro .................... .................... .................... .................... .................... ....................
17528 MT970548.1 |Severe acute respiratory syndrome coro .................... .................... .................... .................... .................... ....................
17529 MT970549.1 |Severe acute respiratory syndrome coro .................... .................... .................... .................... .................... ....................
17530 MT970550.1 |Severe acute respiratory syndrome coro .................... .................... .................... .................... .................... ....................
17531 MT970551.1 |Severe acute respiratory syndrome coro .................... .................... .................... .................... .................... ....................
17532 MT970552.1 |Severe acute respiratory syndrome coro .................... .................... .................... .................... .................... ....................
17533 MT970553.1 |Severe acute respiratory syndrome coro .................... .................... .................... .................... .................... ....................
17534 MT970554.1 |Severe acute respiratory syndrome coro .................... .................... .................... .................... .................... ....................
17535 MT970555.1 |Severe acute respiratory syndrome coro .................... .................... .................... .................... .................... ....................
17536 MT970556.1 |Severe acute respiratory syndrome coro .................... .................... .................... .................... .................... ....................
17537 MT970557.1 |Severe acute respiratory syndrome coro .................... .................... .................... .................... .................... ....................
17538 MT970558.1 |Severe acute respiratory syndrome coro .................... .................... .................... .................... .................... ....................
17539 MT970559.1 |Severe acute respiratory syndrome coro .................... .................... .................... .................... .................... ....................
17540 MT970560.1 |Severe acute respiratory syndrome coro .................... .................... .................... .................... .................... ....................
17541 MT970561.1 |Severe acute respiratory syndrome coro .................... .................... .................... .................... .................... ....................
17542 MT970562.1 |Severe acute respiratory syndrome coro .................... .................... .................... .................... .................... ....................
17543 MT970563.1 |Severe acute respiratory syndrome coro .................... .................... .................... .................... .................... ....................
17544 MT970564.1 |Severe acute respiratory syndrome coro .................... .................... .................... .................... .................... ....................
17545 MT970565.1 |Severe acute respiratory syndrome coro .................... .................... .................... .................... .................... ....................
17546 MT970566.1 |Severe acute respiratory syndrome coro .................... .................... .................... .................... .................... ....................
17547 MT970567.1 |Severe acute respiratory syndrome coro .................... .................... .................... .................... .................... ....................
17548 MT970568.1 |Severe acute respiratory syndrome coro .................... .................... .................... .................... .................... ....................
17549 MT970569.1 |Severe acute respiratory syndrome coro .................... .................... .................... .................... .................... ....................
17550 MT970570.1 |Severe acute respiratory syndrome coro .................... .................... .................... .................... .................... ....................
17551 MT970571.1 |Severe acute respiratory syndrome coro .................... .................... .................... .................... .................... ....................
17552 MT970572.1 |Severe acute respiratory syndrome coro .................... .................... .................... .................... .................... ....................
17553 MT970573.1 |Severe acute respiratory syndrome coro .................... .................... .................... .................... .................... ....................
17554 MT970574.1 |Severe acute respiratory syndrome coro .................... .................... .................... .................... .................... ....................
17555 MT970575.1 |Severe acute respiratory syndrome coro .................... .................... .................... .................... .................... ....................
17556 MT970576.1 |Severe acute respiratory syndrome coro .................... .................... .................... .................... .................... ....................
17557 MT970577.1 |Severe acute respiratory syndrome coro .................... .................... .................... .................... .................... ....................
17558 MT970578.1 |Severe acute respiratory syndrome coro .................... .................... .................... .................... .................... ....................
17559 MT970579.1 |Severe acute respiratory syndrome coro .................... .................... .................... .................... .................... ....................
17560 MT970580.1 |Severe acute respiratory syndrome coro .................... .................... .................... .................... .................... ....................
17561 MT970581.1 |Severe acute respiratory syndrome coro .................... .................... .................... .................... .................... ....................
17562 MT970582.1 |Severe acute respiratory syndrome coro .................... .................... .................... .................... .................... ....................
17563 MT970583.1 |Severe acute respiratory syndrome coro .................... .................... .................... .................... .................... ....................
17564 MT970584.1 |Severe acute respiratory syndrome coro .................... .................... .................... .................... .................... ....................
17565 MT970585.1 |Severe acute respiratory syndrome coro .................... .................... .................... .................... .................... ....................
17566 MT970586.1 |Severe acute respiratory syndrome coro .................... .................... .................... .................... .................... ....................
17567 MT970587.1 |Severe acute respiratory syndrome coro .................... .................... .................... .................... .................... ....................
17568 MT970588.1 |Severe acute respiratory syndrome coro .................... .................... .................... .................... .................... ....................
17569 MT970589.1 |Severe acute respiratory syndrome coro .................... .................... .................... .................... .................... ....................
17570 MT970590.1 |Severe acute respiratory syndrome coro .................... .................... .................... .................... .................... ....................
17571 MT970591.1 |Severe acute respiratory syndrome coro .................... .................... .................... .................... .................... ....................
17572 MT970593.1 |Severe acute respiratory syndrome coro .................... .................... .................... .................... .................... ....................
17573 MT970594.1 |Severe acute respiratory syndrome coro .................... .................... .................... .................... .................... ....................
17574 MT970595.1 |Severe acute respiratory syndrome coro .................... .................... .................... .................... .................... ....................
17575 MT970596.1 |Severe acute respiratory syndrome coro .................... .................... .................... .................... .................... ....................
17576 MT970597.1 |Severe acute respiratory syndrome coro .................... .................... .................... .................... .................... ....................
17577 MT970598.1 |Severe acute respiratory syndrome coro .................... .................... .................... .................... .................... ....................
17578 MT970599.1 |Severe acute respiratory syndrome coro .................... .................... .................... .................... .................... ....................
17579 MT970600.1 |Severe acute respiratory syndrome coro .................... .................... .................... .................... .................... ....................
17580 MT970601.1 |Severe acute respiratory syndrome coro .................... .................... .................... .................... .................... ....................
17581 MT970602.1 |Severe acute respiratory syndrome coro .................... .................... .................... .................... .................... ....................
17582 MT970603.1 |Severe acute respiratory syndrome coro .................... .................... .................... .................... .................... ....................
17583 MT970604.1 |Severe acute respiratory syndrome coro .................... .................... .................... .................... .................... ....................
17584 MT970605.1 |Severe acute respiratory syndrome coro .................... .................... .................... .................... .................... ....................
17585 MT970606.1 |Severe acute respiratory syndrome coro .................... .................... .................... .................... .................... ....................
17586 MT970607.1 |Severe acute respiratory syndrome coro .................... .................... .................... .................... .................... ....................
17587 MT970608.1 |Severe acute respiratory syndrome coro .................... .................... .................... .................... .................... ....................
17588 MT970609.1 |Severe acute respiratory syndrome coro .................... .................... .................... .................... .................... ....................
17589 MT970610.1 |Severe acute respiratory syndrome coro .................... .................... .................... .................... .................... ....................
17590 MT970611.1 |Severe acute respiratory syndrome coro .................... .................... .................... .................... .................... ....................
17591 MT970612.1 |Severe acute respiratory syndrome coro .................... .................... .................... .................... .................... ....................
17592 MT970613.1 |Severe acute respiratory syndrome coro .................... .................... .................... .................... .................... ....................
17593 MT970614.1 |Severe acute respiratory syndrome coro .................... .................... .................... .................... .................... ....................
17594 MT970615.1 |Severe acute respiratory syndrome coro .................... .................... .................... .................... .................... ....................
17595 MT970616.1 |Severe acute respiratory syndrome coro .................... .................... .................... .................... .................... ....................
17596 MT970617.1 |Severe acute respiratory syndrome coro .................... .................... .................... .................... .................... ....................
17597 MT970618.1 |Severe acute respiratory syndrome coro .................... .................... .................... .................... .................... ....................
17598 MT970619.1 |Severe acute respiratory syndrome coro .................... .................... .................... .................... .................... ....................
17599 MT970620.1 |Severe acute respiratory syndrome coro .................... .................... .................... .................... .................... ....................
17600 MT970621.1 |Severe acute respiratory syndrome coro .................... .................... .................... .................... .................... ....................
17601 MT970622.1 |Severe acute respiratory syndrome coro .................... .................... .................... .................... .................... ....................
17602 MT970623.1 |Severe acute respiratory syndrome coro .................... .................... .................... .................... .................... ....................
17603 MT970624.1 |Severe acute respiratory syndrome coro .................... .................... .................... .................... .................... ....................
17604 MT970625.1 |Severe acute respiratory syndrome coro .................... .................... .................... .................... .................... ....................

| | | | | | | | |
|---|---|---|---|---|---|---|---|
| 17605 | MT970626.1 \|Severe acute respiratory syndrome coro | .................... | .................... | ..................... | .................... | .................... | .................... |
| 17606 | MT970627.1 \|Severe acute respiratory syndrome coro | .................... | .................... | ..................... | .................... | .................... | .................... |
| 17607 | MT970628.1 \|Severe acute respiratory syndrome coro | .................... | .................... | ..................... | .................... | .................... | .................... |
| 17608 | MT970629.1 \|Severe acute respiratory syndrome coro | .................... | .................... | ..................... | .................... | .................... | .................... |
| 17609 | MT970630.1 \|Severe acute respiratory syndrome coro | .................... | .................... | ..................... | .................... | .................... | .................... |
| 17610 | MT970631.1 \|Severe acute respiratory syndrome coro | .................... | .................... | ..................... | .................... | .................... | .................... |
| 17611 | MT970632.1 \|Severe acute respiratory syndrome coro | .................... | .................... | ..................... | .................... | .................... | .................... |
| 17612 | MT970633.1 \|Severe acute respiratory syndrome coro | .................... | .................... | ..................... | .................... | .................... | .................... |
| 17613 | MT970634.1 \|Severe acute respiratory syndrome coro | .................... | .................... | ..................... | .................... | .................... | .................... |
| 17614 | MT970635.1 \|Severe acute respiratory syndrome coro | .................... | .................... | ..................... | .................... | .................... | .................... |
| 17615 | MT970636.1 \|Severe acute respiratory syndrome coro | .................... | .................... | ..................... | .................... | .................... | .................... |
| 17616 | MT970637.1 \|Severe acute respiratory syndrome coro | .................... | .................... | ..................... | .................... | .................... | .................... |
| 17617 | MT970638.1 \|Severe acute respiratory syndrome coro | .................... | .................... | ..................... | .................... | .................... | .................... |
| 17618 | MT970639.1 \|Severe acute respiratory syndrome coro | .................... | .................... | ..................... | .................... | .................... | .................... |
| 17619 | MT970640.1 \|Severe acute respiratory syndrome coro | .................... | .................... | ..................... | .................... | .................... | .................... |
| 17620 | MT970641.1 \|Severe acute respiratory syndrome coro | .................... | .................... | ..................... | .................... | .................... | .................... |
| 17621 | MT970642.1 \|Severe acute respiratory syndrome coro | .................... | .................... | ..................... | .................... | .................... | .................... |
| 17622 | MT970643.1 \|Severe acute respiratory syndrome coro | .................... | .................... | ..................... | .................... | .................... | .................... |
| 17623 | MT970644.1 \|Severe acute respiratory syndrome coro | .................... | .................... | ..................... | .................... | .................... | .................... |
| 17624 | MT970645.1 \|Severe acute respiratory syndrome coro | .................... | .................... | ..................... | .................... | .................... | .................... |
| 17625 | MT970646.1 \|Severe acute respiratory syndrome coro | .................... | .................... | ..................... | .................... | .................... | .................... |
| 17626 | MT970647.1 \|Severe acute respiratory syndrome coro | .................... | .................... | ..................... | .................... | .................... | .................... |
| 17627 | MT970648.1 \|Severe acute respiratory syndrome coro | .................... | .................... | ..................... | .................... | .................... | .................... |
| 17628 | MT970649.1 \|Severe acute respiratory syndrome coro | .................... | .................... | ..................... | .................... | .................... | .................... |
| 17629 | MT970650.1 \|Severe acute respiratory syndrome coro | .................... | .................... | ..................... | .................... | .................... | .................... |
| 17630 | MT970651.1 \|Severe acute respiratory syndrome coro | .................... | .................... | ..................... | .................... | .................... | .................... |
| 17631 | MT970652.1 \|Severe acute respiratory syndrome coro | .................... | .................... | ..................... | .................... | .................... | .................... |
| 17632 | MT970653.1 \|Severe acute respiratory syndrome coro | .................... | .................... | ..................... | .................... | .................... | .................... |
| 17633 | MT970654.1 \|Severe acute respiratory syndrome coro | .................... | .................... | ..................... | .................... | .................... | .................... |
| 17634 | MT970655.1 \|Severe acute respiratory syndrome coro | .................... | .................... | ..................... | .................... | .................... | .................... |
| 17635 | MT970656.1 \|Severe acute respiratory syndrome coro | .................... | .................... | ..................... | .................... | .................... | .................... |
| 17636 | MT970657.1 \|Severe acute respiratory syndrome coro | .................... | .................... | ..................... | .................... | .................... | .................... |
| 17637 | MT970658.1 \|Severe acute respiratory syndrome coro | .................... | .................... | ..................... | .................... | .................... | .................... |
| 17638 | MT970659.1 \|Severe acute respiratory syndrome coro | .................... | .................... | ..................... | .................... | .................... | .................... |
| 17639 | MT970660.1 \|Severe acute respiratory syndrome coro | .................... | .................... | ..................... | .................... | .................... | .................... |
| 17640 | MT970661.1 \|Severe acute respiratory syndrome coro | .................... | .................... | ..................... | .................... | .................... | .................... |
| 17641 | MT970662.1 \|Severe acute respiratory syndrome coro | .................... | .................... | ..................... | .................... | .................... | .................... |
| 17642 | MT970663.1 \|Severe acute respiratory syndrome coro | .................... | .................... | ..................... | .................... | .................... | .................... |
| 17643 | MT970664.1 \|Severe acute respiratory syndrome coro | .................... | .................... | ..................... | .................... | .................... | .................... |
| 17644 | MT970665.1 \|Severe acute respiratory syndrome coro | .................... | .................... | ..................... | .................... | .................... | .................... |
| 17645 | MT970666.1 \|Severe acute respiratory syndrome coro | .................... | .................... | ..................... | .................... | .................... | .................... |
| 17646 | MT970667.1 \|Severe acute respiratory syndrome coro | .................... | .................... | ..................... | .................... | .................... | .................... |
| 17647 | MT970668.1 \|Severe acute respiratory syndrome coro | .................... | .................... | ..................... | .................... | .................... | .................... |
| 17648 | MT970669.1 \|Severe acute respiratory syndrome coro | .................... | .................... | ..................... | .................... | .................... | .................... |
| 17649 | MT970670.1 \|Severe acute respiratory syndrome coro | .................... | .................... | ..................... | .................... | .................... | .................... |
| 17650 | MT970671.1 \|Severe acute respiratory syndrome coro | .................... | .................... | ..................... | .................... | .................... | .................... |
| 17651 | MT970672.1 \|Severe acute respiratory syndrome coro | .................... | .................... | ..................... | .................... | .................... | .................... |
| 17652 | MT970673.1 \|Severe acute respiratory syndrome coro | .................... | .................... | ..................... | .................... | .................... | .................... |
| 17653 | MT970674.1 \|Severe acute respiratory syndrome coro | .................... | .................... | ..................... | .................... | .................... | .................... |
| 17654 | MT970675.1 \|Severe acute respiratory syndrome coro | .................... | .................... | ..................... | .................... | .................... | .................... |
| 17655 | MT970676.1 \|Severe acute respiratory syndrome coro | .................... | .................... | ..................... | .................... | .................... | .................... |
| 17656 | MT970677.1 \|Severe acute respiratory syndrome coro | .................... | .................... | ..................... | .................... | .................... | .................... |
| 17657 | MT970678.1 \|Severe acute respiratory syndrome coro | .................... | .................... | ..................... | .................... | .................... | .................... |
| 17658 | MT970679.1 \|Severe acute respiratory syndrome coro | .................... | .................... | ..................... | .................... | .................... | .................... |
| 17659 | MT970680.1 \|Severe acute respiratory syndrome coro | .................... | .................... | ..................... | .................... | .................... | .................... |
| 17660 | MT970681.1 \|Severe acute respiratory syndrome coro | .................... | .................... | ..................... | .................... | .................... | .................... |
| 17661 | MT970682.1 \|Severe acute respiratory syndrome coro | .................... | .................... | ..................... | .................... | .................... | .................... |
| 17662 | MT970683.1 \|Severe acute respiratory syndrome coro | .................... | .................... | ..................... | .................... | .................... | .................... |
| 17663 | MT970684.1 \|Severe acute respiratory syndrome coro | .................... | .................... | ..................... | .................... | .................... | .................... |
| 17664 | MT970685.1 \|Severe acute respiratory syndrome coro | .................... | .................... | ..................... | .................... | .................... | .................... |
| 17665 | MT970686.1 \|Severe acute respiratory syndrome coro | .................... | .................... | ..................... | .................... | .................... | .................... |
| 17666 | MT970687.1 \|Severe acute respiratory syndrome coro | .................... | .................... | ..................... | .................... | .................... | .................... |
| 17667 | MT970688.1 \|Severe acute respiratory syndrome coro | .................... | .................... | ..................... | .................... | .................... | .................... |
| 17668 | MT970689.1 \|Severe acute respiratory syndrome coro | .................... | .................... | ..................... | .................... | .................... | .................... |
| 17669 | MT970690.1 \|Severe acute respiratory syndrome coro | .................... | .................... | ..................... | .................... | .................... | .................... |
| 17670 | MT970691.1 \|Severe acute respiratory syndrome coro | .................... | .................... | ..................... | .................... | .................... | .................... |
| 17671 | MT970692.1 \|Severe acute respiratory syndrome coro | .................... | .................... | ..................... | .................... | .................... | .................... |
| 17672 | MT970693.1 \|Severe acute respiratory syndrome coro | .................... | .................... | ..................... | .................... | .................... | .................... |
| 17673 | MT970694.1 \|Severe acute respiratory syndrome coro | .................... | .................... | ..................... | .................... | .................... | .................... |
| 17674 | MT970695.1 \|Severe acute respiratory syndrome coro | .................... | .................... | ..................... | .................... | .................... | .................... |
| 17675 | MT970696.1 \|Severe acute respiratory syndrome coro | .................... | .................... | ..................... | .................... | .................... | .................... |
| 17676 | MT970697.1 \|Severe acute respiratory syndrome coro | .................... | .................... | ..................... | .................... | .................... | .................... |
| 17677 | MT970698.1 \|Severe acute respiratory syndrome coro | .................... | .................... | ..................... | .................... | .................... | .................... |
| 17678 | MT970699.1 \|Severe acute respiratory syndrome coro | .................... | .................... | ..................... | .................... | .................... | .................... |
| 17679 | MT970700.1 \|Severe acute respiratory syndrome coro | .................... | .................... | ..................... | .................... | .................... | .................... |
| 17680 | MT970701.1 \|Severe acute respiratory syndrome coro | .................... | .................... | ..................... | .................... | .................... | .................... |
| 17681 | MT970702.1 \|Severe acute respiratory syndrome coro | .................... | .................... | ..................... | .................... | .................... | .................... |
| 17682 | MT970703.1 \|Severe acute respiratory syndrome coro | .................... | .................... | ..................... | .................... | .................... | .................... |

| | | | | | | | | | |
|---|---|---|---|---|---|---|---|---|---|
| 17683 | MT970704.1 | Severe acute respiratory syndrome coro | ···················· | ···················· | ···················· | ···················· | ···················· | ···················· | |
| 17684 | MT970705.1 | Severe acute respiratory syndrome coro | ···················· | ···················· | ···················· | ···················· | ···················· | ···················· | |
| 17685 | MT970706.1 | Severe acute respiratory syndrome coro | ···················· | ···················· | ···················· | ···················· | ···················· | ···················· | |
| 17686 | MT970707.1 | Severe acute respiratory syndrome coro | ···················· | ···················· | ···················· | ···················· | ···················· | ···················· | |
| 17687 | MT970708.1 | Severe acute respiratory syndrome coro | ···················· | ···················· | ···················· | ···················· | ···················· | ···················· | |
| 17688 | MT970710.1 | Severe acute respiratory syndrome coro | ···················· | ···················· | ···················· | ···················· | ···················· | ···················· | |
| 17689 | MT970711.1 | Severe acute respiratory syndrome coro | ···················· | ···················· | ···················· | ···················· | ···················· | ···················· | |
| 17690 | MT970712.1 | Severe acute respiratory syndrome coro | ···················· | ···················· | ···················· | ···················· | ···················· | ···················· | |
| 17691 | MT970713.1 | Severe acute respiratory syndrome coro | ···················· | ···················· | ···················· | ···················· | ···················· | ···················· | |
| 17692 | MT970714.1 | Severe acute respiratory syndrome coro | ···················· | ···················· | ···················· | ···················· | ···················· | ···················· | |
| 17693 | MT970715.1 | Severe acute respiratory syndrome coro | ···················· | ···················· | ···················· | ···················· | ···················· | ···················· | |
| 17694 | MT970716.1 | Severe acute respiratory syndrome coro | ···················· | ···················· | ···················· | ···················· | ···················· | ···················· | |
| 17695 | MT970717.1 | Severe acute respiratory syndrome coro | ···················· | ···················· | ···················· | ···················· | ···················· | ···················· | |
| 17696 | MT970718.1 | Severe acute respiratory syndrome coro | ···················· | ···················· | ···················· | ···················· | ···················· | ···················· | |
| 17697 | MT970719.1 | Severe acute respiratory syndrome coro | ···················· | ···················· | ···················· | ···················· | ···················· | ···················· | |
| 17698 | MT970720.1 | Severe acute respiratory syndrome coro | ···················· | ···················· | ···················· | ···················· | ···················· | ···················· | |
| 17699 | MT970721.1 | Severe acute respiratory syndrome coro | ···················· | ···················· | ···················· | ···················· | ···················· | ···················· | |
| 17700 | MT970722.1 | Severe acute respiratory syndrome coro | ···················· | ···················· | ···················· | ···················· | ···················· | ···················· | |
| 17701 | MT970723.1 | Severe acute respiratory syndrome coro | ···················· | ···················· | ···················· | ···················· | ···················· | ···················· | |
| 17702 | MT970724.1 | Severe acute respiratory syndrome coro | ···················· | ···················· | ···················· | ···················· | ···················· | ···················· | |
| 17703 | MT970725.1 | Severe acute respiratory syndrome coro | ···················· | ···················· | ···················· | ···················· | ···················· | ···················· | |
| 17704 | MT970726.1 | Severe acute respiratory syndrome coro | ···················· | ···················· | ···················· | ···················· | ···················· | ···················· | |
| 17705 | MT970727.1 | Severe acute respiratory syndrome coro | ···················· | ···················· | ···················· | ···················· | ···················· | ···················· | |
| 17706 | MT970728.1 | Severe acute respiratory syndrome coro | ···················· | ···················· | ···················· | ···················· | ···················· | ···················· | |
| 17707 | MT970729.1 | Severe acute respiratory syndrome coro | ···················· | ···················· | ···················· | ···················· | ···················· | ···················· | |
| 17708 | MT970730.1 | Severe acute respiratory syndrome coro | ···················· | ···················· | ···················· | ···················· | ···················· | ···················· | |
| 17709 | MT970731.1 | Severe acute respiratory syndrome coro | ···················· | ···················· | ···················· | ···················· | ···················· | ···················· | |
| 17710 | MT970732.1 | Severe acute respiratory syndrome coro | ···················· | ···················· | ···················· | ···················· | ···················· | ···················· | |
| 17711 | MT970733.1 | Severe acute respiratory syndrome coro | ···················· | ···················· | ···················· | ···················· | ···················· | ···················· | |
| 17712 | MT970734.1 | Severe acute respiratory syndrome coro | ···················· | ···················· | ···················· | ···················· | ···················· | ···················· | |
| 17713 | MT970735.1 | Severe acute respiratory syndrome coro | ···················· | ···················· | ···················· | ···················· | ···················· | ···················· | |
| 17714 | MT970736.1 | Severe acute respiratory syndrome coro | ···················· | ···················· | ···················· | ···················· | ···················· | ···················· | |
| 17715 | MT970737.1 | Severe acute respiratory syndrome coro | ···················· | ···················· | ···················· | ···················· | ···················· | ···················· | |
| 17716 | MT970738.1 | Severe acute respiratory syndrome coro | ···················· | ···················· | ···················· | ···················· | ···················· | ···················· | |
| 17717 | MT970739.1 | Severe acute respiratory syndrome coro | ···················· | ···················· | ···················· | ···················· | ···················· | ···················· | |
| 17718 | MT970740.1 | Severe acute respiratory syndrome coro | ···················· | ···················· | ···················· | ···················· | ···················· | ···················· | |
| 17719 | MT970741.1 | Severe acute respiratory syndrome coro | ···················· | ···················· | ···················· | ···················· | ···················· | ···················· | |
| 17720 | MT970742.1 | Severe acute respiratory syndrome coro | ···················· | ···················· | ···················· | ···················· | ···················· | ···················· | |
| 17721 | MT970743.1 | Severe acute respiratory syndrome coro | ···················· | ···················· | ···················· | ···················· | ···················· | ···················· | |
| 17722 | MT970744.1 | Severe acute respiratory syndrome coro | ···················· | ···················· | ···················· | ···················· | ···················· | ···················· | |
| 17723 | MT970745.1 | Severe acute respiratory syndrome coro | ···················· | ···················· | ···················· | ···················· | ···················· | ···················· | |
| 17724 | MT970746.1 | Severe acute respiratory syndrome coro | ···················· | ···················· | ···················· | ···················· | ···················· | ···················· | |
| 17725 | MT970747.1 | Severe acute respiratory syndrome coro | ···················· | ···················· | ···················· | ···················· | ···················· | ···················· | |
| 17726 | MT970748.1 | Severe acute respiratory syndrome coro | ···················· | ···················· | ···················· | ···················· | ···················· | ···················· | |
| 17727 | MT970749.1 | Severe acute respiratory syndrome coro | ···················· | ···················· | ···················· | ···················· | ···················· | ···················· | |
| 17728 | MT970750.1 | Severe acute respiratory syndrome coro | ···················· | ···················· | ···················· | ···················· | ···················· | ···················· | |
| 17729 | MT970751.1 | Severe acute respiratory syndrome coro | ···················· | ···················· | ···················· | ···················· | ···················· | ···················· | |
| 17730 | MT970752.1 | Severe acute respiratory syndrome coro | ···················· | ···················· | ···················· | ···················· | ···················· | ···················· | |
| 17731 | MT970753.1 | Severe acute respiratory syndrome coro | ···················· | ···················· | ···················· | ···················· | ···················· | ···················· | |
| 17732 | MT970754.1 | Severe acute respiratory syndrome coro | ···················· | ···················· | ···················· | ···················· | ···················· | ···················· | |
| 17733 | MT970755.1 | Severe acute respiratory syndrome coro | ···················· | ···················· | ···················· | ···················· | ···················· | ···················· | |
| 17734 | MT970756.1 | Severe acute respiratory syndrome coro | ···················· | ···················· | ···················· | ···················· | ···················· | ···················· | |
| 17735 | MT970757.1 | Severe acute respiratory syndrome coro | ···················· | ···················· | ···················· | ···················· | ···················· | ···················· | |
| 17736 | MT970758.1 | Severe acute respiratory syndrome coro | ···················· | ···················· | ···················· | ···················· | ···················· | ···················· | |
| 17737 | MT970759.1 | Severe acute respiratory syndrome coro | ···················· | ···················· | ···················· | ···················· | ···················· | ···················· | |
| 17738 | MT970760.1 | Severe acute respiratory syndrome coro | ···················· | ···················· | ···················· | ···················· | ···················· | ···················· | |
| 17739 | MT970761.1 | Severe acute respiratory syndrome coro | ···················· | ···················· | ···················· | ···················· | ···················· | ···················· | |
| 17740 | MT970762.1 | Severe acute respiratory syndrome coro | ···················· | ···················· | ···················· | ···················· | ···················· | ···················· | |
| 17741 | MT970763.1 | Severe acute respiratory syndrome coro | ···················· | ···················· | ···················· | ···················· | ···················· | ···················· | |
| 17742 | MT970764.1 | Severe acute respiratory syndrome coro | ···················· | ···················· | ···················· | ···················· | ···················· | ···················· | |
| 17743 | MT970765.1 | Severe acute respiratory syndrome coro | ···················· | ···················· | ···················· | ···················· | ···················· | ···················· | |
| 17744 | MT970766.1 | Severe acute respiratory syndrome coro | ···················· | ···················· | ···················· | ···················· | ···················· | ···················· | |
| 17745 | MT970767.1 | Severe acute respiratory syndrome coro | ···················· | ···················· | ···················· | ···················· | ···················· | ···················· | |
| 17746 | MT970768.1 | Severe acute respiratory syndrome coro | ···················· | ···················· | ···················· | ···················· | ···················· | ···················· | |
| 17747 | MT970769.1 | Severe acute respiratory syndrome coro | ···················· | ···················· | ···················· | ···················· | ···················· | ···················· | |
| 17748 | MT970770.1 | Severe acute respiratory syndrome coro | ···················· | ···················· | ···················· | ···················· | ···················· | ···················· | |
| 17749 | MT970771.1 | Severe acute respiratory syndrome coro | ···················· | ···················· | ···················· | ···················· | ···················· | ···················· | |
| 17750 | MT970772.1 | Severe acute respiratory syndrome coro | ···················· | ···················· | ···················· | ···················· | ···················· | ···················· | |
| 17751 | MT970773.1 | Severe acute respiratory syndrome coro | ···················· | ···················· | ···················· | ···················· | ···················· | ···················· | |
| 17752 | MT970774.1 | Severe acute respiratory syndrome coro | ··················T· | ···················· | ···················· | ···················· | ···················· | ···················· | 1 |
| 17753 | MT970775.1 | Severe acute respiratory syndrome coro | ··················T· | ···················· | ···················· | ···················· | ···················· | ···················· | 1 |
| 17754 | MT970776.1 | Severe acute respiratory syndrome coro | ···················· | ···················· | ···················· | ···················· | ···················· | ···················· | |
| 17755 | MT970777.1 | Severe acute respiratory syndrome coro | ···················· | ···················· | ···················· | ···················· | ···················· | ···················· | |
| 17756 | MT970778.1 | Severe acute respiratory syndrome coro | ···················· | ···················· | ···················· | ···················· | ···················· | ···················· | |
| 17757 | MT970779.1 | Severe acute respiratory syndrome coro | ···················· | ···················· | ···················· | ···················· | ···················· | ···················· | |
| 17758 | MT970780.1 | Severe acute respiratory syndrome coro | ···················· | ···················· | ···················· | ···················· | ···················· | ···················· | |
| 17759 | MT970781.1 | Severe acute respiratory syndrome coro | ···················· | ···················· | ···················· | ···················· | ···················· | ···················· | |
| 17760 | MT970782.1 | Severe acute respiratory syndrome coro | ···················· | ···················· | ···················· | ···················· | ···················· | ···················· | |

```
17761 MT970783.1 |Severe acute respiratory syndrome coro    ....................  ....................  ....................  ....................  ....................  ....................
17762 MT970784.1 |Severe acute respiratory syndrome coro    ....................  ....................  ....................  ....................  ....................  ....................
17763 MT970785.1 |Severe acute respiratory syndrome coro    ....................  ....................  ....................  ....................  ....................  ....................
17764 MT970786.1 |Severe acute respiratory syndrome coro    ....................  ....................  ....................  ....................  ....................  ....................
17765 MT970787.1 |Severe acute respiratory syndrome coro    ....................  ....................  ....................  ....................  ....................  ....................
17766 MT970788.1 |Severe acute respiratory syndrome coro    ....................  ....................  ....................  ....................  ....................  ....................
17767 MT970789.1 |Severe acute respiratory syndrome coro    ....................  ....................  ....................  ....................  ....................  ....................
17768 MT970790.1 |Severe acute respiratory syndrome coro    ....................  ....................  ....................  ....................  ....................  ....................
17769 MT970791.1 |Severe acute respiratory syndrome coro    ....................  ....................  ....................  ....................  ....................  ....................
17770 MT970792.1 |Severe acute respiratory syndrome coro    ....................  ....................  ....................  ....................  ....................  ....................
17771 MT970793.1 |Severe acute respiratory syndrome coro    ....................  ....................  ....................  ....................  ....................  ....................
17772 MT970794.1 |Severe acute respiratory syndrome coro    ....................  ....................  ....................  ....................  ....................  ....................
17773 MT970795.1 |Severe acute respiratory syndrome coro    ....................  ....................  ....................  ....................  ....................  ....................
17774 MT970796.1 |Severe acute respiratory syndrome coro    ....................  ....................  ....................  ....................  ....................  ....................
17775 MT970797.1 |Severe acute respiratory syndrome coro    ....................  ....................  ....................  ....................  ....................  ....................
17776 MT970798.1 |Severe acute respiratory syndrome coro    ....................  ....................  ....................  ....................  ....................  ....................
17777 MT970799.1 |Severe acute respiratory syndrome coro    ....................  ....................  ....................  ....................  ....................  ....................
17778 MT970800.1 |Severe acute respiratory syndrome coro    ....................  ....................  ....................  ....................  ....................  ....................
17779 MT970801.1 |Severe acute respiratory syndrome coro    ....................  ....................  ....................  ....................  ....................  ....................
17780 MT970802.1 |Severe acute respiratory syndrome coro    ....................  ....................  ....................  ....................  ....................  ....................
17781 MT970803.1 |Severe acute respiratory syndrome coro    ....................  ....................  ....................  ....................  ....................  ....................
17782 MT970804.1 |Severe acute respiratory syndrome coro    ....................  ....................  ....................  ....................  ....................  ....................
17783 MT970806.1 |Severe acute respiratory syndrome coro    ....................  ....................  ....................  ....................  ....................  ....................
17784 MT970807.1 |Severe acute respiratory syndrome coro    ....................  ....................  ....................  ....................  ....................  ....................
17785 MT970808.1 |Severe acute respiratory syndrome coro    ....................  ....................  ....................  ....................  ....................  ....................
17786 MT970809.1 |Severe acute respiratory syndrome coro    ....................  ....................  ....................  ....................  ....................  ....................
17787 MT970810.1 |Severe acute respiratory syndrome coro    ....................  ....................  ....................  ....................  ....................  ....................
17788 MT970811.1 |Severe acute respiratory syndrome coro    ....................  ....................  ....................  ....................  ....................  ....................
17789 MT970812.1 |Severe acute respiratory syndrome coro    ....................  ....................  ....................  ....................  ....................  ....................
17790 MT970813.1 |Severe acute respiratory syndrome coro    ....................  ....................  ....................  ....................  ....................  ....................
17791 MT970814.1 |Severe acute respiratory syndrome coro    ....................  ....................  ....................  ....................  ....................  ....................
17792 MT970815.1 |Severe acute respiratory syndrome coro    ....................  ....................  ....................  ....................  ....................  ....................
17793 MT970816.1 |Severe acute respiratory syndrome coro    ....................  ....................  ....................  ....................  ....................  ....................
17794 MT970817.1 |Severe acute respiratory syndrome coro    ....................  ....................  ....................  ....................  ....................  ....................
17795 MT970818.1 |Severe acute respiratory syndrome coro    ....................  ....................  ....................  ....................  ....................  ....................
17796 MT970819.1 |Severe acute respiratory syndrome coro    ....................  ....................  ....................  ....................  ....................  ....................
17797 MT970820.1 |Severe acute respiratory syndrome coro    ....................  ....................  ....................  ....................  ....................  ....................
17798 MT970821.1 |Severe acute respiratory syndrome coro    ....................  ....................  ....................  ....................  ....................  ....................
17799 MT970822.1 |Severe acute respiratory syndrome coro    ....................  ....................  ....................  ....................  ....................  ....................
17800 MT970823.1 |Severe acute respiratory syndrome coro    ....................  ....................  ....................  ....................  ....................  ....................
17801 MT970824.1 |Severe acute respiratory syndrome coro    ....................  ....................  ....................  ....................  ....................  ....................
17802 MT970825.1 |Severe acute respiratory syndrome coro    ....................  ....................  ....................  ....................  ....................  ....................
17803 MT970826.1 |Severe acute respiratory syndrome coro    ....................  ....................  ....................  ....................  ....................  ....................
17804 MT970827.1 |Severe acute respiratory syndrome coro    ....................  ....................  ....................  ....................  ....................  ....................
17805 MT970828.1 |Severe acute respiratory syndrome coro    ....................  ....................  ....................  ....................  ....................  ....................
17806 MT970829.1 |Severe acute respiratory syndrome coro    ....................  ....................  ....................  ....................  ....................  ....................
17807 MT970830.1 |Severe acute respiratory syndrome coro    ....................  ....................  ....................  ....................  ....................  ....................
17808 MT970831.1 |Severe acute respiratory syndrome coro    ....................  ....................  ....................  ....................  ....................  ....................
17809 MT970832.1 |Severe acute respiratory syndrome coro    ....................  ....................  ....................  ....................  ....................  ....................
17810 MT970833.1 |Severe acute respiratory syndrome coro    ....................  ....................  ....................  ....................  ....................  ....................
17811 MT970834.1 |Severe acute respiratory syndrome coro    ....................  ....................  ....................  ....................  ....................  ....................
17812 MT970835.1 |Severe acute respiratory syndrome coro    ....................  ....................  ....................  ....................  ....................  ....................
17813 MT970836.1 |Severe acute respiratory syndrome coro    ....................  ....................  ....................  ....................  ....................  ....................
17814 MT970837.1 |Severe acute respiratory syndrome coro    ....................  ....................  ....................  ....................  ....................  ....................
17815 MT970838.1 |Severe acute respiratory syndrome coro    ....................  ....................  ....................  ....................  ....................  ....................
17816 MT970839.1 |Severe acute respiratory syndrome coro    ....................  ....................  ....................  ....................  ....................  ....................
17817 MT970840.1 |Severe acute respiratory syndrome coro    ....................  ....................  ....................  ....................  ....................  ....................
17818 MT970841.1 |Severe acute respiratory syndrome coro    ....................  ....................  ....................  ....................  ....................  ....................
17819 MT970842.1 |Severe acute respiratory syndrome coro    ....................  ....................  ....................  ....................  ....................  ....................
17820 MT970843.1 |Severe acute respiratory syndrome coro    ....................  ....................  ....................  ....................  ....................  ....................
17821 MT970844.1 |Severe acute respiratory syndrome coro    ....................  ....................  ....................  ....................  ....................  ....................
17822 MT970845.1 |Severe acute respiratory syndrome coro    ....................  ....................  ....................  ....................  ....................  ....................
17823 MT970846.1 |Severe acute respiratory syndrome coro    ....................  ....................  ....................  ....................  ....................  ....................
17824 MT970847.1 |Severe acute respiratory syndrome coro    ....................  ....................  ....................  ....................  ....................  ....................
17825 MT970848.1 |Severe acute respiratory syndrome coro    ....................  ....................  ....................  ....................  ....................  ....................
17826 MT970849.1 |Severe acute respiratory syndrome coro    ....................  ....................  ....................  ....................  ....................  ....................
17827 MT970850.1 |Severe acute respiratory syndrome coro    ....................  ....................  ....................  ....................  ....................  ....................
17828 MT970851.1 |Severe acute respiratory syndrome coro    ....................  ....................  ....................  ....................  ....................  ....................
17829 MT970852.1 |Severe acute respiratory syndrome coro    ....................  ....................  ....................  ....................  ....................  ....................
17830 MT970853.1 |Severe acute respiratory syndrome coro    ....................  ....................  ....................  ....................  ....................  ....................
17831 MT970854.1 |Severe acute respiratory syndrome coro    ....................  ....................  ....................  ....................  ....................  ....................
17832 MT970855.1 |Severe acute respiratory syndrome coro    ....................  ....................  ....................  ....................  ....................  ....................
17833 MT970856.1 |Severe acute respiratory syndrome coro    ....................  ....................  ....................  ....................  ....................  ....................
17834 MT970857.1 |Severe acute respiratory syndrome coro    ....................  ....................  ....................  ....................  ....................  ....................
17835 MT970858.1 |Severe acute respiratory syndrome coro    ....................  ....................  ....................  ....................  ....................  ....................
17836 MT970859.1 |Severe acute respiratory syndrome coro    ....................  ....................  ....................  ....................  ....................  ....................
17837 MT970860.1 |Severe acute respiratory syndrome coro    ....................  ....................  ....................  ....................  ....................  ....................
17838 MT970861.1 |Severe acute respiratory syndrome coro    ....................  ....................  ....................  ....................  ....................  ....................
```

```
17839 MT970862.1 |Severe acute respiratory syndrome coro    ....................  ....................  .....................  ....................  ....................  ....................
17840 MT970863.1 |Severe acute respiratory syndrome coro    ....................  ....................  .....................  ....................  ....................  ....................
17841 MT970864.1 |Severe acute respiratory syndrome coro    ....................  ....................  .....................  ....................  ....................  ....................
17842 MT970865.1 |Severe acute respiratory syndrome coro    ....................  ....................  .....................  ....................  ....................  ....................
17843 MT970866.1 |Severe acute respiratory syndrome coro    ....................  ....................  .....................  ....................  ....................  ....................
17844 MT970868.1 |Severe acute respiratory syndrome coro    ....................  ....................  .....................  ....................  ....................  ....................
17845 MT970869.1 |Severe acute respiratory syndrome coro    ....................  ....................  .....................  ....................  ....................  ....................
17846 MT970870.1 |Severe acute respiratory syndrome coro    ....................  ....................  .....................  ....................  ....................  ....................
17847 MT970871.1 |Severe acute respiratory syndrome coro    ....................  ....................  .....................  ....................  ....................  ....................
17848 MT970872.1 |Severe acute respiratory syndrome coro    ....................  ....................  .....................  ....................  ....................  ....................
17849 MT970873.1 |Severe acute respiratory syndrome coro    ....................  ....................  .....................  ....................  ....................  ....................
17850 MT970874.1 |Severe acute respiratory syndrome coro    ....................  ....................  .....................  ....................  ....................  ....................
17851 MT970875.1 |Severe acute respiratory syndrome coro    ....................  ....................  .....................  ....................  ....................  ....................
17852 MT970876.1 |Severe acute respiratory syndrome coro    ....................  ....................  .....................  ....................  ....................  ....................
17853 MT970878.1 |Severe acute respiratory syndrome coro    ....................  ....................  .....................  ....................  ....................  ....................
17854 MT970879.1 |Severe acute respiratory syndrome coro    ....................  ....................  .....................  ....................  ....................  ....................
17855 MT970880.1 |Severe acute respiratory syndrome coro    ....................  ....................  .....................  ....................  ....................  ....................
17856 MT970881.1 |Severe acute respiratory syndrome coro    ....................  ....................  .....................  ....................  ....................  ....................
17857 MT970882.1 |Severe acute respiratory syndrome coro    ....................  ....................  .....................  ....................  ....................  ....................
17858 MT970883.1 |Severe acute respiratory syndrome coro    ....................  ....................  .....................  ....................  ....................  ....................
17859 MT970884.1 |Severe acute respiratory syndrome coro    ....................  ....................  .....................  ....................  ....................  ....................
17860 MT970885.1 |Severe acute respiratory syndrome coro    ....................  ....................  .....................  ....................  ....................  ....................
17861 MT970886.1 |Severe acute respiratory syndrome coro    ....................  ....................  .....................  ....................  ....................  ....................
17862 MT970887.1 |Severe acute respiratory syndrome coro    ....................  ....................  .....................  ....................  ....................  ....................
17863 MT970888.1 |Severe acute respiratory syndrome coro    ....................  ....................  .....................  ....................  ....................  ....................
17864 MT970889.1 |Severe acute respiratory syndrome coro    ....................  ....................  .....................  ....................  ....................  ....................
17865 MT970891.1 |Severe acute respiratory syndrome coro    ....................  ....................  .....................  ....................  ....................  ....................
17866 MT970892.1 |Severe acute respiratory syndrome coro    ....................  ....................  .....................  ....................  ....................  ....................
17867 MT970893.1 |Severe acute respiratory syndrome coro    ....................  ....................  .....................  ....................  ....................  ....................
17868 MT970894.1 |Severe acute respiratory syndrome coro    ....................  ....................  .....................  ....................  ....................  ....................
17869 MT970895.1 |Severe acute respiratory syndrome coro    ....................  ....................  .....................  ....................  ....................  ....................
17870 MT970896.1 |Severe acute respiratory syndrome coro    ....................  ....................  .....................  ....................  ....................  ....................
17871 MT970897.1 |Severe acute respiratory syndrome coro    ....................  ....................  .....................  ....................  ....................  ....................
17872 MT970898.1 |Severe acute respiratory syndrome coro    ....................  ....................  .....................  ....................  ....................  ....................
17873 MT970899.1 |Severe acute respiratory syndrome coro    ....................  ....................  .....................  ....................  ....................  ....................
17874 MT970900.1 |Severe acute respiratory syndrome coro    ....................  ....................  .....................  ....................  ....................  ....................
17875 MT970901.1 |Severe acute respiratory syndrome coro    ....................  ....................  .....................  ....................  ....................  ....................
17876 MT970902.1 |Severe acute respiratory syndrome coro    ....................  ....................  .....................  ....................  ....................  ....................
17877 MT970903.1 |Severe acute respiratory syndrome coro    ....................  ....................  .....................  ....................  ....................  ....................
17878 MT970904.1 |Severe acute respiratory syndrome coro    ....................  ....................  .....................  ....................  ....................  ....................
17879 MT970905.1 |Severe acute respiratory syndrome coro    ....................  ....................  .....................  ....................  ....................  ....................
17880 MT970906.1 |Severe acute respiratory syndrome coro    ....................  ....................  .....................  ....................  ....................  ....................
17881 MT970907.1 |Severe acute respiratory syndrome coro    ....................  ....................  .....................  ....................  ....................  ....................
17882 MT970908.1 |Severe acute respiratory syndrome coro    ....................  ....................  .....................  ....................  ....................  ....................
17883 MT970909.1 |Severe acute respiratory syndrome coro    ....................  ....................  .....................  ....................  ....................  ....................
17884 MT970910.1 |Severe acute respiratory syndrome coro    ....................  ....................  .....................  ....................  ....................  ....................
17885 MT970911.1 |Severe acute respiratory syndrome coro    ....................  ....................  .....................  ....................  ....................  ....................
17886 MT970912.1 |Severe acute respiratory syndrome coro    ....................  ....................  .....................  ....................  ....................  ....................
17887 MT970913.1 |Severe acute respiratory syndrome coro    ....................  ....................  .....................  ....................  ....................  ....................
17888 MT970914.1 |Severe acute respiratory syndrome coro    ....................  ....................  .....................  ....................  ....................  ....................
17889 MT970915.1 |Severe acute respiratory syndrome coro    ....................  ....................  .....................  ....................  ....................  ....................
17890 MT970916.1 |Severe acute respiratory syndrome coro    ....................  ....................  .....................  ....................  ....................  ....................
17891 MT970917.1 |Severe acute respiratory syndrome coro    ....................  ....................  .....................  ....................  ....................  ....................
17892 MT970918.1 |Severe acute respiratory syndrome coro    ....................  ....................  .....................  ....................  ....................  ....................
17893 MT970919.1 |Severe acute respiratory syndrome coro    ....................  ....................  .....................  ....................  ....................  ....................
17894 MT970920.1 |Severe acute respiratory syndrome coro    ....................  ....................  .....................  ....................  ....................  ....................
17895 MT970921.1 |Severe acute respiratory syndrome coro    ....................  ....................  .....................  ....................  ....................  ....................
17896 MT970922.1 |Severe acute respiratory syndrome coro    ....................  ....................  .....................  ....................  ....................  ....................
17897 MT970923.1 |Severe acute respiratory syndrome coro    ....................  ....................  .....................  ....................  ....................  ....................
17898 MT970924.1 |Severe acute respiratory syndrome coro    ....................  ....................  .....................  ....................  ....................  ....................
17899 MT970925.1 |Severe acute respiratory syndrome coro    ....................  ....................  .....................  ....................  ....................  ....................
17900 MT970926.1 |Severe acute respiratory syndrome coro    ....................  ....................  .....................  ....................  ....................  ....................
17901 MT970927.1 |Severe acute respiratory syndrome coro    ....................  ....................  .....................  ....................  ....................  ....................
17902 MT970928.1 |Severe acute respiratory syndrome coro    ....................  ....................  .....................  ....................  ....................  ....................
17903 MT970929.1 |Severe acute respiratory syndrome coro    ....................  ....................  .....................  ....................  ....................  ....................
17904 MT970930.1 |Severe acute respiratory syndrome coro    ....................  ....................  .....................  ....................  ....................  ....................
17905 MT970931.1 |Severe acute respiratory syndrome coro    ....................  ....................  .....................  ....................  ....................  ....................
17906 MT970932.1 |Severe acute respiratory syndrome coro    ....................  ....................  .....................  ....................  ....................  ....................
17907 MT970934.1 |Severe acute respiratory syndrome coro    ....................  ....................  .....................  ....................  ....................  ....................
17908 MT970935.1 |Severe acute respiratory syndrome coro    ....................  ....................  .....................  ....................  ....................  ....................
17909 MT970936.1 |Severe acute respiratory syndrome coro    ....................  ....................  .....................  ....................  ....................  ....................
17910 MT970937.1 |Severe acute respiratory syndrome coro    ....................  ....................  .....................  ....................  ....................  ....................
17911 MT970938.1 |Severe acute respiratory syndrome coro    ....................  ....................  .....................  ....................  ....................  ....................
17912 MT970939.1 |Severe acute respiratory syndrome coro    ....................  ....................  .....................  ....................  ....................  ....................
17913 MT970940.1 |Severe acute respiratory syndrome coro    ....................  ....................  .....................  ....................  ....................  ....................
17914 MT970941.1 |Severe acute respiratory syndrome coro    ....................  ....................  .....................  ....................  ....................  ....................
17915 MT970942.1 |Severe acute respiratory syndrome coro    ....................  ....................  .....................  ....................  ....................  ....................
17916 MT970943.1 |Severe acute respiratory syndrome coro    ....................  ....................  .....................  ....................  ....................  ....................
```

```
17917 MT970944.1 |Severe acute respiratory syndrome coro    ....................  ....................  ....................  ....................  ....................  ....................
17918 MT970946.1 |Severe acute respiratory syndrome coro    ....................  ....................  ....................  ....................  ....................  ....................
17919 MT970947.1 |Severe acute respiratory syndrome coro    ....................  ....................  ....................  ....................  ....................  ....................
17920 MT970948.1 |Severe acute respiratory syndrome coro    ....................  ....................  ....................  ....................  ....................  ....................
17921 MT970949.1 |Severe acute respiratory syndrome coro    ....................  ....................  ....................  ....................  ....................  ....................
17922 MT970950.1 |Severe acute respiratory syndrome coro    ....................  ....................  ....................  ....................  ....................  ....................
17923 MT970951.1 |Severe acute respiratory syndrome coro    ....................  ....................  ....................  ....................  ....................  ....................
17924 MT970952.1 |Severe acute respiratory syndrome coro    ....................  ....................  ....................  ....................  ....................  ....................
17925 MT970953.1 |Severe acute respiratory syndrome coro    ....................  ....................  ....................  ....................  ....................  ....................
17926 MT970954.1 |Severe acute respiratory syndrome coro    ....................  ....................  ....................  ....................  ....................  ....................
17927 MT970955.1 |Severe acute respiratory syndrome coro    ....................  ....................  ....................  ....................  ....................  ....................
17928 MT970956.1 |Severe acute respiratory syndrome coro    ....................  ....................  ....................  ....................  ....................  ....................
17929 MT970957.1 |Severe acute respiratory syndrome coro    ....................  ....................  ....................  ....................  ....................  ....................
17930 MT970958.1 |Severe acute respiratory syndrome coro    ....................  ....................  ....................  ....................  ....................  ....................
17931 MT970959.1 |Severe acute respiratory syndrome coro    ....................  ....................  ....................  ....................  ....................  ....................
17932 MT970960.1 |Severe acute respiratory syndrome coro    ....................  ....................  ....................  ....................  ....................  ....................
17933 MT970961.1 |Severe acute respiratory syndrome coro    ....................  ....................  ....................  ....................  ....................  ....................
17934 MT970962.1 |Severe acute respiratory syndrome coro    ....................  ....................  ....................  ....................  ....................  ....................
17935 MT970963.1 |Severe acute respiratory syndrome coro    ....................  ....................  ....................  ....................  ....................  ....................
17936 MT970964.1 |Severe acute respiratory syndrome coro    ....................  ....................  ....................  ....................  ....................  ....................
17937 MT970965.1 |Severe acute respiratory syndrome coro    ....................  ....................  ....................  ....................  ....................  ....................
17938 MT970966.1 |Severe acute respiratory syndrome coro    ....................  ....................  ....................  ....................  ....................  ....................
17939 MT970967.1 |Severe acute respiratory syndrome coro    ....................  ....................  ....................  ....................  ....................  ....................
17940 MT970968.1 |Severe acute respiratory syndrome coro    ....................  ....................  ....................  ....................  ....................  ....................
17941 MT970969.1 |Severe acute respiratory syndrome coro    ....................  ....................  ....................  ....................  ....................  ....................
17942 MT970970.1 |Severe acute respiratory syndrome coro    ....................  ....................  ....................  ....................  ....................  ....................
17943 MT970971.1 |Severe acute respiratory syndrome coro    ....................  ....................  ....................  ....................  ....................  ....................
17944 MT970972.1 |Severe acute respiratory syndrome coro    ....................  ....................  ....................  ....................  ....................  ....................
17945 MT970973.1 |Severe acute respiratory syndrome coro    ....................  ....................  ....................  ....................  ....................  ....................
17946 MT970974.1 |Severe acute respiratory syndrome coro    ....................  ....................  ....................  ....................  ....................  ....................
17947 MT970975.1 |Severe acute respiratory syndrome coro    ....................  ....................  ....................  ....................  ....................  ....................
17948 MT970976.1 |Severe acute respiratory syndrome coro    ....................  ....................  ....................  ....................  ....................  ....................
17949 MT970977.1 |Severe acute respiratory syndrome coro    ....................  ....................  ....................  ....................  ....................  ....................
17950 MT970978.1 |Severe acute respiratory syndrome coro    ....................  ....................  ....................  ....................  ....................  ....................
17951 MT970979.1 |Severe acute respiratory syndrome coro    ....................  ....................  ....................  ....................  ....................  ....................
17952 MT970980.1 |Severe acute respiratory syndrome coro    ....................  ....................  ....................  ....................  ....................  ....................
17953 MT970981.1 |Severe acute respiratory syndrome coro    ....................  ....................  ....................  ....................  ....................  ....................
17954 MT970983.1 |Severe acute respiratory syndrome coro    ....................  ....................  ....................  ....................  ....................  ....................
17955 MT970984.1 |Severe acute respiratory syndrome coro    ....................  ....................  ....................  ....................  ....................  ....................
17956 MT970985.1 |Severe acute respiratory syndrome coro    ....................  ....................  ....................  ....................  ....................  ....................
17957 MT970986.1 |Severe acute respiratory syndrome coro    ....................  ....................  ....................  ....................  ....................  ....................
17958 MT970987.1 |Severe acute respiratory syndrome coro    ....................  ....................  ....................  ....................  ....................  ....................
17959 MT970988.1 |Severe acute respiratory syndrome coro    ....................  ....................  ....................  ....................  ....................  ....................
17960 MT970989.1 |Severe acute respiratory syndrome coro    ....................  ....................  ....................  ....................  ....................  ....................
17961 MT970990.1 |Severe acute respiratory syndrome coro    ....................  ....................  ....................  ....................  ....................  ....................
17962 MT970991.1 |Severe acute respiratory syndrome coro    ....................  ....................  ....................  ....................  ....................  ....................
17963 MT970992.1 |Severe acute respiratory syndrome coro    ....................  ....................  ....................  ....................  ....................  ....................
17964 MT970993.1 |Severe acute respiratory syndrome coro    ....................  ....................  ....................  ....................  ....................  ....................
17965 MT970994.1 |Severe acute respiratory syndrome coro    ....................  ....................  ....................  ....................  ....................  ....................
17966 MT970995.1 |Severe acute respiratory syndrome coro    ....................  ....................  ....................  ....................  ....................  ....................
17967 MT970996.1 |Severe acute respiratory syndrome coro    ....................  ....................  ....................  ....................  ....................  ....................
17968 MT970997.1 |Severe acute respiratory syndrome coro    ....................  ....................  ....................  ....................  ....................  ....................
17969 MT970998.1 |Severe acute respiratory syndrome coro    ....................  ....................  ....................  ....................  ....................  ....................
17970 MT970999.1 |Severe acute respiratory syndrome coro    ....................  ....................  ....................  ....................  ....................  ....................
17971 MT971000.1 |Severe acute respiratory syndrome coro    ....................  ....................  ....................  ....................  ....................  ....................
17972 MT971001.1 |Severe acute respiratory syndrome coro    ....................  ....................  ....................  ....................  ....................  ....................
17973 MT971002.1 |Severe acute respiratory syndrome coro    ....................  ....................  ....................  ....................  ....................  ....................
17974 MT971004.1 |Severe acute respiratory syndrome coro    ....................  ....................  ....................  ....................  ....................  ....................
17975 MT971005.1 |Severe acute respiratory syndrome coro    ....................  ....................  ....................  ....................  ....................  ....................
17976 MT971006.1 |Severe acute respiratory syndrome coro    ....................  ....................  ....................  ....................  ....................  ....................
17977 MT971007.1 |Severe acute respiratory syndrome coro    ....................  ....................  ....................  ....................  ....................  ....................
17978 MT971008.1 |Severe acute respiratory syndrome coro    ....................  ....................  ....................  ....................  ....................  ....................
17979 MT971010.1 |Severe acute respiratory syndrome coro    ....................  ....................  ....................  ....................  ....................  ....................
17980 MT971011.1 |Severe acute respiratory syndrome coro    ....................  ....................  ....................  ....................  ....................  ....................
17981 MT971012.1 |Severe acute respiratory syndrome coro    ....................  ....................  ....................  ....................  ....................  ....................
17982 MT971013.1 |Severe acute respiratory syndrome coro    ....................  ....................  ....................  ....................  ....................  ....................
17983 MT971014.1 |Severe acute respiratory syndrome coro    ....................  ....................  ....................  ....................  ....................  ....................
17984 MT971015.1 |Severe acute respiratory syndrome coro    ....................  ....................  ....................  ....................  ....................  ....................
17985 MT971016.1 |Severe acute respiratory syndrome coro    ....................  ....................  ....................  ....................  ....................  ....................
17986 MT971017.1 |Severe acute respiratory syndrome coro    ....................  ....................  ....................  ....................  ....................  ....................
17987 MT971018.1 |Severe acute respiratory syndrome coro    ....................  ....................  ....................  ....................  ....................  ....................
17988 MT971019.1 |Severe acute respiratory syndrome coro    ....................  ....................  ....................  ....................  ....................  ....................
17989 MT971020.1 |Severe acute respiratory syndrome coro    ....................  ....................  ....................  ....................  ....................  ....................
17990 MT971021.1 |Severe acute respiratory syndrome coro    ....................  ....................  ....................  ....................  ....................  ....................
17991 MT971022.1 |Severe acute respiratory syndrome coro    ....................  ....................  ....................  ....................  ....................  ....................
17992 MT971023.1 |Severe acute respiratory syndrome coro    ....................  ....................  ....................  ....................  ....................  ....................
17993 MT971024.1 |Severe acute respiratory syndrome coro    ....................  ....................  ....................  ....................  ....................  ....................
17994 MT971025.1 |Severe acute respiratory syndrome coro    ....................  ....................  ....................  ....................  ....................  ....................
```

| | | | | | | | | |
|---|---|---|---|---|---|---|---|---|
| 17995 | MT971026.1 | \|Severe acute respiratory syndrome coro | .................... | .................... | ..................... | .................... | .................... | .................... |
| 17996 | MT971027.1 | \|Severe acute respiratory syndrome coro | .................... | .................... | ..................... | .................... | .................... | .................... |
| 17997 | MT971028.1 | \|Severe acute respiratory syndrome coro | .................... | .................... | ..................... | .................... | .................... | .................... |
| 17998 | MT971029.1 | \|Severe acute respiratory syndrome coro | .................... | .................... | ..................... | .................... | .................... | .................... |
| 17999 | MT971030.1 | \|Severe acute respiratory syndrome coro | .................... | .................... | ..................... | .................... | .................... | .................... |
| 18000 | MT971031.1 | \|Severe acute respiratory syndrome coro | .................... | .................... | ..................... | .................... | .................... | .................... |
| 18001 | MT971032.1 | \|Severe acute respiratory syndrome coro | .................... | .................... | ..................... | .................... | .................... | .................... |
| 18002 | MT971033.1 | \|Severe acute respiratory syndrome coro | .................... | .................... | ..................... | .................... | .................... | .................... |
| 18003 | MT971034.1 | \|Severe acute respiratory syndrome coro | .................... | .................... | ..................... | .................... | .................... | .................... |
| 18004 | MT971035.1 | \|Severe acute respiratory syndrome coro | .................... | .................... | ..................... | .................... | .................... | .................... |
| 18005 | MT971036.1 | \|Severe acute respiratory syndrome coro | .................... | .................... | ..................... | .................... | .................... | .................... |
| 18006 | MT971037.1 | \|Severe acute respiratory syndrome coro | .................... | .................... | ..................... | .................... | .................... | .................... |
| 18007 | MT971038.1 | \|Severe acute respiratory syndrome coro | .................... | .................... | ..................... | .................... | .................... | .................... |
| 18008 | MT971039.1 | \|Severe acute respiratory syndrome coro | .................... | .................... | ..................... | .................... | .................... | .................... |
| 18009 | MT971040.1 | \|Severe acute respiratory syndrome coro | .................... | .................... | ..................... | .................... | .................... | .................... |
| 18010 | MT971041.1 | \|Severe acute respiratory syndrome coro | .................... | .................... | ..................... | .................... | .................... | .................... |
| 18011 | MT971042.1 | \|Severe acute respiratory syndrome coro | .................... | .................... | ..................... | .................... | .................... | .................... |
| 18012 | MT971043.1 | \|Severe acute respiratory syndrome coro | .................... | .................... | ..................... | .................... | .................... | .................... |
| 18013 | MT971044.1 | \|Severe acute respiratory syndrome coro | .................... | .................... | ..................... | .................... | .................... | .................... |
| 18014 | MT971045.1 | \|Severe acute respiratory syndrome coro | .................... | .................... | ..................... | .................... | .................... | .................... |
| 18015 | MT971046.1 | \|Severe acute respiratory syndrome coro | .................... | .................... | ..................... | .................... | .................... | .................... |
| 18016 | MT971047.1 | \|Severe acute respiratory syndrome coro | .................... | .................... | ..................... | .................... | .................... | .................... |
| 18017 | MT971048.1 | \|Severe acute respiratory syndrome coro | .................... | .................... | ..................... | .................... | .................... | .................... |
| 18018 | MT971049.1 | \|Severe acute respiratory syndrome coro | .................... | .................... | ..................... | .................... | .................... | .................... |
| 18019 | MT971050.1 | \|Severe acute respiratory syndrome coro | .................... | .................... | ..................... | .................... | .................... | .................... |
| 18020 | MT971051.1 | \|Severe acute respiratory syndrome coro | .................... | .................... | ..................... | .................... | .................... | .................... |
| 18021 | MT971052.1 | \|Severe acute respiratory syndrome coro | .................... | .................... | ..................... | .................... | .................... | .................... |
| 18022 | MT971053.1 | \|Severe acute respiratory syndrome coro | .................... | .................... | ..................... | .................... | .................... | .................... |
| 18023 | MT971054.1 | \|Severe acute respiratory syndrome coro | .................... | .................... | ..................... | .................... | .................... | .................... |
| 18024 | MT971055.1 | \|Severe acute respiratory syndrome coro | .................... | .................... | ..................... | .................... | .................... | .................... |
| 18025 | MT971056.1 | \|Severe acute respiratory syndrome coro | .................... | .................... | ..................... | .................... | .................... | .................... |
| 18026 | MT971057.1 | \|Severe acute respiratory syndrome coro | .................... | .................... | ..................... | .................... | .................... | .................... |
| 18027 | MT971058.1 | \|Severe acute respiratory syndrome coro | .................... | .................... | ..................... | .................... | .................... | .................... |
| 18028 | MT971059.1 | \|Severe acute respiratory syndrome coro | .................... | .................... | ..................... | .................... | .................... | .................... |
| 18029 | MT971060.1 | \|Severe acute respiratory syndrome coro | .................... | .................... | ..................... | .................... | .................... | .................... |
| 18030 | MT971061.1 | \|Severe acute respiratory syndrome coro | .................... | .................... | ..................... | .................... | .................... | .................... |
| 18031 | MT971062.1 | \|Severe acute respiratory syndrome coro | .................... | .................... | ..................... | .................... | .................... | .................... |
| 18032 | MT971063.1 | \|Severe acute respiratory syndrome coro | .................... | .................... | ..................... | .................... | .................... | .................... |
| 18033 | MT971064.1 | \|Severe acute respiratory syndrome coro | .................... | .................... | ..................... | .................... | .................... | .................... |
| 18034 | MT971065.1 | \|Severe acute respiratory syndrome coro | .................... | .................... | ..................... | .................... | .................... | .................... |
| 18035 | MT971066.1 | \|Severe acute respiratory syndrome coro | .................... | .................... | ..................... | .................... | .................... | .................... |
| 18036 | MT971067.1 | \|Severe acute respiratory syndrome coro | .................... | .................... | ..................... | .................... | .................... | .................... |
| 18037 | MT971068.1 | \|Severe acute respiratory syndrome coro | .................... | .................... | ..................... | .................... | .................... | .................... |
| 18038 | MT971069.1 | \|Severe acute respiratory syndrome coro | .................... | .................... | ..................... | .................... | .................... | .................... |
| 18039 | MT971070.1 | \|Severe acute respiratory syndrome coro | .................... | .................... | ..................... | .................... | .................... | .................... |
| 18040 | MT971071.1 | \|Severe acute respiratory syndrome coro | .................... | .................... | ..................... | .................... | .................... | .................... |
| 18041 | MT971074.1 | \|Severe acute respiratory syndrome coro | .................... | .................... | ..................... | .................... | .................... | .................... |
| 18042 | MT971075.1 | \|Severe acute respiratory syndrome coro | .................... | .................... | ..................... | .................... | .................... | .................... |
| 18043 | MT971076.1 | \|Severe acute respiratory syndrome coro | .................... | .................... | ..................... | .................... | .................... | .................... |
| 18044 | MT971077.1 | \|Severe acute respiratory syndrome coro | .................... | .................... | ..................... | .................... | .................... | .................... |
| 18045 | MT971078.1 | \|Severe acute respiratory syndrome coro | .................... | .................... | ..................... | .................... | .................... | .................... |
| 18046 | MT971079.1 | \|Severe acute respiratory syndrome coro | .................... | .................... | ..................... | .................... | .................... | .................... |
| 18047 | MT971080.1 | \|Severe acute respiratory syndrome coro | .................... | .................... | ..................... | .................... | .................... | .................... |
| 18048 | MT971081.1 | \|Severe acute respiratory syndrome coro | .................... | .................... | ..................... | .................... | .................... | .................... |
| 18049 | MT971082.1 | \|Severe acute respiratory syndrome coro | .................... | .................... | ..................... | .................... | .................... | .................... |
| 18050 | MT971083.1 | \|Severe acute respiratory syndrome coro | .................... | .................... | ..................... | .................... | .................... | .................... |
| 18051 | MT971084.1 | \|Severe acute respiratory syndrome coro | .................... | .................... | ..................... | .................... | .................... | .................... |
| 18052 | MT971085.1 | \|Severe acute respiratory syndrome coro | .................... | .................... | ..................... | .................... | .................... | .................... |
| 18053 | MT971086.1 | \|Severe acute respiratory syndrome coro | .................... | .................... | ..................... | .................... | .................... | .................... |
| 18054 | MT971087.1 | \|Severe acute respiratory syndrome coro | .................... | .................... | ..................... | .................... | .................... | .................... |
| 18055 | MT971088.1 | \|Severe acute respiratory syndrome coro | .................... | .................... | ..................... | .................... | .................... | .................... |
| 18056 | MT971089.1 | \|Severe acute respiratory syndrome coro | .................... | .................... | ..................... | .................... | .................... | .................... |
| 18057 | MT971090.1 | \|Severe acute respiratory syndrome coro | .................... | .................... | ..................... | .................... | .................... | .................... |
| 18058 | MT971091.1 | \|Severe acute respiratory syndrome coro | .................... | .................... | ..................... | .................... | .................... | .................... |
| 18059 | MT971092.1 | \|Severe acute respiratory syndrome coro | .................... | .................... | ..................... | .................... | .................... | .................... |
| 18060 | MT971093.1 | \|Severe acute respiratory syndrome coro | .................... | .................... | ..................... | .................... | .................... | .................... |
| 18061 | MT971094.1 | \|Severe acute respiratory syndrome coro | .................... | .................... | ..................... | .................... | .................... | .................... |
| 18062 | MT971095.1 | \|Severe acute respiratory syndrome coro | .................... | .................... | ..................... | .................... | .................... | .................... |
| 18063 | MT971096.1 | \|Severe acute respiratory syndrome coro | .................... | .................... | ..................... | .................... | .................... | .................... |
| 18064 | MT971097.1 | \|Severe acute respiratory syndrome coro | .................... | .................... | ..................... | .................... | .................... | .................... |
| 18065 | MT971098.1 | \|Severe acute respiratory syndrome coro | .................... | .................... | ..................... | .................... | .................... | .................... |
| 18066 | MT971099.1 | \|Severe acute respiratory syndrome coro | .................... | .................... | ..................... | .................... | .................... | .................... |
| 18067 | MT971100.1 | \|Severe acute respiratory syndrome coro | .................... | .................... | ..................... | .................... | .................... | .................... |
| 18068 | MT971101.1 | \|Severe acute respiratory syndrome coro | .................... | .................... | ..................... | .................... | .................... | .................... |
| 18069 | MT971102.1 | \|Severe acute respiratory syndrome coro | .................... | .................... | ..................... | .................... | .................... | .................... |
| 18070 | MT971103.1 | \|Severe acute respiratory syndrome coro | .................... | .................... | ..................... | .................... | .................... | .................... |
| 18071 | MT971104.1 | \|Severe acute respiratory syndrome coro | .................... | .................... | ..................... | .................... | .................... | .................... |
| 18072 | MT971105.1 | \|Severe acute respiratory syndrome coro | .................... | .................... | ..................... | .................... | .................... | .................... |

18073 MT971106.1 |Severe acute respiratory syndrome coro .................... .................... ..................... .................... .................... ....................
18074 MT971107.1 |Severe acute respiratory syndrome coro .................... .................... ..................... .................... .................... ....................
18075 MT971108.1 |Severe acute respiratory syndrome coro .................... .................... ..................... .................... .................... ....................
18076 MT971109.1 |Severe acute respiratory syndrome coro .................... .................... ..................... .................... .................... ....................
18077 MT971110.1 |Severe acute respiratory syndrome coro .................... .................... ..................... .................... .................... ....................
18078 MT971111.1 |Severe acute respiratory syndrome coro .................... .................... ..................... .................... .................... ....................
18079 MT971112.1 |Severe acute respiratory syndrome coro .................... .................... ..................... .................... .................... ....................
18080 MT971113.1 |Severe acute respiratory syndrome coro .................... .................... ..................... .................... .................... ....................
18081 MT971114.1 |Severe acute respiratory syndrome coro .................... .................... ..................... .................... .................... ....................
18082 MT971115.1 |Severe acute respiratory syndrome coro .................... .................... ..................... .................... .................... ....................
18083 MT971116.1 |Severe acute respiratory syndrome coro .................... .................... ..................... .................... .................... ....................
18084 MT971117.1 |Severe acute respiratory syndrome coro .................... .................... ..................... .................... .................... ....................
18085 MT971118.1 |Severe acute respiratory syndrome coro .................... .................... ..................... .................... .................... ....................
18086 MT971119.1 |Severe acute respiratory syndrome coro .................... .................... ..................... .................... .................... ....................
18087 MT971120.1 |Severe acute respiratory syndrome coro .................... .................... ..................... .................... .................... ....................
18088 MT971121.1 |Severe acute respiratory syndrome coro .................... .................... ..................... .................... .................... ....................
18089 MT971122.1 |Severe acute respiratory syndrome coro .................... .................... ..................... .................... .................... ....................
18090 MT971123.1 |Severe acute respiratory syndrome coro .................... .................... ..................... .................... .................... ....................
18091 MT971124.1 |Severe acute respiratory syndrome coro .................... .................... ..................... .................... .................... ....................
18092 MT971125.1 |Severe acute respiratory syndrome coro .................... .................... ..................... .................... .................... ....................
18093 MT971126.1 |Severe acute respiratory syndrome coro .................... .................... ..................... .................... .................... ....................
18094 MT971127.1 |Severe acute respiratory syndrome coro .................... .................... ..................... .................... .................... ....................
18095 MT971128.1 |Severe acute respiratory syndrome coro .................... .................... ..................... .................... .................... ....................
18096 MT971129.1 |Severe acute respiratory syndrome coro .................... .................... ..................... .................... .................... ....................
18097 MT971130.1 |Severe acute respiratory syndrome coro .................... .................... ..................... .................... .................... ....................
18098 MT971131.1 |Severe acute respiratory syndrome coro .................... .................... ..................... .................... .................... ....................
18099 MT971132.1 |Severe acute respiratory syndrome coro .................... .................... ..................... .................... .................... ....................
18100 MT971133.1 |Severe acute respiratory syndrome coro .................... .................... ..................... .................... .................... ....................
18101 MT971134.1 |Severe acute respiratory syndrome coro .................... .................... ..................... .................... .................... ....................
18102 MT971135.1 |Severe acute respiratory syndrome coro .................... .................... ..................... .................... .................... ....................
18103 MT971136.1 |Severe acute respiratory syndrome coro .................... .................... ..................... .................... .................... ....................
18104 MT971137.1 |Severe acute respiratory syndrome coro .................... .................... ..................... .................... .................... ....................
18105 MT971138.1 |Severe acute respiratory syndrome coro .................... .................... ..................... .................... .................... ....................
18106 MT971139.1 |Severe acute respiratory syndrome coro .................... .................... ..................... .................... .................... ....................
18107 MT971140.1 |Severe acute respiratory syndrome coro .................... .................... ..................... .................... .................... ....................
18108 MT971141.1 |Severe acute respiratory syndrome coro .................... .................... ..................... .................... .................... ....................
18109 MT971142.1 |Severe acute respiratory syndrome coro .................... .................... ..................... .................... .................... ....................
18110 MT971143.1 |Severe acute respiratory syndrome coro .................... .................... ..................... .................... .................... ....................
18111 MT971144.1 |Severe acute respiratory syndrome coro .................... .................... ..................... .................... .................... ....................
18112 MT971146.1 |Severe acute respiratory syndrome coro .................... .................... ..................... .................... .................... ....................
18113 MT971147.1 |Severe acute respiratory syndrome coro .................... .................... ..................... .................... .................... ....................
18114 MT971148.1 |Severe acute respiratory syndrome coro .................... .................... ..................... .................... .................... ....................
18115 MT971149.1 |Severe acute respiratory syndrome coro .................... .................... ..................... .................... .................... ....................
18116 MT971150.1 |Severe acute respiratory syndrome coro .................... .................... ..................... .................... .................... ....................
18117 MT971151.1 |Severe acute respiratory syndrome coro .................... .................... ..................... .................... .................... ....................
18118 MT971152.1 |Severe acute respiratory syndrome coro .................... .................... ..................... .................... .................... ....................
18119 MT971153.1 |Severe acute respiratory syndrome coro .................... .................... ..................... .................... .................... ....................
18120 MT971155.1 |Severe acute respiratory syndrome coro .................... .................... ..................... .................... .................... ....................
18121 MT971156.1 |Severe acute respiratory syndrome coro .................... .................... ..................... .................... .................... ....................
18122 MT971157.1 |Severe acute respiratory syndrome coro .................... .................... ..................... .................... .................... ....................
18123 MT971158.1 |Severe acute respiratory syndrome coro .................... .................... ..................... .................... .................... ....................
18124 MT971159.1 |Severe acute respiratory syndrome coro .................... .................... ..................... .................... .................... ....................
18125 MT971160.1 |Severe acute respiratory syndrome coro .................... .................... ..................... .................... .................... ....................
18126 MT971161.1 |Severe acute respiratory syndrome coro .................... .................... ..................... .................... .................... ....................
18127 MT971162.1 |Severe acute respiratory syndrome coro .................... .................... ..................... .................... .................... ....................
18128 MT971163.1 |Severe acute respiratory syndrome coro .................... .................... ..................... .................... .................... ....................
18129 MT971164.1 |Severe acute respiratory syndrome coro .................... .................... ..................... .................... .................... ....................
18130 MT971165.1 |Severe acute respiratory syndrome coro .................... .................... ..................... .................... .................... ....................
18131 MT971166.1 |Severe acute respiratory syndrome coro .................... .................... ..................... .................... .................... ....................
18132 MT971167.1 |Severe acute respiratory syndrome coro .................... .................... ..................... .................... .................... ....................
18133 MT971168.1 |Severe acute respiratory syndrome coro .................... .................... ..................... .................... .................... ....................
18134 MT971169.1 |Severe acute respiratory syndrome coro .................... .................... ..................... .................... .................... ....................
18135 MT971170.1 |Severe acute respiratory syndrome coro .................... .................... ..................... .................... .................... ....................
18136 MT971171.1 |Severe acute respiratory syndrome coro .................... .................... ..................... .................... .................... ....................
18137 MT971172.1 |Severe acute respiratory syndrome coro .................... .................... ..................... .................... .................... ....................
18138 MT971173.1 |Severe acute respiratory syndrome coro .................... .................... ..................... .................... .................... ....................
18139 MT971174.1 |Severe acute respiratory syndrome coro .................... .................... ..................... .................... .................... ....................
18140 MT971175.1 |Severe acute respiratory syndrome coro .................... .................... ..................... .................... .................... ....................
18141 MT971176.1 |Severe acute respiratory syndrome coro .................... .................... ..................... .................... .................... ....................
18142 MT971177.1 |Severe acute respiratory syndrome coro .................... .................... ..................... .................... .................... ....................
18143 MT971178.1 |Severe acute respiratory syndrome coro .................... .................... ..................... .................... .................... ....................
18144 MT971179.1 |Severe acute respiratory syndrome coro .................... .................... ..................... .................... .................... ....................
18145 MT971180.1 |Severe acute respiratory syndrome coro .................... .................... ..................... .................... .................... ....................
18146 MT971181.1 |Severe acute respiratory syndrome coro .................... .................... ..................... .................... .................... ....................
18147 MT971182.1 |Severe acute respiratory syndrome coro .................... .................... ..................... .................... .................... ....................
18148 MT971183.1 |Severe acute respiratory syndrome coro .................... .................... ..................... .................... .................... ....................
18149 MT971184.1 |Severe acute respiratory syndrome coro .................... .................... ..................... .................... .................... ....................
18150 MT971185.1 |Severe acute respiratory syndrome coro .................... .................... ..................... .................... .................... ....................

```
18151 MT971186.1 |Severe acute respiratory syndrome coro    ....................  ....................  ....................  ....................  ....................  ....................
18152 MT971187.1 |Severe acute respiratory syndrome coro    ....................  ....................  ....................  ....................  ....................  ....................
18153 MT971188.1 |Severe acute respiratory syndrome coro    ....................  ....................  ....................  ....................  ....................  ....................
18154 MT971189.1 |Severe acute respiratory syndrome coro    ....................  ....................  ....................  ....................  ....................  ....................
18155 MT971190.1 |Severe acute respiratory syndrome coro    ....................  ....................  ....................  ....................  ....................  ....................
18156 MT971191.1 |Severe acute respiratory syndrome coro    ....................  ....................  ....................  ....................  ....................  ....................
18157 MT971192.1 |Severe acute respiratory syndrome coro    ....................  ....................  ....................  ....................  ....................  ....................
18158 MT971193.1 |Severe acute respiratory syndrome coro    ....................  ....................  ....................  ....................  ....................  ....................
18159 MT971194.1 |Severe acute respiratory syndrome coro    ....................  ....................  ....................  ....................  ....................  ....................
18160 MT971195.1 |Severe acute respiratory syndrome coro    ....................  ....................  ....................  ....................  ....................  ....................
18161 MT971196.1 |Severe acute respiratory syndrome coro    ....................  ....................  ....................  ....................  ....................  ....................
18162 MT971197.1 |Severe acute respiratory syndrome coro    ....................  ....................  ....................  ....................  ....................  ....................
18163 MT971198.1 |Severe acute respiratory syndrome coro    ....................  ....................  ....................  ....................  ....................  ....................
18164 MT971199.1 |Severe acute respiratory syndrome coro    ....................  ....................  ....................  ....................  ....................  ....................
18165 MT971200.1 |Severe acute respiratory syndrome coro    ....................  ....................  ....................  ....................  ....................  ....................
18166 MT971201.1 |Severe acute respiratory syndrome coro    ....................  ....................  ....................  ....................  ....................  ....................
18167 MT971202.1 |Severe acute respiratory syndrome coro    ....................  ....................  ....................  ....................  ....................  ....................
18168 MT971203.1 |Severe acute respiratory syndrome coro    ....................  ....................  ....................  ....................  ....................  ....................
18169 MT971204.1 |Severe acute respiratory syndrome coro    ....................  ....................  ....................  ....................  ....................  ....................
18170 MT971205.1 |Severe acute respiratory syndrome coro    ....................  ....................  ....................  ....................  ....................  ....................
18171 MT971206.1 |Severe acute respiratory syndrome coro    ....................  ....................  ....................  ....................  ....................  ....................
18172 MT971207.1 |Severe acute respiratory syndrome coro    ....................  ....................  ....................  ....................  ....................  ....................
18173 MT971208.1 |Severe acute respiratory syndrome coro    ....................  ....................  ....................  ....................  ....................  ....................
18174 MT971209.1 |Severe acute respiratory syndrome coro    ....................  ....................  ....................  ....................  ....................  ....................
18175 MT971210.1 |Severe acute respiratory syndrome coro    ....................  ....................  ....................  ....................  ....................  ....................
18176 MT971211.1 |Severe acute respiratory syndrome coro    ....................  ....................  ....................  ....................  ....................  ....................
18177 MT971212.1 |Severe acute respiratory syndrome coro    ....................  ....................  ....................  ....................  ....................  ....................
18178 MT971213.1 |Severe acute respiratory syndrome coro    ....................  ....................  ....................  ....................  ....................  ....................
18179 MT971214.1 |Severe acute respiratory syndrome coro    ....................  ....................  ....................  ....................  ....................  ....................
18180 MT971215.1 |Severe acute respiratory syndrome coro    ....................  ....................  ....................  ....................  ....................  ....................
18181 MT971216.1 |Severe acute respiratory syndrome coro    ....................  ....................  ....................  ....................  ....................  ....................
18182 MT971217.1 |Severe acute respiratory syndrome coro    ....................  ....................  ....................  ....................  ....................  ....................
18183 MT971218.1 |Severe acute respiratory syndrome coro    ....................  ....................  ....................  ....................  ....................  ....................
18184 MT971219.1 |Severe acute respiratory syndrome coro    ....................  ....................  ....................  ....................  ....................  ....................
18185 MT971220.1 |Severe acute respiratory syndrome coro    ....................  ....................  ....................  ....................  ....................  ....................
18186 MT971221.1 |Severe acute respiratory syndrome coro    ....................  ....................  ....................  ....................  ....................  ....................
18187 MT971222.1 |Severe acute respiratory syndrome coro    ....................  ....................  ....................  ....................  ....................  ....................
18188 MT971223.1 |Severe acute respiratory syndrome coro    ....................  ....................  ....................  ....................  ....................  ....................
18189 MT971224.1 |Severe acute respiratory syndrome coro    ....................  ....................  ....................  ....................  ....................  ....................
18190 MT971225.1 |Severe acute respiratory syndrome coro    ....................  ....................  ....................  ....................  ....................  ....................
18191 MT971226.1 |Severe acute respiratory syndrome coro    ....................  ....................  ....................  ....................  ....................  ....................
18192 MT971227.1 |Severe acute respiratory syndrome coro    ....................  ....................  ....................  ....................  ....................  ....................
18193 MT971228.1 |Severe acute respiratory syndrome coro    ....................  ....................  ....................  ....................  ....................  ....................
18194 MT971229.1 |Severe acute respiratory syndrome coro    ....................  ....................  ....................  ....................  ....................  ....................
18195 MT971230.1 |Severe acute respiratory syndrome coro    ....................  ....................  ....................  ....................  ....................  ....................
18196 MT971231.1 |Severe acute respiratory syndrome coro    ....................  ....................  ....................  ....................  ....................  ....................
18197 MT971232.1 |Severe acute respiratory syndrome coro    ....................  ....................  ....................  ....................  ....................  ....................
18198 MT971233.1 |Severe acute respiratory syndrome coro    ....................  ....................  ....................  ....................  ....................  ....................
18199 MT971234.1 |Severe acute respiratory syndrome coro    ....................  ....................  ....................  ....................  ....................  ....................
18200 MT971235.1 |Severe acute respiratory syndrome coro    ....................  ....................  ....................  ....................  ....................  ....................
18201 MT971237.1 |Severe acute respiratory syndrome coro    ....................  ....................  ....................  ....................  ....................  ....................
18202 MT971238.1 |Severe acute respiratory syndrome coro    ....................  ....................  ....................  ....................  ....................  ....................
18203 MT971239.1 |Severe acute respiratory syndrome coro    ....................  ....................  ....................  ....................  ....................  ....................
18204 MT971240.1 |Severe acute respiratory syndrome coro    ....................  ....................  ....................  ....................  ....................  ....................
18205 MT971241.1 |Severe acute respiratory syndrome coro    ....................  ....................  ....................  ....................  ....................  ....................
18206 MT971242.1 |Severe acute respiratory syndrome coro    ....................  ....................  ....................  ....................  ....................  ....................
18207 MT971243.1 |Severe acute respiratory syndrome coro    ....................  ....................  ....................  ....................  ....................  ....................
18208 MT971244.1 |Severe acute respiratory syndrome coro    ....................  ....................  ....................  ....................  ....................  ....................
18209 MT971245.1 |Severe acute respiratory syndrome coro    ....................  ....................  ....................  ....................  ....................  ....................
18210 MT971246.1 |Severe acute respiratory syndrome coro    ....................  ....................  ....................  ....................  ....................  ....................
18211 MT971247.1 |Severe acute respiratory syndrome coro    ....................  ....................  ....................  ....................  ....................  ....................
18212 MT971248.1 |Severe acute respiratory syndrome coro    ....................  ....................  ....................  ....................  ....................  ....................
18213 MT971249.1 |Severe acute respiratory syndrome coro    ....................  ....................  ....................  ....................  ....................  ....................
18214 MT971250.1 |Severe acute respiratory syndrome coro    ....................  ....................  ....................  ....................  ....................  ....................
18215 MT971251.1 |Severe acute respiratory syndrome coro    ....................  ....................  ....................  ....................  ....................  ....................
18216 MT971252.1 |Severe acute respiratory syndrome coro    ....................  ....................  ....................  ....................  ....................  ....................
18217 MT971254.1 |Severe acute respiratory syndrome coro    ....................  ....................  ....................  ....................  ....................  ....................
18218 MT971255.1 |Severe acute respiratory syndrome coro    ....................  ....................  ....................  ....................  ....................  ....................
18219 MT971256.1 |Severe acute respiratory syndrome coro    ....................  ....................  ....................  ....................  ....................  ....................
18220 MT971257.1 |Severe acute respiratory syndrome coro    ....................  ....................  ....................  ....................  ....................  ....................
18221 MT971258.1 |Severe acute respiratory syndrome coro    ....................  ....................  ....................  ....................  ....................  ....................
18222 MT971259.1 |Severe acute respiratory syndrome coro    ....................  ....................  ....................  ....................  ....................  ....................
18223 MT971260.1 |Severe acute respiratory syndrome coro    ....................  ....................  ....................  ....................  ....................  ....................
18224 MT971261.1 |Severe acute respiratory syndrome coro    ....................  ....................  ....................  ....................  ....................  ....................
18225 MT971262.1 |Severe acute respiratory syndrome coro    ....................  ....................  ....................  ....................  ....................  ....................
18226 MT971263.1 |Severe acute respiratory syndrome coro    ....................  ....................  ....................  ....................  ....................  ....................
18227 MT971264.1 |Severe acute respiratory syndrome coro    ....................  ....................  ....................  ....................  ....................  ....................
18228 MT971265.1 |Severe acute respiratory syndrome coro    ....................  ....................  ....................  ....................  ....................  ....................
```

| | | | | | | | | |
|---|---|---|---|---|---|---|---|---|
| 18229 | MT971266.1 | \|Severe acute respiratory syndrome coro | .................... | .................... | ..................... | .................... | .................... | .................... |
| 18230 | MT971267.1 | \|Severe acute respiratory syndrome coro | .................... | .................... | ..................... | .................... | .................... | .................... |
| 18231 | MT971268.1 | \|Severe acute respiratory syndrome coro | .................... | .................... | ..................... | .................... | .................... | .................... |
| 18232 | MT971269.1 | \|Severe acute respiratory syndrome coro | .................... | .................... | ..................... | .................... | .................... | .................... |
| 18233 | MT971270.1 | \|Severe acute respiratory syndrome coro | .................... | .................... | ..................... | .................... | .................... | .................... |
| 18234 | MT971271.1 | \|Severe acute respiratory syndrome coro | .................... | .................... | ..................... | .................... | .................... | .................... |
| 18235 | MT971272.1 | \|Severe acute respiratory syndrome coro | .................... | .................... | ..................... | .................... | .................... | .................... |
| 18236 | MT971273.1 | \|Severe acute respiratory syndrome coro | .................... | .................... | ..................... | .................... | .................... | .................... |
| 18237 | MT971274.1 | \|Severe acute respiratory syndrome coro | .................... | .................... | ..................... | .................... | .................... | .................... |
| 18238 | MT971275.1 | \|Severe acute respiratory syndrome coro | .................... | .................... | ..................... | .................... | .................... | .................... |
| 18239 | MT971276.1 | \|Severe acute respiratory syndrome coro | .................... | .................... | ..................... | .................... | .................... | .................... |
| 18240 | MT971277.1 | \|Severe acute respiratory syndrome coro | .................... | .................... | ..................... | .................... | .................... | .................... |
| 18241 | MT971278.1 | \|Severe acute respiratory syndrome coro | .................... | .................... | ..................... | .................... | .................... | .................... |
| 18242 | MT971279.1 | \|Severe acute respiratory syndrome coro | .................... | .................... | ..................... | .................... | .................... | .................... |
| 18243 | MT971280.1 | \|Severe acute respiratory syndrome coro | .................... | .................... | ..................... | .................... | .................... | .................... |
| 18244 | MT971281.1 | \|Severe acute respiratory syndrome coro | .................... | .................... | ..................... | .................... | .................... | .................... |
| 18245 | MT971282.1 | \|Severe acute respiratory syndrome coro | .................... | .................... | ..................... | .................... | .................... | .................... |
| 18246 | MT971283.1 | \|Severe acute respiratory syndrome coro | .................... | .................... | ..................... | .................... | .................... | .................... |
| 18247 | MT971284.1 | \|Severe acute respiratory syndrome coro | .................... | .................... | ..................... | .................... | .................... | .................... |
| 18248 | MT971285.1 | \|Severe acute respiratory syndrome coro | .................... | .................... | ..................... | .................... | .................... | .................... |
| 18249 | MT971286.1 | \|Severe acute respiratory syndrome coro | .................... | .................... | ..................... | .................... | .................... | .................... |
| 18250 | MT971287.1 | \|Severe acute respiratory syndrome coro | .................... | .................... | ..................... | .................... | .................... | .................... |
| 18251 | MT971288.1 | \|Severe acute respiratory syndrome coro | .................... | .................... | ..................... | .................... | .................... | .................... |
| 18252 | MT971289.1 | \|Severe acute respiratory syndrome coro | .................... | .................... | ..................... | .................... | .................... | .................... |
| 18253 | MT971290.1 | \|Severe acute respiratory syndrome coro | .................... | .................... | ..................... | .................... | .................... | .................... |
| 18254 | MT971291.1 | \|Severe acute respiratory syndrome coro | .................... | .................... | ..................... | .................... | .................... | .................... |
| 18255 | MT971292.1 | \|Severe acute respiratory syndrome coro | .................... | .................... | ..................... | .................... | .................... | .................... |
| 18256 | MT971293.1 | \|Severe acute respiratory syndrome coro | .................... | .................... | ..................... | .................... | .................... | .................... |
| 18257 | MT971294.1 | \|Severe acute respiratory syndrome coro | .................... | .................... | ..................... | .................... | .................... | .................... |
| 18258 | MT971295.1 | \|Severe acute respiratory syndrome coro | .................... | .................... | ..................... | .................... | .................... | .................... |
| 18259 | MT971296.1 | \|Severe acute respiratory syndrome coro | .................... | .................... | ..................... | .................... | .................... | .................... |
| 18260 | MT971297.1 | \|Severe acute respiratory syndrome coro | .................... | .................... | ..................... | .................... | .................... | .................... |
| 18261 | MT971298.1 | \|Severe acute respiratory syndrome coro | .................... | .................... | ..................... | .................... | .................... | .................... |
| 18262 | MT971299.1 | \|Severe acute respiratory syndrome coro | .................... | .................... | ..................... | .................... | .................... | .................... |
| 18263 | MT971300.1 | \|Severe acute respiratory syndrome coro | .................... | .................... | ..................... | .................... | .................... | .................... |
| 18264 | MT971301.1 | \|Severe acute respiratory syndrome coro | .................... | .................... | ..................... | .................... | .................... | .................... |
| 18265 | MT971302.1 | \|Severe acute respiratory syndrome coro | .................... | .................... | ..................... | .................... | .................... | .................... |
| 18266 | MT971303.1 | \|Severe acute respiratory syndrome coro | .................... | .................... | ..................... | .................... | .................... | .................... |
| 18267 | MT971304.1 | \|Severe acute respiratory syndrome coro | .................... | .................... | ..................... | .................... | .................... | .................... |
| 18268 | MT971305.1 | \|Severe acute respiratory syndrome coro | .................... | .................... | ..................... | .................... | .................... | .................... |
| 18269 | MT971306.1 | \|Severe acute respiratory syndrome coro | .................... | .................... | ..................... | .................... | .................... | .................... |
| 18270 | MT971307.1 | \|Severe acute respiratory syndrome coro | .................... | .................... | ..................... | .................... | .................... | .................... |
| 18271 | MT971308.1 | \|Severe acute respiratory syndrome coro | .................... | .................... | ..................... | .................... | .................... | .................... |
| 18272 | MT971309.1 | \|Severe acute respiratory syndrome coro | .................... | .................... | ..................... | .................... | .................... | .................... |
| 18273 | MT971310.1 | \|Severe acute respiratory syndrome coro | .................... | .................... | ..................... | .................... | .................... | .................... |
| 18274 | MT971311.1 | \|Severe acute respiratory syndrome coro | .................... | .................... | ..................... | .................... | .................... | .................... |
| 18275 | MT971312.1 | \|Severe acute respiratory syndrome coro | .................... | .................... | ..................... | .................... | .................... | .................... |
| 18276 | MT971313.1 | \|Severe acute respiratory syndrome coro | .................... | .................... | ..................... | .................... | .................... | .................... |
| 18277 | MT971314.1 | \|Severe acute respiratory syndrome coro | .................... | .................... | ..................... | .................... | .................... | .................... |
| 18278 | MT971315.1 | \|Severe acute respiratory syndrome coro | .................... | .................... | ..................... | .................... | .................... | .................... |
| 18279 | MT971316.1 | \|Severe acute respiratory syndrome coro | .................... | .................... | ..................... | .................... | .................... | .................... |
| 18280 | MT971317.1 | \|Severe acute respiratory syndrome coro | .................... | .................... | ..................... | .................... | .................... | .................... |
| 18281 | MT971318.1 | \|Severe acute respiratory syndrome coro | .................... | .................... | ..................... | .................... | .................... | .................... |
| 18282 | MT971319.1 | \|Severe acute respiratory syndrome coro | .................... | .................... | ..................... | .................... | .................... | .................... |
| 18283 | MT971320.1 | \|Severe acute respiratory syndrome coro | .................... | .................... | ..................... | .................... | .................... | .................... |
| 18284 | MT971321.1 | \|Severe acute respiratory syndrome coro | .................... | .................... | ..................... | .................... | .................... | .................... |
| 18285 | MT971322.1 | \|Severe acute respiratory syndrome coro | .................... | .................... | ..................... | .................... | .................... | .................... |
| 18286 | MT971323.1 | \|Severe acute respiratory syndrome coro | .................... | .................... | ..................... | .................... | .................... | .................... |
| 18287 | MT971324.1 | \|Severe acute respiratory syndrome coro | .................... | .................... | ..................... | .................... | .................... | .................... |
| 18288 | MT971325.1 | \|Severe acute respiratory syndrome coro | .................... | .................... | ..................... | .................... | .................... | .................... |
| 18289 | MT971326.1 | \|Severe acute respiratory syndrome coro | .................... | .................... | ..................... | .................... | .................... | .................... |
| 18290 | MT971327.1 | \|Severe acute respiratory syndrome coro | .................... | .................... | ..................... | .................... | .................... | .................... |
| 18291 | MT971328.1 | \|Severe acute respiratory syndrome coro | .................... | .................... | ..................... | .................... | .................... | .................... |
| 18292 | MT971329.1 | \|Severe acute respiratory syndrome coro | .................... | .................... | ..................... | .................... | .................... | .................... |
| 18293 | MT971330.1 | \|Severe acute respiratory syndrome coro | .................... | .................... | ..................... | .................... | .................... | .................... |
| 18294 | MT971331.1 | \|Severe acute respiratory syndrome coro | .................... | .................... | ..................... | .................... | .................... | .................... |
| 18295 | MT971332.1 | \|Severe acute respiratory syndrome coro | .................... | .................... | ..................... | .................... | .................... | .................... |
| 18296 | MT971333.1 | \|Severe acute respiratory syndrome coro | .................... | .................... | ..................... | .................... | .................... | .................... |
| 18297 | MT971334.1 | \|Severe acute respiratory syndrome coro | .................... | .................... | ..................... | .................... | .................... | .................... |
| 18298 | MT971335.1 | \|Severe acute respiratory syndrome coro | .................... | .................... | ..................... | .................... | .................... | .................... |
| 18299 | MT971336.1 | \|Severe acute respiratory syndrome coro | .................... | .................... | ..................... | .................... | .................... | .................... |
| 18300 | MT971337.1 | \|Severe acute respiratory syndrome coro | .................... | .................... | ..................... | .................... | .................... | .................... |
| 18301 | MT971338.1 | \|Severe acute respiratory syndrome coro | .................... | .................... | ..................... | .................... | .................... | .................... |
| 18302 | MT971339.1 | \|Severe acute respiratory syndrome coro | .................... | .................... | ..................... | .................... | .................... | .................... |
| 18303 | MT971340.1 | \|Severe acute respiratory syndrome coro | .................... | .................... | ..................... | .................... | .................... | .................... |
| 18304 | MT971341.1 | \|Severe acute respiratory syndrome coro | .................... | .................... | ..................... | .................... | .................... | .................... |
| 18305 | MT971342.1 | \|Severe acute respiratory syndrome coro | .................... | .................... | ..................... | .................... | .................... | .................... |
| 18306 | MT971343.1 | \|Severe acute respiratory syndrome coro | .................... | .................... | ..................... | .................... | .................... | .................... |

```
18307 MT971344.1 |Severe acute respiratory syndrome coro    ....................  ....................  ....................  ....................  ....................  ....................
18308 MT971345.1 |Severe acute respiratory syndrome coro    ....................  ....................  ....................  ....................  ....................  ....................
18309 MT971346.1 |Severe acute respiratory syndrome coro    ....................  ....................  ....................  ....................  ....................  ....................
18310 MT971347.1 |Severe acute respiratory syndrome coro    ....................  ....................  ....................  ....................  ....................  ....................
18311 MT971348.1 |Severe acute respiratory syndrome coro    ....................  ....................  ....................  ....................  ....................  ....................
18312 MT971349.1 |Severe acute respiratory syndrome coro    ....................  ....................  ....................  ....................  ....................  ....................
18313 MT971350.1 |Severe acute respiratory syndrome coro    ....................  ....................  ....................  ....................  ....................  ....................
18314 MT971351.1 |Severe acute respiratory syndrome coro    ....................  ....................  ....................  ....................  ....................  ....................
18315 MT971352.1 |Severe acute respiratory syndrome coro    ....................  ....................  ....................  ....................  ....................  ....................
18316 MT971353.1 |Severe acute respiratory syndrome coro    ....................  ....................  ....................  ....................  ....................  ....................
18317 MT971354.1 |Severe acute respiratory syndrome coro    ....................  ....................  ....................  ....................  ....................  ....................
18318 MT971355.1 |Severe acute respiratory syndrome coro    ....................  ....................  ....................  ....................  ....................  ....................
18319 MT971356.1 |Severe acute respiratory syndrome coro    ....................  ....................  ....................  ....................  ....................  ....................
18320 MT971357.1 |Severe acute respiratory syndrome coro    ....................  ....................  ....................  ....................  ....................  ....................
18321 MT971358.1 |Severe acute respiratory syndrome coro    ....................  ....................  ....................  ....................  ....................  ....................
18322 MT971359.1 |Severe acute respiratory syndrome coro    ....................  ....................  ....................  ....................  ....................  ....................
18323 MT971360.1 |Severe acute respiratory syndrome coro    ....................  ....................  ....................  ....................  ....................  ....................
18324 MT971361.1 |Severe acute respiratory syndrome coro    ....................  ....................  ....................  ....................  ....................  ....................
18325 MT971362.1 |Severe acute respiratory syndrome coro    ....................  ....................  ....................  ....................  ....................  ....................
18326 MT971363.1 |Severe acute respiratory syndrome coro    ....................  ....................  ....................  ....................  ....................  ....................
18327 MT971364.1 |Severe acute respiratory syndrome coro    ....................  ....................  ....................  ....................  ....................  ....................
18328 MT971365.1 |Severe acute respiratory syndrome coro    ....................  ....................  ....................  ....................  ....................  ....................
18329 MT971366.1 |Severe acute respiratory syndrome coro    ....................  ....................  ....................  ....................  ....................  ....................
18330 MT971367.1 |Severe acute respiratory syndrome coro    ....................  ....................  ....................  ....................  ....................  ....................
18331 MT971368.1 |Severe acute respiratory syndrome coro    ....................  ....................  ....................  ....................  ....................  ....................
18332 MT971369.1 |Severe acute respiratory syndrome coro    ....................  ....................  ....................  ....................  ....................  ....................
18333 MT971370.1 |Severe acute respiratory syndrome coro    ....................  ....................  ....................  ....................  ....................  ....................
18334 MT971371.1 |Severe acute respiratory syndrome coro    ....................  ....................  ....................  ....................  ....................  ....................
18335 MT971372.1 |Severe acute respiratory syndrome coro    ....................  ....................  ....................  ....................  ....................  ....................
18336 MT971373.1 |Severe acute respiratory syndrome coro    ....................  ....................  ....................  ....................  ....................  ....................
18337 MT971374.1 |Severe acute respiratory syndrome coro    ....................  ....................  ....................  ....................  ....................  ....................
18338 MT971375.1 |Severe acute respiratory syndrome coro    ....................  ....................  ....................  ....................  ....................  ....................
18339 MT971376.1 |Severe acute respiratory syndrome coro    ....................  ....................  ....................  ....................  ....................  ....................
18340 MT971377.1 |Severe acute respiratory syndrome coro    ....................  ....................  ....................  ....................  ....................  ....................
18341 MT971378.1 |Severe acute respiratory syndrome coro    ....................  ....................  ....................  ....................  ....................  ....................
18342 MT971379.1 |Severe acute respiratory syndrome coro    ....................  ....................  ....................  ....................  ....................  ....................
18343 MT971380.1 |Severe acute respiratory syndrome coro    ....................  ....................  ....................  ....................  ....................  ....................
18344 MT971381.1 |Severe acute respiratory syndrome coro    ....................  ....................  ....................  ....................  ....................  ....................
18345 MT971382.1 |Severe acute respiratory syndrome coro    ....................  ....................  ....................  ....................  ....................  ....................
18346 MT971383.1 |Severe acute respiratory syndrome coro    ....................  ....................  ....................  ....................  ....................  ....................
18347 MT971384.1 |Severe acute respiratory syndrome coro    ....................  ....................  ....................  ....................  ....................  ....................
18348 MT971385.1 |Severe acute respiratory syndrome coro    ....................  ....................  ....................  ....................  ....................  ....................
18349 MT971386.1 |Severe acute respiratory syndrome coro    ....................  ....................  ....................  ....................  ....................  ....................
18350 MT971387.1 |Severe acute respiratory syndrome coro    ....................  ....................  ....................  ....................  ....................  ....................
18351 MT971388.1 |Severe acute respiratory syndrome coro    ....................  ....................  ....................  ....................  ....................  ....................
18352 MT971389.1 |Severe acute respiratory syndrome coro    ....................  ....................  ....................  ....................  ....................  ....................
18353 MT971390.1 |Severe acute respiratory syndrome coro    ....................  ....................  ....................  ....................  ....................  ....................
18354 MT971391.1 |Severe acute respiratory syndrome coro    ....................  ....................  ....................  ....................  ....................  ....................
18355 MT971392.1 |Severe acute respiratory syndrome coro    ....................  ....................  ....................  ....................  ....................  ....................
18356 MT971393.1 |Severe acute respiratory syndrome coro    ....................  ....................  ....................  ....................  ....................  ....................
18357 MT971394.1 |Severe acute respiratory syndrome coro    ....................  ....................  ....................  ....................  ....................  ....................
18358 MT971395.1 |Severe acute respiratory syndrome coro    ....................  ....................  ....................  ....................  ....................  ....................
18359 MT971396.1 |Severe acute respiratory syndrome coro    ....................  ....................  ....................  ....................  ....................  ....................
18360 MT971397.1 |Severe acute respiratory syndrome coro    ....................  ....................  ....................  ....................  ....................  ....................
18361 MT971398.1 |Severe acute respiratory syndrome coro    ....................  ....................  ....................  ....................  ....................  ....................
18362 MT971399.1 |Severe acute respiratory syndrome coro    ....................  ....................  ....................  ....................  ....................  ....................
18363 MT971400.1 |Severe acute respiratory syndrome coro    ....................  ....................  ....................  ....................  ....................  ....................
18364 MT971401.1 |Severe acute respiratory syndrome coro    ....................  ....................  ....................  ....................  ....................  ....................
18365 MT971402.1 |Severe acute respiratory syndrome coro    ....................  ....................  ....................  ....................  ....................  ....................
18366 MT971403.1 |Severe acute respiratory syndrome coro    ....................  ....................  ....................  ....................  ....................  ....................
18367 MT971404.1 |Severe acute respiratory syndrome coro    ....................  ....................  ....................  ....................  ....................  ....................
18368 MT971405.1 |Severe acute respiratory syndrome coro    ....................  ....................  ....................  ....................  ....................  ....................
18369 MT971406.1 |Severe acute respiratory syndrome coro    ....................  ....................  ....................  ....................  ....................  ....................
18370 MT971407.1 |Severe acute respiratory syndrome coro    ....................  ....................  ....................  ....................  ....................  ....................
18371 MT971408.1 |Severe acute respiratory syndrome coro    ....................  ....................  ....................  ....................  ....................  ....................
18372 MT971409.1 |Severe acute respiratory syndrome coro    ....................  ....................  ....................  ....................  ....................  ....................
18373 MT971410.1 |Severe acute respiratory syndrome coro    ....................  ....................  ....................  ....................  ....................  ....................
18374 MT971411.1 |Severe acute respiratory syndrome coro    ....................  ....................  ....................  ....................  ....................  ....................
18375 MT971412.1 |Severe acute respiratory syndrome coro    ....................  ....................  ....................  ....................  ....................  ....................
18376 MT971413.1 |Severe acute respiratory syndrome coro    ....................  ....................  ....................  ....................  ....................  ....................
18377 MT971415.1 |Severe acute respiratory syndrome coro    ....................  ....................  ....................  ....................  ....................  ....................
18378 MT971416.1 |Severe acute respiratory syndrome coro    ....................  ....................  ....................  ....................  ....................  ....................
18379 MT971418.1 |Severe acute respiratory syndrome coro    ....................  ....................  ....................  ....................  ....................  ....................
18380 MT971419.1 |Severe acute respiratory syndrome coro    ....................  ....................  ....................  ....................  ....................  ....................
18381 MT971420.1 |Severe acute respiratory syndrome coro    ....................  ....................  ....................  ....................  ....................  ....................
18382 MT971421.1 |Severe acute respiratory syndrome coro    ....................  ....................  ....................  ....................  ....................  ....................
18383 MT971422.1 |Severe acute respiratory syndrome coro    ....................  ....................  ....................  ....................  ....................  ....................
18384 MT971423.1 |Severe acute respiratory syndrome coro    ....................  ....................  ....................  ....................  ....................  ....................
```

| | | | | | | | | |
|---|---|---|---|---|---|---|---|---|
| 18385 | MT971424.1 | \|Severe acute respiratory syndrome coro | .................... | .................... | ..................... | .................... | .................... | .................... |
| 18386 | MT971426.1 | \|Severe acute respiratory syndrome coro | .................... | .................... | ..................... | .................... | .................... | .................... |
| 18387 | MT971427.1 | \|Severe acute respiratory syndrome coro | .................... | .................... | ..................... | .................... | .................... | .................... |
| 18388 | MT971428.1 | \|Severe acute respiratory syndrome coro | .................... | .................... | ..................... | .................... | .................... | .................... |
| 18389 | MT971429.1 | \|Severe acute respiratory syndrome coro | .................... | .................... | ..................... | .................... | .................... | .................... |
| 18390 | MT971430.1 | \|Severe acute respiratory syndrome coro | .................... | .................... | ..................... | .................... | .................... | .................... |
| 18391 | MT971431.1 | \|Severe acute respiratory syndrome coro | .................... | .................... | ..................... | .................... | .................... | .................... |
| 18392 | MT971432.1 | \|Severe acute respiratory syndrome coro | .................... | .................... | ..................... | .................... | .................... | .................... |
| 18393 | MT971433.1 | \|Severe acute respiratory syndrome coro | .................... | .................... | ..................... | .................... | .................... | .................... |
| 18394 | MT971434.1 | \|Severe acute respiratory syndrome coro | .................... | .................... | ..................... | .................... | .................... | .................... |
| 18395 | MT971435.1 | \|Severe acute respiratory syndrome coro | .................... | .................... | ..................... | .................... | .................... | .................... |
| 18396 | MT971436.1 | \|Severe acute respiratory syndrome coro | .................... | .................... | ..................... | .................... | .................... | .................... |
| 18397 | MT971437.1 | \|Severe acute respiratory syndrome coro | .................... | .................... | ..................... | .................... | .................... | .................... |
| 18398 | MT971438.1 | \|Severe acute respiratory syndrome coro | .................... | .................... | ..................... | .................... | .................... | .................... |
| 18399 | MT971439.1 | \|Severe acute respiratory syndrome coro | .................... | .................... | ..................... | .................... | .................... | .................... |
| 18400 | MT971440.1 | \|Severe acute respiratory syndrome coro | .................... | .................... | ..................... | .................... | .................... | .................... |
| 18401 | MT971441.1 | \|Severe acute respiratory syndrome coro | .................... | .................... | ..................... | .................... | .................... | .................... |
| 18402 | MT971442.1 | \|Severe acute respiratory syndrome coro | .................... | .................... | ..................... | .................... | .................... | .................... |
| 18403 | MT971443.1 | \|Severe acute respiratory syndrome coro | .................... | .................... | ..................... | .................... | .................... | .................... |
| 18404 | MT971444.1 | \|Severe acute respiratory syndrome coro | .................... | .................... | ..................... | .................... | .................... | .................... |
| 18405 | MT971445.1 | \|Severe acute respiratory syndrome coro | .................... | .................... | ..................... | .................... | .................... | .................... |
| 18406 | MT971446.1 | \|Severe acute respiratory syndrome coro | .................... | .................... | ..................... | .................... | .................... | .................... |
| 18407 | MT971447.1 | \|Severe acute respiratory syndrome coro | .................... | .................... | ..................... | .................... | .................... | .................... |
| 18408 | MT971449.1 | \|Severe acute respiratory syndrome coro | .................... | .................... | ..................... | .................... | .................... | .................... |
| 18409 | MT971450.1 | \|Severe acute respiratory syndrome coro | .................... | .................... | ..................... | .................... | .................... | .................... |
| 18410 | MT971451.1 | \|Severe acute respiratory syndrome coro | .................... | .................... | ..................... | .................... | .................... | .................... |
| 18411 | MT971452.1 | \|Severe acute respiratory syndrome coro | .................... | .................... | ..................... | .................... | .................... | .................... |
| 18412 | MT971453.1 | \|Severe acute respiratory syndrome coro | .................... | .................... | ..................... | .................... | .................... | .................... |
| 18413 | MT971454.1 | \|Severe acute respiratory syndrome coro | .................... | .................... | ..................... | .................... | .................... | .................... |
| 18415 | MT971456.1 | \|Severe acute respiratory syndrome coro | .................... | .................... | ..................... | .................... | .................... | .................... |
| 18416 | MT971458.1 | \|Severe acute respiratory syndrome coro | .................... | .................... | ..................... | .................... | .................... | .................... |
| 18417 | MT971459.1 | \|Severe acute respiratory syndrome coro | .................... | .................... | ..................... | .................... | .................... | .................... |
| 18418 | MT971460.1 | \|Severe acute respiratory syndrome coro | .................... | .................... | ..................... | .................... | .................... | .................... |
| 18419 | MT971461.1 | \|Severe acute respiratory syndrome coro | .................... | .................... | ..................... | .................... | .................... | .................... |
| 18420 | MT971462.1 | \|Severe acute respiratory syndrome coro | .................... | .................... | ..................... | .................... | .................... | .................... |
| 18421 | MT971463.1 | \|Severe acute respiratory syndrome coro | .................... | .................... | ..................... | .................... | .................... | .................... |
| 18422 | MT971464.1 | \|Severe acute respiratory syndrome coro | .................... | .................... | ..................... | .................... | .................... | .................... |
| 18423 | MT971465.1 | \|Severe acute respiratory syndrome coro | .................... | .................... | ..................... | .................... | .................... | .................... |
| 18424 | MT971466.1 | \|Severe acute respiratory syndrome coro | .................... | .................... | ..................... | .................... | .................... | .................... |
| 18425 | MT971467.1 | \|Severe acute respiratory syndrome coro | .................... | .................... | ..................... | .................... | .................... | .................... |
| 18426 | MT971468.1 | \|Severe acute respiratory syndrome coro | .................... | .................... | ..................... | .................... | .................... | .................... |
| 18427 | MT971469.1 | \|Severe acute respiratory syndrome coro | .................... | .................... | ..................... | .................... | .................... | .................... |
| 18428 | MT971471.1 | \|Severe acute respiratory syndrome coro | .................... | .................... | ..................... | .................... | .................... | .................... |
| 18429 | MT971472.1 | \|Severe acute respiratory syndrome coro | .................... | .................... | ..................... | .................... | .................... | .................... |
| 18430 | MT971473.1 | \|Severe acute respiratory syndrome coro | .................... | .................... | ..................... | .................... | .................... | .................... |
| 18431 | MT971474.1 | \|Severe acute respiratory syndrome coro | .................... | .................... | ..................... | .................... | .................... | .................... |
| 18432 | MT971475.1 | \|Severe acute respiratory syndrome coro | .................... | .................... | ..................... | .................... | .................... | .................... |
| 18433 | MT971476.1 | \|Severe acute respiratory syndrome coro | .................... | .................... | ..................... | .................... | .................... | .................... |
| 18434 | MT971477.1 | \|Severe acute respiratory syndrome coro | .................... | .................... | ..................... | .................... | .................... | .................... |
| 18435 | MT971478.1 | \|Severe acute respiratory syndrome coro | .................... | .................... | ..................... | .................... | .................... | .................... |
| 18436 | MT971480.1 | \|Severe acute respiratory syndrome coro | .................... | .................... | ..................... | .................... | .................... | .................... |
| 18437 | MT971481.1 | \|Severe acute respiratory syndrome coro | .................... | .................... | ..................... | .................... | .................... | .................... |
| 18438 | MT971482.1 | \|Severe acute respiratory syndrome coro | .................... | .................... | ..................... | .................... | .................... | .................... |
| 18439 | MT971483.1 | \|Severe acute respiratory syndrome coro | .................... | .................... | ..................... | .................... | .................... | .................... |
| 18440 | MT971484.1 | \|Severe acute respiratory syndrome coro | .................... | .................... | ..................... | .................... | .................... | .................... |
| 18441 | MT971485.1 | \|Severe acute respiratory syndrome coro | .................... | .................... | ..................... | .................... | .................... | .................... |
| 18442 | MT971486.1 | \|Severe acute respiratory syndrome coro | .................... | .................... | ..................... | .................... | .................... | .................... |
| 18443 | MT971487.1 | \|Severe acute respiratory syndrome coro | .................... | .................... | ..................... | .................... | .................... | .................... |
| 18444 | MT971488.1 | \|Severe acute respiratory syndrome coro | .................... | .................... | ..................... | .................... | .................... | .................... |
| 18445 | MT971489.1 | \|Severe acute respiratory syndrome coro | .................... | .................... | ..................... | .................... | .................... | .................... |
| 18446 | MT971490.1 | \|Severe acute respiratory syndrome coro | .................... | .................... | ..................... | .................... | .................... | .................... |
| 18447 | MT971491.1 | \|Severe acute respiratory syndrome coro | .................... | .................... | ..................... | .................... | .................... | .................... |
| 18448 | MT971492.1 | \|Severe acute respiratory syndrome coro | .................... | .................... | ..................... | .................... | .................... | .................... |
| 18449 | MT971493.1 | \|Severe acute respiratory syndrome coro | .................... | .................... | ..................... | .................... | .................... | .................... |
| 18450 | MT971495.1 | \|Severe acute respiratory syndrome coro | .................... | .................... | ..................... | .................... | .................... | .................... |
| 18451 | MT971496.1 | \|Severe acute respiratory syndrome coro | .................... | .................... | ..................... | .................... | .................... | .................... |
| 18452 | MT971497.1 | \|Severe acute respiratory syndrome coro | .................... | .................... | ..................... | .................... | .................... | .................... |
| 18453 | MT971498.1 | \|Severe acute respiratory syndrome coro | .................... | .................... | ..................... | .................... | .................... | .................... |
| 18454 | MT971499.1 | \|Severe acute respiratory syndrome coro | .................... | .................... | ..................... | .................... | .................... | .................... |
| 18455 | MT971500.1 | \|Severe acute respiratory syndrome coro | .................... | .................... | ..................... | .................... | .................... | .................... |
| 18456 | MT971501.1 | \|Severe acute respiratory syndrome coro | .................... | .................... | ..................... | .................... | .................... | .................... |
| 18457 | MT971502.1 | \|Severe acute respiratory syndrome coro | .................... | .................... | ..................... | .................... | .................... | .................... |
| 18458 | MT971503.1 | \|Severe acute respiratory syndrome coro | .................... | .................... | ..................... | .................... | .................... | .................... |
| 18459 | MT971505.1 | \|Severe acute respiratory syndrome coro | .................... | .................... | ..................... | .................... | .................... | .................... |
| 18460 | MT971506.1 | \|Severe acute respiratory syndrome coro | .................... | .................... | ..................... | .................... | .................... | .................... |
| 18461 | MT971507.1 | \|Severe acute respiratory syndrome coro | .................... | .................... | ..................... | .................... | .................... | .................... |
| 18462 | MT971508.1 | \|Severe acute respiratory syndrome coro | .................... | .................... | ..................... | .................... | .................... | .................... |
| 18463 | MT971510.1 | \|Severe acute respiratory syndrome coro | .................... | .................... | ..................... | .................... | .................... | .................... |

18464 MT971511.1 |Severe acute respiratory syndrome coro .................... .................... ..................... .................... .................... ....................
18465 MT971512.1 |Severe acute respiratory syndrome coro .................... .................... ..................... .................... .................... ....................
18466 MT971514.1 |Severe acute respiratory syndrome coro .................... .................... ..................... .................... .................... ....................
18467 MT971515.1 |Severe acute respiratory syndrome coro .................... .................... ..................... .................... .................... ....................
18468 MT971517.1 |Severe acute respiratory syndrome coro .................... .................... ..................... .................... .................... ....................
18469 MT971518.1 |Severe acute respiratory syndrome coro .................... .................... ..................... .................... .................... ....................
18470 MT971519.1 |Severe acute respiratory syndrome coro .................... .................... ..................... .................... .................... ....................
18471 MT971520.1 |Severe acute respiratory syndrome coro .................... .................... ..................... .................... .................... ....................
18472 MT971521.1 |Severe acute respiratory syndrome coro .................... .................... ..................... .................... .................... ....................
18473 MT971522.1 |Severe acute respiratory syndrome coro .................... .................... ..................... .................... .................... ....................
18474 MT971523.1 |Severe acute respiratory syndrome coro .................... .................... ..................... .................... .................... ....................
18475 MT971524.1 |Severe acute respiratory syndrome coro .................... .................... ..................... .................... .................... ....................
18476 MT971525.1 |Severe acute respiratory syndrome coro .................... .................... ..................... .................... .................... ....................
18477 MT971526.1 |Severe acute respiratory syndrome coro .................... .................... ..................... .................... .................... ....................
18478 MT971527.1 |Severe acute respiratory syndrome coro .................... .................... ..................... .................... .................... ....................
18479 MT971528.1 |Severe acute respiratory syndrome coro .................... .................... ..................... .................... .................... ....................
18480 MT971529.1 |Severe acute respiratory syndrome coro .................... .................... ..................... .................... .................... ....................
18481 MT971530.1 |Severe acute respiratory syndrome coro .................... .................... ..................... .................... .................... ....................
18482 MT971531.1 |Severe acute respiratory syndrome coro .................... .................... ..................... .................... .................... ....................
18483 MT971532.1 |Severe acute respiratory syndrome coro .................... .................... ..................... .................... .................... ....................
18484 MT971533.1 |Severe acute respiratory syndrome coro .................... .................... ..................... .................... .................... ....................
18485 MT971535.1 |Severe acute respiratory syndrome coro .................... .................... ..................... .................... .................... ....................
18486 MT971536.1 |Severe acute respiratory syndrome coro .................... .................... ..................... .................... .................... ....................
18487 MT971537.1 |Severe acute respiratory syndrome coro .................... .................... ..................... .................... .................... ....................
18488 MT971538.1 |Severe acute respiratory syndrome coro .................... .................... ..................... .................... .................... ....................
18489 MT971539.1 |Severe acute respiratory syndrome coro .................... .................... ..................... .................... .................... ....................
18490 MT971540.1 |Severe acute respiratory syndrome coro .................... .................... ..................... .................... .................... ....................
18491 MT971541.1 |Severe acute respiratory syndrome coro .................... .................... ..................... .................... .................... ....................
18492 MT971542.1 |Severe acute respiratory syndrome coro .................... .................... ..................... .................... .................... ....................
18493 MT971543.1 |Severe acute respiratory syndrome coro .................... .................... ..................... .................... .................... ....................
18494 MT971544.1 |Severe acute respiratory syndrome coro .................... .................... ..................... .................... .................... ....................
18495 MT971545.1 |Severe acute respiratory syndrome coro .................... .................... ..................... .................... .................... ....................
18496 MT971546.1 |Severe acute respiratory syndrome coro .................... .................... ..................... .................... .................... ....................
18497 MT971547.1 |Severe acute respiratory syndrome coro .................... .................... ..................... .................... .................... ....................
18498 MT971548.1 |Severe acute respiratory syndrome coro .................... .................... ..................... .................... .................... ....................
18499 MT971549.1 |Severe acute respiratory syndrome coro .................... .................... ..................... .................... .................... ....................
18500 MT971550.1 |Severe acute respiratory syndrome coro .................... .................... ..................... .................... .................... ....................
18501 MT971551.1 |Severe acute respiratory syndrome coro .................... .................... ..................... .................... .................... ....................
18502 MT971553.1 |Severe acute respiratory syndrome coro .................... .................... ..................... .................... .................... ....................
18503 MT971554.1 |Severe acute respiratory syndrome coro .................... .................... ..................... .................... .................... ....................
18504 MT971555.1 |Severe acute respiratory syndrome coro .................... .................... ..................... .................... .................... ....................
18505 MT971556.1 |Severe acute respiratory syndrome coro .................... .................... ..................... .................... .................... ....................
18506 MT971557.1 |Severe acute respiratory syndrome coro .................... .................... ..................... .................... .................... ....................
18507 MT971558.1 |Severe acute respiratory syndrome coro .................... .................... ..................... .................... .................... ....................
18508 MT971560.1 |Severe acute respiratory syndrome coro .................... .................... ..................... .................... .................... ....................
18509 MT971561.1 |Severe acute respiratory syndrome coro .................... .................... ..................... .................... .................... ....................
18510 MT971562.1 |Severe acute respiratory syndrome coro .................... .................... ..................... .................... .................... ....................
18511 MT971564.1 |Severe acute respiratory syndrome coro .................... .................... ..................... .................... .................... ....................
18512 MT971566.1 |Severe acute respiratory syndrome coro .................... .................... ..................... .................... .................... ....................
18513 MT971568.1 |Severe acute respiratory syndrome coro .................... .................... ..................... .................... .................... ....................
18514 MT971569.1 |Severe acute respiratory syndrome coro .................... .................... ..................... .................... .................... ....................
18515 MT971572.1 |Severe acute respiratory syndrome coro .................... .................... ..................... .................... .................... ....................
18516 MT971573.1 |Severe acute respiratory syndrome coro .................... .................... ..................... .................... .................... ....................
18517 MT971574.1 |Severe acute respiratory syndrome coro .................... .................... ..................... .................... .................... ....................
18518 MT971576.1 |Severe acute respiratory syndrome coro .................... .................... ..................... .................... .................... ....................
18519 MT971577.1 |Severe acute respiratory syndrome coro .................... .................... ..................... .................... .................... ....................
18520 MT971579.1 |Severe acute respiratory syndrome coro .................... .................... ..................... .................... .................... ....................
18521 MT971581.1 |Severe acute respiratory syndrome coro .................... .................... ..................... .................... .................... ....................
18522 MT971582.1 |Severe acute respiratory syndrome coro .................... .................... ..................... .................... .................... ....................
18523 MT971583.1 |Severe acute respiratory syndrome coro .................... .................... ..................... .................... .................... ....................
18524 MT971584.1 |Severe acute respiratory syndrome coro .................... .................... ..................... .................... .................... ....................
18525 MT971585.1 |Severe acute respiratory syndrome coro .................... .................... ..................... .................... .................... ....................
18527 MT971588.1 |Severe acute respiratory syndrome coro .................... .................... ..................... .................... .................... ....................
18528 MT971589.1 |Severe acute respiratory syndrome coro .................... .................... ..................... .................... .................... ....................
18529 MT971590.1 |Severe acute respiratory syndrome coro .................... .................... ..................... .................... .................... ....................
18530 MT971591.1 |Severe acute respiratory syndrome coro .................... .................... ..................... .................... .................... ....................
18531 MT971594.1 |Severe acute respiratory syndrome coro .................... .................... ..................... .................... .................... ....................
18532 MT971595.1 |Severe acute respiratory syndrome coro .................... .................... ..................... .................... .................... ....................
18533 MT971596.1 |Severe acute respiratory syndrome coro .................... .................... ..................... .................... .................... ....................
18534 MT971599.1 |Severe acute respiratory syndrome coro .................... .................... ..................... .................... .................... ....................
18535 MT971600.1 |Severe acute respiratory syndrome coro .................... .................... ..................... .................... .................... ....................
18536 MT971602.1 |Severe acute respiratory syndrome coro .................... .................... ..................... .................... .................... ....................
18537 MT971603.1 |Severe acute respiratory syndrome coro .................... .................... ..................... .................... .................... ....................
18538 MT971604.1 |Severe acute respiratory syndrome coro .................... .................... ..................... .................... .................... ....................
18539 MT971605.1 |Severe acute respiratory syndrome coro .................... .................... ..................... .................... .................... ....................
18540 MT971606.1 |Severe acute respiratory syndrome coro .................... .................... ..................... .................... .................... ....................
18541 MT971607.1 |Severe acute respiratory syndrome coro .................... .................... ..................... .................... .................... ....................
18542 MT971608.1 |Severe acute respiratory syndrome coro .................... .................... ..................... .................... .................... ....................

| | | | | | | | | |
|---|---|---|---|---|---|---|---|---|
| 18543 | MT971609.1 | \|Severe acute respiratory syndrome coro | .................... | .................... | .................... | .................... | .................... | .................... |
| 18544 | MT971610.1 | \|Severe acute respiratory syndrome coro | .................... | .................... | .................... | .................... | .................... | .................... |
| 18545 | MT971611.1 | \|Severe acute respiratory syndrome coro | .................... | .................... | .................... | .................... | .................... | .................... |
| 18546 | MT971612.1 | \|Severe acute respiratory syndrome coro | .................... | .................... | .................... | .................... | .................... | .................... |
| 18547 | MT971613.1 | \|Severe acute respiratory syndrome coro | .................... | .................... | .................... | .................... | .................... | .................... |
| 18548 | MT971615.1 | \|Severe acute respiratory syndrome coro | .................... | .................... | .................... | .................... | .................... | .................... |
| 18549 | MT971616.1 | \|Severe acute respiratory syndrome coro | .................... | .................... | .................... | .................... | .................... | .................... |
| 18550 | MT971617.1 | \|Severe acute respiratory syndrome coro | .................... | .................... | .................... | .................... | .................... | .................... |
| 18551 | MT971618.1 | \|Severe acute respiratory syndrome coro | .................... | .................... | .................... | .................... | .................... | .................... |
| 18552 | MT971619.1 | \|Severe acute respiratory syndrome coro | .................... | .................... | .................... | .................... | .................... | .................... |
| 18553 | MT971621.1 | \|Severe acute respiratory syndrome coro | .................... | .................... | .................... | .................... | .................... | .................... |
| 18554 | MT971622.1 | \|Severe acute respiratory syndrome coro | .................... | .................... | .................... | .................... | .................... | .................... |
| 18555 | MT971627.1 | \|Severe acute respiratory syndrome coro | .................... | .................... | .................... | .................... | .................... | .................... |
| 18556 | MT971629.1 | \|Severe acute respiratory syndrome coro | .................... | .................... | .................... | .................... | .................... | .................... |
| 18557 | MT971630.1 | \|Severe acute respiratory syndrome coro | .................... | .................... | .................... | .................... | .................... | .................... |
| 18558 | MT971631.1 | \|Severe acute respiratory syndrome coro | .................... | .................... | .................... | .................... | .................... | .................... |
| 18559 | MT971632.1 | \|Severe acute respiratory syndrome coro | .................... | .................... | .................... | .................... | .................... | .................... |
| 18560 | MT971634.1 | \|Severe acute respiratory syndrome coro | .................... | .................... | .................... | .................... | .................... | .................... |
| 18561 | MT971635.1 | \|Severe acute respiratory syndrome coro | .................... | .................... | .................... | .................... | .................... | .................... |
| 18562 | MT971636.1 | \|Severe acute respiratory syndrome coro | .................... | .................... | .................... | .................... | .................... | .................... |
| 18563 | MT971637.1 | \|Severe acute respiratory syndrome coro | .................... | .................... | .................... | .................... | .................... | .................... |
| 18564 | MT971638.1 | \|Severe acute respiratory syndrome coro | .................... | .................... | .................... | .................... | .................... | .................... |
| 18565 | MT971639.1 | \|Severe acute respiratory syndrome coro | .................... | .................... | .................... | .................... | .................... | .................... |
| 18566 | MT971640.1 | \|Severe acute respiratory syndrome coro | .................... | .................... | .................... | .................... | .................... | .................... |
| 18567 | MT971641.1 | \|Severe acute respiratory syndrome coro | .................... | .................... | .................... | .................... | .................... | .................... |
| 18568 | MT971642.1 | \|Severe acute respiratory syndrome coro | .................... | .................... | .................... | .................... | .................... | .................... |
| 18569 | MT971643.1 | \|Severe acute respiratory syndrome coro | .................... | .................... | .................... | .................... | .................... | .................... |
| 18570 | MT971644.1 | \|Severe acute respiratory syndrome coro | .................... | .................... | .................... | .................... | .................... | .................... |
| 18571 | MT971645.1 | \|Severe acute respiratory syndrome coro | .................... | .................... | .................... | .................... | .................... | .................... |
| 18572 | MT971646.1 | \|Severe acute respiratory syndrome coro | .................... | .................... | .................... | .................... | .................... | .................... |
| 18573 | MT971647.1 | \|Severe acute respiratory syndrome coro | .................... | .................... | .................... | .................... | .................... | .................... |
| 18574 | MT971649.1 | \|Severe acute respiratory syndrome coro | .................... | .................... | .................... | .................... | .................... | .................... |
| 18575 | MT971650.1 | \|Severe acute respiratory syndrome coro | .................... | .................... | .................... | .................... | .................... | .................... |
| 18577 | MT971652.1 | \|Severe acute respiratory syndrome coro | .................... | .................... | .................... | .................... | .................... | .................... |
| 18578 | MT971653.1 | \|Severe acute respiratory syndrome coro | .................... | .................... | .................... | .................... | .................... | .................... |
| 18579 | MT971654.1 | \|Severe acute respiratory syndrome coro | .................... | .................... | .................... | .................... | .................... | .................... |
| 18580 | MT971655.1 | \|Severe acute respiratory syndrome coro | .................... | .................... | .................... | .................... | .................... | .................... |
| 18581 | MT971656.1 | \|Severe acute respiratory syndrome coro | .................... | .................... | .................... | .................... | .................... | .................... |
| 18582 | MT971657.1 | \|Severe acute respiratory syndrome coro | .................... | .................... | .................... | .................... | .................... | .................... |
| 18583 | MT971658.1 | \|Severe acute respiratory syndrome coro | .................... | .................... | .................... | .................... | .................... | .................... |
| 18584 | MT971659.1 | \|Severe acute respiratory syndrome coro | .................... | .................... | .................... | .................... | .................... | .................... |
| 18585 | MT971660.1 | \|Severe acute respiratory syndrome coro | .................... | .................... | .................... | .................... | .................... | .................... |
| 18586 | MT971661.1 | \|Severe acute respiratory syndrome coro | .................... | .................... | .................... | .................... | .................... | .................... |
| 18587 | MT971662.1 | \|Severe acute respiratory syndrome coro | .................... | .................... | .................... | .................... | .................... | .................... |
| 18588 | MT971663.1 | \|Severe acute respiratory syndrome coro | .................... | .................... | .................... | .................... | .................... | .................... |
| 18589 | MT971664.1 | \|Severe acute respiratory syndrome coro | .................... | .................... | .................... | .................... | .................... | .................... |
| 18590 | MT971665.1 | \|Severe acute respiratory syndrome coro | .................... | .................... | .................... | .................... | .................... | .................... |
| 18591 | MT971666.1 | \|Severe acute respiratory syndrome coro | .................... | .................... | .................... | .................... | .................... | .................... |
| 18592 | MT971667.1 | \|Severe acute respiratory syndrome coro | .................... | .................... | .................... | .................... | .................... | .................... |
| 18593 | MT971670.1 | \|Severe acute respiratory syndrome coro | .................... | .................... | .................... | .................... | .................... | .................... |
| 18594 | MT971671.1 | \|Severe acute respiratory syndrome coro | .................... | .................... | .................... | .................... | .................... | .................... |
| 18596 | MT971673.1 | \|Severe acute respiratory syndrome coro | .................... | .................... | .................... | .................... | .................... | .................... |
| 18597 | MT971675.1 | \|Severe acute respiratory syndrome coro | .................... | .................... | .................... | .................... | .................... | .................... |
| 18598 | MT971676.1 | \|Severe acute respiratory syndrome coro | .................... | .................... | .................... | .................... | .................... | .................... |
| 18599 | MT971677.1 | \|Severe acute respiratory syndrome coro | .................... | .................... | .................... | .................... | .................... | .................... |
| 18600 | MT971678.1 | \|Severe acute respiratory syndrome coro | .................... | .................... | .................... | .................... | .................... | .................... |
| 18601 | MT971679.1 | \|Severe acute respiratory syndrome coro | .................... | .................... | .................... | .................... | .................... | .................... |
| 18602 | MT971680.1 | \|Severe acute respiratory syndrome coro | .................... | .................... | .................... | .................... | .................... | .................... |
| 18603 | MT971681.1 | \|Severe acute respiratory syndrome coro | .................... | .................... | .................... | .................... | .................... | .................... |
| 18604 | MT971682.1 | \|Severe acute respiratory syndrome coro | .................... | .................... | .................... | .................... | .................... | .................... |
| 18605 | MT971683.1 | \|Severe acute respiratory syndrome coro | .................... | .................... | .................... | .................... | .................... | .................... |
| 18606 | MT971684.1 | \|Severe acute respiratory syndrome coro | .................... | .................... | .................... | .................... | .................... | .................... |
| 18607 | MT971685.1 | \|Severe acute respiratory syndrome coro | .................... | .................... | .................... | .................... | .................... | .................... |
| 18608 | MT971686.1 | \|Severe acute respiratory syndrome coro | .................... | .................... | .................... | .................... | .................... | .................... |
| 18609 | MT971687.1 | \|Severe acute respiratory syndrome coro | .................... | .................... | .................... | .................... | .................... | .................... |
| 18610 | MT971688.1 | \|Severe acute respiratory syndrome coro | .................... | .................... | .................... | .................... | .................... | .................... |
| 18611 | MT971689.1 | \|Severe acute respiratory syndrome coro | .................... | .................... | .................... | .................... | .................... | .................... |
| 18612 | MT971691.1 | \|Severe acute respiratory syndrome coro | .................... | .................... | .................... | .................... | .................... | .................... |
| 18613 | MT971692.1 | \|Severe acute respiratory syndrome coro | .................... | .................... | .................... | .................... | .................... | .................... |
| 18614 | MT971693.1 | \|Severe acute respiratory syndrome coro | .................... | .................... | .................... | .................... | .................... | .................... |
| 18615 | MT971694.1 | \|Severe acute respiratory syndrome coro | .................... | .................... | .................... | .................... | .................... | .................... |
| 18616 | MT971695.1 | \|Severe acute respiratory syndrome coro | .................... | .................... | .................... | .................... | .................... | .................... |
| 18617 | MT971696.1 | \|Severe acute respiratory syndrome coro | .................... | .................... | .................... | .................... | .................... | .................... |
| 18618 | MT971697.1 | \|Severe acute respiratory syndrome coro | .................... | .................... | .................... | .................... | .................... | .................... |
| 18619 | MT971698.1 | \|Severe acute respiratory syndrome coro | .................... | .................... | .................... | .................... | .................... | .................... |
| 18620 | MT971700.1 | \|Severe acute respiratory syndrome coro | .................... | .................... | .................... | .................... | .................... | .................... |
| 18621 | MT971701.1 | \|Severe acute respiratory syndrome coro | .................... | .................... | .................... | .................... | .................... | .................... |
| 18622 | MT971702.1 | \|Severe acute respiratory syndrome coro | .................... | .................... | .................... | .................... | .................... | .................... |

```
18623 MT971703.1 |Severe acute respiratory syndrome coro    ....................  ....................  ....................  ....................  ....................  ....................
18624 MT971704.1 |Severe acute respiratory syndrome coro    ....................  ....................  ....................  ....................  ....................  ....................
18625 MT971705.1 |Severe acute respiratory syndrome coro    ....................  ....................  ....................  ....................  ....................  ....................
18626 MT971706.1 |Severe acute respiratory syndrome coro    ....................  ....................  ....................  ....................  ....................  ....................
18627 MT971707.1 |Severe acute respiratory syndrome coro    ....................  ....................  ....................  ....................  ....................  ....................
18628 MT971709.1 |Severe acute respiratory syndrome coro    ....................  ....................  ....................  ....................  ....................  ....................
18629 MT971710.1 |Severe acute respiratory syndrome coro    ....................  ....................  ....................  ....................  ....................  ....................
18630 MT971711.1 |Severe acute respiratory syndrome coro    ....................  ....................  ....................  ....................  ....................  ....................
18631 MT971712.1 |Severe acute respiratory syndrome coro    ....................  ....................  ....................  ....................  ....................  ....................
18632 MT971713.1 |Severe acute respiratory syndrome coro    ....................  ....................  ....................  ....................  ....................  ....................
18633 MT971714.1 |Severe acute respiratory syndrome coro    ....................  ....................  ....................  ....................  ....................  ....................
18634 MT971715.1 |Severe acute respiratory syndrome coro    ....................  ....................  ....................  ....................  ....................  ....................
18635 MT971716.1 |Severe acute respiratory syndrome coro    ....................  ....................  ....................  ....................  ....................  ....................
18636 MT971718.1 |Severe acute respiratory syndrome coro    ....................  ....................  ....................  ....................  ....................  ....................
18637 MT971719.1 |Severe acute respiratory syndrome coro    ....................  ....................  ....................  ....................  ....................  ....................
18638 MT971720.1 |Severe acute respiratory syndrome coro    ....................  ....................  ....................  ....................  ....................  ....................
18639 MT971721.1 |Severe acute respiratory syndrome coro    ....................  ....................  ....................  ....................  ....................  ....................
18640 MT971722.1 |Severe acute respiratory syndrome coro    ....................  ....................  ....................  ....................  ....................  ....................
18641 MT971723.1 |Severe acute respiratory syndrome coro    ....................  ....................  ....................  ....................  ....................  ....................
18642 MT971724.1 |Severe acute respiratory syndrome coro    ....................  ....................  ....................  ....................  ....................  ....................
18643 MT971725.1 |Severe acute respiratory syndrome coro    ....................  ....................  ....................  ....................  ....................  ....................
18644 MT971726.1 |Severe acute respiratory syndrome coro    ....................  ....................  ....................  ....................  ....................  ....................
18645 MT971727.1 |Severe acute respiratory syndrome coro    ....................  ....................  ....................  ....................  ....................  ....................
18646 MT971728.1 |Severe acute respiratory syndrome coro    ....................  ....................  ....................  ....................  ....................  ....................
18647 MT971729.1 |Severe acute respiratory syndrome coro    ....................  ....................  ....................  ....................  ....................  ....................
18648 MT971730.1 |Severe acute respiratory syndrome coro    ....................  ....................  ....................  ....................  ....................  ....................
18649 MT971731.1 |Severe acute respiratory syndrome coro    ....................  ....................  ....................  ....................  ....................  ....................
18650 MT971733.1 |Severe acute respiratory syndrome coro    ....................  ....................  ....................  ....................  ....................  ....................
18651 MT971734.1 |Severe acute respiratory syndrome coro    ....................  ....................  ....................  ....................  ....................  ....................
18652 MT971735.1 |Severe acute respiratory syndrome coro    ....................  ....................  ....................  ....................  ....................  ....................
18653 MT971736.1 |Severe acute respiratory syndrome coro    ....................  ....................  ....................  ....................  ....................  ....................
18654 MT971737.1 |Severe acute respiratory syndrome coro    ....................  ....................  ....................  ....................  ....................  ....................
18655 MT971738.1 |Severe acute respiratory syndrome coro    ....................  ....................  ....................  ....................  ....................  ....................
18656 MT971739.1 |Severe acute respiratory syndrome coro    ....................  ....................  ....................  ....................  ....................  ....................
18657 MT971740.1 |Severe acute respiratory syndrome coro    ....................  ....................  ....................  ....................  ....................  ....................
18658 MT971741.1 |Severe acute respiratory syndrome coro    ....................  ....................  ....................  ....................  ....................  ...A................
18659 MT971742.1 |Severe acute respiratory syndrome coro    ....................  ....................  ....................  ....................  ....................  ....................
18660 MT971743.1 |Severe acute respiratory syndrome coro    ....................  ....................  ....................  ....................  ....................  ....................
18661 MT971744.1 |Severe acute respiratory syndrome coro    ....................  ....................  ....................  ....................  ....................  ....................
18662 MT971745.1 |Severe acute respiratory syndrome coro    ....................  ....................  ....................  ....................  ....................  ....................
18663 MT971746.1 |Severe acute respiratory syndrome coro    ....................  ....................  ....................  ....................  ....................  ....................
18664 MT971747.1 |Severe acute respiratory syndrome coro    ....................  ....................  ....................  ....................  ....................  ....................
18665 MT971748.1 |Severe acute respiratory syndrome coro    ....................  ....................  ....................  ....................  ....................  ....................
18666 MT971749.1 |Severe acute respiratory syndrome coro    ....................  ....................  ....................  ....................  ....................  ....................
18667 MT971750.1 |Severe acute respiratory syndrome coro    ....................  ....................  ....................  ....................  ....................  ....................
18668 MT971751.1 |Severe acute respiratory syndrome coro    ....................  ....................  ....................  ....................  ....................  ....................
18669 MT971752.1 |Severe acute respiratory syndrome coro    ....................  ....................  ....................  ....................  ....................  ....................
18670 MT971753.1 |Severe acute respiratory syndrome coro    ....................  ....................  ....................  ....................  ....................  ....................
18671 MT971754.1 |Severe acute respiratory syndrome coro    ....................  ....................  ....................  ....................  ....................  ....................
18672 MT971755.1 |Severe acute respiratory syndrome coro    ....................  ....................  ....................  ....................  ....................  ....................
18673 MT971756.1 |Severe acute respiratory syndrome coro    ....................  ....................  ....................  ....................  ....................  ....................
18674 MT971757.1 |Severe acute respiratory syndrome coro    ....................  ....................  ....................  ....................  ....................  ....................
18675 MT971758.1 |Severe acute respiratory syndrome coro    ....................  ....................  ....................  ....................  ....................  ....................
18676 MT971759.1 |Severe acute respiratory syndrome coro    ....................  ....................  ....................  ....................  ....................  ....................
18677 MT971760.1 |Severe acute respiratory syndrome coro    ....................  ....................  ....................  ....................  ....................  ....................
18678 MT971761.1 |Severe acute respiratory syndrome coro    ....................  ....................  ....................  ....................  ....................  ....................
18679 MT971762.1 |Severe acute respiratory syndrome coro    ....................  ....................  ....................  ....................  ....................  ....................
18680 MT971763.1 |Severe acute respiratory syndrome coro    ....................  ....................  ....................  ....................  ....................  ....................
18681 MT971764.1 |Severe acute respiratory syndrome coro    ....................  ....................  ....................  ....................  ....................  ....................
18682 MT971765.1 |Severe acute respiratory syndrome coro    ....................  ....................  ....................  ....................  ....................  ....................
18683 MT971766.1 |Severe acute respiratory syndrome coro    ....................  ....................  ....................  ....................  ....................  ....................
18684 MT971767.1 |Severe acute respiratory syndrome coro    ....................  ....................  ....................  ....................  ....................  ....................
18685 MT971768.1 |Severe acute respiratory syndrome coro    ....................  ....................  ....................  ....................  ....................  ....................
18686 MT971769.1 |Severe acute respiratory syndrome coro    ....................  ....................  ....................  ....................  ....................  ....................
18687 MT971770.1 |Severe acute respiratory syndrome coro    ....................  ....................  ....................  ....................  ....................  ....................
18688 MT971771.1 |Severe acute respiratory syndrome coro    ....................  ....................  ....................  ....................  ....................  ....................
18689 MT971772.1 |Severe acute respiratory syndrome coro    ....................  ....................  ....................  ....................  ....................  ....................
18690 MT971773.1 |Severe acute respiratory syndrome coro    ....................  ....................  ....................  ....................  ....................  ....................
18691 MT971774.1 |Severe acute respiratory syndrome coro    ....................  ....................  ....................  ....................  ....................  ....................
18692 MT971775.1 |Severe acute respiratory syndrome coro    ....................  ....................  ....................  ....................  ....................  ....................
18693 MT971776.1 |Severe acute respiratory syndrome coro    ....................  ....................  ....................  ....................  ....................  ....................
18694 MT971777.1 |Severe acute respiratory syndrome coro    ....................  ....................  ....................  ....................  ....................  ....................
18695 MT971778.1 |Severe acute respiratory syndrome coro    ....................  ....................  ....................  ....................  ....................  ....................
18696 MT971779.1 |Severe acute respiratory syndrome coro    ....................  ....................  ....................  ....................  ....................  ....................
18697 MT971780.1 |Severe acute respiratory syndrome coro    ....................  ....................  ....................  ....................  ....................  ....................
18698 MT971781.1 |Severe acute respiratory syndrome coro    ....................  ....................  ....................  ....................  ....................  ....................
18699 MT971782.1 |Severe acute respiratory syndrome coro    ....................  ....................  ....................  ....................  ....................  ....................
18700 MT971783.1 |Severe acute respiratory syndrome coro    ....................  ....................  ....................  ....................  ....................  ....................
```

18701 MT971784.1 |Severe acute respiratory syndrome coro .................... .................... ..................... .................... .................... ....................
18702 MT971785.1 |Severe acute respiratory syndrome coro .................... .................... ..................... .................... .................... ....................
18703 MT971786.1 |Severe acute respiratory syndrome coro .................... .................... ..................... .................... .................... ....................
18704 MT971787.1 |Severe acute respiratory syndrome coro .................... .................... ..................... .................... .................... ....................
18705 MT971788.1 |Severe acute respiratory syndrome coro .................... .................... ..................... .................... .................... ....................
18706 MT971789.1 |Severe acute respiratory syndrome coro .................... .................... ..................... .................... .................... ....................
18707 MT971790.1 |Severe acute respiratory syndrome coro .................... .................... ..................... .................... .................... ....................
18708 MT971791.1 |Severe acute respiratory syndrome coro .................... .................... ..................... .................... .................... ....................
18709 MT971792.1 |Severe acute respiratory syndrome coro .................... .................... ..................... .................... .................... ....................
18710 MT971793.1 |Severe acute respiratory syndrome coro .................... .................... ..................... .................... .................... ....................
18711 MT971794.1 |Severe acute respiratory syndrome coro .................... .................... ..................... .................... .................... ....................
18712 MT971795.1 |Severe acute respiratory syndrome coro .................... .................... ..................... .................... .................... ....................
18713 MT971796.1 |Severe acute respiratory syndrome coro .................... .................... ..................... .................... .................... ....................
18714 MT971797.1 |Severe acute respiratory syndrome coro .................... .................... ..................... .................... .................... ....................
18715 MT971798.1 |Severe acute respiratory syndrome coro .................... .................... ..................... .................... .................... ....................
18716 MT971799.1 |Severe acute respiratory syndrome coro .................... .................... ..................... .................... .................... ....................
18717 MT971800.1 |Severe acute respiratory syndrome coro .................... .................... ..................... .................... .................... ....................
18718 MT971801.1 |Severe acute respiratory syndrome coro .................... .................... ..................... .................... .................... ....................
18719 MT971802.1 |Severe acute respiratory syndrome coro .................... .................... ..................... .................... .................... ....................
18720 MT971803.1 |Severe acute respiratory syndrome coro .................... .................... ..................... .................... .................... ....................
18721 MT971804.1 |Severe acute respiratory syndrome coro .................... .................... ..................... .................... .................... ....................
18722 MT971805.1 |Severe acute respiratory syndrome coro .................... .................... ..................... .................... .................... ....................
18723 MT971806.1 |Severe acute respiratory syndrome coro .................... .................... ..................... .................... .................... ....................
18724 MT971807.1 |Severe acute respiratory syndrome coro .................... .................... ..................... .................... .................... ....................
18725 MT971808.1 |Severe acute respiratory syndrome coro .................... .................... ..................... .................... .................... ....................
18726 MT971809.1 |Severe acute respiratory syndrome coro .................... .................... ..................... .................... .................... ....................
18727 MT971810.1 |Severe acute respiratory syndrome coro .................... .................... ..................... .................... .................... ....................
18728 MT971811.1 |Severe acute respiratory syndrome coro .................... .................... ..................... .................... .................... ....................
18729 MT971812.1 |Severe acute respiratory syndrome coro .................... .................... ..................... .................... .................... ....................
18730 MT971813.1 |Severe acute respiratory syndrome coro .................... .................... ..................... .................... .................... ....................
18731 MT971814.1 |Severe acute respiratory syndrome coro .................... .................... ..................... .................... .................... ....................
18732 MT971815.1 |Severe acute respiratory syndrome coro .................... .................... ..................... .................... .................... ....................
18733 MT971816.1 |Severe acute respiratory syndrome coro .................... .................... ..................... .................... .................... ....................
18734 MT971818.1 |Severe acute respiratory syndrome coro .................... .................... ..................... .................... .................... ....................
18735 MT971819.1 |Severe acute respiratory syndrome coro .................... .................... ..................... .................... .................... ....................
18736 MT971820.1 |Severe acute respiratory syndrome coro .................... .................... ..................... .................... .................... ....................
18737 MT971821.1 |Severe acute respiratory syndrome coro .................... .................... ..................... .................... .................... ....................
18738 MT971822.1 |Severe acute respiratory syndrome coro .................... .................... ..................... .................... .................... ....................
18739 MT971823.1 |Severe acute respiratory syndrome coro .................... .................... ..................... .................... .................... ....................
18740 MT971824.1 |Severe acute respiratory syndrome coro .................... .................... ..................... .................... .................... ....................
18741 MT971826.1 |Severe acute respiratory syndrome coro .................... .................... ..................... .................... .................... ....................
18742 MT971827.1 |Severe acute respiratory syndrome coro .................... .................... ..................... .................... .................... ....................
18743 MT971828.1 |Severe acute respiratory syndrome coro .................... .................... ..................... .................... .................... ....................
18744 MT971829.1 |Severe acute respiratory syndrome coro .................... .................... ..................... .................... .................... ....................
18745 MT971830.1 |Severe acute respiratory syndrome coro .................... .................... ..................... .................... .................... ....................
18746 MT971831.1 |Severe acute respiratory syndrome coro .................... .................... ..................... .................... .................... ....................
18747 MT971832.1 |Severe acute respiratory syndrome coro .................... .................... ..................... .................... .................... ....................
18748 MT971833.1 |Severe acute respiratory syndrome coro .................... ..........A......... ..................... .................... .................... ....................
18749 MT971834.1 |Severe acute respiratory syndrome coro .................... .................... ..................... .................... .................... ....................
18750 MT971835.1 |Severe acute respiratory syndrome coro .................... .................... ..................... .................... .................... ....................
18751 MT971836.1 |Severe acute respiratory syndrome coro .................... .................... ..................... .................... .................... ....................
18752 MT971837.1 |Severe acute respiratory syndrome coro .................... .................... ..................... .................... .................... ....................
18753 MT971838.1 |Severe acute respiratory syndrome coro .................... .................... ..................... .................... .................... ....................
18754 MT971839.1 |Severe acute respiratory syndrome coro .................... .................... ..................... .................... .................... ....................
18755 MT971840.1 |Severe acute respiratory syndrome coro .................... .................... ..................... .................... .................... ....................
18756 MT971841.1 |Severe acute respiratory syndrome coro .................... .................... ..................... .................... .................... ....................
18757 MT971842.1 |Severe acute respiratory syndrome coro .................... .................... ..................... .................... .................... ....................
18758 MT971843.1 |Severe acute respiratory syndrome coro .................... .................... ..................... .................... .................... ....................
18759 MT971844.1 |Severe acute respiratory syndrome coro .................... .................... ..................... .................... .................... ....................
18760 MT971845.1 |Severe acute respiratory syndrome coro .................... .................... ..................... .................... .................... ....................
18761 MT971846.1 |Severe acute respiratory syndrome coro .................... .................... ..................... .................... .................... ....................
18762 MT971847.1 |Severe acute respiratory syndrome coro .................... .................... ..................... .................... .................... ....................
18763 MT971848.1 |Severe acute respiratory syndrome coro .................... .................... ..................... .................... .................... ....................
18764 MT971851.1 |Severe acute respiratory syndrome coro .................... .................... ..................... .................... .................... ....................
18765 MT971852.1 |Severe acute respiratory syndrome coro .................... .................... ..................... .................... .................... ....................
18766 MT971853.1 |Severe acute respiratory syndrome coro .................... .................... ..................... .................... .................... ....................
18767 MT971854.1 |Severe acute respiratory syndrome coro .................... .................... ..................... .................... .................... ....................
18768 MT971855.1 |Severe acute respiratory syndrome coro .................... .................... ..................... .................... .................... ....................
18769 MT971856.1 |Severe acute respiratory syndrome coro .................... .................... ..................... .................... .................... ....................
18770 MT971857.1 |Severe acute respiratory syndrome coro .................... .................... ..................... .................... .................... ....................
18771 MT971858.1 |Severe acute respiratory syndrome coro .................... .................... ..................... .................... .................... ....................
18772 MT971859.1 |Severe acute respiratory syndrome coro .................... .................... ..................... .................... .................... ....................
18773 MT971860.1 |Severe acute respiratory syndrome coro .................... .................... ..................... .................... .................... ....................
18774 MT971861.1 |Severe acute respiratory syndrome coro .................... .................... ..................... .................... .................... ....................
18775 MT971862.1 |Severe acute respiratory syndrome coro .................... .................... ..................... .................... .................... ....................
18776 MT971863.1 |Severe acute respiratory syndrome coro .................... .................... ..................... .................... .................... ....................
18777 MT971864.1 |Severe acute respiratory syndrome coro .................... .................... ..................... .................... .................... ....................
18778 MT971865.1 |Severe acute respiratory syndrome coro .................... .................... ..................... .................... .................... ....................

| | | | | | | | | |
|---|---|---|---|---|---|---|---|---|
| 18779 | MT971866.1 | Severe acute respiratory syndrome coro | .................... | .................... | ..................... | .................... | .................... | .................... |
| 18780 | MT971867.1 | Severe acute respiratory syndrome coro | .................... | .................... | ..................... | .................... | .................... | .................... |
| 18781 | MT971868.1 | Severe acute respiratory syndrome coro | .................... | .................... | ..................... | .................... | .................... | .................... |
| 18782 | MT971870.1 | Severe acute respiratory syndrome coro | .................... | .................... | ..................... | .................... | .................... | .................... |
| 18783 | MT971871.1 | Severe acute respiratory syndrome coro | .................... | .................... | ..................... | .................... | .................... | .................... |
| 18784 | MT971872.1 | Severe acute respiratory syndrome coro | .................... | .................... | ..................... | .................... | .................... | .................... |
| 18785 | MT971873.1 | Severe acute respiratory syndrome coro | .................... | .................... | ..................... | .................... | .................... | .................... |
| 18786 | MT971874.1 | Severe acute respiratory syndrome coro | .................... | .................... | ..................... | .................... | .................... | .................... |
| 18787 | MT971875.1 | Severe acute respiratory syndrome coro | .................... | .................... | ..................... | .................... | .................... | .................... |
| 18788 | MT971876.1 | Severe acute respiratory syndrome coro | .................... | .................... | ..................... | .................... | .................... | .................... |
| 18789 | MT971877.1 | Severe acute respiratory syndrome coro | .................... | .................... | ..................... | .................... | .................... | .................... |
| 18790 | MT971878.1 | Severe acute respiratory syndrome coro | .................... | .................... | ..................... | .................... | .................... | .................... |
| 18791 | MT971879.1 | Severe acute respiratory syndrome coro | .................... | .................... | ..................... | .................... | .................... | .................... |
| 18792 | MT971880.1 | Severe acute respiratory syndrome coro | .................... | .................... | ..................... | .................... | .................... | .................... |
| 18793 | MT971881.1 | Severe acute respiratory syndrome coro | .................... | .................... | ..................... | .................... | .................... | .................... |
| 18794 | MT971882.1 | Severe acute respiratory syndrome coro | .................... | .................... | ..................... | .................... | .................... | .................... |
| 18795 | MT971883.1 | Severe acute respiratory syndrome coro | .................... | .................... | ..................... | .................... | .................... | .................... |
| 18796 | MT971884.1 | Severe acute respiratory syndrome coro | .................... | .................... | ..................... | .................... | .................... | .................... |
| 18797 | MT971885.1 | Severe acute respiratory syndrome coro | .................... | .................... | ..................... | .................... | .................... | .................... |
| 18798 | MT971887.1 | Severe acute respiratory syndrome coro | .................... | .................... | ..................... | .................... | .................... | .................... |
| 18799 | MT971888.1 | Severe acute respiratory syndrome coro | .................... | .................... | ..................... | .................... | .................... | .................... |
| 18800 | MT971889.1 | Severe acute respiratory syndrome coro | .................... | .................... | ..................... | .................... | .................... | .................... |
| 18801 | MT971890.1 | Severe acute respiratory syndrome coro | .................... | .................... | ..................... | .................... | .................... | .................... |
| 18802 | MT971891.1 | Severe acute respiratory syndrome coro | .................... | .................... | ..................... | .................... | .................... | .................... |
| 18803 | MT971892.1 | Severe acute respiratory syndrome coro | .................... | .................... | ..................... | .................... | .................... | .................... |
| 18804 | MT971893.1 | Severe acute respiratory syndrome coro | .................... | .................... | ..................... | .................... | .................... | .................... |
| 18805 | MT971894.1 | Severe acute respiratory syndrome coro | .................... | .................... | ..................... | .................... | .................... | .................... |
| 18806 | MT971895.1 | Severe acute respiratory syndrome coro | .................... | .................... | ..................... | .................... | .................... | .................... |
| 18807 | MT971896.1 | Severe acute respiratory syndrome coro | .................... | .................... | ..................... | .................... | .................... | .................... |
| 18808 | MT971897.1 | Severe acute respiratory syndrome coro | .................... | .................... | ..................... | .................... | .................... | .................... |
| 18809 | MT971898.1 | Severe acute respiratory syndrome coro | .................... | .................... | ..................... | .................... | .................... | .................... |
| 18810 | MT971899.1 | Severe acute respiratory syndrome coro | .................... | .................... | ..................... | .................... | .................... | .................... |
| 18811 | MT971900.1 | Severe acute respiratory syndrome coro | .................... | .................... | ..................... | .................... | .................... | .................... |
| 18812 | MT971901.1 | Severe acute respiratory syndrome coro | .................... | .................... | ..................... | .................... | .................... | .................... |
| 18813 | MT971902.1 | Severe acute respiratory syndrome coro | .................... | .................... | ..................... | .................... | .................... | .................... |
| 18814 | MT971903.1 | Severe acute respiratory syndrome coro | .................... | .................... | ..................... | .................... | .................... | .................... |
| 18815 | MT971905.1 | Severe acute respiratory syndrome coro | .................... | .................... | ..................... | .................... | .................... | .................... |
| 18816 | MT971906.1 | Severe acute respiratory syndrome coro | .................... | .................... | ..................... | .................... | .................... | .................... |
| 18817 | MT971907.1 | Severe acute respiratory syndrome coro | .................... | .................... | ..................... | .................... | .................... | .................... |
| 18818 | MT971908.1 | Severe acute respiratory syndrome coro | .................... | .................... | ..................... | .................... | .................... | .................... |
| 18819 | MT971909.1 | Severe acute respiratory syndrome coro | .................... | .................... | ..................... | .................... | .................... | .................... |
| 18820 | MT971910.1 | Severe acute respiratory syndrome coro | .................... | .................... | ..................... | .................... | .................... | .................... |
| 18821 | MT971912.1 | Severe acute respiratory syndrome coro | .................... | .................... | ..................... | .................... | .................... | .................... |
| 18822 | MT971913.1 | Severe acute respiratory syndrome coro | .................... | .................... | ..................... | .................... | .................... | .................... |
| 18823 | MT971914.1 | Severe acute respiratory syndrome coro | .................... | .................... | ..................... | .................... | .................... | .................... |
| 18824 | MT971915.1 | Severe acute respiratory syndrome coro | .................... | .................... | ..................... | .................... | .................... | .................... |
| 18825 | MT971916.1 | Severe acute respiratory syndrome coro | .................... | .................... | ..................... | .................... | .................... | .................... |
| 18826 | MT971917.1 | Severe acute respiratory syndrome coro | .................... | .................... | ..................... | .................... | .................... | .................... |
| 18827 | MT971918.1 | Severe acute respiratory syndrome coro | .................... | .................... | ..................... | .................... | .................... | .................... |
| 18828 | MT971919.1 | Severe acute respiratory syndrome coro | .................... | .................... | ..................... | .................... | .................... | .................... |
| 18829 | MT971920.1 | Severe acute respiratory syndrome coro | .................... | .................... | ..................... | .................... | .................... | .................... |
| 18830 | MT971921.1 | Severe acute respiratory syndrome coro | .................... | .................... | ..................... | .................... | .................... | .................... |
| 18831 | MT971925.1 | Severe acute respiratory syndrome coro | .................... | .................... | ..................... | .................... | .................... | .................... |
| 18832 | MT971926.1 | Severe acute respiratory syndrome coro | .................... | .................... | ..................... | .................... | .................... | .................... |
| 18833 | MT971927.1 | Severe acute respiratory syndrome coro | .................... | .................... | ..................... | .................... | .................... | .................... |
| 18834 | MT971930.1 | Severe acute respiratory syndrome coro | .................... | .................... | ..................... | .................... | .................... | .................... |
| 18835 | MT971931.1 | Severe acute respiratory syndrome coro | .................... | .................... | ..................... | .................... | .................... | .................... |
| 18836 | MT971932.1 | Severe acute respiratory syndrome coro | .................... | .................... | ..................... | .................... | .................... | .................... |
| 18837 | MT971933.1 | Severe acute respiratory syndrome coro | .................... | .................... | ..................... | .................... | .................... | .................... |
| 18838 | MT971934.1 | Severe acute respiratory syndrome coro | .................... | .................... | ..................... | .................... | .................... | .................... |
| 18839 | MT971935.1 | Severe acute respiratory syndrome coro | .................... | .................... | ..................... | .................... | .................... | .................... |
| 18840 | MT971936.1 | Severe acute respiratory syndrome coro | .................... | .................... | ..................... | .................... | .................... | .................... |
| 18841 | MT971937.1 | Severe acute respiratory syndrome coro | .................... | .................... | ..................... | .................... | .................... | .................... |
| 18842 | MT971938.1 | Severe acute respiratory syndrome coro | .................... | .................... | ..................... | .................... | .................... | .................... |
| 18843 | MT971939.1 | Severe acute respiratory syndrome coro | .................... | .................... | ..................... | .................... | .................... | .................... |
| 18844 | MT971940.1 | Severe acute respiratory syndrome coro | .................... | .................... | ..................... | .................... | .................... | .................... |
| 18845 | MT971942.1 | Severe acute respiratory syndrome coro | .................... | .................... | ..................... | .................... | .................... | .................... |
| 18846 | MT971943.1 | Severe acute respiratory syndrome coro | .................... | .................... | ..................... | .................... | .................... | .................... |
| 18847 | MT971945.1 | Severe acute respiratory syndrome coro | .................... | .................... | ..................... | .................... | .................... | .................... |
| 18848 | MT971946.1 | Severe acute respiratory syndrome coro | .................... | .................... | ..................... | .................... | .................... | .................... |
| 18849 | MT971947.1 | Severe acute respiratory syndrome coro | .................... | .................... | ..................... | .................... | .................... | .................... |
| 18850 | MT971948.1 | Severe acute respiratory syndrome coro | .................... | .................... | ..................... | .................... | .................... | .................... |
| 18851 | MT971949.1 | Severe acute respiratory syndrome coro | .................... | .................... | ..................... | .................... | .................... | .................... |
| 18852 | MT971950.1 | Severe acute respiratory syndrome coro | .................... | .................... | ..................... | .................... | .................... | .................... |
| 18853 | MT971951.1 | Severe acute respiratory syndrome coro | .................... | .................... | ..................... | .................... | .................... | .................... |
| 18854 | MT971952.1 | Severe acute respiratory syndrome coro | .................... | .................... | ..................... | .................... | .................... | .................... |
| 18855 | MT971953.1 | Severe acute respiratory syndrome coro | .................... | .................... | ..................... | .................... | .................... | .................... |
| 18856 | MT971954.1 | Severe acute respiratory syndrome coro | .................... | .................... | ..................... | .................... | .................... | .................... |

18857 MT971957.1 |Severe acute respiratory syndrome coro ···················· ···················· ···················· ···················· ···················· ····················
18858 MT971958.1 |Severe acute respiratory syndrome coro ···················· ···················· ···················· ···················· ···················· ····················
18859 MT971959.1 |Severe acute respiratory syndrome coro ···················· ···················· ···················· ···················· ···················· ····················
18860 MT971960.1 |Severe acute respiratory syndrome coro ···················· ···················· ···················· ···················· ···················· ····················
18861 MT971961.1 |Severe acute respiratory syndrome coro ···················· ···················· ···················· ···················· ···················· ····················
18862 MT971962.1 |Severe acute respiratory syndrome coro ···················· ···················· ···················· ···················· ···················· ····················
18863 MT971963.1 |Severe acute respiratory syndrome coro ···················· ···················· ···················· ···················· ···················· ····················
18864 MT971964.1 |Severe acute respiratory syndrome coro ···················· ···················· ···················· ···················· ···················· ····················
18865 MT971965.1 |Severe acute respiratory syndrome coro ···················· ···················· ···················· ···················· ···················· ····················
18866 MT971966.1 |Severe acute respiratory syndrome coro ···················· ···················· ···················· ···················· ···················· ····················
18867 MT971967.1 |Severe acute respiratory syndrome coro ···················· ···················· ···················· ···················· ···················· ····················
18868 MT971968.1 |Severe acute respiratory syndrome coro ···················· ···················· ···················· ···················· ···················· ····················
18869 MT971969.1 |Severe acute respiratory syndrome coro ···················· ···················· ···················· ···················· ···················· ····················
18870 MT971970.1 |Severe acute respiratory syndrome coro ···················· ···················· ···················· ···················· ···················· ····················
18871 MT971971.1 |Severe acute respiratory syndrome coro ···················· ···················· ···················· ···················· ···················· ····················
18872 MT971972.1 |Severe acute respiratory syndrome coro ···················· ···················· ···················· ···················· ···················· ····················
18873 MT971973.1 |Severe acute respiratory syndrome coro ···················· ···················· ···················· ···················· ···················· ····················
18874 MT971974.1 |Severe acute respiratory syndrome coro ···················· ···················· ···················· ···················· ···················· ····················
18875 MT971975.1 |Severe acute respiratory syndrome coro ···················· ···················· ···················· ···················· ···················· ····················
18876 MT971976.1 |Severe acute respiratory syndrome coro ···················· ···················· ···················· ···················· ···················· ····················
18877 MT971977.1 |Severe acute respiratory syndrome coro ···················· ···················· ···················· ···················· ···················· ····················
18878 MT971978.1 |Severe acute respiratory syndrome coro ···················· ···················· ···················· ···················· ···················· ····················
18879 MT971979.1 |Severe acute respiratory syndrome coro ···················· ···················· ···················· ···················· ···················· ····················
18880 MT971981.1 |Severe acute respiratory syndrome coro ···················· ···················· ···················· ···················· ···················· ····················
18881 MT971982.1 |Severe acute respiratory syndrome coro ···················· ···················· ···················· ···················· ···················· ····················
18882 MT971983.1 |Severe acute respiratory syndrome coro ···················· ···················· ···················· ···················· ···················· ····················
18883 MT971984.1 |Severe acute respiratory syndrome coro ···················· ···················· ···················· ···················· ···················· ····················
18884 MT971985.1 |Severe acute respiratory syndrome coro ···················· ···················· ···················· ···················· ···················· ····················
18885 MT971986.1 |Severe acute respiratory syndrome coro ···················· ···················· ···················· ···················· ···················· ····················
18886 MT971987.1 |Severe acute respiratory syndrome coro ···················· ···················· ···················· ···················· ···················· ····················
18887 MT971988.1 |Severe acute respiratory syndrome coro ···················· ···················· ···················· ···················· ···················· ····················
18888 MT971989.1 |Severe acute respiratory syndrome coro ···················· ···················· ···················· ···················· ···················· ····················
18889 MT971990.1 |Severe acute respiratory syndrome coro ···················· ···················· ···················· ···················· ···················· ····················
18890 MT971991.1 |Severe acute respiratory syndrome coro ···················· ···················· ···················· ···················· ···················· ····················
18891 MT971992.1 |Severe acute respiratory syndrome coro ···················· ···················· ···················· ···················· ···················· ····················
18892 MT971993.1 |Severe acute respiratory syndrome coro ···················· ···················· ···················· ···················· ···················· ····················
18893 MT971994.1 |Severe acute respiratory syndrome coro ···················· ···················· ···················· ···················· ···················· ····················
18894 MT971995.1 |Severe acute respiratory syndrome coro ···················· ···················· ···················· ···················· ···················· ····················
18895 MT971996.1 |Severe acute respiratory syndrome coro ···················· ···················· ···················· ···················· ···················· ····················
18896 MT971997.1 |Severe acute respiratory syndrome coro ···················· ···················· ···················· ···················· ···················· ····················
18897 MT971998.1 |Severe acute respiratory syndrome coro ···················· ···················· ···················· ···················· ···················· ····················
18898 MT971999.1 |Severe acute respiratory syndrome coro ···················· ···················· ···················· ···················· ···················· ····················
18899 MT972000.1 |Severe acute respiratory syndrome coro ···················· ···················· ···················· ···················· ···················· ····················
18900 MT972001.1 |Severe acute respiratory syndrome coro ···················· ···················· ···················· ···················· ···················· ····················
18901 MT972002.1 |Severe acute respiratory syndrome coro ···················· ···················· ···················· ···················· ···················· ····················
18902 MT972003.1 |Severe acute respiratory syndrome coro ···················· ···················· ···················· ···················· ···················· ····················
18903 MT972004.1 |Severe acute respiratory syndrome coro ···················· ···················· ···················· ···················· ···················· ····················
18904 MT972005.1 |Severe acute respiratory syndrome coro ···················· ···················· ···················· ···················· ···················· ····················
18905 MT972006.1 |Severe acute respiratory syndrome coro ···················· ···················· ···················· ···················· ···················· ····················
18906 MT972007.1 |Severe acute respiratory syndrome coro ···················· ···················· ···················· ···················· ···················· ····················
18907 MT972008.1 |Severe acute respiratory syndrome coro ···················· ···················· ···················· ···················· ···················· ····················
18908 MT972009.1 |Severe acute respiratory syndrome coro ···················· ···················· ···················· ···················· ···················· ····················
18909 MT972010.1 |Severe acute respiratory syndrome coro ···················· ···················· ···················· ···················· ···················· ····················
18910 MT972011.1 |Severe acute respiratory syndrome coro ···················· ···················· ···················· ···················· ···················· ····················
18911 MT972012.1 |Severe acute respiratory syndrome coro ···················· ···················· ···················· ···················· ···················· ····················
18912 MT972013.1 |Severe acute respiratory syndrome coro ···················· ···················· ···················· ···················· ···················· ····················
18913 MT972014.1 |Severe acute respiratory syndrome coro ···················· ···················· ···················· ···················· ···················· ····················
18914 MT972015.1 |Severe acute respiratory syndrome coro ···················· ···················· ···················· ···················· ···················· ····················
18915 MT972016.1 |Severe acute respiratory syndrome coro ···················· ···················· ···················· ···················· ···················· ····················
18916 MT972017.1 |Severe acute respiratory syndrome coro ···················· ···················· ···················· ···················· ···················· ····················
18917 MT972018.1 |Severe acute respiratory syndrome coro ···················· ···················· ···················· ···················· ···················· ····················
18918 MT972019.1 |Severe acute respiratory syndrome coro ···················· ···················· ···················· ···················· ···················· ····················
18919 MT972020.1 |Severe acute respiratory syndrome coro ···················· ···················· ···················· ···················· ···················· ····················
18920 MT972022.1 |Severe acute respiratory syndrome coro ···················· ···················· ···················· ···················· ···················· ····················
18921 MT972023.1 |Severe acute respiratory syndrome coro ···················· ···················· ···················· ···················· ···················· ····················
18922 MT972024.1 |Severe acute respiratory syndrome coro ···················· ···················· ···················· ···················· ···················· ····················
18923 MT972026.1 |Severe acute respiratory syndrome coro ···················· ···················· ···················· ···················· ···················· ····················
18924 MT972027.1 |Severe acute respiratory syndrome coro ···················· ······A············· ···················· ···················· ···················· ····················
18925 MT972028.1 |Severe acute respiratory syndrome coro ···················· ···················· ···················· ···················· ···················· ····················
18926 MT972029.1 |Severe acute respiratory syndrome coro ···················· ···················· ···················· ···················· ···················· ····················
18927 MT972030.1 |Severe acute respiratory syndrome coro ···················· ···················· ···················· ···················· ···················· ····················
18928 MT972031.1 |Severe acute respiratory syndrome coro ···················· ···················· ···················· ···················· ···················· ····················
18929 MT972032.1 |Severe acute respiratory syndrome coro ···················· ···················· ···················· ···················· ···················· ····················
18930 MT972033.1 |Severe acute respiratory syndrome coro ···················· ···················· ···················· ···················· ···················· ····················
18931 MT972034.1 |Severe acute respiratory syndrome coro ···················· ···················· ···················· ···················· ···················· ····················
18932 MT972035.1 |Severe acute respiratory syndrome coro ···················· ···················· ···················· ···················· ···················· ····················
18933 MT972036.1 |Severe acute respiratory syndrome coro ···················· ···················· ···················· ···················· ···················· ····················
18934 MT972037.1 |Severe acute respiratory syndrome coro ···················· ···················· ···················· ···················· ···················· ····················

```
18935 MT972038.1 |Severe acute respiratory syndrome coro    ····················  ····················  ····················  ····················  ····················  ····················
18936 MT972039.1 |Severe acute respiratory syndrome coro    ····················  ····················  ····················  ····················  ····················  ····················
18937 MT972040.1 |Severe acute respiratory syndrome coro    ····················  ····················  ····················  ····················  ····················  ····················
18938 MT972041.1 |Severe acute respiratory syndrome coro    ····················  ····················  ····················  ····················  ····················  ····················
18939 MT972042.1 |Severe acute respiratory syndrome coro    ····················  ····················  ····················  ····················  ····················  ····················
18940 MT972044.1 |Severe acute respiratory syndrome coro    ····················  ·····A··············  ····················  ····················  ····················  ····················
18941 MT972045.1 |Severe acute respiratory syndrome coro    ····················  ·····A··············  ····················  ····················  ····················  ····················
18942 MT972046.1 |Severe acute respiratory syndrome coro    ····················  ····················  ····················  ····················  ····················  ····················
18943 MT972047.1 |Severe acute respiratory syndrome coro    ····················  ····················  ····················  ····················  ····················  ····················
18944 MT972048.1 |Severe acute respiratory syndrome coro    ····················  ····················  ····················  ····················  ····················  ····················
18945 MT972049.1 |Severe acute respiratory syndrome coro    ····················  ····················  ····················  ····················  ····················  ····················
18946 MT972050.1 |Severe acute respiratory syndrome coro    ····················  ·····A··············  ····················  ····················  ····················  ····················
18947 MT972051.1 |Severe acute respiratory syndrome coro    ····················  ·····A··············  ····················  ····················  ····················  ····················
18948 MT972052.1 |Severe acute respiratory syndrome coro    ····················  ····················  ····················  ····················  ····················  ····················
18949 MT972053.1 |Severe acute respiratory syndrome coro    ····················  ····················  ····················  ····················  ····················  ····················
18950 MT972054.1 |Severe acute respiratory syndrome coro    ····················  ····················  ····················  ····················  ····················  ····················
18951 MT972056.1 |Severe acute respiratory syndrome coro    ····················  ····················  ····················  ····················  ····················  ····················
18952 MT972057.1 |Severe acute respiratory syndrome coro    ····················  ····················  ····················  ····················  ····················  ····················
18953 MT972058.1 |Severe acute respiratory syndrome coro    ····················  ····················  ····················  ····················  ····················  ····················
18954 MT972059.1 |Severe acute respiratory syndrome coro    ····················  ····················  ····················  ····················  ····················  ····················
18955 MT972060.1 |Severe acute respiratory syndrome coro    ····················  ·····A··············  ····················  ····················  ····················  ····················
18956 MT972061.1 |Severe acute respiratory syndrome coro    ····················  ····················  ····················  ····················  ····················  ····················
18957 MT972062.1 |Severe acute respiratory syndrome coro    ····················  ····················  ····················  ····················  ····················  ····················
18958 MT972063.1 |Severe acute respiratory syndrome coro    ····················  ····················  ····················  ····················  ····················  ····················
18959 MT972064.1 |Severe acute respiratory syndrome coro    ····················  ····················  ····················  ····················  ····················  ····················
18960 MT972065.1 |Severe acute respiratory syndrome coro    ····················  ····················  ····················  ····················  ····················  ····················
18961 MT972066.1 |Severe acute respiratory syndrome coro    ····················  ····················  ····················  ····················  ····················  ····················
18962 MT972067.1 |Severe acute respiratory syndrome coro    ····················  ····················  ····················  ····················  ····················  ····················
18963 MT972068.1 |Severe acute respiratory syndrome coro    ····················  ····················  ····················  ····················  ····················  ····················
18964 MT972069.1 |Severe acute respiratory syndrome coro    ····················  ····················  ····················  ····················  ····················  ····················
18965 MT972070.1 |Severe acute respiratory syndrome coro    ····················  ····················  ····················  ····················  ····················  ····················
18966 MT972071.1 |Severe acute respiratory syndrome coro    ····················  ····················  ····················  ····················  ····················  ····················
18967 MT972072.1 |Severe acute respiratory syndrome coro    ····················  ····················  ····················  ····················  ····················  ····················
18968 MT972073.1 |Severe acute respiratory syndrome coro    ····················  ····················  ····················  ····················  ····················  ····················
18969 MT972075.1 |Severe acute respiratory syndrome coro    ····················  ····················  ····················  ····················  ····················  ····················
18970 MT972076.1 |Severe acute respiratory syndrome coro    ····················  ····················  ····················  ····················  ····················  ····················
18971 MT972077.1 |Severe acute respiratory syndrome coro    ····················  ····················  ····················  ····················  ····················  ····················
18972 MT972078.1 |Severe acute respiratory syndrome coro    ····················  ····················  ····················  ····················  ····················  ····················
18973 MT972079.1 |Severe acute respiratory syndrome coro    ····················  ····················  ····················  ····················  ····················  ····················
18974 MT972080.1 |Severe acute respiratory syndrome coro    ····················  ····················  ····················  ····················  ····················  ····················
18975 MT972081.1 |Severe acute respiratory syndrome coro    ····················  ····················  ····················  ····················  ····················  ····················
18976 MT972083.1 |Severe acute respiratory syndrome coro    ····················  ····················  ····················  ····················  ····················  ····················
18977 MT972085.1 |Severe acute respiratory syndrome coro    ····················  ····················  ····················  ····················  ····················  ····················
18978 MT972086.1 |Severe acute respiratory syndrome coro    ····················  ····················  ····················  ····················  ····················  ····················
18979 MT972087.1 |Severe acute respiratory syndrome coro    ····················  ····················  ····················  ····················  ····················  ····················
18980 MT972088.1 |Severe acute respiratory syndrome coro    ····················  ····················  ····················  ····················  ····················  ····················
18981 MT972089.1 |Severe acute respiratory syndrome coro    ····················  ····················  ····················  ····················  ····················  ····················
18982 MT972090.1 |Severe acute respiratory syndrome coro    ····················  ····················  ····················  ····················  ····················  ····················
18983 MT972091.1 |Severe acute respiratory syndrome coro    ····················  ····················  ····················  ····················  ····················  ····················
18984 MT972092.1 |Severe acute respiratory syndrome coro    ····················  ····················  ····················  ····················  ····················  ····················
18985 MT972093.1 |Severe acute respiratory syndrome coro    ····················  ····················  ····················  ····················  ····················  ····················
18986 MT972094.1 |Severe acute respiratory syndrome coro    ····················  ····················  ····················  ····················  ····················  ····················
18987 MT972095.1 |Severe acute respiratory syndrome coro    ····················  ····················  ····················  ····················  ····················  ····················
18988 MT972096.1 |Severe acute respiratory syndrome coro    ····················  ····················  ····················  ····················  ····················  ····················
18989 MT972098.1 |Severe acute respiratory syndrome coro    ····················  ····················  ····················  ····················  ····················  ····················
18990 MT972099.1 |Severe acute respiratory syndrome coro    ····················  ····················  ····················  ····················  ····················  ····················
18992 MT972111.1 |Severe acute respiratory syndrome coro    ····················  ·A··················  ····················  ····················  ····················  ··A·················
18993 MT972112.1 |Severe acute respiratory syndrome coro    ····················  ····················  ····················  ····················  ····················  ····················
18994 MT972113.1 |Severe acute respiratory syndrome coro    ····················  ·A··················  ····················  ····················  ····················  ··A·················
18995 MT972114.1 |Severe acute respiratory syndrome coro    ····················  ····················  ····················  ····················  ····················  ····················
18996 MT972115.1 |Severe acute respiratory syndrome coro    ····················  ····················  ····················  ····················  ····················  ····················
18997 MT972116.1 |Severe acute respiratory syndrome coro    ····················  ····················  ····················  ····················  ····················  ····················
18998 MT972117.1 |Severe acute respiratory syndrome coro    ····················  ····················  ····················  ····················  ····················  ····················
18999 MT972118.1 |Severe acute respiratory syndrome coro    ····················  ····················  ····················  ····················  ····················  ····················
19000 MT972119.1 |Severe acute respiratory syndrome coro    ····················  ····················  ····················  ····················  ····················  ····················
19001 MT972120.1 |Severe acute respiratory syndrome coro    ····················  ····················  ····················  ····················  ····················  ····················
19002 MT972121.1 |Severe acute respiratory syndrome coro    ····················  ····················  ····················  ····················  ····················  ····················
19003 MT972122.1 |Severe acute respiratory syndrome coro    ····················  ····················  ····················  ····················  ····················  ····················
19004 MT972123.1 |Severe acute respiratory syndrome coro    ····················  ····················  ····················  ····················  ····················  ····················
19006 MT972128.1 |Severe acute respiratory syndrome coro    ····················  ····················  ····················  ····················  ····················  ····················
19007 MT972129.1 |Severe acute respiratory syndrome coro    ····················  ····················  ····················  ····················  ····················  ····················
19008 MT972130.1 |Severe acute respiratory syndrome coro    ····················  ····················  ····················  ····················  ····················  ····················
19010 MT972132.1 |Severe acute respiratory syndrome coro    ····················  ····················  ····················  ····················  ····················  ····················
19011 MT972133.1 |Severe acute respiratory syndrome coro    ····················  ····················  ····················  ····················  ····················  ····················
19012 MT972135.1 |Severe acute respiratory syndrome coro    ····················  ····················  ····················  ····················  ····················  ····················
19013 MT972136.1 |Severe acute respiratory syndrome coro    ····················  ····················  ····················  ····················  ····················  ····················
19014 MT972137.1 |Severe acute respiratory syndrome coro    ····················  ····················  ····················  ····················  ····················  ····················
19015 MT972139.1 |Severe acute respiratory syndrome coro    ····················  ····················  ····················  ····················  ····················  ····················
```

| | | | | | | | | | |
|---|---|---|---|---|---|---|---|---|---|
| 19016 | MT972140.1 | Severe acute respiratory syndrome coro | ···················· | ···················· | ···················· | ···················· | ···················· | ···················· | |
| 19017 | MT972141.1 | Severe acute respiratory syndrome coro | ···················· | ···················· | ···················· | ···················· | ···················· | ···················· | |
| 19018 | MT972142.1 | Severe acute respiratory syndrome coro | ···················· | ···················· | ···················· | ···················· | ···················· | ···················· | |
| 19019 | MT972143.1 | Severe acute respiratory syndrome coro | ···················· | ···················· | ···················· | ···················· | ···················· | ···················· | |
| 19020 | MT972144.1 | Severe acute respiratory syndrome coro | ···················· | ···················· | ···················· | ···················· | ···················· | ···················· | |
| 19021 | MT972145.1 | Severe acute respiratory syndrome coro | ···················· | ···················· | ···················· | ···················· | ···················· | ···················· | |
| 19022 | MT972147.1 | Severe acute respiratory syndrome coro | ···················· | ···················· | ···················· | ···················· | ···················· | ···················· | |
| 19023 | MT972148.1 | Severe acute respiratory syndrome coro | ···················· | ···················· | ···················· | ···················· | ···················· | ···················· | |
| 19024 | MT972149.1 | Severe acute respiratory syndrome coro | ···················· | ···················· | ···················· | ···················· | ···················· | ···················· | |
| 19025 | MT972150.1 | Severe acute respiratory syndrome coro | ···················· | ···················· | ···················· | ···················· | ···················· | ···················· | |
| 19026 | MT972151.1 | Severe acute respiratory syndrome coro | ···················· | ···················· | ···················· | ···················· | ···················· | ···················· | |
| 19027 | MT972152.1 | Severe acute respiratory syndrome coro | ···················· | ···················· | ···················· | ···················· | ···················· | ···················· | |
| 19028 | MT972153.1 | Severe acute respiratory syndrome coro | ···················· | ···················· | ···················· | ···················· | ···················· | ···················· | |
| 19029 | MT972155.1 | Severe acute respiratory syndrome coro | ···················· | ···················· | ···················· | ···················· | ···················· | ···················· | |
| 19030 | MT972158.1 | Severe acute respiratory syndrome coro | ···················· | ···················· | ···················· | ···················· | ···················· | ···················· | |
| 19031 | MT972159.1 | Severe acute respiratory syndrome coro | ···················· | ···················· | ···················· | ···················· | ···················· | ···················· | |
| 19032 | MT972160.1 | Severe acute respiratory syndrome coro | ···················· | ···················· | ···················· | ···················· | ···················· | ···················· | |
| 19033 | MT972161.1 | Severe acute respiratory syndrome coro | ···················· | ·A·················· | ···················· | ···················· | ···················· | ··A················· | |
| 19034 | MT972162.1 | Severe acute respiratory syndrome coro | ···················· | ···················· | ···················· | ···················· | ···················· | ···················· | |
| 19035 | MT972163.1 | Severe acute respiratory syndrome coro | ···················· | ···················· | ···················· | ···················· | ···················· | ···················· | |
| 19036 | MT972164.1 | Severe acute respiratory syndrome coro | ···················· | ···················· | ···················· | ···················· | ···················· | ···················· | |
| 19037 | MT972165.1 | Severe acute respiratory syndrome coro | ···················· | ···················· | ···················· | ···················· | ···················· | ···················· | |
| 19038 | MT972166.1 | Severe acute respiratory syndrome coro | ···················· | ···················· | ···················· | ···················· | ···················· | ···················· | |
| 19039 | MT972168.1 | Severe acute respiratory syndrome coro | ···················· | ···················· | ···················· | ···················· | ···················· | ···················· | |
| 19040 | MT972171.1 | Severe acute respiratory syndrome coro | ···················· | ···················· | ···················· | ···················· | ···················· | ···················· | |
| 19041 | MT972172.1 | Severe acute respiratory syndrome coro | ···················· | ···················· | ···················· | ···················· | ···················· | ···················· | |
| 19042 | MT972173.1 | Severe acute respiratory syndrome coro | ···················· | ···················· | ···················· | ···················· | NNN················· | ···················· | 1 |
| 19043 | MT972174.1 | Severe acute respiratory syndrome coro | ···················· | ···················· | ···················· | ···················· | ··NN················ | ···················· | |
| 19047 | MT972178.1 | Severe acute respiratory syndrome coro | ···················· | ···················· | ···················· | ···················· | ···················· | ···················· | |
| 19048 | MT972181.1 | Severe acute respiratory syndrome coro | ···················· | ···················· | ···················· | ···················· | NNN················· | ···················· | 1 |
| 19049 | MT972182.1 | Severe acute respiratory syndrome coro | ···················· | ·A·················· | ···················· | ···················· | ···················· | ··A················· | |
| 19050 | MT972184.1 | Severe acute respiratory syndrome coro | ···················· | ···················· | ···················· | ···················· | ···················· | ···················· | |
| 19051 | MT972185.1 | Severe acute respiratory syndrome coro | ···················· | ···················· | ···················· | ···················· | ···················· | ·NNN················ | 1 |
| 19052 | MT972186.1 | Severe acute respiratory syndrome coro | ···················· | ···················· | ···················· | ···················· | ···················· | ···················· | |
| 19053 | MT972187.1 | Severe acute respiratory syndrome coro | ···················· | ···················· | ···················· | ···················· | ···················· | ···················· | |
| 19054 | MT972188.1 | Severe acute respiratory syndrome coro | ···················· | ···················· | ···················· | ···················· | ···················· | ···················· | |
| 19055 | MT972189.1 | Severe acute respiratory syndrome coro | ···················· | ···················· | ···················· | ···················· | ···················· | ···················· | |
| 19056 | MT972191.1 | Severe acute respiratory syndrome coro | ···················· | ···················· | ···················· | ···················· | ···················· | ···················· | |
| 19057 | MT972192.1 | Severe acute respiratory syndrome coro | ···················· | ···················· | ···················· | ···················· | ···················· | ···················· | |
| 19058 | MT972193.1 | Severe acute respiratory syndrome coro | ···················· | ···················· | ···················· | ···················· | ···················· | ···················· | |
| 19059 | MT972194.1 | Severe acute respiratory syndrome coro | ···················· | ···················· | ···················· | ···················· | ···················· | ···················· | |
| 19060 | MT972195.1 | Severe acute respiratory syndrome coro | ···················· | ···················· | ···················· | ···················· | ···················· | ···················· | |
| 19061 | MT972196.1 | Severe acute respiratory syndrome coro | ···················· | ···················· | ···················· | ···················· | ···················· | ···················· | |
| 19062 | MT972197.1 | Severe acute respiratory syndrome coro | ···················· | ···················· | ···················· | ···················· | ···················· | ···················· | |
| 19063 | MT972198.1 | Severe acute respiratory syndrome coro | ···················· | ···················· | ···················· | ···················· | ···················· | ···················· | |
| 19064 | MT972199.1 | Severe acute respiratory syndrome coro | ···················· | ···················· | ···················· | ···················· | ···················· | ···················· | |
| 19065 | MT972200.1 | Severe acute respiratory syndrome coro | ···················· | ···················· | ···················· | ···················· | ···················· | ···················· | |
| 19066 | MT972201.1 | Severe acute respiratory syndrome coro | ···················· | ···················· | ···················· | ···················· | ···················· | ···················· | |
| 19067 | MT972202.1 | Severe acute respiratory syndrome coro | ···················· | ···················· | ···················· | ···················· | ···················· | ···················· | |
| 19068 | MT972203.1 | Severe acute respiratory syndrome coro | ···················· | ···················· | ···················· | ···················· | ···················· | ···················· | |
| 19069 | MT972204.1 | Severe acute respiratory syndrome coro | ···················· | ···················· | ···················· | ···················· | NNN················· | ···················· | 1 |
| 19070 | MT972205.1 | Severe acute respiratory syndrome coro | ···················· | ···················· | ···················· | ···················· | ···················· | ···················· | |
| 19072 | MT972207.1 | Severe acute respiratory syndrome coro | ···················· | ···················· | ···················· | ···················· | ···················· | ···················· | |
| 19073 | MT972209.1 | Severe acute respiratory syndrome coro | ···················· | ···················· | ···················· | ···················· | ···················· | ···················· | |
| 19074 | MT972210.1 | Severe acute respiratory syndrome coro | ···················· | ···················· | ···················· | ···················· | ···················· | ···················· | |
| 19075 | MT972211.1 | Severe acute respiratory syndrome coro | ···················· | ···················· | ···················· | ···················· | ···················· | ···················· | |
| 19076 | MT972212.1 | Severe acute respiratory syndrome coro | ···················· | ···················· | ···················· | ···················· | ···················· | ···················· | |
| 19078 | MT972214.1 | Severe acute respiratory syndrome coro | ···················· | ···················· | ···················· | ···················· | ···················· | ···················· | |
| 19079 | MT972215.1 | Severe acute respiratory syndrome coro | ···················· | ···················· | ···················· | ···················· | ···················· | ···················· | |
| 19080 | MT972216.1 | Severe acute respiratory syndrome coro | ···················· | ···················· | ···················· | ···················· | ···················· | ···················· | |
| 19081 | MT972217.1 | Severe acute respiratory syndrome coro | ···················· | ···················· | ···················· | ···················· | ···················· | ···················· | |
| 19082 | MT972220.1 | Severe acute respiratory syndrome coro | ···················· | ···················· | ···················· | ···················· | ···················· | ···················· | |
| 19083 | MT972221.1 | Severe acute respiratory syndrome coro | ···················· | ···················· | ···················· | ···················· | ···················· | ···················· | |
| 19084 | MT972223.1 | Severe acute respiratory syndrome coro | ···················· | ···················· | ···················· | ···················· | ···················· | ···················· | |
| 19085 | MT972224.1 | Severe acute respiratory syndrome coro | ···················· | ···················· | ···················· | ···················· | ···················· | ···················· | |
| 19086 | MT972225.1 | Severe acute respiratory syndrome coro | ···················· | ···················· | ···················· | ···················· | ···················· | ···················· | |
| 19087 | MT972226.1 | Severe acute respiratory syndrome coro | ···················· | ···················· | ···················· | ···················· | ···················· | ···················· | |
| 19088 | MT972227.1 | Severe acute respiratory syndrome coro | ···················· | ···················· | ···················· | ···················· | ···················· | ···················· | |
| 19089 | MT972228.1 | Severe acute respiratory syndrome coro | ···················· | ···················· | ···················· | ···················· | ···················· | ···················· | |
| 19090 | MT972231.1 | Severe acute respiratory syndrome coro | ···················· | ···················· | ···················· | ···················· | ···················· | ···················· | |
| 19091 | MT972232.1 | Severe acute respiratory syndrome coro | ···················· | ···················· | ···················· | ···················· | ···················· | ···················· | |
| 19092 | MT972233.1 | Severe acute respiratory syndrome coro | ···················· | ···················· | ···················· | ···················· | ···················· | ···················· | |
| 19093 | MT972234.1 | Severe acute respiratory syndrome coro | ···················· | ···················· | ···················· | ···················· | ···················· | ···················· | |
| 19094 | MT972235.1 | Severe acute respiratory syndrome coro | ···················· | ···················· | ···················· | ···················· | ···················· | ···················· | |
| 19095 | MT972236.1 | Severe acute respiratory syndrome coro | ···················· | ···················· | ···················· | ···················· | ···················· | ···················· | |
| 19096 | MT972237.1 | Severe acute respiratory syndrome coro | ···················· | ···················· | ···················· | ···················· | ···················· | ···················· | |
| 19097 | MT972238.1 | Severe acute respiratory syndrome coro | ···················· | ·A·················· | ···················· | ···················· | ···················· | ··A················· | |
| 19098 | MT972239.1 | Severe acute respiratory syndrome coro | ···················· | ···················· | ···················· | ···················· | ···················· | ···················· | |

| | | | | | | | | | |
|---|---|---|---|---|---|---|---|---|---|
| 19099 | MT972241.1 | \|Severe acute respiratory syndrome coro | ···················· | ···················· | ···················· | ···················· | ···················· | ···················· | |
| 19100 | MT972242.1 | \|Severe acute respiratory syndrome coro | ···················· | ···················· | ···················· | ···················· | ·NN················· | ···················· | |
| 19101 | MT972243.1 | \|Severe acute respiratory syndrome coro | ···················· | ···················· | ···················· | ···················· | ···················· | ···················· | |
| 19102 | MT972244.1 | \|Severe acute respiratory syndrome coro | ···················· | ···················· | ···················· | ···················· | ···················· | ···················· | |
| 19103 | MT972245.1 | \|Severe acute respiratory syndrome coro | ···················· | ···················· | ···················· | ···················· | ···················· | ···················· | |
| 19104 | MT972246.1 | \|Severe acute respiratory syndrome coro | ···················· | ···················· | ···················· | ···················· | ···················· | ···················· | |
| 19105 | MT972247.1 | \|Severe acute respiratory syndrome coro | ···················· | ···················· | ···················· | ···················· | ···················· | ···················· | |
| 19106 | MT972248.1 | \|Severe acute respiratory syndrome coro | ···················· | ···················· | ···················· | ···················· | ···················· | ···················· | |
| 19107 | MT972249.1 | \|Severe acute respiratory syndrome coro | ···················· | ···················· | ···················· | ···················· | ···················· | ···················· | |
| 19108 | MT972250.1 | \|Severe acute respiratory syndrome coro | ···················· | ···················· | ···················· | ···················· | ···················· | ···················· | |
| 19109 | MT972251.1 | \|Severe acute respiratory syndrome coro | ···················· | ···················· | ···················· | ···················· | ···················· | ···················· | |
| 19110 | MT972252.1 | \|Severe acute respiratory syndrome coro | ···················· | ···················· | ···················· | ···················· | ···················· | ···················· | |
| 19111 | MT972253.1 | \|Severe acute respiratory syndrome coro | ···················· | ···················· | ···················· | ···················· | ···················· | ···················· | |
| 19112 | MT972255.1 | \|Severe acute respiratory syndrome coro | ···················· | ···················· | ···················· | ···················· | ···················· | ···················· | |
| 19113 | MT972256.1 | \|Severe acute respiratory syndrome coro | ···················· | ···················· | ···················· | ···················· | ···················· | ···················· | |
| 19114 | MT972257.1 | \|Severe acute respiratory syndrome coro | ···················· | ···················· | ···················· | ···················· | ···················· | ···················· | |
| 19115 | MT972258.1 | \|Severe acute respiratory syndrome coro | ···················· | ···················· | ···················· | ···················· | ···················· | ···················· | |
| 19116 | MT972259.1 | \|Severe acute respiratory syndrome coro | ···················· | ···················· | ···················· | ···················· | ···················· | ···················· | |
| 19117 | MT972260.1 | \|Severe acute respiratory syndrome coro | ···················· | ···················· | ···················· | ···················· | ···················· | ···················· | |
| 19118 | MT972261.1 | \|Severe acute respiratory syndrome coro | ···················· | ···················· | ···················· | ···················· | ···················· | ···················· | |
| 19119 | MT972262.1 | \|Severe acute respiratory syndrome coro | ···················· | ···················· | ···················· | ···················· | ···················· | ···················· | |
| 19120 | MT972263.1 | \|Severe acute respiratory syndrome coro | ···················· | ···················· | ···················· | ···················· | ···················· | ···················· | |
| 19121 | MT972264.1 | \|Severe acute respiratory syndrome coro | ···················· | ···················· | ···················· | ···················· | ···················· | ···················· | |
| 19122 | MT972265.1 | \|Severe acute respiratory syndrome coro | ···················· | ···················· | ···················· | ···················· | ···················· | ···················· | |
| 19123 | MT972266.1 | \|Severe acute respiratory syndrome coro | ···················· | ···················· | ···················· | ···················· | ···················· | ···················· | |
| 19124 | MT972267.1 | \|Severe acute respiratory syndrome coro | ···················· | ···················· | ···················· | ···················· | ···················· | ···················· | |
| 19125 | MT972268.1 | \|Severe acute respiratory syndrome coro | ···················· | ···················· | ···················· | ···················· | ···················· | ···················· | |
| 19126 | MT972269.1 | \|Severe acute respiratory syndrome coro | ···················· | ···················· | ··A················· | ···················· | ···················· | ···················· | |
| 19127 | MT972270.1 | \|Severe acute respiratory syndrome coro | ···················· | ···················· | ···················· | ···················· | ···················· | ···················· | |
| 19128 | MT972271.1 | \|Severe acute respiratory syndrome coro | ···················· | ···················· | ···················· | ···················· | ···················· | ···················· | |
| 19129 | MT972272.1 | \|Severe acute respiratory syndrome coro | ···················· | ···················· | ···················· | ···················· | ···················· | ···················· | |
| 19130 | MT972273.1 | \|Severe acute respiratory syndrome coro | ···················· | ···················· | ···················· | ···················· | ···················· | ···················· | |
| 19131 | MT972274.1 | \|Severe acute respiratory syndrome coro | ···················· | ···················· | ···················· | ···················· | ···················· | ···················· | |
| 19132 | MT972275.1 | \|Severe acute respiratory syndrome coro | ···················· | ···················· | ···················· | ···················· | ···················· | ···················· | |
| 19133 | MT972276.1 | \|Severe acute respiratory syndrome coro | ···················· | ···················· | ···················· | ···················· | ···················· | ···················· | |
| 19134 | MT972277.1 | \|Severe acute respiratory syndrome coro | ···················· | ···················· | ···················· | ···················· | ···················· | ···················· | |
| 19135 | MT972278.1 | \|Severe acute respiratory syndrome coro | ···················· | ···················· | ···················· | ···················· | ···················· | ···················· | |
| 19136 | MT972279.1 | \|Severe acute respiratory syndrome coro | ···················· | ···················· | ···················· | ···················· | ···················· | ···················· | |
| 19137 | MT972280.1 | \|Severe acute respiratory syndrome coro | ···················· | ···················· | ···················· | ···················· | ···················· | ···················· | |
| 19138 | MT972281.1 | \|Severe acute respiratory syndrome coro | ···················· | ···················· | ···················· | ···················· | ···················· | ···················· | |
| 19139 | MT972282.1 | \|Severe acute respiratory syndrome coro | ···················· | ···················· | ···················· | ···················· | ···················· | ···················· | |
| 19141 | MT972284.1 | \|Severe acute respiratory syndrome coro | ···················· | ···················· | ···················· | ···················· | ···················· | ···················· | |
| 19142 | MT972285.1 | \|Severe acute respiratory syndrome coro | ···················· | ···················· | ···················· | ···················· | ···················· | ···················· | |
| 19143 | MT972286.1 | \|Severe acute respiratory syndrome coro | ···················· | ···················· | ···················· | ···················· | ···················· | ···················· | |
| 19144 | MT972287.1 | \|Severe acute respiratory syndrome coro | ···················· | ···················· | ···················· | ···················· | ···················· | ···················· | |
| 19145 | MT972288.1 | \|Severe acute respiratory syndrome coro | ···················· | ···················· | ···················· | ···················· | ···················· | ···················· | |
| 19146 | MT972289.1 | \|Severe acute respiratory syndrome coro | ···················· | ···················· | ···················· | ···················· | ···················· | ···················· | |
| 19147 | MT972290.1 | \|Severe acute respiratory syndrome coro | ···················· | ···················· | ···················· | ···················· | ···················· | ···················· | |
| 19148 | MT972291.1 | \|Severe acute respiratory syndrome coro | ···················· | ···················· | ···················· | ···················· | ···················· | ···················· | |
| 19149 | MT972292.1 | \|Severe acute respiratory syndrome coro | ···················· | ···················· | ···················· | ···················· | NNN················· | ···················· | 1 |
| 19150 | MT972293.1 | \|Severe acute respiratory syndrome coro | ···················· | ···················· | ···················· | ···················· | ···················· | ···················· | |
| 19151 | MT972294.1 | \|Severe acute respiratory syndrome coro | ···················· | ···················· | ···················· | ···················· | ···················· | ···················· | |
| 19152 | MT972295.1 | \|Severe acute respiratory syndrome coro | ···················· | ···················· | ···················· | ···················· | ···················· | ···················· | |
| 19153 | MT972296.1 | \|Severe acute respiratory syndrome coro | ···················· | ···················· | ···················· | ···················· | ···················· | ···················· | |
| 19154 | MT972297.1 | \|Severe acute respiratory syndrome coro | ···················· | ···················· | ···················· | ···················· | ···················· | ···················· | |
| 19155 | MT972298.1 | \|Severe acute respiratory syndrome coro | ···················· | ···················· | ···················· | ···················· | ···················· | ···················· | |
| 19156 | MT972299.1 | \|Severe acute respiratory syndrome coro | ···················· | ···················· | ···················· | ···················· | ···················· | ···················· | |
| 19157 | MT972300.1 | \|Severe acute respiratory syndrome coro | ···················· | ···················· | ···················· | ···················· | ···················· | ···················· | |
| 19158 | MT972301.1 | \|Severe acute respiratory syndrome coro | ···················· | ···················· | ···················· | ···················· | ···················· | ···················· | |
| 19159 | MT972302.1 | \|Severe acute respiratory syndrome coro | ···················· | ···················· | ···················· | ···················· | ···················· | ···················· | |
| 19160 | MT972303.1 | \|Severe acute respiratory syndrome coro | ···················· | ···················· | ···················· | ···················· | ···················· | ···················· | |
| 19161 | MT972304.1 | \|Severe acute respiratory syndrome coro | ···················· | ···················· | ···················· | ···················· | ···················· | ···················· | |
| 19162 | MT972305.1 | \|Severe acute respiratory syndrome coro | ···················· | ···················· | ···················· | ···················· | ···················· | ···················· | |
| 19163 | MT972306.1 | \|Severe acute respiratory syndrome coro | ···················· | ···················· | ···················· | ···················· | ···················· | ···················· | |
| 19164 | MT972307.1 | \|Severe acute respiratory syndrome coro | ···················· | ···················· | ···················· | ···················· | ···················· | ···················· | |
| 19165 | MT972308.1 | \|Severe acute respiratory syndrome coro | ···················· | ···················· | ···················· | ···················· | ···················· | ···················· | |
| 19166 | MT972310.1 | \|Severe acute respiratory syndrome coro | ···················· | ···················· | ···················· | ···················· | ···················· | ···················· | |
| 19167 | MT972311.1 | \|Severe acute respiratory syndrome coro | ···················· | ···················· | ···················· | ···················· | ···················· | ···················· | |
| 19168 | MT972312.1 | \|Severe acute respiratory syndrome coro | ···················· | ···················· | ···················· | ···················· | N··················· | ···················· | |
| 19169 | MT972313.1 | \|Severe acute respiratory syndrome coro | ···················· | ···················· | ···················· | ···················· | ···················· | ···················· | |
| 19170 | MT972314.1 | \|Severe acute respiratory syndrome coro | ···················· | ···················· | ···················· | ···················· | ···················· | ···················· | |
| 19171 | MT972315.1 | \|Severe acute respiratory syndrome coro | ···················· | ···················· | ···················· | ···················· | ···················· | ···················· | |
| 19172 | MT972316.1 | \|Severe acute respiratory syndrome coro | ···················· | ···················· | ···················· | ···················· | ···················· | ···················· | |
| 19173 | MT972317.1 | \|Severe acute respiratory syndrome coro | ···················· | ···················· | ···················· | ···················· | ···················· | ···················· | |
| 19174 | MT972318.1 | \|Severe acute respiratory syndrome coro | ···················· | ···················· | ···················· | ···················· | ···················· | ···················· | |
| 19175 | MT972319.1 | \|Severe acute respiratory syndrome coro | ···················· | ···················· | ···················· | ···················· | ···················· | ···················· | |
| 19176 | MT972320.1 | \|Severe acute respiratory syndrome coro | ···················· | ···················· | ···················· | ···················· | ···················· | ···················· | |
| 19177 | MT972322.1 | \|Severe acute respiratory syndrome coro | ···················· | ···················· | ···················· | ···················· | ···················· | ···················· | |

```
19178 MT972323.1 |Severe acute respiratory syndrome coro    ....................  ....................  ....................  ....................  ....................  ....................
19179 MT972324.1 |Severe acute respiratory syndrome coro    ....................  ....................  ....................  ....................  ....................  ....................
19180 MT972325.1 |Severe acute respiratory syndrome coro    ....................  ....................  ....................  ....................  ....................  ....................
19181 MT972326.1 |Severe acute respiratory syndrome coro    ....................  ....................  ....................  ....................  ....................  ....................
19182 MT972327.1 |Severe acute respiratory syndrome coro    ....................  ....................  ....................  ....................  ....................  ....................
19183 MT972328.1 |Severe acute respiratory syndrome coro    ....................  ....................  ....................  ....................  ....................  ....................
19184 MT972329.1 |Severe acute respiratory syndrome coro    ....................  ....................  ....................  ....................  ....................  ....................
19185 MT972330.1 |Severe acute respiratory syndrome coro    ....................  ....................  ....................  ....................  ....................  ....................
19186 MT972331.1 |Severe acute respiratory syndrome coro    ....................  ....................  ....................  ....................  ....................  ....................
19187 MT972332.1 |Severe acute respiratory syndrome coro    ....................  ....................  ....................  ....................  ....................  ....................
19188 MT972333.1 |Severe acute respiratory syndrome coro    ....................  ....................  ....................  ....................  ....................  ....................
19189 MT972335.1 |Severe acute respiratory syndrome coro    ....................  ....................  ....................  ....................  ....................  ....................
19190 MT972336.1 |Severe acute respiratory syndrome coro    ....................  ....................  ....................  ....................  ....................  ....................
19191 MT972337.1 |Severe acute respiratory syndrome coro    ....................  ....................  ....................  ....................  ....................  ....................
19192 MT972338.1 |Severe acute respiratory syndrome coro    ....................  ....................  ....................  ....................  ....................  ....................
19194 MT972341.1 |Severe acute respiratory syndrome coro    ....................  ....................  ....................  ....................  ....................  ....................
19195 MT972342.1 |Severe acute respiratory syndrome coro    ....................  ....................  ....................  ....................  ....................  ....................
19196 MT972343.1 |Severe acute respiratory syndrome coro    ....................  ....................  ....................  ....................  ....................  ....................
19197 MT972344.1 |Severe acute respiratory syndrome coro    ....................  ....................  ....................  ....................  ....................  ....................
19198 MT972345.1 |Severe acute respiratory syndrome coro    ....................  ....................  ....................  ....................  ....................  ....................
19199 MT972346.1 |Severe acute respiratory syndrome coro    ....................  ....................  ....................  ....................  ....................  ....................
19200 MT972347.1 |Severe acute respiratory syndrome coro    ....................  ....................  ....................  ....................  ....................  ....................
19201 MT972348.1 |Severe acute respiratory syndrome coro    ....................  ....................  ....................  ....................  ....................  ....................
19202 MT972349.1 |Severe acute respiratory syndrome coro    ....................  ....................  ....................  ....................  ....................  ....................
19203 MT972350.1 |Severe acute respiratory syndrome coro    ....................  ....................  ....................  ....................  ....................  ....................
19204 MT972351.1 |Severe acute respiratory syndrome coro    ....................  ....................  ....................  ....................  ....................  ....................
19205 MT972352.1 |Severe acute respiratory syndrome coro    ....................  ....................  ....................  ....................  ....................  ....................
19206 MT972353.1 |Severe acute respiratory syndrome coro    ....................  ....................  ....................  ....................  ....................  ....................
19207 MT972355.1 |Severe acute respiratory syndrome coro    ....................  ....................  ....................  ....................  ....................  ....................
19208 MT972357.1 |Severe acute respiratory syndrome coro    ....................  ....................  ....................  ....................  ....................  ....................
19209 MT972358.1 |Severe acute respiratory syndrome coro    ....................  ....................  ....................  ....................  ....................  ....................
19210 MT972359.1 |Severe acute respiratory syndrome coro    ....................  ....................  ....................  ....................  ....................  ....................
19211 MT972360.1 |Severe acute respiratory syndrome coro    ....................  ....................  ....................  ....................  ....................  ....................
19212 MT972361.1 |Severe acute respiratory syndrome coro    ....................  ....................  ....................  ....................  ....................  ....................
19213 MT972362.1 |Severe acute respiratory syndrome coro    ....................  ....................  ....................  ....................  ....................  ....................
19214 MT972363.1 |Severe acute respiratory syndrome coro    ....................  ....................  ....................  ....................  ....................  ....................
19215 MT972364.1 |Severe acute respiratory syndrome coro    ....................  ....................  ....................  ....................  ....................  ....................
19216 MT972365.1 |Severe acute respiratory syndrome coro    ....................  ....................  ....................  ....................  ....................  ....................
19217 MT972366.1 |Severe acute respiratory syndrome coro    ....................  ....................  ....................  ....................  ....................  ....................
19218 MT972367.1 |Severe acute respiratory syndrome coro    ....................  ....................  ....................  ....................  ....................  ....................
19219 MT972368.1 |Severe acute respiratory syndrome coro    ....................  ....................  ....................  ....................  ....................  ....................
19220 MT972369.1 |Severe acute respiratory syndrome coro    ....................  ....................  ....................  ....................  ....................  ....................
19221 MT972370.1 |Severe acute respiratory syndrome coro    ....................  ....................  ....................  ....................  ....................  ....................
19222 MT972371.1 |Severe acute respiratory syndrome coro    ....................  ....................  ....................  ....................  ....................  ....................
19223 MT972372.1 |Severe acute respiratory syndrome coro    ....................  ....................  ....................  ....................  ....................  ....................
19224 MT972373.1 |Severe acute respiratory syndrome coro    ....................  ....................  ....................  ....................  ....................  ....................
19225 MT972374.1 |Severe acute respiratory syndrome coro    ....................  ....................  ....................  ....................  ....................  ....................
19226 MT972375.1 |Severe acute respiratory syndrome coro    ....................  ....................  ....................  ....................  ....................  ....................
19227 MT972376.1 |Severe acute respiratory syndrome coro    ....................  ....................  ....................  ....................  ....................  ....................
19228 MT972377.1 |Severe acute respiratory syndrome coro    ....................  ....................  ....................  ....................  ....................  ....................
19229 MT972378.1 |Severe acute respiratory syndrome coro    ....................  ....................  ....................  ....................  ....................  ....................
19230 MT972379.1 |Severe acute respiratory syndrome coro    ....................  ....................  ....................  ....................  ....................  ....................
19231 MT972380.1 |Severe acute respiratory syndrome coro    ....................  ....................  ....................  ....................  ....................  ....................
19232 MT972381.1 |Severe acute respiratory syndrome coro    ....................  ....................  ....................  ....................  ....................  ....................
19233 MT972382.1 |Severe acute respiratory syndrome coro    ....................  ....................  ....................  ....................  ....................  ....................
19234 MT972383.1 |Severe acute respiratory syndrome coro    ....................  ....................  ....................  ....................  ....................  ....................
19235 MT972384.1 |Severe acute respiratory syndrome coro    ....................  ....................  ....................  ....................  ....................  ....................
19236 MT972385.1 |Severe acute respiratory syndrome coro    ....................  ....................  ....................  ....................  ....................  ....................
19237 MT972386.1 |Severe acute respiratory syndrome coro    ....................  ....................  ....................  ....................  ....................  ....................
19238 MT972387.1 |Severe acute respiratory syndrome coro    ....................  ....................  ....................  ....................  ....................  ....................
19239 MT972388.1 |Severe acute respiratory syndrome coro    ....................  ....................  ....................  ....................  ....................  ....................
19240 MT972389.1 |Severe acute respiratory syndrome coro    ....................  ....................  ....................  ....................  ....................  ....................
19241 MT972390.1 |Severe acute respiratory syndrome coro    ....................  ....................  ....................  ....................  ....................  ....................
19242 MT972391.1 |Severe acute respiratory syndrome coro    ....................  ....................  ....................  ....................  ....................  ....................
19243 MT972392.1 |Severe acute respiratory syndrome coro    ....................  ....................  ....................  ....................  ....................  ....................
19244 MT972393.1 |Severe acute respiratory syndrome coro    ....................  ....................  ....................  ....................  ....................  ....................
19245 MT972394.1 |Severe acute respiratory syndrome coro    ....................  ....................  ....................  ....................  ....................  ....................
19246 MT972395.1 |Severe acute respiratory syndrome coro    ....................  ....................  ....................  ....................  ....................  ....................
19247 MT972397.1 |Severe acute respiratory syndrome coro    ....................  ....................  ....................  ....................  ....................  ....................
19248 MT972398.1 |Severe acute respiratory syndrome coro    ....................  ....................  ....................  ....................  ....................  ....................
19249 MT972399.1 |Severe acute respiratory syndrome coro    ....................  ....................  ....................  ....................  ....................  ....................
19250 MT972400.1 |Severe acute respiratory syndrome coro    ....................  ....................  ....................  ....................  ....................  ....................
19251 MT972402.1 |Severe acute respiratory syndrome coro    ....................  ....................  ....................  ....................  ....................  ....................
19252 MT972403.1 |Severe acute respiratory syndrome coro    ....................  ....................  ....................  ....................  ....................  ....................
19253 MT972404.1 |Severe acute respiratory syndrome coro    ....................  ....................  ....................  ....................  ....................  ....................
19254 MT972405.1 |Severe acute respiratory syndrome coro    ....................  ....................  ....................  ....................  ....................  ....................
19255 MT972406.1 |Severe acute respiratory syndrome coro    ....................  ....................  ....................  ....................  ....................  ....................
19256 MT972407.1 |Severe acute respiratory syndrome coro    ....................  ....................  ....................  ....................  ....................  ....................
```

| | | | | | | | | | |
|---|---|---|---|---|---|---|---|---|---|
| 19257 | MT972408.1 | \|Severe acute respiratory syndrome coro | ···················· | ···················· | ···················· | ···················· | ···················· | ···················· | |
| 19258 | MT972409.1 | \|Severe acute respiratory syndrome coro | ···················· | ···················· | ···················· | ···················· | ···················· | ···················· | |
| 19259 | MT972410.1 | \|Severe acute respiratory syndrome coro | ···················· | ···················· | ···················· | ···················· | ···················· | ···················· | |
| 19260 | MT972411.1 | \|Severe acute respiratory syndrome coro | ···················· | ···················· | ···················· | ···················· | ···················· | ···················· | |
| 19261 | MT972412.1 | \|Severe acute respiratory syndrome coro | ···················· | ···················· | ···················· | ···················· | ···················· | ···················· | |
| 19262 | MT972413.1 | \|Severe acute respiratory syndrome coro | ···················· | ···················· | ···················· | ···················· | ···················· | ···················· | |
| 19263 | MT972414.1 | \|Severe acute respiratory syndrome coro | ···················· | ···················· | ···················· | ···················· | ···················· | ···················· | |
| 19264 | MT972415.1 | \|Severe acute respiratory syndrome coro | ···················· | ···················· | ···················· | ···················· | NNN················· | ···················· | 1 |
| 19265 | MT972416.1 | \|Severe acute respiratory syndrome coro | ···················· | ···················· | ···················· | ···················· | ···················· | ···················· | |
| 19266 | MT972417.1 | \|Severe acute respiratory syndrome coro | ···················· | ···················· | ···················· | ···················· | ···················· | ···················· | |
| 19267 | MT972418.1 | \|Severe acute respiratory syndrome coro | ···················· | ···················· | ···················· | ···················· | ···················· | ···················· | |
| 19268 | MT972420.1 | \|Severe acute respiratory syndrome coro | ···················· | ···················· | ···················· | ···················· | ···················· | ···················· | |
| 19269 | MT972421.1 | \|Severe acute respiratory syndrome coro | ···················· | ···················· | ···················· | ···················· | ···················· | ···················· | |
| 19270 | MT972423.1 | \|Severe acute respiratory syndrome coro | ···················· | ···················· | ···················· | ···················· | ···················· | ···················· | |
| 19271 | MT972424.1 | \|Severe acute respiratory syndrome coro | ···················· | ···················· | ···················· | ···················· | ···················· | ···················· | |
| 19272 | MT972425.1 | \|Severe acute respiratory syndrome coro | ···················· | ···················· | ···················· | ···················· | ···················· | ···················· | |
| 19274 | MT972427.1 | \|Severe acute respiratory syndrome coro | ···················· | ···················· | ···················· | ···················· | ···················· | ···················· | |
| 19275 | MT972428.1 | \|Severe acute respiratory syndrome coro | ···················· | ···················· | ···················· | ···················· | ···················· | ···················· | |
| 19276 | MT972429.1 | \|Severe acute respiratory syndrome coro | ···················· | ···················· | ···················· | ···················· | ···················· | ···················· | |
| 19277 | MT972430.1 | \|Severe acute respiratory syndrome coro | ···················· | ···················· | ···················· | ···················· | ···················· | ···················· | |
| 19278 | MT972431.1 | \|Severe acute respiratory syndrome coro | ···················· | ···················· | ···················· | ···················· | ···················· | ···················· | |
| 19279 | MT972432.1 | \|Severe acute respiratory syndrome coro | ···················· | ···················· | ···················· | ···················· | ···················· | ···················· | |
| 19280 | MT972433.1 | \|Severe acute respiratory syndrome coro | ···················· | ···················· | ···················· | ···················· | ···················· | ···················· | |
| 19281 | MT972434.1 | \|Severe acute respiratory syndrome coro | ···················· | ···················· | ···················· | ···················· | ···················· | ···················· | |
| 19282 | MT972435.1 | \|Severe acute respiratory syndrome coro | ···················· | ···················· | ···················· | ···················· | ···················· | ···················· | |
| 19283 | MT972436.1 | \|Severe acute respiratory syndrome coro | ···················· | ···················· | ···················· | ···················· | ···················· | ···················· | |
| 19284 | MT972437.1 | \|Severe acute respiratory syndrome coro | ···················· | ···················· | ···················· | ···················· | ···················· | ···················· | |
| 19285 | MT972438.1 | \|Severe acute respiratory syndrome coro | ···················· | ···················· | ···················· | ···················· | ···················· | ···················· | |
| 19286 | MT972440.1 | \|Severe acute respiratory syndrome coro | ···················· | ···················· | ···················· | ···················· | ···················· | ···················· | |
| 19287 | MT972441.1 | \|Severe acute respiratory syndrome coro | ···················· | ···················· | ···················· | ···················· | ···················· | ···················· | |
| 19288 | MT972442.1 | \|Severe acute respiratory syndrome coro | ···················· | ···················· | ···················· | ···················· | ···················· | ···················· | |
| 19289 | MT972443.1 | \|Severe acute respiratory syndrome coro | ···················· | ···················· | ···················· | ···················· | ···················· | ···················· | |
| 19290 | MT972445.1 | \|Severe acute respiratory syndrome coro | ···················· | ···················· | ···················· | ···················· | ···················· | ···················· | |
| 19291 | MT972446.1 | \|Severe acute respiratory syndrome coro | ···················· | ···················· | ···················· | ···················· | ···················· | ···················· | |
| 19292 | MT972447.1 | \|Severe acute respiratory syndrome coro | ···················· | ···················· | ···················· | ···················· | ···················· | ···················· | |
| 19293 | MT972448.1 | \|Severe acute respiratory syndrome coro | ···················· | ···················· | ···················· | ···················· | ···················· | ···················· | |
| 19294 | MT972449.1 | \|Severe acute respiratory syndrome coro | ···················· | ···················· | ···················· | ···················· | ···················· | ···················· | |
| 19296 | MT972451.1 | \|Severe acute respiratory syndrome coro | ···················· | ···················· | ···················· | ···················· | ···················· | ···················· | |
| 19297 | MT972452.1 | \|Severe acute respiratory syndrome coro | ···················· | ···················· | ···················· | ···················· | ···················· | ···················· | |
| 19298 | MT972453.1 | \|Severe acute respiratory syndrome coro | ···················· | ···················· | ···················· | ···················· | ···················· | ···················· | |
| 19299 | MT972454.1 | \|Severe acute respiratory syndrome coro | ···················· | ···················· | ···················· | ···················· | ···················· | ···················· | |
| 19300 | MT972455.1 | \|Severe acute respiratory syndrome coro | ···················· | ···················· | ···················· | ···················· | ···················· | ···················· | |
| 19301 | MT972456.1 | \|Severe acute respiratory syndrome coro | ···················· | ···················· | ···················· | ···················· | ···················· | ···················· | |
| 19302 | MT972457.1 | \|Severe acute respiratory syndrome coro | ···················· | ···················· | ···················· | ···················· | ···················· | ···················· | |
| 19303 | MT972458.1 | \|Severe acute respiratory syndrome coro | ···················· | ···················· | ···················· | ···················· | ···················· | ···················· | |
| 19304 | MT972459.1 | \|Severe acute respiratory syndrome coro | ···················· | ···················· | ···················· | ···················· | ···················· | ···················· | |
| 19305 | MT972460.1 | \|Severe acute respiratory syndrome coro | ···················· | ···················· | ···················· | ···················· | ···················· | ···················· | |
| 19306 | MT972461.1 | \|Severe acute respiratory syndrome coro | ···················· | ···················· | ···················· | ···················· | ···················· | ···················· | |
| 19307 | MT972462.1 | \|Severe acute respiratory syndrome coro | ···················· | ···················· | ···················· | ···················· | ···················· | ···················· | |
| 19308 | MT972463.1 | \|Severe acute respiratory syndrome coro | ···················· | ···················· | ···················· | ···················· | ···················· | ···················· | |
| 19309 | MT972464.1 | \|Severe acute respiratory syndrome coro | ···················· | ···················· | ···················· | ···················· | ···················· | ···················· | |
| 19310 | MT972465.1 | \|Severe acute respiratory syndrome coro | ···················· | ···················· | ···················· | ···················· | ···················· | ···················· | |
| 19311 | MT972466.1 | \|Severe acute respiratory syndrome coro | ···················· | ···················· | ···················· | ···················· | ···················· | ···················· | |
| 19312 | MT972467.1 | \|Severe acute respiratory syndrome coro | ···················· | ···················· | ···················· | ···················· | ···················· | ···················· | |
| 19313 | MT972468.1 | \|Severe acute respiratory syndrome coro | ···················· | ···················· | ···················· | ···················· | ···················· | ···················· | |
| 19314 | MT972469.1 | \|Severe acute respiratory syndrome coro | ···················· | ···················· | ···················· | ···················· | ···················· | ···················· | |
| 19315 | MT972470.1 | \|Severe acute respiratory syndrome coro | ···················· | ···················· | ···················· | ···················· | NNN················· | ···················· | 1 |
| 19316 | MT972471.1 | \|Severe acute respiratory syndrome coro | ···················· | ···················· | ···················· | ···················· | ···················· | ···················· | |
| 19317 | MT972472.1 | \|Severe acute respiratory syndrome coro | ···················· | ···················· | ···················· | ···················· | ···················· | ···················· | |
| 19318 | MT972473.1 | \|Severe acute respiratory syndrome coro | ···················· | ···················· | ···················· | ···················· | ···················· | ···················· | |
| 19319 | MT972474.1 | \|Severe acute respiratory syndrome coro | ···················· | ···················· | ···················· | ···················· | ···················· | ···················· | |
| 19320 | MT972475.1 | \|Severe acute respiratory syndrome coro | ···················· | ···················· | ···················· | ···················· | ···················· | ···················· | |
| 19321 | MT972476.1 | \|Severe acute respiratory syndrome coro | ···················· | ···················· | ···················· | ···················· | ···················· | ···················· | |
| 19322 | MT972477.1 | \|Severe acute respiratory syndrome coro | ···················· | ···················· | ···················· | ···················· | ···················· | ···················· | |
| 19323 | MT972478.1 | \|Severe acute respiratory syndrome coro | ···················· | ···················· | ···················· | ···················· | ···················· | ···················· | |
| 19324 | MT972479.1 | \|Severe acute respiratory syndrome coro | ···················· | ···················· | ···················· | ···················· | ···················· | ···················· | |
| 19325 | MT972480.1 | \|Severe acute respiratory syndrome coro | ···················· | ···················· | ···················· | ···················· | ···················· | ···················· | |
| 19327 | MT972482.1 | \|Severe acute respiratory syndrome coro | ···················· | ···················· | ···················· | ···················· | ···················· | ···················· | |
| 19328 | MT972483.1 | \|Severe acute respiratory syndrome coro | ···················· | ···················· | ···················· | ···················· | ···················· | ···················· | |
| 19329 | MT972484.1 | \|Severe acute respiratory syndrome coro | ···················· | ···················· | ···················· | ···················· | ···················· | ···················· | |
| 19330 | MT972485.1 | \|Severe acute respiratory syndrome coro | ···················· | ···················· | ···················· | ···················· | ···················· | ···················· | |
| 19331 | MT972486.1 | \|Severe acute respiratory syndrome coro | ···················· | ···················· | ···················· | ···················· | ···················· | ···················· | |
| 19332 | MT972487.1 | \|Severe acute respiratory syndrome coro | ···················· | ···················· | ···················· | ···················· | ···················· | ···················· | |
| 19333 | MT972488.1 | \|Severe acute respiratory syndrome coro | ···················· | ···················· | ···················· | ···················· | ···················· | ···················· | |
| 19334 | MT972489.1 | \|Severe acute respiratory syndrome coro | ···················· | ···················· | ···················· | ···················· | ···················· | ···················· | |
| 19336 | MT972491.1 | \|Severe acute respiratory syndrome coro | ···················· | ···················· | ···················· | ···················· | ···················· | ···················· | |
| 19337 | MT972492.1 | \|Severe acute respiratory syndrome coro | ···················· | ···················· | ···················· | ···················· | ···················· | ···················· | |
| 19338 | MT972493.1 | \|Severe acute respiratory syndrome coro | ···················· | ···················· | ···················· | ···················· | ···················· | ···················· | |

| | | | | | | | | | |
|---|---|---|---|---|---|---|---|---|---|
| 19339 | MT972494.1 | Severe acute respiratory syndrome coro | ···················· | ···················· | ···················· | ···················· | ···················· | ···················· | |
| 19340 | MT972495.1 | Severe acute respiratory syndrome coro | ···················· | ···················· | ···················· | ···················· | ···················· | ···················· | |
| 19341 | MT972496.1 | Severe acute respiratory syndrome coro | ···················· | ···················· | ···················· | ···················· | ···················· | ···················· | |
| 19342 | MT972497.1 | Severe acute respiratory syndrome coro | ···················· | ···················· | ···················· | ···················· | ···················· | ···················· | |
| 19343 | MT972498.1 | Severe acute respiratory syndrome coro | ···················· | ···················· | ···················· | ···················· | ···················· | ···················· | |
| 19344 | MT972500.1 | Severe acute respiratory syndrome coro | ···················· | ···················· | ···················· | ···················· | ···················· | ···················· | |
| 19345 | MT972502.1 | Severe acute respiratory syndrome coro | ···················· | ···················· | ···················· | ···················· | ···················· | ···················· | |
| 19346 | MT972503.1 | Severe acute respiratory syndrome coro | ···················· | ···················· | ···················· | ···················· | ···················· | ···················· | |
| 19347 | MT972504.1 | Severe acute respiratory syndrome coro | ···················· | ···················· | ···················· | ···················· | ···················· | ···················· | |
| 19348 | MT972505.1 | Severe acute respiratory syndrome coro | ···················· | ···················· | ···················· | ···················· | ···················· | ···················· | |
| 19349 | MT972506.1 | Severe acute respiratory syndrome coro | ···················· | ···················· | ···················· | ···················· | ···················· | ···················· | |
| 19350 | MT972507.1 | Severe acute respiratory syndrome coro | ···················· | ···················· | ···················· | ···················· | ···················· | ···················· | |
| 19351 | MT972508.1 | Severe acute respiratory syndrome coro | ···················· | ···················· | ···················· | ···················· | ···················· | ···················· | |
| 19352 | MT972509.1 | Severe acute respiratory syndrome coro | ···················· | ···················· | ···················· | ···················· | ···················· | ···················· | |
| 19353 | MT972510.1 | Severe acute respiratory syndrome coro | ···················· | ···················· | ···················· | ···················· | ···················· | ···················· | |
| 19354 | MT972511.1 | Severe acute respiratory syndrome coro | ···················· | ·········A·········· | ···················· | ···················· | ···················· | ···················· | |
| 19355 | MT972512.1 | Severe acute respiratory syndrome coro | ···················· | ···················· | ···················· | ···················· | ···················· | ···················· | |
| 19356 | MT972513.1 | Severe acute respiratory syndrome coro | ···················· | ···················· | ···················· | ···················· | ···················· | ···················· | |
| 19357 | MT972514.1 | Severe acute respiratory syndrome coro | ···················· | ···················· | ···················· | ···················· | ···················· | ···················· | |
| 19358 | MT972515.1 | Severe acute respiratory syndrome coro | ···················· | ···················· | ···················· | ···················· | ···················· | ···················· | |
| 19359 | MT972516.1 | Severe acute respiratory syndrome coro | ···················· | ···················· | ···················· | ···················· | ···················· | ···················· | |
| 19360 | MT972517.1 | Severe acute respiratory syndrome coro | ···················· | ···················· | ···················· | ···················· | ···················· | ···················· | |
| 19361 | MT972518.1 | Severe acute respiratory syndrome coro | ···················· | ···················· | ···················· | ···················· | ···················· | ···················· | |
| 19363 | MT972520.1 | Severe acute respiratory syndrome coro | ···················· | ···················· | ···················· | ···················· | ···················· | ···················· | |
| 19364 | MT972521.1 | Severe acute respiratory syndrome coro | ···················· | ···················· | ···················· | ···················· | ···················· | ···················· | |
| 19365 | MT972522.1 | Severe acute respiratory syndrome coro | ···················· | ···················· | ···················· | ···················· | ···················· | ···················· | |
| 19366 | MT972523.1 | Severe acute respiratory syndrome coro | ···················· | ···················· | ···················· | ···················· | ···················· | ···················· | |
| 19367 | MT972524.1 | Severe acute respiratory syndrome coro | ···················· | ···················· | ···················· | ···················· | ···················· | ···················· | |
| 19368 | MT972525.1 | Severe acute respiratory syndrome coro | ···················· | ···················· | ···················· | ···················· | ···················· | ···················· | |
| 19369 | MT972526.1 | Severe acute respiratory syndrome coro | ···················· | ···················· | ···················· | ···················· | NNN················· | ···················· | 1 |
| 19370 | MT972527.1 | Severe acute respiratory syndrome coro | ···················· | ···················· | ···················· | ···················· | ···················· | ···················· | |
| 19371 | MT972528.1 | Severe acute respiratory syndrome coro | ···················· | ···················· | ···················· | ···················· | ···················· | ···················· | |
| 19374 | MT972532.1 | Severe acute respiratory syndrome coro | ···················· | ···················· | ···················· | ···················· | ···················· | ···················· | |
| 19375 | MT972533.1 | Severe acute respiratory syndrome coro | ···················· | ···················· | ···················· | ···················· | ···················· | ···················· | |
| 19376 | MT972534.1 | Severe acute respiratory syndrome coro | ···················· | ···················· | ···················· | ···················· | ···················· | ···················· | |
| 19377 | MT972535.1 | Severe acute respiratory syndrome coro | ···················· | ···················· | ···················· | ···················· | ···················· | ···················· | |
| 19378 | MT972536.1 | Severe acute respiratory syndrome coro | ···················· | ···················· | ···················· | ···················· | ···················· | ···················· | |
| 19379 | MT972537.1 | Severe acute respiratory syndrome coro | ···················· | ···················· | ···················· | ···················· | ···················· | ···················· | |
| 19380 | MT972538.1 | Severe acute respiratory syndrome coro | ···················· | ···················· | ···················· | ···················· | ···················· | ···················· | |
| 19382 | MT972540.1 | Severe acute respiratory syndrome coro | ···················· | ···················· | ···················· | ···················· | ···················· | ···················· | |
| 19383 | MT972541.1 | Severe acute respiratory syndrome coro | ···················· | ···················· | ···················· | ···················· | ···················· | ···················· | |
| 19384 | MT972542.1 | Severe acute respiratory syndrome coro | ···················· | ···················· | ···················· | ···················· | ···················· | ···················· | |
| 19385 | MT972543.1 | Severe acute respiratory syndrome coro | ···················· | ···················· | ···················· | ···················· | ···················· | ···················· | |
| 19386 | MT972544.1 | Severe acute respiratory syndrome coro | ···················· | ···················· | ···················· | ···················· | ···················· | ···················· | |
| 19387 | MT972546.1 | Severe acute respiratory syndrome coro | ···················· | ···················· | ···················· | ···················· | ···················· | ···················· | |
| 19388 | MT972548.1 | Severe acute respiratory syndrome coro | ···················· | ···················· | ···················· | ···················· | ···················· | ···················· | |
| 19389 | MT972549.1 | Severe acute respiratory syndrome coro | ···················· | ···················· | ···················· | ···················· | ···················· | ···················· | |
| 19390 | MT972550.1 | Severe acute respiratory syndrome coro | ···················· | ···················· | ···················· | ···················· | ···················· | ···················· | |
| 19391 | MT972551.1 | Severe acute respiratory syndrome coro | ···················· | ···················· | ···················· | ···················· | ···················· | ···················· | |
| 19392 | MT972552.1 | Severe acute respiratory syndrome coro | ···················· | ···················· | ···················· | ···················· | ···················· | ···················· | |
| 19393 | MT972553.1 | Severe acute respiratory syndrome coro | ···················· | ···················· | ···················· | ···················· | ···················· | ···················· | |
| 19394 | MT972554.1 | Severe acute respiratory syndrome coro | ···················· | ···················· | ···················· | ···················· | ···················· | ···················· | |
| 19395 | MT972555.1 | Severe acute respiratory syndrome coro | ···················· | ···················· | ···················· | ···················· | ···················· | ···················· | |
| 19396 | MT972556.1 | Severe acute respiratory syndrome coro | ···················· | ···················· | ···················· | ···················· | ···················· | ···················· | |
| 19397 | MT972557.1 | Severe acute respiratory syndrome coro | ···················· | ···················· | ···················· | ···················· | ···················· | ···················· | |
| 19398 | MT972558.1 | Severe acute respiratory syndrome coro | ···················· | ···················· | ···················· | ···················· | ···················· | ···················· | |
| 19399 | MT972559.1 | Severe acute respiratory syndrome coro | ···················· | ···················· | ···················· | ···················· | ···················· | ···················· | |
| 19400 | MT972560.1 | Severe acute respiratory syndrome coro | ···················· | ···················· | ···················· | ···················· | ···················· | ···················· | |
| 19402 | MT972562.1 | Severe acute respiratory syndrome coro | ···················· | ···················· | ···················· | ···················· | ···················· | ···················· | |
| 19403 | MT972563.1 | Severe acute respiratory syndrome coro | ···················· | ···················· | ···················· | ···················· | ···················· | ···················· | |
| 19404 | MT972564.1 | Severe acute respiratory syndrome coro | ···················· | ···················· | ···················· | ···················· | ···················· | ···················· | |
| 19405 | MT972565.1 | Severe acute respiratory syndrome coro | ···················· | ···················· | ···················· | ···················· | ···················· | ···················· | |
| 19406 | MT972566.1 | Severe acute respiratory syndrome coro | ···················· | ···················· | ···················· | ···················· | ···················· | ···················· | |
| 19407 | MT972567.1 | Severe acute respiratory syndrome coro | ···················· | ···················· | ···················· | ···················· | ···················· | ···················· | |
| 19408 | MT972568.1 | Severe acute respiratory syndrome coro | ···················· | ·A·················· | ···················· | ···················· | ···················· | ··A················· | |
| 19409 | MT972570.1 | Severe acute respiratory syndrome coro | ···················· | ···················· | ···················· | ···················· | ··N················· | ···················· | |
| 19410 | MT972571.1 | Severe acute respiratory syndrome coro | ···················· | ···················· | ···················· | ···················· | ···················· | ···················· | |
| 19411 | MT972573.1 | Severe acute respiratory syndrome coro | ···················· | ···················· | ···················· | ···················· | ···················· | ···················· | |
| 19412 | MT972574.1 | Severe acute respiratory syndrome coro | ···················· | ···················· | ···················· | ···················· | ···················· | ···················· | |
| 19414 | MT972576.1 | Severe acute respiratory syndrome coro | ···················· | ···················· | ···················· | ···················· | ···················· | ···················· | |
| 19415 | MT972577.1 | Severe acute respiratory syndrome coro | ···················· | ···················· | ···················· | ···················· | ···················· | ···················· | |
| 19416 | MT972580.1 | Severe acute respiratory syndrome coro | ···················· | ···················· | ···················· | ···················· | ···················· | ···················· | |
| 19417 | MT972582.1 | Severe acute respiratory syndrome coro | ···················· | ···················· | ···················· | ···················· | ···················· | ···················· | |
| 19418 | MT972584.1 | Severe acute respiratory syndrome coro | ···················· | ···················· | ···················· | ···················· | ···················· | ···················· | |
| 19419 | MT972585.1 | Severe acute respiratory syndrome coro | ···················· | ···················· | ···················· | ···················· | ···················· | ···················· | |
| 19420 | MT972586.1 | Severe acute respiratory syndrome coro | ···················· | ···················· | ···················· | ···················· | ···················· | ···················· | |
| 19421 | MT972587.1 | Severe acute respiratory syndrome coro | ···················· | ···················· | ···················· | ···················· | ···················· | ···················· | |
| 19422 | MT972588.1 | Severe acute respiratory syndrome coro | ···················· | ···················· | ···················· | ···················· | ···················· | ···················· | |

| | | | | | | | | | |
|---|---|---|---|---|---|---|---|---|---|
| 19423 | MT972589.1 | Severe acute respiratory syndrome coro | ···················· | ···················· | ···················· | ···················· | ···················· | ···················· | |
| 19424 | MT972590.1 | Severe acute respiratory syndrome coro | ···················· | ···················· | ···················· | ···················· | NNN················· | ···················· | 1 |
| 19425 | MT972591.1 | Severe acute respiratory syndrome coro | ···················· | ···················· | ···················· | ···················· | NNN················· | ···················· | 1 |
| 19426 | MT972592.1 | Severe acute respiratory syndrome coro | ···················· | ···················· | ···················· | ···················· | ···················· | ···················· | |
| 19427 | MT972594.1 | Severe acute respiratory syndrome coro | ···················· | ···················· | ···················· | ···················· | ···················· | ···················· | |
| 19428 | MT972595.1 | Severe acute respiratory syndrome coro | ···················· | ···················· | ···················· | ···················· | ···················· | ···················· | |
| 19430 | MT972599.1 | Severe acute respiratory syndrome coro | ···················· | ···················· | ···················· | ···················· | ···················· | ···················· | |
| 19431 | MT972602.1 | Severe acute respiratory syndrome coro | ···················· | ···················· | ···················· | ···················· | ···················· | ···················· | |
| 19432 | MT972604.1 | Severe acute respiratory syndrome coro | ···················· | ···················· | ···················· | ···················· | ···················· | ···················· | |
| 19434 | MT972607.1 | Severe acute respiratory syndrome coro | ···················· | ···················· | ···················· | ···················· | ···················· | ···················· | |
| 19435 | MT972608.1 | Severe acute respiratory syndrome coro | ···················· | ···················· | ···················· | ···················· | ···················· | ···················· | |
| 19437 | MT972610.1 | Severe acute respiratory syndrome coro | ···················· | ···················· | ···················· | ···················· | ··NN················ | ···················· | |
| 19438 | MT972611.1 | Severe acute respiratory syndrome coro | ···················· | ···················· | ···················· | ···················· | ···················· | ···················· | |
| 19439 | MT972612.1 | Severe acute respiratory syndrome coro | ···················· | ···················· | ···················· | ···················· | ···················· | ···················· | |
| 19440 | MT972614.1 | Severe acute respiratory syndrome coro | ···················· | ···················· | ···················· | ···················· | ···················· | ···················· | |
| 19441 | MT972615.1 | Severe acute respiratory syndrome coro | ···················· | ···················· | ···················· | ···················· | ···················· | ···················· | |
| 19443 | MT972618.1 | Severe acute respiratory syndrome coro | ···················· | ···················· | ···················· | ···················· | ···················· | ···················· | |
| 19444 | MT972619.1 | Severe acute respiratory syndrome coro | ···················· | ···················· | ···················· | ···················· | ···················· | ···················· | |
| 19445 | MT972621.1 | Severe acute respiratory syndrome coro | ···················· | ···················· | ···················· | ···················· | ···················· | ···················· | |
| 19446 | MT972622.1 | Severe acute respiratory syndrome coro | ···················· | ···················· | ···················· | ···················· | ···················· | ···················· | |
| 19447 | MT972625.1 | Severe acute respiratory syndrome coro | ···················· | ···················· | ···················· | ···················· | ·NNN················ | ···················· | 1 |
| 19448 | MT972626.1 | Severe acute respiratory syndrome coro | ···················· | ···················· | ···················· | ···················· | ···················· | ···················· | |
| 19449 | MT972627.1 | Severe acute respiratory syndrome coro | ···················· | ···················· | ···················· | ···················· | ···················· | ···················· | |
| 19450 | MT972629.1 | Severe acute respiratory syndrome coro | ···················· | ···················· | ···················· | ···················· | ···················· | ···················· | |
| 19451 | MT972630.1 | Severe acute respiratory syndrome coro | ···················· | ···················· | ···················· | ···················· | ···················· | ···················· | |
| 19452 | MT972631.1 | Severe acute respiratory syndrome coro | ···················· | ···················· | ···················· | ···················· | ···················· | ···················· | |
| 19454 | MT972634.1 | Severe acute respiratory syndrome coro | ···················· | ···················· | ···················· | ···················· | ···················· | ···················· | |
| 19455 | MT972635.1 | Severe acute respiratory syndrome coro | ···················· | ···················· | ···················· | ···················· | ···················· | ···················· | |
| 19456 | MT972636.1 | Severe acute respiratory syndrome coro | ···················· | ···················· | ···················· | ···················· | ···················· | ···················· | |
| 19457 | MT972637.1 | Severe acute respiratory syndrome coro | ···················· | ···················· | ···················· | ···················· | ···················· | ···················· | |
| 19458 | MT972638.1 | Severe acute respiratory syndrome coro | ···················· | ···················· | ···················· | ···················· | ···················· | ···················· | |
| 19459 | MT972639.1 | Severe acute respiratory syndrome coro | ···················· | ···················· | ···················· | ···················· | ···················· | ···················· | |
| 19460 | MT972640.1 | Severe acute respiratory syndrome coro | ···················· | ···················· | ···················· | ···················· | ···················· | ···················· | |
| 19461 | MT972642.1 | Severe acute respiratory syndrome coro | ···················· | ···················· | ···················· | ···················· | ···················· | ···················· | |
| 19462 | MT972643.1 | Severe acute respiratory syndrome coro | ···················· | ···················· | ···················· | ···················· | ···················· | ···················· | |
| 19463 | MT972647.1 | Severe acute respiratory syndrome coro | ···················· | ···················· | ···················· | ···················· | ···················· | ···················· | |
| 19464 | MT972648.1 | Severe acute respiratory syndrome coro | ···················· | ···················· | ···················· | ···················· | ···················· | ···················· | |
| 19465 | MT972649.1 | Severe acute respiratory syndrome coro | ···················· | ···················· | ···················· | ···················· | ···················· | ···················· | |
| 19467 | MT972652.1 | Severe acute respiratory syndrome coro | ···················· | ···················· | ···················· | ···················· | ···················· | ···················· | |
| 19468 | MT972653.1 | Severe acute respiratory syndrome coro | ···················· | ···················· | ···················· | ···················· | ···················· | ···················· | |
| 19469 | MT972654.1 | Severe acute respiratory syndrome coro | ···················· | ···················· | ···················· | ···················· | ···················· | ···················· | |
| 19470 | MT972655.1 | Severe acute respiratory syndrome coro | ···················· | ···················· | ···················· | ···················· | ···················· | ···················· | |
| 19471 | MT972656.1 | Severe acute respiratory syndrome coro | ···················· | ···················· | ···················· | ···················· | ···················· | ···················· | |
| 19472 | MT972657.1 | Severe acute respiratory syndrome coro | ···················· | ···················· | ···················· | ···················· | ···················· | ···················· | |
| 19474 | MT972659.1 | Severe acute respiratory syndrome coro | ···················· | ···················· | ···················· | ···················· | ···················· | ···················· | |
| 19476 | MT972664.1 | Severe acute respiratory syndrome coro | ···················· | ···················· | ···················· | ···················· | ··N················· | ···················· | |
| 19477 | MT972665.1 | Severe acute respiratory syndrome coro | ···················· | ···················· | ···················· | ···················· | ···················· | ···················· | |
| 19478 | MT972666.1 | Severe acute respiratory syndrome coro | ···················· | ···················· | ···················· | ···················· | ···················· | ···················· | |
| 19480 | MT972668.1 | Severe acute respiratory syndrome coro | ···················· | ···················· | ···················· | ···················· | ···················· | ···················· | |
| 19481 | MT972669.1 | Severe acute respiratory syndrome coro | ···················· | ···················· | ···················· | ···················· | ···················· | ···················· | |
| 19482 | MT972670.1 | Severe acute respiratory syndrome coro | ···················· | ···················· | ···················· | ···················· | NNN················· | ···················· | 1 |
| 19484 | MT972672.1 | Severe acute respiratory syndrome coro | ···················· | ···················· | ···················· | ···················· | ···················· | ···················· | |
| 19485 | MT972675.1 | Severe acute respiratory syndrome coro | ···················· | ···················· | ···················· | ···················· | ···················· | ···················· | |
| 19486 | MT972677.1 | Severe acute respiratory syndrome coro | ···················· | ···················· | ···················· | ···················· | NNN················· | ···················· | 1 |
| 19487 | MT972678.1 | Severe acute respiratory syndrome coro | ···················· | ···················· | ···················· | ···················· | ···················· | ···················· | |
| 19488 | MT972679.1 | Severe acute respiratory syndrome coro | ···················· | ···················· | ···················· | ···················· | ···················· | ···················· | |
| 19489 | MT972680.1 | Severe acute respiratory syndrome coro | ···················· | ···················· | ···················· | ···················· | ···················· | ···················· | |
| 19490 | MT972681.1 | Severe acute respiratory syndrome coro | ···················· | ···················· | ···················· | ···················· | ···················· | ···················· | |
| 19491 | MT972683.1 | Severe acute respiratory syndrome coro | ···················· | ···················· | ···················· | ···················· | ···················· | ···················· | |
| 19492 | MT972684.1 | Severe acute respiratory syndrome coro | ···················· | ···················· | ···················· | ···················· | ···················· | ···················· | |
| 19493 | MT972686.1 | Severe acute respiratory syndrome coro | ···················· | ···················· | ···················· | ···················· | ···················· | ···················· | |
| 19494 | MT972687.1 | Severe acute respiratory syndrome coro | ···················· | ···················· | ···················· | ···················· | ···················· | ···················· | |
| 19495 | MT972688.1 | Severe acute respiratory syndrome coro | ···················· | ···················· | ···················· | ···················· | ···················· | ···················· | |
| 19496 | MT972689.1 | Severe acute respiratory syndrome coro | ···················· | ···················· | ···················· | ···················· | NNN················· | ···················· | 1 |
| 19497 | MT972690.1 | Severe acute respiratory syndrome coro | ···················· | ···················· | ···················· | ···················· | ···················· | ···················· | |
| 19498 | MT972691.1 | Severe acute respiratory syndrome coro | ···················· | ···················· | ···················· | ···················· | ···················· | ···················· | |
| 19499 | MT972692.1 | Severe acute respiratory syndrome coro | ···················· | ···················· | ···················· | ···················· | ···················· | ···················· | |
| 19500 | MT972695.1 | Severe acute respiratory syndrome coro | ···················· | ···················· | ···················· | ···················· | ···················· | ···················· | |
| 19501 | MT972697.1 | Severe acute respiratory syndrome coro | ···················· | ···················· | ···················· | ···················· | ···················· | ···················· | |
| 19503 | MT972701.1 | Severe acute respiratory syndrome coro | ···················· | ···················· | ···················· | ···················· | ···················· | ···················· | |
| 19504 | MT972702.1 | Severe acute respiratory syndrome coro | ···················· | ···················· | ···················· | ···················· | ···················· | ···················· | |
| 19505 | MT972703.1 | Severe acute respiratory syndrome coro | ···················· | ···················· | ···················· | ···················· | ···················· | ···················· | |
| 19506 | MT972704.1 | Severe acute respiratory syndrome coro | ···················· | ···················· | ···················· | ···················· | ···················· | ···················· | |
| 19507 | MT972705.1 | Severe acute respiratory syndrome coro | ···················· | ···················· | ···················· | ···················· | ···················· | ···················· | |
| 19509 | MT972707.1 | Severe acute respiratory syndrome coro | ···················· | ···················· | ···················· | ···················· | ···················· | ···················· | |
| 19510 | MT972708.1 | Severe acute respiratory syndrome coro | ···················· | ···················· | ···················· | ···················· | ···················· | ···················· | |
| 19511 | MT972709.1 | Severe acute respiratory syndrome coro | ···················· | ···················· | ···················· | ···················· | ···················· | ···················· | |
| 19512 | MT972710.1 | Severe acute respiratory syndrome coro | ···················· | ···················· | ···················· | ···················· | NNN················· | ···················· | 1 |

| | | | | | | | | | |
|---|---|---|---|---|---|---|---|---|---|
| 19513 | MT972711.1 | \|Severe acute respiratory syndrome coro | ···················· | ···················· | ···················· | ···················· | ···················· | ···················· | |
| 19514 | MT972712.1 | \|Severe acute respiratory syndrome coro | ···················· | ···················· | ···················· | ···················· | ···················· | ···················· | |
| 19515 | MT972713.1 | \|Severe acute respiratory syndrome coro | ···················· | ···················· | ···················· | ···················· | NNN················· | ···················· | 1 |
| 19517 | MT972715.1 | \|Severe acute respiratory syndrome coro | ···················· | ···················· | ···················· | ···················· | ···················· | ···················· | |
| 19518 | MT972716.1 | \|Severe acute respiratory syndrome coro | ···················· | ···················· | ···················· | ···················· | ···················· | ···················· | |
| 19519 | MT972717.1 | \|Severe acute respiratory syndrome coro | ···················· | ···················· | ···················· | ···················· | ···················· | ···················· | |
| 19522 | MT972720.1 | \|Severe acute respiratory syndrome coro | ···················· | ···················· | ···················· | ···················· | ···················· | ···················· | |
| 19525 | MT972723.1 | \|Severe acute respiratory syndrome coro | ···················· | ···················· | ···················· | ···················· | ···················· | ···················· | |
| 19526 | MT972725.1 | \|Severe acute respiratory syndrome coro | ···················· | ···················· | ···················· | ···················· | ···················· | ···················· | |
| 19527 | MT972727.1 | \|Severe acute respiratory syndrome coro | ···················· | ···················· | ···················· | ···················· | ···················· | ···················· | |
| 19530 | MT972730.1 | \|Severe acute respiratory syndrome coro | ···················· | ···················· | ···················· | ···················· | ···················· | ···················· | |
| 19532 | MT972733.1 | \|Severe acute respiratory syndrome coro | ···················· | ···················· | ···················· | ···················· | ···················· | ···················· | |
| 19533 | MT972734.1 | \|Severe acute respiratory syndrome coro | ···················· | ···················· | ···················· | ···················· | ···················· | ···················· | |
| 19534 | MT972736.1 | \|Severe acute respiratory syndrome coro | ···················· | ···················· | ···················· | ···················· | ···················· | ···················· | |
| 19535 | MT972737.1 | \|Severe acute respiratory syndrome coro | ···················· | ···················· | ···················· | ···················· | ···················· | ···················· | |
| 19536 | MT972738.1 | \|Severe acute respiratory syndrome coro | ···················· | ···················· | ···················· | ···················· | ···················· | ···················· | |
| 19537 | MT972739.1 | \|Severe acute respiratory syndrome coro | ···················· | ···················· | ···················· | ···················· | ···················· | ···················· | |
| 19538 | MT972740.1 | \|Severe acute respiratory syndrome coro | ···················· | ···················· | ···················· | ···················· | ···················· | ···················· | |
| 19539 | MT972741.1 | \|Severe acute respiratory syndrome coro | ···················· | ···················· | ···················· | ···················· | ···················· | ···················· | |
| 19540 | MT972742.1 | \|Severe acute respiratory syndrome coro | ···················· | ···················· | ···················· | ···················· | ···················· | ···················· | |
| 19541 | MT972743.1 | \|Severe acute respiratory syndrome coro | ···················· | ···················· | ···················· | ···················· | ···················· | ···················· | |
| 19542 | MT972744.1 | \|Severe acute respiratory syndrome coro | ···················· | ···················· | ···················· | ···················· | NNN················· | ···················· | 1 |
| 19544 | MT972746.1 | \|Severe acute respiratory syndrome coro | ···················· | ···················· | ···················· | ···················· | ···················· | ···················· | |
| 19546 | MT972749.1 | \|Severe acute respiratory syndrome coro | ···················· | ···················· | ···················· | ···················· | ···················· | ···················· | |
| 19547 | MT972750.1 | \|Severe acute respiratory syndrome coro | ···················· | ···················· | ···················· | ···················· | ···················· | ···················· | |
| 19549 | MT972753.1 | \|Severe acute respiratory syndrome coro | ···················· | ···················· | ···················· | ···················· | ···················· | ···················· | |
| 19550 | MT972754.1 | \|Severe acute respiratory syndrome coro | ···················· | ···················· | ···················· | ···················· | ···················· | ···················· | |
| 19551 | MT972756.1 | \|Severe acute respiratory syndrome coro | ···················· | ···················· | ···················· | ···················· | ···················· | ···················· | |
| 19553 | MT972760.1 | \|Severe acute respiratory syndrome coro | ···················· | ···················· | ···················· | ···················· | NNN················· | ···················· | 1 |
| 19554 | MT972761.1 | \|Severe acute respiratory syndrome coro | ···················· | ···················· | ···················· | ···················· | ···················· | ···················· | |
| 19555 | MT972762.1 | \|Severe acute respiratory syndrome coro | ···················· | ···················· | ···················· | ···················· | ···················· | ···················· | |
| 19556 | MT972763.1 | \|Severe acute respiratory syndrome coro | ···················· | ···················· | ···················· | ···················· | ···················· | ···················· | |
| 19557 | MT972764.1 | \|Severe acute respiratory syndrome coro | ···················· | ···················· | ···················· | ···················· | ···················· | ···················· | |
| 19558 | MT972765.1 | \|Severe acute respiratory syndrome coro | ···················· | ···················· | ···················· | ···················· | ···················· | ···················· | |
| 19559 | MT972767.1 | \|Severe acute respiratory syndrome coro | ···················· | ···················· | ···················· | ···················· | ···················· | ···················· | |
| 19560 | MT972768.1 | \|Severe acute respiratory syndrome coro | ···················· | ···················· | ···················· | ···················· | ···················· | ···················· | |
| 19561 | MT972769.1 | \|Severe acute respiratory syndrome coro | ···················· | ···················· | ···················· | ···················· | ···················· | ···················· | |
| 19562 | MT972770.1 | \|Severe acute respiratory syndrome coro | ···················· | ···················· | ···················· | ···················· | ···················· | ···················· | |
| 19563 | MT972771.1 | \|Severe acute respiratory syndrome coro | ···················· | ···················· | ···················· | ···················· | ···················· | ···················· | |
| 19564 | MT972772.1 | \|Severe acute respiratory syndrome coro | ···················· | ···················· | ···················· | ···················· | ···················· | ···················· | |
| 19565 | MT972773.1 | \|Severe acute respiratory syndrome coro | ···················· | ···················· | ···················· | ···················· | ···················· | ···················· | |
| 19566 | MT972774.1 | \|Severe acute respiratory syndrome coro | ···················· | ···················· | ···················· | ···················· | ···················· | ···················· | |
| 19567 | MT972775.1 | \|Severe acute respiratory syndrome coro | ···················· | ···················· | ···················· | ···················· | ···················· | ···················· | |
| 19568 | MT972776.1 | \|Severe acute respiratory syndrome coro | ···················· | ···················· | ···················· | ···················· | ···················· | ···················· | |
| 19569 | MT972777.1 | \|Severe acute respiratory syndrome coro | ···················· | ···················· | ···················· | ···················· | ···················· | ···················· | |
| 19570 | MT972778.1 | \|Severe acute respiratory syndrome coro | ···················· | ···················· | ···················· | ···················· | ···················· | ···················· | |
| 19571 | MT972779.1 | \|Severe acute respiratory syndrome coro | ···················· | ···················· | ···················· | ···················· | ···················· | ···················· | |
| 19572 | MT972780.1 | \|Severe acute respiratory syndrome coro | ···················· | ···················· | ···················· | ···················· | ···················· | ···················· | |
| 19573 | MT972781.1 | \|Severe acute respiratory syndrome coro | ···················· | ···················· | ···················· | ···················· | ···················· | ···················· | |
| 19574 | MT972782.1 | \|Severe acute respiratory syndrome coro | ···················· | ···················· | ···················· | ···················· | ···················· | ···················· | |
| 19575 | MT972783.1 | \|Severe acute respiratory syndrome coro | ···················· | ···················· | ···················· | ···················· | ···················· | ···················· | |
| 19576 | MT972784.1 | \|Severe acute respiratory syndrome coro | ···················· | ···················· | ···················· | ···················· | ···················· | ···················· | |
| 19577 | MT972785.1 | \|Severe acute respiratory syndrome coro | ···················· | ···················· | ···················· | ···················· | ···················· | ···················· | |
| 19578 | MT972786.1 | \|Severe acute respiratory syndrome coro | ···················· | ···················· | ···················· | ···················· | ···················· | ···················· | |
| 19579 | MT972787.1 | \|Severe acute respiratory syndrome coro | ···················· | ···················· | ···················· | ···················· | ···················· | ···················· | |
| 19580 | MT972788.1 | \|Severe acute respiratory syndrome coro | ···················· | ···················· | ···················· | ···················· | ···················· | ···················· | |
| 19581 | MT972789.1 | \|Severe acute respiratory syndrome coro | ···················· | ···················· | ···················· | ···················· | ···················· | ···················· | |
| 19582 | MT972790.1 | \|Severe acute respiratory syndrome coro | ···················· | ···················· | ···················· | ···················· | ···················· | ···················· | |
| 19583 | MT972792.1 | \|Severe acute respiratory syndrome coro | ···················· | ···················· | ···················· | ···················· | ···················· | ···················· | |
| 19584 | MT972793.1 | \|Severe acute respiratory syndrome coro | ···················· | ···················· | ···················· | ···················· | ···················· | ···················· | |
| 19585 | MT972794.1 | \|Severe acute respiratory syndrome coro | ···················· | ···················· | ···················· | ···················· | ···················· | ···················· | |
| 19586 | MT972795.1 | \|Severe acute respiratory syndrome coro | ···················· | ···················· | ···················· | ···················· | ···················· | ···················· | |
| 19587 | MT972796.1 | \|Severe acute respiratory syndrome coro | ···················· | ···················· | ···················· | ···················· | ···················· | ···················· | |
| 19588 | MT972799.1 | \|Severe acute respiratory syndrome coro | ···················· | ···················· | ···················· | ···················· | ···················· | ···················· | |
| 19589 | MT972800.1 | \|Severe acute respiratory syndrome coro | ···················· | ···················· | ···················· | ···················· | ···················· | ···················· | |
| 19590 | MT972801.1 | \|Severe acute respiratory syndrome coro | ···················· | ···················· | ···················· | ···················· | ···················· | ···················· | |
| 19591 | MT972802.1 | \|Severe acute respiratory syndrome coro | ···················· | ···················· | ···················· | ···················· | ···················· | ···················· | |
| 19592 | MT972803.1 | \|Severe acute respiratory syndrome coro | ···················· | ···················· | ···················· | ···················· | ···················· | ···················· | |
| 19593 | MT972804.1 | \|Severe acute respiratory syndrome coro | ···················· | ···················· | ···················· | ···················· | ···················· | ···················· | |
| 19594 | MT972805.1 | \|Severe acute respiratory syndrome coro | ···················· | ···················· | ···················· | ···················· | ···················· | ···················· | |
| 19595 | MT972806.1 | \|Severe acute respiratory syndrome coro | ···················· | ···················· | ···················· | ···················· | ···················· | ···················· | |
| 19596 | MT972807.1 | \|Severe acute respiratory syndrome coro | ···················· | ···················· | ···················· | ···················· | ···················· | ···················· | |
| 19597 | MT972808.1 | \|Severe acute respiratory syndrome coro | ···················· | ···················· | ···················· | ···················· | ···················· | ···················· | |
| 19598 | MT972809.1 | \|Severe acute respiratory syndrome coro | ···················· | ···················· | ···················· | ···················· | ···················· | ···················· | |
| 19599 | MT972810.1 | \|Severe acute respiratory syndrome coro | ···················· | ···················· | ···················· | ···················· | ···················· | ···················· | |
| 19600 | MT972811.1 | \|Severe acute respiratory syndrome coro | ···················· | ···················· | ···················· | ···················· | ···················· | ···················· | |
| 19601 | MT972812.1 | \|Severe acute respiratory syndrome coro | ···················· | ···················· | ···················· | ···················· | ···················· | ···················· | |
| 19602 | MT972813.1 | \|Severe acute respiratory syndrome coro | ···················· | ···················· | ···················· | ···················· | ···················· | ···················· | |

19603 MT972814.1 |Severe acute respiratory syndrome coro .................... .................... ..................... .................... .................... ....................
19604 MT972816.1 |Severe acute respiratory syndrome coro .................... .................... ..................... .................... .................... ....................
19605 MT972818.1 |Severe acute respiratory syndrome coro .................... .................... ..................... .................... .................... ....................
19606 MT972819.1 |Severe acute respiratory syndrome coro .................... .................... ..................... .................... .................... ....................
19607 MT972820.1 |Severe acute respiratory syndrome coro .................... .................... ..................... .................... .................... ....................
19608 MT972821.1 |Severe acute respiratory syndrome coro .................... .................... ..................... .................... .................... ....................
19609 MT972822.1 |Severe acute respiratory syndrome coro .................... .................... ..................... .................... .................... ....................
19610 MT972823.1 |Severe acute respiratory syndrome coro .................... .................... ..................... .................... .................... ....................
19611 MT972824.1 |Severe acute respiratory syndrome coro .................... .................... ..................... .................... .................... ....................
19612 MT972825.1 |Severe acute respiratory syndrome coro .................... .................... ..................... .................... .................... ....................
19613 MT972826.1 |Severe acute respiratory syndrome coro .................... .................... ..................... .................... .................... ....................
19614 MT972827.1 |Severe acute respiratory syndrome coro .................... .................... ..................... .................... .................... ....................
19615 MT972828.1 |Severe acute respiratory syndrome coro .................... .................... ..................... .................... .................... ....................
19616 MT972829.1 |Severe acute respiratory syndrome coro .................... .................... ..................... .................... .................... ....................
19617 MT972830.1 |Severe acute respiratory syndrome coro .................... .................... ..................... .................... .................... ....................
19618 MT972831.1 |Severe acute respiratory syndrome coro .................... .................... ..................... .................... .................... ....................
19619 MT972832.1 |Severe acute respiratory syndrome coro .................... .................... ..................... .................... .................... ....................
19620 MT972833.1 |Severe acute respiratory syndrome coro .................... .................... ..................... .................... .................... ....................
19621 MT972834.1 |Severe acute respiratory syndrome coro .................... .................... ..................... .................... .................... ....................
19622 MT972835.1 |Severe acute respiratory syndrome coro .................... .................... ..................... .................... .................... ....................
19623 MT972836.1 |Severe acute respiratory syndrome coro .................... .................... ..................... .................... .................... ....................
19624 MT972837.1 |Severe acute respiratory syndrome coro .................... .................... ..................... .................... .................... ....................
19625 MT972838.1 |Severe acute respiratory syndrome coro .................... .................... ..................... .................... .................... ....................
19626 MT972839.1 |Severe acute respiratory syndrome coro .................... .................... ..................... .................... .................... ....................
19627 MT972840.1 |Severe acute respiratory syndrome coro .................... .................... ..................... .................... .................... ....................
19628 MT972841.1 |Severe acute respiratory syndrome coro .................... .................... ..................... .................... .................... ....................
19629 MT972842.1 |Severe acute respiratory syndrome coro .................... .................... ..................... .................... .................... ....................
19630 MT972843.1 |Severe acute respiratory syndrome coro .................... .................... ..................... .................... .................... ....................
19631 MT972844.1 |Severe acute respiratory syndrome coro .................... .................... ..................... .................... .................... ....................
19632 MT972845.1 |Severe acute respiratory syndrome coro .................... .................... ..................... .................... .................... ....................
19633 MT972846.1 |Severe acute respiratory syndrome coro .................... .................... ..................... .................... .................... ....................
19634 MT972847.1 |Severe acute respiratory syndrome coro .................... .................... ..................... .................... .................... ....................
19635 MT972848.1 |Severe acute respiratory syndrome coro .................... .................... ..................... .................... .................... ....................
19636 MT972849.1 |Severe acute respiratory syndrome coro .................... .................... ..................... .................... .................... ....................
19637 MT972850.1 |Severe acute respiratory syndrome coro .................... .................... ..................... .................... .................... ....................
19638 MT972851.1 |Severe acute respiratory syndrome coro .................... .................... ..................... .................... .................... ....................
19639 MT972853.1 |Severe acute respiratory syndrome coro .................... .................... ..................... .................... .................... ....................
19640 MT972854.1 |Severe acute respiratory syndrome coro .................... .................... ..................... .................... .................... ....................
19641 MT972856.1 |Severe acute respiratory syndrome coro .................... .................... ..................... .................... .................... ....................
19642 MT972857.1 |Severe acute respiratory syndrome coro .................... .................... ..................... .................... .................... ....................
19643 MT972858.1 |Severe acute respiratory syndrome coro .................... .................... ..................... .................... .................... ....................
19644 MT972859.1 |Severe acute respiratory syndrome coro .................... .................... ..................... .................... .................... ....................
19645 MT972860.1 |Severe acute respiratory syndrome coro .................... .................... ..................... .................... .................... ....................
19646 MT972861.1 |Severe acute respiratory syndrome coro .................... .................... ..................... .................... .................... ....................
19647 MT972862.1 |Severe acute respiratory syndrome coro .................... .................... ..................... .................... .................... ....................
19648 MT972863.1 |Severe acute respiratory syndrome coro .................... .................... ..................... .................... .................... ....................
19649 MT972864.1 |Severe acute respiratory syndrome coro .................... .................... ..................... .................... .................... ....................
19650 MT972865.1 |Severe acute respiratory syndrome coro .................... .................... ..................... .................... .................... ....................
19651 MT972866.1 |Severe acute respiratory syndrome coro .................... .................... ..................... .................... .................... ....................
19652 MT972867.1 |Severe acute respiratory syndrome coro .................... .................... ..................... .................... .................... ....................
19653 MT972868.1 |Severe acute respiratory syndrome coro .................... .................... ..................... .................... .................... ....................
19654 MT972869.1 |Severe acute respiratory syndrome coro .................... .................... ..................... .................... .................... ....................
19655 MT972870.1 |Severe acute respiratory syndrome coro .................... .................... ..................... .................... .................... ....................
19656 MT972871.1 |Severe acute respiratory syndrome coro .................... .................... ..................... .................... .................... ....................
19657 MT972872.1 |Severe acute respiratory syndrome coro .................... .................... ..................... .................... .................... ....................
19658 MT972873.1 |Severe acute respiratory syndrome coro .................... .................... ..................... .................... .................... ....................
19659 MT972874.1 |Severe acute respiratory syndrome coro .................... .................... ..................... .................... .................... ....................
19660 MT972876.1 |Severe acute respiratory syndrome coro .................... .................... ..................... .................... .................... ....................
19661 MT972877.1 |Severe acute respiratory syndrome coro .................... .................... ..................... .................... .................... ....................
19662 MT972878.1 |Severe acute respiratory syndrome coro .................... .................... ..................... .................... .................... ....................
19663 MT972879.1 |Severe acute respiratory syndrome coro .................... .................... ..................... .................... .................... ....................
19664 MT972881.1 |Severe acute respiratory syndrome coro .................... .................... ..................... .................... .................... ....................
19665 MT972883.1 |Severe acute respiratory syndrome coro .................... .................... ..................... .................... .................... ....................
19666 MT972884.1 |Severe acute respiratory syndrome coro .................... .................... ..................... .................... .................... ....................
19667 MT972885.1 |Severe acute respiratory syndrome coro .................... .................... ..................... .................... .................... ....................
19668 MT972886.1 |Severe acute respiratory syndrome coro .................... .................... ..................... .................... .................... ....................
19669 MT972887.1 |Severe acute respiratory syndrome coro .................... .................... ..................... .................... .................... ....................
19670 MT972888.1 |Severe acute respiratory syndrome coro .................... .................... ..................... .................... .................... ....................
19671 MT972889.1 |Severe acute respiratory syndrome coro .................... .................... ..................... .................... .................... ....................
19672 MT972890.1 |Severe acute respiratory syndrome coro .................... .................... ..................... .................... .................... ....................
19673 MT972891.1 |Severe acute respiratory syndrome coro .................... .................... ..................... .................... .................... ....................
19674 MT972892.1 |Severe acute respiratory syndrome coro .................... .................... ..................... .................... .................... ....................
19675 MT972893.1 |Severe acute respiratory syndrome coro .................... .................... ..................... .................... .................... ....................
19676 MT972894.1 |Severe acute respiratory syndrome coro .................... .................... ..................... .................... .................... ....................
19677 MT972895.1 |Severe acute respiratory syndrome coro .................... .................... ..................... .................... .................... ....................
19678 MT972896.1 |Severe acute respiratory syndrome coro .................... .................... ..................... .................... .................... ....................
19679 MT972898.1 |Severe acute respiratory syndrome coro .................... .................... ..................... .................... .................... ....................
19680 MT972899.1 |Severe acute respiratory syndrome coro .................... .................... ..................... .................... .................... ....................

| | | | | | | | | |
|---|---|---|---|---|---|---|---|---|
| 19681 | MT972900.1 | \|Severe acute respiratory syndrome coro | .................... | .................... | .................... | .................... | .................... | .................... |
| 19682 | MT972901.1 | \|Severe acute respiratory syndrome coro | .................... | .................... | .................... | .................... | .................... | .................... |
| 19683 | MT972902.1 | \|Severe acute respiratory syndrome coro | .................... | .................... | .................... | .................... | .................... | .................... |
| 19684 | MT972903.1 | \|Severe acute respiratory syndrome coro | .................... | .................... | .................... | .................... | .................... | .................... |
| 19685 | MT972904.1 | \|Severe acute respiratory syndrome coro | .................... | .................... | .................... | .................... | .................... | .................... |
| 19686 | MT972906.1 | \|Severe acute respiratory syndrome coro | .................... | .................... | .................... | .................... | .................... | .................... |
| 19687 | MT972907.1 | \|Severe acute respiratory syndrome coro | .................... | .................... | .................... | .................... | .................... | .................... |
| 19688 | MT972908.1 | \|Severe acute respiratory syndrome coro | .................... | .................... | .................... | .................... | .................... | .................... |
| 19689 | MT972909.1 | \|Severe acute respiratory syndrome coro | .................... | .................... | .................... | .................... | .................... | .................... |
| 19690 | MT972910.1 | \|Severe acute respiratory syndrome coro | .................... | .................... | .................... | .................... | .................... | .................... |
| 19691 | MT972911.1 | \|Severe acute respiratory syndrome coro | .................... | .................... | .................... | .................... | .................... | .................... |
| 19692 | MT972912.1 | \|Severe acute respiratory syndrome coro | .................... | .................... | .................... | .................... | .................... | .................... |
| 19693 | MT972913.1 | \|Severe acute respiratory syndrome coro | .................... | .................... | .................... | .................... | .................... | .................... |
| 19694 | MT972915.1 | \|Severe acute respiratory syndrome coro | .................... | .................... | .................... | .................... | .................... | .................... |
| 19695 | MT972916.1 | \|Severe acute respiratory syndrome coro | .................... | .................... | .................... | .................... | .................... | .................... |
| 19696 | MT972917.1 | \|Severe acute respiratory syndrome coro | .................... | .................... | .................... | .................... | .................... | .................... |
| 19697 | MT972919.1 | \|Severe acute respiratory syndrome coro | .................... | .................... | .................... | .................... | .................... | .................... |
| 19698 | MT972920.1 | \|Severe acute respiratory syndrome coro | .................... | .................... | .................... | .................... | .................... | .................... |
| 19699 | MT972921.1 | \|Severe acute respiratory syndrome coro | .................... | .................... | .................... | .................... | .................... | .................... |
| 19700 | MT972922.1 | \|Severe acute respiratory syndrome coro | .................... | .................... | .................... | .................... | .................... | .................... |
| 19701 | MT972923.1 | \|Severe acute respiratory syndrome coro | .................... | .................... | .................... | .................... | .................... | .................... |
| 19702 | MT972924.1 | \|Severe acute respiratory syndrome coro | .................... | .................... | .................... | .................... | .................... | .................... |
| 19703 | MT972925.1 | \|Severe acute respiratory syndrome coro | .................... | .................... | .................... | .................... | .................... | .................... |
| 19704 | MT972926.1 | \|Severe acute respiratory syndrome coro | .................... | .................... | .................... | .................... | .................... | .................... |
| 19705 | MT972927.1 | \|Severe acute respiratory syndrome coro | .................... | .................... | .................... | .................... | .................... | .................... |
| 19706 | MT972928.1 | \|Severe acute respiratory syndrome coro | .................... | .................... | .................... | .................... | .................... | .................... |
| 19707 | MT972929.1 | \|Severe acute respiratory syndrome coro | .................... | .................... | .................... | .................... | .................... | .................... |
| 19708 | MT972930.1 | \|Severe acute respiratory syndrome coro | .................... | .................... | .................... | .................... | .................... | .................... |
| 19709 | MT972931.1 | \|Severe acute respiratory syndrome coro | .................... | .................... | .................... | .................... | .................... | .................... |
| 19710 | MT972932.1 | \|Severe acute respiratory syndrome coro | .................... | .................... | .................... | .................... | .................... | .................... |
| 19711 | MT972933.1 | \|Severe acute respiratory syndrome coro | .................... | .................... | .................... | .................... | .................... | .................... |
| 19712 | MT972934.1 | \|Severe acute respiratory syndrome coro | .................... | .................... | .................... | .................... | .................... | .................... |
| 19713 | MT972935.1 | \|Severe acute respiratory syndrome coro | .................... | .................... | .................... | .................... | .................... | .................... |
| 19714 | MT972936.1 | \|Severe acute respiratory syndrome coro | .................... | .................... | .................... | .................... | .................... | .................... |
| 19715 | MT972937.1 | \|Severe acute respiratory syndrome coro | .................... | .................... | .................... | .................... | .................... | .................... |
| 19716 | MT972938.1 | \|Severe acute respiratory syndrome coro | .................... | .................... | .................... | .................... | .................... | .................... |
| 19717 | MT972939.1 | \|Severe acute respiratory syndrome coro | .................... | .................... | .................... | .................... | .................... | .................... |
| 19718 | MT972940.1 | \|Severe acute respiratory syndrome coro | .................... | .................... | .................... | .................... | .................... | .................... |
| 19719 | MT972942.1 | \|Severe acute respiratory syndrome coro | .................... | .................... | .................... | .................... | .................... | .................... |
| 19720 | MT972944.1 | \|Severe acute respiratory syndrome coro | .................... | .................... | .................... | .................... | .................... | .................... |
| 19721 | MT972946.1 | \|Severe acute respiratory syndrome coro | .................... | .................... | .................... | .................... | .................... | .................... |
| 19722 | MT972947.1 | \|Severe acute respiratory syndrome coro | .................... | .................... | .................... | .................... | .................... | .................... |
| 19723 | MT972948.1 | \|Severe acute respiratory syndrome coro | .................... | .................... | .................... | .................... | .................... | .................... |
| 19724 | MT972949.1 | \|Severe acute respiratory syndrome coro | .................... | .................... | .................... | .................... | .................... | .................... |
| 19725 | MT972951.1 | \|Severe acute respiratory syndrome coro | .................... | .................... | .................... | .................... | .................... | .................... |
| 19726 | MT972952.1 | \|Severe acute respiratory syndrome coro | .................... | .................... | .................... | .................... | .................... | .................... |
| 19727 | MT972954.1 | \|Severe acute respiratory syndrome coro | .................... | .................... | .................... | .................... | .................... | .................... |
| 19729 | MT972957.1 | \|Severe acute respiratory syndrome coro | .................... | .................... | .................... | .................... | .................... | .................... |
| 19730 | MT972959.1 | \|Severe acute respiratory syndrome coro | .................... | .................... | .................... | .................... | .................... | .................... |
| 19731 | MT972960.1 | \|Severe acute respiratory syndrome coro | .................... | .................... | .................... | .................... | .................... | .................... |
| 19732 | MT972962.1 | \|Severe acute respiratory syndrome coro | .................... | .................... | .................... | .................... | .................... | .................... |
| 19733 | MT972964.1 | \|Severe acute respiratory syndrome coro | .................... | .................... | .................... | .................... | .................... | .................... |
| 19734 | MT972965.1 | \|Severe acute respiratory syndrome coro | .................... | .................... | .................... | .................... | .................... | .................... |
| 19735 | MT972966.1 | \|Severe acute respiratory syndrome coro | .................... | .................... | .................... | .................... | .................... | .................... |
| 19736 | MT972967.1 | \|Severe acute respiratory syndrome coro | .................... | .................... | .................... | .................... | .................... | .................... |
| 19737 | MT972969.1 | \|Severe acute respiratory syndrome coro | .................... | .................... | .................... | .................... | .................... | .................... |
| 19738 | MT972970.1 | \|Severe acute respiratory syndrome coro | .................... | .................... | .................... | .................... | .................... | .................... |
| 19739 | MT972973.1 | \|Severe acute respiratory syndrome coro | .................... | .................... | .................... | .................... | .................... | .................... |
| 19740 | MT972974.1 | \|Severe acute respiratory syndrome coro | .................... | .................... | .................... | .................... | .................... | .................... |
| 19741 | MT972975.1 | \|Severe acute respiratory syndrome coro | .................... | .................... | .................... | .................... | .................... | .................... |
| 19742 | MT972976.1 | \|Severe acute respiratory syndrome coro | .................... | .................... | .................... | .................... | .................... | .................... |
| 19743 | MT972977.1 | \|Severe acute respiratory syndrome coro | .................... | .................... | .................... | .................... | .................... | .................... |
| 19744 | MT972978.1 | \|Severe acute respiratory syndrome coro | .................... | .................... | .................... | .................... | .................... | .................... |
| 19745 | MT972980.1 | \|Severe acute respiratory syndrome coro | .................... | .................... | .................... | .................... | .................... | .................... |
| 19746 | MT972981.1 | \|Severe acute respiratory syndrome coro | .................... | .................... | .................... | .................... | .................... | .................... |
| 19747 | MT972984.1 | \|Severe acute respiratory syndrome coro | .................... | .................... | .................... | .................... | .................... | .................... |
| 19748 | MT972985.1 | \|Severe acute respiratory syndrome coro | .................... | .................... | .................... | .................... | .................... | .................... |
| 19749 | MT972986.1 | \|Severe acute respiratory syndrome coro | .................... | .................... | .................... | .................... | .................... | .................... |
| 19750 | MT972988.1 | \|Severe acute respiratory syndrome coro | .................... | .................... | .................... | .................... | .................... | .................... |
| 19751 | MT972989.1 | \|Severe acute respiratory syndrome coro | .................... | .................... | .................... | .................... | .................... | .................... |
| 19752 | MT972991.1 | \|Severe acute respiratory syndrome coro | .................... | .................... | .................... | .................... | .................... | .................... |
| 19753 | MT972992.1 | \|Severe acute respiratory syndrome coro | .................... | .................... | .................... | .................... | .................... | .................... |
| 19754 | MT972993.1 | \|Severe acute respiratory syndrome coro | .................... | .................... | .................... | .................... | .................... | .................... |
| 19755 | MT972994.1 | \|Severe acute respiratory syndrome coro | .................... | .................... | .................... | .................... | .................... | .................... |
| 19756 | MT972995.1 | \|Severe acute respiratory syndrome coro | .................... | .................... | .................... | .................... | .................... | .................... |
| 19757 | MT972996.1 | \|Severe acute respiratory syndrome coro | .................... | .................... | .................... | .................... | .................... | .................... |
| 19758 | MT972997.1 | \|Severe acute respiratory syndrome coro | .................... | .................... | .................... | .................... | .................... | .................... |
| 19759 | MT972998.1 | \|Severe acute respiratory syndrome coro | .................... | .................... | .................... | .................... | .................... | .................... |

```
19760 MT973000.1 |Severe acute respiratory syndrome coro    ....................  ....................  ....................  ....................  ....................  ....................
19761 MT973001.1 |Severe acute respiratory syndrome coro    ....................  ....................  ....................  ....................  ....................  ....................
19762 MT973002.1 |Severe acute respiratory syndrome coro    ....................  ....................  ....................  ....................  ....................  ....................
19763 MT973003.1 |Severe acute respiratory syndrome coro    ....................  ....................  ....................  ....................  ....................  ....................
19764 MT973004.1 |Severe acute respiratory syndrome coro    ....................  ....................  ....................  ....................  ....................  ....................
19765 MT973005.1 |Severe acute respiratory syndrome coro    ....................  ....................  ....................  ....................  ....................  ....................
19766 MT973006.1 |Severe acute respiratory syndrome coro    ....................  ....................  ....................  ....................  ....................  ....................
19767 MT973008.1 |Severe acute respiratory syndrome coro    ....................  ....................  ....................  ....................  ....................  ....................
19768 MT973009.1 |Severe acute respiratory syndrome coro    ....................  ....................  ....................  ....................  ....................  ....................
19769 MT973011.1 |Severe acute respiratory syndrome coro    ....................  ....................  ....................  ....................  ....................  ....................
19770 MT973013.1 |Severe acute respiratory syndrome coro    ....................  ....................  ....................  ....................  ....................  ....................
19771 MT973014.1 |Severe acute respiratory syndrome coro    ....................  ....................  ....................  ....................  ....................  ....................
19772 MT973015.1 |Severe acute respiratory syndrome coro    ....................  ....................  ....................  ....................  ....................  ....................
19773 MT973016.1 |Severe acute respiratory syndrome coro    ....................  ....................  ....................  ....................  ....................  ....................
19774 MT973017.1 |Severe acute respiratory syndrome coro    ....................  ....................  ....................  ....................  ....................  ....................
19775 MT973018.1 |Severe acute respiratory syndrome coro    ....................  ....................  ....................  ....................  ....................  ....................
19776 MT973019.1 |Severe acute respiratory syndrome coro    ....................  ....................  ....................  ....................  ....................  ....................
19777 MT973020.1 |Severe acute respiratory syndrome coro    ....................  ....................  ....................  ....................  ....................  ....................
19778 MT973022.1 |Severe acute respiratory syndrome coro    ....................  ....................  ....................  ....................  ....................  ....................
19779 MT973023.1 |Severe acute respiratory syndrome coro    ....................  ....................  ....................  ....................  ....................  ....................
19780 MT973024.1 |Severe acute respiratory syndrome coro    ....................  ....................  ....................  ....................  ....................  ....................
19781 MT973025.1 |Severe acute respiratory syndrome coro    ....................  ....................  ....................  ....................  ....................  ....................
19782 MT973026.1 |Severe acute respiratory syndrome coro    ....................  ....................  ....................  ....................  ....................  ....................
19783 MT973027.1 |Severe acute respiratory syndrome coro    ....................  ....................  ....................  ....................  ....................  ....................
19784 MT973028.1 |Severe acute respiratory syndrome coro    ....................  ....................  ....................  ....................  ....................  ....................
19785 MT973029.1 |Severe acute respiratory syndrome coro    ....................  ....................  ....................  ....................  ....................  ....................
19786 MT973030.1 |Severe acute respiratory syndrome coro    ....................  ....................  ....................  ....................  ....................  ....................
19787 MT973031.1 |Severe acute respiratory syndrome coro    ....................  ....................  ....................  ....................  ....................  ....................
19788 MT973032.1 |Severe acute respiratory syndrome coro    ....................  ....................  ....................  ....................  ....................  ....................
19789 MT973033.1 |Severe acute respiratory syndrome coro    ....................  ....................  ....................  ....................  ....................  ....................
19790 MT973034.1 |Severe acute respiratory syndrome coro    ....................  ....................  ....................  ....................  ....................  ....................
19791 MT973035.1 |Severe acute respiratory syndrome coro    ....................  ....................  ....................  ....................  ....................  ....................
19792 MT973037.1 |Severe acute respiratory syndrome coro    ....................  ....................  ....................  ....................  ....................  ....................
19793 MT973038.1 |Severe acute respiratory syndrome coro    ....................  ....................  ....................  ....................  ....................  ....................
19794 MT973039.1 |Severe acute respiratory syndrome coro    ....................  ....................  ....................  ....................  ....................  ....................
19795 MT973040.1 |Severe acute respiratory syndrome coro    ....................  ....................  ....................  ....................  ....................  ....................
19796 MT973041.1 |Severe acute respiratory syndrome coro    ....................  ....................  ....................  ....................  ....................  ....................
19797 MT973042.1 |Severe acute respiratory syndrome coro    ....................  ....................  ....................  ....................  ....................  ....................
19798 MT973043.1 |Severe acute respiratory syndrome coro    ....................  ....................  ....................  ....................  ....................  ....................
19799 MT973044.1 |Severe acute respiratory syndrome coro    ....................  ....................  ....................  ....................  ....................  ....................
19800 MT973045.1 |Severe acute respiratory syndrome coro    ....................  ....................  ....................  ....................  ....................  ....................
19801 MT973046.1 |Severe acute respiratory syndrome coro    ....................  ....................  ....................  ....................  ....................  ....................
19802 MT973048.1 |Severe acute respiratory syndrome coro    ....................  ....................  ....................  ....................  ....................  ....................
19803 MT973049.1 |Severe acute respiratory syndrome coro    ....................  ....................  ....................  ....................  ....................  ....................
19804 MT973050.1 |Severe acute respiratory syndrome coro    ....................  ....................  ....................  ....................  ....................  ....................
19805 MT973051.1 |Severe acute respiratory syndrome coro    ....................  ....................  ....................  ....................  ....................  ....................
19806 MT973052.1 |Severe acute respiratory syndrome coro    ....................  ....................  ....................  ....................  ....................  ....................
19807 MT973053.1 |Severe acute respiratory syndrome coro    ....................  ....................  ....................  ....................  ....................  ....................
19808 MT973054.1 |Severe acute respiratory syndrome coro    ....................  ....................  ....................  ....................  ....................  ....................
19809 MT973055.1 |Severe acute respiratory syndrome coro    ....................  ....................  ....................  ....................  ....................  ....................
19810 MT973056.1 |Severe acute respiratory syndrome coro    ....................  ....................  ....................  ....................  ....................  ....................
19811 MT973057.1 |Severe acute respiratory syndrome coro    ....................  ....................  ....................  ....................  ....................  ....................
19812 MT973059.1 |Severe acute respiratory syndrome coro    ....................  ....................  ....................  ....................  ....................  ....................
19813 MT973061.1 |Severe acute respiratory syndrome coro    ....................  ....................  ....................  ....................  ....................  ....................
19814 MT973062.1 |Severe acute respiratory syndrome coro    ....................  ....................  ....................  ....................  ....................  ....................
19815 MT973063.1 |Severe acute respiratory syndrome coro    ....................  ....................  ....................  ....................  ....................  ....................
19816 MT973065.1 |Severe acute respiratory syndrome coro    ....................  ....................  ....................  ....................  ....................  ....................
19817 MT973066.1 |Severe acute respiratory syndrome coro    ....................  ....................  ....................  ....................  ....................  ....................
19818 MT973067.1 |Severe acute respiratory syndrome coro    ....................  ....................  ....................  ....................  ....................  ....................
19819 MT973068.1 |Severe acute respiratory syndrome coro    ....................  ....................  ....................  ....................  ....................  ....................
19820 MT973069.1 |Severe acute respiratory syndrome coro    ....................  ....................  ....................  ....................  ....................  ....................
19821 MT973070.1 |Severe acute respiratory syndrome coro    ....................  ....................  ....................  ....................  ....................  ....................
19822 MT973071.1 |Severe acute respiratory syndrome coro    ....................  ....................  ....................  ....................  ....................  ....................
19823 MT973072.1 |Severe acute respiratory syndrome coro    ....................  ....................  ....................  ....................  ....................  ....................
19824 MT973073.1 |Severe acute respiratory syndrome coro    ....................  ....................  ....................  ....................  ....................  ....................
19825 MT973074.1 |Severe acute respiratory syndrome coro    ....................  ....................  ....................  ....................  ....................  ....................
19826 MT973075.1 |Severe acute respiratory syndrome coro    ....................  ....................  ....................  ....................  ....................  ....................
19827 MT973077.1 |Severe acute respiratory syndrome coro    ....................  ....................  ....................  ....................  ....................  ....................
19828 MT973080.1 |Severe acute respiratory syndrome coro    ....................  ....................  ....................  ....................  ....................  ....................
19829 MT973081.1 |Severe acute respiratory syndrome coro    ....................  ....................  ....................  ....................  ....................  ....................
19830 MT973082.1 |Severe acute respiratory syndrome coro    ....................  ....................  ....................  ....................  ....................  ....................
19831 MT973083.1 |Severe acute respiratory syndrome coro    ....................  ....................  ....................  ....................  ....................  ....................
19832 MT973084.1 |Severe acute respiratory syndrome coro    ....................  ....................  ....................  ....................  ....................  ....................
19833 MT973085.1 |Severe acute respiratory syndrome coro    ....................  ....................  ....................  ....................  ....................  ....................
19834 MT973086.1 |Severe acute respiratory syndrome coro    ....................  ....................  ....................  ....................  ....................  ....................
19835 MT973088.1 |Severe acute respiratory syndrome coro    ....................  ....................  ....................  ....................  ....................  ....................
19836 MT973089.1 |Severe acute respiratory syndrome coro    ....................  ....................  ....................  ....................  ....................  ....................
19837 MT973090.1 |Severe acute respiratory syndrome coro    ....................  ....................  ....................  ....................  ....................  ....................
```

19838 MT973091.1 |Severe acute respiratory syndrome coro .................... .................... ..................... .................... .................... ....................
19839 MT973092.1 |Severe acute respiratory syndrome coro .................... .................... ..................... .................... .................... ....................
19840 MT973093.1 |Severe acute respiratory syndrome coro .................... .................... ..................... .................... .................... ....................
19841 MT973095.1 |Severe acute respiratory syndrome coro .................... .................... ..................... .................... .................... ....................
19842 MT973096.1 |Severe acute respiratory syndrome coro .................... .................... ..................... .................... .................... ....................
19843 MT973097.1 |Severe acute respiratory syndrome coro .................... .................... ..................... .................... .................... ....................
19844 MT973098.1 |Severe acute respiratory syndrome coro .................... .................... ..................... .................... .................... ....................
19845 MT973100.1 |Severe acute respiratory syndrome coro .................... .................... ..................... .................... .................... ....................
19846 MT973101.1 |Severe acute respiratory syndrome coro .................... .................... ..................... .................... .................... ....................
19847 MT973103.1 |Severe acute respiratory syndrome coro .................... .................... ..................... .................... .................... ....................
19848 MT973104.1 |Severe acute respiratory syndrome coro .................... .................... ..................... .................... .................... ....................
19849 MT973105.1 |Severe acute respiratory syndrome coro .................... .................... ..................... .................... .................... ....................
19850 MT973106.1 |Severe acute respiratory syndrome coro .................... .................... ..................... .................... .................... ....................
19851 MT973107.1 |Severe acute respiratory syndrome coro .................... .................... ..................... .................... .................... ....................
19852 MT973108.1 |Severe acute respiratory syndrome coro .................... .................... ..................... .................... .................... ....................
19853 MT973109.1 |Severe acute respiratory syndrome coro .................... .................... ..................... .................... .................... ....................
19854 MT973110.1 |Severe acute respiratory syndrome coro .................... .................... ..................... .................... .................... ....................
19855 MT973111.1 |Severe acute respiratory syndrome coro .................... .................... ..................... .................... .................... ....................
19856 MT973112.1 |Severe acute respiratory syndrome coro .................... .................... ..................... .................... .................... ....................
19857 MT973113.1 |Severe acute respiratory syndrome coro .................... .................... ..................... .................... .................... ....................
19858 MT973114.1 |Severe acute respiratory syndrome coro .................... .................... ..................... .................... .................... ....................
19859 MT973115.1 |Severe acute respiratory syndrome coro .................... .................... ..................... .................... .................... ....................
19860 MT973116.1 |Severe acute respiratory syndrome coro .................... .................... ..................... .................... .................... ....................
19861 MT973117.1 |Severe acute respiratory syndrome coro .................... .................... ..................... .................... .................... ....................
19862 MT973118.1 |Severe acute respiratory syndrome coro .................... .................... ..................... .................... .................... ....................
19863 MT973119.1 |Severe acute respiratory syndrome coro .................... .................... ..................... .................... .................... ....................
19864 MT973120.1 |Severe acute respiratory syndrome coro .................... .................... ..................... .................... .................... ....................
19865 MT973121.1 |Severe acute respiratory syndrome coro .................... .................... ..................... .................... .................... ....................
19866 MT973123.1 |Severe acute respiratory syndrome coro .................... .................... ..................... .................... .................... ....................
19867 MT973125.1 |Severe acute respiratory syndrome coro .................... .................... ..................... .................... .................... ....................
19868 MT973126.1 |Severe acute respiratory syndrome coro .................... .................... ..................... .................... .................... ....................
19869 MT973127.1 |Severe acute respiratory syndrome coro .................... .................... ..................... .................... .................... ....................
19870 MT973128.1 |Severe acute respiratory syndrome coro .................... .................... ..................... .................... .................... ....................
19871 MT973129.1 |Severe acute respiratory syndrome coro .................... .................... ..................... .................... .................... ....................
19872 MT973130.1 |Severe acute respiratory syndrome coro .................... .................... ..................... .................... .................... ....................
19873 MT973132.1 |Severe acute respiratory syndrome coro .................... .................... ..................... .................... .................... ....................
19874 MT973133.1 |Severe acute respiratory syndrome coro .................... .................... ..................... .................... .................... ....................
19875 MT973134.1 |Severe acute respiratory syndrome coro .................... .................... ..................... .................... .................... ....................
19876 MT973135.1 |Severe acute respiratory syndrome coro .................... .................... ..................... .................... .................... ....................
19877 MT973136.1 |Severe acute respiratory syndrome coro .................... .................... ..................... .................... .................... ....................
19878 MT973137.1 |Severe acute respiratory syndrome coro .................... .................... ..................... .................... .................... ....................
19879 MT973139.1 |Severe acute respiratory syndrome coro .................... .................... ..................... .................... .................... ....................
19880 MT973141.1 |Severe acute respiratory syndrome coro .................... .................... ..................... .................... .................... ....................
19881 MT973142.1 |Severe acute respiratory syndrome coro .................... .................... ..................... .................... .................... ....................
19882 MT973143.1 |Severe acute respiratory syndrome coro .................... .................... ..................... .................... .................... ....................
19883 MT973144.1 |Severe acute respiratory syndrome coro .................... .................... ..................... .................... .................... ....................
19884 MT973145.1 |Severe acute respiratory syndrome coro .................... .................... ..................... .................... .................... ....................
19885 MT973146.1 |Severe acute respiratory syndrome coro .................... .................... ..................... .................... .................... ....................
19886 MT973147.1 |Severe acute respiratory syndrome coro .................... .................... ..................... .................... .................... ....................
19887 MT973148.1 |Severe acute respiratory syndrome coro .................... .................... ..................... .................... .................... ....................
19888 MT973150.1 |Severe acute respiratory syndrome coro .................... .................... ..................... .................... .................... ....................
19889 MT973151.1 |Severe acute respiratory syndrome coro .................... .................... ..................... .................... .................... ....................
19890 MT973152.1 |Severe acute respiratory syndrome coro .................... .................... ..................... .................... .................... ....................
19891 MT973153.1 |Severe acute respiratory syndrome coro .................... .................... ..................... .................... .................... ....................
19892 MT973154.1 |Severe acute respiratory syndrome coro .................... .................... ..................... .................... .................... ....................
19893 MT973155.1 |Severe acute respiratory syndrome coro .................... .................... ..................... .................... .................... ....................
19894 MT973156.1 |Severe acute respiratory syndrome coro .................... .................... ..................... .................... .................... ....................
19895 MT973157.1 |Severe acute respiratory syndrome coro .................... .................... ..................... .................... .................... ....................
19896 MT973158.1 |Severe acute respiratory syndrome coro .................... .................... ..................... .................... .................... ....................
19897 MT973159.1 |Severe acute respiratory syndrome coro .................... .................... ..................... .................... .................... ....................
19898 MT973160.1 |Severe acute respiratory syndrome coro .................... .................... ..................... .................... .................... ....................
19899 MT973161.1 |Severe acute respiratory syndrome coro .................... .................... ..................... .................... .................... ....................
19900 MT973162.1 |Severe acute respiratory syndrome coro .................... .................... ..................... .................... .................... ....................
19901 MT973163.1 |Severe acute respiratory syndrome coro .................... .................... ..................... .................... .................... ....................
19902 MT973164.1 |Severe acute respiratory syndrome coro .................... .................... ..................... .................... .................... ....................
19903 MT973165.1 |Severe acute respiratory syndrome coro .................... .................... ..................... .................... .................... ....................
19904 MT973166.1 |Severe acute respiratory syndrome coro .................... .................... ..................... .................... .................... ....................
19905 MT973167.1 |Severe acute respiratory syndrome coro .................... .................... ..................... .................... .................... ....................
19906 MT973168.1 |Severe acute respiratory syndrome coro .................... .................... ..................... .................... .................... ....................
19907 MT973169.1 |Severe acute respiratory syndrome coro .................... .................... ..................... .................... .................... ....................
19908 MT973170.1 |Severe acute respiratory syndrome coro .................... .................... ..................... .................... .................... ....................
19909 MT973171.1 |Severe acute respiratory syndrome coro .................... .................... ..................... .................... .................... ....................
19910 MT973172.1 |Severe acute respiratory syndrome coro .................... .................... ..................... .................... .................... ....................
19911 MT973173.1 |Severe acute respiratory syndrome coro .................... .................... ..................... .................... .................... ....................
19912 MT973174.1 |Severe acute respiratory syndrome coro .................... .................... ..................... .................... .................... ....................
19913 MT973175.1 |Severe acute respiratory syndrome coro .................... .................... ..................... .................... .................... ....................
19914 MT973176.1 |Severe acute respiratory syndrome coro .................... .................... ..................... .................... .................... ....................
19915 MT973177.1 |Severe acute respiratory syndrome coro .................... .................... ..................... .................... .................... ....................

```
19916 MT973178.1 |Severe acute respiratory syndrome coro    ....................  ....................  .....................  ....................  ....................  ....................
19917 MT973179.1 |Severe acute respiratory syndrome coro    ....................  ....................  .....................  ....................  ....................  ....................
19918 MT973180.1 |Severe acute respiratory syndrome coro    ....................  ....................  .....................  ....................  ....................  ....................
19919 MT973181.1 |Severe acute respiratory syndrome coro    ....................  ....................  .....................  ....................  ....................  ....................
19920 MT973182.1 |Severe acute respiratory syndrome coro    ....................  ....................  .....................  ....................  ....................  ....................
19921 MT973183.1 |Severe acute respiratory syndrome coro    ....................  ....................  .....................  ....................  ....................  ....................
19922 MT973184.1 |Severe acute respiratory syndrome coro    ....................  ....................  .....................  ....................  ....................  ....................
19923 MT973185.1 |Severe acute respiratory syndrome coro    ....................  ....................  .....................  ....................  ....................  ....................
19924 MT973186.1 |Severe acute respiratory syndrome coro    ....................  ....................  .....................  ....................  ....................  ....................
19925 MT973187.1 |Severe acute respiratory syndrome coro    ....................  ....................  .....................  ....................  ....................  ....................
19926 MT973189.1 |Severe acute respiratory syndrome coro    ....................  ....................  .....................  ....................  ....................  ....................
19927 MT973190.1 |Severe acute respiratory syndrome coro    ....................  ....................  .....................  ....................  ....................  ....................
19928 MT973191.1 |Severe acute respiratory syndrome coro    ....................  ....................  .....................  ....................  ....................  ....................
19929 MT973192.1 |Severe acute respiratory syndrome coro    ....................  ....................  .....................  ....................  ....................  ....................
19930 MT973193.1 |Severe acute respiratory syndrome coro    ....................  ....................  .....................  ....................  ....................  ....................
19931 MT973194.1 |Severe acute respiratory syndrome coro    ....................  ....................  .....................  ....................  ....................  ....................
19932 MT973195.1 |Severe acute respiratory syndrome coro    ....................  ....................  .....................  ....................  ....................  ....................
19933 MT973196.1 |Severe acute respiratory syndrome coro    ....................  ....................  .....................  ....................  ....................  ....................
19934 MT973197.1 |Severe acute respiratory syndrome coro    ....................  ....................  .....................  ....................  ....................  ....................
19935 MT973198.1 |Severe acute respiratory syndrome coro    ....................  ....................  .....................  ....................  ....................  ....................
19936 MT973199.1 |Severe acute respiratory syndrome coro    ....................  ....................  .....................  ....................  ....................  ....................
19937 MT973200.1 |Severe acute respiratory syndrome coro    ....................  ....................  .....................  ....................  ....................  ....................
19938 MT973201.1 |Severe acute respiratory syndrome coro    ....................  ....................  .....................  ....................  ....................  ....................
19939 MT973202.1 |Severe acute respiratory syndrome coro    ....................  ....................  .....................  ....................  ....................  ....................
19940 MT973203.1 |Severe acute respiratory syndrome coro    ....................  ....................  .....................  ....................  ....................  ....................
19941 MT973204.1 |Severe acute respiratory syndrome coro    ....................  ....................  .....................  ....................  ....................  ....................
19942 MT973205.1 |Severe acute respiratory syndrome coro    ....................  ....................  .....................  ....................  ....................  ....................
19943 MT973206.1 |Severe acute respiratory syndrome coro    ....................  ....................  .....................  ....................  ....................  ....................
19945 MT973208.1 |Severe acute respiratory syndrome coro    ....................  ....................  .....................  ....................  ....................  ....................
19946 MT973209.1 |Severe acute respiratory syndrome coro    ....................  ....................  .....................  ....................  ....................  ....................
19947 MT973210.1 |Severe acute respiratory syndrome coro    ....................  ....................  .....................  ....................  ....................  ....................
19948 MT973211.1 |Severe acute respiratory syndrome coro    ....................  ....................  .....................  ....................  ....................  ....................
19949 MT973212.1 |Severe acute respiratory syndrome coro    ....................  ....................  .....................  ....................  ....................  ....................
19950 MT973213.1 |Severe acute respiratory syndrome coro    ....................  ....................  .....................  ....................  ....................  ....................
19951 MT973214.1 |Severe acute respiratory syndrome coro    ....................  ....................  .....................  ....................  ....................  ....................
19952 MT973216.1 |Severe acute respiratory syndrome coro    ....................  ....................  .....................  ....................  ....................  ....................
19953 MT973217.1 |Severe acute respiratory syndrome coro    ....................  ....................  .....................  ....................  ....................  ....................
19954 MT973218.1 |Severe acute respiratory syndrome coro    ....................  ....................  .....................  ....................  ....................  ....................
19955 MT973220.1 |Severe acute respiratory syndrome coro    ....................  ....................  .....................  ....................  ....................  ....................
19956 MT973221.1 |Severe acute respiratory syndrome coro    ....................  ....................  .....................  ....................  ....................  ....................
19957 MT973222.1 |Severe acute respiratory syndrome coro    ....................  ....................  .....................  ....................  ....................  ....................
19958 MT973225.1 |Severe acute respiratory syndrome coro    ....................  ....................  .....................  ....................  ....................  ....................
19959 MT973226.1 |Severe acute respiratory syndrome coro    ....................  ....................  .....................  ....................  ....................  ....................
19960 MT973227.1 |Severe acute respiratory syndrome coro    ....................  ....................  .....................  ....................  ....................  ....................
19961 MT973228.1 |Severe acute respiratory syndrome coro    ....................  ....................  .....................  ....................  ....................  ....................
19962 MT973230.1 |Severe acute respiratory syndrome coro    ....................  ....................  .....................  ....................  ....................  ....................
19963 MT973231.1 |Severe acute respiratory syndrome coro    ....................  ....................  .....................  ....................  ....................  ....................
19964 MT973232.1 |Severe acute respiratory syndrome coro    ....................  ....................  .....................  ....................  ....................  ....................
19965 MT973233.1 |Severe acute respiratory syndrome coro    ....................  ....................  .....................  ....................  ....................  ....................
19966 MT973234.1 |Severe acute respiratory syndrome coro    ....................  ....................  .....................  ....................  ....................  ....................
19967 MT973235.1 |Severe acute respiratory syndrome coro    ....................  ....................  .....................  ....................  ....................  ....................
19968 MT973236.1 |Severe acute respiratory syndrome coro    ....................  ....................  .....................  ....................  ....................  ....................
19969 MT973237.1 |Severe acute respiratory syndrome coro    ....................  ....................  .....................  ....................  ....................  ....................
19970 MT973239.1 |Severe acute respiratory syndrome coro    ....................  ....................  .....................  ....................  ....................  ....................
19971 MT973241.1 |Severe acute respiratory syndrome coro    ....................  ....................  .....................  ....................  ....................  ....................
19972 MT973242.1 |Severe acute respiratory syndrome coro    ....................  ....................  .....................  ....................  ....................  ....................
19973 MT973244.1 |Severe acute respiratory syndrome coro    ....................  ....................  .....................  ....................  ....................  ....................
19974 MT973245.1 |Severe acute respiratory syndrome coro    ....................  ....................  .....................  ....................  ....................  ....................
19975 MT973246.1 |Severe acute respiratory syndrome coro    ....................  ....................  .....................  ....................  ....................  ....................
19976 MT973247.1 |Severe acute respiratory syndrome coro    ....................  ....................  .....................  ....................  ....................  ....................
19977 MT973249.1 |Severe acute respiratory syndrome coro    ....................  ....................  .....................  ....................  ....................  ....................
19978 MT973250.1 |Severe acute respiratory syndrome coro    ....................  ....................  .....................  ....................  ....................  ....................
19979 MT973251.1 |Severe acute respiratory syndrome coro    ....................  ....................  .....................  ....................  ....................  ....................
19980 MT973252.1 |Severe acute respiratory syndrome coro    ....................  ....................  .....................  ....................  ....................  ....................
19981 MT973253.1 |Severe acute respiratory syndrome coro    ....................  ....................  .....................  ....................  ....................  ....................
19982 MT973254.1 |Severe acute respiratory syndrome coro    ....................  ....................  .....................  ....................  ....................  ....................
19983 MT973255.1 |Severe acute respiratory syndrome coro    ....................  ....................  .....................  ....................  ....................  ....................
19984 MT973256.1 |Severe acute respiratory syndrome coro    ....................  ....................  .....................  ....................  ....................  ....................
19985 MT973257.1 |Severe acute respiratory syndrome coro    ....................  ....................  .....................  ....................  ....................  ....................
19986 MT973258.1 |Severe acute respiratory syndrome coro    ....................  ....................  .....................  ....................  ....................  ....................
19987 MT973259.1 |Severe acute respiratory syndrome coro    ....................  ....................  .....................  ....................  ....................  ....................
19988 MT973260.1 |Severe acute respiratory syndrome coro    ....................  ....................  .....................  ....................  ....................  ....................
19989 MT973262.1 |Severe acute respiratory syndrome coro    ....................  ....................  .....................  ....................  ....................  ....................
19990 MT973263.1 |Severe acute respiratory syndrome coro    ....................  ....................  .....................  ....................  ....................  ....................
19991 MT973264.1 |Severe acute respiratory syndrome coro    ....................  ....................  .....................  ....................  ....................  ....................
19992 MT973265.1 |Severe acute respiratory syndrome coro    ....................  ....................  .....................  ....................  ....................  ....................
19993 MT973266.1 |Severe acute respiratory syndrome coro    ....................  ....................  .....................  ....................  ....................  ....................
19994 MT973267.1 |Severe acute respiratory syndrome coro    ....................  ....................  .....................  ....................  ....................  ....................
```

| | | | | | | | | | |
|---|---|---|---|---|---|---|---|---|---|
| 19995 | MT973269.1 | \|Severe acute respiratory syndrome coro | ···················· | ···················· | ···················· | ···················· | ···················· | ···················· | |
| 19996 | MT973270.1 | \|Severe acute respiratory syndrome coro | ···················· | ···················· | ···················· | ···················· | ···················· | ···················· | |
| 19997 | MT973271.1 | \|Severe acute respiratory syndrome coro | ···················· | ···················· | ···················· | ···················· | ···················· | ···················· | |
| 19998 | MT973272.1 | \|Severe acute respiratory syndrome coro | ···················· | ···················· | ···················· | ···················· | ···················· | ···················· | |
| 19999 | MT973273.1 | \|Severe acute respiratory syndrome coro | ···················· | ···················· | ···················· | ···················· | ···················· | ···················· | |
| 20000 | MT973274.1 | \|Severe acute respiratory syndrome coro | ···················· | ···················· | ···················· | ···················· | ···················· | ···················· | |
| 20001 | MT973275.1 | \|Severe acute respiratory syndrome coro | ···················· | ···················· | ···················· | ···················· | ···················· | ···················· | |
| 20002 | MT973276.1 | \|Severe acute respiratory syndrome coro | ···················· | ···················· | ···················· | ···················· | ···················· | ···················· | |
| 20003 | MT973278.1 | \|Severe acute respiratory syndrome coro | ···················· | ···················· | ···················· | ···················· | ···················· | ···················· | |
| 20004 | MT973279.1 | \|Severe acute respiratory syndrome coro | ···················· | ···················· | ···················· | ···················· | ···················· | ···················· | |
| 20005 | MT973280.1 | \|Severe acute respiratory syndrome coro | ···················· | ···················· | ···················· | ···················· | ···················· | ···················· | |
| 20006 | MT973281.1 | \|Severe acute respiratory syndrome coro | ···················· | ···················· | ···················· | ···················· | ···················· | ···················· | |
| 20007 | MT973282.1 | \|Severe acute respiratory syndrome coro | ···················· | ···················· | ···················· | ···················· | ···················· | ···················· | |
| 20008 | MT973283.1 | \|Severe acute respiratory syndrome coro | ···················· | ···················· | ···················· | ···················· | ···················· | ···················· | |
| 20009 | MT973284.1 | \|Severe acute respiratory syndrome coro | ···················· | ···················· | ···················· | ···················· | ···················· | ···················· | |
| 20010 | MT973285.1 | \|Severe acute respiratory syndrome coro | ···················· | ···················· | ···················· | ···················· | ···················· | ···················· | |
| 20011 | MT973286.1 | \|Severe acute respiratory syndrome coro | ···················· | ···················· | ···················· | ···················· | ···················· | ···················· | |
| 20012 | MT973288.1 | \|Severe acute respiratory syndrome coro | ···················· | ···················· | ···················· | ···················· | ···················· | ···················· | |
| 20013 | MT973289.1 | \|Severe acute respiratory syndrome coro | ···················· | ···················· | ···················· | ···················· | ···················· | ···················· | |
| 20014 | MT973290.1 | \|Severe acute respiratory syndrome coro | ···················· | ···················· | ···················· | ···················· | ···················· | ···················· | |
| 20015 | MT973291.1 | \|Severe acute respiratory syndrome coro | ···················· | ···················· | ···················· | ···················· | ···················· | ···················· | |
| 20016 | MT973292.1 | \|Severe acute respiratory syndrome coro | ···················· | ···················· | ···················· | ···················· | ···················· | ···················· | |
| 20017 | MT973294.1 | \|Severe acute respiratory syndrome coro | ···················· | ···················· | ···················· | ···················· | ···················· | ···················· | |
| 20018 | MT973295.1 | \|Severe acute respiratory syndrome coro | ···················· | ···················· | ···················· | ···················· | ···················· | ···················· | |
| 20019 | MT973296.1 | \|Severe acute respiratory syndrome coro | ···················· | ···················· | ···················· | ···················· | ···················· | ···················· | |
| 20020 | MT973297.1 | \|Severe acute respiratory syndrome coro | ···················· | ···················· | ···················· | ···················· | ···················· | ···················· | |
| 20021 | MT973298.1 | \|Severe acute respiratory syndrome coro | ···················· | ···················· | ···················· | ···················· | ···················· | ···················· | |
| 20022 | MT973299.1 | \|Severe acute respiratory syndrome coro | ···················· | ···················· | ···················· | ···················· | ···················· | ···················· | |
| 20023 | MT973300.1 | \|Severe acute respiratory syndrome coro | ···················· | ···················· | ···················· | ···················· | ···················· | ···················· | |
| 20024 | MT973301.1 | \|Severe acute respiratory syndrome coro | ···················· | ···················· | ···················· | ···················· | ···················· | ···················· | |
| 20025 | MT973302.1 | \|Severe acute respiratory syndrome coro | ···················· | ···················· | ···················· | ···················· | ···················· | ···················· | |
| 20026 | MT973304.1 | \|Severe acute respiratory syndrome coro | ···················· | ···················· | ···················· | ···················· | NNN················· | ···················· | 1 |
| 20028 | MT973306.1 | \|Severe acute respiratory syndrome coro | ···················· | ···················· | ···················· | ···················· | ···················· | ···················· | |
| 20029 | MT973307.1 | \|Severe acute respiratory syndrome coro | ···················· | ···················· | ···················· | ···················· | ···················· | ···················· | |
| 20030 | MT973308.1 | \|Severe acute respiratory syndrome coro | ···················· | ···················· | ···················· | ···················· | ···················· | ···················· | |
| 20031 | MT973310.1 | \|Severe acute respiratory syndrome coro | ···················· | ···················· | ···················· | ···················· | ···················· | ···················· | |
| 20032 | MT973311.1 | \|Severe acute respiratory syndrome coro | ···················· | ···················· | ···················· | ···················· | ···················· | ···················· | |
| 20034 | MT973313.1 | \|Severe acute respiratory syndrome coro | ···················· | ···················· | ···················· | ···················· | ···················· | ···················· | |
| 20035 | MT973314.1 | \|Severe acute respiratory syndrome coro | ···················· | ···················· | ···················· | ···················· | ···················· | ···················· | |
| 20036 | MT973315.1 | \|Severe acute respiratory syndrome coro | ···················· | ···················· | ···················· | ···················· | ···················· | ···················· | |
| 20037 | MT973316.1 | \|Severe acute respiratory syndrome coro | ···················· | ···················· | ···················· | ···················· | ···················· | ···················· | |
| 20038 | MT973317.1 | \|Severe acute respiratory syndrome coro | ···················· | ···················· | ···················· | ···················· | ···················· | ···················· | |
| 20040 | MT973319.1 | \|Severe acute respiratory syndrome coro | ···················· | ···················· | ···················· | ···················· | ···················· | ···················· | |
| 20041 | MT973320.1 | \|Severe acute respiratory syndrome coro | ···················· | ···················· | ···················· | ···················· | ···················· | ···················· | |
| 20042 | MT973321.1 | \|Severe acute respiratory syndrome coro | ···················· | ···················· | ···················· | ···················· | ···················· | ···················· | |
| 20044 | MT973323.1 | \|Severe acute respiratory syndrome coro | ···················· | ···················· | ···················· | ···················· | ···················· | ···················· | |
| 20045 | MT973324.1 | \|Severe acute respiratory syndrome coro | ···················· | ···················· | ···················· | ···················· | ···················· | ···················· | |
| 20046 | MT973326.1 | \|Severe acute respiratory syndrome coro | ···················· | ···················· | ···················· | ···················· | ···················· | ···················· | |
| 20047 | MT973327.1 | \|Severe acute respiratory syndrome coro | ···················· | ···················· | ···················· | ···················· | ···················· | ···················· | |
| 20048 | MT973328.1 | \|Severe acute respiratory syndrome coro | ···················· | ···················· | ···················· | ···················· | ···················· | ···················· | |
| 20049 | MT973329.1 | \|Severe acute respiratory syndrome coro | ···················· | ···················· | ···················· | ···················· | ···················· | ···················· | |
| 20050 | MT973330.1 | \|Severe acute respiratory syndrome coro | ···················· | ···················· | ···················· | ···················· | ···················· | ···················· | |
| 20051 | MT973331.1 | \|Severe acute respiratory syndrome coro | ···················· | ···················· | ···················· | ···················· | ···················· | ···················· | |
| 20052 | MT973332.1 | \|Severe acute respiratory syndrome coro | ···················· | ···················· | ···················· | ···················· | ···················· | ···················· | |
| 20053 | MT973333.1 | \|Severe acute respiratory syndrome coro | ···················· | ···················· | ···················· | ···················· | ···················· | ···················· | |
| 20054 | MT973334.1 | \|Severe acute respiratory syndrome coro | ···················· | ···················· | ···················· | ···················· | ···················· | ···················· | |
| 20055 | MT973335.1 | \|Severe acute respiratory syndrome coro | ···················· | ···················· | ···················· | ···················· | ···················· | ···················· | |
| 20056 | MT973336.1 | \|Severe acute respiratory syndrome coro | ···················· | ···················· | ···················· | ···················· | ···················· | ···················· | |
| 20057 | MT973337.1 | \|Severe acute respiratory syndrome coro | ···················· | ···················· | ···················· | ···················· | ···················· | ···················· | |
| 20058 | MT973338.1 | \|Severe acute respiratory syndrome coro | ···················· | ···················· | ···················· | ···················· | ···················· | ···················· | |
| 20059 | MT973339.1 | \|Severe acute respiratory syndrome coro | ···················· | ···················· | ···················· | ···················· | ···················· | ···············A···· | |
| 20060 | MT973341.1 | \|Severe acute respiratory syndrome coro | ···················· | ···················· | ···················· | ···················· | ···················· | ···················· | |
| 20061 | MT973342.1 | \|Severe acute respiratory syndrome coro | ···················· | ···················· | ···················· | ···················· | ···················· | ···················· | |
| 20063 | MT973347.1 | \|Severe acute respiratory syndrome coro | ···················· | ···················· | ···················· | ···················· | ···················· | ···················· | |
| 20064 | MT973348.1 | \|Severe acute respiratory syndrome coro | ···················· | ···················· | ···················· | ···················· | ···················· | ···················· | |
| 20068 | MT973366.1 | \|Severe acute respiratory syndrome coro | ···················· | ···················· | ···················· | ···················· | ···················· | ···················· | |
| 20069 | MT973371.1 | \|Severe acute respiratory syndrome coro | ···················· | ···················· | ···················· | ···················· | ···················· | ···················· | |
| 20070 | MT973373.1 | \|Severe acute respiratory syndrome coro | ···················· | ···················· | ···················· | ···················· | ···················· | ···················· | |
| 20073 | MT973380.1 | \|Severe acute respiratory syndrome coro | ···················· | ···················· | ···················· | ···················· | ···················· | ···················· | |
| 20080 | MT973409.1 | \|Severe acute respiratory syndrome coro | ···················· | ···················· | ···················· | ···················· | ···················· | ···················· | |
| 20084 | MT973426.1 | \|Severe acute respiratory syndrome coro | ···················· | ···················· | ···················· | ···················· | ···················· | ···················· | |
| 20085 | MT973427.1 | \|Severe acute respiratory syndrome coro | ···················· | ···················· | ···················· | ···················· | ···················· | ···················· | |
| 20086 | MT973431.1 | \|Severe acute respiratory syndrome coro | ···················· | ···················· | ···················· | ···················· | ···················· | ···················· | |
| 20089 | MT973438.1 | \|Severe acute respiratory syndrome coro | ···················· | ···················· | ···················· | ···················· | ···················· | ···················· | |
| 20090 | MT973448.1 | \|Severe acute respiratory syndrome coro | ···················· | ···················· | ···················· | ···················· | ···················· | ···················· | |
| 20091 | MT973454.1 | \|Severe acute respiratory syndrome coro | ···················· | ···················· | ···················· | ···················· | ···················· | ···················· | |
| 20093 | MT953868.1 | \|Severe acute respiratory syndrome coro | ···················· | ·A·················· | ···················· | ···················· | ···················· | ···A················ | |
| 20094 | MT953874.1 | \|Severe acute respiratory syndrome coro | ···················· | ·A·················· | ···················· | ···················· | ···················· | ···A················ | |

20095 MT953875.1 |Severe acute respiratory syndrome coro ···················· ···················· ···················· ···················· ···················· ····················
20096 MT953876.1 |Severe acute respiratory syndrome coro ···················· ·A·················· ···················· ···················· ···················· ···A················
20097 MT953877.1 |Severe acute respiratory syndrome coro ···················· ·A·················· ···················· ···················· ···················· ···A················
20098 MT953878.1 |Severe acute respiratory syndrome coro ···················· ···················· ···················· ···················· ···················· ····················
20099 MT953879.1 |Severe acute respiratory syndrome coro ···················· ···················· ···················· ···················· ···················· ····················
20100 MT953880.1 |Severe acute respiratory syndrome coro ···················· ···················· ···················· ···················· ···················· ····················
20101 MT953887.1 |Severe acute respiratory syndrome coro ···················· ···················· ···················· ···················· ···················· ····················
20102 MT953890.1 |Severe acute respiratory syndrome coro ···················· ···················· ···················· ···················· ···················· ····················
20103 MT953891.1 |Severe acute respiratory syndrome coro ···················· ·A·················· ···················· ···················· ···················· ···A················
20104 MT953893.1 |Severe acute respiratory syndrome coro ···················· ·A·················· ···················· ···················· ···················· ····················
20105 MT953894.1 |Severe acute respiratory syndrome coro ···················· ·A·················· ···················· ···················· ···················· ···A················
20106 MT953895.1 |Severe acute respiratory syndrome coro ···················· ·A·················· ···················· ···················· ···················· ···A················
20107 MT953925.1 |Severe acute respiratory syndrome coro ···················· ·A·················· ···················· ···················· ···················· ···A················
20108 MT953978.1 |Severe acute respiratory syndrome coro ···················· ·A·················· ···················· ···················· ···················· ···A················
20109 MT953980.1 |Severe acute respiratory syndrome coro ···················· ·A·················· ···················· ···················· ···················· ···A················
20110 MT953981.1 |Severe acute respiratory syndrome coro ···················· ·A·················· ···················· ···················· ···················· ···A················
20111 MT953984.1 |Severe acute respiratory syndrome coro ···················· ·A·················· ···················· ···················· ···················· ···A················
20112 MT953991.1 |Severe acute respiratory syndrome coro ···················· ·A·················· ···················· ···················· ···················· ····················
20113 MT953996.1 |Severe acute respiratory syndrome coro ···················· ·A·················· ···················· ···················· ···················· ···A················
20114 MT954060.1 |Severe acute respiratory syndrome coro ···················· ·A·················· ···················· ···················· ···················· ···A················
20115 MT954065.1 |Severe acute respiratory syndrome coro ···················· ·A·················· ···················· ···················· ···················· ···A················
20116 MT954067.1 |Severe acute respiratory syndrome coro ···················· ···················· ···················· ···················· ···················· ····················
20117 MT954069.1 |Severe acute respiratory syndrome coro ········T··········· ···················· ···················· ···················· ···················· ····················
20118 MT954071.1 |Severe acute respiratory syndrome coro ···················· ·A·················· ···················· ···················· ···················· ···A················
20119 MT954072.1 |Severe acute respiratory syndrome coro ···················· ·A·················· ···················· ···················· ···················· ····················
20120 MT954103.1 |Severe acute respiratory syndrome coro ···················· ·A·················· ···················· ···················· ···················· ···A················
20121 MT954414.1 |Severe acute respiratory syndrome coro ···················· ·A·················· ···················· ···················· ···················· ···A················
20122 MT954416.1 |Severe acute respiratory syndrome coro ···················· ····A··············· ···················· ···················· ···················· ····················
20123 MT954593.1 |Severe acute respiratory syndrome coro ···················· ····A··············· ···················· ···················· ···················· ····················
20124 MT954959.1 |Severe acute respiratory syndrome coro ···················· ···················· ···················· ···················· ···················· ····················
20125 MT954971.1 |Severe acute respiratory syndrome coro ···················· ···················· ···················· ···················· ···················· ····················
20126 MT955114.1 |Severe acute respiratory syndrome coro ···················· ···················· ···················· ···················· ···················· ····················
20127 MT955161.1 |Severe acute respiratory syndrome coro ···················· ···················· ···················· ···················· ···················· ····················
20128 MT955168.1 |Severe acute respiratory syndrome coro ···················· ···················· ···················· ···················· ···················· ····················
20129 MT955169.1 |Severe acute respiratory syndrome coro ···················· ···················· ···················· ···················· ···················· ····················
20130 MT955170.1 |Severe acute respiratory syndrome coro ···················· ···················· ···················· ···················· ···················· ····················
20131 MT955171.1 |Severe acute respiratory syndrome coro ···················· ···················· ···················· ···················· ···················· ····················
20132 MT955172.1 |Severe acute respiratory syndrome coro ···················· ···················· ···················· ···················· ···················· ····················
20133 MT955173.1 |Severe acute respiratory syndrome coro ···················· ·A·················· ···················· ···················· ···················· ···A················
20134 MT955174.1 |Severe acute respiratory syndrome coro ···················· ···················· ···················· ···················· ···················· ····················
20135 MT955326.1 |Severe acute respiratory syndrome coro ············T······· ···················· ···················· ···················· ···················· ····················
20136 MT955347.1 |Severe acute respiratory syndrome coro ···················· ·A·················· ···················· ···················· ···················· ···A················
20137 MT955354.1 |Severe acute respiratory syndrome coro ···················· ·A·················· ···················· ···················· ···················· ···A················
20138 MT955360.1 |Severe acute respiratory syndrome coro ···················· ···················· ···················· ···················· ···················· ····················
20139 MT956642.1 |Severe acute respiratory syndrome coro ···················· ···················· ···················· ···················· ···················· ····················
20140 MT956684.1 |Severe acute respiratory syndrome coro ···················· ····A··············· ···················· ···················· ···················· ····················
20141 MT956686.1 |Severe acute respiratory syndrome coro ···················· ···················· ···················· ···················· ···················· ····················
20142 MT956687.1 |Severe acute respiratory syndrome coro ···················· ···················· ···················· ···················· ···················· ····················
20143 MT956688.1 |Severe acute respiratory syndrome coro ···················· ···················· ···················· ···················· ···················· ····················
20144 MT956693.1 |Severe acute respiratory syndrome coro ···················· ···················· ···················· ···················· ···················· ····················
20145 MT956695.1 |Severe acute respiratory syndrome coro ···················· ···················· ···················· ···················· ···················· ····················
20147 MT956699.1 |Severe acute respiratory syndrome coro ···················· ···················· ···················· ···················· ···················· ····················
20148 MT956700.1 |Severe acute respiratory syndrome coro ···················· ···················· ···················· ···················· ···················· ····················
20150 MT956702.1 |Severe acute respiratory syndrome coro ···················· ···················· ···················· ···················· ···················· ····················
20151 MT956703.1 |Severe acute respiratory syndrome coro ···················· ···················· ···················· ···················· ···················· ····················
20152 MT956704.1 |Severe acute respiratory syndrome coro ···················· ···················· ···················· ···················· ···················· ····················
20153 MT956706.1 |Severe acute respiratory syndrome coro ···················· ···················· ···················· ···················· ···················· ···············A····
20154 MT956709.1 |Severe acute respiratory syndrome coro ···················· ···················· ···················· ···················· ···················· ····················
20155 MT956710.1 |Severe acute respiratory syndrome coro ···················· ···················· ···················· ···················· ···················· ····················
20157 MT956713.1 |Severe acute respiratory syndrome coro ···················· ···················· ···················· ···················· ···················· ····················
20158 MT956714.1 |Severe acute respiratory syndrome coro ···················· ···················· ···················· ···················· ···················· ····················
20159 MT956715.1 |Severe acute respiratory syndrome coro ···················· ···················· ···················· ···················· ···················· ····················
20160 MT956716.1 |Severe acute respiratory syndrome coro ···················· ···················· ···················· ···················· ···················· ····················
20161 MT956717.1 |Severe acute respiratory syndrome coro ···················· ···················· ···················· ···················· ···················· ····················
20162 MT956718.1 |Severe acute respiratory syndrome coro ···················· ···················· ···················· ···················· ···················· ····················
20163 MT956719.1 |Severe acute respiratory syndrome coro ···················· ···················· ···················· ···················· ···················· ····················
20164 MT956720.1 |Severe acute respiratory syndrome coro ···················· ···················· ···················· ···················· ···················· ····················
20165 MT956721.1 |Severe acute respiratory syndrome coro ···················· ···················· ···················· ···················· ···················· ····················
20166 MT956722.1 |Severe acute respiratory syndrome coro ···················· ···················· ···················· ···················· ···················· ····················
20167 MT956723.1 |Severe acute respiratory syndrome coro ···················· ···················· ···················· ···················· ···················· ····················
20168 MT956724.1 |Severe acute respiratory syndrome coro ···················· ···················· ···················· ···················· ···················· ····················
20169 MT956726.1 |Severe acute respiratory syndrome coro ···················· ·A·················· ···················· ···················· ···················· ····················
20170 MT956727.1 |Severe acute respiratory syndrome coro ···················· ·A·················· ···················· ···················· ···················· ····················
20171 MT956729.1 |Severe acute respiratory syndrome coro ···················· ···················· ···················· ···················· ···················· ····················
20172 MT956730.1 |Severe acute respiratory syndrome coro ···················· ···················· ···················· ···················· ···················· ····················
20173 MT956731.1 |Severe acute respiratory syndrome coro ···················· ···················· ···················· ···················· ···················· ····················
20174 MT956732.1 |Severe acute respiratory syndrome coro ···················· ···················· ···················· ···················· ···················· ····················
20175 MT956735.1 |Severe acute respiratory syndrome coro ···················· ···················· ···················· ···················· ···················· ····················

20177 MT956737.1 |Severe acute respiratory syndrome coro ···················· ···················· ···················· ···················· ···················· ····················
20178 MT956738.1 |Severe acute respiratory syndrome coro ···················· ···················· ···················· ···················· ···················· ····················
20179 MT956740.1 |Severe acute respiratory syndrome coro ···················· ···················· ···················· ···················· ···················· ····················
20180 MT956741.1 |Severe acute respiratory syndrome coro ···················· ···················· ···················· ···················· ···················· ····················
20181 MT956742.1 |Severe acute respiratory syndrome coro ···················· ···················· ···················· ···················· ···················· ····················
20182 MT956743.1 |Severe acute respiratory syndrome coro ···················· ···················· ···················· ···················· ···················· ····················
20183 MT956744.1 |Severe acute respiratory syndrome coro ···················· ···················· ···················· ···················· ···················· ····················
20184 MT956745.1 |Severe acute respiratory syndrome coro ···················· ···················· ···················· ···················· ···················· ····················
20185 MT956746.1 |Severe acute respiratory syndrome coro ···················· ···················· ···················· ···················· ···················· ····················
20186 MT956747.1 |Severe acute respiratory syndrome coro ···················· ···················· ···················· ···················· ···················· ····················
20187 MT956748.1 |Severe acute respiratory syndrome coro ···················· ···················· ···················· ···················· ···················· ····················
20188 MT956749.1 |Severe acute respiratory syndrome coro ···················· ···················· ···················· ···················· ···················· ····················
20189 MT956750.1 |Severe acute respiratory syndrome coro ···················· ···················· ···················· ···················· ···················· ····················
20190 MT956751.1 |Severe acute respiratory syndrome coro ···················· ···················· ···················· ···················· ···················· ····················
20191 MT956752.1 |Severe acute respiratory syndrome coro ···················· ···················· ···················· ···················· ···················· ····················
20192 MT956753.1 |Severe acute respiratory syndrome coro ···················· ···················· ···················· ···················· ···················· ····················
20193 MT956754.1 |Severe acute respiratory syndrome coro ···················· ···················· ···················· ···················· ···················· ····················
20194 MT956755.1 |Severe acute respiratory syndrome coro ···················· ···················· ···················· ···················· ···················· ····················
20195 MT956756.1 |Severe acute respiratory syndrome coro ···················· ···················· ···················· ···················· ···················· ····················
20196 MT956757.1 |Severe acute respiratory syndrome coro ···················· ···················· ···················· ···················· ···················· ····················
20198 MT956759.1 |Severe acute respiratory syndrome coro ···················· ···················· ···················· ···················· ···················· ····················
20200 MT956761.1 |Severe acute respiratory syndrome coro ···················· ···················· ···················· ···················· ···················· ····················
20201 MT956762.1 |Severe acute respiratory syndrome coro ···················· ···················· ···················· ···················· ···················· ····················
20202 MT956767.1 |Severe acute respiratory syndrome coro ···················· ···················· ···················· ···················· ···················· ····················
20203 MT956768.1 |Severe acute respiratory syndrome coro ···················· ···················· ···················· ···················· ···················· ····················
20204 MT956769.1 |Severe acute respiratory syndrome coro ···················· ···················· ···················· ···················· ···················· ····················
20205 MT956770.1 |Severe acute respiratory syndrome coro ···················· ···················· ···················· ···················· ···················· ····················
20207 MT956773.1 |Severe acute respiratory syndrome coro ···················· ···················· ···················· ···················· ···················· ····················
20209 MT956776.1 |Severe acute respiratory syndrome coro ···················· ···················· ···················· ···················· ···················· ····················
20210 MT956777.1 |Severe acute respiratory syndrome coro ···················· ···················· ···················· ···················· ···················· ····················
20212 MT956784.1 |Severe acute respiratory syndrome coro ···················· ···················· ···················· ···················· ···················· ····················
20213 MT956785.1 |Severe acute respiratory syndrome coro ···················· ···················· ···················· ···················· ···················· ····················
20214 MT956786.1 |Severe acute respiratory syndrome coro ···················· ···················· ···················· ···················· ···················· ····················
20215 MT956787.1 |Severe acute respiratory syndrome coro ···················· ···················· ···················· ···················· ···················· ····················
20216 MT956788.1 |Severe acute respiratory syndrome coro ···················· ···················· ···················· ···················· ···················· ····················
20217 MT956789.1 |Severe acute respiratory syndrome coro ···················· ···················· ···················· ···················· ···················· ····················
20218 MT956790.1 |Severe acute respiratory syndrome coro ···················· ···················· ···················· ···················· ···················· ····················
20219 MT956791.1 |Severe acute respiratory syndrome coro ···················· ···················· ···················· ···················· ···················· ····················
20220 MT956793.1 |Severe acute respiratory syndrome coro ···················· ···················· ···················· ···················· ···················· ····················
20221 MT956794.1 |Severe acute respiratory syndrome coro ···················· ···················· ···················· ···················· ···················· ····················
20222 MT956795.1 |Severe acute respiratory syndrome coro ···················· ···················· ···················· ···················· ···················· ····················
20223 MT956796.1 |Severe acute respiratory syndrome coro ···················· ···················· ···················· ···················· ···················· ····················
20224 MT956797.1 |Severe acute respiratory syndrome coro ···················· ···················· ···················· ···················· ···················· ····················
20225 MT956798.1 |Severe acute respiratory syndrome coro ···················· ···················· ···················· ···················· ···················· ····················
20226 MT956799.1 |Severe acute respiratory syndrome coro ···················· ·A·················· ···················· ···················· ···················· ····················
20227 MT956801.1 |Severe acute respiratory syndrome coro ···················· ·A·················· ···················· ···················· ···················· ····················
20228 MT956802.1 |Severe acute respiratory syndrome coro ···················· ···················· ···················· ···················· ···················· ····················
20229 MT956803.1 |Severe acute respiratory syndrome coro ···················· ···················· ···················· ···················· ···················· ····················
20230 MT956804.1 |Severe acute respiratory syndrome coro ···················· ···················· ···················· ···················· ···················· ····················
20231 MT956805.1 |Severe acute respiratory syndrome coro ···················· ···················· ···················· ···················· ···················· ····················
20232 MT956806.1 |Severe acute respiratory syndrome coro ···················· ···················· ···················· ···················· ···················· ····················
20233 MT956813.1 |Severe acute respiratory syndrome coro ···················· ···················· ···················· ···················· ···················· ····················
20235 MT956911.1 |Severe acute respiratory syndrome coro ···················· ···················· ···················· ···················· ···················· ····················
20236 MT956912.1 |Severe acute respiratory syndrome coro ···················· ···················· ···················· ···················· ···················· ····················
20237 MT956913.1 |Severe acute respiratory syndrome coro ···················· ···················· ···················· ···················· ···················· ····················
20238 MT956914.1 |Severe acute respiratory syndrome coro ···················· ···················· ···················· ···················· ···················· ····················
20239 MT956915.1 |Severe acute respiratory syndrome coro ···················· ···················· ···················· ···················· ···················· ····················
20240 MT956916.1 |Severe acute respiratory syndrome coro ···················· ···················· ···················· ···················· ···················· ····················
20241 MT956917.1 |Severe acute respiratory syndrome coro ···················· ···················· ···················· ···················· ···················· ····················
20242 MT956918.1 |Severe acute respiratory syndrome coro ···················· ···················· ···················· ···················· ···················· ····················
20243 MT958205.1 |Severe acute respiratory syndrome coro ···················· ···················· ···················· ···················· ···················· ····················
20244 MT958206.1 |Severe acute respiratory syndrome coro ···················· ···················· ···················· ···················· ···················· ····················
20245 MT958207.1 |Severe acute respiratory syndrome coro ···················· ·A·················· ···················· ···················· ···················· ····················
20246 MT958208.1 |Severe acute respiratory syndrome coro ···················· ·A·················· ···················· ···················· ···················· ····················
20247 MT958209.1 |Severe acute respiratory syndrome coro ···················· ·A·················· ···················· ···················· ···················· ····················
20248 MT958210.1 |Severe acute respiratory syndrome coro ···················· ·A·················· ···················· ···················· ···················· ····················
20249 MT958211.1 |Severe acute respiratory syndrome coro ···················· ·A·················· ···················· ···················· ···················· ····················
20250 MT958212.1 |Severe acute respiratory syndrome coro ···················· ·A·················· ···················· ···················· ···················· ····················
20251 MT958213.1 |Severe acute respiratory syndrome coro ···················· ·A·················· ···················· ···················· ···················· ····················
20252 MT958214.1 |Severe acute respiratory syndrome coro ···················· ·A·················· ···················· ···················· ···················· ····················
20253 MT958215.1 |Severe acute respiratory syndrome coro ···················· ·A·················· ···················· ···················· ···················· ····················
20254 MT958216.1 |Severe acute respiratory syndrome coro ···················· ·A·················· ···················· ···················· ···················· ····················
20255 MT958217.1 |Severe acute respiratory syndrome coro ···················· ·A·················· ···················· ···················· ···················· ····················
20256 MT958218.1 |Severe acute respiratory syndrome coro ···················· ·A·················· ···················· ···················· ···················· ····················
20257 MT958219.1 |Severe acute respiratory syndrome coro ···················· ···················· ···················· ···················· ···················· ····················
20258 MT958220.1 |Severe acute respiratory syndrome coro ···················· ·A·················· ···················· ···················· ···················· ····················
20259 MT958221.1 |Severe acute respiratory syndrome coro ···················· ·A·················· ···················· ···················· ···················· ····················
20260 MT958222.1 |Severe acute respiratory syndrome coro ···················· ·A·················· ···················· ···················· ···················· ····················

| | | | | | | | | | |
|---|---|---|---|---|---|---|---|---|---|
| 20261 | MT958223.1 | \|Severe acute respiratory syndrome coro | ···················· | ·A·················· | ···················· | ···················· | ···················· | ···················· | |
| 20262 | MT958224.1 | \|Severe acute respiratory syndrome coro | ···················· | ·A·················· | ···················· | ···················· | ···················· | ···················· | |
| 20263 | MT958225.1 | \|Severe acute respiratory syndrome coro | ···················· | ·A·················· | ···················· | ···················· | ···················· | ···················· | |
| 20264 | MT958226.1 | \|Severe acute respiratory syndrome coro | ···················· | ·A·················· | ···················· | ···················· | ···················· | ···················· | |
| 20265 | MT958227.1 | \|Severe acute respiratory syndrome coro | ···················· | ·A·················· | ···················· | ···················· | ···················· | ···················· | |
| 20266 | MT958228.1 | \|Severe acute respiratory syndrome coro | ···················· | ·A·················· | ···················· | ···················· | ···················· | ···················· | |
| 20267 | MT958229.1 | \|Severe acute respiratory syndrome coro | ···················· | ·A·················· | ···················· | ···················· | ···················· | ···················· | |
| 20268 | MT958230.1 | \|Severe acute respiratory syndrome coro | ···················· | ·A·················· | ···················· | ···················· | ···················· | ···················· | |
| 20269 | MT958231.1 | \|Severe acute respiratory syndrome coro | ···················· | ·A·················· | ···················· | ···················· | ···················· | ···················· | |
| 20270 | MT958232.1 | \|Severe acute respiratory syndrome coro | ···················· | ·A·················· | ···················· | ···················· | ···················· | ···················· | |
| 20271 | MT958233.1 | \|Severe acute respiratory syndrome coro | ···················· | ·A·················· | ···················· | ···················· | ···················· | ···················· | |
| 20272 | MT958234.1 | \|Severe acute respiratory syndrome coro | ···················· | ·A·················· | ···················· | ···················· | ···················· | ···················· | |
| 20273 | MT958235.1 | \|Severe acute respiratory syndrome coro | ···················· | ·A·················· | ···················· | ···················· | ···················· | ···················· | |
| 20274 | MT958236.1 | \|Severe acute respiratory syndrome coro | ···················· | ·A·················· | ···················· | ···················· | ···················· | ···················· | |
| 20275 | MT958237.1 | \|Severe acute respiratory syndrome coro | ···················· | ·A·················· | ···················· | ···················· | ···················· | ···················· | |
| 20276 | MT958238.1 | \|Severe acute respiratory syndrome coro | ···················· | ·A·················· | ···················· | ···················· | ···················· | ···················· | |
| 20277 | MT958239.1 | \|Severe acute respiratory syndrome coro | ···················· | ·A·················· | ···················· | ···················· | ···················· | ···················· | |
| 20278 | MT958240.1 | \|Severe acute respiratory syndrome coro | ···················· | ·A·················· | ···················· | ···················· | ·NNN················ | ···················· | 1 |
| 20279 | MT958241.1 | \|Severe acute respiratory syndrome coro | ···················· | ·A·················· | ···················· | ···················· | ···················· | ···················· | |
| 20280 | MT958242.1 | \|Severe acute respiratory syndrome coro | ···················· | ·A·················· | ···················· | ···················· | ···················· | ···················· | |
| 20281 | MT958243.1 | \|Severe acute respiratory syndrome coro | ···················· | ·A·················· | ···················· | ···················· | ···················· | ···················· | |
| 20282 | MT958244.1 | \|Severe acute respiratory syndrome coro | ···················· | ·A·················· | ···················· | ···················· | ···················· | ···················· | |
| 20283 | MT958245.1 | \|Severe acute respiratory syndrome coro | ···················· | ·A·················· | ···················· | ···················· | ···················· | ···················· | |
| 20284 | MT958246.1 | \|Severe acute respiratory syndrome coro | ···················· | ·A·················· | ···················· | ···················· | ···················· | ···················· | |
| 20285 | MT958247.1 | \|Severe acute respiratory syndrome coro | ···················· | ·A·················· | ···················· | ···················· | ···················· | ···················· | |
| 20286 | MT958248.1 | \|Severe acute respiratory syndrome coro | ···················· | ·A·················· | ···················· | ···················· | ···················· | ···················· | |
| 20287 | MT958249.1 | \|Severe acute respiratory syndrome coro | ···················· | ·A·················· | ···················· | ···················· | ···················· | ···················· | |
| 20288 | MT958250.1 | \|Severe acute respiratory syndrome coro | ···················· | ·A·················· | ···················· | ···················· | ···················· | ···················· | |
| 20289 | MT958251.1 | \|Severe acute respiratory syndrome coro | ···················· | ·A·················· | ···················· | ···················· | ···················· | ···················· | |
| 20290 | MT958252.1 | \|Severe acute respiratory syndrome coro | ···················· | ·A·················· | ···················· | ···················· | ···················· | ···················· | |
| 20291 | MT958253.1 | \|Severe acute respiratory syndrome coro | ···················· | ·A·················· | ···················· | ···················· | ···················· | ···················· | |
| 20292 | MT958254.1 | \|Severe acute respiratory syndrome coro | ···················· | ···················· | ···················· | ···················· | ···················· | ···················· | |
| 20293 | MT958255.1 | \|Severe acute respiratory syndrome coro | ···················· | ···················· | ···················· | ···················· | ···················· | ···················· | |
| 20294 | MT958256.1 | \|Severe acute respiratory syndrome coro | ···················· | ···················· | ···················· | ···················· | ···················· | ···················· | |
| 20296 | MT958258.1 | \|Severe acute respiratory syndrome coro | ···················· | ·A·················· | ···················· | ···················· | ···················· | ···················· | |
| 20297 | MT958259.1 | \|Severe acute respiratory syndrome coro | ···················· | ···················· | ···················· | ···················· | NNN················· | ···················· | 1 |
| 20298 | MT958260.1 | \|Severe acute respiratory syndrome coro | ···················· | ·A·················· | ···················· | ···················· | ···················· | ···················· | |
| 20299 | MT958261.1 | \|Severe acute respiratory syndrome coro | ···················· | ·A·················· | ···················· | ···················· | ·NN················· | ···················· | |
| 20300 | MT958262.1 | \|Severe acute respiratory syndrome coro | ···················· | ···················· | ···················· | ···················· | ···················· | ···················· | |
| 20301 | MT958263.1 | \|Severe acute respiratory syndrome coro | ···················· | ···················· | ···················· | ···················· | ···················· | ···················· | |
| 20302 | MT950187.1 | \|Severe acute respiratory syndrome coro | ···················· | ·A·················· | ···················· | ···················· | ···················· | ···A················ | |
| 20303 | MT950188.1 | \|Severe acute respiratory syndrome coro | ···················· | ·A·················· | ···················· | ···················· | ···················· | ···················· | |
| 20304 | MT950189.1 | \|Severe acute respiratory syndrome coro | ···················· | ···················· | ···················· | ···················· | ···················· | ···················· | |
| 20305 | MT950190.1 | \|Severe acute respiratory syndrome coro | ···················· | ·A·················· | ···················· | ···················· | ···················· | ···A················ | |
| 20306 | MT950191.1 | \|Severe acute respiratory syndrome coro | ···················· | ·A·················· | ···················· | ···················· | ···················· | ···A················ | |
| 20307 | MT950192.1 | \|Severe acute respiratory syndrome coro | ···················· | ·A·················· | ···················· | ···················· | ···················· | ···A················ | |
| 20308 | MT950193.1 | \|Severe acute respiratory syndrome coro | ···················· | ·A·················· | ···················· | ···················· | ···················· | ···A················ | |
| 20309 | MT950195.1 | \|Severe acute respiratory syndrome coro | ···················· | ·A·················· | ···················· | ···················· | ···················· | ···A················ | |
| 20310 | MT950230.1 | \|Severe acute respiratory syndrome coro | ···················· | ·A·················· | ···················· | ···················· | ···················· | ···A················ | |
| 20311 | MT950231.1 | \|Severe acute respiratory syndrome coro | ···················· | ·A·················· | ···················· | ···················· | ···················· | ···A················ | |
| 20312 | MT950348.1 | \|Severe acute respiratory syndrome coro | ···················· | ···················· | ···················· | ···················· | ···················· | ···················· | |
| 20313 | MT950349.1 | \|Severe acute respiratory syndrome coro | ···················· | ·A·················· | ···················· | ···················· | ···················· | ···A················ | |
| 20314 | MT950532.1 | \|Severe acute respiratory syndrome coro | ···················· | ···················· | ···················· | ···················· | ···················· | ···················· | |
| 20315 | MT950684.1 | \|Severe acute respiratory syndrome coro | ···················· | ···················· | ···················· | ···················· | ···················· | ···················· | |
| 20316 | MT950725.1 | \|Severe acute respiratory syndrome coro | ···················· | ·A·················· | ···················· | ···················· | ···················· | ···A················ | |
| 20317 | MT950761.1 | \|Severe acute respiratory syndrome coro | ···················· | ·A·················· | ···················· | ···················· | ···················· | ···A················ | |
| 20318 | MT951157.1 | \|Severe acute respiratory syndrome coro | ···················· | ·A·················· | ···················· | ···················· | ···················· | ···A················ | |
| 20319 | MT951170.1 | \|Severe acute respiratory syndrome coro | ···················· | ···A················ | ···················· | ···················· | ···················· | ···················· | |
| 20320 | MT951177.1 | \|Severe acute respiratory syndrome coro | ···················· | ···A················ | ···················· | ···················· | ···················· | ···················· | |
| 20321 | MT951181.1 | \|Severe acute respiratory syndrome coro | ···················· | ·A·················· | ···················· | ···················· | ···················· | ···A················ | |
| 20322 | MT951208.1 | \|Severe acute respiratory syndrome coro | ···················· | ···················· | ···················· | ···················· | ···················· | ···················· | |
| 20323 | MT951392.1 | \|Severe acute respiratory syndrome coro | ···················· | ·A·················· | ···················· | ···················· | ···················· | ···A················ | |
| 20324 | MT951393.1 | \|Severe acute respiratory syndrome coro | ···················· | ·A·················· | ···················· | ···················· | ···················· | ···A················ | |
| 20325 | MT951924.1 | \|Severe acute respiratory syndrome coro | ···················· | ···················· | ···················· | ···················· | ···················· | ········A··········· | |
| 20326 | MT951925.1 | \|Severe acute respiratory syndrome coro | ···················· | ···················· | ···················· | ···················· | ···················· | ········A··········· | |
| 20327 | MT951926.1 | \|Severe acute respiratory syndrome coro | ···················· | ···················· | ···················· | ···················· | ···················· | ···················· | |
| 20328 | MT951927.1 | \|Severe acute respiratory syndrome coro | ···················· | ···················· | ···················· | ···················· | ···················· | ···················· | |
| 20329 | MT951929.1 | \|Severe acute respiratory syndrome coro | ···················· | ···················· | ···················· | ···················· | ···················· | ···················· | |
| 20330 | MT951930.1 | \|Severe acute respiratory syndrome coro | ···················· | ···················· | ···················· | ···················· | ···················· | ···················· | |
| 20331 | MT951931.1 | \|Severe acute respiratory syndrome coro | ···················· | ···················· | ···················· | ···················· | ···················· | ···················· | |
| 20332 | MT951932.1 | \|Severe acute respiratory syndrome coro | ···················· | ···················· | ···················· | ···················· | ···················· | ···················· | |
| 20333 | MT951934.1 | \|Severe acute respiratory syndrome coro | ···················· | ···················· | ···················· | ···················· | ···················· | ···················· | |
| 20334 | MT951935.1 | \|Severe acute respiratory syndrome coro | ···················· | ···················· | ···················· | ···················· | ···················· | ···················· | |
| 20335 | MT951936.1 | \|Severe acute respiratory syndrome coro | ···················· | ···················· | ···················· | ···················· | ···················· | ···················· | |
| 20336 | MT951937.1 | \|Severe acute respiratory syndrome coro | ···················· | ···················· | ···················· | ···················· | ···················· | ···················· | |
| 20337 | MT951938.1 | \|Severe acute respiratory syndrome coro | ···················· | ···················· | ···················· | ···················· | ···················· | ···················· | |
| 20338 | MT951939.1 | \|Severe acute respiratory syndrome coro | ···················· | ···················· | ···················· | ···················· | ···················· | ···················· | |
| 20339 | MT951940.1 | \|Severe acute respiratory syndrome coro | ···················· | ···················· | ···················· | ···················· | ···················· | ···················· | |

| | | | | | | | | |
|---|---|---|---|---|---|---|---|---|
| 20340 | MT951941.1 | |Severe acute respiratory syndrome coro | ···················· | ···················· | ···················· | ···················· | ···················· | ···················· |
| 20341 | MT951942.1 | |Severe acute respiratory syndrome coro | ···················· | ···················· | ···················· | ···················· | ···················· | ···················· |
| 20342 | MT951944.1 | |Severe acute respiratory syndrome coro | ···················· | ···················· | ···················· | ···················· | ···················· | ···················· |
| 20343 | MT951945.1 | |Severe acute respiratory syndrome coro | ···················· | ···················· | ···················· | ···················· | ···················· | ···················· |
| 20344 | MT951946.1 | |Severe acute respiratory syndrome coro | ···················· | ···················· | ···················· | ···················· | ···················· | ···················· |
| 20345 | MT951947.1 | |Severe acute respiratory syndrome coro | ···················· | ···················· | ···················· | ···················· | ···················· | ···················· |
| 20346 | MT951948.1 | |Severe acute respiratory syndrome coro | ···················· | ···················· | ···················· | ···················· | ···················· | ···················· |
| 20347 | MT951949.1 | |Severe acute respiratory syndrome coro | ···················· | ···················· | ···················· | ···················· | ···················· | ···················· |
| 20348 | MT951950.1 | |Severe acute respiratory syndrome coro | ···················· | ···················· | ···················· | ···················· | ···················· | ···················· |
| 20349 | MT951951.1 | |Severe acute respiratory syndrome coro | ···················· | ···················· | ···················· | ···················· | ···················· | ···················· |
| 20350 | MT951952.1 | |Severe acute respiratory syndrome coro | ···················· | ···················· | ···················· | ···················· | ···················· | ···················· |
| 20351 | MT951953.1 | |Severe acute respiratory syndrome coro | ···················· | ···················· | ···················· | ···················· | ···················· | ···················· |
| 20352 | MT951955.1 | |Severe acute respiratory syndrome coro | ···················· | ···················· | ···················· | ···················· | ···················· | ···················· |
| 20353 | MT951956.1 | |Severe acute respiratory syndrome coro | ···················· | ···················· | ···················· | ···················· | ···················· | ···················· |
| 20354 | MT951957.1 | |Severe acute respiratory syndrome coro | ···················· | ···················· | ···················· | ···················· | ···················· | ···················· |
| 20355 | MT951958.1 | |Severe acute respiratory syndrome coro | ···················· | ···················· | ···················· | ···················· | ···················· | ···················· |
| 20356 | MT951959.1 | |Severe acute respiratory syndrome coro | ···················· | ···················· | ···················· | ···················· | ···················· | ···················· |
| 20357 | MT951960.1 | |Severe acute respiratory syndrome coro | ···················· | ···················· | ···················· | ···················· | ···················· | ···················· |
| 20358 | MT951961.1 | |Severe acute respiratory syndrome coro | ···················· | ···················· | ···················· | ···················· | ···················· | ···················· |
| 20359 | MT951962.1 | |Severe acute respiratory syndrome coro | ···················· | ···················· | ···················· | ···················· | ···················· | ···················· |
| 20360 | MT951963.1 | |Severe acute respiratory syndrome coro | ···················· | ···················· | ···················· | ···················· | ···················· | ···················· |
| 20361 | MT951964.1 | |Severe acute respiratory syndrome coro | ···················· | ···················· | ···················· | ···················· | ···················· | ···················· |
| 20362 | MT951965.1 | |Severe acute respiratory syndrome coro | ···················· | ···················· | ···················· | ···················· | ···················· | ···················· |
| 20363 | MT951966.1 | |Severe acute respiratory syndrome coro | ···················· | ···················· | ···················· | ···················· | ···················· | ···················· |
| 20364 | MT951967.1 | |Severe acute respiratory syndrome coro | ···················· | ···················· | ···················· | ···················· | ···················· | ···················· |
| 20365 | MT951968.1 | |Severe acute respiratory syndrome coro | ···················· | ···················· | ···················· | ···················· | ···················· | ···················· |
| 20366 | MT951969.1 | |Severe acute respiratory syndrome coro | ···················· | ···················· | ···················· | ···················· | ···················· | ···················· |
| 20367 | MT951970.1 | |Severe acute respiratory syndrome coro | ···················· | ···················· | ···················· | ···················· | ···················· | ···················· |
| 20368 | MT951971.1 | |Severe acute respiratory syndrome coro | ···················· | ···················· | ···················· | ···················· | ···················· | ···················· |
| 20369 | MT951972.1 | |Severe acute respiratory syndrome coro | ···················· | ···················· | ···················· | ···················· | ···················· | ···················· |
| 20370 | MT951973.1 | |Severe acute respiratory syndrome coro | ···················· | ···················· | ···················· | ···················· | ···················· | ···················· |
| 20371 | MT951974.1 | |Severe acute respiratory syndrome coro | ···················· | ···················· | ···················· | ···················· | ···················· | ···················· |
| 20372 | MT951975.1 | |Severe acute respiratory syndrome coro | ···················· | ···················· | ···················· | ···················· | ···················· | ···················· |
| 20373 | MT951976.1 | |Severe acute respiratory syndrome coro | ···················· | ···················· | ···················· | ···················· | ···················· | ···················· |
| 20374 | MT951977.1 | |Severe acute respiratory syndrome coro | ···················· | ···················· | ···················· | ···················· | ···················· | ···················· |
| 20375 | MT951978.1 | |Severe acute respiratory syndrome coro | ···················· | ···················· | ···················· | ···················· | ···················· | ···················· |
| 20376 | MT951980.1 | |Severe acute respiratory syndrome coro | ···················· | ···················· | ···················· | ···················· | ···················· | ···················· |
| 20377 | MT952134.1 | |Severe acute respiratory syndrome coro | ···················· | ···················· | ···················· | ···················· | ···················· | ···················· |
| 20378 | MT952603.1 | |Severe acute respiratory syndrome coro | ···················· | ···················· | ···················· | ···················· | ···················· | ···················· |
| 20379 | MT952604.1 | |Severe acute respiratory syndrome coro | ···················· | ···················· | ···················· | ···················· | ···················· | ···················· |
| 20380 | MT952605.1 | |Severe acute respiratory syndrome coro | ···················· | ···················· | ···················· | ···················· | ···················· | ···················· |
| 20381 | MT952606.1 | |Severe acute respiratory syndrome coro | ···················· | ···················· | ···················· | ···················· | ···················· | ···················· |
| 20382 | MT952607.1 | |Severe acute respiratory syndrome coro | ···················· | ···················· | ···················· | ···················· | ···················· | ···················· |
| 20383 | MT952608.1 | |Severe acute respiratory syndrome coro | ···················· | ···················· | ···················· | ···················· | ···················· | ···················· |
| 20384 | MT952609.1 | |Severe acute respiratory syndrome coro | ···················· | ···················· | ···················· | ···················· | ···················· | ···················· |
| 20385 | MT952610.1 | |Severe acute respiratory syndrome coro | ···················· | ···················· | ···················· | ···················· | ···················· | ···················· |
| 20386 | MT952612.1 | |Severe acute respiratory syndrome coro | ···················· | ···················· | ···················· | ···················· | ···················· | ···················· |
| 20387 | MT952613.1 | |Severe acute respiratory syndrome coro | ···········T········ | ···················· | ···················· | ···················· | ···················· | ···················· |
| 20388 | MT952614.1 | |Severe acute respiratory syndrome coro | ···········T········ | ···················· | ···················· | ···················· | ···················· | ···················· |
| 20389 | MT952615.1 | |Severe acute respiratory syndrome coro | ···········T········ | ···················· | ···················· | ···················· | ···················· | ···················· |
| 20390 | MT952616.1 | |Severe acute respiratory syndrome coro | ···········T········ | ···················· | ···················· | ···················· | ···················· | ···················· |
| 20391 | MT952617.1 | |Severe acute respiratory syndrome coro | ···········T········ | ···················· | ···················· | ···················· | ···················· | ···················· |
| 20392 | MT952618.1 | |Severe acute respiratory syndrome coro | ···········T········ | ···················· | ···················· | ···················· | ···················· | ···················· |
| 20393 | MT952619.1 | |Severe acute respiratory syndrome coro | ···········T········ | ···················· | ···················· | ···················· | ···················· | ···················· |
| 20394 | MT952620.1 | |Severe acute respiratory syndrome coro | ···········T········ | ···················· | ···················· | ···················· | ···················· | ···················· |
| 20395 | MT952621.1 | |Severe acute respiratory syndrome coro | ···········T········ | ···················· | ···················· | ···················· | ···················· | ···················· |
| 20396 | MT952622.1 | |Severe acute respiratory syndrome coro | ···········T········ | ···················· | ···················· | ···················· | ···················· | ···················· |
| 20397 | MT952623.1 | |Severe acute respiratory syndrome coro | ···········T········ | ···················· | ···················· | ···················· | ···················· | ···················· |
| 20398 | MT952624.1 | |Severe acute respiratory syndrome coro | ···········T········ | ···················· | ···················· | ···················· | ···················· | ···················· |
| 20399 | MT952625.1 | |Severe acute respiratory syndrome coro | ···················· | ·A·················· | ···················· | ···················· | ···················· | ···················· |
| 20400 | MT952626.1 | |Severe acute respiratory syndrome coro | ···················· | ·A·················· | ···················· | ···················· | ···················· | ···················· |
| 20401 | MT952627.1 | |Severe acute respiratory syndrome coro | ···················· | ·A·················· | ···················· | ···················· | ···················· | ···················· |
| 20402 | MT952628.1 | |Severe acute respiratory syndrome coro | ···················· | ·A·················· | ···················· | ···················· | ···················· | ···················· |
| 20403 | MT952629.1 | |Severe acute respiratory syndrome coro | ···················· | ·A·················· | ···················· | ···················· | ···················· | ···················· |
| 20404 | MT952630.1 | |Severe acute respiratory syndrome coro | ···················· | ·A·················· | ···················· | ···················· | ···················· | ···················· |
| 20405 | MT952631.1 | |Severe acute respiratory syndrome coro | ···················· | ···················· | ···················· | ···················· | ···················· | ···················· |
| 20406 | MT952632.1 | |Severe acute respiratory syndrome coro | ···················· | ···················· | ···················· | ···················· | ···················· | ···················· |
| 20407 | MT952633.1 | |Severe acute respiratory syndrome coro | ···················· | ···················· | ···················· | ···················· | ···················· | ···················· |
| 20408 | MT952634.1 | |Severe acute respiratory syndrome coro | ···················· | ···················· | ···················· | ···················· | ···················· | ···················· |
| 20409 | MT952635.1 | |Severe acute respiratory syndrome coro | ···················· | ···················· | ···················· | ···················· | ···················· | ···················· |
| 20410 | MT952636.1 | |Severe acute respiratory syndrome coro | ···················· | ···················· | ···················· | ···················· | ···················· | ···················· |
| 20411 | MT952637.1 | |Severe acute respiratory syndrome coro | ···················· | ···················· | ···················· | ···················· | ···················· | ···················· |
| 20412 | MT952638.1 | |Severe acute respiratory syndrome coro | ···················· | ···················· | ···················· | ···················· | ···················· | ···················· |
| 20413 | MT952639.1 | |Severe acute respiratory syndrome coro | ···················· | ···················· | ···················· | ···················· | ···················· | ···················· |
| 20414 | MT952640.1 | |Severe acute respiratory syndrome coro | ···················· | ···················· | ···················· | ···················· | ···················· | ···················· |
| 20415 | MT952642.1 | |Severe acute respiratory syndrome coro | ···················· | ···················· | ···················· | ···················· | ···················· | ···················· |
| 20416 | MT952643.1 | |Severe acute respiratory syndrome coro | ···················· | ···················· | ···················· | ···················· | ···················· | ···················· |
| 20417 | MT952644.1 | |Severe acute respiratory syndrome coro | ···················· | ···················· | ···················· | ···················· | ···················· | ···················· |

```
20418 MT952645.1 |Severe acute respiratory syndrome coro    ···················· ···················· ···················· ···················· ···················· ····················
20419 MT952646.1 |Severe acute respiratory syndrome coro    ···················· ···················· ···················· ···················· ···················· ····················
20420 MT952647.1 |Severe acute respiratory syndrome coro    ···················· ···················· ···················· ···················· ···················· ····················
20421 MT952648.1 |Severe acute respiratory syndrome coro    ···················· ···················· ···················· ···················· ···················· ····················
20422 MT952649.1 |Severe acute respiratory syndrome coro    ···················· ···················· ···················· ···················· ···················· ····················
20423 MT952650.1 |Severe acute respiratory syndrome coro    ···················· ···················· ···················· ···················· ···················· ····················
20424 MT952651.1 |Severe acute respiratory syndrome coro    ···················· ···················· ···················· ···················· ···················· ····················
20425 MT952652.1 |Severe acute respiratory syndrome coro    ···················· ···················· ···················· ···················· ···················· ····················
20426 MT952653.1 |Severe acute respiratory syndrome coro    ···················· ···················· ···················· ···················· ···················· ····················
20427 MT952654.1 |Severe acute respiratory syndrome coro    ···················· ···················· ···················· ···················· ···················· ····················
20428 MT952655.1 |Severe acute respiratory syndrome coro    ···················· ···················· ···················· ···················· ···················· ····················
20429 MT952656.1 |Severe acute respiratory syndrome coro    ···················· ···················· ···················· ···················· ···················· ····················
20430 MT952663.1 |Severe acute respiratory syndrome coro    ···················· ···················· ···················· ···················· ···················· ····················
20431 MT952664.1 |Severe acute respiratory syndrome coro    ···················· ···················· ···················· ···················· ···················· ····················
20432 MT952665.1 |Severe acute respiratory syndrome coro    ···················· ···················· ···················· ···················· ···················· ····················
20433 MT952666.1 |Severe acute respiratory syndrome coro    ···················· ···················· ···················· ···················· ···················· ····················
20434 MT952667.1 |Severe acute respiratory syndrome coro    ···················· ···················· ···················· ···················· ···················· ····················
20435 MT952668.1 |Severe acute respiratory syndrome coro    ···················· ···················· ···················· ···················· ···················· ····················
20436 MT952669.1 |Severe acute respiratory syndrome coro    ···················· ···················· ···················· ···················· ···················· ····················
20437 MT952670.1 |Severe acute respiratory syndrome coro    ···················· ···················· ···················· ···················· ···················· ····················
20438 MT952671.1 |Severe acute respiratory syndrome coro    ···················· ···················· ···················· ···················· ···················· ····················
20439 MT952672.1 |Severe acute respiratory syndrome coro    ···················· ···················· ···················· ···················· ···················· ····················
20440 MT952673.1 |Severe acute respiratory syndrome coro    ···················· ···················· ···················· ···················· ···················· ····················
20441 MT952675.1 |Severe acute respiratory syndrome coro    ···················· ···················· ···················· ···················· ···················· ····················
20442 MT952677.1 |Severe acute respiratory syndrome coro    ········T··········· ···················· ···················· ···················· ···················· ····················
20443 MT952678.1 |Severe acute respiratory syndrome coro    ···················· ···················· ···················· ···················· ···················· ····················
20444 MT952679.1 |Severe acute respiratory syndrome coro    ···················· ···················· ···················· ···················· ···················· ····················
20445 MT952680.1 |Severe acute respiratory syndrome coro    ···················· ···················· ···················· ···················· ···················· ····················
20446 MT952681.1 |Severe acute respiratory syndrome coro    ···················· ···················· ···················· ···················· ···················· ····················
20447 MT952682.1 |Severe acute respiratory syndrome coro    ···················· ···················· ···················· ···················· ···················· ····················
20448 MT952683.1 |Severe acute respiratory syndrome coro    ···················· ···················· ···················· ···················· ···················· ····················
20449 MT952684.1 |Severe acute respiratory syndrome coro    ···················· ···················· ···················· ···················· ···················· ····················
20450 MT952685.1 |Severe acute respiratory syndrome coro    ···················· ···················· ···················· ···················· ···················· ····················
20451 MT952687.1 |Severe acute respiratory syndrome coro    ···················· ···················· ···················· ···················· ···················· ····················
20452 MT952688.1 |Severe acute respiratory syndrome coro    ···················· ···················· ···················· ···················· ···················· ····················
20453 MT952690.1 |Severe acute respiratory syndrome coro    ···················· ····A··············· ···················· ···················· ···················· ····················
20454 MT952691.1 |Severe acute respiratory syndrome coro    ···················· ···················· ···················· ···················· ···················· ····················
20455 MT952692.1 |Severe acute respiratory syndrome coro    ···················· ···················· ···················· ···················· ···················· ····················
20456 MT952693.1 |Severe acute respiratory syndrome coro    ···················· ···················· ···················· ···················· ···················· ····················
20457 MT952694.1 |Severe acute respiratory syndrome coro    ···················· ···················· ···················· ···················· ···················· ····················
20458 MT952695.1 |Severe acute respiratory syndrome coro    ···················· ···················· ···················· ···················· ···················· ····················
20459 MT952696.1 |Severe acute respiratory syndrome coro    ···················· ···················· ···················· ···················· ···················· ····················
20460 MT952697.1 |Severe acute respiratory syndrome coro    ···················· ···················· ···················· ···················· ···················· ····················
20461 MT952698.1 |Severe acute respiratory syndrome coro    ···················· ···················· ···················· ···················· ···················· ····················
20462 MT952700.1 |Severe acute respiratory syndrome coro    ···················· ···················· ···················· ···················· ···················· ····················
20463 MT952701.1 |Severe acute respiratory syndrome coro    ···················· ···················· ···················· ···················· ···················· ····················
20464 MT952702.1 |Severe acute respiratory syndrome coro    ···················· ···················· ···················· ···················· ···················· ····················
20465 MT947074.1 |Severe acute respiratory syndrome coro    ···················· ···················· ···················· ···················· ···················· ····················
20466 MT947075.1 |Severe acute respiratory syndrome coro    ···················· ···················· ···················· ···················· ···················· ····················
20467 MT947076.1 |Severe acute respiratory syndrome coro    ···················· ···················· ···················· ···················· ···················· ····················
20468 MT947077.1 |Severe acute respiratory syndrome coro    ···················· ···················· ···················· ···················· ···················· ····················
20469 MT947078.1 |Severe acute respiratory syndrome coro    ···················· ···················· ···················· ···················· ···················· ····················
20470 MT947079.1 |Severe acute respiratory syndrome coro    ···················· ···················· ···················· ···················· ···················· ····················
20471 MT947080.1 |Severe acute respiratory syndrome coro    ···················· ···················· ···················· ···················· ···················· ····················
20472 MT947081.1 |Severe acute respiratory syndrome coro    ···················· ···················· ···················· ···················· ···················· ····················
20473 MT947082.1 |Severe acute respiratory syndrome coro    ···················· ···················· ···················· ···················· ···················· ····················
20474 MT947083.1 |Severe acute respiratory syndrome coro    ···················· ···················· ···················· ···················· ···················· ····················
20475 MT947084.1 |Severe acute respiratory syndrome coro    ···················· ···················· ···················· ···················· ···················· ····················
20476 MT947153.1 |Severe acute respiratory syndrome coro    ···················· ···················· ···················· ···················· ···················· ····················
20477 MT947154.1 |Severe acute respiratory syndrome coro    ···················· ···················· ···················· ···················· ···················· ····················
20478 MT947155.1 |Severe acute respiratory syndrome coro    ···················· ···················· ···················· ···················· ···················· ····················
20479 MT947156.1 |Severe acute respiratory syndrome coro    ···················· ···················· ···················· ···················· ···················· ····················
20480 MT947157.1 |Severe acute respiratory syndrome coro    ···················· ···················· ···················· ···················· ···················· ····················
20481 MT947158.1 |Severe acute respiratory syndrome coro    ···················· ···················· ···················· ···················· ···················· ····················
20482 MT947159.1 |Severe acute respiratory syndrome coro    ···················· ···················· ···················· ···················· ···················· ····················
20483 MT947160.1 |Severe acute respiratory syndrome coro    ···················· ···················· ···················· ···················· ···················· ····················
20484 MT947161.1 |Severe acute respiratory syndrome coro    ···················· ···················· ···················· ···················· ···················· ····················
20485 MT947162.1 |Severe acute respiratory syndrome coro    ···················· ···················· ···················· ···················· ···················· ····················
20486 MT947163.1 |Severe acute respiratory syndrome coro    ···················· ···················· ···················· ···················· ···················· ····················
20487 MT947165.1 |Severe acute respiratory syndrome coro    ···················· ···················· ···················· ···················· ···················· ····················
20488 MT947166.1 |Severe acute respiratory syndrome coro    ···················· ···················· ···················· ···················· ···················· ····················
20489 MT947167.1 |Severe acute respiratory syndrome coro    ···················· ···················· ···················· ···················· ···················· ····················
20490 MT947168.1 |Severe acute respiratory syndrome coro    ···················· ···················· ···················· ···················· ···················· ····················
20491 MT947169.1 |Severe acute respiratory syndrome coro    ···················· ···················· ···················· ···················· ···················· ····················
20492 MT947170.1 |Severe acute respiratory syndrome coro    ···················· ···················· ···················· ···················· ···················· ····················
20493 MT947171.1 |Severe acute respiratory syndrome coro    ···················· ···················· ···················· ···················· ···················· ····················
20494 MT947172.1 |Severe acute respiratory syndrome coro    ···················· ···················· ···················· ···················· ···················· ····················
20495 MT947173.1 |Severe acute respiratory syndrome coro    ···················· ···················· ···················· ···················· ···················· ····················
```

20496 MT947174.1 |Severe acute respiratory syndrome coro ···················· ···················· ···················· ···················· ···················· ····················
20497 MT947175.1 |Severe acute respiratory syndrome coro ···················· ···················· ···················· ···················· ···················· ····················
20498 MT947546.1 |Severe acute respiratory syndrome coro ···················· ···················· ···················· ···················· ···················· ····················
20499 MT947547.1 |Severe acute respiratory syndrome coro ···················· ···················· ···················· ···················· ···················· ····················
20500 MT947548.1 |Severe acute respiratory syndrome coro ···················· ···················· ···················· ···················· ···················· ····················
20501 MT947549.1 |Severe acute respiratory syndrome coro ···················· ···················· ···················· ···················· ···················· ····················
20502 MT947550.1 |Severe acute respiratory syndrome coro ···················· ···················· ···················· ···················· ···················· ····················
20503 MT947551.1 |Severe acute respiratory syndrome coro ···················· ···················· ···················· ···················· ···················· ····················
20504 MT947552.1 |Severe acute respiratory syndrome coro ···················· ···················· ···················· ···················· ···················· ····················
20505 MT947553.1 |Severe acute respiratory syndrome coro ···················· ···················· ···················· ···················· ···················· ····················
20506 MT947554.1 |Severe acute respiratory syndrome coro ···················· ···················· ···················· ···················· ···················· ····················
20507 MT947555.1 |Severe acute respiratory syndrome coro ···················· ···················· ···················· ···················· ···················· ····················
20508 MT947556.1 |Severe acute respiratory syndrome coro ···················· ···················· ···················· ···················· ···················· ····················
20509 MT947557.1 |Severe acute respiratory syndrome coro ···················· ···················· ···················· ···················· ···················· ····················
20510 MT947558.1 |Severe acute respiratory syndrome coro ···················· ···················· ···················· ···················· ···················· ····················
20511 MT947559.1 |Severe acute respiratory syndrome coro ···················· ···················· ···················· ···················· ···················· ····················
20512 MT947560.1 |Severe acute respiratory syndrome coro ···················· ···················· ···················· ···················· ···················· ····················
20513 MT947561.1 |Severe acute respiratory syndrome coro ···················· ·A···A·············· ···················· ···················· ···················· ····················
20514 MT947562.1 |Severe acute respiratory syndrome coro ···················· ···················· ···················· ···················· ···················· ····················
20515 MT947563.1 |Severe acute respiratory syndrome coro ···················· ···················· ···················· ···················· ···················· ····················
20516 MT947564.1 |Severe acute respiratory syndrome coro ···················· ···················· ···················· ···················· ···················· ····················
20517 MT947565.1 |Severe acute respiratory syndrome coro ···················· ···················· ···················· ···················· ···················· ····················
20518 MT947566.1 |Severe acute respiratory syndrome coro ···················· ···················· ···················· ···················· ···················· ····················
20519 MT947568.1 |Severe acute respiratory syndrome coro ···················· ···················· ···················· ···················· ···················· ····················
20520 MT947570.1 |Severe acute respiratory syndrome coro ···················· ···················· ···················· ···················· ···················· ····················
20521 MT947571.1 |Severe acute respiratory syndrome coro ···················· ···················· ···················· ···················· ···················· ····················
20522 MT947572.1 |Severe acute respiratory syndrome coro ···················· ···················· ···················· ···················· ···················· ····················
20523 MT947573.1 |Severe acute respiratory syndrome coro ···················· ···················· ···················· ···················· ···················· ····················
20524 MT947574.1 |Severe acute respiratory syndrome coro ···················· ···················· ···················· ···················· ···················· ····················
20525 MT947575.1 |Severe acute respiratory syndrome coro ···················· ·A·················· ···················· ···················· ···················· ····················
20526 MT947576.1 |Severe acute respiratory syndrome coro ···················· ···················· ···················· ···················· ···················· ····················
20527 MT947577.1 |Severe acute respiratory syndrome coro ···················· ···················· ···················· ···················· ···················· ····················
20528 MT947578.1 |Severe acute respiratory syndrome coro ···················· ···················· ···················· ···················· ···················· ····················
20529 MT947579.1 |Severe acute respiratory syndrome coro ···················· ···················· ···················· ···················· ···················· ····················
20530 MT947580.1 |Severe acute respiratory syndrome coro ···················· ···················· ···················· ···················· ···················· ····················
20531 MT947581.1 |Severe acute respiratory syndrome coro ···················· ···················· ···················· ···················· ···················· ····················
20532 MT947582.1 |Severe acute respiratory syndrome coro ···················· ···················· ···················· ···················· ···················· ····················
20533 MT947583.1 |Severe acute respiratory syndrome coro ···················· ···················· ···················· ···················· ···················· ····················
20534 MT947584.1 |Severe acute respiratory syndrome coro ···················· ···················· ···················· ···················· ···················· ····················
20535 MT947586.1 |Severe acute respiratory syndrome coro ···················· ···················· ···················· ···················· ···················· ····················
20536 MT947587.1 |Severe acute respiratory syndrome coro ···················· ···················· ···················· ···················· ···················· ····················
20537 MT947588.1 |Severe acute respiratory syndrome coro ···················· ···················· ···················· ···················· ···················· ····················
20538 MT947591.1 |Severe acute respiratory syndrome coro ···················· ···················· ···················· ···················· ···················· ····················
20539 MT947592.1 |Severe acute respiratory syndrome coro ···················· ···················· ···················· ···················· ···················· ····················
20540 MT947593.1 |Severe acute respiratory syndrome coro ···················· ·A·················· ···················· ···················· ···················· ····················
20541 MT947595.1 |Severe acute respiratory syndrome coro ···················· ···················· ···················· ···················· ···················· ····················
20542 MT941047.1 |Severe acute respiratory syndrome coro ···················· ·A·················· ···················· ···················· ···················· ··A·················
20543 MT941052.1 |Severe acute respiratory syndrome coro ···················· ·A·················· ···················· ···················· ···················· ··A·················
20544 MT941055.1 |Severe acute respiratory syndrome coro ···················· ·A·················· ···················· ···················· ···················· ··A·················
20545 MT941056.1 |Severe acute respiratory syndrome coro ···················· ·A·················· ···················· ···················· ···················· ··A·················
20546 MT941059.1 |Severe acute respiratory syndrome coro ···················· ·A·················· ···················· ···················· ···················· ··A·················
20547 MT941060.1 |Severe acute respiratory syndrome coro ···················· ·A·················· ···················· ···················· ···················· ··A·················
20548 MT941061.1 |Severe acute respiratory syndrome coro ···················· ·A·················· ···················· ···················· ···················· ··A·················
20549 MT941067.1 |Severe acute respiratory syndrome coro ···················· ·A·················· ···················· ···················· ···················· ··A·················
20550 MT941272.1 |Severe acute respiratory syndrome coro ···················· ··········A········· ···················· ···················· ···················· ····················
20551 MT941274.1 |Severe acute respiratory syndrome coro ···················· ·A·················· ···················· ···················· ···················· ··A·················
20552 MT941276.1 |Severe acute respiratory syndrome coro ···················· ·A·················· ···················· ···················· ···················· ··A·················
20553 MT941403.1 |Severe acute respiratory syndrome coro ···················· ·A·················· ···················· ···················· ···················· ··A·················
20554 MT946869.1 |Severe acute respiratory syndrome coro ···················· ···················· ···················· ···················· ···················· ····················
20555 MT946870.1 |Severe acute respiratory syndrome coro ···················· ···················· ···················· ···················· ···················· ····················
20556 MT946871.1 |Severe acute respiratory syndrome coro ···················· ···················· ···················· ···················· ···················· ····················
20557 MT946872.1 |Severe acute respiratory syndrome coro ···················· ···················· ···················· ···················· ···················· ····················
20558 MT946873.1 |Severe acute respiratory syndrome coro ···················· ···················· ···················· ···················· ···················· ····················
20559 MT946884.1 |Severe acute respiratory syndrome coro ···················· ···················· ···················· ···················· ···················· ····················
20560 MT946885.1 |Severe acute respiratory syndrome coro ···················· ···················· ···················· ···················· ···················· ····················
20561 MT946886.1 |Severe acute respiratory syndrome coro ···················· ···················· ···················· ···················· ···················· ····················
20562 MT946887.1 |Severe acute respiratory syndrome coro ···················· ···················· ···················· ···················· ···················· ····················
20563 MT946888.1 |Severe acute respiratory syndrome coro ···················· ···················· ···················· ···················· ···················· ····················
20564 MT946889.1 |Severe acute respiratory syndrome coro ···················· ···················· ···················· ···················· ···················· ····················
20565 MT946890.1 |Severe acute respiratory syndrome coro ···················· ···················· ···················· ···················· ···················· ····················
20566 MT946891.1 |Severe acute respiratory syndrome coro ···················· ···················· ···················· ···················· ···················· ····················
20567 MT946892.1 |Severe acute respiratory syndrome coro ···················· ···················· ···················· ···················· ···················· ····················
20568 MT946893.1 |Severe acute respiratory syndrome coro ···················· ···················· ···················· ···················· ···················· ····················
20569 MT946894.1 |Severe acute respiratory syndrome coro ···················· ···················· ···················· ···················· ···················· ····················
20570 MT946895.1 |Severe acute respiratory syndrome coro ···················· ·A·················· ···················· ···················· ···················· ····················
20571 MT946896.1 |Severe acute respiratory syndrome coro ···················· ···················· ···················· ···················· ···················· ····················
20572 MT845877.1 |Severe acute respiratory syndrome coro ···················· ···················· ···················· ···················· ···················· ····················
20573 MT845878.1 |Severe acute respiratory syndrome coro ···················· ···················· ···················· ···················· ···················· ····················

| | | | | | | | | |
|---|---|---|---|---|---|---|---|---|
| 20574 | MT940449.1 | \|Severe acute respiratory syndrome coro | ···················· | ·A·················· | ···················· | ···················· | ···················· | ···A················ |
| 20575 | MT940451.1 | \|Severe acute respiratory syndrome coro | ···················· | ·A·················· | ···················· | ···················· | ···················· | ···A················ |
| 20576 | MT940452.1 | \|Severe acute respiratory syndrome coro | ···················· | ·A·················· | ···················· | ···················· | ···················· | ···A················ |
| 20577 | MT940458.1 | \|Severe acute respiratory syndrome coro | ···················· | ·A·················· | ···················· | ···················· | ···················· | ···A················ |
| 20578 | MT940459.1 | \|Severe acute respiratory syndrome coro | ···················· | ·A·················· | ···················· | ···················· | ···················· | ···A················ |
| 20579 | MT940460.1 | \|Severe acute respiratory syndrome coro | ···T················ | ·A·················· | ···················· | ···················· | ···················· | ···A················ |
| 20580 | MT940461.1 | \|Severe acute respiratory syndrome coro | ···················· | ·A·················· | ···················· | ···················· | ···················· | ···A················ |
| 20581 | MT940462.1 | \|Severe acute respiratory syndrome coro | ···················· | ·A·················· | ···················· | ···················· | ···················· | ···A················ |
| 20582 | MT940464.1 | \|Severe acute respiratory syndrome coro | ···················· | ·A·················· | ···················· | ···················· | ···················· | ···A················ |
| 20583 | MT940472.1 | \|Severe acute respiratory syndrome coro | ···················· | ···················· | ···················· | ···················· | ···················· | ···················· |
| 20584 | MT940473.1 | \|Severe acute respiratory syndrome coro | ···················· | ···················· | ···················· | ···················· | ···················· | ···················· |
| 20585 | MT940474.1 | \|Severe acute respiratory syndrome coro | ···················· | ·A·················· | ···················· | ···················· | ···················· | ···A················ |
| 20586 | MT940475.1 | \|Severe acute respiratory syndrome coro | ···················· | ·A·················· | ···················· | ···················· | ···················· | ···A················ |
| 20587 | MT940476.1 | \|Severe acute respiratory syndrome coro | ···················· | ·A·················· | ···················· | ···················· | ···················· | ···A················ |
| 20588 | MT940477.1 | \|Severe acute respiratory syndrome coro | ···················· | ···················· | ···················· | ···················· | ···················· | ···················· |
| 20589 | MT940478.1 | \|Severe acute respiratory syndrome coro | ···················· | ···················· | ···················· | ···················· | ···················· | ···················· |
| 20590 | MT940479.1 | \|Severe acute respiratory syndrome coro | ···················· | ···················· | ···················· | ···················· | ···················· | ···················· |
| 20591 | MT940481.1 | \|Severe acute respiratory syndrome coro | ···················· | ···················· | ···················· | ···················· | ···················· | ···················· |
| 20592 | MT940482.1 | \|Severe acute respiratory syndrome coro | ···················· | ···················· | ···················· | ···················· | ···················· | ···················· |
| 20593 | MT940483.1 | \|Severe acute respiratory syndrome coro | ···················· | ···················· | ···················· | ···················· | ···················· | ···················· |
| 20594 | MT940484.1 | \|Severe acute respiratory syndrome coro | ···················· | ···················· | ···················· | ···················· | ···················· | ···················· |
| 20595 | MT940485.1 | \|Severe acute respiratory syndrome coro | ···················· | ···················· | ···················· | ···················· | ···················· | ···················· |
| 20596 | MT940486.1 | \|Severe acute respiratory syndrome coro | ···················· | ···················· | ···················· | ···················· | ···················· | ···················· |
| 20597 | MT940487.1 | \|Severe acute respiratory syndrome coro | ···················· | ···················· | ···················· | ···················· | ···················· | ···················· |
| 20598 | MT940488.1 | \|Severe acute respiratory syndrome coro | ···················· | ···················· | ···················· | ···················· | ···················· | ···················· |
| 20599 | MT940489.1 | \|Severe acute respiratory syndrome coro | ···················· | ···················· | ···················· | ···················· | ···················· | ···················· |
| 20600 | MT940490.1 | \|Severe acute respiratory syndrome coro | ···················· | ···················· | ···················· | ···················· | ···················· | ···················· |
| 20601 | MT940491.1 | \|Severe acute respiratory syndrome coro | ···················· | ···················· | ···················· | ···················· | ···················· | ···················· |
| 20602 | MT940492.1 | \|Severe acute respiratory syndrome coro | ···················· | ···················· | ···················· | ···················· | ···················· | ···················· |
| 20603 | MT940493.1 | \|Severe acute respiratory syndrome coro | ···················· | ···················· | ···················· | ···················· | ···················· | ···················· |
| 20604 | MT940494.1 | \|Severe acute respiratory syndrome coro | ···················· | ···················· | ···················· | ···················· | ···················· | ···················· |
| 20605 | MT940495.1 | \|Severe acute respiratory syndrome coro | ···················· | ···················· | ···················· | ···················· | ···················· | ···················· |
| 20606 | MT940496.1 | \|Severe acute respiratory syndrome coro | ···················· | ···················· | ···················· | ···················· | ···················· | ···················· |
| 20607 | MT940497.1 | \|Severe acute respiratory syndrome coro | ···················· | ···················· | ···················· | ···················· | ···················· | ···················· |
| 20608 | MT940498.1 | \|Severe acute respiratory syndrome coro | ···················· | ···················· | ···················· | ···················· | ···················· | ···················· |
| 20609 | MT940499.1 | \|Severe acute respiratory syndrome coro | ···················· | ···················· | ···················· | ···················· | ···················· | ···················· |
| 20610 | MT940500.1 | \|Severe acute respiratory syndrome coro | ···················· | ···················· | ···················· | ···················· | ···················· | ···················· |
| 20611 | MT940501.1 | \|Severe acute respiratory syndrome coro | ···················· | ···················· | ···················· | ···················· | ···················· | ···················· |
| 20612 | MT940502.1 | \|Severe acute respiratory syndrome coro | ···················· | ···················· | ···················· | ···················· | ···················· | ···················· |
| 20613 | MT940503.1 | \|Severe acute respiratory syndrome coro | ···················· | ···················· | ···················· | ···················· | ···················· | ···················· |
| 20614 | MT940504.1 | \|Severe acute respiratory syndrome coro | ···················· | ···················· | ···················· | ···················· | ···················· | ···················· |
| 20615 | MT940505.1 | \|Severe acute respiratory syndrome coro | ···················· | ···················· | ···················· | ···················· | ···················· | ···················· |
| 20616 | MT940507.1 | \|Severe acute respiratory syndrome coro | ···················· | ···················· | ···················· | ···················· | ···················· | ···················· |
| 20617 | MT940508.1 | \|Severe acute respiratory syndrome coro | ···················· | ···················· | ···················· | ···················· | ···················· | ···················· |
| 20618 | MT940509.1 | \|Severe acute respiratory syndrome coro | ···················· | ···················· | ···················· | ···················· | ···················· | ···················· |
| 20619 | MT940510.1 | \|Severe acute respiratory syndrome coro | ···················· | ···················· | ···················· | ···················· | ···················· | ···················· |
| 20620 | MT940511.1 | \|Severe acute respiratory syndrome coro | ···················· | ···················· | ···················· | ···················· | ···················· | ···················· |
| 20621 | MT940512.1 | \|Severe acute respiratory syndrome coro | ···················· | ···················· | ···················· | ···················· | ···················· | ···················· |
| 20622 | MT940513.1 | \|Severe acute respiratory syndrome coro | ···················· | ···················· | ···················· | ···················· | ···················· | ···················· |
| 20623 | MT940514.1 | \|Severe acute respiratory syndrome coro | ···················· | ···················· | ···················· | ···················· | ···················· | ···················· |
| 20624 | MT940515.1 | \|Severe acute respiratory syndrome coro | ···················· | ·A·················· | ···················· | ···················· | ···················· | ···················· |
| 20625 | MT940516.1 | \|Severe acute respiratory syndrome coro | ···················· | ·A·················· | ···················· | ···················· | ···················· | ···················· |
| 20626 | MT940517.1 | \|Severe acute respiratory syndrome coro | ···················· | ·A·················· | ···················· | ···················· | ···················· | ···················· |
| 20627 | MT940518.1 | \|Severe acute respiratory syndrome coro | ···················· | ·A·················· | ···················· | ···················· | ···················· | ···················· |
| 20628 | MT940519.1 | \|Severe acute respiratory syndrome coro | ···················· | ·A·················· | ···················· | ···················· | ···················· | ···················· |
| 20629 | MT940520.1 | \|Severe acute respiratory syndrome coro | ···················· | ·A·················· | ···················· | ···················· | ···················· | ···················· |
| 20630 | MT940521.1 | \|Severe acute respiratory syndrome coro | ···················· | ·A·················· | ···················· | ···················· | ···················· | ···················· |
| 20631 | MT940522.1 | \|Severe acute respiratory syndrome coro | ···················· | ···················· | ···················· | ···················· | ···················· | ···················· |
| 20632 | MT940523.1 | \|Severe acute respiratory syndrome coro | ···················· | ···················· | ···················· | ···················· | ···················· | ···················· |
| 20633 | MT940524.1 | \|Severe acute respiratory syndrome coro | ···················· | ···················· | ···················· | ···················· | ···················· | ···················· |
| 20634 | MT940525.1 | \|Severe acute respiratory syndrome coro | ···················· | ···················· | ···················· | ···················· | ···················· | ···················· |
| 20635 | MT940526.1 | \|Severe acute respiratory syndrome coro | ···················· | ···················· | ···················· | ···················· | ···················· | ···················· |
| 20636 | MT937317.1 | \|Severe acute respiratory syndrome coro | ···················· | ···················· | ···················· | ···················· | ···················· | ···················· |
| 20637 | MT937318.1 | \|Severe acute respiratory syndrome coro | ···················· | ···················· | ···················· | ···················· | ···················· | ···················· |
| 20638 | MT937319.1 | \|Severe acute respiratory syndrome coro | ···················· | ···················· | ···················· | ···················· | ···················· | ···················· |
| 20639 | MT937320.1 | \|Severe acute respiratory syndrome coro | ···················· | ···················· | ···················· | ···················· | ···················· | ···················· |
| 20640 | MT937321.1 | \|Severe acute respiratory syndrome coro | ···················· | ···················· | ···················· | ···················· | ···················· | ···················· |
| 20641 | MT937322.1 | \|Severe acute respiratory syndrome coro | ···················· | ···················· | ···················· | ···················· | ···················· | ···················· |
| 20642 | MT937323.1 | \|Severe acute respiratory syndrome coro | ···················· | ···················· | ···················· | ···················· | ···················· | ···················· |
| 20643 | MT937324.1 | \|Severe acute respiratory syndrome coro | ···················· | ···················· | ···················· | ···················· | ···················· | ···················· |
| 20644 | MT937325.1 | \|Severe acute respiratory syndrome coro | ···················· | ···················· | ···················· | ···················· | ···················· | ···················· |
| 20645 | MT937326.1 | \|Severe acute respiratory syndrome coro | ···················· | ···················· | ···················· | ···················· | ···················· | ···················· |
| 20646 | MT937808.1 | \|Severe acute respiratory syndrome coro | ···················· | ···················· | ···················· | ···················· | ···················· | ···················· |
| 20647 | MT937810.1 | \|Severe acute respiratory syndrome coro | ···················· | ···················· | ···················· | ···················· | ···················· | ···················· |
| 20648 | MT937811.1 | \|Severe acute respiratory syndrome coro | ···················· | ···················· | ···················· | ···················· | ···················· | ···················· |
| 20649 | MT937812.1 | \|Severe acute respiratory syndrome coro | ···················· | ···················· | ···················· | ···················· | ···················· | ···················· |
| 20650 | MT937813.1 | \|Severe acute respiratory syndrome coro | ···················· | ···················· | ···················· | ···················· | ···················· | ···················· |
| 20651 | MT937814.1 | \|Severe acute respiratory syndrome coro | ···················· | ···················· | ···················· | ···················· | ···················· | ···················· |

| | | | | | | | | |
|---|---|---|---|---|---|---|---|---|
| 20652 | MT937815.1 | \|Severe acute respiratory syndrome coro | ···················· | ···················· | ···················· | ···················· | ···················· | ···················· |
| 20653 | MT937816.1 | \|Severe acute respiratory syndrome coro | ···················· | ···················· | ···················· | ···················· | ···················· | ···················· |
| 20654 | MT937817.1 | \|Severe acute respiratory syndrome coro | ···················· | ···················· | ···················· | ···················· | ···················· | ···················· |
| 20655 | MT937818.1 | \|Severe acute respiratory syndrome coro | ···················· | ···················· | ···················· | ···················· | ···················· | ···················· |
| 20656 | MT937820.1 | \|Severe acute respiratory syndrome coro | ···················· | ···················· | ···················· | ···················· | ···················· | ···················· |
| 20657 | MT937821.1 | \|Severe acute respiratory syndrome coro | ···················· | ···················· | ···················· | ···················· | ···················· | ···················· |
| 20658 | MT937822.1 | \|Severe acute respiratory syndrome coro | ···················· | ······A············· | ···················· | ···················· | ···················· | ···················· |
| 20659 | MT937823.1 | \|Severe acute respiratory syndrome coro | ···················· | ···················· | ···················· | ···················· | ···················· | ···················· |
| 20660 | MT937824.1 | \|Severe acute respiratory syndrome coro | ···················· | ···················· | ···················· | ···················· | ···················· | ···················· |
| 20661 | MT937825.1 | \|Severe acute respiratory syndrome coro | ···················· | ···················· | ···················· | ···················· | ···················· | ···················· |
| 20662 | MT937826.1 | \|Severe acute respiratory syndrome coro | ···················· | ···················· | ···················· | ···················· | ···················· | ···················· |
| 20663 | MT937827.1 | \|Severe acute respiratory syndrome coro | ···················· | ···················· | ···················· | ···················· | ···················· | ···················· |
| 20664 | MT928960.1 | \|Severe acute respiratory syndrome coro | ···················· | ···················· | ···················· | ···················· | ···················· | ···················· |
| 20665 | MT928961.1 | \|Severe acute respiratory syndrome coro | ···················· | ···················· | ···················· | ···················· | ···················· | ···················· |
| 20666 | MT928962.1 | \|Severe acute respiratory syndrome coro | ···················· | ···················· | ···················· | ···················· | ···················· | ···················· |
| 20667 | MT928963.1 | \|Severe acute respiratory syndrome coro | ···················· | ···················· | ···················· | ···················· | ···················· | ···················· |
| 20668 | MT928964.1 | \|Severe acute respiratory syndrome coro | ···················· | ·A·················· | ···················· | ···················· | ···················· | ···················· |
| 20669 | MT928965.1 | \|Severe acute respiratory syndrome coro | ···················· | ···················· | ···················· | ···················· | ···················· | ···················· |
| 20670 | MT928966.1 | \|Severe acute respiratory syndrome coro | ···················· | ···················· | ···················· | ···················· | ···················· | ···················· |
| 20671 | MT928967.1 | \|Severe acute respiratory syndrome coro | ···················· | ···················· | ···················· | ···················· | ···················· | ···················· |
| 20672 | MT928968.1 | \|Severe acute respiratory syndrome coro | ···················· | ···················· | ···················· | ···················· | ···················· | ···················· |
| 20673 | MT928970.1 | \|Severe acute respiratory syndrome coro | ···················· | ···················· | ···················· | ···················· | ···················· | ···················· |
| 20674 | MT928971.1 | \|Severe acute respiratory syndrome coro | ···················· | ···················· | ···················· | ···················· | ···················· | ···················· |
| 20675 | MT928972.1 | \|Severe acute respiratory syndrome coro | ···················· | ···················· | ···················· | ···················· | ···················· | ···················· |
| 20676 | MT928973.1 | \|Severe acute respiratory syndrome coro | ···················· | ···················· | ···················· | ···················· | ···················· | ···················· |
| 20677 | MT928974.1 | \|Severe acute respiratory syndrome coro | ···················· | ···················· | ···················· | ···················· | ···················· | ···················· |
| 20678 | MT928975.1 | \|Severe acute respiratory syndrome coro | ···················· | ···················· | ···················· | ···················· | ···················· | ···················· |
| 20679 | MT928976.1 | \|Severe acute respiratory syndrome coro | ···················· | ···················· | ···················· | ···················· | ···················· | ···················· |
| 20680 | MT928979.1 | \|Severe acute respiratory syndrome coro | ···················· | ···················· | ···················· | ···················· | ···················· | ···················· |
| 20681 | MT928980.1 | \|Severe acute respiratory syndrome coro | ···················· | ···················· | ···················· | ···················· | ···················· | ···················· |
| 20682 | MT928981.1 | \|Severe acute respiratory syndrome coro | ···················· | ···················· | ···················· | ···················· | ···················· | ···················· |
| 20683 | MT928982.1 | \|Severe acute respiratory syndrome coro | ···················· | ···················· | ···················· | ···················· | ···················· | ···················· |
| 20684 | MT928983.1 | \|Severe acute respiratory syndrome coro | ···················· | ···················· | ···················· | ···················· | ···················· | ···················· |
| 20685 | MT928984.1 | \|Severe acute respiratory syndrome coro | ···················· | ···················· | ···················· | ···················· | ···················· | ···················· |
| 20686 | MT928985.1 | \|Severe acute respiratory syndrome coro | ···················· | ···················· | ···················· | ···················· | ···················· | ···················· |
| 20687 | MT928987.1 | \|Severe acute respiratory syndrome coro | ···················· | ···················· | ···················· | ···················· | ···················· | ···················· |
| 20688 | MT928988.1 | \|Severe acute respiratory syndrome coro | ···················· | ···················· | ···················· | ···················· | ···················· | ···················· |
| 20689 | MT928989.1 | \|Severe acute respiratory syndrome coro | ···················· | ···················· | ···················· | ···················· | ···················· | ···················· |
| 20690 | MT928990.1 | \|Severe acute respiratory syndrome coro | ···················· | ···················· | ···················· | ···················· | ···················· | ···················· |
| 20691 | MT928991.1 | \|Severe acute respiratory syndrome coro | ···················· | ···················· | ···················· | ···················· | ···················· | ···················· |
| 20692 | MT928992.1 | \|Severe acute respiratory syndrome coro | ···················· | ···················· | ···················· | ···················· | ···················· | ···················· |
| 20693 | MT928994.1 | \|Severe acute respiratory syndrome coro | ···················· | ···················· | ···················· | ···················· | ···················· | ···················· |
| 20694 | MT928995.1 | \|Severe acute respiratory syndrome coro | ···················· | ···················· | ···················· | ···················· | ···················· | ···················· |
| 20695 | MT928996.1 | \|Severe acute respiratory syndrome coro | ···················· | ···················· | ···················· | ···················· | ···················· | ···················· |
| 20696 | MT928998.1 | \|Severe acute respiratory syndrome coro | ···················· | ···················· | ···················· | ···················· | ···················· | ···················· |
| 20697 | MT928999.1 | \|Severe acute respiratory syndrome coro | ···················· | ···················· | ···················· | ···················· | ···················· | ···················· |
| 20698 | MT929000.1 | \|Severe acute respiratory syndrome coro | ···················· | ···················· | ···················· | ···················· | ···················· | ···················· |
| 20699 | MT929001.1 | \|Severe acute respiratory syndrome coro | ···················· | ···················· | ···················· | ···················· | ···················· | ···················· |
| 20700 | MT929002.1 | \|Severe acute respiratory syndrome coro | ···················· | ·············A······ | ···················· | ···················· | ···················· | ···················· |
| 20701 | MT929003.1 | \|Severe acute respiratory syndrome coro | ···················· | ···················· | ···················· | ···················· | ···················· | ···················· |
| 20702 | MT929004.1 | \|Severe acute respiratory syndrome coro | ···················· | ···················· | ···················· | ···················· | ···················· | ···················· |
| 20703 | MT929005.1 | \|Severe acute respiratory syndrome coro | ···················· | ···················· | ···················· | ···················· | ···················· | ···················· |
| 20704 | MT929006.1 | \|Severe acute respiratory syndrome coro | ···················· | ···················· | ···················· | ···················· | ···················· | ···················· |
| 20705 | MT929007.1 | \|Severe acute respiratory syndrome coro | ···················· | ···················· | ···················· | ···················· | ···················· | ···················· |
| 20706 | MT929008.1 | \|Severe acute respiratory syndrome coro | ···················· | ···················· | ···················· | ···················· | ···················· | ···················· |
| 20707 | MT929009.1 | \|Severe acute respiratory syndrome coro | ···················· | ···················· | ···················· | ···················· | ···················· | ···················· |
| 20708 | MT929010.1 | \|Severe acute respiratory syndrome coro | ···················· | ···················· | ···················· | ···················· | ···················· | ···················· |
| 20709 | MT929011.1 | \|Severe acute respiratory syndrome coro | ···················· | ···················· | ···················· | ···················· | ···················· | ···················· |
| 20710 | MT929012.1 | \|Severe acute respiratory syndrome coro | ···················· | ···················· | ···················· | ···················· | ···················· | ···················· |
| 20711 | MT929013.1 | \|Severe acute respiratory syndrome coro | ···················· | ·A·················· | ···················· | ···················· | ···················· | ···················· |
| 20712 | MT929014.1 | \|Severe acute respiratory syndrome coro | ···················· | ···················· | ···················· | ···················· | ···················· | ···················· |
| 20713 | MT929015.1 | \|Severe acute respiratory syndrome coro | ···················· | ···················· | ···················· | ···················· | ···················· | ···················· |
| 20714 | MT929016.1 | \|Severe acute respiratory syndrome coro | ···················· | ···················· | ···················· | ···················· | ···················· | ···················· |
| 20715 | MT929017.1 | \|Severe acute respiratory syndrome coro | ···················· | ···················· | ···················· | ···················· | ···················· | ···················· |
| 20716 | MT929018.1 | \|Severe acute respiratory syndrome coro | ···················· | ·············A······ | ···················· | ···················· | ···················· | ···················· |
| 20717 | MT929019.1 | \|Severe acute respiratory syndrome coro | ···················· | ···················· | ···················· | ···················· | ···················· | ···················· |
| 20718 | MT929020.1 | \|Severe acute respiratory syndrome coro | ···················· | ···················· | ···················· | ···················· | ···················· | ···················· |
| 20719 | MT929021.1 | \|Severe acute respiratory syndrome coro | ···················· | ·············A······ | ···················· | ···················· | ···················· | ···················· |
| 20720 | MT929023.1 | \|Severe acute respiratory syndrome coro | ···················· | ···················· | ···················· | ···················· | ···················· | ···················· |
| 20721 | MT929024.1 | \|Severe acute respiratory syndrome coro | ···················· | ···················· | ···················· | ···················· | ···················· | ···················· |
| 20722 | MT929025.1 | \|Severe acute respiratory syndrome coro | ···················· | ···················· | ···················· | ···················· | ···················· | ···················· |
| 20723 | MT929026.1 | \|Severe acute respiratory syndrome coro | ···················· | ···················· | ···················· | ···················· | ···················· | ···················· |
| 20724 | MT929027.1 | \|Severe acute respiratory syndrome coro | ···················· | ···················· | ···················· | ···················· | ···················· | ···················· |
| 20725 | MT929028.1 | \|Severe acute respiratory syndrome coro | ···················· | ···················· | ···················· | ···················· | ···················· | ···················· |
| 20726 | MT929029.1 | \|Severe acute respiratory syndrome coro | ···················· | ···················· | ···················· | ···················· | ···················· | ···················· |
| 20727 | MT929030.1 | \|Severe acute respiratory syndrome coro | ···················· | ·A·················· | ···················· | ···················· | ···················· | ···················· |
| 20728 | MT929031.1 | \|Severe acute respiratory syndrome coro | ···················· | ···················· | ···················· | ···················· | ···················· | ···················· |
| 20729 | MT929032.1 | \|Severe acute respiratory syndrome coro | ···················· | ·A·················· | ···················· | ···················· | ···················· | ···················· |

| | | | | | | | | |
|---|---|---|---|---|---|---|---|---|
| 20730 | MT929033.1 | \|Severe acute respiratory syndrome coro | ···················· | ···················· | ···················· | ···················· | ···················· | ···················· |
| 20731 | MT929035.1 | \|Severe acute respiratory syndrome coro | ···················· | ···················· | ···················· | ···················· | ···················· | ···················· |
| 20732 | MT929036.1 | \|Severe acute respiratory syndrome coro | ···················· | ·A·················· | ···················· | ···················· | ···················· | ···················· |
| 20733 | MT929037.1 | \|Severe acute respiratory syndrome coro | ···················· | ···················· | ···················· | ···················· | ···················· | ···················· |
| 20734 | MT929038.1 | \|Severe acute respiratory syndrome coro | ···················· | ···················· | ···················· | ···················· | ···················· | ···················· |
| 20735 | MT929039.1 | \|Severe acute respiratory syndrome coro | ···················· | ·············A······ | ···················· | ···················· | ···················· | ···················· |
| 20736 | MT929040.1 | \|Severe acute respiratory syndrome coro | ···················· | ···················· | ···················· | ···················· | ···················· | ···················· |
| 20737 | MT929041.1 | \|Severe acute respiratory syndrome coro | ···················· | ···················· | ···················· | ···················· | ···················· | ···················· |
| 20738 | MT929042.1 | \|Severe acute respiratory syndrome coro | ···················· | ···················· | ···················· | ···················· | ···················· | ···················· |
| 20739 | MT929043.1 | \|Severe acute respiratory syndrome coro | ···················· | ···················· | ···················· | ···················· | ···················· | ···················· |
| 20740 | MT929044.1 | \|Severe acute respiratory syndrome coro | ···················· | ···················· | ···················· | ···················· | ···················· | ···················· |
| 20741 | MT929045.1 | \|Severe acute respiratory syndrome coro | ···················· | ···················· | ···················· | ···················· | ···················· | ···················· |
| 20742 | MT929047.1 | \|Severe acute respiratory syndrome coro | ···················· | ···················· | ···················· | ···················· | ···················· | ···················· |
| 20743 | MT929049.1 | \|Severe acute respiratory syndrome coro | ···················· | ···················· | ···················· | ···················· | ···················· | ···················· |
| 20744 | MT929050.1 | \|Severe acute respiratory syndrome coro | ···················· | ···················· | ···················· | ···················· | ···················· | ···················· |
| 20745 | MT929051.1 | \|Severe acute respiratory syndrome coro | ···················· | ·············A······ | ···················· | ···················· | ···················· | ···················· |
| 20746 | MT929052.1 | \|Severe acute respiratory syndrome coro | ···················· | ·············A······ | ···················· | ···················· | ···················· | ···················· |
| 20747 | MT929053.1 | \|Severe acute respiratory syndrome coro | ···················· | ···················· | ···················· | ···················· | ···················· | ···················· |
| 20748 | MT929054.1 | \|Severe acute respiratory syndrome coro | ···················· | ···················· | ···················· | ···················· | ···················· | ···················· |
| 20749 | MT929055.1 | \|Severe acute respiratory syndrome coro | ···················· | ··C················· | ···················· | ···················· | ···················· | ···················· |
| 20750 | MT929056.1 | \|Severe acute respiratory syndrome coro | ···················· | ···················· | ···················· | ···················· | ···················· | ···················· |
| 20751 | MT929057.1 | \|Severe acute respiratory syndrome coro | ···················· | ···················· | ···················· | ···················· | ···················· | ···················· |
| 20752 | MT929058.1 | \|Severe acute respiratory syndrome coro | ···················· | ···················· | ···················· | ···················· | ···················· | ···················· |
| 20753 | MT929059.1 | \|Severe acute respiratory syndrome coro | ···················· | ···················· | ···················· | ···················· | ···················· | ···················· |
| 20754 | MT929060.1 | \|Severe acute respiratory syndrome coro | ···················· | ···················· | ···················· | ···················· | ···················· | ···················· |
| 20755 | MT929061.1 | \|Severe acute respiratory syndrome coro | ···················· | ···················· | ···················· | ···················· | ···················· | ···················· |
| 20756 | MT929062.1 | \|Severe acute respiratory syndrome coro | ···················· | ···················· | ···················· | ···················· | ···················· | ···················· |
| 20757 | MT929063.1 | \|Severe acute respiratory syndrome coro | ···················· | ···················· | ···················· | ···················· | ···················· | ···················· |
| 20758 | MT929064.1 | \|Severe acute respiratory syndrome coro | ···················· | ···················· | ···················· | ···················· | ···················· | ···················· |
| 20759 | MT929065.1 | \|Severe acute respiratory syndrome coro | ···················· | ···················· | ···················· | ···················· | ···················· | ···················· |
| 20760 | MT929066.1 | \|Severe acute respiratory syndrome coro | ···················· | ···················· | ···················· | ···················· | ···················· | ···················· |
| 20761 | MT929067.1 | \|Severe acute respiratory syndrome coro | ···················· | ···················· | ···················· | ···················· | ···················· | ···················· |
| 20762 | MT929069.1 | \|Severe acute respiratory syndrome coro | ···················· | ·A·················· | ···················· | ···················· | ···················· | ···················· |
| 20763 | MT929070.1 | \|Severe acute respiratory syndrome coro | ···················· | ···················· | ···················· | ···················· | ···················· | ···················· |
| 20764 | MT929071.1 | \|Severe acute respiratory syndrome coro | ···················· | ·A·················· | ···················· | ···················· | ···················· | ···················· |
| 20765 | MT929072.1 | \|Severe acute respiratory syndrome coro | ···················· | ···················· | ···················· | ···················· | ···················· | ···················· |
| 20766 | MT929073.1 | \|Severe acute respiratory syndrome coro | ···················· | ···················· | ···················· | ···················· | ···················· | ···················· |
| 20767 | MT929074.1 | \|Severe acute respiratory syndrome coro | ···················· | ···················· | ···················· | ···················· | ···················· | ···················· |
| 20768 | MT929075.1 | \|Severe acute respiratory syndrome coro | ···················· | ···················· | ···················· | ···················· | ···················· | ···················· |
| 20769 | MT929076.1 | \|Severe acute respiratory syndrome coro | ···················· | ···················· | ···················· | ···················· | ···················· | ···················· |
| 20770 | MT929077.1 | \|Severe acute respiratory syndrome coro | ···················· | ···················· | ···················· | ···················· | ···················· | ···················· |
| 20771 | MT929078.1 | \|Severe acute respiratory syndrome coro | ···················· | ···················· | ···················· | ···················· | ···················· | ···················· |
| 20772 | MT929079.1 | \|Severe acute respiratory syndrome coro | ···················· | ···················· | ···················· | ···················· | ···················· | ···················· |
| 20773 | MT929080.1 | \|Severe acute respiratory syndrome coro | ···················· | ···················· | ···················· | ···················· | ···················· | ···················· |
| 20774 | MT929081.1 | \|Severe acute respiratory syndrome coro | ···················· | ···················· | ···················· | ···················· | ···················· | ···················· |
| 20775 | MT929082.1 | \|Severe acute respiratory syndrome coro | ···················· | ···················· | ···················· | ···················· | ···················· | ···················· |
| 20776 | MT929083.1 | \|Severe acute respiratory syndrome coro | ···················· | ····A··············· | ···················· | ···················· | ···················· | ···················· |
| 20777 | MT929085.1 | \|Severe acute respiratory syndrome coro | ···················· | ···················· | ···················· | ···················· | ···················· | ···················· |
| 20778 | MT929086.1 | \|Severe acute respiratory syndrome coro | ···················· | ···················· | ···················· | ···················· | ···················· | ···················· |
| 20779 | MT929087.1 | \|Severe acute respiratory syndrome coro | ···················· | ···················· | ···················· | ···················· | ···················· | ···················· |
| 20780 | MT929088.1 | \|Severe acute respiratory syndrome coro | ···················· | ···················· | ···················· | ···················· | ···················· | ···················· |
| 20781 | MT929089.1 | \|Severe acute respiratory syndrome coro | ···················· | ···················· | ···················· | ···················· | ···················· | ···················· |
| 20782 | MT929090.1 | \|Severe acute respiratory syndrome coro | ···················· | ···················· | ···················· | ···················· | ···················· | ···················· |
| 20783 | MT929091.1 | \|Severe acute respiratory syndrome coro | ···················· | ···················· | ···················· | ···················· | ···················· | ···················· |
| 20784 | MT929092.1 | \|Severe acute respiratory syndrome coro | ···················· | ···················· | ···················· | ···················· | ···················· | ···················· |
| 20785 | MT929093.1 | \|Severe acute respiratory syndrome coro | ···················· | ···················· | ···················· | ···················· | ···················· | ···················· |
| 20786 | MT929094.1 | \|Severe acute respiratory syndrome coro | ···················· | ···················· | ···················· | ···················· | ···················· | ···················· |
| 20787 | MT929095.1 | \|Severe acute respiratory syndrome coro | ···················· | ···················· | ···················· | ···················· | ···················· | ···················· |
| 20788 | MT929096.1 | \|Severe acute respiratory syndrome coro | ···················· | ···················· | ···················· | ···················· | ···················· | ···················· |
| 20789 | MT929097.1 | \|Severe acute respiratory syndrome coro | ···················· | ···················· | ···················· | ···················· | ···················· | ···················· |
| 20790 | MT929098.1 | \|Severe acute respiratory syndrome coro | ···················· | ···················· | ···················· | ···················· | ···················· | ···················· |
| 20791 | MT929099.1 | \|Severe acute respiratory syndrome coro | ···················· | ···················· | ···················· | ···················· | ···················· | ···················· |
| 20792 | MT929100.1 | \|Severe acute respiratory syndrome coro | ···················· | ···················· | ···················· | ···················· | ···················· | ···················· |
| 20793 | MT929101.1 | \|Severe acute respiratory syndrome coro | ···················· | ···················· | ···················· | ···················· | ···················· | ···················· |
| 20794 | MT929104.1 | \|Severe acute respiratory syndrome coro | ···················· | ···················· | ···················· | ···················· | ···················· | ···················· |
| 20795 | MT929105.1 | \|Severe acute respiratory syndrome coro | ···················· | ···················· | ···················· | ···················· | ···················· | ···················· |
| 20796 | MT929106.1 | \|Severe acute respiratory syndrome coro | ···················· | ···················· | ···················· | ···················· | ···················· | ···················· |
| 20797 | MT929107.1 | \|Severe acute respiratory syndrome coro | ···················· | ···················· | ···················· | ···················· | ···················· | ···················· |
| 20798 | MT929110.1 | \|Severe acute respiratory syndrome coro | ···················· | ···················· | ···················· | ···················· | ···················· | ···················· |
| 20799 | MT929111.1 | \|Severe acute respiratory syndrome coro | ···················· | ···················· | ···················· | ···················· | ···················· | ···················· |
| 20800 | MT929112.1 | \|Severe acute respiratory syndrome coro | ···················· | ···················· | ···················· | ···················· | ···················· | ···················· |
| 20801 | MT929115.1 | \|Severe acute respiratory syndrome coro | ···················· | ···················· | ···················· | ···················· | ···················· | ···················· |
| 20802 | MT929116.1 | \|Severe acute respiratory syndrome coro | ···················· | ···················· | ···················· | ···················· | ···················· | ···················· |
| 20803 | MT929118.1 | \|Severe acute respiratory syndrome coro | ···················· | ···················· | ···················· | ···················· | ···················· | ···················· |
| 20804 | MT929119.1 | \|Severe acute respiratory syndrome coro | ···················· | ···················· | ···················· | ···················· | ···················· | ···················· |
| 20805 | MT929123.1 | \|Severe acute respiratory syndrome coro | ···················· | ···················· | ···················· | ···················· | ···················· | ···················· |
| 20806 | MT929124.1 | \|Severe acute respiratory syndrome coro | ···················· | ·············A······ | ···················· | ···················· | ···················· | ···················· |
| 20808 | MT929130.1 | \|Severe acute respiratory syndrome coro | ···················· | ···················· | ···················· | ···················· | ···················· | ···················· |

| | | | | | | | | |
|---|---|---|---|---|---|---|---|---|
| 20809 | MT929134.1 | \|Severe acute respiratory syndrome coro | .................... | .................... | .................... | .................... | .................... | .................... |
| 20810 | MT929135.1 | \|Severe acute respiratory syndrome coro | .................... | .................... | .................... | .................... | .................... | .................... |
| 20811 | MT929148.1 | \|Severe acute respiratory syndrome coro | .................... | .A.................. | .................... | .................... | .................... | .................... |
| 20812 | MT929150.1 | \|Severe acute respiratory syndrome coro | .................... | .................... | .................... | .................... | .................... | .................... |
| 20813 | MT929153.1 | \|Severe acute respiratory syndrome coro | .................... | .................... | .................... | .................... | .................... | .................... |
| 20814 | MT929154.1 | \|Severe acute respiratory syndrome coro | .................... | .................... | .................... | .................... | .................... | .................... |
| 20815 | MT929156.1 | \|Severe acute respiratory syndrome coro | .................... | .................... | .................... | .................... | .................... | .................... |
| 20816 | MT929157.1 | \|Severe acute respiratory syndrome coro | .................... | .................... | .................... | .................... | .................... | .................... |
| 20817 | MT929158.1 | \|Severe acute respiratory syndrome coro | .................... | .................... | .................... | .................... | .................... | .................... |
| 20818 | MT929159.1 | \|Severe acute respiratory syndrome coro | .................... | .................... | .................... | .................... | .................... | .................... |
| 20819 | MT925980.1 | \|Severe acute respiratory syndrome coro | .................... | .................... | .................... | .................... | .................... | .................... |
| 20820 | MT925981.1 | \|Severe acute respiratory syndrome coro | .................... | .................... | .................... | .................... | .................... | .................... |
| 20821 | MT925982.1 | \|Severe acute respiratory syndrome coro | .................... | .................... | .................... | .................... | .................... | .................... |
| 20822 | MT925983.1 | \|Severe acute respiratory syndrome coro | .................... | .................... | .................... | .................... | .................... | .................... |
| 20823 | MT925984.1 | \|Severe acute respiratory syndrome coro | .................... | .................... | .................... | .................... | .................... | .................... |
| 20824 | MT925985.1 | \|Severe acute respiratory syndrome coro | .................... | .................... | .................... | .................... | .................... | .................... |
| 20825 | MT925986.1 | \|Severe acute respiratory syndrome coro | .................... | .................... | .................... | .................... | .................... | .................... |
| 20826 | MT925987.1 | \|Severe acute respiratory syndrome coro | .................... | .................... | .................... | .................... | .................... | .................... |
| 20827 | MT925988.1 | \|Severe acute respiratory syndrome coro | .................... | .................... | .................... | .................... | .................... | .................... |
| 20828 | MT925989.1 | \|Severe acute respiratory syndrome coro | .................... | .................... | .................... | .................... | .................... | .................... |
| 20829 | MT925991.1 | \|Severe acute respiratory syndrome coro | .................... | .................... | .................... | .................... | .................... | .................... |
| 20830 | MT925992.1 | \|Severe acute respiratory syndrome coro | .................... | .................... | .................... | .................... | .................... | .................... |
| 20831 | MT925993.1 | \|Severe acute respiratory syndrome coro | .................... | .................... | .................... | .................... | .................... | .................... |
| 20832 | MT925994.1 | \|Severe acute respiratory syndrome coro | .................... | .................... | .................... | .................... | .................... | .................... |
| 20833 | MT925995.1 | \|Severe acute respiratory syndrome coro | .................... | .................... | .................... | .................... | .................... | .................... |
| 20834 | MT925996.1 | \|Severe acute respiratory syndrome coro | .................... | .................... | .................... | .................... | .................... | .................... |
| 20835 | MT925997.1 | \|Severe acute respiratory syndrome coro | .................... | .................... | .................... | .................... | .................... | .................... |
| 20836 | LR877883.1 | \|Severe acute respiratory syndrome coro | .................... | .................... | .................... | .................... | .................... | .................... |
| 20837 | LR877886.1 | \|Severe acute respiratory syndrome coro | .................... | .................... | .................... | .................... | .................... | .................... |
| 20838 | LR877887.1 | \|Severe acute respiratory syndrome coro | .................... | .................... | .................... | .................... | .................... | .................... |
| 20839 | LR877888.1 | \|Severe acute respiratory syndrome coro | .................... | .................... | .................... | .................... | .................... | .................... |
| 20840 | LR877889.1 | \|Severe acute respiratory syndrome coro | .................... | .................... | .................... | .................... | .................... | .................... |
| 20841 | LR877890.1 | \|Severe acute respiratory syndrome coro | .................... | .................... | .................... | .................... | .................... | .................... |
| 20842 | LR877891.1 | \|Severe acute respiratory syndrome coro | .................... | .................... | .................... | .................... | .................... | .................... |
| 20843 | LR877892.1 | \|Severe acute respiratory syndrome coro | .................... | .................... | .................... | .................... | .................... | .................... |
| 20844 | LR877893.1 | \|Severe acute respiratory syndrome coro | .................... | .................... | .................... | .................... | .................... | .................... |
| 20845 | LR877894.1 | \|Severe acute respiratory syndrome coro | .................... | .................... | .................... | .................... | .................... | .................... |
| 20846 | LR877895.1 | \|Severe acute respiratory syndrome coro | .................... | .................... | .................... | .................... | .................... | .................... |
| 20847 | LR877896.1 | \|Severe acute respiratory syndrome coro | .................... | .................... | .................... | .................... | .................... | .................... |
| 20848 | LR877897.1 | \|Severe acute respiratory syndrome coro | .................... | .................... | .................... | .................... | .................... | .................... |
| 20849 | LR877898.1 | \|Severe acute respiratory syndrome coro | .................... | .................... | .................... | .................... | .................... | .................... |
| 20850 | LR877899.1 | \|Severe acute respiratory syndrome coro | .................... | .................... | .................... | .................... | .................... | .................... |
| 20851 | LR877900.1 | \|Severe acute respiratory syndrome coro | .................... | .................... | .................... | .................... | .................... | .................... |
| 20852 | LR877901.1 | \|Severe acute respiratory syndrome coro | .................... | .................... | .................... | .................... | .................... | .................... |
| 20853 | LR877902.1 | \|Severe acute respiratory syndrome coro | .................... | .................... | .................... | .................... | .................... | .................... |
| 20854 | LR877903.1 | \|Severe acute respiratory syndrome coro | .................... | .................... | .................... | .................... | .................... | .................... |
| 20855 | LR877904.1 | \|Severe acute respiratory syndrome coro | .................... | .................... | .................... | .................... | .................... | .................... |
| 20856 | LR877905.1 | \|Severe acute respiratory syndrome coro | .................... | .................... | .................... | .................... | .................... | .................... |
| 20857 | LR877906.1 | \|Severe acute respiratory syndrome coro | .................... | .................... | .................... | .................... | .................... | .................... |
| 20858 | LR877907.1 | \|Severe acute respiratory syndrome coro | .................... | .................... | .................... | .................... | .................... | .................... |
| 20859 | LR877908.1 | \|Severe acute respiratory syndrome coro | .................... | .................... | .................... | .................... | .................... | .................... |
| 20860 | LR877909.1 | \|Severe acute respiratory syndrome coro | .................... | .................... | .................... | .................... | .................... | .................... |
| 20861 | LR877910.1 | \|Severe acute respiratory syndrome coro | .................... | .................... | .................... | .................... | .................... | .................... |
| 20862 | LR877911.1 | \|Severe acute respiratory syndrome coro | .................... | .................... | .................... | .................... | .................... | .................... |
| 20863 | LR877912.1 | \|Severe acute respiratory syndrome coro | .................... | .................... | .................... | .................... | .................... | .................... |
| 20864 | LR877913.1 | \|Severe acute respiratory syndrome coro | .................... | .................... | .................... | .................... | .................... | .................... |
| 20865 | LR877914.1 | \|Severe acute respiratory syndrome coro | .................... | .................... | .................... | .................... | .................... | .................... |
| 20866 | LR877915.1 | \|Severe acute respiratory syndrome coro | .................... | .................... | .................... | .................... | .................... | .................... |
| 20867 | LR877916.1 | \|Severe acute respiratory syndrome coro | .................... | .................... | .................... | .................... | .................... | .................... |
| 20868 | LR877917.1 | \|Severe acute respiratory syndrome coro | .................... | .................... | .................... | .................... | .................... | .................... |
| 20869 | LR877918.1 | \|Severe acute respiratory syndrome coro | .................... | .................... | .................... | .................... | .................... | .................... |
| 20870 | LR877919.1 | \|Severe acute respiratory syndrome coro | .................... | .................... | .................... | .................... | .................... | .................... |
| 20871 | LR877920.1 | \|Severe acute respiratory syndrome coro | .................... | .................... | .................... | .................... | .................... | .................... |
| 20872 | LR877921.1 | \|Severe acute respiratory syndrome coro | .................... | .................... | .................... | .................... | .................... | .................... |
| 20873 | LR877922.1 | \|Severe acute respiratory syndrome coro | .................... | .................... | .................... | .................... | .................... | .................... |
| 20874 | LR877923.1 | \|Severe acute respiratory syndrome coro | .................... | .................... | .................... | .................... | .................... | .................... |
| 20875 | LR877924.1 | \|Severe acute respiratory syndrome coro | .................... | .................... | .................... | .................... | .................... | .................... |
| 20876 | LR877925.1 | \|Severe acute respiratory syndrome coro | .................... | .................... | .................... | .................... | .................... | .................... |
| 20877 | LR877926.1 | \|Severe acute respiratory syndrome coro | .................... | .................... | .................... | .................... | .................... | .................... |
| 20878 | LR877927.1 | \|Severe acute respiratory syndrome coro | .................... | .................... | .................... | .................... | .................... | .................... |
| 20879 | LR877928.1 | \|Severe acute respiratory syndrome coro | .................... | .................... | .................... | .................... | .................... | .................... |
| 20880 | LR877929.1 | \|Severe acute respiratory syndrome coro | .................... | .................... | .................... | .................... | .................... | .................... |
| 20881 | LR877930.1 | \|Severe acute respiratory syndrome coro | .................... | .A.................. | .................... | .................... | .................... | .................... |
| 20882 | LR877931.1 | \|Severe acute respiratory syndrome coro | .................... | .................... | .................... | .................... | .................... | .................... |
| 20883 | LR877932.1 | \|Severe acute respiratory syndrome coro | .................... | .................... | .................... | .................... | .................... | .................... |
| 20884 | LR877933.1 | \|Severe acute respiratory syndrome coro | .................... | .................... | .................... | .................... | .................... | .................... |
| 20885 | LR877934.1 | \|Severe acute respiratory syndrome coro | .................... | .................... | .................... | .................... | .................... | .................... |
| 20886 | LR877935.1 | \|Severe acute respiratory syndrome coro | .................... | .................... | .................... | .................... | .................... | .................... |

20887 LR877936.1 |Severe acute respiratory syndrome coro ···················· ···················· ····················· ···················· ···················· ····················
20888 LR877937.1 |Severe acute respiratory syndrome coro ···················· ···················· ····················· ···················· ···················· ····················
20889 LR877938.1 |Severe acute respiratory syndrome coro ···················· ···················· ····················· ···················· ···················· ····················
20890 LR877939.1 |Severe acute respiratory syndrome coro ···················· ···················· ····················· ···················· ···················· ····················
20891 LR877940.1 |Severe acute respiratory syndrome coro ···················· ···················· ····················· ···················· ···················· ····················
20892 LR877941.1 |Severe acute respiratory syndrome coro ···················· ···················· ····················· ···················· ···················· ····················
20893 LR877942.1 |Severe acute respiratory syndrome coro ···················· ···················· ····················· ···················· ···················· ····················
20894 LR877943.1 |Severe acute respiratory syndrome coro ···················· ···················· ····················· ···················· ···················· ····················
20895 LR877944.1 |Severe acute respiratory syndrome coro ···················· ···················· ····················· ···················· ···················· ····················
20896 LR877945.1 |Severe acute respiratory syndrome coro ···················· ···················· ····················· ···················· ···················· ····················
20897 LR877946.1 |Severe acute respiratory syndrome coro ···················· ···················· ····················· ···················· ···················· ····················
20898 LR877947.1 |Severe acute respiratory syndrome coro ···················· ···················· ····················· ···················· ···················· ····················
20899 LR877948.1 |Severe acute respiratory syndrome coro ···················· ···················· ····················· ···················· ···················· ····················
20900 LR877949.1 |Severe acute respiratory syndrome coro ···················· ···················· ····················· ···················· ···················· ····················
20901 LR877950.1 |Severe acute respiratory syndrome coro ···················· ···················· ····················· ···················· ···················· ····················
20902 LR877951.1 |Severe acute respiratory syndrome coro ···················· ···················· ····················· ···················· ···················· ····················
20903 LR877952.1 |Severe acute respiratory syndrome coro ···················· ···················· ····················· ···················· ···················· ····················
20904 LR877953.1 |Severe acute respiratory syndrome coro ···················· ···················· ····················· ···················· ···················· ····················
20905 LR877954.1 |Severe acute respiratory syndrome coro ···················· ···················· ····················· ···················· ···················· ····················
20906 LR877955.1 |Severe acute respiratory syndrome coro ···················· ···················· ····················· ···················· ···················· ····················
20907 LR877956.1 |Severe acute respiratory syndrome coro ···················· ···················· ····················· ···················· ···················· ····················
20908 LR877957.1 |Severe acute respiratory syndrome coro ···················· ···················· ····················· ···················· ···················· ····················
20909 LR877958.1 |Severe acute respiratory syndrome coro ···················· ···················· ····················· ···················· ···················· ····················
20910 LR877959.1 |Severe acute respiratory syndrome coro ···················· ···················· ····················· ···················· ···················· ····················
20911 LR877960.1 |Severe acute respiratory syndrome coro ···················· ···················· ····················· ···················· ···················· ····················
20912 LR877961.1 |Severe acute respiratory syndrome coro ···················· ···················· ····················· ···················· ···················· ····················
20913 LR877962.1 |Severe acute respiratory syndrome coro ···················· ···················· ····················· ···················· ···················· ····················
20914 LR877963.1 |Severe acute respiratory syndrome coro ···················· ···················· ····················· ···················· ···················· ····················
20915 LR877964.1 |Severe acute respiratory syndrome coro ···················· ···················· ····················· ···················· ···················· ····················
20917 LR877966.1 |Severe acute respiratory syndrome coro ···················· ···················· ····················· ···················· ···················· ····················
20919 LR877968.1 |Severe acute respiratory syndrome coro ···················· ···················· ····················· ···················· ···················· ····················
20920 LR877969.1 |Severe acute respiratory syndrome coro ···················· ···················· ····················· ···················· ···················· ····················
20921 LR877970.1 |Severe acute respiratory syndrome coro ···················· ···················· ····················· ···················· ···················· ····················
20922 LR877971.1 |Severe acute respiratory syndrome coro ···················· ···················· ····················· ···················· ···················· ····················
20923 LR877972.1 |Severe acute respiratory syndrome coro ···················· ···················· ····················· ···················· ···················· ····················
20924 LR877973.1 |Severe acute respiratory syndrome coro ···················· ···················· ····················· ···················· ···················· ····················
20925 LR877974.1 |Severe acute respiratory syndrome coro ···················· ···················· ····················· ···················· ···················· ····················
20926 LR877975.1 |Severe acute respiratory syndrome coro ···················· ···················· ····················· ···················· ···················· ····················
20927 LR877976.1 |Severe acute respiratory syndrome coro ···················· ···················· ····················· ···················· ···················· ····················
20928 LR877977.1 |Severe acute respiratory syndrome coro ···················· ···················· ····················· ···················· ···················· ····················
20929 LR877978.1 |Severe acute respiratory syndrome coro ···················· ···················· ····················· ···················· ···················· ····················
20930 LR877979.1 |Severe acute respiratory syndrome coro ···················· ···················· ····················· ···················· ···················· ····················
20933 LR877982.1 |Severe acute respiratory syndrome coro ···················· ···················· ····················· ···················· ···················· ····················
20935 LR877984.1 |Severe acute respiratory syndrome coro ···················· ···················· ····················· ···················· ···················· ····················
20936 LR877985.1 |Severe acute respiratory syndrome coro ···················· ···················· ····················· ···················· ···················· ····················
20937 LR877986.1 |Severe acute respiratory syndrome coro ···················· ···················· ····················· ···················· ···················· ····················
20938 LR877987.1 |Severe acute respiratory syndrome coro ···················· ···················· ····················· ···················· ···················· ····················
20939 LR877988.1 |Severe acute respiratory syndrome coro ···················· ···················· ····················· ···················· ···················· ····················
20940 LR877989.1 |Severe acute respiratory syndrome coro ···················· ···················· ····················· ···················· ···················· ····················
20941 LR877990.1 |Severe acute respiratory syndrome coro ···················· ···················· ····················· ···················· ···················· ····················
20942 LR877991.1 |Severe acute respiratory syndrome coro ···················· ···················· ····················· ···················· ···················· ····················
20943 LR877992.1 |Severe acute respiratory syndrome coro ···················· ···················· ····················· ···················· ···················· ····················
20944 LR877993.1 |Severe acute respiratory syndrome coro ···················· ···················· ····················· ···················· ···················· ····················
20945 LR877994.1 |Severe acute respiratory syndrome coro ···················· ···················· ····················· ···················· ···················· ····················
20946 LR877995.1 |Severe acute respiratory syndrome coro ···················· ···················· ····················· ···················· ···················· ····················
20947 LR877996.1 |Severe acute respiratory syndrome coro ···················· ···················· ····················· ···················· ···················· ····················
20948 LR877997.1 |Severe acute respiratory syndrome coro ···················· ···················· ····················· ···················· ···················· ····················
20950 LR877999.1 |Severe acute respiratory syndrome coro ···················· ···················· ····················· ···················· ···················· ····················
20951 LR878000.1 |Severe acute respiratory syndrome coro ···················· ···················· ····················· ···················· ···················· ····················
20952 LR878001.1 |Severe acute respiratory syndrome coro ···················· ···················· ····················· ···················· ···················· ····················
20953 LR878002.1 |Severe acute respiratory syndrome coro ···················· ···················· ····················· ···················· ···················· ····················
20954 LR878003.1 |Severe acute respiratory syndrome coro ···················· ·A·················· ····················· ···················· ···················· ····················
20955 LR878004.1 |Severe acute respiratory syndrome coro ···················· ···················· ····················· ···················· ···················· ····················
20956 LR878005.1 |Severe acute respiratory syndrome coro ···················· ···················· ····················· ···················· ···················· ····················
20957 LR878006.1 |Severe acute respiratory syndrome coro ···················· ···················· ····················· ···················· ···················· ····················
20958 LR878007.1 |Severe acute respiratory syndrome coro ···················· ···················· ····················· ···················· ···················· ····················
20959 LR878008.1 |Severe acute respiratory syndrome coro ···················· ···················· ····················· ···················· ···················· ····················
20960 LR878009.1 |Severe acute respiratory syndrome coro ···················· ···················· ····················· ···················· ···················· ····················
20961 LR878010.1 |Severe acute respiratory syndrome coro ···················· ···················· ····················· ···················· ···················· ····················
20962 LR878011.1 |Severe acute respiratory syndrome coro ···················· ···················· ····················· ···················· ···················· ····················
20963 LR878012.1 |Severe acute respiratory syndrome coro ···················· ···················· ····················· ···················· ···················· ····················
20964 LR878013.1 |Severe acute respiratory syndrome coro ···················· ···················· ····················· ···················· ···················· ····················
20965 LR878014.1 |Severe acute respiratory syndrome coro ···················· ···················· ····················· ···················· ···················· ····················
20966 LR878015.1 |Severe acute respiratory syndrome coro ···················· ···················· ····················· ···················· ···················· ····················
20967 LR878016.1 |Severe acute respiratory syndrome coro ···················· ···················· ····················· ···················· ···················· ····················
20968 LR878017.1 |Severe acute respiratory syndrome coro ···················· ···················· ····················· ···················· ···················· ····················
20969 LR878018.1 |Severe acute respiratory syndrome coro ···················· ···················· ····················· ···················· ···················· ····················
20970 LR878019.1 |Severe acute respiratory syndrome coro ···················· ···················· ····················· ···················· ···················· ····················

```
20971 LR878020.1 |Severe acute respiratory syndrome coro    ····················  ····················  ····················  ····················  ····················  ····················
20972 LR878021.1 |Severe acute respiratory syndrome coro    ····················  ····················  ····················  ····················  ····················  ····················
20973 LR878022.1 |Severe acute respiratory syndrome coro    ····················  ····················  ····················  ····················  ····················  ····················
20974 LR878023.1 |Severe acute respiratory syndrome coro    ····················  ····················  ····················  ····················  ····················  ····················
20975 LR878024.1 |Severe acute respiratory syndrome coro    ····················  ····················  ····················  ····················  ····················  ····················
20976 LR878025.1 |Severe acute respiratory syndrome coro    ····················  ····················  ····················  ····················  ····················  ····················
20977 LR878026.1 |Severe acute respiratory syndrome coro    ····················  ····················  ····················  ····················  ····················  ····················
20978 LR878027.1 |Severe acute respiratory syndrome coro    ····················  ····················  ····················  ····················  ····················  ····················
20979 LR878028.1 |Severe acute respiratory syndrome coro    ····················  ··········A·········  ····················  ····················  ····················  ····················
20980 LR878029.1 |Severe acute respiratory syndrome coro    ····················  ····················  ····················  ····················  ····················  ····················
20981 LR878030.1 |Severe acute respiratory syndrome coro    ····················  ····················  ····················  ····················  ····················  ····················
20982 LR878031.1 |Severe acute respiratory syndrome coro    ····················  ··········A·········  ····················  ····················  ····················  ····················
20983 LR878032.1 |Severe acute respiratory syndrome coro    ····················  ····················  ····················  ····················  ····················  ····················
20984 LR878033.1 |Severe acute respiratory syndrome coro    ····················  ····················  ····················  ····················  ····················  ····················
20985 LR878034.1 |Severe acute respiratory syndrome coro    ····················  ····················  ····················  ····················  ····················  ····················
20986 LR878035.1 |Severe acute respiratory syndrome coro    ····················  ····················  ····················  ····················  ····················  ····················
20987 LR878036.1 |Severe acute respiratory syndrome coro    ····················  ····················  ····················  ····················  ····················  ····················
20988 LR878037.1 |Severe acute respiratory syndrome coro    ····················  ····················  ····················  ····················  ····················  ····················
20989 LR878038.1 |Severe acute respiratory syndrome coro    ····················  ····················  ····················  ····················  ····················  ····················
20990 LR878039.1 |Severe acute respiratory syndrome coro    ····················  ····················  ····················  ····················  ····················  ····················
20991 LR878040.1 |Severe acute respiratory syndrome coro    ····················  ····················  ····················  ····················  ····················  ····················
20992 LR878041.1 |Severe acute respiratory syndrome coro    ····················  ····················  ····················  ····················  ····················  ····················
20993 LR878042.1 |Severe acute respiratory syndrome coro    ····················  ····················  ····················  ····················  ····················  ····················
20994 LR878043.1 |Severe acute respiratory syndrome coro    ····················  ····················  ····················  ····················  ····················  ····················
20995 LR878044.1 |Severe acute respiratory syndrome coro    ····················  ····················  ····················  ····················  ····················  ····················
20996 LR878045.1 |Severe acute respiratory syndrome coro    ····················  ····················  ····················  ····················  ····················  ····················
20997 LR878046.1 |Severe acute respiratory syndrome coro    ····················  ····················  ····················  ····················  ····················  ····················
20998 LR878047.1 |Severe acute respiratory syndrome coro    ····················  ····················  ····················  ····················  ····················  ····················
20999 LR878048.1 |Severe acute respiratory syndrome coro    ····················  ····················  ····················  ····················  ····················  ····················
21000 LR878049.1 |Severe acute respiratory syndrome coro    ····················  ····················  ····················  ····················  ····················  ····················
21001 LR878050.1 |Severe acute respiratory syndrome coro    ····················  ··········A·········  ····················  ····················  ····················  ····················
21002 LR878051.1 |Severe acute respiratory syndrome coro    ····················  ····················  ····················  ····················  ····················  ····················
21003 LR878052.1 |Severe acute respiratory syndrome coro    ····················  ····················  ····················  ····················  ····················  ····················
21004 LR878053.1 |Severe acute respiratory syndrome coro    ····················  ····················  ····················  ····················  ····················  ····················
21005 LR878054.1 |Severe acute respiratory syndrome coro    ····················  ····················  ····················  ····················  ····················  ····················
21006 LR878055.1 |Severe acute respiratory syndrome coro    ····················  ····················  ····················  ····················  ····················  ····················
21007 LR878056.1 |Severe acute respiratory syndrome coro    ····················  ····················  ····················  ····················  ····················  ····················
21008 LR878057.1 |Severe acute respiratory syndrome coro    ····················  ····················  ····················  ····················  ····················  ····················
21011 LR878060.1 |Severe acute respiratory syndrome coro    ····················  ····················  ····················  ····················  ····················  ····················
21012 LR878061.1 |Severe acute respiratory syndrome coro    ····················  ····················  ····················  ····················  ····················  ····················
21013 LR878062.1 |Severe acute respiratory syndrome coro    ····················  ····················  ····················  ····················  ····················  ····················
21014 LR878063.1 |Severe acute respiratory syndrome coro    ····················  ····················  ····················  ····················  ····················  ····················
21015 LR878064.1 |Severe acute respiratory syndrome coro    ····················  ····················  ····················  ····················  ····················  ····················
21016 LR878065.1 |Severe acute respiratory syndrome coro    ····················  ····················  ····················  ····················  ····················  ····················
21017 LR878066.1 |Severe acute respiratory syndrome coro    ····················  ····················  ····················  ····················  ····················  ····················
21018 LR878067.1 |Severe acute respiratory syndrome coro    ····················  ····················  ····················  ····················  ····················  ····················
21019 LR878068.1 |Severe acute respiratory syndrome coro    ····················  ····················  ····················  ····················  ····················  ····················
21021 LR878070.1 |Severe acute respiratory syndrome coro    ····················  ····················  ····················  ····················  ····················  ····················
21022 LR878071.1 |Severe acute respiratory syndrome coro    ····················  ····················  ····················  ····················  ····················  ····················
21023 LR878072.1 |Severe acute respiratory syndrome coro    ····················  ····················  ····················  ····················  ····················  ····················
21024 LR878073.1 |Severe acute respiratory syndrome coro    ····················  ····················  ····················  ····················  ····················  ····················
21025 LR878074.1 |Severe acute respiratory syndrome coro    ····················  ····················  ····················  ····················  ····················  ····················
21026 LR878075.1 |Severe acute respiratory syndrome coro    ····················  ····················  ····················  ····················  ····················  ····················
21027 LR878076.1 |Severe acute respiratory syndrome coro    ····················  ····················  ····················  ····················  ····················  ····················
21028 LR878077.1 |Severe acute respiratory syndrome coro    ····················  ····················  ····················  ····················  ····················  ····················
21029 LR878078.1 |Severe acute respiratory syndrome coro    ····················  ····················  ····················  ····················  ····················  ····················
21030 LR878079.1 |Severe acute respiratory syndrome coro    ····················  ····················  ····················  ····················  ····················  ····················
21031 LR878080.1 |Severe acute respiratory syndrome coro    ····················  ····················  ····················  ····················  ····················  ····················
21032 LR878081.1 |Severe acute respiratory syndrome coro    ····················  ····················  ····················  ····················  ····················  ····················
21033 LR878082.1 |Severe acute respiratory syndrome coro    ····················  ····················  ····················  ····················  ····················  ····················
21034 LR878083.1 |Severe acute respiratory syndrome coro    ····················  ····················  ····················  ····················  ····················  ····················
21035 LR878084.1 |Severe acute respiratory syndrome coro    ····················  ····················  ····················  ····················  ····················  ····················
21036 LR878085.1 |Severe acute respiratory syndrome coro    ····················  ····················  ····················  ····················  ····················  ····················
21037 LR878086.1 |Severe acute respiratory syndrome coro    ····················  ····················  ····················  ····················  ····················  ····················
21038 LR878087.1 |Severe acute respiratory syndrome coro    ····················  ····················  ····················  ····················  ····················  ····················
21039 LR878088.1 |Severe acute respiratory syndrome coro    ····················  ····················  ····················  ····················  ····················  ····················
21040 LR878089.1 |Severe acute respiratory syndrome coro    ····················  ····················  ····················  ····················  ····················  ····················
21041 LR878090.1 |Severe acute respiratory syndrome coro    ····················  ····················  ····················  ····················  ····················  ····················
21042 LR878091.1 |Severe acute respiratory syndrome coro    ····················  ····················  ····················  ····················  ····················  ····················
21043 LR878092.1 |Severe acute respiratory syndrome coro    ····················  ····················  ····················  ····················  ····················  ····················
21044 LR878093.1 |Severe acute respiratory syndrome coro    ····················  ····················  ····················  ····················  ····················  ····················
21045 LR878094.1 |Severe acute respiratory syndrome coro    ····················  ····················  ····················  ····················  ····················  ····················
21046 LR878095.1 |Severe acute respiratory syndrome coro    ····················  ····················  ····················  ····················  ····················  ····················
21047 LR878096.1 |Severe acute respiratory syndrome coro    ····················  ····················  ····················  ····················  ····················  ····················
21048 LR878097.1 |Severe acute respiratory syndrome coro    ····················  ····················  ····················  ····················  ····················  ····················
21049 LR878098.1 |Severe acute respiratory syndrome coro    ····················  ····················  ····················  ····················  ····················  ····················
21050 LR878099.1 |Severe acute respiratory syndrome coro    ····················  ····················  ····················  ····················  ····················  ····················
21051 LR878100.1 |Severe acute respiratory syndrome coro    ····················  ····················  ····················  ····················  ····················  ····················
```

```
21052 LR878101.1 |Severe acute respiratory syndrome coro    ····················  ····················  ····················  ····················  ····················  ····················
21053 LR878102.1 |Severe acute respiratory syndrome coro    ····················  ····················  ····················  ····················  ····················  ····················
21054 LR878103.1 |Severe acute respiratory syndrome coro    ····················  ····················  ····················  ····················  ····················  ····················
21055 LR878104.1 |Severe acute respiratory syndrome coro    ····················  ····················  ····················  ····················  ····················  ····················
21056 LR878105.1 |Severe acute respiratory syndrome coro    ····················  ····················  ····················  ····················  ····················  ····················
21057 LR878106.1 |Severe acute respiratory syndrome coro    ····················  ····················  ····················  ····················  ····················  ····················
21058 LR878107.1 |Severe acute respiratory syndrome coro    ····················  ····················  ····················  ····················  ····················  ····················
21059 LR878108.1 |Severe acute respiratory syndrome coro    ····················  ····················  ····················  ····················  ····················  ····················
21060 LR878109.1 |Severe acute respiratory syndrome coro    ····················  ····················  ····················  ····················  ····················  ····················
21061 LR878110.1 |Severe acute respiratory syndrome coro    ····················  ····················  ····················  ····················  ····················  ····················
21062 LR878111.1 |Severe acute respiratory syndrome coro    ····················  ····················  ····················  ····················  ····················  ····················
21063 LR878112.1 |Severe acute respiratory syndrome coro    ····················  ····················  ····················  ····················  ····················  ····················
21064 LR878113.1 |Severe acute respiratory syndrome coro    ····················  ····················  ····················  ····················  ····················  ····················
21065 LR878114.1 |Severe acute respiratory syndrome coro    ····················  ····················  ····················  ····················  ····················  ····················
21066 LR878115.1 |Severe acute respiratory syndrome coro    ····················  ····················  ····················  ····················  ····················  ····················
21067 LR878116.1 |Severe acute respiratory syndrome coro    ····················  ····················  ····················  ····················  ····················  ····················
21068 LR878117.1 |Severe acute respiratory syndrome coro    ····················  ····················  ····················  ····················  ····················  ····················
21069 LR878118.1 |Severe acute respiratory syndrome coro    ····················  ····················  ····················  ····················  ····················  ····················
21070 LR878119.1 |Severe acute respiratory syndrome coro    ····················  ····················  ····················  ····················  ····················  ····················
21071 LR878120.1 |Severe acute respiratory syndrome coro    ····················  ····················  ····················  ····················  ····················  ····················
21072 LR878121.1 |Severe acute respiratory syndrome coro    ····················  ····················  ····················  ····················  ····················  ····················
21073 LR878122.1 |Severe acute respiratory syndrome coro    ····················  ····················  ····················  ····················  ····················  ····················
21074 LR878123.1 |Severe acute respiratory syndrome coro    ····················  ····················  ····················  ····················  ····················  ····················
21075 LR878124.1 |Severe acute respiratory syndrome coro    ····················  ····················  ····················  ····················  ····················  ····················
21076 LR878125.1 |Severe acute respiratory syndrome coro    ····················  ····················  ····················  ····················  ····················  ····················
21077 LR878126.1 |Severe acute respiratory syndrome coro    ····················  ····················  ····················  ····················  ····················  ····················
21078 LR878127.1 |Severe acute respiratory syndrome coro    ····················  ····················  ····················  ····················  ····················  ····················
21079 LR878128.1 |Severe acute respiratory syndrome coro    ····················  ····················  ····················  ····················  ····················  ····················
21081 LR878130.1 |Severe acute respiratory syndrome coro    ····················  ····················  ····················  ····················  ····················  ····················
21083 LR878132.1 |Severe acute respiratory syndrome coro    ····················  ····················  ····················  ····················  ····················  ····················
21084 LR878133.1 |Severe acute respiratory syndrome coro    ····················  ····················  ····················  ····················  ····················  ····················
21085 LR878134.1 |Severe acute respiratory syndrome coro    ····················  ····················  ····················  ····················  ····················  ····················
21086 LR878135.1 |Severe acute respiratory syndrome coro    ····················  ····················  ····················  ····················  ····················  ····················
21087 LR878136.1 |Severe acute respiratory syndrome coro    ····················  ····················  ····················  ····················  ····················  ····················
21088 LR878137.1 |Severe acute respiratory syndrome coro    ····················  ····················  ····················  ····················  ····················  ····················
21089 LR878138.1 |Severe acute respiratory syndrome coro    ····················  ····················  ····················  ····················  ····················  ····················
21090 LR878139.1 |Severe acute respiratory syndrome coro    ····················  ····················  ····················  ····················  ····················  ····················
21091 LR878140.1 |Severe acute respiratory syndrome coro    ····················  ····················  ····················  ····················  ····················  ····················
21092 LR878141.1 |Severe acute respiratory syndrome coro    ····················  ····················  ····················  ····················  ····················  ····················
21093 LR878142.1 |Severe acute respiratory syndrome coro    ····················  ····················  ····················  ····················  ····················  ····················
21094 LR878143.1 |Severe acute respiratory syndrome coro    ····················  ····················  ····················  ····················  ····················  ····················
21095 LR878144.1 |Severe acute respiratory syndrome coro    ····················  ····················  ····················  ····················  ····················  ····················
21096 LR878145.1 |Severe acute respiratory syndrome coro    ····················  ····················  ····················  ····················  ····················  ····················
21097 LR878146.1 |Severe acute respiratory syndrome coro    ····················  ····················  ····················  ····················  ····················  ····················
21098 LR878147.1 |Severe acute respiratory syndrome coro    ····················  ····················  ····················  ····················  ····················  ····················
21099 LR878148.1 |Severe acute respiratory syndrome coro    ····················  ····················  ····················  ····················  ····················  ····················
21100 LR878149.1 |Severe acute respiratory syndrome coro    ····················  ····················  ····················  ····················  ····················  ····················
21101 LR878150.1 |Severe acute respiratory syndrome coro    ····················  ····················  ····················  ····················  ····················  ····················
21102 LR878151.1 |Severe acute respiratory syndrome coro    ····················  ····················  ····················  ····················  ····················  ····················
21103 LR878152.1 |Severe acute respiratory syndrome coro    ····················  ····················  ····················  ····················  ····················  ····················
21104 LR878153.1 |Severe acute respiratory syndrome coro    ····················  ····················  ····················  ····················  ····················  ····················
21106 LR878155.1 |Severe acute respiratory syndrome coro    ····················  ····················  ····················  ····················  ····················  ····················
21107 LR878156.1 |Severe acute respiratory syndrome coro    ····················  ····················  ····················  ····················  ····················  ····················
21108 LR878157.1 |Severe acute respiratory syndrome coro    ····················  ····················  ····················  ····················  ····················  ····················
21109 LR878158.1 |Severe acute respiratory syndrome coro    ····················  ····················  ····················  ····················  ····················  ····················
21110 LR878159.1 |Severe acute respiratory syndrome coro    ····················  ····················  ····················  ····················  ····················  ····················
21111 LR878160.1 |Severe acute respiratory syndrome coro    ····················  ····················  ····················  ····················  ····················  ····················
21112 LR878161.1 |Severe acute respiratory syndrome coro    ····················  ····················  ····················  ····················  NN··················  ····················
21113 LR878162.1 |Severe acute respiratory syndrome coro    ····················  ····················  ····················  ············A·······  ····················  ····················
21114 LR878163.1 |Severe acute respiratory syndrome coro    ····················  ····················  ····················  ····················  ····················  ····················
21115 LR878164.1 |Severe acute respiratory syndrome coro    ····················  ····················  ····················  ····················  ····················  ····················
21116 LR878165.1 |Severe acute respiratory syndrome coro    ····················  ····················  ····················  ····················  ····················  ····················
21117 LR878166.1 |Severe acute respiratory syndrome coro    ····················  ····················  ····················  ····················  ····················  ····················
21118 LR878167.1 |Severe acute respiratory syndrome coro    ····················  ····················  ····················  ····················  ····················  ····················
21119 LR878168.1 |Severe acute respiratory syndrome coro    ····················  ····················  ····················  ····················  ····················  ····················
21120 LR878169.1 |Severe acute respiratory syndrome coro    ····················  ····················  ····················  ····················  ····················  ····················
21121 LR878170.1 |Severe acute respiratory syndrome coro    ····················  ····················  ····················  ····················  ····················  ····················
21122 LR878171.1 |Severe acute respiratory syndrome coro    ····················  ····················  ····················  ····················  ····················  ····················
21123 LR878172.1 |Severe acute respiratory syndrome coro    ····················  ····················  ····················  ····················  ····················  ····················
21124 LR878173.1 |Severe acute respiratory syndrome coro    ····················  ····················  ····················  ····················  ····················  ····················
21125 LR878174.1 |Severe acute respiratory syndrome coro    ····················  ····················  ····················  ····················  ····················  ····················
21126 LR878175.1 |Severe acute respiratory syndrome coro    ····················  ····················  ····················  ····················  ····················  ····················
21127 LR878176.1 |Severe acute respiratory syndrome coro    ····················  ····················  ····················  ····················  ····················  ····················
21128 LR878177.1 |Severe acute respiratory syndrome coro    ····················  ····················  ····················  ····················  ····················  ····················
21129 LR878178.1 |Severe acute respiratory syndrome coro    ·T··················  ····················  ····················  ····················  ····················  ····················
21130 LR878179.1 |Severe acute respiratory syndrome coro    ····················  ····················  ····················  ····················  ····················  ····················
21131 LR878180.1 |Severe acute respiratory syndrome coro    ····················  ····················  ····················  ····················  ····T···············  ····················
21132 LR878181.1 |Severe acute respiratory syndrome coro    ····················  ····················  ····················  ····················  ····················  ····················
```

21133 LR878182.1 |Severe acute respiratory syndrome coro ···················· ···················· ···················· ···················· ···················· ····················
21134 LR878183.1 |Severe acute respiratory syndrome coro ···················· ···················· ···················· ···················· ···T················ ····················
21135 LR878184.1 |Severe acute respiratory syndrome coro ···················· ···················· ···················· ···················· ···················· ····················
21136 LR878185.1 |Severe acute respiratory syndrome coro ···················· ···················· ···················· ···················· ···················· ····················
21137 LR878186.1 |Severe acute respiratory syndrome coro ···················· ···················· ···················· ···················· ···················· ····················
21138 LR878187.1 |Severe acute respiratory syndrome coro ···················· ···················· ···················· ···················· ···················· ····················
21139 LR878188.1 |Severe acute respiratory syndrome coro ···················· ···················· ···················· ···················· ···················· ····················
21140 LR878189.1 |Severe acute respiratory syndrome coro ···················· ···················· ···················· ···················· ···················· ····················
21141 LR878190.1 |Severe acute respiratory syndrome coro ···················· ···················· ···················· ···················· ···················· ····················
21142 LR878191.1 |Severe acute respiratory syndrome coro ···················· ···················· ···················· ···················· ···················· ····················
21143 LR878192.1 |Severe acute respiratory syndrome coro ···················· ···················· ···················· ···················· ···················· ····················
21144 LR878193.1 |Severe acute respiratory syndrome coro ·T·················· ···················· ···················· ···················· ···················· ····················
21145 LR878194.1 |Severe acute respiratory syndrome coro ···················· ···················· ···················· ···················· ···················· ····················
21146 LR878195.1 |Severe acute respiratory syndrome coro ···················· ···················· ···················· ···················· ···················· ····················
21147 LR878196.1 |Severe acute respiratory syndrome coro ···················· ···················· ···················· ···················· ···················· ····················
21148 LR878197.1 |Severe acute respiratory syndrome coro ···················· ···················· ···················· ···················· ···················· ····················
21149 LR878198.1 |Severe acute respiratory syndrome coro ···················· ···················· ···················· ···················· ···················· ····················
21150 LR878199.1 |Severe acute respiratory syndrome coro ···················· ···················· ···················· ···················· ···················· ····················
21151 LR878200.1 |Severe acute respiratory syndrome coro ···················· ···················· ···················· ···················· ···················· ····················
21152 LR878201.1 |Severe acute respiratory syndrome coro ···················· ···················· ···················· ···················· ···················· ····················
21153 LR878202.1 |Severe acute respiratory syndrome coro ···················· ···················· ···················· ···················· ···················· ····················
21154 LR878203.1 |Severe acute respiratory syndrome coro ···················· ···················· ···················· ···················· ···················· ····················
21155 LR878204.1 |Severe acute respiratory syndrome coro ···················· ···················· ···················· ···················· ···················· ····················
21156 LR878205.1 |Severe acute respiratory syndrome coro ···················· ···················· ···················· ···················· ···················· ····················
21157 LR878206.1 |Severe acute respiratory syndrome coro ···················· ···················· ···················· ···················· ···················· ····················
21158 LR878207.1 |Severe acute respiratory syndrome coro ···················· ···················· ···················· ···················· ···················· ····················
21159 LR878208.1 |Severe acute respiratory syndrome coro ···················· ···················· ···················· ···················· ···················· ····················
21160 LR878209.1 |Severe acute respiratory syndrome coro ···················· ···················· ···················· ···················· ···················· ····················
21161 LR878210.1 |Severe acute respiratory syndrome coro ···················· ···················· ···················· ···················· ···················· ····················
21162 LR878211.1 |Severe acute respiratory syndrome coro ···················· ···················· ···················· ···················· ···················· ····················
21163 LR878212.1 |Severe acute respiratory syndrome coro ···················· ···················· ···················· ···················· ···················· ····················
21164 LR878213.1 |Severe acute respiratory syndrome coro ···················· ···················· ···················· ···················· ···················· ····················
21165 LR878214.1 |Severe acute respiratory syndrome coro ···················· ···················· ···················· ···················· ···················· ····················
21166 LR878215.1 |Severe acute respiratory syndrome coro ···················· ···················· ···················· ···················· ···················· ····················
21167 LR878216.1 |Severe acute respiratory syndrome coro ···················· ···················· ···················· ···················· ···················· ····················
21168 LR878217.1 |Severe acute respiratory syndrome coro ···················· ···················· ···················· ···················· ···················· ····················
21169 LR878218.1 |Severe acute respiratory syndrome coro ···················· ···················· ···················· ···················· ···················· ····················
21170 LR878219.1 |Severe acute respiratory syndrome coro ···················· ···················· ···················· ···················· ···················· ····················
21171 LR878220.1 |Severe acute respiratory syndrome coro ···················· ···················· ···················· ···················· ···················· ····················
21172 LR878221.1 |Severe acute respiratory syndrome coro ···················· ···················· ···················· ···················· ···················· ····················
21173 LR878222.1 |Severe acute respiratory syndrome coro ···················· ···················· ···················· ···················· ···················· ····················
21174 LR878223.1 |Severe acute respiratory syndrome coro ···················· ···················· ···················· ···················· ···················· ····················
21175 LR878224.1 |Severe acute respiratory syndrome coro ···················· ···················· ···················· ···················· ···················· ····················
21176 LR878225.1 |Severe acute respiratory syndrome coro ···················· ···················· ···················· ···················· ···················· ····················
21177 LR878226.1 |Severe acute respiratory syndrome coro ···················· ···················· ···················· ···················· ···················· ····················
21178 LR878227.1 |Severe acute respiratory syndrome coro ···················· ···················· ···················· ···················· ···················· ····················
21180 LR878229.1 |Severe acute respiratory syndrome coro ···················· ···················· ···················· ···················· ···················· ····················
21181 LR878230.1 |Severe acute respiratory syndrome coro ···················· ···················· ···················· ···················· ···················· ····················
21185 LR878234.1 |Severe acute respiratory syndrome coro ···················· ···················· ···················· ···················· ···················· ····················
21187 LR878236.1 |Severe acute respiratory syndrome coro ···················· ···················· ···················· ···················· ···················· ····················
21189 LR878238.1 |Severe acute respiratory syndrome coro ···················· ···················· ···················· ···················· ···················· ····················
21192 LR878241.1 |Severe acute respiratory syndrome coro ···················· ···················· ···················· ···················· ···················· ····················
21194 LR878243.1 |Severe acute respiratory syndrome coro ···················· ···················· ···················· ···················· ···················· ····················
21195 LR878244.1 |Severe acute respiratory syndrome coro ···················· ···················· ···················· ···················· ···················· ····················
21196 LR878245.1 |Severe acute respiratory syndrome coro ···················· ···················· ···················· ···················· ···················· ····················
21198 LR878247.1 |Severe acute respiratory syndrome coro ···················· ···················· ···················· ···················· ···················· ····················
21199 LR878248.1 |Severe acute respiratory syndrome coro ···················· ···················· ···················· ···················· ···················· ····················
21200 LR878249.1 |Severe acute respiratory syndrome coro ···················· ···················· ···················· ···················· ···················· ····················
21201 LR878250.1 |Severe acute respiratory syndrome coro ···················· ···················· ···················· ···················· ···················· ····················
21202 LR878251.1 |Severe acute respiratory syndrome coro ···················· ···················· ···················· ···················· ···················· ····················
21203 LR878252.1 |Severe acute respiratory syndrome coro ···················· ···················· ···················· ···················· ···················· ····················
21204 LR878253.1 |Severe acute respiratory syndrome coro ···················· ···················· ···················· ···················· ···················· ····················
21205 LR878254.1 |Severe acute respiratory syndrome coro ···················· ···················· ···················· ···················· ···················· ····················
21206 LR878255.1 |Severe acute respiratory syndrome coro ···················· ···················· ···················· ···················· ···················· ····················
21207 LR878256.1 |Severe acute respiratory syndrome coro ···················· ···················· ···················· ···················· ···················· ····················
21209 LR878258.1 |Severe acute respiratory syndrome coro ···················· ···················· ···················· ···················· ···················· ····················
21210 LR878259.1 |Severe acute respiratory syndrome coro ···················· ···················· ···················· ···················· ···················· ····················
21211 LR878260.1 |Severe acute respiratory syndrome coro ···················· ···················· ···················· ···················· ···················· ····················
21212 LR878261.1 |Severe acute respiratory syndrome coro ···················· ···················· ···················· ···················· ···················· ····················
21213 LR878262.1 |Severe acute respiratory syndrome coro ···················· ···················· ···················· ···················· ···················· ····················
21214 LR878263.1 |Severe acute respiratory syndrome coro ···················· ···················· ···················· ···················· ···················· ····················
21215 LR878264.1 |Severe acute respiratory syndrome coro ···················· ···················· ···················· ···················· ···················· ····················
21216 LR878265.1 |Severe acute respiratory syndrome coro ···················· ···················· ···················· ···················· ···················· ····················
21217 LR878266.1 |Severe acute respiratory syndrome coro ···················· ···················· ···················· ···················· ···················· ····················
21218 LR878267.1 |Severe acute respiratory syndrome coro ···················· ···················· ···················· ···················· ···················· ····················
21219 LR878268.1 |Severe acute respiratory syndrome coro ···················· ···················· ···················· ···················· ···················· ····················
21220 LR878269.1 |Severe acute respiratory syndrome coro ···················· ···················· ···················· ···················· ···················· ····················
21221 LR878270.1 |Severe acute respiratory syndrome coro ···················· ···················· ···················· ···················· ···················· ····················

| | | | | | | | | |
|---|---|---|---|---|---|---|---|---|
| 21222 | LR878271.1 | \|Severe acute respiratory syndrome coro | ···················· | ···················· | ···················· | ···················· | ···················· | ···················· |
| 21223 | LR878272.1 | \|Severe acute respiratory syndrome coro | ···················· | ···················· | ···················· | ···················· | ···················· | ···················· |
| 21224 | LR878273.1 | \|Severe acute respiratory syndrome coro | ···················· | ···················· | ···················· | ···················· | ···················· | ···················· |
| 21225 | LR878274.1 | \|Severe acute respiratory syndrome coro | ···················· | ···················· | ···················· | ···················· | ···················· | ···················· |
| 21226 | LR878275.1 | \|Severe acute respiratory syndrome coro | ···················· | ···················· | ···················· | ···················· | ···················· | ···················· |
| 21227 | LR878276.1 | \|Severe acute respiratory syndrome coro | ···················· | ···················· | ···················· | ···················· | ···················· | ···················· |
| 21228 | LR878277.1 | \|Severe acute respiratory syndrome coro | ···················· | ···················· | ···················· | ···················· | ···················· | ···················· |
| 21229 | LR878278.1 | \|Severe acute respiratory syndrome coro | ···················· | ···················· | ···················· | ···················· | ···················· | ···················· |
| 21230 | LR878279.1 | \|Severe acute respiratory syndrome coro | ···················· | ···················· | ···················· | ···················· | ···················· | ···················· |
| 21231 | LR878280.1 | \|Severe acute respiratory syndrome coro | ···················· | ···················· | ···················· | ···················· | ···················· | ···················· |
| 21232 | LR878281.1 | \|Severe acute respiratory syndrome coro | ···················· | ···················· | ···················· | ···················· | ···················· | ···················· |
| 21233 | LR878282.1 | \|Severe acute respiratory syndrome coro | ···················· | ···················· | ···················· | ···················· | ···················· | ···················· |
| 21234 | LR878283.1 | \|Severe acute respiratory syndrome coro | ···················· | ···················· | ···················· | ···················· | ···················· | ···················· |
| 21235 | LR878284.1 | \|Severe acute respiratory syndrome coro | ···················· | ···················· | ···················· | ···················· | ···················· | ···················· |
| 21236 | LR878285.1 | \|Severe acute respiratory syndrome coro | ···················· | ···················· | ···················· | ···················· | ···················· | ···················· |
| 21237 | LR878286.1 | \|Severe acute respiratory syndrome coro | ···················· | ···················· | ···················· | ···················· | ···················· | ···················· |
| 21238 | LR878287.1 | \|Severe acute respiratory syndrome coro | ···················· | ···················· | ···················· | ···················· | ···················· | ···················· |
| 21239 | LR878288.1 | \|Severe acute respiratory syndrome coro | ···················· | ···················· | ···················· | ···················· | ···················· | ···················· |
| 21240 | LR878289.1 | \|Severe acute respiratory syndrome coro | ···················· | ···················· | ···················· | ···················· | ···················· | ···················· |
| 21241 | LR878290.1 | \|Severe acute respiratory syndrome coro | ···················· | ···················· | ···················· | ···················· | ···················· | ···················· |
| 21242 | LR878291.1 | \|Severe acute respiratory syndrome coro | ···················· | ···················· | ···················· | ···················· | ···················· | ···················· |
| 21243 | LR878292.1 | \|Severe acute respiratory syndrome coro | ···················· | ···················· | ···················· | ···················· | ···················· | ···················· |
| 21244 | LR878293.1 | \|Severe acute respiratory syndrome coro | ···T················ | ···················· | ···················· | ···················· | ···················· | ···················· |
| 21245 | LR878294.1 | \|Severe acute respiratory syndrome coro | ···················· | ···················· | ···················· | ···················· | ···················· | ···················· |
| 21246 | LR878295.1 | \|Severe acute respiratory syndrome coro | ···················· | ···················· | ···················· | ···················· | ···················· | ···················· |
| 21247 | LR878296.1 | \|Severe acute respiratory syndrome coro | ···················· | ···················· | ···················· | ···················· | ···················· | ···················· |
| 21248 | LR878297.1 | \|Severe acute respiratory syndrome coro | ···················· | ···················· | ···················· | ···················· | ···················· | ···················· |
| 21249 | LR878298.1 | \|Severe acute respiratory syndrome coro | ···················· | ···················· | ···················· | ···················· | ···················· | ···················· |
| 21251 | LR878300.1 | \|Severe acute respiratory syndrome coro | ···················· | ···················· | ···················· | ···················· | ···················· | ···················· |
| 21252 | LR878301.1 | \|Severe acute respiratory syndrome coro | ···T················ | ···················· | ···················· | ···················· | ···················· | ···················· |
| 21253 | LR878302.1 | \|Severe acute respiratory syndrome coro | ···················· | ···················· | ···················· | ···················· | ···················· | ···················· |
| 21254 | LR878303.1 | \|Severe acute respiratory syndrome coro | ···················· | ···················· | ···················· | ···················· | ···················· | ···················· |
| 21255 | LR878304.1 | \|Severe acute respiratory syndrome coro | ···················· | ···················· | ···················· | ···················· | ···················· | ···················· |
| 21256 | LR878305.1 | \|Severe acute respiratory syndrome coro | ···················· | ···················· | ···················· | ···················· | ···················· | ···················· |
| 21257 | LR878306.1 | \|Severe acute respiratory syndrome coro | ···················· | ···················· | ···················· | ···················· | ···················· | ···················· |
| 21258 | LR878307.1 | \|Severe acute respiratory syndrome coro | ···················· | ···················· | ···················· | ···················· | ···················· | ···················· |
| 21259 | LR878308.1 | \|Severe acute respiratory syndrome coro | ···················· | ···················· | ···················· | ···················· | ···················· | ···················· |
| 21260 | LR878309.1 | \|Severe acute respiratory syndrome coro | ···················· | ···················· | ···················· | ···················· | ···················· | ···················· |
| 21261 | LR878310.1 | \|Severe acute respiratory syndrome coro | ···················· | ···················· | ···················· | ···················· | ···················· | ···················· |
| 21262 | LR878311.1 | \|Severe acute respiratory syndrome coro | ···················· | ···················· | ···················· | ···················· | ···················· | ···················· |
| 21263 | LR878312.1 | \|Severe acute respiratory syndrome coro | ···················· | ···················· | ···················· | ···················· | ···················· | ···················· |
| 21264 | LR878313.1 | \|Severe acute respiratory syndrome coro | ···················· | ···················· | ···················· | ···················· | ···················· | ···················· |
| 21265 | LR878314.1 | \|Severe acute respiratory syndrome coro | ···················· | ···················· | ···················· | ···················· | ···················· | ···················· |
| 21266 | LR878315.1 | \|Severe acute respiratory syndrome coro | ···················· | ···················· | ···················· | ···················· | ···················· | ···················· |
| 21267 | LR878316.1 | \|Severe acute respiratory syndrome coro | ···················· | ···················· | ···················· | ···················· | ···················· | ···················· |
| 21268 | LR878317.1 | \|Severe acute respiratory syndrome coro | ···················· | ···················· | ···················· | ···················· | NN·················· | ···················· |
| 21269 | LR878318.1 | \|Severe acute respiratory syndrome coro | ···················· | ···················· | ···················· | ···················· | ···················· | ···················· |
| 21270 | LR878319.1 | \|Severe acute respiratory syndrome coro | ···················· | ···················· | ···················· | ···················· | ···················· | ···················· |
| 21271 | LR878320.1 | \|Severe acute respiratory syndrome coro | ···················· | ···················· | ···················· | ···················· | ···················· | ···················· |
| 21272 | LR878321.1 | \|Severe acute respiratory syndrome coro | ···················· | ···················· | ···················· | ···················· | ···················· | ···················· |
| 21273 | LR878322.1 | \|Severe acute respiratory syndrome coro | ···················· | ···················· | ···················· | ···················· | ···················· | ···················· |
| 21274 | LR878323.1 | \|Severe acute respiratory syndrome coro | ···················· | ···················· | ···················· | ···················· | ···················· | ···················· |
| 21275 | LR878324.1 | \|Severe acute respiratory syndrome coro | ···················· | ···················· | ···················· | ···················· | ···················· | ···················· |
| 21276 | LR878325.1 | \|Severe acute respiratory syndrome coro | ···················· | ···················· | ···················· | ···················· | ···················· | ···················· |
| 21277 | LR878326.1 | \|Severe acute respiratory syndrome coro | ···················· | ···················· | ···················· | ···················· | ···················· | ···················· |
| 21278 | LR878327.1 | \|Severe acute respiratory syndrome coro | ···················· | ···················· | ···················· | ···················· | ···················· | ···················· |
| 21279 | LR878328.1 | \|Severe acute respiratory syndrome coro | ···················· | ···················· | ···················· | ···················· | ···················· | ···················· |
| 21280 | LR878329.1 | \|Severe acute respiratory syndrome coro | ···················· | ···················· | ···················· | ···················· | ···················· | ···················· |
| 21281 | LR878330.1 | \|Severe acute respiratory syndrome coro | ···················· | ···················· | ···················· | ···················· | ···················· | ···················· |
| 21282 | LR878331.1 | \|Severe acute respiratory syndrome coro | ···················· | ···················· | ···················· | ···················· | ···················· | ···················· |
| 21283 | LR878332.1 | \|Severe acute respiratory syndrome coro | ···················· | ···················· | ···············C···· | ···················· | ···················· | ···················· |
| 21284 | LR878333.1 | \|Severe acute respiratory syndrome coro | ···················· | ···················· | ···················· | ···················· | ···················· | ···················· |
| 21285 | LR878334.1 | \|Severe acute respiratory syndrome coro | ···················· | ···················· | ···················· | ···················· | ···················· | ···················· |
| 21286 | LR878335.1 | \|Severe acute respiratory syndrome coro | ···················· | ···················· | ···················· | ···················· | ···················· | ···················· |
| 21287 | LR878336.1 | \|Severe acute respiratory syndrome coro | ···················· | ···················· | ···················· | ···················· | ···················· | ···················· |
| 21288 | LR878337.1 | \|Severe acute respiratory syndrome coro | ···················· | ···················· | ···················· | ···················· | ···················· | ···················· |
| 21289 | LR878338.1 | \|Severe acute respiratory syndrome coro | ···················· | ···················· | ···················· | ···················· | ···················· | ···················· |
| 21290 | LR878339.1 | \|Severe acute respiratory syndrome coro | ···················· | ···················· | ···················· | ···················· | ···················· | ···················· |
| 21291 | LR878340.1 | \|Severe acute respiratory syndrome coro | ···················· | ···················· | ···················· | ···················· | ···················· | ···················· |
| 21292 | LR878341.1 | \|Severe acute respiratory syndrome coro | ···················· | ···················· | ···················· | ···················· | ···················· | ···················· |
| 21293 | LR878342.1 | \|Severe acute respiratory syndrome coro | ···················· | ···················· | ···················· | ···················· | ···················· | ···················· |
| 21294 | LR878343.1 | \|Severe acute respiratory syndrome coro | ···················· | ···················· | ···················· | ···················· | ···················· | ···················· |
| 21295 | LR878344.1 | \|Severe acute respiratory syndrome coro | ···················· | ···················· | ···················· | ···················· | ···················· | ···················· |
| 21296 | LR878345.1 | \|Severe acute respiratory syndrome coro | ···················· | ···················· | ···················· | ···················· | ···················· | ···················· |
| 21297 | LR878346.1 | \|Severe acute respiratory syndrome coro | ···················· | ···················· | ···················· | ···················· | ···················· | ···················· |
| 21298 | LR878347.1 | \|Severe acute respiratory syndrome coro | ···················· | ···················· | ···················· | ···················· | ···················· | ···················· |
| 21299 | LR878348.1 | \|Severe acute respiratory syndrome coro | ···················· | ···················· | ···················· | ···················· | ···················· | ···················· |
| 21300 | LR878349.1 | \|Severe acute respiratory syndrome coro | ···················· | ···················· | ···················· | ···················· | ···················· | ···················· |

```
21301 LR878350.1 |Severe acute respiratory syndrome coro    ....................  ....................  .....................  ....................  ....................  ....................
21302 LR878351.1 |Severe acute respiratory syndrome coro    ....................  ....................  .....................  ....................  ....................  ....................
21303 LR878352.1 |Severe acute respiratory syndrome coro    ....................  ....................  .....................  ....................  ....................  ....................
21304 LR878353.1 |Severe acute respiratory syndrome coro    ....................  ....................  .....................  ....................  ....................  ....................
21305 LR878354.1 |Severe acute respiratory syndrome coro    ....................  ....................  .....................  ....................  ....................  ....................
21306 LR878355.1 |Severe acute respiratory syndrome coro    ....................  ....................  .....................  ....................  ....................  ....................
21307 LR878356.1 |Severe acute respiratory syndrome coro    ....................  ....................  .....................  ....................  ....................  ....................
21308 LR878357.1 |Severe acute respiratory syndrome coro    ....................  ....................  .....................  ....................  ....................  ....................
21309 LR878358.1 |Severe acute respiratory syndrome coro    ....................  ....................  .....................  ....................  ....................  ....................
21310 LR878359.1 |Severe acute respiratory syndrome coro    ....................  ....................  .....................  ....................  ....................  ....................
21311 LR878360.1 |Severe acute respiratory syndrome coro    ....................  ....................  .....................  ....................  ....................  ....................
21312 LR878361.1 |Severe acute respiratory syndrome coro    ....................  ....................  .....................  ....................  ....................  ....................
21313 LR878362.1 |Severe acute respiratory syndrome coro    ....................  ....................  .....................  ....................  ....................  ....................
21314 LR878363.1 |Severe acute respiratory syndrome coro    ....................  ....................  .....................  ....................  ....................  ....................
21315 MT919525.1 |Severe acute respiratory syndrome coro    ....................  ....................  .....................  ....................  ....................  ....................
21316 MT919526.1 |Severe acute respiratory syndrome coro    ....................  ....................  .....................  ....................  ....................  ....................
21317 MT919527.1 |Severe acute respiratory syndrome coro    ....................  ....................  .....................  ....................  ....................  ....................
21318 MT919528.1 |Severe acute respiratory syndrome coro    ....................  ....................  .....................  ....................  ....................  ....................
21319 MT919529.1 |Severe acute respiratory syndrome coro    ....................  ....................  .....................  ....................  ....................  ....................
21320 MT919530.1 |Severe acute respiratory syndrome coro    ....................  ....................  .....................  ....................  ....................  ....................
21321 MT919531.1 |Severe acute respiratory syndrome coro    ....................  ....................  .....................  ....................  ....................  ....................
21322 MT919532.1 |Severe acute respiratory syndrome coro    ....................  ....................  .....................  ....................  ....................  ....................
21323 MT919533.1 |Severe acute respiratory syndrome coro    ....................  ....................  .....................  ....................  ....................  ....................
21324 MT919534.1 |Severe acute respiratory syndrome coro    ....................  ....................  .....................  ....................  ....................  ....................
21325 MT919535.1 |Severe acute respiratory syndrome coro    ....................  ....................  .....................  ....................  ....................  ....................
21326 MT919536.1 |Severe acute respiratory syndrome coro    ....................  ....................  .....................  ....................  ....................  ....................
21327 MT919768.1 |Severe acute respiratory syndrome coro    ....................  ....................  .....................  ....................  ....................  ....................
21328 MT919769.1 |Severe acute respiratory syndrome coro    ....................  ....................  .....................  ....................  ....................  ....................
21329 MT919770.1 |Severe acute respiratory syndrome coro    ....................  ....................  .....................  ....................  ....................  ....................
21330 MT919771.1 |Severe acute respiratory syndrome coro    ....................  ....................  .....................  ....................  ....................  ....................
21331 MT919772.1 |Severe acute respiratory syndrome coro    ....................  ....................  .....................  ....................  ....................  ....................
21332 MT919773.1 |Severe acute respiratory syndrome coro    ....................  ....................  .....................  ....................  ....................  ....................
21333 MT919774.1 |Severe acute respiratory syndrome coro    ....................  ....................  .....................  ....................  ....................  ....................
21334 MT919775.1 |Severe acute respiratory syndrome coro    ....................  ....................  .....................  ....................  ....................  ....................
21335 MT919776.1 |Severe acute respiratory syndrome coro    ....................  ....................  .....................  ....................  ....................  ....................
21336 MT919777.1 |Severe acute respiratory syndrome coro    ....................  ....................  .....................  ....................  ....................  ....................
21337 MT919778.1 |Severe acute respiratory syndrome coro    ....................  ....................  .....................  ....................  ....................  ....................
21338 MT919779.1 |Severe acute respiratory syndrome coro    ....................  ....................  .....................  ....................  ....................  ....................
21339 MT919780.1 |Severe acute respiratory syndrome coro    ....................  ....................  .....................  ....................  ....................  ....................
21340 MT919781.1 |Severe acute respiratory syndrome coro    ....................  ....................  .....................  ....................  ....................  ....................
21341 MT919782.1 |Severe acute respiratory syndrome coro    ....................  ....................  .....................  ....................  ....................  ....................
21342 MT919783.1 |Severe acute respiratory syndrome coro    ....................  ....................  .....................  ....................  ....................  ....................
21343 MT919784.1 |Severe acute respiratory syndrome coro    ....................  ....................  .....................  ....................  ....................  ....................
21344 MT919785.1 |Severe acute respiratory syndrome coro    ....................  ....................  .....................  ....................  ....................  ....................
21345 MT919786.1 |Severe acute respiratory syndrome coro    ....................  ....................  .....................  ....................  ....................  ....................
21346 MT919787.1 |Severe acute respiratory syndrome coro    ....................  ....................  .....................  ....................  ....................  ....................
21347 MT919788.1 |Severe acute respiratory syndrome coro    ....................  ....................  .....................  ....................  ....................  ....................
21348 MT919789.1 |Severe acute respiratory syndrome coro    ....................  ....................  .....................  ....................  ....................  ....................
21349 MT919790.1 |Severe acute respiratory syndrome coro    ....................  ....................  .....................  ....................  ....................  ....................
21350 MT919994.1 |Severe acute respiratory syndrome coro    ....................  ....................  .....................  ....................  ....................  ....................
21351 MT919995.1 |Severe acute respiratory syndrome coro    ....................  ....................  .....................  ....................  ....................  ....................
21352 MT919996.1 |Severe acute respiratory syndrome coro    ....................  ....................  .....................  ....................  ....................  ....................
21353 MT919997.1 |Severe acute respiratory syndrome coro    ....................  ....................  .....................  ....................  ....................  ....................
21354 MT919998.1 |Severe acute respiratory syndrome coro    ....................  ....................  .....................  ....................  ....................  ....................
21355 MT919999.1 |Severe acute respiratory syndrome coro    ....................  ....................  .....................  ....................  ....................  ....................
21356 MT920000.1 |Severe acute respiratory syndrome coro    ....................  ....................  .....................  ....................  ....................  ....................
21357 MT920001.1 |Severe acute respiratory syndrome coro    ....................  ....................  .....................  ....................  ....................  ....................
21358 MT920002.1 |Severe acute respiratory syndrome coro    ....................  ....................  .....................  ....................  ....................  ....................
21359 MT920003.1 |Severe acute respiratory syndrome coro    ....................  ....................  .....................  ....................  ....................  ....................
21360 MT920004.1 |Severe acute respiratory syndrome coro    ....................  ....................  .....................  ....................  ....................  ....................
21361 MT920005.1 |Severe acute respiratory syndrome coro    ....................  ....................  .....................  ....................  ....................  ....................
21362 MT920006.1 |Severe acute respiratory syndrome coro    ....................  ....................  .....................  ....................  ....................  ....................
21363 MT920007.1 |Severe acute respiratory syndrome coro    ....................  ....................  .....................  ....................  ....................  ....................
21364 MT920008.1 |Severe acute respiratory syndrome coro    ....................  ....................  .....................  ....................  ....................  ....................
21365 MT920009.1 |Severe acute respiratory syndrome coro    ....................  ....................  .....................  ....................  ....................  ....................
21366 MT920010.1 |Severe acute respiratory syndrome coro    ....................  ....................  .....................  ....................  ....................  ....................
21367 MT920011.1 |Severe acute respiratory syndrome coro    ....................  ....................  .....................  ....................  ....................  ....................
21368 MT920012.1 |Severe acute respiratory syndrome coro    ....................  ....................  .....................  ....................  ....................  ....................
21369 MT920013.1 |Severe acute respiratory syndrome coro    ....................  ....................  .....................  ....................  ....................  ....................
21370 MT920014.1 |Severe acute respiratory syndrome coro    ....................  ....................  .....................  ....................  ....................  ....................
21371 MT920015.1 |Severe acute respiratory syndrome coro    ....................  ....................  .....................  ....................  ....................  ....................
21372 MT920016.1 |Severe acute respiratory syndrome coro    ....................  ....................  .....................  ....................  ....................  ....................
21373 MT920017.1 |Severe acute respiratory syndrome coro    ....................  ....................  .....................  ....................  ....................  ....................
21374 MT920018.1 |Severe acute respiratory syndrome coro    ....................  ....................  .....................  ....................  ....................  ....................
21375 MT920019.1 |Severe acute respiratory syndrome coro    ....................  ....................  .....................  ....................  ....................  ....................
21376 MT920020.1 |Severe acute respiratory syndrome coro    ....................  ....................  .....................  ....................  ....................  ....................
21377 MT920021.1 |Severe acute respiratory syndrome coro    ....................  ....................  .....................  ....................  ....................  ....................
21378 MT920022.1 |Severe acute respiratory syndrome coro    ....................  ....................  .....................  ....................  ....................  ....................
```

| | | | | | | | | |
|---|---|---|---|---|---|---|---|---|
| 21379 | MT920023.1 | Severe acute respiratory syndrome coro | .................... | .................... | .................... | .................... | .................... | .................... |
| 21380 | MT920024.1 | Severe acute respiratory syndrome coro | .................... | .................... | .................... | .................... | .................... | .................... |
| 21381 | MT920025.1 | Severe acute respiratory syndrome coro | .................... | .................... | .................... | .................... | .................... | .................... |
| 21382 | MT920026.1 | Severe acute respiratory syndrome coro | .................... | .................... | .................... | .................... | .................... | .................... |
| 21383 | MT920027.1 | Severe acute respiratory syndrome coro | .................... | .................... | .................... | .................... | .................... | .................... |
| 21384 | MT920028.1 | Severe acute respiratory syndrome coro | .................... | .................... | .................... | .................... | .................... | .................... |
| 21385 | MT920029.1 | Severe acute respiratory syndrome coro | .................... | .................... | .................... | .................... | .................... | .................... |
| 21386 | MT920030.1 | Severe acute respiratory syndrome coro | .................... | .................... | .................... | .................... | .................... | .................... |
| 21387 | MT920031.1 | Severe acute respiratory syndrome coro | .................... | .................... | .................... | .................... | .................... | .................... |
| 21388 | MT920032.1 | Severe acute respiratory syndrome coro | .................... | .................... | .................... | .................... | .................... | .................... |
| 21389 | MT920033.1 | Severe acute respiratory syndrome coro | .................... | .................... | .................... | .................... | .................... | .................... |
| 21390 | MT920034.1 | Severe acute respiratory syndrome coro | .................... | .................... | .................... | .................... | .................... | .................... |
| 21391 | MT920035.1 | Severe acute respiratory syndrome coro | .................... | .................... | .................... | .................... | .................... | .................... |
| 21392 | MT920036.1 | Severe acute respiratory syndrome coro | .................... | .................... | .................... | .................... | .................... | .................... |
| 21393 | MT920037.1 | Severe acute respiratory syndrome coro | .................... | .................... | .................... | .................... | ......T............. | .................... |
| 21394 | MT920038.1 | Severe acute respiratory syndrome coro | .................... | .................... | .................... | .................... | .................... | .................... |
| 21395 | MT920039.1 | Severe acute respiratory syndrome coro | .................... | .................... | .................... | .................... | .................... | .................... |
| 21396 | MT920040.1 | Severe acute respiratory syndrome coro | .................... | .................... | .................... | .................... | .................... | .................... |
| 21397 | MT921009.1 | Severe acute respiratory syndrome coro | .................... | .................... | .................... | .................... | .................... | .................... |
| 21398 | MT921159.1 | Severe acute respiratory syndrome coro | .................... | .................... | .................... | .................... | .................... | .................... |
| 21399 | MT921165.1 | Severe acute respiratory syndrome coro | .................... | .................... | .................... | .................... | .................... | .................... |
| 21400 | MT921166.1 | Severe acute respiratory syndrome coro | .................... | .................... | .................... | .................... | .................... | .................... |
| 21401 | MT913038.1 | Severe acute respiratory syndrome coro | .................... | .................... | .................... | .................... | .................... | .................... |
| 21402 | MT913039.1 | Severe acute respiratory syndrome coro | .................... | .................... | .................... | .................... | .................... | .................... |
| 21403 | MT913040.1 | Severe acute respiratory syndrome coro | .................... | .................... | .................... | .................... | .................... | .................... |
| 21404 | MT913041.1 | Severe acute respiratory syndrome coro | .................... | .................... | .................... | .................... | .................... | .................... |
| 21405 | MT913042.1 | Severe acute respiratory syndrome coro | .................... | .................... | .................... | .................... | .................... | .................... |
| 21406 | MT913043.1 | Severe acute respiratory syndrome coro | .................... | .................... | .................... | .................... | .................... | .................... |
| 21407 | MT913044.1 | Severe acute respiratory syndrome coro | .................... | .................... | .................... | .................... | .................... | .................... |
| 21408 | MT913045.1 | Severe acute respiratory syndrome coro | .................... | .................... | .................... | .................... | .................... | .................... |
| 21409 | MT913046.1 | Severe acute respiratory syndrome coro | .................... | .................... | .................... | .................... | .................... | .................... |
| 21410 | MT913047.1 | Severe acute respiratory syndrome coro | .................... | .................... | .................... | .................... | .................... | .................... |
| 21411 | MT913048.1 | Severe acute respiratory syndrome coro | .................... | .................... | .................... | .................... | .................... | .................... |
| 21412 | MT913049.1 | Severe acute respiratory syndrome coro | .................... | .................... | .................... | .................... | .................... | .................... |
| 21413 | MT913050.1 | Severe acute respiratory syndrome coro | .................... | .................... | .................... | .................... | .................... | .................... |
| 21414 | MT913051.1 | Severe acute respiratory syndrome coro | .................... | .................... | .................... | .................... | .................... | .................... |
| 21415 | MT913052.1 | Severe acute respiratory syndrome coro | .................... | .................... | .................... | .................... | .................... | .................... |
| 21416 | MT913053.1 | Severe acute respiratory syndrome coro | .................... | .................... | .................... | .................... | .................... | .................... |
| 21417 | MT913054.1 | Severe acute respiratory syndrome coro | .................... | .................... | .................... | .................... | .................... | .................... |
| 21418 | MT913055.1 | Severe acute respiratory syndrome coro | .................... | .................... | .................... | .................... | .................... | .................... |
| 21419 | MT913056.1 | Severe acute respiratory syndrome coro | .................... | .................... | .................... | .................... | .................... | .................... |
| 21420 | MT913057.1 | Severe acute respiratory syndrome coro | .................... | .................... | .................... | .................... | .................... | .................... |
| 21421 | MT913058.1 | Severe acute respiratory syndrome coro | .................... | .................... | .................... | .................... | .................... | .................... |
| 21422 | MT913059.1 | Severe acute respiratory syndrome coro | .................... | .................... | .................... | .................... | .................... | .................... |
| 21423 | MT913061.1 | Severe acute respiratory syndrome coro | .................... | .................... | .................... | .................... | .................... | .................... |
| 21424 | MT913062.1 | Severe acute respiratory syndrome coro | .................... | .................... | .................... | .................... | .................... | .................... |
| 21425 | MT913063.1 | Severe acute respiratory syndrome coro | .................... | .................... | .................... | .................... | .................... | .................... |
| 21426 | MT913064.1 | Severe acute respiratory syndrome coro | .................... | .................... | .................... | .................... | .................... | .................... |
| 21427 | MT913065.1 | Severe acute respiratory syndrome coro | .................... | .................... | .................... | .................... | .................... | .................... |
| 21428 | MT913066.1 | Severe acute respiratory syndrome coro | .................... | .................... | .................... | .................... | .................... | .................... |
| 21429 | MT913067.1 | Severe acute respiratory syndrome coro | .................... | .................... | .................... | .................... | .................... | .................... |
| 21430 | MT913068.1 | Severe acute respiratory syndrome coro | .................... | .................... | .................... | .................... | .................... | .................... |
| 21431 | MT913069.1 | Severe acute respiratory syndrome coro | .................... | .................... | .................... | .................... | .................... | .................... |
| 21432 | MT913070.1 | Severe acute respiratory syndrome coro | .................... | .................... | .................... | .................... | .................... | .................... |
| 21433 | MT913071.1 | Severe acute respiratory syndrome coro | .................... | .................... | .................... | .................... | .................... | .................... |
| 21434 | MT913073.1 | Severe acute respiratory syndrome coro | .................... | .................... | .................... | .................... | .................... | .................... |
| 21435 | MT913074.1 | Severe acute respiratory syndrome coro | .................... | .................... | .................... | .................... | .................... | .................... |
| 21436 | MT913075.1 | Severe acute respiratory syndrome coro | .................... | .................... | .................... | .................... | .................... | .................... |
| 21437 | MT913076.1 | Severe acute respiratory syndrome coro | .................... | .................... | .................... | .................... | .................... | .................... |
| 21438 | MT913077.1 | Severe acute respiratory syndrome coro | .................... | .................... | .................... | .................... | .................... | .................... |
| 21439 | MT913078.1 | Severe acute respiratory syndrome coro | .................... | .................... | .................... | .................... | .................... | .................... |
| 21440 | MT913079.1 | Severe acute respiratory syndrome coro | .................... | .................... | .................... | .................... | .................... | .................... |
| 21441 | MT913080.1 | Severe acute respiratory syndrome coro | .................... | .................... | .................... | .................... | .................... | .................... |
| 21442 | MT913081.1 | Severe acute respiratory syndrome coro | .................... | .................... | .................... | .................... | .................... | .................... |
| 21443 | MT913082.1 | Severe acute respiratory syndrome coro | .................... | .................... | .................... | .................... | .................... | .................... |
| 21444 | MT913083.1 | Severe acute respiratory syndrome coro | .................... | .................... | .................... | .................... | .................... | .................... |
| 21445 | MT913084.1 | Severe acute respiratory syndrome coro | .................... | .................... | .................... | .................... | .................... | .................... |
| 21446 | MT913085.1 | Severe acute respiratory syndrome coro | .................... | .................... | .................... | .................... | .................... | .................... |
| 21447 | MT913086.1 | Severe acute respiratory syndrome coro | .................... | .................... | .................... | .................... | .................... | .................... |
| 21448 | MT913087.1 | Severe acute respiratory syndrome coro | .................... | .................... | .................... | .................... | .................... | .................... |
| 21449 | MT913088.1 | Severe acute respiratory syndrome coro | .................... | .................... | .................... | .................... | .................... | .................... |
| 21450 | MT913089.1 | Severe acute respiratory syndrome coro | .................... | .................... | .................... | .................... | .................... | .................... |
| 21451 | MT913090.1 | Severe acute respiratory syndrome coro | .................... | .................... | .................... | .................... | .................... | .................... |
| 21452 | MT913092.1 | Severe acute respiratory syndrome coro | .................... | .................... | .................... | .................... | .................... | .................... |
| 21453 | MT913093.1 | Severe acute respiratory syndrome coro | .................... | .................... | .................... | .................... | .................... | .................... |
| 21454 | MT913095.1 | Severe acute respiratory syndrome coro | .................... | .................... | .................... | .................... | .................... | .................... |
| 21455 | MT913096.1 | Severe acute respiratory syndrome coro | .................... | .................... | .................... | .................... | .................... | .................... |
| 21456 | MT913098.1 | Severe acute respiratory syndrome coro | .................... | .................... | .................... | .................... | .................... | .................... |

| | | | | | | | | |
|---|---|---|---|---|---|---|---|---|
| 21457 | MT913099.1 | Severe acute respiratory syndrome coro | .................... | .................... | ..................... | .................... | .................... | .................... |
| 21458 | MT913100.1 | Severe acute respiratory syndrome coro | .................... | .................... | ..................... | .................... | .................... | .................... |
| 21459 | MT913101.1 | Severe acute respiratory syndrome coro | .................... | .................... | ..................... | .................... | .................... | .................... |
| 21460 | MT913102.1 | Severe acute respiratory syndrome coro | .................... | .................... | ..................... | .................... | .................... | .................... |
| 21461 | MT913103.1 | Severe acute respiratory syndrome coro | .................... | .................... | ..................... | .................... | .................... | .................... |
| 21462 | MT913104.1 | Severe acute respiratory syndrome coro | .................... | .................... | ..................... | .................... | .................... | .................... |
| 21463 | MT913105.1 | Severe acute respiratory syndrome coro | .................... | .................... | ..................... | .................... | .................... | .................... |
| 21464 | MT913106.1 | Severe acute respiratory syndrome coro | .................... | .................... | ..................... | .................... | .................... | .................... |
| 21465 | MT913107.1 | Severe acute respiratory syndrome coro | .................... | .................... | ..................... | .................... | .................... | .................... |
| 21466 | MT913109.1 | Severe acute respiratory syndrome coro | .................... | .................... | ..................... | .................... | .................... | .................... |
| 21467 | MT913110.1 | Severe acute respiratory syndrome coro | .................... | .................... | ..................... | .................... | .................... | .................... |
| 21469 | MT913112.1 | Severe acute respiratory syndrome coro | .................... | .................... | ..................... | .................... | .................... | .................... |
| 21470 | MT913113.1 | Severe acute respiratory syndrome coro | .................... | .................... | ..................... | .................... | .................... | .................... |
| 21471 | MT913114.1 | Severe acute respiratory syndrome coro | .................... | .................... | ..................... | .................... | .................... | .................... |
| 21472 | MT913116.1 | Severe acute respiratory syndrome coro | .................... | .................... | ..................... | .................... | .................... | .................... |
| 21473 | MT913117.1 | Severe acute respiratory syndrome coro | .................... | .................... | ..................... | .................... | .................... | .................... |
| 21474 | MT913118.1 | Severe acute respiratory syndrome coro | .................... | .................... | ..................... | .................... | .................... | .................... |
| 21475 | MT913119.1 | Severe acute respiratory syndrome coro | .................... | .................... | ..................... | .................... | .................... | .................... |
| 21476 | MT913122.1 | Severe acute respiratory syndrome coro | .................... | .................... | ..................... | .................... | .................... | .................... |
| 21477 | MT913123.1 | Severe acute respiratory syndrome coro | .................... | .................... | ..................... | .................... | .................... | .................... |
| 21478 | MT913124.1 | Severe acute respiratory syndrome coro | .................... | .................... | ..................... | .................... | .................... | .................... |
| 21479 | MT913125.1 | Severe acute respiratory syndrome coro | .................... | .................... | ..................... | .................... | .................... | .................... |
| 21480 | MT913126.1 | Severe acute respiratory syndrome coro | .................... | .................... | ..................... | .................... | .................... | .................... |
| 21481 | MT913127.1 | Severe acute respiratory syndrome coro | .................... | .................... | ..................... | .................... | .................... | .................... |
| 21482 | MT913128.1 | Severe acute respiratory syndrome coro | .................... | .................... | ..................... | .................... | .................... | .................... |
| 21483 | MT913129.1 | Severe acute respiratory syndrome coro | .................... | .................... | ..................... | .................... | .................... | .................... |
| 21484 | MT913130.1 | Severe acute respiratory syndrome coro | .................... | .................... | ..................... | .................... | .................... | .................... |
| 21485 | MT913131.1 | Severe acute respiratory syndrome coro | .................... | .................... | ..................... | .................... | .................... | .................... |
| 21486 | MT913133.1 | Severe acute respiratory syndrome coro | .................... | .................... | ..................... | .................... | .................... | .................... |
| 21487 | MT913134.1 | Severe acute respiratory syndrome coro | .................... | .................... | ..................... | .................... | .................... | .................... |
| 21488 | MT913135.1 | Severe acute respiratory syndrome coro | .................... | .................... | ..................... | .................... | .................... | .................... |
| 21489 | MT913137.1 | Severe acute respiratory syndrome coro | .................... | .................... | ..................... | .................... | .................... | .................... |
| 21490 | MT913138.1 | Severe acute respiratory syndrome coro | .................... | .................... | ..................... | .................... | .................... | .................... |
| 21491 | MT913139.1 | Severe acute respiratory syndrome coro | .................... | .................... | ..................... | .................... | .................... | .................... |
| 21492 | MT913140.1 | Severe acute respiratory syndrome coro | .................... | .................... | ..................... | .................... | .................... | .................... |
| 21493 | MT913141.1 | Severe acute respiratory syndrome coro | .................... | .................... | ..................... | .................... | .................... | .................... |
| 21494 | MT913142.1 | Severe acute respiratory syndrome coro | .................... | .................... | ..................... | .................... | .................... | .................... |
| 21495 | MT913143.1 | Severe acute respiratory syndrome coro | .................... | .................... | ..................... | .................... | .................... | .................... |
| 21496 | MT913144.1 | Severe acute respiratory syndrome coro | .................... | .................... | ..................... | .................... | .................... | .................... |
| 21497 | MT913145.1 | Severe acute respiratory syndrome coro | .................... | .................... | ..................... | .................... | .................... | .................... |
| 21498 | MT913146.1 | Severe acute respiratory syndrome coro | .................... | .................... | ..................... | .................... | .................... | .................... |
| 21499 | MT913147.1 | Severe acute respiratory syndrome coro | .................... | .................... | ..................... | .................... | .................... | .................... |
| 21500 | MT913148.1 | Severe acute respiratory syndrome coro | .................... | .................... | ..................... | .................... | .................... | .................... |
| 21501 | MT913149.1 | Severe acute respiratory syndrome coro | .................... | .................... | ..................... | .................... | .................... | .................... |
| 21502 | MT913150.1 | Severe acute respiratory syndrome coro | .................... | .................... | ..................... | .................... | .................... | .................... |
| 21503 | MT913151.1 | Severe acute respiratory syndrome coro | .................... | ....A............... | ..................... | .................... | .................... | .................... |
| 21504 | MT913152.1 | Severe acute respiratory syndrome coro | .................... | .................... | ..................... | .................... | .................... | .................... |
| 21505 | LR877721.1 | Severe acute respiratory syndrome coro | .................... | .................... | ..................... | .................... | .................... | .................... |
| 21506 | LR877722.1 | Severe acute respiratory syndrome coro | .................... | .................... | ..................... | .................... | .................... | .................... |
| 21507 | LR877723.1 | Severe acute respiratory syndrome coro | .................... | .................... | ..................... | .................... | .................... | .................... |
| 21508 | LR877724.1 | Severe acute respiratory syndrome coro | .................... | .................... | ..................... | .................... | .................... | .................... |
| 21509 | LR877725.1 | Severe acute respiratory syndrome coro | .................... | .................... | ..................... | .................... | .................... | .................... |
| 21510 | LR877726.1 | Severe acute respiratory syndrome coro | .................... | .................... | ..................... | .................... | .................... | .................... |
| 21511 | LR877727.1 | Severe acute respiratory syndrome coro | .................... | .................... | ..................... | .................... | .................... | .................... |
| 21512 | LR877728.1 | Severe acute respiratory syndrome coro | .................... | .................... | ..................... | .................... | .................... | .................... |
| 21513 | LR877729.1 | Severe acute respiratory syndrome coro | .................... | .................... | ..................... | .................... | .................... | .................... |
| 21514 | LR877730.1 | Severe acute respiratory syndrome coro | .................... | .................... | ..................... | .................... | .................... | .................... |
| 21515 | LR877731.1 | Severe acute respiratory syndrome coro | .................... | .................... | ..................... | .................... | .................... | .................... |
| 21516 | LR877732.1 | Severe acute respiratory syndrome coro | .................... | .................... | ..................... | .................... | .................... | .................... |
| 21517 | LR877733.1 | Severe acute respiratory syndrome coro | .................... | .................... | ..................... | .................... | .................... | .................... |
| 21518 | LR877734.1 | Severe acute respiratory syndrome coro | .................... | .................... | ..................... | .................... | .................... | .................... |
| 21519 | LR877735.1 | Severe acute respiratory syndrome coro | .................... | .................... | ..................... | .................... | .................... | .................... |
| 21520 | LR877736.1 | Severe acute respiratory syndrome coro | .................... | .................... | ..................... | .................... | .................... | .................... |
| 21521 | LR877737.1 | Severe acute respiratory syndrome coro | .................... | .................... | ..................... | .................... | .................... | .................... |
| 21522 | LR877738.1 | Severe acute respiratory syndrome coro | .................... | .................... | ..................... | .................... | .................... | .................... |
| 21523 | LR877739.1 | Severe acute respiratory syndrome coro | .................... | .................... | ..................... | .................... | .................... | .................... |
| 21524 | LR877740.1 | Severe acute respiratory syndrome coro | .................... | .................... | ..................... | .................... | .................... | .................... |
| 21525 | LR877741.1 | Severe acute respiratory syndrome coro | .................... | .................... | ..................... | .................... | .................... | .................... |
| 21526 | LR877742.1 | Severe acute respiratory syndrome coro | .................... | .................... | ..................... | .................... | .................... | .................... |
| 21527 | LR877743.1 | Severe acute respiratory syndrome coro | .................... | .................... | ..................... | .................... | .................... | .................... |
| 21528 | LR877744.1 | Severe acute respiratory syndrome coro | .................... | .................... | ..................... | .................... | .................... | .................... |
| 21529 | LR877745.1 | Severe acute respiratory syndrome coro | .................... | .................... | ..................... | .................... | .................... | .................... |
| 21530 | LR877746.1 | Severe acute respiratory syndrome coro | .................... | .................... | ..................... | .................... | .................... | .................... |
| 21531 | LR877747.1 | Severe acute respiratory syndrome coro | .................... | .................... | ..................... | .................... | .................... | .................... |
| 21532 | LR877748.1 | Severe acute respiratory syndrome coro | .................... | .................... | ..................... | .................... | .................... | .................... |
| 21533 | LR877749.1 | Severe acute respiratory syndrome coro | .................... | .................... | ..................... | .................... | .................... | .................... |
| 21534 | LR877750.1 | Severe acute respiratory syndrome coro | .................... | .................... | ..................... | .................... | .................... | .................... |
| 21535 | LR877751.1 | Severe acute respiratory syndrome coro | .................... | .................... | ..................... | .................... | .................... | .................... |

21536 LR877752.1 |Severe acute respiratory syndrome coro .................... .................... .................... .................... .................... ....................
21537 LR877753.1 |Severe acute respiratory syndrome coro .................... .................... .................... .................... .................... ....................
21538 LR877754.1 |Severe acute respiratory syndrome coro .................... .................... .................... .................... .................... ....................
21539 LR877755.1 |Severe acute respiratory syndrome coro .................... .................... .................... .................... .................... ....................
21540 LR877756.1 |Severe acute respiratory syndrome coro .................... .................... .................... .................... .................... ....................
21541 LR877757.1 |Severe acute respiratory syndrome coro .................... .................... .................... .................... .................... ....................
21542 LR877758.1 |Severe acute respiratory syndrome coro .................... .................... .................... .................... .................... ....................
21543 LR877759.1 |Severe acute respiratory syndrome coro .................... .................... .................... .................... .................... ....................
21544 LR877760.1 |Severe acute respiratory syndrome coro .................... .................... .................... .................... .................... ....................
21545 LR877761.1 |Severe acute respiratory syndrome coro .................... .................... .................... .................... .................... ....................
21546 LR877762.1 |Severe acute respiratory syndrome coro .................... .................... .................... .................... .................... ....................
21547 LR877763.1 |Severe acute respiratory syndrome coro .................... .............A...... .................... .................... .................... ....................
21548 LR877764.1 |Severe acute respiratory syndrome coro .................... .................... .................... .................... .................... ....................
21549 LR877765.1 |Severe acute respiratory syndrome coro ........T........... .................... .................... .................... .................... ....................
21550 LR877766.1 |Severe acute respiratory syndrome coro .................... .................... .................... .................... .................... ....................
21551 LR877767.1 |Severe acute respiratory syndrome coro .................... .................... .................... .................... .................... ....................
21552 LR877768.1 |Severe acute respiratory syndrome coro .................... .................... .................... .................... .................... ....................
21553 LR877769.1 |Severe acute respiratory syndrome coro .................... .................... .................... .................... .................... ....................
21554 LR877770.1 |Severe acute respiratory syndrome coro .................... .................... .................... .................... .................... ....................
21555 LR877771.1 |Severe acute respiratory syndrome coro .................... .................... .................... .................... .................... ....................
21556 LR877772.1 |Severe acute respiratory syndrome coro .................... .................... .................... .................... ....T............... ....................
21557 LR877773.1 |Severe acute respiratory syndrome coro .................... .................... .................... .................... .................... ....................
21558 LR877774.1 |Severe acute respiratory syndrome coro .................... .................... .................... .................... .................... ....................
21559 LR877775.1 |Severe acute respiratory syndrome coro .................... .................... .................... .................... .................... ....................
21560 LR877776.1 |Severe acute respiratory syndrome coro .................... .................... .................... .................... .................... ....................
21561 LR877777.1 |Severe acute respiratory syndrome coro .................... .................... .................... .................... .................... ....................
21562 LR877778.1 |Severe acute respiratory syndrome coro .................... .................... .................... .................... .................... ....................
21563 LR877779.1 |Severe acute respiratory syndrome coro .................... .................... .................... .................... .................... ....................
21564 LR877780.1 |Severe acute respiratory syndrome coro .................... .................... .................... .................... ....T............... ....................
21565 LR877781.1 |Severe acute respiratory syndrome coro .................... .................... .................... .................... .................... ....................
21566 LR877782.1 |Severe acute respiratory syndrome coro .................... .................... .................... .................... .................... ....................
21567 LR877783.1 |Severe acute respiratory syndrome coro .................... .................... .................... .................... .................... ....................
21568 LR877784.1 |Severe acute respiratory syndrome coro .................... .................... .................... .................... .................... ....................
21569 LR877785.1 |Severe acute respiratory syndrome coro .................... .................... .................... .................... .................... ....................
21570 LR877786.1 |Severe acute respiratory syndrome coro .................... .................... .................... .................... .................... ....................
21571 LR877787.1 |Severe acute respiratory syndrome coro .................... .................... .................... .................... .................... ....................
21572 LR877788.1 |Severe acute respiratory syndrome coro .................... .................... .................... .................... .................... ....................
21573 LR877789.1 |Severe acute respiratory syndrome coro .................... .................... .................... .................... .................... ....................
21574 LR877790.1 |Severe acute respiratory syndrome coro .................... .................... .................... .................... .................... ....................
21575 LR877791.1 |Severe acute respiratory syndrome coro .................... .................... .................... .................... .................... ....................
21576 LR877792.1 |Severe acute respiratory syndrome coro .................... .................... .................... .................... .................... ....................
21577 LR877793.1 |Severe acute respiratory syndrome coro .................... .................... .................... .................... .................... ....................
21578 LR877794.1 |Severe acute respiratory syndrome coro .................... .................... .................... .................... .................... ....................
21579 LR877795.1 |Severe acute respiratory syndrome coro .................... .................... .................... .................... .................... ....................
21580 LR877796.1 |Severe acute respiratory syndrome coro .................... .................... .................... .................... .................... ....................
21581 LR877797.1 |Severe acute respiratory syndrome coro .................... .................... .................... .................... .................... ....................
21582 LR877798.1 |Severe acute respiratory syndrome coro .................... .................... .................... .................... .................... ....................
21583 LR877799.1 |Severe acute respiratory syndrome coro .................... .................... .................... .................... .................... ....................
21584 LR877800.1 |Severe acute respiratory syndrome coro .................... .................... .................... .................... .................... ....................
21585 LR877801.1 |Severe acute respiratory syndrome coro .................... .................... .................... .................... .................... ....................
21586 LR877802.1 |Severe acute respiratory syndrome coro .................... .................... .................... .................... .................... ....................
21587 LR877803.1 |Severe acute respiratory syndrome coro .................... .................... .................... .................... .................... ....................
21588 LR877804.1 |Severe acute respiratory syndrome coro .................... .................... .................... .................... .................... ....................
21589 LR877805.1 |Severe acute respiratory syndrome coro .................... .................... .................... .................... .................... ....................
21590 LR877806.1 |Severe acute respiratory syndrome coro .................... .................... .................... .................... .................... ....................
21591 LR877807.1 |Severe acute respiratory syndrome coro .................... .................... .................... .................... .................... ....................
21592 LR877808.1 |Severe acute respiratory syndrome coro .................... .................... .................... .................... .................... ....................
21593 LR877809.1 |Severe acute respiratory syndrome coro .................... .................... .................... .................... .................... ....................
21594 LR877810.1 |Severe acute respiratory syndrome coro .................... .................... .................... .................... .................... ....................
21595 LR877811.1 |Severe acute respiratory syndrome coro .................... .................... .................... .................... .................... ....................
21596 LR877812.1 |Severe acute respiratory syndrome coro .................... .................... .................... .................... .................... ....................
21597 LR877813.1 |Severe acute respiratory syndrome coro .................... .................... .................... .................... .................... ....................
21598 LR877814.1 |Severe acute respiratory syndrome coro .................... .................... .................... .................... .................... ....................
21599 LR877815.1 |Severe acute respiratory syndrome coro .................... .................... .................... .................... .................... ....................
21600 LR877816.1 |Severe acute respiratory syndrome coro .................... .................... .................... .................... .................... ....................
21601 LR877817.1 |Severe acute respiratory syndrome coro .................... .................... .................... .................... .................... ....................
21602 LR877818.1 |Severe acute respiratory syndrome coro .................... .................... .................... .................... .................... ....................
21603 LR877819.1 |Severe acute respiratory syndrome coro .................... .................... .................... .................... .................... ....................
21604 LR877820.1 |Severe acute respiratory syndrome coro .................... .................... .................... .................... .................... ....................
21605 LR877821.1 |Severe acute respiratory syndrome coro .................... .................... .................... .................... .................... ....................
21606 LR877822.1 |Severe acute respiratory syndrome coro .................... .................... .................... .................... .................... ....................
21607 LR877823.1 |Severe acute respiratory syndrome coro .................... .................... .................... .................... .................... ....................
21608 LR877824.1 |Severe acute respiratory syndrome coro .................... .................... .................... .................... .................... ....................
21609 LR877825.1 |Severe acute respiratory syndrome coro .................... .................... .................... .................... .................... ....................
21610 MT907515.1 |Severe acute respiratory syndrome coro .................... .................... .................... .................... .................... ....................
21611 MT907516.1 |Severe acute respiratory syndrome coro .................... .................... .................... .................... .................... ....................
21612 MT907517.1 |Severe acute respiratory syndrome coro .................... .................... .................... .................... .................... ....................
21613 MT907518.1 |Severe acute respiratory syndrome coro .................... .................... .................... .................... .................... ....................

| | | | | | | | |
|---|---|---|---|---|---|---|---|
| 21614 MT907519.1 | \|Severe acute respiratory syndrome coro | .................... | .................... | ..................... | .................... | .................... | .................... |
| 21615 MT907520.2 | \|Severe acute respiratory syndrome coro | .................... | .................... | ..................... | .................... | .................... | .................... |
| 21616 MT911534.1 | \|Severe acute respiratory syndrome coro | .................... | .................... | ..................... | .................... | .................... | .................... |
| 21617 MT911538.1 | \|Severe acute respiratory syndrome coro | .................... | .................... | ..................... | .................... | .................... | .................... |
| 21618 MT911539.1 | \|Severe acute respiratory syndrome coro | .................... | .................... | ..................... | .................... | .................... | .................... |
| 21619 MT911540.1 | \|Severe acute respiratory syndrome coro | .................... | .................... | ..................... | .................... | .................... | .................... |
| 21620 MT911541.1 | \|Severe acute respiratory syndrome coro | .................... | .................... | ..................... | .................... | .................... | .................... |
| 21621 MT911542.1 | \|Severe acute respiratory syndrome coro | .................... | .................... | ..................... | .................... | .................... | .................... |
| 21622 MT911543.1 | \|Severe acute respiratory syndrome coro | .................... | .................... | ..................... | .................... | .................... | .................... |
| 21623 MT911544.1 | \|Severe acute respiratory syndrome coro | .................... | .................... | ..................... | .................... | .................... | .................... |
| 21624 MT911545.1 | \|Severe acute respiratory syndrome coro | .................... | .................... | ..................... | .................... | .................... | .................... |
| 21625 MT911546.1 | \|Severe acute respiratory syndrome coro | .................... | .................... | ..................... | .................... | .................... | .................... |
| 21626 MT911547.1 | \|Severe acute respiratory syndrome coro | .................... | .................... | ..................... | .................... | .................... | .................... |
| 21627 MT911548.1 | \|Severe acute respiratory syndrome coro | .................... | .................... | ..................... | .................... | .................... | .................... |
| 21628 MT911549.1 | \|Severe acute respiratory syndrome coro | .................... | .................... | ..................... | .................... | .................... | .................... |
| 21629 MT911550.1 | \|Severe acute respiratory syndrome coro | .................... | .................... | ..................... | .................... | .................... | .................... |
| 21630 MT911551.1 | \|Severe acute respiratory syndrome coro | .................... | .................... | ..................... | .................... | .................... | .................... |
| 21631 MT911552.1 | \|Severe acute respiratory syndrome coro | .................... | .................... | ..................... | .................... | .................... | .................... |
| 21632 MT911553.1 | \|Severe acute respiratory syndrome coro | .................... | .................... | ..................... | .................... | .................... | .................... |
| 21633 MT911554.1 | \|Severe acute respiratory syndrome coro | .................... | .................... | ..................... | .................... | .................... | .................... |
| 21634 MT911555.1 | \|Severe acute respiratory syndrome coro | .................... | .................... | ..................... | .................... | .................... | .................... |
| 21635 MT911556.1 | \|Severe acute respiratory syndrome coro | .................... | .................... | ..................... | .................... | .................... | .................... |
| 21636 MT911557.1 | \|Severe acute respiratory syndrome coro | .................... | .................... | ..................... | .................... | .................... | .................... |
| 21637 MT911558.1 | \|Severe acute respiratory syndrome coro | .................... | .................... | ..................... | .................... | .................... | .................... |
| 21638 MT911559.1 | \|Severe acute respiratory syndrome coro | .................... | .................... | ..................... | .................... | .................... | .................... |
| 21639 MT911560.1 | \|Severe acute respiratory syndrome coro | .................... | .................... | ..................... | .................... | .................... | .................... |
| 21640 MT911561.1 | \|Severe acute respiratory syndrome coro | .................... | .................... | ..................... | .................... | .................... | .................... |
| 21641 MT911562.1 | \|Severe acute respiratory syndrome coro | .................... | .................... | ..................... | .................... | .................... | .................... |
| 21642 MT911563.1 | \|Severe acute respiratory syndrome coro | .................... | .................... | ..................... | .................... | .................... | .................... |
| 21643 MT911564.1 | \|Severe acute respiratory syndrome coro | .................... | .................... | ..................... | .................... | .................... | .................... |
| 21644 MT911565.1 | \|Severe acute respiratory syndrome coro | .................... | .................... | ..................... | .................... | .................... | .................... |
| 21645 MT911566.1 | \|Severe acute respiratory syndrome coro | .................... | .................... | ..................... | .................... | .................... | .................... |
| 21646 MT911567.1 | \|Severe acute respiratory syndrome coro | .................... | .................... | ..................... | .................... | .................... | .................... |
| 21647 MT911568.1 | \|Severe acute respiratory syndrome coro | .................... | .................... | ..................... | .................... | .................... | .................... |
| 21648 MT911796.1 | \|Severe acute respiratory syndrome coro | .................... | .................... | ..................... | .................... | .................... | .................... |
| 21649 MT911797.1 | \|Severe acute respiratory syndrome coro | .................... | .................... | ..................... | .................... | .................... | .................... |
| 21650 MT911798.1 | \|Severe acute respiratory syndrome coro | .................... | .................... | ..................... | .................... | .................... | .................... |
| 21651 MT911799.1 | \|Severe acute respiratory syndrome coro | .................... | .................... | ..................... | .................... | .................... | .................... |
| 21652 MT911800.1 | \|Severe acute respiratory syndrome coro | .................... | .................... | ..................... | .................... | .................... | .................... |
| 21653 MT911801.1 | \|Severe acute respiratory syndrome coro | .................... | .................... | ..................... | .................... | .................... | .................... |
| 21654 MT911803.1 | \|Severe acute respiratory syndrome coro | .................... | .................... | ..................... | .................... | .................... | .................... |
| 21655 MT911804.1 | \|Severe acute respiratory syndrome coro | .................... | .................... | ..................... | .................... | .................... | .................... |
| 21656 MT911805.1 | \|Severe acute respiratory syndrome coro | .................... | .................... | ..................... | .................... | .................... | .................... |
| 21657 MT911806.1 | \|Severe acute respiratory syndrome coro | .................... | .................... | ..................... | .................... | .................... | .................... |
| 21658 MT911807.1 | \|Severe acute respiratory syndrome coro | .................... | .................... | ..................... | .................... | .................... | .................... |
| 21659 MT911808.1 | \|Severe acute respiratory syndrome coro | .................... | .................... | ..................... | .................... | .................... | .................... |
| 21660 MT911809.1 | \|Severe acute respiratory syndrome coro | .................... | .................... | ..................... | .................... | .................... | .................... |
| 21661 MT911810.1 | \|Severe acute respiratory syndrome coro | .................... | .................... | ..................... | .................... | .................... | .................... |
| 21662 MT911811.1 | \|Severe acute respiratory syndrome coro | .................... | .................... | ..................... | .................... | .................... | .................... |
| 21663 MT911812.1 | \|Severe acute respiratory syndrome coro | .................... | .................... | ..................... | .................... | .................... | .................... |
| 21664 MT911813.1 | \|Severe acute respiratory syndrome coro | .................... | .................... | ..................... | .................... | .................... | .................... |
| 21665 MT911814.1 | \|Severe acute respiratory syndrome coro | .................... | .................... | ..................... | .................... | .................... | .................... |
| 21666 MT911815.1 | \|Severe acute respiratory syndrome coro | .................... | .................... | ..................... | .................... | .................... | .................... |
| 21667 MT911816.1 | \|Severe acute respiratory syndrome coro | .................... | .................... | ..................... | .................... | .................... | .................... |
| 21668 MT911817.1 | \|Severe acute respiratory syndrome coro | .................... | .................... | ..................... | .................... | .................... | .................... |
| 21669 MT911818.1 | \|Severe acute respiratory syndrome coro | .................... | .................... | ..................... | .................... | .................... | .................... |
| 21670 MT911819.1 | \|Severe acute respiratory syndrome coro | .................... | .................... | ..................... | .................... | .................... | .................... |
| 21671 MT911820.1 | \|Severe acute respiratory syndrome coro | .................... | .................... | ..................... | .................... | .................... | .................... |
| 21672 MT911821.1 | \|Severe acute respiratory syndrome coro | .................... | .................... | ..................... | .................... | .................... | .................... |
| 21673 MT911822.1 | \|Severe acute respiratory syndrome coro | .................... | .................... | ..................... | .................... | .................... | .................... |
| 21674 MT911823.1 | \|Severe acute respiratory syndrome coro | .................... | .................... | ..................... | .................... | .................... | .................... |
| 21675 MT911824.1 | \|Severe acute respiratory syndrome coro | .................... | .................... | ..................... | .................... | .................... | .................... |
| 21676 MT911825.1 | \|Severe acute respiratory syndrome coro | .................... | .................... | ..................... | .................... | .................... | .................... |
| 21677 MT911827.1 | \|Severe acute respiratory syndrome coro | .................... | .................... | ..................... | .................... | .................... | .................... |
| 21678 MT911828.1 | \|Severe acute respiratory syndrome coro | .................... | .................... | ..................... | .................... | .................... | .................... |
| 21679 LC573273.1 | \|Severe acute respiratory syndrome coro | .................... | .................... | ...........T......... | .................... | .................... | .................... |
| 21680 LC573274.1 | \|Severe acute respiratory syndrome coro | .................... | .................... | ..................... | .................... | .................... | .................... |
| 21681 LC573275.1 | \|Severe acute respiratory syndrome coro | .................... | .................... | ..................... | .................... | .................... | .................... |
| 21682 LC573276.1 | \|Severe acute respiratory syndrome coro | .................... | .................... | ..................... | .................... | .................... | .................... |
| 21683 LC573277.1 | \|Severe acute respiratory syndrome coro | .................... | .................... | ..................... | .................... | .................... | .................... |
| 21684 LC573278.1 | \|Severe acute respiratory syndrome coro | .................... | .................... | ..................... | .................... | .................... | .................... |
| 21685 LC573279.1 | \|Severe acute respiratory syndrome coro | .................... | .................... | ..................... | .................... | .................... | .................... |
| 21686 LC573280.1 | \|Severe acute respiratory syndrome coro | .................... | .................... | ..................... | .................... | .................... | .................... |
| 21687 LC573281.1 | \|Severe acute respiratory syndrome coro | .................... | .................... | ..................... | .................... | .................... | .................... |
| 21688 LC573282.1 | \|Severe acute respiratory syndrome coro | .................... | .................... | ..................... | .................... | .................... | .................... |
| 21689 LC573283.1 | \|Severe acute respiratory syndrome coro | .................... | .................... | ..................... | .................... | .................... | .................... |
| 21690 LC573284.1 | \|Severe acute respiratory syndrome coro | .................... | .................... | ..................... | .................... | .................... | .................... |
| 21691 LC573285.1 | \|Severe acute respiratory syndrome coro | .................... | .................... | ..................... | .................... | .................... | .................... |

21692 LC573286.1 |Severe acute respiratory syndrome coro .................... .................... .................... .................... .................... ....................
21693 MT905416.1 |Severe acute respiratory syndrome coro .................... .................... .................... .................... .................... ....................
21694 MT905417.1 |Severe acute respiratory syndrome coro .................... .................... .................... .................... .................... ....................
21695 MT905418.1 |Severe acute respiratory syndrome coro .................... .................... .................... .................... .................... ....................
21696 MT905419.1 |Severe acute respiratory syndrome coro .................... .................... .................... .................... .................... ....................
21697 MT905420.1 |Severe acute respiratory syndrome coro .................... .................... .................... .................... .................... ....................
21698 MT905421.1 |Severe acute respiratory syndrome coro .................... .................... .................... .................... .................... ....................
21699 MT905422.1 |Severe acute respiratory syndrome coro .................... .................... .................... .................... .................... ....................
21700 MT905423.1 |Severe acute respiratory syndrome coro .................... .................... .................... .................... .................... ....................
21701 MT905424.1 |Severe acute respiratory syndrome coro .................... .................... .................... .................... .................... ....................
21702 MT905425.1 |Severe acute respiratory syndrome coro .................... .................... .................... .................... .................... ....................
21703 MT905426.1 |Severe acute respiratory syndrome coro .................... .................... .................... .................... .................... ....................
21704 MT905427.1 |Severe acute respiratory syndrome coro .................... .................... .................... .................... .................... ....................
21705 MT905428.1 |Severe acute respiratory syndrome coro .................... .................... .................... .................... .................... ....................
21706 MT905429.1 |Severe acute respiratory syndrome coro .................... .................... .................... .................... .................... ....................
21707 MT905430.1 |Severe acute respiratory syndrome coro .................... .................... .................... .................... .................... ....................
21708 MT905431.1 |Severe acute respiratory syndrome coro .................... .................... .................... .................... .................... ....................
21709 LR877186.1 |Severe acute respiratory syndrome coro .................... .................... .................... .................... .................... ....................
21710 MT479216.1 |Severe acute respiratory syndrome coro .................... .................... .................... .................... .................... ....................
21711 MT483867.1 |Severe acute respiratory syndrome coro .................... .................... .................... .................... .................... ....................
21712 MT483874.1 |Severe acute respiratory syndrome coro .................... .................... .................... .................... .................... ....................
21713 MT483875.1 |Severe acute respiratory syndrome coro .................... .................... .................... .................... .................... ....................
21714 MT483876.1 |Severe acute respiratory syndrome coro .................... .................... .................... .................... .................... ....................
21715 MT483877.1 |Severe acute respiratory syndrome coro .................... .................... .................... .................... .................... ....................
21716 MT483878.1 |Severe acute respiratory syndrome coro .................... .................... .................... .................... .................... ....................
21717 MT483879.1 |Severe acute respiratory syndrome coro .................... .................... .................... .................... .................... ....................
21718 MT483880.1 |Severe acute respiratory syndrome coro .................... .................... .................... .................... .................... ....................
21719 MT483881.1 |Severe acute respiratory syndrome coro .................... .................... .................... .................... .................... ....................
21720 MT893659.1 |Severe acute respiratory syndrome coro .................... .................... .................... .................... .................... ....................
21721 MT893661.1 |Severe acute respiratory syndrome coro .................... .................... .................... .................... .................... ....................
21723 MT893664.1 |Severe acute respiratory syndrome coro .................... .................... .................... .................... .................... ....................
21724 MT893667.1 |Severe acute respiratory syndrome coro .................... .................... .................... .................... .................... ....................
21725 MT893668.1 |Severe acute respiratory syndrome coro .................... .................... .................... .................... .................... ....................
21726 MT893669.1 |Severe acute respiratory syndrome coro .................... .................... .................... .................... .................... ....................
21727 MT893670.1 |Severe acute respiratory syndrome coro .................... .................... .................... .................... .................... ....................
21729 MT893672.1 |Severe acute respiratory syndrome coro .................... .................... .................... .................... .................... ....................
21730 MT893673.1 |Severe acute respiratory syndrome coro .................... .................... .................... .................... .................... ....................
21731 MT893674.1 |Severe acute respiratory syndrome coro .................... .................... .................... .................... .................... ....................
21732 MT893675.1 |Severe acute respiratory syndrome coro .................... .................... .................... .................... .................... ....................
21733 MT893677.1 |Severe acute respiratory syndrome coro .................... .................... .................... .................... .................... ....................
21734 MT893679.1 |Severe acute respiratory syndrome coro .................... .................... .................... .................... .................... ....................
21735 MT893680.1 |Severe acute respiratory syndrome coro .................... .................... .................... .................... .................... ....................
21736 MT893681.1 |Severe acute respiratory syndrome coro .................... .................... .................... .................... .................... ....................
21737 MT893683.1 |Severe acute respiratory syndrome coro .................... .................... .................... .................... .................... ....................
21738 MT893684.1 |Severe acute respiratory syndrome coro .................... .................... .................... .................... .................... ....................
21739 MT893685.1 |Severe acute respiratory syndrome coro .................... .................... .................... .................... .................... ....................
21740 MT893686.1 |Severe acute respiratory syndrome coro .................... .................... .................... .................... .................... ....................
21741 MT893688.1 |Severe acute respiratory syndrome coro .................... .................... .................... .................... .................... ....................
21742 MT893692.1 |Severe acute respiratory syndrome coro .................... .................... .................... .................... .................... ....................
21743 MT897260.1 |Severe acute respiratory syndrome coro .................... .................... .................... .................... .................... ....................
21744 MT897261.1 |Severe acute respiratory syndrome coro .................... .................... .................... .................... .................... ....................
21745 MT897459.1 |Severe acute respiratory syndrome coro .................... .................... .................... .................... .................... ....................
21746 MT897460.1 |Severe acute respiratory syndrome coro .................... .................... .................... .................... .................... ....................
21747 MT897461.1 |Severe acute respiratory syndrome coro .................... .................... .................... .................... .................... ....................
21748 MT897462.1 |Severe acute respiratory syndrome coro .................... .................... .................... .................... .................... ....................
21749 MT897463.1 |Severe acute respiratory syndrome coro .................... .................... .................... .................... .................... ....................
21750 MT897464.1 |Severe acute respiratory syndrome coro .................... .................... .................... .................... .................... ....................
21751 MT897465.1 |Severe acute respiratory syndrome coro .................... .................... .................... .................... .................... ....................
21752 MT897466.1 |Severe acute respiratory syndrome coro .................... .................... .................... .................... .................... ....................
21753 MT890210.1 |Severe acute respiratory syndrome coro .................... .................... .................... .................... .................... ....................
21755 MT890230.1 |Severe acute respiratory syndrome coro .................... .................... .................... .................... N................... ....................
21761 MT890277.1 |Severe acute respiratory syndrome coro .................... .................... .................... .................... .................... ....................
21763 MT890280.1 |Severe acute respiratory syndrome coro .................... .................... .................... .................... .................... ....................
21765 MT890285.1 |Severe acute respiratory syndrome coro .................... .................... .................... .................... .................... ....................
21766 MT890286.1 |Severe acute respiratory syndrome coro .................... .................... .................... .................... .................... ....................
21767 MT890288.1 |Severe acute respiratory syndrome coro .................... .................... .................... .................... .................... ....................
21768 MT890289.1 |Severe acute respiratory syndrome coro .................... .................... .................... .................... .................... ....................
21769 MT890290.1 |Severe acute respiratory syndrome coro .................... .................... .................... .................... .................... ....................
21770 MT890292.1 |Severe acute respiratory syndrome coro .................... .................... .................... .................... .................... ....................
21771 MT890293.1 |Severe acute respiratory syndrome coro .................... .................... .................... .................... .................... ....................
21772 MT890294.1 |Severe acute respiratory syndrome coro .................... .................... .................... .................... .................... ....................
21773 MT890295.1 |Severe acute respiratory syndrome coro .................... .................... .................... .................... .................... ....................
21774 MT890296.1 |Severe acute respiratory syndrome coro .................... .................... .................... .................... .................... ....................
21775 MT890301.1 |Severe acute respiratory syndrome coro .................... .................... .................... .................... .................... ....................
21776 MT890302.1 |Severe acute respiratory syndrome coro .................... .................... .................... .................... .................... ....................
21777 MT890309.1 |Severe acute respiratory syndrome coro .................... .................... .................... .................... .................... ....................
21778 MT890310.1 |Severe acute respiratory syndrome coro .................... .................... .................... .................... .................... ....................
21779 MT890311.1 |Severe acute respiratory syndrome coro .................... .................... .................... .................... .................... ....................

21780 MT890313.1 |Severe acute respiratory syndrome coro ···················· ···················· ···················· ···················· ···················· ····················
21781 MT890316.1 |Severe acute respiratory syndrome coro ···················· ···················· ···················· ···················· ···················· ····················
21782 MT890320.1 |Severe acute respiratory syndrome coro ···················· ·············A······ ···················· ···················· ···················· ····················
21783 MT890321.1 |Severe acute respiratory syndrome coro ···················· ···················· ···················· ···················· ···················· ····················
21784 MT890322.1 |Severe acute respiratory syndrome coro ···················· ···················· ···················· ···················· ···················· ····················
21785 MT890330.1 |Severe acute respiratory syndrome coro ···················· ···················· ···················· ···················· ···················· ····················
21786 MT890335.1 |Severe acute respiratory syndrome coro ···················· ···················· ···················· ···················· ···················· ····················
21787 MT890336.1 |Severe acute respiratory syndrome coro ···················· ···················· ···················· ···················· ···················· ····················
21788 MT890337.1 |Severe acute respiratory syndrome coro ···················· ···················· ···················· ···················· ···················· ····················
21789 MT890338.1 |Severe acute respiratory syndrome coro ···················· ···················· ···················· ···················· ···················· ····················
21790 MT890340.1 |Severe acute respiratory syndrome coro ···················· ···················· ···················· ···················· ···················· ····················
21791 MT890344.1 |Severe acute respiratory syndrome coro ···················· ···················· ···················· ···················· ···················· ····················
21792 MT890345.1 |Severe acute respiratory syndrome coro ···················· ···················· ···················· ···················· ···················· ····················
21793 MT890346.1 |Severe acute respiratory syndrome coro ···················· ···················· ···················· ···················· ···················· ····················
21794 MT890347.1 |Severe acute respiratory syndrome coro ···················· ···················· ···················· ···················· ···················· ····················
21795 MT890348.1 |Severe acute respiratory syndrome coro ···················· ···················· ···················· ···················· ···················· ····················
21796 MT890349.1 |Severe acute respiratory syndrome coro ···················· ···················· ···················· ···················· ···················· ····················
21797 MT890352.1 |Severe acute respiratory syndrome coro ···················· ···················· ···················· ···················· ···················· ····················
21798 MT890353.1 |Severe acute respiratory syndrome coro ···················· ···················· ···················· ···················· ···················· ····················
21799 MT890355.1 |Severe acute respiratory syndrome coro ···················· ···················· ···················· ···················· ···················· ····················
21800 MT890356.1 |Severe acute respiratory syndrome coro ···················· ····A··············· ···················· ···················· ···················· ····················
21801 MT890357.1 |Severe acute respiratory syndrome coro ···················· ····A··············· ···················· ···················· ···················· ····················
21802 MT890358.1 |Severe acute respiratory syndrome coro ···················· ···················· ···················· ···················· ···················· ····················
21803 MT890462.1 |Severe acute respiratory syndrome coro ···················· ···················· ···················· ···················· ···················· ····················
21804 MT890669.1 |Severe acute respiratory syndrome coro ···················· ···················· ···················· ···················· ···················· ····················
21805 MT892963.1 |Severe acute respiratory syndrome coro ···················· ···················· ···················· ···················· ···················· ····················
21806 MT892964.1 |Severe acute respiratory syndrome coro ···················· ···················· ···················· ···················· ···················· ····················
21807 MT892965.1 |Severe acute respiratory syndrome coro ···················· ···················· ···················· ···················· ···················· ····················
21808 MT892966.1 |Severe acute respiratory syndrome coro ···················· ···················· ···················· ···················· ···················· ····················
21809 MT892967.1 |Severe acute respiratory syndrome coro ···················· ···················· ···················· ···················· ···················· ····················
21810 MT892968.1 |Severe acute respiratory syndrome coro ···················· ···················· ···················· ···················· ···················· ····················
21811 MT892969.1 |Severe acute respiratory syndrome coro ···················· ···················· ···················· ···················· ···················· ····················
21812 MT892970.1 |Severe acute respiratory syndrome coro ···················· ···················· ···················· ···················· ··N················· ····················
21813 MT892971.1 |Severe acute respiratory syndrome coro ···················· ···················· ···················· ···················· ···················· ····················
21814 MT892972.1 |Severe acute respiratory syndrome coro ···················· ···················· ···················· ···················· ···················· ····················
21815 MT892973.1 |Severe acute respiratory syndrome coro ···················· ···················· ···················· ···················· ···················· ····················
21816 MT892974.1 |Severe acute respiratory syndrome coro ···················· ·A·················· ···················· ···················· ···················· ····················
21817 MT892975.1 |Severe acute respiratory syndrome coro ···················· ···················· ···················· ···················· ···················· ····················
21818 MT892976.1 |Severe acute respiratory syndrome coro ···················· ···················· ···················· ···················· ···················· ····················
21819 MT892977.1 |Severe acute respiratory syndrome coro ···················· ···················· ···················· ···················· ···················· ····················
21820 MT892978.1 |Severe acute respiratory syndrome coro ···················· ···················· ···················· ···················· ···················· ····················
21821 MT892979.1 |Severe acute respiratory syndrome coro ···················· ···················· ···················· ···················· ···················· ····················
21822 MT892980.1 |Severe acute respiratory syndrome coro ···················· ···················· ···················· ···················· ···················· ····················
21823 MT892981.1 |Severe acute respiratory syndrome coro ···················· ···················· ···················· ···················· ···················· ····················
21824 MT892982.1 |Severe acute respiratory syndrome coro ···················· ·A·················· ···················· ···················· ···················· ····················
21825 MT892983.1 |Severe acute respiratory syndrome coro ···················· ···················· ···················· ···················· ···················· ····················
21826 MT892984.1 |Severe acute respiratory syndrome coro ···················· ···················· ···················· ···················· ···················· ····················
21827 MT892985.1 |Severe acute respiratory syndrome coro ···················· ···················· ···················· ···················· ···················· ····················
21828 MT892986.1 |Severe acute respiratory syndrome coro ···················· ···················· ···················· ···················· ···················· ····················
21829 MT892987.1 |Severe acute respiratory syndrome coro ···················· ···················· ···················· ···················· ···················· ····················
21830 MT892988.1 |Severe acute respiratory syndrome coro ···················· ···················· ···················· ···················· ···················· ····················
21831 MT892989.1 |Severe acute respiratory syndrome coro ···················· ···················· ···················· ···················· ···················· ····················
21832 MT892990.1 |Severe acute respiratory syndrome coro ···················· ···················· ···················· ···················· ···················· ····················
21833 MT892991.1 |Severe acute respiratory syndrome coro ···················· ···················· ···················· ···················· ···················· ····················
21834 MT892992.1 |Severe acute respiratory syndrome coro ···················· ···················· ···················· ···················· ···················· ····················
21835 MT892993.1 |Severe acute respiratory syndrome coro ···················· ···················· ···················· ···················· ···················· ····················
21836 MT892994.1 |Severe acute respiratory syndrome coro ···················· ···················· ···················· ···················· ···················· ····················
21837 MT892995.1 |Severe acute respiratory syndrome coro ···················· ···················· ···················· ···················· ···················· ····················
21838 MT892996.1 |Severe acute respiratory syndrome coro ···················· ···················· ···················· ···················· ···················· ····················
21839 MT892997.1 |Severe acute respiratory syndrome coro ···················· ···················· ···················· ···················· ···················· ····················
21840 MT892998.1 |Severe acute respiratory syndrome coro ···················· ···················· ···················· ···················· ···················· ····················
21841 MT892999.1 |Severe acute respiratory syndrome coro ···················· ···················· ···················· ···················· ···················· ····················
21842 MT893000.1 |Severe acute respiratory syndrome coro ···················· ···················· ···················· ···················· ······T············· ····················
21843 MT893001.1 |Severe acute respiratory syndrome coro ···················· ···················· ···················· ···················· ···················· ····················
21844 MT893002.1 |Severe acute respiratory syndrome coro ···················· ···················· ···················· ···················· ···················· ····················
21845 MT893003.1 |Severe acute respiratory syndrome coro ···················· ···················· ···················· ···················· ···················· ····················
21846 MT893004.1 |Severe acute respiratory syndrome coro ···················· ···················· ···················· ···················· ···················· ····················
21847 MT893005.1 |Severe acute respiratory syndrome coro ···················· ···················· ···················· ···················· ···················· ····················
21848 MT893006.1 |Severe acute respiratory syndrome coro ···················· ···················· ···················· ···················· ···················· ····················
21849 MT893007.1 |Severe acute respiratory syndrome coro ···················· ···················· ···················· ···················· ···················· ····················
21850 MT893008.1 |Severe acute respiratory syndrome coro ···················· ···················· ···················· ···················· ···················· ·····M··············
21851 MT893009.1 |Severe acute respiratory syndrome coro ···················· ···················· ···················· ···················· ···················· ····················
21852 MT893010.1 |Severe acute respiratory syndrome coro ···················· ···················· ···················· ···················· ···················· ····················
21853 MT893011.1 |Severe acute respiratory syndrome coro ···················· ····G··············· ···················· ···················· ···················· ····················
21854 MT893012.1 |Severe acute respiratory syndrome coro ···················· ···················· ···················· ···················· ···················· ····················
21855 MT893013.1 |Severe acute respiratory syndrome coro ···················· ···················· ···················· ···················· ···················· ····················
21856 MT893014.1 |Severe acute respiratory syndrome coro ···················· ····G··············· ···················· ···················· ···················· ····················
21857 MT893015.1 |Severe acute respiratory syndrome coro ···················· ···················· ···················· ···················· ···················· ····················

21858 MT893016.1 |Severe acute respiratory syndrome coro .................... .................... ..................... .................... .................... ....................
21859 MT893017.1 |Severe acute respiratory syndrome coro .................... .................... ..................... .................... .................... ....................
21860 MT893018.1 |Severe acute respiratory syndrome coro .................... .................... ..................... .................... .................... ....................
21861 MT893019.1 |Severe acute respiratory syndrome coro .................... .................... ..................... .................... .................... ....................
21862 MT034054.1 |Severe acute respiratory syndrome coro .................... .................... ..................... .................... .................... ....................
21863 MT889692.1 |Severe acute respiratory syndrome coro .................... .A.................. ..................... .................... .................... ....................
21864 MT883499.1 |Severe acute respiratory syndrome coro .................... .................... ..................... .................... .................... ....................
21865 MT883500.1 |Severe acute respiratory syndrome coro .................... .................... ..................... .................... .................... ....................
21866 MT883501.1 |Severe acute respiratory syndrome coro .................... .................... ..................... .................... .................... ....................
21867 MT883502.1 |Severe acute respiratory syndrome coro .................... .................... ..................... .................... .................... ....................
21868 MT883503.1 |Severe acute respiratory syndrome coro .................... .................... ..................... .................... .................... ....................
21869 MT883504.1 |Severe acute respiratory syndrome coro .................... .................... ..................... .................... .................... ....................
21870 MT883505.1 |Severe acute respiratory syndrome coro .................... .................... ..................... .................... .................... ....................
21871 MT886288.1 |Severe acute respiratory syndrome coro .................... .................... ..................... .................... .................... ....................
21872 MT886289.1 |Severe acute respiratory syndrome coro .................... .................... ..................... .................... .................... ....................
21873 MT886290.1 |Severe acute respiratory syndrome coro .................... .................... ..................... .................... .................... ....................
21874 MT886292.1 |Severe acute respiratory syndrome coro .................... .................... ..................... .................... .................... ....................
21875 MT886293.1 |Severe acute respiratory syndrome coro .................... .................... ..................... .................... .................... ....................
21876 MT886294.1 |Severe acute respiratory syndrome coro .................... .................... ..................... .................... .................... ....................
21877 MT886295.1 |Severe acute respiratory syndrome coro .................... .................... ..................... .................... .................... ....................
21878 MT886296.1 |Severe acute respiratory syndrome coro .................... .................... ..................... .................... .................... ....................
21879 MT886297.1 |Severe acute respiratory syndrome coro .................... .................... ..................... .................... .................... ....................
21880 MT886298.1 |Severe acute respiratory syndrome coro .................... .................... ..................... .................... .................... ....................
21881 MT886299.1 |Severe acute respiratory syndrome coro .................... .................... ..................... .................... .................... ....................
21882 MT886300.1 |Severe acute respiratory syndrome coro .................... .................... ..................... .................... .................... ....................
21883 MT886301.1 |Severe acute respiratory syndrome coro .................... .................... ..................... .................... .................... ....................
21884 MT886302.1 |Severe acute respiratory syndrome coro .................... .................... ..................... .................... .................... ....................
21885 MT886303.1 |Severe acute respiratory syndrome coro .................... .................... ..................... .................... .................... ....................
21886 MT886304.1 |Severe acute respiratory syndrome coro .................... .................... ..................... .................... .................... ....................
21887 MT886305.1 |Severe acute respiratory syndrome coro .................... .................... ..................... .................... .................... ....................
21888 MT886306.1 |Severe acute respiratory syndrome coro .................... .................... ..................... .................... .................... ....................
21889 MT886307.1 |Severe acute respiratory syndrome coro .................... .................... ..................... .................... .................... ....................
21890 MT886308.1 |Severe acute respiratory syndrome coro .................... .................... ..................... .................... .................... ....................
21891 MT886309.1 |Severe acute respiratory syndrome coro .................... .................... ..................... .................... .................... ....................
21892 MT886310.1 |Severe acute respiratory syndrome coro .................... .................... ..................... .................... .................... ....................
21893 MT886311.1 |Severe acute respiratory syndrome coro .................... .................... ..................... .................... .................... ....................
21894 MT886312.1 |Severe acute respiratory syndrome coro .................... .................... ..................... .................... .................... ....................
21895 MT886313.1 |Severe acute respiratory syndrome coro .................... .................... ..................... .................... .................... ....................
21896 MT886314.1 |Severe acute respiratory syndrome coro .................... .................... ..................... .................... .................... ....................
21897 MT886315.1 |Severe acute respiratory syndrome coro .................... .................... ..................... .................... .................... ....................
21898 MT886316.1 |Severe acute respiratory syndrome coro .................... .................... ..................... .................... .................... ....................
21899 MT886317.1 |Severe acute respiratory syndrome coro .................... .................... ..................... .................... .................... ....................
21900 MT886318.1 |Severe acute respiratory syndrome coro .................... .................... ..................... .................... .................... ....................
21901 MT886321.1 |Severe acute respiratory syndrome coro .................... .................... ..................... .................... .................... ....................
21902 MT886322.1 |Severe acute respiratory syndrome coro .................... .................... ..................... .................... .................... ....................
21903 MT886323.1 |Severe acute respiratory syndrome coro .................... .................... ..................... .................... .................... ....................
21904 MT886324.1 |Severe acute respiratory syndrome coro .................... .................... ..................... .................... .................... ....................
21905 MT886325.1 |Severe acute respiratory syndrome coro .................... .................... ..................... .................... .................... ....................
21906 MT886326.1 |Severe acute respiratory syndrome coro .................... .................... ..................... .................... .................... ....................
21907 MT886327.1 |Severe acute respiratory syndrome coro .................... .................... ..................... .................... .................... ....................
21908 MT886328.1 |Severe acute respiratory syndrome coro .................... .................... ..................... .................... .................... ....................
21909 MT886329.1 |Severe acute respiratory syndrome coro .................... .................... ..................... .................... .................... ....................
21910 MT886330.1 |Severe acute respiratory syndrome coro .................... .................... ..................... .................... .................... ....................
21911 MT886331.1 |Severe acute respiratory syndrome coro .................... .................... ..................... .................... .................... ....................
21912 MT886332.1 |Severe acute respiratory syndrome coro .................... .................... ..................... .................... .................... ....................
21913 MT886333.1 |Severe acute respiratory syndrome coro .................... .................... ..................... .................... .................... ....................
21914 MT886334.1 |Severe acute respiratory syndrome coro .................... .................... ..................... .................... .................... ....................
21915 MT886335.1 |Severe acute respiratory syndrome coro .................... .................... ..................... .................... .................... ....................
21916 MT886336.1 |Severe acute respiratory syndrome coro .................... .................... ..................... .................... .................... ....................
21917 MT886337.1 |Severe acute respiratory syndrome coro .................... .................... ..................... .................... .................... ....................
21918 MT886338.1 |Severe acute respiratory syndrome coro .................... .................... ..................... .................... .................... ....................
21919 MT886339.1 |Severe acute respiratory syndrome coro .................... .................... ..................... .................... .................... ....................
21920 MT886340.1 |Severe acute respiratory syndrome coro .................... .................... ..................... .................... .................... ....................
21921 MT886341.1 |Severe acute respiratory syndrome coro .................... .................... ..................... .................... .................... ....................
21922 MT886342.1 |Severe acute respiratory syndrome coro .................... .................... ..................... .................... .................... ....................
21923 MT886343.1 |Severe acute respiratory syndrome coro .................... .................... ..................... .................... .................... ....................
21924 MT886344.1 |Severe acute respiratory syndrome coro .................... .................... ..................... .................... .................... ....................
21925 MT886345.1 |Severe acute respiratory syndrome coro .................... .................... ..................... .................... .................... ....................
21926 MT886346.1 |Severe acute respiratory syndrome coro .................... .................... ..................... .................... .................... ....................
21927 MT886347.1 |Severe acute respiratory syndrome coro .................... .................... ..................... .................... .................... ....................
21928 MT886348.1 |Severe acute respiratory syndrome coro .................... .................... ..................... .................... .................... ....................
21929 MT886349.1 |Severe acute respiratory syndrome coro .................... .................... ..................... .................... .................... ....................
21930 MT886350.1 |Severe acute respiratory syndrome coro .................... .................... ..................... .................... .................... ....................
21932 MT886352.1 |Severe acute respiratory syndrome coro .................... .................... ..................... .................... .................... ....................
21933 MT886353.1 |Severe acute respiratory syndrome coro .................... .................... ..................... .................... .................... ....................
21934 MT886354.1 |Severe acute respiratory syndrome coro .................... .................... ..................... .................... .................... ....................
21935 MT886355.1 |Severe acute respiratory syndrome coro .................... .................... ..................... .................... .................... ....................
21936 MT886356.1 |Severe acute respiratory syndrome coro .................... .................... ..................... .................... .................... ....................

| | | | | | | | | |
|---|---|---|---|---|---|---|---|---|
| 21937 | MT886357.1 | Severe acute respiratory syndrome coro | ···················· | ···················· | ····················· | ···················· | ···················· | ···················· |
| 21938 | MT886358.1 | Severe acute respiratory syndrome coro | ···················· | ···················· | ····················· | ···················· | ···················· | ···················· |
| 21939 | MT886359.1 | Severe acute respiratory syndrome coro | ···················· | ···················· | ····················· | ···················· | ···················· | ···················· |
| 21940 | MT886360.1 | Severe acute respiratory syndrome coro | ···················· | ···················· | ····················· | ···················· | ···················· | ···················· |
| 21941 | MT886362.1 | Severe acute respiratory syndrome coro | ···················· | ···················· | ····················· | ···················· | ···················· | ···················· |
| 21942 | MT886363.1 | Severe acute respiratory syndrome coro | ···················· | ···················· | ····················· | ···················· | ···················· | ···················· |
| 21943 | MT886364.1 | Severe acute respiratory syndrome coro | ···················· | ···················· | ····················· | ···················· | ···················· | ···················· |
| 21944 | MT886365.1 | Severe acute respiratory syndrome coro | ···················· | ···················· | ····················· | ···················· | ···················· | ···················· |
| 21945 | MT886366.1 | Severe acute respiratory syndrome coro | ···················· | ···················· | ····················· | ···················· | ···················· | ···················· |
| 21946 | MT886367.1 | Severe acute respiratory syndrome coro | ···················· | ···················· | ····················· | ···················· | ···················· | ···················· |
| 21947 | MT886368.1 | Severe acute respiratory syndrome coro | ···················· | ···················· | ····················· | ···················· | ···················· | ···················· |
| 21948 | MT886369.1 | Severe acute respiratory syndrome coro | ···················· | ···················· | ····················· | ···················· | ···················· | ···················· |
| 21949 | MT886370.1 | Severe acute respiratory syndrome coro | ···················· | ···················· | ····················· | ···················· | ···················· | ···················· |
| 21950 | MT886371.1 | Severe acute respiratory syndrome coro | ···················· | ···················· | ····················· | ···················· | ···················· | ···················· |
| 21951 | MT886372.1 | Severe acute respiratory syndrome coro | ···················· | ···················· | ····················· | ···················· | ···················· | ···················· |
| 21952 | MT886373.1 | Severe acute respiratory syndrome coro | ···················· | ···················· | ····················· | ···················· | ···················· | ···················· |
| 21953 | MT886375.1 | Severe acute respiratory syndrome coro | ···················· | ···················· | ····················· | ···················· | ···················· | ···················· |
| 21954 | MT886376.1 | Severe acute respiratory syndrome coro | ···················· | ···················· | ····················· | ···················· | ···················· | ···················· |
| 21955 | MT886377.1 | Severe acute respiratory syndrome coro | ···················· | ···················· | ····················· | ···················· | ···················· | ···················· |
| 21956 | MT886378.1 | Severe acute respiratory syndrome coro | ···················· | ···················· | ····················· | ···················· | ···················· | ···················· |
| 21957 | MT886379.1 | Severe acute respiratory syndrome coro | ···················· | ···················· | ····················· | ···················· | ···················· | ···················· |
| 21958 | MT886380.1 | Severe acute respiratory syndrome coro | ···················· | ···················· | ····················· | ···················· | ···················· | ···················· |
| 21959 | MT886381.1 | Severe acute respiratory syndrome coro | ···················· | ···················· | ····················· | ···················· | ···················· | ···················· |
| 21960 | MT886382.1 | Severe acute respiratory syndrome coro | ···················· | ···················· | ····················· | ···················· | ···················· | ···················· |
| 21961 | MT886383.1 | Severe acute respiratory syndrome coro | ···················· | ···················· | ····················· | ···················· | ···················· | ···················· |
| 21962 | MT886384.1 | Severe acute respiratory syndrome coro | ···················· | ···················· | ····················· | ···················· | ···················· | ···················· |
| 21963 | MT886385.1 | Severe acute respiratory syndrome coro | ···················· | ···················· | ····················· | ···················· | ···················· | ···················· |
| 21964 | MT886386.1 | Severe acute respiratory syndrome coro | ···················· | ···················· | ····················· | ···················· | ···················· | ···················· |
| 21965 | MT886407.1 | Severe acute respiratory syndrome coro | ···················· | ···················· | ····················· | ···················· | ···················· | ···················· |
| 21966 | MT886409.1 | Severe acute respiratory syndrome coro | ···················· | ···················· | ····················· | ···················· | ···················· | ···················· |
| 21967 | MT886410.1 | Severe acute respiratory syndrome coro | ···················· | ···················· | ····················· | ···················· | ···················· | ···················· |
| 21968 | MT886413.1 | Severe acute respiratory syndrome coro | ···················· | ···················· | ····················· | ···················· | ···················· | ···················· |
| 21969 | MT879619.1 | Severe acute respiratory syndrome coro | ···················· | ···················· | ····················· | ···················· | ···················· | ···················· |
| 21970 | MT880873.1 | Severe acute respiratory syndrome coro | ···················· | ···················· | ····················· | ···················· | ···················· | ···················· |
| 21971 | MT880874.1 | Severe acute respiratory syndrome coro | ···················· | ···················· | ····················· | ···················· | ···················· | ···················· |
| 21972 | MT880875.1 | Severe acute respiratory syndrome coro | ···················· | ···················· | ····················· | ···················· | ···················· | ···················· |
| 21973 | MT880876.1 | Severe acute respiratory syndrome coro | ···················· | ···················· | ····················· | ···················· | ···················· | ···················· |
| 21974 | MT880877.1 | Severe acute respiratory syndrome coro | ···················· | ···················· | ····················· | ···················· | ···················· | ···················· |
| 21975 | MT880878.1 | Severe acute respiratory syndrome coro | ···················· | ···················· | ····················· | ···················· | ···················· | ···················· |
| 21976 | MT880879.1 | Severe acute respiratory syndrome coro | ···················· | ···················· | ····················· | ···················· | ···················· | ···················· |
| 21977 | MT880880.1 | Severe acute respiratory syndrome coro | ···················· | ···················· | ····················· | ···················· | ···················· | ···················· |
| 21978 | MT880881.1 | Severe acute respiratory syndrome coro | ···················· | ···················· | ····················· | ···················· | ···················· | ···················· |
| 21979 | MT880882.1 | Severe acute respiratory syndrome coro | ···················· | ···················· | ····················· | ···················· | ···················· | ···················· |
| 21980 | MT880883.1 | Severe acute respiratory syndrome coro | ···················· | ···················· | ····················· | ···················· | ···················· | ···················· |
| 21981 | MT880884.1 | Severe acute respiratory syndrome coro | ···················· | ···················· | ····················· | ···················· | ···················· | ···················· |
| 21982 | MT880885.1 | Severe acute respiratory syndrome coro | ···················· | ···················· | ····················· | ···················· | ···················· | ···················· |
| 21983 | MT880886.1 | Severe acute respiratory syndrome coro | ···················· | ···················· | ····················· | ···················· | ···················· | ···················· |
| 21984 | MT880887.1 | Severe acute respiratory syndrome coro | ···················· | ···················· | ····················· | ···················· | ···················· | ···················· |
| 21985 | MT880888.1 | Severe acute respiratory syndrome coro | ···················· | ···················· | ····················· | ···················· | ···················· | ···················· |
| 21986 | MT882310.1 | Severe acute respiratory syndrome coro | ···················· | ···················· | ····················· | ···················· | ···················· | ···················· |
| 21987 | MT882311.1 | Severe acute respiratory syndrome coro | ···················· | ···················· | ····················· | ···················· | ···················· | ···················· |
| 21988 | MT882312.1 | Severe acute respiratory syndrome coro | ············Y······· | ···················· | ····················· | ···················· | ···················· | ···················· |
| 21989 | MT882313.1 | Severe acute respiratory syndrome coro | ···················· | ···················· | ····················· | ···················· | ···················· | ···················· |
| 21990 | MT882314.1 | Severe acute respiratory syndrome coro | ···················· | ···················· | ····················· | ···················· | ···················· | ···················· |
| 21991 | MT882315.1 | Severe acute respiratory syndrome coro | ···················· | ···················· | ····················· | ···················· | ···················· | ···················· |
| 21992 | MT882316.1 | Severe acute respiratory syndrome coro | ···················· | ···················· | ····················· | ···················· | ···················· | ···················· |
| 21993 | MT882317.1 | Severe acute respiratory syndrome coro | ···················· | ···················· | ····················· | ···················· | ···················· | ···················· |
| 21994 | MT882318.1 | Severe acute respiratory syndrome coro | ···················· | ···················· | ····················· | ···················· | ···················· | ···················· |
| 21995 | MT882319.1 | Severe acute respiratory syndrome coro | ···················· | ···················· | ····················· | ···················· | ···················· | ···················· |
| 21996 | MT882320.1 | Severe acute respiratory syndrome coro | ···················· | ···················· | ····················· | ···················· | ···················· | ···················· |
| 21997 | MT882321.1 | Severe acute respiratory syndrome coro | ···················· | ···················· | ····················· | ···················· | ···················· | ···················· |
| 21998 | MT882322.1 | Severe acute respiratory syndrome coro | ···················· | ···················· | ····················· | ···················· | ···················· | ···················· |
| 21999 | MT882323.1 | Severe acute respiratory syndrome coro | ···················· | ···················· | ····················· | ···················· | ···················· | ···················· |
| 22000 | MT882324.1 | Severe acute respiratory syndrome coro | ···················· | ···················· | ····················· | ···················· | ···················· | ···················· |
| 22001 | MT882325.1 | Severe acute respiratory syndrome coro | ···················· | ···················· | ····················· | ···················· | ···················· | ···················· |
| 22002 | LR877181.1 | Severe acute respiratory syndrome coro | ···················· | ···················· | ····················· | ···················· | ···················· | ···················· |
| 22003 | LR877182.1 | Severe acute respiratory syndrome coro | ···················· | ···················· | ····················· | ···················· | ···················· | ···················· |
| 22004 | LR877183.1 | Severe acute respiratory syndrome coro | ···················· | ···················· | ····················· | ···················· | ···················· | ···················· |
| 22006 | MT873050.1 | Severe acute respiratory syndrome coro | ···················· | ···················· | ····················· | ···················· | ···················· | ···················· |
| 22007 | MT873051.1 | Severe acute respiratory syndrome coro | ···················· | ···················· | ····················· | ···················· | ···················· | ···················· |
| 22008 | MT873052.1 | Severe acute respiratory syndrome coro | ···················· | ···················· | ····················· | ···················· | ···················· | ···················· |
| 22009 | MT873053.1 | Severe acute respiratory syndrome coro | ···················· | ···················· | ····················· | ···················· | ···················· | ···················· |
| 22010 | MT873055.1 | Severe acute respiratory syndrome coro | ···················· | ···················· | ····················· | ···················· | ···················· | ···················· |
| 22011 | MT873056.1 | Severe acute respiratory syndrome coro | ···················· | ···················· | ····················· | ···················· | ···················· | ···················· |
| 22012 | MT873057.1 | Severe acute respiratory syndrome coro | ···················· | ···················· | ····················· | ···················· | ···················· | ···················· |
| 22013 | MT873058.1 | Severe acute respiratory syndrome coro | ···················· | ···················· | ····················· | ···················· | ···················· | ···················· |
| 22014 | MT873059.1 | Severe acute respiratory syndrome coro | ···················· | ···················· | ····················· | ···················· | ···················· | ···················· |
| 22015 | MT873060.1 | Severe acute respiratory syndrome coro | ···················· | ···················· | ····················· | ···················· | ···················· | ···················· |

| | | | | | | | | | |
|---|---|---|---|---|---|---|---|---|---|
| 22016 | MT873061.1 | \|Severe acute respiratory syndrome coro | .................... | .................... | .................... | .................... | .................... | .................... | |
| 22017 | MT873062.1 | \|Severe acute respiratory syndrome coro | .................... | .................... | .................... | .................... | .................... | .................... | |
| 22018 | MT873063.1 | \|Severe acute respiratory syndrome coro | .................... | .................... | .................... | .................... | .................... | .................... | |
| 22019 | MT873064.1 | \|Severe acute respiratory syndrome coro | .................... | .................... | .................... | .................... | .................... | .................... | |
| 22020 | MT873065.1 | \|Severe acute respiratory syndrome coro | .................... | .................... | .................... | .................... | .................... | .................... | |
| 22021 | MT873066.1 | \|Severe acute respiratory syndrome coro | .................... | .................... | .................... | .................... | .................... | .................... | |
| 22022 | MT873067.1 | \|Severe acute respiratory syndrome coro | .................... | .................... | .................... | .................... | .................... | .................... | |
| 22023 | MT873068.1 | \|Severe acute respiratory syndrome coro | .................... | .................... | .................... | .................... | .................... | .................... | |
| 22024 | MT873069.1 | \|Severe acute respiratory syndrome coro | .................... | .................... | .................... | .................... | .................... | .................... | |
| 22025 | MT873070.1 | \|Severe acute respiratory syndrome coro | .................... | .................... | .................... | .................... | .................... | .................... | |
| 22026 | MT873071.1 | \|Severe acute respiratory syndrome coro | .................... | .................... | .................... | .................... | .................... | .................... | |
| 22027 | MT873072.1 | \|Severe acute respiratory syndrome coro | .................... | .................... | .................... | .................... | .................... | .................... | |
| 22028 | MT873073.1 | \|Severe acute respiratory syndrome coro | .................... | .................... | .................... | .................... | .................... | .................... | |
| 22029 | MT873075.1 | \|Severe acute respiratory syndrome coro | .................... | .................... | .................... | .................... | .................... | .................... | |
| 22030 | MT873076.1 | \|Severe acute respiratory syndrome coro | .................... | .................... | .................... | .................... | .................... | .................... | |
| 22031 | MT873078.1 | \|Severe acute respiratory syndrome coro | .................... | .................... | .................... | .................... | .................... | .................... | |
| 22032 | MT873079.1 | \|Severe acute respiratory syndrome coro | .................... | .................... | .................... | .................... | .................... | .................... | |
| 22033 | MT873080.1 | \|Severe acute respiratory syndrome coro | .................... | .................... | .................... | .................... | .................... | .................... | |
| 22034 | MT873081.1 | \|Severe acute respiratory syndrome coro | .................... | .................... | .................... | .................... | .................... | .................... | |
| 22035 | MT873082.1 | \|Severe acute respiratory syndrome coro | .................... | .................... | .................... | .................... | .................... | .................... | |
| 22036 | MT873083.1 | \|Severe acute respiratory syndrome coro | .................... | .................... | .................... | .................... | .................... | .................... | |
| 22037 | MT873084.1 | \|Severe acute respiratory syndrome coro | .................... | .................... | .................... | .................... | .................... | .................... | |
| 22038 | MT873085.1 | \|Severe acute respiratory syndrome coro | .................... | .................... | .................... | .................... | .................... | .................... | |
| 22039 | MT873086.1 | \|Severe acute respiratory syndrome coro | .................... | .................... | .................... | .................... | .................... | .................... | |
| 22040 | MT873087.1 | \|Severe acute respiratory syndrome coro | .................... | .................... | .................... | .................... | .................... | .................... | |
| 22041 | MT873088.1 | \|Severe acute respiratory syndrome coro | .................... | .................... | .................... | .................... | .................... | .................... | |
| 22042 | MT873089.1 | \|Severe acute respiratory syndrome coro | .................... | .................... | .................... | .................... | .................... | .................... | |
| 22043 | MT873090.1 | \|Severe acute respiratory syndrome coro | .................... | .................... | .................... | .................... | .................... | .................... | |
| 22044 | MT873091.1 | \|Severe acute respiratory syndrome coro | .................... | .................... | .................... | .................... | .................... | .................... | |
| 22045 | MT873092.1 | \|Severe acute respiratory syndrome coro | .................... | .................... | .................... | .................... | .................... | .................... | |
| 22046 | MT873093.1 | \|Severe acute respiratory syndrome coro | .................... | .................... | .................... | .................... | .................... | .................... | |
| 22047 | MT873094.1 | \|Severe acute respiratory syndrome coro | .................... | .................... | .................... | .................... | .................... | .................... | |
| 22048 | MT873095.1 | \|Severe acute respiratory syndrome coro | .................... | .................... | .................... | .................... | .................... | .................... | |
| 22049 | MT873096.1 | \|Severe acute respiratory syndrome coro | .................... | .................... | .................... | .................... | .................... | .................... | |
| 22050 | MT873097.1 | \|Severe acute respiratory syndrome coro | .................... | .................... | .................... | .................... | .................... | .................... | |
| 22051 | MT873098.1 | \|Severe acute respiratory syndrome coro | .................... | .................... | .................... | .................... | .................... | .................... | |
| 22052 | MT873099.1 | \|Severe acute respiratory syndrome coro | .................... | .................... | .................... | .................... | .................... | .................... | |
| 22053 | MT873100.1 | \|Severe acute respiratory syndrome coro | .................... | .................... | .................... | .................... | .................... | .................... | |
| 22054 | MT873102.1 | \|Severe acute respiratory syndrome coro | .................... | .................... | .................... | .................... | .................... | .................... | |
| 22055 | MT873103.1 | \|Severe acute respiratory syndrome coro | .................... | .................... | .................... | .................... | .................... | .................... | |
| 22056 | MT873104.1 | \|Severe acute respiratory syndrome coro | .................... | .................... | .................... | .................... | .................... | .................... | |
| 22057 | MT873105.1 | \|Severe acute respiratory syndrome coro | .................... | .................... | .................... | .................... | .................... | .................... | |
| 22058 | MT873108.1 | \|Severe acute respiratory syndrome coro | .................... | .................... | .................... | .................... | .................... | .................... | |
| 22059 | MT873109.1 | \|Severe acute respiratory syndrome coro | .................... | .................... | .................... | .................... | .................... | .................... | |
| 22060 | MT873110.1 | \|Severe acute respiratory syndrome coro | .................... | .................... | .................... | .................... | .................... | .................... | |
| 22061 | MT873111.1 | \|Severe acute respiratory syndrome coro | .................... | .................... | .................... | .................... | .................... | .................... | |
| 22062 | MT873112.1 | \|Severe acute respiratory syndrome coro | .................... | .................... | .................... | .................... | .................... | .................... | |
| 22063 | MT873113.1 | \|Severe acute respiratory syndrome coro | .................... | .................... | .................... | .................... | .................... | .................... | |
| 22064 | MT873114.1 | \|Severe acute respiratory syndrome coro | .................... | .................... | .................... | .................... | .................... | .................... | |
| 22065 | MT873115.1 | \|Severe acute respiratory syndrome coro | .................... | .................... | .................... | .................... | .................... | .................... | |
| 22066 | MT873116.1 | \|Severe acute respiratory syndrome coro | .................... | ...................A | .................... | .................... | .................... | .................... | 1 |
| 22067 | MT873117.1 | \|Severe acute respiratory syndrome coro | .................... | .................... | .................... | .................... | .................... | .................... | |
| 22068 | MT873118.1 | \|Severe acute respiratory syndrome coro | .................... | .................... | .................... | .................... | .................... | .................... | |
| 22069 | MT873119.1 | \|Severe acute respiratory syndrome coro | .................... | .................... | .................... | .................... | .................... | .................... | |
| 22070 | MT873120.1 | \|Severe acute respiratory syndrome coro | .................... | .................... | .................... | .................... | .................... | .................... | |
| 22071 | MT873121.1 | \|Severe acute respiratory syndrome coro | .................... | .................... | .................... | .................... | .................... | .................... | |
| 22072 | MT873123.1 | \|Severe acute respiratory syndrome coro | .................... | .................... | .................... | .................... | .................... | .................... | |
| 22073 | MT873124.1 | \|Severe acute respiratory syndrome coro | .................... | .................... | .................... | .................... | .................... | .................... | |
| 22074 | MT873125.1 | \|Severe acute respiratory syndrome coro | .................... | .................... | .................... | .................... | .................... | .................... | |
| 22075 | MT873126.1 | \|Severe acute respiratory syndrome coro | .................... | .................... | .................... | .................... | .................... | .................... | |
| 22076 | MT873127.1 | \|Severe acute respiratory syndrome coro | .................... | .................... | .................... | .................... | .................... | .................... | |
| 22077 | MT873128.1 | \|Severe acute respiratory syndrome coro | .................... | .................... | .................... | .................... | .................... | .................... | |
| 22078 | MT873129.1 | \|Severe acute respiratory syndrome coro | .................... | .................... | .................... | .................... | .................... | .................... | |
| 22079 | MT873130.1 | \|Severe acute respiratory syndrome coro | .................... | .................... | .................... | .................... | .................... | .................... | |
| 22080 | MT873131.1 | \|Severe acute respiratory syndrome coro | .................... | .................... | .................... | .................... | .................... | .................... | |
| 22081 | MT873132.1 | \|Severe acute respiratory syndrome coro | .................... | .................... | .................... | .................... | .................... | .................... | |
| 22082 | MT873133.1 | \|Severe acute respiratory syndrome coro | .................... | .................... | .................... | .................... | .................... | .................... | |
| 22083 | MT873134.1 | \|Severe acute respiratory syndrome coro | .................... | .................... | .................... | .................... | .................... | .................... | |
| 22084 | MT873135.1 | \|Severe acute respiratory syndrome coro | .................... | .................... | .................... | .................... | .................... | .................... | |
| 22085 | MT873136.1 | \|Severe acute respiratory syndrome coro | .................... | .................... | .................... | .................... | .................... | .................... | |
| 22086 | MT873137.1 | \|Severe acute respiratory syndrome coro | .................... | .................... | .................... | .................... | .................... | .................... | |
| 22087 | MT873139.1 | \|Severe acute respiratory syndrome coro | .................... | .................... | .................... | .................... | .................... | .................... | |
| 22088 | MT873140.1 | \|Severe acute respiratory syndrome coro | .................... | .................... | .................... | .................... | .................... | .................... | |
| 22089 | MT873141.1 | \|Severe acute respiratory syndrome coro | .................... | .................... | .................... | .................... | .................... | .................... | |
| 22090 | MT873143.1 | \|Severe acute respiratory syndrome coro | .................... | .................... | .................... | .................... | .................... | .................... | |
| 22091 | MT873145.1 | \|Severe acute respiratory syndrome coro | .................... | .................... | .................... | .................... | .................... | .................... | |
| 22092 | MT873146.1 | \|Severe acute respiratory syndrome coro | .................... | .................... | .................... | .................... | .................... | .................... | |
| 22093 | MT873147.1 | \|Severe acute respiratory syndrome coro | .................... | .................... | .................... | .................... | .................... | .................... | |

| | | | | | | | | | |
|---|---|---|---|---|---|---|---|---|---|
| 22094 | MT873148.1 | Severe acute respiratory syndrome coro | ···················· | ···················· | ···················· | ···················· | ···················· | ···················· | |
| 22095 | MT873149.1 | Severe acute respiratory syndrome coro | ···················· | ···················· | ···················· | ···················· | ···················· | ···················· | |
| 22096 | MT873150.1 | Severe acute respiratory syndrome coro | ···················· | ···················· | ···················· | ···················· | ···················· | ···················· | |
| 22097 | MT873151.1 | Severe acute respiratory syndrome coro | ···················· | ···················· | ···················· | ···················· | ···················· | ···················· | |
| 22098 | MT873152.1 | Severe acute respiratory syndrome coro | ···················· | ···················· | ···················· | ···················· | ···················· | ···················· | |
| 22099 | MT873153.1 | Severe acute respiratory syndrome coro | ···················· | ···················· | ···················· | ···················· | ···················· | ···················· | |
| 22100 | MT873154.1 | Severe acute respiratory syndrome coro | ···················· | ···················· | ···················· | ···················· | ···················· | ···················· | |
| 22101 | MT873156.1 | Severe acute respiratory syndrome coro | ···················· | ···················· | ···················· | ···················· | ···················· | ···················· | |
| 22102 | MT873157.1 | Severe acute respiratory syndrome coro | ···················· | ···················· | ···················· | ···················· | ···················· | ···················· | |
| 22103 | MT873158.1 | Severe acute respiratory syndrome coro | ···················· | ···················· | ···················· | ···················· | ···················· | ···················· | |
| 22104 | MT873159.1 | Severe acute respiratory syndrome coro | ···················· | ···················· | ···················· | ···················· | ···················· | ···················· | |
| 22105 | MT873160.1 | Severe acute respiratory syndrome coro | ···················· | ···················· | ···················· | ···················· | ···················· | ···················· | |
| 22106 | MT873161.1 | Severe acute respiratory syndrome coro | ···················· | ···················· | ···················· | ···················· | ···················· | ···················· | |
| 22107 | MT873163.1 | Severe acute respiratory syndrome coro | ···················· | ···················· | ···················· | ···················· | ···················· | ···················· | |
| 22108 | MT873165.1 | Severe acute respiratory syndrome coro | ···················· | ···················· | ···················· | ···················· | ···················· | ···················· | |
| 22109 | MT873166.1 | Severe acute respiratory syndrome coro | ···················· | ···················· | ···················· | ···················· | ···················· | ···················· | |
| 22110 | MT873167.1 | Severe acute respiratory syndrome coro | ···················· | ···················· | ···················· | ···················· | ···················· | ···················· | |
| 22111 | MT873168.1 | Severe acute respiratory syndrome coro | ···················· | ···················· | ···················· | ···················· | ···················· | ···················· | |
| 22112 | MT873169.1 | Severe acute respiratory syndrome coro | ···················· | ···················· | ···················· | ···················· | ···················· | ···················· | |
| 22113 | MT873172.1 | Severe acute respiratory syndrome coro | ···················· | ···················· | ···················· | ···················· | ···················· | ···················· | |
| 22114 | MT873173.1 | Severe acute respiratory syndrome coro | ···················· | ···················· | ···················· | ···················· | ···················· | ···················· | |
| 22115 | MT873174.1 | Severe acute respiratory syndrome coro | ···················· | ···················· | ···················· | ···················· | ···················· | ···················· | |
| 22116 | MT873175.1 | Severe acute respiratory syndrome coro | ···················· | ···················· | ···················· | ···················· | ···················· | ···················· | |
| 22117 | MT873176.1 | Severe acute respiratory syndrome coro | ···················· | ···················· | ···················· | ···················· | ···················· | ···················· | |
| 22118 | MT873177.1 | Severe acute respiratory syndrome coro | ···················· | ···················· | ···················· | ···················· | ···················· | ···················· | |
| 22119 | MT873178.1 | Severe acute respiratory syndrome coro | ···················· | ···················· | ···················· | ···················· | ···················· | ···················· | |
| 22120 | MT873179.1 | Severe acute respiratory syndrome coro | ···················· | ···················· | ···················· | ···················· | ···················· | ···················· | |
| 22121 | MT873180.1 | Severe acute respiratory syndrome coro | ···················· | ···················· | ···················· | ···················· | ···················· | ···················· | |
| 22122 | MT873181.1 | Severe acute respiratory syndrome coro | ···················· | ···················· | ···················· | ···················· | ···················· | ···················· | |
| 22123 | MT873182.1 | Severe acute respiratory syndrome coro | ···················· | ···················· | ···················· | ···················· | ···················· | ···················· | |
| 22124 | MT873183.1 | Severe acute respiratory syndrome coro | ···················· | ···················· | ···················· | ···················· | ···················· | ···················· | |
| 22125 | MT873184.1 | Severe acute respiratory syndrome coro | ···················· | ···················· | ···················· | ···················· | ···················· | ···················· | |
| 22126 | MT873185.1 | Severe acute respiratory syndrome coro | ···················· | ···················· | ···················· | ···················· | ···················· | ···················· | |
| 22127 | MT873187.1 | Severe acute respiratory syndrome coro | ···················· | ···················· | ···················· | ···················· | ···················· | ···················· | |
| 22128 | MT873188.1 | Severe acute respiratory syndrome coro | ···················· | ···················· | ···················· | ···················· | ···················· | ···················· | |
| 22129 | MT873189.1 | Severe acute respiratory syndrome coro | ···················· | ···················· | ···················· | ···················· | ···················· | ···················· | |
| 22130 | MT873190.1 | Severe acute respiratory syndrome coro | ···················· | ···················· | ···················· | ···················· | ···················· | ···················· | |
| 22131 | MT873191.1 | Severe acute respiratory syndrome coro | ···················· | ···················· | ···················· | ···················· | ···················· | ···················· | |
| 22132 | MT873192.1 | Severe acute respiratory syndrome coro | ···················· | ···················· | ···················· | ···················· | ···················· | ···················· | |
| 22133 | MT873194.1 | Severe acute respiratory syndrome coro | ···················· | ···················· | ···················· | ···················· | ···················· | ···················· | |
| 22134 | MT873195.1 | Severe acute respiratory syndrome coro | ···················· | ···················· | ···················· | ···················· | ···················· | ···················· | |
| 22135 | MT873196.1 | Severe acute respiratory syndrome coro | ···················· | ···················A | ···················· | ···················· | ···················· | ···················· | 1 |
| 22136 | MT873197.1 | Severe acute respiratory syndrome coro | ···················· | ···················· | ···················· | ···················· | ···················· | ···················· | |
| 22137 | MT873198.1 | Severe acute respiratory syndrome coro | ···················· | ···················· | ···················· | ···················· | ···················· | ···················· | |
| 22138 | MT873199.1 | Severe acute respiratory syndrome coro | ···················· | ···················· | ···················· | ···················· | ···················· | ···················· | |
| 22139 | MT873200.1 | Severe acute respiratory syndrome coro | ···················· | ···················· | ···················· | ···················· | ···················· | ···················· | |
| 22140 | MT873201.1 | Severe acute respiratory syndrome coro | ···················· | ···················· | ···················· | ···················· | ···················· | ···················· | |
| 22141 | MT873202.1 | Severe acute respiratory syndrome coro | ···················· | ···················· | ···················· | ···················· | ···················· | ···················· | |
| 22142 | MT873203.1 | Severe acute respiratory syndrome coro | ···················· | ···················· | ···················· | ···················· | ···················· | ···················· | |
| 22143 | MT873204.1 | Severe acute respiratory syndrome coro | ···················· | ···················· | ···················· | ···················· | ···················· | ···················· | |
| 22145 | MT873206.1 | Severe acute respiratory syndrome coro | ···················· | ···················· | ···················· | ···················· | ···················· | ···················· | |
| 22146 | MT873207.1 | Severe acute respiratory syndrome coro | ···················· | ···················· | ···················· | ···················· | ···················· | ···················· | |
| 22147 | MT873208.1 | Severe acute respiratory syndrome coro | ···················· | ···················· | ···················· | ···················· | ···················· | ···················· | |
| 22148 | MT873209.1 | Severe acute respiratory syndrome coro | ···················· | ···················· | ···················· | ···················· | ···················· | ···················· | |
| 22149 | MT873210.1 | Severe acute respiratory syndrome coro | ···················· | ···················· | ···················· | ···················· | ···················· | ···················· | |
| 22150 | MT873211.1 | Severe acute respiratory syndrome coro | ···················· | ···················· | ···················· | ···················· | ···················· | ···················· | |
| 22151 | MT873212.1 | Severe acute respiratory syndrome coro | ···················· | ···················· | ···················· | ···················· | ···················· | ···················· | |
| 22152 | MT873213.1 | Severe acute respiratory syndrome coro | ···················· | ···················· | ···················· | ···················· | ···················· | ···················· | |
| 22153 | MT873216.1 | Severe acute respiratory syndrome coro | ···················· | ···················· | ···················· | ···················· | ···················· | ···················· | |
| 22154 | MT873217.1 | Severe acute respiratory syndrome coro | ···················· | ···················· | ···················· | ···················· | ···················· | ···················· | |
| 22155 | MT873218.1 | Severe acute respiratory syndrome coro | ···················· | ···················· | ···················· | ···················· | ···················· | ···················· | |
| 22156 | MT873220.1 | Severe acute respiratory syndrome coro | ···················· | ···················· | ···················· | ···················· | ···················· | ···················· | |
| 22157 | MT873221.1 | Severe acute respiratory syndrome coro | ···················· | ···················· | ···················· | ···················· | ···················· | ···················· | |
| 22158 | MT873222.1 | Severe acute respiratory syndrome coro | ···················· | ···················· | ···················· | ···················· | ···················· | ···················· | |
| 22159 | MT873223.1 | Severe acute respiratory syndrome coro | ···················· | ···················· | ···················· | ···················· | ···················· | ···················· | |
| 22160 | MT873224.1 | Severe acute respiratory syndrome coro | ···················· | ···················· | ···················· | ···················· | ···················· | ···················· | |
| 22161 | MT873225.1 | Severe acute respiratory syndrome coro | ···················· | ···················· | ···················· | ···················· | ···················· | ···················· | |
| 22162 | MT873226.1 | Severe acute respiratory syndrome coro | ···················· | ···················· | ···················· | ···················· | ···················· | ···················· | |
| 22163 | MT873228.1 | Severe acute respiratory syndrome coro | ···················· | ···················· | ···················· | ···················· | ···················· | ···················· | |
| 22164 | MT873230.1 | Severe acute respiratory syndrome coro | ···················· | ···················· | ···················· | ···················· | ···················· | ···················· | |
| 22165 | MT873231.1 | Severe acute respiratory syndrome coro | ···················· | ···················· | ···················· | ···················· | ···················· | ···················· | |
| 22166 | MT873233.1 | Severe acute respiratory syndrome coro | ···················· | ···················· | ···················· | ···················· | ···················· | ···················· | |
| 22167 | MT873235.1 | Severe acute respiratory syndrome coro | ···················· | ···················· | ···················· | ···················· | ···················· | ···················· | |
| 22168 | MT873236.1 | Severe acute respiratory syndrome coro | ···················· | ···················· | ···················· | ···················· | ···················· | ···················· | |
| 22169 | MT873237.1 | Severe acute respiratory syndrome coro | ···················· | ···················· | ···················· | ···················· | ···················· | ···················· | |
| 22170 | MT873238.1 | Severe acute respiratory syndrome coro | ···················· | ···················· | ···················· | ···················· | ···················· | ···················· | |
| 22171 | MT873239.1 | Severe acute respiratory syndrome coro | ···················· | ···················· | ···················· | ···················· | ···················· | ···················· | |
| 22172 | MT873240.1 | Severe acute respiratory syndrome coro | ···················· | ···················· | ···················· | ···················· | ···················· | ···················· | |

| | | | | | | | | |
|---|---|---|---|---|---|---|---|---|
| 22173 | MT873242.1 | \|Severe acute respiratory syndrome coro | .................... | .................... | .................... | .................... | .................... | .................... |
| 22174 | MT873243.1 | \|Severe acute respiratory syndrome coro | .................... | .................... | .................... | .................... | .................... | .................... |
| 22175 | MT873244.1 | \|Severe acute respiratory syndrome coro | .................... | .................... | .................... | .................... | .................... | .................... |
| 22176 | MT873245.1 | \|Severe acute respiratory syndrome coro | .................... | .................... | .................... | .................... | .................... | .................... |
| 22177 | MT873247.1 | \|Severe acute respiratory syndrome coro | .................... | .................... | .................... | .................... | .................... | .................... |
| 22178 | MT873249.1 | \|Severe acute respiratory syndrome coro | .................... | .................... | .................... | .................... | .................... | .................... |
| 22179 | MT873250.1 | \|Severe acute respiratory syndrome coro | .................... | .................... | .................... | .................... | .................... | .................... |
| 22180 | MT873251.1 | \|Severe acute respiratory syndrome coro | .................... | .................... | .................... | .................... | .................... | .................... |
| 22181 | MT873253.1 | \|Severe acute respiratory syndrome coro | .................... | .................... | .................... | .................... | .................... | .................... |
| 22182 | MT873254.1 | \|Severe acute respiratory syndrome coro | .................... | .................... | .................... | .................... | .................... | .................... |
| 22183 | MT873255.1 | \|Severe acute respiratory syndrome coro | .................... | .................... | .................... | .................... | .................... | .................... |
| 22184 | MT873256.1 | \|Severe acute respiratory syndrome coro | .................... | .................... | .................... | .................... | .................... | .................... |
| 22185 | MT873257.1 | \|Severe acute respiratory syndrome coro | .................... | .................... | .................... | .................... | .................... | .................... |
| 22186 | MT873258.1 | \|Severe acute respiratory syndrome coro | .................... | .................... | .................... | .................... | .................... | .................... |
| 22187 | MT873259.1 | \|Severe acute respiratory syndrome coro | .................... | .................... | .................... | .................... | .................... | .................... |
| 22188 | MT873261.1 | \|Severe acute respiratory syndrome coro | .................... | .................... | .................... | .................... | .................... | .................... |
| 22189 | MT873262.1 | \|Severe acute respiratory syndrome coro | .................... | .................... | .................... | .................... | .................... | .................... |
| 22190 | MT873263.1 | \|Severe acute respiratory syndrome coro | .................... | .................... | .................... | .................... | .................... | .................... |
| 22191 | MT873264.1 | \|Severe acute respiratory syndrome coro | .................... | .................... | .................... | .................... | .................... | .................... |
| 22192 | MT873265.1 | \|Severe acute respiratory syndrome coro | .................... | .................... | .................... | .................... | .................... | .................... |
| 22193 | MT873266.1 | \|Severe acute respiratory syndrome coro | .................... | .................... | .................... | .................... | .................... | .................... |
| 22194 | MT873267.1 | \|Severe acute respiratory syndrome coro | .................... | .................... | .................... | .................... | .................... | .................... |
| 22195 | MT873268.1 | \|Severe acute respiratory syndrome coro | .................... | .................... | .................... | .................... | .................... | .................... |
| 22196 | MT873269.1 | \|Severe acute respiratory syndrome coro | .................... | .................... | .................... | .................... | .................... | .................... |
| 22197 | MT873270.1 | \|Severe acute respiratory syndrome coro | .................... | .................... | .................... | .................... | .................... | .................... |
| 22198 | MT873272.1 | \|Severe acute respiratory syndrome coro | .................... | .................... | .................... | .................... | .................... | .................... |
| 22199 | MT873273.1 | \|Severe acute respiratory syndrome coro | .................... | .................... | .................... | .................... | .................... | .................... |
| 22200 | MT873274.1 | \|Severe acute respiratory syndrome coro | .................... | .................... | .................... | .................... | .................... | .................... |
| 22201 | MT873275.1 | \|Severe acute respiratory syndrome coro | .................... | .................... | .................... | .................... | .................... | .................... |
| 22202 | MT873277.1 | \|Severe acute respiratory syndrome coro | .................... | .................... | .................... | .................... | .................... | .................... |
| 22203 | MT873278.1 | \|Severe acute respiratory syndrome coro | .................... | .................... | .................... | .................... | .................... | .................... |
| 22204 | MT873279.1 | \|Severe acute respiratory syndrome coro | .................... | .................... | .................... | .................... | .................... | .................... |
| 22205 | MT873280.1 | \|Severe acute respiratory syndrome coro | .................... | .................... | .................... | .................... | .................... | .................... |
| 22206 | MT873281.1 | \|Severe acute respiratory syndrome coro | .................... | .................... | .................... | .................... | .................... | .................... |
| 22207 | MT873282.1 | \|Severe acute respiratory syndrome coro | .................... | ......A............. | .................... | .................... | .................... | .................... |
| 22208 | MT873283.1 | \|Severe acute respiratory syndrome coro | .................... | .................... | .................... | .................... | .................... | .................... |
| 22209 | MT873284.1 | \|Severe acute respiratory syndrome coro | .................... | .................... | .................... | .................... | .................... | .................... |
| 22210 | MT873285.1 | \|Severe acute respiratory syndrome coro | .................... | .................... | .................... | .................... | .................... | .................... |
| 22211 | MT873286.1 | \|Severe acute respiratory syndrome coro | .................... | .................... | .................... | .................... | .................... | .................... |
| 22212 | MT873287.1 | \|Severe acute respiratory syndrome coro | .................... | .................... | .................... | .................... | .................... | .................... |
| 22213 | MT873289.1 | \|Severe acute respiratory syndrome coro | .................... | .................... | .................... | .................... | .................... | .................... |
| 22214 | MT873290.1 | \|Severe acute respiratory syndrome coro | .................... | .................... | .................... | .................... | .................... | .................... |
| 22215 | MT873292.1 | \|Severe acute respiratory syndrome coro | .................... | .................... | .................... | .................... | .................... | .................... |
| 22216 | MT873293.1 | \|Severe acute respiratory syndrome coro | .................... | .................... | .................... | .................... | .................... | .................... |
| 22217 | MT873294.1 | \|Severe acute respiratory syndrome coro | .................... | .................... | .................... | .................... | .................... | .................... |
| 22218 | MT873295.1 | \|Severe acute respiratory syndrome coro | .................... | .................... | .................... | .................... | .................... | .................... |
| 22219 | MT873296.1 | \|Severe acute respiratory syndrome coro | .................... | .................... | .................... | .................... | .................... | .................... |
| 22220 | MT873298.1 | \|Severe acute respiratory syndrome coro | .................... | .................... | .................... | .................... | .................... | .................... |
| 22221 | MT873299.1 | \|Severe acute respiratory syndrome coro | .................... | .................... | .................... | .................... | .................... | .................... |
| 22222 | MT873300.1 | \|Severe acute respiratory syndrome coro | .................... | .................... | .................... | .................... | .................... | .................... |
| 22223 | MT873301.1 | \|Severe acute respiratory syndrome coro | .................... | .................... | .................... | .................... | .................... | .................... |
| 22224 | MT873302.1 | \|Severe acute respiratory syndrome coro | .................... | .................... | .................... | .................... | .................... | .................... |
| 22225 | MT873303.1 | \|Severe acute respiratory syndrome coro | .................... | .................... | .................... | .................... | .................... | .................... |
| 22226 | MT873304.1 | \|Severe acute respiratory syndrome coro | .................... | .................... | .................... | .................... | .................... | .................... |
| 22227 | MT873305.1 | \|Severe acute respiratory syndrome coro | .................... | .................... | .................... | .................... | .................... | .................... |
| 22228 | MT873306.1 | \|Severe acute respiratory syndrome coro | .................... | .................... | .................... | .................... | .................... | .................... |
| 22229 | MT873307.1 | \|Severe acute respiratory syndrome coro | .................... | .................... | .................... | .................... | .................... | .................... |
| 22230 | MT873308.1 | \|Severe acute respiratory syndrome coro | .................... | .................... | .................... | .................... | .................... | .................... |
| 22231 | MT873309.1 | \|Severe acute respiratory syndrome coro | .................... | .................... | .................... | .................... | .................... | .................... |
| 22232 | MT873311.1 | \|Severe acute respiratory syndrome coro | .................... | .................... | .................... | .................... | .................... | .................... |
| 22233 | MT873312.1 | \|Severe acute respiratory syndrome coro | .................... | .................... | .................... | .................... | .................... | .................... |
| 22234 | MT873314.1 | \|Severe acute respiratory syndrome coro | .................... | .................... | .................... | .................... | .................... | .................... |
| 22235 | MT873316.1 | \|Severe acute respiratory syndrome coro | .................... | .................... | .................... | .................... | .................... | .................... |
| 22236 | MT873317.1 | \|Severe acute respiratory syndrome coro | .................... | .................... | .................... | .................... | .................... | .................... |
| 22237 | MT873318.1 | \|Severe acute respiratory syndrome coro | .................... | .................... | .................... | .................... | .................... | .................... |
| 22238 | MT873319.1 | \|Severe acute respiratory syndrome coro | .................... | .................... | .................... | .................... | .................... | .................... |
| 22239 | MT873320.1 | \|Severe acute respiratory syndrome coro | .................... | .................... | .................... | .................... | .................... | .................... |
| 22240 | MT873321.1 | \|Severe acute respiratory syndrome coro | .................... | .................... | .................... | .................... | .................... | .................... |
| 22241 | MT873323.1 | \|Severe acute respiratory syndrome coro | .................... | .................... | .................... | .................... | .................... | .................... |
| 22242 | MT873324.1 | \|Severe acute respiratory syndrome coro | .................... | .................... | .................... | .................... | .................... | .................... |
| 22243 | MT873326.1 | \|Severe acute respiratory syndrome coro | .................... | .................... | .................... | .................... | .................... | .................... |
| 22244 | MT873327.1 | \|Severe acute respiratory syndrome coro | .................... | .................... | .................... | .................... | .................... | .................... |
| 22245 | MT873328.1 | \|Severe acute respiratory syndrome coro | .................... | .................... | .................... | .................... | .................... | .................... |
| 22246 | MT873329.1 | \|Severe acute respiratory syndrome coro | .................... | .................... | .................... | .................... | .................... | .................... |
| 22247 | MT873330.1 | \|Severe acute respiratory syndrome coro | .................... | .................... | .................... | .................... | .................... | .................... |
| 22248 | MT873331.1 | \|Severe acute respiratory syndrome coro | .................... | .................... | .................... | .................... | .................... | .................... |
| 22249 | MT873332.1 | \|Severe acute respiratory syndrome coro | .................... | .................... | .................... | .................... | .................... | .................... |
| 22250 | MT873333.1 | \|Severe acute respiratory syndrome coro | .................... | .................... | .................... | .................... | .................... | .................... |

```
22251 MT873334.1 |Severe acute respiratory syndrome coro  ····················  ····················  ····················  ····················  ····················  ····················
22252 MT873335.1 |Severe acute respiratory syndrome coro  ····················  ······A·············  ····················  ····················  ····················  ····················
22253 MT873336.1 |Severe acute respiratory syndrome coro  ····················  ····················  ····················  ····················  ····················  ····················
22254 MT873337.1 |Severe acute respiratory syndrome coro  ····················  ····················  ····················  ····················  ····················  ····················
22255 MT873338.1 |Severe acute respiratory syndrome coro  ····················  ····················  ····················  ····················  ····················  ····················
22256 MT873339.1 |Severe acute respiratory syndrome coro  ····················  ····················  ····················  ····················  ····················  ····················
22257 MT873340.1 |Severe acute respiratory syndrome coro  ····················  ····················  ····················  ····················  ····················  ····················
22258 MT873342.1 |Severe acute respiratory syndrome coro  ····················  ····················  ····················  ····················  ····················  ····················
22259 MT873343.1 |Severe acute respiratory syndrome coro  ····················  ····················  ····················  ····················  ····················  ····················
22260 MT873344.1 |Severe acute respiratory syndrome coro  ····················  ····················  ····················  ····················  ····················  ····················
22261 MT873345.1 |Severe acute respiratory syndrome coro  ····················  ····················  ····················  ····················  ····················  ····················
22262 MT873346.1 |Severe acute respiratory syndrome coro  ····················  ····················  ····················  ····················  ····················  ····················
22263 MT873347.1 |Severe acute respiratory syndrome coro  ····················  ····················  ····················  ····················  ····················  ····················
22264 MT873348.1 |Severe acute respiratory syndrome coro  ····················  ····················  ····················  ····················  ····················  ····················
22265 MT873349.1 |Severe acute respiratory syndrome coro  ····················  ····················  ····················  ····················  ····················  ····················
22266 MT873350.1 |Severe acute respiratory syndrome coro  ····················  ····················  ····················  ····················  ····················  ····················
22267 MT873351.1 |Severe acute respiratory syndrome coro  ····················  ····················  ····················  ····················  ····················  ····················
22268 MT873352.1 |Severe acute respiratory syndrome coro  ····················  ····················  ····················  ····················  ····················  ····················
22269 MT873353.1 |Severe acute respiratory syndrome coro  ····················  ····················  ····················  ····················  ····················  ····················
22270 MT873355.1 |Severe acute respiratory syndrome coro  ····················  ····················  ····················  ····················  ····················  ····················
22271 MT873356.1 |Severe acute respiratory syndrome coro  ····················  ····················  ····················  ····················  ····················  ····················
22272 MT873359.1 |Severe acute respiratory syndrome coro  ····················  ····················  ····················  ····················  ····················  ····················
22273 MT873360.1 |Severe acute respiratory syndrome coro  ····················  ····················  ····················  ····················  ····················  ····················
22274 MT873361.1 |Severe acute respiratory syndrome coro  ····················  ····················  ····················  ····················  ····················  ····················
22275 MT873362.1 |Severe acute respiratory syndrome coro  ····················  ····················  ····················  ····················  ····················  ····················
22276 MT873363.1 |Severe acute respiratory syndrome coro  ····················  ····················  ····················  ····················  ····················  ····················
22277 MT873364.1 |Severe acute respiratory syndrome coro  ····················  ···················A  ····················  ····················  ····················  ····················  1
22278 MT873365.1 |Severe acute respiratory syndrome coro  ····················  ····················  ····················  ····················  ····················  ····················
22279 MT873366.1 |Severe acute respiratory syndrome coro  ····················  ····················  ····················  ····················  ····················  ····················
22280 MT873367.1 |Severe acute respiratory syndrome coro  ····················  ····················  ····················  ····················  ····················  ····················
22281 MT873368.1 |Severe acute respiratory syndrome coro  ····················  ····················  ····················  ····················  ····················  ····················
22282 MT873369.1 |Severe acute respiratory syndrome coro  ····················  ····················  ····················  ····················  ····················  ····················
22283 MT873370.1 |Severe acute respiratory syndrome coro  ····················  ····················  ····················  ····················  ····················  ····················
22284 MT873372.1 |Severe acute respiratory syndrome coro  ····················  ····················  ····················  ····················  ····················  ····················
22285 MT873373.1 |Severe acute respiratory syndrome coro  ····················  ····················  ····················  ····················  ····················  ····················
22286 MT873375.1 |Severe acute respiratory syndrome coro  ····················  ····················  ····················  ····················  ····················  ····················
22287 MT873376.1 |Severe acute respiratory syndrome coro  ····················  ····················  ····················  ····················  ····················  ····················
22288 MT873377.1 |Severe acute respiratory syndrome coro  ····················  ····················  ····················  ····················  ····················  ····················
22289 MT873378.1 |Severe acute respiratory syndrome coro  ····················  ····················  ····················  ····················  ····················  ····················
22290 MT873379.1 |Severe acute respiratory syndrome coro  ····················  ···················A  ····················  ····················  ····················  ····················  1
22291 MT873380.1 |Severe acute respiratory syndrome coro  ····················  ····················  ····················  ····················  ····················  ····················
22292 MT873381.1 |Severe acute respiratory syndrome coro  ····················  ····················  ····················  ····················  ····················  ····················
22293 MT873382.1 |Severe acute respiratory syndrome coro  ····················  ····················  ····················  ····················  ····················  ····················
22294 MT873383.1 |Severe acute respiratory syndrome coro  ····················  ····················  ····················  ····················  ····················  ····················
22295 MT873384.1 |Severe acute respiratory syndrome coro  ····················  ···················A  ····················  ····················  ····················  ····················  1
22296 MT873385.1 |Severe acute respiratory syndrome coro  ····················  ····················  ····················  ····················  ····················  ····················
22297 MT873387.1 |Severe acute respiratory syndrome coro  ····················  ····················  ····················  ····················  ····················  ····················
22298 MT873389.1 |Severe acute respiratory syndrome coro  ····················  ····················  ····················  ····················  ····················  ····················
22299 MT873390.1 |Severe acute respiratory syndrome coro  ····················  ····················  ····················  ····················  ····················  ····················
22300 MT873391.1 |Severe acute respiratory syndrome coro  ····················  ····················  ····················  ····················  ····················  ····················
22301 MT873392.1 |Severe acute respiratory syndrome coro  ····················  ····················  ····················  ····················  ····················  ····················
22302 MT873393.1 |Severe acute respiratory syndrome coro  ····················  ····················  ····················  ····················  ····················  ····················
22303 MT873395.1 |Severe acute respiratory syndrome coro  ····················  ····················  ····················  ····················  ····················  ····················
22304 MT873396.1 |Severe acute respiratory syndrome coro  ····················  ····················  ····················  ····················  ····················  ····················
22305 MT873397.1 |Severe acute respiratory syndrome coro  ····················  ····················  ····················  ····················  ····················  ····················
22306 MT873398.1 |Severe acute respiratory syndrome coro  ····················  ····················  ····················  ····················  ····················  ····················
22307 MT873399.1 |Severe acute respiratory syndrome coro  ····················  ····················  ····················  ····················  ····················  ····················
22308 MT873401.1 |Severe acute respiratory syndrome coro  ····················  ····················  ····················  ····················  ····················  ····················
22309 MT873402.1 |Severe acute respiratory syndrome coro  ····················  ····················  ····················  ····················  ····················  ····················
22310 MT873403.1 |Severe acute respiratory syndrome coro  ····················  ····················  ····················  ····················  ····················  ····················
22311 MT873404.1 |Severe acute respiratory syndrome coro  ····················  ····················  ····················  ····················  ····················  ····················
22312 MT873405.1 |Severe acute respiratory syndrome coro  ····················  ····················  ····················  ····················  ····················  ····················
22313 MT873406.1 |Severe acute respiratory syndrome coro  ····················  ····················  ····················  ····················  ····················  ····················
22314 MT873407.1 |Severe acute respiratory syndrome coro  ····················  ····················  ····················  ····················  ····················  ····················
22315 MT873408.1 |Severe acute respiratory syndrome coro  ····················  ····················  ····················  ····················  ····················  ····················
22316 MT873409.1 |Severe acute respiratory syndrome coro  ····················  ····················  ····················  ····················  ····················  ····················
22317 MT873410.1 |Severe acute respiratory syndrome coro  ····················  ····················  ····················  ····················  ····················  ····················
22318 MT873411.1 |Severe acute respiratory syndrome coro  ····················  ····················  ····················  ····················  ····················  ····················
22319 MT873412.1 |Severe acute respiratory syndrome coro  ····················  ····················  ····················  ····················  ····················  ····················
22320 MT873413.1 |Severe acute respiratory syndrome coro  ····················  ····················  ····················  ····················  ····················  ····················
22321 MT873414.1 |Severe acute respiratory syndrome coro  ····················  ····················  ····················  ····················  ····················  ····················
22322 MT873416.1 |Severe acute respiratory syndrome coro  ····················  ····················  ····················  ····················  ····················  ····················
22323 MT873417.1 |Severe acute respiratory syndrome coro  ····················  ····················  ····················  ····················  ····················  ····················
22324 MT873418.1 |Severe acute respiratory syndrome coro  ····················  ····················  ····················  ····················  ····················  ····················
22325 MT873419.1 |Severe acute respiratory syndrome coro  ····················  ····················  ····················  ····················  ····················  ····················
22326 MT873420.1 |Severe acute respiratory syndrome coro  ····················  ····················  ····················  ····················  ····················  ····················
22327 MT873421.1 |Severe acute respiratory syndrome coro  ····················  ····················  ····················  ····················  ····················  ····················
22328 MT873422.1 |Severe acute respiratory syndrome coro  ····················  ····················  ····················  ····················  ····················  ····················
```

| | | | | | | | | |
|---|---|---|---|---|---|---|---|---|
| 22329 | MT873424.1 | \|Severe acute respiratory syndrome coro | ···················· | ···················· | ····················· | ···················· | ···················· | ···················· |
| 22330 | MT873426.1 | \|Severe acute respiratory syndrome coro | ···················· | ···················· | ····················· | ···················· | ···················· | ···················· |
| 22331 | MT873428.1 | \|Severe acute respiratory syndrome coro | ···················· | ···················· | ····················· | ···················· | ···················· | ···················· |
| 22332 | MT873429.1 | \|Severe acute respiratory syndrome coro | ···················· | ···················· | ····················· | ···················· | ···················· | ···················· |
| 22333 | MT873430.1 | \|Severe acute respiratory syndrome coro | ···················· | ···················· | ····················· | ···················· | ···················· | ···················· |
| 22334 | MT873431.1 | \|Severe acute respiratory syndrome coro | ···················· | ···················· | ····················· | ···················· | ···················· | ···················· |
| 22335 | MT873433.1 | \|Severe acute respiratory syndrome coro | ···················· | ···················· | ····················· | ···················· | ···················· | ···················· |
| 22336 | MT873434.1 | \|Severe acute respiratory syndrome coro | ···················· | ···················· | ····················· | ···················· | ···················· | ···················· |
| 22337 | MT873436.1 | \|Severe acute respiratory syndrome coro | ···················· | ···················· | ····················· | ···················· | ···················· | ···················· |
| 22338 | MT873437.1 | \|Severe acute respiratory syndrome coro | ···················· | ···················· | ····················· | ···················· | ···················· | ···················· |
| 22339 | MT873438.1 | \|Severe acute respiratory syndrome coro | ···················· | ···················· | ····················· | ···················· | ···················· | ···················· |
| 22340 | MT873439.1 | \|Severe acute respiratory syndrome coro | ···················· | ···················· | ····················· | ···················· | ···················· | ···················· |
| 22341 | MT873440.1 | \|Severe acute respiratory syndrome coro | ···················· | ···················· | ····················· | ···················· | ···················· | ···················· |
| 22342 | MT873441.1 | \|Severe acute respiratory syndrome coro | ···················· | ···················· | ····················· | ···················· | ···················· | ···················· |
| 22343 | MT873442.1 | \|Severe acute respiratory syndrome coro | ···················· | ···················· | ····················· | ···················· | ···················· | ···················· |
| 22344 | MT873443.1 | \|Severe acute respiratory syndrome coro | ···················· | ···················· | ····················· | ···················· | ···················· | ···················· |
| 22345 | MT873444.1 | \|Severe acute respiratory syndrome coro | ···················· | ···················· | ····················· | ···················· | ···················· | ···················· |
| 22346 | MT873445.1 | \|Severe acute respiratory syndrome coro | ···················· | ···················· | ····················· | ···················· | ···················· | ···················· |
| 22347 | MT873446.1 | \|Severe acute respiratory syndrome coro | ···················· | ···················· | ····················· | ···················· | ···················· | ···················· |
| 22348 | MT873447.1 | \|Severe acute respiratory syndrome coro | ···················· | ···················· | ····················· | ···················· | ···················· | ···················· |
| 22349 | MT873448.1 | \|Severe acute respiratory syndrome coro | ···················· | ···················· | ····················· | ···················· | ···················· | ···················· |
| 22350 | MT873450.1 | \|Severe acute respiratory syndrome coro | ···················· | ···················· | ····················· | ···················· | ···················· | ···················· |
| 22351 | MT873451.1 | \|Severe acute respiratory syndrome coro | ···················· | ···················· | ····················· | ···················· | ···················· | ···················· |
| 22352 | MT873452.1 | \|Severe acute respiratory syndrome coro | ···················· | ···················· | ····················· | ···················· | ···················· | ···················· |
| 22353 | MT873453.1 | \|Severe acute respiratory syndrome coro | ···················· | ···················· | ····················· | ···················· | ···················· | ···················· |
| 22354 | MT873454.1 | \|Severe acute respiratory syndrome coro | ···················· | ···················· | ····················· | ···················· | ···················· | ···················· |
| 22355 | MT873455.1 | \|Severe acute respiratory syndrome coro | ···················· | ···················· | ····················· | ···················· | ···················· | ···················· |
| 22356 | MT873456.1 | \|Severe acute respiratory syndrome coro | ···················· | ···················· | ····················· | ···················· | ···················· | ···················· |
| 22357 | MT873457.1 | \|Severe acute respiratory syndrome coro | ···················· | ···················· | ····················· | ···················· | ···················· | ···················· |
| 22358 | MT873458.1 | \|Severe acute respiratory syndrome coro | ···················· | ···················· | ····················· | ···················· | ···················· | ···················· |
| 22359 | MT873459.1 | \|Severe acute respiratory syndrome coro | ···················· | ···················· | ····················· | ···················· | ···················· | ···················· |
| 22360 | MT873460.1 | \|Severe acute respiratory syndrome coro | ···················· | ···················· | ····················· | ···················· | ···················· | ···················· |
| 22361 | MT873461.1 | \|Severe acute respiratory syndrome coro | ···················· | ···················· | ····················· | ···················· | ···················· | ···················· |
| 22362 | MT873462.1 | \|Severe acute respiratory syndrome coro | ···················· | ···················· | ····················· | ···················· | ···················· | ···················· |
| 22363 | MT873463.1 | \|Severe acute respiratory syndrome coro | ···················· | ···················· | ····················· | ···················· | ···················· | ···················· |
| 22364 | MT873464.1 | \|Severe acute respiratory syndrome coro | ···················· | ···················· | ····················· | ···················· | ···················· | ···················· |
| 22365 | MT873465.1 | \|Severe acute respiratory syndrome coro | ···················· | ···················· | ····················· | ···················· | ···················· | ···················· |
| 22366 | MT873466.1 | \|Severe acute respiratory syndrome coro | ···················· | ···················· | ····················· | ···················· | ···················· | ···················· |
| 22367 | MT873467.1 | \|Severe acute respiratory syndrome coro | ···················· | ···················· | ····················· | ···················· | ···················· | ···················· |
| 22368 | MT873468.1 | \|Severe acute respiratory syndrome coro | ···················· | ···················· | ····················· | ···················· | ···················· | ···················· |
| 22369 | MT873469.1 | \|Severe acute respiratory syndrome coro | ···················· | ···················· | ····················· | ···················· | ···················· | ···················· |
| 22370 | MT873470.1 | \|Severe acute respiratory syndrome coro | ···················· | ···················· | ····················· | ···················· | ···················· | ···················· |
| 22371 | MT873471.1 | \|Severe acute respiratory syndrome coro | ···················· | ···················· | ····················· | ···················· | ···················· | ···················· |
| 22372 | MT873472.1 | \|Severe acute respiratory syndrome coro | ···················· | ···················· | ····················· | ···················· | ···················· | ···················· |
| 22373 | MT873473.1 | \|Severe acute respiratory syndrome coro | ···················· | ···················· | ····················· | ···················· | ···················· | ···················· |
| 22374 | MT873475.1 | \|Severe acute respiratory syndrome coro | ···················· | ···················· | ····················· | ···················· | ···················· | ···················· |
| 22375 | MT873476.1 | \|Severe acute respiratory syndrome coro | ···················· | ···················· | ····················· | ···················· | ···················· | ···················· |
| 22376 | MT873477.1 | \|Severe acute respiratory syndrome coro | ···················· | ···················· | ····················· | ···················· | ···················· | ···················· |
| 22377 | MT873479.1 | \|Severe acute respiratory syndrome coro | ···················· | ···················· | ····················· | ···················· | ···················· | ···················· |
| 22378 | MT873480.1 | \|Severe acute respiratory syndrome coro | ···················· | ···················· | ····················· | ···················· | ···················· | ···················· |
| 22379 | MT873481.1 | \|Severe acute respiratory syndrome coro | ···················· | ·····A·············· | ····················· | ···················· | ···················· | ···················· |
| 22380 | MT873482.1 | \|Severe acute respiratory syndrome coro | ···················· | ···················· | ····················· | ···················· | ···················· | ···················· |
| 22381 | MT873484.1 | \|Severe acute respiratory syndrome coro | ···················· | ···················· | ····················· | ···················· | ···················· | ···················· |
| 22382 | MT873485.1 | \|Severe acute respiratory syndrome coro | ···················· | ···················· | ····················· | ···················· | ···················· | ···················· |
| 22383 | MT873486.1 | \|Severe acute respiratory syndrome coro | ···················· | ···················· | ····················· | ···················· | ···················· | ···················· |
| 22384 | MT873487.1 | \|Severe acute respiratory syndrome coro | ···················· | ···················· | ····················· | ···················· | ···················· | ···················· |
| 22385 | MT873488.1 | \|Severe acute respiratory syndrome coro | ···················· | ···················· | ····················· | ···················· | ···················· | ···················· |
| 22386 | MT873489.1 | \|Severe acute respiratory syndrome coro | ···················· | ···················· | ····················· | ···················· | ···················· | ···················· |
| 22387 | MT873490.1 | \|Severe acute respiratory syndrome coro | ···················· | ···················· | ····················· | ···················· | ···················· | ···················· |
| 22388 | MT873491.1 | \|Severe acute respiratory syndrome coro | ···················· | ···················· | ····················· | ···················· | ···················· | ···················· |
| 22389 | MT873493.1 | \|Severe acute respiratory syndrome coro | ···················· | ···················· | ····················· | ···················· | ···················· | ···················· |
| 22390 | MT873494.1 | \|Severe acute respiratory syndrome coro | ···················· | ···················· | ····················· | ···················· | ···················· | ···················· |
| 22391 | MT873496.1 | \|Severe acute respiratory syndrome coro | ···················· | ···················· | ····················· | ···················· | ···················· | ···················· |
| 22392 | MT873497.1 | \|Severe acute respiratory syndrome coro | ···················· | ···················· | ····················· | ···················· | ···················· | ···················· |
| 22393 | MT873498.1 | \|Severe acute respiratory syndrome coro | ···················· | ···················· | ····················· | ···················· | ···················· | ···················· |
| 22394 | MT873499.1 | \|Severe acute respiratory syndrome coro | ···················· | ···················· | ····················· | ···················· | ···················· | ···················· |
| 22395 | MT873500.1 | \|Severe acute respiratory syndrome coro | ···················· | ···················· | ····················· | ···················· | ···················· | ···················· |
| 22396 | MT873501.1 | \|Severe acute respiratory syndrome coro | ···················· | ···················· | ····················· | ···················· | ···················· | ···················· |
| 22397 | MT873503.1 | \|Severe acute respiratory syndrome coro | ···················· | ···················· | ····················· | ···················· | ···················· | ···················· |
| 22398 | MT873504.1 | \|Severe acute respiratory syndrome coro | ···················· | ···················· | ····················· | ···················· | ···················· | ···················· |
| 22399 | MT873505.1 | \|Severe acute respiratory syndrome coro | ···················· | ···················· | ····················· | ···················· | ···················· | ···················· |
| 22400 | MT873506.1 | \|Severe acute respiratory syndrome coro | ···················· | ···················· | ····················· | ···················· | ···················· | ···················· |
| 22401 | MT873507.1 | \|Severe acute respiratory syndrome coro | ···················· | ···················· | ····················· | ···················· | ···················· | ···················· |
| 22402 | MT873509.1 | \|Severe acute respiratory syndrome coro | ···················· | ···················· | ····················· | ···················· | ···················· | ···················· |
| 22403 | MT873510.1 | \|Severe acute respiratory syndrome coro | ···················· | ···················· | ····················· | ···················· | ···················· | ···················· |
| 22404 | MT875354.1 | \|Severe acute respiratory syndrome coro | ···················· | ·A·················· | ····················· | ···················· | ···················· | ···················· |
| 22405 | MT875355.1 | \|Severe acute respiratory syndrome coro | ···················· | ·A·················· | ····················· | ···················· | ···················· | ···················· |
| 22406 | MT875356.1 | \|Severe acute respiratory syndrome coro | ···················· | ·A·················· | ····················· | ···················· | ···················· | ···················· |

| | | | | | | | | | |
|---|---|---|---|---|---|---|---|---|---|
| 22407 | MT875357.1 | \|Severe acute respiratory syndrome coro | ···················· | ·A·················· | ···················· | ···················· | ···················· | ···················· | |
| 22408 | MT875358.1 | \|Severe acute respiratory syndrome coro | ···················· | ·A·················· | ···················· | ···················· | ···················· | ···················· | |
| 22409 | MT875359.1 | \|Severe acute respiratory syndrome coro | ···················· | ·A·················· | ···················· | ···················· | ···················· | ···················· | |
| 22410 | MT875360.1 | \|Severe acute respiratory syndrome coro | ···················· | ·A·················· | ···················· | ···················· | ···················· | ···················· | |
| 22411 | MT876431.1 | \|Severe acute respiratory syndrome coro | ···················· | ···················· | ···················· | ···················· | ···················· | ···················· | |
| 22412 | MT876432.1 | \|Severe acute respiratory syndrome coro | ···················· | ···················· | ···················· | ···················· | ···················· | ···················· | |
| 22413 | MT876433.1 | \|Severe acute respiratory syndrome coro | ···················· | ···················· | ···················· | ···················· | ···················· | ···················· | |
| 22414 | MT876525.1 | \|Severe acute respiratory syndrome coro | ···················· | ···················· | ···················· | ···················· | ···················· | ···················· | |
| 22415 | MT876526.1 | \|Severe acute respiratory syndrome coro | ···················· | ···················· | ···················· | ···················· | ···················· | ···················· | |
| 22416 | MT876527.1 | \|Severe acute respiratory syndrome coro | ···················· | ···················· | ···················· | ···················· | ···················· | ···················· | |
| 22417 | MT876546.1 | \|Severe acute respiratory syndrome coro | ···················· | ···················· | ···················· | ···················· | ···················· | ···················· | |
| 22418 | MT876547.1 | \|Severe acute respiratory syndrome coro | ···················· | ···················· | ···················· | ···················· | ···················· | ···················· | |
| 22419 | MT876548.1 | \|Severe acute respiratory syndrome coro | ···················· | ···················· | ···················· | ···················· | ···················· | ···················· | |
| 22420 | MT876554.1 | \|Severe acute respiratory syndrome coro | ···················· | ···················· | ···················· | ···················· | ···················· | ···················· | |
| 22421 | MT876555.1 | \|Severe acute respiratory syndrome coro | ···················· | ···················· | ···················· | ···················· | ···················· | ···················· | |
| 22422 | MT876556.1 | \|Severe acute respiratory syndrome coro | ···················· | ···················· | ···················· | ···················· | ···················· | ···················· | |
| 22423 | MT876571.1 | \|Severe acute respiratory syndrome coro | ···················· | ···················· | ···················· | ···················· | ···················· | ···················· | |
| 22424 | MT876572.1 | \|Severe acute respiratory syndrome coro | ···················· | ···················· | ···················· | ···················· | ···················· | ···················· | |
| 22425 | MT876598.1 | \|Severe acute respiratory syndrome coro | ···················· | ···················· | ···················· | ···················· | ···················· | ···················· | |
| 22426 | MT876599.1 | \|Severe acute respiratory syndrome coro | ···················· | ···················· | ···················· | ···················· | ···················· | ···················· | |
| 22427 | MT876606.1 | \|Severe acute respiratory syndrome coro | ···················· | ···················· | ···················· | ···················· | ···················· | ···················· | |
| 22428 | MT876607.1 | \|Severe acute respiratory syndrome coro | ···················· | ···················· | ···················· | ···················· | ···················· | ···················· | |
| 22429 | MT877256.1 | \|Severe acute respiratory syndrome coro | ···················· | ···················· | ···················· | ···················· | ···················· | ···················· | |
| 22430 | MT877296.1 | \|Severe acute respiratory syndrome coro | ···················· | ···················· | ···················· | ···················· | ···················· | ···················· | |
| 22431 | MT877298.1 | \|Severe acute respiratory syndrome coro | ···················· | ···················· | ···················· | ···················· | ···················· | ···················· | |
| 22432 | MT877299.1 | \|Severe acute respiratory syndrome coro | ···················· | ···················· | ···················· | ···················· | ···················· | ···················· | |
| 22433 | MT877300.1 | \|Severe acute respiratory syndrome coro | ···················· | ···················· | ···················· | ···················· | ···················· | ···················· | |
| 22434 | MT877302.1 | \|Severe acute respiratory syndrome coro | ···················· | ···················· | ···················· | ···················· | ···················· | ···················· | |
| 22435 | MT877304.1 | \|Severe acute respiratory syndrome coro | ···················· | ···················· | ···················· | ···················· | ···················· | ···················· | |
| 22436 | MT877305.1 | \|Severe acute respiratory syndrome coro | ···················· | ···················· | ···················· | ···················· | ···················· | ···················· | |
| 22437 | MT877306.1 | \|Severe acute respiratory syndrome coro | ···················· | ···················· | ···················· | ···················· | ···················· | ···················· | |
| 22438 | MT877309.1 | \|Severe acute respiratory syndrome coro | ···················· | ···················· | ···················· | ···················· | ···················· | ···················· | |
| 22439 | MT877315.1 | \|Severe acute respiratory syndrome coro | ···················· | ···················· | ···················· | ···················· | ···················· | ···················· | |
| 22440 | MT877316.1 | \|Severe acute respiratory syndrome coro | ···················· | ···················· | ···················· | ···················· | ···················· | ···················· | |
| 22441 | MT877320.1 | \|Severe acute respiratory syndrome coro | ···················· | ···················· | ···················· | ···················· | ···················· | ···················· | |
| 22442 | MT877324.1 | \|Severe acute respiratory syndrome coro | ···················· | ···················A | ···················· | ···················· | ···················· | ···················· | 1 |
| 22443 | MT877328.1 | \|Severe acute respiratory syndrome coro | ···················· | ···················· | ···················· | ···················· | ···················· | ···················· | |
| 22444 | MT877329.1 | \|Severe acute respiratory syndrome coro | ···················· | ···················· | ···················· | ···················· | ···················· | ···················· | |
| 22445 | MT877330.1 | \|Severe acute respiratory syndrome coro | ···················· | ···················· | ···················· | ···················· | ···················· | ···················· | |
| 22446 | MT877331.1 | \|Severe acute respiratory syndrome coro | ···················· | ···················· | ···················· | ···················· | ···················· | ···················· | |
| 22447 | MT877333.1 | \|Severe acute respiratory syndrome coro | ···················· | ···················· | ···················· | ···················· | ···················· | ···················· | |
| 22448 | MT877335.1 | \|Severe acute respiratory syndrome coro | ···················· | ···················· | ···················· | ···················· | ···················· | ···················· | |
| 22449 | MT877338.1 | \|Severe acute respiratory syndrome coro | ···················· | ···················· | ···················· | ···················· | ···················· | ···················· | |
| 22450 | MT877340.1 | \|Severe acute respiratory syndrome coro | ···················· | ···················· | ···················· | ···················· | ···················· | ···················· | |
| 22451 | MT877357.1 | \|Severe acute respiratory syndrome coro | ···················· | ···················· | ···················· | ···················· | ···················· | ···················· | |
| 22452 | MT877359.1 | \|Severe acute respiratory syndrome coro | ···················· | ···················· | ···················· | ···················· | ···················· | ···················· | |
| 22453 | MT877360.1 | \|Severe acute respiratory syndrome coro | ···················· | ···················A | ···················· | ···················· | ···················· | ···················· | 1 |
| 22454 | MT877361.1 | \|Severe acute respiratory syndrome coro | ···················· | ···················· | ···················· | ···················· | ···················· | ···················· | |
| 22455 | MT877365.1 | \|Severe acute respiratory syndrome coro | ···················· | ···················· | ···················· | ···················· | ···················· | ···················· | |
| 22456 | MT877366.1 | \|Severe acute respiratory syndrome coro | ···················· | ···················· | ···················· | ···················· | ···················· | ···················· | |
| 22457 | MT877379.1 | \|Severe acute respiratory syndrome coro | ···················· | ···················· | ···················· | ···················· | ···················· | ···················· | |
| 22458 | MT877381.1 | \|Severe acute respiratory syndrome coro | ···················· | ···················· | ···················· | ···················· | ···················· | ···················· | |
| 22459 | MT877383.1 | \|Severe acute respiratory syndrome coro | ···················· | ···················· | ···················· | ···················· | ···················· | ···················· | |
| 22460 | MT877409.1 | \|Severe acute respiratory syndrome coro | ···················· | ···················· | ···················· | ···················· | ···················· | ···················· | |
| 22461 | MT877418.1 | \|Severe acute respiratory syndrome coro | ···················· | ···················· | ···················· | ···················· | ···················· | ···················· | |
| 22462 | MT872492.1 | \|Severe acute respiratory syndrome coro | ···················· | ···················· | ···················· | ···················· | ···················· | ···················· | |
| 22463 | MT872493.1 | \|Severe acute respiratory syndrome coro | ···················· | ···················· | ···················· | ···················· | ···················· | ···················· | |
| 22464 | MT872494.1 | \|Severe acute respiratory syndrome coro | ···················· | ···················· | ···················· | ···················· | ···················· | ···················· | |
| 22465 | MT872495.1 | \|Severe acute respiratory syndrome coro | ···················· | ···················· | ···················· | ···················· | ···················· | ···················· | |
| 22466 | MT872496.1 | \|Severe acute respiratory syndrome coro | ···················· | ···················· | ···················· | ···················· | ···················· | ···················· | |
| 22467 | MT872497.1 | \|Severe acute respiratory syndrome coro | ···················· | ···················· | ···················· | ···················· | ···················· | ···················· | |
| 22468 | MT872498.1 | \|Severe acute respiratory syndrome coro | ···················· | ···················· | ···················· | ···················· | ···················· | ···················· | |
| 22469 | MT872499.1 | \|Severe acute respiratory syndrome coro | ···················· | ···················· | ···················· | ···················· | ···················· | ···················· | |
| 22470 | MT872501.1 | \|Severe acute respiratory syndrome coro | ···················· | ···················· | ···················· | ···················· | ···················· | ···················· | |
| 22471 | MT872502.1 | \|Severe acute respiratory syndrome coro | ···················· | ···················· | ···················· | ···················· | ···················· | ···················· | |
| 22472 | MT873801.1 | \|Severe acute respiratory syndrome coro | ···················· | ···················· | ···················· | ···················· | ···················· | ···················· | |
| 22473 | MT873803.1 | \|Severe acute respiratory syndrome coro | ···················· | ···················· | ···················· | ···················· | ···················· | ···················· | |
| 22474 | MT873807.1 | \|Severe acute respiratory syndrome coro | ···················· | ···················· | ···················· | ···················· | ···················· | ···················· | |
| 22475 | MT873812.1 | \|Severe acute respiratory syndrome coro | ···················· | ···················· | ···················· | ···················· | ···················· | ···················· | |
| 22476 | MT873821.1 | \|Severe acute respiratory syndrome coro | ···················· | ···················· | ···················· | ···················· | ···················· | ···················· | |
| 22477 | MT873827.1 | \|Severe acute respiratory syndrome coro | ···················· | ···················· | ···················· | ···················· | ···················· | ···················· | |
| 22478 | MT873890.1 | \|Severe acute respiratory syndrome coro | ···················· | ···················· | ···················· | ···················· | ···················· | ···················· | |
| 22479 | MT873892.1 | \|Severe acute respiratory syndrome coro | ···················· | ···················· | ···················· | ···················· | ···················· | ···················· | |
| 22480 | MT873898.1 | \|Severe acute respiratory syndrome coro | ···················· | ···················· | ···················· | ···················· | ···················· | ···················· | |
| 22481 | MT873923.1 | \|Severe acute respiratory syndrome coro | ···················· | ···················· | ···················· | ···················· | ···················· | ···················· | |
| 22482 | LC572067.1 | \|Severe acute respiratory syndrome coro | ···················· | ···················· | ···················· | ···················· | ···················· | ···················· | |
| 22483 | LC572068.1 | \|Severe acute respiratory syndrome coro | ···················· | ···················· | ···················· | ···················· | ···················· | ···················· | |
| 22484 | LC572069.1 | \|Severe acute respiratory syndrome coro | ···················· | ···················· | ···················· | ···················· | ···················· | ···················· | |

| | | | | | | | | |
|---|---|---|---|---|---|---|---|---|
| 22485 | LC572070.1 | Severe acute respiratory syndrome coro | ···················· | ···················· | ···················· | ···················· | ···················· | ···················· |
| 22486 | MT856452.1 | Severe acute respiratory syndrome coro | ···················· | ···················· | ···················· | ···················· | ···················· | ···················· |
| 22487 | MT856453.1 | Severe acute respiratory syndrome coro | ···················· | ···················· | ···················· | ···················· | ···················· | ···················· |
| 22488 | MT856454.1 | Severe acute respiratory syndrome coro | ···················· | ···················· | ···················· | ···················· | ···················· | ···················· |
| 22489 | MT856455.1 | Severe acute respiratory syndrome coro | ···················· | ···················· | ···················· | ···················· | ···················· | ···················· |
| 22490 | MT856456.1 | Severe acute respiratory syndrome coro | ···················· | ···················· | ···················· | ···················· | ···················· | ···················· |
| 22491 | MT856457.1 | Severe acute respiratory syndrome coro | ···················· | ···················· | ···················· | ···················· | ···················· | ···················· |
| 22492 | MT856458.1 | Severe acute respiratory syndrome coro | ···················· | ···················· | ···················· | ···················· | ···················· | ···················· |
| 22493 | MT856459.1 | Severe acute respiratory syndrome coro | ···················· | ···················· | ···················· | ···················· | ···················· | ···················· |
| 22494 | MT856678.1 | Severe acute respiratory syndrome coro | ···················· | ·········A·········· | ···················· | ···················· | ···················· | ···················· |
| 22495 | MT856679.1 | Severe acute respiratory syndrome coro | ···················· | ·········A·········· | ···················· | ···················· | ···················· | ···················· |
| 22496 | MT856680.1 | Severe acute respiratory syndrome coro | ···················· | ·········A·········· | ···················· | ···················· | ···················· | ···················· |
| 22497 | MT856682.1 | Severe acute respiratory syndrome coro | ···················· | ·········N·········· | ···················· | ···················· | ···················· | ···················· |
| 22498 | MT856683.1 | Severe acute respiratory syndrome coro | ···················· | ·········N·········· | ···················· | ···················· | ···················· | ···················· |
| 22499 | MT856684.1 | Severe acute respiratory syndrome coro | ···················· | ··············C····· | ···················· | ···················· | ···················· | ···················· |
| 22500 | MT856685.1 | Severe acute respiratory syndrome coro | ···················· | ···················· | ···················· | ···················· | ···················· | ···················· |
| 22501 | MT856686.1 | Severe acute respiratory syndrome coro | ···················· | ··············C····· | ···················· | ···················· | ···················· | ···················· |
| 22502 | MT856687.1 | Severe acute respiratory syndrome coro | ···················· | ···················· | ···················· | ···················· | ···················· | ···················· |
| 22503 | MT856688.1 | Severe acute respiratory syndrome coro | ···················· | ···················· | ···················· | ···················· | ···················· | ···················· |
| 22504 | MT856689.1 | Severe acute respiratory syndrome coro | ···················· | ·········A·········· | ···················· | ···················· | ···················· | ···················· |
| 22505 | MT856690.1 | Severe acute respiratory syndrome coro | ···················· | ·········A·········· | ···················· | ···················· | ···················· | ···················· |
| 22506 | MT856691.1 | Severe acute respiratory syndrome coro | ···················· | ·········N·········· | ···················· | ···················· | ···················· | ···················· |
| 22507 | MT846004.1 | Severe acute respiratory syndrome coro | ···················· | ···················· | ···················· | ···················· | ···················· | ···················· |
| 22508 | MT846005.1 | Severe acute respiratory syndrome coro | ···················· | ···················· | ···················· | ···················· | ···················· | ···················· |
| 22509 | MT846006.1 | Severe acute respiratory syndrome coro | ···················· | ···················· | ···················· | ···················· | ···················· | ···················· |
| 22510 | MT846007.1 | Severe acute respiratory syndrome coro | ···················· | ···················· | ···················· | ···················· | ···················· | ···················· |
| 22511 | MT846008.1 | Severe acute respiratory syndrome coro | ···················· | ···················· | ···················· | ···················· | ···················· | ···················· |
| 22512 | MT846009.1 | Severe acute respiratory syndrome coro | ···················· | ···················· | ···················· | ···················· | ···················· | ···················· |
| 22513 | MT846010.1 | Severe acute respiratory syndrome coro | ···················· | ···················· | ···················· | ···················· | ···················· | ···················· |
| 22514 | MT846011.1 | Severe acute respiratory syndrome coro | ···················· | ···················· | ···················· | ···················· | ···················· | ···················· |
| 22515 | MT846012.1 | Severe acute respiratory syndrome coro | ···················· | ···················· | ···················· | ···················· | ···················· | ···················· |
| 22516 | MT846013.1 | Severe acute respiratory syndrome coro | ···················· | ···················· | ···················· | ···················· | ···················· | ···················· |
| 22517 | MT846014.1 | Severe acute respiratory syndrome coro | ···················· | ···················· | ···················· | ···················· | ···················· | ···················· |
| 22518 | MT846015.1 | Severe acute respiratory syndrome coro | ···················· | ···················· | ···················· | ···················· | ···················· | ···················· |
| 22519 | MT846413.1 | Severe acute respiratory syndrome coro | ···················· | ···················· | ···················· | ···················· | ···················· | ···················· |
| 22520 | MT846415.1 | Severe acute respiratory syndrome coro | ···················· | ···················· | ···················· | ···················· | ···················· | ···················· |
| 22521 | MT846417.1 | Severe acute respiratory syndrome coro | ···················· | ···················· | ···················· | ···················· | ···················· | ···················· |
| 22522 | MT846418.1 | Severe acute respiratory syndrome coro | ···················· | ···················· | ···················· | ···················· | ···················· | ···················· |
| 22523 | MT846420.1 | Severe acute respiratory syndrome coro | ···················· | ···················· | ···················· | ···················· | ···················· | ···················· |
| 22524 | MT846421.1 | Severe acute respiratory syndrome coro | ···················· | ···················· | ···················· | ···················· | ···················· | ···················· |
| 22525 | MT846422.1 | Severe acute respiratory syndrome coro | ···················· | ···················· | ···················· | ···················· | ···················· | ···················· |
| 22526 | MT846423.1 | Severe acute respiratory syndrome coro | ···················· | ···················· | ···················· | ···················· | ···················· | ···················· |
| 22527 | MT846425.1 | Severe acute respiratory syndrome coro | ···················· | ···················· | ···················· | ···················· | ···················· | ···················· |
| 22528 | MT846426.1 | Severe acute respiratory syndrome coro | ···················· | ···················· | ···················· | ···················· | ···················· | ···················· |
| 22529 | MT846428.1 | Severe acute respiratory syndrome coro | ···················· | ···················· | ···················· | ···················· | ···················· | ···················· |
| 22530 | MT846429.1 | Severe acute respiratory syndrome coro | ···················· | ···················· | ···················· | ···················· | ···················· | ···················· |
| 22531 | MT846430.1 | Severe acute respiratory syndrome coro | ···················· | ···················· | ···················· | ···················· | ···················· | ···················· |
| 22532 | MT846431.1 | Severe acute respiratory syndrome coro | ···················· | ···················· | ···················· | ···················· | ···················· | ···················· |
| 22533 | MT846432.1 | Severe acute respiratory syndrome coro | ···················· | ···················· | ···················· | ···················· | ···················· | ···················· |
| 22535 | MT846434.1 | Severe acute respiratory syndrome coro | ···················· | ···················· | ···················· | ···················· | ···················· | ···················· |
| 22536 | MT846435.1 | Severe acute respiratory syndrome coro | ···················· | ···················· | ···················· | ···················· | ···················· | ···················· |
| 22537 | MT846436.1 | Severe acute respiratory syndrome coro | ···················· | ···················· | ···················· | ···················· | ···················· | ···················· |
| 22538 | MT846438.1 | Severe acute respiratory syndrome coro | ···················· | ···················· | ···················· | ···················· | ···················· | ···················· |
| 22539 | MT846439.1 | Severe acute respiratory syndrome coro | ···················· | ···················· | ···················· | ···················· | ···················· | ···················· |
| 22540 | MT846440.1 | Severe acute respiratory syndrome coro | ···················· | ···················· | ···················· | ···················· | ···················· | ···················· |
| 22541 | MT846442.1 | Severe acute respiratory syndrome coro | ···················· | ···················· | ···················· | ···················· | ···················· | ···················· |
| 22542 | MT846443.1 | Severe acute respiratory syndrome coro | ···················· | ···················· | ···················· | ···················· | ···················· | ···················· |
| 22543 | MT846444.1 | Severe acute respiratory syndrome coro | ···················· | ···················· | ···················· | ···················· | ···················· | ···················· |
| 22544 | MT846445.1 | Severe acute respiratory syndrome coro | ···················· | ···················· | ···················· | ···················· | ···················· | ···················· |
| 22545 | MT846446.1 | Severe acute respiratory syndrome coro | ···················· | ···················· | ···················· | ···················· | ···················· | ···················· |
| 22546 | MT846449.1 | Severe acute respiratory syndrome coro | ···················· | ···················· | ···················· | ···················· | ···················· | ···················· |
| 22548 | MT846451.1 | Severe acute respiratory syndrome coro | ···················· | ···················· | ···················· | ···················· | ···················· | ···················· |
| 22549 | MT846452.1 | Severe acute respiratory syndrome coro | ···················· | ···················· | ···················· | ···················· | ···················· | ···················· |
| 22551 | MT846455.1 | Severe acute respiratory syndrome coro | ···················· | ···················· | ···················· | ···················· | ···················· | ···················· |
| 22552 | MT846456.1 | Severe acute respiratory syndrome coro | ···················· | ···················· | ···················· | ···················· | ···················· | ···················· |
| 22553 | MT846457.1 | Severe acute respiratory syndrome coro | ···················· | ···················· | ···················· | ···················· | ···················· | ···················· |
| 22554 | MT846460.1 | Severe acute respiratory syndrome coro | ···················· | ···················· | ···················· | ···················· | ···················· | ···················· |
| 22555 | MT846461.1 | Severe acute respiratory syndrome coro | ···················· | ···················· | ···················· | ···················· | ···················· | ···················· |
| 22556 | MT846462.1 | Severe acute respiratory syndrome coro | ···················· | ···················· | ···················· | ···················· | ···················· | ···················· |
| 22557 | MT846463.1 | Severe acute respiratory syndrome coro | ···················· | ···················· | ···················· | ···················· | ···················· | ···················· |
| 22558 | MT846464.1 | Severe acute respiratory syndrome coro | ···················· | ···················· | ···················· | ···················· | ···················· | ···················· |
| 22559 | MT846465.1 | Severe acute respiratory syndrome coro | ···················· | ···················· | ···················· | ···················· | ···················· | ···················· |
| 22560 | MT846467.1 | Severe acute respiratory syndrome coro | ···················· | ···················· | ···················· | ···················· | ···················· | ···················· |
| 22561 | MT846468.1 | Severe acute respiratory syndrome coro | ···················· | ···················· | ···················· | ···················· | ···················· | ···················· |
| 22562 | MT846469.1 | Severe acute respiratory syndrome coro | ···················· | ···················· | ···················· | ···················· | ···················· | ···················· |
| 22563 | MT846470.1 | Severe acute respiratory syndrome coro | ···················· | ···················· | ···················· | ···················· | ···················· | ···················· |
| 22564 | MT846471.1 | Severe acute respiratory syndrome coro | ···················· | ···················· | ···················· | ···················· | ···················· | ···················· |
| 22565 | MT846473.1 | Severe acute respiratory syndrome coro | ···················· | ···················· | ···················· | ···················· | ···················· | ···················· |

| | | | | | | | | |
|---|---|---|---|---|---|---|---|---|
| 22566 | MT846475.1 | \|Severe acute respiratory syndrome coro | .................... | .................... | ..................... | .................... | .................... | .................... |
| 22567 | MT846476.1 | \|Severe acute respiratory syndrome coro | .................... | .................... | ..................... | .................... | .................... | .................... |
| 22568 | MT846477.1 | \|Severe acute respiratory syndrome coro | .................... | .................... | ..................... | .................... | .................... | .................... |
| 22569 | MT846478.1 | \|Severe acute respiratory syndrome coro | .................... | .................... | ..................... | .................... | .................... | .................... |
| 22570 | MT846480.1 | \|Severe acute respiratory syndrome coro | .................... | .................... | ..................... | .................... | .................... | .................... |
| 22571 | MT846481.1 | \|Severe acute respiratory syndrome coro | .................... | .................... | ..................... | .................... | .................... | .................... |
| 22572 | MT846483.1 | \|Severe acute respiratory syndrome coro | .................... | .................... | ..................... | .................... | .................... | .................... |
| 22573 | MT846484.1 | \|Severe acute respiratory syndrome coro | .................... | .................... | ..................... | .................... | .................... | .................... |
| 22574 | MT846485.1 | \|Severe acute respiratory syndrome coro | .................... | .................... | ..................... | .................... | .................... | .................... |
| 22575 | MT846486.1 | \|Severe acute respiratory syndrome coro | .................... | .................... | ..................... | .................... | .................... | .................... |
| 22576 | MT846488.1 | \|Severe acute respiratory syndrome coro | .................... | .................... | ..................... | .................... | .................... | .................... |
| 22577 | MT846489.1 | \|Severe acute respiratory syndrome coro | .................... | .................... | ..................... | .................... | .................... | .................... |
| 22578 | MT846490.1 | \|Severe acute respiratory syndrome coro | .................... | .................... | ..................... | .................... | .................... | .................... |
| 22579 | MT846491.1 | \|Severe acute respiratory syndrome coro | .................... | .................... | ..................... | .................... | .................... | .................... |
| 22580 | MT846492.1 | \|Severe acute respiratory syndrome coro | .................... | .................... | ..................... | .................... | .................... | .................... |
| 22581 | MT846494.1 | \|Severe acute respiratory syndrome coro | .................... | .................... | ..................... | .................... | .................... | .................... |
| 22582 | MT846499.1 | \|Severe acute respiratory syndrome coro | .................... | .................... | ..................... | .................... | .................... | .................... |
| 22583 | MT846500.1 | \|Severe acute respiratory syndrome coro | .................... | .................... | ..................... | .................... | .................... | .................... |
| 22584 | MT846501.1 | \|Severe acute respiratory syndrome coro | .................... | .................... | ..................... | .................... | .................... | .................... |
| 22585 | MT846502.1 | \|Severe acute respiratory syndrome coro | .................... | .................... | ..................... | .................... | .................... | .................... |
| 22586 | MT846503.1 | \|Severe acute respiratory syndrome coro | .................... | .................... | ..................... | .................... | .................... | .................... |
| 22587 | MT846504.1 | \|Severe acute respiratory syndrome coro | .................... | .................... | ..................... | .................... | .................... | .................... |
| 22588 | MT846505.1 | \|Severe acute respiratory syndrome coro | .................... | .................... | ..................... | .................... | .................... | .................... |
| 22589 | MT846506.1 | \|Severe acute respiratory syndrome coro | .................... | .................... | ..................... | .................... | .................... | .................... |
| 22590 | MT846507.1 | \|Severe acute respiratory syndrome coro | .................... | .................... | ..................... | .................... | .................... | .................... |
| 22591 | MT846508.1 | \|Severe acute respiratory syndrome coro | .................... | .................... | ..................... | .................... | .................... | .................... |
| 22592 | MT846509.1 | \|Severe acute respiratory syndrome coro | .................... | .................... | ..................... | .................... | .................... | .................... |
| 22593 | MT846510.1 | \|Severe acute respiratory syndrome coro | .................... | .................... | ..................... | .................... | .................... | .................... |
| 22594 | MT846511.1 | \|Severe acute respiratory syndrome coro | .................... | .................... | ..................... | .................... | .................... | .................... |
| 22595 | MT846512.1 | \|Severe acute respiratory syndrome coro | .................... | .................... | ..................... | .................... | .................... | .................... |
| 22596 | MT846513.1 | \|Severe acute respiratory syndrome coro | .................... | .................... | ..................... | .................... | .................... | .................... |
| 22597 | MT846514.1 | \|Severe acute respiratory syndrome coro | .................... | .................... | ..................... | .................... | .................... | .................... |
| 22598 | MT846515.1 | \|Severe acute respiratory syndrome coro | .................... | .................... | ..................... | .................... | .................... | .................... |
| 22599 | MT846517.1 | \|Severe acute respiratory syndrome coro | .................... | .................... | ..................... | .................... | .................... | .................... |
| 22600 | MT846518.1 | \|Severe acute respiratory syndrome coro | .................... | .................... | ..................... | .................... | .................... | .................... |
| 22601 | MT846519.1 | \|Severe acute respiratory syndrome coro | .................... | .................... | ..................... | .................... | .................... | .................... |
| 22602 | MT846520.1 | \|Severe acute respiratory syndrome coro | .................... | .................... | ..................... | .................... | .................... | .................... |
| 22603 | MT846521.1 | \|Severe acute respiratory syndrome coro | .................... | .................... | ..................... | .................... | .................... | .................... |
| 22604 | MT846522.1 | \|Severe acute respiratory syndrome coro | .................... | .................... | ..................... | .................... | .................... | .................... |
| 22605 | MT846523.1 | \|Severe acute respiratory syndrome coro | .................... | .................... | ..................... | .................... | .................... | .................... |
| 22606 | MT846525.1 | \|Severe acute respiratory syndrome coro | .................... | .................... | ..................... | .................... | .................... | .................... |
| 22607 | MT846526.1 | \|Severe acute respiratory syndrome coro | .................... | .................... | ..................... | .................... | .................... | .................... |
| 22608 | MT846527.1 | \|Severe acute respiratory syndrome coro | .................... | .................... | ..................... | .................... | .................... | .................... |
| 22609 | MT846528.1 | \|Severe acute respiratory syndrome coro | .................... | .................... | ..................... | .................... | .................... | .................... |
| 22610 | MT846529.1 | \|Severe acute respiratory syndrome coro | .................... | .................... | ..................... | .................... | .................... | .................... |
| 22611 | MT846531.1 | \|Severe acute respiratory syndrome coro | .................... | .................... | ..................... | .................... | .................... | .................... |
| 22612 | MT846533.1 | \|Severe acute respiratory syndrome coro | .................... | .................... | ..................... | .................... | .................... | .................... |
| 22613 | MT846534.1 | \|Severe acute respiratory syndrome coro | .................... | .................... | ..................... | .................... | .................... | .................... |
| 22614 | MT846535.1 | \|Severe acute respiratory syndrome coro | .................... | .................... | ..................... | .................... | .................... | .................... |
| 22615 | MT846537.1 | \|Severe acute respiratory syndrome coro | .................... | .................... | ..................... | .................... | .................... | .................... |
| 22616 | MT846538.1 | \|Severe acute respiratory syndrome coro | .................... | .................... | ..................... | .................... | .................... | .................... |
| 22617 | MT846539.1 | \|Severe acute respiratory syndrome coro | .................... | .................... | ..................... | .................... | .................... | .................... |
| 22618 | MT846540.1 | \|Severe acute respiratory syndrome coro | .................... | .................... | ..................... | .................... | .................... | .................... |
| 22619 | MT846541.1 | \|Severe acute respiratory syndrome coro | .................... | .................... | ..................... | .................... | .................... | .................... |
| 22620 | MT846545.1 | \|Severe acute respiratory syndrome coro | .................... | .................... | ..................... | .................... | .................... | .................... |
| 22621 | MT846547.1 | \|Severe acute respiratory syndrome coro | .................... | .................... | ..................... | .................... | .................... | .................... |
| 22622 | MT846553.1 | \|Severe acute respiratory syndrome coro | .................... | .................... | ..................... | .................... | .................... | .................... |
| 22623 | MT846554.1 | \|Severe acute respiratory syndrome coro | .................... | .................... | ..................... | .................... | .................... | .................... |
| 22624 | MT846555.1 | \|Severe acute respiratory syndrome coro | .................... | .................... | ..................... | .................... | .................... | .................... |
| 22625 | MT846556.1 | \|Severe acute respiratory syndrome coro | .................... | .................... | ..................... | .................... | .................... | .................... |
| 22626 | MT846557.1 | \|Severe acute respiratory syndrome coro | .................... | .................... | ..................... | .................... | .................... | .................... |
| 22627 | MT846558.1 | \|Severe acute respiratory syndrome coro | .................... | .................... | ..................... | .................... | .................... | .................... |
| 22628 | MT846559.1 | \|Severe acute respiratory syndrome coro | .................... | .................... | ..................... | .................... | .................... | .................... |
| 22629 | MT846560.1 | \|Severe acute respiratory syndrome coro | .................... | .................... | ..................... | .................... | .................... | .................... |
| 22630 | MT846561.1 | \|Severe acute respiratory syndrome coro | .................... | .................... | ..................... | .................... | .................... | .................... |
| 22631 | MT846563.1 | \|Severe acute respiratory syndrome coro | .................... | .................... | ..................... | .................... | .................... | .................... |
| 22632 | MT846564.1 | \|Severe acute respiratory syndrome coro | .................... | .................... | ..................... | .................... | .................... | .................... |
| 22633 | MT846565.1 | \|Severe acute respiratory syndrome coro | .................... | .................... | ..................... | .................... | .................... | .................... |
| 22634 | MT846567.1 | \|Severe acute respiratory syndrome coro | .................... | .................... | ..................... | .................... | .................... | .................... |
| 22635 | MT846568.1 | \|Severe acute respiratory syndrome coro | .................... | .................... | ..................... | .................... | .................... | .................... |
| 22636 | MT846569.1 | \|Severe acute respiratory syndrome coro | .................... | .A.................. | ..................... | .................... | .................... | .................... |
| 22637 | MT846570.1 | \|Severe acute respiratory syndrome coro | .................... | .................... | ..................... | .................... | .................... | .................... |
| 22638 | MT846571.1 | \|Severe acute respiratory syndrome coro | .................... | .................... | ..................... | .................... | .................... | .................... |
| 22639 | MT846572.1 | \|Severe acute respiratory syndrome coro | .................... | .................... | ..................... | .................... | .................... | .................... |
| 22640 | MT846573.1 | \|Severe acute respiratory syndrome coro | .................... | .................... | ..................... | .................... | .................... | .................... |
| 22641 | MT846574.1 | \|Severe acute respiratory syndrome coro | .................... | .................... | ..................... | .................... | .................... | .................... |
| 22642 | MT846575.1 | \|Severe acute respiratory syndrome coro | .................... | .................... | ..................... | .................... | .................... | .................... |
| 22643 | MT846576.1 | \|Severe acute respiratory syndrome coro | .................... | .................... | ..................... | .................... | .................... | .................... |

| | | | | | | | | |
|---|---|---|---|---|---|---|---|---|
| 22644 | MT846577.1 | \|Severe acute respiratory syndrome coro | .................... | .................... | ..................... | .................... | .................... | .................... |
| 22645 | MT846578.1 | \|Severe acute respiratory syndrome coro | .................... | .................... | ..................... | .................... | .................... | .................... |
| 22646 | MT846579.1 | \|Severe acute respiratory syndrome coro | .................... | .................... | ..................... | .................... | .................... | .................... |
| 22647 | MT846580.1 | \|Severe acute respiratory syndrome coro | .................... | .................... | ..................... | .................... | .................... | .................... |
| 22648 | MT846581.1 | \|Severe acute respiratory syndrome coro | .................... | .................... | ..................... | .................... | .................... | .................... |
| 22649 | MT846583.1 | \|Severe acute respiratory syndrome coro | .................... | .................... | ..................... | .................... | .................... | .................... |
| 22650 | MT846584.1 | \|Severe acute respiratory syndrome coro | .................... | .................... | ..................... | .................... | .................... | .................... |
| 22651 | MT846585.1 | \|Severe acute respiratory syndrome coro | .................... | .................... | ..................... | .................... | .................... | .................... |
| 22652 | MT846586.1 | \|Severe acute respiratory syndrome coro | .................... | .................... | ..................... | .................... | .................... | .................... |
| 22653 | MT846587.1 | \|Severe acute respiratory syndrome coro | .................... | .................... | ..................... | .................... | .................... | .................... |
| 22654 | MT846588.1 | \|Severe acute respiratory syndrome coro | .................... | .................... | ..................... | .................... | .................... | .................... |
| 22655 | MT846589.1 | \|Severe acute respiratory syndrome coro | .................... | .................... | ..................... | .................... | .................... | .................... |
| 22656 | MT846590.1 | \|Severe acute respiratory syndrome coro | .................... | .................... | ..................... | .................... | .................... | .................... |
| 22657 | MT846591.1 | \|Severe acute respiratory syndrome coro | .................... | .................... | ..................... | .................... | .................... | .................... |
| 22658 | MT846592.1 | \|Severe acute respiratory syndrome coro | .................... | .................... | ..................... | .................... | .................... | .................... |
| 22659 | MT846593.1 | \|Severe acute respiratory syndrome coro | .................... | .................... | ..................... | .................... | .................... | .................... |
| 22660 | MT846594.1 | \|Severe acute respiratory syndrome coro | .................... | .................... | ..................... | .................... | .................... | .................... |
| 22661 | MT846595.1 | \|Severe acute respiratory syndrome coro | .................... | .................... | ..................... | .................... | .................... | .................... |
| 22662 | MT846596.1 | \|Severe acute respiratory syndrome coro | .................... | .................... | ..................... | .................... | .................... | .................... |
| 22663 | MT846597.1 | \|Severe acute respiratory syndrome coro | .................... | .................... | ..................... | .................... | .................... | .................... |
| 22664 | MT846598.1 | \|Severe acute respiratory syndrome coro | .................... | .................... | ..................... | .................... | .................... | .................... |
| 22665 | MT846600.1 | \|Severe acute respiratory syndrome coro | .................... | .................... | ..................... | .................... | .................... | .................... |
| 22666 | MT846601.1 | \|Severe acute respiratory syndrome coro | .................... | .................... | ..................... | .................... | .................... | .................... |
| 22667 | MT846602.1 | \|Severe acute respiratory syndrome coro | .................... | .................... | ..................... | .................... | .................... | .................... |
| 22668 | MT846603.1 | \|Severe acute respiratory syndrome coro | .................... | .................... | ..................... | .................... | .................... | .................... |
| 22669 | MT846604.1 | \|Severe acute respiratory syndrome coro | .................... | .................... | ..................... | .................... | .................... | .................... |
| 22670 | MT846605.1 | \|Severe acute respiratory syndrome coro | .................... | .................... | ..................... | .................... | .................... | .................... |
| 22671 | MT846606.1 | \|Severe acute respiratory syndrome coro | .................... | .A.................. | ..................... | .................... | .................... | .................... |
| 22672 | MT846607.1 | \|Severe acute respiratory syndrome coro | .................... | .................... | ..................... | .................... | .................... | .................... |
| 22673 | MT846608.1 | \|Severe acute respiratory syndrome coro | .................... | .................... | ..................... | .................... | .................... | .................... |
| 22674 | MT846609.1 | \|Severe acute respiratory syndrome coro | .................... | .................... | ..................... | .................... | .................... | .................... |
| 22675 | MT846610.1 | \|Severe acute respiratory syndrome coro | .................... | .A.................. | ..................... | .................... | .................... | .................... |
| 22676 | MT846611.1 | \|Severe acute respiratory syndrome coro | .................... | .................... | ..................... | .................... | .................... | .................... |
| 22677 | MT846612.1 | \|Severe acute respiratory syndrome coro | .................... | .................... | ..................... | .................... | .................... | .................... |
| 22678 | MT846613.1 | \|Severe acute respiratory syndrome coro | .................... | .................... | ..................... | .................... | .................... | .................... |
| 22679 | MT846614.1 | \|Severe acute respiratory syndrome coro | .................... | .................... | ..................... | .................... | .................... | .................... |
| 22680 | MT846615.1 | \|Severe acute respiratory syndrome coro | .................... | .................... | ..................... | .................... | .................... | .................... |
| 22681 | MT846616.1 | \|Severe acute respiratory syndrome coro | .................... | .................... | ..................... | .................... | .................... | .................... |
| 22682 | MT846618.1 | \|Severe acute respiratory syndrome coro | .................... | .................... | ..................... | .................... | .................... | .................... |
| 22683 | MT846619.1 | \|Severe acute respiratory syndrome coro | .................... | .................... | ..................... | .................... | .................... | .................... |
| 22684 | MT846620.1 | \|Severe acute respiratory syndrome coro | .................... | .................... | ..................... | .................... | .................... | .................... |
| 22685 | MT846621.1 | \|Severe acute respiratory syndrome coro | .................... | .................... | ..................... | .................... | .................... | .................... |
| 22686 | MT846622.1 | \|Severe acute respiratory syndrome coro | .................... | .................... | ..................... | .................... | .................... | .................... |
| 22687 | MT846623.1 | \|Severe acute respiratory syndrome coro | .................... | .................... | ..................... | .................... | .................... | .................... |
| 22688 | MT846626.1 | \|Severe acute respiratory syndrome coro | .................... | .................... | ..................... | .................... | .................... | .................... |
| 22689 | MT846627.1 | \|Severe acute respiratory syndrome coro | .................... | .................... | ..................... | .................... | .................... | .................... |
| 22690 | MT846628.1 | \|Severe acute respiratory syndrome coro | .................... | .................... | ..................... | .................... | .................... | .................... |
| 22691 | MT846629.1 | \|Severe acute respiratory syndrome coro | .................... | .................... | ..................... | .................... | .................... | .................... |
| 22692 | MT846631.1 | \|Severe acute respiratory syndrome coro | .................... | .................... | ..................... | .................... | .................... | .................... |
| 22693 | MT846632.1 | \|Severe acute respiratory syndrome coro | .................... | .................... | ..................... | .................... | .................... | .................... |
| 22694 | MT846633.1 | \|Severe acute respiratory syndrome coro | .................... | .................... | ..................... | .................... | .................... | .................... |
| 22695 | MT846634.1 | \|Severe acute respiratory syndrome coro | .................... | .................... | ..................... | .................... | .................... | .................... |
| 22696 | MT846635.1 | \|Severe acute respiratory syndrome coro | .................... | .................... | ..................... | .................... | .................... | .................... |
| 22697 | MT846636.1 | \|Severe acute respiratory syndrome coro | .................... | .................... | ..................... | .................... | .................... | .................... |
| 22698 | MT846637.1 | \|Severe acute respiratory syndrome coro | .................... | .................... | ..................... | .................... | .................... | .................... |
| 22699 | MT846638.1 | \|Severe acute respiratory syndrome coro | .................... | .................... | ..................... | .................... | .................... | .................... |
| 22700 | MT846639.1 | \|Severe acute respiratory syndrome coro | .................... | .................... | ..................... | .................... | .................... | .................... |
| 22701 | MT846640.1 | \|Severe acute respiratory syndrome coro | .................... | .................... | ..................... | .................... | .................... | .................... |
| 22702 | MT846641.1 | \|Severe acute respiratory syndrome coro | .................... | .................... | ..................... | .................... | .................... | .................... |
| 22703 | MT846642.1 | \|Severe acute respiratory syndrome coro | .................... | .................... | ..................... | .................... | .................... | .................... |
| 22704 | MT846643.1 | \|Severe acute respiratory syndrome coro | .................... | .................... | ..................... | .................... | .................... | .................... |
| 22705 | MT846644.1 | \|Severe acute respiratory syndrome coro | .................... | .................... | ..................... | .................... | .................... | .................... |
| 22706 | MT846645.1 | \|Severe acute respiratory syndrome coro | .................... | .................... | ..................... | .................... | .................... | .................... |
| 22707 | MT846646.1 | \|Severe acute respiratory syndrome coro | .................... | .................... | ..................... | .................... | .................... | .................... |
| 22708 | MT846647.1 | \|Severe acute respiratory syndrome coro | .................... | .................... | ..................... | .................... | .................... | .................... |
| 22709 | MT846648.1 | \|Severe acute respiratory syndrome coro | .................... | .................... | ..................... | .................... | .................... | .................... |
| 22710 | MT846649.1 | \|Severe acute respiratory syndrome coro | .................... | .................... | ..................... | .................... | .................... | .................... |
| 22711 | MT846650.1 | \|Severe acute respiratory syndrome coro | .................... | .................... | ..................... | .................... | .................... | .................... |
| 22712 | MT846651.1 | \|Severe acute respiratory syndrome coro | .................... | .................... | ..................... | .................... | .................... | .................... |
| 22713 | MT846652.1 | \|Severe acute respiratory syndrome coro | .................... | .................... | ..................... | .................... | .................... | .................... |
| 22714 | MT846653.1 | \|Severe acute respiratory syndrome coro | .................... | .................... | ..................... | .................... | .................... | .................... |
| 22715 | MT846654.1 | \|Severe acute respiratory syndrome coro | .................... | .................... | ..................... | .................... | .................... | .................... |
| 22716 | MT846655.1 | \|Severe acute respiratory syndrome coro | .................... | .................... | ..................... | .................... | .................... | .................... |
| 22717 | MT846657.1 | \|Severe acute respiratory syndrome coro | .................... | .................... | ..................... | .................... | .................... | .................... |
| 22718 | MT846658.1 | \|Severe acute respiratory syndrome coro | .................... | .................... | ..................... | .................... | .................... | .................... |
| 22719 | MT846659.1 | \|Severe acute respiratory syndrome coro | .................... | .................... | ..................... | .................... | .................... | .................... |
| 22720 | MT846660.1 | \|Severe acute respiratory syndrome coro | .................... | .................... | ..................... | .................... | .................... | .................... |
| 22721 | MT846662.1 | \|Severe acute respiratory syndrome coro | .................... | .................... | ..................... | .................... | .................... | .................... |

| | | | | | | | | |
|---|---|---|---|---|---|---|---|---|
| 22722 | MT846663.1 | \|Severe acute respiratory syndrome coro | ···················· | ···················· | ···················· | ···················· | ···················· | ···················· |
| 22723 | MT846664.1 | \|Severe acute respiratory syndrome coro | ···················· | ···················· | ···················· | ···················· | ···················· | ···················· |
| 22724 | MT846665.1 | \|Severe acute respiratory syndrome coro | ···················· | ···················· | ···················· | ···················· | ···················· | ···················· |
| 22725 | MT846666.1 | \|Severe acute respiratory syndrome coro | ···················· | ···················· | ···················· | ···················· | ···················· | ···················· |
| 22726 | MT846667.1 | \|Severe acute respiratory syndrome coro | ···················· | ···················· | ···················· | ···················· | ···················· | ···················· |
| 22727 | MT846668.1 | \|Severe acute respiratory syndrome coro | ···················· | ···················· | ···················· | ···················· | ···················· | ···················· |
| 22728 | MT846669.1 | \|Severe acute respiratory syndrome coro | ···················· | ···················· | ···················· | ···················· | ···················· | ···················· |
| 22729 | MT846670.1 | \|Severe acute respiratory syndrome coro | ···················· | ···················· | ···················· | ···················· | ···················· | ···················· |
| 22730 | MT846671.1 | \|Severe acute respiratory syndrome coro | ···················· | ···················· | ···················· | ···················· | ···················· | ···················· |
| 22731 | MT846673.1 | \|Severe acute respiratory syndrome coro | ···················· | ···················· | ···················· | ···················· | ···················· | ···················· |
| 22732 | MT846674.1 | \|Severe acute respiratory syndrome coro | ···················· | ···················· | ···················· | ···················· | ···················· | ···················· |
| 22733 | MT846675.1 | \|Severe acute respiratory syndrome coro | ···················· | ·A·················· | ···················· | ···················· | ···················· | ···················· |
| 22734 | MT846676.1 | \|Severe acute respiratory syndrome coro | ···················· | ···················· | ···················· | ···················· | ···················· | ···················· |
| 22735 | MT846677.1 | \|Severe acute respiratory syndrome coro | ···················· | ···················· | ···················· | ···················· | ···················· | ···················· |
| 22736 | MT846679.1 | \|Severe acute respiratory syndrome coro | ···················· | ···················· | ···················· | ···················· | ···················· | ···················· |
| 22737 | MT846681.1 | \|Severe acute respiratory syndrome coro | ···················· | ···················· | ···················· | ···················· | ···················· | ···················· |
| 22738 | MT846682.1 | \|Severe acute respiratory syndrome coro | ···················· | ···A················ | ···················· | ···················· | ···················· | ···················· |
| 22739 | MT846684.1 | \|Severe acute respiratory syndrome coro | ···················· | ···················· | ···················· | ···················· | ···················· | ···················· |
| 22740 | MT846686.1 | \|Severe acute respiratory syndrome coro | ···················· | ···················· | ···················· | ···················· | ···················· | ···················· |
| 22741 | MT846687.1 | \|Severe acute respiratory syndrome coro | ···················· | ·A·················· | ···················· | ···················· | ···················· | ···················· |
| 22742 | MT846689.1 | \|Severe acute respiratory syndrome coro | ···················· | ·A·················· | ···················· | ···················· | ···················· | ···················· |
| 22743 | MT846690.1 | \|Severe acute respiratory syndrome coro | ···················· | ···················· | ···················· | ···················· | ···················· | ···················· |
| 22744 | MT846691.1 | \|Severe acute respiratory syndrome coro | ···················· | ···················· | ···················· | ···················· | ···················· | ···················· |
| 22745 | MT847208.1 | \|Severe acute respiratory syndrome coro | ···················· | ·A·················· | ···················· | ···················· | ···················· | ···················· |
| 22746 | MT847209.1 | \|Severe acute respiratory syndrome coro | ···················· | ···················· | ···················· | ···················· | ···················· | ···················· |
| 22747 | MT847210.1 | \|Severe acute respiratory syndrome coro | ···················· | ···················· | ···················· | ···················· | ···················· | ···················· |
| 22748 | MT847211.1 | \|Severe acute respiratory syndrome coro | ···················· | ···················· | ···················· | ···················· | ···················· | ···················· |
| 22749 | MT847212.1 | \|Severe acute respiratory syndrome coro | ···················· | ···················· | ···················· | ···················· | ···················· | ···················· |
| 22750 | MT847213.1 | \|Severe acute respiratory syndrome coro | ···················· | ···················· | ···················· | ···················· | ···················· | ···················· |
| 22751 | MT847214.1 | \|Severe acute respiratory syndrome coro | ···················· | ···················· | ···················· | ···················· | ···················· | ···················· |
| 22752 | MT847215.1 | \|Severe acute respiratory syndrome coro | ···················· | ···················· | ···················· | ···················· | ···················· | ···················· |
| 22753 | MT847216.1 | \|Severe acute respiratory syndrome coro | ···················· | ···················· | ···················· | ···················· | ···················· | ···················· |
| 22754 | MT847217.1 | \|Severe acute respiratory syndrome coro | ···················· | ···················· | ···················· | ···················· | ···················· | ···················· |
| 22755 | MT847218.1 | \|Severe acute respiratory syndrome coro | ···················· | ···················· | ···················· | ···················· | ···················· | ···················· |
| 22756 | MT847219.1 | \|Severe acute respiratory syndrome coro | ···················· | ···················· | ···················· | ···················· | ···················· | ···················· |
| 22757 | MT847220.1 | \|Severe acute respiratory syndrome coro | ···················· | ···················· | ···················· | ···················· | ···················· | ···················· |
| 22758 | MT847221.1 | \|Severe acute respiratory syndrome coro | ···················· | ···················· | ···················· | ···················· | ···················· | ···················· |
| 22759 | MT847222.1 | \|Severe acute respiratory syndrome coro | ···················· | ·A·················· | ···················· | ···················· | ···················· | ··A················· |
| 22760 | MT847223.1 | \|Severe acute respiratory syndrome coro | ···················· | ···················· | ···················· | ···················· | ···················· | ···················· |
| 22761 | MT847224.1 | \|Severe acute respiratory syndrome coro | ···················· | ···················· | ···················· | ···················· | ···················· | ···················· |
| 22762 | MT847225.1 | \|Severe acute respiratory syndrome coro | ···················· | ···················· | ···················· | ···················· | ···················· | ···················· |
| 22763 | MT847226.1 | \|Severe acute respiratory syndrome coro | ···················· | ···················· | ···················· | ···················· | ···················· | ···················· |
| 22764 | MT847227.1 | \|Severe acute respiratory syndrome coro | ···················· | ···················· | ···················· | ···················· | ···················· | ···················· |
| 22765 | MT847228.1 | \|Severe acute respiratory syndrome coro | ···················· | ···················· | ···················· | ···················· | ···················· | ···················· |
| 22766 | MT835383.1 | \|Severe acute respiratory syndrome coro | ···················· | ···················· | ···················· | ···················· | ···················· | ···················· |
| 22767 | MT843285.1 | \|Severe acute respiratory syndrome coro | ···················· | ···················· | ···················· | ···················· | ···················· | ···················· |
| 22768 | MT843286.1 | \|Severe acute respiratory syndrome coro | ···················· | ···················· | ···················· | ···················· | ···················· | ···················· |
| 22769 | MT843287.1 | \|Severe acute respiratory syndrome coro | ···················· | ···················· | ···················· | ···················· | ···················· | ···················· |
| 22770 | MT843288.1 | \|Severe acute respiratory syndrome coro | ···················· | ···················· | ···················· | ···················· | ···················· | ···················· |
| 22771 | MT843289.1 | \|Severe acute respiratory syndrome coro | ···················· | ···················· | ···················· | ···················· | ···················· | ···················· |
| 22772 | MT843290.1 | \|Severe acute respiratory syndrome coro | ···················· | ···················· | ···················· | ···················· | ···················· | ···················· |
| 22773 | MT843291.1 | \|Severe acute respiratory syndrome coro | ···················· | ···················· | ···················· | ···················· | ···················· | ···················· |
| 22774 | MT843292.1 | \|Severe acute respiratory syndrome coro | ···················· | ···················· | ···················· | ···················· | ···················· | ···················· |
| 22775 | MT843293.1 | \|Severe acute respiratory syndrome coro | ···················· | ···················· | ···················· | ···················· | ···················· | ···················· |
| 22776 | MT843294.1 | \|Severe acute respiratory syndrome coro | ···················· | ···················· | ···················· | ···················· | ···················· | ···················· |
| 22777 | MT843295.1 | \|Severe acute respiratory syndrome coro | ···················· | ···················· | ···················· | ···················· | ···················· | ···················· |
| 22778 | MT843297.1 | \|Severe acute respiratory syndrome coro | ···················· | ···················· | ···················· | ···················· | ···················· | ···················· |
| 22779 | MT843298.1 | \|Severe acute respiratory syndrome coro | ···················· | ···················· | ···················· | ···················· | ···················· | ···················· |
| 22780 | MT843299.1 | \|Severe acute respiratory syndrome coro | ···················· | ···················· | ···················· | ···················· | ···················· | ···················· |
| 22781 | MT843300.1 | \|Severe acute respiratory syndrome coro | ···················· | ···················· | ···················· | ···················· | ···················· | ···················· |
| 22782 | MT843301.1 | \|Severe acute respiratory syndrome coro | ···················· | ···················· | ···················· | ···················· | ···················· | ···················· |
| 22783 | MT843303.1 | \|Severe acute respiratory syndrome coro | ···················· | ···················· | ···················· | ···················· | ···················· | ···················· |
| 22784 | MT843304.1 | \|Severe acute respiratory syndrome coro | ···················· | ···················· | ···················· | ···················· | ···················· | ···················· |
| 22785 | MT843305.1 | \|Severe acute respiratory syndrome coro | ···················· | ···················· | ···················· | ···················· | ···················· | ···················· |
| 22786 | MT843306.1 | \|Severe acute respiratory syndrome coro | ···················· | ···················· | ···················· | ···················· | ···················· | ···················· |
| 22787 | MT843307.1 | \|Severe acute respiratory syndrome coro | ···················· | ···················· | ···················· | ···················· | ···················· | ···················· |
| 22788 | MT843309.1 | \|Severe acute respiratory syndrome coro | ···················· | ···················· | ···················· | ···················· | ···················· | ···················· |
| 22789 | MT843310.1 | \|Severe acute respiratory syndrome coro | ···················· | ···················· | ···················· | ···················· | ···················· | ···················· |
| 22790 | MT843311.1 | \|Severe acute respiratory syndrome coro | ···················· | ···················· | ···················· | ···················· | ···················· | ···················· |
| 22791 | MT843312.1 | \|Severe acute respiratory syndrome coro | ···················· | ···················· | ···················· | ···················· | ···················· | ···················· |
| 22792 | MT843313.1 | \|Severe acute respiratory syndrome coro | ···················· | ···················· | ···················· | ···················· | ···················· | ···················· |
| 22793 | MT843314.1 | \|Severe acute respiratory syndrome coro | ···················· | ···················· | ···················· | ···················· | ···················· | ···················· |
| 22794 | MT843315.1 | \|Severe acute respiratory syndrome coro | ···················· | ···················· | ···················· | ···················· | ···················· | ···················· |
| 22795 | MT843316.1 | \|Severe acute respiratory syndrome coro | ·····G·············· | ···················· | ···················· | ···················· | ···················· | ···················· |
| 22796 | MT843317.1 | \|Severe acute respiratory syndrome coro | ···················· | ···················· | ···················· | ···················· | ···················· | ···················· |
| 22797 | MT843318.1 | \|Severe acute respiratory syndrome coro | ···················· | ···················· | ···················· | ···················· | ···················· | ···················· |
| 22798 | MT843319.1 | \|Severe acute respiratory syndrome coro | ···················· | ···················· | ···················· | ···················· | ···················· | ···················· |
| 22799 | MT843320.1 | \|Severe acute respiratory syndrome coro | ···················· | ···················· | ···················· | ···················· | ···················· | ···················· |

22800 MT843737.1 |Severe acute respiratory syndrome coro .................... .A.................. .................... .................... .................... ....................
22801 MT843738.1 |Severe acute respiratory syndrome coro .................... .................... .................... .................... .................... ....................
22802 MT843739.1 |Severe acute respiratory syndrome coro .................... .................... .................... .................... .................... ....................
22803 MT843740.1 |Severe acute respiratory syndrome coro .................... .................... .................... .................... .................... ....................
22804 MT843741.1 |Severe acute respiratory syndrome coro .................... .................... .................... .................... .................... ....................
22805 MT843742.1 |Severe acute respiratory syndrome coro .................... .................... .................... .................... .................... ....................
22806 MT843743.1 |Severe acute respiratory syndrome coro .................... .................... .................... .................... .................... ....................
22807 MT843744.1 |Severe acute respiratory syndrome coro .................... .................... .................... .................... .................... ....................
22808 MT843745.1 |Severe acute respiratory syndrome coro .................... .................... .................... .................... .................... ....................
22809 MT843746.1 |Severe acute respiratory syndrome coro .................... .................... .................... .................... .................... ....................
22810 MT843747.1 |Severe acute respiratory syndrome coro .................... .................... .................... .................... .................... ....................
22811 MT843748.1 |Severe acute respiratory syndrome coro .................... .................... .................... .................... .................... ....................
22812 MT843749.1 |Severe acute respiratory syndrome coro .................... .................... .................... .................... .................... ....................
22813 MT843750.1 |Severe acute respiratory syndrome coro .................... .................... .................... .................... .................... ....................
22814 MT843751.1 |Severe acute respiratory syndrome coro .................... .................... .................... .................... .................... ....................
22815 MT843752.1 |Severe acute respiratory syndrome coro .................... .................... .................... .................... .................... ....................
22816 MT843753.1 |Severe acute respiratory syndrome coro .................... .................... .................... .................... .................... ....................
22817 MT843754.1 |Severe acute respiratory syndrome coro .................... .................... .................... .................... .................... ....................
22818 MT843755.1 |Severe acute respiratory syndrome coro .................... .................... .................... .................... .................... ....................
22819 MT843756.1 |Severe acute respiratory syndrome coro .................... .................... .................... .................... .................... ....................
22820 MT843757.1 |Severe acute respiratory syndrome coro .................... .................... .................... .................... .................... ....................
22821 MT843758.1 |Severe acute respiratory syndrome coro .................... .................... .................... .................... .................... ....................
22822 MT843759.1 |Severe acute respiratory syndrome coro .................... .................... .................... .................... .................... ....................
22823 MT844021.1 |Severe acute respiratory syndrome coro .................... .................... .................... .................... .................... ....................
22824 MT844022.1 |Severe acute respiratory syndrome coro .................... .................... .................... .................... .................... ....................
22825 MT844023.1 |Severe acute respiratory syndrome coro .................... .................... .................... .................... .................... ....................
22826 MT844024.1 |Severe acute respiratory syndrome coro .................... .................... .................... .................... .................... ....................
22827 MT844025.1 |Severe acute respiratory syndrome coro .................... .................... .................... .................... .................... ....................
22828 MT844026.1 |Severe acute respiratory syndrome coro .................... .................... .................... .................... .................... ....................
22829 MT844027.1 |Severe acute respiratory syndrome coro .................... .................... .................... .................... .................... ....................
22830 MT844028.1 |Severe acute respiratory syndrome coro .................... .................... .................... .................... .................... ....................
22831 MT844029.1 |Severe acute respiratory syndrome coro .................... .................... .................... .................... .................... ....................
22832 MT844030.1 |Severe acute respiratory syndrome coro .................... .................... .................... .................... .................... ....................
22833 MT844031.1 |Severe acute respiratory syndrome coro .................... .................... .................... .................... .................... ....................
22834 MT844032.1 |Severe acute respiratory syndrome coro .................... .................... .................... .................... .................... ....................
22835 MT844033.1 |Severe acute respiratory syndrome coro .................... .................... .................... .................... .................... ....................
22836 MT844034.1 |Severe acute respiratory syndrome coro .................... .................... .................... .................... .................... ....................
22837 MT844035.1 |Severe acute respiratory syndrome coro .................... .................... .................... .................... .................... ....................
22838 MT844036.1 |Severe acute respiratory syndrome coro .................... .................... .................... .................... .................... ....................
22839 MT844037.1 |Severe acute respiratory syndrome coro .................... .................... .................... .................... .................... ....................
22840 MT844038.1 |Severe acute respiratory syndrome coro .................... .................... .................... .................... .................... ....................
22841 MT844039.1 |Severe acute respiratory syndrome coro .................... .................... .................... .................... .................... ....................
22842 MT844040.1 |Severe acute respiratory syndrome coro .................... .................... .................... .................... .................... ....................
22843 MT844041.1 |Severe acute respiratory syndrome coro .................... .................... .................... .................... .................... ....................
22844 MT844042.1 |Severe acute respiratory syndrome coro .................... .................... .................... .................... .................... ....................
22845 MT844043.1 |Severe acute respiratory syndrome coro .................... .................... .................... .................... .................... ....................
22846 MT844044.1 |Severe acute respiratory syndrome coro .................... .................... .................... .................... .................... ....................
22847 MT844045.1 |Severe acute respiratory syndrome coro .................... .................... .................... .................... .................... ....................
22848 MT844046.1 |Severe acute respiratory syndrome coro .................... .................... .................... .................... .................... ....................
22849 MT844047.1 |Severe acute respiratory syndrome coro .................... .................... .................... .................... .................... ....................
22850 MT844048.1 |Severe acute respiratory syndrome coro .................... .................... .................... .................... .................... ....................
22851 MT844049.1 |Severe acute respiratory syndrome coro .................... .................... .................... .................... .................... ....................
22852 LC573289.1 |Severe acute respiratory syndrome coro .................... .................... .................... .................... .................... ....................
22853 MT831122.1 |Severe acute respiratory syndrome coro .................... .................... .................... .................... .................... ....................
22854 MT831123.1 |Severe acute respiratory syndrome coro .................... .................... .................... .................... .................... ....................
22855 MT831124.1 |Severe acute respiratory syndrome coro .................... .................... .................... .................... .................... ....................
22856 MT831125.1 |Severe acute respiratory syndrome coro .................... .................... .................... .................... .................... ....................
22857 MT831127.1 |Severe acute respiratory syndrome coro .................... .................... .................... .................... .................... ....................
22858 MT831128.1 |Severe acute respiratory syndrome coro .................... .................... .................... .................... .................... ....................
22859 MT831129.1 |Severe acute respiratory syndrome coro .................... .................... .................... .................... .................... ....................
22860 MT831130.1 |Severe acute respiratory syndrome coro .................... .................... .................... .................... .................... ....................
22861 MT831131.1 |Severe acute respiratory syndrome coro .................... .................... .................... .................... .................... ....................
22862 MT831133.1 |Severe acute respiratory syndrome coro .................... .................... .................... .................... .................... ....................
22863 MT831134.1 |Severe acute respiratory syndrome coro .................... .................... .................... .................... .................... ....................
22864 MT831135.1 |Severe acute respiratory syndrome coro .................... .................... .................... .................... .................... ....................
22865 MT831137.1 |Severe acute respiratory syndrome coro .................... .................... .................... .................... .................... ....................
22866 MT831138.1 |Severe acute respiratory syndrome coro .................... .................... .................... .................... .................... ....................
22867 MT831139.1 |Severe acute respiratory syndrome coro .................... .................... .................... .................... .................... ....................
22868 MT831140.1 |Severe acute respiratory syndrome coro .................... .................... .................... .................... .................... ....................
22869 MT831141.1 |Severe acute respiratory syndrome coro .................... .................... .................... .................... .................... ....................
22870 MT831142.1 |Severe acute respiratory syndrome coro .................... .................... .................... .................... .................... ....................
22871 MT831143.1 |Severe acute respiratory syndrome coro .................... .................... .................... .................... .................... ....................
22872 MT831144.1 |Severe acute respiratory syndrome coro .................... .................... .................... .................... .................... ....................
22873 MT831145.1 |Severe acute respiratory syndrome coro .................... .................... .................... .................... .................... ....................
22874 MT831147.1 |Severe acute respiratory syndrome coro .................... .................... .................... .................... .................... ....................
22875 MT831148.1 |Severe acute respiratory syndrome coro .................... .................... .................... .................... .................... ....................
22876 MT831149.1 |Severe acute respiratory syndrome coro .................... .................... .................... .................... .................... ....................
22877 MT831150.1 |Severe acute respiratory syndrome coro .................... .................... .................... .................... .................... ....................

| | | | | | | | |
|---|---|---|---|---|---|---|---|
| 22878 | MT831151.1 \|Severe acute respiratory syndrome coro | .................... | .................... | .................... | .................... | .................... | .................... |
| 22879 | MT831152.1 \|Severe acute respiratory syndrome coro | .................... | .................... | .................... | .................... | .................... | .................... |
| 22880 | MT831153.1 \|Severe acute respiratory syndrome coro | .................... | .................... | .................... | .................... | .................... | .................... |
| 22881 | MT831154.1 \|Severe acute respiratory syndrome coro | .................... | .................... | .................... | .................... | .................... | .................... |
| 22882 | MT831156.1 \|Severe acute respiratory syndrome coro | .................... | .................... | .................... | .................... | .................... | .................... |
| 22883 | MT831157.1 \|Severe acute respiratory syndrome coro | .................... | .................... | .................... | .................... | .................... | .................... |
| 22884 | MT831158.1 \|Severe acute respiratory syndrome coro | .................... | .................... | .................... | .................... | .................... | .................... |
| 22885 | MT831159.1 \|Severe acute respiratory syndrome coro | .................... | .................... | .................... | .................... | .................... | .................... |
| 22886 | MT831160.1 \|Severe acute respiratory syndrome coro | .................... | .................... | .................... | .................... | .................... | .................... |
| 22887 | MT831161.1 \|Severe acute respiratory syndrome coro | .................... | .................... | .................... | .................... | .................... | .................... |
| 22888 | MT831162.1 \|Severe acute respiratory syndrome coro | .................... | .................... | .................... | .................... | .................... | .................... |
| 22889 | MT831163.1 \|Severe acute respiratory syndrome coro | .................... | .................... | .................... | .................... | .................... | .................... |
| 22890 | MT831164.1 \|Severe acute respiratory syndrome coro | .................... | .................... | .................... | .................... | .................... | .................... |
| 22891 | MT831165.1 \|Severe acute respiratory syndrome coro | .................... | .................... | .................... | .................... | .................... | .................... |
| 22892 | MT831166.1 \|Severe acute respiratory syndrome coro | .................... | .................... | .................... | .................... | .................... | .................... |
| 22893 | MT831167.1 \|Severe acute respiratory syndrome coro | .................... | .................... | .................... | .................... | .................... | .................... |
| 22894 | MT831168.1 \|Severe acute respiratory syndrome coro | .................... | .................... | .................... | .................... | .................... | .................... |
| 22895 | MT831169.1 \|Severe acute respiratory syndrome coro | .................... | .................... | .................... | .................... | .................... | .................... |
| 22896 | MT831170.1 \|Severe acute respiratory syndrome coro | .................... | .................... | .................... | .................... | .................... | .................... |
| 22897 | MT831171.1 \|Severe acute respiratory syndrome coro | .................... | .................... | .................... | .................... | .................... | .................... |
| 22898 | MT831172.1 \|Severe acute respiratory syndrome coro | .................... | .................... | .................... | .................... | .................... | .................... |
| 22899 | MT831173.1 \|Severe acute respiratory syndrome coro | .................... | .................... | .................... | .................... | .................... | .................... |
| 22900 | MT831174.1 \|Severe acute respiratory syndrome coro | .................... | .................... | .................... | .................... | .................... | .................... |
| 22901 | MT831175.1 \|Severe acute respiratory syndrome coro | .................... | .................... | .................... | .................... | .................... | .................... |
| 22902 | MT831176.1 \|Severe acute respiratory syndrome coro | .................... | .................... | .................... | .................... | .................... | .................... |
| 22903 | MT831177.1 \|Severe acute respiratory syndrome coro | .................... | .................... | .................... | .................... | .................... | .................... |
| 22904 | MT831178.1 \|Severe acute respiratory syndrome coro | .................... | .................... | .................... | .................... | .................... | .................... |
| 22905 | MT831179.1 \|Severe acute respiratory syndrome coro | .................... | .................... | .................... | .................... | .................... | .................... |
| 22906 | MT831180.1 \|Severe acute respiratory syndrome coro | .................... | .................... | .................... | .................... | .................... | .................... |
| 22907 | MT831181.1 \|Severe acute respiratory syndrome coro | .................... | .................... | .................... | .................... | .................... | .................... |
| 22908 | MT831182.1 \|Severe acute respiratory syndrome coro | .................... | .................... | .................... | .................... | .................... | .................... |
| 22909 | MT831183.1 \|Severe acute respiratory syndrome coro | .................... | .................... | .................... | .................... | .................... | .................... |
| 22910 | MT831184.1 \|Severe acute respiratory syndrome coro | .................... | .................... | .................... | .................... | .................... | .................... |
| 22911 | MT831186.1 \|Severe acute respiratory syndrome coro | .................... | .................... | .................... | .................... | .................... | .................... |
| 22912 | MT831187.1 \|Severe acute respiratory syndrome coro | .................... | .................... | .................... | .................... | .................... | .................... |
| 22913 | MT831188.1 \|Severe acute respiratory syndrome coro | .................... | .................... | .................... | .................... | .................... | .................... |
| 22914 | MT831189.1 \|Severe acute respiratory syndrome coro | .................... | .................... | .................... | .................... | .................... | .................... |
| 22915 | MT831190.1 \|Severe acute respiratory syndrome coro | .................... | .................... | .................... | .................... | .................... | .................... |
| 22916 | MT831191.1 \|Severe acute respiratory syndrome coro | .................... | .................... | .................... | .................... | .................... | .................... |
| 22917 | MT831192.1 \|Severe acute respiratory syndrome coro | .................... | .................... | .................... | .................... | .................... | .................... |
| 22918 | MT831193.1 \|Severe acute respiratory syndrome coro | .................... | .................... | .................... | .................... | .................... | .................... |
| 22919 | MT831194.1 \|Severe acute respiratory syndrome coro | .................... | .................... | .................... | .................... | .................... | .................... |
| 22920 | MT831195.1 \|Severe acute respiratory syndrome coro | .................... | .................... | .................... | .................... | .................... | .................... |
| 22921 | MT831196.1 \|Severe acute respiratory syndrome coro | .................... | .................... | .................... | .................... | .................... | .................... |
| 22922 | MT831197.1 \|Severe acute respiratory syndrome coro | .................... | .................... | .................... | .................... | .................... | .................... |
| 22923 | MT831198.1 \|Severe acute respiratory syndrome coro | .................... | .................... | .................... | .................... | .................... | .................... |
| 22924 | MT831199.1 \|Severe acute respiratory syndrome coro | .................... | .................... | .................... | .................... | .................... | .................... |
| 22925 | MT831200.1 \|Severe acute respiratory syndrome coro | .................... | .................... | .................... | .................... | .................... | .................... |
| 22926 | MT831201.1 \|Severe acute respiratory syndrome coro | .................... | .................... | .................... | .................... | .................... | .................... |
| 22927 | MT831202.1 \|Severe acute respiratory syndrome coro | .................... | .................... | .................... | .................... | .................... | .................... |
| 22928 | MT831204.1 \|Severe acute respiratory syndrome coro | .................... | .................... | .................... | .................... | .................... | .................... |
| 22929 | MT831205.1 \|Severe acute respiratory syndrome coro | .................... | .................... | .................... | .................... | .................... | .................... |
| 22930 | MT831206.1 \|Severe acute respiratory syndrome coro | .................... | .................... | .................... | .................... | .................... | .................... |
| 22931 | MT831207.1 \|Severe acute respiratory syndrome coro | .................... | .................... | .................... | .................... | .................... | .................... |
| 22932 | MT831208.1 \|Severe acute respiratory syndrome coro | .................... | .................... | .................... | .................... | .................... | .................... |
| 22933 | MT831209.1 \|Severe acute respiratory syndrome coro | .................... | .................... | .................... | .................... | .................... | .................... |
| 22934 | MT831210.1 \|Severe acute respiratory syndrome coro | .................... | .................... | .................... | .................... | .................... | .................... |
| 22935 | MT831211.1 \|Severe acute respiratory syndrome coro | .................... | .................... | .................... | .................... | .................... | .................... |
| 22936 | MT831212.1 \|Severe acute respiratory syndrome coro | .................... | .................... | .................... | .................... | .................... | .................... |
| 22937 | MT831213.1 \|Severe acute respiratory syndrome coro | .................... | .................... | .................... | .................... | .................... | .................... |
| 22938 | MT831214.1 \|Severe acute respiratory syndrome coro | .................... | .................... | .................... | .................... | .................... | .................... |
| 22939 | MT831215.1 \|Severe acute respiratory syndrome coro | .................... | .................... | .................... | .................... | .................... | .................... |
| 22940 | MT831216.1 \|Severe acute respiratory syndrome coro | .................... | .................... | .................... | .................... | .................... | .................... |
| 22941 | MT831217.1 \|Severe acute respiratory syndrome coro | .................... | .................... | .................... | .................... | .................... | .................... |
| 22942 | MT831218.1 \|Severe acute respiratory syndrome coro | .................... | .................... | .................... | .................... | .................... | .................... |
| 22943 | MT831219.1 \|Severe acute respiratory syndrome coro | .................... | .................... | .................... | .................... | .................... | .................... |
| 22944 | MT831220.1 \|Severe acute respiratory syndrome coro | .................... | .................... | .................... | .................... | .................... | .................... |
| 22945 | MT831221.1 \|Severe acute respiratory syndrome coro | .................... | .................... | .................... | .................... | .................... | .................... |
| 22946 | MT831222.1 \|Severe acute respiratory syndrome coro | .................... | .................... | .................... | .................... | .................... | .................... |
| 22947 | MT831223.1 \|Severe acute respiratory syndrome coro | .................... | .................... | .................... | .................... | .................... | .................... |
| 22948 | MT831224.1 \|Severe acute respiratory syndrome coro | .................... | .................... | .................... | .................... | .................... | .................... |
| 22949 | MT831225.1 \|Severe acute respiratory syndrome coro | .................... | .................... | .................... | .................... | .................... | .................... |
| 22950 | MT831226.1 \|Severe acute respiratory syndrome coro | .................... | .................... | .................... | .................... | .................... | .................... |
| 22951 | MT831227.1 \|Severe acute respiratory syndrome coro | .................... | .................... | .................... | .................... | .................... | .................... |
| 22952 | MT831229.1 \|Severe acute respiratory syndrome coro | .................... | .................... | .................... | .................... | .................... | .................... |
| 22953 | MT831230.1 \|Severe acute respiratory syndrome coro | .................... | .................... | .................... | .................... | .................... | .................... |
| 22954 | MT831231.1 \|Severe acute respiratory syndrome coro | .................... | .................... | .................... | .................... | .................... | .................... |
| 22955 | MT831232.1 \|Severe acute respiratory syndrome coro | .................... | .................... | .................... | .................... | .................... | .................... |

```
22956 MT831233.1 |Severe acute respiratory syndrome coro    ....................  ....................  .....................  ....................  ....................  ....................
22957 MT831234.1 |Severe acute respiratory syndrome coro    ....................  ....................  .....................  ....................  ....................  ....................
22958 MT831235.1 |Severe acute respiratory syndrome coro    ....................  ....................  .....................  ....................  ....................  ....................
22959 MT831236.1 |Severe acute respiratory syndrome coro    ....................  ....................  .....................  ....................  ....................  ....................
22960 MT831237.1 |Severe acute respiratory syndrome coro    ....................  ....................  .....................  ....................  ....................  ....................
22961 MT831238.1 |Severe acute respiratory syndrome coro    ....................  ....................  .....................  ....................  ....................  ....................
22962 MT831239.1 |Severe acute respiratory syndrome coro    ....................  ....................  .....................  ....................  ....................  ....................
22963 MT831240.1 |Severe acute respiratory syndrome coro    ....................  ....................  .....................  ....................  ....................  ....................
22964 MT831241.1 |Severe acute respiratory syndrome coro    ....................  ....................  .....................  ....................  ....................  ....................
22965 MT831243.1 |Severe acute respiratory syndrome coro    ....................  ....................  .....................  ....................  ....................  ....................
22966 MT831244.1 |Severe acute respiratory syndrome coro    ....................  ....................  .....................  ....................  ....................  ....................
22967 MT831245.1 |Severe acute respiratory syndrome coro    ....................  ....................  .....................  ....................  ....................  ....................
22968 MT831246.1 |Severe acute respiratory syndrome coro    ....................  ....................  .....................  ....................  ....................  ....................
22969 MT831248.1 |Severe acute respiratory syndrome coro    ....................  ....................  .....................  ....................  ....................  ....................
22970 MT831249.1 |Severe acute respiratory syndrome coro    ....................  ....................  .....................  ....................  ....................  ....................
22971 MT831250.1 |Severe acute respiratory syndrome coro    ....................  ....................  .....................  ....................  ....................  ....................
22972 MT831251.1 |Severe acute respiratory syndrome coro    ....................  ....................  .....................  ....................  ....................  ....................
22973 MT831252.1 |Severe acute respiratory syndrome coro    ....................  ....................  .....................  ....................  ....................  ....................
22974 MT831253.1 |Severe acute respiratory syndrome coro    ....................  ....................  .....................  ....................  ....................  ....................
22975 MT831254.1 |Severe acute respiratory syndrome coro    ....................  ....................  .....................  ....................  ....................  ....................
22976 MT831255.1 |Severe acute respiratory syndrome coro    ....................  ....................  .....................  ....................  ....................  ....................
22977 MT831256.1 |Severe acute respiratory syndrome coro    ....................  ....................  .....................  ....................  ....................  ....................
22978 MT831257.1 |Severe acute respiratory syndrome coro    ....................  ....................  .....................  ....................  ....................  ....................
22979 MT831258.1 |Severe acute respiratory syndrome coro    ....................  ....................  .....................  ....................  ....................  ....................
22980 MT831259.1 |Severe acute respiratory syndrome coro    ....................  ....................  .....................  ....................  ....................  ....................
22981 MT831260.1 |Severe acute respiratory syndrome coro    ....................  ....................  .....................  ....................  ....................  ....................
22982 MT831261.1 |Severe acute respiratory syndrome coro    ....................  ....................  .....................  ....................  ....................  ....................
22983 MT831262.1 |Severe acute respiratory syndrome coro    ....................  ....................  .....................  ....................  ....................  ....................
22984 MT831263.1 |Severe acute respiratory syndrome coro    ....................  ....................  .....................  ....................  ....................  ....................
22985 MT831264.1 |Severe acute respiratory syndrome coro    ....................  ....................  .....................  ....................  ....................  ....................
22986 MT831265.1 |Severe acute respiratory syndrome coro    ....................  ....................  .....................  ....................  ....................  ....................
22987 MT831266.1 |Severe acute respiratory syndrome coro    ....................  ....................  .....................  ....................  ....................  ....................
22988 MT831267.1 |Severe acute respiratory syndrome coro    ....................  ....................  .....................  ....................  ....................  ....................
22989 MT831268.1 |Severe acute respiratory syndrome coro    ....................  ....................  .....................  ....................  ....................  ....................
22990 MT831269.1 |Severe acute respiratory syndrome coro    ....................  ....................  .....................  ....................  ....................  ....................
22991 MT831270.1 |Severe acute respiratory syndrome coro    ....................  ....................  .....................  ....................  ....................  ....................
22992 MT831271.1 |Severe acute respiratory syndrome coro    ....................  ....................  .....................  ....................  ....................  ....................
22993 MT831272.1 |Severe acute respiratory syndrome coro    ....................  ....................  .....................  ....................  ....................  ....................
22994 MT831273.1 |Severe acute respiratory syndrome coro    ....................  ....................  .....................  ....................  ....................  ....................
22995 MT831274.1 |Severe acute respiratory syndrome coro    ....................  ....................  .....................  ....................  ....................  ....................
22996 MT831275.1 |Severe acute respiratory syndrome coro    ....................  ....................  .....................  ....................  ....................  ....................
22997 MT831276.1 |Severe acute respiratory syndrome coro    ....................  ....................  .....................  ....................  ....................  ....................
22998 MT831277.1 |Severe acute respiratory syndrome coro    ....................  ....................  .....................  ....................  ....................  ....................
22999 MT831278.1 |Severe acute respiratory syndrome coro    ....................  ....................  .....................  ....................  ....................  ....................
23000 MT831279.1 |Severe acute respiratory syndrome coro    ....................  ....................  .....................  ....................  ....................  ....................
23001 MT831280.1 |Severe acute respiratory syndrome coro    ....................  ....................  .....................  ....................  ....................  ....................
23002 MT831281.1 |Severe acute respiratory syndrome coro    ....................  ....................  .....................  ....................  ....................  ....................
23003 MT831282.1 |Severe acute respiratory syndrome coro    ....................  ....................  .....................  ....................  ....................  ....................
23004 MT831283.1 |Severe acute respiratory syndrome coro    ....................  ....................  .....................  ....................  ....................  ....................
23005 MT831284.1 |Severe acute respiratory syndrome coro    ....................  ....................  .....................  ....................  ....................  ....................
23006 MT831285.1 |Severe acute respiratory syndrome coro    ....................  ....................  .....................  ....................  ....................  ....................
23007 MT831286.1 |Severe acute respiratory syndrome coro    ....................  ....................  .....................  ....................  ....................  ....................
23008 MT831287.1 |Severe acute respiratory syndrome coro    ....................  ....................  .....................  ....................  ....................  ....................
23009 MT831288.1 |Severe acute respiratory syndrome coro    ....................  ....................  .....................  ....................  ....................  ....................
23010 MT831289.1 |Severe acute respiratory syndrome coro    ....................  ....................  .....................  ....................  ....................  ....................
23011 MT831290.1 |Severe acute respiratory syndrome coro    ....................  ....................  .....................  ....................  ....................  ....................
23012 MT831291.1 |Severe acute respiratory syndrome coro    ....................  ....................  .....................  ....................  ....................  ....................
23013 MT831292.1 |Severe acute respiratory syndrome coro    ....................  ....................  .....................  ....................  ....................  ....................
23014 MT831293.1 |Severe acute respiratory syndrome coro    ....................  ....................  .....................  ....................  ....................  ....................
23015 MT831294.1 |Severe acute respiratory syndrome coro    ....................  ....................  .....................  ....................  ....................  ....................
23016 MT831295.1 |Severe acute respiratory syndrome coro    ....................  ....................  .....................  ....................  ....................  ....................
23017 MT831296.1 |Severe acute respiratory syndrome coro    ....................  ....................  .....................  ....................  ....................  ....................
23018 MT831297.1 |Severe acute respiratory syndrome coro    ....................  ....................  .....................  ....................  ....................  ....................
23019 MT831298.1 |Severe acute respiratory syndrome coro    ....................  ....................  .....................  ....................  ....................  ....................
23020 MT831299.1 |Severe acute respiratory syndrome coro    ....................  ....................  .....................  ....................  ....................  ....................
23021 MT831300.1 |Severe acute respiratory syndrome coro    ....................  ....................  .....................  ....................  ....................  ....................
23022 MT831301.1 |Severe acute respiratory syndrome coro    ....................  ....................  .....................  ....................  ....................  ....................
23023 MT831302.1 |Severe acute respiratory syndrome coro    ....................  ....................  .....................  ....................  ....................  ....................
23024 MT831303.1 |Severe acute respiratory syndrome coro    ....................  ....................  .....................  ....................  ....................  ....................
23025 MT831304.1 |Severe acute respiratory syndrome coro    ....................  ....................  .....................  ....................  ....................  ....................
23026 MT831305.1 |Severe acute respiratory syndrome coro    ....................  ....................  .....................  ....................  ....................  ....................
23027 MT831306.1 |Severe acute respiratory syndrome coro    ....................  ....................  .....................  ....................  ....................  ....................
23028 MT831307.1 |Severe acute respiratory syndrome coro    ....................  ....................  .....................  ....................  ....................  ....................
23029 MT831308.1 |Severe acute respiratory syndrome coro    ....................  ....................  .....................  ....................  ....................  ....................
23030 MT831309.1 |Severe acute respiratory syndrome coro    ....................  ....................  .....................  ....................  ....................  ....................
23031 MT831310.1 |Severe acute respiratory syndrome coro    ....................  ....................  .....................  ....................  ....................  ....................
23032 MT831311.1 |Severe acute respiratory syndrome coro    ....................  ....................  .....................  ....................  ....................  ....................
23033 MT831312.1 |Severe acute respiratory syndrome coro    ....................  ....................  .....................  ....................  ....................  ....................
```

23034 MT831313.1 |Severe acute respiratory syndrome coro .................... .................... ..................... .................... .................... ....................
23035 MT831314.1 |Severe acute respiratory syndrome coro .................... .................... ..................... .................... .................... ....................
23036 MT831315.1 |Severe acute respiratory syndrome coro .................... .................... ..................... .................... .................... ....................
23037 MT831316.1 |Severe acute respiratory syndrome coro .................... .................... ..................... .................... .................... ....................
23038 MT831318.1 |Severe acute respiratory syndrome coro .................... .................... ..................... .................... .................... ....................
23039 MT831319.1 |Severe acute respiratory syndrome coro .................... .................... ..................... .................... .................... ....................
23040 MT831320.1 |Severe acute respiratory syndrome coro .................... .................... ..................... .................... .................... ....................
23041 MT831321.1 |Severe acute respiratory syndrome coro .................... .................... ..................... .................... .................... ....................
23042 MT831322.1 |Severe acute respiratory syndrome coro .................... .................... ..................... .................... .................... ....................
23043 MT831323.1 |Severe acute respiratory syndrome coro .................... .................... ..................... .................... .................... ....................
23044 MT831324.1 |Severe acute respiratory syndrome coro .................... .................... ..................... .................... .................... ....................
23045 MT831325.1 |Severe acute respiratory syndrome coro .................... .................... ..................... .................... .................... ....................
23046 MT831326.1 |Severe acute respiratory syndrome coro .................... .................... ..................... .................... .................... ....................
23047 MT831327.1 |Severe acute respiratory syndrome coro .................... .................... ..................... .................... .................... ....................
23048 MT831328.1 |Severe acute respiratory syndrome coro .................... .................... ..................... .................... .................... ....................
23049 MT831329.1 |Severe acute respiratory syndrome coro .................... .................... ..................... .................... .................... ....................
23050 MT831330.1 |Severe acute respiratory syndrome coro .................... .................... ..................... .................... .................... ....................
23051 MT831331.1 |Severe acute respiratory syndrome coro .................... .................... ..................... .................... .................... ....................
23052 MT831332.1 |Severe acute respiratory syndrome coro .................... .................... ..................... .................... .................... ....................
23053 MT831333.1 |Severe acute respiratory syndrome coro .................... .................... ..................... .................... .................... ....................
23054 MT831334.1 |Severe acute respiratory syndrome coro .................... .................... ..................... .................... .................... ....................
23055 MT831335.1 |Severe acute respiratory syndrome coro .................... .................... ..................... .................... .................... ....................
23056 MT831336.1 |Severe acute respiratory syndrome coro .................... .................... ..................... .................... .................... ....................
23057 MT831337.1 |Severe acute respiratory syndrome coro .................... .................... ..................... .................... .................... ....................
23058 MT831338.1 |Severe acute respiratory syndrome coro .................... .................... ..................... .................... .................... ....................
23059 MT831339.1 |Severe acute respiratory syndrome coro .................... .................... ..................... .................... .................... ....................
23060 MT831340.1 |Severe acute respiratory syndrome coro .................... .................... ..................... .................... .................... ....................
23061 MT831341.1 |Severe acute respiratory syndrome coro .................... .................... ..................... .................... .................... ....................
23062 MT831342.1 |Severe acute respiratory syndrome coro .................... .................... ..................... .................... .................... ....................
23063 MT831343.1 |Severe acute respiratory syndrome coro .................... .................... ..................... .................... .................... ....................
23064 MT831344.1 |Severe acute respiratory syndrome coro .................... .................... ..................... .................... .................... ....................
23065 MT831345.1 |Severe acute respiratory syndrome coro .................... .................... ..................... .................... .................... ....................
23066 MT831346.1 |Severe acute respiratory syndrome coro .................... .................... ..................... .................... .................... ....................
23067 MT831347.1 |Severe acute respiratory syndrome coro .................... .A.................. ..................... .................... .................... ....................
23068 MT831348.1 |Severe acute respiratory syndrome coro .................... .................... ..................... .................... .................... ....................
23069 MT831349.1 |Severe acute respiratory syndrome coro .................... .................... ..................... .................... .................... ....................
23070 MT831350.1 |Severe acute respiratory syndrome coro .................... .................... ..................... .................... .................... ....................
23071 MT831351.1 |Severe acute respiratory syndrome coro .................... .................... ..................... .................... .................... ....................
23072 MT831352.1 |Severe acute respiratory syndrome coro .................... .................... ..................... .................... .................... ....................
23073 MT831353.1 |Severe acute respiratory syndrome coro .................... .................... ..................... .................... .................... ....................
23074 MT831354.1 |Severe acute respiratory syndrome coro .................... .................... ..................... .................... .................... ....................
23075 MT831355.1 |Severe acute respiratory syndrome coro .................... .................... ..................... .................... .................... ....................
23076 MT831356.1 |Severe acute respiratory syndrome coro .................... .................... ..................... .................... .................... ....................
23077 MT831357.1 |Severe acute respiratory syndrome coro .................... .................... ..................... .................... .................... ....................
23078 MT831358.1 |Severe acute respiratory syndrome coro .................... .................... ..................... .................... .................... ....................
23079 MT831359.1 |Severe acute respiratory syndrome coro .................... .................... ..................... .................... .................... ....................
23080 MT831360.1 |Severe acute respiratory syndrome coro .................... .................... ..................... .................... .................... ....................
23081 MT831361.1 |Severe acute respiratory syndrome coro .................... .................... ..................... .................... .................... ....................
23082 MT831362.1 |Severe acute respiratory syndrome coro .................... .................... ..................... .................... .................... ....................
23083 MT831363.1 |Severe acute respiratory syndrome coro .................... .................... ..................... .................... .................... ....................
23084 MT831364.1 |Severe acute respiratory syndrome coro .................... .................... ..................... .................... .................... ....................
23085 MT831365.1 |Severe acute respiratory syndrome coro .................... .................... ..................... .................... .................... ....................
23086 MT831366.1 |Severe acute respiratory syndrome coro .................... .................... ..................... .................... .................... ....................
23087 MT831367.1 |Severe acute respiratory syndrome coro .................... .................... ..................... .................... .................... ....................
23088 MT831368.1 |Severe acute respiratory syndrome coro .................... .................... ..................... .................... .................... ....................
23089 MT831369.1 |Severe acute respiratory syndrome coro .................... .................... ..................... .................... .................... ....................
23090 MT831370.1 |Severe acute respiratory syndrome coro .................... .................... ..................... .................... .................... ....................
23091 MT831371.1 |Severe acute respiratory syndrome coro .................... .................... ..................... .................... .................... ....................
23092 MT831372.1 |Severe acute respiratory syndrome coro .................... .................... ..................... .................... .................... ....................
23093 MT831373.1 |Severe acute respiratory syndrome coro .................... .................... ..................... .................... .................... ....................
23094 MT831374.1 |Severe acute respiratory syndrome coro .................... .................... ..................... .................... .................... ....................
23095 MT831375.1 |Severe acute respiratory syndrome coro .................... .................... ..................... .................... .................... ....................
23096 MT831376.1 |Severe acute respiratory syndrome coro .................... .................... ..................... .................... .................... ....................
23097 MT831377.1 |Severe acute respiratory syndrome coro .................... .................... ..................... .................... .................... ....................
23098 MT831378.1 |Severe acute respiratory syndrome coro .................... .................... ..................... .................... .................... ....................
23099 MT831379.1 |Severe acute respiratory syndrome coro .................... .................... ..................... .................... .................... ....................
23100 MT831380.1 |Severe acute respiratory syndrome coro .................... .................... ..................... .................... .................... ....................
23101 MT831381.1 |Severe acute respiratory syndrome coro .................... .................... ..................... .................... .................... ....................
23102 MT831382.1 |Severe acute respiratory syndrome coro .................... .................... ..................... .................... .................... ....................
23103 MT831385.1 |Severe acute respiratory syndrome coro .................... .................... ..................... .................... .................... ....................
23104 MT831386.1 |Severe acute respiratory syndrome coro .................... .................... ..................... .................... .................... ....................
23105 MT831387.1 |Severe acute respiratory syndrome coro .................... .................... ..................... .................... .................... ....................
23106 MT831389.1 |Severe acute respiratory syndrome coro .................... .................... ..................... .................... .................... ....................
23107 MT831390.1 |Severe acute respiratory syndrome coro .................... .................... ..................... .................... .................... ....................
23108 MT831391.1 |Severe acute respiratory syndrome coro .................... .................... ..................... .................... .................... ....................
23109 MT831392.1 |Severe acute respiratory syndrome coro .................... .................... ..................... .................... .................... ....................
23111 MT831394.1 |Severe acute respiratory syndrome coro .................... .................... ..................... .................... .................... ....................
23112 MT831395.1 |Severe acute respiratory syndrome coro .................... .................... ..................... .................... .................... ....................

```
23113 MT831396.1 |Severe acute respiratory syndrome coro    ....................  ....................  ....................  ....................  ....................  ....................
23114 MT831397.1 |Severe acute respiratory syndrome coro    ....................  ....................  ....................  ....................  ....................  ....................
23115 MT831398.1 |Severe acute respiratory syndrome coro    ....................  ....................  ....................  ....................  ....................  ....................
23116 MT831399.1 |Severe acute respiratory syndrome coro    ....................  ....................  ....................  ....................  ....................  ....................
23117 MT831400.1 |Severe acute respiratory syndrome coro    ....................  ....................  ....................  ....................  ....................  ....................
23118 MT831401.1 |Severe acute respiratory syndrome coro    ....................  ....................  ....................  ....................  ....................  ....................
23119 MT831402.1 |Severe acute respiratory syndrome coro    ....................  ....................  ....................  ....................  ....................  ....................
23120 MT831403.1 |Severe acute respiratory syndrome coro    ....................  ....................  ....................  ....................  ....................  ....................
23121 MT831404.1 |Severe acute respiratory syndrome coro    ....................  ....................  ....................  ....................  ....................  ....................
23122 MT831405.1 |Severe acute respiratory syndrome coro    ....................  ....................  ....................  ....................  ....................  ....................
23123 MT831406.1 |Severe acute respiratory syndrome coro    ....................  ....................  ....................  ....................  ....................  ....................
23124 MT831407.1 |Severe acute respiratory syndrome coro    ....................  ....................  ....................  ....................  ....................  ....................
23125 MT831408.1 |Severe acute respiratory syndrome coro    ....................  ....................  ....................  ....................  ....................  ....................
23126 MT831409.1 |Severe acute respiratory syndrome coro    ....................  ....................  ....................  ....................  ....................  ....................
23127 MT831410.1 |Severe acute respiratory syndrome coro    ....................  ....................  ....................  ....................  ....................  ....................
23128 MT831411.1 |Severe acute respiratory syndrome coro    ....................  ....................  ....................  ....................  ....................  ....................
23129 MT831412.1 |Severe acute respiratory syndrome coro    ....................  ....................  ....................  ....................  ....................  ....................
23130 MT831413.1 |Severe acute respiratory syndrome coro    ....................  ....................  ....................  ....................  ....................  ....................
23131 MT831414.1 |Severe acute respiratory syndrome coro    ....................  ....................  ....................  ....................  ....................  ....................
23132 MT831415.1 |Severe acute respiratory syndrome coro    ....................  ....................  ....................  ....................  ....................  ....................
23133 MT831416.1 |Severe acute respiratory syndrome coro    ....................  ....................  ....................  ....................  ....................  ....................
23134 MT831417.1 |Severe acute respiratory syndrome coro    ....................  ....................  ....................  ....................  ....................  ....................
23135 MT831418.1 |Severe acute respiratory syndrome coro    ....................  ....................  ....................  ....................  ....................  ....................
23136 MT831419.1 |Severe acute respiratory syndrome coro    ....................  ....................  ....................  ....................  ....................  ....................
23137 MT831420.1 |Severe acute respiratory syndrome coro    ....................  ....................  ....................  ....................  ....................  ....................
23138 MT831421.1 |Severe acute respiratory syndrome coro    ....................  ....................  ....................  ....................  ....................  ....................
23139 MT831422.1 |Severe acute respiratory syndrome coro    ....................  ....................  ....................  ....................  ....................  ....................
23140 MT831423.1 |Severe acute respiratory syndrome coro    ....................  ....................  ....................  ....................  ....................  ....................
23141 MT831424.1 |Severe acute respiratory syndrome coro    ....................  ....................  ....................  ....................  ....................  ....................
23142 MT831425.1 |Severe acute respiratory syndrome coro    ....................  ....................  ....................  ....................  ....................  ....................
23143 MT831426.1 |Severe acute respiratory syndrome coro    ....................  ....................  ....................  ....................  ....................  ....................
23144 MT831427.1 |Severe acute respiratory syndrome coro    ....................  ....................  ....................  ....................  ....................  ....................
23145 MT831428.1 |Severe acute respiratory syndrome coro    ....................  ....................  ....................  ....................  ....................  ....................
23146 MT831429.1 |Severe acute respiratory syndrome coro    ....................  ....................  ....................  ....................  ....................  ....................
23147 MT831431.1 |Severe acute respiratory syndrome coro    ....................  ....................  ....................  ....................  ....................  ....................
23148 MT831432.1 |Severe acute respiratory syndrome coro    ....................  ....................  ....................  ....................  ....................  ....................
23149 MT831433.1 |Severe acute respiratory syndrome coro    ....................  ....................  ....................  ....................  ....................  ....................
23150 MT831434.1 |Severe acute respiratory syndrome coro    ....................  ....................  ....................  ....................  ....................  ....................
23151 MT831435.1 |Severe acute respiratory syndrome coro    ....................  ....................  ....................  ....................  ....................  ....................
23152 MT831436.1 |Severe acute respiratory syndrome coro    ....................  ....................  ....................  ....................  ....................  ....................
23153 MT831437.1 |Severe acute respiratory syndrome coro    ....................  ....................  ....................  ....................  ....................  ....................
23154 MT831438.1 |Severe acute respiratory syndrome coro    ....................  ....................  ....................  ....................  ....................  ....................
23155 MT831439.1 |Severe acute respiratory syndrome coro    ....................  ....................  ....................  ....................  ....................  ....................
23156 MT831440.1 |Severe acute respiratory syndrome coro    ....................  ....................  ....................  ....................  ....................  ....................
23157 MT831441.1 |Severe acute respiratory syndrome coro    ....................  ....................  ....................  ....................  ....................  ....................
23158 MT831442.1 |Severe acute respiratory syndrome coro    ....................  ....................  ....................  ....................  ....................  ....................
23159 MT831443.1 |Severe acute respiratory syndrome coro    ....................  ....................  ....................  ....................  ....................  ....................
23160 MT831444.1 |Severe acute respiratory syndrome coro    ....................  ....................  ....................  ....................  ....................  ....................
23161 MT831445.1 |Severe acute respiratory syndrome coro    ....................  ....................  ....................  ....................  ....................  ....................
23162 MT831446.1 |Severe acute respiratory syndrome coro    ....................  ....................  ....................  ....................  ....................  ....................
23163 MT831447.1 |Severe acute respiratory syndrome coro    ....................  ....................  ....................  ....................  ....................  ....................
23164 MT831448.1 |Severe acute respiratory syndrome coro    ....................  ....................  ....................  ....................  ....................  ....................
23165 MT831449.1 |Severe acute respiratory syndrome coro    ....................  ....................  ....................  ....................  ....................  ....................
23166 MT831450.1 |Severe acute respiratory syndrome coro    ....................  ....................  ....................  ....................  ....................  ....................
23167 MT831451.1 |Severe acute respiratory syndrome coro    ....................  ....................  ....................  ....................  ....................  ....................
23168 MT831452.1 |Severe acute respiratory syndrome coro    ....................  ....................  ....................  ....................  ....................  ....................
23169 MT831453.1 |Severe acute respiratory syndrome coro    ....................  ....................  ....................  ....................  ....................  ....................
23170 MT831454.1 |Severe acute respiratory syndrome coro    ....................  ....................  ....................  ....................  ....................  ....................
23171 MT831455.1 |Severe acute respiratory syndrome coro    ....................  ....................  ....................  ....................  ....................  ....................
23172 MT831456.1 |Severe acute respiratory syndrome coro    ....................  ....................  ....................  ....................  ....................  ....................
23173 MT831457.1 |Severe acute respiratory syndrome coro    ....................  ....................  ....................  ....................  ....................  ....................
23174 MT831458.1 |Severe acute respiratory syndrome coro    ....................  ....................  ....................  ....................  ....................  ....................
23175 MT831459.1 |Severe acute respiratory syndrome coro    ....................  ....................  ....................  ....................  ....................  ....................
23176 MT831460.1 |Severe acute respiratory syndrome coro    ....................  ....................  ....................  ....................  ....................  ....................
23177 MT831461.1 |Severe acute respiratory syndrome coro    ....................  ....................  ....................  ....................  ....................  ....................
23178 MT831462.1 |Severe acute respiratory syndrome coro    ....................  ....................  ....................  ....................  ....................  ....................
23179 MT831463.1 |Severe acute respiratory syndrome coro    ....................  ....................  ....................  ....................  ....................  ....................
23180 MT831464.1 |Severe acute respiratory syndrome coro    ....................  ....................  ....................  ....................  ....................  ....................
23181 MT831465.1 |Severe acute respiratory syndrome coro    ....................  ....................  ....................  ....................  ....................  ....................
23182 MT831467.1 |Severe acute respiratory syndrome coro    ....................  ....................  ....................  ....................  ....................  ....................
23183 MT831468.1 |Severe acute respiratory syndrome coro    ....................  ....................  ....................  ....................  ....................  ....................
23184 MT831469.1 |Severe acute respiratory syndrome coro    ....................  ....................  ....................  ....................  ....................  ....................
23185 MT831470.1 |Severe acute respiratory syndrome coro    ....................  ....................  ....................  ....................  ....................  ....................
23186 MT831471.1 |Severe acute respiratory syndrome coro    ....................  ....................  ....................  ....................  ....................  ....................
23187 MT831472.1 |Severe acute respiratory syndrome coro    ....................  ....................  ....................  ....................  ....................  ....................
23188 MT831473.1 |Severe acute respiratory syndrome coro    ....................  ....................  ....................  ....................  ....................  ....................
23189 MT831474.1 |Severe acute respiratory syndrome coro    ....................  ....................  ....................  ....................  ....................  ....................
23190 MT831475.1 |Severe acute respiratory syndrome coro    ....................  ....................  ....................  ....................  ....................  ....................
```

| | | | | | | | | |
|---|---|---|---|---|---|---|---|---|
| 23191 | MT831476.1 | \|Severe acute respiratory syndrome coro | .................... | .................... | .................... | .................... | .................... | .................... |
| 23192 | MT831477.1 | \|Severe acute respiratory syndrome coro | .................... | .................... | .................... | .................... | .................... | .................... |
| 23193 | MT831478.1 | \|Severe acute respiratory syndrome coro | .................... | .................... | .................... | .................... | .................... | .................... |
| 23194 | MT831479.1 | \|Severe acute respiratory syndrome coro | .................... | .................... | .................... | .................... | .................... | .................... |
| 23195 | MT831480.1 | \|Severe acute respiratory syndrome coro | .................... | .................... | .................... | .................... | .................... | .................... |
| 23196 | MT831482.1 | \|Severe acute respiratory syndrome coro | .................... | .................... | .................... | .................... | .................... | .................... |
| 23197 | MT831483.1 | \|Severe acute respiratory syndrome coro | .................... | .................... | .................... | .................... | .................... | .................... |
| 23198 | MT831484.1 | \|Severe acute respiratory syndrome coro | .................... | .................... | .................... | .................... | .................... | .................... |
| 23199 | MT831485.1 | \|Severe acute respiratory syndrome coro | .................... | .................... | .................... | .................... | .................... | .................... |
| 23200 | MT831486.1 | \|Severe acute respiratory syndrome coro | .................... | .................... | .................... | .................... | .................... | .................... |
| 23201 | MT831531.1 | \|Severe acute respiratory syndrome coro | .................... | .................... | .................... | .................... | .................... | .................... |
| 23202 | MT831532.1 | \|Severe acute respiratory syndrome coro | .................... | .................... | .................... | .................... | .................... | .................... |
| 23203 | MT831533.1 | \|Severe acute respiratory syndrome coro | .................... | .................... | .................... | .................... | .................... | .................... |
| 23204 | MT831534.1 | \|Severe acute respiratory syndrome coro | .................... | .................... | .................... | .................... | .................... | .................... |
| 23205 | MT831535.1 | \|Severe acute respiratory syndrome coro | .................... | .................... | .................... | .................... | .................... | .................... |
| 23206 | MT831536.1 | \|Severe acute respiratory syndrome coro | .................... | .................... | .................... | .................... | .................... | .................... |
| 23207 | MT831537.1 | \|Severe acute respiratory syndrome coro | .................... | .................... | .................... | .................... | .................... | .................... |
| 23208 | MT831538.1 | \|Severe acute respiratory syndrome coro | .................... | .................... | .................... | .................... | .................... | .................... |
| 23209 | MT831539.1 | \|Severe acute respiratory syndrome coro | .................... | .................... | .................... | .................... | .................... | .................... |
| 23210 | MT831540.1 | \|Severe acute respiratory syndrome coro | .................... | .................... | .................... | .................... | .................... | .................... |
| 23211 | MT831541.1 | \|Severe acute respiratory syndrome coro | .................... | .................... | .................... | .................... | .................... | .................... |
| 23212 | MT831542.1 | \|Severe acute respiratory syndrome coro | .................... | .................... | .................... | .................... | .................... | .................... |
| 23213 | MT831543.1 | \|Severe acute respiratory syndrome coro | .................... | .................... | .................... | .................... | .................... | .................... |
| 23214 | MT831544.1 | \|Severe acute respiratory syndrome coro | .................... | .................... | .................... | .................... | .................... | .................... |
| 23215 | MT831545.1 | \|Severe acute respiratory syndrome coro | .................... | .................... | .................... | .................... | .................... | .................... |
| 23216 | MT831546.1 | \|Severe acute respiratory syndrome coro | .................... | .................... | .................... | .................... | .................... | .................... |
| 23217 | MT831547.1 | \|Severe acute respiratory syndrome coro | .................... | .................... | .................... | .................... | .................... | .................... |
| 23218 | MT831548.1 | \|Severe acute respiratory syndrome coro | .................... | .................... | .................... | .................... | .................... | .................... |
| 23219 | MT831549.1 | \|Severe acute respiratory syndrome coro | .................... | .................... | .................... | .................... | .................... | .................... |
| 23220 | MT831550.1 | \|Severe acute respiratory syndrome coro | .................... | .................... | .................... | .................... | .................... | .................... |
| 23221 | MT831552.1 | \|Severe acute respiratory syndrome coro | .................... | .................... | .................... | .................... | .................... | .................... |
| 23222 | MT831553.1 | \|Severe acute respiratory syndrome coro | .................... | .................... | .................... | .................... | .................... | .................... |
| 23223 | MT831554.1 | \|Severe acute respiratory syndrome coro | .................... | .................... | .................... | .................... | .................... | .................... |
| 23224 | MT831555.1 | \|Severe acute respiratory syndrome coro | .................... | .................... | .................... | .................... | .................... | .................... |
| 23225 | MT831556.1 | \|Severe acute respiratory syndrome coro | .................... | .................... | .................... | .................... | .................... | .................... |
| 23226 | MT831557.1 | \|Severe acute respiratory syndrome coro | .................... | .................... | .................... | .................... | .................... | .................... |
| 23227 | MT831558.1 | \|Severe acute respiratory syndrome coro | .................... | .................... | .................... | .................... | .................... | .................... |
| 23228 | MT831559.1 | \|Severe acute respiratory syndrome coro | .................... | .................... | .................... | .................... | .................... | .................... |
| 23229 | MT831561.1 | \|Severe acute respiratory syndrome coro | .................... | .................... | .................... | .................... | .................... | .................... |
| 23230 | MT831562.1 | \|Severe acute respiratory syndrome coro | .................... | .................... | .................... | .................... | .................... | .................... |
| 23231 | MT831563.1 | \|Severe acute respiratory syndrome coro | .................... | .................... | .................... | .................... | .................... | .................... |
| 23232 | MT831564.1 | \|Severe acute respiratory syndrome coro | .................... | .................... | .................... | .................... | .................... | .................... |
| 23233 | MT831565.1 | \|Severe acute respiratory syndrome coro | .................... | .................... | .................... | .................... | .................... | .................... |
| 23234 | MT831566.1 | \|Severe acute respiratory syndrome coro | .................... | .................... | .................... | .................... | .................... | .................... |
| 23235 | MT831567.1 | \|Severe acute respiratory syndrome coro | .................... | .................... | .................... | .................... | .................... | .................... |
| 23236 | MT831568.1 | \|Severe acute respiratory syndrome coro | .................... | .................... | .................... | .................... | .................... | .................... |
| 23237 | MT831569.1 | \|Severe acute respiratory syndrome coro | .................... | .................... | .................... | .................... | .................... | .................... |
| 23238 | MT831570.1 | \|Severe acute respiratory syndrome coro | .................... | .................... | .................... | .................... | .................... | .................... |
| 23239 | MT831571.1 | \|Severe acute respiratory syndrome coro | .................... | .................... | .................... | .................... | .................... | .................... |
| 23240 | MT831572.1 | \|Severe acute respiratory syndrome coro | .................... | .................... | .................... | .................... | .................... | .................... |
| 23241 | MT831573.1 | \|Severe acute respiratory syndrome coro | .................... | .................... | .................... | .................... | .................... | .................... |
| 23242 | MT831574.1 | \|Severe acute respiratory syndrome coro | .................... | .................... | .................... | .................... | .................... | .................... |
| 23243 | MT831575.1 | \|Severe acute respiratory syndrome coro | .................... | .................... | .................... | .................... | .................... | .................... |
| 23244 | MT831576.1 | \|Severe acute respiratory syndrome coro | .................... | .................... | .................... | .................... | .................... | .................... |
| 23245 | MT831577.1 | \|Severe acute respiratory syndrome coro | .................... | .................... | .................... | .................... | .................... | .................... |
| 23246 | MT831578.1 | \|Severe acute respiratory syndrome coro | .................... | .................... | .................... | .................... | .................... | .................... |
| 23247 | MT831579.1 | \|Severe acute respiratory syndrome coro | .................... | .................... | .................... | .................... | .................... | .................... |
| 23248 | MT831580.1 | \|Severe acute respiratory syndrome coro | .................... | .................... | .................... | .................... | .................... | .................... |
| 23249 | MT831581.1 | \|Severe acute respiratory syndrome coro | .................... | .................... | .................... | .................... | .................... | .................... |
| 23250 | MT831582.1 | \|Severe acute respiratory syndrome coro | .................... | .................... | .................... | .................... | .................... | .................... |
| 23251 | MT831583.1 | \|Severe acute respiratory syndrome coro | .................... | .................... | .................... | .................... | .................... | .................... |
| 23252 | MT831584.1 | \|Severe acute respiratory syndrome coro | .................... | .................... | .................... | .................... | .................... | .................... |
| 23253 | MT831585.1 | \|Severe acute respiratory syndrome coro | .................... | .................... | .................... | .................... | .................... | .................... |
| 23254 | MT831586.1 | \|Severe acute respiratory syndrome coro | .................... | .................... | .................... | .................... | .................... | .................... |
| 23255 | MT831587.1 | \|Severe acute respiratory syndrome coro | .................... | .................... | .................... | .................... | .................... | .................... |
| 23256 | MT831588.1 | \|Severe acute respiratory syndrome coro | .................... | .................... | .................... | .................... | .................... | .................... |
| 23257 | MT831589.1 | \|Severe acute respiratory syndrome coro | .................... | .................... | .................... | .................... | .................... | .................... |
| 23258 | MT831590.1 | \|Severe acute respiratory syndrome coro | .................... | .................... | .................... | .................... | .................... | .................... |
| 23259 | MT831591.1 | \|Severe acute respiratory syndrome coro | .................... | .................... | .................... | .................... | .................... | .................... |
| 23260 | MT831593.1 | \|Severe acute respiratory syndrome coro | .................... | .................... | .................... | .................... | .................... | .................... |
| 23261 | MT831594.1 | \|Severe acute respiratory syndrome coro | .................... | .................... | .................... | .................... | .................... | .................... |
| 23262 | MT831595.1 | \|Severe acute respiratory syndrome coro | .................... | .................... | .................... | .................... | .................... | .................... |
| 23263 | MT831596.1 | \|Severe acute respiratory syndrome coro | .................... | .................... | .................... | .................... | .................... | .................... |
| 23264 | MT831597.1 | \|Severe acute respiratory syndrome coro | .................... | .................... | .................... | .................... | .................... | .................... |
| 23265 | MT831598.1 | \|Severe acute respiratory syndrome coro | .................... | .................... | .................... | .................... | .................... | .................... |
| 23266 | MT831599.1 | \|Severe acute respiratory syndrome coro | .................... | .................... | .................... | .................... | .................... | .................... |
| 23267 | MT831601.1 | \|Severe acute respiratory syndrome coro | .................... | .................... | .................... | .................... | .................... | .................... |
| 23268 | MT831602.1 | \|Severe acute respiratory syndrome coro | .................... | .................... | .................... | .................... | .................... | .................... |

```
23269 MT831603.1 |Severe acute respiratory syndrome coro    ....................  ....................  ....................  ....................  ....................  ....................
23270 MT831604.1 |Severe acute respiratory syndrome coro    ....................  ....................  ....................  ....................  ....................  ....................
23271 MT831605.1 |Severe acute respiratory syndrome coro    ....................  ....................  ....................  ....................  ....................  ....................
23272 MT831606.1 |Severe acute respiratory syndrome coro    ....................  ....................  ....................  ....................  ....................  ....................
23273 MT831607.1 |Severe acute respiratory syndrome coro    ....................  ....................  ....................  ....................  ....................  ....................
23274 MT831608.1 |Severe acute respiratory syndrome coro    ....................  ....................  ....................  ....................  ....................  ....................
23275 MT831609.1 |Severe acute respiratory syndrome coro    ....................  ....................  ....................  ....................  ....................  ....................
23276 MT831610.1 |Severe acute respiratory syndrome coro    ....................  ....................  ....................  ....................  ....................  ....................
23277 MT831611.1 |Severe acute respiratory syndrome coro    ....................  ....................  ....................  ....................  ....................  ....................
23278 MT831612.1 |Severe acute respiratory syndrome coro    ....................  ....................  ....................  ....................  ....................  ....................
23279 MT831613.1 |Severe acute respiratory syndrome coro    ....................  ....................  ....................  ....................  ....................  ....................
23280 MT831614.1 |Severe acute respiratory syndrome coro    ....................  ....................  ....................  ....................  ....................  ....................
23281 MT831615.1 |Severe acute respiratory syndrome coro    ....................  ....................  ....................  ....................  ....................  ....................
23282 MT831616.1 |Severe acute respiratory syndrome coro    ....................  ....................  ....................  ....................  ....................  ....................
23283 MT831617.1 |Severe acute respiratory syndrome coro    ....................  ....................  ....................  ....................  ....................  ....................
23284 MT831618.1 |Severe acute respiratory syndrome coro    ....................  ....................  ....................  ....................  ....................  ....................
23285 MT831619.1 |Severe acute respiratory syndrome coro    ....................  ....................  ....................  ....................  ....................  ....................
23286 MT831620.1 |Severe acute respiratory syndrome coro    ....................  ....................  ....................  ....................  ....................  ....................
23287 MT831621.1 |Severe acute respiratory syndrome coro    ....................  ....................  ....................  ....................  ....................  ....................
23288 MT831623.1 |Severe acute respiratory syndrome coro    ....................  ....................  ....................  ....................  ....................  ....................
23289 MT831624.1 |Severe acute respiratory syndrome coro    ....................  ....................  ....................  ....................  ....................  ....................
23290 MT831625.1 |Severe acute respiratory syndrome coro    ....................  ....................  ....................  ....................  ....................  ....................
23291 MT831626.1 |Severe acute respiratory syndrome coro    ....................  ....................  ....................  ....................  ....................  ....................
23292 MT831627.1 |Severe acute respiratory syndrome coro    ....................  ....................  ....................  ....................  ....................  ....................
23293 MT831629.1 |Severe acute respiratory syndrome coro    ....................  ....................  ....................  ....................  ....................  ....................
23294 MT831630.1 |Severe acute respiratory syndrome coro    ....................  ....................  ....................  ....................  ....................  ....................
23295 MT831631.1 |Severe acute respiratory syndrome coro    ....................  ....................  ....................  ....................  ....................  ....................
23296 MT831632.1 |Severe acute respiratory syndrome coro    ....................  ....................  ....................  ....................  ....................  ....................
23297 MT831633.1 |Severe acute respiratory syndrome coro    ....................  ....................  ....................  ....................  ....................  ....................
23298 MT831634.1 |Severe acute respiratory syndrome coro    ....................  ....................  ....................  ....................  ....................  ....................
23299 MT831635.1 |Severe acute respiratory syndrome coro    ....................  ....................  ....................  ....................  ....................  ....................
23300 MT831636.1 |Severe acute respiratory syndrome coro    ....................  ....................  ....................  ....................  ....................  ....................
23301 MT831637.1 |Severe acute respiratory syndrome coro    ....................  ....................  ....................  ....................  ....................  ....................
23302 MT831638.1 |Severe acute respiratory syndrome coro    ....................  ....................  ....................  ....................  ....................  ....................
23303 MT831639.1 |Severe acute respiratory syndrome coro    ....................  ....................  ....................  ....................  ....................  ....................
23304 MT831640.1 |Severe acute respiratory syndrome coro    ....................  ....................  ....................  ....................  ....................  ....................
23305 MT831641.1 |Severe acute respiratory syndrome coro    ....................  ....................  ....................  ....................  ....................  ....................
23306 MT831642.1 |Severe acute respiratory syndrome coro    ....................  ....................  ....................  ....................  ....................  ....................
23307 MT831643.1 |Severe acute respiratory syndrome coro    ....................  ....................  ....................  ....................  ....................  ....................
23308 MT831644.1 |Severe acute respiratory syndrome coro    ....................  ....................  ....................  ....................  ....................  ....................
23309 MT831645.1 |Severe acute respiratory syndrome coro    ....................  ....................  ....................  ....................  ....................  ....................
23310 MT831646.1 |Severe acute respiratory syndrome coro    ....................  ....................  ....................  ....................  ....................  ....................
23311 MT831648.1 |Severe acute respiratory syndrome coro    ....................  ....................  ....................  ....................  ....................  ....................
23312 MT831649.1 |Severe acute respiratory syndrome coro    ....................  ....................  ....................  ....................  ....................  ....................
23313 MT831650.1 |Severe acute respiratory syndrome coro    ....................  ....................  ....................  ....................  ....................  ....................
23314 MT831652.1 |Severe acute respiratory syndrome coro    ....................  ....................  ....................  ....................  ....................  ....................
23315 MT831653.1 |Severe acute respiratory syndrome coro    ....................  ....................  ....................  ....................  ....................  ....................
23316 MT831654.1 |Severe acute respiratory syndrome coro    ....................  ....................  ....................  ....................  ....................  ....................
23317 MT831655.1 |Severe acute respiratory syndrome coro    ....................  ....................  ....................  ....................  ....................  ....................
23318 MT831656.1 |Severe acute respiratory syndrome coro    ....................  ....................  ....................  ....................  ....................  ....................
23319 MT831657.1 |Severe acute respiratory syndrome coro    ....................  ....................  ....................  ....................  ....................  ....................
23320 MT831658.1 |Severe acute respiratory syndrome coro    ....................  ....................  ....................  ....................  ....................  ....................
23321 MT831659.1 |Severe acute respiratory syndrome coro    ....................  ....................  ....................  ....................  ....................  ....................
23322 MT831660.1 |Severe acute respiratory syndrome coro    ....................  ....................  ....................  ....................  ....................  ....................
23323 MT831661.1 |Severe acute respiratory syndrome coro    ....................  ....................  ....................  ....................  ....................  ....................
23324 MT831662.1 |Severe acute respiratory syndrome coro    ....................  ....................  ....................  ....................  ....................  ....................
23325 MT831663.1 |Severe acute respiratory syndrome coro    ....................  ....................  ....................  ....................  ....................  ....................
23326 MT831664.1 |Severe acute respiratory syndrome coro    ....................  ....................  ....................  ....................  ....................  ....................
23327 MT831665.1 |Severe acute respiratory syndrome coro    ....................  ....................  ....................  ....................  ....................  ....................
23328 MT831666.1 |Severe acute respiratory syndrome coro    ....................  ....................  ....................  ....................  ....................  ....................
23329 MT831667.1 |Severe acute respiratory syndrome coro    ....................  ....................  ....................  ....................  ....................  ....................
23330 MT831668.1 |Severe acute respiratory syndrome coro    ....................  ....................  ....................  ....................  ....................  ....................
23331 MT831669.1 |Severe acute respiratory syndrome coro    ....................  ....................  ....................  ....................  ....................  ....................
23332 MT831670.1 |Severe acute respiratory syndrome coro    ....................  ....................  ....................  ....................  ....................  ....................
23333 MT831671.1 |Severe acute respiratory syndrome coro    ....................  ....................  ....................  ....................  ....................  ....................
23334 MT831672.1 |Severe acute respiratory syndrome coro    ....................  ....................  ....................  ....................  ....................  ....................
23335 MT831673.1 |Severe acute respiratory syndrome coro    ....................  ....................  ....................  ....................  ....................  ....................
23336 MT831674.1 |Severe acute respiratory syndrome coro    ....................  ....................  ....................  ....................  ....................  ....................
23337 MT831675.1 |Severe acute respiratory syndrome coro    ....................  ....................  ....................  ....................  ....................  ....................
23338 MT831676.1 |Severe acute respiratory syndrome coro    ....................  ....................  ....................  ....................  ....................  ....................
23339 MT831677.1 |Severe acute respiratory syndrome coro    ....................  ....................  ....................  ....................  ....................  ....................
23340 MT831678.1 |Severe acute respiratory syndrome coro    ....................  ....................  ....................  ....................  ....................  ....................
23341 MT831679.1 |Severe acute respiratory syndrome coro    ....................  ....................  ....................  ....................  ....................  ....................
23342 MT831680.1 |Severe acute respiratory syndrome coro    ....................  ....................  ....................  ....................  ....................  ....................
23343 MT831681.1 |Severe acute respiratory syndrome coro    ....................  ....................  ....................  ....................  ....................  ....................
23344 MT831682.1 |Severe acute respiratory syndrome coro    ....................  ....................  ....................  ....................  ....................  ....................
23345 MT831683.1 |Severe acute respiratory syndrome coro    ....................  ....................  ....................  ....................  ....................  ....................
23346 MT831684.1 |Severe acute respiratory syndrome coro    ....................  ....................  ....................  ....................  ....................  ....................
```

23347 MT831685.1 |Severe acute respiratory syndrome coro .................... .................... .................... .................... .................... ....................
23348 MT831686.1 |Severe acute respiratory syndrome coro .................... .................... .................... .................... .................... ....................
23349 MT831687.1 |Severe acute respiratory syndrome coro .................... .................... .................... .................... .................... ....................
23350 MT831688.1 |Severe acute respiratory syndrome coro .................... .................... .................... .................... .................... ....................
23351 MT831689.1 |Severe acute respiratory syndrome coro .................... .................... .................... .................... .................... ....................
23352 MT831690.1 |Severe acute respiratory syndrome coro .................... .................... .................... .................... .................... ....................
23353 MT831691.1 |Severe acute respiratory syndrome coro .................... .................... .................... .................... .................... ....................
23354 MT831692.1 |Severe acute respiratory syndrome coro .................... .................... .................... .................... .................... ....................
23355 MT831693.1 |Severe acute respiratory syndrome coro .................... .................... .................... .................... .................... ....................
23356 MT831694.1 |Severe acute respiratory syndrome coro .................... .................... .................... .................... .................... ....................
23357 MT831695.1 |Severe acute respiratory syndrome coro .................... .................... .................... .................... .................... ....................
23358 MT831696.1 |Severe acute respiratory syndrome coro .................... .................... .................... .................... .................... ....................
23359 MT831697.1 |Severe acute respiratory syndrome coro .................... .................... .................... .................... .................... ....................
23360 MT831698.1 |Severe acute respiratory syndrome coro .................... .................... .................... .................... .................... ....................
23361 MT831699.1 |Severe acute respiratory syndrome coro .................... .................... .................... .................... .................... ....................
23362 MT831700.1 |Severe acute respiratory syndrome coro .................... .................... .................... .................... .................... ....................
23363 MT831701.1 |Severe acute respiratory syndrome coro .................... .................... .................... .................... .................... ....................
23364 MT831702.1 |Severe acute respiratory syndrome coro .................... .................... .................... .................... .................... ....................
23365 MT831703.1 |Severe acute respiratory syndrome coro .................... .................... .................... .................... .................... ....................
23366 MT831704.1 |Severe acute respiratory syndrome coro .................... .................... .................... .................... .................... ....................
23367 MT831705.1 |Severe acute respiratory syndrome coro .................... .................... .................... .................... .................... ....................
23368 MT831706.1 |Severe acute respiratory syndrome coro .................... .................... .................... .................... .................... ....................
23369 MT831707.1 |Severe acute respiratory syndrome coro .................... .................... .................... .................... .................... ....................
23370 MT831708.1 |Severe acute respiratory syndrome coro .................... .................... .................... .................... .................... ....................
23371 MT831709.1 |Severe acute respiratory syndrome coro .................... .................... .................... .................... .................... ....................
23372 MT831710.1 |Severe acute respiratory syndrome coro .................... .................... .................... .................... .................... ....................
23373 MT831711.1 |Severe acute respiratory syndrome coro .................... .................... .................... .................... .................... ....................
23374 MT831713.1 |Severe acute respiratory syndrome coro .................... .................... .................... .................... .................... ....................
23375 MT831714.1 |Severe acute respiratory syndrome coro .................... .................... .................... .................... .................... ....................
23376 MT831716.1 |Severe acute respiratory syndrome coro .................... .................... .................... .................... .................... ....................
23377 MT831718.1 |Severe acute respiratory syndrome coro .................... .................... .................... .................... .................... ....................
23378 MT831719.1 |Severe acute respiratory syndrome coro .................... .................... .................... .................... .................... ....................
23379 MT831720.1 |Severe acute respiratory syndrome coro .................... .................... .................... .................... .................... ....................
23380 MT831721.1 |Severe acute respiratory syndrome coro .................... .................... .................... .................... .................... ....................
23381 MT831722.1 |Severe acute respiratory syndrome coro .................... .................... .................... .................... .................... ....................
23382 MT831723.1 |Severe acute respiratory syndrome coro .................... .................... .................... .................... .................... ....................
23383 MT831724.1 |Severe acute respiratory syndrome coro .................... .................... .................... .................... .................... ....................
23384 MT831725.1 |Severe acute respiratory syndrome coro .................... .................... .................... .................... .................... ....................
23385 MT831726.1 |Severe acute respiratory syndrome coro .................... .................... .................... .................... .................... ....................
23386 MT831727.1 |Severe acute respiratory syndrome coro .................... .................... .................... .................... .................... ....................
23387 MT831728.1 |Severe acute respiratory syndrome coro .................... .................... .................... .................... .................... ....................
23388 MT831729.1 |Severe acute respiratory syndrome coro .................... .................... .................... .................... .................... ....................
23389 MT831730.1 |Severe acute respiratory syndrome coro .................... .................... .................... .................... .................... ....................
23390 MT831731.1 |Severe acute respiratory syndrome coro .................... .................... .................... .................... .................... ....................
23391 MT831732.1 |Severe acute respiratory syndrome coro .................... .................... .................... .................... .................... ....................
23392 MT831733.1 |Severe acute respiratory syndrome coro .................... .................... .................... .................... .................... ....................
23393 MT831734.1 |Severe acute respiratory syndrome coro .................... .................... .................... .................... .................... ....................
23394 MT831735.1 |Severe acute respiratory syndrome coro .................... .................... .................... .................... .................... ....................
23395 MT831736.1 |Severe acute respiratory syndrome coro .................... .................... .................... .................... .................... ....................
23396 MT831737.1 |Severe acute respiratory syndrome coro .................... .................... .................... .................... .................... ....................
23397 MT831738.1 |Severe acute respiratory syndrome coro .................... .................... .................... .................... .................... ....................
23398 MT831739.1 |Severe acute respiratory syndrome coro .................... .................... .................... .................... .................... ....................
23399 MT831740.1 |Severe acute respiratory syndrome coro .................... .................... .................... .................... .................... ....................
23400 MT831741.1 |Severe acute respiratory syndrome coro .................... .................... .................... .................... .................... ....................
23401 MT831742.1 |Severe acute respiratory syndrome coro .................... .................... .................... .................... .................... ....................
23402 MT831743.1 |Severe acute respiratory syndrome coro .................... .................... .................... .................... .................... ....................
23403 MT831744.1 |Severe acute respiratory syndrome coro .................... .................... .................... .................... .................... ....................
23404 MT831745.1 |Severe acute respiratory syndrome coro .................... .................... .................... .................... .................... ....................
23405 MT831746.1 |Severe acute respiratory syndrome coro .................... .................... .................... .................... .................... ....................
23406 MT831747.1 |Severe acute respiratory syndrome coro .................... .................... .................... .................... .................... ....................
23407 MT831748.1 |Severe acute respiratory syndrome coro .................... .................... .................... .................... .................... ....................
23408 MT831749.1 |Severe acute respiratory syndrome coro .................... .................... .................... .................... .................... ....................
23409 MT831750.1 |Severe acute respiratory syndrome coro .................... .................... .................... .................... .................... ....................
23410 MT831751.1 |Severe acute respiratory syndrome coro .................... .................... .................... .................... .................... ....................
23411 MT831752.1 |Severe acute respiratory syndrome coro .................... .................... .................... .................... .................... ....................
23412 MT831753.1 |Severe acute respiratory syndrome coro .................... .................... .................... .................... .................... ....................
23413 MT831754.1 |Severe acute respiratory syndrome coro .................... .A.................. .................... .................... .................... ....................
23414 MT831755.1 |Severe acute respiratory syndrome coro .................... .................... .................... .................... .................... ....................
23415 MT831756.1 |Severe acute respiratory syndrome coro .................... .................... .................... .................... .................... ....................
23416 MT831757.1 |Severe acute respiratory syndrome coro .................... .................... .................... .................... .................... ....................
23417 MT831760.1 |Severe acute respiratory syndrome coro .................... .................... .................... .................... .................... ....................
23418 MT831761.1 |Severe acute respiratory syndrome coro .................... .................... .................... .................... .................... ....................
23419 MT831762.1 |Severe acute respiratory syndrome coro .................... .................... .................... .................... .................... ....................
23420 MT831763.1 |Severe acute respiratory syndrome coro .................... .................... .................... .................... .................... ....................
23421 MT831764.1 |Severe acute respiratory syndrome coro .................... .................... .................... .................... .................... ....................
23422 MT831765.1 |Severe acute respiratory syndrome coro .................... .................... .................... .................... .................... ....................
23423 MT831766.1 |Severe acute respiratory syndrome coro .................... .................... .................... .................... .................... ....................
23424 MT831767.1 |Severe acute respiratory syndrome coro .................... .................... .................... .................... .................... ....................

```
23425 MT831768.1 |Severe acute respiratory syndrome coro   ....................  ....................  .....................  ....................  ....................  ....................
23426 MT831769.1 |Severe acute respiratory syndrome coro   ....................  ....................  .....................  ....................  ....................  ....................
23427 MT831770.1 |Severe acute respiratory syndrome coro   ....................  ....................  .....................  ....................  ....................  ....................
23428 MT831771.1 |Severe acute respiratory syndrome coro   ....................  ....................  .....................  ....................  ....................  ....................
23429 MT831772.1 |Severe acute respiratory syndrome coro   ....................  ....................  .....................  ....................  ....................  ....................
23430 MT831773.1 |Severe acute respiratory syndrome coro   ....................  ....................  .....................  ....................  ....................  ....................
23431 MT831774.1 |Severe acute respiratory syndrome coro   ....................  ....................  .....................  ....................  ....................  ....................
23432 MT831775.1 |Severe acute respiratory syndrome coro   ....................  ....................  .....................  ....................  ....................  ....................
23433 MT831776.1 |Severe acute respiratory syndrome coro   ....................  ....................  .....................  ....................  ....................  ....................
23434 MT831777.1 |Severe acute respiratory syndrome coro   ....................  ....................  .....................  ....................  ....................  ....................
23435 MT831778.1 |Severe acute respiratory syndrome coro   ....................  ....................  .....................  ....................  ....................  ....................
23436 MT831779.1 |Severe acute respiratory syndrome coro   ....................  ....................  .....................  ....................  ....................  ....................
23437 MT831780.1 |Severe acute respiratory syndrome coro   ....................  ....................  .....................  ....................  ....................  ....................
23438 MT831781.1 |Severe acute respiratory syndrome coro   ....................  ....................  .....................  ....................  ....................  ....................
23439 MT831782.1 |Severe acute respiratory syndrome coro   ....................  ....................  .....................  ....................  ....................  ....................
23440 MT831783.1 |Severe acute respiratory syndrome coro   ....................  ....................  .....................  ....................  ....................  ....................
23441 MT831784.1 |Severe acute respiratory syndrome coro   ....................  ....................  .....................  ....................  ....................  ....................
23442 MT831785.1 |Severe acute respiratory syndrome coro   ....................  ....................  .....................  ....................  ....................  ....................
23443 MT831786.1 |Severe acute respiratory syndrome coro   ....................  ....................  .....................  ....................  ....................  ....................
23444 MT831787.1 |Severe acute respiratory syndrome coro   ....................  ....................  .....................  ....................  ....................  ....................
23445 MT831788.1 |Severe acute respiratory syndrome coro   ....................  ....................  .....................  ....................  ....................  ....................
23446 MT831789.1 |Severe acute respiratory syndrome coro   ....................  ....................  .....................  ....................  ....................  ....................
23447 MT831790.1 |Severe acute respiratory syndrome coro   ....................  ....................  .....................  ....................  ....................  ....................
23448 MT831791.1 |Severe acute respiratory syndrome coro   ....................  ....................  .....................  ....................  ....................  ....................
23449 MT831792.1 |Severe acute respiratory syndrome coro   ....................  ....................  .....................  ....................  ....................  ....................
23450 MT831793.1 |Severe acute respiratory syndrome coro   ....................  ....................  .....................  ....................  ....................  ....................
23451 MT831794.1 |Severe acute respiratory syndrome coro   ....................  ....................  .....................  ....................  ....................  ....................
23452 MT831795.1 |Severe acute respiratory syndrome coro   ....................  ....................  .....................  ....................  ....................  ....................
23453 MT831796.1 |Severe acute respiratory syndrome coro   ....................  ....................  .....................  ....................  ....................  ....................
23454 MT831798.1 |Severe acute respiratory syndrome coro   ....................  ....................  .....................  ....................  ....................  ....................
23455 MT831799.1 |Severe acute respiratory syndrome coro   ....................  ....................  .....................  ....................  ....................  ....................
23456 MT831800.1 |Severe acute respiratory syndrome coro   ....................  ....................  .....................  ....................  ....................  ....................
23457 MT831802.1 |Severe acute respiratory syndrome coro   ....................  ....................  .....................  ....................  ....................  ....................
23458 MT831804.1 |Severe acute respiratory syndrome coro   ....................  ....................  .....................  ....................  ....................  ....................
23459 MT831805.1 |Severe acute respiratory syndrome coro   ....................  ....................  .....................  ....................  ....................  ....................
23460 MT831806.1 |Severe acute respiratory syndrome coro   ....................  ....................  .....................  ....................  ....................  ....................
23461 MT831807.1 |Severe acute respiratory syndrome coro   ....................  ....................  .....................  ....................  ....................  ....................
23462 MT831808.1 |Severe acute respiratory syndrome coro   ....................  ....................  .....................  ....................  ....................  ....................
23463 MT831809.1 |Severe acute respiratory syndrome coro   ....................  ....................  .....................  ....................  ....................  ....................
23464 MT831811.1 |Severe acute respiratory syndrome coro   ....................  ....................  .....................  ....................  ....................  ....................
23465 MT831813.1 |Severe acute respiratory syndrome coro   ....................  ....................  .....................  ....................  ....................  ....................
23466 MT831815.1 |Severe acute respiratory syndrome coro   ....................  ....................  .....................  ....................  ....................  ....................
23467 MT831816.1 |Severe acute respiratory syndrome coro   ....................  ....................  .....................  ....................  ....................  ....................
23468 MT831818.1 |Severe acute respiratory syndrome coro   ....................  ....................  .....................  ....................  ....................  ....................
23469 MT831819.1 |Severe acute respiratory syndrome coro   ....................  ....................  .....................  ....................  ....................  ....................
23470 MT831820.1 |Severe acute respiratory syndrome coro   ....................  ....................  .....................  ....................  ....................  ....................
23471 MT831821.1 |Severe acute respiratory syndrome coro   ....................  ....................  .....................  ....................  ....................  ....................
23472 MT831822.1 |Severe acute respiratory syndrome coro   ....................  ....................  .....................  ....................  ....................  ....................
23473 MT831823.1 |Severe acute respiratory syndrome coro   ....................  ....................  .....................  ....................  ....................  ....................
23474 MT831824.1 |Severe acute respiratory syndrome coro   ....................  ....................  .....................  ....................  ....................  ....................
23475 MT831825.1 |Severe acute respiratory syndrome coro   ....................  ....................  .....................  ....................  ....................  ....................
23476 MT831826.1 |Severe acute respiratory syndrome coro   ....................  ....................  .....................  ....................  ....................  ....................
23477 MT831827.1 |Severe acute respiratory syndrome coro   ....................  ....................  .....................  ....................  ....................  ....................
23478 MT831828.1 |Severe acute respiratory syndrome coro   ....................  ....................  .....................  ....................  ....................  ....................
23479 MT831829.1 |Severe acute respiratory syndrome coro   ....................  ....................  .....................  ....................  ....................  ....................
23480 MT831830.1 |Severe acute respiratory syndrome coro   ....................  ....................  .....................  ....................  ....................  ....................
23481 MT831832.1 |Severe acute respiratory syndrome coro   ....................  ....................  .....................  ....................  ....................  ....................
23482 MT831833.1 |Severe acute respiratory syndrome coro   ....................  ....................  .....................  ....................  ....................  ....................
23483 MT831834.1 |Severe acute respiratory syndrome coro   ....................  ....................  .....................  ....................  ....................  ....................
23484 MT831835.1 |Severe acute respiratory syndrome coro   ....................  ....................  .....................  ....................  ....................  ....................
23485 MT831836.1 |Severe acute respiratory syndrome coro   ....................  ....................  .....................  ....................  ....................  ....................
23486 MT831837.1 |Severe acute respiratory syndrome coro   ....................  ....................  .....................  ....................  ....................  ....................
23487 MT831838.1 |Severe acute respiratory syndrome coro   ....................  ....................  .....................  ....................  ....................  ....................
23488 MT831839.1 |Severe acute respiratory syndrome coro   ....................  ....................  .....................  ....................  ....................  ....................
23489 MT831840.1 |Severe acute respiratory syndrome coro   ....................  ....................  .....................  ....................  ....................  ....................
23490 MT831841.1 |Severe acute respiratory syndrome coro   ....................  ....................  .....................  ....................  ....................  ....................
23491 MT831842.1 |Severe acute respiratory syndrome coro   ....................  ....................  .....................  ....................  ....................  ....................
23492 MT831843.1 |Severe acute respiratory syndrome coro   ....................  ....................  .....................  ....................  ....................  ....................
23493 MT831844.1 |Severe acute respiratory syndrome coro   ....................  ....................  .....................  ....................  ....................  ....................
23494 MT831845.1 |Severe acute respiratory syndrome coro   ....................  ....................  .....................  ....................  ....................  ....................
23495 MT831846.1 |Severe acute respiratory syndrome coro   ....................  ....................  .....................  ....................  ....................  ....................
23496 MT831847.1 |Severe acute respiratory syndrome coro   ....................  ....................  .....................  ....................  ....................  ....................
23497 MT831848.1 |Severe acute respiratory syndrome coro   ....................  ....................  .....................  ....................  ....................  ....................
23498 MT831850.1 |Severe acute respiratory syndrome coro   ....................  ....................  .....................  ....................  ....................  ....................
23499 MT831851.1 |Severe acute respiratory syndrome coro   ....................  ....................  .....................  ....................  ....................  ....................
23500 MT831852.1 |Severe acute respiratory syndrome coro   ....................  ....................  .....................  ....................  ....................  ....................
23501 MT831853.1 |Severe acute respiratory syndrome coro   ....................  ....................  .....................  ....................  ....................  ....................
23502 MT831854.1 |Severe acute respiratory syndrome coro   ....................  ....................  .....................  ....................  ....................  ....................
```

| | | | | | | | | |
|---|---|---|---|---|---|---|---|---|
| 23503 | MT831855.1 | Severe acute respiratory syndrome coro | ···················· | ···················· | ···················· | ···················· | ···················· | ···················· |
| 23504 | MT831856.1 | Severe acute respiratory syndrome coro | ···················· | ···················· | ···················· | ···················· | ···················· | ···················· |
| 23505 | MT831857.1 | Severe acute respiratory syndrome coro | ···················· | ···················· | ···················· | ···················· | ···················· | ···················· |
| 23506 | MT831858.1 | Severe acute respiratory syndrome coro | ···················· | ···················· | ···················· | ···················· | ···················· | ···················· |
| 23507 | MT831859.1 | Severe acute respiratory syndrome coro | ···················· | ···················· | ···················· | ···················· | ···················· | ···················· |
| 23508 | MT831860.1 | Severe acute respiratory syndrome coro | ···················· | ···················· | ···················· | ···················· | ···················· | ···················· |
| 23509 | MT831861.1 | Severe acute respiratory syndrome coro | ···················· | ···················· | ···················· | ···················· | ···················· | ···················· |
| 23510 | MT831862.1 | Severe acute respiratory syndrome coro | ···················· | ···················· | ···················· | ···················· | ···················· | ···················· |
| 23511 | MT831863.1 | Severe acute respiratory syndrome coro | ···················· | ···················· | ···················· | ···················· | ···················· | ···················· |
| 23512 | MT831865.1 | Severe acute respiratory syndrome coro | ···················· | ···················· | ···················· | ···················· | ···················· | ···················· |
| 23513 | MT831866.1 | Severe acute respiratory syndrome coro | ···················· | ···················· | ···················· | ···················· | ···················· | ···················· |
| 23514 | MT831867.1 | Severe acute respiratory syndrome coro | ···················· | ···················· | ···················· | ···················· | ···················· | ···················· |
| 23515 | MT831868.1 | Severe acute respiratory syndrome coro | ···················· | ···················· | ···················· | ···················· | ···················· | ···················· |
| 23516 | MT831869.1 | Severe acute respiratory syndrome coro | ···················· | ···················· | ···················· | ···················· | ···················· | ···················· |
| 23517 | MT831870.1 | Severe acute respiratory syndrome coro | ···················· | ···················· | ···················· | ···················· | ···················· | ···················· |
| 23518 | MT831872.1 | Severe acute respiratory syndrome coro | ···················· | ···················· | ···················· | ···················· | ···················· | ···················· |
| 23519 | MT831873.1 | Severe acute respiratory syndrome coro | ···················· | ···················· | ···················· | ···················· | ···················· | ···················· |
| 23520 | MT831874.1 | Severe acute respiratory syndrome coro | ···················· | ·A·················· | ···················· | ···················· | ···················· | ···················· |
| 23521 | MT831875.1 | Severe acute respiratory syndrome coro | ···················· | ···················· | ···················· | ···················· | ···················· | ···················· |
| 23522 | MT831876.1 | Severe acute respiratory syndrome coro | ···················· | ···················· | ···················· | ···················· | ···················· | ···················· |
| 23523 | MT831877.1 | Severe acute respiratory syndrome coro | ···················· | ···················· | ···················· | ···················· | ···················· | ···················· |
| 23524 | MT831878.1 | Severe acute respiratory syndrome coro | ···················· | ···················· | ···················· | ···················· | ···················· | ···················· |
| 23525 | MT831879.1 | Severe acute respiratory syndrome coro | ···················· | ···················· | ···················· | ···················· | ···················· | ···················· |
| 23526 | MT831880.1 | Severe acute respiratory syndrome coro | ···················· | ·A·················· | ···················· | ···················· | ···················· | ···················· |
| 23527 | MT831881.1 | Severe acute respiratory syndrome coro | ···················· | ···················· | ···················· | ···················· | ···················· | ···················· |
| 23528 | MT831883.1 | Severe acute respiratory syndrome coro | ···················· | ···················· | ···················· | ···················· | ···················· | ···················· |
| 23529 | MT831884.1 | Severe acute respiratory syndrome coro | ···················· | ···················· | ···················· | ···················· | ···················· | ···················· |
| 23530 | MT831885.1 | Severe acute respiratory syndrome coro | ···················· | ···················· | ···················· | ···················· | ···················· | ···················· |
| 23531 | MT831886.1 | Severe acute respiratory syndrome coro | ···················· | ···················· | ···················· | ···················· | ···················· | ···················· |
| 23532 | MT831887.1 | Severe acute respiratory syndrome coro | ···················· | ···················· | ···················· | ···················· | ···················· | ···················· |
| 23533 | MT831888.1 | Severe acute respiratory syndrome coro | ···················· | ···················· | ···················· | ···················· | ···················· | ···················· |
| 23534 | MT831889.1 | Severe acute respiratory syndrome coro | ···················· | ···················· | ···················· | ···················· | ···················· | ···················· |
| 23535 | MT831890.1 | Severe acute respiratory syndrome coro | ···················· | ···················· | ···················· | ···················· | ···················· | ···················· |
| 23536 | MT831891.1 | Severe acute respiratory syndrome coro | ···················· | ···················· | ···················· | ···················· | ···················· | ···················· |
| 23537 | MT831892.1 | Severe acute respiratory syndrome coro | ···················· | ···················· | ···················· | ···················· | ···················· | ···················· |
| 23538 | MT831894.1 | Severe acute respiratory syndrome coro | ···················· | ···················· | ···················· | ···················· | ···················· | ···················· |
| 23539 | MT831895.1 | Severe acute respiratory syndrome coro | ···················· | ···················· | ···················· | ···················· | ···················· | ···················· |
| 23540 | MT831896.1 | Severe acute respiratory syndrome coro | ···················· | ···················· | ···················· | ···················· | ···················· | ···················· |
| 23541 | MT831897.1 | Severe acute respiratory syndrome coro | ···················· | ···················· | ···················· | ···················· | ···················· | ···················· |
| 23542 | MT831898.1 | Severe acute respiratory syndrome coro | ···················· | ···················· | ···················· | ···················· | ···················· | ···················· |
| 23543 | MT831899.1 | Severe acute respiratory syndrome coro | ···················· | ···················· | ···················· | ···················· | ···················· | ···················· |
| 23544 | MT831900.1 | Severe acute respiratory syndrome coro | ···················· | ···················· | ···················· | ···················· | ···················· | ···················· |
| 23545 | MT833958.1 | Severe acute respiratory syndrome coro | ···················· | ···················· | ···················· | ···················· | ···················· | ···················· |
| 23546 | MT833960.1 | Severe acute respiratory syndrome coro | ···················· | ···················· | ···················· | ···················· | ···················· | ···················· |
| 23547 | MT833961.1 | Severe acute respiratory syndrome coro | ···················· | ···················· | ···················· | ···················· | ···················· | ···················· |
| 23548 | MT833962.1 | Severe acute respiratory syndrome coro | ···················· | ···················· | ···················· | ···················· | ···················· | ···················· |
| 23549 | MT833964.1 | Severe acute respiratory syndrome coro | ···················· | ···················· | ···················· | ···················· | ···················· | ···················· |
| 23550 | MT833965.1 | Severe acute respiratory syndrome coro | ···················· | ···················· | ···················· | ···················· | ···················· | ···················· |
| 23551 | MT833966.1 | Severe acute respiratory syndrome coro | ···················· | ···················· | ···················· | ···················· | ···················· | ···················· |
| 23552 | MT833967.1 | Severe acute respiratory syndrome coro | ···················· | ···················· | ···················· | ···················· | ···················· | ···················· |
| 23553 | MT833968.1 | Severe acute respiratory syndrome coro | ···················· | ···················· | ···················· | ···················· | ···················· | ···················· |
| 23554 | MT833969.1 | Severe acute respiratory syndrome coro | ···················· | ···················· | ···················· | ···················· | ···················· | ···················· |
| 23555 | MT833972.1 | Severe acute respiratory syndrome coro | ···················· | ···················· | ···················· | ···················· | ···················· | ···················· |
| 23556 | MT833974.1 | Severe acute respiratory syndrome coro | ···················· | ···················· | ···················· | ···················· | ···················· | ···················· |
| 23557 | MT833975.1 | Severe acute respiratory syndrome coro | ···················· | ···················· | ···················· | ···················· | ···················· | ···················· |
| 23558 | MT833976.1 | Severe acute respiratory syndrome coro | ···················· | ···················· | ···················· | ···················· | ···················· | ···················· |
| 23559 | MT833977.1 | Severe acute respiratory syndrome coro | ···················· | ···················· | ···················· | ···················· | ···················· | ···················· |
| 23560 | MT833978.1 | Severe acute respiratory syndrome coro | ···················· | ···················· | ···················· | ···················· | ···················· | ···················· |
| 23561 | MT833979.1 | Severe acute respiratory syndrome coro | ···················· | ···················· | ···················· | ···················· | ···················· | ···················· |
| 23562 | MT833980.1 | Severe acute respiratory syndrome coro | ···················· | ···················· | ···················· | ···················· | ···················· | ···················· |
| 23563 | MT833981.1 | Severe acute respiratory syndrome coro | ···················· | ···················· | ···················· | ···················· | ···················· | ···················· |
| 23564 | MT833982.1 | Severe acute respiratory syndrome coro | ···················· | ···················· | ···················· | ···················· | ···················· | ···················· |
| 23565 | MT833985.1 | Severe acute respiratory syndrome coro | ···················· | ···················· | ···················· | ···················· | ···················· | ···················· |
| 23566 | MT833986.1 | Severe acute respiratory syndrome coro | ···················· | ···················· | ···················· | ···················· | ···················· | ···················· |
| 23567 | MT833987.1 | Severe acute respiratory syndrome coro | ···················· | ···················· | ···················· | ···················· | ···················· | ···················· |
| 23568 | MT833988.1 | Severe acute respiratory syndrome coro | ···················· | ···················· | ···················· | ···················· | ···················· | ···················· |
| 23569 | MT833989.1 | Severe acute respiratory syndrome coro | ···················· | ···················· | ···················· | ···················· | ···················· | ···················· |
| 23570 | MT833990.1 | Severe acute respiratory syndrome coro | ···················· | ···················· | ···················· | ···················· | ···················· | ···················· |
| 23571 | MT833991.1 | Severe acute respiratory syndrome coro | ···················· | ···················· | ···················· | ···················· | ···················· | ···················· |
| 23572 | MT833992.1 | Severe acute respiratory syndrome coro | ···················· | ···················· | ···················· | ···················· | ···················· | ···················· |
| 23573 | MT833993.1 | Severe acute respiratory syndrome coro | ···················· | ···················· | ···················· | ···················· | ···················· | ···················· |
| 23574 | MT833994.1 | Severe acute respiratory syndrome coro | ···················· | ···················· | ···················· | ···················· | ···················· | ···················· |
| 23575 | MT833995.1 | Severe acute respiratory syndrome coro | ···················· | ···················· | ···················· | ···················· | ···················· | ···················· |
| 23576 | MT833996.1 | Severe acute respiratory syndrome coro | ···················· | ···················· | ···················· | ···················· | ···················· | ···················· |
| 23577 | MT833997.1 | Severe acute respiratory syndrome coro | ···················· | ···················· | ···················· | ···················· | ···················· | ···················· |
| 23578 | MT833998.1 | Severe acute respiratory syndrome coro | ···················· | ···················· | ···················· | ···················· | ···················· | ···················· |
| 23579 | MT833999.1 | Severe acute respiratory syndrome coro | ···················· | ···················· | ···················· | ···················· | ···················· | ···················· |
| 23580 | MT834000.1 | Severe acute respiratory syndrome coro | ···················· | ···················· | ···················· | ···················· | ···················· | ···················· |

| | | | | | | | | |
|---|---|---|---|---|---|---|---|---|
| 23581 | MT834001.1 | \|Severe acute respiratory syndrome coro | ···················· | ···················· | ···················· | ···················· | ···················· | ···················· |
| 23582 | MT834002.1 | \|Severe acute respiratory syndrome coro | ·T·················· | ···················· | ···················· | ···················· | ···················· | ···················· |
| 23583 | MT834003.1 | \|Severe acute respiratory syndrome coro | ···················· | ···················· | ···················· | ···················· | ···················· | ···················· |
| 23584 | MT834004.1 | \|Severe acute respiratory syndrome coro | ···················· | ···················· | ···················· | ···················· | ···················· | ···················· |
| 23585 | MT834005.1 | \|Severe acute respiratory syndrome coro | ···················· | ···················· | ···················· | ···················· | ···················· | ···················· |
| 23586 | MT834006.1 | \|Severe acute respiratory syndrome coro | ···················· | ···················· | ···················· | ···················· | ···················· | ···················· |
| 23587 | MT834007.1 | \|Severe acute respiratory syndrome coro | ···················· | ···················· | ···················· | ···················· | ···················· | ···················· |
| 23588 | MT834008.1 | \|Severe acute respiratory syndrome coro | ···················· | ···················· | ···················· | ···················· | ···················· | ···················· |
| 23589 | MT834009.1 | \|Severe acute respiratory syndrome coro | ···················· | ···················· | ···················· | ···················· | ···················· | ···················· |
| 23590 | MT834010.1 | \|Severe acute respiratory syndrome coro | ···················· | ···················· | ···················· | ···················· | ···················· | ···················· |
| 23591 | MT834011.1 | \|Severe acute respiratory syndrome coro | ···················· | ···················· | ···················· | ···················· | ···················· | ···················· |
| 23592 | MT834012.1 | \|Severe acute respiratory syndrome coro | ···················· | ···················· | ···················· | ···················· | ···················· | ···················· |
| 23593 | MT834013.1 | \|Severe acute respiratory syndrome coro | ···················· | ···················· | ···················· | ···················· | ···················· | ···················· |
| 23594 | MT834014.1 | \|Severe acute respiratory syndrome coro | ···················· | ···················· | ···················· | ···················· | ···················· | ···················· |
| 23595 | MT834015.1 | \|Severe acute respiratory syndrome coro | ···················· | ···················· | ···················· | ···················· | ···················· | ···················· |
| 23596 | MT834016.1 | \|Severe acute respiratory syndrome coro | ···················· | ···················· | ···················· | ···················· | ···················· | ···················· |
| 23597 | MT834017.1 | \|Severe acute respiratory syndrome coro | ···················· | ···················· | ···················· | ···················· | ···················· | ···················· |
| 23598 | MT834019.1 | \|Severe acute respiratory syndrome coro | ···················· | ···················· | ···················· | ···················· | ···················· | ···················· |
| 23599 | MT834020.1 | \|Severe acute respiratory syndrome coro | ···················· | ···················· | ···················· | ···················· | ···················· | ···················· |
| 23600 | MT834021.1 | \|Severe acute respiratory syndrome coro | ···················· | ···················· | ···················· | ···················· | ···················· | ···················· |
| 23601 | MT834022.1 | \|Severe acute respiratory syndrome coro | ···················· | ···················· | ···················· | ···················· | ···················· | ···················· |
| 23602 | MT834023.1 | \|Severe acute respiratory syndrome coro | ···················· | ···················· | ···················· | ···················· | ···················· | ···················· |
| 23603 | MT834024.1 | \|Severe acute respiratory syndrome coro | ···················· | ···················· | ···················· | ···················· | ···················· | ···················· |
| 23604 | MT834025.1 | \|Severe acute respiratory syndrome coro | ···················· | ···················· | ···················· | ···················· | ···················· | ···················· |
| 23605 | MT834026.1 | \|Severe acute respiratory syndrome coro | ···················· | ···················· | ···················· | ···················· | ···················· | ···················· |
| 23606 | MT834027.1 | \|Severe acute respiratory syndrome coro | ···················· | ···················· | ···················· | ···················· | ···················· | ···················· |
| 23607 | MT834028.1 | \|Severe acute respiratory syndrome coro | ···················· | ···················· | ···················· | ···················· | ···················· | ···················· |
| 23608 | MT834029.1 | \|Severe acute respiratory syndrome coro | ···················· | ···················· | ···················· | ···················· | ···················· | ···················· |
| 23609 | MT834030.1 | \|Severe acute respiratory syndrome coro | ···················· | ···················· | ···················· | ···················· | ···················· | ···················· |
| 23610 | MT834031.1 | \|Severe acute respiratory syndrome coro | ···················· | ···················· | ···················· | ···················· | ···················· | ···················· |
| 23611 | MT834032.1 | \|Severe acute respiratory syndrome coro | ···················· | ···················· | ···················· | ···················· | ···················· | ···················· |
| 23612 | MT834033.1 | \|Severe acute respiratory syndrome coro | ···················· | ···················· | ···················· | ···················· | ···················· | ···················· |
| 23613 | MT834034.1 | \|Severe acute respiratory syndrome coro | ···················· | ···················· | ···················· | ···················· | ···················· | ···················· |
| 23614 | MT834035.1 | \|Severe acute respiratory syndrome coro | ···················· | ···················· | ···················· | ···················· | ···················· | ···················· |
| 23615 | MT834036.1 | \|Severe acute respiratory syndrome coro | ···················· | ···················· | ···················· | ···················· | ···················· | ···················· |
| 23616 | MT834037.1 | \|Severe acute respiratory syndrome coro | ···················· | ···················· | ···················· | ···················· | ···················· | ···················· |
| 23617 | MT834038.1 | \|Severe acute respiratory syndrome coro | ···················· | ···················· | ···················· | ···················· | ···················· | ···················· |
| 23618 | MT834039.1 | \|Severe acute respiratory syndrome coro | ···················· | ···················· | ···················· | ···················· | ···················· | ···················· |
| 23619 | MT834040.1 | \|Severe acute respiratory syndrome coro | ···················· | ···················· | ···················· | ···················· | ···················· | ···················· |
| 23620 | MT834041.1 | \|Severe acute respiratory syndrome coro | ···················· | ···················· | ···················· | ···················· | ···················· | ···················· |
| 23621 | MT834042.1 | \|Severe acute respiratory syndrome coro | ···················· | ···················· | ···················· | ···················· | ···················· | ···················· |
| 23622 | MT834043.1 | \|Severe acute respiratory syndrome coro | ···················· | ·A·················· | ···················· | ···················· | ···················· | ···················· |
| 23623 | MT834044.1 | \|Severe acute respiratory syndrome coro | ···················· | ···················· | ···················· | ···················· | ···················· | ···················· |
| 23624 | MT834045.1 | \|Severe acute respiratory syndrome coro | ···················· | ···················· | ···················· | ···················· | ···················· | ···················· |
| 23625 | MT834046.1 | \|Severe acute respiratory syndrome coro | ···················· | ···················· | ···················· | ···················· | ···················· | ···················· |
| 23626 | MT834047.1 | \|Severe acute respiratory syndrome coro | ···················· | ···················· | ···················· | ···················· | ···················· | ···················· |
| 23627 | MT834048.1 | \|Severe acute respiratory syndrome coro | ···················· | ···················· | ···················· | ···················· | ···················· | ···················· |
| 23628 | MT834049.1 | \|Severe acute respiratory syndrome coro | ···················· | ···················· | ···················· | ···················· | ···················· | ···················· |
| 23629 | MT834050.1 | \|Severe acute respiratory syndrome coro | ···················· | ···················· | ···················· | ···················· | ···················· | ···················· |
| 23630 | MT834052.1 | \|Severe acute respiratory syndrome coro | ···················· | ···················· | ···················· | ···················· | ···················· | ···················· |
| 23631 | MT834053.1 | \|Severe acute respiratory syndrome coro | ···················· | ···················· | ···················· | ···················· | ···················· | ···················· |
| 23632 | MT834054.1 | \|Severe acute respiratory syndrome coro | ···················· | ···················· | ···················· | ···················· | ···················· | ···················· |
| 23633 | MT834055.1 | \|Severe acute respiratory syndrome coro | ···················· | ···················· | ···················· | ···················· | ···················· | ···················· |
| 23634 | MT834057.1 | \|Severe acute respiratory syndrome coro | ···················· | ···················· | ···················· | ···················· | ···················· | ···················· |
| 23635 | MT834058.1 | \|Severe acute respiratory syndrome coro | ···················· | ···················· | ···················· | ···················· | ···················· | ···················· |
| 23636 | MT834059.1 | \|Severe acute respiratory syndrome coro | ···················· | ···················· | ···················· | ···················· | ···················· | ···················· |
| 23637 | MT834060.1 | \|Severe acute respiratory syndrome coro | ···················· | ···················· | ···················· | ···················· | ···················· | ···················· |
| 23638 | MT834061.1 | \|Severe acute respiratory syndrome coro | ···················· | ···················· | ···················· | ···················· | ···················· | ···················· |
| 23639 | MT834062.1 | \|Severe acute respiratory syndrome coro | ···················· | ···················· | ···················· | ···················· | ···················· | ···················· |
| 23640 | MT834064.1 | \|Severe acute respiratory syndrome coro | ···················· | ···················· | ···················· | ···················· | ···················· | ···················· |
| 23641 | MT834065.1 | \|Severe acute respiratory syndrome coro | ···················· | ···················· | ···················· | ···················· | ···················· | ···················· |
| 23642 | MT834066.1 | \|Severe acute respiratory syndrome coro | ···················· | ···················· | ···················· | ···················· | ···················· | ···················· |
| 23643 | MT834067.1 | \|Severe acute respiratory syndrome coro | ···················· | ···················· | ···················· | ···················· | ···················· | ···················· |
| 23644 | MT834068.1 | \|Severe acute respiratory syndrome coro | ···················· | ···················· | ···················· | ···················· | ···················· | ···················· |
| 23645 | MT834069.1 | \|Severe acute respiratory syndrome coro | ···················· | ···················· | ···················· | ···················· | ···················· | ···················· |
| 23646 | MT834070.1 | \|Severe acute respiratory syndrome coro | ···················· | ···················· | ···················· | ···················· | ···················· | ···················· |
| 23647 | MT834071.1 | \|Severe acute respiratory syndrome coro | ···················· | ···················· | ···················· | ···················· | ···················· | ···················· |
| 23648 | MT834072.1 | \|Severe acute respiratory syndrome coro | ···················· | ···················· | ···················· | ···················· | ···················· | ···················· |
| 23649 | MT834073.1 | \|Severe acute respiratory syndrome coro | ···················· | ···················· | ···················· | ···················· | ···················· | ···················· |
| 23650 | MT834074.1 | \|Severe acute respiratory syndrome coro | ···················· | ···················· | ···················· | ···················· | ···················· | ···················· |
| 23651 | MT834075.1 | \|Severe acute respiratory syndrome coro | ···················· | ···················· | ···················· | ···················· | ···················· | ···················· |
| 23652 | MT834076.1 | \|Severe acute respiratory syndrome coro | ···················· | ···················· | ···················· | ···················· | ···················· | ···················· |
| 23653 | MT834077.1 | \|Severe acute respiratory syndrome coro | ···················· | ···················· | ···················· | ···················· | ···················· | ···················· |
| 23654 | MT834078.1 | \|Severe acute respiratory syndrome coro | ···················· | ···················· | ···················· | ···················· | ···················· | ···················· |
| 23655 | MT834079.1 | \|Severe acute respiratory syndrome coro | ···················· | ···················· | ···················· | ···················· | ···················· | ···················· |
| 23656 | MT834080.1 | \|Severe acute respiratory syndrome coro | ···················· | C··················· | ···················· | ···················· | ···················· | ···················· |
| 23657 | MT834082.1 | \|Severe acute respiratory syndrome coro | ···················· | ···················· | ···················· | ···················· | ···················· | ···················· |
| 23658 | MT834083.1 | \|Severe acute respiratory syndrome coro | ···················· | ···················· | ···················· | ···················· | ···················· | ···················· |

23659 MT834084.1 |Severe acute respiratory syndrome coro ···················· ···················· ···················· ···················· ···················· ····················
23660 MT834086.1 |Severe acute respiratory syndrome coro ···················· ···················· ···················· ···················· ···················· ····················
23661 MT834087.1 |Severe acute respiratory syndrome coro ···················· ···················· ···················· ···················· ···················· ····················
23662 MT834088.1 |Severe acute respiratory syndrome coro ···················· ···················· ···················· ···················· ···················· ····················
23663 MT834089.1 |Severe acute respiratory syndrome coro ···················· ···················· ···················· ···················· ···················· ····················
23664 MT834090.1 |Severe acute respiratory syndrome coro ···················· ···················· ···················· ···················· ···················· ····················
23665 MT834091.1 |Severe acute respiratory syndrome coro ···················· ···················· ···················· ···················· ···················· ····················
23666 MT834092.1 |Severe acute respiratory syndrome coro ···················· ···················· ···················· ···················· ···················· ····················
23667 MT834093.1 |Severe acute respiratory syndrome coro ···················· ···················· ···················· ···················· ···················· ····················
23668 MT834094.1 |Severe acute respiratory syndrome coro ···················· ···················· ···················· ···················· ···················· ····················
23669 MT834095.1 |Severe acute respiratory syndrome coro ···················· ···················· ···················· ···················· ···················· ····················
23670 MT834096.1 |Severe acute respiratory syndrome coro ···················· ···················· ···················· ···················· ···················· ····················
23671 MT834097.1 |Severe acute respiratory syndrome coro ···················· ···················· ···················· ···················· ···················· ····················
23672 MT834099.1 |Severe acute respiratory syndrome coro ···················· ···················· ···················· ···················· ···················· ····················
23673 MT834100.1 |Severe acute respiratory syndrome coro ···················· ···················· ···················· ···················· ···················· ····················
23674 MT834101.1 |Severe acute respiratory syndrome coro ···················· ···················· ···················· ···················· ···················· ····················
23675 MT834102.1 |Severe acute respiratory syndrome coro ···················· ···················· ···················· ···················· ···················· ····················
23676 MT834103.1 |Severe acute respiratory syndrome coro ···················· ···················· ···················· ···················· ···················· ····················
23677 MT834104.1 |Severe acute respiratory syndrome coro ···················· ···················· ···················· ···················· ···················· ····················
23678 MT834105.1 |Severe acute respiratory syndrome coro ···················· ···················· ···················· ···················· ···················· ····················
23679 MT834106.1 |Severe acute respiratory syndrome coro ···················· ···················· ···················· ···················· ···················· ····················
23680 MT834107.1 |Severe acute respiratory syndrome coro ···················· ···················· ···················· ···················· ···················· ····················
23681 MT834108.1 |Severe acute respiratory syndrome coro ···················· ···················· ···················· ···················· ···················· ····················
23682 MT834109.1 |Severe acute respiratory syndrome coro ···················· ···················· ···················· ···················· ···················· ····················
23683 MT834110.1 |Severe acute respiratory syndrome coro ···················· ···················· ···················· ···················· ···················· ····················
23684 MT834111.1 |Severe acute respiratory syndrome coro ···················· ···················· ···················· ···················· ···················· ····················
23685 MT834112.1 |Severe acute respiratory syndrome coro ···················· ···················· ···················· ···················· ···················· ····················
23686 MT834113.1 |Severe acute respiratory syndrome coro ···················· ···················· ···················· ···················· ···················· ····················
23687 MT834114.1 |Severe acute respiratory syndrome coro ···················· ···················· ···················· ···················· ···················· ····················
23688 MT834115.1 |Severe acute respiratory syndrome coro ···················· ···················· ···················· ···················· ···················· ····················
23689 MT834116.1 |Severe acute respiratory syndrome coro ···················· ···················· ···················· ···················· ···················· ····················
23690 MT834117.1 |Severe acute respiratory syndrome coro ···················· ···················· ···················· ···················· ···················· ····················
23691 MT834118.1 |Severe acute respiratory syndrome coro ···················· ···················· ···················· ···················· ···················· ····················
23692 MT834119.1 |Severe acute respiratory syndrome coro ···················· ···················· ···················· ···················· ···················· ····················
23693 MT834120.1 |Severe acute respiratory syndrome coro ···················· ···················· ···················· ···················· ···················· ····················
23694 MT834121.1 |Severe acute respiratory syndrome coro ···················· ···················· ···················· ···················· ···················· ····················
23695 MT834123.1 |Severe acute respiratory syndrome coro ···················· ···················· ···················· ···················· ···················· ····················
23696 MT834124.1 |Severe acute respiratory syndrome coro ···················· ···················· ···················· ···················· ···················· ····················
23697 MT834125.1 |Severe acute respiratory syndrome coro ···················· ···················· ···················· ···················· ···················· ····················
23698 MT834126.1 |Severe acute respiratory syndrome coro ···················· ···················· ···················· ···················· ···················· ····················
23699 MT834127.1 |Severe acute respiratory syndrome coro ···················· ···················· ···················· ···················· ···················· ····················
23700 MT834128.1 |Severe acute respiratory syndrome coro ···················· ···················· ···················· ···················· ···················· ····················
23701 MT834129.1 |Severe acute respiratory syndrome coro ···················· ···················· ···················· ···················· ···················· ····················
23702 MT834130.1 |Severe acute respiratory syndrome coro ···················· ···················· ···················· ···················· ···················· ····················
23703 MT834132.1 |Severe acute respiratory syndrome coro ···················· ···················· ···················· ···················· ···················· ····················
23704 MT834133.1 |Severe acute respiratory syndrome coro ···················· ···················· ···················· ···················· ···················· ····················
23705 MT834134.1 |Severe acute respiratory syndrome coro ···················· ···················· ···················· ···················· ···················· ····················
23706 MT834135.1 |Severe acute respiratory syndrome coro ···················· ···················· ···················· ···················· ···················· ····················
23707 MT834136.1 |Severe acute respiratory syndrome coro ···················· ···················· ···················· ···················· ···················· ····················
23708 MT834137.1 |Severe acute respiratory syndrome coro ···················· ···················· ···················· ···················· ···················· ····················
23709 MT834138.1 |Severe acute respiratory syndrome coro ···················· ···················· ···················· ···················· ···················· ····················
23710 MT834139.1 |Severe acute respiratory syndrome coro ···················· ···················· ···················· ···················· ··C················· ····················
23711 MT834141.1 |Severe acute respiratory syndrome coro ···················· ···················· ···················· ···················· ···················· ····················
23712 MT834142.1 |Severe acute respiratory syndrome coro ···················· ···················· ···················· ···················· ···················· ····················
23713 MT834143.1 |Severe acute respiratory syndrome coro ···················· ···················· ···················· ···················· ···················· ····················
23714 MT834144.1 |Severe acute respiratory syndrome coro ···················· ···················· ···················· ···················· ···················· ····················
23715 MT834145.1 |Severe acute respiratory syndrome coro ···················· ···················· ···················· ···················· ···················· ····················
23716 MT834146.1 |Severe acute respiratory syndrome coro ···················· ···················· ···················· ···················· ···················· ····················
23717 MT834147.1 |Severe acute respiratory syndrome coro ···················· ···················· ···················· ···················· ···················· ····················
23718 MT834148.1 |Severe acute respiratory syndrome coro ···················· ···················· ···················· ···················· ···················· ····················
23719 MT834150.1 |Severe acute respiratory syndrome coro ···················· ···················· ···················· ···················· ···················· ····················
23720 MT834151.1 |Severe acute respiratory syndrome coro ···················· ···················· ···················· ···················· ···················· ····················
23721 MT834152.1 |Severe acute respiratory syndrome coro ···················· ···················· ···················· ···················· ···················· ····················
23722 MT834153.1 |Severe acute respiratory syndrome coro ···················· ···················· ···················· ···················· ···················· ····················
23723 MT834154.1 |Severe acute respiratory syndrome coro ···················· ···················· ···················· ···················· ···················· ····················
23724 MT834155.1 |Severe acute respiratory syndrome coro ···················· ···················· ···················· ···················· ···················· ····················
23725 MT834156.1 |Severe acute respiratory syndrome coro ···················· ···················· ···················· ···················· ···················· ····················
23726 MT834157.1 |Severe acute respiratory syndrome coro ···················· ···················· ···················· ···················· ···················· ····················
23727 MT834158.1 |Severe acute respiratory syndrome coro ···················· ···················· ···················· ···················· ···················· ····················
23728 MT834159.1 |Severe acute respiratory syndrome coro ···················· ···················· ···················· ···················· ···················· ····················
23729 MT834160.1 |Severe acute respiratory syndrome coro ···················· ···················· ···················· ···················· ···················· ····················
23730 MT834161.1 |Severe acute respiratory syndrome coro ···················· ···················· ···················· ···················· ···················· ····················
23731 MT834162.1 |Severe acute respiratory syndrome coro ···················· ···················· ···················· ···················· ···················· ····················
23732 MT834163.1 |Severe acute respiratory syndrome coro ···················· ···················· ···················· ···················· ···················· ····················
23733 MT834164.1 |Severe acute respiratory syndrome coro ···················· ···················· ···················· ···················· ···················· ····················
23734 MT834165.1 |Severe acute respiratory syndrome coro ···················· ···················· ···················· ···················· ···················· ····················
23735 MT834166.1 |Severe acute respiratory syndrome coro ···················· ···················· ···················· ···················· ···················· ····················
23736 MT834167.1 |Severe acute respiratory syndrome coro ···················· ···················· ···················· ···················· ···················· ····················

| | | | | | | | | |
|---|---|---|---|---|---|---|---|---|
| 23737 | MT834168.1 | \|Severe acute respiratory syndrome coro | ···················· | ···················· | ···················· | ···················· | ···················· | ···················· |
| 23738 | MT834169.1 | \|Severe acute respiratory syndrome coro | ···················· | ···················· | ···················· | ···················· | ···················· | ···················· |
| 23739 | MT834170.1 | \|Severe acute respiratory syndrome coro | ···················· | ···················· | ···················· | ···················· | ···················· | ···················· |
| 23740 | MT834171.1 | \|Severe acute respiratory syndrome coro | ···················· | ···················· | ···················· | ···················· | ··C················· | ···················· |
| 23741 | MT834172.1 | \|Severe acute respiratory syndrome coro | ···················· | ···················· | ···················· | ···················· | ···················· | ···················· |
| 23742 | MT834173.1 | \|Severe acute respiratory syndrome coro | ···················· | ···················· | ···················· | ···················· | ···················· | ···················· |
| 23743 | MT834174.1 | \|Severe acute respiratory syndrome coro | ···················· | ···················· | ···················· | ···················· | ···················· | ···················· |
| 23744 | MT834175.1 | \|Severe acute respiratory syndrome coro | ···················· | ···················· | ···················· | ···················· | ···················· | ···················· |
| 23745 | MT834176.1 | \|Severe acute respiratory syndrome coro | ···················· | ···················· | ···················· | ···················· | ···················· | ···················· |
| 23746 | MT834177.1 | \|Severe acute respiratory syndrome coro | ···················· | ···················· | ···················· | ···················· | ···················· | ···················· |
| 23747 | MT834178.1 | \|Severe acute respiratory syndrome coro | ···················· | ···················· | ···················· | ···················· | ···················· | ···················· |
| 23748 | MT834180.1 | \|Severe acute respiratory syndrome coro | ···················· | ···················· | ···················· | ···················· | ···················· | ···················· |
| 23749 | MT834181.1 | \|Severe acute respiratory syndrome coro | ···················· | ···················· | ···················· | ···················· | ···················· | ···················· |
| 23750 | MT834182.1 | \|Severe acute respiratory syndrome coro | ···················· | ···················· | ···················· | ···················· | ···················· | ···················· |
| 23751 | MT834183.1 | \|Severe acute respiratory syndrome coro | ···················· | ···················· | ···················· | ···················· | ···················· | ···················· |
| 23752 | MT834184.1 | \|Severe acute respiratory syndrome coro | ···················· | ···················· | ···················· | ···················· | ···················· | ···················· |
| 23753 | MT834185.1 | \|Severe acute respiratory syndrome coro | ···················· | ···················· | ···················· | ···················· | ···················· | ···················· |
| 23754 | MT834186.1 | \|Severe acute respiratory syndrome coro | ···················· | ···················· | ···················· | ···················· | ···················· | ···················· |
| 23755 | MT834187.1 | \|Severe acute respiratory syndrome coro | ···················· | ···················· | ···················· | ···················· | ···················· | ···················· |
| 23756 | MT834188.1 | \|Severe acute respiratory syndrome coro | ···················· | ···················· | ···················· | ···················· | ···················· | ···················· |
| 23757 | MT834189.1 | \|Severe acute respiratory syndrome coro | ···················· | ···················· | ···················· | ···················· | ···················· | ···················· |
| 23758 | MT834190.1 | \|Severe acute respiratory syndrome coro | ···················· | ···················· | ···················· | ···················· | ···················· | ···················· |
| 23759 | MT834191.1 | \|Severe acute respiratory syndrome coro | ···················· | ···················· | ···················· | ···················· | ···················· | ···················· |
| 23760 | MT834192.1 | \|Severe acute respiratory syndrome coro | ···················· | ···················· | ···················· | ···················· | ···················· | ···················· |
| 23761 | MT834193.1 | \|Severe acute respiratory syndrome coro | ···················· | ···················· | ···················· | ···················· | ···················· | ···················· |
| 23762 | MT834195.1 | \|Severe acute respiratory syndrome coro | ···················· | ···················· | ···················· | ···················· | ···················· | ···················· |
| 23763 | MT834196.1 | \|Severe acute respiratory syndrome coro | ···················· | ···················· | ···················· | ···················· | ···················· | ···················· |
| 23764 | MT834197.1 | \|Severe acute respiratory syndrome coro | ···················· | ···················· | ···················· | ···················· | ···················· | ···················· |
| 23765 | MT834198.1 | \|Severe acute respiratory syndrome coro | ···················· | ···················· | ···················· | ···················· | ···················· | ···················· |
| 23766 | MT834199.1 | \|Severe acute respiratory syndrome coro | ···················· | ···················· | ···················· | ···················· | ···················· | ···················· |
| 23767 | MT834201.1 | \|Severe acute respiratory syndrome coro | ···················· | ···················· | ···················· | ···················· | ···················· | ···················· |
| 23768 | MT834203.1 | \|Severe acute respiratory syndrome coro | ···················· | ···················· | ···················· | ···················· | ···················· | ···················· |
| 23769 | MT834204.1 | \|Severe acute respiratory syndrome coro | ·T·················· | ···················· | ···················· | ···················· | ···················· | ···················· |
| 23770 | MT834206.1 | \|Severe acute respiratory syndrome coro | ···················· | ···················· | ···················· | ···················· | ···················· | ···················· |
| 23771 | MT834207.1 | \|Severe acute respiratory syndrome coro | ···················· | ···················· | ···················· | ···················· | ···················· | ···················· |
| 23772 | MT834208.1 | \|Severe acute respiratory syndrome coro | ···················· | ···················· | ···················· | ···················· | ···················· | ···················· |
| 23773 | MT834209.1 | \|Severe acute respiratory syndrome coro | ···················· | ···················· | ···················· | ···················· | ···················· | ···················· |
| 23774 | MT834210.1 | \|Severe acute respiratory syndrome coro | ···················· | ···················· | ···················· | ···················· | ···················· | ···················· |
| 23775 | MT834211.1 | \|Severe acute respiratory syndrome coro | ···················· | ···················· | ···················· | ···················· | ···················· | ···················· |
| 23776 | MT834212.1 | \|Severe acute respiratory syndrome coro | ···················· | ···················· | ···················· | ···················· | ···················· | ···················· |
| 23777 | MT834213.1 | \|Severe acute respiratory syndrome coro | ···················· | ···················· | ···················· | ···················· | ···················· | ···················· |
| 23778 | MT834214.1 | \|Severe acute respiratory syndrome coro | ···················· | ···················· | ···················· | ···················· | ···················· | ···················· |
| 23779 | MT834216.1 | \|Severe acute respiratory syndrome coro | ···················· | ···················· | ···················· | ···················· | ···················· | ···················· |
| 23780 | MT834217.1 | \|Severe acute respiratory syndrome coro | ···················· | ···················· | ···················· | ···················· | ···················· | ···················· |
| 23781 | MT834218.1 | \|Severe acute respiratory syndrome coro | ···················· | ···················· | ···················· | ···················· | ···················· | ···················· |
| 23782 | MT834219.1 | \|Severe acute respiratory syndrome coro | ···················· | ···················· | ···················· | ···················· | ···················· | ···················· |
| 23783 | MT834220.1 | \|Severe acute respiratory syndrome coro | ···················· | ···················· | ···················· | ···················· | ···················· | ···················· |
| 23784 | MT834221.1 | \|Severe acute respiratory syndrome coro | ···················· | ···················· | ···················· | ···················· | ···················· | ···················· |
| 23785 | MT834222.1 | \|Severe acute respiratory syndrome coro | ···················· | ···················· | ···················· | ···················· | ···················· | ···················· |
| 23786 | MT834223.1 | \|Severe acute respiratory syndrome coro | ···················· | ···················· | ···················· | ···················· | ···················· | ···················· |
| 23787 | MT834224.1 | \|Severe acute respiratory syndrome coro | ···················· | ···················· | ···················· | ···················· | ···················· | ···················· |
| 23788 | MT834225.1 | \|Severe acute respiratory syndrome coro | ···················· | ···················· | ···················· | ···················· | ···················· | ···················· |
| 23789 | MT834226.1 | \|Severe acute respiratory syndrome coro | ···················· | ···················· | ···················· | ···················· | ···················· | ···················· |
| 23790 | MT834227.1 | \|Severe acute respiratory syndrome coro | ···················· | ···················· | ···················· | ···················· | ···················· | ···················· |
| 23791 | MT834228.1 | \|Severe acute respiratory syndrome coro | ···················· | ···················· | ···················· | ···················· | ···················· | ···················· |
| 23792 | MT834229.1 | \|Severe acute respiratory syndrome coro | ···················· | ···················· | ···················· | ···················· | ···················· | ···················· |
| 23793 | MT834230.1 | \|Severe acute respiratory syndrome coro | ···················· | ···················· | ···················· | ···················· | ···················· | ···················· |
| 23794 | MT834231.1 | \|Severe acute respiratory syndrome coro | ···················· | ···················· | ···················· | ···················· | ···················· | ···················· |
| 23795 | MT834232.1 | \|Severe acute respiratory syndrome coro | ···················· | ···················· | ···················· | ···················· | ···················· | ···················· |
| 23796 | MT834233.1 | \|Severe acute respiratory syndrome coro | ···················· | ···················· | ···················· | ···················· | ···················· | ···················· |
| 23797 | MT834234.1 | \|Severe acute respiratory syndrome coro | ···················· | ···················· | ···················· | ···················· | ···················· | ···················· |
| 23798 | MT834235.1 | \|Severe acute respiratory syndrome coro | ···················· | ···················· | ···················· | ···················· | ···················· | ···················· |
| 23799 | MT834236.1 | \|Severe acute respiratory syndrome coro | ···················· | ···················· | ···················· | ···················· | ···················· | ···················· |
| 23800 | MT834237.1 | \|Severe acute respiratory syndrome coro | ···················· | ···················· | ···················· | ···················· | ···················· | ···················· |
| 23801 | MT834238.1 | \|Severe acute respiratory syndrome coro | ···················· | ···················· | ···················· | ···················· | ···················· | ···················· |
| 23802 | MT834239.1 | \|Severe acute respiratory syndrome coro | ···················· | ···················· | ···················· | ···················· | ···················· | ···················· |
| 23803 | MT834241.1 | \|Severe acute respiratory syndrome coro | ···················· | ···················· | ···················· | ···················· | ···················· | ···················· |
| 23804 | MT834242.1 | \|Severe acute respiratory syndrome coro | ···················· | ···················· | ···················· | ···················· | ···················· | ···················· |
| 23805 | MT834243.1 | \|Severe acute respiratory syndrome coro | ···················· | ···················· | ···················· | ···················· | ···················· | ···················· |
| 23806 | MT834244.1 | \|Severe acute respiratory syndrome coro | ···················· | ···················· | ···················· | ···················· | ···················· | ···················· |
| 23807 | MT834245.1 | \|Severe acute respiratory syndrome coro | ···················· | ···················· | ···················· | ···················· | ···················· | ···················· |
| 23808 | MT834246.1 | \|Severe acute respiratory syndrome coro | ···················· | ···················· | ···················· | ···················· | ···················· | ···················· |
| 23809 | MT834247.1 | \|Severe acute respiratory syndrome coro | ···················· | ···················· | ···················· | ···················· | ···················· | ···················· |
| 23810 | MT834249.1 | \|Severe acute respiratory syndrome coro | ···················· | ···················· | ···················· | ···················· | ···················· | ···················· |
| 23811 | MT834250.1 | \|Severe acute respiratory syndrome coro | ···················· | ···················· | ···················· | ···················· | ···················· | ···················· |
| 23812 | MT834251.1 | \|Severe acute respiratory syndrome coro | ···················· | ···················· | ···················· | ···················· | ···················· | ···················· |
| 23813 | MT834252.1 | \|Severe acute respiratory syndrome coro | ···················· | ···················· | ···················· | ···················· | ···················· | ···················· |
| 23814 | MT834253.1 | \|Severe acute respiratory syndrome coro | ···················· | ···················· | ···················· | ···················· | ···················· | ···················· |

| | | | | | | | |
|---|---|---|---|---|---|---|---|
| 23815 | MT834254.1 \|Severe acute respiratory syndrome coro | .................... | .................... | .................... | .................... | .................... | .................... |
| 23816 | MT834255.1 \|Severe acute respiratory syndrome coro | .................... | .................... | .................... | .................... | .................... | .................... |
| 23817 | MT834256.1 \|Severe acute respiratory syndrome coro | .................... | .................... | .................... | .................... | .................... | .................... |
| 23818 | MT834258.1 \|Severe acute respiratory syndrome coro | .................... | .................... | .................... | .................... | .................... | .................... |
| 23819 | MT834259.1 \|Severe acute respiratory syndrome coro | .................... | .................... | .................... | .................... | .................... | .................... |
| 23820 | MT834260.1 \|Severe acute respiratory syndrome coro | .................... | .................... | .................... | .................... | .................... | .................... |
| 23821 | MT834261.1 \|Severe acute respiratory syndrome coro | .................... | .................... | .................... | .................... | .................... | .................... |
| 23822 | MT834262.1 \|Severe acute respiratory syndrome coro | .................... | .................... | .................... | .................... | .................... | .................... |
| 23823 | MT834263.1 \|Severe acute respiratory syndrome coro | .................... | .................... | .................... | .................... | .................... | .................... |
| 23824 | MT834264.1 \|Severe acute respiratory syndrome coro | .................... | .................... | .................... | .................... | .................... | .................... |
| 23825 | MT834265.1 \|Severe acute respiratory syndrome coro | .................... | .................... | .................... | .................... | .................... | .................... |
| 23826 | MT834266.1 \|Severe acute respiratory syndrome coro | .................... | .................... | .................... | .................... | .................... | .................... |
| 23827 | MT834267.1 \|Severe acute respiratory syndrome coro | .................... | .................... | .................... | .................... | .................... | .................... |
| 23828 | MT834268.1 \|Severe acute respiratory syndrome coro | .................... | .................... | .................... | .................... | .................... | .................... |
| 23829 | MT834269.1 \|Severe acute respiratory syndrome coro | .................... | .................... | .................... | .................... | .................... | .................... |
| 23830 | MT834270.1 \|Severe acute respiratory syndrome coro | .................... | .................... | .................... | .................... | .................... | .................... |
| 23831 | MT834271.1 \|Severe acute respiratory syndrome coro | .................... | .................... | .................... | .................... | .................... | .................... |
| 23832 | MT834272.1 \|Severe acute respiratory syndrome coro | .................... | .................... | .................... | .................... | .................... | .................... |
| 23833 | MT834273.1 \|Severe acute respiratory syndrome coro | .................... | .................... | .................... | .................... | .................... | .................... |
| 23834 | MT834274.1 \|Severe acute respiratory syndrome coro | .................... | .................... | .................... | .................... | .................... | .................... |
| 23835 | MT834275.1 \|Severe acute respiratory syndrome coro | .................... | .................... | .................... | .................... | .................... | .................... |
| 23836 | MT834276.1 \|Severe acute respiratory syndrome coro | .................... | .................... | .................... | .................... | .................... | .................... |
| 23837 | MT834277.1 \|Severe acute respiratory syndrome coro | .................... | .................... | .................... | .................... | .................... | .................... |
| 23838 | MT834278.1 \|Severe acute respiratory syndrome coro | .................... | .................... | .................... | .................... | .................... | .................... |
| 23839 | MT834280.1 \|Severe acute respiratory syndrome coro | .................... | .................... | .................... | .................... | .................... | .................... |
| 23840 | MT834281.1 \|Severe acute respiratory syndrome coro | .................... | .................... | .................... | .................... | .................... | .................... |
| 23841 | MT834282.1 \|Severe acute respiratory syndrome coro | .................... | .................... | .................... | .................... | .................... | .................... |
| 23842 | MT834284.1 \|Severe acute respiratory syndrome coro | .................... | .................... | .................... | .................... | .................... | .................... |
| 23843 | MT834285.1 \|Severe acute respiratory syndrome coro | .................... | .................... | .................... | .................... | .................... | .................... |
| 23844 | MT834286.1 \|Severe acute respiratory syndrome coro | .................... | .................... | .................... | .................... | .................... | .................... |
| 23845 | MT834287.1 \|Severe acute respiratory syndrome coro | .................... | .................... | .................... | .................... | .................... | .................... |
| 23846 | MT834288.1 \|Severe acute respiratory syndrome coro | .................... | .................... | .................... | .................... | .................... | .................... |
| 23847 | MT834289.1 \|Severe acute respiratory syndrome coro | .................... | .................... | .................... | .................... | .................... | .................... |
| 23848 | MT834290.1 \|Severe acute respiratory syndrome coro | .................... | .................... | .................... | .................... | .................... | .................... |
| 23849 | MT834291.1 \|Severe acute respiratory syndrome coro | .................... | .................... | .................... | .................... | .................... | .................... |
| 23850 | MT834292.1 \|Severe acute respiratory syndrome coro | .................... | .................... | .................... | .................... | .................... | .................... |
| 23851 | MT834293.1 \|Severe acute respiratory syndrome coro | .................... | .................... | .................... | .................... | .................... | .................... |
| 23852 | MT834295.1 \|Severe acute respiratory syndrome coro | .................... | .................... | .................... | .................... | .................... | .................... |
| 23853 | MT834296.1 \|Severe acute respiratory syndrome coro | .................... | .................... | .................... | .................... | .................... | .................... |
| 23854 | MT834297.1 \|Severe acute respiratory syndrome coro | .................... | .................... | .................... | .................... | .................... | .................... |
| 23855 | MT834298.1 \|Severe acute respiratory syndrome coro | .................... | .................... | .................... | .................... | .................... | .................... |
| 23856 | MT834299.1 \|Severe acute respiratory syndrome coro | .................... | .................... | .................... | .................... | .................... | .................... |
| 23857 | MT834300.1 \|Severe acute respiratory syndrome coro | .................... | .................... | .................... | .................... | .................... | .................... |
| 23858 | MT834301.1 \|Severe acute respiratory syndrome coro | .................... | .................... | .................... | .................... | .................... | .................... |
| 23859 | MT834302.1 \|Severe acute respiratory syndrome coro | .................... | .................... | .................... | .................... | .................... | .................... |
| 23860 | MT834303.1 \|Severe acute respiratory syndrome coro | .................... | .................... | .................... | .................... | .................... | .................... |
| 23861 | MT834304.1 \|Severe acute respiratory syndrome coro | .................... | .................... | .................... | .................... | .................... | .................... |
| 23862 | MT834305.1 \|Severe acute respiratory syndrome coro | .................... | .................... | .................... | .................... | .................... | .................... |
| 23863 | MT834306.1 \|Severe acute respiratory syndrome coro | .................... | .................... | .................... | .................... | .................... | .................... |
| 23864 | MT834307.1 \|Severe acute respiratory syndrome coro | .................... | .................... | .................... | .................... | .................... | .................... |
| 23865 | MT834308.1 \|Severe acute respiratory syndrome coro | .................... | .................... | .................... | .................... | .................... | .................... |
| 23866 | MT834309.1 \|Severe acute respiratory syndrome coro | .................... | .................... | .................... | .................... | .................... | .................... |
| 23867 | MT834310.1 \|Severe acute respiratory syndrome coro | .................... | .................... | .................... | .................... | .................... | .................... |
| 23868 | MT834311.1 \|Severe acute respiratory syndrome coro | .................... | .................... | .................... | .................... | .................... | .................... |
| 23869 | MT834312.1 \|Severe acute respiratory syndrome coro | .................... | .................... | .................... | .................... | .................... | .................... |
| 23870 | MT834313.1 \|Severe acute respiratory syndrome coro | .................... | .................... | .................... | .................... | .................... | .................... |
| 23871 | MT834315.1 \|Severe acute respiratory syndrome coro | .................... | .................... | .................... | .................... | .................... | .................... |
| 23872 | MT834316.1 \|Severe acute respiratory syndrome coro | .................... | .................... | .................... | .................... | .................... | .................... |
| 23873 | MT834317.1 \|Severe acute respiratory syndrome coro | .................... | .................... | .................... | .................... | .................... | .................... |
| 23874 | MT834318.1 \|Severe acute respiratory syndrome coro | .................... | .................... | .................... | .................... | .................... | .................... |
| 23875 | MT834319.1 \|Severe acute respiratory syndrome coro | .................... | .................... | .................... | .................... | .................... | .................... |
| 23876 | MT834320.1 \|Severe acute respiratory syndrome coro | .................... | .................... | .................... | .................... | .................... | .................... |
| 23877 | MT834321.1 \|Severe acute respiratory syndrome coro | .................... | .................... | .................... | .................... | .................... | .................... |
| 23878 | MT834322.1 \|Severe acute respiratory syndrome coro | .................... | .................... | .................... | .................... | .................... | .................... |
| 23879 | MT834323.1 \|Severe acute respiratory syndrome coro | .................... | C................... | .................... | .................... | .................... | .................... |
| 23880 | MT834324.1 \|Severe acute respiratory syndrome coro | .................... | .................... | .................... | .................... | .................... | .................... |
| 23881 | MT834325.1 \|Severe acute respiratory syndrome coro | .................... | .................... | .................... | .................... | .................... | .................... |
| 23882 | MT834326.1 \|Severe acute respiratory syndrome coro | .................... | .................... | .................... | .................... | .................... | .................... |
| 23883 | MT834327.1 \|Severe acute respiratory syndrome coro | .................... | .................... | .................... | .................... | .................... | .................... |
| 23884 | MT834328.1 \|Severe acute respiratory syndrome coro | .................... | .................... | .................... | .................... | .................... | .................... |
| 23885 | MT834329.1 \|Severe acute respiratory syndrome coro | .................... | .................... | .................... | .................... | .................... | .................... |
| 23886 | MT834330.1 \|Severe acute respiratory syndrome coro | .................... | .................... | .................... | .................... | .................... | .................... |
| 23887 | MT834331.1 \|Severe acute respiratory syndrome coro | .................... | .................... | .................... | .................... | .................... | .................... |
| 23888 | MT834335.1 \|Severe acute respiratory syndrome coro | .................... | .................... | .................... | .................... | .................... | .................... |
| 23889 | MT834336.1 \|Severe acute respiratory syndrome coro | .................... | .................... | .................... | .................... | .................... | .................... |
| 23890 | MT834337.1 \|Severe acute respiratory syndrome coro | .................... | .................... | .................... | .................... | .................... | .................... |
| 23891 | MT834338.1 \|Severe acute respiratory syndrome coro | .................... | .................... | .................... | .................... | .................... | .................... |
| 23892 | MT834339.1 \|Severe acute respiratory syndrome coro | .................... | .................... | .................... | .................... | .................... | .................... |

| | | | | | | | | |
|---|---|---|---|---|---|---|---|---|
| 23893 | MT834341.1 | Severe acute respiratory syndrome coro | ···················· | ···················· | ···················· | ···················· | ···················· | ···················· |
| 23894 | MT834343.1 | Severe acute respiratory syndrome coro | ···················· | ···················· | ···················· | ···················· | ···················· | ···················· |
| 23895 | MT834344.1 | Severe acute respiratory syndrome coro | ···················· | ···················· | ···················· | ···················· | ···················· | ···················· |
| 23896 | MT834345.1 | Severe acute respiratory syndrome coro | ···················· | ···················· | ···················· | ···················· | ···················· | ···················· |
| 23897 | MT834347.1 | Severe acute respiratory syndrome coro | ···················· | ···················· | ···················· | ···················· | ···················· | ···················· |
| 23898 | MT834348.1 | Severe acute respiratory syndrome coro | ···················· | ···················· | ···················· | ···················· | ···················· | ···················· |
| 23899 | MT834350.1 | Severe acute respiratory syndrome coro | ···················· | ···················· | ···················· | ···················· | ···················· | ···················· |
| 23900 | MT834352.1 | Severe acute respiratory syndrome coro | ···················· | ···················· | ···················· | ···················· | ···················· | ···················· |
| 23901 | MT834353.1 | Severe acute respiratory syndrome coro | ···················· | ···················· | ···················· | ···················· | ···················· | ···················· |
| 23902 | MT834354.1 | Severe acute respiratory syndrome coro | ···················· | ···················· | ···················· | ···················· | ···················· | ···················· |
| 23903 | MT834355.1 | Severe acute respiratory syndrome coro | ···················· | ···················· | ···················· | ···················· | ···················· | ···················· |
| 23904 | MT834356.1 | Severe acute respiratory syndrome coro | ···················· | ···················· | ···················· | ···················· | ···················· | ···················· |
| 23905 | MT834357.1 | Severe acute respiratory syndrome coro | ···················· | ···················· | ···················· | ···················· | ···················· | ···················· |
| 23906 | MT834358.1 | Severe acute respiratory syndrome coro | ···················· | ···················· | ···················· | ···················· | ···················· | ···················· |
| 23907 | MT834359.1 | Severe acute respiratory syndrome coro | ···················· | ···················· | ···················· | ···················· | ···················· | ···················· |
| 23908 | MT834360.1 | Severe acute respiratory syndrome coro | ···················· | ···················· | ···················· | ···················· | ···················· | ···················· |
| 23909 | MT834607.1 | Severe acute respiratory syndrome coro | ···················· | ···················· | ···················· | ···················· | ···················· | ···················· |
| 23910 | MT834608.1 | Severe acute respiratory syndrome coro | ···················· | ···················· | ···················· | ···················· | ···················· | ···················· |
| 23911 | MT834609.1 | Severe acute respiratory syndrome coro | ···················· | ···················· | ···················· | ···················· | ···················· | ···················· |
| 23912 | MT834611.1 | Severe acute respiratory syndrome coro | ···················· | ···················· | ···················· | ···················· | ···················· | ···················· |
| 23913 | MT834613.1 | Severe acute respiratory syndrome coro | ···················· | ···················· | ···················· | ···················· | ···················· | ···················· |
| 23914 | MT834614.1 | Severe acute respiratory syndrome coro | ···················· | ···················· | ···················· | ···················· | ···················· | ···················· |
| 23915 | MT834615.1 | Severe acute respiratory syndrome coro | ···················· | ···················· | ···················· | ···················· | ···················· | ···················· |
| 23917 | MT834617.1 | Severe acute respiratory syndrome coro | ···················· | ···················· | ···················· | ···················· | ···················· | ···················· |
| 23918 | MT834619.1 | Severe acute respiratory syndrome coro | ···················· | ···················· | ···················· | ···················· | ···················· | ···················· |
| 23919 | MT834620.1 | Severe acute respiratory syndrome coro | ···················· | ···················· | ···················· | ···················· | ···················· | ···················· |
| 23920 | MT834622.1 | Severe acute respiratory syndrome coro | ···················· | ···················· | ···················· | ···················· | ···················· | ···················· |
| 23921 | MT834623.1 | Severe acute respiratory syndrome coro | ···················· | ···················· | ···················· | ···················· | ···················· | ···················· |
| 23922 | MT834624.1 | Severe acute respiratory syndrome coro | ···················· | ···················· | ···················· | ···················· | ···················· | ···················· |
| 23923 | MT834625.1 | Severe acute respiratory syndrome coro | ···················· | ···················· | ···················· | ···················· | ···················· | ···················· |
| 23924 | MT834626.1 | Severe acute respiratory syndrome coro | ···················· | ···················· | ···················· | ···················· | ···················· | ···················· |
| 23925 | MT834628.1 | Severe acute respiratory syndrome coro | ···················· | ···················· | ···················· | ···················· | N··················· | ···················· |
| 23926 | MT834630.1 | Severe acute respiratory syndrome coro | ···················· | ···················· | ···················· | ···················· | ···················· | ···················· |
| 23927 | MT834631.1 | Severe acute respiratory syndrome coro | ···················· | ···················· | ···················· | ···················· | ···················· | ···················· |
| 23928 | MT834632.1 | Severe acute respiratory syndrome coro | ···················· | ···················· | ···················· | ···················· | ···················· | ···················· |
| 23929 | MT834634.1 | Severe acute respiratory syndrome coro | ···················· | ···················· | ···················· | ···················· | ···················· | ···················· |
| 23930 | MT834635.1 | Severe acute respiratory syndrome coro | ···················· | ···················· | ···················· | ···················· | ···················· | ···················· |
| 23931 | MT834636.1 | Severe acute respiratory syndrome coro | ···················· | ···················· | ···················· | ···················· | ···················· | ···················· |
| 23932 | MT834637.1 | Severe acute respiratory syndrome coro | ···················· | ·········A·········· | ···················· | ···················· | ···················· | ···················· |
| 23933 | MT834638.1 | Severe acute respiratory syndrome coro | ···················· | ···················· | ···················· | ···················· | ···················· | ···················· |
| 23934 | MT834639.1 | Severe acute respiratory syndrome coro | ···················· | ···················· | ···················· | ···················· | ···················· | ···················· |
| 23935 | MT834640.1 | Severe acute respiratory syndrome coro | ···················· | ···················· | ···················· | ···················· | ···················· | ···················· |
| 23936 | MT834641.1 | Severe acute respiratory syndrome coro | ···················· | ···················· | ···················· | ···················· | ···················· | ···················· |
| 23937 | MT834642.1 | Severe acute respiratory syndrome coro | ···················· | ···················· | ···················· | ···················· | ···················· | ···················· |
| 23938 | MT834643.1 | Severe acute respiratory syndrome coro | ···················· | ···················· | ···················· | ···················· | ···················· | ···················· |
| 23939 | MT834644.1 | Severe acute respiratory syndrome coro | ···················· | ···················· | ···················· | ···················· | ···················· | ···················· |
| 23940 | MT834645.1 | Severe acute respiratory syndrome coro | ···················· | ···················· | ···················· | ···················· | ···················· | ···················· |
| 23941 | MT834646.1 | Severe acute respiratory syndrome coro | ···················· | ···················· | ···················· | ···················· | ···················· | ···················· |
| 23942 | MT834647.1 | Severe acute respiratory syndrome coro | ···················· | ···················· | ···················· | ···················· | ···················· | ···················· |
| 23943 | MT834648.1 | Severe acute respiratory syndrome coro | ···················· | ···················· | ···················· | ···················· | ···················· | ···················· |
| 23944 | MT834649.1 | Severe acute respiratory syndrome coro | ···················· | ···················· | ···················· | ···················· | ···················· | ···················· |
| 23945 | MT834651.1 | Severe acute respiratory syndrome coro | ···················· | ···················· | ···················· | ···················· | ···················· | ···················· |
| 23946 | MT834652.1 | Severe acute respiratory syndrome coro | ···················· | ···················· | ···················· | ···················· | ···················· | ···················· |
| 23947 | MT834653.1 | Severe acute respiratory syndrome coro | ···················· | ···················· | ···················· | ···················· | ···················· | ···················· |
| 23948 | MT834656.1 | Severe acute respiratory syndrome coro | ···················· | ···················· | ···················· | ···················· | ···················· | ···················· |
| 23949 | MT834657.1 | Severe acute respiratory syndrome coro | ···················· | ···················· | ···················· | ···················· | ···················· | ···················· |
| 23950 | MT834660.1 | Severe acute respiratory syndrome coro | ···················· | ···················· | ···················· | ···················· | ···················· | ···················· |
| 23951 | MT834661.1 | Severe acute respiratory syndrome coro | ···················· | ···················· | ···················· | ···················· | ···················· | ···················· |
| 23952 | MT834662.1 | Severe acute respiratory syndrome coro | ···················· | ···················· | ···················· | ···················· | ···················· | ···················· |
| 23953 | MT834664.1 | Severe acute respiratory syndrome coro | ···················· | ···················· | ···················· | ···················· | ···················· | ···················· |
| 23954 | MT834667.1 | Severe acute respiratory syndrome coro | ···················· | ···················· | ···················· | ···················· | ···················· | ···················· |
| 23956 | MT834671.1 | Severe acute respiratory syndrome coro | ···················· | ···················· | ···················· | ···················· | ···················· | ···················· |
| 23957 | MT834672.1 | Severe acute respiratory syndrome coro | ···················· | ···················· | ···················· | ···················· | ···················· | ···················· |
| 23958 | MT834673.1 | Severe acute respiratory syndrome coro | ···················· | ···················· | ···················· | ···················· | ···················· | ···················· |
| 23959 | MT834674.1 | Severe acute respiratory syndrome coro | ···················· | ···················· | ···················· | ···················· | ···················· | ···················· |
| 23961 | MT834676.1 | Severe acute respiratory syndrome coro | ···················· | ···················· | ···················· | ···················· | ···················· | ···················· |
| 23962 | MT834678.1 | Severe acute respiratory syndrome coro | ···················· | ···················· | ···················· | ···················· | ···················· | ···················· |
| 23963 | MT834679.1 | Severe acute respiratory syndrome coro | ···················· | ···················· | ···················· | ···················· | ···················· | ···················· |
| 23964 | MT834680.1 | Severe acute respiratory syndrome coro | ···················· | ···················· | ···················· | ···················· | ···················· | ···················· |
| 23965 | MT834681.1 | Severe acute respiratory syndrome coro | ···················· | ···················· | ···················· | ···················· | ···················· | ···················· |
| 23966 | MT834682.1 | Severe acute respiratory syndrome coro | ···················· | ···················· | ···················· | ···················· | ···················· | ···················· |
| 23967 | MT834684.1 | Severe acute respiratory syndrome coro | ···················· | ···················· | ···················· | ···················· | ···················· | ···················· |
| 23968 | MT834685.1 | Severe acute respiratory syndrome coro | ···················· | ···················· | ···················· | ···················· | ···················· | ···················· |
| 23969 | MT834686.1 | Severe acute respiratory syndrome coro | ···················· | ···················· | ···················· | ···················· | ···················· | ···················· |
| 23970 | MT834687.1 | Severe acute respiratory syndrome coro | ···················· | ···················· | ···················· | ···················· | ···················· | ···················· |
| 23971 | MT834688.1 | Severe acute respiratory syndrome coro | ···················· | ···················· | ···················· | ···················· | ···················· | ···················· |
| 23972 | MT834689.1 | Severe acute respiratory syndrome coro | ···················· | ·············T······ | ···················· | ···················· | ···················· | ···················· |
| 23973 | MT834690.1 | Severe acute respiratory syndrome coro | ···················· | ···················· | ···················· | ···················· | ···················· | ···················· |

```
23974 MT834691.1 |Severe acute respiratory syndrome coro    ....................  ....................  .....................  ....................  ....................  ....................
23975 MT834692.1 |Severe acute respiratory syndrome coro    ....................  ....................  .....................  ....................  ....................  ....................
23976 MT834693.1 |Severe acute respiratory syndrome coro    ....................  ....................  .....................  ....................  ....................  ....................
23977 MT834694.1 |Severe acute respiratory syndrome coro    ....................  ....................  .....................  ....................  ....................  ....................
23978 MT834695.1 |Severe acute respiratory syndrome coro    ....................  ....................  .....................  ....................  ....................  ....................
23979 MT834696.1 |Severe acute respiratory syndrome coro    ....................  ....................  .....................  ....................  ....................  ....................
23980 MT834697.1 |Severe acute respiratory syndrome coro    ....................  ....................  .....................  ....................  ....................  ....................
23981 MT834699.1 |Severe acute respiratory syndrome coro    ....................  ....................  .....................  ....................  ....................  ....................
23982 MT834715.1 |Severe acute respiratory syndrome coro    ....................  ....................  .....................  ....................  ....................  ....................
23984 MT834731.1 |Severe acute respiratory syndrome coro    ....................  ....................  .....................  ....................  ....................  ....................
23985 MT834733.1 |Severe acute respiratory syndrome coro    ....................  ....................  .....................  ....................  ....................  ....................
23986 MT834734.1 |Severe acute respiratory syndrome coro    ....................  ....................  .....................  ....................  ....................  ....................
23987 MT834735.1 |Severe acute respiratory syndrome coro    ....................  ....................  .....................  ....................  ....................  ....................
23988 MT834849.1 |Severe acute respiratory syndrome coro    ....................  ....................  .....................  ....................  ....................  ....................
23989 MT834850.1 |Severe acute respiratory syndrome coro    ....................  ....................  .....................  ....................  ....................  ....................
23990 MT835026.1 |Severe acute respiratory syndrome coro    ....................  ....................  .....................  ....................  ....................  ....................
23991 MT835027.1 |Severe acute respiratory syndrome coro    ....................  ....................  .....................  ....................  ....................  ....................
23992 MT704816.1 |Severe acute respiratory syndrome coro    ....................  ....................  .....................  ....................  ....................  ....................
23993 MT704817.1 |Severe acute respiratory syndrome coro    ....................  ....................  .....................  ....................  ....................  ....................
23994 MT704818.1 |Severe acute respiratory syndrome coro    ....................  ....................  .....................  ....................  ....................  ....................
23995 MT704819.1 |Severe acute respiratory syndrome coro    ....................  ....................  .....................  ....................  ....................  ....................
23996 MT704820.1 |Severe acute respiratory syndrome coro    ....................  ....................  .....................  ....................  ....................  ....................
23997 MT704821.1 |Severe acute respiratory syndrome coro    ....................  ....................  .....................  ....................  ....................  ....................
23998 MT704822.1 |Severe acute respiratory syndrome coro    ....................  ....................  .....................  ....................  ....................  ....................
23999 MT704823.1 |Severe acute respiratory syndrome coro    ....................  ....................  .....................  ....................  ....................  ....................
24000 MT704824.1 |Severe acute respiratory syndrome coro    ....................  ....................  .....................  ....................  ....................  ....................
24001 MT704825.1 |Severe acute respiratory syndrome coro    ....................  ....................  .....................  ....................  ....................  ....................
24002 MT827074.1 |Severe acute respiratory syndrome coro    ....................  ....................  .....................  ....................  ....................  ....................
24003 MT827075.1 |Severe acute respiratory syndrome coro    ....................  ....................  .....................  ....................  ....................  ....................
24004 MT827190.1 |Severe acute respiratory syndrome coro    ....................  ....................  .....................  ....................  ....................  ....................
24005 MT827202.1 |Severe acute respiratory syndrome coro    ....................  ....................  .....................  ....................  ....................  ....................
24006 MT827203.1 |Severe acute respiratory syndrome coro    ....................  ....................  .....................  ....................  ....................  ....................
24007 MT827206.1 |Severe acute respiratory syndrome coro    ....................  ....................  .....................  ....................  ....................  ....................
24008 MT827207.1 |Severe acute respiratory syndrome coro    ....................  ....................  .....................  ....................  ....................  ....................
24009 MT827208.1 |Severe acute respiratory syndrome coro    ....................  ....................  .....................  ....................  ....................  ....................
24010 MT827209.1 |Severe acute respiratory syndrome coro    ....................  ....................  .....................  ....................  ....................  ....................
24011 MT827210.1 |Severe acute respiratory syndrome coro    ....................  ....................  .....................  ....................  ....................  ....................
24012 MT827211.1 |Severe acute respiratory syndrome coro    ....................  ....................  .....................  ....................  ....................  ....................
24013 MT827212.1 |Severe acute respiratory syndrome coro    ....................  ....................  .....................  ....................  ....................  ....................
24014 MT827213.1 |Severe acute respiratory syndrome coro    ....................  ....................  .....................  ....................  ....................  ....................
24015 MT827214.1 |Severe acute respiratory syndrome coro    ....................  ....................  .....................  ....................  ....................  ....................
24016 MT827215.1 |Severe acute respiratory syndrome coro    ....................  ....................  .....................  ....................  ....................  ....................
24017 MT827216.1 |Severe acute respiratory syndrome coro    ....................  ....................  .....................  ....................  ....................  ....................
24018 MT827217.1 |Severe acute respiratory syndrome coro    ....................  ....................  .....................  ....................  ....................  ....................
24019 MT827218.1 |Severe acute respiratory syndrome coro    ....................  ....................  .....................  ....................  ....................  ....................
24020 MT827219.1 |Severe acute respiratory syndrome coro    ....................  ....................  .....................  ....................  ....................  ....................
24021 MT827220.1 |Severe acute respiratory syndrome coro    ....................  ....................  .....................  ....................  ....................  ....................
24022 MT827221.1 |Severe acute respiratory syndrome coro    ....................  ....................  .....................  ....................  ....................  ....................
24023 MT827222.1 |Severe acute respiratory syndrome coro    ....................  ....................  .....................  ....................  ....................  ....................
24024 MT827223.1 |Severe acute respiratory syndrome coro    ....................  ....................  .....................  ....................  ....................  ....................
24025 MT827224.1 |Severe acute respiratory syndrome coro    ....................  ....................  .....................  ....................  ....................  ....................
24026 MT827225.1 |Severe acute respiratory syndrome coro    ....................  ....................  .....................  ....................  ....................  ....................
24027 MT827226.1 |Severe acute respiratory syndrome coro    ....................  ....................  .....................  ....................  ....................  ....................
24028 MT827227.1 |Severe acute respiratory syndrome coro    ....................  ....................  .....................  ....................  ....................  ....................
24029 MT827228.1 |Severe acute respiratory syndrome coro    ....................  ....................  .....................  ....................  ....................  ....................
24030 MT827229.1 |Severe acute respiratory syndrome coro    ....................  ....................  .....................  ....................  ....................  ....................
24031 MT827230.1 |Severe acute respiratory syndrome coro    ....................  ....................  .....................  ....................  ....................  ....................
24032 MT827231.1 |Severe acute respiratory syndrome coro    ....................  ....................  .....................  ....................  ....................  ....................
24033 MT827232.1 |Severe acute respiratory syndrome coro    ....................  ....................  .....................  ....................  ....................  ....................
24034 MT827233.1 |Severe acute respiratory syndrome coro    ....................  ....................  .....................  ....................  ....................  ....................
24035 MT827234.1 |Severe acute respiratory syndrome coro    ....................  ....................  .....................  ....................  ....................  ....................
24036 MT827235.1 |Severe acute respiratory syndrome coro    ....................  ....................  .....................  ....................  ....................  ....................
24037 MT827236.1 |Severe acute respiratory syndrome coro    ....................  ....................  .....................  ....................  ....................  ....................
24038 MT827237.1 |Severe acute respiratory syndrome coro    ....................  ....................  .....................  ....................  ....................  ....................
24039 MT827238.1 |Severe acute respiratory syndrome coro    ....................  ....................  .....................  ....................  ....................  ....................
24040 MT827239.1 |Severe acute respiratory syndrome coro    ....................  ....................  .....................  ....................  ....................  ....................
24041 MT827240.1 |Severe acute respiratory syndrome coro    ....................  ....................  .....................  ....................  ....................  ....................
24042 MT827241.1 |Severe acute respiratory syndrome coro    ....................  ....................  .....................  ....................  ....................  ....................
24043 MT827242.1 |Severe acute respiratory syndrome coro    ....................  ....................  .....................  ....................  ....................  ....................
24044 MT827243.1 |Severe acute respiratory syndrome coro    ....................  ....................  .....................  ....................  ....................  ....................
24045 MT827244.1 |Severe acute respiratory syndrome coro    ....................  ....................  .....................  ....................  ....................  ....................
24046 MT827245.1 |Severe acute respiratory syndrome coro    ....................  ....................  .....................  ....................  ....................  ....................
24047 MT827246.1 |Severe acute respiratory syndrome coro    ....................  ....................  .....................  ....................  ....................  ....................
24048 MT827247.1 |Severe acute respiratory syndrome coro    ....................  ....................  .....................  ....................  ....................  ....................
24049 MT827248.1 |Severe acute respiratory syndrome coro    ....................  ....................  .....................  ....................  ....................  ....................
24050 MT827249.1 |Severe acute respiratory syndrome coro    ....................  ....................  .....................  ....................  ....................  ....................
24051 MT827250.1 |Severe acute respiratory syndrome coro    ....................  ....................  .....................  ....................  ....................  ....................
24052 MT827251.1 |Severe acute respiratory syndrome coro    ....................  ....................  .....................  ....................  ....................  ....................
```

24053 MT827252.1 |Severe acute respiratory syndrome coro ···················· ···················· ···················· ···················· ···················· ····················
24054 MT827253.1 |Severe acute respiratory syndrome coro ···················· ···················· ···················· ···················· ···················· ····················
24055 MT827809.1 |Severe acute respiratory syndrome coro ···················· ···················· ···················· ···················· ···················· ····················
24056 MT827810.1 |Severe acute respiratory syndrome coro ···················· ···················· ···················· ···················· ···················· ····················
24057 MT827811.1 |Severe acute respiratory syndrome coro ···················· ···················· ···················· ···················· ···················· ····················
24058 MT827812.1 |Severe acute respiratory syndrome coro ···················· ···················· ···················· ···················· ···················· ····················
24059 MT827813.1 |Severe acute respiratory syndrome coro ···················· ···················· ···················· ···················· ···················· ····················
24060 MT827815.1 |Severe acute respiratory syndrome coro ···················· ···················· ···················· ···················· ···················· ····················
24061 MT827816.1 |Severe acute respiratory syndrome coro ···················· ···················· ···················· ···················· ···················· ····················
24062 MT827817.1 |Severe acute respiratory syndrome coro ···················· ···················· ···················· ···················· ···················· ····················
24063 MT827818.1 |Severe acute respiratory syndrome coro ···················· ···················· ···················· ···················· ···················· ····················
24064 MT827819.1 |Severe acute respiratory syndrome coro ···················· ···················· ···················· ···················· ···················· ····················
24065 MT827820.1 |Severe acute respiratory syndrome coro ···················· ···················· ···················· ···················· ···················· ····················
24066 MT827821.1 |Severe acute respiratory syndrome coro ···················· ···················· ···················· ···················· ···················· ····················
24067 MT827822.1 |Severe acute respiratory syndrome coro ···················· ···················· ···················· ···················· ···················· ····················
24068 MT827823.1 |Severe acute respiratory syndrome coro ···················· ···················· ···················· ···················· ···················· ····················
24069 MT827824.1 |Severe acute respiratory syndrome coro ···················· ···················· ···················· ···················· ···················· ····················
24070 MT827825.1 |Severe acute respiratory syndrome coro ···················· ···················· ···················· ···················· ···················· ····················
24071 MT827826.1 |Severe acute respiratory syndrome coro ···················· ···················· ···················· ···················· ···················· ····················
24072 MT827827.1 |Severe acute respiratory syndrome coro ···················· ···················· ···················· ···················· ···················· ····················
24073 MT827828.1 |Severe acute respiratory syndrome coro ···················· ···················· ···················· ···················· ···················· ····················
24074 MT827829.1 |Severe acute respiratory syndrome coro ···················· ···················· ···················· ···················· ···················· ····················
24075 MT827831.1 |Severe acute respiratory syndrome coro ···················· ···················· ··········T········· ···················· ···················· ····················
24076 MT827832.1 |Severe acute respiratory syndrome coro ···················· ···················· ···················· ···················· ···················· ····················
24077 MT827833.1 |Severe acute respiratory syndrome coro ···················· ···················· ···················· ···················· ···················· ····················
24078 MT827834.1 |Severe acute respiratory syndrome coro ···················· ···················· ···················· ···················· ···················· ····················
24079 MT827835.1 |Severe acute respiratory syndrome coro ···················· ···················· ···················· ···················· ···················· ····················
24080 MT827836.1 |Severe acute respiratory syndrome coro ···················· ···················· ···················· ···················· ···················· ····················
24081 MT827837.1 |Severe acute respiratory syndrome coro ···················· ···················· ···················· ···················· ···················· ····················
24082 MT827839.1 |Severe acute respiratory syndrome coro ···················· ···················· ···················· ···················· ···················· ····················
24083 MT827840.1 |Severe acute respiratory syndrome coro ···················· ···················· ···················· ···················· ···················· ····················
24084 MT827841.1 |Severe acute respiratory syndrome coro ···················· ···················· ···················· ···················· ···················· ····················
24085 MT827842.1 |Severe acute respiratory syndrome coro ···················· ···················· ···················· ···················· ···················· ····················
24086 MT827843.1 |Severe acute respiratory syndrome coro ···················· ···················· ···················· ···················· ···················· ····················
24087 MT827845.1 |Severe acute respiratory syndrome coro ···················· ···················· ···················· ···················· ···················· ····················
24088 MT827872.1 |Severe acute respiratory syndrome coro ···················· ···················· ···················· ···················· ···················· ····················
24089 MT827940.1 |Severe acute respiratory syndrome coro ···················· ···················· ···················· ···················· ···················· ····················
24090 MT821531.1 |Severe acute respiratory syndrome coro ···················· ···················· ···················· ···················· ···················· ····················
24091 MT821532.1 |Severe acute respiratory syndrome coro ···················· ···················· ···················· ···················· ···················· ····················
24092 MT821533.1 |Severe acute respiratory syndrome coro ···················· ···················· ···················· ···················· ···················· ····················
24093 MT821535.1 |Severe acute respiratory syndrome coro ···················· ···················· ···················· ···················· ···················· ····················
24094 MT821536.1 |Severe acute respiratory syndrome coro ···················· ···················· ···················· ···················· ···················· ····················
24095 MT821537.1 |Severe acute respiratory syndrome coro ···················· ···················· ···················· ···················· ···················· ····················
24096 MT821538.1 |Severe acute respiratory syndrome coro ···················· ···················· ···················· ···················· ···················· ····················
24097 MT821539.1 |Severe acute respiratory syndrome coro ···················· ···················· ···················· ···················· ············T······· ····················
24098 MT821540.1 |Severe acute respiratory syndrome coro ···················· ···················· ···················· ···················· ···················· ····················
24099 MT821541.1 |Severe acute respiratory syndrome coro ···················· ···················· ···················· ···················· ···················· ····················
24100 MT821542.1 |Severe acute respiratory syndrome coro ···················· ···················· ···················· ···················· ···················· ····················
24101 MT821543.1 |Severe acute respiratory syndrome coro ···················· ···················· ···················· ···················· ···················· ····················
24102 MT821545.1 |Severe acute respiratory syndrome coro ···················· ···················· ···················· ···················· ···················· ····················
24103 MT821546.1 |Severe acute respiratory syndrome coro ···················· ···················· ···················· ···················· ···················· ····················
24104 MT821547.1 |Severe acute respiratory syndrome coro ···················· ···················· ···················· ···················· ···················· ····················
24105 MT821548.1 |Severe acute respiratory syndrome coro ···················· ···················· ···················· ···················· ···················· ····················
24106 MT821549.1 |Severe acute respiratory syndrome coro ···················· ···················· ···················· ···················· ···················· ····················
24107 MT821550.1 |Severe acute respiratory syndrome coro ···················· ···················· ···················· ···················· ···················· ····················
24108 MT821551.1 |Severe acute respiratory syndrome coro ···················· ···················· ···················· ···················· ···················· ····················
24109 MT821552.1 |Severe acute respiratory syndrome coro ···················· ···················· ···················· ···················· ···················· ····················
24110 MT821553.1 |Severe acute respiratory syndrome coro ···················· ···················· ···················· ···················· ···················· ····················
24111 MT821554.1 |Severe acute respiratory syndrome coro ···················· ···················· ···················· ···················· ···················· ····················
24112 MT821555.1 |Severe acute respiratory syndrome coro ···················· ···················· ···················· ···················· ···················· ····················
24113 MT821556.1 |Severe acute respiratory syndrome coro ···················· ···················· ···················· ···················· ···················· ····················
24114 MT821557.1 |Severe acute respiratory syndrome coro ···················· ···················· ···················· ···················· ···················· ····················
24115 MT821558.1 |Severe acute respiratory syndrome coro ···················· ···················· ···················· ···················· ···················· ····················
24116 MT821559.1 |Severe acute respiratory syndrome coro ···················· ···················· ···················· ···················· ···················· ····················
24117 MT821560.1 |Severe acute respiratory syndrome coro ···················· ···················· ···················· ···················· ···················· ····················
24118 MT821561.1 |Severe acute respiratory syndrome coro ···················· ···················· ···················· ···················· ···················· ····················
24119 MT821562.1 |Severe acute respiratory syndrome coro ···················· ···················· ···················· ···················· ···················· ····················
24120 MT821563.1 |Severe acute respiratory syndrome coro ···················· ···················· ···················· ···················· ···················· ····················
24121 MT821564.1 |Severe acute respiratory syndrome coro ···················· ···················· ···················· ···················· ···················· ····················
24122 MT821565.1 |Severe acute respiratory syndrome coro ···················· ···················· ···················· ···················· ···················· ····················
24123 MT821566.1 |Severe acute respiratory syndrome coro ···················· ···················· ···················· ···················· ···················· ····················
24124 MT821567.1 |Severe acute respiratory syndrome coro ···················· ···················· ···················· ···················· ···················· ····················
24125 MT821568.1 |Severe acute respiratory syndrome coro ···················· ···················· ···················· ···················· ···················· ····················
24126 MT821569.1 |Severe acute respiratory syndrome coro ···················· ···················· ···················· ···················· ···················· ····················
24127 MT821570.1 |Severe acute respiratory syndrome coro ···················· ·A·················· ···················· ···················· ···················· ····················
24128 MT821571.1 |Severe acute respiratory syndrome coro ···················· ···················· ···················· ···················· ···················· ····················
24129 MT821572.1 |Severe acute respiratory syndrome coro ···················· ···················· ···················· ···················· ···················· ····················
24130 MT821573.1 |Severe acute respiratory syndrome coro ···················· ···················· ···················· ···················· ···················· ····················

```
24131 MT821574.1 |Severe acute respiratory syndrome coro    ....................  ....................  .....................  ....................  ....................  ....................
24132 MT821575.1 |Severe acute respiratory syndrome coro    ....................  ....................  .....................  ....................  ....................  ....................
24133 MT821577.1 |Severe acute respiratory syndrome coro    ....................  ....................  .....................  ....................  ....................  ....................
24134 MT821578.1 |Severe acute respiratory syndrome coro    ....................  ....................  .....................  ....................  ....................  ....................
24135 MT821579.1 |Severe acute respiratory syndrome coro    ....................  ....................  .....................  ....................  ....................  ....................
24136 MT821580.1 |Severe acute respiratory syndrome coro    ....................  ....................  .....................  ....................  ....................  ....................
24137 MT821581.1 |Severe acute respiratory syndrome coro    ....................  ....................  .....................  ....................  ....................  ....................
24138 MT821582.1 |Severe acute respiratory syndrome coro    ....................  ....................  .....................  ....................  ....................  ....................
24139 MT821584.1 |Severe acute respiratory syndrome coro    ....................  ....................  .....................  ....................  ....................  ....................
24140 MT821585.1 |Severe acute respiratory syndrome coro    ....................  ....................  .....................  ....................  ....................  ....................
24141 MT821586.1 |Severe acute respiratory syndrome coro    ....................  ....................  .....................  ....................  ....................  ....................
24142 MT821587.1 |Severe acute respiratory syndrome coro    ....................  ....................  .....................  ....................  ....................  ....................
24143 MT821588.1 |Severe acute respiratory syndrome coro    ....................  ....................  .....................  ....................  ....................  ....................
24144 MT821589.1 |Severe acute respiratory syndrome coro    ....................  ....................  .....................  ....................  ....................  ....................
24145 MT821590.1 |Severe acute respiratory syndrome coro    ....................  ....................  .....................  ....................  ....................  ....................
24146 MT821591.1 |Severe acute respiratory syndrome coro    ....................  ....................  .....................  ....................  ....................  ....................
24147 MT821592.1 |Severe acute respiratory syndrome coro    ....................  ....................  .....................  ....................  ....................  ....................
24148 MT821593.1 |Severe acute respiratory syndrome coro    ....................  ....................  .....................  ....................  ....................  ....................
24149 MT821594.1 |Severe acute respiratory syndrome coro    ....................  ....................  .....................  ....................  ....................  ....................
24150 MT821595.1 |Severe acute respiratory syndrome coro    ....................  ....................  .....................  ....................  ....................  ....................
24151 MT821596.1 |Severe acute respiratory syndrome coro    ....................  ....................  .....................  ....................  ....................  ....................
24152 MT821597.1 |Severe acute respiratory syndrome coro    ....................  ....................  .....................  ....................  ....................  ....................
24153 MT821598.1 |Severe acute respiratory syndrome coro    ....................  ....................  .....................  ....................  ....................  ....................
24154 MT821599.1 |Severe acute respiratory syndrome coro    ....................  ....................  .....................  ....................  ....................  ....................
24155 MT821600.1 |Severe acute respiratory syndrome coro    ....................  ....................  .....................  ....................  ....................  ....................
24156 MT821601.1 |Severe acute respiratory syndrome coro    ....................  ....................  .....................  ....................  ....................  ....................
24157 MT821602.1 |Severe acute respiratory syndrome coro    ....................  ....................  .....................  ....................  ....................  ....................
24158 MT821603.1 |Severe acute respiratory syndrome coro    ....................  ....................  .....................  ....................  ....................  ....................
24159 MT821604.1 |Severe acute respiratory syndrome coro    ....................  ....................  .....................  ....................  ....................  ....................
24160 MT821605.1 |Severe acute respiratory syndrome coro    ....................  ....................  .....................  ....................  ....................  ....................
24161 MT821606.1 |Severe acute respiratory syndrome coro    ....................  ....................  .....................  ....................  ....................  ....................
24162 MT821607.1 |Severe acute respiratory syndrome coro    ....................  ....................  .....................  ....................  ....................  ....................
24163 MT821608.1 |Severe acute respiratory syndrome coro    ....................  ....................  .....................  ....................  ....................  ....................
24164 MT821609.1 |Severe acute respiratory syndrome coro    ....................  ....................  .....................  ....................  ....................  ....................
24165 MT821610.1 |Severe acute respiratory syndrome coro    ....................  ....................  .....................  ....................  ....................  ....................
24166 MT821611.1 |Severe acute respiratory syndrome coro    ....................  ....................  .....................  ....................  ....................  ....................
24167 MT821612.1 |Severe acute respiratory syndrome coro    ....................  ....................  .....................  ....................  ....................  ....................
24168 MT821613.1 |Severe acute respiratory syndrome coro    ....................  ....................  .....................  ....................  ....................  ....................
24169 MT821614.1 |Severe acute respiratory syndrome coro    ....................  ....................  .....................  ....................  ....................  ....................
24170 MT821615.1 |Severe acute respiratory syndrome coro    ....................  ....................  .....................  ....................  ....................  ....................
24171 MT821616.1 |Severe acute respiratory syndrome coro    ....................  ....................  .....................  ....................  ....................  ....................
24172 MT821617.1 |Severe acute respiratory syndrome coro    ....................  ....................  .....................  ....................  ....................  ....................
24173 MT821618.1 |Severe acute respiratory syndrome coro    ....................  ....................  .....................  ....................  ....................  ....................
24174 MT821619.1 |Severe acute respiratory syndrome coro    ....................  ....................  .....................  ....................  ....................  ....................
24175 MT821620.1 |Severe acute respiratory syndrome coro    ....................  ....................  .....................  ....................  ....................  ....................
24176 MT821621.1 |Severe acute respiratory syndrome coro    ....................  ....................  .....................  ....................  ....................  ....................
24177 MT821622.1 |Severe acute respiratory syndrome coro    ....................  ....................  .....................  ....................  ....................  ....................
24178 MT821623.1 |Severe acute respiratory syndrome coro    ....................  ....................  .....................  ....................  ....................  ....................
24179 MT821624.1 |Severe acute respiratory syndrome coro    ....................  ....................  .....................  ....................  ....................  ....................
24180 MT821625.1 |Severe acute respiratory syndrome coro    ....................  ....................  .....................  ....................  ....................  ....................
24181 MT821626.1 |Severe acute respiratory syndrome coro    ....................  ....................  .....................  ....................  ....................  ....................
24182 MT821627.1 |Severe acute respiratory syndrome coro    ....................  ....................  .....................  ....................  ....................  ....................
24183 MT821628.1 |Severe acute respiratory syndrome coro    ....................  ....................  .....................  ....................  ....................  ....................
24184 MT821629.1 |Severe acute respiratory syndrome coro    ....................  ....................  .....................  ....................  ....................  ....................
24185 MT821630.1 |Severe acute respiratory syndrome coro    ....................  ....................  .....................  ....................  ....................  ....................
24186 MT821631.1 |Severe acute respiratory syndrome coro    ....................  ....................  .....................  ....................  ....................  ....................
24187 MT821632.1 |Severe acute respiratory syndrome coro    ....................  ....................  .....................  ....................  ....................  ....................
24188 MT821634.1 |Severe acute respiratory syndrome coro    ....................  ....................  .....................  ....................  ....................  ....................
24189 MT821635.1 |Severe acute respiratory syndrome coro    ....................  ....................  .....................  ....................  ....................  ....................
24190 MT821636.1 |Severe acute respiratory syndrome coro    ....................  ....................  .....................  ....................  ....................  ....................
24191 MT821637.1 |Severe acute respiratory syndrome coro    ....................  ....................  .....................  ....................  ....................  ....................
24192 MT821638.1 |Severe acute respiratory syndrome coro    ....................  ....................  .....................  ....................  ....................  ....................
24193 MT821639.1 |Severe acute respiratory syndrome coro    ....................  ....................  .....................  ....................  ....................  ....................
24194 MT821640.1 |Severe acute respiratory syndrome coro    ....................  ....................  .....................  ....................  ....................  ....................
24195 MT821641.1 |Severe acute respiratory syndrome coro    ....................  ....................  .....................  ....................  ....................  ....................
24196 MT821642.1 |Severe acute respiratory syndrome coro    ....................  ....................  .....................  ....................  ....................  ....................
24197 MT821643.1 |Severe acute respiratory syndrome coro    ....................  ....................  .....................  ....................  ....................  ....................
24198 MT821644.1 |Severe acute respiratory syndrome coro    ....................  ....................  .....................  ....................  ....................  ....................
24199 MT821645.1 |Severe acute respiratory syndrome coro    ....................  ....................  .....................  ....................  ....................  ....................
24200 MT821646.1 |Severe acute respiratory syndrome coro    ....................  ....................  .....................  ....................  ....................  ....................
24201 MT821647.1 |Severe acute respiratory syndrome coro    ....................  ....................  .....................  ....................  ....................  ....................
24202 MT821648.1 |Severe acute respiratory syndrome coro    ....................  ....................  .....................  ....................  ....................  ....................
24203 MT821650.1 |Severe acute respiratory syndrome coro    ....................  ....................  .....................  ....................  ....................  ....................
24204 MT821652.1 |Severe acute respiratory syndrome coro    ....................  ....................  .....................  ....................  ....................  ....................
24205 MT821653.1 |Severe acute respiratory syndrome coro    ....................  ....................  .....................  ....................  ....................  ....................
24206 MT821654.1 |Severe acute respiratory syndrome coro    ....................  ....................  .....................  ....................  ....................  ....................
24207 MT821655.1 |Severe acute respiratory syndrome coro    ....................  ....................  .....................  ....................  ....................  ....................
24208 MT821656.1 |Severe acute respiratory syndrome coro    ....................  ....................  .....................  ....................  ....................  ....................
```

24209 MT821658.1 |Severe acute respiratory syndrome coro .................... .................... ..................... .................... .................... ....................
24210 MT821659.1 |Severe acute respiratory syndrome coro .................... .................... ..................... .................... .................... ....................
24211 MT821660.1 |Severe acute respiratory syndrome coro .................... .................... ..................... .................... .................... ....................
24212 MT821661.1 |Severe acute respiratory syndrome coro .................... .................... ..................... .................... .................... ....................
24213 MT821662.1 |Severe acute respiratory syndrome coro .................... .................... ..................... .................... .................... ....................
24214 MT821663.1 |Severe acute respiratory syndrome coro .................... .................... ..................... .................... .................... ....................
24215 MT821664.1 |Severe acute respiratory syndrome coro .................... .................... ..................... .................... .................... ....................
24216 MT821665.1 |Severe acute respiratory syndrome coro .................... .................... ..................... .................... .................... ....................
24217 MT821666.1 |Severe acute respiratory syndrome coro .................... .................... ..................... .................... .................... ....................
24218 MT821668.1 |Severe acute respiratory syndrome coro .................... .................... ..................... .................... .................... ....................
24219 MT821669.1 |Severe acute respiratory syndrome coro .................... .................... ..................... .................... .................... ....................
24220 MT821670.1 |Severe acute respiratory syndrome coro .................... .................... ..................... .................... .................... ....................
24221 MT821671.1 |Severe acute respiratory syndrome coro .................... .................... ..................... .................... .................... ....................
24222 MT821673.1 |Severe acute respiratory syndrome coro .................... .................... ..................... .................... .................... ....................
24223 MT821674.1 |Severe acute respiratory syndrome coro .................... .................... ..................... .................... .................... ....................
24224 MT821676.1 |Severe acute respiratory syndrome coro .................... .................... ..................... .................... .................... ....................
24225 MT821677.1 |Severe acute respiratory syndrome coro .................... .................... ..................... .................... .................... ....................
24226 MT821678.1 |Severe acute respiratory syndrome coro .................... .................... ..................... .................... .................... ....................
24227 MT821679.1 |Severe acute respiratory syndrome coro .................... .................... ..................... .................... .................... ....................
24228 MT821680.1 |Severe acute respiratory syndrome coro .................... .................... ..................... .................... .................... ....................
24229 MT821681.1 |Severe acute respiratory syndrome coro .................... .................... ..................... .................... .................... ....................
24230 MT821682.1 |Severe acute respiratory syndrome coro .................... .................... ..................... .................... .................... ....................
24231 MT821683.1 |Severe acute respiratory syndrome coro .................... .................... ..................... .................... .................... ....................
24232 MT821684.1 |Severe acute respiratory syndrome coro .................... .................... ..................... .................... .................... ....................
24233 MT821685.1 |Severe acute respiratory syndrome coro .................... .................... ..................... .................... .................... ....................
24234 MT821686.1 |Severe acute respiratory syndrome coro .................... .................... ..................... .................... .................... ....................
24235 MT821687.1 |Severe acute respiratory syndrome coro .................... .................... ..................... .................... .................... ....................
24236 MT821688.1 |Severe acute respiratory syndrome coro .................... .................... ..................... .................... .................... ....................
24237 MT821689.1 |Severe acute respiratory syndrome coro .................... .................... ..................... .................... .................... ....................
24238 MT821690.1 |Severe acute respiratory syndrome coro .................... .................... ..................... .................... .................... ....................
24239 MT821691.1 |Severe acute respiratory syndrome coro .................... .................... ..................... .................... .................... ....................
24240 MT821692.1 |Severe acute respiratory syndrome coro .................... .................... ..................... .................... .................... ....................
24241 MT821693.1 |Severe acute respiratory syndrome coro .................... .................... ..................... .................... .................... ....................
24242 MT821694.1 |Severe acute respiratory syndrome coro .................... .................... ..................... .................... .................... ....................
24243 MT821695.1 |Severe acute respiratory syndrome coro .................... .................... ..................... .................... .................... ....................
24244 MT821696.1 |Severe acute respiratory syndrome coro .................... .................... ..................... .................... .................... ....................
24245 MT821698.1 |Severe acute respiratory syndrome coro .................... .................... ..................... .................... .................... ....................
24246 MT821699.1 |Severe acute respiratory syndrome coro .................... .................... ..................... .................... .................... ....................
24247 MT821700.1 |Severe acute respiratory syndrome coro .................... .................... ..................... .................... .................... ....................
24248 MT821701.1 |Severe acute respiratory syndrome coro .................... .................... ..................... .................... .................... ....................
24249 MT821702.1 |Severe acute respiratory syndrome coro .................... .................... ..................... .................... .................... ....................
24250 MT821703.1 |Severe acute respiratory syndrome coro .................... .................... ..................... .................... .................... ....................
24251 MT821704.1 |Severe acute respiratory syndrome coro .................... .................... ..................... .................... .................... ....................
24252 MT821705.1 |Severe acute respiratory syndrome coro .................... .................... ..................... .................... .................... ....................
24253 MT821707.1 |Severe acute respiratory syndrome coro .................... .................... ..................... .................... .................... ....................
24254 MT821709.1 |Severe acute respiratory syndrome coro .................... .................... ..................... .................... .................... ....................
24255 MT821710.1 |Severe acute respiratory syndrome coro .................... .................... ..................... .................... .................... ....................
24256 MT821711.1 |Severe acute respiratory syndrome coro .................... .................... ..................... .................... .................... ....................
24257 MT821714.1 |Severe acute respiratory syndrome coro .................... .................... ..................... .................... .................... ....................
24258 MT821715.1 |Severe acute respiratory syndrome coro .................... .................... ..................... .................... .................... ....................
24259 MT821716.1 |Severe acute respiratory syndrome coro .................... .................... ..................... .................... .................... ....................
24260 MT821717.1 |Severe acute respiratory syndrome coro .................... .................... ..................... .................... .................... ....................
24261 MT821718.1 |Severe acute respiratory syndrome coro .................... .................... ..................... .................... .................... ....................
24262 MT821720.1 |Severe acute respiratory syndrome coro .................... .................... ..................... .................... .................... ....................
24263 MT821721.1 |Severe acute respiratory syndrome coro .................... .................... ..................... .................... .................... ....................
24264 MT821723.1 |Severe acute respiratory syndrome coro .................... .................... ..................... .................... .................... ....................
24265 MT821724.1 |Severe acute respiratory syndrome coro .................... .................... ..................... .................... .................... ....................
24266 MT821725.1 |Severe acute respiratory syndrome coro .................... .................... ..................... .................... .................... ....................
24267 MT821726.1 |Severe acute respiratory syndrome coro .................... .................... ..................... .................... .................... ....................
24268 MT821727.1 |Severe acute respiratory syndrome coro .................... .................... ..................... .................... .................... ....................
24269 MT821728.1 |Severe acute respiratory syndrome coro .................... .................... ..................... .................... .................... ....................
24270 MT821729.1 |Severe acute respiratory syndrome coro .................... .................... ..................... .................... .................... ....................
24271 MT821730.1 |Severe acute respiratory syndrome coro .................... .................... ..................... .................... .................... ....................
24272 MT821731.1 |Severe acute respiratory syndrome coro .................... .................... ..................... .................... .................... ....................
24273 MT821732.1 |Severe acute respiratory syndrome coro .................... .................... ..................... .................... .................... ....................
24274 MT821733.1 |Severe acute respiratory syndrome coro .................... .................... ..................... .................... .................... ....................
24275 MT821735.1 |Severe acute respiratory syndrome coro .................... .................... ..................... .................... .................... ....................
24276 MT821737.1 |Severe acute respiratory syndrome coro .................... .................... ..................... .................... .................... ....................
24277 MT821738.1 |Severe acute respiratory syndrome coro .................... .................... ..................... .................... .................... ....................
24278 MT821739.1 |Severe acute respiratory syndrome coro .................... .................... ..................... .................... .................... ....................
24279 MT821740.1 |Severe acute respiratory syndrome coro .................... .................... ..................... .................... .................... ....................
24280 MT821741.1 |Severe acute respiratory syndrome coro .................... .................... ..................... .................... .................... ....................
24281 MT821742.1 |Severe acute respiratory syndrome coro .................... .................... ..................... .................... .................... ....................
24282 MT821743.1 |Severe acute respiratory syndrome coro .................... .................... ..................... .................... .................... ....................
24283 MT821744.1 |Severe acute respiratory syndrome coro .................... .................... ..................... .................... .................... ....................
24284 MT821745.1 |Severe acute respiratory syndrome coro .................... .................... ..................... .................... .................... ....................
24285 MT821746.1 |Severe acute respiratory syndrome coro .................... .................... ..................... .................... .................... ....................
24286 MT821747.1 |Severe acute respiratory syndrome coro .................... .................... ..................... .................... .................... ....................

24287 MT821748.1 |Severe acute respiratory syndrome coro .................... .................... .................... .................... .................... ....................
24288 MT821749.1 |Severe acute respiratory syndrome coro .................... .................... .................... .................... .................... ....................
24289 MT821750.1 |Severe acute respiratory syndrome coro .................... .................... .................... .................... .................... ....................
24290 MT821752.1 |Severe acute respiratory syndrome coro .................... .................... .................... .................... .................... ....................
24291 MT821753.1 |Severe acute respiratory syndrome coro .................... .................... .................... .................... .................... ....................
24292 MT821754.1 |Severe acute respiratory syndrome coro .................... .................... .................... .................... .................... ....................
24293 MT821755.1 |Severe acute respiratory syndrome coro .................... .................... .................... .................... ............T....... ....................
24294 MT821756.1 |Severe acute respiratory syndrome coro .................... .................... .................... .................... .................... ....................
24295 MT821758.1 |Severe acute respiratory syndrome coro .................... .................... .................... .................... .................... ....................
24296 MT821759.1 |Severe acute respiratory syndrome coro .................... .................... .................... .................... .................... ....................
24297 MT821760.1 |Severe acute respiratory syndrome coro .................... .................... .................... .................... .................... ....................
24298 MT821761.1 |Severe acute respiratory syndrome coro .................... .................... .................... .................... .................... ....................
24299 MT821762.1 |Severe acute respiratory syndrome coro .................... .................... .................... .................... .................... ....................
24300 MT821763.1 |Severe acute respiratory syndrome coro .................... .................... .................... .................... .................... ....................
24301 MT821764.1 |Severe acute respiratory syndrome coro .................... .................... .................... .................... .................... ....................
24302 MT821765.1 |Severe acute respiratory syndrome coro .................... .................... .................... .................... .................... ....................
24303 MT821766.1 |Severe acute respiratory syndrome coro .................... .................... .................... .................... .................... ....................
24304 MT821767.1 |Severe acute respiratory syndrome coro .................... .................... .................... .................... .................... ....................
24305 MT821768.1 |Severe acute respiratory syndrome coro .................... .................... .................... .................... .................... ....................
24306 MT821769.1 |Severe acute respiratory syndrome coro .................... .................... .................... .................... .................... ....................
24307 MT821770.1 |Severe acute respiratory syndrome coro .................... .................... .................... .................... .................... ....................
24308 MT821771.1 |Severe acute respiratory syndrome coro .................... .................... .................... .................... .................... ....................
24309 MT821772.1 |Severe acute respiratory syndrome coro .................... .................... .................... .................... .................... ....................
24310 MT821773.1 |Severe acute respiratory syndrome coro .................... .................... .................... .................... .................... ....................
24311 MT821774.1 |Severe acute respiratory syndrome coro .................... .................... .................... .................... .................... ....................
24312 MT821775.1 |Severe acute respiratory syndrome coro .................... .................... .................... .................... .................... ....................
24313 MT821776.1 |Severe acute respiratory syndrome coro .................... .................... .................... .................... .................... ....................
24314 MT821777.1 |Severe acute respiratory syndrome coro .................... .................... .................... .................... .................... ....................
24315 MT821778.1 |Severe acute respiratory syndrome coro .................... .................... .................... .................... .................... ....................
24316 MT821780.1 |Severe acute respiratory syndrome coro .................... .................... .................... .................... .................... ....................
24317 MT821781.1 |Severe acute respiratory syndrome coro .................... .................... .................... .................... .................... ....................
24318 MT821782.1 |Severe acute respiratory syndrome coro .................... .................... .................... .................... .................... ....................
24319 MT821783.1 |Severe acute respiratory syndrome coro .................... .................... .................... .................... .................... ....................
24320 MT821784.1 |Severe acute respiratory syndrome coro .................... .................... .................... .................... .................... ....................
24321 MT821785.1 |Severe acute respiratory syndrome coro .................... .................... .................... .................... .................... ....................
24322 MT821786.1 |Severe acute respiratory syndrome coro .................... .................... .................... .................... .................... ....................
24323 MT821787.1 |Severe acute respiratory syndrome coro .................... .................... .................... .................... .................... ....................
24324 MT821788.1 |Severe acute respiratory syndrome coro .................... .................... .................... .................... .................... ....................
24325 MT821789.1 |Severe acute respiratory syndrome coro .................... .................... .................... .................... .................... ....................
24326 MT821790.1 |Severe acute respiratory syndrome coro .................... .................... .................... .................... .................... ....................
24327 MT821791.1 |Severe acute respiratory syndrome coro .................... .................... .................... .................... .................... ....................
24328 MT821792.1 |Severe acute respiratory syndrome coro .................... .................... .................... .................... .................... ....................
24329 MT821793.1 |Severe acute respiratory syndrome coro .................... .................... .................... .................... .................... ....................
24330 MT821795.1 |Severe acute respiratory syndrome coro .................... .................... .................... .................... .................... ....................
24331 MT820127.1 |Severe acute respiratory syndrome coro .................... .................... .................... .................... .................... ....................
24332 MT820128.1 |Severe acute respiratory syndrome coro .................... .................... .................... .................... .................... ....................
24333 MT820129.1 |Severe acute respiratory syndrome coro .................... .................... .................... .................... .................... ....................
24334 MT820130.1 |Severe acute respiratory syndrome coro .................... .................... .................... .................... .................... ....................
24335 MT820131.1 |Severe acute respiratory syndrome coro .................... .................... .................... .................... .................... ....................
24336 MT820132.1 |Severe acute respiratory syndrome coro .................... .................... .................... .................... .................... ....................
24337 MT820461.1 |Severe acute respiratory syndrome coro .................... .................... .................... .................... .................... ....................
24338 MT820462.1 |Severe acute respiratory syndrome coro .................... .................... .................... .................... .................... ....................
24339 MT820463.1 |Severe acute respiratory syndrome coro .................... .................... .................... .................... .................... ....................
24340 MT820464.1 |Severe acute respiratory syndrome coro .................... .................... .................... .................... .................... ....................
24341 MT820465.1 |Severe acute respiratory syndrome coro .................... .................... .................... .................... .................... ....................
24342 MT820466.1 |Severe acute respiratory syndrome coro .................... .................... .................... .................... .................... ....................
24343 MT820467.1 |Severe acute respiratory syndrome coro .................... .................... .................... .................... .................... ....................
24344 MT820468.1 |Severe acute respiratory syndrome coro .................... .................... .................... .................... .................... ....................
24345 MT820469.1 |Severe acute respiratory syndrome coro .................... .................... .................... .................... .................... ....................
24346 MT820470.1 |Severe acute respiratory syndrome coro .................... .................... .................... .................... .................... ....................
24347 MT820471.1 |Severe acute respiratory syndrome coro .................... .................... .................... .................... .................... ....................
24348 MT820472.1 |Severe acute respiratory syndrome coro .................... .................... .................... .................... .................... ....................
24349 MT820473.1 |Severe acute respiratory syndrome coro .................... .................... .................... .................... .................... ....................
24350 MT820474.1 |Severe acute respiratory syndrome coro .................... .................... .................... .................... .................... ....................
24351 MT820475.1 |Severe acute respiratory syndrome coro .................... .................... .................... .................... .................... ....................
24352 MT820476.1 |Severe acute respiratory syndrome coro .................... .................... .................... .................... .................... ....................
24353 MT820477.1 |Severe acute respiratory syndrome coro .................... .................... .................... .................... .................... ....................
24354 MT820478.1 |Severe acute respiratory syndrome coro .................... .................... .................... .................... .................... ....................
24355 MT820479.1 |Severe acute respiratory syndrome coro .................... .................... .................... .................... .................... ....................
24356 MT820480.1 |Severe acute respiratory syndrome coro .................... .................... .................... .................... .................... ....................
24357 MT820481.1 |Severe acute respiratory syndrome coro .................... .................... .................... .................... .................... ....................
24358 MT820482.1 |Severe acute respiratory syndrome coro .................... .................... .................... .................... .................... ....................
24359 MT820483.1 |Severe acute respiratory syndrome coro .................... .................... .................... .................... .................... ....................
24360 MT820484.1 |Severe acute respiratory syndrome coro .................... .................... .................... .................... .................... ....................
24361 MT820486.1 |Severe acute respiratory syndrome coro .................... .................... .................... .................... .................... ....................
24362 MT820487.1 |Severe acute respiratory syndrome coro .................... .................... .................... .................... .................... ....................
24363 MT820488.1 |Severe acute respiratory syndrome coro .................... .................... .................... .................... .................... ....................
24364 MT820489.1 |Severe acute respiratory syndrome coro .................... .................... .................... .................... .................... ....................

| | | | | | | | | |
|---|---|---|---|---|---|---|---|---|
| 24365 | MT810084.1 | \|Severe acute respiratory syndrome coro | ···················· | ···················· | ···················· | ···················· | ···················· | ···················· |
| 24366 | MT810086.1 | \|Severe acute respiratory syndrome coro | ···················· | ·A·················· | ···················· | ···················· | ···················· | ···················· |
| 24367 | MT810087.1 | \|Severe acute respiratory syndrome coro | ···················· | ···················· | ···················· | ···················· | ···················· | ···················· |
| 24368 | MT810088.1 | \|Severe acute respiratory syndrome coro | ···················· | ·A·················· | ···················· | ···················· | ···················· | ···················· |
| 24369 | MT810089.1 | \|Severe acute respiratory syndrome coro | ···················· | ·A·················· | ···················· | ···················· | ···················· | ···················· |
| 24370 | MT810477.1 | \|Severe acute respiratory syndrome coro | ···················· | ···················· | ···················· | ···················· | ···················· | ···················· |
| 24371 | MT810478.1 | \|Severe acute respiratory syndrome coro | ···················· | ···················· | ···················· | ···················· | ···················· | ···················· |
| 24372 | MT810479.1 | \|Severe acute respiratory syndrome coro | ···················· | ···················· | ···················· | ···················· | ···················· | ···················· |
| 24373 | MT810480.1 | \|Severe acute respiratory syndrome coro | ···················· | ···················· | ···················· | ···················· | ···················· | ···················· |
| 24374 | MT810481.1 | \|Severe acute respiratory syndrome coro | ···················· | ···················· | ···················· | ···················· | ···················· | ···················· |
| 24375 | MT810483.1 | \|Severe acute respiratory syndrome coro | ···················· | ···················· | ···················· | ···················· | ···················· | ···················· |
| 24376 | MT810485.1 | \|Severe acute respiratory syndrome coro | ···················· | ·A·················· | ···················· | ···················· | ···················· | ···················· |
| 24377 | MT810486.1 | \|Severe acute respiratory syndrome coro | ···················· | ·A·················· | ···················· | ···················· | ···················· | ···················· |
| 24378 | MT810487.1 | \|Severe acute respiratory syndrome coro | ···················· | ·A·················· | ···················· | ···················· | ···················· | ···················· |
| 24379 | MT810488.1 | \|Severe acute respiratory syndrome coro | ···················· | ·A·················· | ···················· | ···················· | ···················· | ···················· |
| 24380 | MT810489.1 | \|Severe acute respiratory syndrome coro | ···················· | ···················· | ···················· | ···················· | ···················· | ···················· |
| 24381 | MT810491.1 | \|Severe acute respiratory syndrome coro | ···················· | ·A·················· | ···················· | ···················· | ···················· | ···················· |
| 24382 | MT810492.1 | \|Severe acute respiratory syndrome coro | ···················· | ·A·················· | ···················· | ···················· | ···················· | ···················· |
| 24383 | MT810493.1 | \|Severe acute respiratory syndrome coro | ···················· | ·A·················· | ···················· | ···················· | ···················· | ···················· |
| 24384 | MT810494.1 | \|Severe acute respiratory syndrome coro | ···················· | ·A·················· | ···················· | ···················· | ···················· | ···················· |
| 24385 | MT810495.1 | \|Severe acute respiratory syndrome coro | ···················· | ·A·················· | ···················· | ···················· | ···················· | ···················· |
| 24386 | MT810496.1 | \|Severe acute respiratory syndrome coro | ···················· | ·A·················· | ···················· | ···················· | ···················· | ···················· |
| 24387 | MT810497.1 | \|Severe acute respiratory syndrome coro | ···················· | ·A·················· | ···················· | ···················· | ···················· | ···················· |
| 24388 | MT810498.1 | \|Severe acute respiratory syndrome coro | ···················· | ·A·················· | ···················· | ···················· | ···················· | ···················· |
| 24389 | MT810499.1 | \|Severe acute respiratory syndrome coro | ···················· | ·A·················· | ···················· | ···················· | ···················· | ···················· |
| 24390 | MT810500.1 | \|Severe acute respiratory syndrome coro | ···················· | ···················· | ···················· | ···················· | ···················· | ···················· |
| 24391 | MT810501.1 | \|Severe acute respiratory syndrome coro | ···················· | ·A·················· | ···················· | ···················· | ···················· | ···················· |
| 24392 | MT810502.1 | \|Severe acute respiratory syndrome coro | ···················· | ·A·················· | ···················· | ···················· | ···················· | ···················· |
| 24393 | MT810503.1 | \|Severe acute respiratory syndrome coro | ···················· | ·A·················· | ···················· | ···················· | ···················· | ···················· |
| 24394 | MT810504.1 | \|Severe acute respiratory syndrome coro | ···················· | ···················· | ···················· | ···················· | ···················· | ···················· |
| 24395 | MT810505.1 | \|Severe acute respiratory syndrome coro | ···················· | ·A·················· | ···················· | ···················· | ···················· | ···················· |
| 24396 | MT810506.1 | \|Severe acute respiratory syndrome coro | ···················· | ···················· | ···················· | ···················· | ···················· | ···················· |
| 24397 | MT810507.1 | \|Severe acute respiratory syndrome coro | ···················· | ···················· | ···················· | ···················· | ···················· | ···················· |
| 24398 | MT810508.1 | \|Severe acute respiratory syndrome coro | ···················· | ···················· | ···················· | ···················· | ···················· | ···················· |
| 24399 | MT810509.1 | \|Severe acute respiratory syndrome coro | ···················· | ·A·················· | ···················· | ···················· | ···················· | ···················· |
| 24400 | MT810510.1 | \|Severe acute respiratory syndrome coro | ···················· | ···················· | ···················· | ···················· | ···················· | ···················· |
| 24401 | MT810511.1 | \|Severe acute respiratory syndrome coro | ···················· | ···················· | ···················· | ···················· | ···················· | ···················· |
| 24402 | MT810512.1 | \|Severe acute respiratory syndrome coro | ···················· | ···················· | ···················· | ···················· | ···················· | ···················· |
| 24403 | MT810513.1 | \|Severe acute respiratory syndrome coro | ···················· | ···················· | ···················· | ···················· | ···················· | ···················· |
| 24404 | MT810514.1 | \|Severe acute respiratory syndrome coro | ···················· | ···················· | ···················· | ···················· | ···················· | ···················· |
| 24405 | MT810516.1 | \|Severe acute respiratory syndrome coro | ···················· | ···················· | ···················· | ···················· | ···················· | ···················· |
| 24406 | MT810518.1 | \|Severe acute respiratory syndrome coro | ···················· | ·A·················· | ···················· | ···················· | ···················· | ···················· |
| 24407 | MT810519.1 | \|Severe acute respiratory syndrome coro | ···················· | ·A·················· | ···················· | ···················· | ···················· | ···················· |
| 24408 | MT810521.1 | \|Severe acute respiratory syndrome coro | ···················· | ···················· | ···················· | ···················· | ···················· | ···················· |
| 24409 | MT810522.1 | \|Severe acute respiratory syndrome coro | ···················· | ···················· | ···················· | ···················· | ···················· | ···················· |
| 24410 | MT810523.1 | \|Severe acute respiratory syndrome coro | ···················· | ···················· | ···················· | ···················· | ···················· | ···················· |
| 24411 | MT810524.1 | \|Severe acute respiratory syndrome coro | ···················· | ···················· | ···················· | ···················· | ···················· | ···················· |
| 24412 | MT810525.1 | \|Severe acute respiratory syndrome coro | ···················· | ···················· | ···················· | ···················· | ···················· | ···················· |
| 24413 | MT810526.1 | \|Severe acute respiratory syndrome coro | ···················· | ···················· | ···················· | ···················· | ···················· | ···················· |
| 24414 | MT810527.1 | \|Severe acute respiratory syndrome coro | ···················· | ···················· | ···················· | ···················· | ···················· | ···················· |
| 24415 | MT810528.1 | \|Severe acute respiratory syndrome coro | ···················· | ···················· | ···················· | ···················· | ···················· | ···················· |
| 24416 | MT810529.1 | \|Severe acute respiratory syndrome coro | ···················· | ···················· | ···················· | ···················· | ···················· | ···················· |
| 24417 | MT810530.1 | \|Severe acute respiratory syndrome coro | ···················· | ···················· | ···················· | ···················· | ···················· | ···················· |
| 24418 | MT810531.1 | \|Severe acute respiratory syndrome coro | ···················· | ···················· | ···················· | ···················· | ···················· | ···················· |
| 24419 | MT810532.1 | \|Severe acute respiratory syndrome coro | ···················· | ···················· | ···················· | ···················· | ···················· | ···················· |
| 24420 | MT810533.1 | \|Severe acute respiratory syndrome coro | ···················· | ···················· | ···················· | ···················· | ···················· | ···················· |
| 24421 | MT810534.1 | \|Severe acute respiratory syndrome coro | ···················· | ···················· | ···················· | ···················· | ···················· | ···················· |
| 24422 | MT810535.1 | \|Severe acute respiratory syndrome coro | ···················· | ···················· | ···················· | ···················· | ···················· | ···················· |
| 24423 | MT810536.1 | \|Severe acute respiratory syndrome coro | ···················· | ···················· | ···················· | ···················· | ···················· | ···················· |
| 24424 | MT810537.1 | \|Severe acute respiratory syndrome coro | ···················· | ···················· | ···················· | ···················· | ···················· | ···················· |
| 24425 | MT810538.1 | \|Severe acute respiratory syndrome coro | ···················· | ···················· | ···················· | ···················· | ···················· | ···················· |
| 24426 | MT810539.1 | \|Severe acute respiratory syndrome coro | ···················· | ···················· | ···················· | ···················· | ···················· | ···················· |
| 24427 | MT810540.1 | \|Severe acute respiratory syndrome coro | ···················· | ···················· | ···················· | ···················· | ···················· | ···················· |
| 24428 | MT810541.1 | \|Severe acute respiratory syndrome coro | ···················· | ···················· | ···················· | ···················· | ···················· | ···················· |
| 24429 | MT810542.1 | \|Severe acute respiratory syndrome coro | ···················· | ···················· | ···················· | ···················· | ···················· | ···················· |
| 24430 | MT810544.1 | \|Severe acute respiratory syndrome coro | ···················· | ···················· | ···················· | ···················· | ···················· | ···················· |
| 24431 | MT810545.1 | \|Severe acute respiratory syndrome coro | ···················· | ···················· | ···················· | ···················· | ···················· | ···················· |
| 24432 | MT810546.1 | \|Severe acute respiratory syndrome coro | ···················· | ···················· | ···················· | ···················· | ···················· | ···················· |
| 24433 | MT810547.1 | \|Severe acute respiratory syndrome coro | ···················· | ···················· | ···················· | ···················· | ···················· | ···················· |
| 24434 | MT810548.1 | \|Severe acute respiratory syndrome coro | ···················· | ···················· | ···················· | ···················· | ···················· | ···················· |
| 24435 | MT810549.1 | \|Severe acute respiratory syndrome coro | ···················· | ···················· | ···················· | ···················· | ···················· | ···················· |
| 24436 | MT810550.1 | \|Severe acute respiratory syndrome coro | ···················· | ·A·················· | ···················· | ···················· | ···················· | ···················· |
| 24437 | MT810551.1 | \|Severe acute respiratory syndrome coro | ···················· | ·A·················· | ···················· | ···················· | ···················· | ···················· |
| 24438 | MT810552.1 | \|Severe acute respiratory syndrome coro | ···················· | ···················· | ···················· | ···················· | ···················· | ···················· |
| 24439 | MT810553.1 | \|Severe acute respiratory syndrome coro | ···················· | ···················· | ···················· | ···················· | ···················· | ···················· |
| 24440 | MT810554.1 | \|Severe acute respiratory syndrome coro | ···················· | ···················· | ···················· | ···················· | ···················· | ···················· |
| 24441 | MT810555.1 | \|Severe acute respiratory syndrome coro | ···················· | ·A·················· | ···················· | ···················· | ···················· | ···················· |
| 24442 | MT810556.1 | \|Severe acute respiratory syndrome coro | ···················· | ···················· | ···················· | ···················· | ···················· | ···················· |

24443 MT810557.1 |Severe acute respiratory syndrome coro ···················· ···················· ···················· ···················· ···················· ····················
24444 MT810558.1 |Severe acute respiratory syndrome coro ···················· ···················· ···················· ···················· ···················· ····················
24445 MT810559.1 |Severe acute respiratory syndrome coro ···················· ···················· ···················· ···················· ···················· ····················
24446 MT810560.1 |Severe acute respiratory syndrome coro ···················· ···················· ···················· ···················· ···················· ····················
24447 MT810561.1 |Severe acute respiratory syndrome coro ···················· ···················· ···················· ···················· ···················· ····················
24448 MT810562.1 |Severe acute respiratory syndrome coro ···················· ···················· ···················· ···················· ···················· ····················
24449 MT810563.1 |Severe acute respiratory syndrome coro ···················· ···················· ···················· ···················· ···················· ····················
24450 MT810564.1 |Severe acute respiratory syndrome coro ···················· ···················· ···················· ···················· ···················· ····················
24451 MT810565.1 |Severe acute respiratory syndrome coro ···················· ···················· ···················· ···················· ···················· ····················
24452 MT810566.1 |Severe acute respiratory syndrome coro ···················· ···················· ···················· ···················· ···················· ····················
24453 MT810567.1 |Severe acute respiratory syndrome coro ···················· ···················· ···················· ···················· ···················· ····················
24454 MT810568.1 |Severe acute respiratory syndrome coro ···················· ···················· ···················· ···················· ···················· ····················
24455 MT810570.1 |Severe acute respiratory syndrome coro ···················· ···················· ···················· ···················· ···················· ····················
24456 MT810571.1 |Severe acute respiratory syndrome coro ···················· ···················· ···················· ···················· ···················· ····················
24457 MT810572.1 |Severe acute respiratory syndrome coro ···················· ···················· ···················· ···················· ···················· ····················
24458 MT810573.1 |Severe acute respiratory syndrome coro ···················· ···················· ···················· ···················· ···················· ····················
24459 MT810574.1 |Severe acute respiratory syndrome coro ···················· ···················· ···················· ···················· ···················· ····················
24460 MT810575.1 |Severe acute respiratory syndrome coro ···················· ···················· ···················· ···················· ···················· ····················
24461 MT810577.1 |Severe acute respiratory syndrome coro ···················· ···················· ···················· ···················· ···················· ····················
24462 MT810578.1 |Severe acute respiratory syndrome coro ···················· ···················· ···················· ···················· ···················· ····················
24463 MT810579.1 |Severe acute respiratory syndrome coro ···················· ···················· ···················· ···················· ···················· ····················
24464 MT810580.1 |Severe acute respiratory syndrome coro ···················· ···················· ···················· ···················· ···················· ····················
24465 MT810581.1 |Severe acute respiratory syndrome coro ···················· ···················· ···················· ···················· ···················· ····················
24466 MT810582.1 |Severe acute respiratory syndrome coro ···················· ···················· ···················· ···················· ···················· ····················
24467 MT810584.1 |Severe acute respiratory syndrome coro ···················· ···················· ···················· ···················· ···················· ····················
24468 MT810585.1 |Severe acute respiratory syndrome coro ···················· ·A·················· ···················· ···················· ···················· ····················
24469 MT810586.1 |Severe acute respiratory syndrome coro ···················· ·A·················· ···················· ···················· ···················· ····················
24470 MT810587.1 |Severe acute respiratory syndrome coro ···················· ·A·················· ···················· ···················· ···················· ····················
24471 MT810588.1 |Severe acute respiratory syndrome coro ···················· ·A·················· ···················· ···················· ···················· ····················
24472 MT810589.1 |Severe acute respiratory syndrome coro ···················· ·A·················· ···················· ···················· ···················· ····················
24473 MT810590.1 |Severe acute respiratory syndrome coro ···················· ·A·················· ···················· ···················· ···················· ····················
24474 MT810591.1 |Severe acute respiratory syndrome coro ···················· ·A·················· ···················· ···················· ···················· ····················
24475 MT810592.1 |Severe acute respiratory syndrome coro ···················· ·A·················· ···················· ···················· ···················· ····················
24476 MT810593.1 |Severe acute respiratory syndrome coro ···················· ·A·················· ···················· ···················· ···················· ····················
24477 MT810594.1 |Severe acute respiratory syndrome coro ···················· ·A·················· ···················· ···················· ···················· ····················
24478 MT810595.1 |Severe acute respiratory syndrome coro ···················· ·A·················· ···················· ···················· ···················· ····················
24479 MT810596.1 |Severe acute respiratory syndrome coro ···················· ·A·················· ···················· ···················· ···················· ····················
24480 MT810597.1 |Severe acute respiratory syndrome coro ···················· ·A·················· ···················· ···················· ···················· ····················
24481 MT810598.1 |Severe acute respiratory syndrome coro ···················· ·A·················· ···················· ···················· ···················· ····················
24482 MT810599.1 |Severe acute respiratory syndrome coro ···················· ···················· ···················· ···················· ···················· ····················
24483 MT810600.1 |Severe acute respiratory syndrome coro ···················· ···················· ···················· ···················· ···················· ····················
24484 MT810601.1 |Severe acute respiratory syndrome coro ···················· ···················· ···················· ···················· ···················· ····················
24485 MT810602.1 |Severe acute respiratory syndrome coro ···················· ···················· ···················· ···················· ···················· ····················
24486 MT810603.1 |Severe acute respiratory syndrome coro ···················· ·A·················· ···················· ···················· ···················· ····················
24487 MT810604.1 |Severe acute respiratory syndrome coro ···················· ···················· ···················· ···················· ···················· ····················
24488 MT810605.1 |Severe acute respiratory syndrome coro ···················· ···················· ···················· ···················· ···················· ····················
24489 MT810606.1 |Severe acute respiratory syndrome coro ···················· ···················· ···················· ···················· ···················· ····················
24490 MT810607.1 |Severe acute respiratory syndrome coro ···················· ···················· ···················· ···················· ···················· ····················
24491 MT810608.1 |Severe acute respiratory syndrome coro ···················· ···················· ···················· ···················· ···················· ····················
24492 MT810609.1 |Severe acute respiratory syndrome coro ···················· ·A·················· ···················· ···················· ···················· ····················
24493 MT810610.1 |Severe acute respiratory syndrome coro ···················· ···················· ···················· ···················· ···················· ····················
24494 MT810611.1 |Severe acute respiratory syndrome coro ···················· ···················· ···················· ···················· ···················· ····················
24495 MT810612.1 |Severe acute respiratory syndrome coro ···················· ···················· ···················· ···················· ···················· ····················
24496 MT810613.1 |Severe acute respiratory syndrome coro ···················· ·A·················· ···················· ···················· ···················· ····················
24497 MT810614.1 |Severe acute respiratory syndrome coro ···················· ···················· ···················· ···················· ···················· ····················
24498 MT810615.1 |Severe acute respiratory syndrome coro ···················· ·A·················· ···················· ···················· ···················· ····················
24499 MT810616.1 |Severe acute respiratory syndrome coro ···················· ·A·················· ···················· ···················· ···················· ····················
24500 MT810617.1 |Severe acute respiratory syndrome coro ···················· ···················· ···················· ···················· ···················· ····················
24501 MT810618.1 |Severe acute respiratory syndrome coro ···················· ···················· ···················· ···················· ···················· ····················
24502 MT810619.1 |Severe acute respiratory syndrome coro ···················· ···················· ···················· ···················· ···················· ····················
24503 MT810620.1 |Severe acute respiratory syndrome coro ···················· ·A·················· ···················· ···················· ···················· ····················
24504 MT810621.1 |Severe acute respiratory syndrome coro ···················· ···················· ···················· ···················· ···················· ····················
24505 MT810623.1 |Severe acute respiratory syndrome coro ···················· ·A·················· ········T··········· ···················· ···················· ····················
24506 MT810624.1 |Severe acute respiratory syndrome coro ···················· ·A·················· ···················· ···················· ···················· ····················
24507 MT810625.1 |Severe acute respiratory syndrome coro ···················· ···················· ···················· ···················· ···················· ····················
24508 MT810626.1 |Severe acute respiratory syndrome coro ···················· ·A·················· ···················· ···················· ···················· ····················
24509 MT810627.1 |Severe acute respiratory syndrome coro ···················· ···················· ···················· ···················· ···················· ····················
24510 MT810628.1 |Severe acute respiratory syndrome coro ···················· ·A·················· ···················· ···················· ···················· ····················
24511 MT810629.1 |Severe acute respiratory syndrome coro ···················· ···················· ···················· ···················· ···················· ····················
24512 MT810630.1 |Severe acute respiratory syndrome coro ···················· ·A·················· ···················· ···················· ···················· ····················
24513 MT810631.1 |Severe acute respiratory syndrome coro ···················· ···················· ···················· ···················· ···················· ····················
24514 MT810632.1 |Severe acute respiratory syndrome coro ···················· ···················· ···················· ···················· ···················· ····················
24515 MT810633.1 |Severe acute respiratory syndrome coro ···················· ···················· ···················· ···················· ···················· ····················
24516 MT810634.1 |Severe acute respiratory syndrome coro ···················· ···················· ···················· ···················· ···················· ····················
24517 MT810635.1 |Severe acute respiratory syndrome coro ···················· ···················· ···················· ···················· ···················· ····················
24518 MT810636.1 |Severe acute respiratory syndrome coro ···················· ···················· ···················· ···················· ···················· ····················
24519 MT810637.1 |Severe acute respiratory syndrome coro ···················· ···················· ···················· ···················· ···················· ····················
24520 MT810638.1 |Severe acute respiratory syndrome coro ···················· ···················· ···················· ···················· ···················· ····················

| | | | | | | | | |
|---|---|---|---|---|---|---|---|---|
| 24521 | MT810639.1 | \|Severe acute respiratory syndrome coro | .................... | .................... | .................... | .................... | .................... | .................... |
| 24522 | MT810640.1 | \|Severe acute respiratory syndrome coro | .................... | .................... | .................... | .................... | .................... | .................... |
| 24523 | MT810641.1 | \|Severe acute respiratory syndrome coro | .................... | .A.................. | .................... | .................... | .................... | .................... |
| 24524 | MT810642.1 | \|Severe acute respiratory syndrome coro | .................... | .A.................. | .................... | .................... | .................... | .................... |
| 24525 | MT810643.1 | \|Severe acute respiratory syndrome coro | .................... | .................... | .................... | .................... | .................... | .................... |
| 24526 | MT810644.1 | \|Severe acute respiratory syndrome coro | .................... | .................... | .................... | .................... | .................... | .................... |
| 24527 | MT810645.1 | \|Severe acute respiratory syndrome coro | .................... | .................... | .................... | .................... | .................... | .................... |
| 24528 | MT810646.1 | \|Severe acute respiratory syndrome coro | .................... | .................... | .................... | .................... | .................... | .................... |
| 24529 | MT810647.1 | \|Severe acute respiratory syndrome coro | .................... | .................... | .................... | .................... | .................... | .................... |
| 24530 | MT810648.1 | \|Severe acute respiratory syndrome coro | .................... | .................... | .................... | .................... | .................... | .................... |
| 24531 | MT810649.1 | \|Severe acute respiratory syndrome coro | .................... | .A.................. | .................... | .................... | .................... | .................... |
| 24532 | MT810650.1 | \|Severe acute respiratory syndrome coro | .................... | .A.................. | .................... | .................... | .................... | .................... |
| 24533 | MT810651.1 | \|Severe acute respiratory syndrome coro | .................... | .................... | .................... | .................... | .................... | .................... |
| 24534 | MT810652.1 | \|Severe acute respiratory syndrome coro | .................... | .A.................. | .................... | .................... | .................... | .................... |
| 24535 | MT810654.1 | \|Severe acute respiratory syndrome coro | .................... | .................... | .................... | .................... | .................... | .................... |
| 24536 | MT810655.1 | \|Severe acute respiratory syndrome coro | .................... | .A.................. | .................... | .................... | .................... | .................... |
| 24537 | MT810657.1 | \|Severe acute respiratory syndrome coro | .................... | .A.................. | .................... | .................... | .................... | .................... |
| 24538 | MT810658.1 | \|Severe acute respiratory syndrome coro | .................... | .A.................. | .................... | .................... | .................... | .................... |
| 24539 | MT810659.1 | \|Severe acute respiratory syndrome coro | .................... | .A.................. | .................... | .................... | .................... | .................... |
| 24540 | MT810660.1 | \|Severe acute respiratory syndrome coro | .................... | .A.................. | .................... | .................... | .................... | .................... |
| 24541 | MT810661.1 | \|Severe acute respiratory syndrome coro | .................... | .................... | .................... | .................... | .................... | .................... |
| 24542 | MT810662.1 | \|Severe acute respiratory syndrome coro | .................... | .A.................. | .................... | .................... | .................... | .................... |
| 24543 | MT810663.1 | \|Severe acute respiratory syndrome coro | .................... | .A.................. | .................... | .................... | .................... | .................... |
| 24544 | MT810664.1 | \|Severe acute respiratory syndrome coro | .................... | .A.................. | .................... | .................... | .................... | .................... |
| 24545 | MT810665.1 | \|Severe acute respiratory syndrome coro | .................... | .A.................. | .................... | .................... | .................... | .................... |
| 24546 | MT810666.1 | \|Severe acute respiratory syndrome coro | .................... | .A.................. | .................... | .................... | .................... | .................... |
| 24547 | MT810667.1 | \|Severe acute respiratory syndrome coro | .................... | .A.................. | .................... | .................... | .................... | .................... |
| 24548 | MT810668.1 | \|Severe acute respiratory syndrome coro | .................... | .A.................. | .................... | .................... | .................... | .................... |
| 24549 | MT810669.1 | \|Severe acute respiratory syndrome coro | .................... | .A.................. | .................... | .................... | .................... | .................... |
| 24550 | MT810670.1 | \|Severe acute respiratory syndrome coro | .................... | .A.................. | .................... | .................... | .................... | .................... |
| 24551 | MT810671.1 | \|Severe acute respiratory syndrome coro | .................... | .A.................. | .................... | .................... | .................... | .................... |
| 24552 | MT810672.1 | \|Severe acute respiratory syndrome coro | .................... | .................... | .................... | .................... | .................... | .................... |
| 24553 | MT810673.1 | \|Severe acute respiratory syndrome coro | .................... | .A.................. | .................... | .................... | .................... | .................... |
| 24554 | MT810675.1 | \|Severe acute respiratory syndrome coro | .................... | .A.................. | .................... | .................... | .................... | .................... |
| 24555 | MT810676.1 | \|Severe acute respiratory syndrome coro | .................... | .A.................. | .................... | .................... | .................... | .................... |
| 24556 | MT810677.1 | \|Severe acute respiratory syndrome coro | .................... | .A.................. | .................... | .................... | .................... | .................... |
| 24557 | MT810678.1 | \|Severe acute respiratory syndrome coro | .................... | .A.................. | .................... | .................... | .................... | .................... |
| 24558 | MT810679.1 | \|Severe acute respiratory syndrome coro | .................... | .................... | .................... | .................... | .................... | .................... |
| 24559 | MT810680.1 | \|Severe acute respiratory syndrome coro | .................... | .A.................. | .................... | .................... | .................... | .................... |
| 24560 | MT810681.1 | \|Severe acute respiratory syndrome coro | .................... | .A.................. | .................... | .................... | .................... | .................... |
| 24561 | MT810682.1 | \|Severe acute respiratory syndrome coro | .................... | .A.................. | .................... | .................... | .................... | .................... |
| 24562 | MT810683.1 | \|Severe acute respiratory syndrome coro | .................... | .A.................. | .................... | .................... | .................... | .................... |
| 24563 | MT810684.1 | \|Severe acute respiratory syndrome coro | .................... | .A.................. | .................... | .................... | .................... | .................... |
| 24564 | MT810685.1 | \|Severe acute respiratory syndrome coro | .................... | .................... | .................... | .................... | .................... | .................... |
| 24565 | MT810686.1 | \|Severe acute respiratory syndrome coro | .................... | .A.................. | .................... | .................... | .................... | .................... |
| 24566 | MT810687.1 | \|Severe acute respiratory syndrome coro | .................... | .A.................. | .................... | .................... | .................... | .................... |
| 24567 | MT810688.1 | \|Severe acute respiratory syndrome coro | .................... | .A.................. | .................... | .................... | .................... | .................... |
| 24568 | MT810690.1 | \|Severe acute respiratory syndrome coro | .................... | .A.................. | .................... | .................... | .................... | .................... |
| 24569 | MT810691.1 | \|Severe acute respiratory syndrome coro | .................... | .A.................. | .................... | .................... | .................... | .................... |
| 24570 | MT810693.1 | \|Severe acute respiratory syndrome coro | .................... | .A.................. | .................... | .................... | .................... | .................... |
| 24571 | MT810694.1 | \|Severe acute respiratory syndrome coro | .................... | .A.................. | .................... | .................... | .................... | .................... |
| 24572 | MT810695.1 | \|Severe acute respiratory syndrome coro | .................... | .A.................. | .................... | .................... | .................... | .................... |
| 24573 | MT810697.1 | \|Severe acute respiratory syndrome coro | .................... | .A.................. | .................... | .................... | .................... | .................... |
| 24574 | MT810698.1 | \|Severe acute respiratory syndrome coro | .................... | .A.................. | .................... | .................... | .................... | .................... |
| 24575 | MT810699.1 | \|Severe acute respiratory syndrome coro | .................... | .................... | .................... | .................... | .................... | .................... |
| 24576 | MT810700.1 | \|Severe acute respiratory syndrome coro | .................... | .................... | .................... | .................... | .................... | .................... |
| 24577 | MT810701.1 | \|Severe acute respiratory syndrome coro | .................... | .A.................. | .................... | .................... | .................... | .................... |
| 24578 | MT810702.1 | \|Severe acute respiratory syndrome coro | .................... | .A.................. | .................... | .................... | .................... | .................... |
| 24579 | MT810703.1 | \|Severe acute respiratory syndrome coro | .................... | .A.................. | .................... | .................... | .................... | .................... |
| 24580 | MT810704.1 | \|Severe acute respiratory syndrome coro | .................... | .A.................. | .................... | .................... | .................... | .................... |
| 24581 | MT810705.1 | \|Severe acute respiratory syndrome coro | .................... | .A.................. | .................... | .................... | .................... | .................... |
| 24583 | MT810707.1 | \|Severe acute respiratory syndrome coro | .................... | .................... | .................... | .................... | .................... | .................... |
| 24584 | MT810708.1 | \|Severe acute respiratory syndrome coro | .................... | .A.................. | .................... | .................... | .................... | .................... |
| 24585 | MT810709.1 | \|Severe acute respiratory syndrome coro | .................... | .A.................. | .................... | .................... | .................... | .................... |
| 24586 | MT810710.1 | \|Severe acute respiratory syndrome coro | .................... | .A.................. | .................... | .................... | .................... | .................... |
| 24587 | MT810712.1 | \|Severe acute respiratory syndrome coro | .................... | .................... | .................... | .................... | .................... | .................... |
| 24589 | MT810714.1 | \|Severe acute respiratory syndrome coro | .................... | .................... | .................... | .................... | .................... | .................... |
| 24590 | MT810716.1 | \|Severe acute respiratory syndrome coro | .................... | .A.................. | .................... | .................... | .................... | .................... |
| 24591 | MT810717.1 | \|Severe acute respiratory syndrome coro | .................... | .A.................. | .................... | .................... | .................... | .................... |
| 24592 | MT810718.1 | \|Severe acute respiratory syndrome coro | .................... | .A.................. | .................... | .................... | .................... | .................... |
| 24593 | MT810719.1 | \|Severe acute respiratory syndrome coro | .................... | .A.................. | .................... | .................... | .................... | .................... |
| 24594 | MT810720.1 | \|Severe acute respiratory syndrome coro | .................... | .................... | .................... | .................... | .................... | .................... |
| 24595 | MT810721.1 | \|Severe acute respiratory syndrome coro | .................... | .A.................. | .................... | .................... | .................... | .................... |
| 24596 | MT810722.1 | \|Severe acute respiratory syndrome coro | .................... | .A.................. | .................... | .................... | .................... | .................... |
| 24597 | MT810723.1 | \|Severe acute respiratory syndrome coro | .................... | .................... | .................... | .................... | .................... | .................... |
| 24598 | MT810724.1 | \|Severe acute respiratory syndrome coro | .................... | .................... | .................... | .................... | .................... | .................... |
| 24599 | MT810725.1 | \|Severe acute respiratory syndrome coro | .................... | .................... | .................... | .................... | .................... | .................... |
| 24600 | MT810727.1 | \|Severe acute respiratory syndrome coro | .................... | .................... | .................... | .................... | .................... | .................... |

```
24601 MT810728.1 |Severe acute respiratory syndrome coro    ····················  ····················  ·····················  ····················  ····················  ····················
24602 MT810730.1 |Severe acute respiratory syndrome coro    ····················  ·A··················  ·····················  ····················  ····················  ····················
24603 MT810731.1 |Severe acute respiratory syndrome coro    ····················  ·A··················  ·····················  ····················  ····················  ····················
24604 MT810758.1 |Severe acute respiratory syndrome coro    ····················  ·A··················  ·····················  ····················  ····················  ····················
24605 MT810761.1 |Severe acute respiratory syndrome coro    ····················  ·A··················  ·····················  ····················  ····················  ····················
24606 MT810764.1 |Severe acute respiratory syndrome coro    ····················  ·A··················  ·····················  ····················  ····················  ····················
24607 MT810765.1 |Severe acute respiratory syndrome coro    ····················  ·A··················  ·····················  ····················  ····················  ····················
24608 MT810766.1 |Severe acute respiratory syndrome coro    ····················  ·A··················  ·····················  ····················  ····················  ····················
24609 MT810767.1 |Severe acute respiratory syndrome coro    ····················  ·A··················  ·····················  ····················  ····················  ····················
24610 MT810768.1 |Severe acute respiratory syndrome coro    ····················  ·A··················  ·····················  ····················  ····················  ····················
24611 MT810769.1 |Severe acute respiratory syndrome coro    ····················  ·A··················  ·····················  ····················  ····················  ····················
24612 MT810771.1 |Severe acute respiratory syndrome coro    ····················  ·A··················  ·····················  ····················  ····················  ····················
24613 MT810772.1 |Severe acute respiratory syndrome coro    ····················  ·A··················  ·····················  ····················  ····················  ····················
24614 MT810773.1 |Severe acute respiratory syndrome coro    ····················  ····················  ·····················  ····················  ····················  ····················
24615 MT810774.1 |Severe acute respiratory syndrome coro    ····················  ·A··················  ·····················  ····················  ····················  ····················
24616 MT810775.1 |Severe acute respiratory syndrome coro    ····················  ·A··················  ·····················  ····················  ····················  ····················
24617 MT810776.1 |Severe acute respiratory syndrome coro    ····················  ·A··················  ·····················  ····················  ····················  ····················
24618 MT810778.1 |Severe acute respiratory syndrome coro    ····················  ·A··················  ·····················  ····················  ····················  ····················
24619 MT810779.1 |Severe acute respiratory syndrome coro    ····················  ·A··················  ·····················  ····················  ····················  ····················
24620 MT810780.1 |Severe acute respiratory syndrome coro    ····················  ·A··················  ·····················  ····················  ····················  ····················
24621 MT810781.1 |Severe acute respiratory syndrome coro    ····················  ····················  ·····················  ····················  ····················  ····················
24622 MT810782.1 |Severe acute respiratory syndrome coro    ····················  ·A··················  ·····················  ····················  ····················  ····················
24623 MT810783.1 |Severe acute respiratory syndrome coro    ····················  ····················  ·····················  ····················  ····················  ····················
24624 MT810784.1 |Severe acute respiratory syndrome coro    ····················  ····················  ·····················  ····················  ····················  ····················
24625 MT810786.1 |Severe acute respiratory syndrome coro    ····················  ·A··················  ·····················  ····················  ····················  ····················
24626 MT810787.1 |Severe acute respiratory syndrome coro    ····················  ·A··················  ·····················  ····················  ····················  ····················
24627 MT810788.1 |Severe acute respiratory syndrome coro    ····················  ·A··················  ·····················  ····················  ····················  ····················
24628 MT810789.1 |Severe acute respiratory syndrome coro    ····················  ·A··················  ·····················  ····················  ····················  ····················
24629 MT810814.1 |Severe acute respiratory syndrome coro    ····················  ····················  ·····················  ····················  ····················  ····················
24630 MT810815.1 |Severe acute respiratory syndrome coro    ····················  ····················  ·····················  ····················  ····················  ····················
24631 MT810816.1 |Severe acute respiratory syndrome coro    ····················  ····················  ·····················  ····················  ····················  ····················
24632 MT810819.1 |Severe acute respiratory syndrome coro    ····················  ····················  ·····················  ····················  ····················  ····················
24633 MT810821.1 |Severe acute respiratory syndrome coro    ····················  ····················  ·····················  ····················  ····················  ····················
24634 MT810822.1 |Severe acute respiratory syndrome coro    ····················  ····················  ·····················  ····················  ····················  ····················
24635 MT810823.1 |Severe acute respiratory syndrome coro    ····················  ····················  ·····················  ····················  ····················  ····················
24636 MT810824.1 |Severe acute respiratory syndrome coro    ····················  ····················  ·····················  ····················  ····················  ····················
24637 MT810825.1 |Severe acute respiratory syndrome coro    ····················  ····················  ·····················  ····················  ····················  ····················
24638 MT810826.1 |Severe acute respiratory syndrome coro    ····················  ····················  ·····················  ····················  ····················  ····················
24639 MT810827.1 |Severe acute respiratory syndrome coro    ····················  ····················  ·····················  ····················  ····················  ····················
24640 MT810828.1 |Severe acute respiratory syndrome coro    ····················  ····················  ·····················  ····················  ····················  ····················
24641 MT810829.1 |Severe acute respiratory syndrome coro    ····················  ····················  ·····················  ····················  ····················  ····················
24642 MT810830.1 |Severe acute respiratory syndrome coro    ····················  ····················  ·····················  ····················  ····················  ····················
24643 MT810831.1 |Severe acute respiratory syndrome coro    ····················  ····················  ·····················  ····················  ····················  ····················
24644 MT810832.1 |Severe acute respiratory syndrome coro    ····················  ····················  ·····················  ····················  ····················  ····················
24645 MT810833.1 |Severe acute respiratory syndrome coro    ····················  ····················  ·····················  ····················  ····················  ····················
24646 MT810835.1 |Severe acute respiratory syndrome coro    ····················  ····················  ·····················  ····················  ····················  ····················
24647 MT810836.1 |Severe acute respiratory syndrome coro    ····················  ·············A······  ·····················  ····················  ····················  ····················
24648 MT810837.1 |Severe acute respiratory syndrome coro    ····················  ····················  ·····················  ····················  ····················  ····················
24649 MT810838.1 |Severe acute respiratory syndrome coro    ····················  ·A··················  ·····················  ····················  ····················  ····················
24650 MT810839.1 |Severe acute respiratory syndrome coro    ····················  ·A··················  ·····················  ····················  ····················  ····················
24651 MT810841.1 |Severe acute respiratory syndrome coro    ····················  ·A··················  ·····················  ····················  ····················  ····················
24652 MT810842.1 |Severe acute respiratory syndrome coro    ····················  ·A··················  ·····················  ····················  ····················  ····················
24653 MT810843.1 |Severe acute respiratory syndrome coro    ····················  ·A··················  ·····················  ····················  ····················  ····················
24654 MT810846.1 |Severe acute respiratory syndrome coro    ····················  ····················  ·····················  ····················  ····················  ····················
24655 MT810847.1 |Severe acute respiratory syndrome coro    ····················  ····················  ·····················  ····················  ····················  ····················
24656 MT810848.1 |Severe acute respiratory syndrome coro    ····················  ·A··················  ·····················  ····················  ····················  ····················
24657 MT810850.1 |Severe acute respiratory syndrome coro    ····················  ·A··················  ·····················  ····················  ····················  ····················
24658 MT810851.1 |Severe acute respiratory syndrome coro    ····················  ····················  ·····················  ····················  ····················  ····················
24659 MT810852.1 |Severe acute respiratory syndrome coro    ····················  ·A··················  ·····················  ····················  ····················  ····················
24660 MT810855.1 |Severe acute respiratory syndrome coro    ····················  ····················  ·····················  ····················  ····················  ····················
24661 MT810856.1 |Severe acute respiratory syndrome coro    ····················  ·A··················  ·····················  ····················  ····················  ····················
24662 MT810857.1 |Severe acute respiratory syndrome coro    ····················  ·A··················  ·····················  ····················  ····················  ····················
24663 MT810858.1 |Severe acute respiratory syndrome coro    ····················  ·A··················  ·····················  ····················  ····················  ····················
24664 MT810859.1 |Severe acute respiratory syndrome coro    ····················  ·A··················  ·····················  ····················  ····················  ····················
24665 MT810861.1 |Severe acute respiratory syndrome coro    ····················  ·A··················  ·····················  ····················  ····················  ····················
24666 MT810863.1 |Severe acute respiratory syndrome coro    ····················  ·A··················  ·····················  ····················  ····················  ····················
24667 MT810865.1 |Severe acute respiratory syndrome coro    ····················  ·A··················  ·····················  ····················  ····················  ····················
24668 MT810866.1 |Severe acute respiratory syndrome coro    ····················  ····················  ·····················  ····················  ····················  ····················
24669 MT810867.1 |Severe acute respiratory syndrome coro    ····················  ····················  ·····················  ····················  ····················  ····················
24670 MT810869.1 |Severe acute respiratory syndrome coro    ····················  ····················  ·····················  ····················  ····················  ····················
24671 MT810870.1 |Severe acute respiratory syndrome coro    ····················  ·A··················  ·····················  ····················  ····················  ····················
24672 MT810872.1 |Severe acute respiratory syndrome coro    ····················  ·A··················  ·····················  ····················  ····················  ····················
24673 MT810873.1 |Severe acute respiratory syndrome coro    ····················  ·A··················  ·····················  ····················  ····················  ····················
24674 MT810874.1 |Severe acute respiratory syndrome coro    ····················  ·A··················  ·····················  ····················  ····················  ····················
24675 MT810875.1 |Severe acute respiratory syndrome coro    ····················  ····················  ·····················  ····················  ····················  ····················
24676 MT810876.1 |Severe acute respiratory syndrome coro    ····················  ····················  ·····················  ····················  ····················  ····················
24677 MT810877.1 |Severe acute respiratory syndrome coro    ····················  ····················  ·····················  ····················  ····················  ····················
24678 MT810879.1 |Severe acute respiratory syndrome coro    ····················  ····················  ·····················  ····················  ····················  ····················
```

```
24679 MT810881.1 |Severe acute respiratory syndrome coro    ····················  ····················  ····················  ····················  ····················  ····················
24680 MT810882.1 |Severe acute respiratory syndrome coro    ····················  ····················  ····················  ····················  ····················  ····················
24681 MT810883.1 |Severe acute respiratory syndrome coro    ····················  ·A··················  ····················  ····················  ····················  ····················
24682 MT810884.1 |Severe acute respiratory syndrome coro    ····················  ·A··················  ····················  ····················  ····················  ····················
24683 MT810885.1 |Severe acute respiratory syndrome coro    ····················  ····················  ····················  ····················  ····················  ····················
24684 MT810886.1 |Severe acute respiratory syndrome coro    ····················  ·A··················  ····················  ····················  ····················  ····················
24685 MT810888.1 |Severe acute respiratory syndrome coro    ····················  ·A··················  ····················  ····················  ····················  ····················
24686 MT810889.1 |Severe acute respiratory syndrome coro    ····················  ····················  ····················  ····················  ····················  ····················
24687 MT810890.1 |Severe acute respiratory syndrome coro    ····················  ····················  ····················  ····················  ····················  ····················
24688 MT810891.1 |Severe acute respiratory syndrome coro    ····················  ····················  ····················  ····················  ····················  ····················
24689 MT810892.1 |Severe acute respiratory syndrome coro    ····················  ····················  ····················  ····················  ····················  ····················
24690 MT810893.1 |Severe acute respiratory syndrome coro    ····················  ····················  ····················  ····················  ····················  ····················
24691 MT810894.1 |Severe acute respiratory syndrome coro    ····················  ·A··················  ····················  ····················  ····················  ····················
24692 MT810895.1 |Severe acute respiratory syndrome coro    ····················  ·A··················  ····················  ····················  ····················  ····················
24693 MT810896.1 |Severe acute respiratory syndrome coro    ····················  ····················  ····················  ····················  ····················  ····················
24694 MT810897.1 |Severe acute respiratory syndrome coro    ····················  ·A··················  ····················  ····················  ····················  ····················
24695 MT810898.1 |Severe acute respiratory syndrome coro    ····················  ····················  ····················  ····················  ····················  ····················
24696 MT810899.1 |Severe acute respiratory syndrome coro    ····················  ····················  ····················  ····················  ····················  ····················
24697 MT810900.1 |Severe acute respiratory syndrome coro    ····················  ····················  ····················  ····················  ····················  ····················
24698 MT810901.1 |Severe acute respiratory syndrome coro    ····················  ····················  ····················  ····················  ····················  ····················
24699 MT810902.1 |Severe acute respiratory syndrome coro    ····················  ·A··················  ····················  ····················  ····················  ····················
24700 MT810903.1 |Severe acute respiratory syndrome coro    ····················  ·A··················  ····················  ····················  ····················  ····················
24701 MT810905.1 |Severe acute respiratory syndrome coro    ····················  ····················  ····················  ····················  ····················  ····················
24702 MT810906.1 |Severe acute respiratory syndrome coro    ····················  ····················  ····················  ····················  ····················  ····················
24703 MT810907.1 |Severe acute respiratory syndrome coro    ····················  ····················  ····················  ····················  ····················  ····················
24704 MT810908.1 |Severe acute respiratory syndrome coro    ····················  ····················  ····················  ····················  ····················  ····················
24705 MT810909.1 |Severe acute respiratory syndrome coro    ····················  ····················  ····················  ····················  ····················  ····················
24706 MT810910.1 |Severe acute respiratory syndrome coro    ····················  ····················  ····················  ····················  ····················  ····················
24707 MT810911.1 |Severe acute respiratory syndrome coro    ····················  ····················  ····················  ····················  ····················  ····················
24708 MT810913.1 |Severe acute respiratory syndrome coro    ····················  ····················  ····················  ····················  ····················  ····················
24709 MT810915.1 |Severe acute respiratory syndrome coro    ····················  ····················  ····················  ····················  ····················  ····················
24710 MT810916.1 |Severe acute respiratory syndrome coro    ····················  ····················  ····················  ····················  ····················  ····················
24711 MT810917.1 |Severe acute respiratory syndrome coro    ····················  ····················  ····················  ····················  ····················  ····················
24712 MT810919.1 |Severe acute respiratory syndrome coro    ····················  ····················  ····················  ····················  ····················  ····················
24713 MT810920.1 |Severe acute respiratory syndrome coro    ····················  ····················  ····················  ····················  ····················  ····················
24714 MT810922.1 |Severe acute respiratory syndrome coro    ····················  ····················  ····················  ····················  ····················  ····················
24715 MT810924.1 |Severe acute respiratory syndrome coro    ····················  ····················  ····················  ····················  ····················  ····················
24716 MT810926.1 |Severe acute respiratory syndrome coro    ····················  ····················  ····················  ····················  ····················  ····················
24717 MT810927.1 |Severe acute respiratory syndrome coro    ····················  ····················  ····················  ····················  ····················  ····················
24718 MT810928.1 |Severe acute respiratory syndrome coro    ····················  ····················  ····················  ····················  ····················  ····················
24719 MT810929.1 |Severe acute respiratory syndrome coro    ····················  ····················  ····················  ····················  ····················  ····················
24720 MT810930.1 |Severe acute respiratory syndrome coro    ····················  ····················  ····················  ····················  ····················  ····················
24721 MT810932.1 |Severe acute respiratory syndrome coro    ····················  ····················  ····················  ····················  ····················  ····················
24722 MT810933.1 |Severe acute respiratory syndrome coro    ····················  ····················  ····················  ····················  ····················  ····················
24723 MT810936.1 |Severe acute respiratory syndrome coro    ····················  ····················  ····················  ····················  ····················  ····················
24724 MT810941.1 |Severe acute respiratory syndrome coro    ····················  ····················  ····················  ····················  ····················  ····················
24725 MT810942.1 |Severe acute respiratory syndrome coro    ····················  ·A··················  ····················  ····················  ····················  ····················
24726 MT810943.1 |Severe acute respiratory syndrome coro    ····················  ····················  ····················  ····················  ····················  ····················
24727 MT810946.1 |Severe acute respiratory syndrome coro    ····················  ·A··················  ····················  ····················  ····················  ····················
24728 MT810947.1 |Severe acute respiratory syndrome coro    ····················  ·A··················  ····················  ····················  ····················  ····················
24729 MT810949.1 |Severe acute respiratory syndrome coro    ····················  ····················  ····················  ····················  ····················  ····················
24730 MT810950.1 |Severe acute respiratory syndrome coro    ····················  ·A··················  ····················  ····················  ····················  ····················
24731 MT810951.1 |Severe acute respiratory syndrome coro    ····················  ····················  ····················  ····················  ····················  ····················
24732 MT810952.1 |Severe acute respiratory syndrome coro    ····················  ····················  ····················  ····················  ····················  ····················
24733 MT810953.1 |Severe acute respiratory syndrome coro    ····················  ····················  ····················  ····················  ····················  ····················
24734 MT810955.1 |Severe acute respiratory syndrome coro    ····················  ····················  ····················  ····················  ····················  ····················
24735 MT810957.1 |Severe acute respiratory syndrome coro    ····················  ····················  ····················  ····················  ····················  ····················
24736 MT810958.1 |Severe acute respiratory syndrome coro    ····················  ····················  ····················  ····················  ····················  ····················
24737 MT810961.1 |Severe acute respiratory syndrome coro    ····················  ····················  ····················  ····················  ····················  ····················
24738 MT810962.1 |Severe acute respiratory syndrome coro    ····················  ····················  ····················  ····················  ····················  ····················
24739 MT810963.1 |Severe acute respiratory syndrome coro    ····················  ····················  ····················  ····················  ····················  ····················
24740 MT810964.1 |Severe acute respiratory syndrome coro    ····················  ····················  ····················  ····················  ····················  ····················
24741 MT810966.1 |Severe acute respiratory syndrome coro    ····················  ····················  ····················  ····················  ····················  ····················
24742 MT810967.1 |Severe acute respiratory syndrome coro    ····················  ····················  ····················  ····················  ····················  ····················
24743 MT810969.1 |Severe acute respiratory syndrome coro    ····················  ····················  ····················  ····················  ····················  ····················
24744 MT810971.1 |Severe acute respiratory syndrome coro    ····················  ····················  ····················  ····················  ····················  ····················
24745 MT810972.1 |Severe acute respiratory syndrome coro    ····················  ····················  ····················  ····················  ····················  ····················
24746 MT810973.1 |Severe acute respiratory syndrome coro    ····················  ····················  ····················  ····················  ····················  ····················
24747 MT810975.1 |Severe acute respiratory syndrome coro    ····················  ····················  ····················  ····················  ····················  ····················
24748 MT810976.1 |Severe acute respiratory syndrome coro    ····················  ·A··················  ····················  ····················  ····················  ····················
24749 MT810977.1 |Severe acute respiratory syndrome coro    ····················  ·A··················  ····················  ····················  ····················  ····················
24750 MT810978.1 |Severe acute respiratory syndrome coro    ····················  ····················  ····················  ····················  ····················  ····················
24751 MT810979.1 |Severe acute respiratory syndrome coro    ····················  ····················  ····················  ····················  ····················  ····················
24752 MT810980.1 |Severe acute respiratory syndrome coro    ····················  ····················  ····················  ····················  ····················  ····················
24753 MT810981.1 |Severe acute respiratory syndrome coro    ····················  ····················  ····················  ····················  ····················  ····················
24754 MT810982.1 |Severe acute respiratory syndrome coro    ····················  ·A··················  ····················  ····················  ····················  ····················
24755 MT810983.1 |Severe acute respiratory syndrome coro    ····················  ····················  ····················  ····················  ····················  ····················
24756 MT810984.1 |Severe acute respiratory syndrome coro    ····················  ·A··················  ····················  ····················  ····················  ····················
```

| | | | | | | | | |
|---|---|---|---|---|---|---|---|---|
| 24757 | MT810985.1 | Severe acute respiratory syndrome coro | ···················· | ·A·················· | ···················· | ···················· | ···················· | ···················· |
| 24758 | MT810986.1 | Severe acute respiratory syndrome coro | ···················· | ···················· | ···················· | ···················· | ···················· | ···················· |
| 24759 | MT810987.1 | Severe acute respiratory syndrome coro | ···················· | ···················· | ···················· | ···················· | ···················· | ···················· |
| 24760 | MT810988.1 | Severe acute respiratory syndrome coro | ···················· | ···················· | ···················· | ···················· | ···················· | ···················· |
| 24761 | MT810989.1 | Severe acute respiratory syndrome coro | ···················· | ···················· | ···················· | ···················· | ···················· | ···················· |
| 24762 | MT810990.1 | Severe acute respiratory syndrome coro | ···················· | ···················· | ···················· | ···················· | ···················· | ···················· |
| 24763 | MT810991.1 | Severe acute respiratory syndrome coro | ···················· | ·A·················· | ···················· | ···················· | ···················· | ···················· |
| 24764 | MT810992.1 | Severe acute respiratory syndrome coro | ···················· | ···················· | ···················· | ···················· | ···················· | ···················· |
| 24765 | MT810993.1 | Severe acute respiratory syndrome coro | ···················· | ···················· | ···················· | ···················· | ···················· | ···················· |
| 24766 | MT810994.1 | Severe acute respiratory syndrome coro | ···················· | ···················· | ···················· | ···················· | ···················· | ···················· |
| 24767 | MT810995.1 | Severe acute respiratory syndrome coro | ···················· | ···················· | ···················· | ···················· | ···················· | ···················· |
| 24768 | MT810996.1 | Severe acute respiratory syndrome coro | ···················· | ···················· | ···················· | ···················· | ···················· | ···················· |
| 24769 | MT810997.1 | Severe acute respiratory syndrome coro | ···················· | ···················· | ···················· | ···················· | ···················· | ···················· |
| 24770 | MT810998.1 | Severe acute respiratory syndrome coro | ···················· | ···················· | ···················· | ···················· | ···················· | ···················· |
| 24771 | MT811000.1 | Severe acute respiratory syndrome coro | ···················· | ···················· | ···················· | ···················· | ···················· | ···················· |
| 24772 | MT811001.1 | Severe acute respiratory syndrome coro | ···················· | ···················· | ···················· | ···················· | ···················· | ···················· |
| 24773 | MT811002.1 | Severe acute respiratory syndrome coro | ···················· | ···················· | ···················· | ···················· | ···················· | ···················· |
| 24774 | MT811003.1 | Severe acute respiratory syndrome coro | ···················· | ···················· | ···················· | ···················· | ···················· | ···················· |
| 24775 | MT811006.1 | Severe acute respiratory syndrome coro | ···················· | ···················· | ···················· | ···················· | ···················· | ···················· |
| 24776 | MT811007.1 | Severe acute respiratory syndrome coro | ···················· | ·A·················· | ···················· | ···················· | ···················· | ···················· |
| 24777 | MT811008.1 | Severe acute respiratory syndrome coro | ···················· | ·A·················· | ···················· | ···················· | ···················· | ···················· |
| 24778 | MT811010.1 | Severe acute respiratory syndrome coro | ···················· | ···················· | ···················· | ···················· | ···················· | ···················· |
| 24779 | MT811011.1 | Severe acute respiratory syndrome coro | ···················· | ···················· | ···················· | ···················· | ···················· | ···················· |
| 24780 | MT811012.1 | Severe acute respiratory syndrome coro | ···················· | ···················· | ···················· | ···················· | ···················· | ···················· |
| 24781 | MT811013.1 | Severe acute respiratory syndrome coro | ···················· | ···················· | ···················· | ···················· | ···················· | ···················· |
| 24782 | MT811014.1 | Severe acute respiratory syndrome coro | ···················· | ···················· | ···················· | ···················· | ···················· | ···················· |
| 24783 | MT811015.1 | Severe acute respiratory syndrome coro | ···················· | ···················· | ···················· | ···················· | ···················· | ···················· |
| 24784 | MT811016.1 | Severe acute respiratory syndrome coro | ···················· | ···················· | ···················· | ···················· | ···················· | ···················· |
| 24785 | MT811017.1 | Severe acute respiratory syndrome coro | ···················· | ···················· | ···················· | ···················· | ···················· | ···················· |
| 24786 | MT811018.1 | Severe acute respiratory syndrome coro | ···················· | ···················· | ···················· | ···················· | ···················· | ···················· |
| 24787 | MT811019.1 | Severe acute respiratory syndrome coro | ···················· | ···················· | ···················· | ···················· | ···················· | ···················· |
| 24788 | MT811020.1 | Severe acute respiratory syndrome coro | ···················· | ·A·················· | ···················· | ···················· | ···················· | ···················· |
| 24789 | MT811021.1 | Severe acute respiratory syndrome coro | ···················· | ···················· | ···················· | ···················· | ···················· | ···················· |
| 24790 | MT811022.1 | Severe acute respiratory syndrome coro | ···················· | ···················· | ···················· | ···················· | ···················· | ···················· |
| 24791 | MT811162.1 | Severe acute respiratory syndrome coro | ···················· | ···················· | ···················· | ···················· | ···················· | ···················· |
| 24792 | MT811164.1 | Severe acute respiratory syndrome coro | ···················· | ···················· | ···················· | ···················· | ···················· | ···················· |
| 24793 | MT811165.1 | Severe acute respiratory syndrome coro | ···················· | ···················· | ···················· | ···················· | ···················· | ···················· |
| 24794 | MT811166.1 | Severe acute respiratory syndrome coro | ···················· | ···················· | ···················· | ···················· | ···················· | ···················· |
| 24795 | MT811169.1 | Severe acute respiratory syndrome coro | ···················· | ···················· | ···················· | ···················· | ···················· | ···················· |
| 24796 | MT811171.1 | Severe acute respiratory syndrome coro | ···················· | ···················· | ···················· | ···················· | ···················· | ···················· |
| 24797 | MT811173.1 | Severe acute respiratory syndrome coro | ···················· | ···················· | ···················· | ···················· | ···················· | ···················· |
| 24798 | MT811175.1 | Severe acute respiratory syndrome coro | ···················· | ···················· | ···················· | ···················· | ···················· | ···················· |
| 24799 | MT811176.1 | Severe acute respiratory syndrome coro | ···················· | ···················· | ···················· | ···················· | ···················· | ···················· |
| 24800 | MT811177.1 | Severe acute respiratory syndrome coro | ···················· | ···················· | ···················· | ···················· | ···················· | ···················· |
| 24801 | MT811179.1 | Severe acute respiratory syndrome coro | ···················· | ···················· | ···················· | ···················· | ···················· | ···················· |
| 24802 | MT811180.1 | Severe acute respiratory syndrome coro | ···················· | ···················· | ···················· | ···················· | ···················· | ···················· |
| 24803 | MT811181.1 | Severe acute respiratory syndrome coro | ···················· | ···················· | ···················· | ···················· | ···················· | ···················· |
| 24804 | MT811182.1 | Severe acute respiratory syndrome coro | ···················· | ···················· | ···················· | ···················· | ···················· | ···················· |
| 24805 | MT811183.1 | Severe acute respiratory syndrome coro | ···················· | ···················· | ···················· | ···················· | ···················· | ···················· |
| 24806 | MT811184.1 | Severe acute respiratory syndrome coro | ···················· | ···················· | ···················· | ···················· | ···················· | ···················· |
| 24807 | MT811185.1 | Severe acute respiratory syndrome coro | ···················· | ···················· | ···················· | ···················· | ···················· | ···················· |
| 24808 | MT811186.1 | Severe acute respiratory syndrome coro | ···················· | ···················· | ···················· | ···················· | ···················· | ···················· |
| 24809 | MT811187.1 | Severe acute respiratory syndrome coro | ···················· | ···················· | ···················· | ···················· | ···················· | ···················· |
| 24810 | MT811188.1 | Severe acute respiratory syndrome coro | ···················· | ···················· | ···················· | ···················· | ···················· | ···················· |
| 24811 | MT811190.1 | Severe acute respiratory syndrome coro | ···················· | ···················· | ···················· | ···················· | ···················· | ···················· |
| 24812 | MT811191.1 | Severe acute respiratory syndrome coro | ···················· | ···················· | ···················· | ···················· | ···················· | ···················· |
| 24813 | MT811194.1 | Severe acute respiratory syndrome coro | ···················· | ·A·················· | ···················· | ···················· | ···················· | ···················· |
| 24814 | MT811196.1 | Severe acute respiratory syndrome coro | ···················· | ·A·················· | ···················· | ···················· | ···················· | ···················· |
| 24815 | MT811197.1 | Severe acute respiratory syndrome coro | ···················· | ···················· | ···················· | ···················· | ···················· | ···················· |
| 24816 | MT811198.1 | Severe acute respiratory syndrome coro | ···················· | ···················· | ···················· | ···················· | ···················· | ···················· |
| 24817 | MT811199.1 | Severe acute respiratory syndrome coro | ···················· | ···················· | ···················· | ···················· | ···················· | ···················· |
| 24818 | MT811200.1 | Severe acute respiratory syndrome coro | ···················· | ·A·················· | ···················· | ···················· | ···················· | ···················· |
| 24819 | MT811201.1 | Severe acute respiratory syndrome coro | ···················· | ···················· | ···················· | ···················· | ···················· | ···················· |
| 24820 | MT811202.1 | Severe acute respiratory syndrome coro | ···················· | ···················· | ···················· | ···················· | ···················· | ···················· |
| 24821 | MT811203.1 | Severe acute respiratory syndrome coro | ···················· | ···················· | ···················· | ···················· | ···················· | ···················· |
| 24822 | MT811204.1 | Severe acute respiratory syndrome coro | ···················· | ···················· | ···················· | ···················· | ···················· | ···················· |
| 24823 | MT811205.1 | Severe acute respiratory syndrome coro | ···················· | ···················· | ···················· | ···················· | ···················· | ···················· |
| 24824 | MT811206.1 | Severe acute respiratory syndrome coro | ···················· | ·A·················· | ···················· | ···················· | ···················· | ···················· |
| 24825 | MT811207.1 | Severe acute respiratory syndrome coro | ···················· | ···················· | ···················· | ···················· | ···················· | ···················· |
| 24826 | MT811208.1 | Severe acute respiratory syndrome coro | ···················· | ···················· | ···················· | ···················· | ···················· | ···················· |
| 24827 | MT811209.1 | Severe acute respiratory syndrome coro | ···················· | ···················· | ···················· | ···················· | ···················· | ···················· |
| 24828 | MT811210.1 | Severe acute respiratory syndrome coro | ···················· | ···················· | ···················· | ···················· | ···················· | ···················· |
| 24829 | MT811212.1 | Severe acute respiratory syndrome coro | ···················· | ·A·················· | ···················· | ···················· | ···················· | ···················· |
| 24830 | MT811215.1 | Severe acute respiratory syndrome coro | ···················· | ···················· | ···················· | ···················· | ···················· | ···················· |
| 24831 | MT811216.1 | Severe acute respiratory syndrome coro | ···················· | ···················· | ···················· | ···················· | ···················· | ···················· |
| 24832 | MT811217.1 | Severe acute respiratory syndrome coro | ···················· | ···················· | ···················· | ···················· | ···················· | ···················· |
| 24833 | MT811218.1 | Severe acute respiratory syndrome coro | ···················· | ···················· | ···················· | ···················· | ···················· | ···················· |
| 24834 | MT811219.1 | Severe acute respiratory syndrome coro | ···················· | ·A·················· | ···················· | ···················· | ···················· | ···················· |

| | | | | | | | | |
|---|---|---|---|---|---|---|---|---|
| 24835 | MT811220.1 | \|Severe acute respiratory syndrome coro | .................... | .................... | .................... | .................... | .................... | .................... |
| 24836 | MT811221.1 | \|Severe acute respiratory syndrome coro | .................... | .A.................. | .................... | .................... | .................... | .................... |
| 24837 | MT811222.1 | \|Severe acute respiratory syndrome coro | .................... | .................... | .................... | .................... | .................... | .................... |
| 24838 | MT811224.1 | \|Severe acute respiratory syndrome coro | .................... | .A.................. | .................... | .................... | .................... | .................... |
| 24839 | MT811226.1 | \|Severe acute respiratory syndrome coro | .................... | .A.................. | .................... | .................... | .................... | .................... |
| 24840 | MT811227.1 | \|Severe acute respiratory syndrome coro | .................... | .A.................. | .................... | .................... | .................... | .................... |
| 24841 | MT811228.1 | \|Severe acute respiratory syndrome coro | .................... | .................... | .................... | .................... | .................... | .................... |
| 24842 | MT811229.1 | \|Severe acute respiratory syndrome coro | .................... | .A.................. | .................... | .................... | .................... | .................... |
| 24843 | MT811230.1 | \|Severe acute respiratory syndrome coro | .................... | .................... | .................... | .................... | .................... | .................... |
| 24844 | MT811231.1 | \|Severe acute respiratory syndrome coro | .................... | .A.................. | .................... | .................... | .................... | .................... |
| 24845 | MT811233.1 | \|Severe acute respiratory syndrome coro | .................... | .................... | .................... | .................... | .................... | .................... |
| 24846 | MT811234.1 | \|Severe acute respiratory syndrome coro | .................... | .A.................. | .................... | .................... | .................... | .................... |
| 24847 | MT811235.1 | \|Severe acute respiratory syndrome coro | .................... | .A.................. | .................... | .................... | .................... | .................... |
| 24848 | MT811236.1 | \|Severe acute respiratory syndrome coro | .................... | .................... | .................... | .................... | .................... | .................... |
| 24849 | MT811237.1 | \|Severe acute respiratory syndrome coro | .................... | .A.................. | .................... | .................... | .................... | .................... |
| 24850 | MT811238.1 | \|Severe acute respiratory syndrome coro | .................... | .A.................. | .................... | .................... | .................... | .................... |
| 24851 | MT811240.1 | \|Severe acute respiratory syndrome coro | .................... | .A.................. | .................... | .................... | .................... | .................... |
| 24852 | MT811241.1 | \|Severe acute respiratory syndrome coro | .................... | .A.................. | .................... | .................... | .................... | .................... |
| 24853 | MT811242.1 | \|Severe acute respiratory syndrome coro | .................... | .................... | .................... | .................... | .................... | .................... |
| 24854 | MT811244.1 | \|Severe acute respiratory syndrome coro | .................... | .A.................. | .................... | .................... | .................... | .................... |
| 24855 | MT811245.1 | \|Severe acute respiratory syndrome coro | .................... | .A.................. | .................... | .................... | .................... | .................... |
| 24856 | MT811246.1 | \|Severe acute respiratory syndrome coro | .................... | .A.................. | .................... | .................... | .................... | .................... |
| 24857 | MT811247.1 | \|Severe acute respiratory syndrome coro | .................... | .A.................. | .................... | .................... | .................... | .................... |
| 24858 | MT811248.1 | \|Severe acute respiratory syndrome coro | .................... | .A.................. | .................... | .................... | .................... | .................... |
| 24859 | MT811249.1 | \|Severe acute respiratory syndrome coro | .................... | .A.................. | .................... | .................... | .................... | .................... |
| 24860 | MT811250.1 | \|Severe acute respiratory syndrome coro | .................... | .................... | .................... | .................... | .................... | .................... |
| 24861 | MT811251.1 | \|Severe acute respiratory syndrome coro | .................... | .A.................. | .................... | .................... | .................... | .................... |
| 24862 | MT811252.1 | \|Severe acute respiratory syndrome coro | .................... | .A.................. | .................... | .................... | .................... | .................... |
| 24863 | MT811253.1 | \|Severe acute respiratory syndrome coro | .................... | .A.................. | .................... | .................... | .................... | .................... |
| 24864 | MT811254.1 | \|Severe acute respiratory syndrome coro | .................... | .A.................. | .................... | .................... | .................... | .................... |
| 24865 | MT811255.1 | \|Severe acute respiratory syndrome coro | .................... | .................... | .................... | .................... | .................... | .................... |
| 24866 | MT811256.1 | \|Severe acute respiratory syndrome coro | .................... | .................... | .................... | .................... | .................... | .................... |
| 24867 | MT811257.1 | \|Severe acute respiratory syndrome coro | .................... | .A.................. | .................... | .................... | .................... | .................... |
| 24868 | MT811258.1 | \|Severe acute respiratory syndrome coro | .................... | .................... | .................... | .................... | .................... | .................... |
| 24869 | MT811259.1 | \|Severe acute respiratory syndrome coro | .................... | .................... | .................... | .................... | .................... | .................... |
| 24870 | MT811261.1 | \|Severe acute respiratory syndrome coro | .................... | .................... | .................... | .................... | .................... | .................... |
| 24871 | MT811262.1 | \|Severe acute respiratory syndrome coro | .................... | .A.................. | .................... | .................... | .................... | .................... |
| 24872 | MT811263.1 | \|Severe acute respiratory syndrome coro | .................... | .................... | .................... | .................... | .................... | .................... |
| 24873 | MT811264.1 | \|Severe acute respiratory syndrome coro | .................... | .A.................. | .................... | .................... | .................... | .................... |
| 24874 | MT811265.1 | \|Severe acute respiratory syndrome coro | .................... | .A.................. | .................... | .................... | .................... | .................... |
| 24875 | MT811266.1 | \|Severe acute respiratory syndrome coro | .................... | .A.................. | .................... | .................... | .................... | .................... |
| 24876 | MT811267.1 | \|Severe acute respiratory syndrome coro | .................... | .................... | .................... | .................... | .................... | .................... |
| 24877 | MT811268.1 | \|Severe acute respiratory syndrome coro | .................... | .................... | .................... | .................... | .................... | .................... |
| 24878 | MT811269.1 | \|Severe acute respiratory syndrome coro | .................... | .A.................. | .................... | .................... | .................... | .................... |
| 24879 | MT811270.1 | \|Severe acute respiratory syndrome coro | .................... | .................... | .................... | .................... | .................... | .................... |
| 24880 | MT811271.1 | \|Severe acute respiratory syndrome coro | .................... | .................... | .................... | .................... | .................... | .................... |
| 24881 | MT811272.1 | \|Severe acute respiratory syndrome coro | .................... | .................... | .................... | .................... | .................... | .................... |
| 24882 | MT811274.1 | \|Severe acute respiratory syndrome coro | .................... | .................... | .................... | .................... | .................... | .................... |
| 24883 | MT811275.1 | \|Severe acute respiratory syndrome coro | .................... | .A.................. | .................... | .................... | .................... | .................... |
| 24884 | MT811276.1 | \|Severe acute respiratory syndrome coro | .................... | .................... | .................... | .................... | .................... | .................... |
| 24885 | MT811277.1 | \|Severe acute respiratory syndrome coro | .................... | .................... | .................... | .................... | .................... | .................... |
| 24886 | MT811278.1 | \|Severe acute respiratory syndrome coro | .................... | .A.................. | .................... | .................... | .................... | .................... |
| 24887 | MT811279.1 | \|Severe acute respiratory syndrome coro | .................... | .................... | .................... | .................... | .................... | .................... |
| 24888 | MT811280.1 | \|Severe acute respiratory syndrome coro | .................... | .................... | .................... | .................... | .................... | .................... |
| 24889 | MT811281.1 | \|Severe acute respiratory syndrome coro | .................... | .................... | .................... | .................... | .................... | .................... |
| 24890 | MT811282.1 | \|Severe acute respiratory syndrome coro | .................... | .................... | .................... | .................... | .................... | .................... |
| 24891 | MT811283.1 | \|Severe acute respiratory syndrome coro | .................... | .A.................. | .................... | .................... | .................... | .................... |
| 24892 | MT811286.1 | \|Severe acute respiratory syndrome coro | .................... | .A.................. | .................... | .................... | .................... | .................... |
| 24893 | MT811288.1 | \|Severe acute respiratory syndrome coro | .................... | .................... | .................... | .................... | .................... | .................... |
| 24894 | MT811289.1 | \|Severe acute respiratory syndrome coro | .................... | .A.................. | .................... | .................... | .................... | .................... |
| 24895 | MT811290.1 | \|Severe acute respiratory syndrome coro | .................... | .A.................. | .................... | .................... | .................... | .................... |
| 24896 | MT811292.1 | \|Severe acute respiratory syndrome coro | .................... | .................... | .................... | .................... | .................... | .................... |
| 24897 | MT811294.1 | \|Severe acute respiratory syndrome coro | .................... | .A.................. | .................... | .................... | .................... | .................... |
| 24898 | MT811295.1 | \|Severe acute respiratory syndrome coro | .................... | .A.................. | .................... | .................... | .................... | .................... |
| 24899 | MT811296.1 | \|Severe acute respiratory syndrome coro | .................... | .................... | .................... | .................... | .................... | .................... |
| 24900 | MT811297.1 | \|Severe acute respiratory syndrome coro | .................... | .................... | .................... | .................... | .................... | .................... |
| 24901 | MT811298.1 | \|Severe acute respiratory syndrome coro | .................... | .A.................. | .................... | .................... | .................... | .................... |
| 24902 | MT811299.1 | \|Severe acute respiratory syndrome coro | .................... | .................... | .................... | .................... | .................... | .................... |
| 24903 | MT811300.1 | \|Severe acute respiratory syndrome coro | .................... | .A.................. | .................... | .................... | .................... | .................... |
| 24904 | MT811301.1 | \|Severe acute respiratory syndrome coro | .................... | .A.................. | .................... | .................... | .................... | .................... |
| 24905 | MT811302.1 | \|Severe acute respiratory syndrome coro | .................... | .................... | .................... | .................... | .................... | .................... |
| 24906 | MT811303.1 | \|Severe acute respiratory syndrome coro | .................... | .A.................. | .................... | .................... | .................... | .................... |
| 24907 | MT811304.1 | \|Severe acute respiratory syndrome coro | .................... | .................... | .................... | .................... | .................... | .................... |
| 24908 | MT811305.1 | \|Severe acute respiratory syndrome coro | .................... | .................... | .................... | .................... | .................... | .................... |
| 24909 | MT811306.1 | \|Severe acute respiratory syndrome coro | .................... | .A.................. | .................... | .................... | .................... | .................... |
| 24910 | MT811307.1 | \|Severe acute respiratory syndrome coro | .................... | .................... | .................... | .................... | .................... | .................... |
| 24911 | MT811308.1 | \|Severe acute respiratory syndrome coro | .................... | .A.................. | .................... | .................... | .................... | .................... |
| 24912 | MT811309.1 | \|Severe acute respiratory syndrome coro | .................... | .................... | .................... | .................... | .................... | .................... |

24913 MT811310.1 |Severe acute respiratory syndrome coro ···················· ···················· ···················· ···················· ···················· ····················
24914 MT811311.1 |Severe acute respiratory syndrome coro ···················· ···················· ···················· ···················· ···················· ····················
24915 MT811312.1 |Severe acute respiratory syndrome coro ···················· ···················· ···················· ···················· ···················· ····················
24916 MT811313.1 |Severe acute respiratory syndrome coro ···················· ···················· ···················· ···················· ···················· ····················
24917 MT811314.1 |Severe acute respiratory syndrome coro ···················· ···················· ···················· ···················· ···················· ····················
24918 MT811315.1 |Severe acute respiratory syndrome coro ···················· ···················· ···················· ···················· ···················· ····················
24919 MT811316.1 |Severe acute respiratory syndrome coro ···················· ···················· ···················· ···················· ···················· ····················
24920 MT811317.1 |Severe acute respiratory syndrome coro ···················· ···················· ···················· ···················· ···················· ····················
24921 MT811318.1 |Severe acute respiratory syndrome coro ···················· ·A·················· ···················· ···················· ···················· ····················
24922 MT811320.1 |Severe acute respiratory syndrome coro ···················· ···················· ···················· ···················· ···················· ····················
24923 MT811321.1 |Severe acute respiratory syndrome coro ···················· ···················· ···················· ···················· ···················· ····················
24924 MT811322.1 |Severe acute respiratory syndrome coro ···················· ···················· ···················· ···················· ···················· ····················
24925 MT811323.1 |Severe acute respiratory syndrome coro ···················· ···················· ···················· ···················· ···················· ····················
24926 MT811324.1 |Severe acute respiratory syndrome coro ···················· ···················· ···················· ···················· ···················· ····················
24927 MT811325.1 |Severe acute respiratory syndrome coro ···················· ···················· ···················· ···················· ···················· ····················
24928 MT811326.1 |Severe acute respiratory syndrome coro ···················· ···················· ···················· ···················· ···················· ····················
24929 MT811328.1 |Severe acute respiratory syndrome coro ···················· ···················· ···················· ···················· ···················· ····················
24930 MT811329.1 |Severe acute respiratory syndrome coro ···················· ···················· ···················· ···················· ···················· ····················
24931 MT811330.1 |Severe acute respiratory syndrome coro ···················· ···················· ···················· ···················· ···················· ····················
24932 MT811331.1 |Severe acute respiratory syndrome coro ···················· ···················· ···················· ···················· ···················· ····················
24933 MT811332.1 |Severe acute respiratory syndrome coro ···················· ···················· ···················· ···················· ···················· ····················
24934 MT811333.1 |Severe acute respiratory syndrome coro ···················· ···················· ···················· ···················· ···················· ····················
24935 MT811334.1 |Severe acute respiratory syndrome coro ···················· ···················· ···················· ···················· ···················· ····················
24936 MT811335.1 |Severe acute respiratory syndrome coro ···················· ···················· ···················· ···················· ···················· ····················
24937 MT811336.1 |Severe acute respiratory syndrome coro ···················· ···················· ···················· ···················· ···················· ····················
24938 MT811337.1 |Severe acute respiratory syndrome coro ···················· ·A·················· ···················· ···················· ···················· ····················
24939 MT811338.1 |Severe acute respiratory syndrome coro ···················· ·A·················· ···················· ···················· ···················· ····················
24940 MT811339.1 |Severe acute respiratory syndrome coro ···················· ···················· ···················· ···················· ···················· ····················
24941 MT811340.1 |Severe acute respiratory syndrome coro ···················· ·A·················· ···················· ···················· ···················· ····················
24942 MT811341.1 |Severe acute respiratory syndrome coro ···················· ·A·················· ···················· ···················· ···················· ····················
24943 MT811343.1 |Severe acute respiratory syndrome coro ···················· ·A·················· ···················· ···················· ···················· ····················
24944 MT811344.1 |Severe acute respiratory syndrome coro ···················· ···················· ···················· ···················· ···················· ····················
24945 MT811345.1 |Severe acute respiratory syndrome coro ···················· ·A·················· ···················· ···················· ···················· ····················
24946 MT811346.1 |Severe acute respiratory syndrome coro ···················· ·A·················· ···················· ···················· ···················· ····················
24947 MT811347.1 |Severe acute respiratory syndrome coro ···················· ·A·················· ···················· ···················· ···················· ····················
24948 MT811348.1 |Severe acute respiratory syndrome coro ···················· ···················· ···················· ···················· ···················· ····················
24949 MT811349.1 |Severe acute respiratory syndrome coro ···················· ···················· ···················· ···················· ···················· ····················
24950 MT811350.1 |Severe acute respiratory syndrome coro ···················· ·A·················· ···················· ···················· ···················· ····················
24951 MT811351.1 |Severe acute respiratory syndrome coro ···················· ·A·················· ···················· ···················· ···················· ····················
24952 MT811352.1 |Severe acute respiratory syndrome coro ···················· ·A·················· ···················· ···················· ···················· ····················
24953 MT811353.1 |Severe acute respiratory syndrome coro ···················· ···················· ···················· ···················· ···················· ····················
24954 MT811354.1 |Severe acute respiratory syndrome coro ···················· ···················· ···················· ···················· ···················· ····················
24955 MT811355.1 |Severe acute respiratory syndrome coro ···················· ···················· ···················· ···················· ···················· ····················
24956 MT811356.1 |Severe acute respiratory syndrome coro ···················· ···················· ···················· ···················· ···················· ····················
24957 MT811357.1 |Severe acute respiratory syndrome coro ···················· ···················· ···················· ···················· ···················· ····················
24958 MT811358.1 |Severe acute respiratory syndrome coro ···················· ·A·················· ···················· ···················· ···················· ····················
24959 MT811359.1 |Severe acute respiratory syndrome coro ···················· ·A·················· ···················· ···················· ···················· ····················
24960 MT811360.1 |Severe acute respiratory syndrome coro ···················· ···················· ···················· ···················· ···················· ····················
24961 MT811361.1 |Severe acute respiratory syndrome coro ···················· ·A·················· ···················· ···················· ···················· ····················
24962 MT811362.1 |Severe acute respiratory syndrome coro ···················· ···················· ···················· ···················· ···················· ····················
24963 MT811363.1 |Severe acute respiratory syndrome coro ···················· ···················· ···················· ···················· ···················· ····················
24964 MT811364.1 |Severe acute respiratory syndrome coro ···················· ·A·················· ···················· ···················· ···················· ····················
24965 MT811365.1 |Severe acute respiratory syndrome coro ···················· ···················· ···················· ···················· ···················· ····················
24966 MT811366.1 |Severe acute respiratory syndrome coro ···················· ···················· ···················· ···················· ···················· ····················
24967 MT811367.1 |Severe acute respiratory syndrome coro ···················· ···················· ···················· ···················· ···················· ····················
24968 MT811368.1 |Severe acute respiratory syndrome coro ···················· ···················· ···················· ···················· ···················· ····················
24969 MT811369.1 |Severe acute respiratory syndrome coro ···················· ···················· ···················· ···················· ···················· ····················
24970 MT811371.1 |Severe acute respiratory syndrome coro ···················· ···················· ···················· ···················· ···················· ····················
24971 MT811372.1 |Severe acute respiratory syndrome coro ···················· ···················· ···················· ···················· ···················· ····················
24972 MT811374.1 |Severe acute respiratory syndrome coro ···················· ···················· ···················· ···················· ···················· ····················
24973 MT811375.1 |Severe acute respiratory syndrome coro ···················· ···················· ···················· ···················· ···················· ····················
24974 MT811378.1 |Severe acute respiratory syndrome coro ···················· ···················· ···················· ···················· ···················· ····················
24975 MT811380.1 |Severe acute respiratory syndrome coro ···················· ···················· ···················· ···················· ···················· ····················
24976 MT811382.1 |Severe acute respiratory syndrome coro ···················· ···················· ···················· ···················· ···················· ····················
24977 MT811383.1 |Severe acute respiratory syndrome coro ···················· ···················· ···················· ···················· ···················· ····················
24978 MT811384.1 |Severe acute respiratory syndrome coro ···················· ···················· ···················· ···················· ···················· ····················
24979 MT811385.1 |Severe acute respiratory syndrome coro ···················· ···················· ···················· ···················· ···················· ····················
24980 MT811386.1 |Severe acute respiratory syndrome coro ···················· ···················· ···················· ···················· ···················· ····················
24981 MT811387.1 |Severe acute respiratory syndrome coro ···················· ···················· ···················· ···················· ···················· ····················
24982 MT811388.1 |Severe acute respiratory syndrome coro ···················· ···················· ···················· ···················· ···················· ····················
24983 MT811389.1 |Severe acute respiratory syndrome coro ···················· ···················· ···················· ···················· ···················· ····················
24984 MT811390.1 |Severe acute respiratory syndrome coro ···················· ···················· ···················· ···················· ···················· ····················
24985 MT811392.1 |Severe acute respiratory syndrome coro ···················· ···················· ···················· ···················· ···················· ····················
24986 MT811393.1 |Severe acute respiratory syndrome coro ···················· ···················· ···················· ···················· ···················· ····················
24987 MT811394.1 |Severe acute respiratory syndrome coro ···················· ···················· ···················· ···················· ···················· ····················
24988 MT811395.1 |Severe acute respiratory syndrome coro ···················· ···················· ···················· ···················· ···················· ····················
24989 MT811397.1 |Severe acute respiratory syndrome coro ···················· ···················· ···················· ···················· ···················· ····················
24990 MT811398.1 |Severe acute respiratory syndrome coro ···················· ···················· ···················· ···················· ···················· ····················

| | | | | | | | | |
|---|---|---|---|---|---|---|---|---|
| 24991 | MT811399.1 | Severe acute respiratory syndrome coro | ···················· | ···················· | ···················· | ···················· | ···················· | ···················· |
| 24992 | MT811400.1 | Severe acute respiratory syndrome coro | ···················· | ···················· | ···················· | ···················· | ···················· | ···················· |
| 24993 | MT811401.1 | Severe acute respiratory syndrome coro | ···················· | ···················· | ···················· | ···················· | ···················· | ···················· |
| 24994 | MT811403.1 | Severe acute respiratory syndrome coro | ···················· | ···················· | ···················· | ···················· | ···················· | ···················· |
| 24995 | MT811404.1 | Severe acute respiratory syndrome coro | ···················· | ···················· | ···················· | ···················· | ···················· | ···················· |
| 24996 | MT811405.1 | Severe acute respiratory syndrome coro | ···················· | ···················· | ···················· | ···················· | ···················· | ···················· |
| 24997 | MT811406.1 | Severe acute respiratory syndrome coro | ···················· | ·A·················· | ···················· | ···················· | ···················· | ···················· |
| 24998 | MT811407.1 | Severe acute respiratory syndrome coro | ···················· | ·A·················· | ···················· | ···················· | ···················· | ···················· |
| 24999 | MT811408.1 | Severe acute respiratory syndrome coro | ···················· | ···················· | ···················· | ···················· | ···················· | ···················· |
| 25000 | MT811409.1 | Severe acute respiratory syndrome coro | ···················· | ···················· | ···················· | ···················· | ···················· | ···················· |
| 25001 | MT811410.1 | Severe acute respiratory syndrome coro | ···················· | ·A·················· | ···················· | ···················· | ···················· | ···················· |
| 25002 | MT811411.1 | Severe acute respiratory syndrome coro | ···················· | ·A·················· | ···················· | ···················· | ···················· | ···················· |
| 25003 | MT811412.1 | Severe acute respiratory syndrome coro | ···················· | ·A·················· | ···················· | ···················· | ···················· | ···················· |
| 25004 | MT811413.1 | Severe acute respiratory syndrome coro | ···················· | ·A·················· | ···················· | ···················· | ···················· | ···················· |
| 25005 | MT811414.1 | Severe acute respiratory syndrome coro | ···················· | ···················· | ···················· | ···················· | ···················· | ···················· |
| 25006 | MT811416.1 | Severe acute respiratory syndrome coro | ···················· | ·A·················· | ···················· | ···················· | ···················· | ···················· |
| 25007 | MT811420.1 | Severe acute respiratory syndrome coro | ···················· | ·A·················· | ···················· | ···················· | ···················· | ···················· |
| 25008 | MT811421.1 | Severe acute respiratory syndrome coro | ···················· | ·A·················· | ···················· | ···················· | ···················· | ···················· |
| 25009 | MT811422.1 | Severe acute respiratory syndrome coro | ···················· | ·A·················· | ···················· | ···················· | ···················· | ···················· |
| 25010 | MT811423.1 | Severe acute respiratory syndrome coro | ···················· | ·A·················· | ···················· | ···················· | ···················· | ···················· |
| 25011 | MT811424.1 | Severe acute respiratory syndrome coro | ···················· | ·A·················· | ···················· | ···················· | ···················· | ···················· |
| 25012 | MT811426.1 | Severe acute respiratory syndrome coro | ···················· | ···················· | ···················· | ···················· | ···················· | ···················· |
| 25013 | MT811428.1 | Severe acute respiratory syndrome coro | ···················· | ···················· | ···················· | ···················· | ···················· | ···················· |
| 25014 | MT811429.1 | Severe acute respiratory syndrome coro | ···················· | ···················· | ···················· | ···················· | ···················· | ···················· |
| 25015 | MT811430.1 | Severe acute respiratory syndrome coro | ···················· | ·A·················· | ···················· | ···················· | ···················· | ···················· |
| 25016 | MT811432.1 | Severe acute respiratory syndrome coro | ···················· | ···················· | ···················· | ···················· | ···················· | ···················· |
| 25017 | MT811433.1 | Severe acute respiratory syndrome coro | ···················· | ···················· | ···················· | ···················· | ···················· | ···················· |
| 25018 | MT811434.1 | Severe acute respiratory syndrome coro | ···················· | ···················· | ···················· | ···················· | ···················· | ···················· |
| 25019 | MT811435.1 | Severe acute respiratory syndrome coro | ···················· | ···················· | ···················· | ···················· | ···················· | ···················· |
| 25020 | MT811436.1 | Severe acute respiratory syndrome coro | ···················· | ···················· | ···················· | ···················· | ···················· | ···················· |
| 25021 | MT811437.1 | Severe acute respiratory syndrome coro | ···················· | ···················· | ···················· | ···················· | ···················· | ···················· |
| 25022 | MT811438.1 | Severe acute respiratory syndrome coro | ···················· | ···················· | ···················· | ···················· | ···················· | ···················· |
| 25023 | MT811439.1 | Severe acute respiratory syndrome coro | ···················· | ···················· | ···················· | ···················· | ···················· | ···················· |
| 25024 | MT811440.1 | Severe acute respiratory syndrome coro | ···················· | ···················· | ···················· | ···················· | ···················· | ···················· |
| 25025 | MT811441.1 | Severe acute respiratory syndrome coro | ···················· | ···················· | ···················· | ···················· | ···················· | ···················· |
| 25026 | MT811442.1 | Severe acute respiratory syndrome coro | ···················· | ···················· | ···················· | ···················· | ···················· | ···················· |
| 25027 | MT811443.1 | Severe acute respiratory syndrome coro | ···················· | ···················· | ···················· | ···················· | ···················· | ···················· |
| 25028 | MT811445.1 | Severe acute respiratory syndrome coro | ···················· | ···················· | ···················· | ···················· | ···················· | ···················· |
| 25029 | MT811446.1 | Severe acute respiratory syndrome coro | ···················· | ···················· | ···················· | ···················· | ···················· | ···················· |
| 25030 | MT811447.1 | Severe acute respiratory syndrome coro | ···················· | ·A·················· | ···················· | ···················· | ···················· | ···················· |
| 25031 | MT811448.1 | Severe acute respiratory syndrome coro | ···················· | ···················· | ···················· | ···················· | ···················· | ···················· |
| 25032 | MT811451.1 | Severe acute respiratory syndrome coro | ···················· | ···················· | ···················· | ···················· | ···················· | ···················· |
| 25033 | MT811453.1 | Severe acute respiratory syndrome coro | ···················· | ···················· | ···················· | ···················· | ···················· | ···················· |
| 25034 | MT811454.1 | Severe acute respiratory syndrome coro | ···················· | ···················· | ···················· | ···················· | ···················· | ···················· |
| 25035 | MT811456.1 | Severe acute respiratory syndrome coro | ···················· | ·A·················· | ···················· | ···················· | ···················· | ···················· |
| 25036 | MT811458.1 | Severe acute respiratory syndrome coro | ···················· | ·A·················· | ···················· | ···················· | ···················· | ···················· |
| 25037 | MT811459.1 | Severe acute respiratory syndrome coro | ···················· | ·A·················· | ···················· | ···················· | ···················· | ···················· |
| 25038 | MT811460.1 | Severe acute respiratory syndrome coro | ···················· | ·A·················· | ···················· | ···················· | ···················· | ···················· |
| 25039 | MT811461.1 | Severe acute respiratory syndrome coro | ···················· | ·A·················· | ···················· | ···················· | ···················· | ···················· |
| 25040 | MT811463.1 | Severe acute respiratory syndrome coro | ···················· | ···················· | ···················· | ···················· | ···················· | ···················· |
| 25041 | MT811464.1 | Severe acute respiratory syndrome coro | ···················· | ·A·················· | ···················· | ···················· | ···················· | ···················· |
| 25042 | MT811465.1 | Severe acute respiratory syndrome coro | ···················· | ·A·················· | ···················· | ···················· | ···················· | ···················· |
| 25043 | MT811466.1 | Severe acute respiratory syndrome coro | ···················· | ·A·················· | ···················· | ···················· | ···················· | ···················· |
| 25044 | MT811467.1 | Severe acute respiratory syndrome coro | ···················· | ···················· | ···················· | ···················· | ···················· | ···················· |
| 25045 | MT811468.1 | Severe acute respiratory syndrome coro | ···················· | ···················· | ···················· | ···················· | ···················· | ···················· |
| 25046 | MT811469.1 | Severe acute respiratory syndrome coro | ···················· | ·A·················· | ···················· | ···················· | ···················· | ···················· |
| 25047 | MT811470.1 | Severe acute respiratory syndrome coro | ···················· | ···················· | ···················· | ···················· | ···················· | ···················· |
| 25048 | MT811471.1 | Severe acute respiratory syndrome coro | ···················· | ·A·················· | ···················· | ···················· | ···················· | ···················· |
| 25049 | MT811472.1 | Severe acute respiratory syndrome coro | ···················· | ···················· | ···················· | ···················· | ···················· | ···················· |
| 25050 | MT811473.1 | Severe acute respiratory syndrome coro | ···················· | ·A·················· | ···················· | ···················· | ···················· | ···················· |
| 25051 | MT811474.1 | Severe acute respiratory syndrome coro | ···················· | ·A·················· | ···················· | ···················· | ···················· | ···················· |
| 25052 | MT811475.1 | Severe acute respiratory syndrome coro | ···················· | ·A·················· | ···················· | ···················· | ···················· | ···················· |
| 25053 | MT811476.1 | Severe acute respiratory syndrome coro | ···················· | ·A·················· | ···················· | ···················· | ···················· | ···················· |
| 25054 | MT811477.1 | Severe acute respiratory syndrome coro | ···················· | ···················· | ···················· | ···················· | ···················· | ···················· |
| 25055 | MT811478.1 | Severe acute respiratory syndrome coro | ···················· | ·A·················· | ···················· | ···················· | ···················· | ···················· |
| 25056 | MT811479.1 | Severe acute respiratory syndrome coro | ···················· | ·A·················· | ···················· | ···················· | ···················· | ···················· |
| 25057 | MT811480.1 | Severe acute respiratory syndrome coro | ···················· | ···················· | ···················· | ···················· | ···················· | ···················· |
| 25058 | MT811481.1 | Severe acute respiratory syndrome coro | ···················· | ·A·················· | ···················· | ···················· | ···················· | ···················· |
| 25059 | MT811482.1 | Severe acute respiratory syndrome coro | ···················· | ···················· | ···················· | ···················· | ···················· | ···················· |
| 25060 | MT811483.1 | Severe acute respiratory syndrome coro | ···················· | ·A·················· | ···················· | ···················· | ···················· | ···················· |
| 25061 | MT811484.1 | Severe acute respiratory syndrome coro | ···················· | ·A·················· | ···················· | ···················· | ···················· | ···················· |
| 25062 | MT811485.1 | Severe acute respiratory syndrome coro | ···················· | ···················· | ···················· | ···················· | ···················· | ···················· |
| 25063 | MT811486.1 | Severe acute respiratory syndrome coro | ···················· | ·A·················· | ···················· | ···················· | ···················· | ···················· |
| 25064 | MT811487.1 | Severe acute respiratory syndrome coro | ···················· | ·A·················· | ···················· | ···················· | ···················· | ···················· |
| 25065 | MT811488.1 | Severe acute respiratory syndrome coro | ···················· | ·A·················· | ···················· | ···················· | ···················· | ···················· |
| 25066 | MT811489.1 | Severe acute respiratory syndrome coro | ···················· | ···················· | ···················· | ···················· | ···················· | ···················· |
| 25067 | MT811490.1 | Severe acute respiratory syndrome coro | ···················· | ···················· | ···················· | ···················· | ···················· | ···················· |
| 25068 | MT811491.1 | Severe acute respiratory syndrome coro | ···················· | ·A·················· | ···················· | ···················· | ···················· | ···················· |

25069 MT811492.1 |Severe acute respiratory syndrome coro ···················· ·A·················· ···················· ···················· ···················· ····················
25070 MT811493.1 |Severe acute respiratory syndrome coro ···················· ···················· ···················· ···················· ···················· ····················
25071 MT811494.1 |Severe acute respiratory syndrome coro ···················· ·A·················· ···················· ···················· ···················· ····················
25072 MT811495.1 |Severe acute respiratory syndrome coro ···················· ···················· ···················· ···················· ···················· ····················
25073 MT811496.1 |Severe acute respiratory syndrome coro ···················· ·A·················· ···················· ···················· ···················· ····················
25074 MT811497.1 |Severe acute respiratory syndrome coro ···················· ·A·················· ···················· ···················· ···················· ····················
25075 MT811498.1 |Severe acute respiratory syndrome coro ···················· ·A·················· ···················· ···················· ···················· ····················
25076 MT811499.1 |Severe acute respiratory syndrome coro ···················· ···················· ···················· ···················· ···················· ····················
25077 MT811500.1 |Severe acute respiratory syndrome coro ···················· ·A·················· ···················· ···················· ···················· ····················
25078 MT811501.1 |Severe acute respiratory syndrome coro ···················· ·A·················· ···················· ···················· ···················· ····················
25079 MT811502.1 |Severe acute respiratory syndrome coro ···················· ···················· ···················· ···················· ···················· ····················
25080 MT811503.1 |Severe acute respiratory syndrome coro ···················· ·A·················· ···················· ···················· ···················· ····················
25081 MT811504.1 |Severe acute respiratory syndrome coro ···················· ···················· ···················· ···················· ···················· ····················
25082 MT811505.1 |Severe acute respiratory syndrome coro ···················· ·A·················· ···················· ···················· ···················· ····················
25083 MT811506.1 |Severe acute respiratory syndrome coro ···················· ·A·················· ···················· ···················· ···················· ····················
25084 MT811507.1 |Severe acute respiratory syndrome coro ···················· ···················· ···················· ···················· ···················· ····················
25085 MT811508.1 |Severe acute respiratory syndrome coro ···················· ·A·················· ···················· ···················· ···················· ····················
25086 MT811509.1 |Severe acute respiratory syndrome coro ···················· ···················· ···················· ···················· ···················· ····················
25087 MT811510.1 |Severe acute respiratory syndrome coro ···················· ···················· ···················· ···················· ···················· ····················
25088 MT811511.1 |Severe acute respiratory syndrome coro ···················· ···················· ···················· ···················· ···················· ····················
25089 MT811512.1 |Severe acute respiratory syndrome coro ···················· ···················· ···················· ···················· ···················· ····················
25090 MT811513.1 |Severe acute respiratory syndrome coro ···················· ·A·················· ···················· ···················· ···················· ····················
25091 MT811514.1 |Severe acute respiratory syndrome coro ···················· ··C················· ···················· ···················· ···················· ····················
25092 MT811515.1 |Severe acute respiratory syndrome coro ···················· ·A·················· ···················· ···················· ···················· ····················
25093 MT811516.1 |Severe acute respiratory syndrome coro ···················· ·A·················· ···················· ···················· ···················· ····················
25094 MT811517.1 |Severe acute respiratory syndrome coro ···················· ·A·················· ···················· ···················· ···················· ····················
25095 MT811518.1 |Severe acute respiratory syndrome coro ···················· ···················· ···················· ···················· ···················· ····················
25096 MT811519.1 |Severe acute respiratory syndrome coro ········T··········· ···················· ···················· ···················· ···················· ····················
25097 MT811520.1 |Severe acute respiratory syndrome coro ···················· ·A·················· ···················· ···················· ···················· ····················
25098 MT811521.1 |Severe acute respiratory syndrome coro ···················· ···················· ···················· ···················· ···················· ····················
25099 MT811522.1 |Severe acute respiratory syndrome coro ···················· ·A·················· ···················· ···················· ···················· ····················
25100 MT811523.1 |Severe acute respiratory syndrome coro ···················· ·A·················· ···················· ···················· ···················· ····················
25101 MT811525.1 |Severe acute respiratory syndrome coro ···················· ·A·················· ···················· ···················· ···················· ····················
25102 MT811526.1 |Severe acute respiratory syndrome coro ···················· ·A·················· ···················· ···················· ···················· ····················
25103 MT811527.1 |Severe acute respiratory syndrome coro ···················· ···················· ···················· ···················· ···················· ····················
25104 MT811528.1 |Severe acute respiratory syndrome coro ···················· ···················· ···················· ···················· ···················· ····················
25105 MT811529.1 |Severe acute respiratory syndrome coro ···················· ·A·················· ···················· ···················· ···················· ····················
25106 MT811530.1 |Severe acute respiratory syndrome coro ···················· ···················· ···················· ···················· ···················· ····················
25107 MT811531.1 |Severe acute respiratory syndrome coro ···················· ···················· ···················· ···················· ···················· ····················
25108 MT811532.1 |Severe acute respiratory syndrome coro ···················· ·A·················· ···················· ···················· ···················· ····················
25109 MT811533.1 |Severe acute respiratory syndrome coro ········T··········· ·A·················· ···················· ···················· ···················· ····················
25110 MT811534.1 |Severe acute respiratory syndrome coro ···················· ·A·················· ···················· ···················· ···················· ····················
25111 MT811535.1 |Severe acute respiratory syndrome coro ···················· ···················· ···················· ···················· ···················· ····················
25112 MT811544.1 |Severe acute respiratory syndrome coro ···················· ···················· ···················· ···················· ···················· ····················
25113 MT811545.1 |Severe acute respiratory syndrome coro ···················· ···················· ···················· ···················· ···················· ····················
25114 MT811546.1 |Severe acute respiratory syndrome coro ···················· ···················· ···················· ···················· ···················· ····················
25115 MT811548.1 |Severe acute respiratory syndrome coro ···················· ···················· ···················· ···················· ···················· ····················
25116 MT811549.1 |Severe acute respiratory syndrome coro ···················· ·············A······ ···················· ···················· ···················· ····················
25117 MT811550.1 |Severe acute respiratory syndrome coro ···················· ···················· ···················· ···················· ···················· ····················
25118 MT811551.1 |Severe acute respiratory syndrome coro ···················· ···················· ···················· ···················· ···················· ····················
25119 MT811552.1 |Severe acute respiratory syndrome coro ···················· ···················· ···················· ···················· ···················· ····················
25120 MT811553.1 |Severe acute respiratory syndrome coro ···················· ···················· ···················· ···················· ···················· ····················
25121 MT811554.1 |Severe acute respiratory syndrome coro ···················· ···················· ···················· ···················· ···················· ····················
25122 MT811555.1 |Severe acute respiratory syndrome coro ···················· ···················· ···················· ···················· ···················· ····················
25123 MT811556.1 |Severe acute respiratory syndrome coro ···················· ···················· ···················· ···················· ···················· ····················
25124 MT811557.1 |Severe acute respiratory syndrome coro ···················· ···················· ···················· ···················· ···················· ····················
25125 MT811558.1 |Severe acute respiratory syndrome coro ···················· ···················· ···················· ···················· ···················· ····················
25126 MT811560.1 |Severe acute respiratory syndrome coro ···················· ···················· ···················· ···················· ···················· ····················
25127 MT811561.1 |Severe acute respiratory syndrome coro ···················· ···················· ···················· ···················· ···················· ····················
25128 MT811562.1 |Severe acute respiratory syndrome coro ···················· ···················· ···················· ···················· ···················· ····················
25129 MT811564.1 |Severe acute respiratory syndrome coro ···················· ·············A······ ···················· ···················· ···················· ····················
25130 MT811565.1 |Severe acute respiratory syndrome coro ···················· ·············A······ ···················· ···················· ···················· ····················
25131 MT811566.1 |Severe acute respiratory syndrome coro ···················· ···················· ···················· ···················· ···················· ····················
25132 MT811567.1 |Severe acute respiratory syndrome coro ···················· ·············A······ ···················· ···················· ···················· ····················
25133 MT811568.1 |Severe acute respiratory syndrome coro ···················· ···················· ···················· ···················· ···················· ····················
25134 MT811569.1 |Severe acute respiratory syndrome coro ···················· ···················· ···················· ···················· ···················· ····················
25136 MT811641.1 |Severe acute respiratory syndrome coro ···················· ···················· ···················· ···················· ···················· ····················
25137 MT811649.1 |Severe acute respiratory syndrome coro ···················· ···················· ···················· ···················· ···················· ····················
25138 MT811654.1 |Severe acute respiratory syndrome coro ···················· ···················· ···················· ···················· ···················· ····················
25139 MT811655.1 |Severe acute respiratory syndrome coro ···················· ···················· ···················· ···················· ···················· ····················
25140 MT811657.1 |Severe acute respiratory syndrome coro ···················· ···················· ···················· ···················· ···················· ····················
25141 MT811658.1 |Severe acute respiratory syndrome coro ···················· ···················· ···················· ···················· ···················· ····················
25142 MT811659.1 |Severe acute respiratory syndrome coro ···················· ···················· ···················· ···················· ···················· ····················
25143 MT811660.1 |Severe acute respiratory syndrome coro ···················· ···················· ···················· ···················· ···················· ····················
25144 MT811662.1 |Severe acute respiratory syndrome coro ···················· ···················· ···················· ···················· ···················· ····················
25145 MT811664.1 |Severe acute respiratory syndrome coro ···················· ···················· ···················· ···················· ···················· ····················
25146 MT811667.1 |Severe acute respiratory syndrome coro ···················· ···················· ···················· ···················· ···················· ····················
25147 MT811669.1 |Severe acute respiratory syndrome coro ···················· ···················· ···················· ···················· ···················· ····················

```
25148 MT811670.1 |Severe acute respiratory syndrome coro  ....................  ....................  ....................  ....................  ....................  ....................
25149 MT811671.1 |Severe acute respiratory syndrome coro  ....................  ....................  ....................  ....................  ....................  ....................
25150 MT811672.1 |Severe acute respiratory syndrome coro  ....................  ....................  ....................  ....................  ....................  ....................
25151 MT811673.1 |Severe acute respiratory syndrome coro  ....................  ....................  ....................  ....................  ....................  ....................
25152 MT811675.1 |Severe acute respiratory syndrome coro  ....................  ....................  ....................  ....................  ....................  ....................
25153 MT811676.1 |Severe acute respiratory syndrome coro  ....................  ....................  ....................  ....................  ....................  ....................
25154 MT811677.1 |Severe acute respiratory syndrome coro  ....................  ....................  ....................  ....................  ....................  ....................
25155 MT811678.1 |Severe acute respiratory syndrome coro  ....................  ....................  ....................  ....................  ....................  ....................
25156 MT811679.1 |Severe acute respiratory syndrome coro  ....................  ....................  ....................  ....................  ....................  ....................
25157 MT811680.1 |Severe acute respiratory syndrome coro  ....................  ....................  ....................  ....................  ....................  ....................
25158 MT811681.1 |Severe acute respiratory syndrome coro  ....................  ....................  ....................  ....................  ....................  ....................
25159 MT811682.1 |Severe acute respiratory syndrome coro  ....................  ....................  ....................  ....................  ....................  ....................
25160 MT811683.1 |Severe acute respiratory syndrome coro  ....................  ....................  ....................  ....................  ....................  ....................
25161 MT811688.1 |Severe acute respiratory syndrome coro  ....................  ....................  ....................  ....................  ....................  ....................
25162 MT811689.1 |Severe acute respiratory syndrome coro  ....................  ....................  ....................  ....................  ....................  ....................
25163 MT811693.1 |Severe acute respiratory syndrome coro  ....................  ....................  ....................  ....................  ....................  ....................
25164 MT811695.1 |Severe acute respiratory syndrome coro  ....................  ....................  ....................  ....................  ....................  ....................
25165 MT811696.1 |Severe acute respiratory syndrome coro  ....................  ....................  ....................  ....................  ....................  ....................
25166 MT811697.1 |Severe acute respiratory syndrome coro  ....................  ....................  ....................  ....................  ....................  ....................
25167 MT811698.1 |Severe acute respiratory syndrome coro  ....................  ....................  ....................  ....................  ....................  ....................
25168 MT811699.1 |Severe acute respiratory syndrome coro  ....................  ....................  ....................  ....................  ....................  ....................
25169 MT811700.1 |Severe acute respiratory syndrome coro  ....................  ....................  ....................  ....................  ....................  ....................
25170 MT811701.1 |Severe acute respiratory syndrome coro  ....................  ....................  ....................  ....................  ....................  ....................
25171 MT811702.1 |Severe acute respiratory syndrome coro  ....................  ....................  ....................  ....................  ....................  ....................
25172 MT811703.1 |Severe acute respiratory syndrome coro  ....................  ....................  ....................  ....................  ....................  ....................
25173 MT811705.1 |Severe acute respiratory syndrome coro  ....................  ....................  ....................  ....................  ....................  ....................
25174 MT811706.1 |Severe acute respiratory syndrome coro  ....................  ....................  ....................  ....................  ....................  ....................
25175 MT811707.1 |Severe acute respiratory syndrome coro  ....................  ....................  ....................  ....................  ....................  ....................
25176 MT811708.1 |Severe acute respiratory syndrome coro  ....................  ....................  ....................  ....................  ....................  ....................
25177 MT811709.1 |Severe acute respiratory syndrome coro  ....................  ....................  ....................  ....................  ....................  ....................
25178 MT811710.1 |Severe acute respiratory syndrome coro  ....................  ....................  ....................  ....................  ....................  ....................
25179 MT811711.1 |Severe acute respiratory syndrome coro  ....................  ....................  ....................  ....................  ....................  ....................
25180 MT811712.1 |Severe acute respiratory syndrome coro  ....................  ....................  ....................  ....................  ....................  ....................
25181 MT811713.1 |Severe acute respiratory syndrome coro  ....................  ....................  ....................  ....................  ....................  ....................
25182 MT811714.1 |Severe acute respiratory syndrome coro  ....................  ....................  ....................  ....................  ....................  ....................
25183 MT811715.1 |Severe acute respiratory syndrome coro  ....................  ....................  ....................  ....................  ....................  ....................
25184 MT811716.1 |Severe acute respiratory syndrome coro  ....................  ....................  ....................  ....................  ....................  ....................
25185 MT811718.1 |Severe acute respiratory syndrome coro  ....................  ....................  ....................  ....................  ....................  ....................
25186 MT811719.1 |Severe acute respiratory syndrome coro  ....................  ....................  ....................  ....................  ....................  ....................
25187 MT811720.1 |Severe acute respiratory syndrome coro  ....................  ....................  ....................  ....................  ....................  ....................
25188 MT811721.1 |Severe acute respiratory syndrome coro  ....................  ....................  ....................  ....................  ....................  ....................
25189 MT811722.1 |Severe acute respiratory syndrome coro  ....................  ....................  ....................  ....................  ....................  ....................
25190 MT811723.1 |Severe acute respiratory syndrome coro  ....................  ....................  ....................  ....................  ....................  ....................
25191 MT811724.1 |Severe acute respiratory syndrome coro  ....................  ....................  ....................  ....................  ....................  ....................
25192 MT811725.1 |Severe acute respiratory syndrome coro  ....................  ....................  ....................  ....................  ....................  ....................
25193 MT811726.1 |Severe acute respiratory syndrome coro  ....................  ....................  ....................  ....................  ....................  ....................
25194 MT811727.1 |Severe acute respiratory syndrome coro  ....................  ....................  ....................  ....................  ....................  ....................
25195 MT811728.1 |Severe acute respiratory syndrome coro  ....................  ....................  ....................  ....................  ....................  ....................
25196 MT811729.1 |Severe acute respiratory syndrome coro  ....................  ....................  ....................  ....................  ....................  ....................
25197 MT811731.1 |Severe acute respiratory syndrome coro  ....................  ....................  ....................  ....................  ....................  ....................
25198 MT811732.1 |Severe acute respiratory syndrome coro  ....................  ....................  ....................  ....................  ....................  ....................
25199 MT811734.1 |Severe acute respiratory syndrome coro  ....................  ....................  ....................  ....................  ....................  ....................
25200 MT811737.1 |Severe acute respiratory syndrome coro  ....................  ....................  ....................  ....................  ....................  ....................
25201 MT811738.1 |Severe acute respiratory syndrome coro  ....................  ....................  ....................  ....................  ....................  ....................
25202 MT811740.1 |Severe acute respiratory syndrome coro  ....................  ....................  ....................  ....................  ....................  ....................
25203 MT811741.1 |Severe acute respiratory syndrome coro  ....................  ....................  ....................  ....................  ....................  ....................
25204 MT811743.1 |Severe acute respiratory syndrome coro  ....................  ....................  ....................  ....................  ....................  ....................
25205 MT811744.1 |Severe acute respiratory syndrome coro  ....................  ....................  ....................  ....................  ....................  ....................
25206 MT811745.1 |Severe acute respiratory syndrome coro  ....................  ....................  ....................  ....................  ....................  ....................
25207 MT811747.1 |Severe acute respiratory syndrome coro  ....................  ....................  ....................  ....................  ....................  ....................
25208 MT811748.1 |Severe acute respiratory syndrome coro  ....................  ....................  ....................  ....................  ....................  ....................
25209 MT811749.1 |Severe acute respiratory syndrome coro  ....................  ....................  ....................  ....................  ....................  ....................
25210 MT811751.1 |Severe acute respiratory syndrome coro  ....................  ....................  ....................  ....................  ....................  ....................
25211 MT811752.1 |Severe acute respiratory syndrome coro  ....................  ....................  ....................  ....................  ....................  ....................
25212 MT811753.1 |Severe acute respiratory syndrome coro  ....................  ....................  ....................  ....................  ....................  ....................
25213 MT811755.1 |Severe acute respiratory syndrome coro  ....................  ....................  ....................  ....................  ....................  ....................
25214 MT811758.1 |Severe acute respiratory syndrome coro  ....................  ....................  ....................  ....................  ....................  ....................
25215 MT814636.1 |Severe acute respiratory syndrome coro  ....................  ....................  ....................  ....................  ....................  ....................
25216 MT814698.1 |Severe acute respiratory syndrome coro  ....................  ....................  ....................  ....................  ....................  ....................
25217 MT806104.1 |Severe acute respiratory syndrome coro  ....................  ....................  ....................  ....................  ....................  ....................
25218 MT806176.1 |Severe acute respiratory syndrome coro  ....................  ....................  ....................  ....................  ....................  ....................
25219 MT806191.1 |Severe acute respiratory syndrome coro  ....................  ....................  ....................  ....................  ....................  ....................
25220 MT806192.1 |Severe acute respiratory syndrome coro  ....................  ....................  ....................  ....................  ....................  ...A................
25221 MT806756.1 |Severe acute respiratory syndrome coro  ....................  ....................  ....................  ....................  ....................  ....................
25222 MT806757.1 |Severe acute respiratory syndrome coro  ....................  ....................  ....................  ....................  ....................  ....................
25223 MT806758.1 |Severe acute respiratory syndrome coro  ....................  ....................  ....................  ....................  ....................  ....................
25224 MT806759.1 |Severe acute respiratory syndrome coro  ....................  ....................  ....................  ....................  ....................  ....................
25225 MT806760.1 |Severe acute respiratory syndrome coro  ....................  ....................  ....................  ....................  ....................  ....................
```

```
25226 MT806761.1 |Severe acute respiratory syndrome coro  ....................  ....................  ....................  ....................  ....................  ....................
25227 MT806762.1 |Severe acute respiratory syndrome coro  ....................  ....................  ....................  ....................  ....................  ....................
25228 MT806763.1 |Severe acute respiratory syndrome coro  ....................  ....................  ....................  ....................  ....................  ....................
25229 MT806764.1 |Severe acute respiratory syndrome coro  ....................  ....................  ....................  ....................  ....................  ....................
25230 MT806765.1 |Severe acute respiratory syndrome coro  ....................  ....................  ....................  ....................  ....................  ....................
25231 MT806766.1 |Severe acute respiratory syndrome coro  ....................  ....................  ....................  ....................  ....................  ....................
25232 MT806767.1 |Severe acute respiratory syndrome coro  ....................  ....................  ....................  ....................  ....................  ....................
25233 MT806768.1 |Severe acute respiratory syndrome coro  ....................  ....................  ....................  ....................  ....................  ....................
25234 MT806769.1 |Severe acute respiratory syndrome coro  ....................  ....................  ....................  ....................  ....................  ....................
25235 MT806770.1 |Severe acute respiratory syndrome coro  ....................  ....................  ....................  ....................  ....................  ....................
25236 MT806771.1 |Severe acute respiratory syndrome coro  ....................  ....................  ....................  ....................  ....................  ....................
25237 MT806772.1 |Severe acute respiratory syndrome coro  ....................  ....................  ....................  ....................  ....................  ....................
25238 MT806773.1 |Severe acute respiratory syndrome coro  ....................  ....................  ....................  ....................  ....................  ....................
25239 MT806774.1 |Severe acute respiratory syndrome coro  ....................  ....................  ....................  ....................  ....................  ....................
25240 MT806775.1 |Severe acute respiratory syndrome coro  ....................  ....................  ....................  ....................  ....................  ....................
25241 MT806776.1 |Severe acute respiratory syndrome coro  ....................  ....................  ....................  ....................  ....................  ....................
25242 MT806777.1 |Severe acute respiratory syndrome coro  ....................  ....A...............  ....................  ....................  ....................  ....................
25243 MT806778.1 |Severe acute respiratory syndrome coro  ....................  ....................  ....................  ....................  ....................  ....................
25244 MT806779.1 |Severe acute respiratory syndrome coro  ....................  .........A..........  ....................  ....................  ....................  ....................
25245 MT806780.1 |Severe acute respiratory syndrome coro  ....................  ....................  ....................  ....................  ....................  ....................
25246 MT806781.1 |Severe acute respiratory syndrome coro  ....................  ....................  ....................  ....................  ....................  ....................
25247 MT806782.1 |Severe acute respiratory syndrome coro  ....................  ....................  ....................  ....................  ....................  ....................
25248 MT806783.1 |Severe acute respiratory syndrome coro  ....................  ....................  ....................  ....................  ....................  ....................
25249 MT806785.1 |Severe acute respiratory syndrome coro  ....................  ....................  ....................  ....................  ....................  ....................
25250 MT806786.1 |Severe acute respiratory syndrome coro  ....................  ....................  ....................  ....................  ....................  ....................
25251 MT806787.1 |Severe acute respiratory syndrome coro  ....................  ....................  ....................  ....................  ....................  ....................
25252 MT806788.1 |Severe acute respiratory syndrome coro  ....................  ....................  ....................  ....................  ....................  ....................
25253 MT806789.1 |Severe acute respiratory syndrome coro  ....................  ....................  ....................  ....................  ....................  ....................
25254 MT806790.1 |Severe acute respiratory syndrome coro  ....................  ....................  ....................  ....................  ....................  ....................
25255 MT806791.1 |Severe acute respiratory syndrome coro  ....................  ....................  ....................  ....................  ....................  ....................
25256 MT806792.1 |Severe acute respiratory syndrome coro  ....................  ....................  ....................  ....................  ....................  ....................
25257 MT806793.1 |Severe acute respiratory syndrome coro  ....................  ....................  ....................  ....................  ....................  ....................
25258 MT806794.1 |Severe acute respiratory syndrome coro  ....................  ....................  ....................  ....................  ....................  ....................
25259 MT806795.1 |Severe acute respiratory syndrome coro  ....................  ....................  ....................  ....................  ....................  ....................
25260 MT806796.1 |Severe acute respiratory syndrome coro  ....................  ....................  ....................  ....................  ....................  ....................
25261 MT806797.1 |Severe acute respiratory syndrome coro  ....................  ....................  ....................  ....................  ....................  ....................
25262 MT806798.1 |Severe acute respiratory syndrome coro  ....................  ....................  ....................  ....................  ....................  ....................
25263 MT806799.1 |Severe acute respiratory syndrome coro  ....................  ....................  ....................  ....................  ....................  ....................
25264 MT806800.1 |Severe acute respiratory syndrome coro  ....................  ....................  ....................  ....................  ....................  ....................
25265 MT806801.1 |Severe acute respiratory syndrome coro  ....................  ....................  ....................  ....................  ....................  ....................
25266 MT806802.1 |Severe acute respiratory syndrome coro  ....................  ....................  ....................  ....................  ....................  ....................
25267 MT806803.1 |Severe acute respiratory syndrome coro  ....................  ....................  ....................  ....................  ....................  ....................
25268 MT806804.1 |Severe acute respiratory syndrome coro  ....................  ....................  ....................  ....................  ....................  ....................
25269 MT806805.1 |Severe acute respiratory syndrome coro  ....................  ....................  ....................  ....................  ....................  ....................
25270 MT806806.1 |Severe acute respiratory syndrome coro  ....................  ....................  ....................  ....................  ....................  ....................
25271 MT806807.1 |Severe acute respiratory syndrome coro  ....................  ....................  ....................  ....................  ....................  ....................
25272 MT806808.1 |Severe acute respiratory syndrome coro  ....................  ....................  ....................  ....................  ....................  ....................
25273 MT806809.1 |Severe acute respiratory syndrome coro  ....................  ....................  ....................  ....................  ....................  ....................
25274 MT806884.1 |Severe acute respiratory syndrome coro  ....................  ....................  ....................  ....................  ....................  ....................
25275 MT807936.1 |Severe acute respiratory syndrome coro  ....................  ....................  ....................  ....................  ....................  ....................
25276 MT808071.1 |Severe acute respiratory syndrome coro  ....................  ....................  ....................  ....................  ....................  ....................
25277 MT799967.1 |Severe acute respiratory syndrome coro  ....................  ....................  ....................  ....................  ....................  ...A................
25278 MT799969.1 |Severe acute respiratory syndrome coro  ....................  ....................  ....................  ....................  ....................  ....................
25279 MT799970.1 |Severe acute respiratory syndrome coro  ....................  .A..................  ....................  ....................  ....................  ...A................
25280 MT799972.1 |Severe acute respiratory syndrome coro  ....................  .A..................  ....................  ....................  ....................  ...A................
25281 MT799974.1 |Severe acute respiratory syndrome coro  ....................  ....................  ....................  ....................  ....................  ....................
25282 MT799976.1 |Severe acute respiratory syndrome coro  ....................  .A..................  ....................  ....................  ....................  ...A................
25283 MT799991.1 |Severe acute respiratory syndrome coro  ....................  ....................  ....................  ....................  ....................  ....................
25284 MT799998.1 |Severe acute respiratory syndrome coro  ....................  ....................  ....................  ....................  ....................  ....................
25285 MT800002.1 |Severe acute respiratory syndrome coro  ....................  ....................  ....................  ....................  ....................  ....................
25286 MT800005.1 |Severe acute respiratory syndrome coro  ....................  ....................  ....................  ....................  ....................  ....................
25287 MT800010.1 |Severe acute respiratory syndrome coro  ....................  ....................  ....................  ....................  ....................  ....................
25288 MT800013.1 |Severe acute respiratory syndrome coro  ....................  ....................  ....................  ....................  ....................  ....................
25289 MT800014.1 |Severe acute respiratory syndrome coro  ....................  ....................  ....................  ....................  ....................  ....................
25290 MT800015.1 |Severe acute respiratory syndrome coro  ....................  ....................  ....................  ....................  ....................  ....................
25291 MT800016.1 |Severe acute respiratory syndrome coro  ....................  ....................  ....................  ....................  ....................  ....................
25292 MT800021.1 |Severe acute respiratory syndrome coro  ....................  ....................  ....................  ....................  ....................  ....................
25293 MT800023.1 |Severe acute respiratory syndrome coro  ....................  ......A.............  ....................  ....................  ....................  ....................
25294 MT800024.1 |Severe acute respiratory syndrome coro  ....................  ....................  ....................  ....................  ....................  ....................
25295 MT800029.1 |Severe acute respiratory syndrome coro  ....................  ....................  ....................  ....................  ....................  ....................
25296 MT800030.1 |Severe acute respiratory syndrome coro  ....................  ....................  ....................  ....................  ....................  ....................
25297 MT800031.1 |Severe acute respiratory syndrome coro  ....................  ....................  ....................  ....................  ....................  ....................
25298 MT800035.1 |Severe acute respiratory syndrome coro  ....................  ....................  ....................  ....................  ....................  ....................
25299 MT800036.1 |Severe acute respiratory syndrome coro  ....................  ....................  ....................  ....................  ....................  ....................
25300 MT800037.1 |Severe acute respiratory syndrome coro  ....................  ....................  ....................  ....................  ....................  ....................
25301 MT800041.1 |Severe acute respiratory syndrome coro  ....................  ....................  ....................  ....................  ....................  ....................
25302 MT800043.1 |Severe acute respiratory syndrome coro  ....................  ....................  ....................  ....................  ....................  ...A................
25303 MT800045.1 |Severe acute respiratory syndrome coro  ....................  ....................  ....................  ....................  ....................  ....................
```

| | | | | | | | | |
|---|---|---|---|---|---|---|---|---|
| 25304 | MT800047.1 |Severe acute respiratory syndrome coro | ···················· | ···················· | ···················· | ···················· | ···················· | ···················· |
| 25305 | MT800050.1 |Severe acute respiratory syndrome coro | ···················· | ···················· | ···················· | ···················· | ···················· | ···················· |
| 25306 | MT800105.1 |Severe acute respiratory syndrome coro | ···················· | ···················· | ···················· | ···················· | ···················· | ···················· |
| 25307 | MT800282.1 |Severe acute respiratory syndrome coro | ···················· | ···················· | ···················· | ···················· | ···················· | ···················· |
| 25308 | MT800284.1 |Severe acute respiratory syndrome coro | ···················· | ···················· | ···················· | ···················· | ···················· | ···················· |
| 25309 | MT800505.1 |Severe acute respiratory syndrome coro | ···················· | ·A·················· | ···················· | ···················· | ···················· | ··A················· |
| 25310 | MT800515.1 |Severe acute respiratory syndrome coro | ···················· | ···················· | ···················· | ···················· | ···················· | ···················· |
| 25311 | MT800542.1 |Severe acute respiratory syndrome coro | ···················· | ···················· | ···················· | ···················· | ···················· | ···················· |
| 25312 | MT800753.1 |Severe acute respiratory syndrome coro | ···················· | ···················· | ···················· | ···················· | ···················· | ···················· |
| 25313 | MT800758.1 |Severe acute respiratory syndrome coro | ···················· | ···················· | ···················· | ···················· | ···················· | ··A················· |
| 25314 | MT800794.1 |Severe acute respiratory syndrome coro | ···················· | ···················· | ···················· | ···················· | ···················· | ···················· |
| 25315 | MT800816.1 |Severe acute respiratory syndrome coro | ···················· | ·A·················· | ···················· | ···················· | ···················· | ··A················· |
| 25316 | MT800818.1 |Severe acute respiratory syndrome coro | ···················· | ·A·················· | ···················· | ···················· | ···················· | ··A················· |
| 25317 | MT800820.1 |Severe acute respiratory syndrome coro | ···················· | ·A·················· | ···················· | ···················· | ···················· | ··A················· |
| 25318 | MT800845.1 |Severe acute respiratory syndrome coro | ···················· | ···················· | ···················· | ···················· | ···················· | ···················· |
| 25319 | MT800873.1 |Severe acute respiratory syndrome coro | ···················· | ···················· | ···················· | ···················· | ···················· | ···················· |
| 25320 | MT800895.1 |Severe acute respiratory syndrome coro | ···················· | ···················· | ···················· | ···················· | ···················· | ···················· |
| 25321 | MT800896.1 |Severe acute respiratory syndrome coro | ···················· | ···················· | ···················· | ···················· | ···················· | ··A················· |
| 25322 | MT800923.1 |Severe acute respiratory syndrome coro | ···················· | ·A·················· | ···················· | ···················· | ···················· | ··A················· |
| 25323 | MT800924.1 |Severe acute respiratory syndrome coro | ···················· | ···················· | ···················· | ···················· | ···················· | ···················· |
| 25324 | MT800925.1 |Severe acute respiratory syndrome coro | ···················· | ·A·················· | ···················· | ···················· | ···················· | ··A················· |
| 25325 | MT800928.1 |Severe acute respiratory syndrome coro | ···················· | ·A·················· | ···················· | ···················· | ···················· | ··A················· |
| 25326 | MT800934.1 |Severe acute respiratory syndrome coro | ···················· | ···················· | ···················· | ···················· | ···················· | ··A················· |
| 25327 | MT800936.1 |Severe acute respiratory syndrome coro | ···················· | ·A·················· | ···················· | ···················· | ···················· | ··A················· |
| 25328 | MT800939.1 |Severe acute respiratory syndrome coro | ···················· | ···················· | ···················· | ···················· | ···················· | ···················· |
| 25329 | MT800940.1 |Severe acute respiratory syndrome coro | ···················· | ···················· | ···················· | ···················· | ···················· | ··A················· |
| 25330 | MT800942.1 |Severe acute respiratory syndrome coro | ···················· | ···················· | ···················· | ···················· | ···················· | ···················· |
| 25331 | MT800977.1 |Severe acute respiratory syndrome coro | ···················· | ···················· | ···················· | ···················· | ···················· | ··A················· |
| 25332 | MT800978.1 |Severe acute respiratory syndrome coro | ···················· | ·A·················· | ···················· | ···················· | ···················· | ··A················· |
| 25333 | MT800994.1 |Severe acute respiratory syndrome coro | ···················· | ·A·················· | ···················· | ···················· | ···················· | ··A················· |
| 25334 | MT800995.1 |Severe acute respiratory syndrome coro | ···················· | ···················· | ···················· | ···················· | ···················· | ··A················· |
| 25335 | MT800997.1 |Severe acute respiratory syndrome coro | ···················· | ···················· | ···················· | ···················· | ···················· | ···················· |
| 25336 | MT800999.1 |Severe acute respiratory syndrome coro | ···················· | ·A·················· | ···················· | ···················· | ···················· | ··A················· |
| 25337 | MT801000.1 |Severe acute respiratory syndrome coro | ···················· | ···················· | ···················· | ···················· | ···················· | ···················· |
| 25338 | MT801001.1 |Severe acute respiratory syndrome coro | ···················· | ···················· | ···················· | ···················· | ···················· | ···················· |
| 25339 | MT801002.1 |Severe acute respiratory syndrome coro | ···················· | ···················· | ···················· | ···················· | ···················· | ···················· |
| 25340 | MT801003.1 |Severe acute respiratory syndrome coro | ···················· | ···················· | ···················· | ···················· | ···················· | ···················· |
| 25341 | MT801004.1 |Severe acute respiratory syndrome coro | ···················· | ·A·················· | ···················· | ···················· | ···················· | ··A················· |
| 25342 | MT801006.1 |Severe acute respiratory syndrome coro | ···················· | ···················· | ···················· | ···················· | ···················· | ···················· |
| 25343 | MT801043.1 |Severe acute respiratory syndrome coro | ···················· | ···················· | ···················· | ···················· | ···················· | ···················· |
| 25344 | MT801048.1 |Severe acute respiratory syndrome coro | ···················· | ···················· | ···················· | ···················· | ···················· | ··A················· |
| 25345 | MT801051.1 |Severe acute respiratory syndrome coro | ···················· | ···················· | ···················· | ···················· | ···················· | ···················· |
| 25346 | MT252714.1 |Severe acute respiratory syndrome coro | ···················· | ···················· | ···················· | ···················· | ···················· | ···················· |
| 25347 | MT252718.1 |Severe acute respiratory syndrome coro | ···················· | ···················· | ···················· | ···················· | ···················· | ···················· |
| 25348 | MT252720.1 |Severe acute respiratory syndrome coro | ···················· | ···················· | ···················· | ···················· | ···················· | ···················· |
| 25349 | MT252722.1 |Severe acute respiratory syndrome coro | ···················· | ···················· | ···················· | ···················· | ···················· | ···················· |
| 25350 | MT252724.1 |Severe acute respiratory syndrome coro | ···················· | ···················· | ···················· | ···················· | ···················· | ···················· |
| 25351 | MT252727.1 |Severe acute respiratory syndrome coro | ···················· | ···················· | ···················· | ···················· | ···················· | ···················· |
| 25352 | MT252731.1 |Severe acute respiratory syndrome coro | ···················· | ···················· | ···················· | ···················· | ···················· | ···················· |
| 25353 | MT252732.1 |Severe acute respiratory syndrome coro | ···················· | ···················· | ···················· | ···················· | ···················· | ···················· |
| 25354 | MT252743.1 |Severe acute respiratory syndrome coro | ···················· | ···················· | ···················· | ···················· | ···················· | ···················· |
| 25355 | MT252744.1 |Severe acute respiratory syndrome coro | ···················· | ···················· | ···················· | ···················· | ···················· | ···················· |
| 25356 | MT252750.1 |Severe acute respiratory syndrome coro | ···················· | ···················· | ···················· | ···················· | ···················· | ···················· |
| 25357 | MT252752.1 |Severe acute respiratory syndrome coro | ···················· | ···················· | ···················· | ···················· | ···················· | ···················· |
| 25358 | MT252753.1 |Severe acute respiratory syndrome coro | ···················· | ···················· | ···················· | ···················· | ···················· | ···················· |
| 25359 | MT252754.1 |Severe acute respiratory syndrome coro | ···················· | ···················· | ···················· | ···················· | ···················· | ···················· |
| 25360 | MT252755.1 |Severe acute respiratory syndrome coro | ···················· | ···················· | ···················· | ···················· | ···················· | ···················· |
| 25361 | MT252759.1 |Severe acute respiratory syndrome coro | ···················· | ···················· | ···················· | ···················· | ···················· | ···················· |
| 25362 | MT252760.1 |Severe acute respiratory syndrome coro | ···················· | ···················· | ···················· | ···················· | ···················· | ···················· |
| 25363 | MT252762.1 |Severe acute respiratory syndrome coro | ···················· | ···················· | ···················· | ···················· | ···················· | ···················· |
| 25364 | MT252776.1 |Severe acute respiratory syndrome coro | ···················· | ···················· | ···················· | ···················· | ···················· | ···················· |
| 25365 | MT252777.1 |Severe acute respiratory syndrome coro | ···················· | ···················· | ···················· | ···················· | ···················· | ···················· |
| 25366 | MT252786.1 |Severe acute respiratory syndrome coro | ···················· | ···················· | ···················· | ···················· | ···················· | ···················· |
| 25367 | MT252789.1 |Severe acute respiratory syndrome coro | ···················· | ···················· | ···················· | ···················· | ···················· | ···················· |
| 25368 | MT252791.1 |Severe acute respiratory syndrome coro | ···················· | ···················· | ···················· | ···················· | ···················· | ···················· |
| 25369 | MT252796.1 |Severe acute respiratory syndrome coro | ···················· | ···················· | ···················· | ···················· | ···················· | ···················· |
| 25370 | MT252812.1 |Severe acute respiratory syndrome coro | ···················· | ···················· | ···················· | ···················· | ···················· | ···················· |
| 25371 | MT252813.1 |Severe acute respiratory syndrome coro | ···················· | ···················· | ···················· | ···················· | ···················· | ···················· |
| 25372 | MT252814.1 |Severe acute respiratory syndrome coro | ···················· | ···················· | ···················· | ···················· | ···················· | ···················· |
| 25373 | MT252815.1 |Severe acute respiratory syndrome coro | ···················· | ···················· | ···················· | ···················· | ···················· | ···················· |
| 25374 | MT252816.1 |Severe acute respiratory syndrome coro | ···················· | ···················· | ···················· | ···················· | ···················· | ···················· |
| 25375 | MT252817.1 |Severe acute respiratory syndrome coro | ···················· | ···················· | ···················· | ···················· | ···················· | ···················· |
| 25376 | MT252818.1 |Severe acute respiratory syndrome coro | ···················· | ···················· | ···················· | ···················· | ···················· | ···················· |
| 25377 | MT252819.1 |Severe acute respiratory syndrome coro | ···················· | ···················· | ···················· | ···················· | ···················· | ···················· |
| 25378 | MT252820.1 |Severe acute respiratory syndrome coro | ···················· | ···················· | ···················· | ···················· | ···················· | ···················· |
| 25379 | MT798592.1 |Severe acute respiratory syndrome coro | ···················· | ···················· | ···················· | ···················· | ···················· | ···················· |
| 25380 | MT795870.1 |Severe acute respiratory syndrome coro | ···················· | ···················· | ···················· | ···················· | ···················· | ···················· |
| 25381 | MT795871.1 |Severe acute respiratory syndrome coro | ···················· | ···················· | ···················· | ···················· | ···················· | ···················· |

25382 MT795872.1 |Severe acute respiratory syndrome coro ···················· ···················· ···················· ···················· ···················· ····················
25383 MT795873.1 |Severe acute respiratory syndrome coro ···················· ···················· ···················· ···················· ···················· ····················
25384 MT795874.1 |Severe acute respiratory syndrome coro ···················· ···················· ···················· ···················· ···················· ····················
25385 MT795875.1 |Severe acute respiratory syndrome coro ···················· ···················· ···················· ···················· ···················· ····················
25386 MT795876.1 |Severe acute respiratory syndrome coro ···················· ···················· ···················· ···················· ···················· ····················
25387 MT795878.1 |Severe acute respiratory syndrome coro ···················· ···················· ···················· ···················· ···················· ····················
25388 MT795879.1 |Severe acute respiratory syndrome coro ···················· ···················· ···················· ···················· ···················· ····················
25389 MT795880.1 |Severe acute respiratory syndrome coro ···················· ···················· ···················· ···················· ···················· ····················
25390 MT795881.1 |Severe acute respiratory syndrome coro ···················· ···················· ···················· ···················· ···················· ····················
25391 MT795882.1 |Severe acute respiratory syndrome coro ···················· ···················· ···················· ···················· ···················· ····················
25392 MT795883.1 |Severe acute respiratory syndrome coro ···················· ···················· ···················· ···················· ···················· ····················
25393 MT795884.1 |Severe acute respiratory syndrome coro ············T······· ···················· ···················· ···················· ···················· ····················
25394 MT795887.1 |Severe acute respiratory syndrome coro ···················· ···················· ···················· ···················· ···················· ····················
25395 MT795888.1 |Severe acute respiratory syndrome coro ···················· ···················· ···················· ···················· ···················· ····················
25396 MT795889.1 |Severe acute respiratory syndrome coro ···················· ···················· ···················· ···················· ···················· ····················
25397 MT795890.1 |Severe acute respiratory syndrome coro ···················· ···················· ···················· ···················· ···················· ····················
25398 MT795891.1 |Severe acute respiratory syndrome coro ···················· ···················· ···················· ···················· ···················· ····················
25399 MT795892.1 |Severe acute respiratory syndrome coro ···················· ···················· ···················· ···················· ···················· ····················
25400 MT795893.1 |Severe acute respiratory syndrome coro ···················· ···················· ···················· ···················· ···················· ····················
25401 MT795894.1 |Severe acute respiratory syndrome coro ···················· ···················· ···················· ···················· ···················· ····················
25402 MT795895.1 |Severe acute respiratory syndrome coro ···················· ···················· ···················· ···················· ···················· ····················
25403 MT795896.1 |Severe acute respiratory syndrome coro ···················· ···················· ···················· ···················· ···················· ····················
25404 MT795898.1 |Severe acute respiratory syndrome coro ···················· ···················· ···················· ···················· ···················· ····················
25405 MT795899.1 |Severe acute respiratory syndrome coro ···················· ···················· ···················· ···················· ···················· ····················
25406 MT795900.1 |Severe acute respiratory syndrome coro ···················· ···················· ···················· ···················· ···················· ····················
25407 MT795901.1 |Severe acute respiratory syndrome coro ···················· ···················· ···················· ···················· ···················· ····················
25408 MT795902.1 |Severe acute respiratory syndrome coro ···················· ···················· ···················· ···················· ···················· ····················
25409 MT795904.1 |Severe acute respiratory syndrome coro ···················· ···················· ···················· ···················· ···················· ····················
25410 MT795905.1 |Severe acute respiratory syndrome coro ···················· ···················· ···················· ···················· ···················· ····················
25411 MT795906.1 |Severe acute respiratory syndrome coro ···················· ···················· ···················· ···················· ···················· ····················
25412 MT795907.1 |Severe acute respiratory syndrome coro ···················· ···················· ···················· ···················· ···················· ····················
25413 MT795909.1 |Severe acute respiratory syndrome coro ···················· ···················· ···················· ···················· ···················· ····················
25414 MT795910.1 |Severe acute respiratory syndrome coro ···················· ···················· ···················· ···················· ···················· ····················
25415 MT795911.1 |Severe acute respiratory syndrome coro ···················· ···················· ···················· ···················· ···················· ····················
25416 MT795912.1 |Severe acute respiratory syndrome coro ···················· ···················· ···················· ···················· ···················· ····················
25417 MT795913.1 |Severe acute respiratory syndrome coro ···················· ···················· ···················· ···················· ···················· ····················
25418 MT797758.1 |Severe acute respiratory syndrome coro ···················· ·········A·········· ···················· ···················· ···················· ····················
25419 MT797759.1 |Severe acute respiratory syndrome coro ···················· ·········A·········· ···················· ···················· ···················· ····················
25420 MT797760.1 |Severe acute respiratory syndrome coro ···················· ·········A·········· ···················· ···················· ···················· ····················
25421 MT797761.1 |Severe acute respiratory syndrome coro ···················· ·········A·········· ···················· ···················· ···················· ····················
25422 MT797762.1 |Severe acute respiratory syndrome coro ···················· ·········A·········· ···················· ···················· ···················· ····················
25423 MT797764.1 |Severe acute respiratory syndrome coro ···················· ·········A·········· ···················· ···················· ···················· ····················
25424 MT797765.1 |Severe acute respiratory syndrome coro ···················· ···················· ···················· ···················· ···················· ····················
25425 MT797768.1 |Severe acute respiratory syndrome coro ···················· ···················· ···················· ···················· ···················· ····················
25426 MT797769.1 |Severe acute respiratory syndrome coro ···················· ···················· ···················· ···················· ···················· ····················
25427 MT797771.1 |Severe acute respiratory syndrome coro ···················· ·············C······ ···················· ···················· ···················· ····················
25428 MT797772.1 |Severe acute respiratory syndrome coro ···················· ·········A·········· ···················· ···················· ···················· ····················
25429 LC570961.1 |Severe acute respiratory syndrome coro ···················· ···················· ···················· ···················· ···················· ····················
25430 LC570962.1 |Severe acute respiratory syndrome coro ···················· ···················· ···················· ···················· ···················· ····················
25431 LC570963.1 |Severe acute respiratory syndrome coro ···················· ···················· ···················· ···················· ···················· ····················
25432 LC570967.1 |Severe acute respiratory syndrome coro ···················· ···················· ···················· ···················· ···················· ····················
25433 LC570968.1 |Severe acute respiratory syndrome coro ···················· ···················· ···················· ···················· ···················· ····················
25434 LC570969.1 |Severe acute respiratory syndrome coro ···················· ···················· ···················· ···················· ···················· ····················
25435 LC570970.1 |Severe acute respiratory syndrome coro ···················· ···················· ···················· ···················· ···················· ····················
25436 LC570971.1 |Severe acute respiratory syndrome coro ···················· ···················· ···················· ···················· ···················· ····················
25437 LC570972.1 |Severe acute respiratory syndrome coro ···················· ···················· ···················· ···················· ···················· ····················
25438 LC570973.1 |Severe acute respiratory syndrome coro ···················· ···················· ···················· ···················· ···················· ····················
25439 LC570974.1 |Severe acute respiratory syndrome coro ···················· ···················· ···················· ···················· ···················· ····················
25440 LC570975.1 |Severe acute respiratory syndrome coro ···················· ···················· ···················· ···················· ···················· ····················
25441 LC570976.1 |Severe acute respiratory syndrome coro ···················· ···················· ···················· ···················· ···················· ····················
25442 LC570977.1 |Severe acute respiratory syndrome coro ···················· ···················· ···················· ···················· ···················· ····················
25443 LC570978.1 |Severe acute respiratory syndrome coro ···················· ···················· ···················· ···················· ···················· ····················
25444 LC570979.1 |Severe acute respiratory syndrome coro ···················· ···················· ···················· ···················· ···················· ····················
25445 LC570980.1 |Severe acute respiratory syndrome coro ···················· ···················· ···················· ···················· ···················· ····················
25446 LC570981.1 |Severe acute respiratory syndrome coro ···················· ···················· ···················· ···················· ···················· ····················
25447 LC570982.1 |Severe acute respiratory syndrome coro ···················· ···················· ···················· ···················· ···················· ····················
25448 LC570983.1 |Severe acute respiratory syndrome coro ···················· ···················· ···················· ···················· ···················· ····················
25449 LC570984.1 |Severe acute respiratory syndrome coro ···················· ···················· ···················· ···················· ···················· ····················
25450 LC570985.1 |Severe acute respiratory syndrome coro ···················· ···················· ···················· ···················· ···················· ····················
25451 LC570986.1 |Severe acute respiratory syndrome coro ···················· ···················· ···················· ···················· ···················· ····················
25452 LC570987.1 |Severe acute respiratory syndrome coro ···················· ···················· ···················· ···················· ···················· ····················
25453 LC570988.1 |Severe acute respiratory syndrome coro ···················· ···················· ···················· ···················· ···················· ····················
25454 LC570989.1 |Severe acute respiratory syndrome coro ···················· ···················· ···················· ···················· ···················· ····················
25455 LC570990.1 |Severe acute respiratory syndrome coro ···················· ···················· ···················· ···················· ···················· ····················
25456 LC570991.1 |Severe acute respiratory syndrome coro ···················· ···················· ···················· ···················· ···················· ····················
25457 LC570992.1 |Severe acute respiratory syndrome coro ···················· ···················· ···················· ···················· ···················· ····················
25458 LC570993.1 |Severe acute respiratory syndrome coro ···················· ···················· ···················· ···················· ···················· ····················
25459 LC570994.1 |Severe acute respiratory syndrome coro ···················· ···················· ···················· ···················· ···················· ····················

| | | | | | | | | | |
|---|---|---|---|---|---|---|---|---|---|
| 25460 | LC570995.1 | \|Severe acute respiratory syndrome coro | ···················· | ···················· | ···················· | ···················· | ···················· | ···················· | |
| 25461 | LC570996.1 | \|Severe acute respiratory syndrome coro | ···················· | ···················· | ···················· | ···················· | ···················· | ···················· | |
| 25462 | LC570997.1 | \|Severe acute respiratory syndrome coro | ···················· | ···················· | ···················· | ···················· | ···················· | ···················· | |
| 25463 | LC570998.1 | \|Severe acute respiratory syndrome coro | ···················· | ···················· | ···················· | ···················· | ···················· | ···················· | |
| 25464 | LC570999.1 | \|Severe acute respiratory syndrome coro | ···················· | ···················· | ···················· | ···················· | ···················· | ···················· | |
| 25465 | LC571002.1 | \|Severe acute respiratory syndrome coro | ···················· | ···················· | ···················· | ···················· | ···················· | ···················· | |
| 25466 | LC571003.1 | \|Severe acute respiratory syndrome coro | ···················· | ···················· | ···················· | ···················· | ···················· | ···················· | |
| 25467 | LC571004.1 | \|Severe acute respiratory syndrome coro | ···················· | ···················· | ···················· | ···················· | ···················· | ···················· | |
| 25468 | LC571005.1 | \|Severe acute respiratory syndrome coro | ···················· | ···················· | ···················· | ···················· | ···················· | ···················· | |
| 25469 | LC571006.1 | \|Severe acute respiratory syndrome coro | ···················· | ···················· | ···················· | ···················· | ···················· | ···················· | |
| 25470 | LC571007.1 | \|Severe acute respiratory syndrome coro | ···················· | ···················· | ···················· | ···················· | ···················· | ···················· | |
| 25471 | LC571008.1 | \|Severe acute respiratory syndrome coro | ···················· | ···················· | ···················· | ···················· | ···················· | ···················· | |
| 25472 | LC571009.1 | \|Severe acute respiratory syndrome coro | ···················· | ···················· | ···················· | ···················· | ···················· | ···················· | |
| 25473 | LC571010.1 | \|Severe acute respiratory syndrome coro | ···················· | ···················· | ···················· | ···················· | ···················· | ···················· | |
| 25474 | LC571011.1 | \|Severe acute respiratory syndrome coro | ···················· | ···················· | ···················· | ···················· | ···················· | ···················· | |
| 25475 | LC571012.1 | \|Severe acute respiratory syndrome coro | ···················· | ···················· | ···················· | ···················· | ···················· | ···················· | |
| 25476 | LC571013.1 | \|Severe acute respiratory syndrome coro | ···················· | ···················· | ···················· | ···················· | ···················· | ···················· | |
| 25477 | LC571019.1 | \|Severe acute respiratory syndrome coro | ···················· | ···················· | ···················· | ···················· | ···················· | ···················· | |
| 25478 | LC571022.1 | \|Severe acute respiratory syndrome coro | ···················· | ···················· | ···················· | ···················· | ···················· | ···················· | |
| 25479 | LC571023.1 | \|Severe acute respiratory syndrome coro | ···················· | ···················· | ···················· | ···················· | ···················· | ···················· | |
| 25480 | LC571024.1 | \|Severe acute respiratory syndrome coro | ···················· | ···················· | ···················· | ···················· | ···················· | ···················· | |
| 25481 | LC571025.1 | \|Severe acute respiratory syndrome coro | ···················· | ···················· | ···················· | ···················· | ···················· | ···················· | |
| 25482 | LC571030.1 | \|Severe acute respiratory syndrome coro | ···················· | ···················· | ···················· | ···················· | ···················· | ···················· | |
| 25483 | LC571031.1 | \|Severe acute respiratory syndrome coro | ···················· | ···················· | ···················· | ···················· | ···················· | ···················· | |
| 25484 | LC571034.1 | \|Severe acute respiratory syndrome coro | ···················· | ···················· | ···················· | ···················· | ···················· | ···················· | |
| 25485 | LC571035.1 | \|Severe acute respiratory syndrome coro | ···················· | ···················· | ···················· | ···················· | ···················· | ···················· | |
| 25486 | LC571036.1 | \|Severe acute respiratory syndrome coro | ···················· | ···················· | ···················· | ···················· | ···················· | ···················· | |
| 25487 | LC571037.1 | \|Severe acute respiratory syndrome coro | ···················· | ···················· | ···················· | ···················· | ···················· | ···················· | |
| 25488 | LC571038.1 | \|Severe acute respiratory syndrome coro | ···················· | ···················· | ···················· | ···················· | ···················· | ···················· | |
| 25489 | LC571039.1 | \|Severe acute respiratory syndrome coro | ···················· | ···················· | ···················· | ···················· | ···················· | ···················· | |
| 25490 | LC571040.1 | \|Severe acute respiratory syndrome coro | ···················· | ···················· | ···················· | ···················· | ···················· | ···················· | |
| 25491 | LC571041.1 | \|Severe acute respiratory syndrome coro | ···················· | ···················· | ···················· | ···················· | ···················· | ···················· | |
| 25492 | MT791844.1 | \|Severe acute respiratory syndrome coro | ···················· | ···················· | ···················· | ···················· | ···················· | ···················· | |
| 25493 | MT791845.1 | \|Severe acute respiratory syndrome coro | ···················· | ···················· | ···················· | ···················· | ···················· | ···················· | |
| 25494 | MT791850.1 | \|Severe acute respiratory syndrome coro | ···················· | ···················· | ···················· | ···················· | ···················· | ···················· | |
| 25495 | MT791851.1 | \|Severe acute respiratory syndrome coro | ···················· | ···················· | ···················· | ···················· | ···················· | ···················· | |
| 25496 | MT791852.1 | \|Severe acute respiratory syndrome coro | ···················· | ···················· | ···················· | ···················· | ··················NN | ···················· | 1 |
| 25497 | MT791854.1 | \|Severe acute respiratory syndrome coro | ···················· | ···················· | ···················· | ···················· | ···················· | ···················· | |
| 25498 | MT791855.1 | \|Severe acute respiratory syndrome coro | ···················· | ···················· | ···················· | ···················· | ···················· | ···················· | |
| 25499 | MT791857.1 | \|Severe acute respiratory syndrome coro | ···················· | ···················· | ···················· | ···················· | ···················· | ···················· | |
| 25500 | MT791858.1 | \|Severe acute respiratory syndrome coro | ···················· | ···················· | ···················· | ···················· | ···················· | ···················· | |
| 25501 | MT791861.1 | \|Severe acute respiratory syndrome coro | ···················· | ···················· | ···················· | ···················· | ···················· | ···················· | |
| 25502 | MT791862.1 | \|Severe acute respiratory syndrome coro | ···················· | ···················· | ···················· | ···················· | ···················· | ···················· | |
| 25503 | MT791864.1 | \|Severe acute respiratory syndrome coro | ···················· | ···················· | ···················· | ···················· | ···················· | ···················· | |
| 25504 | MT791868.1 | \|Severe acute respiratory syndrome coro | ···················· | ···················· | ···················· | ···················· | ···················· | ···················· | |
| 25505 | MT791869.1 | \|Severe acute respiratory syndrome coro | ···················· | ···················· | ···················· | ···················· | ···················· | ···················· | |
| 25506 | MT791870.1 | \|Severe acute respiratory syndrome coro | ···················· | ···················· | ···················· | ···················· | ···················· | ···················· | |
| 25507 | MT791878.1 | \|Severe acute respiratory syndrome coro | ···················· | ···················· | ···················· | ···················· | ···················· | ···················· | |
| 25508 | MT791880.1 | \|Severe acute respiratory syndrome coro | ···················· | ···················· | ···················· | ···················· | ···················· | ···················· | |
| 25509 | MT791883.1 | \|Severe acute respiratory syndrome coro | ···················· | ···················· | ···················· | ···················· | ···················· | ···················· | |
| 25511 | MT791891.1 | \|Severe acute respiratory syndrome coro | ···················· | ···················· | ···················· | ···················· | ···················· | ···················· | |
| 25512 | MT791895.1 | \|Severe acute respiratory syndrome coro | ···················· | ···················· | ···················· | ···················· | ···················· | ···················· | |
| 25513 | MT791896.1 | \|Severe acute respiratory syndrome coro | ···················· | ···················· | ···················· | ···················· | ···················· | ···················· | |
| 25514 | MT791899.1 | \|Severe acute respiratory syndrome coro | ···················· | ···················· | ···················· | ···················· | ···················· | ···················· | |
| 25515 | MT791907.1 | \|Severe acute respiratory syndrome coro | ···················· | ···················· | ···················· | ···················· | ···················· | ···················· | |
| 25516 | MT791914.1 | \|Severe acute respiratory syndrome coro | ···················· | ···················· | ···················· | ···················· | ···················· | ···················· | |
| 25517 | MT791915.1 | \|Severe acute respiratory syndrome coro | ···················· | ···················· | ···················· | ···················· | ···················· | ···················· | |
| 25518 | MT791920.1 | \|Severe acute respiratory syndrome coro | ···················· | ···················· | ···················· | ···················· | ···················· | ···················· | |
| 25519 | MT791923.1 | \|Severe acute respiratory syndrome coro | ···················· | ···················· | ···················· | ···················· | ···················· | ···················· | |
| 25520 | MT791926.1 | \|Severe acute respiratory syndrome coro | ···················· | ···················· | ···················· | ···················· | ···················· | ···················· | |
| 25521 | MT791936.1 | \|Severe acute respiratory syndrome coro | ···················· | ···················· | ···················· | ···················· | ···················· | ···················· | |
| 25522 | MT791937.1 | \|Severe acute respiratory syndrome coro | ···················· | ···················· | ···················· | ···················· | ···················· | ···················· | |
| 25523 | MT791938.1 | \|Severe acute respiratory syndrome coro | ···················· | ···················· | ···················· | ···················· | ···················· | ···················· | |
| 25524 | MT791939.1 | \|Severe acute respiratory syndrome coro | ···················· | ···················· | ···················· | ···················· | ···················· | ···················· | |
| 25525 | MT791940.1 | \|Severe acute respiratory syndrome coro | ···················· | ···················· | ···················· | ···················· | ···················· | ···················· | |
| 25526 | MT791941.1 | \|Severe acute respiratory syndrome coro | ···················· | ···················· | ···················· | ···················· | ···················· | ···················· | |
| 25527 | MT791942.1 | \|Severe acute respiratory syndrome coro | ···················· | ···················· | ···················· | ···················· | ···················· | ···················· | |
| 25528 | MT791943.1 | \|Severe acute respiratory syndrome coro | ···················· | ···················· | ···················· | ···················· | ···················· | ···················· | |
| 25529 | MT791944.1 | \|Severe acute respiratory syndrome coro | ···················· | ···················· | ···················· | ···················· | ···················· | ···················· | |
| 25530 | MT791945.1 | \|Severe acute respiratory syndrome coro | ···················· | ···················· | ···················· | ···················· | ···················· | ···················· | |
| 25531 | MT791946.1 | \|Severe acute respiratory syndrome coro | ···················· | ···················· | ···················· | ···················· | ···················· | ···················· | |
| 25532 | MT791947.1 | \|Severe acute respiratory syndrome coro | ···················· | ···················· | ···················· | ···················· | ···················· | ···················· | |
| 25533 | MT791948.1 | \|Severe acute respiratory syndrome coro | ···················· | ···················· | ···················· | ···················· | ···················· | ···················· | |
| 25534 | MT791949.1 | \|Severe acute respiratory syndrome coro | ···················· | ···················· | ···················· | ···················· | ···················· | ···················· | |
| 25535 | MT791950.1 | \|Severe acute respiratory syndrome coro | ···················· | ···················· | ···················· | ···················· | ···················· | ···················· | |
| 25536 | MT791953.1 | \|Severe acute respiratory syndrome coro | ···················· | ···················· | ···················· | ···················· | ···················· | ···················· | |
| 25537 | MT791954.1 | \|Severe acute respiratory syndrome coro | ···················· | ···················· | ···················· | ···················· | ···················· | ···················· | |
| 25538 | MT791955.1 | \|Severe acute respiratory syndrome coro | ···················· | ···················· | ···················· | ···················· | ···················· | ···················· | |

```
25539 MT791956.1 |Severe acute respiratory syndrome coro    ....................  ....................  ....................  ....................  ....................  ....................
25540 MT791957.1 |Severe acute respiratory syndrome coro    ....................  ....................  ....................  ....................  ....................  ....................
25541 MT791958.1 |Severe acute respiratory syndrome coro    ....................  ....................  ....................  ....................  ....................  ....................
25542 MT791959.1 |Severe acute respiratory syndrome coro    ....................  ....................  ....................  ....................  ....................  ....................
25543 MT791960.1 |Severe acute respiratory syndrome coro    ....................  ....................  ....................  ....................  ....................  ....................
25544 MT791961.1 |Severe acute respiratory syndrome coro    ....................  ....................  ....................  ....................  ....................  ....................
25545 MT791962.1 |Severe acute respiratory syndrome coro    ....................  ....................  ....................  ....................  ....................  ....................
25546 MT791963.1 |Severe acute respiratory syndrome coro    ....................  ....................  ....................  ....................  ....................  ....................
25547 MT786422.1 |Severe acute respiratory syndrome coro    ....................  ....................  ....................  ....................  ....................  ....................
25548 MT786438.1 |Severe acute respiratory syndrome coro    ....................  ....................  ....................  ....................  ....................  ....................
25549 MT786442.1 |Severe acute respiratory syndrome coro    ....................  ....................  ....................  ....................  ....................  ....................
25550 MT786444.1 |Severe acute respiratory syndrome coro    ....................  ....................  ....................  ....................  ....................  ....................
25551 MT786455.1 |Severe acute respiratory syndrome coro    ....................  ....................  ....................  ....................  ....................  ....................
25552 MT786798.1 |Severe acute respiratory syndrome coro    ....................  ....................  ....................  ....................  ....................  ....................
25553 MT786799.1 |Severe acute respiratory syndrome coro    ....................  ....................  ....................  ....................  ....................  ....................
25554 MT786800.1 |Severe acute respiratory syndrome coro    ....................  ....................  ....................  ....................  ....................  ....................
25555 MT786801.1 |Severe acute respiratory syndrome coro    ....................  ....................  ....................  ....................  ....................  ....................
25556 MT786802.1 |Severe acute respiratory syndrome coro    ....................  ....................  ....................  ....................  ....................  ....................
25557 MT786804.1 |Severe acute respiratory syndrome coro    ....................  .A..................  ....................  ....................  ....................  ....................
25558 MT786809.1 |Severe acute respiratory syndrome coro    ....................  ....................  ....................  ....................  ....................  ....................
25559 MT786810.1 |Severe acute respiratory syndrome coro    ....................  ....................  ....................  ....................  ....................  ....................
25560 MT786811.1 |Severe acute respiratory syndrome coro    ....................  ....................  ....................  ....................  ....................  ....................
25561 MT786812.1 |Severe acute respiratory syndrome coro    ....................  ....................  ....................  ....................  ....................  ....................
25562 MT786813.1 |Severe acute respiratory syndrome coro    ....................  ....................  ....................  ....................  ....................  ....................
25563 MT786814.1 |Severe acute respiratory syndrome coro    ....................  ....................  ....................  ....................  ....................  ....................
25564 MT786815.1 |Severe acute respiratory syndrome coro    ....................  ....................  ....................  ....................  ....................  ....................
25565 MT786817.1 |Severe acute respiratory syndrome coro    ....................  ....................  ....................  ....................  ....................  ....................
25566 MT786818.1 |Severe acute respiratory syndrome coro    ....................  ....................  ....................  ....................  ....................  ....................
25567 MT786819.1 |Severe acute respiratory syndrome coro    ....................  ....................  ....................  ....................  ....................  ....................
25568 MT786820.1 |Severe acute respiratory syndrome coro    ....................  ....................  ....................  ....................  ....................  ....................
25569 MT786821.1 |Severe acute respiratory syndrome coro    ....................  ....................  ....................  ....................  ....................  ....................
25570 MT786822.1 |Severe acute respiratory syndrome coro    ....................  ....................  ....................  ....................  ....................  ....................
25571 MT786823.1 |Severe acute respiratory syndrome coro    ....................  ....................  ....................  ....................  ....................  ....................
25572 MT786824.1 |Severe acute respiratory syndrome coro    ....................  ....................  ....................  ....................  ....................  ....................
25573 MT786825.1 |Severe acute respiratory syndrome coro    ....................  ....................  ....................  ....................  ....................  ....................
25574 MT786826.1 |Severe acute respiratory syndrome coro    ....................  ....................  ....................  ....................  ....................  ....................
25575 MT786827.1 |Severe acute respiratory syndrome coro    ....................  ....................  ....................  ....................  ....................  ....................
25576 MT786828.1 |Severe acute respiratory syndrome coro    ....................  ....................  ....................  ....................  ....................  ....................
25577 MT786829.1 |Severe acute respiratory syndrome coro    ....................  ....................  ....................  ....................  ....................  ....................
25578 MT786830.1 |Severe acute respiratory syndrome coro    ....................  ....................  ....................  ....................  ....................  ....................
25579 MT786831.1 |Severe acute respiratory syndrome coro    ....................  ....................  ....................  ....................  ....................  ....................
25580 MT786832.1 |Severe acute respiratory syndrome coro    ....................  ....................  ....................  ....................  ....................  ....................
25581 MT786833.1 |Severe acute respiratory syndrome coro    ....................  ....................  ....................  ....................  ....................  ....................
25582 MT786834.1 |Severe acute respiratory syndrome coro    ....................  ....................  ....................  ....................  ....................  ....................
25583 MT786835.1 |Severe acute respiratory syndrome coro    ....................  ....................  ....................  ....................  ....................  ....................
25584 MT786836.1 |Severe acute respiratory syndrome coro    ....................  ....................  ....................  ....................  ....................  ....................
25585 MT786837.1 |Severe acute respiratory syndrome coro    ....................  ....................  ....................  ....................  ....................  ....................
25586 MT786838.1 |Severe acute respiratory syndrome coro    ....................  ....................  ....................  ....................  ....................  ....................
25587 MT786839.1 |Severe acute respiratory syndrome coro    ....................  ....................  ....................  ....................  ....................  ....................
25588 MT786840.1 |Severe acute respiratory syndrome coro    ....................  ....................  ....................  ....................  ....................  ....................
25589 MT786842.1 |Severe acute respiratory syndrome coro    ....................  ....................  ....................  ....................  ....................  ....................
25590 MT786844.1 |Severe acute respiratory syndrome coro    ....................  ....................  ....................  ....................  ....................  ....................
25591 MT786845.1 |Severe acute respiratory syndrome coro    ....................  ....................  ....................  ....................  ....................  ....................
25592 MT786846.1 |Severe acute respiratory syndrome coro    ....................  ....................  ....................  ....................  ....................  ....................
25593 MT786847.1 |Severe acute respiratory syndrome coro    ....................  ....................  ....................  ....................  ....................  ....................
25594 MT786848.1 |Severe acute respiratory syndrome coro    ....................  ....................  ....................  ....................  ....................  ....................
25595 MT786849.1 |Severe acute respiratory syndrome coro    ....................  ....................  ....................  ....................  ....................  ....................
25596 MT786850.1 |Severe acute respiratory syndrome coro    ....................  ....................  ....................  ....................  ....................  ....................
25597 MT786851.1 |Severe acute respiratory syndrome coro    ....................  ....................  ....................  ....................  ....................  ....................
25598 MT786859.1 |Severe acute respiratory syndrome coro    ....................  ....................  ....................  ....................  ....................  ....................
25599 MT786865.1 |Severe acute respiratory syndrome coro    ....................  ....................  ....................  ....................  ....................  ....................
25600 MT786868.1 |Severe acute respiratory syndrome coro    ....................  ....................  ....................  ....................  ....................  ....................
25601 MT786941.1 |Severe acute respiratory syndrome coro    ....................  .........A..........  ....................  ....................  ....................  ....................
25602 MT786943.1 |Severe acute respiratory syndrome coro    ....................  .........A..........  ....................  ....................  ....................  ....................
25603 MT786946.1 |Severe acute respiratory syndrome coro    ....................  .........A..........  ....................  ....................  ....................  ....................
25604 MT787473.1 |Severe acute respiratory syndrome coro    ....................  ....................  ....................  ....................  ....................  ....................
25605 MT787486.1 |Severe acute respiratory syndrome coro    ....................  ....................  ....................  ....................  ....................  ....................
25606 MT787500.1 |Severe acute respiratory syndrome coro    ....................  ....................  ....................  ....................  ....................  ....................
25607 MT787503.1 |Severe acute respiratory syndrome coro    ....................  ....................  ....................  ....................  ....................  ....................
25608 MT787553.1 |Severe acute respiratory syndrome coro    ....................  ....................  ....................  ....................  ....................  ....................
25609 MT787564.1 |Severe acute respiratory syndrome coro    ....................  ....................  ....................  ....................  ....................  ....................
25610 MT787577.1 |Severe acute respiratory syndrome coro    ....................  ....................  ....................  ....................  ....................  ....................
25611 MT787646.1 |Severe acute respiratory syndrome coro    ....................  ....................  ....................  ....................  ....................  ....................
25612 MT787647.1 |Severe acute respiratory syndrome coro    ....................  ....................  ....................  ....................  ....................  ....................
25613 MT787648.1 |Severe acute respiratory syndrome coro    ....................  ....................  ....................  ....................  ....................  ....................
25614 MT787650.1 |Severe acute respiratory syndrome coro    ....................  ....................  ....................  ....................  ....................  ....................
25615 MT787742.1 |Severe acute respiratory syndrome coro    ....................  ....................  ....................  ....................  ....................  ....................
25616 MT787745.1 |Severe acute respiratory syndrome coro    ....................  ....................  ....................  ....................  ....................  ....................
```

| | | | | | | | | |
|---|---|---|---|---|---|---|---|---|
| 25617 | MT787746.1 | \|Severe acute respiratory syndrome coro | ···················· | ···················· | ···················· | ···················· | ···················· | ···················· |
| 25618 | MT787749.1 | \|Severe acute respiratory syndrome coro | ···················· | ···················· | ···················· | ···················· | ···················· | ···················· |
| 25619 | MT789688.1 | \|Severe acute respiratory syndrome coro | ···················· | ···················· | ···················· | ···················· | ···················· | ···················· |
| 25620 | MT789690.1 | \|Severe acute respiratory syndrome coro | ···················· | ·A·················· | ···················· | ···················· | ···················· | ···················· |
| 25621 | MT789712.1 | \|Severe acute respiratory syndrome coro | ···················· | ···················· | ···················· | ···················· | ···················· | ···················· |
| 25622 | MT790522.1 | \|Severe acute respiratory syndrome coro | ···················· | ···················· | ···················· | ···················· | ···················· | ···················· |
| 25623 | MT791312.1 | \|Severe acute respiratory syndrome coro | ···················· | ···················· | ···················· | ···················· | ···················· | ···················· |
| 25624 | MT781414.1 | \|Severe acute respiratory syndrome coro | ···················· | ···················· | ···················· | ···················· | ···················· | ···················· |
| 25625 | MT781415.1 | \|Severe acute respiratory syndrome coro | ···················· | ···················· | ···················· | ···················· | ···················· | ···················· |
| 25626 | MT782348.1 | \|Severe acute respiratory syndrome coro | ···················· | ···················· | ···················· | ···················· | ···················· | ···················· |
| 25627 | MT782361.1 | \|Severe acute respiratory syndrome coro | ···················· | ···················· | ···················· | ···················· | ···················· | ···················· |
| 25628 | MT782364.1 | \|Severe acute respiratory syndrome coro | ···················· | ···················· | ···················· | ···················· | ···················· | ···················· |
| 25629 | MT782369.1 | \|Severe acute respiratory syndrome coro | ···················· | ···················· | ···················· | ···················· | ···················· | ···················· |
| 25630 | MT782370.1 | \|Severe acute respiratory syndrome coro | ···················· | ···················· | ···················· | ···················· | ···················· | ···················· |
| 25631 | MT782371.1 | \|Severe acute respiratory syndrome coro | ···················· | ···················· | ···················· | ···················· | ···················· | ···················· |
| 25632 | MT782377.1 | \|Severe acute respiratory syndrome coro | ···················· | ···················· | ···················· | ···················· | ···················· | ···················· |
| 25633 | MT782378.1 | \|Severe acute respiratory syndrome coro | ···················· | ···················· | ···················· | ···················· | ···················· | ···················· |
| 25634 | MT782381.1 | \|Severe acute respiratory syndrome coro | ···················· | ···················· | ···················· | ···················· | ···················· | ···················· |
| 25635 | MT782382.1 | \|Severe acute respiratory syndrome coro | ···················· | ···················· | ···················· | ···················· | ···················· | ···················· |
| 25636 | MT786327.1 | \|Severe acute respiratory syndrome coro | ···················· | ···················· | ···················· | ···················· | ···················· | ···················· |
| 25637 | MT786329.1 | \|Severe acute respiratory syndrome coro | ···················· | ···················· | ···················· | ···················· | ···················· | ···················· |
| 25638 | MT786331.1 | \|Severe acute respiratory syndrome coro | ···················· | ···················· | ···················· | ···················· | ···················· | ···················· |
| 25639 | MT786334.1 | \|Severe acute respiratory syndrome coro | ···················· | ···················· | ···················· | ···················· | ···················· | ···················· |
| 25640 | MT786335.1 | \|Severe acute respiratory syndrome coro | ···················· | ···················· | ···················· | ···················· | ···················· | ···················· |
| 25641 | MT786336.1 | \|Severe acute respiratory syndrome coro | ···················· | ···················· | ···················· | ···················· | ···················· | ···················· |
| 25642 | MT786337.1 | \|Severe acute respiratory syndrome coro | ···················· | ···················· | ···················· | ···················· | ···················· | ···················· |
| 25643 | MT775826.1 | \|Severe acute respiratory syndrome coro | ···················· | ···················· | ···················· | ···················· | ···················· | ···················· |
| 25644 | MT775827.1 | \|Severe acute respiratory syndrome coro | ···················· | ···················· | ···················· | ···················· | ···················· | ···················· |
| 25645 | MT775828.1 | \|Severe acute respiratory syndrome coro | ···················· | ···················· | ···················· | ···················· | ···················· | ···················· |
| 25646 | MT775829.1 | \|Severe acute respiratory syndrome coro | ···················· | ···················· | ···················· | ···················· | ···················· | ···················· |
| 25647 | MT775830.1 | \|Severe acute respiratory syndrome coro | ···················· | ···················· | ···················· | ···················· | ···················· | ···················· |
| 25648 | MT775831.1 | \|Severe acute respiratory syndrome coro | ···················· | ···················· | ···················· | ···················· | ···················· | ···················· |
| 25649 | MT775832.1 | \|Severe acute respiratory syndrome coro | ···················· | ···················· | ···················· | ···················· | ···················· | ···················· |
| 25650 | MT775833.1 | \|Severe acute respiratory syndrome coro | ···················· | ···················· | ···················· | ···················· | ···················· | ···················· |
| 25651 | MT776401.1 | \|Severe acute respiratory syndrome coro | ···················· | ···················· | ···················· | ···················· | ···················· | ···················· |
| 25652 | MT776402.1 | \|Severe acute respiratory syndrome coro | ···················· | ···················· | ···················· | ···················· | ···················· | ···················· |
| 25653 | MT776403.1 | \|Severe acute respiratory syndrome coro | ···················· | ···················· | ···················· | ···················· | ···················· | ···················· |
| 25654 | MT776404.1 | \|Severe acute respiratory syndrome coro | ···················· | ···················· | ···················· | ···················· | ···················· | ···················· |
| 25655 | MT776405.1 | \|Severe acute respiratory syndrome coro | ···················· | ···················· | ···················· | ···················· | ···················· | ···················· |
| 25656 | MT776406.1 | \|Severe acute respiratory syndrome coro | ···················· | ···················· | ···················· | ···················· | ···················· | ···················· |
| 25657 | MT776407.1 | \|Severe acute respiratory syndrome coro | ···················· | ···················· | ···················· | ···················· | ···················· | ···················· |
| 25658 | MT776408.1 | \|Severe acute respiratory syndrome coro | ···················· | ···················· | ···················· | ···················· | ···················· | ···················· |
| 25659 | MT776409.1 | \|Severe acute respiratory syndrome coro | ···················· | ···················· | ···················· | ···················· | ···················· | ···················· |
| 25660 | MT776410.1 | \|Severe acute respiratory syndrome coro | ···················· | ···················· | ···················· | ···················· | ···················· | ···················· |
| 25661 | MT776411.1 | \|Severe acute respiratory syndrome coro | ···················· | ···················· | ···················· | ···················· | ···················· | ···················· |
| 25662 | MT776412.1 | \|Severe acute respiratory syndrome coro | ···················· | ···················· | ···················· | ···················· | ···················· | ···················· |
| 25663 | MT776413.1 | \|Severe acute respiratory syndrome coro | ···················· | ···················· | ···················· | ···················· | ···················· | ···················· |
| 25664 | MT776414.1 | \|Severe acute respiratory syndrome coro | ···················· | ···················· | ···················· | ···················· | ···················· | ···················· |
| 25665 | MT776415.1 | \|Severe acute respiratory syndrome coro | ···················· | ···················· | ···················· | ···················· | ···················· | ···················· |
| 25666 | MT776416.1 | \|Severe acute respiratory syndrome coro | ···················· | ···················· | ···················· | ···················· | ···················· | ···················· |
| 25667 | MT776417.1 | \|Severe acute respiratory syndrome coro | ···················· | ···················· | ···················· | ···················· | ···················· | ···················· |
| 25668 | MT776418.1 | \|Severe acute respiratory syndrome coro | ···················· | ···················· | ···················· | ···················· | ···················· | ···················· |
| 25669 | MT776419.1 | \|Severe acute respiratory syndrome coro | ···················· | ···················· | ···················· | ···················· | ···················· | ···················· |
| 25670 | MT776420.1 | \|Severe acute respiratory syndrome coro | ···················· | ···················· | ···················· | ···················· | ···················· | ···················· |
| 25671 | MT776421.1 | \|Severe acute respiratory syndrome coro | ···················· | ···················· | ···················· | ···················· | ···················· | ···················· |
| 25672 | MT776422.1 | \|Severe acute respiratory syndrome coro | ···················· | ···················· | ···················· | ···················· | ···················· | ···················· |
| 25673 | MT776423.1 | \|Severe acute respiratory syndrome coro | ···················· | ···················· | ···················· | ···················· | ···················· | ···················· |
| 25674 | MT776424.1 | \|Severe acute respiratory syndrome coro | ···················· | ···················· | ···················· | ···················· | ···················· | ···················· |
| 25675 | MT776425.1 | \|Severe acute respiratory syndrome coro | ···················· | ···················· | ···················· | ···················· | ···················· | ···················· |
| 25676 | MT776426.1 | \|Severe acute respiratory syndrome coro | ···················· | ···················· | ···················· | ···················· | ···················· | ···················· |
| 25677 | MT776427.1 | \|Severe acute respiratory syndrome coro | ···················· | ···················· | ···················· | ···················· | ···················· | ···················· |
| 25678 | MT776904.1 | \|Severe acute respiratory syndrome coro | ···················· | ···················· | ···················· | ···················· | ···················· | ···················· |
| 25679 | MT777677.1 | \|Severe acute respiratory syndrome coro | ···················· | ···················· | ···················· | ···················· | ···················· | ···················· |
| 25680 | MT778827.1 | \|Severe acute respiratory syndrome coro | ···················· | ··············C····· | ···················· | ···················· | ···················· | ···················· |
| 25681 | MT778828.1 | \|Severe acute respiratory syndrome coro | ···················· | ··············C····· | ···················· | ···················· | ···················· | ···················· |
| 25682 | MT778829.1 | \|Severe acute respiratory syndrome coro | ···················· | ···················· | ···················· | ···················· | ···················· | ···················· |
| 25683 | MT772210.1 | \|Severe acute respiratory syndrome coro | ···················· | ·A·················· | ···················· | ···················· | ···················· | ···A················ |
| 25684 | MT772215.1 | \|Severe acute respiratory syndrome coro | ···················· | ···················· | ···················· | ···················· | ···················· | ···················· |
| 25685 | MT772237.1 | \|Severe acute respiratory syndrome coro | ···················· | ···················· | ···················· | ···················· | ···················· | ···A················ |
| 25686 | MT772240.1 | \|Severe acute respiratory syndrome coro | ···················· | ···················· | ···················· | ···················· | ···················· | ···················· |
| 25687 | MT772252.1 | \|Severe acute respiratory syndrome coro | ···················· | ···················· | ···················· | ···················· | ···················· | ···················· |
| 25688 | MT772253.1 | \|Severe acute respiratory syndrome coro | ···················· | ···················· | ···················· | ···················· | ···················· | ···················· |
| 25689 | MT772256.1 | \|Severe acute respiratory syndrome coro | ···················· | ···················· | ···················· | ···················· | ···················· | ···A················ |
| 25690 | MT772271.1 | \|Severe acute respiratory syndrome coro | ···················· | ···················· | ···················· | ···················· | ···················· | ···················· |
| 25691 | MT772293.1 | \|Severe acute respiratory syndrome coro | ···················· | ···················· | ···················· | ···················· | ···················· | ···················· |
| 25692 | MT772294.1 | \|Severe acute respiratory syndrome coro | ···················· | ···················· | ···················· | ···················· | ···················· | ···················· |
| 25693 | MT772297.1 | \|Severe acute respiratory syndrome coro | ···················· | ···················· | ···················· | ···················· | ···A················ | ···················· |
| 25694 | MT772403.1 | \|Severe acute respiratory syndrome coro | ···················· | ···················· | ···················· | ···················· | ···················· | ···················· |

```
25695 MT772404.1 |Severe acute respiratory syndrome coro    ....................  ....................  .....................  ....................  ....................  ....................
25696 MT772405.1 |Severe acute respiratory syndrome coro    ....................  ....................  .....................  ....................  ....................  ....................
25697 MT772407.1 |Severe acute respiratory syndrome coro    ....................  ....................  .....................  ....................  ....................  ....................
25698 MT772408.1 |Severe acute respiratory syndrome coro    ....................  ....................  .....................  ....................  ....................  ....................
25699 MT772409.1 |Severe acute respiratory syndrome coro    ....................  ....................  .....................  ....................  ....................  ....................
25700 MT772410.1 |Severe acute respiratory syndrome coro    ....................  ....................  .....................  ....................  ....................  ....................
25701 MT772411.1 |Severe acute respiratory syndrome coro    ....................  ....................  .....................  ....................  ....................  ....................
25702 MT772412.1 |Severe acute respiratory syndrome coro    ....................  ....................  .....................  ....................  ....................  ....................
25703 MT772416.1 |Severe acute respiratory syndrome coro    ....................  ....................  .....................  ....................  ....................  ....................
25704 MT772417.1 |Severe acute respiratory syndrome coro    ....................  ....................  .....................  ....................  ....................  ....................
25705 MT772418.1 |Severe acute respiratory syndrome coro    ....................  ....................  .....................  ....................  ....................  ....................
25706 MT772419.1 |Severe acute respiratory syndrome coro    ....................  ....................  .....................  ....................  ....................  ....................
25707 MT772421.1 |Severe acute respiratory syndrome coro    ....................  ....................  .....................  ....................  ....................  ....................
25708 MT772422.1 |Severe acute respiratory syndrome coro    ....................  ....................  .....................  ....................  ....................  ....................
25709 MT772423.1 |Severe acute respiratory syndrome coro    ....................  ....................  .....................  ....................  ....................  ....................
25710 MT772425.1 |Severe acute respiratory syndrome coro    ....................  ....................  .....................  ....................  ....................  ....................
25711 MT772426.1 |Severe acute respiratory syndrome coro    ....................  ....................  .....................  ....................  ....................  ....................
25712 MT772427.1 |Severe acute respiratory syndrome coro    ....................  ....................  .....................  ....................  ....................  ....................
25713 MT772428.1 |Severe acute respiratory syndrome coro    ....................  ....................  .....................  ....................  ....................  ....................
25714 MT772430.1 |Severe acute respiratory syndrome coro    ....................  ....................  .....................  ....................  ....................  ....................
25715 MT772432.1 |Severe acute respiratory syndrome coro    ....................  ....................  .....................  ....................  ....................  ....................
25716 MT772433.1 |Severe acute respiratory syndrome coro    ....................  ....................  .....................  ....................  ....................  ....................
25717 MT772434.1 |Severe acute respiratory syndrome coro    ....................  ....................  .....................  ....................  ....................  ....................
25718 MT772435.1 |Severe acute respiratory syndrome coro    ....................  ....................  .....................  ....................  ....................  ....................
25719 MT772436.1 |Severe acute respiratory syndrome coro    ....................  ....................  .....................  ....................  ....................  ....................
25720 MT772437.1 |Severe acute respiratory syndrome coro    ....................  ....................  .....................  ....................  ....................  ....................
25721 MT772438.1 |Severe acute respiratory syndrome coro    ....................  ....................  .....................  ....................  ....................  ....................
25722 MT772440.1 |Severe acute respiratory syndrome coro    ....................  ....................  .....................  ....................  ....................  ....................
25723 MT772441.1 |Severe acute respiratory syndrome coro    ....................  ....................  .....................  ....................  ....................  ....................
25724 MT772443.1 |Severe acute respiratory syndrome coro    ....................  ....................  .....................  ....................  ....................  ....................
25725 MT772444.1 |Severe acute respiratory syndrome coro    ....................  ....................  .....................  ....................  ....................  ....................
25726 MT772445.1 |Severe acute respiratory syndrome coro    ....................  ....................  .....................  ....................  ....................  ....................
25727 MT772447.1 |Severe acute respiratory syndrome coro    ....................  ....................  .....................  ....................  ....................  ....................
25728 MT772448.1 |Severe acute respiratory syndrome coro    ....................  ....................  .....................  ....................  ....................  ....................
25729 MT772449.1 |Severe acute respiratory syndrome coro    ....................  ....................  .....................  ....................  ....................  ....................
25730 MT772450.1 |Severe acute respiratory syndrome coro    ....................  ....................  .....................  ....................  ....................  ....................
25731 MT772452.1 |Severe acute respiratory syndrome coro    ....................  ....................  .....................  ....................  ....................  ....................
25732 MT772453.1 |Severe acute respiratory syndrome coro    ....................  ....................  .....................  ....................  ....................  ....................
25733 MT772454.1 |Severe acute respiratory syndrome coro    ....................  ....................  .....................  ....................  ....................  ....................
25734 MT772455.1 |Severe acute respiratory syndrome coro    ....................  ....................  .....................  ....................  ....................  ....................
25735 MT772457.1 |Severe acute respiratory syndrome coro    ....................  ....................  .....................  ....................  ....................  ....................
25736 MT772458.1 |Severe acute respiratory syndrome coro    ....................  ....................  .....................  ....................  ....................  ....................
25737 MT772459.1 |Severe acute respiratory syndrome coro    ....................  ....................  .....................  ....................  ....................  ....................
25738 MT772460.1 |Severe acute respiratory syndrome coro    ....................  ....................  .....................  ....................  ....................  ....................
25739 MT772461.1 |Severe acute respiratory syndrome coro    ....................  ....................  .....................  ....................  ....................  ....................
25740 MT772462.1 |Severe acute respiratory syndrome coro    ....................  ....................  .....................  ....................  ....................  ....................
25741 MT772463.1 |Severe acute respiratory syndrome coro    ....................  ....................  .....................  ....................  ....................  ....................
25742 MT772464.1 |Severe acute respiratory syndrome coro    ....................  ....................  .....................  ....................  ....................  ....................
25743 MT772465.1 |Severe acute respiratory syndrome coro    ....................  ....................  .....................  ....................  ....................  ....................
25744 MT772466.1 |Severe acute respiratory syndrome coro    ....................  ....................  .....................  ....................  ....................  ....................
25745 MT772467.1 |Severe acute respiratory syndrome coro    ....................  ....................  .....................  ....................  ....................  ....................
25746 MT772469.1 |Severe acute respiratory syndrome coro    ....................  ....................  .....................  ....................  ....................  ....................
25747 MT772471.1 |Severe acute respiratory syndrome coro    ....................  ....................  .....................  ....................  ....................  ....................
25748 MT772472.1 |Severe acute respiratory syndrome coro    ....................  ....................  .....................  ....................  ....................  ....................
25749 MT772473.1 |Severe acute respiratory syndrome coro    ....................  ....................  .....................  ....................  ....................  ....................
25750 MT772474.1 |Severe acute respiratory syndrome coro    ....................  ....................  .....................  ....................  ....................  ....................
25751 MT772475.1 |Severe acute respiratory syndrome coro    ....................  ....................  .....................  ....................  ....................  ....................
25752 MT772476.1 |Severe acute respiratory syndrome coro    ....................  ....................  .....................  ....................  ....................  ....................
25753 MT772477.1 |Severe acute respiratory syndrome coro    ....................  ....................  .....................  ....................  ....................  ....................
25754 MT772479.1 |Severe acute respiratory syndrome coro    ....................  ....................  .....................  ....................  ....................  ....................
25755 MT772480.1 |Severe acute respiratory syndrome coro    ....................  ....................  .....................  ....................  ....................  ....................
25756 MT772481.1 |Severe acute respiratory syndrome coro    ....................  ....................  .....................  ....................  ....................  ....................
25757 MT772482.1 |Severe acute respiratory syndrome coro    ....................  ....................  .....................  ....................  ....................  ....................
25758 MT772483.1 |Severe acute respiratory syndrome coro    ....................  ....................  .....................  ....................  ....................  ....................
25759 MT772484.1 |Severe acute respiratory syndrome coro    ....................  ....................  .....................  ....................  ....................  ....................
25760 MT772485.1 |Severe acute respiratory syndrome coro    ....................  ....................  .....................  ....................  ....................  ....................
25761 MT772486.1 |Severe acute respiratory syndrome coro    ....................  ....................  .....................  ....................  ....................  ....................
25762 MT772487.1 |Severe acute respiratory syndrome coro    ....................  ....................  .....................  ....................  ....................  ....................
25763 MT772488.1 |Severe acute respiratory syndrome coro    ....................  ....................  .....................  ....................  ....................  ....................
25764 MT772489.1 |Severe acute respiratory syndrome coro    ....................  ....................  .....................  ....................  ....................  ....................
25765 MT772490.1 |Severe acute respiratory syndrome coro    ....................  ....................  .....................  ....................  ....................  ....................
25766 MT772491.1 |Severe acute respiratory syndrome coro    ....................  ....................  .....................  ....................  ....................  ....................
25767 MT772494.1 |Severe acute respiratory syndrome coro    ....................  ....................  .....................  ....................  ....................  ....................
25768 MT772495.1 |Severe acute respiratory syndrome coro    ....................  ....................  .....................  ....................  ....................  ....................
25769 MT772496.1 |Severe acute respiratory syndrome coro    ....................  ....................  .....................  ....................  ....................  ....................
25770 MT772497.1 |Severe acute respiratory syndrome coro    ....................  ....................  .....................  ....................  ....................  ....................
25771 MT772498.1 |Severe acute respiratory syndrome coro    ....................  ....................  .....................  ....................  ....................  ....................
25772 MT772499.1 |Severe acute respiratory syndrome coro    ....................  ....................  .....................  ....................  ....................  ....................
```

| | | | | | | | | |
|---|---|---|---|---|---|---|---|---|
| 25773 | MT772500.1 | \|Severe acute respiratory syndrome coro | ···················· | ···················· | ···················· | ···················· | ···················· | ···················· |
| 25774 | MT772501.1 | \|Severe acute respiratory syndrome coro | ···················· | ···················· | ···················· | ···················· | ···················· | ···················· |
| 25775 | MT772502.1 | \|Severe acute respiratory syndrome coro | ···················· | ···················· | ···················· | ···················· | ···················· | ···················· |
| 25776 | MT772503.1 | \|Severe acute respiratory syndrome coro | ···················· | ···················· | ···················· | ···················· | ···················· | ···················· |
| 25777 | MT772504.1 | \|Severe acute respiratory syndrome coro | ···················· | ···················· | ···················· | ···················· | ···················· | ···················· |
| 25778 | MT772505.1 | \|Severe acute respiratory syndrome coro | ···················· | ···················· | ···················· | ···················· | ···················· | ···················· |
| 25779 | MT772506.1 | \|Severe acute respiratory syndrome coro | ···················· | ···················· | ···················· | ···················· | ···················· | ···················· |
| 25780 | MT772507.1 | \|Severe acute respiratory syndrome coro | ···················· | ···················· | ···················· | ···················· | ···················· | ···················· |
| 25781 | MT772509.1 | \|Severe acute respiratory syndrome coro | ···················· | ···················· | ···················· | ···················· | ···················· | ···················· |
| 25782 | MT772510.1 | \|Severe acute respiratory syndrome coro | ···················· | ···················· | ···················· | ···················· | ···················· | ···················· |
| 25783 | MT772512.1 | \|Severe acute respiratory syndrome coro | ···················· | ···················· | ···················· | ···················· | ···················· | ···················· |
| 25784 | MT772514.1 | \|Severe acute respiratory syndrome coro | ···················· | ···················· | ···················· | ···················· | ···················· | ···················· |
| 25785 | MT772515.1 | \|Severe acute respiratory syndrome coro | ···················· | ···················· | ···················· | ···················· | ···················· | ···················· |
| 25786 | MT772516.1 | \|Severe acute respiratory syndrome coro | ···················· | ···················· | ···················· | ···················· | ···················· | ···················· |
| 25787 | MT772518.1 | \|Severe acute respiratory syndrome coro | ···················· | ···················· | ···················· | ···················· | ···················· | ···················· |
| 25788 | MT772519.1 | \|Severe acute respiratory syndrome coro | ···················· | ···················· | ···················· | ···················· | ···················· | ···················· |
| 25789 | MT772520.1 | \|Severe acute respiratory syndrome coro | ···················· | ···················· | ···················· | ···················· | ···················· | ···················· |
| 25790 | MT772521.1 | \|Severe acute respiratory syndrome coro | ···················· | ···················· | ···················· | ···················· | ···················· | ···················· |
| 25791 | MT772522.1 | \|Severe acute respiratory syndrome coro | ···················· | ···················· | ···················· | ···················· | ···················· | ···················· |
| 25792 | MT772523.1 | \|Severe acute respiratory syndrome coro | ···················· | ···················· | ···················· | ···················· | ···················· | ···················· |
| 25793 | MT772524.1 | \|Severe acute respiratory syndrome coro | ···················· | ···················· | ···················· | ···················· | ···················· | ···················· |
| 25794 | MT772525.1 | \|Severe acute respiratory syndrome coro | ···················· | ···················· | ···················· | ···················· | ···················· | ···················· |
| 25795 | MT772526.1 | \|Severe acute respiratory syndrome coro | ···················· | ···················· | ···················· | ···················· | ···················· | ···················· |
| 25796 | MT772527.1 | \|Severe acute respiratory syndrome coro | ···················· | ···················· | ···················· | ···················· | ···················· | ···················· |
| 25797 | MT772528.1 | \|Severe acute respiratory syndrome coro | ···················· | ···················· | ···················· | ···················· | ···················· | ···················· |
| 25798 | MT772529.1 | \|Severe acute respiratory syndrome coro | ···················· | ···················· | ···················· | ···················· | ···················· | ···················· |
| 25799 | MT772531.1 | \|Severe acute respiratory syndrome coro | ···················· | ···················· | ···················· | ···················· | ···················· | ···················· |
| 25800 | MT772532.1 | \|Severe acute respiratory syndrome coro | ···················· | ···················· | ···················· | ···················· | ···················· | ···················· |
| 25801 | MT772533.1 | \|Severe acute respiratory syndrome coro | ···················· | ···················· | ···················· | ···················· | ···················· | ···················· |
| 25802 | MT772534.1 | \|Severe acute respiratory syndrome coro | ···················· | ···················· | ···················· | ···················· | ···················· | ···················· |
| 25803 | MT772535.1 | \|Severe acute respiratory syndrome coro | ···················· | ···················· | ···················· | ···················· | ···················· | ···················· |
| 25804 | MT772536.1 | \|Severe acute respiratory syndrome coro | ···················· | ···················· | ···················· | ···················· | ···················· | ···················· |
| 25805 | MT772537.1 | \|Severe acute respiratory syndrome coro | ···················· | ···················· | ···················· | ···················· | ···················· | ···················· |
| 25806 | MT772538.1 | \|Severe acute respiratory syndrome coro | ···················· | ···················· | ···················· | ···················· | ···················· | ···················· |
| 25807 | MT772539.1 | \|Severe acute respiratory syndrome coro | ···················· | ···················· | ···················· | ···················· | ···················· | ···················· |
| 25808 | MT772541.1 | \|Severe acute respiratory syndrome coro | ···················· | ···················· | ···················· | ···················· | ···················· | ···················· |
| 25809 | MT772543.1 | \|Severe acute respiratory syndrome coro | ···················· | ···················· | ···················· | ···················· | ···················· | ···················· |
| 25810 | MT772544.1 | \|Severe acute respiratory syndrome coro | ···················· | ···················· | ···················· | ···················· | ···················· | ···················· |
| 25811 | MT772545.1 | \|Severe acute respiratory syndrome coro | ···················· | ···················· | ···················· | ···················· | ···················· | ···················· |
| 25812 | MT772546.1 | \|Severe acute respiratory syndrome coro | ···················· | ···················· | ···················· | ···················· | ···················· | ···················· |
| 25813 | MT772547.1 | \|Severe acute respiratory syndrome coro | ···················· | ···················· | ···················· | ···················· | ···················· | ···················· |
| 25814 | MT772548.1 | \|Severe acute respiratory syndrome coro | ···················· | ···················· | ···················· | ···················· | ···················· | ···················· |
| 25815 | MT772549.1 | \|Severe acute respiratory syndrome coro | ···················· | ···················· | ···················· | ···················· | ···················· | ···················· |
| 25816 | MT772550.1 | \|Severe acute respiratory syndrome coro | ···················· | ···················· | ···················· | ···················· | ···················· | ···················· |
| 25817 | MT772551.1 | \|Severe acute respiratory syndrome coro | ···················· | ···················· | ···················· | ···················· | ···················· | ···················· |
| 25818 | MT772552.1 | \|Severe acute respiratory syndrome coro | ···················· | ···················· | ···················· | ···················· | ···················· | ···················· |
| 25819 | MT772553.1 | \|Severe acute respiratory syndrome coro | ···················· | ···················· | ···················· | ···················· | ···················· | ···················· |
| 25820 | MT772554.1 | \|Severe acute respiratory syndrome coro | ···················· | ···················· | ···················· | ···················· | ···················· | ···················· |
| 25821 | MT772555.1 | \|Severe acute respiratory syndrome coro | ···················· | ···················· | ···················· | ···················· | ···················· | ···················· |
| 25822 | MT772556.1 | \|Severe acute respiratory syndrome coro | ···················· | ···················· | ···················· | ···················· | ···················· | ···················· |
| 25823 | MT772557.1 | \|Severe acute respiratory syndrome coro | ···················· | ···················· | ···················· | ···················· | ···················· | ···················· |
| 25824 | MT772558.1 | \|Severe acute respiratory syndrome coro | ···················· | ···················· | ···················· | ···················· | ···················· | ···················· |
| 25825 | MT772559.1 | \|Severe acute respiratory syndrome coro | ···················· | ···················· | ···················· | ···················· | ···················· | ···················· |
| 25826 | MT772561.1 | \|Severe acute respiratory syndrome coro | ···················· | ···················· | ···················· | ···················· | ···················· | ···················· |
| 25827 | MT772562.1 | \|Severe acute respiratory syndrome coro | ···················· | ···················· | ···················· | ···················· | ···················· | ···················· |
| 25828 | MT772563.1 | \|Severe acute respiratory syndrome coro | ···················· | ···················· | ···················· | ···················· | ···················· | ···················· |
| 25829 | MT772564.1 | \|Severe acute respiratory syndrome coro | ···················· | ···················· | ···················· | ···················· | ···················· | ···················· |
| 25830 | MT772565.1 | \|Severe acute respiratory syndrome coro | ···················· | ···················· | ···················· | ···················· | ···················· | ···················· |
| 25831 | MT772566.1 | \|Severe acute respiratory syndrome coro | ···················· | ···················· | ···················· | ···················· | ···················· | ···················· |
| 25832 | MT772567.1 | \|Severe acute respiratory syndrome coro | ···················· | ···················· | ···················· | ···················· | ··············N····· | ·······N······N····· |
| 25833 | MT772568.1 | \|Severe acute respiratory syndrome coro | ···················· | ···················· | ···················· | ···················· | ···················· | ···················· |
| 25834 | MT772569.1 | \|Severe acute respiratory syndrome coro | ···················· | ···················· | ···················· | ···················· | ···················· | ···················· |
| 25835 | MT772570.1 | \|Severe acute respiratory syndrome coro | ···················· | ···················· | ···················· | ···················· | ···················· | ···················· |
| 25836 | MT772571.1 | \|Severe acute respiratory syndrome coro | ···················· | ···················· | ···················· | ···················· | ···················· | ···················· |
| 25837 | MT772573.1 | \|Severe acute respiratory syndrome coro | ···················· | ···················· | ···················· | ···················· | ···················· | ···················· |
| 25838 | MT772574.1 | \|Severe acute respiratory syndrome coro | ···················· | ···················· | ···················· | ···················· | ···················· | ···················· |
| 25839 | MT772575.1 | \|Severe acute respiratory syndrome coro | ···················· | ···················· | ···················· | ···················· | ···················· | ···················· |
| 25840 | MT772577.1 | \|Severe acute respiratory syndrome coro | ···················· | ···················· | ···················· | ···················· | ··············N····· | ··NN················ |
| 25841 | MT772578.1 | \|Severe acute respiratory syndrome coro | ···················· | ···················· | ···················· | ···················· | ···················· | ···················· |
| 25842 | MT772579.1 | \|Severe acute respiratory syndrome coro | ···················· | ···················· | ···················· | ···················· | ···················· | ···················· |
| 25843 | MT772580.1 | \|Severe acute respiratory syndrome coro | ···················· | ···················· | ···················· | ···················· | ···················· | ···················· |
| 25844 | MT773133.1 | \|Severe acute respiratory syndrome coro | ···················· | ···················· | ···················· | ···················· | ···················· | ···················· |
| 25845 | MT773134.1 | \|Severe acute respiratory syndrome coro | ···················· | ·A·················· | ···················· | ···················· | ···················· | ···A················ |
| 25846 | MT766907.1 | \|Severe acute respiratory syndrome coro | ···················· | ···················· | ···················· | ···················· | ···················· | ···················· |
| 25847 | MT766908.1 | \|Severe acute respiratory syndrome coro | ···················· | ···················· | ···················· | ···················· | ···················· | ···················· |
| 25848 | MT772085.1 | \|Severe acute respiratory syndrome coro | ···················· | ···················· | ···················· | ···················· | ···················· | ···················· |
| 25849 | MT772086.1 | \|Severe acute respiratory syndrome coro | ···················· | ···················· | ···················· | ···················· | ···················· | ···················· |
| 25850 | MT772087.1 | \|Severe acute respiratory syndrome coro | ···················· | ···················· | ···················· | ···················· | ···················· | ···················· |

| | | | | | | | | |
|---|---|---|---|---|---|---|---|---|
| 25851 | MT772088.1 | \|Severe acute respiratory syndrome coro | ···················· | ···················· | ···················· | ···················· | ···················· | ···················· |
| 25852 | MT772089.1 | \|Severe acute respiratory syndrome coro | ···················· | ···················· | ···················· | ···················· | ···················· | ···················· |
| 25853 | MT772090.1 | \|Severe acute respiratory syndrome coro | ···················· | ···················· | ···················· | ···················· | ···················· | ···················· |
| 25854 | MT772091.1 | \|Severe acute respiratory syndrome coro | ···················· | ···················· | ···················· | ···················· | ···················· | ···················· |
| 25855 | MT758193.1 | \|Severe acute respiratory syndrome coro | ···················· | ···················· | ···················· | ···················· | ···················· | ···················· |
| 25856 | MT758197.1 | \|Severe acute respiratory syndrome coro | ········T··········· | ····A··············· | ···················· | ···················· | ···················· | ···················· |
| 25857 | MT758202.1 | \|Severe acute respiratory syndrome coro | ········T··········· | ····A··············· | ···················· | ···················· | ···················· | ···················· |
| 25858 | MT758203.1 | \|Severe acute respiratory syndrome coro | ········T··········· | ···················· | ···················· | ···················· | ···················· | ···················· |
| 25859 | MT758213.1 | \|Severe acute respiratory syndrome coro | ···················· | ·A·················· | ···················· | ···················· | ···················· | ···A················ |
| 25860 | MT758215.1 | \|Severe acute respiratory syndrome coro | ···················· | ·A·················· | ···················· | ···················· | ···················· | ···A················ |
| 25861 | MT758240.1 | \|Severe acute respiratory syndrome coro | ···················· | ···················· | ···················· | ···················· | ···················· | ···················· |
| 25862 | MT758314.1 | \|Severe acute respiratory syndrome coro | ···················· | ···················· | ···················· | ···················· | ···················· | ···A················ |
| 25863 | MT758346.1 | \|Severe acute respiratory syndrome coro | ···················· | ·A·················· | ···················· | ···················· | ···················· | ···A················ |
| 25864 | MT758375.1 | \|Severe acute respiratory syndrome coro | ···················· | ·A·················· | ···················· | ···················· | ···················· | ···A················ |
| 25865 | MT758402.1 | \|Severe acute respiratory syndrome coro | ···················· | ·A·················· | ···················· | ···················· | ···················· | ···················· |
| 25866 | MT758421.1 | \|Severe acute respiratory syndrome coro | ···················· | ···················· | ···················· | ···················· | ···················· | ···················· |
| 25867 | MT758436.1 | \|Severe acute respiratory syndrome coro | ···················· | ···················· | ···················· | ···················· | ···················· | ···A················ |
| 25868 | MT758439.1 | \|Severe acute respiratory syndrome coro | ···················· | ···················· | ···················· | ···················· | ···················· | ···················· |
| 25869 | MT758471.1 | \|Severe acute respiratory syndrome coro | ···················· | ·A·················· | ···················· | ···················· | ···················· | ···A················ |
| 25870 | MT758683.1 | \|Severe acute respiratory syndrome coro | ···················· | ···················· | ···················· | ···················· | ···················· | ···················· |
| 25871 | MT758701.1 | \|Severe acute respiratory syndrome coro | ···················· | ···················· | ···················· | ···················· | ···················· | ···················· |
| 25872 | MT759582.1 | \|Severe acute respiratory syndrome coro | ···················· | ···················· | ···················· | ···················· | ···················· | ···A················ |
| 25873 | MT759588.1 | \|Severe acute respiratory syndrome coro | ···················· | ·A·················· | ···················· | ···················· | ···················· | ···················· |
| 25874 | MT759714.1 | \|Severe acute respiratory syndrome coro | ···················· | ···················· | ···················· | ···················· | ···················· | ···················· |
| 25875 | MT759742.1 | \|Severe acute respiratory syndrome coro | ···················· | ···················· | ···················· | ···················· | ···················· | ···A················ |
| 25876 | MT759820.1 | \|Severe acute respiratory syndrome coro | ···················· | ···················· | ···················· | ···················· | ···················· | ···················· |
| 25877 | MT759844.1 | \|Severe acute respiratory syndrome coro | ···················· | ···················· | ···················· | ···················· | ···················· | ···················· |
| 25878 | MT760790.1 | \|Severe acute respiratory syndrome coro | ···················· | ·A·················· | ···················· | ···················· | ···················· | ···A················ |
| 25879 | MT762396.1 | \|Severe acute respiratory syndrome coro | ···················· | ···················· | ···················· | ···················· | ···················· | ···················· |
| 25880 | MT762397.1 | \|Severe acute respiratory syndrome coro | ···················· | ···················· | ···················· | ···················· | ···················· | ···················· |
| 25881 | MT762398.1 | \|Severe acute respiratory syndrome coro | ···················· | ·A·················· | ···················· | ···················· | ···················· | ···A················ |
| 25882 | MT762942.1 | \|Severe acute respiratory syndrome coro | ···················· | ···················· | ···················· | ···················· | ···················· | ···················· |
| 25883 | MT762943.1 | \|Severe acute respiratory syndrome coro | ···················· | ···················· | ···················· | ···················· | ···················· | ···················· |
| 25884 | MT762944.1 | \|Severe acute respiratory syndrome coro | ···················· | ···················· | ···················· | ···················· | ···················· | ···················· |
| 25885 | MT764166.1 | \|Severe acute respiratory syndrome coro | ···················· | ···················· | ···················· | ···················· | ···················· | ···················· |
| 25886 | MT764170.1 | \|Severe acute respiratory syndrome coro | ···················· | ···················· | ···················· | ···················· | ···················· | ···················· |
| 25887 | MT764172.1 | \|Severe acute respiratory syndrome coro | ···················· | ···················· | ···················· | ···················· | ···················· | ···················· |
| 25888 | MT764174.1 | \|Severe acute respiratory syndrome coro | ···················· | ···················· | ···················· | ···················· | ···················· | ···················· |
| 25889 | MT764177.1 | \|Severe acute respiratory syndrome coro | ···················· | ···················· | ···················· | ···················· | ···················· | ···················· |
| 25890 | MT764178.1 | \|Severe acute respiratory syndrome coro | ···················· | ···················· | ···················· | ···················· | ···················· | ···················· |
| 25891 | MT764179.1 | \|Severe acute respiratory syndrome coro | ···················· | ···················· | ···················· | ···················· | ···················· | ···················· |
| 25892 | MT764180.1 | \|Severe acute respiratory syndrome coro | ···················· | ··············C····· | ···················· | ···················· | ···················· | ···················· |
| 25893 | MT764181.1 | \|Severe acute respiratory syndrome coro | ···················· | ··············C····· | ···················· | ···················· | ···················· | ···················· |
| 25894 | MT755582.1 | \|Severe acute respiratory syndrome coro | ···················· | ···················· | ···················· | ···················· | ···················· | ···················· |
| 25895 | MT755585.1 | \|Severe acute respiratory syndrome coro | ···················· | ···················· | ···················· | ···················· | ···················· | ···················· |
| 25896 | MT755587.1 | \|Severe acute respiratory syndrome coro | ···················· | ···················· | ···················· | ···················· | ···················· | ···················· |
| 25897 | MT755593.1 | \|Severe acute respiratory syndrome coro | ···················· | ···················· | ···················· | ···················· | ···················· | ···················· |
| 25898 | MT755594.1 | \|Severe acute respiratory syndrome coro | ···················· | ···················· | ···················· | ···················· | ···················· | ···················· |
| 25899 | MT755599.1 | \|Severe acute respiratory syndrome coro | ···················· | ···················· | ···················· | ···················· | ···················· | ···················· |
| 25900 | MT755600.1 | \|Severe acute respiratory syndrome coro | ···················· | ···················· | ···················· | ···················· | ···················· | ···················· |
| 25901 | MT755602.1 | \|Severe acute respiratory syndrome coro | ···················· | ···················· | ···················· | ···················· | ···················· | ···················· |
| 25902 | MT755827.1 | \|Severe acute respiratory syndrome coro | ···················· | ···················· | ···················· | ···················· | ···················· | ···················· |
| 25903 | MT755828.1 | \|Severe acute respiratory syndrome coro | ···················· | ···················· | ···················· | ···················· | ···················· | ···················· |
| 25904 | MT755829.1 | \|Severe acute respiratory syndrome coro | ···················· | ···················· | ···················· | ···················· | ···················· | ···················· |
| 25905 | MT755830.1 | \|Severe acute respiratory syndrome coro | ···················· | ···················· | ···················· | ···················· | ···················· | ···················· |
| 25906 | MT755831.1 | \|Severe acute respiratory syndrome coro | ···················· | ···················· | ···················· | ···················· | ···················· | ···················· |
| 25907 | MT755832.1 | \|Severe acute respiratory syndrome coro | ···················· | ···················· | ···················· | ···················· | ···················· | ···················· |
| 25908 | MT755833.1 | \|Severe acute respiratory syndrome coro | ···················· | ···················· | ···················· | ···················· | ···················· | ···················· |
| 25909 | MT755834.1 | \|Severe acute respiratory syndrome coro | ···················· | ···················· | ···················· | ···················· | ···················· | ···················· |
| 25910 | MT755835.1 | \|Severe acute respiratory syndrome coro | ···················· | ···················· | ···················· | ···················· | ···················· | ···················· |
| 25911 | MT755836.1 | \|Severe acute respiratory syndrome coro | ···················· | ···················· | ···················· | ···················· | ···················· | ···················· |
| 25912 | MT755837.1 | \|Severe acute respiratory syndrome coro | ···················· | ···················· | ···················· | ···················· | ···················· | ···················· |
| 25913 | MT755838.1 | \|Severe acute respiratory syndrome coro | ···················· | ···················· | ···················· | ···················· | ···················· | ···················· |
| 25914 | MT755857.1 | \|Severe acute respiratory syndrome coro | ···················· | ·A·················· | ···················· | ···················· | ···················· | ···A················ |
| 25915 | MT755858.1 | \|Severe acute respiratory syndrome coro | ···················· | ···················· | ···················· | ···················· | ···················· | ···················· |
| 25916 | MT755883.1 | \|Severe acute respiratory syndrome coro | ···················· | ···················· | ···················· | ···················· | ···················· | ···················· |
| 25917 | MT755884.1 | \|Severe acute respiratory syndrome coro | ···················· | ···················· | ···················· | ···················· | ···················· | ···················· |
| 25918 | MT755885.1 | \|Severe acute respiratory syndrome coro | ···················· | ···················· | ···················· | ···················· | ···················· | ···················· |
| 25919 | MT755886.1 | \|Severe acute respiratory syndrome coro | ···················· | ···················· | ···················· | ···················· | ···················· | ···················· |
| 25920 | MT755887.1 | \|Severe acute respiratory syndrome coro | ···················· | ···················· | ···················· | ···················· | ···················· | ···················· |
| 25921 | MT755888.1 | \|Severe acute respiratory syndrome coro | ···················· | ···················· | ···················· | ···················· | ···················· | ···A················ |
| 25922 | MT755889.1 | \|Severe acute respiratory syndrome coro | ···················· | ···················· | ···················· | ···················· | ···················· | ···················· |
| 25923 | MT755890.1 | \|Severe acute respiratory syndrome coro | ···················· | ·A·················· | ···················· | ···················· | ···················· | ···A················ |
| 25924 | MT755891.1 | \|Severe acute respiratory syndrome coro | ···················· | ·A·················· | ···················· | ···················· | ···················· | ···A················ |
| 25925 | MT755892.1 | \|Severe acute respiratory syndrome coro | ···················· | ···················· | ···················· | ···················· | ···················· | ···················· |
| 25926 | MT755893.1 | \|Severe acute respiratory syndrome coro | ···················· | ···················· | ···················· | ···················· | ···················· | ···A················ |
| 25927 | MT755894.1 | \|Severe acute respiratory syndrome coro | ···················· | ·A·················· | ···················· | ···················· | ···················· | ···················· |
| 25928 | MT755895.1 | \|Severe acute respiratory syndrome coro | ···················· | ···················· | ···················· | ···················· | ···················· | ···················· |

| | | | | | | | | | |
|---|---|---|---|---|---|---|---|---|---|
| 25929 | MT755896.1 | Severe acute respiratory syndrome coro | ···················· | ···················· | ···················· | ···················· | ···················· | ···················· | |
| 25930 | MT755897.1 | Severe acute respiratory syndrome coro | ···················· | ···················· | ···················· | ···················· | ···················· | ···················· | |
| 25931 | MT755898.1 | Severe acute respiratory syndrome coro | ···················· | ···················· | ···················· | ···················· | ···················· | ···················· | |
| 25932 | MT755899.1 | Severe acute respiratory syndrome coro | ···················· | ···················· | ···················· | ···················· | ···················· | ···················· | |
| 25933 | MT755900.1 | Severe acute respiratory syndrome coro | ···················· | ···················· | ···················· | ···················· | ···················· | ···················· | |
| 25934 | MT755901.1 | Severe acute respiratory syndrome coro | ···················· | ···················· | ···················· | ···················· | ···················· | ···················· | |
| 25935 | MT755916.1 | Severe acute respiratory syndrome coro | ···················· | ···················· | ···················· | ···················· | ···················· | ···················· | |
| 25936 | MT755917.1 | Severe acute respiratory syndrome coro | ···················· | ···················· | ···················· | ···················· | ···················· | ···················· | |
| 25937 | MT755955.1 | Severe acute respiratory syndrome coro | ···················· | ···················· | ···················· | ···················· | ···················· | ···················· | |
| 25938 | MT755956.1 | Severe acute respiratory syndrome coro | ···················· | ···················· | ···················· | ···················· | ···················· | ···················· | |
| 25939 | MT755973.1 | Severe acute respiratory syndrome coro | ···················· | ···················· | ···················· | ···················· | ···················· | ··A················· | |
| 25941 | MT757012.1 | Severe acute respiratory syndrome coro | ···················· | ···················· | ···················· | ···················· | ···················· | ···················· | |
| 25942 | MT757013.1 | Severe acute respiratory syndrome coro | ···················· | ···················· | ···················· | ···················· | ···················· | ···················· | |
| 25943 | MT757016.1 | Severe acute respiratory syndrome coro | ···················· | ···················· | ···················· | ···················· | ···················· | ···················· | |
| 25944 | MT757018.1 | Severe acute respiratory syndrome coro | ···················· | ···················· | ···················· | ···················· | ···················· | ···················· | |
| 25945 | MT757019.1 | Severe acute respiratory syndrome coro | ···················· | ···················· | ···················· | ···················· | ···················· | ···················· | |
| 25946 | MT757020.1 | Severe acute respiratory syndrome coro | ···················· | ···················· | ···················· | ···················· | ···················· | ···················· | |
| 25947 | MT757021.1 | Severe acute respiratory syndrome coro | ···················· | ···················· | ···················· | ···················· | ···················· | ···················· | |
| 25948 | MT757022.1 | Severe acute respiratory syndrome coro | ···················· | ···················· | ···················· | ···················· | ···················· | ···················· | |
| 25949 | MT757024.1 | Severe acute respiratory syndrome coro | ···················· | ·············A······ | ···················· | ···················· | ···················· | ···················· | |
| 25950 | MT757025.1 | Severe acute respiratory syndrome coro | ···················· | ···················· | ···················· | ···················· | ···················· | ···················· | |
| 25951 | MT757027.1 | Severe acute respiratory syndrome coro | ···················· | ···················· | ···················· | ···················· | ···················· | ···················· | |
| 25952 | MT757029.1 | Severe acute respiratory syndrome coro | ···················· | ···················· | ···················· | ···················· | ···················· | ···················· | |
| 25953 | MT757030.1 | Severe acute respiratory syndrome coro | ···················· | ···················· | ···················· | ···················· | ···················· | ···················· | |
| 25954 | MT757032.1 | Severe acute respiratory syndrome coro | ···················· | ···················· | ···················· | ···················· | ···················· | ···················· | |
| 25955 | MT757033.1 | Severe acute respiratory syndrome coro | ···················· | ···················· | ···················· | ···················· | ···················· | ···················· | |
| 25956 | MT757037.1 | Severe acute respiratory syndrome coro | ···················· | ···················· | ···················· | ···················· | ···················· | ···················· | |
| 25957 | MT757038.1 | Severe acute respiratory syndrome coro | ···················· | ···················· | ···················· | ···················· | ···················· | ···················· | |
| 25958 | MT757039.1 | Severe acute respiratory syndrome coro | ···················· | ···················· | ···················· | ···················· | ···················· | ···················· | |
| 25959 | MT757040.1 | Severe acute respiratory syndrome coro | ···················· | ···················· | ···················· | ···················· | ···················· | ···················· | |
| 25960 | MT757041.1 | Severe acute respiratory syndrome coro | ···················· | ···················· | ···················· | ···················· | ···················· | ···················· | |
| 25961 | MT757043.1 | Severe acute respiratory syndrome coro | ···················· | ···················· | ···················· | ···················· | ···················· | ···················· | |
| 25962 | MT757044.1 | Severe acute respiratory syndrome coro | ···················· | ···················· | ···················· | ···················· | ···················· | ···················· | |
| 25963 | MT757047.1 | Severe acute respiratory syndrome coro | ···················· | ···················· | ···················· | ···················· | ···················· | ···················· | |
| 25964 | MT757049.1 | Severe acute respiratory syndrome coro | ···················· | ···················· | ···················· | ···················· | ···················· | ···················· | |
| 25966 | MT757054.1 | Severe acute respiratory syndrome coro | ···················· | ···················· | ···················· | ···················· | ···················· | ···················· | |
| 25967 | MT757056.1 | Severe acute respiratory syndrome coro | ···················· | ···················· | ···················· | ···················· | ···················· | ···················· | |
| 25968 | MT757058.1 | Severe acute respiratory syndrome coro | ···················· | ···················· | ···················· | ···················· | ···················· | ···················· | |
| 25969 | MT757063.1 | Severe acute respiratory syndrome coro | ···················· | ···················· | ···················· | ···················· | NNN················· | ···················· | 1 |
| 25970 | MT757064.1 | Severe acute respiratory syndrome coro | ···················· | ···················· | ···················· | ···················· | ···················· | ···················· | |
| 25971 | MT757066.1 | Severe acute respiratory syndrome coro | ···················· | ···················· | ···················· | ···················· | ···················· | ···················· | |
| 25972 | MT757067.1 | Severe acute respiratory syndrome coro | ···················· | ···················· | ···················· | ···················· | ···················· | ···················· | |
| 25973 | MT757068.1 | Severe acute respiratory syndrome coro | ···················· | ···················· | ···················· | ···················· | ···················· | ···················· | |
| 25974 | MT757070.1 | Severe acute respiratory syndrome coro | ···················· | ···················· | ···················· | ···················· | ···················· | ···················· | |
| 25975 | MT757071.1 | Severe acute respiratory syndrome coro | ···················· | ···················· | ···················· | ···················· | ···················· | ···················· | |
| 25976 | MT757072.1 | Severe acute respiratory syndrome coro | ···················· | ···················· | ···················· | ···················· | ···················· | ···················· | |
| 25977 | MT757073.1 | Severe acute respiratory syndrome coro | ···················· | ···················· | ···················· | ···················· | ···················· | ··A················· | |
| 25978 | MT757074.1 | Severe acute respiratory syndrome coro | ···················· | ···················· | ···················· | ···················· | ···················· | ··A················· | |
| 25979 | MT757075.1 | Severe acute respiratory syndrome coro | ···················· | ···················· | ···················· | ···················· | ···················· | ··A················· | |
| 25980 | MT757076.1 | Severe acute respiratory syndrome coro | ···················· | ···················· | ···················· | ···················· | ···················· | ···················· | |
| 25981 | MT757077.1 | Severe acute respiratory syndrome coro | ···················· | ···················· | ···················· | ···················· | ···················· | ···················· | |
| 25982 | MT757078.1 | Severe acute respiratory syndrome coro | ···················· | ···················· | ···················· | ···················· | ···················· | ···················· | |
| 25983 | MT757079.1 | Severe acute respiratory syndrome coro | ···················· | ···················· | ···················· | ···················· | ···················· | ···················· | |
| 25984 | MT757080.1 | Severe acute respiratory syndrome coro | ···················· | ···················· | ···················· | ···················· | ···················· | ···················· | |
| 25985 | MT757081.1 | Severe acute respiratory syndrome coro | ···················· | ···················· | ···················· | ···················· | ···················· | ···················· | |
| 25986 | MT757082.1 | Severe acute respiratory syndrome coro | ···················· | ···················· | ···················· | ···················· | ···················· | ··A················· | |
| 25987 | MT757083.1 | Severe acute respiratory syndrome coro | ···················· | ···················· | ···················· | ···················· | ···················· | ··A················· | |
| 25988 | MT757085.1 | Severe acute respiratory syndrome coro | ···················· | ···················· | ···················· | ···················· | ···················· | ···················· | |
| 25991 | MT757096.1 | Severe acute respiratory syndrome coro | ···················· | ···················· | ···················· | ···················· | ···················· | ···················· | |
| 25992 | MT757100.1 | Severe acute respiratory syndrome coro | ···················· | ···················· | ···················· | ···················· | ···················· | ···················· | |
| 25993 | MT757104.1 | Severe acute respiratory syndrome coro | ···················· | ···················· | ···················· | ···················· | ···················· | ···················· | |
| 25994 | MT757105.1 | Severe acute respiratory syndrome coro | ···················· | ···················· | ···················· | ···················· | ···················· | ···················· | |
| 25995 | MT757110.1 | Severe acute respiratory syndrome coro | ···················· | ···················· | ···················· | ···················· | ···················· | ···················· | |
| 25996 | MT757111.1 | Severe acute respiratory syndrome coro | ···················· | ···················· | ···················· | ···················· | ···················· | ···················· | |
| 25997 | MT757115.1 | Severe acute respiratory syndrome coro | ···················· | ···················· | ···················· | ···················· | ···················· | ···················· | |
| 25998 | MT757116.1 | Severe acute respiratory syndrome coro | ···················· | ···················· | ···················· | ···················· | ···················· | ···················· | |
| 25999 | MT757117.1 | Severe acute respiratory syndrome coro | ···················· | ···················· | ···················· | ···················· | ···················· | ···················· | |
| 26000 | MT757119.1 | Severe acute respiratory syndrome coro | ···················· | ···················· | ···················· | ···················· | ···················· | ···················· | |
| 26001 | MT757121.1 | Severe acute respiratory syndrome coro | ···················· | ···················· | ···················· | ···················· | ···················· | ···················· | |
| 26002 | MT757122.1 | Severe acute respiratory syndrome coro | ···················· | ···················· | ···················· | ···················· | ···················· | ···················· | |
| 26003 | MT757123.1 | Severe acute respiratory syndrome coro | ···················· | ···················· | ···················· | ···················· | ···················· | ···················· | |
| 26004 | MT745584.1 | Severe acute respiratory syndrome coro | ···················· | ···················· | ···················· | ···················· | ···················· | ···················· | |
| 26005 | MT745601.1 | Severe acute respiratory syndrome coro | ···················· | ···················· | ···················· | ···················· | ···················· | ···················· | |
| 26006 | MT745605.1 | Severe acute respiratory syndrome coro | ···················· | ···················· | ···················· | ···················· | ··N················· | ···················· | |
| 26007 | MT745608.1 | Severe acute respiratory syndrome coro | ···················· | ···················· | ···················· | ···················· | ···················· | ···················· | |
| 26008 | MT745610.1 | Severe acute respiratory syndrome coro | ···················· | ···················· | ···················· | ···················· | ···················· | ···················· | |
| 26009 | MT745611.1 | Severe acute respiratory syndrome coro | ···················· | ···················· | ···················· | ···················· | ···················· | ···················· | |
| 26010 | MT745629.1 | Severe acute respiratory syndrome coro | ···················· | ···················· | ···················· | ···················· | ···················· | ···················· | |

| | | | | | | | | | |
|---|---|---|---|---|---|---|---|---|---|
| 26012 | MT745631.1 | \|Severe acute respiratory syndrome coro | ···················· | ···················· | ···················· | ···················· | ···················· | ···················· | |
| 26013 | MT745639.1 | \|Severe acute respiratory syndrome coro | ···················· | ···················· | ···················· | ···················· | ···················· | ···················· | |
| 26014 | MT745640.1 | \|Severe acute respiratory syndrome coro | ···················· | ···················· | ···················· | ···················· | ·NN················· | ···················· | |
| 26015 | MT745642.1 | \|Severe acute respiratory syndrome coro | ···················· | ···················· | ···················· | ···················· | ···················· | ···················· | |
| 26016 | MT745644.1 | \|Severe acute respiratory syndrome coro | ···················· | ···················· | ···················· | ···················· | ···················· | ···················· | |
| 26017 | MT745645.1 | \|Severe acute respiratory syndrome coro | ···················· | ···················· | ···················· | ···················· | ···················· | ···················· | |
| 26018 | MT745646.1 | \|Severe acute respiratory syndrome coro | ···················· | ···················· | ···················· | ···················· | ···················· | ···················· | |
| 26019 | MT745648.1 | \|Severe acute respiratory syndrome coro | ···················· | ···················· | ···················· | ···················· | ···················· | ···················· | |
| 26020 | MT745649.1 | \|Severe acute respiratory syndrome coro | ···················· | ···················· | ···················· | ···················· | ···················· | ···················· | |
| 26021 | MT745650.1 | \|Severe acute respiratory syndrome coro | ···················· | ···················· | ···················· | ···················· | ···················· | ···················· | |
| 26022 | MT745651.1 | \|Severe acute respiratory syndrome coro | ···················· | ···················· | ···················· | ···················· | ···················· | ···················· | |
| 26023 | MT745652.1 | \|Severe acute respiratory syndrome coro | ···················· | ···················· | ···················· | ···················· | ···················· | ···················· | |
| 26024 | MT745653.1 | \|Severe acute respiratory syndrome coro | ···················· | ···················· | ···················· | ···················· | ···················· | ···················· | |
| 26025 | MT745654.1 | \|Severe acute respiratory syndrome coro | ···················· | ···················· | ···················· | ···················· | ···················· | ···················· | |
| 26026 | MT745655.1 | \|Severe acute respiratory syndrome coro | ···················· | ···················· | ···················· | ···················· | ···················· | ···················· | |
| 26027 | MT745656.1 | \|Severe acute respiratory syndrome coro | ···················· | ···················· | ···················· | ···················· | ···················· | ···················· | |
| 26028 | MT745657.1 | \|Severe acute respiratory syndrome coro | ···················· | ···················· | ···················· | ···················· | ···················· | ···················· | |
| 26029 | MT745658.1 | \|Severe acute respiratory syndrome coro | ···················· | ···················· | ···················· | ···················· | ···················· | ···················· | |
| 26030 | MT745659.1 | \|Severe acute respiratory syndrome coro | ···················· | ···················· | ···················· | ···················· | ···················· | ···················· | |
| 26031 | MT745660.1 | \|Severe acute respiratory syndrome coro | ···················· | ···················· | ···················· | ···················· | ···················· | ···················· | |
| 26032 | MT745661.1 | \|Severe acute respiratory syndrome coro | ···················· | ···················· | ···················· | ···················· | ···················· | ···················· | |
| 26033 | MT745662.1 | \|Severe acute respiratory syndrome coro | ···················· | ···················· | ···················· | ···················· | ···················· | ···················· | |
| 26034 | MT745663.1 | \|Severe acute respiratory syndrome coro | ···················· | ···················· | ···················· | ···················· | ···················· | ···················· | |
| 26035 | MT745664.1 | \|Severe acute respiratory syndrome coro | ···················· | ···················· | ···················· | ···················· | ···················· | ···················· | |
| 26036 | MT745665.1 | \|Severe acute respiratory syndrome coro | ···················· | ···················· | ···················· | ···················· | ···················· | ···················· | |
| 26037 | MT745666.1 | \|Severe acute respiratory syndrome coro | ···················· | ···················· | ···················· | ···················· | ···················· | ···················· | |
| 26038 | MT745667.1 | \|Severe acute respiratory syndrome coro | ···················· | ···················· | ···················· | ···················· | ···················· | ···················· | |
| 26039 | MT745668.1 | \|Severe acute respiratory syndrome coro | ···················· | ···················· | ···················· | ···················· | ···················· | ···················· | |
| 26040 | MT745669.1 | \|Severe acute respiratory syndrome coro | ···················· | ···················· | ···················· | ···················· | ···················· | ···················· | |
| 26041 | MT745670.1 | \|Severe acute respiratory syndrome coro | ···················· | ···················· | ···················· | ···················· | ···················· | ···················· | |
| 26042 | MT745671.1 | \|Severe acute respiratory syndrome coro | ···················· | ···················· | ···················· | ···················· | ···················· | ···················· | |
| 26043 | MT745672.1 | \|Severe acute respiratory syndrome coro | ···················· | ···················· | ···················· | ···················· | NNN················· | ···················· | 1 |
| 26044 | MT745673.1 | \|Severe acute respiratory syndrome coro | ···················· | ···················· | ···················· | ···················· | ···················· | ···················· | |
| 26045 | MT745674.1 | \|Severe acute respiratory syndrome coro | ···················· | ···················· | ···················· | ···················· | ···················· | ···················· | |
| 26046 | MT745676.1 | \|Severe acute respiratory syndrome coro | ···················· | ···················· | ···················· | ···················· | ···················· | ···················· | |
| 26047 | MT745677.1 | \|Severe acute respiratory syndrome coro | ···················· | ···················· | ···················· | ···················· | ···················· | ···················· | |
| 26048 | MT745678.1 | \|Severe acute respiratory syndrome coro | ···················· | ···················· | ···················· | ···················· | ·NN················· | ···················· | |
| 26050 | MT745680.1 | \|Severe acute respiratory syndrome coro | ···················· | ···················· | ···················· | ···················· | ···················· | ···················· | |
| 26051 | MT745681.1 | \|Severe acute respiratory syndrome coro | ···················· | ···················· | ···················· | ···················· | ···················· | ···················· | |
| 26052 | MT745682.1 | \|Severe acute respiratory syndrome coro | ···················· | ···················· | ···················· | ···················· | ···················· | ···················· | |
| 26053 | MT745683.1 | \|Severe acute respiratory syndrome coro | ···················· | ···················· | ···················· | ···················· | ···················· | ···················· | |
| 26054 | MT745684.1 | \|Severe acute respiratory syndrome coro | ···················· | ···················· | ···················· | ···················· | ···················· | ···················· | |
| 26055 | MT745685.1 | \|Severe acute respiratory syndrome coro | ···················· | ···················· | ···················· | ···················· | ···················· | ···················· | |
| 26056 | MT745686.1 | \|Severe acute respiratory syndrome coro | ···················· | ···················· | ···················· | ···················· | ···················· | ···················· | |
| 26057 | MT745687.1 | \|Severe acute respiratory syndrome coro | ···················· | ···················· | ···················· | ···················· | ···················· | ···················· | |
| 26058 | MT745688.1 | \|Severe acute respiratory syndrome coro | ···················· | ···················· | ···················· | ···················· | ···················· | ···················· | |
| 26059 | MT745689.1 | \|Severe acute respiratory syndrome coro | ···················· | ···················· | ···················· | ···················· | ···················· | ···················· | |
| 26060 | MT745690.1 | \|Severe acute respiratory syndrome coro | ···················· | ···················· | ···················· | ···················· | ···················· | ···················· | |
| 26061 | MT745691.1 | \|Severe acute respiratory syndrome coro | ···················· | ···················· | ···················· | ···················· | ···················· | ···················· | |
| 26062 | MT745692.1 | \|Severe acute respiratory syndrome coro | ···················· | ···················· | ···················· | ···················· | ···················· | ···················· | |
| 26063 | MT745693.1 | \|Severe acute respiratory syndrome coro | ···················· | ···················· | ···················· | ···················· | ···················· | ···················· | |
| 26064 | MT745694.1 | \|Severe acute respiratory syndrome coro | ···················· | ·A·················· | ···················· | ···················· | ···················· | ··A················· | |
| 26065 | MT745695.1 | \|Severe acute respiratory syndrome coro | ···················· | ···················· | ···················· | ···················· | ···················· | ···················· | |
| 26066 | MT745696.1 | \|Severe acute respiratory syndrome coro | ···················· | ···················· | ···················· | ···················· | ···················· | ···················· | |
| 26067 | MT745697.1 | \|Severe acute respiratory syndrome coro | ···················· | ···················· | ···················· | ···················· | ···················· | ···················· | |
| 26068 | MT745698.1 | \|Severe acute respiratory syndrome coro | ···················· | ···················· | ···················· | ···················· | ···················· | ···················· | |
| 26069 | MT745699.1 | \|Severe acute respiratory syndrome coro | ···················· | ···················· | ···················· | ···················· | ···················· | ···················· | |
| 26071 | MT745701.1 | \|Severe acute respiratory syndrome coro | ···················· | ···················· | ···················· | ···················· | ···················· | ···················· | |
| 26072 | MT745702.1 | \|Severe acute respiratory syndrome coro | ···················· | ···················· | ···················· | ···················· | ···················· | ···················· | |
| 26073 | MT745703.1 | \|Severe acute respiratory syndrome coro | ···················· | ···················· | ···················· | ···················· | ···················· | ···················· | |
| 26074 | MT745704.1 | \|Severe acute respiratory syndrome coro | ···················· | ···················· | ···················· | ···················· | ···················· | ···················· | |
| 26075 | MT745705.1 | \|Severe acute respiratory syndrome coro | ···················· | ···················· | ···················· | ···················· | ···················· | ···················· | |
| 26076 | MT745706.1 | \|Severe acute respiratory syndrome coro | ···················· | ···················· | ···················· | ···················· | ···················· | ···················· | |
| 26077 | MT745707.1 | \|Severe acute respiratory syndrome coro | ···················· | ···················· | ···················· | ···················· | ···················· | ···················· | |
| 26078 | MT745708.1 | \|Severe acute respiratory syndrome coro | ···················· | ···················· | ···················· | ···················· | ···················· | ···················· | |
| 26079 | MT745709.1 | \|Severe acute respiratory syndrome coro | ···················· | ···················· | ···················· | ···················· | ···················· | ···················· | |
| 26080 | MT745710.1 | \|Severe acute respiratory syndrome coro | ···················· | ···················· | ···················· | ···················· | ···················· | ···················· | |
| 26081 | MT745711.1 | \|Severe acute respiratory syndrome coro | ···················· | ·A·················· | ···················· | ···················· | ···················· | ··A················· | |
| 26082 | MT745712.1 | \|Severe acute respiratory syndrome coro | ···················· | ·A·················· | ···················· | ···················· | ···················· | ··A················· | |
| 26083 | MT745713.1 | \|Severe acute respiratory syndrome coro | ···················· | ···················· | ···················· | ···················· | ···················· | ···················· | |
| 26084 | MT745714.1 | \|Severe acute respiratory syndrome coro | ···················· | ···················· | ···················· | ···················· | ···················· | ···················· | |
| 26085 | MT745715.1 | \|Severe acute respiratory syndrome coro | ···················· | ···················· | ···················· | ···················· | ···················· | NN·················· | |
| 26086 | MT745716.1 | \|Severe acute respiratory syndrome coro | ···················· | ···················· | ···················· | ···················· | ···················· | ···················· | |
| 26087 | MT745717.1 | \|Severe acute respiratory syndrome coro | ···················· | ···················· | ···················· | ···················· | ···················· | ···················· | |
| 26088 | MT745718.1 | \|Severe acute respiratory syndrome coro | ···················· | ···················· | ···················· | ···················· | ···················· | ···················· | |
| 26089 | MT745719.1 | \|Severe acute respiratory syndrome coro | ···················· | ···················· | ···················· | ···················· | ···················· | ···················· | |
| 26090 | MT745720.1 | \|Severe acute respiratory syndrome coro | ···················· | ···················· | ···················· | ···················· | ···················· | ···················· | |
| 26091 | MT745721.1 | \|Severe acute respiratory syndrome coro | ···················· | ···················· | ···················· | ···················· | ···················· | ···················· | |

| | | | | | | | | | |
|---|---|---|---|---|---|---|---|---|---|
| 26093 | MT745723.1 | \|Severe acute respiratory syndrome coro | ···················· | ···················· | ···················· | ···················· | ···················· | ···················· | |
| 26094 | MT745724.1 | \|Severe acute respiratory syndrome coro | ···················· | ···················· | ···················· | ···················· | ···················· | ···················· | |
| 26095 | MT745725.1 | \|Severe acute respiratory syndrome coro | ···················· | ···················· | ···················· | ···················· | ···················· | ···················· | |
| 26096 | MT745726.1 | \|Severe acute respiratory syndrome coro | ···················· | ···················· | ···················· | ···················· | ···················· | ···················· | |
| 26097 | MT745727.1 | \|Severe acute respiratory syndrome coro | ···················· | ···················· | ···················· | ···················· | ···················· | ···················· | |
| 26098 | MT745728.1 | \|Severe acute respiratory syndrome coro | ···················· | ···················· | ···················· | ···················· | ···················· | ···················· | |
| 26099 | MT745729.1 | \|Severe acute respiratory syndrome coro | ···················· | ···················· | ···················· | ···················· | ···················· | ···················· | |
| 26100 | MT745730.1 | \|Severe acute respiratory syndrome coro | ···················· | ···················· | ···················· | ···················· | ···················· | ···················· | |
| 26101 | MT745731.1 | \|Severe acute respiratory syndrome coro | ···················· | ···················· | ···················· | ···················· | ···················· | ···················· | |
| 26102 | MT745732.1 | \|Severe acute respiratory syndrome coro | ···················· | ···················· | ···················· | ···················· | ···················· | ···················· | |
| 26103 | MT745733.1 | \|Severe acute respiratory syndrome coro | ···················· | ···················· | ···················· | ···················· | ···················· | ···················· | |
| 26105 | MT745735.1 | \|Severe acute respiratory syndrome coro | ···················· | ···················· | ···················· | ···················· | ···················· | ···················· | |
| 26106 | MT745736.1 | \|Severe acute respiratory syndrome coro | ···················· | ···················· | ···················· | ···················· | ···················· | ···················· | |
| 26107 | MT745737.1 | \|Severe acute respiratory syndrome coro | ···················· | ···················· | ···················· | ···················· | GCA················· | ···················· | 1 |
| 26108 | MT745738.1 | \|Severe acute respiratory syndrome coro | ···················· | ···················· | ···················· | ···················· | NNN················· | ···················· | 1 |
| 26109 | MT745739.1 | \|Severe acute respiratory syndrome coro | ···················· | ···················· | ···················· | ···················· | ···················· | ···················· | |
| 26110 | MT745740.1 | \|Severe acute respiratory syndrome coro | ···················· | ···················· | ···················· | ···················· | ···················· | ···················· | |
| 26111 | MT745741.1 | \|Severe acute respiratory syndrome coro | ···················· | ···················· | ···················· | ···················· | ···················· | ···················· | |
| 26112 | MT745742.1 | \|Severe acute respiratory syndrome coro | ···················· | ···················· | ···················· | ···················· | ···················· | ···················· | |
| 26113 | MT745743.1 | \|Severe acute respiratory syndrome coro | ···················· | ···················· | ···················· | ···················· | ···················· | ···················· | |
| 26114 | MT745744.1 | \|Severe acute respiratory syndrome coro | ···················· | ···················· | ···················· | ···················· | ···················· | ···················· | |
| 26116 | MT745746.1 | \|Severe acute respiratory syndrome coro | ···················· | ···················· | ···················· | ···················· | ···················· | ···················· | |
| 26117 | MT745747.1 | \|Severe acute respiratory syndrome coro | ···················· | ···················· | ···················· | ···················· | ···················· | ···················· | |
| 26118 | MT745748.1 | \|Severe acute respiratory syndrome coro | ···················· | ···················· | ···················· | ···················· | ···················· | ···················· | |
| 26119 | MT745749.1 | \|Severe acute respiratory syndrome coro | ···················· | ···················· | ···················· | ···················· | ···················· | ···················· | |
| 26120 | MT745767.1 | \|Severe acute respiratory syndrome coro | ···················· | ···················· | ···················· | ···················· | ···················· | ···················· | |
| 26121 | MT745836.1 | \|Severe acute respiratory syndrome coro | ···················· | ···················· | ···················· | ···················· | ···················· | ···················· | |
| 26122 | MT745875.1 | \|Severe acute respiratory syndrome coro | ···················· | ···················· | ···················· | ···················· | ···················· | ···················· | |
| 26123 | MT747438.1 | \|Severe acute respiratory syndrome coro | ···················· | ···················· | ···················· | ···················· | ···················· | ···················· | |
| 26124 | MT748758.1 | \|Severe acute respiratory syndrome coro | ···················· | ···················· | ···················· | ···················· | ···················· | ···················· | |
| 26125 | MT750020.1 | \|Severe acute respiratory syndrome coro | ···················· | ···················· | ···················· | ···················· | ···················· | ···················· | |
| 26126 | MT750021.1 | \|Severe acute respiratory syndrome coro | ···················· | ···················· | ···················· | ···················· | ···················· | ···················· | |
| 26127 | MT750022.1 | \|Severe acute respiratory syndrome coro | ···················· | ···················· | ···················· | ···················· | ···················· | ···················· | |
| 26128 | MT750023.1 | \|Severe acute respiratory syndrome coro | ···················· | ···················· | ···················· | ···················· | ···················· | ···················· | |
| 26129 | MT750024.1 | \|Severe acute respiratory syndrome coro | ···················· | ···················· | ···················· | ···················· | ···················· | ···················· | |
| 26130 | MT750025.1 | \|Severe acute respiratory syndrome coro | ·T·················· | ···················· | ···················· | ···················· | ···················· | ···················· | |
| 26131 | MT750026.1 | \|Severe acute respiratory syndrome coro | ···················· | ···················· | ···················· | ···················· | ···················· | ···················· | |
| 26132 | MT750027.1 | \|Severe acute respiratory syndrome coro | ···················· | ···················· | ···················· | ···················· | ···················· | ···················· | |
| 26133 | MT750028.1 | \|Severe acute respiratory syndrome coro | ···················· | ···················· | ···················· | ···················· | ···················· | ···················· | |
| 26134 | MT750029.1 | \|Severe acute respiratory syndrome coro | ···················· | ···················· | ···················· | ···················· | ···················· | ···················· | |
| 26135 | MT750030.1 | \|Severe acute respiratory syndrome coro | ···················· | ···················· | ···················· | ···················· | ···················· | ···················· | |
| 26136 | MT750031.1 | \|Severe acute respiratory syndrome coro | ···················· | ···················· | ···················· | ···················· | ···················· | ···················· | |
| 26137 | MT750032.1 | \|Severe acute respiratory syndrome coro | ···················· | ···················· | ···················· | ···················· | ···················· | ···················· | |
| 26138 | MT750033.1 | \|Severe acute respiratory syndrome coro | ···················· | ···················· | ···················· | ···················· | ···················· | ···················· | |
| 26139 | MT750034.1 | \|Severe acute respiratory syndrome coro | ···················· | ···················· | ···················· | ···················· | ···················· | ···················· | |
| 26140 | MT750035.1 | \|Severe acute respiratory syndrome coro | ···················· | ···················· | ···················· | ···················· | ···················· | ···················· | |
| 26141 | MT750036.1 | \|Severe acute respiratory syndrome coro | ···················· | ···················· | ···················· | ···················· | ···················· | ···················· | |
| 26142 | MT750037.1 | \|Severe acute respiratory syndrome coro | ···················· | ···················· | ···················· | ···················· | ···················· | ···················· | |
| 26143 | MT750038.1 | \|Severe acute respiratory syndrome coro | ···················· | ···················· | ···················· | ···················· | ···················· | ···················· | |
| 26144 | MT750039.1 | \|Severe acute respiratory syndrome coro | ···················· | ···················· | ···················· | ···················· | ···················· | ···················· | |
| 26145 | MT750040.1 | \|Severe acute respiratory syndrome coro | ···················· | ···················· | ···················· | ···················· | ···················· | ···················· | |
| 26146 | MT750041.1 | \|Severe acute respiratory syndrome coro | ·T·················· | ···················· | ···················· | ···················· | ···················· | ···················· | |
| 26147 | MT750042.1 | \|Severe acute respiratory syndrome coro | ·T·················· | ···················· | ···················· | ···················· | ···················· | ···················· | |
| 26148 | MT750043.1 | \|Severe acute respiratory syndrome coro | ···················· | ···················· | ···················· | ···················· | ···················· | ···················· | |
| 26149 | MT750044.1 | \|Severe acute respiratory syndrome coro | ···················· | ···················· | ···················· | ···················· | ···················· | ···················· | |
| 26150 | MT750045.1 | \|Severe acute respiratory syndrome coro | ···················· | ···················· | ···················· | ···················· | ···················· | ···················· | |
| 26151 | MT750046.1 | \|Severe acute respiratory syndrome coro | ···················· | ···················· | ···················· | ···················· | ···················· | ···················· | |
| 26152 | MT750047.1 | \|Severe acute respiratory syndrome coro | ···················· | ···················· | ···················· | ···················· | ···················· | ···················· | |
| 26153 | MT750048.1 | \|Severe acute respiratory syndrome coro | ···················· | ···················· | ···················· | ···················· | ···················· | ···················· | |
| 26154 | MT750049.1 | \|Severe acute respiratory syndrome coro | ···················· | ···················· | ···················· | ···················· | ···················· | ···················· | |
| 26155 | MT750050.1 | \|Severe acute respiratory syndrome coro | ···················· | ···················· | ···················· | ···················· | ···················· | ···················· | |
| 26156 | MT750051.1 | \|Severe acute respiratory syndrome coro | ···················· | ···················· | ···················· | ···················· | ···················· | ···················· | |
| 26157 | MT750052.1 | \|Severe acute respiratory syndrome coro | ···················· | ···················· | ···················· | ···················· | ···················· | ···················· | |
| 26158 | MT750053.1 | \|Severe acute respiratory syndrome coro | ···················· | ···················· | ···················· | ···················· | ···················· | ···················· | |
| 26159 | MT750054.1 | \|Severe acute respiratory syndrome coro | ···················· | ···················· | ···················· | ···················· | ···················· | ···················· | |
| 26160 | MT750055.1 | \|Severe acute respiratory syndrome coro | ···················· | ···················· | ···················· | ···················· | ···················· | ···················· | |
| 26161 | MT750056.1 | \|Severe acute respiratory syndrome coro | ···················· | ···················· | ···················· | ···················· | ···················· | ···················· | |
| 26162 | MT750057.1 | \|Severe acute respiratory syndrome coro | ···················· | ···················· | ···················· | ···················· | ···················· | ···················· | |
| 26163 | MT750058.1 | \|Severe acute respiratory syndrome coro | ···················· | ···················· | ···················· | ···················· | ···················· | ···················· | |
| 26164 | MT750059.1 | \|Severe acute respiratory syndrome coro | ···················· | ···················· | ···················· | ···················· | ···················· | ···················· | |
| 26165 | MT750060.1 | \|Severe acute respiratory syndrome coro | ···················· | ···················· | ···················· | ···················· | ···················· | ···················· | |
| 26166 | MT750061.1 | \|Severe acute respiratory syndrome coro | ···················· | ···················· | ···················· | ···················· | ···················· | ···················· | |
| 26167 | MT750062.1 | \|Severe acute respiratory syndrome coro | ···················· | ···················· | ···················· | ···················· | ···················· | ···················· | |
| 26168 | MT750063.1 | \|Severe acute respiratory syndrome coro | ···················· | ···················· | ···················· | ···················· | ···················· | ···················· | |
| 26169 | MT750064.1 | \|Severe acute respiratory syndrome coro | ···················· | ···················· | ···················· | ···················· | ···················· | ···················· | |
| 26170 | MT750065.1 | \|Severe acute respiratory syndrome coro | ···················· | ···················· | ···················· | ···················· | ···················· | ···················· | |
| 26171 | MT750066.1 | \|Severe acute respiratory syndrome coro | ···················· | ···················· | ···················· | ···················· | ···················· | ···················· | |
| 26172 | MT750067.1 | \|Severe acute respiratory syndrome coro | ···················· | ···················· | ···················· | ···················· | ···················· | ···················· | |

| | | | | | | | | |
|---|---|---|---|---|---|---|---|---|
| 26173 | MT750069.1 | \|Severe acute respiratory syndrome coro | .................... | .................... | ..................... | .................... | .................... | .................... |
| 26174 | MT750070.1 | \|Severe acute respiratory syndrome coro | .................... | .................... | ..................... | .................... | .................... | .................... |
| 26175 | MT750071.1 | \|Severe acute respiratory syndrome coro | .................... | .................... | ..................... | .................... | .................... | .................... |
| 26176 | MT750072.1 | \|Severe acute respiratory syndrome coro | .................... | .................... | ..................... | .................... | .................... | .................... |
| 26177 | MT750073.1 | \|Severe acute respiratory syndrome coro | .................... | .................... | ..................... | .................... | .................... | .................... |
| 26178 | MT750074.1 | \|Severe acute respiratory syndrome coro | .................... | .................... | ..................... | .................... | .................... | .................... |
| 26179 | MT750075.1 | \|Severe acute respiratory syndrome coro | .................... | .................... | ..................... | .................... | .................... | .................... |
| 26180 | MT750078.1 | \|Severe acute respiratory syndrome coro | .................... | .................... | ..................... | .................... | .................... | .................... |
| 26181 | MT750079.1 | \|Severe acute respiratory syndrome coro | .................... | .................... | ..................... | .................... | .................... | .................... |
| 26182 | MT750080.1 | \|Severe acute respiratory syndrome coro | .................... | .................... | ..................... | .................... | .................... | .................... |
| 26183 | MT750081.1 | \|Severe acute respiratory syndrome coro | .................... | .................... | ..................... | .................... | .................... | .................... |
| 26184 | MT750083.1 | \|Severe acute respiratory syndrome coro | .................... | .................... | ..................... | .................... | .................... | .................... |
| 26185 | MT750084.1 | \|Severe acute respiratory syndrome coro | .................... | .................... | ..................... | .................... | .................... | .................... |
| 26186 | MT750087.1 | \|Severe acute respiratory syndrome coro | .................... | .................... | ..................... | .................... | .................... | .................... |
| 26187 | MT750088.1 | \|Severe acute respiratory syndrome coro | .................... | .................... | ..................... | .................... | .................... | .................... |
| 26188 | MT750089.1 | \|Severe acute respiratory syndrome coro | .................... | .................... | ..................... | .................... | .................... | .................... |
| 26189 | MT750090.1 | \|Severe acute respiratory syndrome coro | .................... | .................... | ..................... | .................... | .................... | .................... |
| 26190 | MT750091.1 | \|Severe acute respiratory syndrome coro | .................... | .................... | ..................... | .................... | .................... | .................... |
| 26191 | MT750092.1 | \|Severe acute respiratory syndrome coro | .................... | .................... | ..................... | .................... | .................... | .................... |
| 26192 | MT750093.1 | \|Severe acute respiratory syndrome coro | .................... | .................... | ..................... | .................... | .................... | .................... |
| 26193 | MT750094.1 | \|Severe acute respiratory syndrome coro | .................... | .................... | ..................... | .................... | .................... | .................... |
| 26194 | MT750095.1 | \|Severe acute respiratory syndrome coro | .................... | .................... | ..................... | .................... | .................... | .................... |
| 26195 | MT750096.1 | \|Severe acute respiratory syndrome coro | .................... | .................... | ..................... | .................... | .................... | .................... |
| 26196 | MT750097.1 | \|Severe acute respiratory syndrome coro | .................... | .................... | ..................... | .................... | .................... | .................... |
| 26197 | MT750098.1 | \|Severe acute respiratory syndrome coro | .................... | .................... | ..................... | .................... | .................... | .................... |
| 26198 | MT750099.1 | \|Severe acute respiratory syndrome coro | .................... | .................... | ..................... | .................... | .................... | .................... |
| 26199 | MT750100.1 | \|Severe acute respiratory syndrome coro | .................... | .................... | ..................... | .................... | .................... | .................... |
| 26200 | MT750101.1 | \|Severe acute respiratory syndrome coro | .................... | .................... | ..................... | .................... | .................... | .................... |
| 26201 | MT750102.1 | \|Severe acute respiratory syndrome coro | .................... | .................... | ..................... | .................... | .................... | .................... |
| 26202 | MT750103.1 | \|Severe acute respiratory syndrome coro | .................... | .................... | ..................... | .................... | .................... | .................... |
| 26203 | MT750104.1 | \|Severe acute respiratory syndrome coro | .................... | .................... | ..................... | .................... | .................... | .................... |
| 26204 | MT750105.1 | \|Severe acute respiratory syndrome coro | .................... | .................... | ..................... | .................... | .................... | .................... |
| 26205 | MT750106.1 | \|Severe acute respiratory syndrome coro | .................... | .................... | ..................... | .................... | .................... | .................... |
| 26206 | MT750107.1 | \|Severe acute respiratory syndrome coro | .................... | .................... | ..................... | .................... | .................... | .................... |
| 26207 | MT750108.1 | \|Severe acute respiratory syndrome coro | .................... | .................... | ..................... | .................... | .................... | .................... |
| 26208 | MT750109.1 | \|Severe acute respiratory syndrome coro | .................... | .................... | ..................... | .................... | .................... | .................... |
| 26209 | MT750110.1 | \|Severe acute respiratory syndrome coro | .................... | .................... | ..................... | .................... | .................... | .................... |
| 26210 | MT750111.1 | \|Severe acute respiratory syndrome coro | .................... | .................... | ..................... | .................... | .................... | .................... |
| 26211 | MT750112.1 | \|Severe acute respiratory syndrome coro | .................... | .................... | ..................... | .................... | .................... | .................... |
| 26212 | MT750113.1 | \|Severe acute respiratory syndrome coro | .................... | .................... | ..................... | .................... | .................... | .................... |
| 26213 | MT750114.1 | \|Severe acute respiratory syndrome coro | .................... | .................... | ..................... | .................... | .................... | .................... |
| 26214 | MT750115.1 | \|Severe acute respiratory syndrome coro | .................... | .................... | ..................... | .................... | .................... | .................... |
| 26215 | MT750116.1 | \|Severe acute respiratory syndrome coro | .................... | .................... | ..................... | .................... | .................... | .................... |
| 26216 | MT750121.1 | \|Severe acute respiratory syndrome coro | .................... | .................... | ..................... | .................... | .................... | .................... |
| 26217 | MT750122.1 | \|Severe acute respiratory syndrome coro | .................... | .................... | ..................... | .................... | .................... | .................... |
| 26218 | MT750124.1 | \|Severe acute respiratory syndrome coro | .................... | .................... | ..................... | .................... | .................... | .................... |
| 26219 | MT750131.1 | \|Severe acute respiratory syndrome coro | .................... | .................... | ..................... | .................... | .................... | .................... |
| 26220 | MT750132.1 | \|Severe acute respiratory syndrome coro | .................... | .................... | ..................... | .................... | .................... | .................... |
| 26221 | MT750135.1 | \|Severe acute respiratory syndrome coro | .................... | .................... | ..................... | .................... | .................... | .................... |
| 26222 | MT750136.1 | \|Severe acute respiratory syndrome coro | .................... | .................... | ..................... | .................... | .................... | .................... |
| 26223 | MT750139.1 | \|Severe acute respiratory syndrome coro | .................... | .................... | ..................... | .................... | .................... | .................... |
| 26224 | MT750142.1 | \|Severe acute respiratory syndrome coro | .................... | .................... | ..................... | .................... | .................... | .................... |
| 26225 | MT750143.1 | \|Severe acute respiratory syndrome coro | .................... | .................... | ..................... | .................... | .................... | .................... |
| 26226 | MT750144.1 | \|Severe acute respiratory syndrome coro | .................... | .................... | ..................... | .................... | .................... | .................... |
| 26227 | MT750145.1 | \|Severe acute respiratory syndrome coro | .................... | .................... | ..................... | .................... | .................... | .................... |
| 26228 | MT750148.1 | \|Severe acute respiratory syndrome coro | .................... | .................... | ..................... | .................... | .................... | .................... |
| 26229 | MT750149.1 | \|Severe acute respiratory syndrome coro | .................... | .................... | ..................... | .................... | .................... | .................... |
| 26230 | MT750151.1 | \|Severe acute respiratory syndrome coro | .................... | .................... | ..................... | .................... | .................... | .................... |
| 26231 | MT750152.1 | \|Severe acute respiratory syndrome coro | .................... | .................... | ..................... | .................... | .................... | .................... |
| 26232 | MT750153.1 | \|Severe acute respiratory syndrome coro | .................... | .................... | ..................... | .................... | .................... | .................... |
| 26233 | MT750154.1 | \|Severe acute respiratory syndrome coro | .................... | .................... | ..................... | .................... | .................... | .................... |
| 26234 | MT750155.1 | \|Severe acute respiratory syndrome coro | .................... | .................... | ..................... | .................... | .................... | .................... |
| 26235 | MT750156.1 | \|Severe acute respiratory syndrome coro | .................... | .................... | ..................... | .................... | .................... | .................... |
| 26236 | MT750157.1 | \|Severe acute respiratory syndrome coro | .................... | .................... | ..................... | .................... | .................... | .................... |
| 26237 | MT750158.1 | \|Severe acute respiratory syndrome coro | .................... | .................... | ..................... | .................... | .................... | .................... |
| 26238 | MT750159.1 | \|Severe acute respiratory syndrome coro | .................... | .................... | ..................... | .................... | .................... | .................... |
| 26239 | MT750160.1 | \|Severe acute respiratory syndrome coro | .................... | .................... | ..................... | .................... | .................... | .................... |
| 26240 | MT750161.1 | \|Severe acute respiratory syndrome coro | .................... | .................... | ..................... | .................... | .................... | .................... |
| 26241 | MT750162.1 | \|Severe acute respiratory syndrome coro | .................... | .................... | ..................... | .................... | .................... | .................... |
| 26242 | MT750163.1 | \|Severe acute respiratory syndrome coro | .................... | .................... | ..................... | .................... | .................... | .................... |
| 26243 | MT750164.1 | \|Severe acute respiratory syndrome coro | .................... | .................... | ..................... | .................... | .................... | .................... |
| 26244 | MT750165.1 | \|Severe acute respiratory syndrome coro | .................... | .................... | ..................... | .................... | .................... | .................... |
| 26245 | MT750166.1 | \|Severe acute respiratory syndrome coro | .................... | .................... | ..................... | .................... | .................... | .................... |
| 26246 | MT750167.1 | \|Severe acute respiratory syndrome coro | .................... | .................... | ..................... | .................... | .................... | .................... |
| 26247 | MT750168.1 | \|Severe acute respiratory syndrome coro | .................... | .................... | ..................... | .................... | .................... | .................... |
| 26248 | MT750169.1 | \|Severe acute respiratory syndrome coro | .................... | .................... | ..................... | .................... | .................... | .................... |
| 26249 | MT750170.1 | \|Severe acute respiratory syndrome coro | .................... | .................... | ..................... | .................... | .................... | .................... |
| 26250 | MT750171.1 | \|Severe acute respiratory syndrome coro | .................... | .................... | ..................... | .................... | .................... | .................... |

```
26251 MT750172.1 |Severe acute respiratory syndrome coro   ....................  ....................  ....................  ....................  ....................  ....................
26252 MT750173.1 |Severe acute respiratory syndrome coro   ....................  ....................  ....................  ....................  ....................  ....................
26253 MT750174.1 |Severe acute respiratory syndrome coro   ....................  ....................  ....................  ....................  ....................  ....................
26254 MT750175.1 |Severe acute respiratory syndrome coro   ....................  ....................  ....................  ....................  ....................  ....................
26255 MT750176.1 |Severe acute respiratory syndrome coro   ....................  ....................  ....................  ....................  ....................  ....................
26256 MT750177.1 |Severe acute respiratory syndrome coro   ....................  ....................  ....................  ....................  ....................  ....................
26257 MT750178.1 |Severe acute respiratory syndrome coro   ....................  ....................  ....................  ....................  ....................  ....................
26258 MT750179.1 |Severe acute respiratory syndrome coro   ....................  ....................  ....................  ....................  ....................  ....................
26259 MT750180.1 |Severe acute respiratory syndrome coro   ....................  ....................  ....................  ....................  ....................  ....................
26260 MT750181.1 |Severe acute respiratory syndrome coro   ....................  ....................  ....................  ....................  ....................  ....................
26261 MT750182.1 |Severe acute respiratory syndrome coro   ....................  ....................  ....................  ....................  ....................  ....................
26262 MT750186.1 |Severe acute respiratory syndrome coro   ....................  ....................  ....................  ....................  ....................  ....................
26263 MT750187.1 |Severe acute respiratory syndrome coro   ....................  ....................  ....................  ....................  ....................  ....................
26264 MT750188.1 |Severe acute respiratory syndrome coro   ....................  ....................  ....................  ....................  ....................  ....................
26265 MT750189.1 |Severe acute respiratory syndrome coro   ....................  ....................  ....................  ....................  ....................  ....................
26266 MT750190.1 |Severe acute respiratory syndrome coro   ....................  ....................  ....................  ....................  ....................  ....................
26267 MT750191.1 |Severe acute respiratory syndrome coro   ....................  ....................  ....................  ....................  ....................  ....................
26268 MT750192.1 |Severe acute respiratory syndrome coro   ....................  ....................  ....................  ....................  ....................  ....................
26269 MT750194.1 |Severe acute respiratory syndrome coro   ....................  ....................  ....................  ....................  ....................  ....................
26270 MT750195.1 |Severe acute respiratory syndrome coro   ....................  ....................  ....................  ....................  ....................  ....................
26271 MT750197.1 |Severe acute respiratory syndrome coro   ....................  .A..................  ....................  ....................  ....................  ....................
26272 MT750198.1 |Severe acute respiratory syndrome coro   ....................  ....................  ....................  ....................  ....................  ....................
26273 MT750199.1 |Severe acute respiratory syndrome coro   ....................  ....................  ....................  ....................  ....................  ....................
26274 MT750200.1 |Severe acute respiratory syndrome coro   ....................  ....................  ....................  ....................  ....................  ....................
26275 MT750201.1 |Severe acute respiratory syndrome coro   ....................  ....................  ....................  ....................  ....................  ....................
26276 MT750333.1 |Severe acute respiratory syndrome coro   ....................  .A..................  ....................  ....................  ....................  ....................
26277 MT750334.1 |Severe acute respiratory syndrome coro   ....................  ....................  ....................  ....................  ....................  ....................
26278 MT750335.1 |Severe acute respiratory syndrome coro   ....................  ....................  ....................  ....................  ....................  ....................
26279 MT750336.1 |Severe acute respiratory syndrome coro   ....................  ....................  ....................  ....................  ....................  ....................
26280 MT750337.1 |Severe acute respiratory syndrome coro   ....................  ....................  ....................  ....................  ....................  ....................
26281 MT750338.1 |Severe acute respiratory syndrome coro   ....................  ....................  ....................  ....................  ....................  ....................
26282 MT750339.1 |Severe acute respiratory syndrome coro   ....................  ....................  ....................  ....................  ....................  ....................
26283 MT750340.1 |Severe acute respiratory syndrome coro   ....................  ....................  ....................  ....................  ....................  ....................
26284 MT750341.1 |Severe acute respiratory syndrome coro   ....................  ....................  ....................  ....................  ....................  ....................
26285 MT750342.1 |Severe acute respiratory syndrome coro   ....................  ....................  ....................  ....................  ....................  ....................
26286 MT750343.1 |Severe acute respiratory syndrome coro   ....................  ....................  ....................  ....................  ....................  ....................
26287 MT750344.1 |Severe acute respiratory syndrome coro   ....................  ....................  ....................  ....................  ....................  ....................
26288 MT750346.1 |Severe acute respiratory syndrome coro   ....................  ....................  ....................  ....................  ....................  ....................
26289 MT750347.1 |Severe acute respiratory syndrome coro   ....................  ....................  ....................  ....................  ....................  ....................
26290 MT750348.1 |Severe acute respiratory syndrome coro   ....................  ....................  ....................  ....................  ....................  ....................
26291 MT750350.1 |Severe acute respiratory syndrome coro   ....................  ....................  ....................  ....................  ....................  ....................
26292 MT750351.1 |Severe acute respiratory syndrome coro   ....................  ....................  ....................  ....................  ....................  ....................
26293 MT750352.1 |Severe acute respiratory syndrome coro   ....................  ....................  ....................  ....................  ....................  ....................
26294 MT750353.1 |Severe acute respiratory syndrome coro   ....................  ....................  ....................  ....................  ....................  ....................
26295 MT750354.1 |Severe acute respiratory syndrome coro   ....................  ....................  ....................  ....................  ....................  ....................
26296 MT750355.1 |Severe acute respiratory syndrome coro   ....................  ....................  ....................  ....................  ....................  ....................
26297 MT750356.1 |Severe acute respiratory syndrome coro   ....................  ....................  ....................  ....................  ....................  ....................
26298 MT750357.1 |Severe acute respiratory syndrome coro   ....................  ....................  ....................  ....................  ....................  ....................
26299 MT750358.1 |Severe acute respiratory syndrome coro   ....................  ....................  ....................  ....................  ....................  ....................
26300 MT750359.1 |Severe acute respiratory syndrome coro   ....................  ....................  ....................  ....................  ....................  ....................
26301 MT750361.1 |Severe acute respiratory syndrome coro   ....................  ....................  ....................  ....................  ....................  ....................
26302 MT750362.1 |Severe acute respiratory syndrome coro   ....................  ....................  ....................  ....................  ....................  ....................
26303 MT750363.1 |Severe acute respiratory syndrome coro   ....................  ....................  ....................  ....................  ....................  ....................
26304 MT750364.1 |Severe acute respiratory syndrome coro   ....................  ....................  ....................  ....................  ....................  ....................
26305 MT750365.1 |Severe acute respiratory syndrome coro   ....................  ....................  ....................  ....................  ....................  ....................
26306 MT750366.1 |Severe acute respiratory syndrome coro   ....................  ....................  ....................  ....................  ....................  ....................
26307 MT750367.1 |Severe acute respiratory syndrome coro   ....................  ....................  ....................  ....................  ....................  ....................
26308 MT750368.1 |Severe acute respiratory syndrome coro   ....................  ....................  ....................  ....................  ....................  ....................
26309 MT750369.1 |Severe acute respiratory syndrome coro   ....................  ....................  ....................  ....................  ....................  ....................
26310 MT750370.1 |Severe acute respiratory syndrome coro   ....................  ....................  ....................  ....................  ....................  ....................
26311 MT750371.1 |Severe acute respiratory syndrome coro   ....................  ....................  ....................  ....................  ......K.............  ....................
26312 MT750372.1 |Severe acute respiratory syndrome coro   ....................  ....................  ....................  ....................  ....................  ....................
26313 MT750373.1 |Severe acute respiratory syndrome coro   ....................  ....................  ....................  ....................  ....................  ....................
26314 MT750374.1 |Severe acute respiratory syndrome coro   ....................  ....................  ....................  ....................  ....................  ....................
26315 MT750377.1 |Severe acute respiratory syndrome coro   ....................  ....................  ....................  ....................  ....................  ....................
26316 MT750378.1 |Severe acute respiratory syndrome coro   ....................  ....................  ....................  ....................  ....................  ....................
26317 MT750379.1 |Severe acute respiratory syndrome coro   ....................  ....................  ....................  ....................  ....................  ....................
26318 MT750380.1 |Severe acute respiratory syndrome coro   ....................  ....................  ....................  ....................  ....................  ....................
26319 MT750381.1 |Severe acute respiratory syndrome coro   ....................  ....................  ....................  ....................  ....................  ....................
26320 MT750383.1 |Severe acute respiratory syndrome coro   ....................  ....................  ....................  ....................  ....................  ....................
26321 MT750384.1 |Severe acute respiratory syndrome coro   ....................  ....................  ....................  ....................  ....................  ....................
26322 MT750385.1 |Severe acute respiratory syndrome coro   ....................  ....................  ....................  ....................  ....................  ....................
26323 MT750386.1 |Severe acute respiratory syndrome coro   ....................  ....................  ....................  ....................  ....................  ....................
26324 MT750387.1 |Severe acute respiratory syndrome coro   ....................  ....................  ....................  ....................  ....................  ....................
26325 MT750389.1 |Severe acute respiratory syndrome coro   ....................  ....................  ....................  ....................  ....................  ....................
26326 MT750390.1 |Severe acute respiratory syndrome coro   ....................  ....................  ....................  ....................  ....................  ....................
26327 MT750393.1 |Severe acute respiratory syndrome coro   ....................  ....................  ....................  ....................  ....................  ....................
26328 MT750394.1 |Severe acute respiratory syndrome coro   ....................  ....................  ....................  ....................  ....................  ....................
```

```
26329 MT750395.1 |Severe acute respiratory syndrome coro ···················· ···················· ···················· ···················· ···················· ····················
26330 MT750396.1 |Severe acute respiratory syndrome coro ···················· ···················· ···················· ···················· ···················· ····················
26331 MT750397.1 |Severe acute respiratory syndrome coro ···················· ···················· ···················· ···················· ···················· ····················
26332 MT750401.1 |Severe acute respiratory syndrome coro ···················· ···················· ···················· ···················· ···················· ····················
26333 MT750402.1 |Severe acute respiratory syndrome coro ···················· ···················· ···················· ···················· ···················· ····················
26334 MT750404.1 |Severe acute respiratory syndrome coro ···················· ···················· ···················· ···················· ···················· ····················
26335 MT750405.1 |Severe acute respiratory syndrome coro ···················· ···················· ···················· ···················· ···················· ····················
26336 MT750406.1 |Severe acute respiratory syndrome coro ···················· ···················· ···················· ···················· ···················· ····················
26337 MT750407.1 |Severe acute respiratory syndrome coro ···················· ···················· ···················· ···················· ···················· ····················
26338 MT750408.1 |Severe acute respiratory syndrome coro ···················· ···················· ···················· ···················· ···················· ····················
26339 MT750409.1 |Severe acute respiratory syndrome coro ···················· ···················· ···················· ···················· ···················· ····················
26340 MT750410.1 |Severe acute respiratory syndrome coro ···················· ···················· ···················· ···················· ···················· ····················
26341 MT750411.1 |Severe acute respiratory syndrome coro ···················· ···················· ···················· ···················· ···················· ····················
26342 MT750412.1 |Severe acute respiratory syndrome coro ···················· ···················· ···················· ···················· ···················· ····················
26343 MT750413.1 |Severe acute respiratory syndrome coro ···T················ ···················· ···················· ···················· ···················· ····················
26344 MT750414.1 |Severe acute respiratory syndrome coro ···················· ···················· ···················· ···················· ···················· ····················
26345 MT750415.1 |Severe acute respiratory syndrome coro ···················· ···················· ···················· ···················· ···················· ····················
26346 MT750416.1 |Severe acute respiratory syndrome coro ···················· ···················· ···················· ···················· ···················· ····················
26347 MT750417.1 |Severe acute respiratory syndrome coro ···················· ···················· ···················· ···················· ···················· ····················
26348 MT750418.1 |Severe acute respiratory syndrome coro ···················· ···················· ···················· ···················· ···················· ····················
26349 MT750419.1 |Severe acute respiratory syndrome coro ···················· ···················· ···················· ···················· ···················· ····················
26350 MT750420.1 |Severe acute respiratory syndrome coro ···················· ···················· ···················· ···················· ···················· ····················
26351 MT750421.1 |Severe acute respiratory syndrome coro ···················· ···················· ···················· ···················· ···················· ····················
26352 MT750422.1 |Severe acute respiratory syndrome coro ···················· ···················· ···················· ···················· ···················· ····················
26353 MT750423.1 |Severe acute respiratory syndrome coro ···················· ···················· ···················· ···················· ···················· ····················
26354 MT750424.1 |Severe acute respiratory syndrome coro ···················· ···················· ···················· ···················· ···················· ····················
26355 MT750425.1 |Severe acute respiratory syndrome coro ···················· ···················· ···················· ···················· ···················· ····················
26356 MT750426.1 |Severe acute respiratory syndrome coro ···················· ···················· ···················· ···················· ···················· ····················
26357 MT750427.1 |Severe acute respiratory syndrome coro ···T················ ···················· ···················· ···················· ···················· ····················
26358 MT750429.1 |Severe acute respiratory syndrome coro ···················· ···················· ···················· ···················· ···················· ····················
26359 MT750430.1 |Severe acute respiratory syndrome coro ···················· ···················· ···················· ···················· ···················· ····················
26360 MT750431.1 |Severe acute respiratory syndrome coro ···················· ···················· ···················· ···················· ···················· ····················
26361 MT750432.1 |Severe acute respiratory syndrome coro ···················· ···················· ···················· ···················· ···················· ····················
26362 MT750433.1 |Severe acute respiratory syndrome coro ···················· ···················· ···················· ···················· ···················· ····················
26363 MT750434.1 |Severe acute respiratory syndrome coro ···················· ···················· ···················· ···················· ···················· ····················
26364 MT750436.1 |Severe acute respiratory syndrome coro ···················· ···················· ···················· ···················· ···················· ····················
26365 MT750437.1 |Severe acute respiratory syndrome coro ···················· ···················· ···················· ···················· ···················· ····················
26366 MT750439.1 |Severe acute respiratory syndrome coro ···················· ···················· ···················· ···················· ···················· ····················
26367 MT750440.1 |Severe acute respiratory syndrome coro ···················· ···················· ···················· ···················· ···················· ····················
26368 MT750441.1 |Severe acute respiratory syndrome coro ···················· ···················· ···················· ···················· ···················· ····················
26369 MT750442.1 |Severe acute respiratory syndrome coro ···················· ···················· ···················· ···················· ···················· ····················
26370 MT750443.1 |Severe acute respiratory syndrome coro ···················· ···················· ···················· ···················· ···················· ····················
26371 MT750444.1 |Severe acute respiratory syndrome coro ···················· ···················· ···················· ···················· ···················· ····················
26372 MT750446.1 |Severe acute respiratory syndrome coro ···················· ···················· ···················· ···················· ···················· ····················
26373 MT750447.1 |Severe acute respiratory syndrome coro ···················· ···················· ···················· ···················· ···················· ····················
26374 MT750448.1 |Severe acute respiratory syndrome coro ···················· ···················· ···················· ···················· ···················· ····················
26375 MT750450.1 |Severe acute respiratory syndrome coro ···················· ···················· ···················· ···················· ···················· ····················
26376 MT750451.1 |Severe acute respiratory syndrome coro ···················· ···················· ···················· ···················· ···················· ····················
26377 MT750452.1 |Severe acute respiratory syndrome coro ···················· ···················· ···················· ···················· ···················· ····················
26378 MT750453.1 |Severe acute respiratory syndrome coro ···················· ···················· ···················· ···················· ···················· ····················
26379 MT750455.1 |Severe acute respiratory syndrome coro ···················· ···················· ···················· ···················· ···················· ····················
26380 MT750456.1 |Severe acute respiratory syndrome coro ···················· ···················· ···················· ···················· ···················· ····················
26381 MT750457.1 |Severe acute respiratory syndrome coro ···················· ···················· ···················· ···················· ···················· ····················
26382 MT750458.1 |Severe acute respiratory syndrome coro ···················· ···················· ···················· ···················· ···················· ····················
26383 MT750459.1 |Severe acute respiratory syndrome coro ···················· ···················· ···················· ···················· ···················· ····················
26384 MT750460.1 |Severe acute respiratory syndrome coro ···················· ···················· ···················· ···················· ···················· ····················
26385 MT750462.1 |Severe acute respiratory syndrome coro ···················· ···················· ···················· ···················· ···················· ····················
26386 MT750463.1 |Severe acute respiratory syndrome coro ···················· ···················· ···················· ···················· ···················· ····················
26387 MT750464.1 |Severe acute respiratory syndrome coro ···················· ···················· ···················· ···················· ···················· ····················
26388 MT750465.1 |Severe acute respiratory syndrome coro ···················· ···················· ···················· ···················· ···················· ····················
26389 MT750466.1 |Severe acute respiratory syndrome coro ···················· ···················· ···················· ···················· ···················· ····················
26390 MT750467.1 |Severe acute respiratory syndrome coro ···················· ···················· ···················· ···················· ···················· ····················
26391 MT750468.1 |Severe acute respiratory syndrome coro ···················· ···················· ···················· ···················· ···················· ····················
26392 MT750469.1 |Severe acute respiratory syndrome coro ···················· ···················· ···················· ···················· ···················· ····················
26393 MT750470.1 |Severe acute respiratory syndrome coro ···················· ···················· ···················· ···················· ···················· ····················
26394 MT750471.1 |Severe acute respiratory syndrome coro ···················· ···················· ···················· ···················· ···················· ····················
26395 MT750472.1 |Severe acute respiratory syndrome coro ···················· ···················· ···················· ···················· ···················· ····················
26396 MT750473.1 |Severe acute respiratory syndrome coro ···················· ···················· ···················· ···················· ···················· ····················
26397 MT750474.1 |Severe acute respiratory syndrome coro ···················· ···················· ···················· ···················· ···················· ····················
26398 MT750475.1 |Severe acute respiratory syndrome coro ···················· ···················· ···················· ···················· ···················· ····················
26399 MT750476.1 |Severe acute respiratory syndrome coro ···················· ···················· ···················· ···················· ···················· ····················
26400 MT750477.1 |Severe acute respiratory syndrome coro ···················· ···················· ···················· ···················· ···················· ····················
26401 MT750478.1 |Severe acute respiratory syndrome coro ···················· ···················· ···················· ···················· ···················· ····················
26402 MT750479.1 |Severe acute respiratory syndrome coro ···················· ···················· ···················· ···················· ···················· ····················
26403 MT750480.1 |Severe acute respiratory syndrome coro ···················· ···················· ···················· ···················· ···················· ····················
26404 MT750481.1 |Severe acute respiratory syndrome coro ···················· ···················· ···················· ···················· ···················· ····················
26405 MT750482.1 |Severe acute respiratory syndrome coro ···················· ···················· ···················· ···················· ···················· ····················
26406 MT750483.1 |Severe acute respiratory syndrome coro ···················· ···················· ···················· ···················· ···················· ····················
```

| | | | | | | | | |
|---|---|---|---|---|---|---|---|---|
| 26407 | MT750484.1 | \|Severe acute respiratory syndrome coro | ···················· | ···················· | ···················· | ···················· | ···················· | ···················· |
| 26408 | MT750485.1 | \|Severe acute respiratory syndrome coro | ···················· | ···················· | ···················· | ···················· | ···················· | ···················· |
| 26409 | MT750486.1 | \|Severe acute respiratory syndrome coro | ···················· | ···················· | ···················· | ···················· | ···················· | ···················· |
| 26410 | MT750487.1 | \|Severe acute respiratory syndrome coro | ···················· | ···················· | ···················· | ···················· | ···················· | ···················· |
| 26411 | MT750488.1 | \|Severe acute respiratory syndrome coro | ···················· | ···················· | ···················· | ···················· | ···················· | ···················· |
| 26412 | MT750490.1 | \|Severe acute respiratory syndrome coro | ···················· | ···················· | ···················· | ···················· | ···················· | ···················· |
| 26413 | MT750491.1 | \|Severe acute respiratory syndrome coro | ···················· | ···················· | ···················· | ···················· | ···················· | ···················· |
| 26414 | MT750492.1 | \|Severe acute respiratory syndrome coro | ···················· | ···················· | ···················· | ···················· | ···················· | ···················· |
| 26415 | MT750493.1 | \|Severe acute respiratory syndrome coro | ···················· | ···················· | ···················· | ···················· | ···················· | ···················· |
| 26416 | MT750495.1 | \|Severe acute respiratory syndrome coro | ···················· | ···················· | ···················· | ···················· | ···················· | ···················· |
| 26417 | MT750497.1 | \|Severe acute respiratory syndrome coro | ···················· | ···················· | ···················· | ···················· | ···················· | ···················· |
| 26418 | MT750498.1 | \|Severe acute respiratory syndrome coro | ···················· | ···················· | ···················· | ···················· | ···················· | ···················· |
| 26419 | MT750500.1 | \|Severe acute respiratory syndrome coro | ···················· | ···················· | ···················· | ···················· | ···················· | ···················· |
| 26420 | MT750501.1 | \|Severe acute respiratory syndrome coro | ···················· | ···················· | ···················· | ···················· | ···················· | ···················· |
| 26421 | MT750502.1 | \|Severe acute respiratory syndrome coro | ···················· | ···················· | ···················· | ···················· | ···················· | ···················· |
| 26422 | MT750503.1 | \|Severe acute respiratory syndrome coro | ···················· | ···················· | ···················· | ···················· | ···················· | ···················· |
| 26423 | MT750504.1 | \|Severe acute respiratory syndrome coro | ···················· | ···················· | ···················· | ···················· | ···················· | ···················· |
| 26424 | MT750505.1 | \|Severe acute respiratory syndrome coro | ···················· | ···················· | ···················· | ···················· | ···················· | ···················· |
| 26425 | MT750506.1 | \|Severe acute respiratory syndrome coro | ···················· | ···················· | ···················· | ···················· | ···················· | ···················· |
| 26426 | MT750507.1 | \|Severe acute respiratory syndrome coro | ···················· | ···················· | ···················· | ···················· | ···················· | ···················· |
| 26427 | MT750509.1 | \|Severe acute respiratory syndrome coro | ···················· | ···················· | ···················· | ···················· | ···················· | ···················· |
| 26428 | MT750510.1 | \|Severe acute respiratory syndrome coro | ···················· | ···················· | ···················· | ···················· | ···················· | ···················· |
| 26429 | MT742782.1 | \|Severe acute respiratory syndrome coro | ···················· | ···················· | ···················· | ···················· | ···················· | ···················· |
| 26430 | MT742783.1 | \|Severe acute respiratory syndrome coro | ···················· | ···················· | ···················· | ···················· | ···················· | ···················· |
| 26431 | MT742784.1 | \|Severe acute respiratory syndrome coro | ···················· | ···················· | ···················· | ···················· | ···················· | ···················· |
| 26432 | LC565407.1 | \|Severe acute respiratory syndrome coro | ···················· | ···················· | ···················· | ···················· | ···················· | ···················· |
| 26433 | LC565408.1 | \|Severe acute respiratory syndrome coro | ···················· | ···················· | ···················· | ···················· | ···················· | ···················· |
| 26434 | LC565409.1 | \|Severe acute respiratory syndrome coro | ···················· | ···················· | ···················· | ···················· | ···················· | ···················· |
| 26435 | LC565410.1 | \|Severe acute respiratory syndrome coro | ···················· | ···················· | ···················· | ···················· | ···················· | ···················· |
| 26436 | LC565411.1 | \|Severe acute respiratory syndrome coro | ···················· | ···················· | ···················· | ···················· | ···················· | ···················· |
| 26437 | LC565412.1 | \|Severe acute respiratory syndrome coro | ···················· | ···················· | ···················· | ···················· | ···················· | ···················· |
| 26438 | LC565413.1 | \|Severe acute respiratory syndrome coro | ···················· | ···················· | ···················· | ···················· | ···················· | ···················· |
| 26439 | LC565414.1 | \|Severe acute respiratory syndrome coro | ···················· | ···················· | ···················· | ···················· | ···················· | ···················· |
| 26440 | LC567845.1 | \|Severe acute respiratory syndrome coro | ···················· | ···················· | ···················· | ···················· | ···················· | ···················· |
| 26441 | LC567846.1 | \|Severe acute respiratory syndrome coro | ···················· | ···················· | ···················· | ···················· | ···················· | ···················· |
| 26442 | LC567847.1 | \|Severe acute respiratory syndrome coro | ···················· | ···················· | ···················· | ···················· | ···················· | ···················· |
| 26443 | LC567848.1 | \|Severe acute respiratory syndrome coro | ···················· | ···················· | ···················· | ···················· | ···················· | ···················· |
| 26444 | LC567849.1 | \|Severe acute respiratory syndrome coro | ···················· | ···················· | ···················· | ···················· | ···················· | ···················· |
| 26445 | LC567850.1 | \|Severe acute respiratory syndrome coro | ···················· | ···················· | ···················· | ···················· | ···················· | ···················· |
| 26446 | LC567851.1 | \|Severe acute respiratory syndrome coro | ···················· | ···················· | ···················· | ···················· | ···················· | ···················· |
| 26447 | LC567852.1 | \|Severe acute respiratory syndrome coro | ···················· | ···················· | ···················· | ···················· | ···················· | ···················· |
| 26448 | LC567853.1 | \|Severe acute respiratory syndrome coro | ···················· | ···················· | ···················· | ···················· | ···················· | ···················· |
| 26449 | LC567854.1 | \|Severe acute respiratory syndrome coro | ···················· | ···················· | ···················· | ···················· | ···················· | ···················· |
| 26450 | LC567855.1 | \|Severe acute respiratory syndrome coro | ···················· | ···················· | ···················· | ···················· | ···················· | ···················· |
| 26451 | LC567856.1 | \|Severe acute respiratory syndrome coro | ···················· | ···················· | ···················· | ···················· | ···················· | ···················· |
| 26452 | LC567857.1 | \|Severe acute respiratory syndrome coro | ···················· | ···················· | ···················· | ···················· | ···················· | ···················· |
| 26453 | LC567858.1 | \|Severe acute respiratory syndrome coro | ···················· | ···················· | ···················· | ···················· | ···················· | ···················· |
| 26454 | LC567859.1 | \|Severe acute respiratory syndrome coro | ···················· | ···················· | ···················· | ···················· | ···················· | ···················· |
| 26455 | LR824570.1 | \|Severe acute respiratory syndrome coro | ···················· | ···················· | ···················· | ···················· | ···················· | ···················· |
| 26456 | LR824571.1 | \|Severe acute respiratory syndrome coro | ···················· | ···················· | ···················· | ···················· | ···················· | ···················· |
| 26457 | MT740381.1 | \|Severe acute respiratory syndrome coro | ···················· | ···················· | ···················· | ···················· | ···················· | ···················· |
| 26458 | MT740382.1 | \|Severe acute respiratory syndrome coro | ···················· | ···················· | ···················· | ···················· | ···················· | ···················· |
| 26459 | MT740383.1 | \|Severe acute respiratory syndrome coro | ···················· | ···················· | ···················· | ···················· | ···················· | ···················· |
| 26460 | MT740384.1 | \|Severe acute respiratory syndrome coro | ···················· | ···················· | ···················· | ···················· | ···················· | ···················· |
| 26461 | MT740390.1 | \|Severe acute respiratory syndrome coro | ···················· | ···················· | ···················· | ···················· | ···················· | ···················· |
| 26462 | MT740391.1 | \|Severe acute respiratory syndrome coro | ···················· | ···················· | ···················· | ···················· | ···················· | ···················· |
| 26463 | MT740392.1 | \|Severe acute respiratory syndrome coro | ···················· | ···················· | ···················· | ···················· | ···················· | ···················· |
| 26464 | MT740393.1 | \|Severe acute respiratory syndrome coro | ···················· | ···················· | ···················· | ···················· | ···················· | ···················· |
| 26465 | MT740394.1 | \|Severe acute respiratory syndrome coro | ···················· | ···················· | ···················· | ···················· | ···················· | ···················· |
| 26466 | MT740395.1 | \|Severe acute respiratory syndrome coro | ···················· | ···················· | ···················· | ···················· | ···················· | ···················· |
| 26467 | MT740396.1 | \|Severe acute respiratory syndrome coro | ···················· | ···················· | ···················· | ···················· | ···················· | ···················· |
| 26468 | MT740397.1 | \|Severe acute respiratory syndrome coro | ···················· | ···················· | ···················· | ···················· | ···················· | ···················· |
| 26469 | MT740401.1 | \|Severe acute respiratory syndrome coro | ···················· | ···················· | ···················· | ···················· | ···················· | ···················· |
| 26470 | MT740402.1 | \|Severe acute respiratory syndrome coro | ···················· | ···················· | ···················· | ···················· | ···················· | ···················· |
| 26471 | MT740403.1 | \|Severe acute respiratory syndrome coro | ···················· | ···················· | ···················· | ···················· | ···················· | ···················· |
| 26472 | MT740404.1 | \|Severe acute respiratory syndrome coro | ···················· | ···················· | ···················· | ···················· | ···················· | ···················· |
| 26473 | MT740431.1 | \|Severe acute respiratory syndrome coro | ···················· | ···················· | ···················· | ···················· | ···················· | ···················· |
| 26474 | MT740432.1 | \|Severe acute respiratory syndrome coro | ···················· | ···················· | ···················· | ···················· | ···················· | ···················· |
| 26475 | MT740434.1 | \|Severe acute respiratory syndrome coro | ···················· | ···················· | ···················· | ···················· | ···················· | ···················· |
| 26476 | MT740435.1 | \|Severe acute respiratory syndrome coro | ···················· | ···················· | ···················· | ···················· | ···················· | ···················· |
| 26477 | MT740436.1 | \|Severe acute respiratory syndrome coro | ···················· | ···················· | ···················· | ···················· | ···················· | ···················· |
| 26478 | MT740438.1 | \|Severe acute respiratory syndrome coro | ···················· | ···················· | ···················· | ···················· | ···················· | ···················· |
| 26479 | MT740439.1 | \|Severe acute respiratory syndrome coro | ···················· | ···················· | ···················· | ···················· | ···················· | ···················· |
| 26480 | MT740440.1 | \|Severe acute respiratory syndrome coro | ···················· | ···················· | ···················· | ···················· | ···················· | ···················· |
| 26481 | MT740441.1 | \|Severe acute respiratory syndrome coro | ···················· | ····A··············· | ···················· | ···················· | ···················· | ···················· |
| 26482 | MT740444.1 | \|Severe acute respiratory syndrome coro | ···T················ | ·A·················· | ···················· | ···················· | ···················· | ···A················ |
| 26483 | MT740476.1 | \|Severe acute respiratory syndrome coro | ···················· | ···················· | ···················· | ···················· | ···················· | ···················· |
| 26484 | MT740477.1 | \|Severe acute respiratory syndrome coro | ···················· | ···················· | ···················· | ···················· | ···················· | ···················· |

| | | | | | | | | |
|---|---|---|---|---|---|---|---|---|
| 26485 | MT740478.1 | \|Severe acute respiratory syndrome coro | ···················· | ···················· | ···················· | ···················· | ···················· | ···················· |
| 26486 | MT740479.1 | \|Severe acute respiratory syndrome coro | ···················· | ···················· | ···················· | ···················· | ···················· | ···················· |
| 26487 | MT740480.1 | \|Severe acute respiratory syndrome coro | ···················· | ···················· | ···················· | ···················· | ···················· | ···················· |
| 26488 | MT740483.1 | \|Severe acute respiratory syndrome coro | ···················· | ···················· | ···················· | ···················· | ···················· | ···················· |
| 26489 | MT740485.1 | \|Severe acute respiratory syndrome coro | ···················· | ·A·················· | ···················· | ···················· | ···················· | ··A················· |
| 26490 | MT740617.1 | \|Severe acute respiratory syndrome coro | ···················· | ·A·················· | ···················· | ···················· | ···················· | ··A················· |
| 26491 | MT740618.1 | \|Severe acute respiratory syndrome coro | ···················· | ·A·················· | ···················· | ···················· | ···················· | ··A················· |
| 26492 | MT740619.1 | \|Severe acute respiratory syndrome coro | ···················· | ·A·················· | ···················· | ···················· | ···················· | ··A················· |
| 26493 | MT740684.1 | \|Severe acute respiratory syndrome coro | ···················· | ·A·················· | ···················· | ···················· | ···················· | ··A················· |
| 26494 | MT740686.1 | \|Severe acute respiratory syndrome coro | ···················· | ···················· | ···················· | ···················· | ···················· | ···················· |
| 26495 | MT740688.1 | \|Severe acute respiratory syndrome coro | ···················· | ···················· | ···················· | ···················· | ···················· | ···················· |
| 26496 | MT740706.1 | \|Severe acute respiratory syndrome coro | ···················· | ·A·················· | ···················· | ···················· | ···················· | ··A················· |
| 26497 | MT740708.1 | \|Severe acute respiratory syndrome coro | ···················· | ·A·················· | ···················· | ···················· | ···················· | ··A················· |
| 26498 | MT740717.1 | \|Severe acute respiratory syndrome coro | ···················· | ·A·················· | ···················· | ···················· | ···················· | ··A················· |
| 26499 | MT740723.1 | \|Severe acute respiratory syndrome coro | ···················· | ·A·················· | ···················· | ···················· | ···················· | ··A················· |
| 26500 | MT740724.1 | \|Severe acute respiratory syndrome coro | ···················· | ·A·················· | ···················· | ···················· | ···················· | ··A················· |
| 26501 | MT740820.1 | \|Severe acute respiratory syndrome coro | ···················· | ···················· | ···················· | ···················· | ···················· | ··A················· |
| 26502 | MT740896.1 | \|Severe acute respiratory syndrome coro | ···················· | ·A·················· | ···················· | ···················· | ···················· | ···················· |
| 26503 | MT740923.1 | \|Severe acute respiratory syndrome coro | ···················· | ···················· | ···················· | ···················· | ···················· | ···················· |
| 26504 | MT741096.1 | \|Severe acute respiratory syndrome coro | ···················· | ···················· | ···················· | ···················· | ···················· | ···················· |
| 26505 | MT741490.1 | \|Severe acute respiratory syndrome coro | ···················· | ···················· | ···················· | ···················· | ···················· | ··A················· |
| 26506 | MT742087.1 | \|Severe acute respiratory syndrome coro | ···················· | ···················· | ···················· | ···················· | ···················· | ···················· |
| 26507 | MT742088.1 | \|Severe acute respiratory syndrome coro | ···················· | ···················· | ···················· | ···················· | ·····T·············· | ···················· |
| 26508 | MT742089.1 | \|Severe acute respiratory syndrome coro | ···················· | ···················· | ···················· | ···················· | ···················· | ···················· |
| 26509 | MT742090.1 | \|Severe acute respiratory syndrome coro | ···················· | ···················· | ···················· | ···················· | ···················· | ···················· |
| 26510 | MT742091.1 | \|Severe acute respiratory syndrome coro | ···················· | ···················· | ···················· | ···················· | ···················· | ···················· |
| 26511 | MT742092.1 | \|Severe acute respiratory syndrome coro | ···················· | ···················· | ···················· | ···················· | ···················· | ···················· |
| 26512 | MT734046.1 | \|Severe acute respiratory syndrome coro | ···················· | ···················· | ···················· | ···················· | ···················· | ···················· |
| 26513 | MT734055.1 | \|Severe acute respiratory syndrome coro | ···················· | ···················· | ···················· | ···················· | ···················· | ···················· |
| 26514 | MT738101.1 | \|Severe acute respiratory syndrome coro | ···················· | ···················· | ···················· | ···················· | ···················· | ···················· |
| 26515 | MT738173.1 | \|Severe acute respiratory syndrome coro | ···················· | ···················· | ···················· | ···················· | ···················· | ···················· |
| 26516 | MT739425.1 | \|Severe acute respiratory syndrome coro | ···················· | ···················· | ···················· | ···················· | ···················· | ···················· |
| 26517 | MT739426.1 | \|Severe acute respiratory syndrome coro | ···················· | ···················· | ···················· | ···················· | ···················· | ···················· |
| 26518 | MT739427.1 | \|Severe acute respiratory syndrome coro | ···················· | ···················· | ···················· | ···················· | ···················· | ···················· |
| 26519 | MT739428.1 | \|Severe acute respiratory syndrome coro | ···················· | ···················· | ···················· | ···················· | ···················· | ···················· |
| 26520 | MT739429.1 | \|Severe acute respiratory syndrome coro | ···················· | ···················· | ···················· | ···················· | ···················· | ···················· |
| 26521 | MT739430.1 | \|Severe acute respiratory syndrome coro | ···················· | ···················· | ···················· | ···················· | ···················· | ···················· |
| 26522 | MT739431.1 | \|Severe acute respiratory syndrome coro | ···················· | ···················· | ···················· | ···················· | ···················· | ···················· |
| 26523 | MT739432.1 | \|Severe acute respiratory syndrome coro | ···················· | ···················· | ···················· | ···················· | ···················· | ···················· |
| 26524 | MT739433.1 | \|Severe acute respiratory syndrome coro | ···················· | ···················· | ···················· | ···················· | ···················· | ···················· |
| 26525 | MT739434.1 | \|Severe acute respiratory syndrome coro | ···················· | ···················· | ···················· | ···················· | ···················· | ···················· |
| 26526 | MT739435.1 | \|Severe acute respiratory syndrome coro | ···················· | ···················· | ···················· | ···················· | ···················· | ···················· |
| 26527 | MT739436.1 | \|Severe acute respiratory syndrome coro | ···················· | ···················· | ···················· | ···················· | ···················· | ···················· |
| 26528 | MT739437.1 | \|Severe acute respiratory syndrome coro | ···················· | ···················· | ···················· | ···················· | ···················· | ···················· |
| 26529 | MT739438.1 | \|Severe acute respiratory syndrome coro | ···················· | ···················· | ···················· | ···················· | ···················· | ···················· |
| 26530 | MT739439.1 | \|Severe acute respiratory syndrome coro | ···················· | ···················· | ···················· | ···················· | ···················· | ···················· |
| 26531 | MT739440.1 | \|Severe acute respiratory syndrome coro | ···················· | ···················· | ···················· | ···················· | ···················· | ···················· |
| 26532 | MT739441.1 | \|Severe acute respiratory syndrome coro | ···················· | ···················· | ···················· | ···················· | ···················· | ···················· |
| 26533 | MT739442.1 | \|Severe acute respiratory syndrome coro | ···················· | ···················· | ···················· | ···················· | ···················· | ···················· |
| 26534 | MT739443.1 | \|Severe acute respiratory syndrome coro | ···················· | ···················· | ···················· | ···················· | ···················· | ···················· |
| 26535 | MT739444.2 | \|Severe acute respiratory syndrome coro | ···················· | ···················· | ···················· | ···················· | ···················· | ···················· |
| 26536 | MT739445.1 | \|Severe acute respiratory syndrome coro | ···················· | ···················· | ···················· | ···················· | ···················· | ···················· |
| 26537 | MT739446.1 | \|Severe acute respiratory syndrome coro | ···················· | ···················· | ···················· | ···················· | ···················· | ···················· |
| 26538 | MT739448.1 | \|Severe acute respiratory syndrome coro | ···················· | ···················· | ···················· | ···················· | ···················· | ···················· |
| 26539 | MT739449.1 | \|Severe acute respiratory syndrome coro | ···················· | ···················· | ···················· | ···················· | ···················· | ···················· |
| 26540 | MT739450.1 | \|Severe acute respiratory syndrome coro | ···················· | ···················· | ···················· | ···················· | ···················· | ···················· |
| 26541 | MT739451.1 | \|Severe acute respiratory syndrome coro | ···················· | ···················· | ···················· | ···················· | ···················· | ···················· |
| 26542 | MT739452.1 | \|Severe acute respiratory syndrome coro | ···················· | ···················· | ···················· | ···················· | ···················· | ···················· |
| 26543 | MT739453.1 | \|Severe acute respiratory syndrome coro | ···················· | ···················· | ···················· | ···················· | ···················· | ···················· |
| 26544 | MT739454.1 | \|Severe acute respiratory syndrome coro | ···················· | ···················· | ···················· | ···················· | ···················· | ···················· |
| 26545 | MT739455.1 | \|Severe acute respiratory syndrome coro | ···················· | ···················· | ···················· | ···················· | ···················· | ···················· |
| 26546 | MT739456.1 | \|Severe acute respiratory syndrome coro | ···················· | ···················· | ···················· | ···················· | ···················· | ···················· |
| 26547 | MT739457.1 | \|Severe acute respiratory syndrome coro | ···················· | ···················· | ···················· | ···················· | ···················· | ···················· |
| 26548 | MT739458.1 | \|Severe acute respiratory syndrome coro | ···················· | ···················· | ···················· | ···················· | ···················· | ···················· |
| 26549 | MT739460.1 | \|Severe acute respiratory syndrome coro | ···················· | ···················· | ···················· | ···················· | ···················· | ···················· |
| 26550 | MT739461.1 | \|Severe acute respiratory syndrome coro | ···················· | ···················· | ···················· | ···················· | ···················· | ···················· |
| 26551 | MT739462.1 | \|Severe acute respiratory syndrome coro | ···················· | ···················· | ···················· | ···················· | ···················· | ···················· |
| 26552 | MT739463.1 | \|Severe acute respiratory syndrome coro | ···················· | ···················· | ···················· | ···················· | ···················· | ···················· |
| 26553 | MT739464.1 | \|Severe acute respiratory syndrome coro | ···················· | ···················· | ···················· | ···················· | ···················· | ···················· |
| 26554 | MT739465.1 | \|Severe acute respiratory syndrome coro | ···················· | ···················· | ···················· | ···················· | ···················· | ···················· |
| 26555 | MT739466.1 | \|Severe acute respiratory syndrome coro | ···················· | ···················· | ···················· | ···················· | ···················· | ···················· |
| 26556 | MT739467.1 | \|Severe acute respiratory syndrome coro | ···················· | ···················· | ···················· | ···················· | ···················· | ···················· |
| 26557 | MT739468.1 | \|Severe acute respiratory syndrome coro | ···················· | ···················· | ···················· | ···················· | ···················· | ···················· |
| 26558 | MT739469.1 | \|Severe acute respiratory syndrome coro | ···················· | ···················· | ···················· | ···················· | ···················· | ···················· |
| 26559 | MT739470.1 | \|Severe acute respiratory syndrome coro | ···················· | ···················· | ···················· | ···················· | ···················· | ···················· |
| 26560 | MT739471.1 | \|Severe acute respiratory syndrome coro | ···················· | ···················· | ···················· | ···················· | ···················· | ···················· |
| 26561 | MT739472.1 | \|Severe acute respiratory syndrome coro | ···················· | ···················· | ···················· | ···················· | ···················· | ···················· |
| 26562 | MT739473.1 | \|Severe acute respiratory syndrome coro | ···················· | ···················· | ···················· | ···················· | ···················· | ···················· |

```
26563 MT739474.1 |Severe acute respiratory syndrome coro   ····················  ····················  ·····················  ····················  ····················  ····················
26564 MT739475.1 |Severe acute respiratory syndrome coro   ····················  ····················  ·····················  ····················  ····················  ····················
26565 MT739476.1 |Severe acute respiratory syndrome coro   ····················  ····················  ·····················  ····················  ····················  ····················
26566 MT739477.1 |Severe acute respiratory syndrome coro   ····················  ····················  ·····················  ····················  ····················  ····················
26567 MT730002.1 |Severe acute respiratory syndrome coro   ····················  ····················  ·····················  ····················  ····················  ····················
26568 MT731285.1 |Severe acute respiratory syndrome coro   ····················  ····················  ·····················  ····················  ····················  ····················
26569 MT731292.1 |Severe acute respiratory syndrome coro   ····················  ····················  ·····················  ····················  ····················  ····················
26570 MT731327.1 |Severe acute respiratory syndrome coro   ····················  ····················  ·····················  ····················  ····················  ····················
26571 MT731346.1 |Severe acute respiratory syndrome coro   ····················  ····················  ·····················  ····················  ····················  ····················
26572 MT731468.1 |Severe acute respiratory syndrome coro   ····················  ····················  ·····················  ····················  ····················  ····················
26573 MT731653.1 |Severe acute respiratory syndrome coro   ····················  ····················  ·····················  ····················  ····················  ····················
26574 MT731654.1 |Severe acute respiratory syndrome coro   ····················  ····················  ·····················  ····················  ····················  ····················
26575 MT731655.1 |Severe acute respiratory syndrome coro   ····················  ····················  ·····················  ····················  ····················  ····················
26576 MT731656.1 |Severe acute respiratory syndrome coro   ····················  ·A··················  ·····················  ····················  ····················  ··A·················
26577 MT731657.1 |Severe acute respiratory syndrome coro   ····················  ····················  ·····················  ····················  ····················  ····················
26578 MT731658.1 |Severe acute respiratory syndrome coro   ····················  ····················  ·····················  ····················  ····················  ····················
26579 MT731659.1 |Severe acute respiratory syndrome coro   ····················  ····················  ·····················  ····················  ····················  ····················
26580 MT731660.1 |Severe acute respiratory syndrome coro   ····················  ····················  ·····················  ····················  ····················  ····················
26581 MT731661.1 |Severe acute respiratory syndrome coro   ····················  ····················  ·····················  ····················  ····················  ····················
26582 MT731662.1 |Severe acute respiratory syndrome coro   ····················  ·A··················  ·····················  ····················  ····················  ··A·················
26583 MT731663.1 |Severe acute respiratory syndrome coro   ····················  ····················  ·····················  ····················  ····················  ····················
26584 MT731664.1 |Severe acute respiratory syndrome coro   ····················  ····················  ·····················  ····················  ····················  ····················
26585 MT731665.1 |Severe acute respiratory syndrome coro   ····················  ····················  ·····················  ····················  ····················  ····················
26586 MT731666.1 |Severe acute respiratory syndrome coro   ····················  ·A··················  ·····················  ····················  ····················  ··A·················
26587 MT731667.1 |Severe acute respiratory syndrome coro   ····················  ····················  ·····················  ····················  ····················  ····················
26588 MT731668.1 |Severe acute respiratory syndrome coro   ····················  ····················  ·····················  ····················  ····················  ····················
26589 MT731669.1 |Severe acute respiratory syndrome coro   ····················  ·A··················  ·····················  ····················  ····················  ··A·················
26590 MT731673.1 |Severe acute respiratory syndrome coro   ····················  ····················  ·····················  ····················  ····················  ····················
26591 MT731733.1 |Severe acute respiratory syndrome coro   ····················  ····················  ·········T···········  ····················  ····················  ····················
26592 MT731734.1 |Severe acute respiratory syndrome coro   ····················  ····················  ·····················  ····················  ····················  ····················
26593 MT731735.1 |Severe acute respiratory syndrome coro   ····················  ····················  ·····················  ····················  ····················  ····················
26594 MT731736.1 |Severe acute respiratory syndrome coro   ····················  ····················  ·····················  ····················  ····················  ····················
26595 MT731737.1 |Severe acute respiratory syndrome coro   ····················  ····················  ·····················  ····················  ····················  ····················
26596 MT731738.1 |Severe acute respiratory syndrome coro   ····················  ·A··················  ·····················  ····················  ····················  ··A·················
26597 MT731739.1 |Severe acute respiratory syndrome coro   ····················  ····················  ·····················  ····················  ····················  ····················
26598 MT731740.1 |Severe acute respiratory syndrome coro   ····················  ····················  ·····················  ····················  ····················  ····················
26599 MT731741.1 |Severe acute respiratory syndrome coro   ····················  ····················  ·····················  ····················  ····················  ····················
26600 MT731742.1 |Severe acute respiratory syndrome coro   ····················  ····················  ·····················  ····················  ····················  ····················
26601 MT731743.1 |Severe acute respiratory syndrome coro   ····················  ····················  ·····················  ····················  ····················  ····················
26602 MT731744.1 |Severe acute respiratory syndrome coro   ····················  ····················  ·····················  ····················  ····················  ····················
26603 MT731746.1 |Severe acute respiratory syndrome coro   ····················  ····················  ·····················  ····················  ····················  ····················
26604 MT731764.1 |Severe acute respiratory syndrome coro   ····················  ····················  ·····················  ····················  ····················  ····················
26605 MT731931.1 |Severe acute respiratory syndrome coro   ····················  ····················  ·····················  ····················  ····················  ····················
26606 MT731932.1 |Severe acute respiratory syndrome coro   ····················  ····················  ·····················  ····················  ····················  ····················
26607 MT731933.1 |Severe acute respiratory syndrome coro   ····················  ····················  ·····················  ····················  ····················  ····················
26608 MT731934.1 |Severe acute respiratory syndrome coro   ····················  ····················  ·····················  ····················  ····················  ····················
26609 MT731935.1 |Severe acute respiratory syndrome coro   ····················  ····················  ·····················  ····················  ····················  ····················
26610 MT731936.1 |Severe acute respiratory syndrome coro   ····················  ····················  ·····················  ····················  ····················  ····················
26611 MT731937.1 |Severe acute respiratory syndrome coro   ····················  ····················  ·····················  ····················  ····················  ····················
26612 MT731938.1 |Severe acute respiratory syndrome coro   ····················  ····················  ·····················  ····················  ····················  ····················
26613 MT733120.1 |Severe acute respiratory syndrome coro   ····················  ····················  ·····················  ····················  ····················  ····················
26614 MT712859.1 |Severe acute respiratory syndrome coro   ····················  ····················  ·····················  ····················  ····················  ····················
26615 MT712861.1 |Severe acute respiratory syndrome coro   ····················  ····················  ·····················  ····················  ····················  ····················
26616 MT712863.1 |Severe acute respiratory syndrome coro   ····················  ····················  ·····················  ····················  ····················  ····················
26617 MT706050.1 |Severe acute respiratory syndrome coro   ····················  ····················  ·····················  ····················  ····················  ····················
26618 MT706132.1 |Severe acute respiratory syndrome coro   ····················  ····················  ·····················  ····················  ····················  ····················
26619 MT706133.1 |Severe acute respiratory syndrome coro   ····················  ····················  ·····················  ····················  ····················  ····················
26620 MT706134.1 |Severe acute respiratory syndrome coro   ····················  ····················  ·····················  ····················  ····················  ····················
26621 MT706135.1 |Severe acute respiratory syndrome coro   ····················  ····················  ·····················  ····················  ····················  ····················
26622 MT706136.1 |Severe acute respiratory syndrome coro   ····················  ····················  ·····················  ····················  ····················  ····················
26623 MT706137.1 |Severe acute respiratory syndrome coro   ····················  ····················  ·····················  ····················  ····················  ····················
26624 MT706138.1 |Severe acute respiratory syndrome coro   ····················  ····················  ·····················  ····················  ····················  ····················
26625 MT706140.1 |Severe acute respiratory syndrome coro   ····················  ····················  ·····················  ····················  ····················  ····················
26626 MT706141.1 |Severe acute respiratory syndrome coro   ····················  ····················  ·····················  ····················  ····················  ····················
26627 MT706142.1 |Severe acute respiratory syndrome coro   ····················  ····················  ·····················  ····················  ····················  ····················
26628 MT706143.1 |Severe acute respiratory syndrome coro   ····················  ····················  ·····················  ····················  ····················  ····················
26629 MT706144.1 |Severe acute respiratory syndrome coro   ····················  ····················  ·····················  ····················  ····················  ····················
26630 MT706145.1 |Severe acute respiratory syndrome coro   ····················  ····················  ·····················  ····················  ····················  ····················
26631 MT706146.1 |Severe acute respiratory syndrome coro   ····················  ····················  ·····················  ····················  ····················  ····················
26632 MT706148.1 |Severe acute respiratory syndrome coro   ····················  ····················  ·····················  ····················  ····················  ····················
26633 MT706149.1 |Severe acute respiratory syndrome coro   ····················  ····················  ·····················  ····················  ····················  ····················
26634 MT706150.1 |Severe acute respiratory syndrome coro   ····················  ····················  ·····················  ····················  ····················  ····················
26635 MT706151.1 |Severe acute respiratory syndrome coro   ····················  ····················  ·····················  ····················  ····················  ····················
26636 MT706152.1 |Severe acute respiratory syndrome coro   ····················  ····················  ·····················  ····················  ····················  ····················
26637 MT706153.1 |Severe acute respiratory syndrome coro   ····················  ····················  ·····················  ····················  ····················  ····················
26638 MT706154.1 |Severe acute respiratory syndrome coro   ····················  ····················  ·····················  ····················  ····················  ····················
26639 MT706155.1 |Severe acute respiratory syndrome coro   ····················  ····················  ·····················  ····················  ····················  ····················
26640 MT706156.1 |Severe acute respiratory syndrome coro   ····················  ····················  ·····················  ····················  ····················  ····················
```

26641 MT706157.1 |Severe acute respiratory syndrome coro .................... .................... .................... .................... .................... ....................
26642 MT706158.1 |Severe acute respiratory syndrome coro .................... .................... .................... .................... .................... ....................
26643 MT706159.1 |Severe acute respiratory syndrome coro .................... .................... .................... .................... .................... ....................
26644 MT706160.1 |Severe acute respiratory syndrome coro .................... .................... .................... .................... .................... ....................
26645 MT706161.1 |Severe acute respiratory syndrome coro .................... .................... .................... .................... .................... ....................
26646 MT706162.1 |Severe acute respiratory syndrome coro .................... .................... .................... .................... .................... ....................
26647 MT706163.1 |Severe acute respiratory syndrome coro .................... .................... .................... .................... .................... ....................
26648 MT706164.1 |Severe acute respiratory syndrome coro .................... .................... .................... .................... .................... ....................
26649 MT706165.1 |Severe acute respiratory syndrome coro .................... .................... .................... .................... .................... ....................
26650 MT706166.1 |Severe acute respiratory syndrome coro .................... .................... .................... .................... .................... ....................
26651 MT706167.1 |Severe acute respiratory syndrome coro .................... .................... .................... .................... .................... ....................
26652 MT706168.1 |Severe acute respiratory syndrome coro .................... .................... .................... .................... .................... ....................
26653 MT706169.1 |Severe acute respiratory syndrome coro .................... .................... .................... .................... .................... ....................
26654 MT706170.1 |Severe acute respiratory syndrome coro .................... .................... .................... .................... .................... ....................
26655 MT706171.1 |Severe acute respiratory syndrome coro .................... .................... .................... .................... .................... ....................
26656 MT706172.1 |Severe acute respiratory syndrome coro .................... .................... .................... .................... .................... ....................
26657 MT706173.1 |Severe acute respiratory syndrome coro .................... .................... .................... .................... .................... ....................
26658 MT706174.1 |Severe acute respiratory syndrome coro .................... .................... .................... .................... .................... ....................
26659 MT706175.1 |Severe acute respiratory syndrome coro .................... .................... .................... .................... .................... ....................
26660 MT706176.1 |Severe acute respiratory syndrome coro .................... .................... .................... .................... .................... ....................
26661 MT706177.1 |Severe acute respiratory syndrome coro .................... .................... .................... .................... .................... ....................
26662 MT706178.1 |Severe acute respiratory syndrome coro .................... .................... .................... .................... .................... ....................
26663 MT706179.1 |Severe acute respiratory syndrome coro .................... .................... .................... .................... .................... ....................
26664 MT706180.1 |Severe acute respiratory syndrome coro .................... .................... .................... .................... .................... ....................
26665 MT706181.1 |Severe acute respiratory syndrome coro .................... .................... .................... .................... .................... ....................
26666 MT706182.1 |Severe acute respiratory syndrome coro .................... .................... .................... .................... .................... ....................
26667 MT706183.1 |Severe acute respiratory syndrome coro .................... .................... .................... .................... .................... ....................
26668 MT706184.1 |Severe acute respiratory syndrome coro .................... .................... .................... .................... .................... ....................
26669 MT706185.1 |Severe acute respiratory syndrome coro .................... .................... .................... .................... .................... ....................
26670 MT706186.1 |Severe acute respiratory syndrome coro .................... .................... .................... .................... .................... ....................
26671 MT706187.1 |Severe acute respiratory syndrome coro .................... .................... .................... .................... .................... N...................
26672 MT706188.1 |Severe acute respiratory syndrome coro .................... .................... .................... .................... .................... ....................
26673 MT706189.1 |Severe acute respiratory syndrome coro .................... .................... .................... .................... .................... ....................
26674 MT706190.1 |Severe acute respiratory syndrome coro .................... .................... .................... .................... .................... ....................
26675 MT706191.1 |Severe acute respiratory syndrome coro .................... .................... .................... .................... .................... ....................
26676 MT706192.1 |Severe acute respiratory syndrome coro .................... .................... .................... .................... .................... ....................
26677 MT706193.1 |Severe acute respiratory syndrome coro .................... .................... .................... .................... .................... ....................
26678 MT706194.1 |Severe acute respiratory syndrome coro .................... .................... .................... .................... .................... ....................
26679 MT706195.1 |Severe acute respiratory syndrome coro .................... .................... .................... .................... .................... ....................
26680 MT706196.1 |Severe acute respiratory syndrome coro .................... .................... .................... .................... .................... ....................
26681 MT706197.1 |Severe acute respiratory syndrome coro .................... .................... .................... .................... .................... ....................
26682 MT706198.1 |Severe acute respiratory syndrome coro .................... .................... .................... .................... .................... ....................
26683 MT706199.1 |Severe acute respiratory syndrome coro .................... .................... .................... .................... .................... ....................
26684 MT706200.1 |Severe acute respiratory syndrome coro .................... .................... .................... .................... .................... ....................
26685 MT706201.1 |Severe acute respiratory syndrome coro .................... .................... .................... .................... .................... ....................
26686 MT706202.1 |Severe acute respiratory syndrome coro .................... .................... .................... .................... .................... ....................
26687 MT706203.1 |Severe acute respiratory syndrome coro .................... .................... .................... .................... .................... ....................
26688 MT706204.1 |Severe acute respiratory syndrome coro .................... .................... .................... .................... .................... ....................
26689 MT706205.1 |Severe acute respiratory syndrome coro .................... .................... .................... .................... .................... ....................
26690 MT706206.1 |Severe acute respiratory syndrome coro .................... .................... .................... .................... .................... ....................
26691 MT706207.1 |Severe acute respiratory syndrome coro .................... .................... .................... .................... .................... ....................
26692 MT706208.1 |Severe acute respiratory syndrome coro .................... .................... .................... .................... .................... ....................
26693 MT706210.1 |Severe acute respiratory syndrome coro .................... .................... .................... .................... .................... ....................
26694 MT706212.1 |Severe acute respiratory syndrome coro .................... .................... .................... .................... .................... ....................
26695 MT706213.1 |Severe acute respiratory syndrome coro .................... .................... .................... .................... .................... ....................
26696 MT706214.1 |Severe acute respiratory syndrome coro .................... .................... .................... .................... .................... ....................
26697 MT706215.1 |Severe acute respiratory syndrome coro .................... .................... .................... .................... .................... ....................
26698 MT706216.1 |Severe acute respiratory syndrome coro .................... .................... .................... .................... .................... ....................
26699 MT706218.1 |Severe acute respiratory syndrome coro .................... .................... .................... .................... .................... ....................
26700 MT706219.1 |Severe acute respiratory syndrome coro .................... .................... .................... .................... .................... ....................
26701 MT706220.1 |Severe acute respiratory syndrome coro .................... .................... .................... .................... .................... ....................
26702 MT706221.1 |Severe acute respiratory syndrome coro .................... .................... .................... .................... .................... ....................
26703 MT706222.1 |Severe acute respiratory syndrome coro .................... .................... .................... .................... .................... ....................
26704 MT706223.1 |Severe acute respiratory syndrome coro .................... .................... .................... .................... .................... ....................
26705 MT706224.1 |Severe acute respiratory syndrome coro .................... .................... .................... .................... .................... ....................
26706 MT706225.1 |Severe acute respiratory syndrome coro .................... .................... .................... .................... .................... ....................
26707 MT706226.1 |Severe acute respiratory syndrome coro .................... .................... .................... .................... .................... ....................
26708 MT706227.1 |Severe acute respiratory syndrome coro .................... .................... .................... .................... .................... ....................
26709 MT706228.1 |Severe acute respiratory syndrome coro .................... .................... .................... .................... .................... ....................
26710 MT706229.1 |Severe acute respiratory syndrome coro .................... .................... .................... .................... .................... ....................
26711 MT706230.1 |Severe acute respiratory syndrome coro .................... .................... .................... .................... .................... ....................
26712 MT706231.1 |Severe acute respiratory syndrome coro .................... .................... .................... .................... .................... ....................
26713 MT706232.1 |Severe acute respiratory syndrome coro .................... .................... .................... .................... .................... ....................
26714 MT706233.1 |Severe acute respiratory syndrome coro .................... .................... .................... .................... .................... ....................
26715 MT706234.1 |Severe acute respiratory syndrome coro .................... .................... .................... .................... .................... ....................
26716 MT706235.1 |Severe acute respiratory syndrome coro .................... .................... .................... .................... .................... ....................
26717 MT706236.1 |Severe acute respiratory syndrome coro .................... .................... .................... .................... .................... ....................
26718 MT706237.1 |Severe acute respiratory syndrome coro .................... .................... .................... .................... .................... ....................

| | | | | | | | | |
|---|---|---|---|---|---|---|---|---|
| 26719 | MT706238.1 | \|Severe acute respiratory syndrome coro | .................... | .................... | .................... | .................... | .................... | .................... |
| 26720 | MT706239.1 | \|Severe acute respiratory syndrome coro | .................... | .................... | .................... | .................... | .................... | .................... |
| 26721 | MT706240.1 | \|Severe acute respiratory syndrome coro | .................... | .................... | .................... | .................... | .................... | .................... |
| 26722 | MT706241.1 | \|Severe acute respiratory syndrome coro | .................... | .................... | .................... | .................... | .................... | .................... |
| 26723 | MT706243.1 | \|Severe acute respiratory syndrome coro | .................... | .................... | .................... | .................... | .................... | .................... |
| 26724 | MT706244.1 | \|Severe acute respiratory syndrome coro | .................... | .................... | .................... | .................... | .................... | .................... |
| 26725 | MT706245.1 | \|Severe acute respiratory syndrome coro | .................... | .................... | .................... | .................... | .................... | .................... |
| 26726 | MT706246.1 | \|Severe acute respiratory syndrome coro | .................... | .................... | .................... | .................... | .................... | .................... |
| 26727 | MT706247.1 | \|Severe acute respiratory syndrome coro | .................... | .................... | .................... | .................... | .................... | .................... |
| 26728 | MT706248.1 | \|Severe acute respiratory syndrome coro | .................... | .................... | .................... | .................... | .................... | .................... |
| 26729 | MT706249.1 | \|Severe acute respiratory syndrome coro | .................... | .................... | .................... | .................... | .................... | .................... |
| 26730 | MT706250.1 | \|Severe acute respiratory syndrome coro | .................... | .................... | .................... | .................... | .................... | .................... |
| 26731 | MT706251.1 | \|Severe acute respiratory syndrome coro | .................... | .................... | .................... | .................... | .................... | .................... |
| 26732 | MT706252.1 | \|Severe acute respiratory syndrome coro | .................... | .................... | .................... | .................... | .................... | .................... |
| 26733 | MT706253.1 | \|Severe acute respiratory syndrome coro | .................... | .................... | .................... | .................... | .................... | .................... |
| 26734 | MT706254.1 | \|Severe acute respiratory syndrome coro | .................... | .................... | .................... | .................... | .................... | .................... |
| 26735 | MT706255.1 | \|Severe acute respiratory syndrome coro | .................... | .................... | .................... | .................... | .................... | .................... |
| 26736 | MT706256.1 | \|Severe acute respiratory syndrome coro | .................... | .................... | .................... | .................... | .................... | .................... |
| 26737 | MT706258.1 | \|Severe acute respiratory syndrome coro | .................... | .................... | .................... | .................... | .................... | .................... |
| 26738 | MT706259.1 | \|Severe acute respiratory syndrome coro | .................... | .................... | .................... | .................... | .................... | .................... |
| 26739 | MT706260.1 | \|Severe acute respiratory syndrome coro | .................... | .................... | .................... | .................... | .................... | .................... |
| 26740 | MT706262.1 | \|Severe acute respiratory syndrome coro | .................... | .................... | .................... | .................... | .................... | .................... |
| 26741 | MT706263.1 | \|Severe acute respiratory syndrome coro | .................... | .................... | .................... | .................... | .................... | .................... |
| 26742 | MT706264.1 | \|Severe acute respiratory syndrome coro | .................... | .................... | .................... | .................... | .................... | .................... |
| 26743 | MT706265.1 | \|Severe acute respiratory syndrome coro | .................... | .................... | .................... | .................... | .................... | .................... |
| 26744 | MT706266.1 | \|Severe acute respiratory syndrome coro | .................... | .................... | .................... | .................... | .................... | .................... |
| 26745 | MT706268.1 | \|Severe acute respiratory syndrome coro | .................... | .................... | .................... | .................... | .................... | .................... |
| 26746 | MT706269.1 | \|Severe acute respiratory syndrome coro | .................... | .................... | .................... | .................... | .................... | .................... |
| 26747 | MT706270.1 | \|Severe acute respiratory syndrome coro | .................... | .................... | .................... | .................... | .................... | .................... |
| 26748 | MT706271.1 | \|Severe acute respiratory syndrome coro | .................... | .................... | .................... | .................... | .................... | .................... |
| 26749 | MT706272.1 | \|Severe acute respiratory syndrome coro | .................... | .................... | .................... | .................... | .................... | .................... |
| 26750 | MT706273.1 | \|Severe acute respiratory syndrome coro | .................... | .................... | .................... | .................... | .................... | .................... |
| 26751 | MT706275.1 | \|Severe acute respiratory syndrome coro | .................... | .................... | .................... | .................... | .................... | .................... |
| 26752 | MT706276.1 | \|Severe acute respiratory syndrome coro | .................... | .................... | .................... | .................... | .................... | .................... |
| 26753 | MT706277.1 | \|Severe acute respiratory syndrome coro | .................... | .................... | .................... | .................... | .................... | .................... |
| 26754 | MT706279.1 | \|Severe acute respiratory syndrome coro | .................... | .................... | .................... | .................... | .................... | .................... |
| 26755 | MT706280.1 | \|Severe acute respiratory syndrome coro | .................... | .................... | .................... | .................... | .................... | .................... |
| 26756 | MT706281.1 | \|Severe acute respiratory syndrome coro | .................... | .................... | .................... | .................... | .................... | .................... |
| 26757 | MT706282.1 | \|Severe acute respiratory syndrome coro | .................... | .................... | .................... | .................... | .................... | .................... |
| 26758 | MT706284.1 | \|Severe acute respiratory syndrome coro | .................... | .................... | .................... | .................... | .................... | .................... |
| 26759 | MT706285.1 | \|Severe acute respiratory syndrome coro | .................... | .................... | .................... | .................... | .................... | .................... |
| 26760 | MT706286.1 | \|Severe acute respiratory syndrome coro | .................... | .................... | .................... | .................... | .................... | .................... |
| 26761 | MT706287.1 | \|Severe acute respiratory syndrome coro | .................... | .................... | .................... | .................... | .................... | .................... |
| 26762 | MT706288.1 | \|Severe acute respiratory syndrome coro | .................... | .................... | .................... | .................... | .................... | .................... |
| 26763 | MT706289.1 | \|Severe acute respiratory syndrome coro | .................... | .................... | .................... | .................... | .................... | .................... |
| 26764 | MT706290.1 | \|Severe acute respiratory syndrome coro | .................... | .................... | .................... | .................... | .................... | .................... |
| 26765 | MT706291.1 | \|Severe acute respiratory syndrome coro | .................... | .................... | .................... | .................... | .................... | .................... |
| 26766 | MT706292.1 | \|Severe acute respiratory syndrome coro | .................... | .................... | .................... | .................... | .................... | .................... |
| 26767 | MT706293.1 | \|Severe acute respiratory syndrome coro | .................... | .................... | .................... | .................... | .................... | .................... |
| 26768 | MT706294.1 | \|Severe acute respiratory syndrome coro | .................... | .................... | .................... | .................... | .................... | .................... |
| 26769 | MT706295.1 | \|Severe acute respiratory syndrome coro | .................... | .................... | .................... | .................... | .................... | .................... |
| 26770 | MT706296.1 | \|Severe acute respiratory syndrome coro | .................... | .................... | .................... | .................... | .................... | .................... |
| 26771 | MT706297.1 | \|Severe acute respiratory syndrome coro | .................... | .................... | .................... | .................... | .................... | .................... |
| 26772 | MT706298.1 | \|Severe acute respiratory syndrome coro | .................... | .................... | .................... | .................... | .................... | .................... |
| 26773 | MT706299.1 | \|Severe acute respiratory syndrome coro | .................... | .................... | .................... | .................... | .................... | .................... |
| 26774 | MT706300.1 | \|Severe acute respiratory syndrome coro | .................... | .................... | .................... | .................... | .................... | .................... |
| 26775 | MT706301.1 | \|Severe acute respiratory syndrome coro | .................... | .................... | .................... | .................... | .................... | .................... |
| 26776 | MT706302.1 | \|Severe acute respiratory syndrome coro | .................... | .................... | .................... | .................... | .................... | .................... |
| 26777 | MT706303.1 | \|Severe acute respiratory syndrome coro | .................... | .................... | .................... | .................... | .................... | .................... |
| 26778 | MT706304.1 | \|Severe acute respiratory syndrome coro | .................... | .................... | .................... | .................... | .................... | .................... |
| 26779 | MT706305.1 | \|Severe acute respiratory syndrome coro | .................... | .................... | .................... | .................... | .................... | .................... |
| 26780 | MT706306.1 | \|Severe acute respiratory syndrome coro | .................... | .................... | .................... | .................... | .................... | .................... |
| 26781 | MT706307.1 | \|Severe acute respiratory syndrome coro | .................... | .................... | .................... | .................... | .................... | .................... |
| 26782 | MT706308.1 | \|Severe acute respiratory syndrome coro | .................... | .................... | .................... | .................... | .................... | .................... |
| 26783 | MT706309.1 | \|Severe acute respiratory syndrome coro | .................... | .................... | .................... | .................... | .................... | .................... |
| 26784 | MT706310.1 | \|Severe acute respiratory syndrome coro | .................... | .................... | .................... | .................... | .................... | .................... |
| 26785 | MT706311.1 | \|Severe acute respiratory syndrome coro | .................... | .................... | .................... | .................... | .................... | .................... |
| 26786 | MT706312.1 | \|Severe acute respiratory syndrome coro | .................... | .................... | .................... | .................... | .................... | .................... |
| 26787 | MT706313.1 | \|Severe acute respiratory syndrome coro | .................... | .................... | .................... | .................... | .................... | .................... |
| 26788 | MT706314.1 | \|Severe acute respiratory syndrome coro | .................... | .................... | .................... | .................... | .................... | .................... |
| 26789 | MT706315.1 | \|Severe acute respiratory syndrome coro | .................... | .................... | .................... | .................... | .................... | .................... |
| 26790 | MT706317.1 | \|Severe acute respiratory syndrome coro | .................... | .................... | .................... | .................... | .................... | .................... |
| 26791 | MT706318.1 | \|Severe acute respiratory syndrome coro | .................... | .........A.......... | .................... | .................... | .................... | .................... |
| 26792 | MT706319.1 | \|Severe acute respiratory syndrome coro | .................... | .................... | .................... | .................... | .................... | .................... |
| 26793 | MT706320.1 | \|Severe acute respiratory syndrome coro | .................... | .................... | .................... | .................... | .................... | .................... |
| 26794 | MT706321.1 | \|Severe acute respiratory syndrome coro | .................... | .................... | .................... | .................... | .................... | .................... |
| 26795 | MT706322.1 | \|Severe acute respiratory syndrome coro | .................... | .................... | .................... | .................... | .................... | .................... |
| 26796 | MT706323.1 | \|Severe acute respiratory syndrome coro | .................... | .................... | .................... | .................... | .................... | .................... |

| | | | | | | | | |
|---|---|---|---|---|---|---|---|---|
| 26797 | MT706324.1 | Severe acute respiratory syndrome coro | ···················· | ···················· | ····················· | ···················· | ···················· | ···················· |
| 26798 | MT706325.1 | Severe acute respiratory syndrome coro | ···················· | ···················· | ····················· | ···················· | ···················· | ···················· |
| 26799 | MT706326.1 | Severe acute respiratory syndrome coro | ···················· | ···················· | ····················· | ···················· | ···················· | ···················· |
| 26800 | MT706327.1 | Severe acute respiratory syndrome coro | ···················· | ···················· | ····················· | ···················· | ···················· | ···················· |
| 26801 | MT706328.1 | Severe acute respiratory syndrome coro | ···················· | ···················· | ····················· | ···················· | ···················· | ···················· |
| 26802 | MT706329.1 | Severe acute respiratory syndrome coro | ···················· | ···················· | ····················· | ···················· | ···················· | ···················· |
| 26803 | MT706330.1 | Severe acute respiratory syndrome coro | ···················· | ···················· | ····················· | ···················· | ···················· | ···················· |
| 26804 | MT706331.1 | Severe acute respiratory syndrome coro | ···················· | ···················· | ····················· | ···················· | ···················· | ···················· |
| 26805 | MT706332.1 | Severe acute respiratory syndrome coro | ···················· | ···················· | ····················· | ···················· | ···················· | ···················· |
| 26806 | MT706333.1 | Severe acute respiratory syndrome coro | ···················· | ···················· | ····················· | ···················· | ···················· | ···················· |
| 26807 | MT706334.1 | Severe acute respiratory syndrome coro | ···················· | ···················· | ····················· | ···················· | ···················· | ···················· |
| 26808 | MT706335.1 | Severe acute respiratory syndrome coro | ···················· | ···················· | ····················· | ···················· | ···················· | ···················· |
| 26809 | MT706336.1 | Severe acute respiratory syndrome coro | ···················· | ···················· | ····················· | ···················· | ···················· | ···················· |
| 26810 | MT706337.1 | Severe acute respiratory syndrome coro | ···················· | ···················· | ····················· | ···················· | ···················· | ···················· |
| 26811 | MT706338.1 | Severe acute respiratory syndrome coro | ···················· | ···················· | ····················· | ···················· | ···················· | ···················· |
| 26812 | MT706339.1 | Severe acute respiratory syndrome coro | ···················· | ···················· | ····················· | ···················· | ···················· | ···················· |
| 26813 | MT706340.1 | Severe acute respiratory syndrome coro | ···················· | ···················· | ····················· | ···················· | ···················· | ···················· |
| 26814 | MT706341.1 | Severe acute respiratory syndrome coro | ···················· | ···················· | ····················· | ···················· | ···················· | ···················· |
| 26815 | MT706342.1 | Severe acute respiratory syndrome coro | ···················· | ···················· | ····················· | ···················· | ···················· | ···················· |
| 26816 | MT706343.1 | Severe acute respiratory syndrome coro | ···················· | ···················· | ····················· | ···················· | ···················· | ···················· |
| 26817 | MT706344.1 | Severe acute respiratory syndrome coro | ···················· | ···················· | ····················· | ···················· | ···················· | ···················· |
| 26818 | MT706345.1 | Severe acute respiratory syndrome coro | ···················· | ···················· | ····················· | ···················· | ···················· | ···················· |
| 26819 | MT706346.1 | Severe acute respiratory syndrome coro | ···················· | ···················· | ····················· | ···················· | ···················· | ···················· |
| 26820 | MT706347.1 | Severe acute respiratory syndrome coro | ···················· | ···················· | ····················· | ···················· | ···················· | ···················· |
| 26821 | MT706348.1 | Severe acute respiratory syndrome coro | ···················· | ···················· | ····················· | ···················· | ···················· | ···················· |
| 26822 | MT706349.1 | Severe acute respiratory syndrome coro | ···················· | ···················· | ····················· | ···················· | ···················· | ···················· |
| 26823 | MT706350.1 | Severe acute respiratory syndrome coro | ···················· | ···················· | ····················· | ···················· | ···················· | ···················· |
| 26824 | MT706351.1 | Severe acute respiratory syndrome coro | ···················· | ···················· | ····················· | ···················· | ···················· | ···················· |
| 26825 | MT706352.1 | Severe acute respiratory syndrome coro | ···················· | ···················· | ····················· | ···················· | ···················· | ···················· |
| 26826 | MT706353.1 | Severe acute respiratory syndrome coro | ···················· | ···················· | ····················· | ···················· | ···················· | ···················· |
| 26827 | MT706354.1 | Severe acute respiratory syndrome coro | ···················· | ···················· | ····················· | ···················· | ···················· | ···················· |
| 26828 | MT706355.1 | Severe acute respiratory syndrome coro | ···················· | ···················· | ····················· | ···················· | ···················· | ···················· |
| 26829 | MT706356.1 | Severe acute respiratory syndrome coro | ···················· | ···················· | ····················· | ···················· | ···················· | ···················· |
| 26830 | MT706357.1 | Severe acute respiratory syndrome coro | ···················· | ···················· | ····················· | ···················· | ···················· | ···················· |
| 26831 | MT706358.1 | Severe acute respiratory syndrome coro | ···················· | ···················· | ····················· | ···················· | ···················· | ···················· |
| 26832 | MT706359.1 | Severe acute respiratory syndrome coro | ···················· | ···················· | ····················· | ···················· | ···················· | ···················· |
| 26833 | MT706360.1 | Severe acute respiratory syndrome coro | ···················· | ···················· | ····················· | ···················· | ···················· | ···················· |
| 26834 | MT706361.1 | Severe acute respiratory syndrome coro | ···················· | ···················· | ····················· | ···················· | ···················· | ···················· |
| 26835 | MT706362.1 | Severe acute respiratory syndrome coro | ···················· | ···················· | ····················· | ···················· | ···················· | ···················· |
| 26836 | MT706363.1 | Severe acute respiratory syndrome coro | ···················· | ···················· | ····················· | ···················· | ···················· | ···················· |
| 26837 | MT706364.1 | Severe acute respiratory syndrome coro | ···················· | ···················· | ····················· | ···················· | ···················· | ···················· |
| 26838 | MT706365.1 | Severe acute respiratory syndrome coro | ···················· | ···················· | ····················· | ···················· | ···················· | ···················· |
| 26839 | MT706366.1 | Severe acute respiratory syndrome coro | ···················· | ···················· | ····················· | ···················· | ···················· | ···················· |
| 26840 | MT706367.1 | Severe acute respiratory syndrome coro | ···················· | ···················· | ····················· | ···················· | ···················· | ···················· |
| 26841 | MT706368.1 | Severe acute respiratory syndrome coro | ···················· | ···················· | ····················· | ···················· | ···················· | ···················· |
| 26842 | MT706369.1 | Severe acute respiratory syndrome coro | ···················· | ···················· | ····················· | ···················· | ···················· | ···················· |
| 26843 | MT706370.1 | Severe acute respiratory syndrome coro | ···················· | ···················· | ····················· | ···················· | ···················· | ···················· |
| 26844 | MT706371.1 | Severe acute respiratory syndrome coro | ···················· | ···················· | ····················· | ···················· | ···················· | ···················· |
| 26845 | MT706372.1 | Severe acute respiratory syndrome coro | ···················· | ···················· | ····················· | ···················· | ···················· | ···················· |
| 26846 | MT706373.1 | Severe acute respiratory syndrome coro | ···················· | ···················· | ····················· | ···················· | ···················· | ···················· |
| 26847 | MT706374.1 | Severe acute respiratory syndrome coro | ···················· | ···················· | ····················· | ···················· | ···················· | ···················· |
| 26848 | MT706375.1 | Severe acute respiratory syndrome coro | ···················· | ···················· | ····················· | ···················· | ···················· | ···················· |
| 26849 | MT706376.1 | Severe acute respiratory syndrome coro | ···················· | ···················· | ····················· | ···················· | ···················· | ···················· |
| 26850 | MT706379.1 | Severe acute respiratory syndrome coro | ···················· | ···················· | ····················· | ···················· | ···················· | ···················· |
| 26851 | MT706380.1 | Severe acute respiratory syndrome coro | ···················· | ···················· | ····················· | ···················· | ···················· | ···················· |
| 26852 | MT706381.1 | Severe acute respiratory syndrome coro | ···················· | ···················· | ····················· | ···················· | ···················· | ···················· |
| 26853 | MT706382.1 | Severe acute respiratory syndrome coro | ···················· | ···················· | ····················· | ···················· | ···················· | ···················· |
| 26854 | MT706383.1 | Severe acute respiratory syndrome coro | ···················· | ···················· | ····················· | ···················· | ···················· | ···················· |
| 26855 | MT706384.1 | Severe acute respiratory syndrome coro | ···················· | ···················· | ····················· | ···················· | ···················· | ···················· |
| 26856 | MT706385.1 | Severe acute respiratory syndrome coro | ···················· | ···················· | ····················· | ···················· | ···················· | ···················· |
| 26857 | MT706386.1 | Severe acute respiratory syndrome coro | ···················· | ···················· | ····················· | ···················· | ···················· | ···················· |
| 26858 | MT706387.1 | Severe acute respiratory syndrome coro | ···················· | ···················· | ····················· | ···················· | ···················· | ···················· |
| 26859 | MT706389.1 | Severe acute respiratory syndrome coro | ···················· | ···················· | ····················· | ···················· | ···················· | ···················· |
| 26860 | MT706390.1 | Severe acute respiratory syndrome coro | ···················· | ···················· | ····················· | ···················· | ···················· | ···················· |
| 26861 | MT706391.1 | Severe acute respiratory syndrome coro | ···················· | ···················· | ····················· | ···················· | ···················· | ···················· |
| 26862 | MT706392.1 | Severe acute respiratory syndrome coro | ···················· | ···················· | ····················· | ···················· | ···················· | ···················· |
| 26863 | MT706393.1 | Severe acute respiratory syndrome coro | ···················· | ···················· | ····················· | ···················· | ···················· | ···················· |
| 26864 | MT706394.1 | Severe acute respiratory syndrome coro | ···················· | ···················· | ····················· | ···················· | ···················· | ···················· |
| 26865 | MT706395.1 | Severe acute respiratory syndrome coro | ···················· | ···················· | ····················· | ···················· | ···················· | ···················· |
| 26866 | MT706396.1 | Severe acute respiratory syndrome coro | ···················· | ···················· | ····················· | ···················· | ···················· | ···················· |
| 26867 | MT706397.1 | Severe acute respiratory syndrome coro | ···················· | ···················· | ····················· | ···················· | ···················· | ···················· |
| 26868 | MT706398.1 | Severe acute respiratory syndrome coro | ···················· | ······A············· | ····················· | ···················· | ···················· | ···················· |
| 26869 | MT706399.1 | Severe acute respiratory syndrome coro | ···················· | ···················· | ····················· | ···················· | ···················· | ···················· |
| 26870 | MT706400.1 | Severe acute respiratory syndrome coro | ···················· | ···················· | ····················· | ···················· | ···················· | ···················· |
| 26871 | MT706401.1 | Severe acute respiratory syndrome coro | ···················· | ···················· | ····················· | ···················· | ···················· | ···················· |
| 26872 | MT706402.1 | Severe acute respiratory syndrome coro | ···················· | ···················· | ····················· | ···················· | ···················· | ···················· |
| 26873 | MT706403.1 | Severe acute respiratory syndrome coro | ···················· | ···················· | ····················· | ···················· | ···················· | ···················· |
| 26874 | MT706404.1 | Severe acute respiratory syndrome coro | ···················· | ···················· | ····················· | ···················· | ···················· | ···················· |

| | | | | | | | | | |
|---|---|---|---|---|---|---|---|---|---|
| 26875 | MT706405.1 | \|Severe acute respiratory syndrome coro | .................... | ......A............. | .................... | .................... | .................... | .................... | |
| 26876 | MT706406.1 | \|Severe acute respiratory syndrome coro | .................... | .................... | .................... | .................... | .................... | .................... | |
| 26877 | MT706407.1 | \|Severe acute respiratory syndrome coro | .................... | .................... | .................... | .................... | .................... | .................... | |
| 26878 | MT706408.1 | \|Severe acute respiratory syndrome coro | .................... | ......A............. | .................... | .................... | .................... | .................... | |
| 26879 | MT706409.1 | \|Severe acute respiratory syndrome coro | .................... | ...................A | .................... | .................... | .................... | .................... | 1 |
| 26880 | MT706410.1 | \|Severe acute respiratory syndrome coro | .................... | .................... | .................... | .................... | .................... | .................... | |
| 26881 | MT706411.1 | \|Severe acute respiratory syndrome coro | .................... | ......A............. | .................... | .................... | .................... | .................... | |
| 26882 | MT706412.1 | \|Severe acute respiratory syndrome coro | .................... | .................... | .................... | .................... | .................... | .................... | |
| 26883 | MT706413.1 | \|Severe acute respiratory syndrome coro | .................... | .................... | .................... | .................... | .................... | .................... | |
| 26884 | MT706414.1 | \|Severe acute respiratory syndrome coro | .................... | .................... | .................... | .................... | .................... | .................... | |
| 26885 | MT706415.1 | \|Severe acute respiratory syndrome coro | .................... | .................... | .................... | .................... | .................... | .................... | |
| 26886 | MT706416.1 | \|Severe acute respiratory syndrome coro | .................... | .................... | .................... | .................... | .................... | .................... | |
| 26887 | MT706417.1 | \|Severe acute respiratory syndrome coro | .................... | .................... | .................... | .................... | .................... | .................... | |
| 26888 | MT706418.1 | \|Severe acute respiratory syndrome coro | .................... | .................... | .................... | .................... | .................... | .................... | |
| 26889 | MT706419.1 | \|Severe acute respiratory syndrome coro | .................... | .................... | .................... | .................... | .................... | .................... | |
| 26890 | MT706420.1 | \|Severe acute respiratory syndrome coro | .................... | .................... | .................... | .................... | .................... | .................... | |
| 26891 | MT706421.1 | \|Severe acute respiratory syndrome coro | .................... | .................... | .................... | .................... | .................... | .................... | |
| 26892 | MT706422.1 | \|Severe acute respiratory syndrome coro | .................... | .................... | .................... | .................... | .................... | .................... | |
| 26893 | MT706423.1 | \|Severe acute respiratory syndrome coro | .................... | .................... | .................... | .................... | .................... | .................... | |
| 26894 | MT706424.1 | \|Severe acute respiratory syndrome coro | .................... | .................... | .................... | .................... | .................... | .................... | |
| 26895 | MT706425.1 | \|Severe acute respiratory syndrome coro | .................... | .................... | .................... | .................... | .................... | .................... | |
| 26896 | MT706426.1 | \|Severe acute respiratory syndrome coro | .................... | .................... | .................... | .................... | .................... | .................... | |
| 26897 | MT706427.1 | \|Severe acute respiratory syndrome coro | .................... | .................... | .................... | .................... | .................... | .................... | |
| 26898 | MT706428.1 | \|Severe acute respiratory syndrome coro | .................... | .................... | .................... | .................... | .................... | .................... | |
| 26899 | MT706429.1 | \|Severe acute respiratory syndrome coro | .................... | .................... | .................... | .................... | .................... | .................... | |
| 26900 | MT706430.1 | \|Severe acute respiratory syndrome coro | .................... | .................... | .................... | .................... | .................... | .................... | |
| 26901 | MT706431.1 | \|Severe acute respiratory syndrome coro | .................... | .................... | .................... | .................... | .................... | .................... | |
| 26902 | MT706432.1 | \|Severe acute respiratory syndrome coro | .................... | .A.................. | .................... | .................... | .................... | .................... | |
| 26903 | MT706433.1 | \|Severe acute respiratory syndrome coro | .................... | .................... | .................... | .................... | .................... | .................... | |
| 26904 | MT706434.1 | \|Severe acute respiratory syndrome coro | .................... | .................... | .................... | .................... | .................... | .................... | |
| 26905 | MT706435.1 | \|Severe acute respiratory syndrome coro | .................... | .................... | .................... | .................... | .................... | .................... | |
| 26906 | MT706436.1 | \|Severe acute respiratory syndrome coro | .................... | .................... | .................... | .................... | .................... | .................... | |
| 26907 | MT706437.1 | \|Severe acute respiratory syndrome coro | .................... | .................... | .................... | .................... | .................... | .................... | |
| 26908 | MT706438.1 | \|Severe acute respiratory syndrome coro | .................... | .................... | .................... | .................... | .................... | .................... | |
| 26909 | MT706439.1 | \|Severe acute respiratory syndrome coro | .................... | .................... | .................... | .................... | .................... | .................... | |
| 26910 | MT706440.1 | \|Severe acute respiratory syndrome coro | .................... | .................... | .................... | .................... | .................... | .................... | |
| 26911 | MT706441.1 | \|Severe acute respiratory syndrome coro | .................... | .................... | .................... | .................... | .................... | .................... | |
| 26912 | MT706442.1 | \|Severe acute respiratory syndrome coro | .................... | .................... | .................... | .................... | .................... | .................... | |
| 26913 | MT706443.1 | \|Severe acute respiratory syndrome coro | .................... | .................... | .................... | .................... | .................... | .................... | |
| 26914 | MT706444.1 | \|Severe acute respiratory syndrome coro | .................... | .................... | .................... | .................... | .................... | .................... | |
| 26915 | MT706446.1 | \|Severe acute respiratory syndrome coro | .................... | .................... | .................... | .................... | .................... | .................... | |
| 26916 | MT706447.1 | \|Severe acute respiratory syndrome coro | .................... | .................... | .................... | .................... | .................... | .................... | |
| 26917 | MT706448.1 | \|Severe acute respiratory syndrome coro | .................... | .................... | .................... | .................... | .................... | .................... | |
| 26918 | MT706449.1 | \|Severe acute respiratory syndrome coro | .................... | .................... | .................... | .................... | .................... | .................... | |
| 26919 | MT706450.1 | \|Severe acute respiratory syndrome coro | .................... | .................... | .................... | .................... | .................... | .................... | |
| 26920 | MT706451.1 | \|Severe acute respiratory syndrome coro | .................... | .................... | .................... | .................... | .................... | .................... | |
| 26921 | MT706453.1 | \|Severe acute respiratory syndrome coro | .................... | .................... | .................... | .................... | .................... | .................... | |
| 26922 | MT709104.1 | \|Severe acute respiratory syndrome coro | .................... | .................... | .................... | .................... | .................... | .................... | |
| 26923 | MT709105.1 | \|Severe acute respiratory syndrome coro | .................... | .................... | .................... | .................... | .................... | .................... | |
| 26924 | MT710714.1 | \|Severe acute respiratory syndrome coro | .................... | .................... | .................... | .................... | .................... | .................... | |
| 26925 | MT711872.1 | \|Severe acute respiratory syndrome coro | .................... | .................... | .................... | .................... | .................... | .................... | |
| 26926 | MT711873.1 | \|Severe acute respiratory syndrome coro | .................... | .................... | .................... | .................... | .................... | .................... | |
| 26927 | MT711880.1 | \|Severe acute respiratory syndrome coro | .................... | .................... | .................... | .................... | .................... | .................... | |
| 26928 | MT705205.1 | \|Severe acute respiratory syndrome coro | .................... | .................... | .................... | .................... | .................... | .................... | |
| 26929 | MT705206.1 | \|Severe acute respiratory syndrome coro | .................... | .................... | .................... | .................... | .................... | .................... | |
| 26930 | MT704812.1 | \|Severe acute respiratory syndrome coro | .................... | .................... | .................... | .................... | .................... | .................... | |
| 26931 | MT703677.1 | \|Severe acute respiratory syndrome coro | .................... | .................... | .................... | .................... | .................... | .................... | |
| 26932 | MT703883.1 | \|Severe acute respiratory syndrome coro | .................... | .................... | .................... | .................... | .................... | .................... | |
| 26933 | MT703884.1 | \|Severe acute respiratory syndrome coro | .................... | .................... | .................... | .................... | .................... | .................... | |
| 26934 | MT703958.1 | \|Severe acute respiratory syndrome coro | .................... | .................... | .................... | .................... | .................... | .................... | |
| 26935 | MT703963.1 | \|Severe acute respiratory syndrome coro | .................... | .................... | .................... | .................... | .................... | .................... | |
| 26937 | MT703973.1 | \|Severe acute respiratory syndrome coro | .................... | .................... | .................... | .................... | .................... | .................... | |
| 26938 | MT703975.1 | \|Severe acute respiratory syndrome coro | .................... | .................... | .................... | .................... | .................... | .................... | |
| 26940 | MT703977.1 | \|Severe acute respiratory syndrome coro | .................... | .................... | .................... | .................... | .................... | .................... | |
| 26941 | MT703978.1 | \|Severe acute respiratory syndrome coro | .................... | .................... | .................... | .................... | .................... | .................... | |
| 26942 | MT703979.1 | \|Severe acute respiratory syndrome coro | .................... | .................... | .................... | .................... | .................... | .................... | |
| 26943 | MT703982.1 | \|Severe acute respiratory syndrome coro | .................... | .................... | .................... | .................... | .................... | .................... | |
| 26944 | MT703983.1 | \|Severe acute respiratory syndrome coro | .................... | .................... | .................... | .................... | .................... | .................... | |
| 26945 | MT703984.1 | \|Severe acute respiratory syndrome coro | .................... | .................... | .................... | .................... | .................... | .................... | |
| 26946 | MT703985.1 | \|Severe acute respiratory syndrome coro | .................... | .................... | .................... | .................... | .................... | .................... | |
| 26947 | MT703989.1 | \|Severe acute respiratory syndrome coro | .................... | .................... | .................... | .................... | .................... | .................... | |
| 26948 | MT703990.1 | \|Severe acute respiratory syndrome coro | .................... | .................... | .................... | .................... | .................... | .................... | |
| 26949 | MT703991.1 | \|Severe acute respiratory syndrome coro | .................... | .................... | .................... | .................... | .................... | .................... | |
| 26950 | MT703992.1 | \|Severe acute respiratory syndrome coro | .................... | .................... | .................... | .................... | .................... | .................... | |
| 26951 | MT703993.1 | \|Severe acute respiratory syndrome coro | .................... | .................... | .................... | .................... | .................... | .................... | |
| 26952 | MT703994.1 | \|Severe acute respiratory syndrome coro | .................... | .................... | .................... | .................... | .................... | .................... | |
| 26953 | MT703995.1 | \|Severe acute respiratory syndrome coro | .................... | .................... | .................... | .................... | .................... | .................... | |
| 26954 | MT703996.1 | \|Severe acute respiratory syndrome coro | .................... | .................... | .................... | .................... | .................... | .................... | |

26955 MT703997.1 |Severe acute respiratory syndrome coro ···················· ···················· ···················· ···················· ···················· ····················
26956 MT703998.1 |Severe acute respiratory syndrome coro ···················· ···················· ···················· ···················· ···················· ····················
26957 MT703999.1 |Severe acute respiratory syndrome coro ···················· ···················· ···················· ···················· ···················· ····················
26958 MT704000.1 |Severe acute respiratory syndrome coro ···················· ···················· ···················· ···················· ···················· ····················
26959 MT704001.1 |Severe acute respiratory syndrome coro ···················· ···················· ···················· ···················· ···················· ····················
26960 MT704003.1 |Severe acute respiratory syndrome coro ···················· ···················· ···················· ···················· ···················· ····················
26961 MT704004.1 |Severe acute respiratory syndrome coro ···················· ···················· ···················· ···················· ···················· ····················
26962 MT704005.1 |Severe acute respiratory syndrome coro ···················· ···················· ···················· ···················· ···················· ····················
26963 MT704006.1 |Severe acute respiratory syndrome coro ···················· ···················· ···················· ···················· ···················· ····················
26964 MT704007.1 |Severe acute respiratory syndrome coro ···················· ···················· ···················· ···················· ···················· ····················
26965 MT704008.1 |Severe acute respiratory syndrome coro ···················· ···················· ···················· ···················· ···················· ····················
26966 MT704009.1 |Severe acute respiratory syndrome coro ···················· ···················· ···················· ···················· ···················· ····················
26967 MT704011.1 |Severe acute respiratory syndrome coro ···················· ···················· ···················· ···················· ···················· ····················
26968 MT704012.1 |Severe acute respiratory syndrome coro ···················· ···················· ···················· ···················· ···················· ····················
26969 MT704014.1 |Severe acute respiratory syndrome coro ···················· ···················· ···················· ···················· ···················· ····················
26970 MT704017.1 |Severe acute respiratory syndrome coro ···················· ···················· ···················· ···················· ···················· ····················
26971 MT704018.1 |Severe acute respiratory syndrome coro ···················· ···················· ···················· ···················· ···················· ····················
26972 MT704019.1 |Severe acute respiratory syndrome coro ···················· ···················· ···················· ···················· ···················· ····················
26973 MT704022.1 |Severe acute respiratory syndrome coro ···················· ···················· ···················· ···················· ···················· ····················
26974 MT704023.1 |Severe acute respiratory syndrome coro ···················· ···················· ···················· ···················· ···················· ····················
26975 MT704024.1 |Severe acute respiratory syndrome coro ···················· ···················· ···················· ···················· ···················· ····················
26976 MT704025.1 |Severe acute respiratory syndrome coro ···················· ···················· ···················· ···················· ···················· ····················
26977 MT704026.1 |Severe acute respiratory syndrome coro ···················· ···················· ···················· ···················· ···················· ····················
26978 MT704029.1 |Severe acute respiratory syndrome coro ···················· ···················· ···················· ···················· ···················· ····················
26979 MT704030.1 |Severe acute respiratory syndrome coro ···················· ···················· ···················· ···················· ···················· ····················
26980 MT704033.1 |Severe acute respiratory syndrome coro ···················· ···················· ···················· ···················· ···················· ····················
26981 MT704037.1 |Severe acute respiratory syndrome coro ···················· ···················· ···················· ···················· ···················· ····················
26982 MT704038.1 |Severe acute respiratory syndrome coro ···················· ···················· ···················· ···················· ···················· ····················
26983 MT704039.1 |Severe acute respiratory syndrome coro ···················· ···················· ···················· ···················· ···················· ···A················
26984 MT704040.1 |Severe acute respiratory syndrome coro ···················· ···················· ···················· ···················· ···················· ····················
26985 MT704041.1 |Severe acute respiratory syndrome coro ···················· ···················· ···················· ···················· ···················· ····················
26986 MT704042.1 |Severe acute respiratory syndrome coro ···················· ···················· ···················· ···················· ···················· ····················
26987 MT704043.1 |Severe acute respiratory syndrome coro ···················· ···················· ···················· ···················· ···················· ····················
26988 MT704044.1 |Severe acute respiratory syndrome coro ···················· ···················· ···················· ···················· ···················· ····················
26989 MT704045.1 |Severe acute respiratory syndrome coro ···················· ···················· ···················· ···················· ···················· ····················
26990 MT704046.1 |Severe acute respiratory syndrome coro ···················· ···················· ···················· ···················· ···················· ····················
26991 MT704047.1 |Severe acute respiratory syndrome coro ···················· ···················· ···················· ···················· ···················· ····················
26992 MT704049.1 |Severe acute respiratory syndrome coro ···················· ···················· ···················· ···················· ···················· ····················
26993 MT704050.1 |Severe acute respiratory syndrome coro ···················· ···················· ···················· ···················· ···················· ···A················
26994 MT704051.1 |Severe acute respiratory syndrome coro ···················· ···················· ···················· ···················· ···················· ····················
26995 MT704052.1 |Severe acute respiratory syndrome coro ···················· ···················· ···················· ···················· ···················· ····················
26996 MT704055.1 |Severe acute respiratory syndrome coro ···················· ···················· ···················· ···················· ···················· ····················
26997 MT704056.1 |Severe acute respiratory syndrome coro ···················· ···················· ···················· ···················· ···················· ····················
26998 MT704060.1 |Severe acute respiratory syndrome coro ···················· ···················· ···················· ···················· ···················· ····················
26999 MT704061.1 |Severe acute respiratory syndrome coro ···················· ···················· ···················· ···················· ···················· ····················
27000 MT704063.1 |Severe acute respiratory syndrome coro ···················· ···················· ···················· ···················· ···················· ····················
27001 MT704064.1 |Severe acute respiratory syndrome coro ···················· ···················· ···················· ···················· ···················· ····················
27002 MT704065.1 |Severe acute respiratory syndrome coro ···················· ···················· ···················· ···················· ···················· ····················
27003 MT704066.1 |Severe acute respiratory syndrome coro ···················· ···················· ···················· ···················· ···················· ····················
27004 MT704067.1 |Severe acute respiratory syndrome coro ···················· ···················· ···················· ···················· ···················· ····················
27005 MT704068.1 |Severe acute respiratory syndrome coro ···················· ···················· ···················· ···················· ···················· ····················
27006 MT704069.1 |Severe acute respiratory syndrome coro ···················· ···················· ···················· ···················· ···················· ····················
27007 MT704070.1 |Severe acute respiratory syndrome coro ···················· ···················· ···················· ···················· ···················· ····················
27008 MT704071.1 |Severe acute respiratory syndrome coro ···················· ···················· ···················· ···················· ···················· ····················
27009 MT704072.1 |Severe acute respiratory syndrome coro ···················· ···················· ···················· ···················· ···················· ····················
27010 MT704073.1 |Severe acute respiratory syndrome coro ···················· ···················· ···················· ···················· ···················· ····················
27011 MT704074.1 |Severe acute respiratory syndrome coro ···················· ···················· ···················· ···················· ···················· ····················
27012 MT704075.1 |Severe acute respiratory syndrome coro ···················· ···················· ···················· ···················· ···················· ····················
27013 MT704077.1 |Severe acute respiratory syndrome coro ···················· ···················· ···················· ···················· ···················· ····················
27014 MT704080.1 |Severe acute respiratory syndrome coro ···················· ···················· ···················· ···················· ···················· ····················
27015 MT704081.1 |Severe acute respiratory syndrome coro ···················· ···················· ···················· ···················· ···················· ····················
27016 MT704084.1 |Severe acute respiratory syndrome coro ···················· ···················· ···················· ···················· ···················· ····················
27017 MT704085.1 |Severe acute respiratory syndrome coro ···················· ···················· ···················· ···················· ···················· ····················
27018 MT704088.1 |Severe acute respiratory syndrome coro ···················· ···················· ···················· ···················· ···················· ····················
27019 MT704094.1 |Severe acute respiratory syndrome coro ···················· ···················· ···················· ···················· ···················· ····················
27020 MT704095.1 |Severe acute respiratory syndrome coro ···················· ···················· ···················· ···················· ···················· ····················
27021 MT704100.1 |Severe acute respiratory syndrome coro ···················· ···················· ···················· ···················· ···················· ····················
27022 MT704101.1 |Severe acute respiratory syndrome coro ···················· ···················· ···················· ···················· ···················· ····················
27023 MT704102.1 |Severe acute respiratory syndrome coro ···················· ···················· ···················· ···················· ···················· ····················
27024 MT704111.1 |Severe acute respiratory syndrome coro ···················· ···················· ···················· ···················· ···················· ····················
27025 MT704112.1 |Severe acute respiratory syndrome coro ···················· ····T··············· ···················· ···················· ···················· ····················
27026 MT704114.1 |Severe acute respiratory syndrome coro ···················· ···················· ···················· ···················· ···················· ····················
27027 MT704115.1 |Severe acute respiratory syndrome coro ···················· ···················· ···················· ···················· ···················· ····················
27028 MT704116.1 |Severe acute respiratory syndrome coro ···················· ···················· ···················· ···················· ···················· ····················
27029 MT704117.1 |Severe acute respiratory syndrome coro ···················· ····T··············· ···················· ···················· ···················· ····················
27030 MT704119.1 |Severe acute respiratory syndrome coro ···················· ···················· ···················· ···················· ···················· ····················
27031 MT704120.1 |Severe acute respiratory syndrome coro ···················· ···················· ···················· ···················· ···················· ····················
27032 MT704121.1 |Severe acute respiratory syndrome coro ···················· ···················· ···················· ···················· ···················· ····················

| | | | | | | | | | |
|---|---|---|---|---|---|---|---|---|---|
| 27033 | MT704122.1 | \|Severe acute respiratory syndrome coro | ···················· | ···················· | ···················· | ···················· | ···················· | ···················· | |
| 27034 | MT704123.1 | \|Severe acute respiratory syndrome coro | ···················· | ···················· | ···················· | ···················· | ···················· | ···················· | |
| 27035 | MT704124.1 | \|Severe acute respiratory syndrome coro | ···················· | ···················· | ···················· | ···················· | ···················· | ···················· | |
| 27036 | MT704125.1 | \|Severe acute respiratory syndrome coro | ···················· | ···················· | ···················· | ···················· | ···················· | ···················· | |
| 27037 | MT704126.1 | \|Severe acute respiratory syndrome coro | ···················· | ···················· | ···················· | ···················· | ···················· | ···················· | |
| 27038 | MT704127.1 | \|Severe acute respiratory syndrome coro | ···················· | ···················· | ···················· | ···················· | ······T············· | ···················· | |
| 27039 | MT704128.1 | \|Severe acute respiratory syndrome coro | ···················· | ···················· | ···················· | ···················· | ······T············· | ···················· | |
| 27040 | MT704129.1 | \|Severe acute respiratory syndrome coro | ···················· | ···················· | ···················· | ···················· | ···················· | ···················· | |
| 27041 | MT704131.1 | \|Severe acute respiratory syndrome coro | ···················· | ···················· | ···················· | ···················· | ···················· | ···················· | |
| 27042 | MT704132.1 | \|Severe acute respiratory syndrome coro | ···················· | ···················· | ···················· | ···················· | ···················· | ···················· | |
| 27043 | MT704133.1 | \|Severe acute respiratory syndrome coro | ···················· | ···················· | ···················· | ···················· | ···················· | ···················· | |
| 27044 | MT704134.1 | \|Severe acute respiratory syndrome coro | ···················· | ···················· | ···················· | ···················· | ···················· | ···················· | |
| 27045 | MT704135.1 | \|Severe acute respiratory syndrome coro | ···················· | ···················· | ···················· | ···················· | ···················· | ···················· | |
| 27046 | MT704136.1 | \|Severe acute respiratory syndrome coro | ···················· | ···················· | ···················· | ···················· | ···················· | ···················· | |
| 27047 | MT682732.1 | \|Severe acute respiratory syndrome coro | ···················· | ···················· | ···················· | ···················· | ···················· | ···················· | |
| 27048 | MT683386.1 | \|Severe acute respiratory syndrome coro | ···················· | ···················· | ···················· | ···················· | ···················· | ···················· | |
| 27049 | MT683387.1 | \|Severe acute respiratory syndrome coro | ···················· | ···················· | ···················· | ···················· | ···················· | ···················· | |
| 27050 | MT683388.1 | \|Severe acute respiratory syndrome coro | ···················· | ···················· | ···················· | ···················· | ···················· | ···················· | |
| 27051 | MT683389.1 | \|Severe acute respiratory syndrome coro | ···················· | ···················· | ···················· | ···················· | ···················· | ···················· | |
| 27052 | MT683390.1 | \|Severe acute respiratory syndrome coro | ···················· | ···················· | ···················· | ···················· | ···················· | ···················· | |
| 27053 | MT683391.1 | \|Severe acute respiratory syndrome coro | ···················· | ···················A | ···················· | ···················· | ···················· | ···················· | 1 |
| 27054 | MT683392.1 | \|Severe acute respiratory syndrome coro | ···················· | ···················· | ···················· | ···················· | ···················· | ···················· | |
| 27055 | MT683393.1 | \|Severe acute respiratory syndrome coro | ···················· | ···················· | ···················· | ···················· | ···················· | ···················· | |
| 27056 | MT683394.1 | \|Severe acute respiratory syndrome coro | ···················· | ···················· | ···················· | ···················· | ···················· | ···················· | |
| 27057 | MT683395.1 | \|Severe acute respiratory syndrome coro | ···················· | ···················· | ···················· | ···················· | ···················· | ···················· | |
| 27058 | MT683397.1 | \|Severe acute respiratory syndrome coro | ···················· | ···················· | ···················· | ···················· | ···················· | ···················· | |
| 27059 | MT683399.1 | \|Severe acute respiratory syndrome coro | ···················· | ···················· | ···················· | ···················· | ···················· | ···················· | |
| 27060 | MT683403.1 | \|Severe acute respiratory syndrome coro | ···················· | ···················· | ···················· | ···················· | ···················· | ···················· | |
| 27061 | MT683404.1 | \|Severe acute respiratory syndrome coro | ···················· | ···················· | ···················· | ···················· | ···················· | ···················· | |
| 27062 | MT683406.1 | \|Severe acute respiratory syndrome coro | ···················· | ···················· | ···················· | ···················· | ···················· | ···················· | |
| 27063 | MT683407.1 | \|Severe acute respiratory syndrome coro | ···················· | ···················· | ···················· | ···················· | ···················· | ···················· | |
| 27064 | MT683408.1 | \|Severe acute respiratory syndrome coro | ···················· | ···················· | ···················· | ···················· | ···················· | ···················· | |
| 27065 | MT683409.1 | \|Severe acute respiratory syndrome coro | ···················· | ···················· | ···················· | ···················· | ···················· | ···················· | |
| 27066 | MT683410.1 | \|Severe acute respiratory syndrome coro | ···················· | ···················· | ···················· | ···················· | ···················· | ···················· | |
| 27067 | MT683411.1 | \|Severe acute respiratory syndrome coro | ···················· | ···················· | ···················· | ···················· | ···················· | ···················· | |
| 27068 | MT683412.1 | \|Severe acute respiratory syndrome coro | ···················· | ···················· | ···················· | ···················· | ···················· | ···················· | |
| 27069 | MT683413.1 | \|Severe acute respiratory syndrome coro | ···················· | ···················· | ···················· | ···················· | ···················· | ···················· | |
| 27070 | MT683416.1 | \|Severe acute respiratory syndrome coro | ···················· | ···················· | ···················· | ···················· | ···················· | ···················· | |
| 27071 | MT683417.1 | \|Severe acute respiratory syndrome coro | ···················· | ···················· | ···················· | ···················· | ···················· | ···················· | |
| 27072 | MT683418.1 | \|Severe acute respiratory syndrome coro | ···················· | ···················· | ···················· | ···················· | ···················· | ···················· | |
| 27073 | LC556315.1 | \|Severe acute respiratory syndrome coro | ···················· | ···················· | ···················· | ···················· | ···················· | ···················· | |
| 27074 | LC556316.1 | \|Severe acute respiratory syndrome coro | ···················· | ···················· | ···················· | ···················· | ···················· | ···················· | |
| 27075 | LC556317.1 | \|Severe acute respiratory syndrome coro | ···················· | ···················· | ···················· | ···················· | ···················· | ···················· | |
| 27076 | LC556318.1 | \|Severe acute respiratory syndrome coro | ···················· | ···················· | ···················· | ···················· | ···················· | ···················· | |
| 27077 | LC556319.1 | \|Severe acute respiratory syndrome coro | ···················· | ···················· | ···················· | ···················· | ···················· | ···················· | |
| 27078 | LC556320.1 | \|Severe acute respiratory syndrome coro | ···················· | ···················· | ···················· | ···················· | ···················· | ···················· | |
| 27079 | LC556321.1 | \|Severe acute respiratory syndrome coro | ···················· | ···················· | ···················· | ···················· | ···················· | ···················· | |
| 27080 | LC556322.1 | \|Severe acute respiratory syndrome coro | ···················· | ···················· | ···················· | ···················· | ···················· | ···················· | |
| 27081 | LC556323.1 | \|Severe acute respiratory syndrome coro | ···················· | ···················· | ···················· | ···················· | ···················· | ···················· | |
| 27082 | LC556324.1 | \|Severe acute respiratory syndrome coro | ···················· | ···················· | ···················· | ···················· | ···················· | ···················· | |
| 27083 | MT624728.1 | \|Severe acute respiratory syndrome coro | ···················· | ······R············· | ···················· | ···················· | ···················· | ···················· | |
| 27084 | MT624733.1 | \|Severe acute respiratory syndrome coro | ···················· | ······A············· | ···················· | ···················· | ···················· | ···················· | |
| 27085 | MT627389.1 | \|Severe acute respiratory syndrome coro | ···················· | ······A············· | ···················· | ···················· | ···················· | ···················· | |
| 27086 | MT627390.1 | \|Severe acute respiratory syndrome coro | ···················· | ······A············· | ···················· | ···················· | ···················· | ···················· | |
| 27087 | MT627391.1 | \|Severe acute respiratory syndrome coro | ···················· | ···················· | ···················· | ···················· | ···················· | ···················· | |
| 27088 | MT627392.1 | \|Severe acute respiratory syndrome coro | ···················· | ···················· | ···················· | ···················· | ···················· | ···················· | |
| 27089 | MT627393.1 | \|Severe acute respiratory syndrome coro | ···················· | ···················· | ···················· | ···················· | ···················· | ···················· | |
| 27090 | MT627394.1 | \|Severe acute respiratory syndrome coro | ···················· | ···················· | ···················· | ···················· | ···················· | ···················· | |
| 27091 | MT627395.1 | \|Severe acute respiratory syndrome coro | ···················· | ···················· | ···················· | ···················· | ···················· | ···················· | |
| 27092 | MT627396.1 | \|Severe acute respiratory syndrome coro | ···················· | ······R············· | ···················· | ···················· | ···················· | ···················· | |
| 27093 | MT627404.1 | \|Severe acute respiratory syndrome coro | ···················· | ···················· | ···················· | ···················· | ···················· | ···················· | |
| 27094 | MT627405.1 | \|Severe acute respiratory syndrome coro | ···················· | ···················· | ···················· | ···················· | ···················· | ···················· | |
| 27095 | MT627406.1 | \|Severe acute respiratory syndrome coro | ···················· | ······A············· | ···················· | ···················· | ···················· | ···················· | |
| 27096 | MT678839.1 | \|Severe acute respiratory syndrome coro | ···················· | ···················· | ···················· | ···················· | ···················· | ···················· | |
| 27097 | MT679155.1 | \|Severe acute respiratory syndrome coro | ···················· | ···················· | ···················· | ···················· | ···················· | ···················· | |
| 27098 | MT679156.1 | \|Severe acute respiratory syndrome coro | ···················· | ···················· | ···················· | ···················· | ···················· | ···················· | |
| 27099 | MT679157.1 | \|Severe acute respiratory syndrome coro | ···················· | ···················· | ···················· | ···················· | ···················· | ···················· | |
| 27100 | MT679158.1 | \|Severe acute respiratory syndrome coro | ···················· | ···················· | ···················· | ···················· | ···················· | ···················· | |
| 27101 | MT679159.1 | \|Severe acute respiratory syndrome coro | ···················· | ···················· | ···················· | ···················· | ···················· | ···················· | |
| 27102 | MT679160.1 | \|Severe acute respiratory syndrome coro | ···················· | ···················· | ···················· | ···················· | ···················· | ···················· | |
| 27103 | MT679161.1 | \|Severe acute respiratory syndrome coro | ···················· | ···················· | ···················· | ···················· | ···················· | ···················· | |
| 27104 | MT679162.1 | \|Severe acute respiratory syndrome coro | ···················· | ···················· | ···················· | ···················· | ···················· | ···················· | |
| 27105 | MT679163.1 | \|Severe acute respiratory syndrome coro | ···················· | ···················· | ···················· | ···················· | ···················· | ···················· | |
| 27106 | MT679164.1 | \|Severe acute respiratory syndrome coro | ···················· | ···················· | ···················· | ···················· | ···················· | ···················· | |
| 27107 | MT679165.1 | \|Severe acute respiratory syndrome coro | ···················· | ···················· | ···················· | ···················· | ···················· | ···················· | |
| 27108 | MT679166.1 | \|Severe acute respiratory syndrome coro | ···················· | ···················· | ···················· | ···················· | ···················· | ···················· | |
| 27109 | MT679167.1 | \|Severe acute respiratory syndrome coro | ···················· | ···················· | ···················· | ···················· | ···················· | ···················· | |
| 27110 | MT679168.1 | \|Severe acute respiratory syndrome coro | ···················· | ···················· | ···················· | ···················· | ···················· | ···················· | |

27111 MT679169.1 |Severe acute respiratory syndrome coro ···················· ···················· ···················· ···················· ···················· ····················
27112 MT679170.1 |Severe acute respiratory syndrome coro ···················· ···················· ···················· ···················· ···················· ····················
27113 MT679171.1 |Severe acute respiratory syndrome coro ···················· ···················· ···················· ···················· ···················· ····················
27114 MT679172.1 |Severe acute respiratory syndrome coro ···················· ···················· ···················· ···················· ···················· ····················
27115 MT679173.1 |Severe acute respiratory syndrome coro ···················· ···················· ···················· ···················· ···················· ····················
27116 MT679174.1 |Severe acute respiratory syndrome coro ···················· ···················· ···················· ···················· ···················· ····················
27117 MT679175.1 |Severe acute respiratory syndrome coro ···················· ···················· ···················· ···················· ···················· ····················
27118 MT679176.1 |Severe acute respiratory syndrome coro ···················· ···················· ···················· ···················· ···················· ····················
27119 MT679177.1 |Severe acute respiratory syndrome coro ···················· ···················· ···················· ···················· ···················· ····················
27120 MT679178.1 |Severe acute respiratory syndrome coro ···················· ···················· ···················· ···················· ···················· ····················
27121 MT679179.1 |Severe acute respiratory syndrome coro ···················· ···················· ···················· ···················· ···················· ····················
27122 MT679180.1 |Severe acute respiratory syndrome coro ···················· ···················· ···················· ···················· ···················· ····················
27123 MT679181.1 |Severe acute respiratory syndrome coro ···················· ···················· ···················· ···················· ···················· ····················
27124 MT679182.1 |Severe acute respiratory syndrome coro ···················· ···················· ···················· ···················· ···················· ····················
27125 MT679183.1 |Severe acute respiratory syndrome coro ···················· ···················· ···················· ···················· ···················· ····················
27126 MT679184.1 |Severe acute respiratory syndrome coro ···················· ···················· ···················· ···················· ···················· ····················
27127 MT679185.1 |Severe acute respiratory syndrome coro ···················· ···················· ···················· ···················· ···················· ····················
27128 MT679186.1 |Severe acute respiratory syndrome coro ···················· ···················· ···················· ···················· ···················· ····················
27129 MT679187.1 |Severe acute respiratory syndrome coro ···················· ···················· ···················· ···················· ···················· ····················
27130 MT679188.1 |Severe acute respiratory syndrome coro ···················· ···················· ···················· ···················· ···················· ····················
27131 MT679189.1 |Severe acute respiratory syndrome coro ···················· ···················· ···················· ···················· ···················· ····················
27132 MT679190.1 |Severe acute respiratory syndrome coro ···················· ···················· ···················· ···················· ···················· ····················
27133 MT679191.1 |Severe acute respiratory syndrome coro ···················· ···················· ···················· ···················· ···················· ····················
27134 MT679192.1 |Severe acute respiratory syndrome coro ···················· ···················· ···················· ···················· ···················· ····················
27135 MT679193.1 |Severe acute respiratory syndrome coro ···················· ···················· ···················· ···················· ···················· ····················
27136 MT679194.1 |Severe acute respiratory syndrome coro ···················· ···················· ···················· ···················· ···················· ····················
27137 MT679195.1 |Severe acute respiratory syndrome coro ···················· ···················· ···················· ···················· ···················· ····················
27138 MT679196.1 |Severe acute respiratory syndrome coro ···················· ···················· ···················· ···················· ···················· ·········A··········
27139 MT679198.1 |Severe acute respiratory syndrome coro ···················· ···················· ···················· ···················· ···················· ····················
27140 MT679201.1 |Severe acute respiratory syndrome coro ···················· ···················· ···················· ···················· ···················· ····················
27141 MT679202.1 |Severe acute respiratory syndrome coro ···················· ···················· ···················· ···················· ···················· ····················
27142 MT679205.1 |Severe acute respiratory syndrome coro ···················· ···················· ···················· ···················· ···················· ····················
27143 MT679206.1 |Severe acute respiratory syndrome coro ···················· ···················· ···················· ···················· ···················· ····················
27144 MT679208.1 |Severe acute respiratory syndrome coro ···················· ···················· ···················· ···················· ···················· ····················
27145 MT680217.1 |Severe acute respiratory syndrome coro ···················· ···················· ···················· ···················· ···················· ····················
27146 MT680218.1 |Severe acute respiratory syndrome coro ···················· ···················· ···················· ···················· ···················· ····················
27147 MT680219.1 |Severe acute respiratory syndrome coro ···················· ···················· ···················· ···················· ···················· ····················
27148 MT680220.1 |Severe acute respiratory syndrome coro ···················· ···················· ···················· ···················· ···················· ····················
27149 MT680221.1 |Severe acute respiratory syndrome coro ···················· ···················· ···················· ···················· ···················· ····················
27150 MT680222.1 |Severe acute respiratory syndrome coro ···················· ···················· ···················· ···················· ···················· ····················
27151 MT680223.1 |Severe acute respiratory syndrome coro ···················· ···················· ···················· ···················· ···················· ····················
27152 MT680224.1 |Severe acute respiratory syndrome coro ···················· ···················· ···················· ···················· ···················· ····················
27153 MT680225.1 |Severe acute respiratory syndrome coro ···················· ···················· ···················· ···················· ···················· ····················
27154 MT680226.1 |Severe acute respiratory syndrome coro ···················· ···················· ···················· ···················· ···················· ····················
27155 MT680227.1 |Severe acute respiratory syndrome coro ···················· ···················· ···················· ···················· ···················· ····················
27156 MT680228.1 |Severe acute respiratory syndrome coro ···················· ···················· ···················· ···················· ···················· ·········A··········
27157 MT680229.1 |Severe acute respiratory syndrome coro ···················· ···················· ···················· ···················· ···················· ····················
27158 MT680230.1 |Severe acute respiratory syndrome coro ···················· ···················· ···················· ···················· ···················· ····················
27159 MT680231.1 |Severe acute respiratory syndrome coro ···················· ···················· ···················· ···················· ···················· ····················
27160 MT680232.1 |Severe acute respiratory syndrome coro ···················· ···················· ···················· ···················· ···················· ····················
27161 MT680233.1 |Severe acute respiratory syndrome coro ···················· ···················· ···················· ···················· ···················· ····················
27162 MT680234.1 |Severe acute respiratory syndrome coro ···················· ···················· ···················· ···················· ···················· ····················
27163 MT680235.1 |Severe acute respiratory syndrome coro ···················· ···················· ···················· ···················· ···················· ····················
27164 MT680236.1 |Severe acute respiratory syndrome coro ···················· ···················· ···················· ···················· ···················· ····················
27165 MT680237.1 |Severe acute respiratory syndrome coro ···················· ···················· ···················· ···················· ···················· ····················
27166 MT680238.1 |Severe acute respiratory syndrome coro ···················· ···················· ···················· ···················· ···················· ····················
27167 MT680239.1 |Severe acute respiratory syndrome coro ···················· ···················· ···················· ···················· ···················· ····················
27168 MT680240.1 |Severe acute respiratory syndrome coro ···················· ···················· ···················· ···················· ···················· ····················
27169 MT680241.1 |Severe acute respiratory syndrome coro ···················· ···················· ···················· ···················· ···················· ····················
27170 MT680242.1 |Severe acute respiratory syndrome coro ···················· ···················· ···················· ···················· ···················· ····················
27171 MT609918.1 |Severe acute respiratory syndrome coro ···················· ···················· ···················· ···················· ···················· ····················
27172 MT610913.1 |Severe acute respiratory syndrome coro ···················· ···················· ···················· ···················· ···················· ····················
27173 MT611440.1 |Severe acute respiratory syndrome coro ···················· ···················· ···················· ···················· ···················· ····················
27174 MT611441.1 |Severe acute respiratory syndrome coro ···················· ······A············· ···················· ···················· ···················· ····················
27175 MT611442.1 |Severe acute respiratory syndrome coro ···················· ···················· ···················· ···················· ···················· ····················
27176 MT611443.1 |Severe acute respiratory syndrome coro ···················· ··············Y····· ···················· ···················· ···················· ····················
27177 MT611444.1 |Severe acute respiratory syndrome coro ···················· ···················· ···················· ···················· ···················· ····················
27178 MT611448.1 |Severe acute respiratory syndrome coro ···················· ···················· ···················· ···················· ···················· ····················
27179 MT611449.1 |Severe acute respiratory syndrome coro ···················· ···················· ···················· ···················· ···················· ····················
27180 MT611450.1 |Severe acute respiratory syndrome coro ···················· ···················· ···················· ···················· ···················· ····················
27181 MT611451.1 |Severe acute respiratory syndrome coro ···················· ···················· ···················· ···················· ···················· ····················
27182 MT611527.1 |Severe acute respiratory syndrome coro ···················· ···················· ···················· ···················· ···················· ····················
27183 MT611530.1 |Severe acute respiratory syndrome coro ···················· ···················· ···················· ···················· ···················· ····················
27184 MT611536.1 |Severe acute respiratory syndrome coro ···················· ···················· ···················· ···················· ···················· ····················
27185 MT611537.1 |Severe acute respiratory syndrome coro ···················· ···················· ···················· ···················· ···················· ····················
27186 MT611538.1 |Severe acute respiratory syndrome coro ···················· ···················· ···················· ···················· ···················· ····················
27187 MT611539.1 |Severe acute respiratory syndrome coro ···················· ···················· ···················· ···················· ···················· ····················
27188 MT611968.1 |Severe acute respiratory syndrome coro ···················· ·R·················· ···················· ···················· ···················· ···R················

27189 MT614347.1 |Severe acute respiratory syndrome coro ···················· ···················· ···················· ···················· ···················· ····················
27190 MT614348.1 |Severe acute respiratory syndrome coro ···················· ···················· ···················· ···················· ···················· ····················
27191 MT614349.1 |Severe acute respiratory syndrome coro ···················· ·········A·········· ···················· ···················· ···················· ····················
27192 MT614356.1 |Severe acute respiratory syndrome coro ···················· ···················· ···················· ···················· ···················· ····················
27193 MT614357.1 |Severe acute respiratory syndrome coro ···················· ···················· ···················· ···················· ···················· ····················
27194 MT614595.1 |Severe acute respiratory syndrome coro ···················· ···················· ···················· ···················· ···················· ····················
27195 MT614597.1 |Severe acute respiratory syndrome coro ···················· ·········A·········· ···················· ···················· ···················· ····················
27196 MT614598.1 |Severe acute respiratory syndrome coro ···················· ·········A·········· ···················· ···················· ···················· ····················
27197 MT614599.1 |Severe acute respiratory syndrome coro ···················· ···················· ···················· ···················· ···················· ····················
27198 MT614600.1 |Severe acute respiratory syndrome coro ···················· ·········A·········· ···················· ···················· ···················· ····················
27199 MT614601.1 |Severe acute respiratory syndrome coro ···················· ···················· ···················· ···················· ···················· ····················
27200 MT648830.1 |Severe acute respiratory syndrome coro ···················· ···················· ···················· ···················· ···················· ····················
27201 MT658264.1 |Severe acute respiratory syndrome coro ···················· ···················· ···················· ···················· ···················· ····················
27202 MT658500.1 |Severe acute respiratory syndrome coro ···················· ···················· ···················· ···················· ···················· ····················
27203 MT658501.1 |Severe acute respiratory syndrome coro ···················· ···················· ···················· ···················· ···················· ····················
27204 MT658502.1 |Severe acute respiratory syndrome coro ···················· ···················· ···················· ···················· ···················· ····················
27205 MT658503.1 |Severe acute respiratory syndrome coro ···················· ·········A·········· ···················· ···················· ···················· ····················
27206 MT658504.1 |Severe acute respiratory syndrome coro ···················· ······R············· ···················· ···················· ···················· ····················
27207 MT658505.1 |Severe acute respiratory syndrome coro ···················· ···················· ···················· ···················· ···················· ····················
27208 MT675289.1 |Severe acute respiratory syndrome coro ···················· ···················· ···················· ···················· ···················· ····················
27209 MT675517.1 |Severe acute respiratory syndrome coro ···················· ···················· ···················· ···················· ···················· ····················
27210 MT675933.1 |Severe acute respiratory syndrome coro ···················· ·A·················· ···················· ···················· ···················· ··A·················
27211 MT675937.1 |Severe acute respiratory syndrome coro ···················· ·A·················· ···················· ···················· ···················· ··A·················
27212 MT675938.1 |Severe acute respiratory syndrome coro ···················· ·A·················· ···················· ···················· ···················· ··A·················
27213 MT675939.1 |Severe acute respiratory syndrome coro ···················· ·A·················· ···················· ···················· ···················· ··A·················
27214 MT675940.1 |Severe acute respiratory syndrome coro ···················· ·A·················· ···················· ···················· ···················· ··A·················
27215 MT675941.1 |Severe acute respiratory syndrome coro ···················· ·A·················· ···················· ···················· ···················· ··A·················
27216 MT675942.1 |Severe acute respiratory syndrome coro ···················· ·A·················· ···················· ···················· ···················· ··A·················
27217 MT675943.1 |Severe acute respiratory syndrome coro ···················· ·A·················· ···················· ···················· ···················· ··A·················
27218 MT675944.1 |Severe acute respiratory syndrome coro ···················· ·A·················· ···················· ···················· ···················· ··A·················
27219 MT675945.1 |Severe acute respiratory syndrome coro ···················· ·A·················· ···················· ···················· ···················· ··A·················
27220 MT675950.1 |Severe acute respiratory syndrome coro ···················· ·A·················· ···················· ···················· ···················· ··A·················
27221 MT675951.1 |Severe acute respiratory syndrome coro ···················· ···················· ···················· ···················· ···················· ····················
27222 MT675952.1 |Severe acute respiratory syndrome coro ···················· ·A·················· ···················· ···················· ···················· ··A·················
27223 MT675954.1 |Severe acute respiratory syndrome coro ···················· ·A·················· ···················· ···················· ···················· ··A·················
27224 MT675955.1 |Severe acute respiratory syndrome coro ···················· ·A·················· ···················· ···················· ···················· ··A·················
27225 MT675956.1 |Severe acute respiratory syndrome coro ···················· ···················· ···················· ···················· ···················· ····················
27226 MT675957.1 |Severe acute respiratory syndrome coro ···················· ·A·················· ···················· ···················· ···················· ··A·················
27227 MT675958.1 |Severe acute respiratory syndrome coro ···················· ···················· ···················· ···················· ···················· ····················
27228 MT675959.1 |Severe acute respiratory syndrome coro ···················· ·A·················· ···················· ···················· ···················· ··A·················
27229 MT676013.1 |Severe acute respiratory syndrome coro ···················· ···················· ···················· ···················· ···················· ····················
27230 MT676014.1 |Severe acute respiratory syndrome coro ···················· ···················· ···················· ···················· ···················· ····················
27231 MT676288.1 |Severe acute respiratory syndrome coro ···················· ···················· ···················· ···················· ···················· ····················
27232 MT676289.1 |Severe acute respiratory syndrome coro ···················· ···················· ···················· ···················· ···················· ····················
27233 MT676290.1 |Severe acute respiratory syndrome coro ···················· ···················· ···················· ···················· ···················· ····················
27234 MT676291.1 |Severe acute respiratory syndrome coro ···················· ···················· ···················· ···················· ···················· ··A·················
27235 MT676366.1 |Severe acute respiratory syndrome coro ···················· ···················· ···················· ···················· ···················· ····················
27236 MT676367.1 |Severe acute respiratory syndrome coro ···················· ···················· ···················· ···················· ···················· ··A·················
27237 MT676368.1 |Severe acute respiratory syndrome coro ···················· ·A·················· ···················· ···················· ···················· ··A·················
27238 MT676369.1 |Severe acute respiratory syndrome coro ···················· ···················· ···················· ···················· ···················· ····················
27239 MT676388.1 |Severe acute respiratory syndrome coro ···················· ···················· ···················· ···················· ···················· ····················
27240 MT676389.1 |Severe acute respiratory syndrome coro ···················· ···················· ···················· ···················· ···················· ····················
27241 MT676390.1 |Severe acute respiratory syndrome coro ···················· ·A·················· ···················· ···················· ···················· ····················
27242 MT676397.1 |Severe acute respiratory syndrome coro ···················· ···················· ···················· ···················· ···················· ····················
27243 MT614596.1 |Severe acute respiratory syndrome coro ···················· ···················· ···················· ···················· ···················· ····················
27244 MT669321.1 |Severe acute respiratory syndrome coro ···················· ···················· ···················· ···················· ···················· ····················
27245 MT669322.1 |Severe acute respiratory syndrome coro ···················· ·A·················· ···················· ···················· ···················· ··A·················
27246 MT670008.1 |Severe acute respiratory syndrome coro ···················· ···················· ···················· ···················· ···················· ····················
27247 MT670013.1 |Severe acute respiratory syndrome coro ···················· ···················· ···················· ···················· ···················· ····················
27248 MT670014.1 |Severe acute respiratory syndrome coro ···················· ···················· ···················· ···················· ···················· ····················
27249 MT670015.1 |Severe acute respiratory syndrome coro ···················· ···················· ···················· ···················· ···················· ····················
27250 MT670017.1 |Severe acute respiratory syndrome coro ···················· ···················· ···················· ···················· ···················· ····················
27251 MT670018.1 |Severe acute respiratory syndrome coro ···················· ···················· ···················· ···················· ···················· ····················
27252 MT670019.1 |Severe acute respiratory syndrome coro ···················· ···················· ···················· ···················· ···················· ····················
27253 MT670021.1 |Severe acute respiratory syndrome coro ···················· ···················· ···················· ···················· ···················· ····················
27254 MT670022.1 |Severe acute respiratory syndrome coro ···················· ···················· ···················· ···················· ···················· ····················
27255 MT670023.1 |Severe acute respiratory syndrome coro ···················· ···················· ···················· ···················· ···················· ····················
27256 MT671810.1 |Severe acute respiratory syndrome coro ···················· ···················· ···················· ···················· ···················· ····················
27257 MT671811.1 |Severe acute respiratory syndrome coro ···················· ···················· ···················· ···················· ···················· ····················
27258 MT671812.1 |Severe acute respiratory syndrome coro ···················· ···················· ···················· ···················· ···················· ····················
27259 MT671813.1 |Severe acute respiratory syndrome coro ···················· ···················· ···················· ···················· ···················· ····················
27260 MT671814.1 |Severe acute respiratory syndrome coro ···················· ···················· ···················· ···················· ···················· ····················
27261 MT671815.1 |Severe acute respiratory syndrome coro ···················· ···················· ···················· ···················· ···················· ····················
27262 MT671816.1 |Severe acute respiratory syndrome coro ···················· ···················· ···················· N··················· ···················· ····················
27263 MT671817.1 |Severe acute respiratory syndrome coro ···················· ···················· ···················· ···················· ···················· ····················
27264 MT671818.1 |Severe acute respiratory syndrome coro ···················· ···················· ···················· ···················· ···················· ····················
27265 MT671819.1 |Severe acute respiratory syndrome coro ···················· ···················· ···················· ···················· ···················· ····················
27266 MT671820.1 |Severe acute respiratory syndrome coro ···················· ···················· ···················· ···················· ···················· ····················

| | | | | | | | | |
|---|---|---|---|---|---|---|---|---|
| 27267 | MT671821.1 | Severe acute respiratory syndrome coro | .................... | .................... | .................... | .................... | .................... | .................... |
| 27268 | MT671822.1 | Severe acute respiratory syndrome coro | .................... | .................... | .................... | .................... | .................... | .................... |
| 27269 | MT671823.1 | Severe acute respiratory syndrome coro | .................... | .................... | .................... | .................... | .................... | .................... |
| 27270 | MT671824.1 | Severe acute respiratory syndrome coro | .................... | .................... | .................... | .................... | .................... | .................... |
| 27271 | MT671825.1 | Severe acute respiratory syndrome coro | .................... | .................... | .................... | .................... | .................... | .................... |
| 27272 | MT671826.1 | Severe acute respiratory syndrome coro | .................... | .................... | .................... | .................... | .................... | .................... |
| 27273 | MT671827.1 | Severe acute respiratory syndrome coro | .................... | .................... | .................... | .................... | .................... | .................... |
| 27274 | MT671828.1 | Severe acute respiratory syndrome coro | .................... | .................... | .................... | .................... | .................... | .................... |
| 27275 | MT664105.1 | Severe acute respiratory syndrome coro | .................... | .................... | .................... | .................... | .................... | .................... |
| 27276 | MT664106.1 | Severe acute respiratory syndrome coro | .................... | .................... | .................... | .................... | .................... | .................... |
| 27277 | MT664107.1 | Severe acute respiratory syndrome coro | .................... | .A.................. | .................... | .................... | .................... | ...A................ |
| 27278 | MT664109.1 | Severe acute respiratory syndrome coro | .................... | .................... | .................... | .................... | .................... | .................... |
| 27279 | MT664117.1 | Severe acute respiratory syndrome coro | .................... | .A.................. | .................... | .................... | .................... | ...A................ |
| 27280 | MT664118.1 | Severe acute respiratory syndrome coro | ...T................ | .A.................. | .................... | .................... | .................... | ...A................ |
| 27281 | MT664143.1 | Severe acute respiratory syndrome coro | .................... | .A.................. | .................... | .................... | .................... | ...A................ |
| 27282 | MT664161.1 | Severe acute respiratory syndrome coro | .................... | .A.................. | .................... | .................... | .................... | ...A................ |
| 27283 | MT664169.1 | Severe acute respiratory syndrome coro | .................... | .................... | .................... | .................... | .................... | .................... |
| 27284 | MT664170.1 | Severe acute respiratory syndrome coro | .................... | .A.................. | .................... | .................... | .................... | ...A................ |
| 27285 | MT664171.1 | Severe acute respiratory syndrome coro | .................... | .................... | .................... | .................... | .................... | .................... |
| 27286 | MT664172.1 | Severe acute respiratory syndrome coro | .................... | .................... | .................... | .................... | .................... | ...A................ |
| 27287 | MT664175.1 | Severe acute respiratory syndrome coro | .................... | .................... | .................... | .................... | .................... | .................... |
| 27288 | MT664197.1 | Severe acute respiratory syndrome coro | .................... | .................... | .................... | .................... | .................... | .................... |
| 27289 | MT664201.1 | Severe acute respiratory syndrome coro | .................... | .A.................. | .................... | .................... | .................... | ...A................ |
| 27290 | MT664202.1 | Severe acute respiratory syndrome coro | .................... | .................... | .................... | .................... | .................... | .................... |
| 27291 | MT664203.1 | Severe acute respiratory syndrome coro | .................... | .A.................. | .................... | .................... | .................... | ...A................ |
| 27292 | MT664205.1 | Severe acute respiratory syndrome coro | .................... | .A.................. | .................... | .................... | .................... | ...A................ |
| 27293 | MT664209.1 | Severe acute respiratory syndrome coro | .................... | .................... | .................... | .................... | .................... | .................... |
| 27294 | MT664727.1 | Severe acute respiratory syndrome coro | .................... | .................... | .................... | .................... | ......T............. | .................... |
| 27295 | MT664729.1 | Severe acute respiratory syndrome coro | .................... | .A.................. | .................... | .................... | .................... | ...A................ |
| 27296 | MT664774.1 | Severe acute respiratory syndrome coro | .................... | .................... | .................... | .................... | .................... | ...A................ |
| 27297 | MT664793.1 | Severe acute respiratory syndrome coro | .................... | .................... | .................... | .................... | .................... | ...A................ |
| 27298 | MT664795.1 | Severe acute respiratory syndrome coro | .................... | .................... | .................... | .................... | .................... | .................... |
| 27299 | MT664796.1 | Severe acute respiratory syndrome coro | .................... | .A.................. | .................... | .................... | .................... | ...A................ |
| 27300 | MT664807.1 | Severe acute respiratory syndrome coro | .................... | .A.................. | .................... | .................... | .................... | ...A................ |
| 27301 | MT664808.1 | Severe acute respiratory syndrome coro | .................... | .A.................. | .................... | .................... | .................... | ...A................ |
| 27302 | MT664822.1 | Severe acute respiratory syndrome coro | .................... | .A.................. | .................... | .................... | .................... | ...A................ |
| 27303 | MT664986.1 | Severe acute respiratory syndrome coro | .................... | .A.................. | .................... | .................... | .................... | ...A................ |
| 27304 | MT664990.1 | Severe acute respiratory syndrome coro | .................... | .................... | .................... | .................... | .................... | ...A................ |
| 27305 | MT665006.1 | Severe acute respiratory syndrome coro | .................... | .A.................. | .................... | .................... | .................... | ...A................ |
| 27306 | MT665028.1 | Severe acute respiratory syndrome coro | .................... | .A.................. | .................... | .................... | .................... | ...A................ |
| 27307 | MT665970.1 | Severe acute respiratory syndrome coro | .................... | .A.................. | .................... | .................... | .................... | ...A................ |
| 27308 | MT665972.1 | Severe acute respiratory syndrome coro | .................... | .A.................. | .................... | .................... | .................... | ...A................ |
| 27309 | MT665974.1 | Severe acute respiratory syndrome coro | .................... | .................... | .................... | .................... | .................... | .................... |
| 27310 | MT666042.1 | Severe acute respiratory syndrome coro | .................... | .................... | .................... | .................... | .................... | .................... |
| 27311 | MT666068.1 | Severe acute respiratory syndrome coro | .................... | .................... | .................... | .................... | .................... | .................... |
| 27312 | MT666099.1 | Severe acute respiratory syndrome coro | .................... | .................... | .................... | .................... | .................... | .................... |
| 27313 | MT667351.1 | Severe acute respiratory syndrome coro | .................... | .................... | .................... | .................... | .................... | .................... |
| 27314 | MT668716.1 | Severe acute respiratory syndrome coro | .................... | .................... | .................... | .................... | .................... | .................... |
| 27315 | MT661524.1 | Severe acute respiratory syndrome coro | .................... | .................... | .................... | .................... | .................... | .................... |
| 27316 | MT653094.1 | Severe acute respiratory syndrome coro | .................... | .................... | .................... | .................... | .................... | .................... |
| 27317 | MT653097.1 | Severe acute respiratory syndrome coro | .................... | .................... | .................... | .................... | .................... | .................... |
| 27318 | MT653099.1 | Severe acute respiratory syndrome coro | .................... | .................... | .................... | .................... | .................... | .................... |
| 27319 | MT655131.1 | Severe acute respiratory syndrome coro | .................... | .................... | .................... | .................... | .................... | .................... |
| 27320 | MT655132.1 | Severe acute respiratory syndrome coro | .................... | .................... | .................... | .................... | .................... | .................... |
| 27321 | MT655133.1 | Severe acute respiratory syndrome coro | .................... | .................... | .................... | .................... | .................... | .................... |
| 27322 | MT655134.1 | Severe acute respiratory syndrome coro | .................... | .................... | .................... | .................... | .................... | .................... |
| 27323 | MT655135.1 | Severe acute respiratory syndrome coro | .................... | .................... | .................... | .................... | .................... | .................... |
| 27324 | MT655742.1 | Severe acute respiratory syndrome coro | .................... | .................... | .................... | .................... | ...C................ | .................... |
| 27325 | MT655743.1 | Severe acute respiratory syndrome coro | .................... | .................... | .................... | .................... | .................... | .................... |
| 27326 | MT655744.1 | Severe acute respiratory syndrome coro | .................... | .................... | .................... | .................... | .................... | .................... |
| 27327 | MT655745.1 | Severe acute respiratory syndrome coro | .................... | .................... | .................... | .................... | .................... | .................... |
| 27328 | MT655746.1 | Severe acute respiratory syndrome coro | .................... | .................... | .................... | .................... | .................... | .................... |
| 27329 | MT655750.1 | Severe acute respiratory syndrome coro | .................... | .................... | .................... | .................... | .................... | .................... |
| 27330 | MT655751.1 | Severe acute respiratory syndrome coro | .................... | .................... | .................... | .................... | .................... | .................... |
| 27331 | MT655948.1 | Severe acute respiratory syndrome coro | .................... | .................... | .................... | .................... | .................... | .................... |
| 27332 | MT656010.1 | Severe acute respiratory syndrome coro | .................... | .................... | .................... | .................... | .................... | .................... |
| 27333 | MT656011.1 | Severe acute respiratory syndrome coro | .................... | .................... | .................... | .................... | .................... | .................... |
| 27334 | MT656256.1 | Severe acute respiratory syndrome coro | .................... | .................... | .................... | .................... | .................... | .................... |
| 27335 | MT656257.1 | Severe acute respiratory syndrome coro | .................... | .................... | .................... | .................... | .................... | .................... |
| 27336 | MT656590.1 | Severe acute respiratory syndrome coro | .................... | .................... | .................... | .................... | .................... | .................... |
| 27337 | MT656591.1 | Severe acute respiratory syndrome coro | ........T........... | .................... | .................... | .................... | .................... | .................... |
| 27338 | MT657271.1 | Severe acute respiratory syndrome coro | .................... | .................... | .................... | .................... | .................... | .................... |
| 27339 | MT657275.1 | Severe acute respiratory syndrome coro | .................... | .................... | .................... | .................... | .................... | .................... |
| 27340 | MT657958.1 | Severe acute respiratory syndrome coro | .................... | .................... | .................... | .................... | .................... | .................... |
| 27341 | MT658507.1 | Severe acute respiratory syndrome coro | .................... | .................... | .................... | .................... | .................... | .................... |
| 27342 | MT658508.1 | Severe acute respiratory syndrome coro | .................... | .A.................. | .................... | .................... | .................... | ...A................ |
| 27343 | MT658509.1 | Severe acute respiratory syndrome coro | .................... | ........A........... | .................... | .................... | .................... | .................... |
| 27344 | MT646053.1 | Severe acute respiratory syndrome coro | .................... | .................... | .................... | .................... | .................... | .................... |

27345 MT646054.1 |Severe acute respiratory syndrome coro .................... .................... ..................... .................... .................... ....................
27346 MT646064.1 |Severe acute respiratory syndrome coro .................... .................... ..................... .................... .................... ....................
27347 MT646065.1 |Severe acute respiratory syndrome coro .................... .................... ..................... .................... .................... ....................
27348 MT646068.1 |Severe acute respiratory syndrome coro .................... .................... ..................... .................... .................... ....................
27349 MT646069.1 |Severe acute respiratory syndrome coro .................... .................... ..................... .................... .................... ....................
27350 MT646070.1 |Severe acute respiratory syndrome coro .................... .................... ..................... .................... .................... ....................
27351 MT646071.1 |Severe acute respiratory syndrome coro .................... .................... ..................... .................... .................... ....................
27352 MT646072.1 |Severe acute respiratory syndrome coro .................... .................... ..................... .................... .................... ....................
27353 MT646073.1 |Severe acute respiratory syndrome coro .................... .................... ..................... .................... .................... ....................
27354 MT646074.1 |Severe acute respiratory syndrome coro .................... .................... ..................... .................... .................... ....................
27355 MT646075.1 |Severe acute respiratory syndrome coro .................... .................... ..................... .................... .................... ....................
27356 MT646076.1 |Severe acute respiratory syndrome coro .................... .................... ..................... .................... .................... ....................
27357 MT646077.1 |Severe acute respiratory syndrome coro .................... .................... ..................... .................... .................... ....................
27358 MT646078.1 |Severe acute respiratory syndrome coro .................... .................... ..................... .................... .................... ....................
27359 MT646079.1 |Severe acute respiratory syndrome coro .................... .................... ..................... .................... .................... ....................
27360 MT646080.1 |Severe acute respiratory syndrome coro .................... .................... ..................... .................... .................... ....................
27361 MT646081.1 |Severe acute respiratory syndrome coro .................... .................... ..................... .................... .................... ....................
27362 MT646082.1 |Severe acute respiratory syndrome coro .................... .................... ..................... .................... .................... ....................
27363 MT646083.1 |Severe acute respiratory syndrome coro .................... .................... ..................... .................... .................... ....................
27364 MT646084.1 |Severe acute respiratory syndrome coro .................... .................... ..................... .................... .................... ....................
27365 MT646086.1 |Severe acute respiratory syndrome coro .................... .................... ..................... .................... .................... ....................
27366 MT646087.1 |Severe acute respiratory syndrome coro .................... .................... ..................... .................... .................... ....................
27367 MT646088.1 |Severe acute respiratory syndrome coro .................... .................... ..................... .................... .................... ....................
27368 MT646089.1 |Severe acute respiratory syndrome coro .................... .................... ..................... .................... .................... ....................
27369 MT646090.1 |Severe acute respiratory syndrome coro .................... .................... ..................... .................... .................... ....................
27370 MT646091.1 |Severe acute respiratory syndrome coro .................... .................... ..................... .................... .................... ....................
27371 MT646092.1 |Severe acute respiratory syndrome coro .................... .................... ..................... .................... .................... ....................
27372 MT646093.1 |Severe acute respiratory syndrome coro .................... .................... ..................... .................... .................... ....................
27373 MT646094.1 |Severe acute respiratory syndrome coro .................... .................... ..................... .................... .................... ....................
27374 MT646096.1 |Severe acute respiratory syndrome coro .................... .................... ..................... .................... .................... ....................
27375 MT646097.1 |Severe acute respiratory syndrome coro .................... .................... ..................... .................... .................... ....................
27376 MT646098.1 |Severe acute respiratory syndrome coro .................... .................... ..................... .................... .................... ....................
27377 MT646101.1 |Severe acute respiratory syndrome coro .................... .................... ..................... .................... .................... ....................
27378 MT646106.1 |Severe acute respiratory syndrome coro .................... .................... ..................... .................... .................... ....................
27379 MT646112.1 |Severe acute respiratory syndrome coro .................... .................... ..................... .................... .................... ....................
27380 MT646116.1 |Severe acute respiratory syndrome coro .................... .................... ..................... .................... .................... ....................
27381 MT646117.1 |Severe acute respiratory syndrome coro .................... .................... ..................... .................... .................... ....................
27382 MT646120.1 |Severe acute respiratory syndrome coro .................... .................... ..................... .................... .................... ....................
27383 MT648676.1 |Severe acute respiratory syndrome coro .................... .................... ..................... .................... .................... ....................
27384 MT641485.1 |Severe acute respiratory syndrome coro .................... .................... ..................... .................... .................... ....................
27385 MT641486.1 |Severe acute respiratory syndrome coro .................... .................... ..................... .................... .................... ....................
27386 MT641488.1 |Severe acute respiratory syndrome coro .................... .................... ..................... .................... .................... ....................
27387 MT641489.1 |Severe acute respiratory syndrome coro .................... .................... ..................... .................... .................... ....................
27388 MT641490.1 |Severe acute respiratory syndrome coro .................... .................... ..................... .................... .................... ....................
27389 MT641492.1 |Severe acute respiratory syndrome coro .................... .................... ..................... .................... .................... ....................
27390 MT641493.1 |Severe acute respiratory syndrome coro .................... .................... ..................... .................... .................... ....................
27391 MT641494.1 |Severe acute respiratory syndrome coro .................... .................... ..................... .................... .................... ....................
27392 MT641495.1 |Severe acute respiratory syndrome coro .................... .................... ..................... .................... .................... ....................
27393 MT641496.1 |Severe acute respiratory syndrome coro .................... .................... ..................... .................... .................... ....................
27394 MT641497.1 |Severe acute respiratory syndrome coro .................... .................... ..................... .................... .................... ....................
27395 MT641498.1 |Severe acute respiratory syndrome coro .................... .................... ..................... .................... .................... ....................
27396 MT641499.1 |Severe acute respiratory syndrome coro .................... .................... ..................... .................... .................... ....................
27397 MT641500.1 |Severe acute respiratory syndrome coro .................... .................... ..................... .................... .................... ....................
27398 MT641502.1 |Severe acute respiratory syndrome coro .................... .................... ..................... .................... .................... ....................
27399 MT641503.1 |Severe acute respiratory syndrome coro .................... .................... ..................... .................... .................... ....................
27400 MT641504.1 |Severe acute respiratory syndrome coro .................... .................... ..................... .................... .................... ....................
27401 MT641505.1 |Severe acute respiratory syndrome coro .................... .................... ..................... .................... .................... ....................
27402 MT641506.1 |Severe acute respiratory syndrome coro .................... .................... ..................... .................... .................... ....................
27403 MT641507.1 |Severe acute respiratory syndrome coro .................... .................... ..................... .................... .................... ....................
27404 MT641508.1 |Severe acute respiratory syndrome coro .................... .................... ..................... .................... .................... ....................
27405 MT641509.1 |Severe acute respiratory syndrome coro .................... .................... ..................... .................... .................... ....................
27406 MT641510.1 |Severe acute respiratory syndrome coro .................... .................... ..................... .................... .................... ....................
27407 MT641511.1 |Severe acute respiratory syndrome coro .................... .................... ..................... .................... .................... ....................
27408 MT641512.1 |Severe acute respiratory syndrome coro .................... .................... ..................... .................... .................... ....................
27409 MT641513.1 |Severe acute respiratory syndrome coro .................... .................... ..................... .................... .................... ....................
27410 MT641514.1 |Severe acute respiratory syndrome coro .................... .................... ..................... .................... .................... ....................
27411 MT641515.1 |Severe acute respiratory syndrome coro .................... .................... ..................... .................... .................... ....................
27412 MT641516.1 |Severe acute respiratory syndrome coro .................... .................... ..................... .................... .................... ....................
27413 MT641517.1 |Severe acute respiratory syndrome coro .................... .................... ..................... .................... .................... ....................
27414 MT641518.1 |Severe acute respiratory syndrome coro .................... .................... ..................... .................... .................... ....................
27415 MT641519.1 |Severe acute respiratory syndrome coro .................... .................... ..................... .................... .................... ....................
27416 MT641520.1 |Severe acute respiratory syndrome coro .................... .................... ..................... .................... .................... ....................
27417 MT641521.1 |Severe acute respiratory syndrome coro .................... .................... ..................... .................... .................... ....................
27418 MT641522.1 |Severe acute respiratory syndrome coro .................... .................... ..................... .................... .................... ....................
27419 MT641523.1 |Severe acute respiratory syndrome coro .................... .................... ..................... .................... .................... ....................
27420 MT641524.1 |Severe acute respiratory syndrome coro .................... .................... ..................... .................... .................... ....................
27421 MT641525.1 |Severe acute respiratory syndrome coro .................... .................... ..................... .................... .................... ....................
27422 MT641526.1 |Severe acute respiratory syndrome coro .................... .................... ..................... .................... .................... ....................

27423 MT641527.1 |Severe acute respiratory syndrome coro .................... .................... .................... .................... .................... ....................
27424 MT641528.1 |Severe acute respiratory syndrome coro .................... .................... .................... .................... .................... ....................
27425 MT641529.1 |Severe acute respiratory syndrome coro .................... .................... .................... .................... .................... ....................
27426 MT641530.1 |Severe acute respiratory syndrome coro .................... .................... .................... .................... .................... ....................
27427 MT641532.1 |Severe acute respiratory syndrome coro .................... .................... .................... .................... .................... ....................
27428 MT641533.1 |Severe acute respiratory syndrome coro .................... .................... .................... .................... .................... ....................
27429 MT641534.1 |Severe acute respiratory syndrome coro .................... .................... .................... .................... .................... ....................
27430 MT641535.1 |Severe acute respiratory syndrome coro .................... .................... .................... .................... .................... ....................
27431 MT641536.1 |Severe acute respiratory syndrome coro .................... .................... .................... .................... .................... ....................
27432 MT641537.1 |Severe acute respiratory syndrome coro .................... .................... .................... .................... .................... ....................
27433 MT641538.1 |Severe acute respiratory syndrome coro .................... .................... .................... .................... .................... ....................
27434 MT641539.1 |Severe acute respiratory syndrome coro .................... .................... .................... .................... .................... ....................
27435 MT641540.1 |Severe acute respiratory syndrome coro .................... .................... .................... .................... .................... ....................
27436 MT641541.1 |Severe acute respiratory syndrome coro .................... .................... .................... .................... .................... ....................
27437 MT641542.1 |Severe acute respiratory syndrome coro .................... .................... .................... .................... .................... ....................
27438 MT641543.1 |Severe acute respiratory syndrome coro .................... .................... .................... .................... .................... ....................
27439 MT641544.1 |Severe acute respiratory syndrome coro .................... .................... .................... .................... .................... ....................
27440 MT641545.1 |Severe acute respiratory syndrome coro .................... .................... .................... .................... .................... ....................
27441 MT641546.1 |Severe acute respiratory syndrome coro .................... .................... .................... .................... .................... ....................
27442 MT641547.1 |Severe acute respiratory syndrome coro .................... .................... .................... .................... .................... ....................
27443 MT641548.1 |Severe acute respiratory syndrome coro .................... .................... .................... .................... .................... ....................
27444 MT641549.1 |Severe acute respiratory syndrome coro .................... .................... .................... .................... .................... ....................
27445 MT641550.1 |Severe acute respiratory syndrome coro .................... .................... .................... .................... .................... ....................
27446 MT641551.1 |Severe acute respiratory syndrome coro .................... .................... .................... .................... .................... ....................
27447 MT641552.1 |Severe acute respiratory syndrome coro .................... .................... .................... .................... .................... ....................
27448 MT641553.1 |Severe acute respiratory syndrome coro .................... .................... .................... .................... .................... ....................
27449 MT641554.1 |Severe acute respiratory syndrome coro .................... .................... .................... .................... .................... ....................
27450 MT641557.1 |Severe acute respiratory syndrome coro .................... .................... .................... .................... .................... ....................
27451 MT641558.1 |Severe acute respiratory syndrome coro .................... .................... .................... .................... .................... ....................
27452 MT641559.1 |Severe acute respiratory syndrome coro .................... .................... .................... .................... .................... ....................
27453 MT641560.1 |Severe acute respiratory syndrome coro .................... .................... .................... .................... .................... ....................
27454 MT641561.1 |Severe acute respiratory syndrome coro .................... .................... .................... .................... .................... ....................
27455 MT641562.1 |Severe acute respiratory syndrome coro .................... .................... .................... .................... .................... ....................
27456 MT641563.1 |Severe acute respiratory syndrome coro .................... .................... .................... .................... .................... ....................
27457 MT641564.1 |Severe acute respiratory syndrome coro .................... .................... .................... .................... .................... ....................
27458 MT641565.1 |Severe acute respiratory syndrome coro .................... .................... .................... .................... .................... ....................
27459 MT641566.1 |Severe acute respiratory syndrome coro .................... .................... .................... .................... .................... ....................
27460 MT641567.1 |Severe acute respiratory syndrome coro .................... .................... .................... .................... .................... ....................
27461 MT641568.1 |Severe acute respiratory syndrome coro .................... .................... .................... .................... .................... ....................
27462 MT641569.1 |Severe acute respiratory syndrome coro .................... .................... .................... .................... .................... ....................
27463 MT641570.1 |Severe acute respiratory syndrome coro .................... .................... .................... .................... .................... ....................
27464 MT641571.1 |Severe acute respiratory syndrome coro .................... .................... .................... .................... .................... ....................
27465 MT641572.1 |Severe acute respiratory syndrome coro .................... .................... .................... .................... .................... ....................
27466 MT641573.1 |Severe acute respiratory syndrome coro .................... .................... .................... .................... .................... ....................
27467 MT641574.1 |Severe acute respiratory syndrome coro .................... .................... .................... .................... .................... ....................
27468 MT641575.1 |Severe acute respiratory syndrome coro .................... .................... .................... .................... .................... ....................
27469 MT641576.1 |Severe acute respiratory syndrome coro .................... .................... .................... .................... .................... ....................
27470 MT641577.1 |Severe acute respiratory syndrome coro .................... .................... .................... .................... .................... ....................
27471 MT641578.1 |Severe acute respiratory syndrome coro .................... .................... .................... .................... .................... ....................
27472 MT641579.1 |Severe acute respiratory syndrome coro .................... .................... .................... .................... .................... ....................
27473 MT641580.1 |Severe acute respiratory syndrome coro ....T............... .................... .................... .................... .................... ....................
27474 MT641581.1 |Severe acute respiratory syndrome coro .................... .................... .................... .................... .................... ....................
27475 MT641582.1 |Severe acute respiratory syndrome coro .................... .................... .................... .................... .................... ....................
27476 MT641583.1 |Severe acute respiratory syndrome coro .................... .................... .................... .................... .................... ....................
27477 MT641584.1 |Severe acute respiratory syndrome coro .................... .................... .................... .................... .................... ....................
27478 MT641585.1 |Severe acute respiratory syndrome coro .................... .................... .................... .................... .................... ....................
27479 MT641586.1 |Severe acute respiratory syndrome coro .................... .................... .................... .................... .................... ....................
27480 MT641587.1 |Severe acute respiratory syndrome coro .................... .................... .................... .................... .................... ....................
27481 MT641588.1 |Severe acute respiratory syndrome coro .................... .................... .................... .................... .................... ....................
27482 MT641589.1 |Severe acute respiratory syndrome coro .................... .................... .................... .................... .................... ....................
27483 MT641590.1 |Severe acute respiratory syndrome coro .................... .................... .................... .................... .................... ....................
27484 MT641591.1 |Severe acute respiratory syndrome coro .................... .................... .................... .................... .................... ....................
27485 MT641592.1 |Severe acute respiratory syndrome coro .................... .................... .................... .................... .................... ....................
27486 MT641593.1 |Severe acute respiratory syndrome coro .................... .................... .................... .................... .................... ....................
27487 MT641594.1 |Severe acute respiratory syndrome coro .................... .................... .................... .................... .................... ....................
27488 MT641595.1 |Severe acute respiratory syndrome coro .................... .................... .................... .................... .................... ....................
27489 MT641596.1 |Severe acute respiratory syndrome coro .................... .................... .................... .................... .................... ....................
27490 MT641598.1 |Severe acute respiratory syndrome coro .................... .................... .................... .................... .................... ....................
27491 MT641599.1 |Severe acute respiratory syndrome coro .................... .................... .................... .................... .................... ....................
27492 MT641600.1 |Severe acute respiratory syndrome coro .................... .................... .................... .................... .................... ....................
27493 MT641601.1 |Severe acute respiratory syndrome coro .................... .................... .................... .................... .................... ....................
27494 MT641602.1 |Severe acute respiratory syndrome coro .................... .................... .................... .................... .................... ....................
27495 MT641603.1 |Severe acute respiratory syndrome coro .................... .................... .................... .................... .................... ....................
27496 MT641604.1 |Severe acute respiratory syndrome coro .................... .................... .................... .................... .................... ....................
27497 MT641605.1 |Severe acute respiratory syndrome coro .................... .................... .................... .................... .................... ....................
27498 MT641606.1 |Severe acute respiratory syndrome coro .................... .................... .................... .................... .................... ....................
27499 MT641607.1 |Severe acute respiratory syndrome coro .................... .................... .................... .................... .................... ....................
27500 MT641608.1 |Severe acute respiratory syndrome coro .................... .................... .................... .................... .................... ....................

27501 MT641609.1 |Severe acute respiratory syndrome coro .................... .................... .................... .................... .................... ....................
27502 MT641610.1 |Severe acute respiratory syndrome coro .................... .................... .................... .................... .................... ....................
27503 MT641611.1 |Severe acute respiratory syndrome coro .................... .................... .................... .................... .................... ....................
27504 MT641612.1 |Severe acute respiratory syndrome coro .................... .................... .................... .................... .................... ....................
27505 MT641613.1 |Severe acute respiratory syndrome coro .................... .................... .................... .................... .................... ....................
27506 MT641614.1 |Severe acute respiratory syndrome coro .................... .................... .................... .................... .................... ....................
27507 MT641615.1 |Severe acute respiratory syndrome coro .................... .................... .................... .................... .................... ....................
27508 MT641617.1 |Severe acute respiratory syndrome coro .................... .................... .................... .................... .................... ....................
27509 MT641618.1 |Severe acute respiratory syndrome coro .................... .................... .................... .................... .................... ....................
27510 MT641619.1 |Severe acute respiratory syndrome coro .................... .................... .................... .................... .................... ....................
27511 MT641620.1 |Severe acute respiratory syndrome coro .................... .................... .................... .................... .................... ....................
27512 MT641621.1 |Severe acute respiratory syndrome coro .................... .................... .................... .................... .................... ....................
27513 MT641622.1 |Severe acute respiratory syndrome coro .................... .................... .................... .................... .................... ....................
27514 MT641623.1 |Severe acute respiratory syndrome coro .................... .................... .................... .................... .................... ....................
27515 MT641624.1 |Severe acute respiratory syndrome coro .................... .................... .................... .................... .................... ....................
27516 MT641625.1 |Severe acute respiratory syndrome coro .................... .................... .................... .................... .................... ....................
27517 MT641626.1 |Severe acute respiratory syndrome coro .................... .................... .................... .................... .................... ....................
27518 MT641627.1 |Severe acute respiratory syndrome coro .................... .................... .................... .................... .................... ....................
27519 MT641628.1 |Severe acute respiratory syndrome coro .................... .................... .................... .................... .................... ....................
27520 MT641629.1 |Severe acute respiratory syndrome coro .................... .................... .................... .................... .................... ....................
27521 MT641630.1 |Severe acute respiratory syndrome coro .................... .................... .................... .................... .................... ....................
27522 MT641631.1 |Severe acute respiratory syndrome coro .................... .................... .................... .................... .................... ....................
27523 MT641632.1 |Severe acute respiratory syndrome coro .................... .................... .................... .................... .................... ....................
27524 MT641633.1 |Severe acute respiratory syndrome coro .................... .................... .................... .................... .................... ....................
27525 MT641634.1 |Severe acute respiratory syndrome coro .................... .................... .................... .................... .................... ....................
27526 MT641635.1 |Severe acute respiratory syndrome coro .................... .................... .................... .................... .................... ....................
27527 MT641636.1 |Severe acute respiratory syndrome coro .................... .................... .................... .................... .................... ....................
27528 MT641639.1 |Severe acute respiratory syndrome coro .................... .................... .................... .................... .................... ....................
27529 MT641640.1 |Severe acute respiratory syndrome coro .................... .................... .................... .................... .................... ....................
27530 MT641641.1 |Severe acute respiratory syndrome coro .................... .................... .................... .................... .................... ....................
27531 MT641642.1 |Severe acute respiratory syndrome coro .................... .................... .................... .................... .................... ....................
27532 MT641643.1 |Severe acute respiratory syndrome coro .................... .................... .................... .................... .................... ....................
27533 MT641644.1 |Severe acute respiratory syndrome coro .................... .................... .................... .................... .................... ....................
27534 MT641645.1 |Severe acute respiratory syndrome coro .................... .................... .................... .................... .................... ....................
27535 MT641646.1 |Severe acute respiratory syndrome coro .................... .................... .................... .................... .................... ....................
27536 MT641648.1 |Severe acute respiratory syndrome coro .................... .................... .................... .................... .................... ....................
27537 MT641651.1 |Severe acute respiratory syndrome coro .................... .................... .................... .................... .................... ....................
27539 MT641661.1 |Severe acute respiratory syndrome coro .................... .................... .................... .................... .................... ....................
27541 MT641663.1 |Severe acute respiratory syndrome coro .................... .................... .................... .................... .................... ....................
27542 MT641664.1 |Severe acute respiratory syndrome coro .................... .................... .................... .................... .................... ....................
27543 MT641665.1 |Severe acute respiratory syndrome coro .................... .................... .................... .................... .................... ....................
27544 MT641666.1 |Severe acute respiratory syndrome coro .................... .................... .................... .................... .................... ....................
27545 MT641667.1 |Severe acute respiratory syndrome coro .................... .................... .................... .................... .................... ....................
27546 MT641669.1 |Severe acute respiratory syndrome coro .................... .................... .................... .................... .................... ....................
27547 MT641674.1 |Severe acute respiratory syndrome coro .................... .................... .................... .................... .................... ...............R....
27548 MT641676.1 |Severe acute respiratory syndrome coro .................... .................... .................... .................... .................... ....................
27549 MT641677.1 |Severe acute respiratory syndrome coro .................... .................... .................... .................... .................... ....................
27550 MT641678.1 |Severe acute respiratory syndrome coro .................... .................... .................... .................... .................... ....................
27552 MT641681.1 |Severe acute respiratory syndrome coro .................... .................... .................... .................... .................... ....................
27553 MT641682.1 |Severe acute respiratory syndrome coro .................... .................... .................... .................... .................... ....................
27554 MT641686.1 |Severe acute respiratory syndrome coro .................... .................... .................... .................... .................... ....................
27555 MT641689.1 |Severe acute respiratory syndrome coro .................... .................... .................... .................... .................... ....................
27556 MT641692.1 |Severe acute respiratory syndrome coro .................... .................... .................... .................... .................... ....................
27557 MT641693.1 |Severe acute respiratory syndrome coro .................... .................... .................... .................... .................... ....................
27559 MT641699.1 |Severe acute respiratory syndrome coro .................... .................... .................... .................... .............Y...... ....................
27560 MT641708.1 |Severe acute respiratory syndrome coro .................... .................... .................... .................... .................... ....................
27561 MT641710.1 |Severe acute respiratory syndrome coro .................... .................... .................... .................... .................... ....................
27562 MT641720.1 |Severe acute respiratory syndrome coro .................... .................... .................... .................... .................... ....................
27563 MT641721.1 |Severe acute respiratory syndrome coro .................... .................... .................... .................... .................... ....................
27567 MT641728.1 |Severe acute respiratory syndrome coro .................... .................... .................... .................... .................... ....................
27568 MT641729.1 |Severe acute respiratory syndrome coro .................... .................... .................... .................... .................... ....................
27569 MT641730.1 |Severe acute respiratory syndrome coro .................... .................... .................... .................... .................... ....................
27570 MT641731.1 |Severe acute respiratory syndrome coro .................... .................... .................... .................... .................... ....................
27571 MT641733.1 |Severe acute respiratory syndrome coro .................... .................... .................... .................... .................... ....................
27572 MT641735.1 |Severe acute respiratory syndrome coro .................... .................... .................... .................... .................... ....................
27573 MT641736.1 |Severe acute respiratory syndrome coro .................... .................... .................... .................... .................... ....................
27574 MT641737.1 |Severe acute respiratory syndrome coro .................... .................... .................... .................... .................... ....................
27575 MT641738.1 |Severe acute respiratory syndrome coro .................... .................... .................... .................... .................... ....................
27576 MT641739.1 |Severe acute respiratory syndrome coro .................... .................... .................... .................... .................... ....................
27577 MT641740.1 |Severe acute respiratory syndrome coro .................... .................... .................... .................... .................... ....................
27578 MT641741.1 |Severe acute respiratory syndrome coro .................... .................... .................... .................... .................... ....................
27579 MT641742.1 |Severe acute respiratory syndrome coro .................... .................... .................... .................... .................... ....................
27580 MT641743.1 |Severe acute respiratory syndrome coro .................... .................... .................... .................... .................... ....................
27581 MT641744.1 |Severe acute respiratory syndrome coro .................... .................... .................... .................... .................... ....................
27582 MT641745.1 |Severe acute respiratory syndrome coro .................... .................... .................... .................... .................... ....................
27583 MT641746.1 |Severe acute respiratory syndrome coro .................... .................... .................... .................... .................... ....................
27584 MT641748.1 |Severe acute respiratory syndrome coro .................... .................... .................... .................... .................... ....................
27585 MT641750.1 |Severe acute respiratory syndrome coro .................... .................... .................... .................... .................... ....................

| | | | | | | | | | |
|---|---|---|---|---|---|---|---|---|---|
| 27586 | MT641751.1 | \|Severe acute respiratory syndrome coro | .................... | .................... | .................... | .................... | .................... | .................... | |
| 27587 | MT641752.1 | \|Severe acute respiratory syndrome coro | .................... | .................... | .................... | .................... | .................... | .................... | |
| 27588 | MT641753.1 | \|Severe acute respiratory syndrome coro | .................... | .................... | .................... | .................... | .................... | .................... | |
| 27589 | MT641754.1 | \|Severe acute respiratory syndrome coro | .................... | .................... | .................... | .................... | .................... | .................... | |
| 27590 | MT641757.1 | \|Severe acute respiratory syndrome coro | .................... | .................... | .................... | .................... | .................... | .................... | |
| 27591 | MT641758.1 | \|Severe acute respiratory syndrome coro | .................... | .................... | .................... | .................... | .................... | .................... | |
| 27592 | MT641759.1 | \|Severe acute respiratory syndrome coro | .................... | .................... | .................... | .................... | .................... | .................... | |
| 27593 | MT641760.1 | \|Severe acute respiratory syndrome coro | .................... | .................... | .................... | .................... | .................... | .................... | |
| 27594 | MT641761.1 | \|Severe acute respiratory syndrome coro | .................... | .................... | .................... | .................... | .................... | .................... | |
| 27595 | MT641762.1 | \|Severe acute respiratory syndrome coro | .................... | .................... | .................... | .................... | .................... | .................... | |
| 27597 | MT641764.1 | \|Severe acute respiratory syndrome coro | .................... | .................... | .................... | .................... | .................... | .................... | |
| 27598 | MT641766.1 | \|Severe acute respiratory syndrome coro | .................... | .................... | .................... | .................... | .................... | .................... | |
| 27599 | MT641767.1 | \|Severe acute respiratory syndrome coro | .................... | .................... | .................... | .................... | .................... | N................... | |
| 27601 | MT641774.1 | \|Severe acute respiratory syndrome coro | .................... | .................... | .................... | .................... | NNN................. | .................... | 1 |
| 27602 | MT641776.1 | \|Severe acute respiratory syndrome coro | .................... | .................... | .................... | .................... | NNN................. | .................... | 1 |
| 27604 | MT641778.1 | \|Severe acute respiratory syndrome coro | .................... | .................... | .................... | .................... | .................... | .................... | |
| 27605 | MT642068.1 | \|Severe acute respiratory syndrome coro | .................... | .................... | .................... | .................... | .................... | .................... | |
| 27606 | MT642069.1 | \|Severe acute respiratory syndrome coro | .................... | .................... | .................... | .................... | .................... | .................... | |
| 27607 | MT642072.1 | \|Severe acute respiratory syndrome coro | .................... | .................... | .................... | .................... | .................... | .................... | |
| 27608 | MT642073.1 | \|Severe acute respiratory syndrome coro | .................... | .................... | .................... | .................... | .................... | .................... | |
| 27609 | MT642074.1 | \|Severe acute respiratory syndrome coro | .................... | .................... | .................... | .................... | .................... | .................... | |
| 27610 | MT642078.1 | \|Severe acute respiratory syndrome coro | .................... | .................... | .................... | .................... | .................... | .................... | |
| 27611 | MT642079.1 | \|Severe acute respiratory syndrome coro | .................... | .................... | .................... | .................... | .................... | .................... | |
| 27612 | MT642080.1 | \|Severe acute respiratory syndrome coro | .................... | .A.................. | .................... | .................... | .................... | .................... | |
| 27613 | MT642081.1 | \|Severe acute respiratory syndrome coro | .................... | .................... | .................... | .................... | .................... | .................... | |
| 27614 | MT642082.1 | \|Severe acute respiratory syndrome coro | .................... | .................... | .................... | .................... | .................... | .................... | |
| 27615 | MT642083.1 | \|Severe acute respiratory syndrome coro | .................... | .................... | .................... | .................... | .................... | .................... | |
| 27616 | MT642085.1 | \|Severe acute respiratory syndrome coro | .................... | .................... | .................... | .................... | .................... | .................... | |
| 27617 | MT642086.1 | \|Severe acute respiratory syndrome coro | .................... | .................... | .................... | .................... | .................... | .................... | |
| 27618 | MT642087.1 | \|Severe acute respiratory syndrome coro | .................... | .................... | .................... | .................... | .................... | .................... | |
| 27619 | MT642088.1 | \|Severe acute respiratory syndrome coro | .................... | .................... | .................... | .................... | .................... | .................... | |
| 27620 | MT642089.1 | \|Severe acute respiratory syndrome coro | .................... | .................... | .................... | .................... | .................... | .................... | |
| 27621 | MT642091.1 | \|Severe acute respiratory syndrome coro | .................... | .................... | .................... | .................... | .................... | .................... | |
| 27622 | MT642092.1 | \|Severe acute respiratory syndrome coro | .................... | .................... | .................... | .................... | .................... | .................... | |
| 27623 | MT642093.1 | \|Severe acute respiratory syndrome coro | .................... | .................... | .................... | .................... | .................... | .................... | |
| 27624 | MT642094.1 | \|Severe acute respiratory syndrome coro | .................... | .................... | .................... | .................... | .................... | .................... | |
| 27625 | MT642095.1 | \|Severe acute respiratory syndrome coro | .................... | .................... | .................... | .................... | .................... | .................... | |
| 27626 | MT642096.1 | \|Severe acute respiratory syndrome coro | .................... | .................... | .................... | .................... | .................... | .................... | |
| 27627 | MT642097.1 | \|Severe acute respiratory syndrome coro | .................... | .................... | .................... | .................... | .................... | .................... | |
| 27628 | MT642098.1 | \|Severe acute respiratory syndrome coro | .................... | .................... | .................... | .................... | .................... | .................... | |
| 27629 | MT642099.1 | \|Severe acute respiratory syndrome coro | .................... | .A.................. | .................... | .................... | .................... | .................... | |
| 27630 | MT642100.1 | \|Severe acute respiratory syndrome coro | .................... | .................... | .................... | .................... | .................... | .................... | |
| 27631 | MT642101.1 | \|Severe acute respiratory syndrome coro | .................... | .................... | .................... | .................... | .................... | .................... | |
| 27632 | MT642102.1 | \|Severe acute respiratory syndrome coro | .................... | .................... | .................... | .................... | .................... | .................... | |
| 27633 | MT642103.1 | \|Severe acute respiratory syndrome coro | .................... | .................... | .................... | .................... | .................... | .................... | |
| 27634 | MT642104.1 | \|Severe acute respiratory syndrome coro | .................... | .................... | .................... | .................... | .................... | .................... | |
| 27635 | MT642105.1 | \|Severe acute respiratory syndrome coro | .................... | .................... | .................... | .................... | .................... | .................... | |
| 27636 | MT642106.1 | \|Severe acute respiratory syndrome coro | .................... | .................... | .................... | .................... | .................... | .................... | |
| 27637 | MT642107.1 | \|Severe acute respiratory syndrome coro | .................... | .................... | .................... | .................... | .................... | .................... | |
| 27638 | MT642109.1 | \|Severe acute respiratory syndrome coro | .................... | .................... | .................... | .................... | .................... | .................... | |
| 27639 | MT642111.1 | \|Severe acute respiratory syndrome coro | .................... | .................... | .................... | .................... | .................... | .................... | |
| 27640 | MT642112.1 | \|Severe acute respiratory syndrome coro | .................... | .................... | .................... | .................... | .................... | .................... | |
| 27641 | MT642113.1 | \|Severe acute respiratory syndrome coro | .................... | .................... | .................... | .................... | .................... | .................... | |
| 27642 | MT642114.1 | \|Severe acute respiratory syndrome coro | .................... | .................... | .................... | .................... | .................... | .................... | |
| 27643 | MT642115.1 | \|Severe acute respiratory syndrome coro | .................... | .................... | .................... | .................... | .................... | .................... | |
| 27644 | MT642116.1 | \|Severe acute respiratory syndrome coro | .................... | .................... | .................... | .................... | .................... | .................... | |
| 27645 | MT642117.1 | \|Severe acute respiratory syndrome coro | .................... | .................... | .................... | .................... | .................... | .................... | |
| 27646 | MT642118.1 | \|Severe acute respiratory syndrome coro | .................... | .................... | .................... | .................... | .................... | .................... | |
| 27647 | MT642119.1 | \|Severe acute respiratory syndrome coro | .................... | .................... | .................... | .................... | .................... | .................... | |
| 27648 | MT642120.1 | \|Severe acute respiratory syndrome coro | .................... | .................... | .................... | .................... | .................... | .................... | |
| 27649 | MT642121.1 | \|Severe acute respiratory syndrome coro | .................... | .................... | .................... | .................... | .................... | .................... | |
| 27650 | MT642122.1 | \|Severe acute respiratory syndrome coro | .................... | .................... | .................... | .................... | .................... | .................... | |
| 27651 | MT642123.1 | \|Severe acute respiratory syndrome coro | .................... | .................... | .................... | .................... | .................... | .................... | |
| 27652 | MT642124.1 | \|Severe acute respiratory syndrome coro | .................... | .................... | .................... | .................... | .................... | .................... | |
| 27653 | MT642125.1 | \|Severe acute respiratory syndrome coro | .................... | .................... | .................... | .................... | .................... | .................... | |
| 27654 | MT642126.1 | \|Severe acute respiratory syndrome coro | .................... | .................... | .................... | .................... | .................... | .................... | |
| 27655 | MT642127.1 | \|Severe acute respiratory syndrome coro | .................... | .................... | .................... | .................... | .................... | .................... | |
| 27656 | MT642128.1 | \|Severe acute respiratory syndrome coro | .................... | .................... | .................... | .................... | .................... | .................... | |
| 27657 | MT642129.1 | \|Severe acute respiratory syndrome coro | .................... | .................... | .................... | .................... | .................... | .................... | |
| 27658 | MT642130.1 | \|Severe acute respiratory syndrome coro | .................... | .................... | .................... | .................... | .................... | .................... | |
| 27659 | MT642131.1 | \|Severe acute respiratory syndrome coro | .................... | .................... | .................... | .................... | .................... | .................... | |
| 27660 | MT642132.1 | \|Severe acute respiratory syndrome coro | .................... | .................... | .................... | .................... | .................... | .................... | |
| 27661 | MT642133.1 | \|Severe acute respiratory syndrome coro | .................... | .................... | .................... | .................... | .................... | .................... | |
| 27662 | MT642134.1 | \|Severe acute respiratory syndrome coro | .................... | .................... | .................... | .................... | .................... | .................... | |
| 27663 | MT642136.1 | \|Severe acute respiratory syndrome coro | .................... | .................... | .................... | .................... | .................... | .................... | |
| 27664 | MT642137.1 | \|Severe acute respiratory syndrome coro | .................... | .................... | .................... | .................... | .................... | .................... | |
| 27665 | MT642138.1 | \|Severe acute respiratory syndrome coro | .................... | .................... | .................... | .................... | .................... | .................... | |
| 27666 | MT642139.1 | \|Severe acute respiratory syndrome coro | .................... | .................... | .................... | .................... | .................... | .................... | |

27667 MT642140.1 |Severe acute respiratory syndrome coro ···················· ···················· ···················· ···················· ···················· ····················
27668 MT642141.1 |Severe acute respiratory syndrome coro ···················· ···················· ···················· ···················· ···················· ····················
27669 MT642142.1 |Severe acute respiratory syndrome coro ···················· ···················· ···················· ···················· ···················· ····················
27670 MT642143.1 |Severe acute respiratory syndrome coro ···················· ···················· ···················· ···················· ···················· ····················
27671 MT642144.1 |Severe acute respiratory syndrome coro ···················· ···················· ···················· ···················· ···················· ····················
27672 MT642145.1 |Severe acute respiratory syndrome coro ···················· ···················· ···················· ···················· ···················· ····················
27673 MT642146.1 |Severe acute respiratory syndrome coro ···················· ···················· ···················· ···················· ···················· ····················
27674 MT642147.1 |Severe acute respiratory syndrome coro ···················· ···················· ···················· ···················· ···················· ····················
27675 MT642148.1 |Severe acute respiratory syndrome coro ···················· ···················· ···················· ···················· ···················· ····················
27676 MT642149.1 |Severe acute respiratory syndrome coro ···················· ···················· ···················· ···················· ···················· ····················
27677 MT642150.1 |Severe acute respiratory syndrome coro ···················· ···················· ···················· ···················· ···················· ····················
27678 MT642151.1 |Severe acute respiratory syndrome coro ···················· ···················· ···················· ···················· ···················· ····················
27679 MT642152.1 |Severe acute respiratory syndrome coro ···················· ···················· ···················· ···················· ···················· ····················
27680 MT642153.1 |Severe acute respiratory syndrome coro ···················· ···················· ···················· ···················· ···················· ····················
27681 MT642154.1 |Severe acute respiratory syndrome coro ···················· ···················· ···················· ···················· ···················· ····················
27682 MT642155.1 |Severe acute respiratory syndrome coro ···················· ···················· ···················· ···················· ···················· ····················
27683 MT642156.1 |Severe acute respiratory syndrome coro ···················· ···················· ···················· ···················· ···················· ····················
27684 MT642157.1 |Severe acute respiratory syndrome coro ···················· ·A·················· ···················· ···················· ···················· ····················
27685 MT642158.1 |Severe acute respiratory syndrome coro ···················· ···················· ···················· ···················· ···················· ····················
27686 MT642159.1 |Severe acute respiratory syndrome coro ···················· ···················· ···················· ···················· ···················· ····················
27687 MT642160.1 |Severe acute respiratory syndrome coro ···················· ···················· ···················· ···················· ···················· ····················
27688 MT642161.1 |Severe acute respiratory syndrome coro ···················· ···················· ···················· ···················· ···················· ····················
27689 MT642162.1 |Severe acute respiratory syndrome coro ···················· ···················· ···················· ···················· ···················· ····················
27690 MT642163.1 |Severe acute respiratory syndrome coro ···················· ···················· ···················· ···················· ···················· ····················
27691 MT642164.1 |Severe acute respiratory syndrome coro ···················· ···················· ···················· ···················· ···················· ····················
27692 MT642165.1 |Severe acute respiratory syndrome coro ···················· ···················· ···················· ···················· ···················· ····················
27693 MT642166.1 |Severe acute respiratory syndrome coro ···················· ···················· ···················· ···················· ···················· ····················
27694 MT642167.1 |Severe acute respiratory syndrome coro ···················· ···················· ···················· ···················· ···················· ····················
27695 MT642168.1 |Severe acute respiratory syndrome coro ···················· ···················· ···················· ···················· ···················· ····················
27696 MT642169.1 |Severe acute respiratory syndrome coro ···················· ···················· ···················· ···················· ···················· ····················
27697 MT642170.1 |Severe acute respiratory syndrome coro ···················· ···················· ···················· ···················· ···················· ····················
27698 MT642171.1 |Severe acute respiratory syndrome coro ···················· ···················· ···················· ···················· ···················· ····················
27699 MT642172.1 |Severe acute respiratory syndrome coro ···················· ···················· ···················· ···················· ···················· ····················
27700 MT642173.1 |Severe acute respiratory syndrome coro ···················· ·A·················· ···················· ···················· ···················· ····················
27701 MT642174.1 |Severe acute respiratory syndrome coro ···················· ···················· ···················· ···················· ···················· ····················
27702 MT642175.1 |Severe acute respiratory syndrome coro ···················· ···················· ···················· ···················· ···················· ····················
27703 MT642176.1 |Severe acute respiratory syndrome coro ···················· ···················· ···················· ···················· ···················· ····················
27704 MT642177.1 |Severe acute respiratory syndrome coro ···················· ···················· ···················· ···················· ···················· ····················
27705 MT642178.1 |Severe acute respiratory syndrome coro ···················· ···················· ···················· ···················· ···················· ····················
27706 MT642179.1 |Severe acute respiratory syndrome coro ···················· ···················· ···················· ···················· ···················· ····················
27707 MT642180.1 |Severe acute respiratory syndrome coro ···················· ···················· ···················· ···················· ···················· ····················
27708 MT642181.1 |Severe acute respiratory syndrome coro ···················· ···················· ···················· ···················· ···················· ····················
27709 MT642182.1 |Severe acute respiratory syndrome coro ···················· ···················· ···················· ···················· ···················· ····················
27710 MT642183.1 |Severe acute respiratory syndrome coro ···················· ···················· ···················· ···················· ···················· ····················
27711 MT642184.1 |Severe acute respiratory syndrome coro ···················· ···················· ···················· ···················· ···················· ····················
27712 MT642185.1 |Severe acute respiratory syndrome coro ···················· ···················· ···················· ···················· ···················· ····················
27713 MT642186.1 |Severe acute respiratory syndrome coro ···················· ···················· ···················· ···················· ···················· ····················
27714 MT642187.1 |Severe acute respiratory syndrome coro ···················· ···················· ···················· ···················· ···················· ····················
27715 MT642188.1 |Severe acute respiratory syndrome coro ···················· ···················· ···················· ···················· ···················· ····················
27716 MT642189.1 |Severe acute respiratory syndrome coro ···················· N······N············ ···················· ···················· ···················· ····················
27717 MT642190.1 |Severe acute respiratory syndrome coro ···················· ···················· ···················· ···················· ···················· ····················
27718 MT642191.1 |Severe acute respiratory syndrome coro ···················· ···················· ···················· ···················· ···················· ····················
27719 MT642192.1 |Severe acute respiratory syndrome coro ···················· ···················· ···················· ···················· ···················· ····················
27720 MT642193.1 |Severe acute respiratory syndrome coro ···················· ···················· ···················· ···················· ···················· ····················
27721 MT642194.1 |Severe acute respiratory syndrome coro ···················· ···················· ···················· ···················· ···················· ····················
27722 MT642195.1 |Severe acute respiratory syndrome coro ···················· ···················· ···················· ···················· ···················· ····················
27723 MT642196.1 |Severe acute respiratory syndrome coro ···················· ···················· ···················· ···················· ···················· ····················
27724 MT642197.1 |Severe acute respiratory syndrome coro ···················· ···················· ···················· ···················· ···················· ····················
27725 MT642198.1 |Severe acute respiratory syndrome coro ···················· ···················· ···················· ···················· ···················· ····················
27726 MT642199.1 |Severe acute respiratory syndrome coro ···················· ···················· ···················· ···················· ···················· ····················
27727 MT642200.1 |Severe acute respiratory syndrome coro ···················· ···················· ···················· ···················· ···················· ····················
27728 MT642201.1 |Severe acute respiratory syndrome coro ···················· ···················· ···················· ···················· ···················· ····················
27729 MT642202.1 |Severe acute respiratory syndrome coro ···················· ···················· ···················· ···················· ···················· ····················
27730 MT642203.1 |Severe acute respiratory syndrome coro ···················· ···················· ···················· ···················· ···················· ····················
27731 MT642204.1 |Severe acute respiratory syndrome coro ···················· ···················· ···················· ···················· ···················· ····················
27732 MT642205.1 |Severe acute respiratory syndrome coro ···················· ···················· ···················· ···················· ···················· ····················
27733 MT642206.1 |Severe acute respiratory syndrome coro ···················· ···················· ···················· ···················· ···················· ····················
27734 MT642207.1 |Severe acute respiratory syndrome coro ···················· ···················· ···················· ···················· ···················· ····················
27735 MT642209.1 |Severe acute respiratory syndrome coro ···················· ···················· ···················· ···················· ···················· ····················
27736 MT642210.1 |Severe acute respiratory syndrome coro ···················· ···················· ···················· ···················· ···················· ····················
27737 MT642211.1 |Severe acute respiratory syndrome coro ···················· ···················· ···················· ···················· ···················· ····················
27738 MT642212.1 |Severe acute respiratory syndrome coro ···················· ···················· ···················· ···················· ···················· ····················
27739 MT642213.1 |Severe acute respiratory syndrome coro ···················· ···················· ···················· ···················· ···················· ····················
27740 MT642214.1 |Severe acute respiratory syndrome coro ···················· ···················· ···················· ···················· ···················· ····················
27741 MT642215.1 |Severe acute respiratory syndrome coro ···················· ···················· ···················· ···················· ···················· ····················
27742 MT642216.1 |Severe acute respiratory syndrome coro ···················· ···················· ···················· ···················· ···················· ····················
27743 MT642217.1 |Severe acute respiratory syndrome coro ···················· ···················· ···················· ···················· ···················· ····················
27744 MT642218.1 |Severe acute respiratory syndrome coro ···················· ···················· ···················· ···················· ···················· ····················

| | | | | | | | | |
|---|---|---|---|---|---|---|---|---|
| 27745 | MT642219.1 | Severe acute respiratory syndrome coro | .................... | .................... | .................... | .................... | .................... | .................... |
| 27746 | MT642220.1 | Severe acute respiratory syndrome coro | .................... | .................... | .................... | .................... | .................... | .................... |
| 27747 | MT642221.1 | Severe acute respiratory syndrome coro | .................... | .................... | .................... | .................... | .................... | .................... |
| 27748 | MT642222.1 | Severe acute respiratory syndrome coro | .................... | .................... | .................... | .................... | .................... | .................... |
| 27749 | MT642223.1 | Severe acute respiratory syndrome coro | .................... | .................... | .................... | .................... | .................... | .................... |
| 27750 | MT642224.1 | Severe acute respiratory syndrome coro | .................... | .................... | .................... | .................... | .................... | .................... |
| 27751 | MT642225.1 | Severe acute respiratory syndrome coro | .................... | .................... | .................... | .................... | .................... | .................... |
| 27752 | MT642226.1 | Severe acute respiratory syndrome coro | .................... | .................... | .................... | .................... | .................... | .................... |
| 27753 | MT642227.1 | Severe acute respiratory syndrome coro | .................... | .................... | .................... | .................... | .................... | .................... |
| 27754 | MT642228.1 | Severe acute respiratory syndrome coro | .................... | .................... | .................... | .................... | .................... | .................... |
| 27755 | MT642230.1 | Severe acute respiratory syndrome coro | .................... | .................... | .................... | .................... | .................... | .................... |
| 27756 | MT642231.1 | Severe acute respiratory syndrome coro | .................... | .................... | .................... | .................... | .................... | .................... |
| 27757 | MT642232.1 | Severe acute respiratory syndrome coro | .................... | .................... | .................... | .................... | .................... | .................... |
| 27758 | MT642233.1 | Severe acute respiratory syndrome coro | .................... | .................... | .................... | .................... | .................... | .................... |
| 27759 | MT642234.1 | Severe acute respiratory syndrome coro | .................... | .................... | .................... | .................... | .................... | .................... |
| 27760 | MT642235.1 | Severe acute respiratory syndrome coro | .................... | .................... | .................... | .................... | .................... | .................... |
| 27761 | MT642238.1 | Severe acute respiratory syndrome coro | .................... | .................... | .................... | .................... | .................... | .................... |
| 27762 | MT642239.1 | Severe acute respiratory syndrome coro | .................... | .................... | .................... | .................... | .................... | .................... |
| 27763 | MT642240.1 | Severe acute respiratory syndrome coro | .................... | .A.................. | .................... | .................... | .................... | .................... |
| 27764 | MT642242.1 | Severe acute respiratory syndrome coro | .................... | .................... | .................... | .................... | .................... | .................... |
| 27765 | MT642243.1 | Severe acute respiratory syndrome coro | .................... | .................... | .................... | .................... | .................... | .................... |
| 27766 | MT642244.1 | Severe acute respiratory syndrome coro | .................... | .................... | .................... | .................... | .................... | .................... |
| 27767 | MT642245.1 | Severe acute respiratory syndrome coro | .................... | .................... | .................... | .................... | .................... | .................... |
| 27768 | MT642246.1 | Severe acute respiratory syndrome coro | .................... | .................... | .................... | .................... | .................... | .................... |
| 27769 | MT642247.1 | Severe acute respiratory syndrome coro | .................... | .................... | .................... | .................... | .................... | .................... |
| 27770 | MT642248.1 | Severe acute respiratory syndrome coro | .................... | .................... | .................... | .................... | .................... | .................... |
| 27771 | MT642249.1 | Severe acute respiratory syndrome coro | .................... | .................... | .................... | .................... | .................... | .................... |
| 27772 | MT642250.1 | Severe acute respiratory syndrome coro | .................... | .................... | .................... | .................... | .................... | .................... |
| 27773 | MT642251.1 | Severe acute respiratory syndrome coro | .................... | .................... | .................... | .................... | .................... | .................... |
| 27774 | MT642252.1 | Severe acute respiratory syndrome coro | .................... | .................... | .................... | .................... | .................... | .................... |
| 27775 | MT642253.1 | Severe acute respiratory syndrome coro | .................... | .................... | .................... | .................... | .................... | .................... |
| 27776 | MT642255.1 | Severe acute respiratory syndrome coro | .................... | .................... | .................... | .................... | .................... | .................... |
| 27777 | MT642256.1 | Severe acute respiratory syndrome coro | .................... | .................... | .................... | .................... | .................... | .................... |
| 27778 | MT642257.1 | Severe acute respiratory syndrome coro | .................... | .................... | .................... | .................... | .................... | .................... |
| 27779 | MT642258.1 | Severe acute respiratory syndrome coro | .................... | .................... | .................... | .................... | .................... | .................... |
| 27780 | MT642259.1 | Severe acute respiratory syndrome coro | .................... | .................... | .................... | .................... | .................... | .................... |
| 27781 | MT642260.1 | Severe acute respiratory syndrome coro | .................... | .................... | .................... | .................... | .................... | .................... |
| 27782 | MT642261.1 | Severe acute respiratory syndrome coro | .................... | .................... | .................... | .................... | .................... | .................... |
| 27783 | MT642262.1 | Severe acute respiratory syndrome coro | .................... | .................... | .................... | .................... | .................... | .................... |
| 27784 | MT642263.1 | Severe acute respiratory syndrome coro | .................... | .................... | .................... | .................... | .................... | .................... |
| 27785 | MT642264.1 | Severe acute respiratory syndrome coro | .................... | .................... | .................... | .................... | .................... | .................... |
| 27786 | MT642265.1 | Severe acute respiratory syndrome coro | .................... | .................... | .................... | .................... | .................... | .................... |
| 27787 | MT642266.1 | Severe acute respiratory syndrome coro | .................... | .................... | .................... | .................... | .................... | .................... |
| 27788 | MT642267.1 | Severe acute respiratory syndrome coro | .................... | .................... | .................... | .................... | .................... | .................... |
| 27789 | MT642268.1 | Severe acute respiratory syndrome coro | .................... | .................... | .................... | .................... | .................... | .................... |
| 27790 | MT642269.1 | Severe acute respiratory syndrome coro | .................... | .................... | .................... | .................... | .................... | .................... |
| 27791 | MT642270.1 | Severe acute respiratory syndrome coro | .................... | .................... | .................... | .................... | .................... | .................... |
| 27792 | MT642271.1 | Severe acute respiratory syndrome coro | .................... | .................... | .................... | .................... | .................... | .................... |
| 27793 | MT642272.1 | Severe acute respiratory syndrome coro | .................... | .................... | .................... | .................... | .................... | .................... |
| 27794 | MT642273.1 | Severe acute respiratory syndrome coro | .................... | .................... | .................... | .................... | .................... | .................... |
| 27795 | MT642275.1 | Severe acute respiratory syndrome coro | .................... | .................... | .................... | .................... | .................... | .................... |
| 27796 | MT642276.1 | Severe acute respiratory syndrome coro | .................... | .................... | .................... | .................... | .................... | .................... |
| 27797 | MT642277.1 | Severe acute respiratory syndrome coro | .................... | .................... | .................... | .................... | .................... | .................... |
| 27798 | MT642278.1 | Severe acute respiratory syndrome coro | .................... | .................... | .................... | .................... | .................... | .................... |
| 27799 | MT642279.1 | Severe acute respiratory syndrome coro | .................... | .................... | .................... | .................... | .................... | .................... |
| 27800 | MT642280.1 | Severe acute respiratory syndrome coro | .................... | .................... | .................... | .................... | .................... | .................... |
| 27801 | MT642281.1 | Severe acute respiratory syndrome coro | .................... | .................... | .................... | .................... | .................... | .................... |
| 27802 | MT642282.1 | Severe acute respiratory syndrome coro | .................... | .................... | .................... | .................... | .................... | .................... |
| 27803 | MT642283.1 | Severe acute respiratory syndrome coro | .................... | .................... | .................... | .................... | .................... | .................... |
| 27804 | MT642284.1 | Severe acute respiratory syndrome coro | .................... | .................... | .................... | .................... | .................... | .................... |
| 27805 | MT642285.1 | Severe acute respiratory syndrome coro | .................... | .................... | .................... | .................... | .................... | .................... |
| 27806 | MT642286.1 | Severe acute respiratory syndrome coro | .................... | .................... | .................... | .................... | .................... | .................... |
| 27807 | MT642288.1 | Severe acute respiratory syndrome coro | .................... | .................... | .................... | .................... | .................... | .................... |
| 27808 | MT642289.1 | Severe acute respiratory syndrome coro | .................... | .................... | .................... | .................... | .................... | .................... |
| 27809 | MT642290.1 | Severe acute respiratory syndrome coro | .................... | .................... | .................... | .................... | .................... | .................... |
| 27810 | MT642291.1 | Severe acute respiratory syndrome coro | .................... | .................... | .................... | .................... | .................... | .................... |
| 27811 | MT642292.1 | Severe acute respiratory syndrome coro | .................... | .................... | .................... | .................... | .................... | .................... |
| 27812 | MT642293.1 | Severe acute respiratory syndrome coro | .................... | .................... | .................... | .................... | .................... | .................... |
| 27813 | MT642294.1 | Severe acute respiratory syndrome coro | .................... | .................... | .................... | .................... | .................... | .................... |
| 27814 | MT642295.1 | Severe acute respiratory syndrome coro | .................... | .................... | .................... | .................... | .................... | .................... |
| 27815 | MT642296.1 | Severe acute respiratory syndrome coro | .................... | .................... | .................... | .................... | .................... | .................... |
| 27816 | MT642297.1 | Severe acute respiratory syndrome coro | .................... | .................... | .................... | .................... | .................... | .................... |
| 27817 | MT642298.1 | Severe acute respiratory syndrome coro | .................... | .................... | .................... | .................... | .................... | .................... |
| 27818 | MT642299.1 | Severe acute respiratory syndrome coro | .................... | .................... | .................... | .................... | .................... | .................... |
| 27819 | MT642300.1 | Severe acute respiratory syndrome coro | .................... | .................... | .................... | .................... | .................... | .................... |
| 27820 | MT642301.1 | Severe acute respiratory syndrome coro | .................... | .................... | .................... | .................... | .................... | .................... |
| 27821 | MT642303.1 | Severe acute respiratory syndrome coro | .................... | .................... | .................... | .................... | .................... | .................... |
| 27822 | MT642304.1 | Severe acute respiratory syndrome coro | .................... | .................... | .................... | .................... | .................... | .................... |

```
27823 MT642305.1 |Severe acute respiratory syndrome coro    .................... .................... ..................... .................... .................... ....................
27824 MT642306.1 |Severe acute respiratory syndrome coro    .................... .................... ..................... .................... .................... ....................
27825 MT642307.1 |Severe acute respiratory syndrome coro    .................... .................... ..................... .................... .................... ....................
27826 MT642308.1 |Severe acute respiratory syndrome coro    .................... .................... ..................... .................... .................... ....................
27827 MT642309.1 |Severe acute respiratory syndrome coro    .................... .................... ..................... .................... .................... ....................
27828 MT642310.1 |Severe acute respiratory syndrome coro    .................... .................... ..................... .................... .................... ....................
27829 MT642311.1 |Severe acute respiratory syndrome coro    .................... .................... ..................... .................... .................... ....................
27830 MT642312.1 |Severe acute respiratory syndrome coro    .................... .................... ..................... .................... .................... ....................
27831 MT642313.1 |Severe acute respiratory syndrome coro    .................... .................... ..................... .................... .................... ....................
27832 MT642314.1 |Severe acute respiratory syndrome coro    .................... .................... ..................... .................... .................... ....................
27833 MT642315.1 |Severe acute respiratory syndrome coro    .................... .................... ..................... .................... .................... ....................
27834 MT642317.1 |Severe acute respiratory syndrome coro    .................... .................... ..................... .................... .................... ....................
27835 MT642318.1 |Severe acute respiratory syndrome coro    .................... .................... ..................... .................... .................... ....................
27836 MT642319.1 |Severe acute respiratory syndrome coro    .................... .................... ..................... .................... .................... ....................
27837 MT642320.1 |Severe acute respiratory syndrome coro    .................... .................... ..................... .................... .................... ....................
27838 MT642321.1 |Severe acute respiratory syndrome coro    .................... .................... ..................... .................... .................... ....................
27839 MT642323.1 |Severe acute respiratory syndrome coro    .................... .................... ..................... .................... .................... ....................
27840 MT642324.1 |Severe acute respiratory syndrome coro    .................... .................... ..................... .................... .................... ....................
27841 MT642325.1 |Severe acute respiratory syndrome coro    .................... .................... ..................... .................... .................... ....................
27842 MT642326.1 |Severe acute respiratory syndrome coro    .................... .................... ..................... .................... .................... ....................
27843 MT642328.1 |Severe acute respiratory syndrome coro    .................... .................... ..................... .................... .................... ....................
27844 MT642329.1 |Severe acute respiratory syndrome coro    .................... .................... ..................... .................... .................... ....................
27845 MT642330.1 |Severe acute respiratory syndrome coro    .................... .................... ..................... .................... .................... ....................
27846 MT642331.1 |Severe acute respiratory syndrome coro    .................... .................... ..................... .................... .................... ....................
27847 MT642332.1 |Severe acute respiratory syndrome coro    .................... .................... ..................... .................... .................... ....................
27848 MT642333.1 |Severe acute respiratory syndrome coro    .................... .................... ..................... .................... .................... ....................
27849 MT642334.1 |Severe acute respiratory syndrome coro    .................... .................... ..................... .................... .................... ....................
27850 MT642336.1 |Severe acute respiratory syndrome coro    .................... .................... ..................... .................... .................... ....................
27851 MT642337.1 |Severe acute respiratory syndrome coro    .................... .................... ..................... .................... .................... ....................
27852 MT642338.1 |Severe acute respiratory syndrome coro    .................... .................... ..................... .................... .................... ....................
27853 MT642339.1 |Severe acute respiratory syndrome coro    .................... .................... ..................... .................... .................... ....................
27854 MT642340.1 |Severe acute respiratory syndrome coro    .................... .................... ..................... .................... .................... ....................
27855 MT642341.1 |Severe acute respiratory syndrome coro    .................... .................... ..................... .................... .................... ....................
27856 MT642342.1 |Severe acute respiratory syndrome coro    .................... .................... ..................... .................... .................... ....................
27857 MT642343.1 |Severe acute respiratory syndrome coro    .................... .................... ..................... .................... .................... ....................
27858 MT642344.1 |Severe acute respiratory syndrome coro    .................... .................... ..................... .................... .................... ....................
27859 MT642345.1 |Severe acute respiratory syndrome coro    .................... .................... ..................... .................... .................... ....................
27860 MT642346.1 |Severe acute respiratory syndrome coro    .................... .................... ..................... .................... .................... ....................
27861 MT642347.1 |Severe acute respiratory syndrome coro    .................... .................... ..................... .................... .................... ....................
27862 MT642348.1 |Severe acute respiratory syndrome coro    .................... .................... ..................... .................... .................... ....................
27863 MT642349.1 |Severe acute respiratory syndrome coro    .................... .................... ..................... .................... .................... ....................
27864 MT642350.1 |Severe acute respiratory syndrome coro    .................... .................... ..................... .................... .................... ....................
27865 MT642351.1 |Severe acute respiratory syndrome coro    .................... .................... ..................... .................... .................... ....................
27866 MT642352.1 |Severe acute respiratory syndrome coro    .................... .................... ..................... .................... .................... ....................
27867 MT642353.1 |Severe acute respiratory syndrome coro    .................... .................... ..................... .................... .................... ....................
27868 MT642357.1 |Severe acute respiratory syndrome coro    .................... .................... ..................... .................... .................... ....................
27869 MT642358.1 |Severe acute respiratory syndrome coro    .................... .................... ..................... .................... .................... ....................
27870 MT642359.1 |Severe acute respiratory syndrome coro    .................... .................... ..................... .................... .................... ....................
27871 MT642360.1 |Severe acute respiratory syndrome coro    .................... .................... ..................... .................... .................... ....................
27872 MT642361.1 |Severe acute respiratory syndrome coro    .................... .................... ..................... .................... .................... ....................
27873 MT642362.1 |Severe acute respiratory syndrome coro    .................... .................... ..................... .................... .................... ....................
27874 MT642363.1 |Severe acute respiratory syndrome coro    .................... .................... ..................... .................... .................... ....................
27875 MT642364.1 |Severe acute respiratory syndrome coro    .................... .................... ..................... .................... .................... ....................
27876 MT642365.1 |Severe acute respiratory syndrome coro    .................... .................... ..................... .................... .................... ....................
27877 MT642366.1 |Severe acute respiratory syndrome coro    .................... .................... ..................... .................... .................... ....................
27878 MT642367.1 |Severe acute respiratory syndrome coro    .................... .................... ..................... .................... .................... ....................
27879 MT642369.1 |Severe acute respiratory syndrome coro    .................... .................... ..................... .................... .................... ....................
27880 MT642370.1 |Severe acute respiratory syndrome coro    .................... .................... ..................... .................... .................... ....................
27881 MT642371.1 |Severe acute respiratory syndrome coro    .................... .................... ..................... .................... .................... ....................
27882 MT642372.1 |Severe acute respiratory syndrome coro    .................... .................... ..................... .................... .................... ....................
27883 MT642373.1 |Severe acute respiratory syndrome coro    .................... .................... ..................... .................... .................... ....................
27884 MT642374.1 |Severe acute respiratory syndrome coro    .................... .................... ..................... .................... .................... ....................
27885 MT642375.1 |Severe acute respiratory syndrome coro    .................... .................... ..................... .................... .................... ....................
27886 MT642377.1 |Severe acute respiratory syndrome coro    .................... .................... ..................... .................... .................... ....................
27887 MT642378.1 |Severe acute respiratory syndrome coro    .................... .................... ..................... .................... .................... ....................
27888 MT642379.1 |Severe acute respiratory syndrome coro    .................... .................... ..................... .................... .................... ....................
27889 MT642381.1 |Severe acute respiratory syndrome coro    .................... .................... ..................... .................... .................... ....................
27890 MT642382.1 |Severe acute respiratory syndrome coro    .................... .................... ..................... .................... .................... ....................
27891 MT642383.1 |Severe acute respiratory syndrome coro    .................... .................... ..................... .................... .................... ....................
27892 MT642384.1 |Severe acute respiratory syndrome coro    .................... .................... ..................... .................... .................... ....................
27893 MT642385.1 |Severe acute respiratory syndrome coro    .................... .................... ..................... .................... .................... ....................
27894 MT642387.1 |Severe acute respiratory syndrome coro    .................... .................... ..................... .................... .................... ....................
27895 MT642388.1 |Severe acute respiratory syndrome coro    .................... .................... ..................... .................... .................... ....................
27896 MT642389.1 |Severe acute respiratory syndrome coro    .................... .................... ..................... .................... .................... ....................
27897 MT642391.1 |Severe acute respiratory syndrome coro    .................... .................... ..................... .................... .................... ....................
27898 MT642392.1 |Severe acute respiratory syndrome coro    .................... .................... ..................... .................... .................... ....................
27899 MT642393.1 |Severe acute respiratory syndrome coro    .................... .................... ..................... .................... .................... ....................
27900 MT642394.1 |Severe acute respiratory syndrome coro    .................... .................... ..................... .................... .................... ....................
```

```
27901 MT642396.1 |Severe acute respiratory syndrome coro    ····················  ····················  ····················  ····················  ····················  ····················
27902 MT642397.1 |Severe acute respiratory syndrome coro    ····················  ····················  ····················  ····················  ····················  ····················
27903 MT642398.1 |Severe acute respiratory syndrome coro    ····················  ····················  ····················  ····················  ····················  ····················
27904 MT642399.1 |Severe acute respiratory syndrome coro    ····················  ····················  ····················  ····················  ····················  ····················
27905 MT642400.1 |Severe acute respiratory syndrome coro    ····················  ····················  ····················  ····················  ····················  ····················
27906 MT642401.1 |Severe acute respiratory syndrome coro    ····················  ····················  ····················  ····················  ····················  ····················
27907 MT642402.1 |Severe acute respiratory syndrome coro    ····················  ····················  ····················  ····················  ····················  ····················
27908 MT642403.1 |Severe acute respiratory syndrome coro    ····················  ····················  ····················  ····················  ····················  ····················
27909 MT642404.1 |Severe acute respiratory syndrome coro    ····················  ····················  ····················  ····················  ····················  ····················
27910 MT642405.1 |Severe acute respiratory syndrome coro    ····················  ····················  ····················  ····················  ····················  ····················
27911 MT642406.1 |Severe acute respiratory syndrome coro    ····················  ····················  ····················  ····················  ····················  ····················
27912 MT642407.1 |Severe acute respiratory syndrome coro    ····················  ····················  ····················  ····················  ····················  ····················
27913 MT642408.1 |Severe acute respiratory syndrome coro    ····················  ····················  ····················  ····················  ····················  ····················
27914 MT642409.1 |Severe acute respiratory syndrome coro    ····················  ····················  ····················  ····················  ····················  ····················
27915 MT642410.1 |Severe acute respiratory syndrome coro    ····················  ····················  ····················  ····················  ····················  ····················
27916 MT642411.1 |Severe acute respiratory syndrome coro    ····················  ····················  ····················  ····················  ····················  ····················
27917 MT642412.1 |Severe acute respiratory syndrome coro    ····················  ····················  ····················  ····················  ····················  ····················
27918 MT642413.1 |Severe acute respiratory syndrome coro    ····················  ····················  ····················  ····················  ····················  ····················
27919 MT642414.1 |Severe acute respiratory syndrome coro    ····················  ····················  ····················  ····················  ····················  ····················
27920 MT642415.1 |Severe acute respiratory syndrome coro    ····················  ····················  ····················  ····················  ····················  ····················
27921 MT642418.1 |Severe acute respiratory syndrome coro    ····················  ····················  ····················  ····················  ····················  ····················
27922 MT642420.1 |Severe acute respiratory syndrome coro    ····················  ····················  ····················  ····················  ····················  ····················
27923 MT642421.1 |Severe acute respiratory syndrome coro    ····················  ····················  ····················  ····················  ····················  ····················
27924 MT642422.1 |Severe acute respiratory syndrome coro    ····················  ····················  ····················  ····················  ····················  ····················
27925 MT642423.1 |Severe acute respiratory syndrome coro    ····················  ····················  ····················  ····················  ····················  ····················
27926 MT642424.1 |Severe acute respiratory syndrome coro    ····················  ····················  ····················  ····················  ····················  ····················
27927 MT642425.1 |Severe acute respiratory syndrome coro    ····················  ····················  ····················  ····················  ····················  ····················
27928 MT642426.1 |Severe acute respiratory syndrome coro    ····················  ····················  ····················  ····················  ····················  ····················
27929 MT642427.1 |Severe acute respiratory syndrome coro    ····················  ····················  ····················  ····················  ····················  ····················
27930 MT642428.1 |Severe acute respiratory syndrome coro    ····················  ····················  ····················  ····················  ····················  ····················
27931 MT642429.1 |Severe acute respiratory syndrome coro    ····················  ····················  ····················  ····················  ····················  ····················
27932 MT642430.1 |Severe acute respiratory syndrome coro    ····················  ····················  ····················  ····················  ····················  ····················
27933 MT642431.1 |Severe acute respiratory syndrome coro    ····················  ····················  ····················  ····················  ····················  ····················
27934 MT642432.1 |Severe acute respiratory syndrome coro    ····················  ····················  ····················  ····················  ····················  ····················
27935 MT642433.1 |Severe acute respiratory syndrome coro    ····················  ····················  ····················  ····················  ····················  ····················
27936 MT642434.1 |Severe acute respiratory syndrome coro    ····················  ····················  ····················  ····················  ····················  ····················
27937 MT642435.1 |Severe acute respiratory syndrome coro    ····················  ····················  ····················  ····················  ····················  ····················
27938 MT644268.1 |Severe acute respiratory syndrome coro    ····················  ····················  ····················  ····················  ····················  ····················
27939 MT635255.1 |Severe acute respiratory syndrome coro    ····················  ····················  ····················  ····················  ····················  ····················
27940 MT635269.1 |Severe acute respiratory syndrome coro    ····················  ····················  ····················  ····················  ····················  ··A·················
27941 MT635270.1 |Severe acute respiratory syndrome coro    ····················  ·A··················  ····················  ····················  ····················  ··A·················
27942 MT635271.1 |Severe acute respiratory syndrome coro    ····················  ····················  ····················  ····················  ····················  ····················
27943 MT635272.1 |Severe acute respiratory syndrome coro    ····················  ·A··················  ····················  ····················  ····················  ····················
27944 MT635328.1 |Severe acute respiratory syndrome coro    ····················  ····················  ····················  ····················  ····················  ····················
27945 MT635339.1 |Severe acute respiratory syndrome coro    ····················  ····················  ····················  ····················  ····················  ····················
27946 MT635391.1 |Severe acute respiratory syndrome coro    ····················  ····················  ····················  ····················  ····················  ····················
27947 MT635392.1 |Severe acute respiratory syndrome coro    ····················  ····················  ····················  ····················  ····················  ··A·················
27948 MT635393.1 |Severe acute respiratory syndrome coro    ····················  ····················  ····················  ····················  ····················  ····················
27949 MT635397.1 |Severe acute respiratory syndrome coro    ····················  ····················  ····················  ····················  ····················  ····················
27950 MT635403.1 |Severe acute respiratory syndrome coro    ····················  ····················  ····················  ····················  ····················  ····················
27951 MT635404.1 |Severe acute respiratory syndrome coro    ····················  ····················  ····················  ····················  ····················  ····················
27952 MT635405.1 |Severe acute respiratory syndrome coro    ····················  ····················  ····················  ····················  ····················  ····················
27953 MT635406.1 |Severe acute respiratory syndrome coro    ····················  ····················  ····················  ····················  ····················  ····················
27954 MT635407.1 |Severe acute respiratory syndrome coro    ····················  ····················  ····················  ····················  ····················  ····················
27955 MT635408.1 |Severe acute respiratory syndrome coro    ····················  ····················  ····················  ····················  ····················  ····················
27956 MT635409.1 |Severe acute respiratory syndrome coro    ····················  ····················  ····················  ····················  ····················  ····················
27957 MT635410.1 |Severe acute respiratory syndrome coro    ····················  ····················  ····················  ····················  ····················  ····················
27958 MT635445.1 |Severe acute respiratory syndrome coro    ····················  ····················  ····················  ····················  ····················  ····················
27959 MT635641.1 |Severe acute respiratory syndrome coro    ····················  ····················  ····················  ····················  ····················  ····················
27960 MT635643.1 |Severe acute respiratory syndrome coro    ····················  ····················  ····················  ····················  ····················  ····················
27961 MT635645.1 |Severe acute respiratory syndrome coro    ····················  ····················  ····················  ····················  ····················  ····················
27962 MT635646.1 |Severe acute respiratory syndrome coro    ····················  ····················  ····················  ····················  ····················  ····················
27963 MT635647.1 |Severe acute respiratory syndrome coro    ····················  ····················  ····················  ····················  ····················  ····················
27964 MT635648.1 |Severe acute respiratory syndrome coro    ····················  ····················  ····················  ····················  ····················  ····················
27965 MT635649.1 |Severe acute respiratory syndrome coro    ····················  ····················  ····················  ····················  ····················  ····················
27966 MT635650.1 |Severe acute respiratory syndrome coro    ····················  ····················  ····················  ····················  ····················  ····················
27967 MT635651.1 |Severe acute respiratory syndrome coro    ····················  ····················  ····················  ····················  ····················  ····················
27968 MT635652.1 |Severe acute respiratory syndrome coro    ····················  ····················  ····················  ····················  ····················  ····················
27969 MT635653.1 |Severe acute respiratory syndrome coro    ····················  ····················  ····················  ····················  ····················  ····················
27970 MT635654.1 |Severe acute respiratory syndrome coro    ····················  ····················  ····················  ····················  ····················  ····················
27971 MT635655.1 |Severe acute respiratory syndrome coro    ····················  ····················  ····················  ····················  ····················  ····················
27972 MT635672.1 |Severe acute respiratory syndrome coro    ····················  ····················  ····················  ····················  ····················  ····················
27973 MT635855.1 |Severe acute respiratory syndrome coro    ····················  ·A··················  ····················  ····················  ····················  ··A·················
27974 MT635856.1 |Severe acute respiratory syndrome coro    ····················  ····················  ····················  ····················  ····················  ····················
27975 MT635857.1 |Severe acute respiratory syndrome coro    ····················  ·A··················  ····················  ····················  ····················  ··A·················
27976 MT635858.1 |Severe acute respiratory syndrome coro    ····················  ·A··················  ····················  ····················  ····················  ··A·················
27977 MT637143.1 |Severe acute respiratory syndrome coro    ····················  ····················  ····················  ····················  ····················  ····················
27978 MT627213.1 |Severe acute respiratory syndrome coro    ····················  ····················  ····················  ····················  ····················  ····················
```

```
27979 MT627216.1 |Severe acute respiratory syndrome coro    .................... .................... .................... .................... .................... ....................
27980 MT627217.1 |Severe acute respiratory syndrome coro    .................... .................... .................... .................... .................... ....................
27981 MT627219.1 |Severe acute respiratory syndrome coro    .................... .................... .................... .................... .................... ....................
27982 MT627221.1 |Severe acute respiratory syndrome coro    .................... .................... .................... .................... .................... ....................
27983 MT627223.1 |Severe acute respiratory syndrome coro    .................... .................... .................... .................... .................... ....................
27984 MT627225.1 |Severe acute respiratory syndrome coro    .................... .................... .................... .................... .................... ....................
27985 MT627226.1 |Severe acute respiratory syndrome coro    .................... .................... .................... .................... .................... ....................
27986 MT627227.1 |Severe acute respiratory syndrome coro    .................... .................... .................... .................... .................... ....................
27987 MT627228.1 |Severe acute respiratory syndrome coro    .................... .................... .................... .................... .................... ....................
27988 MT627229.1 |Severe acute respiratory syndrome coro    .................... .................... .................... .................... .................... ....................
27989 MT627231.1 |Severe acute respiratory syndrome coro    .................... .................... .................... .................... .................... ....................
27990 MT627232.1 |Severe acute respiratory syndrome coro    .................... .................... .................... .................... .................... ....................
27991 MT627233.1 |Severe acute respiratory syndrome coro    .................... .................... .................... .................... .................... ....................
27992 MT627234.1 |Severe acute respiratory syndrome coro    .................... .................... .................... .................... .................... ....................
27993 MT627235.1 |Severe acute respiratory syndrome coro    .................... .................... .................... .................... .................... ....................
27994 MT627236.1 |Severe acute respiratory syndrome coro    .................... .................... .................... .................... .................... ....................
27995 MT627237.1 |Severe acute respiratory syndrome coro    .................... .................... .................... .................... .................... ....................
27996 MT627238.1 |Severe acute respiratory syndrome coro    .................... .................... .................... .................... .................... ....................
27997 MT627240.1 |Severe acute respiratory syndrome coro    .................... .................... .................... .................... .................... ....................
27998 MT627241.1 |Severe acute respiratory syndrome coro    .................... .................... .................... .................... .................... ....................
27999 MT627242.1 |Severe acute respiratory syndrome coro    .................... .................... .................... .................... .................... ....................
28000 MT627243.1 |Severe acute respiratory syndrome coro    .................... .................... .................... .................... .................... ....................
28001 MT627244.1 |Severe acute respiratory syndrome coro    .................... .................... .................... .................... .................... ....................
28002 MT627245.1 |Severe acute respiratory syndrome coro    .................... .................... .................... .................... .................... ....................
28003 MT627246.1 |Severe acute respiratory syndrome coro    .................... .................... .................... .................... .................... ....................
28004 MT627247.1 |Severe acute respiratory syndrome coro    .................... .................... .................... .................... .................... ....................
28005 MT627248.1 |Severe acute respiratory syndrome coro    .................... .................... .................... .................... .................... ....................
28006 MT627249.1 |Severe acute respiratory syndrome coro    .................... .................... .................... .................... .................... ....................
28007 MT627250.1 |Severe acute respiratory syndrome coro    .................... .................... .................... .................... .................... ....................
28008 MT627251.1 |Severe acute respiratory syndrome coro    .................... .................... .................... .................... .................... ....................
28009 MT627252.1 |Severe acute respiratory syndrome coro    .................... .................... .................... .................... .................... ....................
28010 MT627253.1 |Severe acute respiratory syndrome coro    .................... .................... .................... .................... .................... ....................
28011 MT627254.1 |Severe acute respiratory syndrome coro    .................... .................... .................... .................... .................... ....................
28012 MT627255.1 |Severe acute respiratory syndrome coro    .................... .................... .................... .................... .................... ....................
28013 MT627256.1 |Severe acute respiratory syndrome coro    .................... .................... .................... .................... .................... ....................
28014 MT627257.1 |Severe acute respiratory syndrome coro    .................... .................... .................... .................... .................... ....................
28015 MT627258.1 |Severe acute respiratory syndrome coro    .................... .................... .................... .................... .................... ....................
28016 MT627259.1 |Severe acute respiratory syndrome coro    .................... .................... .................... .................... .................... ....................
28017 MT627260.1 |Severe acute respiratory syndrome coro    .................... .................... .................... .................... .................... ....................
28018 MT627261.1 |Severe acute respiratory syndrome coro    .................... .................... .................... .................... .................... ....................
28019 MT627262.1 |Severe acute respiratory syndrome coro    .................... .................... .................... .................... .................... ....................
28020 MT627263.1 |Severe acute respiratory syndrome coro    .................... .................... .................... .................... .................... ....................
28021 MT627264.1 |Severe acute respiratory syndrome coro    .................... .................... .................... .................... .................... ....................
28022 MT627265.1 |Severe acute respiratory syndrome coro    .................... .................... .................... .................... .................... ....................
28023 MT627266.1 |Severe acute respiratory syndrome coro    .................... .................... .................... .................... .................... ....................
28024 MT627267.1 |Severe acute respiratory syndrome coro    .................... .................... .................... .................... .................... ....................
28025 MT627268.1 |Severe acute respiratory syndrome coro    .................... .................... .................... .................... .................... ....................
28026 MT627269.1 |Severe acute respiratory syndrome coro    .................... .................... .................... .................... .................... ....................
28027 MT627270.1 |Severe acute respiratory syndrome coro    .................... .................... .................... .................... .................... ....................
28028 MT627271.1 |Severe acute respiratory syndrome coro    .................... .................... .................... .................... .................... ....................
28029 MT627272.1 |Severe acute respiratory syndrome coro    .................... .................... .................... .................... .................... ....................
28030 MT627273.1 |Severe acute respiratory syndrome coro    .................... .................... .................... .................... .................... ....................
28031 MT627274.1 |Severe acute respiratory syndrome coro    .................... .................... .................... .................... .................... ....................
28032 MT627275.1 |Severe acute respiratory syndrome coro    .................... .................... .................... .................... .................... ....................
28033 MT627276.1 |Severe acute respiratory syndrome coro    .................... .................... .................... .................... .................... ....................
28034 MT627277.1 |Severe acute respiratory syndrome coro    .................... .................... .................... .................... .................... ....................
28035 MT627278.1 |Severe acute respiratory syndrome coro    .................... .................... .................... .................... .................... ....................
28036 MT627279.1 |Severe acute respiratory syndrome coro    .................... .................... .................... .................... .................... ....................
28037 MT627280.1 |Severe acute respiratory syndrome coro    .................... .................... .................... .................... .................... ....................
28038 MT627281.1 |Severe acute respiratory syndrome coro    .................... .................... .................... .................... .................... ....................
28039 MT627282.1 |Severe acute respiratory syndrome coro    .................... .................... .................... .................... .................... ....................
28040 MT627283.1 |Severe acute respiratory syndrome coro    .................... .................... .................... .................... .................... ....................
28041 MT627284.1 |Severe acute respiratory syndrome coro    .................... .................... .................... .................... .................... ....................
28042 MT627285.1 |Severe acute respiratory syndrome coro    .................... .................... .................... .................... .................... ....................
28043 MT627286.1 |Severe acute respiratory syndrome coro    .................... .................... .................... .................... .................... ....................
28044 MT627287.1 |Severe acute respiratory syndrome coro    .................... .................... .................... .................... .................... ....................
28045 MT627288.1 |Severe acute respiratory syndrome coro    .................... .................... .................... .................... .................... ....................
28046 MT627289.1 |Severe acute respiratory syndrome coro    .................... .................... .................... .................... .................... ....................
28047 MT627290.1 |Severe acute respiratory syndrome coro    .................... .................... .................... .................... .................... ....................
28048 MT627291.1 |Severe acute respiratory syndrome coro    .................... .................... .................... .................... .................... ....................
28049 MT627292.1 |Severe acute respiratory syndrome coro    .................... .................... .................... .................... .................... ....................
28050 MT627293.1 |Severe acute respiratory syndrome coro    .................... .................... .................... .................... .................... ....................
28051 MT627294.1 |Severe acute respiratory syndrome coro    .................... .................... .................... .................... .................... ....................
28052 MT627295.1 |Severe acute respiratory syndrome coro    .................... .................... .................... .................... .................... ....................
28053 MT627296.1 |Severe acute respiratory syndrome coro    .................... .................... .................... .................... .................... ....................
28054 MT627297.1 |Severe acute respiratory syndrome coro    .................... .................... .................... .................... .................... ....................
28055 MT627298.1 |Severe acute respiratory syndrome coro    .................... .................... .................... .................... .................... ....................
28056 MT627299.1 |Severe acute respiratory syndrome coro    .................... .................... .................... .................... .................... ....................
```

28057 MT627300.1 |Severe acute respiratory syndrome coro ···················· ···················· ···················· ···················· ···················· ····················
28058 MT627302.1 |Severe acute respiratory syndrome coro ···················· ···················· ···················· ···················· ···················· ····················
28059 MT627303.1 |Severe acute respiratory syndrome coro ···················· ···················· ···················· ···················· ···················· ····················
28060 MT627304.1 |Severe acute respiratory syndrome coro ···················· ···················· ···················· ···················· ···················· ····················
28061 MT627305.1 |Severe acute respiratory syndrome coro ···················· ···················· ···················· ···················· ···················· ····················
28062 MT627306.1 |Severe acute respiratory syndrome coro ···················· ···················· ···················· ···················· ···················· ····················
28063 MT627307.1 |Severe acute respiratory syndrome coro ···················· ···················· ···················· ···················· ···················· ····················
28064 MT627308.1 |Severe acute respiratory syndrome coro ···················· ···················· ···················· ···················· ···················· ····················
28065 MT627309.1 |Severe acute respiratory syndrome coro ···················· ···················· ···················· ···················· ···················· ····················
28066 MT627310.1 |Severe acute respiratory syndrome coro ···················· ···················· ···················· ···················· ···················· ····················
28067 MT627311.1 |Severe acute respiratory syndrome coro ···················· ···················· ···················· ···················· ···················· ····················
28068 MT627312.1 |Severe acute respiratory syndrome coro ···················· ···················· ···················· ···················· ···················· ····················
28069 MT627314.1 |Severe acute respiratory syndrome coro ···················· ···················· ···················· ···················· ···················· ····················
28070 MT627315.1 |Severe acute respiratory syndrome coro ···················· ···················· ···················· ···················· ···················· ····················
28071 MT627316.1 |Severe acute respiratory syndrome coro ···················· ···················· ···················· ···················· ···················· ····················
28072 MT627317.1 |Severe acute respiratory syndrome coro ···················· ···················· ···················· ···················· ···················· ····················
28073 MT627318.1 |Severe acute respiratory syndrome coro ···················· ···················· ···················· ···················· ···················· ····················
28074 MT627325.1 |Severe acute respiratory syndrome coro ···················· ···················· ···················· ···················· ···················· ····················
28075 MT627407.1 |Severe acute respiratory syndrome coro ···················· ···················· ···················· ···················· ···················· ····················
28076 MT627408.1 |Severe acute respiratory syndrome coro ···················· ···················· ···················· ···················· ···················· ····················
28077 MT627409.1 |Severe acute respiratory syndrome coro ···················· ···················· ···················· ···················· ···················· ····················
28078 MT627410.1 |Severe acute respiratory syndrome coro ···················· ···················· ···················· ···················· ···················· ····················
28079 MT627411.1 |Severe acute respiratory syndrome coro ···················· ···················· ···················· ···················· ···················· ····················
28080 MT627412.1 |Severe acute respiratory syndrome coro ···················· ···················· ···················· ···················· ···················· ····················
28081 MT627413.1 |Severe acute respiratory syndrome coro ···················· ···················· ···················· ···················· ···················· ····················
28082 MT627414.1 |Severe acute respiratory syndrome coro ···················· ···················· ···················· ···················· ···················· ····················
28083 MT627415.1 |Severe acute respiratory syndrome coro ···················· ···················· ···················· ···················· ···················· ····················
28084 MT627416.1 |Severe acute respiratory syndrome coro ···················· ···················· ···················· ···················· ···················· ····················
28085 MT627417.1 |Severe acute respiratory syndrome coro ···················· ···················· ···················· ···················· ···················· ····················
28086 MT627418.1 |Severe acute respiratory syndrome coro ···················· ···················· ···················· ···················· ···················· ····················
28087 MT627419.1 |Severe acute respiratory syndrome coro ···················· ···················· ···················· ···················· ···················· ····················
28088 MT627420.1 |Severe acute respiratory syndrome coro ···················· ···················· ···················· ···················· ···················· ····················
28089 MT627421.1 |Severe acute respiratory syndrome coro ···················· ···················· ···················· ···················· ···················· ····················
28090 MT627422.1 |Severe acute respiratory syndrome coro ···················· ···················· ···················· ···················· ···················· ····················
28091 MT627423.1 |Severe acute respiratory syndrome coro ···················· ···················· ···················· ···················· ···················· ····················
28092 MT627424.1 |Severe acute respiratory syndrome coro ···················· ···················· ···················· ···················· ···················· ····················
28093 MT627425.1 |Severe acute respiratory syndrome coro ···················· ···················· ···················· ···················· ···················· ····················
28094 MT627426.1 |Severe acute respiratory syndrome coro ···················· ···················· ···················· ···················· ···················· ····················
28095 MT627427.1 |Severe acute respiratory syndrome coro ···················· ···················· ···················· ···················· ···················· ····················
28096 MT627428.1 |Severe acute respiratory syndrome coro ···················· ···················· ···················· ···················· ···················· ····················
28097 MT627429.1 |Severe acute respiratory syndrome coro ···················· ···················· ···················· ···················· ···················· ····················
28098 MT627430.1 |Severe acute respiratory syndrome coro ···················· ···················· ···················· ···················· ···················· ····················
28099 MT627431.1 |Severe acute respiratory syndrome coro ···················· ···················· ···················· ···················· ···················· ····················
28100 MT627432.1 |Severe acute respiratory syndrome coro ···················· ·A·················· ···················· ···················· ···················· ····················
28101 MT627433.1 |Severe acute respiratory syndrome coro ···················· ···················· ···················· ···················· ···················· ····················
28102 MT627434.1 |Severe acute respiratory syndrome coro ···················· ···················· ···················· ···················· ···················· ····················
28103 MT627435.1 |Severe acute respiratory syndrome coro ···················· ···················· ···················· ···················· ···················· ····················
28104 MT627436.1 |Severe acute respiratory syndrome coro ···················· ···················· ···················· ···················· ···················· ····················
28105 MT627437.1 |Severe acute respiratory syndrome coro ···················· ···················· ···················· ···················· ···················· ····················
28106 MT627607.1 |Severe acute respiratory syndrome coro ···················· ···················· ···················· ···················· ···················· ····················
28107 MT627609.1 |Severe acute respiratory syndrome coro ···················· ···················· ···················· ···················· ···················· ····················
28108 MT627612.1 |Severe acute respiratory syndrome coro ···················· ···················· ···················· ···················· ···················· ····················
28109 MT627613.1 |Severe acute respiratory syndrome coro ···················· ···················· ···················· ···················· ···················· ····················
28110 MT627614.1 |Severe acute respiratory syndrome coro ···················· ···················· ···················· ···················· ···················· ····················
28111 MT627615.1 |Severe acute respiratory syndrome coro ···················· ···················· ···················· ···················· ···················· ····················
28112 MT627616.1 |Severe acute respiratory syndrome coro ···················· ···················· ···················· ···················· ···················· ····················
28113 MT627618.1 |Severe acute respiratory syndrome coro ···················· ···················· ···················· ···················· ···················· ····················
28114 MT627619.1 |Severe acute respiratory syndrome coro ···················· ···················· ···················· A··················· ···················· ····················
28115 MT627620.1 |Severe acute respiratory syndrome coro ···················· ···················· ···················· ···················· ···················· ····················
28116 MT627621.1 |Severe acute respiratory syndrome coro ···················· ···················· ···················· ···················· ···················· ····················
28117 MT627622.1 |Severe acute respiratory syndrome coro ···················· ···················· ···················· ···················· ···················· ····················
28118 MT627623.1 |Severe acute respiratory syndrome coro ···················· ···················· ···················· ···················· ···················· ····················
28119 MT627624.1 |Severe acute respiratory syndrome coro ···················· ···················· ···················· ···················· ···················· ····················
28120 MT627625.1 |Severe acute respiratory syndrome coro ···················· ···················· ···················· ···················· ···················· ····················
28121 MT627626.1 |Severe acute respiratory syndrome coro ···················· ···················· ···················· ···················· ···················· ····················
28122 MT627627.1 |Severe acute respiratory syndrome coro ···················· ···················· ···················· ···················· ···················· ····················
28123 MT627628.1 |Severe acute respiratory syndrome coro ···················· ···················· ···················· ···················· ···················· ····················
28124 MT627629.1 |Severe acute respiratory syndrome coro ···················· ···················· ···················· ···················· ···················· ····················
28125 MT627630.1 |Severe acute respiratory syndrome coro ···················· ···················· ···················· ···················· ···················· ····················
28126 MT627631.1 |Severe acute respiratory syndrome coro ···················· ···················· ···················· ···················· ···················· ····················
28127 MT627632.1 |Severe acute respiratory syndrome coro ···················· ···················· ···················· ···················· ···················· ····················
28128 MT627633.1 |Severe acute respiratory syndrome coro ···················· ···················· ···················· ···················· ···················· ····················
28129 MT627634.1 |Severe acute respiratory syndrome coro ···················· ···················· ···················· ···················· ···················· ····················
28130 MT627635.1 |Severe acute respiratory syndrome coro ···················· ···················· ···················· ···················· ···················· ····················
28131 MT627636.1 |Severe acute respiratory syndrome coro ···················· ···················· ···················· ···················· ···················· ····················
28132 MT627637.1 |Severe acute respiratory syndrome coro ···················· ···················· ···················· ···················· ···················· ····················
28133 MT627638.1 |Severe acute respiratory syndrome coro ···················· ···················· ···················· ···················· ···················· ····················
28134 MT627745.1 |Severe acute respiratory syndrome coro ···················· ···················· ···················· ···················· ···················· ····················

```
28135 MT627746.1 |Severe acute respiratory syndrome coro    ....................  ....................  ....................  ....................  ....................  ....................
28136 MT627747.1 |Severe acute respiratory syndrome coro    ....................  ....................  ....................  ....................  ....................  ....................
28137 MT627748.1 |Severe acute respiratory syndrome coro    ....................  ....................  ....................  ....................  ....................  ....................
28138 MT627749.1 |Severe acute respiratory syndrome coro    ....................  ....................  ....................  ....................  ....................  ....................
28139 MT627750.1 |Severe acute respiratory syndrome coro    ....................  ....................  ....................  ....................  ....................  ....................
28140 MT627751.1 |Severe acute respiratory syndrome coro    ....................  ....................  ....................  ....................  ....................  ....................
28141 MT627752.1 |Severe acute respiratory syndrome coro    ....................  ....................  ....................  ....................  ....................  ....................
28142 MT627754.1 |Severe acute respiratory syndrome coro    ....................  ....................  ....................  ....................  ....................  ....................
28143 MT627755.1 |Severe acute respiratory syndrome coro    ....................  ....................  ....................  ....................  ....................  ....................
28144 MT627756.1 |Severe acute respiratory syndrome coro    ....................  ....................  ....................  ....................  ....................  ....................
28145 MT627757.1 |Severe acute respiratory syndrome coro    ....................  ....................  ....................  ....................  ....................  ....................
28146 MT627758.1 |Severe acute respiratory syndrome coro    ....................  ....................  ....................  ....................  ....................  ....................
28147 MT627759.1 |Severe acute respiratory syndrome coro    ....................  ....................  ....................  ....................  ....................  ....................
28148 MT627760.1 |Severe acute respiratory syndrome coro    ....................  ....................  ....................  ....................  ....................  ....................
28149 MT627761.1 |Severe acute respiratory syndrome coro    ....................  ....................  ....................  ....................  ....................  ....................
28150 MT627762.1 |Severe acute respiratory syndrome coro    ....................  ....................  ....................  ....................  ....................  ....................
28151 MT627763.1 |Severe acute respiratory syndrome coro    ....................  ....................  ....................  ....................  ....................  ....................
28152 MT627764.1 |Severe acute respiratory syndrome coro    ....................  ....................  ....................  ....................  ....................  ....................
28153 MT627765.1 |Severe acute respiratory syndrome coro    ....................  ....................  ....................  ....................  ....................  ....................
28154 MT627766.1 |Severe acute respiratory syndrome coro    ....................  ....................  ....................  ....................  ....................  ....................
28155 MT627767.1 |Severe acute respiratory syndrome coro    ....................  ....................  ....................  ....................  ....................  ....................
28156 MT627920.1 |Severe acute respiratory syndrome coro    ....................  ....................  ....................  ....................  ....................  ....................
28157 MT627921.1 |Severe acute respiratory syndrome coro    ....................  ....................  ....................  ....................  ....................  ....................
28158 MT627922.1 |Severe acute respiratory syndrome coro    ....................  ....................  ....................  ....................  ....................  ....................
28159 MT627923.1 |Severe acute respiratory syndrome coro    ....................  ....................  ....................  ....................  ....................  ....................
28160 MT627924.1 |Severe acute respiratory syndrome coro    ....................  ....................  ....................  ....................  ....................  ....................
28162 MT627926.1 |Severe acute respiratory syndrome coro    ....................  ....................  ....................  ....................  ....................  ....................
28163 MT627927.1 |Severe acute respiratory syndrome coro    ....................  ....................  ....................  ....................  ....................  ....................
28164 MT627928.1 |Severe acute respiratory syndrome coro    ....................  ....................  ....................  ....................  ....................  ....................
28165 MT627929.1 |Severe acute respiratory syndrome coro    ....................  ....................  ....................  ....................  ....................  ....................
28166 MT627930.1 |Severe acute respiratory syndrome coro    ....................  ....................  ....................  ....................  ....................  ....................
28167 MT627931.1 |Severe acute respiratory syndrome coro    ....................  ....................  ....................  ....................  ....................  ....................
28168 MT627932.1 |Severe acute respiratory syndrome coro    ....................  ....................  ....................  ....................  ....................  ....................
28169 MT628037.1 |Severe acute respiratory syndrome coro    ....................  ....................  ....................  ....................  ....................  ....................
28170 MT628038.1 |Severe acute respiratory syndrome coro    ....................  ....................  ....................  ....................  ....................  ....................
28171 MT628039.1 |Severe acute respiratory syndrome coro    ....................  ....................  ....................  ....................  ....................  ....................
28172 MT628040.1 |Severe acute respiratory syndrome coro    ....................  ....................  ....................  ....................  ....................  ....................
28173 MT628041.1 |Severe acute respiratory syndrome coro    ....................  ....................  ....................  ....................  ....................  ....................
28174 MT628042.1 |Severe acute respiratory syndrome coro    ....................  ....................  ....................  ....................  ....................  ....................
28175 MT628043.1 |Severe acute respiratory syndrome coro    ....................  ....................  ....................  ....................  ....................  ....................
28176 MT628044.1 |Severe acute respiratory syndrome coro    ....................  ....................  ....................  ....................  ....................  ....................
28177 MT628045.1 |Severe acute respiratory syndrome coro    ....................  ....................  ....................  ....................  ....................  ....................
28178 MT628046.1 |Severe acute respiratory syndrome coro    ....................  ....................  ....................  ....................  ....................  ....................
28179 MT628047.1 |Severe acute respiratory syndrome coro    ....................  ....................  ....................  ....................  ....................  ....................
28180 MT628048.1 |Severe acute respiratory syndrome coro    ....................  ....................  ....................  ....................  ....................  ....................
28181 MT628049.1 |Severe acute respiratory syndrome coro    ....................  ....................  ....................  ....................  ....................  ....................
28182 MT628050.1 |Severe acute respiratory syndrome coro    ....................  ....................  ....................  ....................  ....................  ....................
28183 MT628051.1 |Severe acute respiratory syndrome coro    ....................  ....................  ....................  ....................  ....................  ....................
28184 MT628052.1 |Severe acute respiratory syndrome coro    ....................  ....................  ....................  ....................  ....................  ....................
28185 MT628053.1 |Severe acute respiratory syndrome coro    ....................  ....................  ....................  ....................  ....................  ....................
28186 MT628054.1 |Severe acute respiratory syndrome coro    ....................  ....................  ....................  ....................  ....................  ....................
28187 MT628055.1 |Severe acute respiratory syndrome coro    ....................  ....................  ....................  ....................  ....................  ....................
28188 MT628056.1 |Severe acute respiratory syndrome coro    ....................  ....................  ....................  ....................  ....................  ....................
28189 MT628057.1 |Severe acute respiratory syndrome coro    ....................  ....................  ....................  ....................  ....................  ....................
28190 MT628058.1 |Severe acute respiratory syndrome coro    ....................  ....................  ....................  ....................  ....................  ....................
28191 MT628059.1 |Severe acute respiratory syndrome coro    ....................  ....................  ....................  ....................  ....................  ....................
28192 MT628060.1 |Severe acute respiratory syndrome coro    ....................  ....................  ....................  ....................  ....................  ....................
28193 MT628061.1 |Severe acute respiratory syndrome coro    ....................  ....................  ....................  ....................  ....................  ....................
28194 MT628062.1 |Severe acute respiratory syndrome coro    ....................  ....................  ....................  ....................  ....................  ....................
28195 MT628063.1 |Severe acute respiratory syndrome coro    ....................  ....................  ....................  ....................  ....................  ....................
28196 MT628064.1 |Severe acute respiratory syndrome coro    ....................  ....................  ....................  ....................  ....................  ....................
28197 MT628065.1 |Severe acute respiratory syndrome coro    ....................  ....................  ....................  ....................  ....................  ....................
28198 MT628066.1 |Severe acute respiratory syndrome coro    ....................  ....................  ....................  ....................  ....................  ....................
28199 MT628067.1 |Severe acute respiratory syndrome coro    ....................  ....................  ....................  ....................  ....................  ....................
28200 MT628068.1 |Severe acute respiratory syndrome coro    ....................  ....................  ....................  ....................  ....................  ....................
28201 MT628069.1 |Severe acute respiratory syndrome coro    ....................  ....................  ....................  ....................  ....................  ....................
28202 MT628070.1 |Severe acute respiratory syndrome coro    ....................  ....................  ....................  ....................  ....................  ....................
28203 MT628071.1 |Severe acute respiratory syndrome coro    ....................  ....................  ....................  ....................  ....................  ....................
28204 MT628073.1 |Severe acute respiratory syndrome coro    ....................  ....................  ....................  ....................  ....................  ....................
28205 MT628074.1 |Severe acute respiratory syndrome coro    ....................  ....................  ....................  ....................  ....................  ....................
28206 MT628075.1 |Severe acute respiratory syndrome coro    ....................  ....................  ....................  ....................  ....................  ....................
28207 MT628076.1 |Severe acute respiratory syndrome coro    ....................  ....................  ....................  ....................  ....................  ....................
28208 MT628077.1 |Severe acute respiratory syndrome coro    ....................  ....................  ....................  ....................  ....................  ....................
28209 MT628078.1 |Severe acute respiratory syndrome coro    ....................  ....................  ....................  ....................  ....................  ....................
28210 MT628079.1 |Severe acute respiratory syndrome coro    ....................  ....................  ....................  ....................  ....................  ....................
28211 MT628080.1 |Severe acute respiratory syndrome coro    ....................  ....................  ....................  ....................  ....................  ....................
28212 MT628081.1 |Severe acute respiratory syndrome coro    ....................  ....................  ....................  ....................  ....................  ....................
28213 MT628082.1 |Severe acute respiratory syndrome coro    ....................  ....................  ....................  ....................  ....................  ....................
```

28214 MT628084.1 |Severe acute respiratory syndrome coro ···················· ···················· ····················· ···················· ···················· ····················
28215 MT628085.1 |Severe acute respiratory syndrome coro ···················· ···················· ····················· ···················· ···················· ····················
28216 MT628087.1 |Severe acute respiratory syndrome coro ···················· ···················· ····················· ···················· ···················· ····················
28217 MT628088.1 |Severe acute respiratory syndrome coro ···················· ···················· ····················· ···················· ···················· ····················
28218 MT628089.1 |Severe acute respiratory syndrome coro ···················· ···················· ····················· ···················· ···················· ····················
28219 MT628090.1 |Severe acute respiratory syndrome coro ···················· ···················· ····················· ···················· ···················· ····················
28220 MT628091.1 |Severe acute respiratory syndrome coro ···················· ···················· ····················· ···················· ···················· ····················
28221 MT628092.1 |Severe acute respiratory syndrome coro ···················· ···················· ····················· ···················· ···················· ····················
28222 MT628093.1 |Severe acute respiratory syndrome coro ···················· ···················· ····················· ···················· ···················· ····················
28223 MT628095.1 |Severe acute respiratory syndrome coro ···················· ···················· ····················· ···················· ···················· ····················
28224 MT628097.1 |Severe acute respiratory syndrome coro ···················· ···················· ··········T········· ···················· ···················· ····················
28225 MT628098.1 |Severe acute respiratory syndrome coro ···················· ···················· ····················· ···················· ···················· ····················
28226 MT628100.1 |Severe acute respiratory syndrome coro ···················· ···················· ····················· ···················· ···················· ····················
28227 MT628101.1 |Severe acute respiratory syndrome coro ···················· ···················· ····················· ···················· ···················· ····················
28228 MT628102.1 |Severe acute respiratory syndrome coro ···················· ···················· ····················· ···················· ···················· ····················
28229 MT628104.1 |Severe acute respiratory syndrome coro ···················· ···················· ····················· ···················· ···················· ····················
28230 MT628105.1 |Severe acute respiratory syndrome coro ···················· ···················· ····················· ···················· ···················· ····················
28231 MT628110.1 |Severe acute respiratory syndrome coro ···················· ···················· ····················· ···················· ···················· ····················
28232 MT628113.1 |Severe acute respiratory syndrome coro ···················· ···················· ····················· ···················· ···················· ····················
28233 MT628116.1 |Severe acute respiratory syndrome coro ···R················ ···················· ····················· ···················· ···················· ····················
28234 MT628118.1 |Severe acute respiratory syndrome coro ···················· ···················· ····················· ···················· ···················· ····················
28235 MT628119.1 |Severe acute respiratory syndrome coro ···················· ···················· ····················· ···················· ···················· ····················
28236 MT628121.1 |Severe acute respiratory syndrome coro ···················· ···················· ····················· ···················· ···················· ····················
28237 MT628123.1 |Severe acute respiratory syndrome coro ···················· ···················· ····················· ···················· ···················· ····················
28238 MT628124.1 |Severe acute respiratory syndrome coro ···················· ···················· ····················· ···················· ···················· ····················
28239 MT628125.1 |Severe acute respiratory syndrome coro ···················· ···················· ····················· ···················· ···················· ····················
28240 MT628126.1 |Severe acute respiratory syndrome coro ···················· ···················· ····················· ···················· ···················· ····················
28241 MT628127.1 |Severe acute respiratory syndrome coro ···················· ···················· ····················· ···················· ···················· ····················
28242 MT628128.1 |Severe acute respiratory syndrome coro ···················· ···················· ····················· ···················· ···················· ····················
28243 MT628129.1 |Severe acute respiratory syndrome coro ···················· ···················· ····················· ···················· ···················· ····················
28244 MT628130.1 |Severe acute respiratory syndrome coro ···················· ···················· ····················· ···················· ···················· ····················
28245 MT628131.1 |Severe acute respiratory syndrome coro ···················· ···················· ····················· ···················· ···················· ····················
28246 MT628132.1 |Severe acute respiratory syndrome coro ···················· ···················· ····················· ···················· ···················· ····················
28247 MT628133.1 |Severe acute respiratory syndrome coro ···················· ···················· ····················· ···················· ···················· ····················
28248 MT628134.1 |Severe acute respiratory syndrome coro ···················· ···················· ····················· ···················· ···················· ····················
28249 MT628135.1 |Severe acute respiratory syndrome coro ···················· ···················· ····················· ···················· ···················· ····················
28250 MT628136.1 |Severe acute respiratory syndrome coro ···················· ···················· ····················· ···················· ···················· ····················
28251 MT628137.1 |Severe acute respiratory syndrome coro ···················· ···················· ····················· ···················· ···················· ····················
28252 MT628138.1 |Severe acute respiratory syndrome coro ···················· ···················· ····················· ···················· ···················· ····················
28253 MT628139.1 |Severe acute respiratory syndrome coro ···················· ···················· ····················· ···················· ···················· ····················
28254 MT628140.1 |Severe acute respiratory syndrome coro ···················· ···················· ····················· ···················· ···················· ····················
28255 MT628141.1 |Severe acute respiratory syndrome coro ···················· ···················· ····················· ···················· ···················· ····················
28256 MT628142.1 |Severe acute respiratory syndrome coro ···················· ···················· ····················· ···················· ···················· ····················
28257 MT628143.1 |Severe acute respiratory syndrome coro ···················· ···················· ····················· ···················· ···················· ····················
28258 MT628145.1 |Severe acute respiratory syndrome coro ···················· ···················· ····················· ···················· ···················· ····················
28259 MT628146.1 |Severe acute respiratory syndrome coro ···················· ···················· ····················· ···················· ···················· ····················
28260 MT628147.1 |Severe acute respiratory syndrome coro ···················· ···················· ····················· ···················· ···················· ····················
28261 MT628148.1 |Severe acute respiratory syndrome coro ···················· ···················· ····················· ···················· ···················· ····················
28262 MT628149.1 |Severe acute respiratory syndrome coro ···················· ···················· ····················· ···················· ···················· ····················
28263 MT628150.1 |Severe acute respiratory syndrome coro ···················· ···················· ····················· ···················· ···················· ····················
28264 MT628152.1 |Severe acute respiratory syndrome coro ···················· ···················· ····················· ···················· ···················· ····················
28265 MT628153.1 |Severe acute respiratory syndrome coro ···················· ···················· ····················· ···················· ···················· ····················
28266 MT628154.1 |Severe acute respiratory syndrome coro ···················· ···················· ····················· ···················· ···················· ····················
28267 MT628155.1 |Severe acute respiratory syndrome coro ···················· ···················· ····················· ···················· ···················· ····················
28268 MT628156.1 |Severe acute respiratory syndrome coro ···················· ···················· ····················· ···················· ···················· ····················
28269 MT628157.1 |Severe acute respiratory syndrome coro ···················· ···················· ····················· ···················· ···················· ····················
28270 MT628158.1 |Severe acute respiratory syndrome coro ···················· ···················· ····················· ···················· ···················· ····················
28271 MT628160.1 |Severe acute respiratory syndrome coro ···················· ···················· ····················· ···················· ···················· ····················
28272 MT628163.1 |Severe acute respiratory syndrome coro ···················· ···················· ····················· ···················· ···················· ····················
28273 MT628164.1 |Severe acute respiratory syndrome coro ···················· ···················· ····················· ···················· ···················· ····················
28274 MT628165.1 |Severe acute respiratory syndrome coro ···················· ···················· ····················· ···················· ···················· ····················
28275 MT628166.1 |Severe acute respiratory syndrome coro ···················· ···················· ····················· ···················· ···················· ····················
28276 MT628167.1 |Severe acute respiratory syndrome coro ···················· ···················· ····················· ···················· ···················· ····················
28277 MT628169.1 |Severe acute respiratory syndrome coro ···················· ···················· ····················· ···················· ···················· ····················
28278 MT628170.1 |Severe acute respiratory syndrome coro ···················· ···················· ····················· ···················· ···················· ····················
28279 MT628171.1 |Severe acute respiratory syndrome coro ···················· ···················· ····················· ···················· ···················· ····················
28280 MT628172.1 |Severe acute respiratory syndrome coro ···················· ···················· ····················· ···················· ···················· ····················
28281 MT628173.1 |Severe acute respiratory syndrome coro ···················· ···················· ····················· ···················· ···················· ····················
28282 MT628174.1 |Severe acute respiratory syndrome coro ···················· ···················· ····················· ···················· ···················· ····················
28283 MT628175.1 |Severe acute respiratory syndrome coro ···················· ···················· ····················· ···················· ···················· ····················
28284 MT628176.1 |Severe acute respiratory syndrome coro ···················· ···················· ····················· ···················· ···················· ····················
28285 MT628177.1 |Severe acute respiratory syndrome coro ···················· ···················· ····················· ···················· ···················· ····················
28286 MT628178.1 |Severe acute respiratory syndrome coro ···················· ···················· ····················· ···················· ···················· ····················
28287 MT628180.1 |Severe acute respiratory syndrome coro ···················· ···················· ····················· ···················· ···················· ····················
28288 MT628181.1 |Severe acute respiratory syndrome coro ···················· ···················· ····················· ···················· ···················· ····················
28289 MT628184.1 |Severe acute respiratory syndrome coro ···················· ···················· ····················· ···················· ···················· ····················
28290 MT628186.1 |Severe acute respiratory syndrome coro ···················· ···················· ····················· ···················· ···················· ····················
28291 MT628187.1 |Severe acute respiratory syndrome coro ···················· ···················· ····················· ···················· ···················· ····················

| | | | | | | | | |
|---|---|---|---|---|---|---|---|---|
| 28292 | MT628190.1 | \|Severe acute respiratory syndrome coro | ···················· | ···················· | ····················· | ···················· | ···················· | ···················· |
| 28293 | MT628191.1 | \|Severe acute respiratory syndrome coro | ···················· | ···················· | ····················· | ···················· | ···················· | ···················· |
| 28294 | MT628192.1 | \|Severe acute respiratory syndrome coro | ···················· | ···················· | ····················· | ···················· | ···················· | ···················· |
| 28295 | MT628193.1 | \|Severe acute respiratory syndrome coro | ···················· | ···················· | ····················· | ···················· | ···················· | ···················· |
| 28296 | MT628194.1 | \|Severe acute respiratory syndrome coro | ···················· | ···················· | ····················· | ···················· | ···················· | ···················· |
| 28297 | MT628195.1 | \|Severe acute respiratory syndrome coro | ···················· | ···················· | ····················· | ···················· | ···················· | ···················· |
| 28298 | MT628196.1 | \|Severe acute respiratory syndrome coro | ···················· | ···················· | ····················· | ···················· | ···················· | ···················· |
| 28299 | MT628197.1 | \|Severe acute respiratory syndrome coro | ···················· | ···················· | ····················· | ···················· | ···················· | ···················· |
| 28300 | MT628198.1 | \|Severe acute respiratory syndrome coro | ···················· | ···················· | ····················· | ···················· | ···················· | ···················· |
| 28301 | MT628199.1 | \|Severe acute respiratory syndrome coro | ···················· | ···················· | ····················· | ···················· | ···················· | ···················· |
| 28302 | MT628200.1 | \|Severe acute respiratory syndrome coro | ···················· | ·A·················· | ····················· | ···················· | ···················· | ···················· |
| 28303 | MT628201.1 | \|Severe acute respiratory syndrome coro | ···················· | ···················· | ····················· | ···················· | ···················· | ···················· |
| 28304 | MT628202.1 | \|Severe acute respiratory syndrome coro | ···················· | ·A·················· | ····················· | ···················· | ···················· | ···················· |
| 28305 | MT628203.1 | \|Severe acute respiratory syndrome coro | ···················· | ···················· | ····················· | ···················· | ···················· | ···················· |
| 28306 | MT628204.1 | \|Severe acute respiratory syndrome coro | ···················· | ···················· | ····················· | ···················· | ···················· | ···················· |
| 28307 | MT628205.1 | \|Severe acute respiratory syndrome coro | ···················· | ·A·················· | ····················· | ···················· | ···················· | ···················· |
| 28308 | MT628206.1 | \|Severe acute respiratory syndrome coro | ···················· | ···················· | ····················· | ···················· | ···················· | ···················· |
| 28309 | MT628207.1 | \|Severe acute respiratory syndrome coro | ···················· | ···················· | ····················· | ···················· | ···················· | ···················· |
| 28310 | MT628208.1 | \|Severe acute respiratory syndrome coro | ···················· | ···················· | ····················· | ···················· | ···················· | ···················· |
| 28311 | MT628209.1 | \|Severe acute respiratory syndrome coro | ···················· | ···················· | ····················· | ···················· | ···················· | ···················· |
| 28312 | MT628210.1 | \|Severe acute respiratory syndrome coro | ···················· | ···················· | ····················· | ···················· | ···················· | ···················· |
| 28313 | MT628211.1 | \|Severe acute respiratory syndrome coro | ···················· | ···················· | ····················· | ···················· | ···················· | ···················· |
| 28314 | MT628212.1 | \|Severe acute respiratory syndrome coro | ···················· | ···················· | ····················· | ···················· | ···················· | ···················· |
| 28315 | MT628213.1 | \|Severe acute respiratory syndrome coro | ···················· | ···················· | ····················· | ···················· | ···················· | ···················· |
| 28316 | MT628214.1 | \|Severe acute respiratory syndrome coro | ···················· | ···················· | ····················· | ···················· | ···················· | ···················· |
| 28317 | MT628215.1 | \|Severe acute respiratory syndrome coro | ···················· | ···················· | ····················· | ···················· | ···················· | ···················· |
| 28318 | MT628216.1 | \|Severe acute respiratory syndrome coro | ···················· | ···················· | ····················· | ···················· | ···················· | ···················· |
| 28319 | MT628217.1 | \|Severe acute respiratory syndrome coro | ···················· | ···················· | ····················· | ···················· | ···················· | ···················· |
| 28320 | MT628218.1 | \|Severe acute respiratory syndrome coro | ···················· | ···················· | ····················· | ···················· | ···················· | ···················· |
| 28321 | MT628219.1 | \|Severe acute respiratory syndrome coro | ···················· | ···················· | ····················· | ···················· | ···················· | ···················· |
| 28322 | MT628220.1 | \|Severe acute respiratory syndrome coro | ···················· | ···················· | ····················· | ···················· | ···················· | ···················· |
| 28323 | MT628221.1 | \|Severe acute respiratory syndrome coro | ···················· | ···················· | ····················· | ···················· | ···················· | ···················· |
| 28324 | MT628222.1 | \|Severe acute respiratory syndrome coro | ···················· | ···················· | ····················· | ···················· | ···················· | ···················· |
| 28325 | MT628223.1 | \|Severe acute respiratory syndrome coro | ···················· | ···················· | ····················· | ···················· | ···················· | ···················· |
| 28326 | MT628224.1 | \|Severe acute respiratory syndrome coro | ···················· | ···················· | ····················· | ···················· | ···················· | ···················· |
| 28327 | MT628225.1 | \|Severe acute respiratory syndrome coro | ···················· | ···················· | ····················· | ···················· | ···················· | ···················· |
| 28328 | MT628227.1 | \|Severe acute respiratory syndrome coro | ···················· | ···················· | ····················· | ···················· | ···················· | ···················· |
| 28329 | MT628228.1 | \|Severe acute respiratory syndrome coro | ···················· | ···················· | ····················· | ···················· | ···················· | ···················· |
| 28330 | MT628230.1 | \|Severe acute respiratory syndrome coro | ···················· | ···················· | ····················· | ···················· | ···················· | ···················· |
| 28331 | MT628231.1 | \|Severe acute respiratory syndrome coro | ···················· | ···················· | ····················· | ···················· | ···················· | ···················· |
| 28332 | MT628232.1 | \|Severe acute respiratory syndrome coro | ···················· | ···················· | ····················· | ···················· | ···················· | ···················· |
| 28333 | MT628233.1 | \|Severe acute respiratory syndrome coro | ···················· | ···················· | ····················· | ···················· | ···················· | ···················· |
| 28334 | MT628234.1 | \|Severe acute respiratory syndrome coro | ···················· | ···················· | ····················· | ···················· | ···················· | ···················· |
| 28335 | MT628236.1 | \|Severe acute respiratory syndrome coro | ···················· | ···················· | ····················· | ···················· | ···················· | ···················· |
| 28336 | MT628238.1 | \|Severe acute respiratory syndrome coro | ···················· | ···················· | ····················· | ···················· | ···················· | ···················· |
| 28337 | MT628241.1 | \|Severe acute respiratory syndrome coro | ···················· | ···················· | ····················· | ···················· | ···················· | ···················· |
| 28338 | MT628242.1 | \|Severe acute respiratory syndrome coro | ···················· | ··············C····· | ····················· | ···················· | ···················· | ···················· |
| 28339 | MT628243.1 | \|Severe acute respiratory syndrome coro | ···················· | ···················· | ····················· | ···················· | ···················· | ···················· |
| 28340 | MT628244.1 | \|Severe acute respiratory syndrome coro | ···················· | ···················· | ····················· | ···················· | ···················· | ···················· |
| 28341 | MT628245.1 | \|Severe acute respiratory syndrome coro | ···················· | ···················· | ····················· | ···················· | ···················· | ···················· |
| 28342 | MT628246.1 | \|Severe acute respiratory syndrome coro | ···················· | ···················· | ····················· | ···················· | ···················· | ···················· |
| 28343 | MT628247.1 | \|Severe acute respiratory syndrome coro | ···················· | ···················· | ····················· | ···················· | ···················· | ···················· |
| 28344 | MT628248.1 | \|Severe acute respiratory syndrome coro | ···················· | ···················· | ····················· | ···················· | ···················· | ···················· |
| 28345 | MT628249.1 | \|Severe acute respiratory syndrome coro | ···················· | ···················· | ····················· | ···················· | ···················· | ···················· |
| 28346 | MT628250.1 | \|Severe acute respiratory syndrome coro | ···················· | ···················· | ····················· | ···················· | ···················· | ···················· |
| 28347 | MT628251.1 | \|Severe acute respiratory syndrome coro | ···················· | ···················· | ····················· | ···················· | ···················· | ···················· |
| 28348 | MT628252.1 | \|Severe acute respiratory syndrome coro | ···················· | ···················· | ····················· | ···················· | ···················· | ···················· |
| 28349 | MT628253.1 | \|Severe acute respiratory syndrome coro | ···················· | ···················· | ····················· | ···················· | ···················· | ···················· |
| 28350 | MT628254.1 | \|Severe acute respiratory syndrome coro | ···················· | ···················· | ····················· | ···················· | ···················· | ···················· |
| 28351 | MT628255.1 | \|Severe acute respiratory syndrome coro | ···················· | ···················· | ····················· | ···················· | ···················· | ···················· |
| 28352 | MT628256.1 | \|Severe acute respiratory syndrome coro | ···················· | ···················· | ····················· | ···················· | ···················· | ···················· |
| 28353 | MT628257.1 | \|Severe acute respiratory syndrome coro | ···················· | ···················· | ····················· | ···················· | ···················· | ···················· |
| 28354 | MT628258.1 | \|Severe acute respiratory syndrome coro | ···················· | ···················· | ····················· | ···················· | ···················· | ···················· |
| 28355 | MT628259.1 | \|Severe acute respiratory syndrome coro | ···················· | ···················· | ····················· | ···················· | ···················· | ···················· |
| 28356 | MT628260.1 | \|Severe acute respiratory syndrome coro | ···················· | ···················· | ····················· | ···················· | ···················· | ···················· |
| 28357 | MT628261.1 | \|Severe acute respiratory syndrome coro | ···················· | ···················· | ····················· | ···················· | ···················· | ···················· |
| 28358 | MT628262.1 | \|Severe acute respiratory syndrome coro | ···················· | ···················· | ····················· | ···················· | ···················· | ···················· |
| 28359 | MT628263.1 | \|Severe acute respiratory syndrome coro | ···················· | ···················· | ····················· | ···················· | ···················· | ···················· |
| 28360 | MT628264.1 | \|Severe acute respiratory syndrome coro | ···················· | ···················· | ····················· | ···················· | ···················· | ···················· |
| 28361 | MT628265.1 | \|Severe acute respiratory syndrome coro | ···················· | ···················· | ····················· | ···················· | ···················· | ···················· |
| 28362 | MT628266.1 | \|Severe acute respiratory syndrome coro | ···················· | ···················· | ····················· | ···················· | ···················· | ···················· |
| 28363 | MT628267.1 | \|Severe acute respiratory syndrome coro | ···················· | ···················· | ····················· | ···················· | ···················· | ···················· |
| 28364 | MT628268.1 | \|Severe acute respiratory syndrome coro | ···················· | ···················· | ····················· | ···················· | ···················· | ···················· |
| 28365 | MT628269.1 | \|Severe acute respiratory syndrome coro | ···················· | ···················· | ····················· | ···················· | ···················· | ···················· |
| 28366 | MT628270.1 | \|Severe acute respiratory syndrome coro | ···················· | ···················· | ····················· | ···················· | ···················· | ···················· |
| 28367 | MT628271.1 | \|Severe acute respiratory syndrome coro | ···················· | ···················· | ····················· | ···················· | ···················· | ···················· |
| 28368 | MT628272.1 | \|Severe acute respiratory syndrome coro | ···················· | ···················· | ····················· | ···················· | ···················· | ···················· |
| 28369 | MT628273.1 | \|Severe acute respiratory syndrome coro | ···················· | ···················· | ····················· | ···················· | ···················· | ···················· |

```
28370 MT628274.1 |Severe acute respiratory syndrome coro    ···················· ···················· ···················· ···················· ···················· ····················
28371 MT628275.1 |Severe acute respiratory syndrome coro    ···················· ···················· ···················· ···················· ···················· ····················
28372 MT628276.1 |Severe acute respiratory syndrome coro    ···················· ···················· ···················· ···················· ···················· ····················
28373 MT628279.1 |Severe acute respiratory syndrome coro    ···················· ·A·················· ···················· ···················· ···················· ····················
28374 MT628280.1 |Severe acute respiratory syndrome coro    ···················· ···················· ···················· ···················· ···················· ····················
28375 MT630421.1 |Severe acute respiratory syndrome coro    ···················· ·A·················· ···················· ···················· ···················· ··A·················
28376 MT630422.1 |Severe acute respiratory syndrome coro    ···················· ···················· ···················· ···················· ···················· ····················
28377 MT630423.1 |Severe acute respiratory syndrome coro    ···················· ···················· ···················· ···················· ···················· ····················
28378 MT630424.1 |Severe acute respiratory syndrome coro    ···················· ···················· ···················· ···················· ···················· ····················
28379 MT630425.1 |Severe acute respiratory syndrome coro    ···················· ···················· ···················· ···················· ···················· ····················
28380 MT630426.1 |Severe acute respiratory syndrome coro    ···················· ···················· ···················· ···················· ···················· ····················
28381 MT630427.1 |Severe acute respiratory syndrome coro    ···················· ···················· ···················· ···················· ···················· ····················
28382 MT630428.1 |Severe acute respiratory syndrome coro    ···················· ···················· ···················· ···················· ···················· ····················
28383 MT630429.1 |Severe acute respiratory syndrome coro    ···················· ···················· ···················· ···················· ···················· ····················
28384 MT630430.1 |Severe acute respiratory syndrome coro    ···················· ···················· ···················· ···················· ···················· ····················
28385 MT630431.1 |Severe acute respiratory syndrome coro    ···················· ···················· ···················· ···················· ···················· ····················
28386 MT630432.1 |Severe acute respiratory syndrome coro    ···················· ···················· ···················· ···················· ···················· ····················
28387 MT631784.1 |Severe acute respiratory syndrome coro    ···················· ···················· ···················· ···················· ···················· ····················
28388 MT631785.1 |Severe acute respiratory syndrome coro    ···················· ···················· ···················· ···················· ···················· ····················
28389 MT631786.1 |Severe acute respiratory syndrome coro    ···················· ···················· ···················· ···················· ···················· ····················
28390 MT631787.1 |Severe acute respiratory syndrome coro    ···················· ···················· ···················· ···················· ···················· ····················
28391 MT631788.1 |Severe acute respiratory syndrome coro    ···················· ···················· ···················· ···················· ···················· ····················
28392 MT631789.1 |Severe acute respiratory syndrome coro    ···················· ···················· ···················· ···················· ···················· ····················
28393 MT631790.1 |Severe acute respiratory syndrome coro    ···················· ···················· ···················· ···················· ···················· ····················
28394 MT631791.1 |Severe acute respiratory syndrome coro    ···················· ···················· ···················· ···················· ···················· ····················
28395 MT631792.1 |Severe acute respiratory syndrome coro    ···················· ···················· ···················· ···················· ···················· ····················
28396 MT631793.1 |Severe acute respiratory syndrome coro    ···················· ···················· ···················· ···················· ···················· ····················
28397 MT631794.1 |Severe acute respiratory syndrome coro    ···················· ···················· ···················· ···················· ···················· ····················
28398 MT631795.1 |Severe acute respiratory syndrome coro    ···················· ···················· ···················· ···················· ···················· ····················
28399 MT631797.1 |Severe acute respiratory syndrome coro    ···················· ···················· ···················· ···················· ···················· ····················
28400 MT631799.1 |Severe acute respiratory syndrome coro    ···················· ···················· ···················· ···················· ···················· ····················
28401 MT631800.1 |Severe acute respiratory syndrome coro    ···················· ···················· ···················· ···················· ···················· ····················
28402 MT631802.1 |Severe acute respiratory syndrome coro    ···················· ···················· ···················· ···················· ···················· ····················
28403 MT631803.1 |Severe acute respiratory syndrome coro    ···················· ···················· ···················· ···················· ···················· ····················
28404 MT631805.1 |Severe acute respiratory syndrome coro    ···················· ···················· ···················· ···················· ···················· ····················
28405 MT631807.1 |Severe acute respiratory syndrome coro    ···················· ···················· ···················· ···················· ···················· ····················
28406 MT631808.1 |Severe acute respiratory syndrome coro    ···················· ···················· ···················· ···················· ···················· ····················
28407 MT631809.1 |Severe acute respiratory syndrome coro    ···················· ···················· ···················· ···················· ···················· ····················
28408 MT631813.1 |Severe acute respiratory syndrome coro    ···················· ···················· ···················· ···················· ···················· ····················
28409 MT631815.1 |Severe acute respiratory syndrome coro    ···················· ···················· ···················· ···················· ···················· ····················
28410 MT631816.1 |Severe acute respiratory syndrome coro    ···················· ···················· ···················· ···················· ···················· ····················
28411 MT631817.1 |Severe acute respiratory syndrome coro    ···················· ···················· ···················· ···················· ···················· ····················
28412 MT631818.1 |Severe acute respiratory syndrome coro    ···················· ···················· ···················· ···················· ···················· ····················
28413 MT631819.1 |Severe acute respiratory syndrome coro    ···················· ···················· ···················· ···················· ···················· ····················
28414 MT631820.1 |Severe acute respiratory syndrome coro    ···················· ···················· ···················· ···················· ···················· ····················
28415 MT631821.1 |Severe acute respiratory syndrome coro    ···················· ···················· ···················· ···················· ···················· ····················
28416 MT631822.1 |Severe acute respiratory syndrome coro    ···················· ···················· ···················· ···················· ···················· ····················
28417 MT631823.1 |Severe acute respiratory syndrome coro    ···················· ···················· ···················· ···················· ···················· ····················
28418 MT631824.1 |Severe acute respiratory syndrome coro    ···················· ···················· ···················· ···················· ···················· ····················
28419 MT631825.1 |Severe acute respiratory syndrome coro    ···················· ···················· ···················· ···················· ···················· ····················
28420 MT631827.1 |Severe acute respiratory syndrome coro    ···················· ···················· ···················· ···················· ···················· ····················
28421 MT631828.1 |Severe acute respiratory syndrome coro    ···················· ···················· ···················· ···················· ···················· ····················
28422 MT631829.1 |Severe acute respiratory syndrome coro    ···················· ···················· ···················· ···················· ···················· ····················
28423 MT631830.1 |Severe acute respiratory syndrome coro    ···················· ···················· ···················· ···················· ···················· ····················
28424 MT631831.1 |Severe acute respiratory syndrome coro    ···················· ···················· ···················· ···················· ···················· ····················
28425 MT631833.1 |Severe acute respiratory syndrome coro    ···················· ···················· ···················· ···················· ···················· ····················
28426 MT631837.1 |Severe acute respiratory syndrome coro    ···················· ···················· ···················· ···················· ···················· ····················
28427 MT631838.1 |Severe acute respiratory syndrome coro    ···················· ···················· ···················· ···················· ···················· ····················
28428 MT631839.1 |Severe acute respiratory syndrome coro    ···················· ···················· ···················· ···················· ···················· ····················
28429 MT631840.1 |Severe acute respiratory syndrome coro    ···················· ···················· ···················· ···················· ···················· ····················
28430 MT631841.1 |Severe acute respiratory syndrome coro    ···················· ···················· ···················· ···················· ···················· ····················
28431 MT631842.1 |Severe acute respiratory syndrome coro    ···················· ···················· ···················· ···················· ···················· ····················
28432 MT632491.1 |Severe acute respiratory syndrome coro    ···················· ···················· ····C··············· ···················· ···················· ····················
28433 MT632493.1 |Severe acute respiratory syndrome coro    ···················· ···················· ···················· ···················· ···················· ····················
28434 MT632494.1 |Severe acute respiratory syndrome coro    ···················· ···················· ···················· ···················· ···················· ····················
28435 MT632495.1 |Severe acute respiratory syndrome coro    ···················· ···················· ···················· ···················· ···················· ····················
28436 MT632496.1 |Severe acute respiratory syndrome coro    ···················· ···················· ···················· ···················· ···················· ····················
28437 MT632497.1 |Severe acute respiratory syndrome coro    ···················· ···················· ···················· ···················· ···················· ····················
28438 MT632498.1 |Severe acute respiratory syndrome coro    ···················· ···················· ···················· ···················· ···················· ····················
28439 MT632499.1 |Severe acute respiratory syndrome coro    ···················· ···················· ···················· ···················· ···················· ····················
28440 MT632500.1 |Severe acute respiratory syndrome coro    ···················· ···················· ···················· ···················· ···················· ····················
28441 MT632501.1 |Severe acute respiratory syndrome coro    ···················· ···················· ···················· ···················· ···················· ····················
28442 MT632502.1 |Severe acute respiratory syndrome coro    ···················· ···················· ···················· ···················· ···················· ····················
28443 MT632503.1 |Severe acute respiratory syndrome coro    ···················· ···················· ···················· ···················· ···················· ····················
28444 MT632504.1 |Severe acute respiratory syndrome coro    ···················· ···················· ···················· ···················· ···················· ····················
28445 MT632505.1 |Severe acute respiratory syndrome coro    ···················· ···················· ···················· ···················· ···················· ····················
28446 MT632506.1 |Severe acute respiratory syndrome coro    ···················· ···················· ···················· ···················· ···················· ····················
28447 MT632507.1 |Severe acute respiratory syndrome coro    ···················· ···················· ···················· ···················· ···················· ····················
```

28448 MT632508.1 |Severe acute respiratory syndrome coro .................... .................... ..................... .................... .................... ....................
28449 MT632509.1 |Severe acute respiratory syndrome coro .................... .................... ..................... .................... .................... ....................
28450 MT632510.1 |Severe acute respiratory syndrome coro .................... .................... ..................... .................... .................... ....................
28451 MT632511.1 |Severe acute respiratory syndrome coro .................... .................... ..................... .................... .................... ....................
28452 MT632512.1 |Severe acute respiratory syndrome coro .................... .................... ..................... .................... .................... ....................
28453 MT632513.1 |Severe acute respiratory syndrome coro .................... .................... ..................... .................... .................... ....................
28454 MT632514.1 |Severe acute respiratory syndrome coro .................... .................... ..................... .................... .................... ....................
28455 MT632515.1 |Severe acute respiratory syndrome coro .................... .................... ..................... .................... .................... ....................
28456 MT632516.1 |Severe acute respiratory syndrome coro .................... .................... ..................... .................... .................... ....................
28457 MT632517.1 |Severe acute respiratory syndrome coro .................... .................... ..................... .................... .................... ....................
28458 MT632518.1 |Severe acute respiratory syndrome coro .................... .................... ..................... .................... .................... ....................
28459 MT632519.1 |Severe acute respiratory syndrome coro .................... .................... ..................... .................... .................... ....................
28460 MT632520.1 |Severe acute respiratory syndrome coro .................... .................... ..................... .................... .................... ....................
28461 MT632521.1 |Severe acute respiratory syndrome coro .................... .................... ..................... .................... .................... ....................
28462 MT632522.1 |Severe acute respiratory syndrome coro .................... .................... ..................... .................... .................... ....................
28463 MT632523.1 |Severe acute respiratory syndrome coro .................... .................... ..................... .................... .................... ....................
28464 MT632524.1 |Severe acute respiratory syndrome coro .................... .................... ..................... .................... .................... ....................
28465 MT632525.1 |Severe acute respiratory syndrome coro .................... .................... ..................... .................... .................... ....................
28466 MT632526.1 |Severe acute respiratory syndrome coro .................... .................... ..................... .................... .................... ....................
28467 MT632527.1 |Severe acute respiratory syndrome coro .................... .................... ..................... .................... .................... ....................
28468 MT632528.1 |Severe acute respiratory syndrome coro .................... .................... ..................... .................... .................... ....................
28469 MT632529.1 |Severe acute respiratory syndrome coro .................... .................... ..................... .................... .................... ....................
28470 MT632531.1 |Severe acute respiratory syndrome coro .................... .................... ..................... .................... .................... ....................
28471 MT632532.1 |Severe acute respiratory syndrome coro .................... .................... ..................... .................... .................... ....................
28472 MT632533.1 |Severe acute respiratory syndrome coro .................... .................... ..................... .................... .................... ....................
28473 MT632534.1 |Severe acute respiratory syndrome coro .................... .................... ..................... .................... .................... ....................
28474 MT632535.1 |Severe acute respiratory syndrome coro .................... .................... ..................... .................... .................... ....................
28475 MT632536.1 |Severe acute respiratory syndrome coro .................... .................... ..................... .................... .................... ....................
28476 MT632537.1 |Severe acute respiratory syndrome coro .................... .................... ..................... .................... .................... ....................
28477 MT632538.1 |Severe acute respiratory syndrome coro .................... .................... ..................... .................... .................... ....................
28478 MT632539.1 |Severe acute respiratory syndrome coro .................... .................... ..................... .................... .................... ....................
28479 MT632540.1 |Severe acute respiratory syndrome coro .................... .................... ..................... .................... .................... ....................
28480 MT632541.1 |Severe acute respiratory syndrome coro .................... .................... ..................... .................... .................... ....................
28481 MT632542.1 |Severe acute respiratory syndrome coro .................... .................... ..................... .................... .................... ....................
28482 MT632543.1 |Severe acute respiratory syndrome coro .................... .................... ..................... .................... .................... ....................
28483 MT632544.1 |Severe acute respiratory syndrome coro .................... .................... ..................... .................... .................... ....................
28484 MT632545.1 |Severe acute respiratory syndrome coro .................... .................... ..................... .................... .................... ....................
28485 MT632546.1 |Severe acute respiratory syndrome coro .................... .................... ..................... .................... .................... ....................
28486 MT632547.1 |Severe acute respiratory syndrome coro .................... .................... ..................... .................... .................... ....................
28487 MT632548.1 |Severe acute respiratory syndrome coro .................... .................... ..................... .................... .................... ....................
28488 MT632550.1 |Severe acute respiratory syndrome coro .................... .................... ..................... .................... .................... ....................
28489 MT632551.1 |Severe acute respiratory syndrome coro .................... .................... ..................... .................... .................... ....................
28490 MT632552.1 |Severe acute respiratory syndrome coro .................... .................... ..................... .................... .................... ....................
28491 MT632553.1 |Severe acute respiratory syndrome coro .................... .................... ..................... .................... .................... ....................
28492 MT632554.1 |Severe acute respiratory syndrome coro .................... .................... ..................... .................... .................... ....................
28493 MT632555.1 |Severe acute respiratory syndrome coro .................... .................... ..................... .................... .................... ....................
28494 MT632556.1 |Severe acute respiratory syndrome coro .................... .................... ..................... .................... .................... ....................
28495 MT632557.1 |Severe acute respiratory syndrome coro .................... .................... ..................... .................... .................... ....................
28496 MT632558.1 |Severe acute respiratory syndrome coro .................... .................... ..................... .................... .................... ....................
28497 MT632559.1 |Severe acute respiratory syndrome coro .................... .................... ..................... .................... .................... ....................
28498 MT632560.1 |Severe acute respiratory syndrome coro .................... .................... ..................... .................... .................... ....................
28499 MT632561.1 |Severe acute respiratory syndrome coro .................... .................... ..................... .................... .................... ....................
28500 MT632562.1 |Severe acute respiratory syndrome coro .................... .................... ..................... .................... .................... ....................
28501 MT632563.1 |Severe acute respiratory syndrome coro .................... .................... ..................... .................... .................... ....................
28503 MT632565.1 |Severe acute respiratory syndrome coro .................... .................... ..................... .................... .................... ....................
28504 MT632566.1 |Severe acute respiratory syndrome coro .................... .................... ..................... .................... .................... ....................
28505 MT632567.1 |Severe acute respiratory syndrome coro .................... .................... ..................... .................... .................... ....................
28506 MT632569.1 |Severe acute respiratory syndrome coro .................... .................... ..................... .................... .................... ....................
28507 MT632570.1 |Severe acute respiratory syndrome coro .................... .................... ..................... .................... .................... ....................
28508 MT632572.1 |Severe acute respiratory syndrome coro .................... .................... ..................... .................... .................... ....................
28509 MT632573.1 |Severe acute respiratory syndrome coro .................... .................... ..................... .................... .................... ....................
28510 MT632574.1 |Severe acute respiratory syndrome coro .................... .................... ..................... .................... .................... ....................
28511 MT632575.1 |Severe acute respiratory syndrome coro .................... .................... ..................... .................... .................... ....................
28512 MT632576.1 |Severe acute respiratory syndrome coro .................... .................... ..................... .................... .................... ....................
28513 MT632577.1 |Severe acute respiratory syndrome coro .................... .................... ..................... .................... .................... ....................
28514 MT632578.1 |Severe acute respiratory syndrome coro .................... .................... ..................... .................... .................... ....................
28515 MT632579.1 |Severe acute respiratory syndrome coro .................... .................... ..................... .................... .................... ....................
28516 MT632580.1 |Severe acute respiratory syndrome coro .................... .................... ..................... .................... .................... ....................
28517 MT632581.1 |Severe acute respiratory syndrome coro .................... .................... ..................... .................... .................... ....................
28518 MT632582.1 |Severe acute respiratory syndrome coro .................... .................... ..................... .................... .................... ....................
28519 MT632583.1 |Severe acute respiratory syndrome coro .................... .................... ..................... .................... .................... ....................
28520 MT632584.1 |Severe acute respiratory syndrome coro .................... .................... ..................... .................... .................... ....................
28521 MT632585.1 |Severe acute respiratory syndrome coro .................... .................... ..................... .................... .................... ....................
28522 MT632586.1 |Severe acute respiratory syndrome coro .................... .................... ..................... .................... .................... ....................
28523 MT632588.1 |Severe acute respiratory syndrome coro .................... .................... ..................... .................... .................... ....................
28524 MT632589.1 |Severe acute respiratory syndrome coro .................... .................... ..................... .................... .................... ....................
28525 MT632590.1 |Severe acute respiratory syndrome coro .................... .................... ..................... .................... .................... ....................
28526 MT632591.1 |Severe acute respiratory syndrome coro .................... .................... ..................... .................... .................... ....................

| | | | | | | | | |
|---|---|---|---|---|---|---|---|---|
| 28527 | MT632593.1 | \|Severe acute respiratory syndrome coro | ···················· | ···················· | ···················· | ···················· | ···················· | ···················· |
| 28528 | MT632594.1 | \|Severe acute respiratory syndrome coro | ···················· | ···················· | ···················· | ···················· | ···················· | ···················· |
| 28529 | MT632595.1 | \|Severe acute respiratory syndrome coro | ···················· | ···················· | ···················· | ···················· | ···················· | ···················· |
| 28530 | MT632596.1 | \|Severe acute respiratory syndrome coro | ···················· | ···················· | ···················· | ···················· | ···················· | ···················· |
| 28531 | MT632597.1 | \|Severe acute respiratory syndrome coro | ···················· | ···················· | ···················· | ···················· | ···················· | ···················· |
| 28532 | MT632598.1 | \|Severe acute respiratory syndrome coro | ···················· | ···················· | ···················· | ···················· | ···················· | ···················· |
| 28533 | MT632599.1 | \|Severe acute respiratory syndrome coro | ···················· | ···················· | ···················· | ···················· | ···················· | ···················· |
| 28534 | MT632600.1 | \|Severe acute respiratory syndrome coro | ···················· | ···················· | ···················· | ···················· | ···················· | ···················· |
| 28535 | MT632601.1 | \|Severe acute respiratory syndrome coro | ···················· | ···················· | ···················· | ···················· | ···················· | ···················· |
| 28536 | MT632602.1 | \|Severe acute respiratory syndrome coro | ···················· | ···················· | ···················· | ···················· | ···················· | ···················· |
| 28537 | MT632603.1 | \|Severe acute respiratory syndrome coro | ···················· | ···················· | ···················· | ···················· | ···················· | ···················· |
| 28538 | MT632605.1 | \|Severe acute respiratory syndrome coro | ···················· | ···················· | ···················· | ···················· | ···················· | ···················· |
| 28539 | MT632606.1 | \|Severe acute respiratory syndrome coro | ···················· | ···················· | ···················· | ···················· | ···················· | ···················· |
| 28540 | MT632607.1 | \|Severe acute respiratory syndrome coro | ···················· | ···················· | ···················· | ···················· | ···················· | ···················· |
| 28541 | MT632608.1 | \|Severe acute respiratory syndrome coro | ···················· | ···················· | ···················· | ···················· | ···················· | ···················· |
| 28542 | MT632609.1 | \|Severe acute respiratory syndrome coro | ···················· | ···················· | ···················· | ···················· | ···················· | ···················· |
| 28543 | MT632610.1 | \|Severe acute respiratory syndrome coro | ···················· | ···················· | ···················· | ···················· | ···················· | ···················· |
| 28544 | MT632611.1 | \|Severe acute respiratory syndrome coro | ···················· | ···················· | ···················· | ···················· | ···················· | ···················· |
| 28545 | MT632612.1 | \|Severe acute respiratory syndrome coro | ···················· | ···················· | ···················· | ···················· | ···················· | ···················· |
| 28546 | MT632613.1 | \|Severe acute respiratory syndrome coro | ···················· | ···················· | ···················· | ···················· | ···················· | ···················· |
| 28547 | MT632614.1 | \|Severe acute respiratory syndrome coro | ···················· | ···················· | ···················· | ···················· | ···················· | ···················· |
| 28548 | MT632615.1 | \|Severe acute respiratory syndrome coro | ···················· | ···················· | ···················· | ···················· | ···················· | ···················· |
| 28549 | MT632616.1 | \|Severe acute respiratory syndrome coro | ···················· | ···················· | ···················· | ···················· | ···················· | ···················· |
| 28550 | MT632618.1 | \|Severe acute respiratory syndrome coro | ···················· | ···················· | ···················· | ···················· | ···················· | ···················· |
| 28551 | MT632619.1 | \|Severe acute respiratory syndrome coro | ···················· | ···················· | ···················· | ···················· | ···················· | ···················· |
| 28552 | MT632620.1 | \|Severe acute respiratory syndrome coro | ···················· | ···················· | ···················· | ···················· | ···················· | ···················· |
| 28553 | MT632621.1 | \|Severe acute respiratory syndrome coro | ···················· | ···················· | ···················· | ···················· | ···················· | ···················· |
| 28554 | MT632622.1 | \|Severe acute respiratory syndrome coro | ···················· | ···················· | ···················· | ···················· | ···················· | ···················· |
| 28555 | MT632623.1 | \|Severe acute respiratory syndrome coro | ···················· | ···················· | ···················· | ···················· | ···················· | ···················· |
| 28556 | MT632624.1 | \|Severe acute respiratory syndrome coro | ···················· | ···················· | ···················· | ···················· | ···················· | ···················· |
| 28557 | MT632625.1 | \|Severe acute respiratory syndrome coro | ···················· | ···················· | ···················· | ···················· | ···················· | ···················· |
| 28558 | MT632626.1 | \|Severe acute respiratory syndrome coro | ···················· | ···················· | ···················· | ···················· | ···················· | ···················· |
| 28559 | MT632627.1 | \|Severe acute respiratory syndrome coro | ···················· | ···················· | ···················· | ···················· | ···················· | ···················· |
| 28560 | MT632629.1 | \|Severe acute respiratory syndrome coro | ···················· | ···················· | ···················· | ···················· | ···················· | ···················· |
| 28561 | MT632630.1 | \|Severe acute respiratory syndrome coro | ···················· | ···················· | ···················· | ···················· | ···················· | ···················· |
| 28562 | MT632631.1 | \|Severe acute respiratory syndrome coro | ···················· | ···················· | ···················· | ···················· | ···················· | ···················· |
| 28563 | MT632632.1 | \|Severe acute respiratory syndrome coro | ···················· | ···················· | ···················· | ···················· | ···················· | ···················· |
| 28564 | MT632766.1 | \|Severe acute respiratory syndrome coro | ···················· | ···················· | ···················· | ···················· | ···················· | ···················· |
| 28565 | MT632769.1 | \|Severe acute respiratory syndrome coro | ···················· | ···················· | ···················· | ···················· | ···················· | ···················· |
| 28566 | MT632770.1 | \|Severe acute respiratory syndrome coro | ···················· | ···················· | ···················· | ···················· | ···················· | ···················· |
| 28567 | MT632771.1 | \|Severe acute respiratory syndrome coro | ···················· | ···················· | ···················· | ···················· | ···················· | ···················· |
| 28568 | MT632772.1 | \|Severe acute respiratory syndrome coro | ···················· | ···················· | ···················· | ···················· | ···················· | ···················· |
| 28569 | MT632773.1 | \|Severe acute respiratory syndrome coro | ···················· | ···················· | ···················· | ···················· | ···················· | ···················· |
| 28570 | MT632775.1 | \|Severe acute respiratory syndrome coro | ···················· | ···················· | ···················· | ···················· | ···················· | ···················· |
| 28571 | MT632776.1 | \|Severe acute respiratory syndrome coro | ···················· | ···················· | ···················· | ···················· | ···················· | ···················· |
| 28572 | MT632777.1 | \|Severe acute respiratory syndrome coro | ···················· | ···················· | ···················· | ···················· | ···················· | ···················· |
| 28573 | MT632778.1 | \|Severe acute respiratory syndrome coro | ···················· | ···················· | ···················· | ···················· | ···················· | ···················· |
| 28574 | MT632780.1 | \|Severe acute respiratory syndrome coro | ···················· | ···················· | ···················· | ···················· | ···················· | ···················· |
| 28575 | MT632781.1 | \|Severe acute respiratory syndrome coro | ···················· | ···················· | ···················· | ···················· | ···················· | ···················· |
| 28576 | MT632782.1 | \|Severe acute respiratory syndrome coro | ···················· | ···················· | ···················· | ···················· | ···················· | ···················· |
| 28577 | MT632783.1 | \|Severe acute respiratory syndrome coro | ···················· | ···················· | ···················· | ···················· | ···················· | ···················· |
| 28578 | MT632784.1 | \|Severe acute respiratory syndrome coro | ···················· | ···················· | ···················· | ···················· | ···················· | ···················· |
| 28579 | MT632785.1 | \|Severe acute respiratory syndrome coro | ···················· | ···················· | ···················· | ···················· | ···················· | ···················· |
| 28580 | MT632786.1 | \|Severe acute respiratory syndrome coro | ···················· | ···················· | ···················· | ···················· | ···················· | ···················· |
| 28581 | MT632787.1 | \|Severe acute respiratory syndrome coro | ···················· | ···················· | ···················· | ···················· | ···················· | ···················· |
| 28582 | MT632788.1 | \|Severe acute respiratory syndrome coro | ···················· | ···················· | ···················· | ···················· | ···················· | ···················· |
| 28583 | MT632789.1 | \|Severe acute respiratory syndrome coro | ···················· | ···················· | ···················· | ···················· | ···················· | ···················· |
| 28584 | MT632790.1 | \|Severe acute respiratory syndrome coro | ···················· | ···················· | ···················· | ···················· | ················G··· | ···················· |
| 28585 | MT632791.1 | \|Severe acute respiratory syndrome coro | ···················· | ···················· | ···················· | ···················· | ···················· | ···················· |
| 28586 | MT632792.1 | \|Severe acute respiratory syndrome coro | ···················· | ···················· | ···················· | ···················· | ···················· | ···················· |
| 28587 | MT632793.1 | \|Severe acute respiratory syndrome coro | ···················· | ···················· | ···················· | ···················· | ···················· | ···················· |
| 28588 | MT632794.1 | \|Severe acute respiratory syndrome coro | ···················· | ···················· | ···················· | ···················· | ···················· | ···················· |
| 28589 | MT632795.1 | \|Severe acute respiratory syndrome coro | ···················· | ···················· | ···················· | ···················· | ···················· | ···················· |
| 28590 | MT632796.1 | \|Severe acute respiratory syndrome coro | ···················· | ···················· | ···················· | ···················· | ···················· | ···················· |
| 28591 | MT632797.1 | \|Severe acute respiratory syndrome coro | ···················· | ···················· | ···················· | ···················· | ···················· | ···················· |
| 28592 | MT632798.1 | \|Severe acute respiratory syndrome coro | ···················· | ···················· | ···················· | ···················· | ···················· | ···················· |
| 28593 | MT632799.1 | \|Severe acute respiratory syndrome coro | ···················· | ···················· | ···················· | ···················· | ···················· | ···················· |
| 28594 | MT632800.1 | \|Severe acute respiratory syndrome coro | ···················· | ···················· | ···················· | ···················· | ···················· | ···················· |
| 28595 | MT632801.1 | \|Severe acute respiratory syndrome coro | ···················· | ···················· | ···················· | ···················· | ···················· | ···················· |
| 28596 | MT632802.1 | \|Severe acute respiratory syndrome coro | ···················· | ···················· | ···················· | ···················· | ···················· | ···················· |
| 28597 | MT632803.1 | \|Severe acute respiratory syndrome coro | ···················· | ···················· | ···················· | ···················· | ···················· | ···················· |
| 28598 | MT632804.1 | \|Severe acute respiratory syndrome coro | ···················· | ···················· | ···················· | ···················· | ···················· | ···················· |
| 28599 | MT632805.1 | \|Severe acute respiratory syndrome coro | ···················· | ···················· | ···················· | ···················· | ···················· | ···················· |
| 28600 | MT632806.1 | \|Severe acute respiratory syndrome coro | ···················· | ···················· | ···················· | ···················· | ···················· | ···················· |
| 28601 | MT632807.1 | \|Severe acute respiratory syndrome coro | ···················· | ···················· | ···················· | ···················· | ···················· | ···················· |
| 28602 | MT632808.1 | \|Severe acute respiratory syndrome coro | ···················· | ···················· | ···················· | ···················· | ···················· | ···················· |
| 28603 | MT632810.1 | \|Severe acute respiratory syndrome coro | ···················· | ···················· | ···················· | ···················· | ···················· | ···················· |
| 28604 | MT632813.1 | \|Severe acute respiratory syndrome coro | ···················· | ···················· | ···················· | ···················· | ···················· | ···················· |

28605 MT632814.1 |Severe acute respiratory syndrome coro ···················· ···················· ····················· ···················· ···················· ····················
28606 MT632815.1 |Severe acute respiratory syndrome coro ···················· ···················· ····················· ···················· ···················· ····················
28607 MT632816.1 |Severe acute respiratory syndrome coro ···················· ···················· ····················· ···················· ···················· ····················
28608 MT632817.1 |Severe acute respiratory syndrome coro ···················· ···················· ····················· ···················· ···················· ····················
28609 MT632818.1 |Severe acute respiratory syndrome coro ···················· ···················· ····················· ···················· ···················· ····················
28610 MT632819.1 |Severe acute respiratory syndrome coro ···················· ···················· ····················· ···················· ···················· ····················
28611 MT632820.1 |Severe acute respiratory syndrome coro ···················· ···················· ····················· ···················· ···················· ····················
28612 MT632821.1 |Severe acute respiratory syndrome coro ···················· ···················· ····················· ···················· ···················· ····················
28613 MT632823.1 |Severe acute respiratory syndrome coro ···················· ···················· ····················· ···················· ···················· ····················
28614 MT632824.1 |Severe acute respiratory syndrome coro ···················· ···················· ····················· ···················· ···················· ····················
28615 MT632825.1 |Severe acute respiratory syndrome coro ···················· ···················· ····················· ···················· ···················· ····················
28616 MT632826.1 |Severe acute respiratory syndrome coro ···················· ···················· ····················· ···················· ···················· ····················
28617 MT632827.1 |Severe acute respiratory syndrome coro ···················· ···················· ····················· ···················· ···················· ····················
28618 MT632828.1 |Severe acute respiratory syndrome coro ···················· ···················· ····················· ···················· ···················· ····················
28619 MT632829.1 |Severe acute respiratory syndrome coro ···················· ···················· ····················· ···················· ···················· ····················
28620 MT632830.1 |Severe acute respiratory syndrome coro ···················· ···················· ····················· ···················· ···················· ····················
28621 MT632831.1 |Severe acute respiratory syndrome coro ···················· ···················· ····················· ···················· ···················· ····················
28622 MT632832.1 |Severe acute respiratory syndrome coro ···················· ···················· ····················· ···················· ···················· ····················
28623 MT632833.1 |Severe acute respiratory syndrome coro ···················· ···················· ····················· ···················· ···················· ····················
28624 MT632835.1 |Severe acute respiratory syndrome coro ···················· ···················· ····················· ···················· ···················· ····················
28625 MT632836.1 |Severe acute respiratory syndrome coro ···················· ···················· ····················· ···················· ···················· ····················
28626 MT632838.1 |Severe acute respiratory syndrome coro ···················· ···················· ····················· ···················· ···················· ····················
28627 MT632839.1 |Severe acute respiratory syndrome coro ···················· ···················· ····················· ···················· ···················· ····················
28628 MT632840.1 |Severe acute respiratory syndrome coro ···················· ···················· ····················· ···················· ···················· ····················
28629 MT632841.1 |Severe acute respiratory syndrome coro ···················· ···················· ····················· ···················· ···················· ····················
28630 MT632842.1 |Severe acute respiratory syndrome coro ···················· ···················· ····················· ···················· ···················· ····················
28631 MT632843.1 |Severe acute respiratory syndrome coro ···················· ···················· ····················· ···················· ···················· ····················
28632 MT632844.1 |Severe acute respiratory syndrome coro ···················· ···················· ····················· ···················· ···················· ····················
28633 MT632845.1 |Severe acute respiratory syndrome coro ···················· ···················· ····················· ···················· ···················· ····················
28634 MT632846.1 |Severe acute respiratory syndrome coro ···················· ···················· ····················· ···················· ···················· ····················
28635 MT632847.1 |Severe acute respiratory syndrome coro ···················· ···················· ····················· ···················· ···················· ····················
28636 MT632848.1 |Severe acute respiratory syndrome coro ···················· ···················· ····················· ···················· ···················· ····················
28637 MT632849.1 |Severe acute respiratory syndrome coro ···················· ···················· ····················· ···················· ···················· ····················
28638 MT632850.1 |Severe acute respiratory syndrome coro ···················· ···················· ····················· ···················· ···················· ····················
28639 MT632851.1 |Severe acute respiratory syndrome coro ···················· ···················· ····················· ···················· ···················· ····················
28640 MT632852.1 |Severe acute respiratory syndrome coro ···················· ···················· ····················· ···················· ···················· ····················
28641 MT632853.1 |Severe acute respiratory syndrome coro ···················· ···················· ····················· ···················· ···················· ····················
28642 MT632854.1 |Severe acute respiratory syndrome coro ···················· ···················· ····················· ···················· ···················· ····················
28643 MT632855.1 |Severe acute respiratory syndrome coro ···················· ···················· ····················· ···················· ···················· ····················
28644 MT632856.1 |Severe acute respiratory syndrome coro ···················· ···················· ····················· ···················· ···················· ····················
28645 MT632857.1 |Severe acute respiratory syndrome coro ···················· ···················· ····················· ···················· ···················· ····················
28646 MT632858.1 |Severe acute respiratory syndrome coro ···················· ···················· ····················· ···················· ···················· ····················
28647 MT632859.1 |Severe acute respiratory syndrome coro ···················· ···················· ····················· ···················· ···················· ····················
28648 MT632860.1 |Severe acute respiratory syndrome coro ···················· ···················· ····················· ···················· ···················· ····················
28649 MT632861.1 |Severe acute respiratory syndrome coro ···················· ···················· ····················· ···················· ···················· ····················
28650 MT632863.1 |Severe acute respiratory syndrome coro ···················· ···················· ····················· ···················· ···················· ····················
28651 MT632865.1 |Severe acute respiratory syndrome coro ···················· ···················· ····················· ···················· ···················· ····················
28652 MT632866.1 |Severe acute respiratory syndrome coro ···················· ···················· ····················· ···················· ···················· ····················
28653 MT632867.1 |Severe acute respiratory syndrome coro ···················· ···················· ····················· ···················· ···················· ····················
28654 MT632868.1 |Severe acute respiratory syndrome coro ···················· ···················· ····················· ···················· ···················· ····················
28655 MT632869.1 |Severe acute respiratory syndrome coro ···················· ···················· ····················· ···················· ···················· ····················
28656 MT632871.1 |Severe acute respiratory syndrome coro ···················· ···················· ····················· ···················· ···················· ····················
28657 MT632872.1 |Severe acute respiratory syndrome coro ···················· ···················· ····················· ···················· ···················· ····················
28658 MT632874.1 |Severe acute respiratory syndrome coro ···················· ···················· ····················· ···················· ···················· ····················
28659 MT632875.1 |Severe acute respiratory syndrome coro ···················· ···················· ····················· ···················· ···················· ····················
28660 MT632876.1 |Severe acute respiratory syndrome coro ···················· ···················· ····················· ···················· ···················· ····················
28661 MT632877.1 |Severe acute respiratory syndrome coro ···················· ···················· ····················· ···················· ···················· ····················
28662 MT632878.1 |Severe acute respiratory syndrome coro ···················· ···················· ····················· ···················· ···················· ····················
28663 MT632879.1 |Severe acute respiratory syndrome coro ···················· ···················· ····················· ···················· ···················· ····················
28664 MT632880.1 |Severe acute respiratory syndrome coro ···················· ···················· ····················· ···················· ···················· ····················
28665 MT632881.1 |Severe acute respiratory syndrome coro ···················· ···················· ····················· ···················· ···················· ····················
28666 MT632882.1 |Severe acute respiratory syndrome coro ···················· ···················· ····················· ···················· ···················· ····················
28667 MT632883.1 |Severe acute respiratory syndrome coro ···················· ···················· ····················· ···················· ···················· ····················
28668 MT632884.1 |Severe acute respiratory syndrome coro ········T··········· ···················· ····················· ···················· ···················· ····················
28669 MT632885.1 |Severe acute respiratory syndrome coro ···················· ···················· ····················· ···················· ···················· ····················
28670 MT632886.1 |Severe acute respiratory syndrome coro ···················· ···················· ····················· ···················· ···················· ····················
28671 MT632887.1 |Severe acute respiratory syndrome coro ···················· ···················· ····················· ···················· ···················· ····················
28672 MT632888.1 |Severe acute respiratory syndrome coro ···················· ···················· ····················· ···················· ···················· ····················
28673 MT632889.1 |Severe acute respiratory syndrome coro ···················· ···················· ····················· ···················· ···················· ····················
28674 MT632890.1 |Severe acute respiratory syndrome coro ···················· ···················· ····················· ···················· ···················· ····················
28675 MT632892.1 |Severe acute respiratory syndrome coro ···················· ···················· ····················· ···················· ···················· ····················
28676 MT632893.1 |Severe acute respiratory syndrome coro ···················· ···················· ····················· ···················· ···················· ····················
28677 MT632894.1 |Severe acute respiratory syndrome coro ···················· ···················· ····················· ···················· ···················· ····················
28678 MT632895.1 |Severe acute respiratory syndrome coro ···················· ···················· ····················· ···················· ···················· ····················
28679 MT632896.1 |Severe acute respiratory syndrome coro ···················· ···················· ····················· ···················· ···················· ····················
28680 MT632897.1 |Severe acute respiratory syndrome coro ···················· ···················· ····················· ···················· ···················· ····················
28681 MT632900.1 |Severe acute respiratory syndrome coro ···················· ···················· ····················· ···················· ···················· ····················
28682 MT632901.1 |Severe acute respiratory syndrome coro ···················· ···················· ····················· ···················· ···················· ····················

28683 MT632902.1 |Severe acute respiratory syndrome coro .................... .................... .................... .................... .................... ....................
28684 MT632903.1 |Severe acute respiratory syndrome coro .................... .................... .................... .................... .................... ....................
28685 MT632904.1 |Severe acute respiratory syndrome coro .................... .................... .................... .................... .................... ....................
28686 MT632905.1 |Severe acute respiratory syndrome coro .................... .................... .................... .................... .................... ....................
28687 MT632907.1 |Severe acute respiratory syndrome coro .................... .................... .................... .................... .................... ....................
28688 MT632908.1 |Severe acute respiratory syndrome coro .................... .................... .................... .................... .................... ....................
28689 MT632909.1 |Severe acute respiratory syndrome coro .................... .................... .................... .................... .................... ....................
28690 MT632910.1 |Severe acute respiratory syndrome coro .................... .................... .................... .................... .................... ....................
28691 MT632911.1 |Severe acute respiratory syndrome coro .................... .................... .................... .................... .................... ....................
28692 MT632912.1 |Severe acute respiratory syndrome coro .................... .................... .................... .................... .................... ....................
28693 MT632914.1 |Severe acute respiratory syndrome coro .................... .................... .................... .................... .................... ....................
28694 MT632916.1 |Severe acute respiratory syndrome coro .................... .................... .................... .................... .................... ....................
28695 MT632917.1 |Severe acute respiratory syndrome coro .................... .................... .................... .................... .................... ....................
28696 MT632918.1 |Severe acute respiratory syndrome coro .................... .................... .................... .................... .................... ....................
28697 MT632919.1 |Severe acute respiratory syndrome coro .................... .................... .................... .................... .................... ....................
28698 MT632920.1 |Severe acute respiratory syndrome coro .................... .................... .................... .................... .................... ....................
28699 MT632921.1 |Severe acute respiratory syndrome coro .................... .................... .................... .................... .................... ....................
28700 MT632922.1 |Severe acute respiratory syndrome coro .................... .................... .................... .................... .................... ....................
28701 MT632923.1 |Severe acute respiratory syndrome coro .................... .................... .................... .................... .................... ....................
28702 MT632925.1 |Severe acute respiratory syndrome coro .................... .................... .................... .................... .................... ....................
28703 MT632926.1 |Severe acute respiratory syndrome coro .................... .................... .................... .................... .................... ....................
28704 MT632927.1 |Severe acute respiratory syndrome coro .................... .................... .................... .................... .................... ....................
28705 MT632928.1 |Severe acute respiratory syndrome coro .................... .................... .................... .................... .................... ....................
28706 MT632929.1 |Severe acute respiratory syndrome coro .................... .................... .................... .................... .................... ....................
28707 MT632930.1 |Severe acute respiratory syndrome coro .................... .................... .................... .................... .................... ....................
28708 MT632931.1 |Severe acute respiratory syndrome coro .................... .................... .................... .................... .................... ....................
28709 MT632932.1 |Severe acute respiratory syndrome coro .................... .................... .................... .................... .................... ....................
28710 MT632933.1 |Severe acute respiratory syndrome coro .................... .................... .................... .................... .................... ....................
28711 MT632934.1 |Severe acute respiratory syndrome coro .................... .................... .................... .................... .................... ....................
28712 MT632935.1 |Severe acute respiratory syndrome coro .................... .................... .................... .................... .................... ....................
28713 MT632936.1 |Severe acute respiratory syndrome coro .................... .................... .................... .................... .................... ....................
28714 MT632937.1 |Severe acute respiratory syndrome coro .................... .................... .................... .................... .................... ....................
28715 MT632939.1 |Severe acute respiratory syndrome coro .................... .................... .................... .................... .................... ....................
28716 MT632940.1 |Severe acute respiratory syndrome coro .................... .................... .................... .................... .................... ....................
28717 MT632941.1 |Severe acute respiratory syndrome coro .................... .................... .................... .................... .................... ....................
28718 MT632942.1 |Severe acute respiratory syndrome coro .................... .................... .................... .................... .................... ....................
28719 MT632943.1 |Severe acute respiratory syndrome coro .................... .................... .................... .................... .................... ....................
28720 MT632944.1 |Severe acute respiratory syndrome coro .................... .................... .................... .................... .................... ....................
28721 MT632945.1 |Severe acute respiratory syndrome coro .................... .................... .................... .................... .................... ....................
28722 MT632946.1 |Severe acute respiratory syndrome coro .................... .................... .................... .................... .................... ....................
28723 MT632947.1 |Severe acute respiratory syndrome coro .................... .................... .................... .................... .................... ....................
28724 MT632948.1 |Severe acute respiratory syndrome coro .................... .................... .................... .................... .................... ....................
28725 MT632949.1 |Severe acute respiratory syndrome coro .................... .................... .................... .................... .................... ....................
28726 MT632950.1 |Severe acute respiratory syndrome coro .................... .................... .................... .................... .................... ....................
28727 MT632952.1 |Severe acute respiratory syndrome coro .................... .................... .................... .................... .................... ....................
28728 MT632953.1 |Severe acute respiratory syndrome coro .................... .................... .................... .................... .................... ....................
28729 MT632954.1 |Severe acute respiratory syndrome coro .................... .................... .................... .................... .................... ....................
28730 MT632955.1 |Severe acute respiratory syndrome coro .................... .................... .................... .................... .................... ....................
28731 MT632956.1 |Severe acute respiratory syndrome coro .................... .................... .................... .................... .................... ....................
28732 MT632957.1 |Severe acute respiratory syndrome coro .................... .................... .................... .................... .................... ....................
28733 MT632958.1 |Severe acute respiratory syndrome coro .................... .................... .................... .................... .................... ....................
28734 MT632959.1 |Severe acute respiratory syndrome coro .................... .................... .................... .................... .................... ....................
28735 MT632960.1 |Severe acute respiratory syndrome coro .................... .................... .................... .................... .................... ....................
28736 MT632961.1 |Severe acute respiratory syndrome coro .................... .................... .................... .................... .................... ....................
28737 MT632962.1 |Severe acute respiratory syndrome coro .................... .................... .................... .................... .................... ....................
28738 MT632963.1 |Severe acute respiratory syndrome coro .................... .................... .................... .................... .................... ....................
28739 MT632964.1 |Severe acute respiratory syndrome coro .................... .................... .................... .................... .................... ....................
28740 MT632965.1 |Severe acute respiratory syndrome coro .................... .................... .................... .................... .................... ....................
28741 MT632966.1 |Severe acute respiratory syndrome coro .................... .................... .................... .................... .................... ....................
28742 MT632967.1 |Severe acute respiratory syndrome coro .................... .................... .................... .................... .................... ....................
28743 MT632968.1 |Severe acute respiratory syndrome coro .................... .................... .................... .................... .................... ....................
28744 MT632969.1 |Severe acute respiratory syndrome coro .................... .................... .................... .................... .................... ....................
28745 MT632970.1 |Severe acute respiratory syndrome coro .................... .................... .................... .................... .................... ....................
28746 MT632971.1 |Severe acute respiratory syndrome coro .................... .................... .................... .................... .................... ....................
28747 MT632972.1 |Severe acute respiratory syndrome coro .................... .................... .................... .................... .................... ....................
28748 MT632973.1 |Severe acute respiratory syndrome coro .................... .................... .................... .................... .................... ....................
28749 MT632974.1 |Severe acute respiratory syndrome coro .................... .................... .................... .................... .................... ....................
28750 MT632975.1 |Severe acute respiratory syndrome coro .................... .................... .................... .................... .................... ....................
28751 MT632976.1 |Severe acute respiratory syndrome coro .................... .................... .................... .................... .................... ....................
28752 MT632977.1 |Severe acute respiratory syndrome coro .................... .................... .................... .................... .................... ....................
28753 MT632978.1 |Severe acute respiratory syndrome coro .................... .................... .................... .................... .................... ....................
28754 MT632979.1 |Severe acute respiratory syndrome coro .................... .................... .................... .................... .................... ....................
28755 MT632981.1 |Severe acute respiratory syndrome coro .................... .................... .................... .................... .................... ....................
28756 MT632982.1 |Severe acute respiratory syndrome coro .................... .................... .................... .................... .................... ....................
28757 MT632983.1 |Severe acute respiratory syndrome coro .................... .................... .................... .................... .................... ....................
28758 MT632984.1 |Severe acute respiratory syndrome coro .................... .................... .................... .................... .................... ....................
28759 MT632985.1 |Severe acute respiratory syndrome coro .................... .................... .................... .................... .................... ....................
28760 MT632986.1 |Severe acute respiratory syndrome coro .................... .................... .................... .................... .................... ....................

28761 MT632987.1 |Severe acute respiratory syndrome coro .................... .................... ..................... .................... .................... ....................
28762 MT632988.1 |Severe acute respiratory syndrome coro .................... .................... ..................... .................... .................... ....................
28763 MT632989.1 |Severe acute respiratory syndrome coro .................... .................... ..................... .................... .................... ....................
28764 MT632991.1 |Severe acute respiratory syndrome coro .................... .................... ..................... .................... .................... ....................
28765 MT632992.1 |Severe acute respiratory syndrome coro .................... .................... ..................... .................... .................... ....................
28766 MT632993.1 |Severe acute respiratory syndrome coro .................... .................... ..................... .................... .................... ....................
28767 MT632994.1 |Severe acute respiratory syndrome coro .................... .................... ..................... .................... .................... ....................
28768 MT632995.1 |Severe acute respiratory syndrome coro .................... .................... ..................... .................... .................... ....................
28769 MT632996.1 |Severe acute respiratory syndrome coro .................... .................... ..................... .................... .................... ....................
28770 MT632997.1 |Severe acute respiratory syndrome coro .................... .................... ..................... .................... .................... ....................
28771 MT632998.1 |Severe acute respiratory syndrome coro .................... .................... ..................... .................... .................... ....................
28772 MT632999.1 |Severe acute respiratory syndrome coro .................... .................... ..................... .................... .................... ....................
28773 MT633000.1 |Severe acute respiratory syndrome coro .................... .................... ..................... .................... .................... ....................
28774 MT633001.1 |Severe acute respiratory syndrome coro .................... .................... ..................... .................... .................... ....................
28775 MT633002.1 |Severe acute respiratory syndrome coro .................... .................... ..................... .................... .................... ....................
28776 MT633003.1 |Severe acute respiratory syndrome coro .................... .................... ..................... .................... .................... ....................
28777 MT633004.1 |Severe acute respiratory syndrome coro .................... .................... ..................... .................... .................... ....................
28778 MT633005.1 |Severe acute respiratory syndrome coro .................... .................... ..................... .................... .................... ....................
28779 MT633006.1 |Severe acute respiratory syndrome coro .................... .................... ..................... .................... .................... ....................
28780 MT633007.1 |Severe acute respiratory syndrome coro .................... .................... ..................... .................... .................... ....................
28781 MT633008.1 |Severe acute respiratory syndrome coro .................... .................... ..................... .................... .................... ....................
28782 MT633009.1 |Severe acute respiratory syndrome coro .................... .................... ..................... .................... .................... ....................
28783 MT633010.1 |Severe acute respiratory syndrome coro .................... .................... ..................... .................... .................... ....................
28784 MT633011.1 |Severe acute respiratory syndrome coro .................... .................... ..................... .................... .................... ....................
28785 MT633013.1 |Severe acute respiratory syndrome coro .................... .................... ..................... .................... .................... ....................
28786 MT633014.1 |Severe acute respiratory syndrome coro .................... .................... ..................... .................... .................... ....................
28787 MT633015.1 |Severe acute respiratory syndrome coro .................... .................... ..................... .................... .................... ....................
28788 MT633016.1 |Severe acute respiratory syndrome coro .................... .................... ..................... .................... .................... ....................
28789 MT633017.1 |Severe acute respiratory syndrome coro .................... .................... ..................... .................... .................... ....................
28790 MT633018.1 |Severe acute respiratory syndrome coro .................... .................... ..................... .................... .................... ....................
28791 MT633019.1 |Severe acute respiratory syndrome coro .................... .................... ..................... .................... .................... ....................
28792 MT633020.1 |Severe acute respiratory syndrome coro .................... .................... ..................... .................... .................... ....................
28793 MT633021.1 |Severe acute respiratory syndrome coro .................... .................... ..................... .................... .................... ....................
28794 MT633022.1 |Severe acute respiratory syndrome coro .................... .................... ..................... .................... .................... ....................
28795 MT633023.1 |Severe acute respiratory syndrome coro .................... .................... ..................... .................... .................... ....................
28796 MT633024.1 |Severe acute respiratory syndrome coro .................... .................... ..................... .................... .................... ....................
28797 MT633025.1 |Severe acute respiratory syndrome coro .................... .................... ..................... .................... .................... ....................
28798 MT633026.1 |Severe acute respiratory syndrome coro .................... .................... ..................... .................... .................... ....................
28799 MT633027.1 |Severe acute respiratory syndrome coro .................... .................... ..................... .................... .................... ....................
28800 MT633028.1 |Severe acute respiratory syndrome coro .................... .................... ..................... .................... .................... ....................
28801 MT633029.1 |Severe acute respiratory syndrome coro .................... .................... ..................... .................... .................... ....................
28802 MT633030.1 |Severe acute respiratory syndrome coro .................... .................... ..................... .................... .................... ....................
28803 MT633031.1 |Severe acute respiratory syndrome coro .................... .................... ..................... .................... .................... ....................
28804 MT633032.1 |Severe acute respiratory syndrome coro .................... .................... ..................... .................... .................... ....................
28805 MT633033.1 |Severe acute respiratory syndrome coro .................... .................... ..................... .................... .................... ....................
28806 MT633034.1 |Severe acute respiratory syndrome coro .................... .................... ..................... .................... .................... ....................
28807 MT633035.1 |Severe acute respiratory syndrome coro .................... .................... ..................... .................... .................... ....................
28808 MT633036.1 |Severe acute respiratory syndrome coro .................... .................... ..................... .................... .................... ....................
28809 MT633037.1 |Severe acute respiratory syndrome coro .................... .................... ..................... .................... .................... ....................
28810 MT633038.1 |Severe acute respiratory syndrome coro .................... .................... ..................... .................... .................... ....................
28811 MT633039.1 |Severe acute respiratory syndrome coro .................... .................... ..................... .................... .................... ....................
28812 MT633041.1 |Severe acute respiratory syndrome coro .................... .................... ..................... .................... .................... ....................
28813 MT633044.1 |Severe acute respiratory syndrome coro .................... .................... ..................... .................... .................... ....................
28814 MT633045.1 |Severe acute respiratory syndrome coro .................... .................... ..................... .................... .................... ....................
28815 MT633046.1 |Severe acute respiratory syndrome coro .................... .................... ..................... .................... .................... ....................
28816 LC553269.1 |Severe acute respiratory syndrome coro .................... .................... ..................... .................... .................... ....................
28817 LC553270.1 |Severe acute respiratory syndrome coro .................... .................... ..................... .................... .................... ....................
28818 MT614447.1 |Severe acute respiratory syndrome coro .................... .................... ..................... .................... .................... ....................
28819 MT614449.1 |Severe acute respiratory syndrome coro .................... .................... ..................... .................... .................... ....................
28820 MT614450.1 |Severe acute respiratory syndrome coro .................... .................... ..................... .................... .................... ....................
28821 MT614452.1 |Severe acute respiratory syndrome coro .................... .................... ..................... .................... .................... ....................
28822 MT614453.1 |Severe acute respiratory syndrome coro .................... .................... ..................... .................... .................... ....................
28823 MT614454.1 |Severe acute respiratory syndrome coro .................... .................... ..................... .................... .................... ....................
28824 MT614455.1 |Severe acute respiratory syndrome coro .................... .................... ..................... .................... .................... ....................
28825 MT614456.1 |Severe acute respiratory syndrome coro .................... .................... ..................... .................... .................... ....................
28826 MT614457.1 |Severe acute respiratory syndrome coro .................... .................... ..................... .................... .................... ....................
28827 MT614458.1 |Severe acute respiratory syndrome coro .................... .................... ..................... .................... .................... ....................
28828 MT614459.1 |Severe acute respiratory syndrome coro .................... .................... ..................... .................... .................... ....................
28829 MT614460.1 |Severe acute respiratory syndrome coro .................... .................... ..................... .................... .................... ....................
28830 MT614461.1 |Severe acute respiratory syndrome coro .................... .................... ..................... .................... .................... ....................
28831 MT614462.1 |Severe acute respiratory syndrome coro .................... .................... ..................... .................... .................... ....................
28832 MT614463.1 |Severe acute respiratory syndrome coro .................... .................... ..................... .................... .................... ....................
28833 MT614464.1 |Severe acute respiratory syndrome coro .................... .................... ..................... .................... .................... ....................
28834 MT614465.1 |Severe acute respiratory syndrome coro .................... .................... ..................... .................... .................... ....................
28835 MT614466.1 |Severe acute respiratory syndrome coro .................... .................... ..................... .................... .................... ....................
28836 MT614467.1 |Severe acute respiratory syndrome coro .................... .................... ..................... .................... .................... ....................
28837 MT614468.1 |Severe acute respiratory syndrome coro .................... .................... ..................... .................... .................... ....................
28838 MT614469.1 |Severe acute respiratory syndrome coro .................... .................... ..................... .................... .................... ....................

28839 MT614470.1 |Severe acute respiratory syndrome coro .................... .................... .................... .................... .................... ....................
28840 MT614471.1 |Severe acute respiratory syndrome coro .................... .................... .................... .................... .................... ....................
28841 MT614472.1 |Severe acute respiratory syndrome coro .................... .................... .................... .................... .................... ....................
28842 MT614473.1 |Severe acute respiratory syndrome coro .................... .................... .................... .................... .................... ....................
28843 MT614474.1 |Severe acute respiratory syndrome coro .................... .................... .................... .................... .................... ....................
28844 MT614475.1 |Severe acute respiratory syndrome coro .................... .................... .................... .................... .................... ....................
28845 MT614476.1 |Severe acute respiratory syndrome coro .................... ......A............. .................... .................... .................... ....................
28846 MT614477.1 |Severe acute respiratory syndrome coro .................... .................... .................... .................... .................... ....................
28847 MT614478.1 |Severe acute respiratory syndrome coro .................... .................... .................... .................... .................... ....................
28848 MT614479.1 |Severe acute respiratory syndrome coro .................... .................... .................... .................... .................... ....................
28849 MT614480.1 |Severe acute respiratory syndrome coro .................... .................... .................... .................... .................... ....................
28850 MT614481.1 |Severe acute respiratory syndrome coro .................... .................... .................... .................... .................... ....................
28851 MT614482.1 |Severe acute respiratory syndrome coro .................... .................... .................... .................... .................... ....................
28852 MT614483.1 |Severe acute respiratory syndrome coro .................... .................... .................... .................... .................... ....................
28853 MT614484.1 |Severe acute respiratory syndrome coro .................... .................... .................... .................... .................... ....................
28854 MT614485.1 |Severe acute respiratory syndrome coro .................... .................... .................... .................... .................... ....................
28855 MT614486.1 |Severe acute respiratory syndrome coro ...T................ .................... .................... .................... .................... ....................
28856 MT614487.1 |Severe acute respiratory syndrome coro .................... .................... .................... .................... .................... ....................
28857 MT614488.1 |Severe acute respiratory syndrome coro .................... .................... .................... .................... .................... ....................
28858 MT614489.1 |Severe acute respiratory syndrome coro .................... .................... .................... .................... .................... ....................
28859 MT614490.1 |Severe acute respiratory syndrome coro .................... .................... .................... .................... .................... ....................
28860 MT614491.1 |Severe acute respiratory syndrome coro .................... .................... .................... .................... .................... ....................
28861 MT614492.1 |Severe acute respiratory syndrome coro .................... .................... .................... .................... .................... ....................
28862 MT614493.1 |Severe acute respiratory syndrome coro .................... .................... .................... .................... .................... ....................
28863 MT614494.1 |Severe acute respiratory syndrome coro .................... .................... .................... .................... .................... ....................
28864 MT614495.1 |Severe acute respiratory syndrome coro .................... .................... .................... .................... .................... ....................
28865 MT614496.1 |Severe acute respiratory syndrome coro .................... .................... .................... .................... .................... ....................
28866 MT614497.1 |Severe acute respiratory syndrome coro .................... .................... .................... .................... .................... ....................
28867 MT614498.1 |Severe acute respiratory syndrome coro .................... .................... .................... .................... .................... ....................
28868 MT614499.1 |Severe acute respiratory syndrome coro .................... .................... .................... .................... .................... ....................
28869 MT614500.1 |Severe acute respiratory syndrome coro .................... .................... .................... .................... .................... ....................
28870 MT614501.1 |Severe acute respiratory syndrome coro .................... .................... .................... .................... .................... ....................
28871 MT614502.1 |Severe acute respiratory syndrome coro .................... .................... .................... .................... .................... ....................
28872 MT614503.1 |Severe acute respiratory syndrome coro .................... .................... .................... .................... .................... ....................
28873 MT614504.1 |Severe acute respiratory syndrome coro .................... .................... .................... .................... .................... ....................
28874 MT614505.1 |Severe acute respiratory syndrome coro .................... .................... .................... .................... .................... ....................
28875 MT614506.1 |Severe acute respiratory syndrome coro .................... .................... .................... .................... .................... ....................
28876 MT614507.1 |Severe acute respiratory syndrome coro .................... .................... .................... .................... .................... ....................
28877 MT614508.1 |Severe acute respiratory syndrome coro .................... .................... .................... .................... .................... ....................
28878 MT614509.1 |Severe acute respiratory syndrome coro .................... .................... .................... .................... .................... ....................
28879 MT614510.1 |Severe acute respiratory syndrome coro .................... .................... .................... .................... .................... ....................
28880 MT614512.1 |Severe acute respiratory syndrome coro .................... .................... .................... .................... .................... ....................
28881 MT614513.1 |Severe acute respiratory syndrome coro .................... .................... .................... .................... .................... ....................
28882 MT614514.1 |Severe acute respiratory syndrome coro .................... .................... .................... .................... .................... ....................
28883 MT614515.1 |Severe acute respiratory syndrome coro .................... .................... .................... .................... .................... ....................
28884 MT614516.1 |Severe acute respiratory syndrome coro .................... .................... .................... .................... .................... ....................
28885 MT614517.1 |Severe acute respiratory syndrome coro .................... .................... .................... .................... .................... ....................
28886 MT614519.1 |Severe acute respiratory syndrome coro .................... .................... .................... .................... .................... ....................
28887 MT614520.1 |Severe acute respiratory syndrome coro .................... .................... .................... .................... .................... ....................
28888 MT614521.1 |Severe acute respiratory syndrome coro .................... .................... .................... .................... .................... ....................
28889 MT614522.1 |Severe acute respiratory syndrome coro .................... .................... .................... .................... .................... ....................
28890 MT614523.1 |Severe acute respiratory syndrome coro .................... .................... .................... .................... .................... ....................
28891 MT614524.1 |Severe acute respiratory syndrome coro .................... .................... .................... .................... .................... ....................
28892 MT614525.1 |Severe acute respiratory syndrome coro .................... .................... .................... .................... .................... ....................
28893 MT614526.1 |Severe acute respiratory syndrome coro .................... .................... .................... .................... .................... ....................
28894 MT614527.1 |Severe acute respiratory syndrome coro .................... .................... .................... .................... .................... ....................
28895 MT614528.1 |Severe acute respiratory syndrome coro .................... .................... .................... .................... .................... ....................
28896 MT614529.1 |Severe acute respiratory syndrome coro .................... .................... .................... .................... .................... ....................
28897 MT614530.1 |Severe acute respiratory syndrome coro .................... .................... .................... .................... .................... ....................
28898 MT614531.1 |Severe acute respiratory syndrome coro .................... .................... .................... .................... .................... ....................
28899 MT614532.1 |Severe acute respiratory syndrome coro .................... .................... .................... .................... .................... ....................
28900 MT614533.1 |Severe acute respiratory syndrome coro .................... .................... .................... .................... .................... ....................
28901 MT614534.1 |Severe acute respiratory syndrome coro .................... .................... .................... .................... .................... ....................
28902 MT614535.1 |Severe acute respiratory syndrome coro .................... .................... .................... .................... .................... ....................
28903 MT614536.1 |Severe acute respiratory syndrome coro .................... .................... .................... .................... .................... ....................
28904 MT614537.1 |Severe acute respiratory syndrome coro .................... .................... .................... .................... .................... ....................
28905 MT614538.1 |Severe acute respiratory syndrome coro .................... .................... .................... .................... .................... ....................
28906 MT614539.1 |Severe acute respiratory syndrome coro .................... .................... .................... .................... .................... ....................
28907 MT614540.1 |Severe acute respiratory syndrome coro .................... .................... .................... .................... .................... ....................
28908 MT614541.1 |Severe acute respiratory syndrome coro .................... .................... .................... .................... .................... ....................
28909 MT614542.1 |Severe acute respiratory syndrome coro .................... .................... .................... .................... .................... ....................
28910 MT614543.1 |Severe acute respiratory syndrome coro .................... .................... .................... .................... .................... ....................
28911 MT614544.1 |Severe acute respiratory syndrome coro .................... .................... .................... .................... .................... ....................
28912 MT614545.1 |Severe acute respiratory syndrome coro .................... .................... .................... .................... .................... ....................
28913 MT614546.1 |Severe acute respiratory syndrome coro .................... .................... .................... .................... .................... ....................
28914 MT614547.1 |Severe acute respiratory syndrome coro .................... .................... .................... .................... .................... ....................
28915 MT614548.1 |Severe acute respiratory syndrome coro .................... .................... .................... .................... .................... ....................
28916 MT614549.1 |Severe acute respiratory syndrome coro .................... .................... .................... .................... .................... ....................

```
28917 MT614550.1 |Severe acute respiratory syndrome coro    ····················  ····················  ····················  ····················  ····················  ····················
28918 MT614551.1 |Severe acute respiratory syndrome coro    ····················  ····················  ····················  ····················  ····················  ····················
28919 MT614552.1 |Severe acute respiratory syndrome coro    ····················  ····················  ····················  ····················  ····················  ····················
28920 MT614553.1 |Severe acute respiratory syndrome coro    ····················  ····················  ····················  ····················  ····················  ····················
28921 MT614554.1 |Severe acute respiratory syndrome coro    ····················  ····················  ····················  ····················  ····················  ····················
28922 MT614555.1 |Severe acute respiratory syndrome coro    ····················  ····················  ····················  ····················  ····················  ····················
28923 MT614556.1 |Severe acute respiratory syndrome coro    ····················  ····················  ····················  ····················  ····················  ····················
28924 MT614557.1 |Severe acute respiratory syndrome coro    ····················  ····················  ····················  ····················  ····················  ····················
28925 MT614558.1 |Severe acute respiratory syndrome coro    ····················  ····················  ····················  ····················  ····················  ····················
28926 MT614559.1 |Severe acute respiratory syndrome coro    ····················  ····················  ····················  ····················  ····················  ····················
28927 MT614560.1 |Severe acute respiratory syndrome coro    ····················  ····················  ····················  ····················  ····················  ····················
28928 MT614561.1 |Severe acute respiratory syndrome coro    ····················  ····················  ····················  ····················  ····················  ····················
28929 MT614563.1 |Severe acute respiratory syndrome coro    ····················  ····················  ····················  ····················  ····················  ····················
28930 MT620766.1 |Severe acute respiratory syndrome coro    ····················  ····················  ····················  ····················  ····················  ····················
28931 MT620767.1 |Severe acute respiratory syndrome coro    ····················  ····················  ····················  ····················  ····················  ····················
28932 MT620769.1 |Severe acute respiratory syndrome coro    ····················  ····················  ····················  ····················  ····················  ····················
28933 MT620770.1 |Severe acute respiratory syndrome coro    ····················  ····················  ····················  ····················  ····················  ····················
28934 MT620771.1 |Severe acute respiratory syndrome coro    ····················  ····················  ····················  ····················  ····················  ····················
28935 MT620772.1 |Severe acute respiratory syndrome coro    ····················  ····················  ····················  ····················  ····················  ····················
28936 MT621560.1 |Severe acute respiratory syndrome coro    ····················  ····················  ····················  ····················  ····················  ····················
28937 MT622319.1 |Severe acute respiratory syndrome coro    ····················  ····················  ····················  ····················  ····················  ····················
28938 MT622321.1 |Severe acute respiratory syndrome coro    ····················  ····················  ····················  ····················  ····················  ····················
28939 MT365025.1 |Severe acute respiratory syndrome coro    ····················  ····················  ····················  ····················  ····················  ····················
28940 MT365026.1 |Severe acute respiratory syndrome coro    ····················  ····················  ····················  ····················  ····················  ····················
28941 MT365027.1 |Severe acute respiratory syndrome coro    ····················  ····················  ····················  ····················  ····················  ····················
28942 MT510690.1 |Severe acute respiratory syndrome coro    ····················  ······A·············  ····················  ····················  ····················  ····················
28943 MT510691.1 |Severe acute respiratory syndrome coro    ····················  ······R·············  ····················  ····················  ····················  ····················
28944 MT510692.1 |Severe acute respiratory syndrome coro    ····················  ······A·············  ····················  ····················  ····················  ····················
28945 MT510693.1 |Severe acute respiratory syndrome coro    ····················  ····················  ····················  ····················  ····················  ····················
28946 MT510694.1 |Severe acute respiratory syndrome coro    ····················  ····················  ····················  ····················  ····················  ····················
28947 MT510695.1 |Severe acute respiratory syndrome coro    ····················  ······A·············  ····················  ····················  ····················  ····················
28948 MT510696.1 |Severe acute respiratory syndrome coro    ····················  ······A·············  ····················  ····················  ····················  ····················
28949 MT510697.1 |Severe acute respiratory syndrome coro    ····················  ····················  ····················  ····················  ····················  ····················
28950 MT510698.1 |Severe acute respiratory syndrome coro    ····················  ····················  ····················  ····················  ····················  ····················
28951 MT510699.1 |Severe acute respiratory syndrome coro    ····················  ····················  ····················  ····················  ····················  ····················
28952 MT510700.1 |Severe acute respiratory syndrome coro    ····················  ······A·············  ····················  ····················  ····················  ····················
28953 MT510701.1 |Severe acute respiratory syndrome coro    ····················  ····················  ····················  ····················  ····················  ····················
28954 MT510702.1 |Severe acute respiratory syndrome coro    ····················  ····················  ····················  ····················  ····················  ····················
28955 MT510703.1 |Severe acute respiratory syndrome coro    ····················  ······R·············  ····················  ····················  ····················  ····················
28956 MT511059.1 |Severe acute respiratory syndrome coro    ····················  ····················  ····················  ····················  ····················  ····················
28957 MT511065.1 |Severe acute respiratory syndrome coro    ····················  ····················  ····················  ····················  ····················  ····················
28958 MT511083.1 |Severe acute respiratory syndrome coro    ····················  ····················  ····················  ····················  ····················  ····················
28959 MT511084.1 |Severe acute respiratory syndrome coro    ····················  ····················  ····················  ····················  ····················  ····················
28960 MT517443.1 |Severe acute respiratory syndrome coro    ····················  ····················  ····················  ····················  ····················  ····················
28961 MT605815.1 |Severe acute respiratory syndrome coro    ····················  ····················  ····················  ····················  ····················  ····················
28962 MT605816.1 |Severe acute respiratory syndrome coro    ····················  ····················  ····················  ····················  ····················  ····················
28963 MT605817.1 |Severe acute respiratory syndrome coro    ····················  ····················  ····················  ····················  ····················  ····················
28964 MT605818.1 |Severe acute respiratory syndrome coro    ····················  ····················  ····················  ····················  ····················  ····················
28965 MT607244.1 |Severe acute respiratory syndrome coro    ····················  ·A··················  ····················  ····················  ····················  ····················
28966 MT607245.1 |Severe acute respiratory syndrome coro    ····················  ····················  ····················  ····················  ····················  ····················
28967 MT607246.1 |Severe acute respiratory syndrome coro    ····················  ····················  ····················  ····················  ····················  ····················
28968 MT607247.1 |Severe acute respiratory syndrome coro    ····················  ····················  ····················  ····················  ····················  ····················
28969 MT607248.1 |Severe acute respiratory syndrome coro    ····················  ····················  ····················  ····················  ····················  ····················
28970 MT607249.1 |Severe acute respiratory syndrome coro    ····················  ····················  ····················  ····················  ····················  ····················
28971 MT607250.1 |Severe acute respiratory syndrome coro    ····················  ·A··················  ····················  ····················  ····················  ····················
28972 MT607251.1 |Severe acute respiratory syndrome coro    ····················  ····················  ····················  ····················  ····················  ····················
28973 MT607252.1 |Severe acute respiratory syndrome coro    ····················  ····················  ····················  ····················  ····················  ····················
28974 MT607253.1 |Severe acute respiratory syndrome coro    ····················  ····················  ····················  ····················  ····················  ····················
28975 MT607254.1 |Severe acute respiratory syndrome coro    ····················  ····················  ····················  ····················  ····················  ····················
28976 MT607255.1 |Severe acute respiratory syndrome coro    ····················  ····················  ····················  ····················  ····················  ····················
28977 MT607601.1 |Severe acute respiratory syndrome coro    ····················  ····················  ····················  ····················  ····················  ····················
28978 MT607602.1 |Severe acute respiratory syndrome coro    ····················  ·A··················  ····················  ····················  ····················  ···A················
28979 MT607603.1 |Severe acute respiratory syndrome coro    ····················  ·A··················  ····················  ····················  ····················  ····················
28980 MT607604.1 |Severe acute respiratory syndrome coro    ····················  ····················  ····················  ····················  ····················  ···A················
28981 MT607605.1 |Severe acute respiratory syndrome coro    ····················  ·A··················  ····················  ····················  ····················  ···A················
28982 MT607606.1 |Severe acute respiratory syndrome coro    ····················  ····················  ····················  ····················  ····················  ····················
28983 MT607607.1 |Severe acute respiratory syndrome coro    ····················  ····················  ····················  ····················  ····················  ···A················
28984 MT607608.1 |Severe acute respiratory syndrome coro    ····················  ·A··················  ····················  ····················  ····················  ···A················
28985 MT607609.1 |Severe acute respiratory syndrome coro    ····················  ·A··················  ····················  ····················  ····················  ···A················
28986 MT607610.1 |Severe acute respiratory syndrome coro    ····················  ·A··················  ····················  ····················  ····················  ···A················
28987 MT607611.1 |Severe acute respiratory syndrome coro    ····················  ·A··················  ····················  ····················  ····················  ···A················
28988 MT607612.1 |Severe acute respiratory syndrome coro    ····················  ····················  ····················  ····················  ····················  ····················
28989 MT607613.1 |Severe acute respiratory syndrome coro    ····················  ····················  ····················  ····················  ····················  ····················
28990 MT607614.1 |Severe acute respiratory syndrome coro    ····················  ·A··················  ····················  ····················  ····················  ···A················
28991 MT607615.1 |Severe acute respiratory syndrome coro    ····················  ····················  ····················  ····················  ····················  ····················
28992 MT607616.1 |Severe acute respiratory syndrome coro    ····················  ····················  ····················  ····················  ····················  ····················
28993 MT607617.1 |Severe acute respiratory syndrome coro    ····················  ·A··················  ····················  ····················  ····················  ···A················
28994 MT607618.1 |Severe acute respiratory syndrome coro    ····················  ····················  ····················  ····················  ····················  ····················
```

28995 MT607619.1 |Severe acute respiratory syndrome coro ···················· ···················· ····················· ···················· ···················· ····················
28996 MT607620.1 |Severe acute respiratory syndrome coro ···················· ·A·················· ····················· ···················· ···················· ····················
28997 MT607621.1 |Severe acute respiratory syndrome coro ···················· ···················· ····················· ···················· ···················· ····················
28998 MT608648.1 |Severe acute respiratory syndrome coro ···················· ·A·················· ····················· ···················· ···················· ···A················
28999 MT609558.1 |Severe acute respiratory syndrome coro ···················· ···················· ····················· ···················· ···················· ····················
29000 MT609559.1 |Severe acute respiratory syndrome coro ···················· ···················· ····················· ···················· ···················· ····················
29001 MT609560.1 |Severe acute respiratory syndrome coro ···················· ···················· ····················· ···················· ···················· ····················
29002 MT609561.1 |Severe acute respiratory syndrome coro ···················· ···················· ····················· ···················· ···················· ····················
29003 MT609562.1 |Severe acute respiratory syndrome coro ···················· ···················· ····················· ···················· ···················· ····················
29004 MT609563.1 |Severe acute respiratory syndrome coro ···················· ···················· ····················· ···················· ···················· ····················
29005 MT609564.1 |Severe acute respiratory syndrome coro ···················· ···················· ····················· ···················· ···················· ····················
29006 MT609565.1 |Severe acute respiratory syndrome coro ···················· ···················· ····················· ···················· ···················· ····················
29007 MT609566.1 |Severe acute respiratory syndrome coro ···················· ···················· ····················· ···················· ···················· ····················
29008 MT609567.1 |Severe acute respiratory syndrome coro ···················· ···················· ····················· ···················· ···················· ····················
29009 MT609568.1 |Severe acute respiratory syndrome coro ···················· ···················· ····················· ···················· ···················· ····················
29010 MT609569.1 |Severe acute respiratory syndrome coro ···················· ···················· ····················· ···················· ···················· ····················
29011 MT609570.1 |Severe acute respiratory syndrome coro ···················· ···················· ····················· ···················· ···················· ····················
29012 MT609571.1 |Severe acute respiratory syndrome coro ···················· ···················· ····················· ···················· ···················· ····················
29013 MT609572.1 |Severe acute respiratory syndrome coro ···················· ···················· ····················· ···················· ···················· ····················
29014 MT609573.1 |Severe acute respiratory syndrome coro ···················· ···················· ····················· ···················· ···················· ····················
29015 MT609574.1 |Severe acute respiratory syndrome coro ···················· ···················· ····················· ···················· ···················· ····················
29016 MT609575.1 |Severe acute respiratory syndrome coro ···················· ···················· ····················· ···················· ···················· ····················
29017 MT609576.1 |Severe acute respiratory syndrome coro ···················· ···················· ····················· ···················· ···················· ····················
29018 MT609577.1 |Severe acute respiratory syndrome coro ···················· ···················· ····················· ···················· ···················· ····················
29019 MT609578.1 |Severe acute respiratory syndrome coro ···················· ···················· ····················· ···················· ···················· ····················
29020 MT609579.1 |Severe acute respiratory syndrome coro ···················· ···················· ····················· ···················· ···················· ····················
29021 MT609580.1 |Severe acute respiratory syndrome coro ···················· ···················· ····················· ···················· ···················· ····················
29022 MT609581.1 |Severe acute respiratory syndrome coro ···················· ···················· ····················· ···················· ···················· ····················
29023 MT609582.1 |Severe acute respiratory syndrome coro ···················· ···················· ····················· ···················· ···················· ····················
29024 MT609583.1 |Severe acute respiratory syndrome coro ···················· ···················· ····················· ···················· ···················· ····················
29025 MT609584.1 |Severe acute respiratory syndrome coro ···················· ···················· ····················· ···················· ···················· ····················
29026 MT609585.1 |Severe acute respiratory syndrome coro ···················· ···················· ····················· ···················· ···················· ····················
29027 MT609586.1 |Severe acute respiratory syndrome coro ···················· ···················· ····················· ···················· ···················· ····················
29028 MT609587.1 |Severe acute respiratory syndrome coro ···················· ···················· ····················· ···················· ···················· ····················
29029 MT609588.1 |Severe acute respiratory syndrome coro ···················· ···················· ····················· ···················· ···················· ····················
29030 MT609589.1 |Severe acute respiratory syndrome coro ···················· ···················· ····················· ···················· ···················· ····················
29031 MT609590.1 |Severe acute respiratory syndrome coro ···················· ···················· ····················· ···················· ···················· ····················
29032 MT609591.1 |Severe acute respiratory syndrome coro ···················· ···················· ····················· ···················· ···················· ····················
29033 MT609592.1 |Severe acute respiratory syndrome coro ···················· ···················· ····················· ···················· ···················· ····················
29034 MT609593.1 |Severe acute respiratory syndrome coro ···················· ···················· ····················· ···················· ···················· ····················
29035 MT609594.1 |Severe acute respiratory syndrome coro ···················· ···················· ····················· ···················· ···················· ····················
29036 MT609595.1 |Severe acute respiratory syndrome coro ···················· ···················· ····················· ···················· ···················· ····················
29037 MT609596.1 |Severe acute respiratory syndrome coro ···················· ···················· ····················· ···················· ···················· ····················
29038 MT609597.1 |Severe acute respiratory syndrome coro ···················· ···················· ····················· ···················· ···················· ····················
29039 MT609598.1 |Severe acute respiratory syndrome coro ···················· ···················· ····················· ···················· ···················· ····················
29040 MT609599.1 |Severe acute respiratory syndrome coro ···················· ···················· ····················· ···················· ···················· ····················
29041 MT609600.1 |Severe acute respiratory syndrome coro ···················· ···················· ····················· ···················· ···················· ····················
29047 MT612138.1 |Severe acute respiratory syndrome coro ···················· ···················· ····················· ···················· ···················· ····················
29053 MT612153.1 |Severe acute respiratory syndrome coro ···················· ···················· ····················· ···················· ···················· ····················
29054 MT612154.1 |Severe acute respiratory syndrome coro ···················· ···················· ····················· ···················· ···················· ····················
29058 MT612170.1 |Severe acute respiratory syndrome coro ···················· ···················· ····················· ···················· ···················· ····················
29064 MT612253.1 |Severe acute respiratory syndrome coro ···················· ···················· ····················· ···················· ···················· ····················
29066 MT577627.1 |Severe acute respiratory syndrome coro ···················· ···················· ····················· ···················· ···················· ····················
29067 MT598171.1 |Severe acute respiratory syndrome coro ···················· ···················· ····················· ···················· ···················· ····················
29068 MT598172.1 |Severe acute respiratory syndrome coro ···················· ···················· ····················· ···················· ···················· ····················
29069 MT598173.1 |Severe acute respiratory syndrome coro ···················· ···················· ·······A············· ···················· ···················· ····················
29070 MT598174.1 |Severe acute respiratory syndrome coro ···················· ···················· ····················· ···················· ···················· ····················
29071 MT598175.1 |Severe acute respiratory syndrome coro ···················· ···················· ····················· ···················· ···················· ····················
29072 MT598176.1 |Severe acute respiratory syndrome coro ···················· ·A·················· ····················· ···················· ···················· ····················
29073 MT598177.1 |Severe acute respiratory syndrome coro ···················· ·A·················· ····················· ···················· ···················· ····················
29074 MT598178.1 |Severe acute respiratory syndrome coro ···················· ·A·················· ····················· ···················· ···················· ····················
29075 MT598633.1 |Severe acute respiratory syndrome coro ···················· ···················· ····················· ···················· ···················· ····················
29076 MT598634.1 |Severe acute respiratory syndrome coro ···················· ···················· ····················· ···················· ···················· ····················
29077 MT598636.1 |Severe acute respiratory syndrome coro ···················· ···················· ····················· ···················· ···················· ····················
29078 MT598639.1 |Severe acute respiratory syndrome coro ···················· ···················· ····················· ···················· ···················· ····················
29079 MT598640.1 |Severe acute respiratory syndrome coro ···················· ···················· ····················· ···················· ···················· ····················
29080 MT598641.1 |Severe acute respiratory syndrome coro ···················· ···················· ····················· ···················· ···················· ····················
29081 MT598642.1 |Severe acute respiratory syndrome coro ···················· ···················· ····················· ···················· ···················· ····················
29082 MT601275.1 |Severe acute respiratory syndrome coro ···················· ···················· ····················· ···················· ···················· ····················
29083 MT601276.1 |Severe acute respiratory syndrome coro ···················· ···················· ····················· ···················· ···················· ····················
29084 MT601277.1 |Severe acute respiratory syndrome coro ···················· ···················· ····················· ···················· ···················· ····················
29085 MT601278.1 |Severe acute respiratory syndrome coro ···················· ···················· ····················· ···················· ···················· ····················
29086 MT601279.1 |Severe acute respiratory syndrome coro ···················· ···················· ····················· ···················· ···················· ····················
29087 MT601280.1 |Severe acute respiratory syndrome coro ···················· ···················· ····················· ···················· ···················· ····················
29088 MT601281.1 |Severe acute respiratory syndrome coro ···················· ·A·················· ····················· ···················· ···················· ···A················
29089 MT601282.1 |Severe acute respiratory syndrome coro ···················· ·A·················· ····················· ···················· ···················· ···A················
29090 MT601283.1 |Severe acute respiratory syndrome coro ···················· ·A·················· ····················· ···················· ···················· ···A················
29091 MT601284.1 |Severe acute respiratory syndrome coro ···················· ···················· ····················· ···················· ···················· ····················

```
29092 MT601285.1 |Severe acute respiratory syndrome coro    ....................  ....................  ....................  ....................  ....................  ....................
29093 MT601286.1 |Severe acute respiratory syndrome coro    ....................  ....................  ....................  ....................  ....................  ....................
29094 MT601287.1 |Severe acute respiratory syndrome coro    ....................  ....................  ....................  ....................  ....................  ....................
29095 MT601288.1 |Severe acute respiratory syndrome coro    ....................  ....................  ....................  ....................  ....................  ....................
29096 MT601289.1 |Severe acute respiratory syndrome coro    ....................  ....................  ....................  ....................  ....................  ....................
29097 MT601290.1 |Severe acute respiratory syndrome coro    ....................  ....................  ....................  ....................  ....................  ....................
29098 MT601291.1 |Severe acute respiratory syndrome coro    ....................  ....................  ....................  ....................  ....................  ....................
29099 MT601292.1 |Severe acute respiratory syndrome coro    ....................  ....................  ....................  ....................  ....................  ....................
29100 MT601293.1 |Severe acute respiratory syndrome coro    ....................  ....................  ....................  ....................  ....................  ....................
29101 MT601294.1 |Severe acute respiratory syndrome coro    ....................  ....................  ....................  ....................  ....................  ....................
29102 MT601295.1 |Severe acute respiratory syndrome coro    ....................  ....................  ....................  ....................  ....................  ....................
29103 MT590598.1 |Severe acute respiratory syndrome coro    ....................  ....................  ....................  ....................  ....................  ....................
29104 MT590599.1 |Severe acute respiratory syndrome coro    ....................  ....................  ....................  ....................  ....................  ....................
29105 MT590600.1 |Severe acute respiratory syndrome coro    ....................  .A..................  ....................  ....................  ....................  ....................
29106 MT594109.1 |Severe acute respiratory syndrome coro    ....................  ....................  ....................  ....................  ....................  ....................
29107 MT594112.1 |Severe acute respiratory syndrome coro    ....................  .A..................  ....................  ....................  ....................  ...A................
29108 MT479223.1 |Severe acute respiratory syndrome coro    ....................  ....................  ....................  ....................  ....................  ....................
29109 MT479224.1 |Severe acute respiratory syndrome coro    ....................  ....................  ....................  ....................  ....................  ....................
29110 MT479225.1 |Severe acute respiratory syndrome coro    ....................  ....................  ....................  ....................  ....................  ....................
29111 MT479226.1 |Severe acute respiratory syndrome coro    ....................  ....................  ....................  ....................  ....................  ....................
29112 MT479227.1 |Severe acute respiratory syndrome coro    ....................  ....................  ....................  ....................  ....................  ....................
29113 MT584985.1 |Severe acute respiratory syndrome coro    ....................  ....................  ....................  ....................  ....................  ....................
29114 MT584997.1 |Severe acute respiratory syndrome coro    ....................  ....................  ....................  ....................  ....................  ....................
29115 MT584998.1 |Severe acute respiratory syndrome coro    ....................  ....................  ....................  ....................  ....................  ....................
29116 MT585003.1 |Severe acute respiratory syndrome coro    ....................  ....................  ....................  ....................  ....................  ....................
29117 MT585010.1 |Severe acute respiratory syndrome coro    ....................  ....................  ....................  ....................  ....................  ....................
29118 MT585011.1 |Severe acute respiratory syndrome coro    ....................  ....................  ....................  ....................  ....................  ....................
29119 MT585014.1 |Severe acute respiratory syndrome coro    ....................  ....................  ....................  ....................  ....................  ....................
29120 MT585019.1 |Severe acute respiratory syndrome coro    ....................  ....................  ....................  ....................  ....................  ....................
29121 MT585027.1 |Severe acute respiratory syndrome coro    ....................  ....................  ....................  ....................  ....................  ....................
29122 MT585032.1 |Severe acute respiratory syndrome coro    ....................  ....................  ....................  ....................  ....................  ....................
29123 MT585036.1 |Severe acute respiratory syndrome coro    ....................  ....................  ....................  ....................  ....................  ....................
29124 MT585039.1 |Severe acute respiratory syndrome coro    ....................  ....................  ....................  ....................  ....................  ....................
29125 MT585042.1 |Severe acute respiratory syndrome coro    ....................  ....................  ....................  ....................  ....................  ....................
29126 MT585044.1 |Severe acute respiratory syndrome coro    ....................  ....................  ....................  ....................  ....................  ....................
29127 MT585045.1 |Severe acute respiratory syndrome coro    ....................  ....................  ....................  ....................  ....................  ....................
29128 MT585048.1 |Severe acute respiratory syndrome coro    ....................  ....................  ....................  ....................  ....................  ....................
29129 MT585050.1 |Severe acute respiratory syndrome coro    ....................  ....................  ....................  ....................  ....................  ....................
29130 MT585052.1 |Severe acute respiratory syndrome coro    ....................  ....................  ....................  ....................  ....................  ....................
29131 MT585058.1 |Severe acute respiratory syndrome coro    ....................  ....................  ....................  ....................  ....................  ....................
29132 MT585059.1 |Severe acute respiratory syndrome coro    ....................  ....................  ....................  ....................  ....................  ....................
29133 MT585060.1 |Severe acute respiratory syndrome coro    ....................  ....................  ....................  ....................  ....................  ....................
29134 MT585061.1 |Severe acute respiratory syndrome coro    ....................  ....................  ....................  ....................  ....................  ....................
29135 MT585064.1 |Severe acute respiratory syndrome coro    ....................  ....................  ....................  ....................  ....................  ....................
29136 MT585066.1 |Severe acute respiratory syndrome coro    ....................  ....................  ....................  ....................  ....................  ....................
29137 MT585067.1 |Severe acute respiratory syndrome coro    ....................  ....................  ....................  ....................  ....................  ....................
29138 MT585071.1 |Severe acute respiratory syndrome coro    ....................  ....................  ....................  ....................  ....................  ....................
29139 MT585077.1 |Severe acute respiratory syndrome coro    ....................  ....................  ....................  ....................  ....................  ....................
29140 MT585082.1 |Severe acute respiratory syndrome coro    ....................  ....................  ....................  ....................  ....................  ....................
29141 MT585083.1 |Severe acute respiratory syndrome coro    ....................  ....................  ....................  ....................  ....................  ....................
29142 MT585084.1 |Severe acute respiratory syndrome coro    ....................  ....................  ....................  ....................  ....................  ....................
29143 MT585085.1 |Severe acute respiratory syndrome coro    ....................  ....................  ....................  ....................  ....................  ....................
29144 MT585086.1 |Severe acute respiratory syndrome coro    ....................  ....................  ....................  ....................  ....................  ....................
29145 MT585087.1 |Severe acute respiratory syndrome coro    ....................  ....................  ....................  ....................  ....................  ....................
29146 MT585088.1 |Severe acute respiratory syndrome coro    ....................  ....................  ....................  ....................  ....................  ....................
29147 MT585089.1 |Severe acute respiratory syndrome coro    ....................  ....................  ....................  ....................  ....................  ....................
29148 MT585090.1 |Severe acute respiratory syndrome coro    ....................  ....................  ....................  ....................  ....................  ....................
29149 MT585091.1 |Severe acute respiratory syndrome coro    ....................  ....................  ....................  ....................  ....................  ....................
29150 MT585092.1 |Severe acute respiratory syndrome coro    ....................  ....................  ....................  ....................  ....................  ....................
29151 MT585093.1 |Severe acute respiratory syndrome coro    ....................  ....................  ....................  ....................  ....................  ....................
29152 MT585094.1 |Severe acute respiratory syndrome coro    ....................  ....................  ....................  ....................  ....................  ....................
29153 MT585095.1 |Severe acute respiratory syndrome coro    ....................  ....................  ....................  ....................  ....................  ....................
29154 MT585096.1 |Severe acute respiratory syndrome coro    ....................  ....................  ....................  ....................  ....................  ....................
29155 MT585097.1 |Severe acute respiratory syndrome coro    ....................  ....................  ....................  ....................  ....................  ....................
29156 MT585098.1 |Severe acute respiratory syndrome coro    ....................  ....................  ....................  ....................  ....................  ....................
29157 MT585099.1 |Severe acute respiratory syndrome coro    ....................  ....................  ....................  ....................  ....................  ....................
29158 MT585100.1 |Severe acute respiratory syndrome coro    ....................  ....................  ....................  ....................  ....................  ....................
29159 MT585101.1 |Severe acute respiratory syndrome coro    ....................  ....................  ....................  ....................  ....................  ....................
29160 MT585102.1 |Severe acute respiratory syndrome coro    ....................  ....................  ....................  ....................  ....................  ....................
29161 MT585103.1 |Severe acute respiratory syndrome coro    ....................  ....................  ....................  ....................  ....................  ....................
29162 MT585104.1 |Severe acute respiratory syndrome coro    ....................  ....................  ....................  ....................  ....T...............  ....................
29163 MT577359.1 |Severe acute respiratory syndrome coro    ....................  ....................  ....................  ....................  ....................  ....................
29164 MT582453.1 |Severe acute respiratory syndrome coro    ....................  ....................  ....................  ....................  ....................  ....................
29165 MT582454.1 |Severe acute respiratory syndrome coro    ....................  ....................  ....................  ....................  ....................  ....................
29166 MT582456.1 |Severe acute respiratory syndrome coro    ....................  ....................  ....................  ....................  ....................  ....................
29167 MT582458.1 |Severe acute respiratory syndrome coro    ....................  ....................  ....................  ....................  ....................  ....................
29168 MT582459.1 |Severe acute respiratory syndrome coro    ....................  ....................  ....................  ....................  ....................  ....................
29169 MT582460.1 |Severe acute respiratory syndrome coro    ....................  ....................  ....................  ....................  ....................  ....................
```

| | | | | | | | | |
|---|---|---|---|---|---|---|---|---|
| 29170 | MT582461.1 | Severe acute respiratory syndrome coro | .................... | .................... | .................... | .................... | .................... | G................... |
| 29171 | MT582462.1 | Severe acute respiratory syndrome coro | .................... | .................... | .................... | .................... | .................... | .................... |
| 29172 | MT582463.1 | Severe acute respiratory syndrome coro | .................... | .................... | .................... | .................... | .................... | .................... |
| 29173 | MT582465.1 | Severe acute respiratory syndrome coro | .................... | .................... | .................... | .................... | .................... | .................... |
| 29174 | MT582466.1 | Severe acute respiratory syndrome coro | .................... | .................... | .................... | .................... | .................... | .................... |
| 29175 | MT582467.1 | Severe acute respiratory syndrome coro | .................... | .................... | .................... | .................... | .................... | .................... |
| 29176 | MT582468.1 | Severe acute respiratory syndrome coro | .................... | .................... | .................... | .................... | .................... | .................... |
| 29177 | MT582469.1 | Severe acute respiratory syndrome coro | .................... | .................... | .................... | .................... | .................... | .................... |
| 29178 | MT582471.1 | Severe acute respiratory syndrome coro | .................... | .................... | .................... | .................... | .................... | .................... |
| 29179 | MT582472.1 | Severe acute respiratory syndrome coro | .................... | .................... | .................... | .................... | .................... | .................... |
| 29180 | MT582473.1 | Severe acute respiratory syndrome coro | .................... | .................... | .................... | .................... | .................... | .................... |
| 29181 | MT582474.1 | Severe acute respiratory syndrome coro | .................... | .................... | .................... | .................... | .................... | .................... |
| 29182 | MT582475.1 | Severe acute respiratory syndrome coro | .................... | .................... | .................... | .................... | .................... | .................... |
| 29183 | MT582476.1 | Severe acute respiratory syndrome coro | .................... | .................... | .................... | .................... | .................... | .................... |
| 29184 | MT582477.1 | Severe acute respiratory syndrome coro | .................... | .................... | .................... | .................... | .................... | .................... |
| 29185 | MT582478.1 | Severe acute respiratory syndrome coro | .................... | .................... | .................... | .................... | .................... | .................... |
| 29186 | MT582479.1 | Severe acute respiratory syndrome coro | .................... | .................... | .................... | .................... | .................... | .................... |
| 29187 | MT582480.1 | Severe acute respiratory syndrome coro | .................... | .................... | .................... | .................... | .................... | .................... |
| 29188 | MT582481.1 | Severe acute respiratory syndrome coro | .................... | .................... | .................... | .................... | .................... | .................... |
| 29189 | MT582482.1 | Severe acute respiratory syndrome coro | .................... | .................... | .................... | .................... | .................... | .................... |
| 29190 | MT582483.1 | Severe acute respiratory syndrome coro | .................... | .................... | .................... | .................... | .................... | .................... |
| 29191 | MT582484.1 | Severe acute respiratory syndrome coro | .................... | .................... | .................... | .................... | .................... | .................... |
| 29192 | MT582485.1 | Severe acute respiratory syndrome coro | .................... | .................... | .................... | .................... | .................... | .................... |
| 29193 | MT582486.1 | Severe acute respiratory syndrome coro | .................... | .................... | .................... | .................... | .................... | .................... |
| 29194 | MT582487.1 | Severe acute respiratory syndrome coro | .................... | .................... | .................... | .................... | .................... | .................... |
| 29195 | MT582488.1 | Severe acute respiratory syndrome coro | .................... | .................... | .................... | .................... | .................... | .................... |
| 29196 | MT582491.1 | Severe acute respiratory syndrome coro | .................... | .................... | .................... | .................... | .................... | .................... |
| 29197 | MT582494.1 | Severe acute respiratory syndrome coro | .................... | .................... | .................... | .................... | .................... | .................... |
| 29198 | MT582496.1 | Severe acute respiratory syndrome coro | .................... | .................... | .................... | .................... | .................... | .................... |
| 29199 | MT582498.1 | Severe acute respiratory syndrome coro | .................... | .................... | .................... | .................... | .................... | .................... |
| 29200 | MT582499.1 | Severe acute respiratory syndrome coro | .................... | .................... | .................... | .................... | .................... | .................... |
| 29201 | MT499215.1 | Severe acute respiratory syndrome coro | .................... | .................... | .................... | .................... | .................... | .................... |
| 29202 | MT499217.1 | Severe acute respiratory syndrome coro | .................... | .................... | .................... | .................... | .................... | ............G....... |
| 29203 | MT499218.1 | Severe acute respiratory syndrome coro | .................... | .................... | .................... | .................... | .................... | .................... |
| 29204 | MT499219.1 | Severe acute respiratory syndrome coro | .................... | .................... | .................... | .................... | .................... | .................... |
| 29205 | MT499220.1 | Severe acute respiratory syndrome coro | .................... | .................... | .................... | .................... | .................... | .................... |
| 29206 | MT576025.1 | Severe acute respiratory syndrome coro | .................... | .................... | .................... | .................... | .................... | .................... |
| 29207 | MT576026.1 | Severe acute respiratory syndrome coro | .................... | .................... | .................... | .................... | .................... | .................... |
| 29208 | MT576027.1 | Severe acute respiratory syndrome coro | .................... | .................... | .................... | .................... | .................... | .................... |
| 29209 | MT576028.1 | Severe acute respiratory syndrome coro | .................... | .................... | .................... | .................... | .................... | .................... |
| 29210 | MT576029.1 | Severe acute respiratory syndrome coro | .................... | .................... | .................... | .................... | .................... | .................... |
| 29211 | MT576030.1 | Severe acute respiratory syndrome coro | .................... | .................... | .................... | .................... | .................... | .................... |
| 29212 | MT576031.1 | Severe acute respiratory syndrome coro | .................... | .................... | .................... | .................... | .................... | .................... |
| 29213 | MT576032.1 | Severe acute respiratory syndrome coro | .................... | .A.................. | .................... | .................... | .................... | .................... |
| 29214 | MT576033.1 | Severe acute respiratory syndrome coro | .................... | .A.................. | .................... | .................... | .................... | ...A................ |
| 29215 | MT576034.1 | Severe acute respiratory syndrome coro | .................... | .................... | .................... | .................... | .................... | .................... |
| 29216 | MT576035.1 | Severe acute respiratory syndrome coro | .................... | .................... | .................... | .................... | .................... | .................... |
| 29217 | MT576037.1 | Severe acute respiratory syndrome coro | .................... | .................... | .................... | .................... | .................... | .................... |
| 29218 | MT576038.1 | Severe acute respiratory syndrome coro | .................... | .................... | .................... | .................... | .................... | .................... |
| 29219 | MT576039.1 | Severe acute respiratory syndrome coro | .................... | .................... | .................... | .................... | .................... | .................... |
| 29220 | MT576040.1 | Severe acute respiratory syndrome coro | .................... | .................... | .................... | .................... | .................... | .................... |
| 29221 | MT576041.1 | Severe acute respiratory syndrome coro | .................... | .................... | .................... | .................... | .................... | .................... |
| 29222 | MT576042.1 | Severe acute respiratory syndrome coro | .................... | .................... | .................... | .................... | .................... | .................... |
| 29223 | MT576043.1 | Severe acute respiratory syndrome coro | .................... | .A.................. | .................... | .................... | .................... | ...A................ |
| 29224 | MT576044.1 | Severe acute respiratory syndrome coro | .................... | .A.................. | .................... | .................... | .................... | ...A................ |
| 29225 | MT576045.1 | Severe acute respiratory syndrome coro | .................... | .................... | .................... | .................... | .................... | .................... |
| 29226 | MT576046.1 | Severe acute respiratory syndrome coro | .................... | .................... | .................... | .................... | .................... | .................... |
| 29227 | MT576047.1 | Severe acute respiratory syndrome coro | .................... | .A.................. | .................... | .................... | .................... | .................... |
| 29228 | MT576048.1 | Severe acute respiratory syndrome coro | .................... | .A.................. | .................... | .................... | .................... | ...A................ |
| 29229 | MT576049.1 | Severe acute respiratory syndrome coro | .................... | .................... | .................... | .................... | .................... | .................... |
| 29230 | MT576052.1 | Severe acute respiratory syndrome coro | .................... | ...A................ | .................... | .................... | .................... | .................... |
| 29231 | MT576053.1 | Severe acute respiratory syndrome coro | .................... | .................... | .................... | .................... | .................... | ...A................ |
| 29232 | MT576054.1 | Severe acute respiratory syndrome coro | .................... | .A.................. | .................... | .................... | .................... | .................... |
| 29233 | MT576055.1 | Severe acute respiratory syndrome coro | .................... | .................... | .................... | .................... | .................... | .................... |
| 29234 | MT576056.1 | Severe acute respiratory syndrome coro | .................... | .A.................. | .................... | .................... | .................... | ...A................ |
| 29235 | MT576057.1 | Severe acute respiratory syndrome coro | .................... | .A.................. | .................... | .................... | .................... | ...A................ |
| 29236 | MT576058.1 | Severe acute respiratory syndrome coro | .................... | .A.................. | .................... | .................... | .................... | ...A................ |
| 29237 | MT576059.1 | Severe acute respiratory syndrome coro | .................... | .A.................. | .................... | .................... | .................... | ...A................ |
| 29238 | MT576060.1 | Severe acute respiratory syndrome coro | .................... | .A.................. | .................... | .................... | .................... | ...A................ |
| 29239 | MT576061.1 | Severe acute respiratory syndrome coro | .................... | .A.................. | .................... | .................... | .................... | ...A................ |
| 29240 | MT576529.1 | Severe acute respiratory syndrome coro | .................... | .A.................. | .................... | .................... | .................... | ...A................ |
| 29241 | MT576530.1 | Severe acute respiratory syndrome coro | .................... | .................... | .................... | .................... | .................... | .................... |
| 29242 | MT576531.1 | Severe acute respiratory syndrome coro | .................... | .A.................. | .................... | .................... | .................... | ...A................ |
| 29243 | MT576532.1 | Severe acute respiratory syndrome coro | .................... | .A.................. | .................... | .................... | .................... | ...A................ |
| 29244 | MT576584.1 | Severe acute respiratory syndrome coro | .................... | .................... | .................... | .................... | .................... | .................... |
| 29245 | MT576639.1 | Severe acute respiratory syndrome coro | .................... | .................... | .................... | .................... | .................... | .................... |
| 29246 | MT576640.1 | Severe acute respiratory syndrome coro | .................... | .................... | .................... | .................... | .................... | .................... |
| 29247 | MT576641.1 | Severe acute respiratory syndrome coro | .................... | .................... | .................... | .................... | .................... | .................... |

```
29248 MT576642.1 |Severe acute respiratory syndrome coro    ....................  ....................  ....................  ....................  ....................  ....................
29249 MT576643.1 |Severe acute respiratory syndrome coro    ....................  ....................  ....................  ....................  ....................  ....................
29250 MT576644.1 |Severe acute respiratory syndrome coro    ....................  ....................  ....................  ....................  ....................  ....................
29251 MT576645.1 |Severe acute respiratory syndrome coro    ....................  ....................  ....................  ....................  ....................  ....................
29252 MT576689.1 |Severe acute respiratory syndrome coro    ....................  ....................  ....................  ....................  ....................  ....................
29253 MT578015.1 |Severe acute respiratory syndrome coro    ....................  ....................  ....................  ....................  ....................  ....................
29254 MT578016.1 |Severe acute respiratory syndrome coro    ....................  ....................  ....................  ....................  ....................  ....................
29255 MT578017.1 |Severe acute respiratory syndrome coro    ....................  ....................  ....................  ....................  ....................  ....................
29256 LC553257.1 |Severe acute respiratory syndrome coro    ....................  ....................  ....................  ....................  ....................  ....................
29257 LC553258.1 |Severe acute respiratory syndrome coro    ....................  ....................  ....................  ....................  ....................  ....................
29258 LC553259.2 |Severe acute respiratory syndrome coro    ....................  ....................  ....................  ....................  ....................  ....................
29259 LC553260.1 |Severe acute respiratory syndrome coro    ....................  ....................  ....................  ....................  ....................  ....................
29260 LC553261.1 |Severe acute respiratory syndrome coro    ....................  ....................  ....................  ....................  ....................  ....................
29261 LC553262.1 |Severe acute respiratory syndrome coro    ....................  ....................  ....................  ....................  ....................  ....................
29262 LC553263.1 |Severe acute respiratory syndrome coro    ....................  ....................  ....................  ....................  ....................  ....................
29263 LC549340.1 |Severe acute respiratory syndrome coro    ....................  ....................  ....................  ....................  ....................  ....................
29264 MT568634.1 |Severe acute respiratory syndrome coro    ....................  ....................  ....................  ....................  ....................  ....................
29265 MT568635.1 |Severe acute respiratory syndrome coro    ....................  ....................  ....................  ....................  ....................  ....................
29266 MT568636.1 |Severe acute respiratory syndrome coro    ....................  ....................  ....................  ....................  ....................  ....................
29267 MT568637.1 |Severe acute respiratory syndrome coro    ....................  ....................  ....................  ....................  ....................  ....................
29268 MT568638.1 |Severe acute respiratory syndrome coro    ....................  ....................  ....................  ....................  ....................  ....................
29269 MT568639.1 |Severe acute respiratory syndrome coro    ....................  ....................  ....................  ....................  ....................  ....................
29270 MT568640.1 |Severe acute respiratory syndrome coro    ....................  ....................  ....................  ....................  ....................  ....................
29271 MT568641.1 |Severe acute respiratory syndrome coro    ....................  ....................  ....................  ....................  ....................  ....................
29272 MT568645.1 |Severe acute respiratory syndrome coro    ....................  ....................  ....................  ....................  ....................  ....................
29273 MT569470.1 |Severe acute respiratory syndrome coro    ....................  ....................  ....................  ....................  ....................  ....................
29274 MT569471.1 |Severe acute respiratory syndrome coro    ....................  ....................  ....................  ....................  ....................  ....................
29275 MT569472.1 |Severe acute respiratory syndrome coro    ....................  ....................  ....................  ....................  ....................  ....................
29276 MT569473.1 |Severe acute respiratory syndrome coro    ....................  ....................  ....................  ....................  ....................  ....................
29277 MT569474.1 |Severe acute respiratory syndrome coro    ....................  ....................  ....................  ....................  ....................  ....................
29278 MT520172.1 |Severe acute respiratory syndrome coro    ....................  ....................  ....................  ....................  ....................  ....................
29279 MT520173.1 |Severe acute respiratory syndrome coro    ....................  ....................  ....................  ....................  ....................  ....................
29280 MT520174.1 |Severe acute respiratory syndrome coro    ....................  ....................  ....................  ....................  ....................  ....................
29281 MT520176.1 |Severe acute respiratory syndrome coro    ....................  ....................  ....................  ....................  ....................  ....................
29282 MT520177.1 |Severe acute respiratory syndrome coro    ....................  ....................  ....................  ....................  ....................  ....................
29283 MT520178.1 |Severe acute respiratory syndrome coro    ....................  ....................  ....................  ....................  ....................  ....................
29284 MT520180.1 |Severe acute respiratory syndrome coro    ....................  ....................  ....................  ....................  ....................  ....................
29285 MT520181.1 |Severe acute respiratory syndrome coro    ....................  ....................  ....................  ....................  ....................  ....................
29286 MT520182.1 |Severe acute respiratory syndrome coro    ....................  ....................  ....................  ....................  ....................  ....................
29287 MT520183.1 |Severe acute respiratory syndrome coro    ....................  ....................  ....................  ....................  ....................  ....................
29288 MT520184.1 |Severe acute respiratory syndrome coro    ....................  ....................  ....................  ....................  ....................  ....................
29289 MT520185.1 |Severe acute respiratory syndrome coro    ....................  ....................  ....................  ....................  ....................  ....................
29290 MT520186.1 |Severe acute respiratory syndrome coro    ....................  ....................  ....................  ....................  ....................  ....................
29291 MT520187.1 |Severe acute respiratory syndrome coro    ....................  ....................  ....................  ....................  ....................  ....................
29292 MT520188.1 |Severe acute respiratory syndrome coro    ....................  ....................  ....................  ....................  ....................  ....................
29293 MT520189.1 |Severe acute respiratory syndrome coro    ....................  ....................  ....................  ....................  ....................  ....................
29294 MT520191.1 |Severe acute respiratory syndrome coro    ....................  ....................  ....................  ....................  ....................  ....................
29295 MT520192.1 |Severe acute respiratory syndrome coro    ....................  ....................  ....................  ....................  ....................  ....................
29296 MT520193.1 |Severe acute respiratory syndrome coro    ....................  ....................  ....................  ....................  ....................  ....................
29297 MT520194.1 |Severe acute respiratory syndrome coro    ....................  ....................  ....................  ....................  ....................  ....................
29298 MT520195.1 |Severe acute respiratory syndrome coro    ....................  ....................  ....................  ....................  ....................  ....................
29299 MT520196.1 |Severe acute respiratory syndrome coro    ....................  ....................  ....................  ....................  ....................  ....................
29300 MT520197.1 |Severe acute respiratory syndrome coro    ....................  ....................  ....................  ....................  ....................  ....................
29301 MT520198.1 |Severe acute respiratory syndrome coro    ....................  ....................  ....................  ....................  ....................  ....................
29302 MT520199.1 |Severe acute respiratory syndrome coro    ....................  ....................  ....................  ....................  ....................  ....................
29303 MT520200.1 |Severe acute respiratory syndrome coro    ....................  ....................  ....................  ....................  ....................  ....................
29304 MT520201.1 |Severe acute respiratory syndrome coro    ....................  ....................  ....................  ....................  ....................  ....................
29305 MT520203.1 |Severe acute respiratory syndrome coro    ....................  ....................  ....................  ....................  ....................  ....................
29306 MT520204.1 |Severe acute respiratory syndrome coro    ....................  ....................  ....................  ....................  ....................  ....................
29307 MT520205.1 |Severe acute respiratory syndrome coro    ....................  ....................  ....................  ....................  ....................  ....................
29308 MT520206.1 |Severe acute respiratory syndrome coro    ....................  ....................  ....................  ....................  ....................  ....................
29309 MT520207.1 |Severe acute respiratory syndrome coro    ....................  ....................  ....................  ....................  ....................  ....................
29310 MT520208.1 |Severe acute respiratory syndrome coro    ....................  ....................  ....................  ....................  ....................  ....................
29311 MT520209.1 |Severe acute respiratory syndrome coro    ....................  ....................  ....................  ....................  ....................  ....................
29312 MT520210.1 |Severe acute respiratory syndrome coro    ....................  ....................  ....................  ....................  ....................  ....................
29313 MT520212.1 |Severe acute respiratory syndrome coro    ....................  ....................  ....................  ....................  ....................  ....................
29314 MT520213.1 |Severe acute respiratory syndrome coro    ....................  ....................  ....................  ....................  ....................  ....................
29315 MT520214.1 |Severe acute respiratory syndrome coro    ....................  ....................  ....................  ....................  ....................  ....................
29316 MT520215.1 |Severe acute respiratory syndrome coro    ....................  ....................  ....................  ....................  ....................  ....................
29317 MT520216.1 |Severe acute respiratory syndrome coro    ....................  ....................  ....................  ....................  ....................  ....................
29318 MT520217.1 |Severe acute respiratory syndrome coro    ....................  ....................  ....................  ....................  ....................  ....................
29319 MT520218.1 |Severe acute respiratory syndrome coro    ....................  ....................  ....................  ....................  ....................  ....................
29320 MT520219.1 |Severe acute respiratory syndrome coro    ....................  ....................  ....................  ....................  ....................  ....................
29321 MT520220.1 |Severe acute respiratory syndrome coro    ....................  ....................  ....................  ....................  ....................  ....................
29322 MT520221.1 |Severe acute respiratory syndrome coro    ....................  ....................  ....................  ....................  ....................  ....................
29323 MT520222.1 |Severe acute respiratory syndrome coro    ....................  ....................  ....................  ....................  ....................  ....................
29324 MT520223.1 |Severe acute respiratory syndrome coro    ....................  ....................  ....................  ....................  ....................  ....................
29325 MT520224.1 |Severe acute respiratory syndrome coro    ....................  ....................  ....................  ....................  ....................  ....................
```

29326 MT520225.1 |Severe acute respiratory syndrome coro .................... .................... ..................... .................... .................... ....................
29327 MT520228.1 |Severe acute respiratory syndrome coro .................... ......A............. ..................... .................... .................... ....................
29328 MT520229.1 |Severe acute respiratory syndrome coro .................... .................... ..................... .................... .................... ....................
29329 MT520230.1 |Severe acute respiratory syndrome coro .................... .................... ..................... .................... .................... ....................
29330 MT520231.1 |Severe acute respiratory syndrome coro .................... .................... ..................... .................... .................... ....................
29331 MT520232.1 |Severe acute respiratory syndrome coro .................... .................... ..................... .................... .................... ....................
29332 MT520233.1 |Severe acute respiratory syndrome coro .................... .................... ..................... .................... .................... ....................
29333 MT520234.1 |Severe acute respiratory syndrome coro .................... .................... ..................... .................... .................... ....................
29334 MT520235.1 |Severe acute respiratory syndrome coro .................... .................... ..................... .................... .................... ....................
29335 MT520236.1 |Severe acute respiratory syndrome coro .................... .................... ..................... .................... .................... ....................
29336 MT520237.1 |Severe acute respiratory syndrome coro .................... .................... ..................... .................... .................... ....................
29337 MT520238.1 |Severe acute respiratory syndrome coro .................... .................... ..................... .................... .................... ....................
29338 MT520239.1 |Severe acute respiratory syndrome coro .................... .................... ..................... .................... .................... ....................
29339 MT520240.1 |Severe acute respiratory syndrome coro .................... .................... ..................... .................... .................... ....................
29340 MT520243.1 |Severe acute respiratory syndrome coro .................... .................... ..................... .................... .................... ....................
29341 MT520244.1 |Severe acute respiratory syndrome coro .................... .................... ..................... .................... .................... ....................
29342 MT520245.1 |Severe acute respiratory syndrome coro .................... .................... ..................... .................... .................... ....................
29343 MT520246.1 |Severe acute respiratory syndrome coro .................... .................... ..................... .................... .................... ....................
29344 MT520247.1 |Severe acute respiratory syndrome coro .................... .................... ..................... .................... .................... ....................
29345 MT520248.1 |Severe acute respiratory syndrome coro .................... .................... ..................... .................... .................... ....................
29346 MT520249.1 |Severe acute respiratory syndrome coro .................... .................... ..................... .................... .................... ....................
29347 MT520250.1 |Severe acute respiratory syndrome coro .................... .................... ..................... .................... .................... ....................
29348 MT520251.1 |Severe acute respiratory syndrome coro .................... .................... ..................... .................... .................... ....................
29349 MT520252.1 |Severe acute respiratory syndrome coro .................... .................... ..................... .................... .................... ....................
29350 MT520253.1 |Severe acute respiratory syndrome coro .................... .................... ..................... .................... .................... ....................
29351 MT520254.1 |Severe acute respiratory syndrome coro .................... .................... ..................... .................... .................... ....................
29352 MT520255.1 |Severe acute respiratory syndrome coro .................... .................... ..................... .................... .................... ....................
29353 MT520256.1 |Severe acute respiratory syndrome coro .................... .................... ..................... .................... .................... ....................
29354 MT520257.1 |Severe acute respiratory syndrome coro .................... .................... ..................... .................... .................... ....................
29355 MT520258.1 |Severe acute respiratory syndrome coro .................... .................... ..................... .................... .................... ....................
29356 MT520259.1 |Severe acute respiratory syndrome coro .................... .................... ..................... .................... .................... ....................
29357 MT520260.1 |Severe acute respiratory syndrome coro .................... .................... ..................... .................... .................... ....................
29358 MT520261.1 |Severe acute respiratory syndrome coro .................... .................... ..................... .................... .................... ....................
29359 MT520262.1 |Severe acute respiratory syndrome coro .................... .................... ..................... .................... .................... ....................
29360 MT520263.1 |Severe acute respiratory syndrome coro .................... .................... ..................... .................... .................... ....................
29361 MT520264.1 |Severe acute respiratory syndrome coro .................... .................... ..................... .................... .................... ....................
29362 MT520265.1 |Severe acute respiratory syndrome coro .................... .................... ..................... .................... .................... ....................
29363 MT520266.1 |Severe acute respiratory syndrome coro .................... .................... ..................... .................... .................... ....................
29364 MT520269.1 |Severe acute respiratory syndrome coro .................... .................... ..................... .................... .................... ....................
29365 MT520270.1 |Severe acute respiratory syndrome coro .................... .................... ..................... .................... .................... ....................
29366 MT520271.1 |Severe acute respiratory syndrome coro .................... .................... ..................... .................... .................... ....................
29367 MT520272.1 |Severe acute respiratory syndrome coro .................... .................... ..................... .................... .................... ....................
29368 MT520273.1 |Severe acute respiratory syndrome coro .................... .................... ..................... .................... .................... ....................
29369 MT520276.1 |Severe acute respiratory syndrome coro .................... .................... ..................... .................... .................... ....................
29370 MT520277.1 |Severe acute respiratory syndrome coro .................... .................... ..................... .................... .................... ....................
29371 MT520278.1 |Severe acute respiratory syndrome coro .................... .................... ..................... .................... .................... ....................
29372 MT520279.1 |Severe acute respiratory syndrome coro .................... .................... ..................... .................... .................... ....................
29373 MT520280.1 |Severe acute respiratory syndrome coro .................... .................... ..................... .................... .................... ....................
29374 MT520281.1 |Severe acute respiratory syndrome coro .................... .................... ..................... .................... .................... ....................
29375 MT520283.1 |Severe acute respiratory syndrome coro .................... .................... ..................... .................... .................... ....................
29376 MT520284.1 |Severe acute respiratory syndrome coro .................... .................... ..................... .................... .................... ....................
29377 MT520285.1 |Severe acute respiratory syndrome coro .................... .................... ..................... .................... .................... ....................
29378 MT520287.1 |Severe acute respiratory syndrome coro .................... .................... ..................... .................... .................... ....................
29379 MT520288.1 |Severe acute respiratory syndrome coro .................... .................... ..................... .................... .................... ....................
29380 MT520289.1 |Severe acute respiratory syndrome coro .................... .................... ..................... .................... .................... ....................
29381 MT520290.1 |Severe acute respiratory syndrome coro .................... .................... ..................... .................... .................... ....................
29382 MT520291.1 |Severe acute respiratory syndrome coro .................... .................... ..................... .................... .................... ....................
29383 MT520292.1 |Severe acute respiratory syndrome coro .................... .................... ..................... .................... .................... ....................
29384 MT520295.1 |Severe acute respiratory syndrome coro .................... .................... ..................... .................... .................... ....................
29385 MT520296.1 |Severe acute respiratory syndrome coro .................... .................... ..................... .................... .................... ....................
29386 MT520298.1 |Severe acute respiratory syndrome coro .................... .................... ..................... .................... .................... ....................
29387 MT520299.1 |Severe acute respiratory syndrome coro .................... .................... ..................... .................... .................... ....................
29388 MT520300.1 |Severe acute respiratory syndrome coro .................... .................... ..................... .................... .................... ....................
29389 MT520301.1 |Severe acute respiratory syndrome coro .................... .................... ..................... .................... .................... ....................
29390 MT520302.1 |Severe acute respiratory syndrome coro .................... .................... ..................... .................... .................... ....................
29391 MT520304.1 |Severe acute respiratory syndrome coro .................... .................... ..................... .................... .................... ....................
29392 MT520305.1 |Severe acute respiratory syndrome coro .................... .................... ..................... .................... .................... ....................
29393 MT520306.1 |Severe acute respiratory syndrome coro .................... .................... ..................... .................... .................... ....................
29394 MT520307.1 |Severe acute respiratory syndrome coro .................... .................... ..................... .................... .................... ....................
29395 MT520308.1 |Severe acute respiratory syndrome coro .................... .................... ..................... .................... .................... ....................
29396 MT520309.1 |Severe acute respiratory syndrome coro .................... .................... ..................... .................... .................... ....................
29397 MT520310.1 |Severe acute respiratory syndrome coro .................... .................... ..................... .................... .................... ....................
29398 MT520312.1 |Severe acute respiratory syndrome coro .................... .................... ..................... .................... .................... ....................
29399 MT520313.1 |Severe acute respiratory syndrome coro .................... .................... ..................... .................... .................... ....................
29400 MT520314.1 |Severe acute respiratory syndrome coro .................... .................... ..................... .................... .................... ....................
29401 MT520315.1 |Severe acute respiratory syndrome coro .................... .................... ..................... .................... .................... ....................
29402 MT520316.1 |Severe acute respiratory syndrome coro .................... .................... ..................... .................... .................... ....................
29403 MT520317.1 |Severe acute respiratory syndrome coro .................... .................... ..................... .................... .................... ....................

```
29404 MT520318.1 |Severe acute respiratory syndrome coro    ....................  ....................  .....................  ....................  ....................  ....................
29405 MT520319.1 |Severe acute respiratory syndrome coro    ....................  ....................  .....................  ....................  ....................  ....................
29406 MT520320.1 |Severe acute respiratory syndrome coro    ....................  ....................  .....................  ....................  ....................  ....................
29407 MT520321.1 |Severe acute respiratory syndrome coro    ....................  ....................  .....................  ....................  ....................  ....................
29408 MT520323.1 |Severe acute respiratory syndrome coro    ....................  ....................  .....................  ....................  ....................  ....................
29409 MT520324.1 |Severe acute respiratory syndrome coro    ....................  ....................  .....................  ....................  ....................  ....................
29410 MT520325.1 |Severe acute respiratory syndrome coro    ....................  ....................  .....................  ....................  ....................  ....................
29411 MT520327.1 |Severe acute respiratory syndrome coro    ....................  ....................  .....................  ....................  ....................  ....................
29412 MT520328.1 |Severe acute respiratory syndrome coro    ....................  ....................  .....................  ....................  ....................  ....................
29413 MT520329.1 |Severe acute respiratory syndrome coro    ....................  ....................  .....................  ....................  ....................  ....................
29414 MT520330.1 |Severe acute respiratory syndrome coro    ....................  ....................  .....................  ....................  ....................  ....................
29415 MT520331.1 |Severe acute respiratory syndrome coro    ....................  ....................  .....................  ....................  ....................  ....................
29416 MT520332.1 |Severe acute respiratory syndrome coro    ....................  ....................  .....................  ....................  ....................  ....................
29417 MT520333.1 |Severe acute respiratory syndrome coro    ....................  ....................  .....................  ....................  ....................  ....................
29418 MT520336.1 |Severe acute respiratory syndrome coro    ....................  ....................  .....................  ....................  ....................  ....................
29419 MT520337.1 |Severe acute respiratory syndrome coro    ....................  ....................  .....................  ....................  ....................  ....................
29420 MT520338.1 |Severe acute respiratory syndrome coro    ....................  ....................  .....................  ....................  ....................  ....................
29421 MT520339.1 |Severe acute respiratory syndrome coro    ....................  ....................  .....................  ....................  ....................  ....................
29422 MT520340.1 |Severe acute respiratory syndrome coro    ....................  ....................  .....................  ....................  ....................  ....................
29423 MT520341.1 |Severe acute respiratory syndrome coro    ....................  ....................  .....................  ....................  ....................  ....................
29424 MT520342.1 |Severe acute respiratory syndrome coro    ....................  ....................  .....................  ....................  ....................  ....................
29425 MT520343.1 |Severe acute respiratory syndrome coro    ....................  ....................  .....................  ....................  ....................  ....................
29426 MT520344.1 |Severe acute respiratory syndrome coro    ....................  ....................  .....................  ....................  ....................  ....................
29427 MT520345.1 |Severe acute respiratory syndrome coro    ....................  ....................  .....................  ....................  ....................  ....................
29428 MT520346.1 |Severe acute respiratory syndrome coro    ....................  ....................  .....................  ....................  ....................  ....................
29429 MT520347.1 |Severe acute respiratory syndrome coro    ....................  ....................  .....................  ....................  ....................  ....................
29430 MT520348.1 |Severe acute respiratory syndrome coro    ....................  ....................  .....................  ....................  ....................  ....................
29431 MT520349.1 |Severe acute respiratory syndrome coro    ....................  ....................  .....................  ....................  ....................  ....................
29432 MT520350.1 |Severe acute respiratory syndrome coro    ....................  ....................  .....................  ....................  ....................  ....................
29433 MT520351.1 |Severe acute respiratory syndrome coro    ....................  ....................  .....................  ....................  ....................  ....................
29434 MT520352.1 |Severe acute respiratory syndrome coro    ....................  ....................  .....................  ....................  ....................  ....................
29435 MT520353.1 |Severe acute respiratory syndrome coro    ....................  ....................  .....................  ....................  ....................  ....................
29436 MT520355.1 |Severe acute respiratory syndrome coro    ....................  ....................  .....................  ....................  ....................  ....................
29437 MT520356.1 |Severe acute respiratory syndrome coro    ....................  ....................  .....................  ....................  ....................  ....................
29438 MT520357.1 |Severe acute respiratory syndrome coro    ....................  ....................  .....................  ....................  ....................  ....................
29439 MT520358.1 |Severe acute respiratory syndrome coro    ....................  ....................  .....................  ....................  ....................  ....................
29440 MT520359.1 |Severe acute respiratory syndrome coro    ....................  ....................  .....................  ....................  ....................  ....................
29441 MT520360.1 |Severe acute respiratory syndrome coro    ....................  ....................  .....................  ....................  ....................  ....................
29442 MT520361.1 |Severe acute respiratory syndrome coro    ....................  ....................  .....................  ....................  ....................  ....................
29443 MT520362.1 |Severe acute respiratory syndrome coro    ....................  ....................  .....................  ....................  ....................  ....................
29444 MT520363.1 |Severe acute respiratory syndrome coro    ....................  ....................  .....................  ....................  ....................  ....................
29445 MT520364.1 |Severe acute respiratory syndrome coro    ....................  ....................  .....................  ....................  ....................  ....................
29446 MT520365.1 |Severe acute respiratory syndrome coro    ....................  ....................  .....................  ....................  ....................  ....................
29447 MT520366.1 |Severe acute respiratory syndrome coro    ....................  ....................  .....................  ....................  ....................  ....................
29448 MT520367.1 |Severe acute respiratory syndrome coro    ....................  ....................  .....................  ....................  ....................  ....................
29449 MT520368.1 |Severe acute respiratory syndrome coro    ....................  ....................  .....................  ....................  ....................  ....................
29450 MT520369.1 |Severe acute respiratory syndrome coro    ....................  ....................  .....................  ....................  ....................  ....................
29451 MT520370.1 |Severe acute respiratory syndrome coro    ....................  ....................  .....................  ....................  ....................  ....................
29452 MT520371.1 |Severe acute respiratory syndrome coro    ....................  ....................  .....................  ....................  ....................  ....................
29453 MT520372.1 |Severe acute respiratory syndrome coro    ....................  ....................  .....................  ....................  ....................  ....................
29454 MT520373.1 |Severe acute respiratory syndrome coro    ....................  ....................  .....................  ....................  ....................  ....................
29455 MT520374.1 |Severe acute respiratory syndrome coro    ....................  ....................  .....................  ....................  ....................  ....................
29456 MT520375.1 |Severe acute respiratory syndrome coro    ....................  ....................  .....................  ....................  ....................  ....................
29457 MT520376.1 |Severe acute respiratory syndrome coro    ....................  ....................  .....................  ....................  ....................  ....................
29458 MT520377.1 |Severe acute respiratory syndrome coro    ....................  ....................  .....................  ....................  ....................  ....................
29459 MT520379.1 |Severe acute respiratory syndrome coro    ....................  ....................  .....................  ....................  ....................  ....................
29460 MT520380.1 |Severe acute respiratory syndrome coro    ....................  ....................  .....................  ....................  ....................  ....................
29461 MT520381.1 |Severe acute respiratory syndrome coro    ....................  ....................  .....................  ....................  ....................  ....................
29462 MT520382.1 |Severe acute respiratory syndrome coro    ....................  ....................  .....................  ....................  ....................  ....................
29463 MT520383.1 |Severe acute respiratory syndrome coro    ....................  ....................  .....................  ....................  ....................  ....................
29464 MT520384.1 |Severe acute respiratory syndrome coro    ....................  ....................  .....................  ....................  ....................  ....................
29465 MT520385.1 |Severe acute respiratory syndrome coro    ....................  ....................  .....................  ....................  ....................  ....................
29466 MT520386.1 |Severe acute respiratory syndrome coro    ....................  ....................  .....................  ....................  ....................  ....................
29467 MT520387.1 |Severe acute respiratory syndrome coro    ....................  ....................  .....................  ....................  ....................  ....................
29468 MT520388.1 |Severe acute respiratory syndrome coro    ....................  ....................  .....................  ....................  ....................  ....................
29469 MT520389.1 |Severe acute respiratory syndrome coro    ....................  ....................  .....................  ....................  ....................  ....................
29470 MT520390.1 |Severe acute respiratory syndrome coro    ....................  ....................  .....................  ....................  ....................  ....................
29471 MT520391.1 |Severe acute respiratory syndrome coro    ....................  ....................  .....................  ....................  ....................  ....................
29472 MT520392.1 |Severe acute respiratory syndrome coro    ....................  ....................  .....................  ....................  ....................  ....................
29473 MT520394.1 |Severe acute respiratory syndrome coro    ....................  ....................  .....................  ....................  ....................  ....................
29474 MT520396.1 |Severe acute respiratory syndrome coro    ....................  ....................  .....................  ....................  ....................  ....................
29475 MT520397.1 |Severe acute respiratory syndrome coro    ....................  ....................  .....................  ....................  ....................  ....................
29476 MT520398.1 |Severe acute respiratory syndrome coro    ....................  ....................  .....................  ....................  ....................  ....................
29477 MT520399.1 |Severe acute respiratory syndrome coro    ....................  ....................  .....................  ....................  ....................  ....................
29478 MT520400.1 |Severe acute respiratory syndrome coro    ....................  ....................  .....................  ....................  ....................  ....................
29479 MT520401.1 |Severe acute respiratory syndrome coro    ....................  ....................  .....................  ....................  ....................  ....................
29480 MT520402.1 |Severe acute respiratory syndrome coro    ....................  ....................  .....................  ....................  ....................  ....................
29481 MT520403.1 |Severe acute respiratory syndrome coro    ....................  ....................  .....................  ....................  ....................  ....................
```

29482 MT520404.1 |Severe acute respiratory syndrome coro .................... .................... .................... .................... .................... ....................
29483 MT520405.1 |Severe acute respiratory syndrome coro .................... .................... .................... .................... .................... ....................
29484 MT520406.1 |Severe acute respiratory syndrome coro .................... .................... .................... .................... .................... ....................
29485 MT520407.1 |Severe acute respiratory syndrome coro .................... .................... .................... .................... .................... ....................
29486 MT520408.1 |Severe acute respiratory syndrome coro .................... .................... .................... .................... .................... ....................
29487 MT520409.1 |Severe acute respiratory syndrome coro .................... .................... .................... .................... .................... ....................
29488 MT520410.1 |Severe acute respiratory syndrome coro .................... .................... .................... .................... .................... ....................
29489 MT520411.1 |Severe acute respiratory syndrome coro .................... .................... .................... .................... .................... ....................
29490 MT520412.1 |Severe acute respiratory syndrome coro .................... .................... .................... .................... .................... ....................
29491 MT520413.1 |Severe acute respiratory syndrome coro .................... .................... .................... .................... .................... ....................
29492 MT520414.1 |Severe acute respiratory syndrome coro .................... .................... .................... .................... .................... ....................
29493 MT520415.1 |Severe acute respiratory syndrome coro .................... .................... .................... .................... .................... ....................
29494 MT520416.1 |Severe acute respiratory syndrome coro .................... .................... .................... .................... .................... ....................
29495 MT520417.1 |Severe acute respiratory syndrome coro .................... .................... .................... .................... .................... ....................
29496 MT520418.1 |Severe acute respiratory syndrome coro .................... .................... .................... .................... .................... ....................
29497 MT520419.1 |Severe acute respiratory syndrome coro .................... .................... .................... .................... .................... ....................
29498 MT520420.1 |Severe acute respiratory syndrome coro .................... .................... .................... .................... .................... ....................
29499 MT520421.1 |Severe acute respiratory syndrome coro .................... .................... .................... .................... .................... ....................
29500 MT520422.1 |Severe acute respiratory syndrome coro .................... .................... .................... .................... .................... ....................
29502 MT520424.1 |Severe acute respiratory syndrome coro .................... .................... .................... .................... .................... ....................
29503 MT520426.1 |Severe acute respiratory syndrome coro .................... .................... .................... .................... .................... ....................
29504 MT520427.1 |Severe acute respiratory syndrome coro .................... .................... .................... .................... .................... ....................
29505 MT520428.1 |Severe acute respiratory syndrome coro .................... .................... .................... .................... .................... ....................
29506 MT520429.1 |Severe acute respiratory syndrome coro .................... .................... .................... .................... .................... ....................
29507 MT520431.1 |Severe acute respiratory syndrome coro .................... .................... .................... .................... .................... ....................
29508 MT520432.1 |Severe acute respiratory syndrome coro .................... .................... .................... .................... .................... ....................
29509 MT520433.1 |Severe acute respiratory syndrome coro .................... .................... .................... .................... .................... ....................
29510 MT520434.1 |Severe acute respiratory syndrome coro .................... .................... .................... .................... .................... ....................
29511 MT520435.1 |Severe acute respiratory syndrome coro .................... .................... .................... .................... .................... ....................
29512 MT520436.1 |Severe acute respiratory syndrome coro .................... .................... .................... .................... .................... ....................
29513 MT520437.1 |Severe acute respiratory syndrome coro .................... .................... .................... .................... .................... ....................
29514 MT520439.1 |Severe acute respiratory syndrome coro .................... .................... .................... .................... .................... ....................
29515 MT520440.1 |Severe acute respiratory syndrome coro .................... .................... .................... .................... .................... ....................
29516 MT520441.1 |Severe acute respiratory syndrome coro .................... .................... .................... .................... .................... ....................
29517 MT520443.1 |Severe acute respiratory syndrome coro .................... .................... .................... .................... .................... ....................
29518 MT520444.1 |Severe acute respiratory syndrome coro .................... .................... .................... .................... .................... ....................
29519 MT520445.1 |Severe acute respiratory syndrome coro .................... .................... .................... .................... .................... ....................
29520 MT520446.1 |Severe acute respiratory syndrome coro .................... .................... .................... .................... .................... ....................
29521 MT520447.1 |Severe acute respiratory syndrome coro .................... .................... .................... .................... .................... ....................
29522 MT520448.1 |Severe acute respiratory syndrome coro .................... .................... .................... .................... .................... ....................
29523 MT520451.1 |Severe acute respiratory syndrome coro .................... .................... .................... .................... .................... ....................
29524 MT520452.1 |Severe acute respiratory syndrome coro .................... .................... .................... .................... .................... ....................
29525 MT520453.1 |Severe acute respiratory syndrome coro .................... .................... .................... .................... .................... ....................
29526 MT520454.1 |Severe acute respiratory syndrome coro .................... .................... .................... .................... .................... ....................
29527 MT520455.1 |Severe acute respiratory syndrome coro .................... .................... .................... .................... .................... ....................
29528 MT520457.1 |Severe acute respiratory syndrome coro .................... .................... .................... .................... .................... ....................
29529 MT520458.1 |Severe acute respiratory syndrome coro .................... .................... .................... .................... .................... ....................
29530 MT520459.1 |Severe acute respiratory syndrome coro .................... .................... .................... .................... .................... ....................
29531 MT520460.1 |Severe acute respiratory syndrome coro .................... .................... .................... .................... .................... ....................
29532 MT520461.1 |Severe acute respiratory syndrome coro .................... .................... .................... .................... .................... ....................
29533 MT520463.1 |Severe acute respiratory syndrome coro .................... .................... .................... .................... .................... ....................
29534 MT520464.1 |Severe acute respiratory syndrome coro .................... .................... .................... .................... .................... ....................
29535 MT520465.1 |Severe acute respiratory syndrome coro .................... .................... .................... .................... .................... ....................
29536 MT520466.1 |Severe acute respiratory syndrome coro .................... .................... .................... .................... .................... ....................
29537 MT520467.1 |Severe acute respiratory syndrome coro .................... .................... .................... .................... .................... ....................
29538 MT520469.1 |Severe acute respiratory syndrome coro .................... .................... .................... .................... .................... ....................
29539 MT520471.1 |Severe acute respiratory syndrome coro .................... .................... .................... .................... .................... ....................
29540 MT520472.1 |Severe acute respiratory syndrome coro .................... .................... .................... .................... .................... ....................
29541 MT520473.1 |Severe acute respiratory syndrome coro .................... .................... .................... .................... .................... ....................
29542 MT520474.1 |Severe acute respiratory syndrome coro .................... .................... .................... .................... .................... ....................
29543 MT520475.1 |Severe acute respiratory syndrome coro .................... .................... .................... .................... .................... ....................
29544 MT520477.1 |Severe acute respiratory syndrome coro .................... .................... .................... .................... .................... ....................
29545 MT520478.1 |Severe acute respiratory syndrome coro .................... .................... .................... .................... .................... ....................
29546 MT520479.1 |Severe acute respiratory syndrome coro .................... .................... .................... .................... .................... ....................
29547 MT520481.1 |Severe acute respiratory syndrome coro .................... .................... .................... .................... .................... ....................
29548 MT520482.1 |Severe acute respiratory syndrome coro .................... .................... .................... .................... .................... ....................
29549 MT520483.1 |Severe acute respiratory syndrome coro .................... .................... .................... .................... .................... ....................
29550 MT520484.1 |Severe acute respiratory syndrome coro .................... .................... .................... .................... .................... ....................
29551 MT520485.1 |Severe acute respiratory syndrome coro .................... .................... .................... .................... .................... ....................
29552 MT520486.1 |Severe acute respiratory syndrome coro .................... .................... .................... .................... .................... ....................
29553 MT520487.1 |Severe acute respiratory syndrome coro .................... .................... .................... .................... .................... ....................
29554 MT520488.1 |Severe acute respiratory syndrome coro .................... .................... .................... .................... .................... ....................
29555 MT520489.1 |Severe acute respiratory syndrome coro .................... .................... .................... .................... .................... ....................
29556 MT520490.1 |Severe acute respiratory syndrome coro .................... .................... .................... .................... .................... ....................
29557 MT520492.1 |Severe acute respiratory syndrome coro .................... .................... .................... .................... .................... ....................
29558 MT520493.1 |Severe acute respiratory syndrome coro .................... .................... .................... .................... .................... ....................
29559 MT520494.1 |Severe acute respiratory syndrome coro .................... .................... .................... .................... .................... ....................
29560 MT520495.1 |Severe acute respiratory syndrome coro .................... .................... .................... .................... .................... ....................

| | | | | | | | | |
|---|---|---|---|---|---|---|---|---|
| 29561 | MT520496.1 | \|Severe acute respiratory syndrome coro | ···················· | ···················· | ···················· | ···················· | ···················· | ···················· |
| 29562 | MT520497.1 | \|Severe acute respiratory syndrome coro | ···················· | ···················· | ···················· | ···················· | ···················· | ···················· |
| 29563 | MT520498.1 | \|Severe acute respiratory syndrome coro | ···················· | ···················· | ···················· | ···················· | ···················· | ···················· |
| 29564 | MT520500.1 | \|Severe acute respiratory syndrome coro | ···················· | ···················· | ···················· | ···················· | ···················· | ···················· |
| 29565 | MT520501.1 | \|Severe acute respiratory syndrome coro | ···················· | ···················· | ···················· | ···················· | ···················· | ···················· |
| 29566 | MT520502.1 | \|Severe acute respiratory syndrome coro | ···················· | ···················· | ···················· | ···················· | ···················· | ···················· |
| 29567 | MT520503.1 | \|Severe acute respiratory syndrome coro | ···················· | ···················· | ···················· | ···················· | ···················· | ···················· |
| 29568 | MT520504.1 | \|Severe acute respiratory syndrome coro | ···················· | ···················· | ···················· | ···················· | ···················· | ···················· |
| 29569 | MT520505.1 | \|Severe acute respiratory syndrome coro | ···················· | ···················· | ···················· | ···················· | ···················· | ···················· |
| 29570 | MT520506.1 | \|Severe acute respiratory syndrome coro | ···················· | ···················· | ···················· | ···················· | ···················· | ···················· |
| 29571 | MT520507.1 | \|Severe acute respiratory syndrome coro | ···················· | ···················· | ···················· | ···················· | ···················· | ···················· |
| 29572 | MT520508.1 | \|Severe acute respiratory syndrome coro | ···················· | ···················· | ···················· | ···················· | ···················· | ···················· |
| 29573 | MT520509.1 | \|Severe acute respiratory syndrome coro | ···················· | ···················· | ···················· | ···················· | ···················· | ···················· |
| 29574 | MT520510.1 | \|Severe acute respiratory syndrome coro | ···················· | ···················· | ···················· | ···················· | ···················· | ···················· |
| 29575 | MT520511.1 | \|Severe acute respiratory syndrome coro | ···················· | ···················· | ···················· | ···················· | ···················· | ···················· |
| 29576 | MT520512.1 | \|Severe acute respiratory syndrome coro | ···················· | ···················· | ···················· | ···················· | ···················· | ···················· |
| 29577 | MT520515.1 | \|Severe acute respiratory syndrome coro | ···················· | ···················· | ···················· | ···················· | ···················· | ···················· |
| 29578 | MT520516.1 | \|Severe acute respiratory syndrome coro | ···················· | ···················· | ···················· | ···················· | ···················· | ···················· |
| 29579 | MT520517.1 | \|Severe acute respiratory syndrome coro | ···················· | ···················· | ···················· | ···················· | ···················· | ···················· |
| 29580 | MT520518.1 | \|Severe acute respiratory syndrome coro | ···················· | ···················· | ···················· | ···················· | ···················· | ···················· |
| 29581 | MT520519.1 | \|Severe acute respiratory syndrome coro | ···················· | ···················· | ···················· | ···················· | ···················· | ···················· |
| 29582 | MT520520.1 | \|Severe acute respiratory syndrome coro | ···················· | ···················· | ···················· | ···················· | ···················· | ···················· |
| 29583 | MT520521.1 | \|Severe acute respiratory syndrome coro | ···················· | ···················· | ···················· | ···················· | ···················· | ···················· |
| 29584 | MT520522.1 | \|Severe acute respiratory syndrome coro | ···················· | ···················· | ···················· | ···················· | ···················· | ···················· |
| 29585 | MT520523.1 | \|Severe acute respiratory syndrome coro | ···················· | ···················· | ···················· | ···················· | ···················· | ···················· |
| 29586 | MT520525.1 | \|Severe acute respiratory syndrome coro | ···················· | ···················· | ···················· | ···················· | ···················· | ···················· |
| 29587 | MT520526.1 | \|Severe acute respiratory syndrome coro | ···················· | ···················· | ···················· | ···················· | ···················· | ···················· |
| 29588 | MT520528.1 | \|Severe acute respiratory syndrome coro | ···················· | ···················· | ···················· | ···················· | ···················· | ···················· |
| 29589 | MT520529.1 | \|Severe acute respiratory syndrome coro | ···················· | ···················· | ···················· | ···················· | ···················· | ···················· |
| 29590 | MT520530.1 | \|Severe acute respiratory syndrome coro | ···················· | ···················· | ···················· | ···················· | ···················· | ···················· |
| 29591 | MT520532.1 | \|Severe acute respiratory syndrome coro | ···················· | ···················· | ···················· | ···················· | ···················· | ···················· |
| 29592 | MT520533.1 | \|Severe acute respiratory syndrome coro | ···················· | ···················· | ···················· | ···················· | ···················· | ···················· |
| 29593 | MT520534.1 | \|Severe acute respiratory syndrome coro | ···················· | ···················· | ···················· | ···················· | ···················· | ···················· |
| 29594 | MT520535.1 | \|Severe acute respiratory syndrome coro | ···················· | ···················· | ···················· | ···················· | ···················· | ···················· |
| 29595 | MT520536.1 | \|Severe acute respiratory syndrome coro | ···················· | ···················· | ···················· | ···················· | ···················· | ···················· |
| 29596 | MT520537.1 | \|Severe acute respiratory syndrome coro | ···················· | ···················· | ···················· | ···················· | ···················· | ···················· |
| 29597 | MT520538.1 | \|Severe acute respiratory syndrome coro | ···················· | ···················· | ···················· | ···················· | ···················· | ···················· |
| 29598 | MT520539.1 | \|Severe acute respiratory syndrome coro | ···················· | ···················· | ···················· | ···················· | ···················· | ···················· |
| 29599 | MT520540.1 | \|Severe acute respiratory syndrome coro | ···················· | ···A················ | ···················· | ···················· | ···················· | ···················· |
| 29600 | MT520541.1 | \|Severe acute respiratory syndrome coro | ···················· | ···················· | ···················· | ···················· | ···················· | ···················· |
| 29601 | MT520542.1 | \|Severe acute respiratory syndrome coro | ···················· | ···················· | ···················· | ···················· | ···················· | ···················· |
| 29602 | MT520543.1 | \|Severe acute respiratory syndrome coro | ···················· | ···················· | ···················· | ···················· | ···················· | ···················· |
| 29603 | MT520544.1 | \|Severe acute respiratory syndrome coro | ···················· | ···················· | ···················· | ···················· | ···················· | ···················· |
| 29604 | MT520546.1 | \|Severe acute respiratory syndrome coro | ···················· | ···················· | ···················· | ···················· | ···················· | ···················· |
| 29605 | MT520547.1 | \|Severe acute respiratory syndrome coro | ···················· | ···················· | ···················· | ···················· | ···················· | ···················· |
| 29606 | MT560525.1 | \|Severe acute respiratory syndrome coro | ···················· | ···················· | ···················· | ···················· | ···················· | ···················· |
| 29607 | MT560530.1 | \|Severe acute respiratory syndrome coro | ···················· | ···················· | ···················· | ···················· | ···················· | ···················· |
| 29608 | MT560531.1 | \|Severe acute respiratory syndrome coro | ···················· | ···················· | ···················· | ···················· | ···················· | ···················· |
| 29609 | MT560656.1 | \|Severe acute respiratory syndrome coro | ···················· | ·A·················· | ···················· | ···················· | ···················· | ··A················· |
| 29610 | MT560657.1 | \|Severe acute respiratory syndrome coro | ···················· | ···················· | ···················· | ···················· | ···················· | ···················· |
| 29611 | MT560667.1 | \|Severe acute respiratory syndrome coro | ···················· | ·A·················· | ···················· | ···················· | ···················· | ··A················· |
| 29612 | MT560668.1 | \|Severe acute respiratory syndrome coro | ···················· | ·A·················· | ···················· | ···················· | ···················· | ··A················· |
| 29613 | MT560669.1 | \|Severe acute respiratory syndrome coro | ···················· | ···················· | ···················· | ···················· | ···················· | ···················· |
| 29614 | MT560670.1 | \|Severe acute respiratory syndrome coro | ···················· | ·A·················· | ···················· | ···················· | ···················· | ··A················· |
| 29615 | MT560671.1 | \|Severe acute respiratory syndrome coro | ···················· | ·A·················· | ···················· | ···················· | ···················· | ··A················· |
| 29616 | MT560672.1 | \|Severe acute respiratory syndrome coro | ···················· | ···················· | ···················· | ···················· | ···················· | ···················· |
| 29617 | MT560673.1 | \|Severe acute respiratory syndrome coro | ···················· | ···················· | ···················· | ···················· | ···················· | ···················· |
| 29618 | MT560674.1 | \|Severe acute respiratory syndrome coro | ···················· | ·A·················· | ···················· | ···················· | ···················· | ··A················· |
| 29619 | MT560675.1 | \|Severe acute respiratory syndrome coro | ···················· | ···················· | ···················· | ···················· | ···················· | ···················· |
| 29620 | MT560677.1 | \|Severe acute respiratory syndrome coro | ···················· | ···················· | ···················· | ···················· | ···················· | ···················· |
| 29621 | MT560679.1 | \|Severe acute respiratory syndrome coro | ···················· | ·A·················· | ···················· | ···················· | ···················· | ··A················· |
| 29622 | MT560680.1 | \|Severe acute respiratory syndrome coro | ···················· | ·A·················· | ···················· | ···················· | ···················· | ··A················· |
| 29623 | MT560682.1 | \|Severe acute respiratory syndrome coro | ···················· | ·A·················· | ···················· | ···················· | ···················· | ···················· |
| 29624 | MT560689.1 | \|Severe acute respiratory syndrome coro | ···················· | ·A·················· | ···················· | ···················· | ···················· | ··A················· |
| 29625 | MT560690.1 | \|Severe acute respiratory syndrome coro | ···················· | ···················· | ···················· | ···················· | ···················· | ···················· |
| 29626 | MT560692.1 | \|Severe acute respiratory syndrome coro | ···················· | ·A·················· | ···················· | ···················· | ···················· | ··A················· |
| 29627 | MT560693.1 | \|Severe acute respiratory syndrome coro | ···················· | ·A·················· | ···················· | ···················· | ···················· | ··A················· |
| 29628 | MT560694.1 | \|Severe acute respiratory syndrome coro | ···················· | ···················· | ···················· | ···················· | ···················· | ···················· |
| 29629 | MT560704.1 | \|Severe acute respiratory syndrome coro | ···················· | ·A·················· | ···················· | ···················· | ···················· | ··A················· |
| 29630 | MT560705.1 | \|Severe acute respiratory syndrome coro | ···················· | ·A·················· | ···················· | ···················· | ···················· | ···················· |
| 29631 | MT560706.1 | \|Severe acute respiratory syndrome coro | ···················· | ···················· | ···················· | ···················· | ···················· | ···················· |
| 29632 | MT560827.1 | \|Severe acute respiratory syndrome coro | ···················· | ···················· | ···················· | ···················· | ···················· | ···················· |
| 29633 | MT565495.1 | \|Severe acute respiratory syndrome coro | ···················· | ···················· | ···················· | ···················· | ···················· | ···················· |
| 29634 | MT565496.1 | \|Severe acute respiratory syndrome coro | ···················· | ···················· | ···················· | ···················· | ···················· | ···················· |
| 29635 | MT565497.1 | \|Severe acute respiratory syndrome coro | ···················· | ···················· | ···················· | ···················· | ···················· | ···················· |
| 29636 | MT565498.1 | \|Severe acute respiratory syndrome coro | ···················· | ···················· | ···················· | ···················· | ···················· | ···················· |
| 29637 | MT565499.1 | \|Severe acute respiratory syndrome coro | ···················· | ···················· | ···················· | ···················· | ···················· | ···················· |
| 29638 | MT566435.1 | \|Severe acute respiratory syndrome coro | ···················· | ···················· | ···················· | ···················· | ···················· | ···················· |

29639 MT566436.1 |Severe acute respiratory syndrome coro ···················· ···················· ···················· ···················· ···················· ····················
29640 MT566437.1 |Severe acute respiratory syndrome coro ···················· ···················· ···················· ···················· ···················· ····················
29641 MT513758.1 |Severe acute respiratory syndrome coro ··A················· ···················· ···················· ···················· ···················· ····················
29642 MT557568.1 |Severe acute respiratory syndrome coro ···················· ···················· ···················· ···················· ···················· ····················
29643 MT557569.1 |Severe acute respiratory syndrome coro ···················· ···················· ···················· ···················· ···················· ····················
29644 MT557570.1 |Severe acute respiratory syndrome coro ···················· ···················· ···················· ···················· ···················· ····················
29645 MT558647.1 |Severe acute respiratory syndrome coro ···················· ···················· ···················· ···················· ···················· ····················
29646 MT558648.1 |Severe acute respiratory syndrome coro ···················· ···················· ···················· ···················· ···················· ····················
29647 MT558649.1 |Severe acute respiratory syndrome coro ···················· ···················· ···················· ···················· ···················· ····················
29648 MT558650.1 |Severe acute respiratory syndrome coro ···················· ···················· ···················· ···················· ···················· ····················
29649 MT558651.1 |Severe acute respiratory syndrome coro ···················· ···················· ···················· ···················· ···················· ····················
29650 MT558652.1 |Severe acute respiratory syndrome coro ···················· ···················· ···················· ···················· ···················· ····················
29651 MT558653.1 |Severe acute respiratory syndrome coro ···················· ···················· ···················· ···················· ···················· ····················
29652 MT558654.1 |Severe acute respiratory syndrome coro ···················· ···················· ···················· ···················· ···················· ····················
29653 MT558655.1 |Severe acute respiratory syndrome coro ···················· ···················· ···················· ···················· ···················· ····················
29654 MT558656.1 |Severe acute respiratory syndrome coro ···················· ···················· ···················· ···················· ···················· ····················
29655 MT558657.1 |Severe acute respiratory syndrome coro ···················· ···················· ···················· ···················· ···················· ····················
29656 MT558658.1 |Severe acute respiratory syndrome coro ···················· ···················· ···················· ···················· ···················· ····················
29657 MT558659.1 |Severe acute respiratory syndrome coro ···················· ···················· ···················· ···················· ···················· ····················
29658 MT558660.1 |Severe acute respiratory syndrome coro ···················· ···················· ···················· ···················· ···················· ····················
29659 MT558661.1 |Severe acute respiratory syndrome coro ···················· ···················· ···················· ···················· ···················· ····················
29660 MT558662.1 |Severe acute respiratory syndrome coro ···················· ···················· ···················· ···················· ···················· ····················
29661 MT558663.1 |Severe acute respiratory syndrome coro ···················· ···················· ···················· ···················· ···················· ····················
29662 MT558664.1 |Severe acute respiratory syndrome coro ···················· ···················· ···················· ···················· ···················· ····················
29663 MT558665.1 |Severe acute respiratory syndrome coro ···················· ···················· ···················· ···················· ···················· ····················
29664 MT558666.1 |Severe acute respiratory syndrome coro ···················· ···················· ···················· ···················· ···················· ····················
29665 MT558667.1 |Severe acute respiratory syndrome coro ···················· ···················· ···················· ···················· ···················· ····················
29666 MT558668.1 |Severe acute respiratory syndrome coro ···················· ···················· ···················· ···················· ···················· ····················
29667 MT558669.1 |Severe acute respiratory syndrome coro ···················· ···················· ···················· ···················· ···················· ····················
29668 MT558670.1 |Severe acute respiratory syndrome coro ···················· ···················· ···················· ···················· ···················· ····················
29669 MT558671.1 |Severe acute respiratory syndrome coro ···················· ···················· ···················· ···················· ···················· ····················
29670 MT558672.1 |Severe acute respiratory syndrome coro ···················· ···················· ···················· ···················· ···················· ····················
29671 MT558673.1 |Severe acute respiratory syndrome coro ···················· ···················· ···················· ···················· ···················· ····················
29672 MT558674.1 |Severe acute respiratory syndrome coro ···················· ···················· ···················· ···················· ···················· ····················
29673 MT558675.1 |Severe acute respiratory syndrome coro ···················· ···················· ···················· ···················· ···················· ····················
29674 MT558676.1 |Severe acute respiratory syndrome coro ···················· ···················· ···················· ···················· ···················· ····················
29675 MT558677.1 |Severe acute respiratory syndrome coro ···················· ···················· ···················· ···················· ···················· ····················
29676 MT558678.1 |Severe acute respiratory syndrome coro ···················· ···················· ···················· ···················· ···················· ····················
29677 MT558679.1 |Severe acute respiratory syndrome coro ···················· ···················· ···················· ···················· ···················· ····················
29678 MT558680.1 |Severe acute respiratory syndrome coro ···················· ···················· ···················· ···················· ···················· ····················
29679 MT558681.1 |Severe acute respiratory syndrome coro ···················· ···················· ···················· ···················· ···················· ····················
29680 MT558682.1 |Severe acute respiratory syndrome coro ···················· ···················· ···················· ···················· ···················· ····················
29681 MT558683.1 |Severe acute respiratory syndrome coro ···················· ···················· ···················· ···················· ···················· ····················
29682 MT558684.1 |Severe acute respiratory syndrome coro ···················· ···················· ···················· ···················· ···················· ····················
29683 MT558685.1 |Severe acute respiratory syndrome coro ···················· ···················· ···················· ···················· ···················· ····················
29684 MT558686.1 |Severe acute respiratory syndrome coro ···················· ···················· ···················· ···················· ···················· ····················
29685 MT558687.1 |Severe acute respiratory syndrome coro ···················· ···················· ···················· ···················· ···················· ····················
29686 MT558688.1 |Severe acute respiratory syndrome coro ···················· ···················· ···················· ···················· ···················· ····················
29687 MT558689.1 |Severe acute respiratory syndrome coro ···················· ···················· ···················· ···················· ···················· ····················
29688 MT558690.1 |Severe acute respiratory syndrome coro ···················· ···················· ···················· ···················· ···················· ····················
29689 MT558691.1 |Severe acute respiratory syndrome coro ···················· ···················· ···················· ···················· ···················· ····················
29690 MT558692.1 |Severe acute respiratory syndrome coro ···················· ···················· ···················· ···················· ···················· ····················
29691 MT558693.1 |Severe acute respiratory syndrome coro ···················· ···················· ···················· ···················· ···················· ····················
29692 MT558694.1 |Severe acute respiratory syndrome coro ···················· ···················· ···················· ···················· ···················· ····················
29693 MT558695.1 |Severe acute respiratory syndrome coro ···················· ···················· ···················· ···················· ···················· ····················
29694 MT558696.1 |Severe acute respiratory syndrome coro ···················· ···················· ···················· ···················· ···················· ····················
29695 MT558697.1 |Severe acute respiratory syndrome coro ···················· ···················· ···················· ···················· ···················· ····················
29696 MT558698.1 |Severe acute respiratory syndrome coro ···················· ···················· ···················· ···················· ···················· ····················
29697 MT558699.1 |Severe acute respiratory syndrome coro ···················· ···················· ···················· ···················· ···················· ····················
29698 MT558700.1 |Severe acute respiratory syndrome coro ···················· ···················· ···················· ···················· ···················· ····················
29699 MT558701.1 |Severe acute respiratory syndrome coro ···················· ···················· ···················· ···················· ···················· ····················
29700 MT558702.1 |Severe acute respiratory syndrome coro ···················· ···················· ···················· ···················· ···················· ····················
29701 MT558703.1 |Severe acute respiratory syndrome coro ···················· ···················· ···················· ···················· ···················· ····················
29702 MT558704.1 |Severe acute respiratory syndrome coro ···················· ···················· ···················· ···················· ···················· ····················
29703 MT558705.1 |Severe acute respiratory syndrome coro ···················· ···················· ···················· ···················· ···················· ····················
29704 MT558706.1 |Severe acute respiratory syndrome coro ···················· ···················· ···················· ···················· ···················· ····················
29705 MT558707.1 |Severe acute respiratory syndrome coro ···················· ···················· ···················· ···················· ···················· ····················
29706 MT559037.1 |Severe acute respiratory syndrome coro ···················· ···················· ···················· ···················· ···················· ····················
29707 MT559038.1 |Severe acute respiratory syndrome coro ···················· ···················· ···················· ···················· ···················· ····················
29708 MT559800.1 |Severe acute respiratory syndrome coro ···················· ···················· ···················· ···················· ···················· ····················
29709 MT559801.1 |Severe acute respiratory syndrome coro ···················· ···················· ···················· ···················· ···················· ····················
29710 MT559802.1 |Severe acute respiratory syndrome coro ···················· ···················· ···················· ···················· ···················· ····················
29711 MT559803.1 |Severe acute respiratory syndrome coro ···················· ···················· ···················· ···················· ···················· ····················
29712 MT551604.1 |Severe acute respiratory syndrome coro ···················· ···················· ···················· ···················· ···················· ····················
29713 MT551605.1 |Severe acute respiratory syndrome coro ···················· ···················· ···················· ···················· ···················· ····················
29714 MT551606.1 |Severe acute respiratory syndrome coro ···················· ···················· ···················· ···················· ···················· ····················
29715 MT551607.1 |Severe acute respiratory syndrome coro ···················· ···················· ···················· ···················· ···················· ····················
29716 MT551608.1 |Severe acute respiratory syndrome coro ···················· ···················· ···················· ···················· ···················· ····················

| | | | | | | | | |
|---|---|---|---|---|---|---|---|---|
| 29717 | MT551609.1 | \|Severe acute respiratory syndrome coro | .................... | .................... | .................... | .................... | .................... | .................... |
| 29718 | MT554052.1 | \|Severe acute respiratory syndrome coro | .................... | .................... | .................... | .................... | .................... | .................... |
| 29719 | MT547814.1 | \|Severe acute respiratory syndrome coro | .................... | .................... | .................... | .................... | .................... | .................... |
| 29720 | MT539726.1 | \|Severe acute respiratory syndrome coro | .................... | .................... | .................... | .................... | .................... | .................... |
| 29721 | MT539729.1 | \|Severe acute respiratory syndrome coro | .................... | .................... | .................... | .................... | .................... | .................... |
| 29722 | MT539158.1 | \|Severe acute respiratory syndrome coro | .................... | .................... | .................... | .................... | .................... | .................... |
| 29723 | MT539159.1 | \|Severe acute respiratory syndrome coro | .................... | .................... | .................... | .................... | .................... | .................... |
| 29724 | MT539160.1 | \|Severe acute respiratory syndrome coro | .................... | .................... | .................... | .................... | .................... | .................... |
| 29725 | MT539162.1 | \|Severe acute respiratory syndrome coro | .................... | .................... | .................... | .................... | .................... | .................... |
| 29726 | MT539163.1 | \|Severe acute respiratory syndrome coro | .................... | .................... | .................... | .................... | .................... | .................... |
| 29727 | MT539164.1 | \|Severe acute respiratory syndrome coro | .................... | .................... | .................... | .................... | .................... | .................... |
| 29728 | MT539165.1 | \|Severe acute respiratory syndrome coro | .................... | .................... | .................... | .................... | .................... | .................... |
| 29729 | MT539166.1 | \|Severe acute respiratory syndrome coro | .................... | .................... | .................... | .................... | .................... | .................... |
| 29730 | MT539167.1 | \|Severe acute respiratory syndrome coro | .................... | .................... | .................... | .................... | .................... | .................... |
| 29731 | MT539168.1 | \|Severe acute respiratory syndrome coro | .................... | .................... | .................... | .................... | .................... | .................... |
| 29732 | MT539169.1 | \|Severe acute respiratory syndrome coro | .................... | .................... | .................... | .................... | .................... | .................... |
| 29733 | MT539170.1 | \|Severe acute respiratory syndrome coro | .................... | .................... | .................... | .................... | .................... | .................... |
| 29734 | MT539171.1 | \|Severe acute respiratory syndrome coro | .................... | .................... | .................... | .................... | .................... | .................... |
| 29735 | MT539172.1 | \|Severe acute respiratory syndrome coro | .................... | .................... | .................... | .................... | .................... | .................... |
| 29736 | MT539173.1 | \|Severe acute respiratory syndrome coro | .................... | .................... | .................... | .................... | .................... | .................... |
| 29737 | MT539174.1 | \|Severe acute respiratory syndrome coro | .................... | .................... | .................... | .................... | .................... | .................... |
| 29738 | MT539175.1 | \|Severe acute respiratory syndrome coro | .................... | .................... | .................... | .................... | .................... | .................... |
| 29739 | MT539176.1 | \|Severe acute respiratory syndrome coro | .................... | .................... | .................... | .................... | .................... | .................... |
| 29740 | MT407649.1 | \|Severe acute respiratory syndrome coro | .................... | .................... | .................... | .................... | .................... | .................... |
| 29741 | MT407650.1 | \|Severe acute respiratory syndrome coro | .................... | .................... | .................... | .................... | .................... | .................... |
| 29742 | MT407651.1 | \|Severe acute respiratory syndrome coro | .................... | .................... | .................... | .................... | .................... | .................... |
| 29743 | MT407652.1 | \|Severe acute respiratory syndrome coro | .................... | .................... | .................... | .................... | .................... | .................... |
| 29744 | MT407653.1 | \|Severe acute respiratory syndrome coro | .................... | .................... | .................... | .................... | .................... | .................... |
| 29745 | MT407654.1 | \|Severe acute respiratory syndrome coro | .................... | .................... | .................... | .................... | .................... | .................... |
| 29746 | MT407655.1 | \|Severe acute respiratory syndrome coro | .................... | .................... | .................... | .................... | .................... | .................... |
| 29747 | MT407656.1 | \|Severe acute respiratory syndrome coro | .................... | .................... | .................... | .................... | .................... | .................... |
| 29748 | MT407657.1 | \|Severe acute respiratory syndrome coro | .................... | .................... | .................... | .................... | .................... | .................... |
| 29749 | MT407658.1 | \|Severe acute respiratory syndrome coro | .................... | .................... | .................... | .................... | .................... | .................... |
| 29750 | MT407659.1 | \|Severe acute respiratory syndrome coro | .................... | .................... | .................... | .................... | .................... | .................... |
| 29751 | MT534285.1 | \|Severe acute respiratory syndrome coro | .................... | .................... | .................... | .................... | .................... | .................... |
| 29752 | MT534304.1 | \|Severe acute respiratory syndrome coro | .................... | .................... | .................... | .................... | .................... | .................... |
| 29753 | MT534305.1 | \|Severe acute respiratory syndrome coro | .................... | .................... | .................... | .................... | .................... | .................... |
| 29754 | MT534306.1 | \|Severe acute respiratory syndrome coro | .................... | .................... | .................... | .................... | .................... | .................... |
| 29755 | MT534307.1 | \|Severe acute respiratory syndrome coro | .................... | .................... | .................... | .................... | .................... | .................... |
| 29756 | MT534308.1 | \|Severe acute respiratory syndrome coro | .................... | .................... | .................... | .................... | .................... | .................... |
| 29757 | MT534309.1 | \|Severe acute respiratory syndrome coro | .................... | .................... | .................... | .................... | .................... | .................... |
| 29758 | MT534311.1 | \|Severe acute respiratory syndrome coro | .................... | .................... | .................... | .................... | .................... | .................... |
| 29759 | MT534312.1 | \|Severe acute respiratory syndrome coro | .................... | .................... | .................... | .................... | .................... | .................... |
| 29760 | MT534313.1 | \|Severe acute respiratory syndrome coro | .................... | .................... | .................... | .................... | .................... | .................... |
| 29761 | MT534314.1 | \|Severe acute respiratory syndrome coro | .................... | .................... | .................... | .................... | .................... | .................... |
| 29762 | MT534315.1 | \|Severe acute respiratory syndrome coro | .................... | .................... | .................... | .................... | .................... | .................... |
| 29763 | MT534316.1 | \|Severe acute respiratory syndrome coro | .................... | .................... | .................... | .................... | .................... | .................... |
| 29764 | MT534317.1 | \|Severe acute respiratory syndrome coro | .................... | .................... | .................... | .................... | .................... | .................... |
| 29765 | MT534318.1 | \|Severe acute respiratory syndrome coro | .................... | .................... | .................... | .................... | .................... | .................... |
| 29766 | MT534320.1 | \|Severe acute respiratory syndrome coro | .................... | .................... | .................... | .................... | .................... | .................... |
| 29767 | MT534322.1 | \|Severe acute respiratory syndrome coro | .................... | .................... | .................... | .................... | .................... | .................... |
| 29768 | MT534323.1 | \|Severe acute respiratory syndrome coro | .................... | .................... | .................... | .................... | .................... | .................... |
| 29769 | MT534324.1 | \|Severe acute respiratory syndrome coro | .................... | .................... | .................... | .................... | .................... | .................... |
| 29770 | MT534325.1 | \|Severe acute respiratory syndrome coro | .................... | .................... | .................... | .................... | .................... | .................... |
| 29771 | MT534326.1 | \|Severe acute respiratory syndrome coro | .................... | .................... | .................... | .................... | .................... | .................... |
| 29772 | MT534327.1 | \|Severe acute respiratory syndrome coro | .................... | .................... | .................... | .................... | .................... | .................... |
| 29773 | MT534328.1 | \|Severe acute respiratory syndrome coro | .................... | .................... | .................... | .................... | .................... | .................... |
| 29774 | MT534329.1 | \|Severe acute respiratory syndrome coro | .................... | .................... | .................... | .................... | .................... | .................... |
| 29775 | MT534330.1 | \|Severe acute respiratory syndrome coro | .................... | .................... | .................... | .................... | .................... | .................... |
| 29776 | MT534331.1 | \|Severe acute respiratory syndrome coro | .................... | .................... | .................... | .................... | .................... | .................... |
| 29777 | MT534332.1 | \|Severe acute respiratory syndrome coro | .................... | .................... | .................... | .................... | .................... | .................... |
| 29778 | MT534333.1 | \|Severe acute respiratory syndrome coro | .................... | .................... | .................... | .................... | .................... | .................... |
| 29779 | MT534334.1 | \|Severe acute respiratory syndrome coro | .................... | .................... | .................... | .................... | .................... | .................... |
| 29780 | MT534335.1 | \|Severe acute respiratory syndrome coro | .................... | .................... | .................... | .................... | .................... | .................... |
| 29781 | MT534336.1 | \|Severe acute respiratory syndrome coro | .................... | .................... | .................... | .................... | .................... | .................... |
| 29782 | MT534337.1 | \|Severe acute respiratory syndrome coro | .................... | .................... | .................... | .................... | .................... | .................... |
| 29783 | MT534338.1 | \|Severe acute respiratory syndrome coro | .................... | .................... | .................... | .................... | .................... | .................... |
| 29784 | MT534339.1 | \|Severe acute respiratory syndrome coro | .................... | .................... | .................... | .................... | .................... | .................... |
| 29785 | MT534630.1 | \|Severe acute respiratory syndrome coro | .................... | .................... | .................... | .................... | .................... | .................... |
| 29786 | MT535472.1 | \|Severe acute respiratory syndrome coro | .................... | .................... | .................... | .................... | .................... | .................... |
| 29787 | MT535481.1 | \|Severe acute respiratory syndrome coro | .................... | .................... | .................... | .................... | .................... | .................... |
| 29788 | MT535486.1 | \|Severe acute respiratory syndrome coro | .................... | .................... | .................... | .................... | .................... | .................... |
| 29789 | MT535487.1 | \|Severe acute respiratory syndrome coro | .................... | .................... | .................... | .................... | .................... | .................... |
| 29790 | MT535488.1 | \|Severe acute respiratory syndrome coro | .................... | .................... | .................... | .................... | .................... | .................... |
| 29791 | MT535489.1 | \|Severe acute respiratory syndrome coro | .................... | .................... | .................... | .................... | .................... | .................... |
| 29792 | MT535490.1 | \|Severe acute respiratory syndrome coro | .................... | .................... | .................... | .................... | .................... | .................... |
| 29793 | MT535491.1 | \|Severe acute respiratory syndrome coro | .................... | .................... | .................... | .................... | .................... | .................... |
| 29794 | MT535492.1 | \|Severe acute respiratory syndrome coro | .................... | .................... | .................... | .................... | .................... | .................... |

```
29795 MT535493.1 |Severe acute respiratory syndrome coro    ....................  ....................  ....................  ....................  ....................  ....................
29796 MT535494.1 |Severe acute respiratory syndrome coro    ....................  ....................  ....................  ....................  ....................  ....................
29797 MT535495.1 |Severe acute respiratory syndrome coro    ....................  ....................  ....................  ....................  ....................  ....................
29798 MT535496.1 |Severe acute respiratory syndrome coro    ....................  ....................  ....................  ....................  ....................  ....................
29799 MT535497.1 |Severe acute respiratory syndrome coro    ....................  ....................  ....................  ....................  ....................  ....................
29800 MT535499.1 |Severe acute respiratory syndrome coro    ....................  ....................  ....................  ....................  ....................  ....................
29801 MT535500.1 |Severe acute respiratory syndrome coro    ....................  ....................  ....................  ....................  ....................  ....................
29802 MT535501.1 |Severe acute respiratory syndrome coro    ....................  ....................  ....................  ....................  ....................  ....................
29803 MT535502.1 |Severe acute respiratory syndrome coro    ....................  ....................  ....................  ....................  ....................  ....................
29804 MT535503.1 |Severe acute respiratory syndrome coro    ....................  ....................  ....................  ....................  ....................  ....................
29805 MT535505.1 |Severe acute respiratory syndrome coro    ....................  ....................  ....................  ....................  ....................  ....................
29806 MT535508.1 |Severe acute respiratory syndrome coro    ....................  ....................  ....................  ....................  ....................  ....................
29807 MT535509.1 |Severe acute respiratory syndrome coro    ....................  ....................  ....................  ....................  ....................  ....................
29808 MT536176.1 |Severe acute respiratory syndrome coro    ....................  ....................  ....................  ....................  ....................  ....................
29809 MT536177.1 |Severe acute respiratory syndrome coro    ....................  ....................  ....................  ....................  ....................  ....................
29810 MT536178.1 |Severe acute respiratory syndrome coro    ....................  ....................  ....................  ....................  ....................  ....................
29811 MT536179.1 |Severe acute respiratory syndrome coro    ....................  ....................  ....................  ....................  ....................  ....................
29812 MT536180.1 |Severe acute respiratory syndrome coro    ....................  ....................  ....................  ....................  ....................  ....................
29813 MT536181.1 |Severe acute respiratory syndrome coro    ....................  ....................  ....................  ....................  ....................  ....................
29814 MT536182.1 |Severe acute respiratory syndrome coro    ....................  ....................  ....................  ....................  ....................  ....................
29815 MT536183.1 |Severe acute respiratory syndrome coro    ....................  ....................  ....................  ....................  ....................  ....................
29816 MT536184.1 |Severe acute respiratory syndrome coro    ....................  ....................  ....................  ....................  ....................  ....................
29817 MT536185.1 |Severe acute respiratory syndrome coro    ....................  ....................  ....................  ....................  ....................  ....................
29818 MT536186.1 |Severe acute respiratory syndrome coro    ....................  ....................  ....................  ....................  ....................  ....................
29819 MT536187.1 |Severe acute respiratory syndrome coro    ....................  ....................  ....................  ....................  ....................  ....................
29820 MT536188.1 |Severe acute respiratory syndrome coro    ....................  ....................  ....................  ....................  ....................  ....................
29821 MT536189.1 |Severe acute respiratory syndrome coro    ....................  ....................  ....................  ....................  ....................  ....................
29822 MT536190.1 |Severe acute respiratory syndrome coro    ....................  ....................  ....................  ....................  ....................  ....................
29823 MT536956.1 |Severe acute respiratory syndrome coro    ....................  ....................  ....................  ....................  ....................  ....................
29824 MT536961.1 |Severe acute respiratory syndrome coro    ....................  ....................  ....................  ....................  ....................  ....................
29825 MT536962.1 |Severe acute respiratory syndrome coro    ....................  ....................  ....................  ....................  ....................  ....................
29826 MT536964.1 |Severe acute respiratory syndrome coro    ....................  ....................  ....................  ....................  ....................  ....................
29827 MT536965.1 |Severe acute respiratory syndrome coro    ....................  ....................  ....................  ....................  ....................  ....................
29828 MT536970.1 |Severe acute respiratory syndrome coro    ....................  ....................  ....................  ....................  ....................  ....................
29829 MT536971.1 |Severe acute respiratory syndrome coro    ....................  ....................  ....................  ....................  ....................  ....................
29830 MT536972.1 |Severe acute respiratory syndrome coro    ....................  ....................  ....................  ....................  ....................  ....................
29831 MT536974.1 |Severe acute respiratory syndrome coro    ....................  ....................  ....................  ....................  ....................  ....................
29832 MT536975.1 |Severe acute respiratory syndrome coro    ....................  ....................  ....................  ....................  ....................  ....................
29833 MT536976.1 |Severe acute respiratory syndrome coro    ....................  ....................  ....................  ....................  ....................  ....................
29834 MT536977.1 |Severe acute respiratory syndrome coro    ....................  ....................  ....................  ....................  ....................  ....................
29835 MT525950.1 |Severe acute respiratory syndrome coro    ....................  ....................  ....................  ....................  ....................  ....................
29836 MT527178.1 |Severe acute respiratory syndrome coro    ....................  ....................  ....................  ....................  ....................  ....................
29837 MT527184.1 |Severe acute respiratory syndrome coro    ....................  ....................  ....................  ....................  ....................  ....................
29838 MT528235.1 |Severe acute respiratory syndrome coro    ....................  ....................  ....................  ....................  ....................  ....................
29839 MT528237.1 |Severe acute respiratory syndrome coro    ....................  ....................  ....................  ....................  ....................  ....................
29840 MT528238.1 |Severe acute respiratory syndrome coro    ....................  ....................  ....................  ....................  ....................  ....................
29841 MT528239.1 |Severe acute respiratory syndrome coro    ....................  ....................  ....................  ....................  ....................  ....................
29842 MT528598.1 |Severe acute respiratory syndrome coro    ....................  ....................  ....................  ....................  ....................  ....................
29843 MT528599.1 |Severe acute respiratory syndrome coro    ....................  ....................  ....................  ....................  ....................  ....................
29844 MT528600.1 |Severe acute respiratory syndrome coro    ....................  ....................  ....................  ....................  ....................  ....................
29845 MT528601.1 |Severe acute respiratory syndrome coro    ....................  ....................  ....................  ....................  ....................  ....................
29846 MT528602.1 |Severe acute respiratory syndrome coro    ....................  ....................  ....................  ....................  ....................  ....................
29847 MT528603.1 |Severe acute respiratory syndrome coro    ....................  ....................  ....................  ....................  ....................  ....................
29848 MT528604.1 |Severe acute respiratory syndrome coro    ....................  ....................  ....................  ....................  ....................  ....................
29849 MT528605.1 |Severe acute respiratory syndrome coro    ....................  ....................  ....................  ....................  ....................  ....................
29850 MT528606.1 |Severe acute respiratory syndrome coro    ....................  ....................  ....................  ....................  ....................  ....................
29851 MT528607.1 |Severe acute respiratory syndrome coro    ....................  ....................  ....................  ....................  ....................  ....................
29852 MT528608.1 |Severe acute respiratory syndrome coro    ....................  ....................  ....................  ....................  ....................  ....................
29853 MT528609.1 |Severe acute respiratory syndrome coro    ....................  ....................  ....................  ....................  ....................  ....................
29854 MT528610.1 |Severe acute respiratory syndrome coro    ....................  ....................  ....................  ....................  ....................  ....................
29855 MT528611.1 |Severe acute respiratory syndrome coro    ....................  ....................  ....................  ....................  ....................  ....................
29856 MT528612.1 |Severe acute respiratory syndrome coro    ....................  ....................  ....................  ....................  ....................  ....................
29857 MT528613.1 |Severe acute respiratory syndrome coro    ....................  ....................  ....................  ....................  ....................  ....................
29858 MT528614.1 |Severe acute respiratory syndrome coro    ....................  ....................  ....................  ....................  ....................  ....................
29859 MT528615.1 |Severe acute respiratory syndrome coro    ....................  ....................  ....................  ....................  ....................  ....................
29860 MT528616.1 |Severe acute respiratory syndrome coro    ....................  ....................  ....................  ....................  ....................  ....................
29861 MT528617.1 |Severe acute respiratory syndrome coro    ....................  ....................  ....................  ....................  ....................  ....................
29862 MT528618.1 |Severe acute respiratory syndrome coro    ....................  ....................  ....................  ....................  ....................  ....................
29863 MT528619.1 |Severe acute respiratory syndrome coro    ....................  ....................  ....................  ....................  ....................  ....................
29864 MT528620.1 |Severe acute respiratory syndrome coro    ....................  ....................  ....................  ....................  ....................  ....................
29865 MT528621.1 |Severe acute respiratory syndrome coro    ....................  ....................  ....................  ....................  ....................  ....................
29866 MT528622.1 |Severe acute respiratory syndrome coro    ....................  ....................  ....................  ....................  ....................  ....................
29867 MT528623.1 |Severe acute respiratory syndrome coro    ....................  ....................  ....................  ....................  ....................  ....................
29868 MT528624.1 |Severe acute respiratory syndrome coro    ....................  ....................  ....................  ....................  ....................  ....................
29869 MT528625.1 |Severe acute respiratory syndrome coro    ....................  ....................  ....................  ....................  ....................  ....................
29870 MT528626.1 |Severe acute respiratory syndrome coro    ....................  ....................  ....................  ....................  ....................  ....................
29871 MT528627.1 |Severe acute respiratory syndrome coro    ....................  ....................  ....................  ....................  ....................  ....................
29872 MT528628.1 |Severe acute respiratory syndrome coro    ....................  ....................  ....................  ....................  ....................  ....................
```

| | | | | | | | | |
|---|---|---|---|---|---|---|---|---|
| 29873 | MT528629.1 | Severe acute respiratory syndrome coro | ···················· | ···················· | ···················· | ···················· | ···················· | ···················· |
| 29874 | MT528630.1 | Severe acute respiratory syndrome coro | ···················· | ···················· | ···················· | ···················· | ···················· | ···················· |
| 29875 | MT528631.1 | Severe acute respiratory syndrome coro | ···················· | ···················· | ···················· | ···················· | ···················· | ···················· |
| 29876 | MT531537.2 | Severe acute respiratory syndrome coro | ···················· | ···················· | ···················· | ···················· | ···················· | ···················· |
| 29877 | MT532435.1 | Severe acute respiratory syndrome coro | ···················· | ···················· | ···················· | ···················· | ···················· | ···················· |
| 29878 | MT532436.1 | Severe acute respiratory syndrome coro | ···················· | ···················· | ···················· | ···················· | ···················· | ···················· |
| 29879 | MT532437.1 | Severe acute respiratory syndrome coro | ···················· | ···················· | ···················· | ···················· | ···················· | ···················· |
| 29880 | MT532438.1 | Severe acute respiratory syndrome coro | ···················· | ···················· | ···················· | ···················· | ···················· | ···················· |
| 29881 | MT532557.1 | Severe acute respiratory syndrome coro | ···················· | ···················· | ···················· | ···················· | ···················· | ···················· |
| 29882 | MT533197.1 | Severe acute respiratory syndrome coro | ···················· | ···················· | ···················· | ···················· | ···················· | ···················· |
| 29883 | MT533198.1 | Severe acute respiratory syndrome coro | ···················· | ···················· | ···················· | ···················· | ···················· | ···················· |
| 29884 | MT533199.1 | Severe acute respiratory syndrome coro | ···················· | ···················· | ···················· | ···················· | ···················· | ···················· |
| 29885 | MT533200.1 | Severe acute respiratory syndrome coro | ···················· | ···················· | ···················· | ···················· | ···················· | ···················· |
| 29886 | MT533201.1 | Severe acute respiratory syndrome coro | ···················· | ···················· | ···················· | ···················· | ···················· | ···················· |
| 29887 | MT533202.1 | Severe acute respiratory syndrome coro | ···················· | ···················· | ···················· | ···················· | ···················· | ···················· |
| 29888 | MT533203.1 | Severe acute respiratory syndrome coro | ···················· | ···················· | ···················· | ···················· | ···················· | ···················· |
| 29889 | MT533204.1 | Severe acute respiratory syndrome coro | ···················· | ···················· | ···················· | ···················· | ···················· | ···················· |
| 29890 | MT533205.1 | Severe acute respiratory syndrome coro | ···················· | ···················· | ···················· | ···················· | ···················· | ···················· |
| 29891 | MT533206.1 | Severe acute respiratory syndrome coro | ···················· | ···················· | ···················· | ···················· | ···················· | ···················· |
| 29892 | MT533208.1 | Severe acute respiratory syndrome coro | ···················· | ···················· | ···················· | ···················· | ···················· | ···················· |
| 29893 | MT533210.1 | Severe acute respiratory syndrome coro | ···················· | ···················· | ···················· | ···················· | ···················· | ···················· |
| 29894 | MT533212.1 | Severe acute respiratory syndrome coro | ···················· | ···················· | ···················· | ···················· | ···················· | ···················· |
| 29895 | MT533215.1 | Severe acute respiratory syndrome coro | ···················· | ···················· | ···················· | ···················· | ···················· | ···················· |
| 29896 | MT533217.1 | Severe acute respiratory syndrome coro | ···················· | ···················· | ···················· | ···················· | ···················· | ···················· |
| 29897 | MT533218.1 | Severe acute respiratory syndrome coro | ···················· | ···················· | ···················· | ···················· | ···················· | ···················· |
| 29898 | MT533219.1 | Severe acute respiratory syndrome coro | ···················· | ···················· | ···················· | ···················· | ···················· | ···················· |
| 29899 | MT533221.1 | Severe acute respiratory syndrome coro | ···················· | ···················· | ···················· | ···················· | ···················· | ···················· |
| 29900 | MT533222.1 | Severe acute respiratory syndrome coro | ···················· | ···················· | ···················· | ···················· | ···················· | ···················· |
| 29901 | MT533223.1 | Severe acute respiratory syndrome coro | ···················· | ···················· | ···················· | ···················· | ···················· | ···················· |
| 29902 | MT533225.1 | Severe acute respiratory syndrome coro | ···················· | ···················· | ···················· | ···················· | ···················· | ···················· |
| 29903 | MT533226.1 | Severe acute respiratory syndrome coro | ···················· | ···················· | ···················· | ···················· | ···················· | ···················· |
| 29904 | MT533227.1 | Severe acute respiratory syndrome coro | ···················· | ···················· | ···················· | ···················· | ···················· | ···················· |
| 29905 | MT533228.1 | Severe acute respiratory syndrome coro | ···················· | ···················· | ···················· | ···················· | ···················· | ···················· |
| 29906 | MT533229.1 | Severe acute respiratory syndrome coro | ···················· | ···················· | ···················· | ···················· | ···················· | ···················· |
| 29907 | MT533230.1 | Severe acute respiratory syndrome coro | ···················· | ···················· | ···················· | ···················· | ···················· | ···················· |
| 29908 | MT533232.1 | Severe acute respiratory syndrome coro | ···················· | ···················· | ···················· | ···················· | ···················· | ···················· |
| 29909 | MT533233.1 | Severe acute respiratory syndrome coro | ···················· | ···················· | ···················· | ···················· | ···················· | ···················· |
| 29910 | MT533234.1 | Severe acute respiratory syndrome coro | ···················· | ···················· | ···················· | ···················· | ···················· | ···················· |
| 29911 | MT533235.1 | Severe acute respiratory syndrome coro | ···················· | ···················· | ···················· | ···················· | ···················· | ···················· |
| 29912 | MT533236.1 | Severe acute respiratory syndrome coro | ···················· | ···················· | ···················· | ···················· | ···················· | ···················· |
| 29913 | MT533237.1 | Severe acute respiratory syndrome coro | ···················· | ···················· | ···················· | ···················· | ···················· | ···················· |
| 29914 | MT533238.1 | Severe acute respiratory syndrome coro | ···················· | ···················· | ···················· | ···················· | ···················· | ···················· |
| 29915 | MT506981.1 | Severe acute respiratory syndrome coro | ···················· | ···················· | ···················· | ···················· | ···················· | ···················· |
| 29916 | MT507018.1 | Severe acute respiratory syndrome coro | ···················· | ···················· | ···················· | ···················· | ···················· | ···················· |
| 29917 | MT507032.2 | Severe acute respiratory syndrome coro | ···················· | ···················· | ···················· | ···················· | ···················· | ···················· |
| 29918 | MT517420.1 | Severe acute respiratory syndrome coro | ···················· | ···················· | ···················· | ···················· | ···················· | ···················· |
| 29919 | MT517421.1 | Severe acute respiratory syndrome coro | ···················· | ···················· | ···················· | ···················· | ···················· | ···················· |
| 29920 | MT517422.1 | Severe acute respiratory syndrome coro | ···················· | ···················· | ···················· | ···················· | ···················· | ···················· |
| 29921 | MT517423.1 | Severe acute respiratory syndrome coro | ···················· | ···················· | ···················· | ···················· | ···················· | ···················· |
| 29922 | MT517424.1 | Severe acute respiratory syndrome coro | ···················· | ···················· | ···················· | ···················· | ···················· | ···················· |
| 29923 | MT517425.1 | Severe acute respiratory syndrome coro | ···················· | ···················· | ···················· | ···················· | ···················· | ···················· |
| 29924 | MT517426.1 | Severe acute respiratory syndrome coro | ···················· | ···················· | ···················· | ···················· | ···················· | ···············A···· |
| 29925 | MT517427.1 | Severe acute respiratory syndrome coro | ···················· | ···················· | ···················· | ···················· | ···················· | ···················· |
| 29926 | MT517428.1 | Severe acute respiratory syndrome coro | ···················· | ···················· | ···················· | ···················· | ···················· | ···················· |
| 29927 | MT517429.1 | Severe acute respiratory syndrome coro | ···················· | ···················· | ···················· | ···················· | ···················· | ···················· |
| 29928 | MT517430.1 | Severe acute respiratory syndrome coro | ···················· | ···················· | ···················· | ···················· | ···················· | ···················· |
| 29929 | MT517431.1 | Severe acute respiratory syndrome coro | ···················· | ···················· | ···················· | ···················· | ···················· | ···················· |
| 29930 | MT517432.1 | Severe acute respiratory syndrome coro | ···················· | ···················· | ···················· | ···················· | ···················· | ···················· |
| 29931 | MT517433.1 | Severe acute respiratory syndrome coro | ···················· | ···················· | ···················· | ···················· | ···················· | ···················· |
| 29932 | MT517434.1 | Severe acute respiratory syndrome coro | ···················· | ···················· | ···················· | ···················· | ···················· | ···················· |
| 29933 | MT517436.1 | Severe acute respiratory syndrome coro | ···················· | ···················· | ···················· | ···················· | ···················· | ···················· |
| 29934 | MT517437.1 | Severe acute respiratory syndrome coro | ···················· | ···················· | ···················· | ···················· | ···················· | ···················· |
| 29935 | MT522852.1 | Severe acute respiratory syndrome coro | ···················· | ···················· | ···················· | ···················· | ···················· | ···················· |
| 29936 | MT522853.1 | Severe acute respiratory syndrome coro | ···················· | ···················· | ···················· | ···················· | ···················· | ···················· |
| 29937 | MT522854.1 | Severe acute respiratory syndrome coro | ···················· | ···················· | ···················· | ···················· | ···················· | ···················· |
| 29938 | MT522855.1 | Severe acute respiratory syndrome coro | ···················· | ···················· | ···················· | ···················· | ···················· | ···················· |
| 29939 | MT510718.1 | Severe acute respiratory syndrome coro | ···················· | ···················· | ···················· | ···················· | ···················· | ···················· |
| 29940 | MT510719.1 | Severe acute respiratory syndrome coro | ···················· | ···················· | ···················· | ···················· | ···················· | ···················· |
| 29941 | MT510721.1 | Severe acute respiratory syndrome coro | ···················· | ···················· | ···················· | ···················· | ···················· | ···················· |
| 29942 | MT510722.1 | Severe acute respiratory syndrome coro | ···················· | ···················· | ···················· | ···················· | ···················· | ···················· |
| 29943 | MT510725.1 | Severe acute respiratory syndrome coro | ···················· | ·A·················· | ···················· | ···················· | ···················· | ···················· |
| 29944 | MT510726.1 | Severe acute respiratory syndrome coro | ···················· | ···················· | ···················· | ···················· | ···················· | ···················· |
| 29945 | MT510727.1 | Severe acute respiratory syndrome coro | ···················· | ···················· | ···················· | ···················· | ···················· | ···················· |
| 29946 | MT510728.1 | Severe acute respiratory syndrome coro | ···················· | ···················· | ···················· | ···················· | ···················· | ···················· |
| 29947 | MT510999.1 | Severe acute respiratory syndrome coro | ···················· | ···················· | ···················· | ···················· | ···················· | ···················· |
| 29948 | MT511066.1 | Severe acute respiratory syndrome coro | ···················· | ···················· | ···················· | ···················· | ···················· | ···················· |
| 29949 | MT511067.1 | Severe acute respiratory syndrome coro | ···················· | ···················· | ···················· | ···················· | ···················· | ···················· |
| 29950 | MT511068.1 | Severe acute respiratory syndrome coro | ···················· | ···················· | ···················· | ···················· | ···················· | ···················· |

```
29951 MT511069.1 |Severe acute respiratory syndrome coro    ····················  ····················  ····················  ····················  ····················  ····················
29952 MT511070.1 |Severe acute respiratory syndrome coro    ····················  ····················  ····················  ····················  ····················  ····················
29953 MT511071.1 |Severe acute respiratory syndrome coro    ····················  ····················  ····················  ····················  ····················  ····················
29954 MT511072.1 |Severe acute respiratory syndrome coro    ····················  ····················  ····················  ····················  ····················  ····················
29955 MT511073.1 |Severe acute respiratory syndrome coro    ····················  ····················  ····················  ····················  ····················  ····················
29956 MT511074.1 |Severe acute respiratory syndrome coro    ····················  ····················  ····················  ····················  ····················  ····················
29957 MT511075.1 |Severe acute respiratory syndrome coro    ····················  ····················  ····················  ····················  ····················  ····················
29958 MT511076.1 |Severe acute respiratory syndrome coro    ····················  ····················  ····················  ····················  ····················  ····················
29959 MT511077.1 |Severe acute respiratory syndrome coro    ····················  ····················  ····················  ····················  ····················  ····················
29960 MT511078.1 |Severe acute respiratory syndrome coro    ····················  ····················  ····················  ····················  ····················  ····················
29961 MT511079.1 |Severe acute respiratory syndrome coro    ····················  ····················  ····················  ····················  ····················  ····················
29962 MT511080.1 |Severe acute respiratory syndrome coro    ····················  ····················  ····················  ····················  ····················  ····················
29963 MT511081.1 |Severe acute respiratory syndrome coro    ····················  ····················  ····················  ····················  ····················  ····················
29964 MT511082.1 |Severe acute respiratory syndrome coro    ····················  ····················  ····················  ····················  ····················  ····················
29965 MT512415.1 |Severe acute respiratory syndrome coro    ····················  ····················  ····················  ····················  ····················  ····················
29966 MT512416.1 |Severe acute respiratory syndrome coro    ····················  ····················  ····················  ····················  ····················  ····················
29967 MT512417.1 |Severe acute respiratory syndrome coro    ····················  ····················  ····················  ····················  ····················  ····················
29968 MT512418.1 |Severe acute respiratory syndrome coro    ····················  ····················  ····················  ····················  ····················  ····················
29969 MT512419.1 |Severe acute respiratory syndrome coro    ····················  ····················  ····················  ····················  ····················  ····················
29970 MT512420.1 |Severe acute respiratory syndrome coro    ····················  ····················  ····················  ····················  ····················  ····················
29971 MT512421.1 |Severe acute respiratory syndrome coro    ····················  ····················  ····················  ····················  ····················  ····················
29972 MT512422.1 |Severe acute respiratory syndrome coro    ····················  ····················  ····················  ····················  ····················  ····················
29973 MT512423.1 |Severe acute respiratory syndrome coro    ····················  ····················  ····················  ····················  ····················  ····················
29974 MT512424.1 |Severe acute respiratory syndrome coro    ····················  ····················  ····················  ····················  ····················  ····················
29975 MT512425.1 |Severe acute respiratory syndrome coro    ····················  ····················  ····················  ····················  ····················  ····················
29976 MT512426.1 |Severe acute respiratory syndrome coro    ····················  ····················  ····················  ····················  ····················  ····················
29977 MT512427.1 |Severe acute respiratory syndrome coro    ····················  ····················  ····················  ····················  ····················  ····················
29978 MT512428.1 |Severe acute respiratory syndrome coro    ····················  ····················  ····················  ····················  ····················  ····················
29979 MT512429.1 |Severe acute respiratory syndrome coro    ····················  ····················  ····················  ····················  ····················  ····················
29980 MT512430.1 |Severe acute respiratory syndrome coro    ····················  ····················  ····················  ····················  ····················  ····················
29981 MT512431.1 |Severe acute respiratory syndrome coro    ····················  ····················  ····················  ····················  ····················  ····················
29982 MT512433.1 |Severe acute respiratory syndrome coro    ····················  ····················  ····················  ····················  ····················  ····················
29983 MT512434.1 |Severe acute respiratory syndrome coro    ····················  ····················  ····················  ····················  ····················  ····················
29984 MT512435.1 |Severe acute respiratory syndrome coro    ····················  ····················  ····················  ····················  ····················  ····················
29985 MT512436.1 |Severe acute respiratory syndrome coro    ····················  ····················  ····················  ····················  ····················  ····················
29986 MT512437.1 |Severe acute respiratory syndrome coro    ····················  ····················  ····················  ····················  ····················  ····················
29987 MT512438.1 |Severe acute respiratory syndrome coro    ····················  ····················  ····················  ····················  ····················  ····················
29988 MT512439.1 |Severe acute respiratory syndrome coro    ····················  ····················  ····················  ····················  ····················  ····················
29989 MT512440.1 |Severe acute respiratory syndrome coro    ····················  ····················  ····················  ····················  ····················  ····················
29990 MT512441.1 |Severe acute respiratory syndrome coro    ····················  ····················  ····················  ····················  ····················  ····················
29991 MT512442.1 |Severe acute respiratory syndrome coro    ····················  ····················  ····················  ····················  ····················  ····················
29992 MT512443.1 |Severe acute respiratory syndrome coro    ····················  ····················  ····················  ····················  ····················  ····················
29993 MT512444.1 |Severe acute respiratory syndrome coro    ····················  ····················  ····················  ····················  ····················  ····················
29994 MT512445.1 |Severe acute respiratory syndrome coro    ····················  ····················  ····················  ····················  ····················  ····················
29995 MT512446.1 |Severe acute respiratory syndrome coro    ····················  ····················  ····················  ····················  ····················  ····················
29996 MT512447.1 |Severe acute respiratory syndrome coro    ····················  ····················  ····················  ····················  ····················  ····················
29997 MT512644.1 |Severe acute respiratory syndrome coro    ····················  ····················  ····················  ····················  ····················  ····················
29998 MT512645.1 |Severe acute respiratory syndrome coro    ····················  ····················  ····················  ····················  ····················  ····················
29999 MT509958.1 |Severe acute respiratory syndrome coro    ····················  ····················  ····················  ····················  ····················  ····················
30000 MT509959.1 |Severe acute respiratory syndrome coro    ····················  ····················  ····················  ····················  ····················  ····················
30001 MT509649.1 |Severe acute respiratory syndrome coro    ····················  ····················  ····················  ····················  ····················  ····················
30002 MT509650.1 |Severe acute respiratory syndrome coro    ····················  ····················  ····················  ····················  ····················  ····················
30003 MT509651.1 |Severe acute respiratory syndrome coro    ····················  ·A··················  ····················  ····················  ····················  ··A·················
30004 MT509656.1 |Severe acute respiratory syndrome coro    ····················  ····················  ····················  ····················  ····················  ····················
30005 MT509657.1 |Severe acute respiratory syndrome coro    ····················  ····················  ····················  ····················  ····················  ····················
30006 MT509658.1 |Severe acute respiratory syndrome coro    ····················  ····················  ····················  ····················  ····················  ····················
30007 MT509659.1 |Severe acute respiratory syndrome coro    ····················  ····················  ····················  ····················  ····················  ····················
30008 MT509669.1 |Severe acute respiratory syndrome coro    ····················  ····················  ····················  ····················  ····················  ····················
30009 MT509674.1 |Severe acute respiratory syndrome coro    ····················  ····················  ····················  ····················  ····················  ····················
30010 MT509683.1 |Severe acute respiratory syndrome coro    ····················  ····················  ····················  ····················  ····················  ····················
30011 MT509685.1 |Severe acute respiratory syndrome coro    ····················  ····················  ····················  ····················  ····················  ····················
30012 MT509702.1 |Severe acute respiratory syndrome coro    ····················  ····················  ····················  ····················  ····················  ····················
30013 MT509703.1 |Severe acute respiratory syndrome coro    ····················  ····················  ····················  ····················  ····················  ····················
30014 MT509452.1 |Severe acute respiratory syndrome coro    ····················  ····················  ····················  ····················  ····················  ····················
30015 MT509453.1 |Severe acute respiratory syndrome coro    ····················  ····················  ····················  ····················  ····················  ····················
30016 MT509454.1 |Severe acute respiratory syndrome coro    ····················  ····················  ····················  ····················  ····················  ····················
30017 MT509455.1 |Severe acute respiratory syndrome coro    ····················  ····················  ····················  ····················  ····················  ····················
30018 MT509456.1 |Severe acute respiratory syndrome coro    ····················  ····················  ····················  ····················  ····················  ····················
30019 MT509457.1 |Severe acute respiratory syndrome coro    ····················  ····················  ····················  ····················  ····················  ····················
30020 MT509458.1 |Severe acute respiratory syndrome coro    ····················  ····················  ····················  ····················  ····················  ····················
30021 MT509459.1 |Severe acute respiratory syndrome coro    ····················  ····················  ····················  ····················  ····················  ····················
30022 MT509460.1 |Severe acute respiratory syndrome coro    ····················  ····················  ····················  ····················  ····················  ····················
30023 MT509461.1 |Severe acute respiratory syndrome coro    ····················  ····················  ····················  ····················  ····················  ····················
30024 MT509462.1 |Severe acute respiratory syndrome coro    ····················  ····················  ····················  ····················  ····················  ····················
30025 MT509463.1 |Severe acute respiratory syndrome coro    ····················  ····················  ····················  ····················  ····················  ····················
30026 MT509464.1 |Severe acute respiratory syndrome coro    ····················  ····················  ····················  ····················  ····················  ····················
30027 MT509465.1 |Severe acute respiratory syndrome coro    ····················  ····················  ····················  ····················  ····················  ····················
30028 MT509466.1 |Severe acute respiratory syndrome coro    ····················  ····················  ····················  ····················  ····················  ····················
```

```
30029 MT509467.1 |Severe acute respiratory syndrome coro    ....................  ....................  ....................  ....................  ....................  ....................
30030 MT509468.1 |Severe acute respiratory syndrome coro    ....................  ....................  ....................  ....................  ....................  ....................
30031 MT509471.1 |Severe acute respiratory syndrome coro    ....................  ....................  ....................  ....................  ....................  ....................
30032 MT509472.1 |Severe acute respiratory syndrome coro    ....................  ....................  ....................  ....................  ....................  ....................
30033 MT509474.1 |Severe acute respiratory syndrome coro    ....................  ....................  ....................  ....................  ....................  ....................
30034 MT509475.1 |Severe acute respiratory syndrome coro    ....................  ....................  ....................  ....................  ....................  ....................
30035 MT509477.1 |Severe acute respiratory syndrome coro    ....................  ....................  ....................  ....................  ....................  ....................
30036 MT509480.1 |Severe acute respiratory syndrome coro    ....................  ....................  ....................  ....................  ....................  ....................
30037 MT509486.1 |Severe acute respiratory syndrome coro    ....................  ....................  ....................  ....................  ....................  ....................
30038 MT509488.1 |Severe acute respiratory syndrome coro    ....................  ....................  ....................  ....................  ....................  ....................
30039 MT509489.1 |Severe acute respiratory syndrome coro    ....................  ....................  ....................  ....................  ....................  ....................
30040 MT509490.1 |Severe acute respiratory syndrome coro    ....................  ....................  ....................  ....................  ....................  ....................
30041 MT509491.1 |Severe acute respiratory syndrome coro    ....................  ....................  ....................  ....................  ....................  ....................
30042 MT509492.1 |Severe acute respiratory syndrome coro    ....................  ....................  ....................  ....................  ....................  ....................
30043 MT509493.1 |Severe acute respiratory syndrome coro    ....................  ....................  ....................  ....................  ....................  ....................
30044 MT509494.1 |Severe acute respiratory syndrome coro    ....................  ....................  ....................  ....................  ....................  ....................
30045 MT509495.1 |Severe acute respiratory syndrome coro    ....................  ....................  ....................  ....................  ....................  ....................
30046 MT509496.1 |Severe acute respiratory syndrome coro    ....................  ....................  ....................  ....................  ....................  ....................
30047 MT509497.1 |Severe acute respiratory syndrome coro    ....................  ....................  ....................  ....................  ....................  ....................
30048 MT509498.1 |Severe acute respiratory syndrome coro    ....................  .A..................  ....................  ....................  ....................  ..A.................
30049 MT509499.1 |Severe acute respiratory syndrome coro    ....................  .A..................  ....................  ....................  ....................  ..A.................
30050 MT509500.1 |Severe acute respiratory syndrome coro    ....................  ....................  ....................  ....................  ....................  ....................
30051 MT509501.1 |Severe acute respiratory syndrome coro    ....................  ....................  ....................  ....................  ....................  ....................
30052 MT509502.1 |Severe acute respiratory syndrome coro    ....................  .A..................  ....................  ....................  ....................  ..A.................
30053 MT509503.1 |Severe acute respiratory syndrome coro    ....................  ....................  ....................  ....................  ....................  ....................
30054 MT509504.1 |Severe acute respiratory syndrome coro    ....................  ....................  ....................  ....................  ....................  ....................
30055 MT509505.1 |Severe acute respiratory syndrome coro    ....................  .A..................  ....................  ....................  ....................  ..A.................
30056 MT509506.1 |Severe acute respiratory syndrome coro    ....................  ....................  ....................  ....................  ....................  ....................
30057 MT509507.1 |Severe acute respiratory syndrome coro    ....................  ....................  ....................  ....................  ....................  ....................
30058 MT509508.1 |Severe acute respiratory syndrome coro    ....................  .A..................  ....................  ....................  ....................  ..A.................
30059 MT509509.1 |Severe acute respiratory syndrome coro    ....................  ....................  ....................  ....................  ....................  ....................
30060 MT509510.1 |Severe acute respiratory syndrome coro    ....................  ....................  ....................  ....................  ....................  ....................
30061 MT509511.1 |Severe acute respiratory syndrome coro    ....................  ....................  ....................  ....................  ....................  ....................
30062 MT509512.1 |Severe acute respiratory syndrome coro    ....................  ....................  ....................  ....................  ....................  ....................
30063 MT506201.1 |Severe acute respiratory syndrome coro    ....................  ....................  ....................  ....................  ....................  ....................
30064 MT506203.1 |Severe acute respiratory syndrome coro    ....................  ....................  ....................  ....................  ....................  ....................
30065 MT506205.1 |Severe acute respiratory syndrome coro    ....................  ....................  ....................  ....................  ....................  ....................
30066 MT506209.1 |Severe acute respiratory syndrome coro    ....................  ....................  ....................  ....................  ....................  ....................
30067 MT506221.1 |Severe acute respiratory syndrome coro    ....................  ....................  ....................  ....................  ....................  ....................
30068 MT506530.1 |Severe acute respiratory syndrome coro    ....................  ....................  ....................  ....................  ....................  ....................
30069 MT506531.1 |Severe acute respiratory syndrome coro    ....................  ....................  ....................  ....................  ....................  ....................
30070 MT506532.1 |Severe acute respiratory syndrome coro    ....................  ....................  ....................  ....................  ....................  ....................
30071 MT506533.1 |Severe acute respiratory syndrome coro    ....................  ....................  ....................  ....................  ....................  ....................
30072 MT506534.1 |Severe acute respiratory syndrome coro    ....................  ....................  ....................  ....................  ....................  ....................
30073 MT506535.1 |Severe acute respiratory syndrome coro    ....................  ....................  ....................  ....................  ....................  ....................
30074 MT506537.1 |Severe acute respiratory syndrome coro    ....................  ....................  ....................  ....................  ....................  ....................
30075 MT506539.1 |Severe acute respiratory syndrome coro    ....................  ....................  ....................  ....................  ....................  ....................
30076 MT506540.1 |Severe acute respiratory syndrome coro    ....................  ....................  ....................  ....................  ....................  ....................
30077 MT506541.1 |Severe acute respiratory syndrome coro    ....................  ....................  ....................  ....................  ....................  ....................
30078 MT506544.1 |Severe acute respiratory syndrome coro    ....................  ....................  ....................  ....................  ....................  ....................
30079 MT506546.1 |Severe acute respiratory syndrome coro    ....................  ....................  ....................  ....................  ....................  ....................
30080 MT506565.1 |Severe acute respiratory syndrome coro    ....................  ....................  ....................  ....................  ....................  ....................
30081 MT506566.1 |Severe acute respiratory syndrome coro    ....................  ....................  ....................  ....................  ....................  ....................
30082 MT506567.1 |Severe acute respiratory syndrome coro    ....................  ....................  ....................  ....................  ....................  ....................
30083 MT506568.1 |Severe acute respiratory syndrome coro    ....................  ....................  ....................  ....................  ....................  ....................
30084 MT506570.1 |Severe acute respiratory syndrome coro    ....................  ....................  ....................  ....................  ....................  ....................
30085 MT506571.1 |Severe acute respiratory syndrome coro    ....................  ....................  ....................  ....................  ....................  ....................
30086 MT506572.1 |Severe acute respiratory syndrome coro    ....................  ....................  ....................  ....................  ....................  ....................
30087 MT506573.1 |Severe acute respiratory syndrome coro    ....................  ....................  ....................  ....................  ....................  ....................
30088 MT506581.1 |Severe acute respiratory syndrome coro    ....................  ....................  ....................  ....................  ....................  ....................
30089 MT506642.1 |Severe acute respiratory syndrome coro    ....................  ....................  ....................  ....................  ....................  ....................
30090 MT506643.1 |Severe acute respiratory syndrome coro    ....................  ....................  ....................  ....................  ....................  ....................
30091 MT506648.1 |Severe acute respiratory syndrome coro    ....................  ....................  ....................  ....................  ....................  ....................
30092 MT506649.1 |Severe acute respiratory syndrome coro    ....................  ....................  ....................  ....................  ....................  ....................
30093 MT506650.1 |Severe acute respiratory syndrome coro    ....................  ....................  ....................  ....................  ....................  ....................
30094 MT506651.1 |Severe acute respiratory syndrome coro    ....................  ....................  ....................  ....................  ....................  ....................
30095 MT506652.1 |Severe acute respiratory syndrome coro    ....................  ....................  ....................  ....................  ....................  ....................
30096 MT506654.1 |Severe acute respiratory syndrome coro    ....................  ....................  ....................  ....................  ....................  ....................
30097 MT506658.1 |Severe acute respiratory syndrome coro    ....................  ....................  ....................  ....................  ....................  ....................
30098 MT506659.1 |Severe acute respiratory syndrome coro    ....................  ....................  ....................  ....................  ....................  ....................
30099 MT506660.1 |Severe acute respiratory syndrome coro    ....................  ....................  ....................  ....................  ....................  ....................
30100 MT506663.1 |Severe acute respiratory syndrome coro    ....................  ....................  ....................  ....................  ....................  ....................
30101 MT506664.1 |Severe acute respiratory syndrome coro    ....................  ....................  ....................  ....................  ....................  ....................
30102 MT506666.1 |Severe acute respiratory syndrome coro    ....................  ....................  ....................  ....................  ....................  ....................
30103 MT506668.1 |Severe acute respiratory syndrome coro    ....................  ....................  ....................  ....................  ....................  ....................
30104 MT506670.1 |Severe acute respiratory syndrome coro    ....................  ....................  ....................  ....................  ....................  ....................
30105 MT506674.1 |Severe acute respiratory syndrome coro    ....................  ....................  ....................  ....................  ....................  ....................
30106 MT506675.1 |Severe acute respiratory syndrome coro    ....................  ....................  ....................  ....................  ....................  ....................
```

30107 MT506677.1 |Severe acute respiratory syndrome coro .................... .................... .................... .................... .................... ....................
30108 MT506679.1 |Severe acute respiratory syndrome coro .................... .................... .................... .................... .................... ....................
30109 MT506680.1 |Severe acute respiratory syndrome coro .................... .................... .................... .................... .................... ....................
30110 MT506681.1 |Severe acute respiratory syndrome coro .................... .................... .................... .................... .................... ....................
30111 MT506683.1 |Severe acute respiratory syndrome coro .................... .................... .................... .................... .................... ....................
30112 MT506685.1 |Severe acute respiratory syndrome coro .................... .................... .................... .................... .................... ....................
30113 MT506687.1 |Severe acute respiratory syndrome coro .................... .................... .................... .................... .................... ....................
30114 MT506689.1 |Severe acute respiratory syndrome coro .................... .................... .................... .................... .................... ....................
30115 MT506690.1 |Severe acute respiratory syndrome coro .................... .................... .................... .................... .................... ....................
30116 MT506693.1 |Severe acute respiratory syndrome coro .................... .................... .................... .................... .................... ....................
30117 MT506694.1 |Severe acute respiratory syndrome coro .................... .................... .................... .................... .................... ....................
30118 MT506695.1 |Severe acute respiratory syndrome coro .................... .................... .................... .................... .................... ....................
30119 MT506696.1 |Severe acute respiratory syndrome coro .................... .................... .................... .................... .................... ....................
30120 MT506697.1 |Severe acute respiratory syndrome coro .................... .................... .................... .................... .................... ....................
30121 MT506700.1 |Severe acute respiratory syndrome coro .................... .................... .................... .................... .................... ....................
30122 MT506701.1 |Severe acute respiratory syndrome coro .................... .................... .................... .................... .................... ....................
30123 MT506707.1 |Severe acute respiratory syndrome coro .................... .................... .................... .................... .................... ....................
30124 MT506885.1 |Severe acute respiratory syndrome coro .................... .................... .................... .................... .................... ....................
30125 MT506886.1 |Severe acute respiratory syndrome coro .................... .................... .................... .................... .................... ....................
30126 MT506887.1 |Severe acute respiratory syndrome coro .................... .................... .................... .................... .................... ....................
30127 MT506888.1 |Severe acute respiratory syndrome coro .................... .................... .................... .................... .................... ....................
30128 MT506889.1 |Severe acute respiratory syndrome coro .................... .................... .................... .................... .................... ....................
30129 MT506890.1 |Severe acute respiratory syndrome coro .................... .................... .................... .................... .................... ....................
30130 MT506891.1 |Severe acute respiratory syndrome coro .................... .................... .................... .................... .................... ....................
30131 MT506892.1 |Severe acute respiratory syndrome coro .................... .................... .................... .................... .................... ....................
30132 MT506893.1 |Severe acute respiratory syndrome coro .................... .................... .................... .................... .................... ....................
30133 MT506894.1 |Severe acute respiratory syndrome coro .................... .................... .................... .................... .................... ....................
30134 MT506895.1 |Severe acute respiratory syndrome coro .................... .................... .................... .................... .................... ....................
30135 MT506896.1 |Severe acute respiratory syndrome coro .................... .................... .................... .................... .................... ....................
30136 MT506897.1 |Severe acute respiratory syndrome coro .................... .................... .................... .................... .................... ....................
30137 MT506898.1 |Severe acute respiratory syndrome coro .................... .................... .................... .................... .................... ....................
30138 MT506899.1 |Severe acute respiratory syndrome coro .................... .................... .................... .................... .................... ....................
30139 MT506900.1 |Severe acute respiratory syndrome coro .................... .................... .................... .................... .................... ....................
30140 MT506901.1 |Severe acute respiratory syndrome coro .................... .................... .................... .................... .................... ....................
30141 MT506902.1 |Severe acute respiratory syndrome coro .................... .................... .................... .................... .................... ....................
30142 MT506903.1 |Severe acute respiratory syndrome coro .................... .................... .................... .................... .................... ....................
30143 MT506904.1 |Severe acute respiratory syndrome coro .................... .................... .................... .................... .................... ....................
30144 MT506905.1 |Severe acute respiratory syndrome coro .................... .................... .................... .................... .................... ....................
30145 MT506906.1 |Severe acute respiratory syndrome coro .................... .................... .................... .................... .................... ....................
30146 MT506907.1 |Severe acute respiratory syndrome coro .................... .................... .................... .................... .................... ....................
30147 MT507271.1 |Severe acute respiratory syndrome coro .................... .................... .................... .................... .................... ....................
30148 MT507272.1 |Severe acute respiratory syndrome coro .................... .................... .................... .................... .................... ....................
30149 MT507273.1 |Severe acute respiratory syndrome coro .................... .................... .................... .................... .................... ....................
30150 MT507274.1 |Severe acute respiratory syndrome coro .................... .................... .................... .................... .................... ....................
30151 MT507275.1 |Severe acute respiratory syndrome coro .................... .................... .................... .................... .................... ....................
30152 MT507276.1 |Severe acute respiratory syndrome coro .................... .................... .................... .................... .................... ....................
30153 MT507277.1 |Severe acute respiratory syndrome coro .................... .................... .................... .................... .................... ....................
30154 MT507278.1 |Severe acute respiratory syndrome coro .................... .................... .................... .................... .................... ....................
30155 MT507279.1 |Severe acute respiratory syndrome coro .................... .................... .................... .................... .................... ....................
30156 MT507280.1 |Severe acute respiratory syndrome coro .................... .................... .................... .................... .................... ....................
30157 MT507281.1 |Severe acute respiratory syndrome coro .................... .................... .................... .................... .................... ....................
30158 MT507282.1 |Severe acute respiratory syndrome coro .................... .................... .................... .................... .................... ....................
30159 MT507793.1 |Severe acute respiratory syndrome coro .................... .................... .................... .................... .................... ....................
30160 MT507794.1 |Severe acute respiratory syndrome coro .................... .................... .................... .................... .................... ....................
30161 MT507795.1 |Severe acute respiratory syndrome coro .................... .................... .................... .................... .................... ....................
30162 MT507796.1 |Severe acute respiratory syndrome coro .................... .................... .................... .................... .................... ....................
30163 MT499171.1 |Severe acute respiratory syndrome coro .................... .................... .................... .................... .................... ....................
30164 MT499172.1 |Severe acute respiratory syndrome coro .................... .................... .................... .................... .................... ....................
30165 MT499173.1 |Severe acute respiratory syndrome coro .................... .................... .................... .................... .................... ....................
30166 MT499174.1 |Severe acute respiratory syndrome coro .................... .................... .................... .................... .................... ....................
30167 MT499175.1 |Severe acute respiratory syndrome coro .................... .................... .................... .................... .................... ....................
30168 MT499176.1 |Severe acute respiratory syndrome coro .................... .................... .................... .................... .................... ....................
30169 MT499177.1 |Severe acute respiratory syndrome coro .................... .................... .................... .................... .................... ....................
30170 MT499178.1 |Severe acute respiratory syndrome coro .................... .................... .................... .................... .................... ....................
30171 MT499179.1 |Severe acute respiratory syndrome coro .................... .................... .................... .................... .................... ....................
30172 MT499180.1 |Severe acute respiratory syndrome coro .................... .................... .................... .................... .................... ....................
30173 MT499181.1 |Severe acute respiratory syndrome coro .................... .................... .................... .................... .................... ....................
30174 MT499182.1 |Severe acute respiratory syndrome coro .................... .................... .................... .................... .................... ....................
30175 MT499183.1 |Severe acute respiratory syndrome coro .................... .................... .................... .................... .................... ....................
30176 MT499184.1 |Severe acute respiratory syndrome coro .................... .................... .................... .................... .................... ....................
30177 MT499185.1 |Severe acute respiratory syndrome coro .................... .................... .................... .................... .................... ....................
30178 MT499186.1 |Severe acute respiratory syndrome coro .................... .................... .................... .................... .................... ....................
30179 MT499187.1 |Severe acute respiratory syndrome coro .................... .................... .................... .................... .................... ....................
30180 MT499188.1 |Severe acute respiratory syndrome coro .................... .................... .................... .................... .................... ....................
30181 MT499189.1 |Severe acute respiratory syndrome coro .................... .................... .................... .................... .................... ....................
30182 MT499190.1 |Severe acute respiratory syndrome coro .................... .................... .................... .................... .................... ....................
30183 MT499191.1 |Severe acute respiratory syndrome coro .................... .................... .................... .................... .................... ....................
30184 MT499192.1 |Severe acute respiratory syndrome coro .................... .................... .................... .................... .................... ....................

| | | | | | | | | |
|---|---|---|---|---|---|---|---|---|
| 30185 | MT499193.1 | \|Severe acute respiratory syndrome coro | ···················· | ···················· | ····················· | ···················· | ···················· | ···················· |
| 30186 | MT499194.1 | \|Severe acute respiratory syndrome coro | ···················· | ···················· | ····················· | ···················· | ···················· | ···················· |
| 30187 | MT499195.1 | \|Severe acute respiratory syndrome coro | ···················· | ···················· | ····················· | ···················· | ···················· | ···················· |
| 30188 | MT499196.1 | \|Severe acute respiratory syndrome coro | ···················· | ···················· | ····················· | ···················· | ···················· | ···················· |
| 30189 | MT499197.1 | \|Severe acute respiratory syndrome coro | ···················· | ···················· | ····················· | ···················· | ···················· | ···················· |
| 30190 | MT499198.1 | \|Severe acute respiratory syndrome coro | ···················· | ···················· | ····················· | ···················· | ···················· | ···················· |
| 30191 | MT499199.1 | \|Severe acute respiratory syndrome coro | ···················· | ···················· | ····················· | ···················· | ···················· | ···················· |
| 30192 | MT499200.1 | \|Severe acute respiratory syndrome coro | ···················· | ···················· | ····················· | ···················· | ···················· | ···················· |
| 30193 | MT499201.1 | \|Severe acute respiratory syndrome coro | ···················· | ···················· | ····················· | ···················· | ···················· | ···················· |
| 30194 | MT499202.1 | \|Severe acute respiratory syndrome coro | ···················· | ···················· | ····················· | ···················· | ···················· | ···················· |
| 30195 | MT499203.1 | \|Severe acute respiratory syndrome coro | ···················· | ···················· | ····················· | ···················· | ···················· | ···················· |
| 30196 | MT499204.1 | \|Severe acute respiratory syndrome coro | ···················· | ···················· | ····················· | ···················· | ···················· | ···················· |
| 30197 | MT499205.1 | \|Severe acute respiratory syndrome coro | ···················· | ···················· | ····················· | ···················· | ···················· | ···················· |
| 30198 | MT499206.1 | \|Severe acute respiratory syndrome coro | ···················· | ···················· | ····················· | ···················· | ···················· | ···················· |
| 30199 | MT499207.1 | \|Severe acute respiratory syndrome coro | ···················· | ···················· | ····················· | ···················· | ···················· | ···················· |
| 30200 | MT499208.1 | \|Severe acute respiratory syndrome coro | ···················· | ···················· | ····················· | ···················· | ···················· | ···················· |
| 30201 | MT499209.1 | \|Severe acute respiratory syndrome coro | ···················· | ···················· | ····················· | ···················· | ···················· | ···················· |
| 30202 | MT499210.1 | \|Severe acute respiratory syndrome coro | ···················· | ···················· | ····················· | ···················· | ···················· | ···················· |
| 30203 | MT500122.1 | \|Severe acute respiratory syndrome coro | ···················· | ···················· | ····················· | ···················· | ···················· | ···················· |
| 30204 | MT502774.1 | \|Severe acute respiratory syndrome coro | ···················· | ···················· | ····················· | ···················· | ···················· | ···················· |
| 30205 | MT481895.1 | \|Severe acute respiratory syndrome coro | ···················· | ·A·················· | ····················· | ···················· | ···················· | ··A················· |
| 30206 | MT481896.1 | \|Severe acute respiratory syndrome coro | ···················· | ···················· | ····················· | ···················· | ···················· | ··A················· |
| 30207 | MT481897.1 | \|Severe acute respiratory syndrome coro | ···················· | ···················· | ····················· | ···················· | ···················· | ···················· |
| 30208 | MT481898.1 | \|Severe acute respiratory syndrome coro | ···················· | ·A·················· | ····················· | ···················· | ···················· | ··A················· |
| 30209 | MT481899.1 | \|Severe acute respiratory syndrome coro | ···················· | ···················· | ····················· | ···················· | ···················· | ···················· |
| 30210 | MT481900.1 | \|Severe acute respiratory syndrome coro | ···················· | ···················· | ····················· | ···················· | ···················· | ···················· |
| 30211 | MT481901.1 | \|Severe acute respiratory syndrome coro | ···················· | ·A·················· | ····················· | ···················· | ···················· | ··A················· |
| 30212 | MT481902.1 | \|Severe acute respiratory syndrome coro | ···················· | ···················· | ····················· | ···················· | ···················· | ···················· |
| 30213 | MT481903.1 | \|Severe acute respiratory syndrome coro | ···················· | ···················· | ····················· | ···················· | ···················· | ···················· |
| 30214 | MT481904.1 | \|Severe acute respiratory syndrome coro | ···················· | ···················· | ····················· | ···················· | ···················· | ···················· |
| 30215 | MT481905.1 | \|Severe acute respiratory syndrome coro | ···················· | ···················· | ····················· | ···················· | ···················· | ···················· |
| 30216 | MT481906.1 | \|Severe acute respiratory syndrome coro | ···················· | ·A·················· | ····················· | ···················· | ···················· | ··A················· |
| 30217 | MT481907.1 | \|Severe acute respiratory syndrome coro | ···················· | ···················· | ····················· | ···················· | ···················· | ···················· |
| 30218 | MT481908.1 | \|Severe acute respiratory syndrome coro | ···················· | ···················· | ····················· | ···················· | ···················· | ···················· |
| 30219 | MT481909.1 | \|Severe acute respiratory syndrome coro | ···················· | ···················· | ····················· | ···················· | ···················· | ···················· |
| 30220 | MT496972.1 | \|Severe acute respiratory syndrome coro | ···················· | ···················· | ····················· | ···················· | ···················· | ···················· |
| 30221 | MT496973.1 | \|Severe acute respiratory syndrome coro | ···················· | ···················· | ····················· | ···················· | ···················· | ···················· |
| 30222 | MT496974.1 | \|Severe acute respiratory syndrome coro | ···················· | ·A·················· | ····················· | ···················· | ···················· | ··A················· |
| 30223 | MT496975.1 | \|Severe acute respiratory syndrome coro | ···················· | ···················· | ····················· | ···················· | ···················· | ···················· |
| 30224 | MT496976.1 | \|Severe acute respiratory syndrome coro | ···················· | ··········A········· | ····················· | ···················· | ···················· | ···················· |
| 30225 | MT496977.1 | \|Severe acute respiratory syndrome coro | ···················· | ···················· | ····················· | ···················· | ···················· | ···················· |
| 30226 | MT496979.1 | \|Severe acute respiratory syndrome coro | ···················· | ···················· | ····················· | ···················· | ···················· | ···················· |
| 30227 | MT496980.1 | \|Severe acute respiratory syndrome coro | ···················· | ·A·················· | ····················· | ···················· | ···················· | ··A················· |
| 30228 | MT496981.1 | \|Severe acute respiratory syndrome coro | ···················· | ·A·················· | ····················· | ···················· | ···················· | ··A················· |
| 30229 | MT496982.1 | \|Severe acute respiratory syndrome coro | ···················· | ·A·················· | ····················· | ···················· | ···················· | ··A················· |
| 30230 | MT496983.1 | \|Severe acute respiratory syndrome coro | ···················· | ·A·················· | ····················· | ···················· | ···················· | ··A················· |
| 30231 | MT496984.1 | \|Severe acute respiratory syndrome coro | ···················· | ···················· | ····················· | ···················· | ···················· | ···················· |
| 30232 | MT496985.1 | \|Severe acute respiratory syndrome coro | ···················· | ·A·················· | ····················· | ···················· | ···················· | ··A················· |
| 30233 | MT496986.1 | \|Severe acute respiratory syndrome coro | ···················· | ·A·················· | ····················· | ···················· | ···················· | ··A················· |
| 30234 | MT496987.1 | \|Severe acute respiratory syndrome coro | ···················· | ·A·················· | ····················· | ···················· | ···················· | ··A················· |
| 30235 | MT496988.1 | \|Severe acute respiratory syndrome coro | ···················· | ···················· | ····················· | ···················· | ···················· | ···················· |
| 30236 | MT496989.1 | \|Severe acute respiratory syndrome coro | ···················· | ···················· | ····················· | ···················· | ···················· | ···················· |
| 30237 | MT496990.1 | \|Severe acute respiratory syndrome coro | ···················· | ···················· | ····················· | ···················· | ···················· | ···················· |
| 30238 | MT496991.1 | \|Severe acute respiratory syndrome coro | ···················· | ·A·················· | ····················· | ···················· | ···················· | ··A················· |
| 30239 | MT496992.1 | \|Severe acute respiratory syndrome coro | ···················· | ·A·················· | ····················· | ···················· | ···················· | ··A················· |
| 30240 | MT496993.1 | \|Severe acute respiratory syndrome coro | ···················· | ·A·················· | ····················· | ···················· | ···················· | ··A················· |
| 30241 | MT496994.1 | \|Severe acute respiratory syndrome coro | ···················· | ·A·················· | ····················· | ···················· | ···················· | ··A················· |
| 30242 | MT496995.1 | \|Severe acute respiratory syndrome coro | ···················· | ·A·················· | ····················· | ···················· | ···················· | ··A················· |
| 30243 | MT496996.1 | \|Severe acute respiratory syndrome coro | ···················· | ···················· | ····················· | ···················· | ···················· | ···················· |
| 30244 | MT496997.1 | \|Severe acute respiratory syndrome coro | ···················· | ···················· | ····················· | ···················· | ···················· | ···················· |
| 30245 | MT270101.1 | \|Severe acute respiratory syndrome coro | ···················· | ···················· | ····················· | ···················· | ···················· | ···················· |
| 30246 | MT270102.1 | \|Severe acute respiratory syndrome coro | ···················· | ···················· | ····················· | ···················· | ···················· | ···················· |
| 30247 | MT270103.1 | \|Severe acute respiratory syndrome coro | ···················· | ···················· | ····················· | ···················· | ···················· | ···················· |
| 30248 | MT270104.1 | \|Severe acute respiratory syndrome coro | ···················· | ···················· | ····················· | ···················· | ···················· | ···················· |
| 30249 | MT270105.1 | \|Severe acute respiratory syndrome coro | ···················· | ···················· | ····················· | ···················· | ···················· | ···················· |
| 30250 | MT270106.1 | \|Severe acute respiratory syndrome coro | ···················· | ···················· | ····················· | ···················· | ···················· | ···················· |
| 30251 | MT270107.1 | \|Severe acute respiratory syndrome coro | ···················· | ···················· | ····················· | ···················· | ···················· | ···················· |
| 30252 | MT270108.1 | \|Severe acute respiratory syndrome coro | ···················· | ···················· | ····················· | ···················· | ···················· | ···················· |
| 30253 | MT270109.1 | \|Severe acute respiratory syndrome coro | ···················· | ···················· | ····················· | ···················· | ···················· | ···················· |
| 30254 | MT270110.1 | \|Severe acute respiratory syndrome coro | ···················· | ···················· | ····················· | ···················· | ···················· | ···················· |
| 30255 | MT270111.1 | \|Severe acute respiratory syndrome coro | ···················· | ···················· | ····················· | ···················· | ···················· | ···················· |
| 30256 | MT270112.1 | \|Severe acute respiratory syndrome coro | ···················· | ···················· | ····················· | ···················· | ···················· | ···················· |
| 30257 | MT270113.1 | \|Severe acute respiratory syndrome coro | ···················· | ···················· | ····················· | ···················· | ···················· | ···················· |
| 30258 | MT270114.1 | \|Severe acute respiratory syndrome coro | ···················· | ···················· | ····················· | ···················· | ···················· | ···················· |
| 30259 | MT483553.1 | \|Severe acute respiratory syndrome coro | ···················· | ···················· | ····················· | ···················· | ···················· | ···················· |
| 30260 | MT483554.1 | \|Severe acute respiratory syndrome coro | ···················· | ···················· | ····················· | ···················· | ···················· | ···················· |
| 30261 | MT483555.1 | \|Severe acute respiratory syndrome coro | ···················· | ···················· | ····················· | ···················· | ···················· | ···················· |
| 30262 | MT483556.1 | \|Severe acute respiratory syndrome coro | ···················· | ···················· | ····················· | ···················· | ···················· | ···················· |

```
30263 MT483557.1 |Severe acute respiratory syndrome coro    ....................  ....................  ....................  ....................  ....................  ....................
30264 MT483558.1 |Severe acute respiratory syndrome coro    ....................  ....................  ....................  ....................  ....................  ....................
30265 MT483559.1 |Severe acute respiratory syndrome coro    ....................  ....................  ....................  ....................  ....................  ....................
30266 MT483560.1 |Severe acute respiratory syndrome coro    ....................  ....................  ....................  ....................  ....................  ....................
30267 MT483563.1 |Severe acute respiratory syndrome coro    ....................  ....................  ....................  ....................  ....................  ....................
30268 MT483702.1 |Severe acute respiratory syndrome coro    ....................  ....................  ....................  ....................  ....................  ....................
30269 MT477835.1 |Severe acute respiratory syndrome coro    ....................  ....................  ....................  ....................  ....................  ....................
30270 MT477836.1 |Severe acute respiratory syndrome coro    ....................  ....................  ....................  ....................  ....................  ....................
30271 MT477837.1 |Severe acute respiratory syndrome coro    ....................  ....................  ....................  ....................  ....................  ....................
30272 MT477838.1 |Severe acute respiratory syndrome coro    ....................  ....................  ....................  ....................  ....................  ....................
30273 MT477839.1 |Severe acute respiratory syndrome coro    ....................  ....................  ....................  ....................  ....................  ....................
30274 MT477840.1 |Severe acute respiratory syndrome coro    ....................  ....................  ....................  ....................  ....................  ....................
30275 MT477841.1 |Severe acute respiratory syndrome coro    ....................  ....................  ....................  ....................  ....................  ....................
30276 MT477842.1 |Severe acute respiratory syndrome coro    ....................  ....................  ....................  ....................  ....................  ....................
30277 MT477843.1 |Severe acute respiratory syndrome coro    ....................  ....................  ....................  ....................  ....................  ....................
30278 MT477844.1 |Severe acute respiratory syndrome coro    ....................  ....................  ....................  ....................  ....................  ....................
30279 MT477845.1 |Severe acute respiratory syndrome coro    ....................  ..........A.........  ....................  ....................  ....................  ....................
30280 MT477846.1 |Severe acute respiratory syndrome coro    ....................  ....................  ....................  ....................  ....................  ....................
30281 MT477847.1 |Severe acute respiratory syndrome coro    ....................  ....................  ....................  ....................  ....................  ....................
30282 MT477848.1 |Severe acute respiratory syndrome coro    ....................  ....................  ....................  ....................  ....................  ....................
30283 MT477849.1 |Severe acute respiratory syndrome coro    ....................  ....................  ....................  ....................  ....................  ....................
30284 MT477850.1 |Severe acute respiratory syndrome coro    ....................  ....................  ....................  ....................  ....................  ....................
30285 MT477851.1 |Severe acute respiratory syndrome coro    ....................  ....................  ....................  ....................  ....................  ....................
30286 MT477852.1 |Severe acute respiratory syndrome coro    ....................  ....................  ....................  ....................  ....................  ....................
30287 MT477853.1 |Severe acute respiratory syndrome coro    ....................  ....................  ....................  ....................  ....................  ....................
30288 MT477854.1 |Severe acute respiratory syndrome coro    ....................  ....................  ....................  ....................  ....................  ....................
30289 MT477855.1 |Severe acute respiratory syndrome coro    ....................  ....................  ....................  ....................  ....................  ....................
30290 MT477856.1 |Severe acute respiratory syndrome coro    ....................  ....................  ....................  ....................  ....................  ....................
30291 MT477857.1 |Severe acute respiratory syndrome coro    ....................  ....................  ....................  ....................  ....................  ....................
30292 MT477858.1 |Severe acute respiratory syndrome coro    ....................  ....................  ....................  ....................  ....................  ....................
30293 MT477859.1 |Severe acute respiratory syndrome coro    ....................  ....................  ....................  ....................  ....................  ....................
30294 MT477860.1 |Severe acute respiratory syndrome coro    ....................  ....................  ....................  ....................  ....................  ....................
30295 MT477861.1 |Severe acute respiratory syndrome coro    ....................  ....................  ....................  ....................  ....................  ....................
30296 MT477862.1 |Severe acute respiratory syndrome coro    ....................  ....................  ....................  ....................  ....................  ....................
30297 MT477885.1 |Severe acute respiratory syndrome coro    ....................  ....................  ....................  ....................  ....................  ....................
30298 MT477903.1 |Severe acute respiratory syndrome coro    ....................  ....................  ....................  ....................  ....................  ....................
30299 MT477904.1 |Severe acute respiratory syndrome coro    ....................  ....................  ....................  ....................  ....................  ....................
30300 MT481927.1 |Severe acute respiratory syndrome coro    ....................  ....................  ....................  ....................  ....................  ....................
30301 MT481928.1 |Severe acute respiratory syndrome coro    ....................  ....................  ....................  ....................  ....................  ....................
30302 MT481929.1 |Severe acute respiratory syndrome coro    ....................  ....................  ....................  ....................  ....................  ....................
30303 MT481930.1 |Severe acute respiratory syndrome coro    ....................  ....................  ....................  ....................  ....................  ....................
30304 MT481931.1 |Severe acute respiratory syndrome coro    ....................  ....................  ....................  ....................  ....................  ....................
30305 MT481932.1 |Severe acute respiratory syndrome coro    ....................  ....................  ....................  ....................  ....................  ....................
30306 MT481933.1 |Severe acute respiratory syndrome coro    ....................  ....................  ....................  ....................  ....................  ....................
30307 MT481934.1 |Severe acute respiratory syndrome coro    ....................  ....................  ....................  ....................  ....................  ....................
30308 MT481935.1 |Severe acute respiratory syndrome coro    ....................  ....................  ....................  ....................  ....................  ....................
30309 MT481936.1 |Severe acute respiratory syndrome coro    ....................  ....................  ....................  ....................  ....................  ....................
30310 MT481992.1 |Severe acute respiratory syndrome coro    ....................  ....................  ....................  ....................  ....................  ....................
30311 MT482110.1 |Severe acute respiratory syndrome coro    ....................  ....................  ....................  ....................  ....................  ....................
30312 MT482111.1 |Severe acute respiratory syndrome coro    ....................  ....................  ....................  ....................  ....................  ....................
30313 MT482113.1 |Severe acute respiratory syndrome coro    ....................  ....................  ....................  ....................  ....................  ....................
30314 MT482114.1 |Severe acute respiratory syndrome coro    ....................  ....................  ....................  ....................  ....................  ....................
30315 MT482115.1 |Severe acute respiratory syndrome coro    ....................  ....................  ....................  ....................  ....................  ....................
30316 MT482116.1 |Severe acute respiratory syndrome coro    ....................  ....................  ....................  ....................  ....................  ....................
30317 MT482117.1 |Severe acute respiratory syndrome coro    ....................  ....................  ....................  ....................  ....................  ....................
30318 MT482118.1 |Severe acute respiratory syndrome coro    ....................  ....................  ....................  ....................  ....................  ....................
30319 MT482119.1 |Severe acute respiratory syndrome coro    ....................  ....................  ....................  ....................  ....................  ....................
30320 MT482121.1 |Severe acute respiratory syndrome coro    ....................  ....................  ....................  ....................  ....................  ....................
30321 MT482122.1 |Severe acute respiratory syndrome coro    ....................  ....................  ....................  ....................  ....................  ....................
30322 MT482123.1 |Severe acute respiratory syndrome coro    ....................  ....................  ....................  ....................  ....................  ....................
30323 MT482124.1 |Severe acute respiratory syndrome coro    ....................  ....................  ....................  ....................  ....................  ....................
30324 MT482125.1 |Severe acute respiratory syndrome coro    ....................  ....................  ....................  ....................  ....................  ....................
30325 MT482126.1 |Severe acute respiratory syndrome coro    ....................  ....................  ....................  ....................  ....................  ....................
30326 MT482127.1 |Severe acute respiratory syndrome coro    ....................  ....................  ....................  ....................  ....................  ....................
30327 MT482128.1 |Severe acute respiratory syndrome coro    ....................  ....................  ....................  ....................  ....................  ....................
30328 MT482129.1 |Severe acute respiratory syndrome coro    ....................  ....................  ....................  ....................  ....................  ....................
30329 MT482130.1 |Severe acute respiratory syndrome coro    ....................  ....................  ....................  ....................  ....................  ....................
30330 MT482131.1 |Severe acute respiratory syndrome coro    ....................  ....................  ....................  ....................  ....................  ....................
30331 MT482132.1 |Severe acute respiratory syndrome coro    ....................  ....................  ....................  ....................  ....................  ....................
30332 MT482133.1 |Severe acute respiratory syndrome coro    ....................  ....................  ....................  ....................  ....................  ....................
30333 MT482134.1 |Severe acute respiratory syndrome coro    ....................  ....................  ....................  ....................  ....................  ....................
30334 MT482135.1 |Severe acute respiratory syndrome coro    ....................  ....................  ....................  ....................  ....................  ....................
30335 MT482136.1 |Severe acute respiratory syndrome coro    ....................  ....................  ....................  ....................  ....................  ....................
30336 MT482137.1 |Severe acute respiratory syndrome coro    ....................  ....................  ....................  ....................  ....................  ....................
30337 MT482138.1 |Severe acute respiratory syndrome coro    ....................  ....................  ....................  ....................  ....................  ....................
30338 MT482139.1 |Severe acute respiratory syndrome coro    ....................  ....................  ....................  ....................  ....................  ....................
30339 MT482140.1 |Severe acute respiratory syndrome coro    ....................  ....................  ....................  ....................  ....................  ....................
30340 MT482141.1 |Severe acute respiratory syndrome coro    ....................  ....................  ....................  ....................  ....................  ....................
```

30341 MT482142.1 |Severe acute respiratory syndrome coro ···················· ···················· ···················· ···················· ···················· ····················
30342 MT482143.1 |Severe acute respiratory syndrome coro ···················· ···················· ···················· ···················· ···················· ····················
30343 MT482144.1 |Severe acute respiratory syndrome coro ···················· ···················· ···················· ···················· ···················· ····················
30344 MT482145.1 |Severe acute respiratory syndrome coro ···················· ···················· ···················· ···················· ···················· ····················
30345 MT482146.1 |Severe acute respiratory syndrome coro ···················· ···················· ···················· ···················· ···················· ····················
30346 MT482147.1 |Severe acute respiratory syndrome coro ···················· ···················· ···················· ···················· ···················· ····················
30347 MT482148.1 |Severe acute respiratory syndrome coro ···················· ···················· ···················· ···················· ···················· ····················
30348 MT482149.1 |Severe acute respiratory syndrome coro ···················· ···················· ···················· ···················· ···················· ····················
30349 LC547518.1 |Severe acute respiratory syndrome coro ···················· ···················· ···················· ···················· ···················· ····················
30350 LC547519.1 |Severe acute respiratory syndrome coro ···················· ···················· ···················· ···················· ···················· ····················
30351 LC547520.1 |Severe acute respiratory syndrome coro ···················· ···················· ···················· ···················· ···················· ····················
30352 LC547521.1 |Severe acute respiratory syndrome coro ···················· ···················· ···················· ···················· ···················· ····················
30353 LC547522.1 |Severe acute respiratory syndrome coro ···················· ···················· ···················· ···················· ···················· ····················
30354 LC547523.1 |Severe acute respiratory syndrome coro ···················· ···················· ···················· ···················· ···················· ····················
30355 LC547524.1 |Severe acute respiratory syndrome coro ···················· ···················· ···················· ···················· ···················· ····················
30356 LC547525.1 |Severe acute respiratory syndrome coro ···················· ···················· ···················· ···················· ···················· ····················
30357 LC547526.1 |Severe acute respiratory syndrome coro ···················· ···················· ···················· ···················· ···················· ····················
30358 LC547527.1 |Severe acute respiratory syndrome coro ···················· ···················· ···················· ···················· ···················· ····················
30359 LC547528.1 |Severe acute respiratory syndrome coro ···················· ···················· ···················· ···················· ···················· ····················
30360 LC547529.1 |Severe acute respiratory syndrome coro ···················· ···················· ···················· ···················· ···················· ····················
30361 LC547530.1 |Severe acute respiratory syndrome coro ···················· ···················· ···················· ···················· ···················· ····················
30362 LC547531.1 |Severe acute respiratory syndrome coro ···················· ···················· ···················· ···················· ···················· ····················
30363 LC547532.1 |Severe acute respiratory syndrome coro ···················· ···················· ···················· ···················· ···················· ····················
30364 LC547533.1 |Severe acute respiratory syndrome coro ···················· ···················· ···················· ···················· ···················· ····················
30365 LC546038.1 |Severe acute respiratory syndrome coro ···················· ···················· ···················· ···················· ···················· ····················
30366 MT079843.1 |Severe acute respiratory syndrome coro ···················· ···················· ···················· ···················· ···················· ····················
30367 MT079844.1 |Severe acute respiratory syndrome coro ···················· ···················· ···················· ···················· ···················· ····················
30368 MT079845.1 |Severe acute respiratory syndrome coro ···················· ···················· ···················· ···················· ···················· ····················
30369 MT079846.1 |Severe acute respiratory syndrome coro ···················· ···················· ···················· ···················· ···················· ····················
30370 MT079847.1 |Severe acute respiratory syndrome coro ···················· ···················· ···················· ···················· ···················· ····················
30371 MT079848.1 |Severe acute respiratory syndrome coro ···················· ···················· ···················· ···················· ···················· ····················
30372 MT079849.1 |Severe acute respiratory syndrome coro ···················· ···················· ···················· ···················· ···················· ····················
30373 MT079850.1 |Severe acute respiratory syndrome coro ···················· ···················· ···················· ···················· ···················· ····················
30374 MT079851.1 |Severe acute respiratory syndrome coro ···················· ···················· ···················· ···················· ···················· ····················
30375 MT079852.1 |Severe acute respiratory syndrome coro ···················· ···················· ···················· ···················· ···················· ····················
30376 MT079853.1 |Severe acute respiratory syndrome coro ···················· ···················· ···················· ···················· ···················· ····················
30377 MT079854.1 |Severe acute respiratory syndrome coro ···················· ···················· ···················· ···················· ···················· ····················
30378 MT472621.1 |Severe acute respiratory syndrome coro ···················· ···················· ···················· ···················· ···················· ····················
30379 MT472622.1 |Severe acute respiratory syndrome coro ···················· ···················· ···················· ···················· ···················· ····················
30380 MT472623.1 |Severe acute respiratory syndrome coro ···················· ···················· ···················· ···················· ···················· ····················
30381 MT472624.1 |Severe acute respiratory syndrome coro ···················· ···················· ···················· ···················· ···················· ····················
30382 MT472625.1 |Severe acute respiratory syndrome coro ···················· ···················· ···················· ···················· ···················· ····················
30383 MT472626.1 |Severe acute respiratory syndrome coro ···················· ···················· ···················· ···················· ···················· ····················
30384 MT472627.1 |Severe acute respiratory syndrome coro ···················· ···················· ···················· ···················· ···················· ····················
30385 MT474126.1 |Severe acute respiratory syndrome coro ···················· ···················· ···················· ···················· ···················· ····················
30386 MT474127.1 |Severe acute respiratory syndrome coro ···················· ···················· ···················· ···················· ···················· ····················
30387 MT474128.1 |Severe acute respiratory syndrome coro ···················· ···················· ···················· ···················· ···················· ····················
30388 MT474129.1 |Severe acute respiratory syndrome coro ···················· ···················· ···················· ···················· ···················· ····················
30389 MT474130.1 |Severe acute respiratory syndrome coro ···················· ···················· ···················· ···················· ···················· ····················
30390 MT474132.1 |Severe acute respiratory syndrome coro ···················· ···················· ···················· ···················· ···················· ····················
30391 MT474133.1 |Severe acute respiratory syndrome coro ···················· ···················· ···················· ···················· ···················· ····················
30392 MT474134.1 |Severe acute respiratory syndrome coro ···················· ···················· ···················· ···················· ···················· ····················
30393 MT474135.1 |Severe acute respiratory syndrome coro ···················· ···················· ···················· ···················· ···················· ····················
30394 MT474136.1 |Severe acute respiratory syndrome coro ···················· ···················· ···················· ···················· ···················· ····················
30395 MT474137.1 |Severe acute respiratory syndrome coro ···················· ···················· ···················· ···················· ···················· ····················
30396 MT474138.1 |Severe acute respiratory syndrome coro ···················· ···················· ···················· ···················· ···················· ····················
30397 MT476384.1 |Severe acute respiratory syndrome coro ···················· ···················· ···················· ···················· ···················· ····················
30398 MT476385.1 |Severe acute respiratory syndrome coro ···················· ···················· ···················· ···················· ···················· ····················
30399 MT358638.1 |Severe acute respiratory syndrome coro ···················· ·A·················· ···················· ···················· ···················· ····················
30400 MT358639.1 |Severe acute respiratory syndrome coro ···················· ···················· ···················· ···················· ···················· ····················
30401 MT358640.1 |Severe acute respiratory syndrome coro ···················· ···················· ···················· ···················· ···················· ····················
30402 MT358641.1 |Severe acute respiratory syndrome coro ···················· ···················· ···················· ···················· ···················· ····················
30403 MT358642.1 |Severe acute respiratory syndrome coro ···················· ···················· ···················· ···················· ···················· ····················
30404 MT358643.1 |Severe acute respiratory syndrome coro ···················· ···················· ···················· ···················· ···················· ····················
30405 MT466071.1 |Severe acute respiratory syndrome coro ···················· ···················· ···················· ···················· ···················· ····················
30406 MT466583.1 |Severe acute respiratory syndrome coro ···················· ···················· ···················· ···················· ···················· ····················
30407 MT467237.1 |Severe acute respiratory syndrome coro ···················· ···················· ···················· ···················· ···················· ····················
30408 MT467238.1 |Severe acute respiratory syndrome coro ···················· ···················· ···················· ···················· ···················· ····················
30409 MT467239.1 |Severe acute respiratory syndrome coro ···················· ···················· ···················· ···················· ···················· ····················
30410 MT467240.1 |Severe acute respiratory syndrome coro ···················· ···················· ···················· ···················· ···················· ····················
30411 MT467241.1 |Severe acute respiratory syndrome coro ···················· ···················· ···················· ···················· ···················· ····················
30412 MT467242.1 |Severe acute respiratory syndrome coro ···················· ···················· ···················· ···················· ···················· ····················
30413 MT467243.1 |Severe acute respiratory syndrome coro ···················· ·A·················· ···················· ···················· ···················· ··A·················
30414 MT467244.1 |Severe acute respiratory syndrome coro ···················· ···················· ···················· ···················· ···················· ····················
30415 MT467245.1 |Severe acute respiratory syndrome coro ···················· ·A·················· ···················· ···················· ···················· ··A·················
30416 MT467246.1 |Severe acute respiratory syndrome coro ···················· ···················· ···················· ···················· ···················· ····················
30417 MT467247.1 |Severe acute respiratory syndrome coro ···················· ···················· ···················· ···················· ···················· ····················
30418 MT467248.1 |Severe acute respiratory syndrome coro ···················· ·A·················· ···················· ···················· ···················· ··A·················

30419 MT467249.1 |Severe acute respiratory syndrome coro .................... .................... .................... .................... .................... ....................
30420 MT467250.1 |Severe acute respiratory syndrome coro .................... .................... .................... .................... .................... ....................
30421 MT467251.1 |Severe acute respiratory syndrome coro .................... .................... .................... .................... .................... ....................
30422 MT467252.1 |Severe acute respiratory syndrome coro .................... .................... .................... .................... .................... ....................
30423 MT467253.1 |Severe acute respiratory syndrome coro .................... .A.................. .................... .................... .................... ...A................
30424 MT467254.1 |Severe acute respiratory syndrome coro ...T................ .A.................. .................... .................... .................... ...A................
30425 MT467255.1 |Severe acute respiratory syndrome coro .................... .................... .................... .................... .................... ....................
30426 MT467256.1 |Severe acute respiratory syndrome coro .................... .A.................. .................... .................... .................... ...A................
30427 MT467257.1 |Severe acute respiratory syndrome coro .................... .................... .................... .................... .................... ....................
30428 MT467258.1 |Severe acute respiratory syndrome coro .................... .A.................. .................... .................... .................... ...A................
30429 MT467259.1 |Severe acute respiratory syndrome coro .................... .................... .................... .................... .................... ....................
30430 MT467260.1 |Severe acute respiratory syndrome coro .................... .................... .................... .................... .................... ....................
30431 MT467261.1 |Severe acute respiratory syndrome coro .................... .................... .................... .................... .................... ....................
30432 MT467262.1 |Severe acute respiratory syndrome coro .................... .................... .................... .................... .................... ....................
30433 MT467263.1 |Severe acute respiratory syndrome coro .................... .A.................. .................... .................... .................... ...A................
30434 MT470100.1 |Severe acute respiratory syndrome coro .................... .................... .................... .................... .................... ....................
30435 MT470101.1 |Severe acute respiratory syndrome coro .................... .................... .................... .................... .................... ....................
30436 MT470102.1 |Severe acute respiratory syndrome coro .................... .................... .................... .................... .................... ....................
30437 MT470103.1 |Severe acute respiratory syndrome coro .................... .................... .................... .................... .................... ....................
30438 MT470104.1 |Severe acute respiratory syndrome coro .................... .................... .................... .................... .................... ....................
30439 MT470105.1 |Severe acute respiratory syndrome coro .................... .................... .................... .................... .................... ....................
30440 MT470106.1 |Severe acute respiratory syndrome coro .................... .................... .................... .................... .................... ....................
30441 MT470107.1 |Severe acute respiratory syndrome coro .................... .................... .................... .................... .................... ....................
30442 MT470108.1 |Severe acute respiratory syndrome coro .................... .................... .................... .................... .................... ....................
30443 MT470109.1 |Severe acute respiratory syndrome coro .................... .................... .................... .................... .................... ....................
30444 MT470110.1 |Severe acute respiratory syndrome coro .................... .................... .................... .................... .................... ....................
30445 MT470111.1 |Severe acute respiratory syndrome coro .................... .................... .................... .................... .................... ....................
30446 MT470112.1 |Severe acute respiratory syndrome coro .................... .................... .................... .................... .................... ....................
30447 MT470113.1 |Severe acute respiratory syndrome coro .................... .................... .................... .................... .................... ....................
30448 MT470114.1 |Severe acute respiratory syndrome coro .................... .................... .................... .................... .................... ....................
30449 MT470115.1 |Severe acute respiratory syndrome coro .................... .................... .................... .................... .................... ....................
30450 MT470116.1 |Severe acute respiratory syndrome coro .................... .................... .................... .................... .................... ....................
30451 MT470117.1 |Severe acute respiratory syndrome coro .................... .................... .................... .................... .................... ....................
30452 MT470118.1 |Severe acute respiratory syndrome coro .................... .................... .................... .................... .................... ....................
30453 MT470119.1 |Severe acute respiratory syndrome coro .................... .................... .................... .................... .................... ....................
30454 MT470120.1 |Severe acute respiratory syndrome coro .................... .................... .................... .................... .................... ....................
30455 MT470121.1 |Severe acute respiratory syndrome coro .................... .................... .................... .................... .................... ....................
30456 MT470122.1 |Severe acute respiratory syndrome coro .................... .................... .................... .................... .................... ....................
30457 MT470123.1 |Severe acute respiratory syndrome coro .................... .................... .................... .................... .................... ....................
30458 MT470124.1 |Severe acute respiratory syndrome coro .................... .................... .................... .................... .................... ....................
30459 MT470125.1 |Severe acute respiratory syndrome coro .................... .................... .................... .................... .................... ....................
30460 MT470126.1 |Severe acute respiratory syndrome coro .................... .................... .................... .................... .................... ....................
30461 MT470127.1 |Severe acute respiratory syndrome coro .................... .................... .................... .................... .................... ....................
30462 MT470128.1 |Severe acute respiratory syndrome coro .................... .................... .................... .................... .................... ....................
30463 MT470129.1 |Severe acute respiratory syndrome coro .................... .................... .................... .................... .................... ....................
30464 MT470130.1 |Severe acute respiratory syndrome coro .................... .................... .................... .................... .................... ....................
30465 MT470131.1 |Severe acute respiratory syndrome coro .................... .................... .................... .................... .................... ....................
30466 MT470132.1 |Severe acute respiratory syndrome coro .................... .................... .................... .................... .................... ....................
30467 MT470133.1 |Severe acute respiratory syndrome coro .................... .................... .................... .................... .................... ....................
30468 MT470134.1 |Severe acute respiratory syndrome coro .................... .................... .................... .................... .................... ....................
30469 MT470135.1 |Severe acute respiratory syndrome coro .................... .................... .................... .................... .................... ....................
30470 MT470136.1 |Severe acute respiratory syndrome coro .................... .................... .................... .................... .................... ....................
30471 MT470137.1 |Severe acute respiratory syndrome coro .................... .................... .................... .................... .................... ....................
30472 MT470138.1 |Severe acute respiratory syndrome coro .................... .................... .................... .................... .................... ....................
30473 MT470139.1 |Severe acute respiratory syndrome coro .................... .................... .................... .................... .................... ....................
30474 MT470140.1 |Severe acute respiratory syndrome coro .................... .................... .................... .................... .................... ....................
30475 MT470141.1 |Severe acute respiratory syndrome coro .................... .................... .................... .................... .................... ....................
30476 MT470142.1 |Severe acute respiratory syndrome coro .................... .................... .................... .................... .................... ....................
30477 MT470143.1 |Severe acute respiratory syndrome coro .................... .................... .................... .................... .................... ....................
30478 MT470144.1 |Severe acute respiratory syndrome coro .................... .................... .................... .................... .................... ....................
30479 MT470145.1 |Severe acute respiratory syndrome coro .................... .................... .................... .................... .................... ....................
30480 MT470146.1 |Severe acute respiratory syndrome coro .................... .................... .................... .................... .................... ....................
30481 MT470147.1 |Severe acute respiratory syndrome coro .................... .................... .................... .................... .................... ....................
30482 MT470148.1 |Severe acute respiratory syndrome coro .................... .................... .................... .................... .................... ....................
30483 MT470149.1 |Severe acute respiratory syndrome coro .................... .................... .................... .................... .................... ....................
30484 MT470150.1 |Severe acute respiratory syndrome coro .................... .................... .................... .................... .................... ....................
30485 MT470151.1 |Severe acute respiratory syndrome coro .................... .................... .................... .................... .................... ....................
30486 MT470152.1 |Severe acute respiratory syndrome coro .................... .................... .................... .................... .................... ....................
30487 MT470153.1 |Severe acute respiratory syndrome coro .................... .................... .................... .................... .................... ....................
30488 MT470154.1 |Severe acute respiratory syndrome coro .................... .................... .................... .................... .................... ....................
30489 MT470155.1 |Severe acute respiratory syndrome coro .................... .................... .................... .................... .................... ....................
30490 MT470156.1 |Severe acute respiratory syndrome coro .................... .................... .................... .................... .................... ....................
30491 MT470157.1 |Severe acute respiratory syndrome coro .................... .................... .................... .................... .................... ....................
30492 MT470158.1 |Severe acute respiratory syndrome coro .................... .................... .................... .................... .................... ....................
30493 MT470159.1 |Severe acute respiratory syndrome coro .................... .................... .................... .................... .................... ....................
30494 MT470160.1 |Severe acute respiratory syndrome coro .................... .................... .................... .................... .................... ....................
30495 MT470161.1 |Severe acute respiratory syndrome coro .................... .................... .................... .................... .................... ....................
30496 MT470162.1 |Severe acute respiratory syndrome coro .................... .................... .................... .................... .................... ....................

30497 MT470163.1 |Severe acute respiratory syndrome coro ···················· ···················· ····················· ···················· ···················· ····················
30498 MT470164.1 |Severe acute respiratory syndrome coro ···················· ···················· ····················· ···················· ···················· ····················
30499 MT470165.1 |Severe acute respiratory syndrome coro ···················· ···················· ····················· ···················· ···················· ····················
30500 MT470166.1 |Severe acute respiratory syndrome coro ···················· ···················· ····················· ···················· ···················· ····················
30501 MT470167.1 |Severe acute respiratory syndrome coro ···················· ···················· ····················· ···················· ···················· ····················
30502 MT470168.1 |Severe acute respiratory syndrome coro ···················· ···················· ····················· ···················· ···················· ····················
30503 MT470169.1 |Severe acute respiratory syndrome coro ···················· ···················· ····················· ···················· ···················· ····················
30504 MT470170.1 |Severe acute respiratory syndrome coro ···················· ···················· ····················· ···················· ···················· ····················
30505 MT470171.1 |Severe acute respiratory syndrome coro ···················· ···················· ····················· ···················· ···················· ····················
30506 MT470172.1 |Severe acute respiratory syndrome coro ···················· ···················· ····················· ···················· ···················· ····················
30507 MT470173.1 |Severe acute respiratory syndrome coro ···················· ···················· ····················· ···················· ···················· ····················
30508 MT470174.1 |Severe acute respiratory syndrome coro ···················· ···················· ····················· ···················· ···················· ····················
30509 MT470175.1 |Severe acute respiratory syndrome coro ···················· ···················· ····················· ···················· ···················· ····················
30510 MT470176.1 |Severe acute respiratory syndrome coro ···················· ···················· ····················· ···················· ···················· ····················
30511 MT470177.1 |Severe acute respiratory syndrome coro ···················· ···················· ····················· ···················· ···················· ····················
30512 MT470178.1 |Severe acute respiratory syndrome coro ···················· ···················· ····················· ···················· ···················· ····················
30513 MT470179.1 |Severe acute respiratory syndrome coro ···················· ···················· ····················· ···················· ···················· ····················
30514 MT470219.1 |Severe acute respiratory syndrome coro ···················· ···················· ····················· ···················· ···················· ····················
30515 MT459832.1 |Severe acute respiratory syndrome coro ···················· ···················· ····················· ···················· ···················· ····················
30516 MT459833.1 |Severe acute respiratory syndrome coro ···················· ···················· ····················· ······T············· ···················· ····················
30517 MT459834.1 |Severe acute respiratory syndrome coro ···················· ···················· ····················· ···················· ···················· ····················
30518 MT459835.1 |Severe acute respiratory syndrome coro ···················· ···················· ····················· ···················· ···················· ····················
30519 MT459836.1 |Severe acute respiratory syndrome coro ···················· ···················· ····················· ···················· ···················· ····················
30520 MT459837.1 |Severe acute respiratory syndrome coro ···················· ···················· ····················· ···················· ···················· ····················
30521 MT459838.1 |Severe acute respiratory syndrome coro ···················· ···················· ····················· ···················· ···················· ····················
30522 MT459839.1 |Severe acute respiratory syndrome coro ···················· ···················· ····················· ···················· ···················· ····················
30523 MT459840.1 |Severe acute respiratory syndrome coro ···················· ···················· ····················· ···················· ···················· ····················
30524 MT459841.1 |Severe acute respiratory syndrome coro ···················· ···················· ····················· ···················· ···················· ····················
30525 MT459842.1 |Severe acute respiratory syndrome coro ···················· ···················· ····················· ···················· ···················· ····················
30526 MT459843.1 |Severe acute respiratory syndrome coro ···················· ···················· ····················· ···················· ···················· ····················
30527 MT459844.1 |Severe acute respiratory syndrome coro ···················· ···················· ····················· ···················· ···················· ····················
30528 MT459845.1 |Severe acute respiratory syndrome coro ···················· ···················· ····················· ···················· ···················· ····················
30529 MT459846.1 |Severe acute respiratory syndrome coro ···················· ···················· ····················· ···················· ···················· ····················
30530 MT459847.1 |Severe acute respiratory syndrome coro ···················· ···················· ····················· ···················· ···················· ····················
30531 MT459848.1 |Severe acute respiratory syndrome coro ···················· ···················· ····················· ···················· ···················· ····················
30532 MT459849.1 |Severe acute respiratory syndrome coro ···················· ···················· ····················· ···················· ···················· ····················
30533 MT459850.1 |Severe acute respiratory syndrome coro ···················· ·············A······ ····················· ···················· ···················· ····················
30534 MT459851.1 |Severe acute respiratory syndrome coro ···················· ···················· ····················· ···················· ···················· ····················
30535 MT459852.1 |Severe acute respiratory syndrome coro ···················· ···················· ····················· ···················· ···················· ····················
30536 MT459853.1 |Severe acute respiratory syndrome coro ···················· ···················· ····················· ···················· ···················· ····················
30537 MT459854.1 |Severe acute respiratory syndrome coro ···················· ···················· ····················· ···················· ···················· ····················
30538 MT459855.1 |Severe acute respiratory syndrome coro ···················· ···················· ····················· ···················· ···················· ····················
30539 MT459856.1 |Severe acute respiratory syndrome coro ···················· ···················· ····················· ···················· ···················· ····················
30540 MT459857.1 |Severe acute respiratory syndrome coro ···················· ···················· ····················· ···················· ···················· ····················
30541 MT459858.1 |Severe acute respiratory syndrome coro ···················· ···················· ····················· ···················· ···················· ····················
30542 MT459859.1 |Severe acute respiratory syndrome coro ···················· ···················· ····················· ···················· ···················· ····················
30543 MT459860.1 |Severe acute respiratory syndrome coro ···················· ···················· ····················· ···················· ···················· ····················
30544 MT459861.1 |Severe acute respiratory syndrome coro ···················· ···················· ····················· ···················· ···················· ····················
30545 MT459862.1 |Severe acute respiratory syndrome coro ···················· ···················· ····················· ···················· ···················· ····················
30546 MT459863.1 |Severe acute respiratory syndrome coro ···················· ···················· ····················· ···················· ···················· ····················
30547 MT459864.1 |Severe acute respiratory syndrome coro ···················· ···················· ····················· ···················· ···················· ····················
30548 MT459865.1 |Severe acute respiratory syndrome coro ···················· ···················· ····················· ···················· ···················· ····················
30549 MT459866.1 |Severe acute respiratory syndrome coro ···················· ···················· ····················· ···················· ···················· ····················
30550 MT459867.1 |Severe acute respiratory syndrome coro ···················· ···················· ····················· ···················· ···················· ····················
30551 MT459868.1 |Severe acute respiratory syndrome coro ···················· ···················· ····················· ···················· ···················· ····················
30552 MT459869.1 |Severe acute respiratory syndrome coro ···················· ···················· ····················· ···················· ···················· ····················
30553 MT459870.1 |Severe acute respiratory syndrome coro ···················· ···················· ····················· ···················· ···················· ····················
30554 MT459871.1 |Severe acute respiratory syndrome coro ···················· ···················· ····················· ···················· ···················· ····················
30555 MT459872.1 |Severe acute respiratory syndrome coro ···················· ···················· ····················· ···················· ···················· ····················
30556 MT459873.1 |Severe acute respiratory syndrome coro ···················· ···················· ····················· ···················· ···················· ····················
30557 MT459874.1 |Severe acute respiratory syndrome coro ···················· ···················· ····················· ···················· ···················· ····················
30558 MT459875.1 |Severe acute respiratory syndrome coro ···················· ···················· ····················· ···················· ···················· ····················
30559 MT459876.1 |Severe acute respiratory syndrome coro ···················· ···················· ····················· ···················· ···················· ····················
30560 MT459877.1 |Severe acute respiratory syndrome coro ···················· ···················· ····················· ···················· ···················· ····················
30561 MT459878.1 |Severe acute respiratory syndrome coro ···················· ···················· ····················· ···················· ···················· ····················
30562 MT459879.1 |Severe acute respiratory syndrome coro ···················· ···················· ····················· ···················· ···················· ····················
30563 MT459880.1 |Severe acute respiratory syndrome coro ···················· ···················· ····················· ···················· ···················· ····················
30564 MT459881.1 |Severe acute respiratory syndrome coro ···················· ···················· ····················· ···················· ···················· ····················
30565 MT459882.1 |Severe acute respiratory syndrome coro ···················· ···················· ····················· ···················· ···················· ····················
30566 MT459883.1 |Severe acute respiratory syndrome coro ···················· ···················· ····················· ···················· ···················· ····················
30567 MT459884.1 |Severe acute respiratory syndrome coro ···················· ···················· ····················· ···················· ···················· ····················
30568 MT459885.1 |Severe acute respiratory syndrome coro ···················· ···················· ····················· ···················· ···················· ····················
30569 MT459886.1 |Severe acute respiratory syndrome coro ···················· ···················· ····················· ···················· ···················· ····················
30570 MT459887.1 |Severe acute respiratory syndrome coro ···················· ···················· ····················· ···················· ···················· ····················
30571 MT459888.1 |Severe acute respiratory syndrome coro ···················· ···················· ····················· ···················· ···················· ····················
30572 MT459889.1 |Severe acute respiratory syndrome coro ···················· ···················· ····················· ···················· ···················· ····················
30573 MT459890.1 |Severe acute respiratory syndrome coro ···················· ···················· ····················· ···················· ···················· ····················
30574 MT459891.1 |Severe acute respiratory syndrome coro ···················· ···················· ····················· ···················· ···················· ····················

```
30575 MT459892.1 |Severe acute respiratory syndrome coro    .................... .................... ..................... .................... .................... ....................
30576 MT459893.1 |Severe acute respiratory syndrome coro    .................... .................... ..................... .................... .................... ....................
30577 MT459894.1 |Severe acute respiratory syndrome coro    .................... .................... ..................... .................... .................... ....................
30578 MT459895.1 |Severe acute respiratory syndrome coro    .................... .................... ..................... .................... .................... ....................
30579 MT459896.1 |Severe acute respiratory syndrome coro    .................... .................... ..................... .................... .................... ....................
30580 MT459897.1 |Severe acute respiratory syndrome coro    .................... .................... ..................... .................... .................... ....................
30581 MT459898.1 |Severe acute respiratory syndrome coro    .................... .................... ..................... .................... .................... ....................
30582 MT459899.1 |Severe acute respiratory syndrome coro    .................... .................... ..................... .................... .................... ....................
30583 MT459900.1 |Severe acute respiratory syndrome coro    .................... .................... ..................... .................... .................... ....................
30584 MT459901.1 |Severe acute respiratory syndrome coro    .................... .................... ..................... .................... .................... ....................
30585 MT459902.1 |Severe acute respiratory syndrome coro    .................... .................... ..................... .................... .................... ....................
30586 MT459903.1 |Severe acute respiratory syndrome coro    .................... .................... ..................... .................... .................... ....................
30587 MT459904.1 |Severe acute respiratory syndrome coro    .................... .................... ..................... .................... .................... ....................
30588 MT459905.1 |Severe acute respiratory syndrome coro    .................... .................... ..................... .................... .................... ....................
30589 MT459906.1 |Severe acute respiratory syndrome coro    .................... .................... ..................... .................... .................... ....................
30590 MT459907.1 |Severe acute respiratory syndrome coro    .................... .................... ..................... .................... .................... ....................
30591 MT459908.1 |Severe acute respiratory syndrome coro    .................... .................... ..................... .................... .................... ....................
30592 MT459909.1 |Severe acute respiratory syndrome coro    .................... .................... ..................... .................... .................... ....................
30593 MT459910.1 |Severe acute respiratory syndrome coro    .................... .................... ..................... .................... .................... ....................
30594 MT459911.1 |Severe acute respiratory syndrome coro    .................... .................... ..................... .................... .................... ....................
30595 MT459912.1 |Severe acute respiratory syndrome coro    .................... .................... ..................... .................... .................... ....................
30596 MT459913.1 |Severe acute respiratory syndrome coro    .................... .................... ..................... .................... .................... ....................
30597 MT459914.1 |Severe acute respiratory syndrome coro    .................... .................... ..................... .................... .................... ....................
30598 MT459915.1 |Severe acute respiratory syndrome coro    .................... .................... ..................... .................... .................... ....................
30599 MT459916.1 |Severe acute respiratory syndrome coro    .................... .................... ..................... .................... .................... ....................
30600 MT459917.1 |Severe acute respiratory syndrome coro    .................... .................... ..................... .................... .................... ....................
30601 MT459919.1 |Severe acute respiratory syndrome coro    .................... .................... ..................... .................... .................... ....................
30602 MT459920.1 |Severe acute respiratory syndrome coro    .................... .................... ..................... .................... .................... ....................
30603 MT459921.1 |Severe acute respiratory syndrome coro    .................... .................... ..................... .................... .................... ....................
30604 MT459922.1 |Severe acute respiratory syndrome coro    .................... .................... ..................... .................... .................... ....................
30605 MT459923.1 |Severe acute respiratory syndrome coro    .................... .................... ..................... .................... .................... ....................
30606 MT459924.1 |Severe acute respiratory syndrome coro    .................... .................... ..................... .................... .................... ....................
30607 MT459925.1 |Severe acute respiratory syndrome coro    .................... .................... ..................... .................... .................... ....................
30608 MT459979.1 |Severe acute respiratory syndrome coro    .................... .................... ..................... .................... .................... ....................
30609 MT459985.1 |Severe acute respiratory syndrome coro    .................... .................... ..................... .................... .................... ....................
30610 MT459986.1 |Severe acute respiratory syndrome coro    .................... .................... ..................... .................... .................... ....................
30611 MT459987.1 |Severe acute respiratory syndrome coro    .................... .................... ..................... .................... .................... ....................
30612 MT459988.1 |Severe acute respiratory syndrome coro    .................... .................... ..................... .................... .................... ....................
30613 MT459989.1 |Severe acute respiratory syndrome coro    .................... .................... ..................... .................... .................... ....................
30614 MT459990.1 |Severe acute respiratory syndrome coro    .................... .................... ..................... .................... .................... ....................
30615 MT460089.1 |Severe acute respiratory syndrome coro    .................... .................... ..................... .................... .................... ....................
30616 MT460090.1 |Severe acute respiratory syndrome coro    .................... .................... ..................... .................... .................... ....................
30617 MT460091.1 |Severe acute respiratory syndrome coro    .................... .................... ..................... .................... .................... ....................
30618 MT460092.1 |Severe acute respiratory syndrome coro    .................... .................... ..................... .................... .................... ....................
30619 MT460113.1 |Severe acute respiratory syndrome coro    .................... .................... ..................... .................... .................... ....................
30620 MT460114.1 |Severe acute respiratory syndrome coro    .................... .................... ..................... .................... .................... ....................
30621 MT460115.1 |Severe acute respiratory syndrome coro    .................... .................... ..................... .................... .................... ....................
30622 MT460116.1 |Severe acute respiratory syndrome coro    .................... .................... ..................... .................... .................... ....................
30623 MT460117.1 |Severe acute respiratory syndrome coro    .................... .................... ..................... .................... .................... ....................
30624 MT460119.1 |Severe acute respiratory syndrome coro    .................... .................... ..................... .................... .................... ....................
30625 MT460120.1 |Severe acute respiratory syndrome coro    .................... .................... ..................... .................... .................... ....................
30626 MT460121.1 |Severe acute respiratory syndrome coro    .................... .................... ..................... .................... .................... ....................
30627 MT460124.1 |Severe acute respiratory syndrome coro    .................... .................... ..................... .................... .................... ....................
30628 MT460125.1 |Severe acute respiratory syndrome coro    .................... .................... ..................... .................... .................... ....................
30629 MT460127.1 |Severe acute respiratory syndrome coro    .................... .................... ..................... .................... .................... ....................
30630 MT460128.1 |Severe acute respiratory syndrome coro    .................... .................... ..................... .................... .................... ....................
30631 MT460129.1 |Severe acute respiratory syndrome coro    .................... .................... ..................... .................... .................... ....................
30632 MT460130.1 |Severe acute respiratory syndrome coro    .................... .................... ..................... .................... .................... ....................
30633 MT460132.1 |Severe acute respiratory syndrome coro    .................... .................... ..................... .................... .................... ....................
30634 MT460133.1 |Severe acute respiratory syndrome coro    .................... .................... ..................... .................... .................... ....................
30635 MT460135.1 |Severe acute respiratory syndrome coro    .................... .................... ..................... .................... .................... ....................
30636 MT460136.1 |Severe acute respiratory syndrome coro    .................... .................... ..................... .................... .................... ....................
30637 MT460137.1 |Severe acute respiratory syndrome coro    .................... .................... ..................... .................... .................... ....................
30638 MT460138.1 |Severe acute respiratory syndrome coro    .................... .................... ..................... .................... .................... ....................
30639 MT460139.1 |Severe acute respiratory syndrome coro    .................... .................... ..................... .................... .................... ....................
30640 MT460140.1 |Severe acute respiratory syndrome coro    .................... .................... ..................... .................... .................... ....................
30641 MT461603.1 |Severe acute respiratory syndrome coro    .................... .................... ..................... .................... .................... ....................
30642 MT461604.1 |Severe acute respiratory syndrome coro    .................... .................... ..................... .................... .................... ....................
30643 MT461605.1 |Severe acute respiratory syndrome coro    .................... .................... ..................... .................... .................... ....................
30644 MT461606.1 |Severe acute respiratory syndrome coro    .................... .................... ..................... .................... .................... ....................
30645 MT461607.1 |Severe acute respiratory syndrome coro    .................... .................... ..................... .................... .................... ....................
30646 MT461608.1 |Severe acute respiratory syndrome coro    .................... .................... ..................... .................... .................... ....................
30647 MT461609.1 |Severe acute respiratory syndrome coro    .................... .................... ..................... .................... .................... ....................
30648 MT461610.1 |Severe acute respiratory syndrome coro    .................... .................... ..................... .................... .................... ....................
30649 MT461611.1 |Severe acute respiratory syndrome coro    .................... .................... ..................... .................... .................... ....................
30650 MT461612.1 |Severe acute respiratory syndrome coro    .................... .................... ..................... .................... .................... ....................
30651 MT461613.1 |Severe acute respiratory syndrome coro    .................... .................... ..................... .................... .................... ....................
30652 MT461614.1 |Severe acute respiratory syndrome coro    .................... .................... ..................... .................... .................... ....................
```

```
30653 MT461615.1 |Severe acute respiratory syndrome coro    ....................  ....................  ....................  ....................  ....................  ....................
30654 MT461616.1 |Severe acute respiratory syndrome coro    ....................  ....................  ....................  ....................  ....................  ....................
30655 MT461617.1 |Severe acute respiratory syndrome coro    ....................  ....................  ....................  ....................  ....................  ....................
30656 MT461618.1 |Severe acute respiratory syndrome coro    ....................  ....................  ....................  ....................  ....................  ....................
30657 MT461619.1 |Severe acute respiratory syndrome coro    ....................  ....................  ....................  ....................  ....................  ....................
30658 MT461620.1 |Severe acute respiratory syndrome coro    ....................  ....................  ....................  ....................  ....................  ....................
30659 MT461621.1 |Severe acute respiratory syndrome coro    ....................  ....................  ....................  ....................  ....................  ....................
30660 MT461622.1 |Severe acute respiratory syndrome coro    ....................  ....................  ....................  ....................  ....................  ....................
30661 MT461623.1 |Severe acute respiratory syndrome coro    ....................  ....................  ....................  ....................  ....................  ....................
30662 MT461624.1 |Severe acute respiratory syndrome coro    ....................  ....................  ....................  ....................  ....................  ....................
30663 MT461625.1 |Severe acute respiratory syndrome coro    ....................  ....................  ....................  ....................  ....................  ....................
30664 MT461626.1 |Severe acute respiratory syndrome coro    ....................  ....................  ....................  ....................  ....................  ....................
30665 MT461627.1 |Severe acute respiratory syndrome coro    ....................  ....................  ....................  ....................  ....................  ....................
30666 MT461628.1 |Severe acute respiratory syndrome coro    ....................  ....................  ....................  ....................  ....................  ....................
30667 MT461629.1 |Severe acute respiratory syndrome coro    ....................  ....................  ....................  ....................  ....................  ....................
30668 MT461630.1 |Severe acute respiratory syndrome coro    ....................  ....................  ....................  ....................  ....................  ....................
30669 MT461631.1 |Severe acute respiratory syndrome coro    ....................  ....................  ....................  ....................  ....................  ....................
30670 MT461632.1 |Severe acute respiratory syndrome coro    ....................  ....................  ....................  ....................  ....................  ....................
30671 MT461633.1 |Severe acute respiratory syndrome coro    ....................  ....................  ....................  ....................  ....................  ....................
30672 MT461634.1 |Severe acute respiratory syndrome coro    ....................  ....................  ....................  ....................  ....................  ....................
30673 MT461635.1 |Severe acute respiratory syndrome coro    ....................  ....................  ....................  ....................  ....................  ....................
30674 MT461636.1 |Severe acute respiratory syndrome coro    ....................  ....................  ....................  ....................  ....................  ....................
30675 MT461637.1 |Severe acute respiratory syndrome coro    ....................  ....................  ....................  ....................  ....................  ....................
30676 MT461638.1 |Severe acute respiratory syndrome coro    ....................  ....................  ....................  ....................  ....................  ....................
30677 MT461639.1 |Severe acute respiratory syndrome coro    ....................  ....................  ....................  ....................  ....................  ....................
30678 MT461640.1 |Severe acute respiratory syndrome coro    ....................  ....................  ....................  ....................  ....................  ....................
30679 MT461641.1 |Severe acute respiratory syndrome coro    ....................  ....................  ....................  ....................  ....................  ....................
30680 MT461642.1 |Severe acute respiratory syndrome coro    ....................  ....................  ....................  ....................  ....................  ....................
30681 MT461643.1 |Severe acute respiratory syndrome coro    ....................  ....................  ....................  ....................  ....................  ....................
30682 MT461644.1 |Severe acute respiratory syndrome coro    ....................  ....................  ....................  ....................  ....................  ....................
30683 MT461645.1 |Severe acute respiratory syndrome coro    ....................  ....................  ....................  ....................  ....................  ....................
30684 MT461646.1 |Severe acute respiratory syndrome coro    ....................  ....................  ....................  ....................  ....................  ....................
30685 MT461647.1 |Severe acute respiratory syndrome coro    ....................  ....................  ....................  ....................  ....................  ....................
30686 MT461648.1 |Severe acute respiratory syndrome coro    ....................  ....................  ....................  ....................  ....................  ....................
30687 MT461649.1 |Severe acute respiratory syndrome coro    ....................  ....................  ....................  ....................  ....................  ....................
30688 MT461650.1 |Severe acute respiratory syndrome coro    ....................  ....................  ....................  ....................  ....................  ....................
30689 MT461651.1 |Severe acute respiratory syndrome coro    ....................  ....................  ....................  ....................  ....................  ....................
30690 MT461652.1 |Severe acute respiratory syndrome coro    ....................  ....................  ....................  ....................  ....................  ....................
30691 MT461653.1 |Severe acute respiratory syndrome coro    ....................  ....................  ....................  ....................  ....................  ....................
30692 MT461654.1 |Severe acute respiratory syndrome coro    ....................  ....................  ....................  ....................  ....................  ....................
30693 MT461655.1 |Severe acute respiratory syndrome coro    ....................  ....................  ....................  ....................  ....................  ....................
30694 MT461656.1 |Severe acute respiratory syndrome coro    ....................  ....................  ....................  ....................  ....................  ....................
30695 MT461657.1 |Severe acute respiratory syndrome coro    ....................  ....................  ....................  ....................  ....................  ....................
30696 MT461658.1 |Severe acute respiratory syndrome coro    ....................  ....................  ....................  ....................  ....................  ....................
30697 MT461659.1 |Severe acute respiratory syndrome coro    ....................  ....................  ....................  ....................  ....................  ....................
30698 MT461660.1 |Severe acute respiratory syndrome coro    ....................  ....................  ....................  ....................  ....................  ....................
30699 MT461662.1 |Severe acute respiratory syndrome coro    ....................  ....................  ....................  ....................  ....................  ....................
30700 MT461663.1 |Severe acute respiratory syndrome coro    ....................  ....A...............  ....................  ....................  ....................  ....................
30701 MT461664.1 |Severe acute respiratory syndrome coro    ....................  ....................  ....................  ....................  ....................  ....................
30702 MT461666.1 |Severe acute respiratory syndrome coro    ....................  ...................A  ....................  ....................  ....................  ....................        1
30703 MT461667.1 |Severe acute respiratory syndrome coro    ....................  ....................  ....................  ....................  ....................  ....................
30704 MT461668.1 |Severe acute respiratory syndrome coro    ....................  ....................  ....................  ....................  ....................  ....................
30705 MT457390.1 |Severe acute respiratory syndrome coro    ....................  ....................  ....................  ....................  ....................  ....................
30706 MT457391.1 |Severe acute respiratory syndrome coro    ....................  ....................  ....................  ....................  ....................  ....................
30707 MT457392.1 |Severe acute respiratory syndrome coro    ....................  ....................  ....................  ....................  ....................  ....................
30708 MT457393.1 |Severe acute respiratory syndrome coro    ....................  ....................  ....................  ....................  ....................  ....................
30709 MT457394.1 |Severe acute respiratory syndrome coro    ....................  ....................  ....................  ....................  ....A...............  ....................
30710 MT457395.1 |Severe acute respiratory syndrome coro    ....................  ....................  ....................  ....................  ....A...............  ....................
30711 MT457396.1 |Severe acute respiratory syndrome coro    ....................  ....................  ....................  ....................  ....................  ....................
30712 MT457397.1 |Severe acute respiratory syndrome coro    ....................  ....................  ....................  ....................  ....................  ....................
30713 MT457398.1 |Severe acute respiratory syndrome coro    ....................  ....................  ....................  ....................  ....A...............  ....................
30714 MT457399.1 |Severe acute respiratory syndrome coro    ....................  ....................  ....................  ....................  ....A...............  ....................
30715 MT457400.1 |Severe acute respiratory syndrome coro    ....................  ....................  ....................  ....................  ....................  ....................
30716 MT457401.1 |Severe acute respiratory syndrome coro    ....................  ....................  ....................  ....................  ....................  ....................
30717 MT457402.1 |Severe acute respiratory syndrome coro    ....................  ....................  ....................  ....................  ....................  ....................
30718 MT457403.1 |Severe acute respiratory syndrome coro    ....................  ....................  ....................  ....................  ....................  ....................
30719 MT449636.1 |Severe acute respiratory syndrome coro    ....................  ....A...............  ....................  ....................  ....................  ....................
30720 MT449637.1 |Severe acute respiratory syndrome coro    ....................  ....................  ....................  ....................  ....................  ....................
30721 MT449639.1 |Severe acute respiratory syndrome coro    ....................  ....................  ....................  ....................  ....................  ....................
30722 MT449641.1 |Severe acute respiratory syndrome coro    ....................  ....................  ....................  ....................  ....................  ....................
30723 MT449644.1 |Severe acute respiratory syndrome coro    ....................  ....................  ....................  ....................  ....................  ....................
30724 MT449647.1 |Severe acute respiratory syndrome coro    ....................  ....................  ....................  ....................  ....................  ....................
30725 MT449651.1 |Severe acute respiratory syndrome coro    ....................  ....................  ....................  ....................  ....................  ....................
30726 MT449656.1 |Severe acute respiratory syndrome coro    ....................  ....................  ....................  ....................  ....................  ....................
30727 MT449664.1 |Severe acute respiratory syndrome coro    ....................  ....................  ....................  ....................  ....................  ....................
30728 MT449665.1 |Severe acute respiratory syndrome coro    ....................  ....................  ....................  ....................  ....................  ....................
30729 MT449670.1 |Severe acute respiratory syndrome coro    ....................  ....................  ....................  ....................  ....................  ....................
30730 MT449672.1 |Severe acute respiratory syndrome coro    ....................  ....................  ....................  ....................  ....................  ....................
```

30731 MT449673.1 |Severe acute respiratory syndrome coro .................... .................... ..................... .................... .................... ....................
30732 MT450872.1 |Severe acute respiratory syndrome coro .................... .................... ..................... .................... .................... ....................
30733 MT450873.1 |Severe acute respiratory syndrome coro .................... .................... ..................... .................... .................... ....................
30734 MT450874.1 |Severe acute respiratory syndrome coro .................... .................... ..................... .................... .................... ....................
30735 MT450875.1 |Severe acute respiratory syndrome coro .................... .................... ..................... .................... .................... ....................
30736 MT450876.1 |Severe acute respiratory syndrome coro .................... .................... ..................... .................... .................... ....................
30737 MT450877.1 |Severe acute respiratory syndrome coro .................... .................... ..................... .................... .................... ....................
30738 MT450878.1 |Severe acute respiratory syndrome coro .................... .................... ..................... .................... .................... ....................
30739 MT450879.1 |Severe acute respiratory syndrome coro .................... .................... ..................... .................... .................... ....................
30740 MT450880.1 |Severe acute respiratory syndrome coro .................... .................... ..................... .................... .................... ....................
30741 MT450881.1 |Severe acute respiratory syndrome coro .................... .................... ..................... .................... .................... ....................
30742 MT450882.1 |Severe acute respiratory syndrome coro .................... .................... ..................... .................... .................... ....................
30743 MT450883.1 |Severe acute respiratory syndrome coro .................... .................... ..................... .................... .................... ....................
30744 MT450884.1 |Severe acute respiratory syndrome coro .................... .................... ..................... .................... .................... ....................
30745 MT450885.1 |Severe acute respiratory syndrome coro .................... .................... ..................... .................... .................... ....................
30746 MT450886.1 |Severe acute respiratory syndrome coro .................... .................... ..................... .................... .................... ....................
30747 MT450887.1 |Severe acute respiratory syndrome coro .................... .................... ..................... .................... .................... ....................
30748 MT450889.1 |Severe acute respiratory syndrome coro .................... .................... ..................... .................... .................... ....................
30749 MT450890.1 |Severe acute respiratory syndrome coro .................... .................... ..................... .................... .................... ....................
30750 MT450891.1 |Severe acute respiratory syndrome coro .................... .................... ..................... .................... .................... ....................
30751 MT450892.1 |Severe acute respiratory syndrome coro .................... .................... ..................... .................... .................... ....................
30752 MT450893.1 |Severe acute respiratory syndrome coro .................... .................... ..................... .................... .................... ....................
30753 MT450894.1 |Severe acute respiratory syndrome coro .................... .................... ..................... .................... .................... ....................
30754 MT450895.1 |Severe acute respiratory syndrome coro .................... .................... ..................... .................... .................... ....................
30755 MT450896.1 |Severe acute respiratory syndrome coro .................... .................... ..................... .................... .................... ....................
30756 MT450897.1 |Severe acute respiratory syndrome coro .................... .................... ..................... .................... .................... ....................
30757 MT450898.1 |Severe acute respiratory syndrome coro .................... .................... ..................... .................... .................... ....................
30758 MT450899.1 |Severe acute respiratory syndrome coro .................... .................... ..................... .................... .................... ....................
30759 MT450900.1 |Severe acute respiratory syndrome coro .................... .................... ..................... .................... .................... ....................
30760 MT450901.1 |Severe acute respiratory syndrome coro .................... .................... ..................... .................... .................... ....................
30761 MT450902.1 |Severe acute respiratory syndrome coro .................... .................... ..................... .................... .................... ....................
30762 MT450903.1 |Severe acute respiratory syndrome coro .................... .................... ..................... .................... .................... ....................
30763 MT450904.1 |Severe acute respiratory syndrome coro .................... .................... ..................... .................... .................... ....................
30764 MT450905.1 |Severe acute respiratory syndrome coro .................... .................... ..................... .................... .................... ....................
30765 MT450906.1 |Severe acute respiratory syndrome coro .................... .................... ..................... .................... .................... ....................
30766 MT450907.1 |Severe acute respiratory syndrome coro .................... .................... ..................... .................... .................... ....................
30767 MT450909.1 |Severe acute respiratory syndrome coro .................... .................... ..................... .................... .................... ....................
30768 MT450910.1 |Severe acute respiratory syndrome coro .................... .................... ..................... .................... .................... ....................
30769 MT450911.1 |Severe acute respiratory syndrome coro .................... .................... ..................... .................... .................... ....................
30770 MT450912.1 |Severe acute respiratory syndrome coro .................... .................... ..................... .................... .................... ....................
30771 MT450913.1 |Severe acute respiratory syndrome coro .................... .................... ..................... .................... .................... ....................
30772 MT450914.1 |Severe acute respiratory syndrome coro .................... .................... ..................... .................... .................... ....................
30773 MT450915.1 |Severe acute respiratory syndrome coro .................... .................... ..................... .................... .................... ....................
30774 MT450917.1 |Severe acute respiratory syndrome coro .................... .................... ..................... .................... .................... ....................
30775 MT450918.1 |Severe acute respiratory syndrome coro .................... .................... ..................... .................... .................... ....................
30776 MT450932.1 |Severe acute respiratory syndrome coro .................... .................... ..................... .................... .................... ....................
30777 MT450935.1 |Severe acute respiratory syndrome coro .................... .................... ..................... .................... .................... ....................
30778 MT450936.1 |Severe acute respiratory syndrome coro .................... .................... ..................... .................... .................... ....................
30779 MT450938.1 |Severe acute respiratory syndrome coro .................... .................... ..................... .................... .................... ....................
30780 MT450941.1 |Severe acute respiratory syndrome coro .................... .................... ..................... .................... .................... ....................
30781 MT450942.1 |Severe acute respiratory syndrome coro .................... .................... ..................... .................... .................... ....................
30782 MT450943.1 |Severe acute respiratory syndrome coro .................... .................... ..................... .................... .................... ....................
30783 MT450944.1 |Severe acute respiratory syndrome coro .................... .................... ..................... .................... .................... ....................
30784 MT450946.1 |Severe acute respiratory syndrome coro .................... .................... ..................... .................... .................... ....................
30785 MT450948.1 |Severe acute respiratory syndrome coro .................... .................... ..................... .................... .................... ....................
30786 MT450949.1 |Severe acute respiratory syndrome coro .................... .................... ..................... .................... .................... ....................
30787 MT450950.1 |Severe acute respiratory syndrome coro .................... .................... ..................... .................... .................... ....................
30788 MT450951.1 |Severe acute respiratory syndrome coro .................... .................... ..................... .................... .................... ....................
30789 MT450954.1 |Severe acute respiratory syndrome coro .................... .................... ..................... .................... .................... ....................
30790 MT450955.1 |Severe acute respiratory syndrome coro .................... .................... ..................... .................... .................... ....................
30791 MT450956.1 |Severe acute respiratory syndrome coro .................... .................... ..................... .................... .................... ....................
30792 MT450957.1 |Severe acute respiratory syndrome coro .................... .................... ..................... .................... .................... ....................
30793 MT450960.1 |Severe acute respiratory syndrome coro .................... .................... ..................... .................... .................... ....................
30794 MT450961.1 |Severe acute respiratory syndrome coro .................... .................... ..................... .................... .................... ....................
30795 MT450962.1 |Severe acute respiratory syndrome coro .................... .................... ..................... .................... .................... ....................
30796 MT450963.1 |Severe acute respiratory syndrome coro .................... .................... ..................... .................... .................... ....................
30797 MT450965.1 |Severe acute respiratory syndrome coro .................... .................... ..................... .................... .................... ....................
30798 MT450966.1 |Severe acute respiratory syndrome coro .................... .................... ..................... .................... .................... ....................
30799 MT450968.1 |Severe acute respiratory syndrome coro .................... .................... ..................... .................... .................... ....................
30800 MT450969.1 |Severe acute respiratory syndrome coro .................... .................... ..................... .................... .................... ....................
30801 MT450970.1 |Severe acute respiratory syndrome coro .................... .................... ..................... .................... .................... ....................
30802 MT450971.1 |Severe acute respiratory syndrome coro .................... .................... ..................... .................... .................... ....................
30803 MT450972.1 |Severe acute respiratory syndrome coro .................... .................... ..................... .................... .................... ....................
30804 MT450973.1 |Severe acute respiratory syndrome coro .................... .................... ..................... .................... .................... ....................
30805 MT450974.1 |Severe acute respiratory syndrome coro .................... .................... ..................... .................... .................... ....................
30806 MT450977.1 |Severe acute respiratory syndrome coro .................... .................... ..................... .................... .................... ....................
30807 MT450980.1 |Severe acute respiratory syndrome coro .................... .................... ..................... .................... .................... ....................
30808 MT450981.1 |Severe acute respiratory syndrome coro .................... .................... ..................... .................... .................... ....................

| | | | | | | | | |
|---|---|---|---|---|---|---|---|---|
| 30809 | MT450982.1 | \|Severe acute respiratory syndrome coro | .................... | .................... | .................... | .................... | .................... | .................... |
| 30810 | MT450983.1 | \|Severe acute respiratory syndrome coro | .................... | .................... | .................... | .................... | .................... | .................... |
| 30811 | MT450984.1 | \|Severe acute respiratory syndrome coro | .................... | .................... | .................... | .................... | .................... | .................... |
| 30812 | MT450985.1 | \|Severe acute respiratory syndrome coro | .................... | .................... | .................... | .................... | .................... | .................... |
| 30813 | MT450988.1 | \|Severe acute respiratory syndrome coro | .................... | .................... | .................... | .................... | .................... | .................... |
| 30814 | MT450989.1 | \|Severe acute respiratory syndrome coro | .................... | .................... | .................... | .................... | .................... | .................... |
| 30815 | MT450990.1 | \|Severe acute respiratory syndrome coro | .................... | .................... | .................... | .................... | .................... | .................... |
| 30816 | MT450991.1 | \|Severe acute respiratory syndrome coro | .................... | .................... | .................... | .................... | .................... | .................... |
| 30817 | MT450992.1 | \|Severe acute respiratory syndrome coro | .................... | .................... | .................... | .................... | .................... | .................... |
| 30818 | MT450993.1 | \|Severe acute respiratory syndrome coro | .................... | .................... | .................... | .................... | .................... | .................... |
| 30819 | MT450994.1 | \|Severe acute respiratory syndrome coro | .................... | .................... | .................... | .................... | .................... | .................... |
| 30820 | MT450995.1 | \|Severe acute respiratory syndrome coro | .................... | .................... | .................... | .................... | .................... | .................... |
| 30821 | MT450996.1 | \|Severe acute respiratory syndrome coro | .................... | .................... | .................... | .................... | .................... | .................... |
| 30822 | MT450997.1 | \|Severe acute respiratory syndrome coro | .................... | .................... | .................... | .................... | .................... | .................... |
| 30823 | MT450998.1 | \|Severe acute respiratory syndrome coro | .................... | .................... | .................... | .................... | .................... | .................... |
| 30824 | MT450999.1 | \|Severe acute respiratory syndrome coro | .................... | .................... | .................... | .................... | .................... | .................... |
| 30825 | MT451000.1 | \|Severe acute respiratory syndrome coro | .................... | .................... | .................... | .................... | .................... | .................... |
| 30826 | MT451002.1 | \|Severe acute respiratory syndrome coro | .................... | .................... | .................... | .................... | .................... | .................... |
| 30827 | MT451004.1 | \|Severe acute respiratory syndrome coro | .................... | .................... | .................... | .................... | .................... | .................... |
| 30828 | MT451005.1 | \|Severe acute respiratory syndrome coro | .................... | .................... | .................... | .................... | .................... | .................... |
| 30829 | MT451006.1 | \|Severe acute respiratory syndrome coro | .................... | .................... | .................... | .................... | .................... | .................... |
| 30830 | MT451007.1 | \|Severe acute respiratory syndrome coro | .................... | .................... | .................... | .................... | .................... | .................... |
| 30831 | MT451008.1 | \|Severe acute respiratory syndrome coro | .................... | .................... | .................... | .................... | .................... | .................... |
| 30832 | MT451009.1 | \|Severe acute respiratory syndrome coro | .................... | .................... | .................... | .................... | .................... | .................... |
| 30833 | MT451013.1 | \|Severe acute respiratory syndrome coro | .................... | .................... | .................... | .................... | .................... | .................... |
| 30834 | MT451015.1 | \|Severe acute respiratory syndrome coro | .................... | .................... | .................... | .................... | .................... | .................... |
| 30835 | MT451017.1 | \|Severe acute respiratory syndrome coro | .................... | .................... | .................... | .................... | .................... | .................... |
| 30836 | MT451018.1 | \|Severe acute respiratory syndrome coro | .................... | .................... | .................... | .................... | .................... | .................... |
| 30837 | MT451019.1 | \|Severe acute respiratory syndrome coro | .................... | .................... | .................... | .................... | .................... | .................... |
| 30838 | MT451020.1 | \|Severe acute respiratory syndrome coro | .................... | .................... | .................... | .................... | .................... | .................... |
| 30839 | MT451021.1 | \|Severe acute respiratory syndrome coro | .................... | .................... | .................... | .................... | .................... | .................... |
| 30840 | MT451022.1 | \|Severe acute respiratory syndrome coro | .................... | .................... | .................... | .................... | .................... | .................... |
| 30841 | MT451024.1 | \|Severe acute respiratory syndrome coro | .................... | .................... | .................... | .................... | .................... | .................... |
| 30842 | MT451026.1 | \|Severe acute respiratory syndrome coro | .................... | .................... | .................... | .................... | .................... | .................... |
| 30843 | MT451033.1 | \|Severe acute respiratory syndrome coro | .................... | .................... | .................... | .................... | .................... | .................... |
| 30844 | MT451035.1 | \|Severe acute respiratory syndrome coro | .................... | .................... | .................... | .................... | .................... | .................... |
| 30845 | MT451036.1 | \|Severe acute respiratory syndrome coro | .................... | .................... | .................... | .................... | .................... | .................... |
| 30846 | MT451037.1 | \|Severe acute respiratory syndrome coro | .................... | .................... | .................... | .................... | .................... | .................... |
| 30847 | MT451038.1 | \|Severe acute respiratory syndrome coro | .................... | .................... | .................... | .................... | .................... | .................... |
| 30848 | MT451039.1 | \|Severe acute respiratory syndrome coro | .................... | .................... | .................... | .................... | .................... | .................... |
| 30849 | MT451040.1 | \|Severe acute respiratory syndrome coro | .................... | .................... | .................... | .................... | .................... | .................... |
| 30850 | MT451041.1 | \|Severe acute respiratory syndrome coro | .................... | .................... | .................... | .................... | .................... | .................... |
| 30851 | MT451042.1 | \|Severe acute respiratory syndrome coro | .................... | .................... | .................... | .................... | .................... | .................... |
| 30852 | MT451044.1 | \|Severe acute respiratory syndrome coro | .................... | .................... | .................... | .................... | .................... | .................... |
| 30853 | MT451045.1 | \|Severe acute respiratory syndrome coro | .................... | .................... | .................... | .................... | .................... | .................... |
| 30854 | MT451046.1 | \|Severe acute respiratory syndrome coro | .................... | .................... | .................... | .................... | .................... | .................... |
| 30855 | MT451047.1 | \|Severe acute respiratory syndrome coro | .................... | .................... | .................... | .................... | .................... | .................... |
| 30856 | MT451048.1 | \|Severe acute respiratory syndrome coro | .................... | .................... | .................... | .................... | .................... | .................... |
| 30857 | MT451049.1 | \|Severe acute respiratory syndrome coro | .................... | .................... | .................... | .................... | .................... | .................... |
| 30858 | MT451051.1 | \|Severe acute respiratory syndrome coro | .................... | .................... | .................... | .................... | .................... | .................... |
| 30859 | MT451052.1 | \|Severe acute respiratory syndrome coro | .................... | .................... | .................... | .................... | .................... | .................... |
| 30860 | MT451056.1 | \|Severe acute respiratory syndrome coro | .................... | .................... | .................... | .................... | .................... | .................... |
| 30861 | MT451057.1 | \|Severe acute respiratory syndrome coro | .................... | .................... | .................... | .................... | .................... | .................... |
| 30862 | MT451058.1 | \|Severe acute respiratory syndrome coro | .................... | .................... | .................... | .................... | .................... | .................... |
| 30863 | MT451059.1 | \|Severe acute respiratory syndrome coro | .................... | .................... | .................... | .................... | .................... | .................... |
| 30864 | MT451060.1 | \|Severe acute respiratory syndrome coro | .................... | .................... | .................... | .................... | .................... | .................... |
| 30865 | MT451061.1 | \|Severe acute respiratory syndrome coro | .................... | .................... | .................... | .................... | .................... | .................... |
| 30866 | MT451062.1 | \|Severe acute respiratory syndrome coro | .................... | .................... | .................... | .................... | .................... | .................... |
| 30867 | MT451065.1 | \|Severe acute respiratory syndrome coro | .................... | .................... | .................... | .................... | .................... | .................... |
| 30868 | MT451066.1 | \|Severe acute respiratory syndrome coro | .................... | .................... | .................... | .................... | .................... | .................... |
| 30869 | MT451072.1 | \|Severe acute respiratory syndrome coro | .................... | .............A...... | .................... | .................... | .................... | .................... |
| 30870 | MT451074.1 | \|Severe acute respiratory syndrome coro | .................... | .................... | .................... | .................... | .................... | .................... |
| 30871 | MT451076.1 | \|Severe acute respiratory syndrome coro | .................... | .................... | .................... | .................... | .................... | .................... |
| 30872 | MT451077.1 | \|Severe acute respiratory syndrome coro | .................... | .................... | .................... | .................... | .................... | .................... |
| 30873 | MT451079.1 | \|Severe acute respiratory syndrome coro | .................... | .................... | .................... | .................... | .................... | .................... |
| 30874 | MT451081.1 | \|Severe acute respiratory syndrome coro | .................... | .................... | .................... | .................... | .................... | .................... |
| 30875 | MT451086.1 | \|Severe acute respiratory syndrome coro | .................... | .................... | .................... | .................... | .................... | .................... |
| 30876 | MT451087.1 | \|Severe acute respiratory syndrome coro | .................... | .................... | .................... | .................... | .................... | .................... |
| 30877 | MT451088.1 | \|Severe acute respiratory syndrome coro | .................... | .................... | .................... | .................... | .................... | .................... |
| 30878 | MT451090.1 | \|Severe acute respiratory syndrome coro | .................... | .................... | .................... | .................... | .................... | .................... |
| 30879 | MT451092.1 | \|Severe acute respiratory syndrome coro | .................... | .................... | .................... | .................... | .................... | .................... |
| 30880 | MT451093.1 | \|Severe acute respiratory syndrome coro | .................... | .................... | .................... | .................... | .................... | .................... |
| 30881 | MT451094.1 | \|Severe acute respiratory syndrome coro | .................... | .................... | .................... | .................... | .................... | .................... |
| 30882 | MT451098.1 | \|Severe acute respiratory syndrome coro | .................... | .................... | .................... | .................... | .................... | .................... |
| 30884 | MT451101.1 | \|Severe acute respiratory syndrome coro | .................... | .................... | .................... | .................... | .................... | .................... |
| 30885 | MT451102.1 | \|Severe acute respiratory syndrome coro | .................... | .................... | .................... | .................... | .................... | .................... |
| 30886 | MT451103.1 | \|Severe acute respiratory syndrome coro | .................... | .................... | .................... | .................... | .................... | .................... |
| 30887 | MT451104.1 | \|Severe acute respiratory syndrome coro | .................... | .................... | .................... | .................... | .................... | .................... |

30888 MT451105.1 |Severe acute respiratory syndrome coro ···················· ···················· ···················· ···················· ···················· ····················
30889 MT451108.1 |Severe acute respiratory syndrome coro ···················· ···················· ···················· ···················· ···················· ····················
30890 MT451109.1 |Severe acute respiratory syndrome coro ···················· ···················· ···················· ···················· ···················· ····················
30891 MT451114.1 |Severe acute respiratory syndrome coro ···················· ···················· ···················· ···················· ···················· ····················
30892 MT451115.1 |Severe acute respiratory syndrome coro ···················· ···················· ···················· ···················· ···················· ····················
30893 MT451116.1 |Severe acute respiratory syndrome coro ···················· ···················· ···················· ···················· ···················· ····················
30894 MT451117.1 |Severe acute respiratory syndrome coro ···················· ···················· ···················· ···················· ···················· ····················
30895 MT451118.1 |Severe acute respiratory syndrome coro ···················· ···················· ···················· ···················· ······A············· ····················
30896 MT451119.1 |Severe acute respiratory syndrome coro ···················· ···················· ···················· ···················· ···················· ····················
30897 MT451121.1 |Severe acute respiratory syndrome coro ···················· ···················· ···················· ···················· ···················· ····················
30898 MT451122.1 |Severe acute respiratory syndrome coro ···················· ···················· ···················· ···················· ···················· ····················
30899 MT451123.1 |Severe acute respiratory syndrome coro ···················· ···················· ···················· ···················· ···················· ····················
30900 MT451124.1 |Severe acute respiratory syndrome coro ···················· ···················· ···················· ···················· ······A············· ····················
30901 MT451125.1 |Severe acute respiratory syndrome coro ···················· ···················· ···················· ···················· ···················· ····················
30902 MT451126.1 |Severe acute respiratory syndrome coro ···················· ···················· ···················· ···················· ···················· ····················
30903 MT451127.1 |Severe acute respiratory syndrome coro ···················· ···················· ···················· ···················· ···················· ····················
30904 MT451128.1 |Severe acute respiratory syndrome coro ···················· ···················· ···················· ···················· ···················· ····················
30905 MT451129.1 |Severe acute respiratory syndrome coro ···················· ···················· ···················· ···················· ···················· ····················
30906 MT451130.1 |Severe acute respiratory syndrome coro ···················· ···················· ···················· ···················· ···················· ····················
30907 MT451131.1 |Severe acute respiratory syndrome coro ···················· ···················· ···················· ···················· ···················· ····················
30908 MT451132.1 |Severe acute respiratory syndrome coro ···················· ···················· ···················· ···················· ···················· ····················
30909 MT451133.1 |Severe acute respiratory syndrome coro ···················· ···················· ···················· ···················· ···················· ····················
30910 MT451134.1 |Severe acute respiratory syndrome coro ···················· ···················· ···················· ···················· ···················· ····················
30911 MT451135.1 |Severe acute respiratory syndrome coro ···················· ···················· ···················· ···················· ···················· ····················
30912 MT451136.1 |Severe acute respiratory syndrome coro ···················· ···················· ···················· ···················· ···················· ····················
30913 MT451137.1 |Severe acute respiratory syndrome coro ···················· ···················· ···················· ···················· ···················· ····················
30914 MT451138.1 |Severe acute respiratory syndrome coro ···················· ···················· ···················· ···················· ···················· ····················
30915 MT451139.1 |Severe acute respiratory syndrome coro ···················· ···················· ···················· ···················· ···················· ····················
30916 MT451140.1 |Severe acute respiratory syndrome coro ···················· ···················· ···················· ···················· ···················· ····················
30917 MT451141.1 |Severe acute respiratory syndrome coro ···················· ···················· ···················· ···················· ···················· ····················
30918 MT451142.1 |Severe acute respiratory syndrome coro ···················· ···················· ···················· ···················· ···················· ····················
30919 MT451143.1 |Severe acute respiratory syndrome coro ···················· ···················· ···················· ···················· ···················· ····················
30920 MT451144.1 |Severe acute respiratory syndrome coro ···················· ···················· ···················· ···················· ···················· ····················
30921 MT451145.1 |Severe acute respiratory syndrome coro ···················· ···················· ···················· ···················· ···················· ····················
30922 MT451146.1 |Severe acute respiratory syndrome coro ···················· ···················· ···················· ···················· ···················· ····················
30923 MT451147.1 |Severe acute respiratory syndrome coro ···················· ···················· ···················· ···················· ···················· ····················
30924 MT451148.1 |Severe acute respiratory syndrome coro ···················· ···················· ···················· ···················· ···················· ····················
30925 MT451149.1 |Severe acute respiratory syndrome coro ···················· ···················· ···················· ···················· ···················· ····················
30926 MT451150.1 |Severe acute respiratory syndrome coro ···················· ···················· ···················· ···················· ···················· ····················
30927 MT451151.1 |Severe acute respiratory syndrome coro ···················· ·A·················· ···················· ···················· ···················· ····················
30928 MT451152.1 |Severe acute respiratory syndrome coro ···················· ···················· ···················· ···················· ···················· ····················
30929 MT451153.1 |Severe acute respiratory syndrome coro ···················· ·A·················· ···················· ···················· ···················· ····················
30930 MT451154.1 |Severe acute respiratory syndrome coro ···················· ···················· ···················· ···················· ···················· ····················
30931 MT451155.1 |Severe acute respiratory syndrome coro ···················· ···················· ···················· ···················· ···················· ····················
30932 MT451156.1 |Severe acute respiratory syndrome coro ···················· ···················· ···················· ···················· ···················· ····················
30933 MT451157.1 |Severe acute respiratory syndrome coro ···················· ···················· ···················· ···················· ···················· ····················
30934 MT451158.1 |Severe acute respiratory syndrome coro ···················· ···················· ···················· ···················· ···················· ····················
30935 MT451159.1 |Severe acute respiratory syndrome coro ···················· ···················· ···················· ···················· ···················· ····················
30936 MT451160.1 |Severe acute respiratory syndrome coro ···················· ···················· ···················· ···················· ······A············· ····················
30937 MT451161.1 |Severe acute respiratory syndrome coro ···················· ··M················· ···················· ···················· ···················· ····················
30938 MT451162.1 |Severe acute respiratory syndrome coro ···················· ···················· ···················· ···················· ······A············· ····················
30939 MT451163.1 |Severe acute respiratory syndrome coro ···················· ···················· ···················· ···················· ···················· ····················
30940 MT451164.1 |Severe acute respiratory syndrome coro ···················· ···················· ···················· ···················· ···················· ····················
30941 MT451165.1 |Severe acute respiratory syndrome coro ···················· ···················· ···················· ···················· ···················· ····················
30942 MT451166.1 |Severe acute respiratory syndrome coro ···················· ···················· ···················· ···················· ···················· ····················
30943 MT451167.1 |Severe acute respiratory syndrome coro ···················· ···················· ···················· ···················· ···················· ····················
30944 MT451168.1 |Severe acute respiratory syndrome coro ···················· ···················· ···················· ···················· ···················· ····················
30945 MT451169.1 |Severe acute respiratory syndrome coro ···················· ···················· ···················· ···················· ···················· ····················
30946 MT451170.1 |Severe acute respiratory syndrome coro ···················· ···················· ···················· ···················· ···················· ····················
30947 MT451171.1 |Severe acute respiratory syndrome coro ···················· ···················· ···················· ···················· ···················· ····················
30948 MT451172.1 |Severe acute respiratory syndrome coro ···················· ···················· ···················· ···················· ···················· ····················
30949 MT451173.1 |Severe acute respiratory syndrome coro ···················· ···················· ···················· ···················· ···················· ····················
30950 MT451174.1 |Severe acute respiratory syndrome coro ···················· ···················· ···················· ···················· ···················· ····················
30951 MT451175.1 |Severe acute respiratory syndrome coro ···················· ···················· ···················· ···················· ···················· ····················
30952 MT451176.1 |Severe acute respiratory syndrome coro ···················· ···················· ···················· ···················· ···················· ····················
30953 MT451177.1 |Severe acute respiratory syndrome coro ···················· ···················· ···················· ···················· ···················· ····················
30954 MT451178.1 |Severe acute respiratory syndrome coro ···················· ···················· ···················· ···················· ···················· ····················
30955 MT451179.1 |Severe acute respiratory syndrome coro ···················· ···················· ···················· ···················· ···················· ····················
30956 MT451180.1 |Severe acute respiratory syndrome coro ···················· ···················· ···················· ···················· ···················· ····················
30957 MT451181.1 |Severe acute respiratory syndrome coro ···················· ···················· ···················· ···················· ···················· ····················
30958 MT451182.1 |Severe acute respiratory syndrome coro ···················· ···················· ···················· ···················· ···················· ····················
30959 MT451183.1 |Severe acute respiratory syndrome coro ···················· ···················· ···················· ···················· ···················· ····················
30960 MT451184.1 |Severe acute respiratory syndrome coro ···················· ···················· ···················· ···················· ···················· ····················
30961 MT451185.1 |Severe acute respiratory syndrome coro ···················· ···················· ···················· ···················· ···················· ····················
30962 MT451186.1 |Severe acute respiratory syndrome coro ···················· ···················· ···················· ···················· ···················· ····················
30963 MT451187.1 |Severe acute respiratory syndrome coro ···················· ···················· ···················· ···················· ···················· ····················
30964 MT451188.1 |Severe acute respiratory syndrome coro ···················· ···················· ···················· ···················· ···················· ····················
30965 MT451189.1 |Severe acute respiratory syndrome coro ···················· ···················· ···················· ···················· ···················· ····················

| | | | | | | | | |
|---|---|---|---|---|---|---|---|---|
| 30966 | MT451190.1 | \|Severe acute respiratory syndrome coro | .................... | .................... | ..................... | .................... | .................... | .................... |
| 30967 | MT451191.1 | \|Severe acute respiratory syndrome coro | .................... | .................... | ..................... | .................... | .................... | .................... |
| 30968 | MT451192.1 | \|Severe acute respiratory syndrome coro | .................... | .................... | ..................... | .................... | .................... | .................... |
| 30969 | MT451194.1 | \|Severe acute respiratory syndrome coro | .................... | .................... | ..................... | .................... | .................... | .................... |
| 30970 | MT451195.1 | \|Severe acute respiratory syndrome coro | .................... | .................... | ..................... | .................... | .................... | .................... |
| 30971 | MT451196.1 | \|Severe acute respiratory syndrome coro | .................... | .................... | ..................... | .................... | .................... | .................... |
| 30972 | MT451197.1 | \|Severe acute respiratory syndrome coro | .................... | .................... | ..................... | .................... | .................... | .................... |
| 30973 | MT451198.1 | \|Severe acute respiratory syndrome coro | .................... | .................... | ..................... | .................... | .................... | .................... |
| 30974 | MT451199.1 | \|Severe acute respiratory syndrome coro | .................... | .................... | ..................... | .................... | .................... | .................... |
| 30975 | MT451201.1 | \|Severe acute respiratory syndrome coro | .................... | .................... | ..................... | .................... | .................... | .................... |
| 30976 | MT451202.1 | \|Severe acute respiratory syndrome coro | .................... | .................... | ..................... | .................... | .................... | .................... |
| 30977 | MT451203.1 | \|Severe acute respiratory syndrome coro | .................... | .................... | ..................... | .................... | .................... | .................... |
| 30978 | MT451204.1 | \|Severe acute respiratory syndrome coro | .................... | .................... | ..................... | .................... | .................... | .................... |
| 30979 | MT451207.1 | \|Severe acute respiratory syndrome coro | .................... | .................... | ..................... | .................... | .................... | .................... |
| 30980 | MT451208.1 | \|Severe acute respiratory syndrome coro | .................... | .................... | ..................... | .................... | .................... | .................... |
| 30981 | MT451209.1 | \|Severe acute respiratory syndrome coro | .................... | .................... | ..................... | .................... | .................... | .................... |
| 30982 | MT451210.1 | \|Severe acute respiratory syndrome coro | .................... | .................... | ..................... | .................... | .................... | .................... |
| 30983 | MT451211.1 | \|Severe acute respiratory syndrome coro | .................... | .................... | ..................... | .................... | .................... | .................... |
| 30984 | MT451212.1 | \|Severe acute respiratory syndrome coro | .................... | .................... | ..................... | .................... | .................... | .................... |
| 30985 | MT451213.1 | \|Severe acute respiratory syndrome coro | .................... | .................... | ..................... | .................... | .................... | .................... |
| 30986 | MT451214.1 | \|Severe acute respiratory syndrome coro | .................... | .................... | ..................... | .................... | .................... | .................... |
| 30987 | MT451215.1 | \|Severe acute respiratory syndrome coro | .................... | .................... | ..................... | .................... | .................... | .................... |
| 30988 | MT451216.1 | \|Severe acute respiratory syndrome coro | .................... | .................... | ..................... | .................... | .................... | .................... |
| 30989 | MT451217.1 | \|Severe acute respiratory syndrome coro | .................... | .................... | ..................... | .................... | .................... | .................... |
| 30990 | MT451218.1 | \|Severe acute respiratory syndrome coro | .................... | .................... | ..................... | .................... | .................... | .................... |
| 30991 | MT451219.1 | \|Severe acute respiratory syndrome coro | .................... | .................... | ..................... | .................... | .................... | .................... |
| 30992 | MT451220.1 | \|Severe acute respiratory syndrome coro | .................... | .................... | ..................... | .................... | .................... | .................... |
| 30993 | MT451224.1 | \|Severe acute respiratory syndrome coro | .................... | .................... | ..................... | .................... | .................... | .................... |
| 30994 | MT451225.1 | \|Severe acute respiratory syndrome coro | .................... | .................... | ..................... | .................... | .................... | .................... |
| 30995 | MT451226.1 | \|Severe acute respiratory syndrome coro | .................... | .................... | ..................... | .................... | .................... | .................... |
| 30996 | MT451227.1 | \|Severe acute respiratory syndrome coro | .................... | .................... | ..................... | .................... | .................... | .................... |
| 30997 | MT451229.1 | \|Severe acute respiratory syndrome coro | .................... | .................... | ..................... | .................... | .................... | .................... |
| 30998 | MT451230.1 | \|Severe acute respiratory syndrome coro | .................... | .................... | ..................... | .................... | .................... | .................... |
| 30999 | MT451231.1 | \|Severe acute respiratory syndrome coro | .................... | .................... | ..................... | .................... | .................... | .................... |
| 31000 | MT451232.1 | \|Severe acute respiratory syndrome coro | .................... | .A.................. | ..................... | .................... | .................... | .................... |
| 31001 | MT451233.1 | \|Severe acute respiratory syndrome coro | .................... | .................... | ..................... | .................... | .................... | .................... |
| 31002 | MT451235.1 | \|Severe acute respiratory syndrome coro | .................... | .................... | ..................... | .................... | .................... | .................... |
| 31003 | MT451236.1 | \|Severe acute respiratory syndrome coro | .................... | .................... | ..................... | .................... | .................... | .................... |
| 31004 | MT451237.1 | \|Severe acute respiratory syndrome coro | .................... | .................... | ..................... | .................... | .................... | .................... |
| 31005 | MT451238.1 | \|Severe acute respiratory syndrome coro | .................... | .................... | ..................... | .................... | .................... | .................... |
| 31006 | MT451239.1 | \|Severe acute respiratory syndrome coro | .................... | .................... | ..................... | .................... | .................... | .................... |
| 31007 | MT451240.1 | \|Severe acute respiratory syndrome coro | .................... | .................... | ..................... | .................... | .................... | .................... |
| 31008 | MT451241.1 | \|Severe acute respiratory syndrome coro | .................... | .................... | ..................... | .................... | .................... | .................... |
| 31009 | MT451242.1 | \|Severe acute respiratory syndrome coro | .................... | .................... | ..................... | .................... | .................... | .................... |
| 31010 | MT451244.1 | \|Severe acute respiratory syndrome coro | .................... | .................... | ..................... | .................... | .................... | .................... |
| 31011 | MT451245.1 | \|Severe acute respiratory syndrome coro | .................... | .................... | ..................... | .................... | .................... | .................... |
| 31012 | MT451246.1 | \|Severe acute respiratory syndrome coro | .................... | .................... | ..................... | .................... | .................... | .................... |
| 31013 | MT451247.1 | \|Severe acute respiratory syndrome coro | .................... | .................... | ..................... | .................... | .................... | .................... |
| 31014 | MT451248.1 | \|Severe acute respiratory syndrome coro | .................... | .................... | ..................... | .................... | .................... | .................... |
| 31015 | MT451253.1 | \|Severe acute respiratory syndrome coro | .................... | .................... | ..................... | .................... | .................... | .................... |
| 31016 | MT451255.1 | \|Severe acute respiratory syndrome coro | .................... | .................... | ..................... | .................... | .................... | .................... |
| 31017 | MT451256.1 | \|Severe acute respiratory syndrome coro | .................... | .................... | ..................... | .................... | .................... | .................... |
| 31018 | MT451257.1 | \|Severe acute respiratory syndrome coro | .................... | .................... | ..................... | .................... | .................... | .................... |
| 31019 | MT451258.1 | \|Severe acute respiratory syndrome coro | .................... | .................... | ..................... | .................... | .................... | .................... |
| 31020 | MT451259.1 | \|Severe acute respiratory syndrome coro | .................... | .................... | ..K.................. | .................... | .................... | .................... |
| 31021 | MT451261.1 | \|Severe acute respiratory syndrome coro | .................... | .................... | ..................... | .................... | .................... | .................... |
| 31022 | MT451263.1 | \|Severe acute respiratory syndrome coro | .................... | .................... | ..................... | .................... | .................... | .................... |
| 31023 | MT451264.1 | \|Severe acute respiratory syndrome coro | .................... | .................... | ..................... | .................... | .................... | .................... |
| 31024 | MT451265.1 | \|Severe acute respiratory syndrome coro | .................... | .................... | ..................... | .................... | .................... | .................... |
| 31025 | MT451269.1 | \|Severe acute respiratory syndrome coro | .................... | .................... | ..K.................. | .................... | .................... | .................... |
| 31026 | MT451272.1 | \|Severe acute respiratory syndrome coro | .................... | .................... | ..................... | .................... | .................... | .................... |
| 31027 | MT451274.1 | \|Severe acute respiratory syndrome coro | .................... | .................... | ..................... | .................... | .................... | .................... |
| 31028 | MT451276.1 | \|Severe acute respiratory syndrome coro | .................... | .................... | ..K.................. | .................... | .................... | .................... |
| 31029 | MT451278.1 | \|Severe acute respiratory syndrome coro | .................... | .................... | ..................... | .................... | .................... | .................... |
| 31030 | MT451279.1 | \|Severe acute respiratory syndrome coro | .................... | .................... | ..K.................. | .................... | .................... | .................... |
| 31031 | MT451281.1 | \|Severe acute respiratory syndrome coro | .................... | .................... | ..................... | .................... | .................... | .................... |
| 31032 | MT451283.1 | \|Severe acute respiratory syndrome coro | .................... | .................... | ..N.................. | .................... | .................... | .................... |
| 31033 | MT451284.1 | \|Severe acute respiratory syndrome coro | .................... | .................... | ..................... | .................... | .................... | .................... |
| 31035 | MT451354.1 | \|Severe acute respiratory syndrome coro | .................... | .................... | ..................... | .................... | .................... | .................... |
| 31036 | MT451355.1 | \|Severe acute respiratory syndrome coro | .................... | .................... | ..................... | .................... | .................... | .................... |
| 31040 | MT451371.1 | \|Severe acute respiratory syndrome coro | .................... | .................... | ..................... | .................... | .................... | .................... |
| 31041 | MT451375.1 | \|Severe acute respiratory syndrome coro | .................... | .................... | ..................... | .................... | .................... | .................... |
| 31046 | MT451392.1 | \|Severe acute respiratory syndrome coro | .................... | .................... | ..................... | .................... | .................... | .................... |
| 31047 | MT451410.1 | \|Severe acute respiratory syndrome coro | .................... | .................... | ..................... | .................... | .................... | .................... |
| 31052 | MT451436.1 | \|Severe acute respiratory syndrome coro | .................... | .................... | ..K.................. | .................... | .................... | .................... |
| 31054 | MT451441.1 | \|Severe acute respiratory syndrome coro | .................... | .................... | ..................... | .................... | .................... | .................... |
| 31055 | MT451442.1 | \|Severe acute respiratory syndrome coro | .................... | .................... | ..................... | .................... | .................... | .................... |
| 31056 | MT451447.1 | \|Severe acute respiratory syndrome coro | .................... | .................... | ..................... | .................... | .................... | .................... |

| | | | | | | | | | | |
|---|---|---|---|---|---|---|---|---|---|---|
| 31057 | MT451448.1 | \|Severe acute respiratory syndrome coro | ···················· | ···················· | ···················· | ···················· | ···················· | ···················· | | |
| 31058 | MT451449.1 | \|Severe acute respiratory syndrome coro | ···················· | ···················· | ···················· | ···················· | ···················· | ···················· | | |
| 31060 | MT451456.1 | \|Severe acute respiratory syndrome coro | ···················· | ···················· | ···················· | ···················· | ···················· | ···················· | | |
| 31061 | MT451464.1 | \|Severe acute respiratory syndrome coro | ···················· | ···················· | ···················· | ···················· | ···················· | ···················· | | |
| 31063 | MT451470.1 | \|Severe acute respiratory syndrome coro | ···················· | ···················· | ···················· | ···················· | ···················· | ···················· | | |
| 31069 | MT451569.1 | \|Severe acute respiratory syndrome coro | ···················· | ···················· | ···················· | ···················· | ···················· | ···················· | | |
| 31071 | MT451580.1 | \|Severe acute respiratory syndrome coro | ···················· | ···················· | ···················· | ···················· | ···················· | ···················· | | |
| 31072 | MT451581.1 | \|Severe acute respiratory syndrome coro | ···················· | ···················· | ···················· | ···················· | ···················· | ···················· | | |
| 31073 | MT451582.1 | \|Severe acute respiratory syndrome coro | ···················· | ···················· | ···················· | ···················· | ···················· | ···················· | | |
| 31075 | MT451586.1 | \|Severe acute respiratory syndrome coro | ···················· | ···················· | ···················· | ···················· | ···················· | ·····M···W·······M·· | 1 | 1 |
| 31076 | MT451590.1 | \|Severe acute respiratory syndrome coro | ···················· | ···················· | ···················· | ···················· | ···················· | ···················· | | |
| 31077 | MT451592.1 | \|Severe acute respiratory syndrome coro | ···················· | ···················· | ···················· | ···················· | ···················· | ···················· | | |
| 31078 | MT451597.1 | \|Severe acute respiratory syndrome coro | ···················· | ···················· | ···················· | ···················· | ···················· | ···················· | | |
| 31081 | MT451602.1 | \|Severe acute respiratory syndrome coro | ···················· | ···················· | ···················· | ···················· | ···················· | ···················· | | |
| 31082 | MT451603.1 | \|Severe acute respiratory syndrome coro | ···················· | ···················· | ···················· | ···················· | ···················· | ···················· | | |
| 31084 | MT451607.1 | \|Severe acute respiratory syndrome coro | ···················· | ···················· | ···················· | ···················· | ···················· | ···················· | | |
| 31085 | MT451608.1 | \|Severe acute respiratory syndrome coro | ···················· | ···················· | ···················· | ···················· | ···················· | ···················· | | |
| 31087 | MT451611.1 | \|Severe acute respiratory syndrome coro | ···················· | ···················· | ···················· | ···················· | ···················· | ···················· | | |
| 31090 | MT451641.1 | \|Severe acute respiratory syndrome coro | ···················· | ···················· | ···················· | ···················· | ···················· | ···················· | | |
| 31091 | MT451646.1 | \|Severe acute respiratory syndrome coro | ···················· | ···················· | ···················· | ···················· | ···················· | NN·················· | | |
| 31094 | MT451655.1 | \|Severe acute respiratory syndrome coro | ···················· | ···················· | ···················· | ···················· | ···················· | ···················· | | |
| 31096 | MT451666.1 | \|Severe acute respiratory syndrome coro | ···················· | ···················· | ···················· | ···················· | ···················· | ···················· | | |
| 31097 | MT451667.1 | \|Severe acute respiratory syndrome coro | ···················· | ···················· | ···················· | ···················· | ···················· | ···················· | | |
| 31098 | MT451668.1 | \|Severe acute respiratory syndrome coro | ···················· | ···················· | ···················· | ···················· | ···················· | ···················· | | |
| 31099 | MT451674.1 | \|Severe acute respiratory syndrome coro | ···················· | ···················· | ···················· | ···················· | ···················· | ···················· | | |
| 31100 | MT451675.1 | \|Severe acute respiratory syndrome coro | ···················· | ···················· | ···················· | ···················· | ···················· | ···················· | | |
| 31107 | MT451708.1 | \|Severe acute respiratory syndrome coro | ···················· | ···················· | ···················· | ···················· | ···················· | ···················· | | |
| 31109 | MT451711.1 | \|Severe acute respiratory syndrome coro | ···················· | ···················· | ···················· | ···················· | ···················· | ···················· | | |
| 31110 | MT451712.1 | \|Severe acute respiratory syndrome coro | ···················· | ···················· | ···················· | ···················· | ···················· | ···················· | | |
| 31111 | MT451713.1 | \|Severe acute respiratory syndrome coro | ···················· | ···················· | ···················· | ···················· | ···················· | ···················· | | |
| 31112 | MT451716.1 | \|Severe acute respiratory syndrome coro | ···················· | ···················· | ···················· | ···················· | ···················· | ···················· | | |
| 31115 | MT451726.1 | \|Severe acute respiratory syndrome coro | ···················· | ···················· | ···················· | ···················· | ···················· | ···················· | | |
| 31116 | MT451728.1 | \|Severe acute respiratory syndrome coro | ···················· | ···················· | ···················· | ···················· | ···················· | ···················· | | |
| 31118 | MT451733.1 | \|Severe acute respiratory syndrome coro | ···················· | ···················· | ···················· | ···················· | ···················· | ···················· | | |
| 31126 | MT451751.1 | \|Severe acute respiratory syndrome coro | ···················· | ···················· | ···················· | ···················· | ···················· | ···················· | | |
| 31127 | MT451754.1 | \|Severe acute respiratory syndrome coro | ···················· | ···················· | ···················· | ···················· | ···················· | ···················· | | |
| 31129 | MT451762.1 | \|Severe acute respiratory syndrome coro | ···················· | ···················· | ···················· | ···················· | ···················· | ···················· | | |
| 31131 | MT451775.1 | \|Severe acute respiratory syndrome coro | ···················· | ···················· | ···················· | ···················· | ···················· | ···················· | | |
| 31133 | MT451777.1 | \|Severe acute respiratory syndrome coro | ···················· | ···················· | ···················· | ···················· | ···················· | ···················· | | |
| 31136 | MT451786.1 | \|Severe acute respiratory syndrome coro | ···················· | ···················· | ···················· | ···················· | ···················· | ······R············· | | |
| 31137 | MT451874.1 | \|Severe acute respiratory syndrome coro | ···················· | ···················· | ···················· | ···················· | ···················· | ···················· | | |
| 31138 | MT451876.1 | \|Severe acute respiratory syndrome coro | ···················· | ···················· | ···················· | ···················· | ···················· | ···················· | | |
| 31139 | MT451877.1 | \|Severe acute respiratory syndrome coro | ···················· | ···················· | ···················· | ···················· | ···················· | ···················· | | |
| 31140 | MT451878.1 | \|Severe acute respiratory syndrome coro | ···················· | ···················· | ···················· | ···················· | ···················· | ···················· | | |
| 31141 | MT451880.1 | \|Severe acute respiratory syndrome coro | ···················· | ···················· | ···················· | ···················· | ···················· | ···················· | | |
| 31142 | MT451881.1 | \|Severe acute respiratory syndrome coro | ···················· | ·A·················· | ···················· | ···················· | ···················· | ··A················· | | |
| 31143 | MT451882.1 | \|Severe acute respiratory syndrome coro | ···················· | ·A·················· | ···················· | ···················· | ···················· | ··A················· | | |
| 31144 | MT451883.1 | \|Severe acute respiratory syndrome coro | ···················· | ···················· | ···················· | ···················· | ···················· | ···················· | | |
| 31145 | MT451884.1 | \|Severe acute respiratory syndrome coro | ···················· | ···················· | ···················· | ···················· | ···················· | ···················· | | |
| 31146 | MT451885.1 | \|Severe acute respiratory syndrome coro | ···················· | ···················· | ···················· | ···················· | ···················· | ···················· | | |
| 31147 | MT451886.1 | \|Severe acute respiratory syndrome coro | ···················· | ···················· | ···················· | ···················· | ···················· | ···················· | | |
| 31148 | MT451887.1 | \|Severe acute respiratory syndrome coro | ···················· | ···················· | ···················· | ···················· | ···················· | ···················· | | |
| 31149 | MT451888.1 | \|Severe acute respiratory syndrome coro | ···················· | ·A·················· | ···················· | ···················· | ···················· | ··A················· | | |
| 31150 | MT451889.1 | \|Severe acute respiratory syndrome coro | ···················· | ···················· | ···················· | ···················· | ···················· | ···················· | | |
| 31151 | MT451890.1 | \|Severe acute respiratory syndrome coro | ···················· | ···················· | ···················· | ···················· | ···················· | ··A················· | | |
| 31152 | MT452574.1 | \|Severe acute respiratory syndrome coro | ···················· | ···················· | ···················· | ···················· | ···················· | ···················· | | |
| 31153 | MT452575.1 | \|Severe acute respiratory syndrome coro | ···················· | ···················· | ···················· | ···················· | ···················· | ···················· | | |
| 31154 | MT452576.1 | \|Severe acute respiratory syndrome coro | ···················· | ···················· | ···················· | ···················· | ···················· | ···················· | | |
| 31155 | MT444148.1 | \|Severe acute respiratory syndrome coro | ···················· | ···················· | ···················· | ···················· | ···················· | ···················· | | |
| 31156 | MT444540.1 | \|Severe acute respiratory syndrome coro | ···················· | ···················· | ···················· | ···················· | ···················· | ···················· | | |
| 31157 | MT444619.1 | \|Severe acute respiratory syndrome coro | ···················· | ···················· | ···················· | ···················· | ···················· | ···················· | | |
| 31158 | MT444624.1 | \|Severe acute respiratory syndrome coro | ···················· | ···················· | ···················· | ···················· | ···················· | ···················· | | |
| 31159 | MT444625.1 | \|Severe acute respiratory syndrome coro | ···················· | ···················· | ···················· | ···················· | ···················· | ···················· | | |
| 31160 | MT444626.1 | \|Severe acute respiratory syndrome coro | ···················· | ···················· | ···················· | ···················· | ···················· | ···················· | | |
| 31161 | MT444633.1 | \|Severe acute respiratory syndrome coro | ···················· | ···················· | ···················· | ···················· | ···················· | ···················· | | |
| 31162 | MT444635.1 | \|Severe acute respiratory syndrome coro | ···················· | ···················· | ···················· | ···················· | ···················· | ···················· | | |
| 31163 | MT446312.1 | \|Severe acute respiratory syndrome coro | ···················· | ···················· | ···················· | ···················· | ···················· | ···················· | | |
| 31164 | MT446339.1 | \|Severe acute respiratory syndrome coro | ···················· | ···················· | ···················· | ···················· | ···················· | ···················· | | |
| 31165 | MT446342.1 | \|Severe acute respiratory syndrome coro | ···················· | ···················· | ···················· | ···················· | ···················· | ···················· | | |
| 31166 | MT446343.1 | \|Severe acute respiratory syndrome coro | ···················· | ···················· | ···················· | ···················· | ···················· | ···················· | | |
| 31167 | MT446344.1 | \|Severe acute respiratory syndrome coro | ···················· | ···················· | ···················· | ···················· | ···················· | ···················· | | |
| 31168 | MT446345.1 | \|Severe acute respiratory syndrome coro | ···················· | ···················· | ···················· | ···················· | ···················· | ···················· | | |
| 31169 | MT446346.1 | \|Severe acute respiratory syndrome coro | ···················· | ···················· | ···················· | ···················· | ···················· | ···················· | | |
| 31170 | MT446347.1 | \|Severe acute respiratory syndrome coro | ···················· | ···················· | ···················· | ···················· | ···················· | ···················· | | |
| 31171 | MT446348.1 | \|Severe acute respiratory syndrome coro | ···················· | ···················· | ···················· | ···················· | ···················· | ···················· | | |
| 31172 | MT446349.1 | \|Severe acute respiratory syndrome coro | ···················· | ···················· | ···················· | ···················· | ···················· | ···················· | | |
| 31173 | MT446350.1 | \|Severe acute respiratory syndrome coro | ···················· | ···················· | ···················· | ···················· | ···················· | ···················· | | |
| 31174 | MT446352.1 | \|Severe acute respiratory syndrome coro | ···················· | ···················· | ···················· | ···················· | ···················· | ···················· | | |

```
31175 MT446353.1 |Severe acute respiratory syndrome coro    ....................  ....................  .....................  ....................  ....................  ....................
31176 MT446354.1 |Severe acute respiratory syndrome coro    ....................  ....................  .....................  ....................  ....................  ....................
31177 MT446356.1 |Severe acute respiratory syndrome coro    ....................  ....................  .....................  ....................  ....................  ....................
31178 MT446357.1 |Severe acute respiratory syndrome coro    ....................  ....................  .....................  ....................  ....................  ....................
31179 MT446358.1 |Severe acute respiratory syndrome coro    ....................  ....................  .....................  ....................  ....................  ....................
31180 MT446359.1 |Severe acute respiratory syndrome coro    ....................  ....................  .....................  ....................  ....................  ....................
31181 MT447154.1 |Severe acute respiratory syndrome coro    ....................  ....................  .....................  ....................  ....................  ....................
31182 MT447155.1 |Severe acute respiratory syndrome coro    ....................  ....................  .....................  ....................  ....................  ....................
31183 MT447156.1 |Severe acute respiratory syndrome coro    ....................  ....................  .....................  ....................  ....................  ....................
31184 MT447157.1 |Severe acute respiratory syndrome coro    ....................  ....................  .....................  ....................  ....................  ....................
31185 MT447158.1 |Severe acute respiratory syndrome coro    ....................  ....................  .....................  ....................  ....................  ....................
31186 MT447159.1 |Severe acute respiratory syndrome coro    ....................  ....................  .....................  ....................  ....................  ....................
31187 MT447161.1 |Severe acute respiratory syndrome coro    ....................  ....................  .....................  ....................  ....................  ....................
31188 MT447162.1 |Severe acute respiratory syndrome coro    ....................  ....................  .....................  ....................  ....................  ....................
31189 MT447163.1 |Severe acute respiratory syndrome coro    ....................  ....................  .....................  ....................  ....................  ....................
31190 MT447165.1 |Severe acute respiratory syndrome coro    ....................  ....................  .....................  ....................  ....................  ....................
31191 MT447166.1 |Severe acute respiratory syndrome coro    ....................  ....................  .....................  ....................  ....................  ....................
31192 MT447169.1 |Severe acute respiratory syndrome coro    ....................  ....................  .....................  ....................  ....................  ....................
31193 MT447171.1 |Severe acute respiratory syndrome coro    ....................  ....................  .....................  ....................  ....................  ....................
31194 MT447172.1 |Severe acute respiratory syndrome coro    ....................  ....................  .....................  ....................  ....................  ....................
31195 MT447173.1 |Severe acute respiratory syndrome coro    ....................  ....................  .....................  ....................  ....................  ....................
31196 MT447174.1 |Severe acute respiratory syndrome coro    ....................  ....................  .....................  ....................  ....................  ....................
31197 MT447175.1 |Severe acute respiratory syndrome coro    ....................  ....................  .....................  ....................  ....................  ....................
31198 MT447176.1 |Severe acute respiratory syndrome coro    ....................  ....................  .....................  ....................  ....................  ....................
31199 MT447177.1 |Severe acute respiratory syndrome coro    ....................  ....................  .....................  ....................  ....................  ....................
31200 MT438718.1 |Severe acute respiratory syndrome coro    ....................  ....................  .....................  ....................  ....................  ....................
31201 MT438719.1 |Severe acute respiratory syndrome coro    ....................  ....................  .....................  ....................  ....................  ....................
31202 MT438720.1 |Severe acute respiratory syndrome coro    ....................  ....................  .....................  ....................  ....................  ....................
31203 MT438721.1 |Severe acute respiratory syndrome coro    ....................  ....................  .....................  ....................  ....................  ....................
31204 MT438722.1 |Severe acute respiratory syndrome coro    ....................  ....................  .....................  ....................  ....................  ....................
31205 MT438723.1 |Severe acute respiratory syndrome coro    ....................  ....................  .....................  ....................  ....................  ....................
31206 MT438724.1 |Severe acute respiratory syndrome coro    ....................  ....................  .....................  ....................  ....................  ....................
31207 MT438725.1 |Severe acute respiratory syndrome coro    ....................  ....................  .....................  ....................  ....................  ....................
31208 MT438727.1 |Severe acute respiratory syndrome coro    ....................  ....................  .....................  ....................  ....................  ....................
31209 MT438728.1 |Severe acute respiratory syndrome coro    ....................  ....................  .....................  ....................  ....................  ....................
31210 MT438729.1 |Severe acute respiratory syndrome coro    ....................  ....................  .....................  ....................  ....................  ....................
31211 MT438730.1 |Severe acute respiratory syndrome coro    ....................  ....................  .....................  ....................  ....................  ....................
31212 MT438731.1 |Severe acute respiratory syndrome coro    ....................  ....................  .....................  ....................  ....................  ....................
31213 MT438732.1 |Severe acute respiratory syndrome coro    ....................  ....................  .....................  ....................  ....................  ....................
31214 MT438734.1 |Severe acute respiratory syndrome coro    ....................  ....................  .....................  ....................  ....................  ....................
31215 MT438735.1 |Severe acute respiratory syndrome coro    ....................  ....................  .....................  ....................  ....................  ....................
31216 MT438736.1 |Severe acute respiratory syndrome coro    ....................  ....................  .....................  ....................  ....................  ....................
31217 MT438737.1 |Severe acute respiratory syndrome coro    ....................  ....................  .....................  ....................  ....................  ....................
31218 MT438738.1 |Severe acute respiratory syndrome coro    ....................  ....................  .....................  ....................  ....................  ....................
31219 MT438739.1 |Severe acute respiratory syndrome coro    ....................  ....................  .....................  ....................  ....................  ....................
31220 MT438740.1 |Severe acute respiratory syndrome coro    ....................  ....................  .....................  ....................  ....................  ....................
31221 MT438741.1 |Severe acute respiratory syndrome coro    ....................  ....................  .....................  ....................  ....................  ....................
31222 MT438742.1 |Severe acute respiratory syndrome coro    ....................  ....................  .....................  ....................  ....................  ....................
31223 MT438743.1 |Severe acute respiratory syndrome coro    ....................  ....................  .....................  ....................  ....................  ....................
31224 MT438744.1 |Severe acute respiratory syndrome coro    ....................  ....................  .....................  ....................  ....................  ....................
31225 MT438746.1 |Severe acute respiratory syndrome coro    ....................  ....................  .....................  ....................  ....................  ....................
31226 MT438747.1 |Severe acute respiratory syndrome coro    ....................  ....................  .....................  ....................  ....................  ....................
31227 MT438748.1 |Severe acute respiratory syndrome coro    ....................  ....................  .....................  ....................  ....................  ....................
31228 MT438749.1 |Severe acute respiratory syndrome coro    ....................  ....................  .....................  ....................  ....................  ....................
31229 MT438750.1 |Severe acute respiratory syndrome coro    ....................  ....................  .....................  ....................  ....................  ....................
31230 MT438751.1 |Severe acute respiratory syndrome coro    ....................  ....................  .....................  ....................  ....................  ....................
31231 MT438752.1 |Severe acute respiratory syndrome coro    ....................  ....................  .....................  ....................  ....................  ....................
31232 MT438753.1 |Severe acute respiratory syndrome coro    ....................  ....................  .....................  ....................  ....................  ....................
31233 MT438754.1 |Severe acute respiratory syndrome coro    ....................  ....................  .....................  ....................  ....................  ....................
31234 MT438755.1 |Severe acute respiratory syndrome coro    ....................  ....................  .....................  ....................  ....................  ....................
31235 MT438756.1 |Severe acute respiratory syndrome coro    ....................  ....................  .....................  ....................  ....................  ....................
31236 MT438757.1 |Severe acute respiratory syndrome coro    ....................  ....................  .....................  ....................  ....................  ....................
31237 MT438758.1 |Severe acute respiratory syndrome coro    ....................  ....................  .....................  ....................  ....................  ....................
31238 MT438759.1 |Severe acute respiratory syndrome coro    ....................  ....................  .....................  ....................  ....................  ....................
31239 MT438760.1 |Severe acute respiratory syndrome coro    ....................  ....................  .....................  ....................  ....................  ....................
31240 MT439233.1 |Severe acute respiratory syndrome coro    ....................  ....................  .....................  ....................  ....................  ....................
31241 MT439234.1 |Severe acute respiratory syndrome coro    ....................  ....................  .....................  ....................  ....................  ....................
31242 MT439237.1 |Severe acute respiratory syndrome coro    ....................  ....................  .....................  ....................  ....................  ....................
31243 MT439239.1 |Severe acute respiratory syndrome coro    ....................  ....................  .....................  ....................  ....................  ....................
31244 MT439242.1 |Severe acute respiratory syndrome coro    ....................  ....................  .....................  ....................  ....................  ....................
31245 MT439244.1 |Severe acute respiratory syndrome coro    ....................  ....................  .....................  ....................  ....................  ....................
31246 MT439245.1 |Severe acute respiratory syndrome coro    ....................  ....................  .....................  ....................  ....................  ....................
31247 MT439247.1 |Severe acute respiratory syndrome coro    ....................  ....................  .....................  ....................  ....................  ....................
31248 MT439249.1 |Severe acute respiratory syndrome coro    ....................  ....................  .....................  ....................  ....................  ....................
31249 MT439250.1 |Severe acute respiratory syndrome coro    ....................  ....................  .....................  ....................  ....................  ....................
31250 MT439251.1 |Severe acute respiratory syndrome coro    ....................  ....................  .....................  ....................  ....................  ....................
31251 MT439252.1 |Severe acute respiratory syndrome coro    ....................  ....................  .....................  ....................  ....................  ....................
31252 MT439253.1 |Severe acute respiratory syndrome coro    ....................  ....................  .....................  ....................  ....................  ....................
```

| | | | | | | | |
|---|---|---|---|---|---|---|---|
| 31253 | MT439254.1 \|Severe acute respiratory syndrome coro | .................... | .................... | ..................... | .................... | .................... | .................... |
| 31254 | MT439256.1 \|Severe acute respiratory syndrome coro | .................... | .................... | ..................... | .................... | .................... | .................... |
| 31255 | MT439258.1 \|Severe acute respiratory syndrome coro | .................... | .................... | ..................... | .................... | .................... | .................... |
| 31256 | MT439260.1 \|Severe acute respiratory syndrome coro | .................... | .................... | ..................... | .................... | .................... | .................... |
| 31257 | MT439261.1 \|Severe acute respiratory syndrome coro | .................... | .................... | ..................... | .................... | .................... | .................... |
| 31258 | MT439262.1 \|Severe acute respiratory syndrome coro | .................... | .................... | ..................... | .................... | .................... | .................... |
| 31259 | MT439263.1 \|Severe acute respiratory syndrome coro | .................... | .................... | ..................... | .................... | .................... | .................... |
| 31260 | MT439265.1 \|Severe acute respiratory syndrome coro | .................... | .................... | ..................... | .................... | .................... | .................... |
| 31261 | MT439267.1 \|Severe acute respiratory syndrome coro | .................... | .................... | ..................... | .................... | .................... | .................... |
| 31262 | MT439268.1 \|Severe acute respiratory syndrome coro | .................... | .................... | ..................... | .................... | .................... | .................... |
| 31263 | MT439270.1 \|Severe acute respiratory syndrome coro | .................... | .................... | ..................... | .................... | .................... | .................... |
| 31264 | MT439271.1 \|Severe acute respiratory syndrome coro | .................... | .................... | ..................... | .................... | .................... | .................... |
| 31265 | MT439273.1 \|Severe acute respiratory syndrome coro | .................... | .................... | ..................... | .................... | .................... | .................... |
| 31266 | MT439279.1 \|Severe acute respiratory syndrome coro | .................... | .................... | ..................... | .................... | .................... | .................... |
| 31267 | MT439280.1 \|Severe acute respiratory syndrome coro | .................... | .................... | ..................... | .................... | .................... | .................... |
| 31268 | MT439282.1 \|Severe acute respiratory syndrome coro | .................... | .................... | ..................... | .................... | .................... | .................... |
| 31269 | MT439283.1 \|Severe acute respiratory syndrome coro | .................... | .................... | ..................... | .................... | .................... | .................... |
| 31270 | MT439284.1 \|Severe acute respiratory syndrome coro | .................... | .................... | ..................... | .................... | .................... | .................... |
| 31271 | MT439287.1 \|Severe acute respiratory syndrome coro | .................... | .................... | ..................... | .................... | .................... | .................... |
| 31272 | MT439288.1 \|Severe acute respiratory syndrome coro | .................... | .................... | ..................... | .................... | .................... | .................... |
| 31273 | MT439289.1 \|Severe acute respiratory syndrome coro | .................... | .................... | ..................... | .................... | .................... | .................... |
| 31274 | MT439290.1 \|Severe acute respiratory syndrome coro | .................... | .................... | ..................... | .................... | .................... | .................... |
| 31275 | MT439291.1 \|Severe acute respiratory syndrome coro | .................... | .................... | ..................... | .................... | .................... | .................... |
| 31276 | MT439292.1 \|Severe acute respiratory syndrome coro | .................... | .................... | ..................... | .................... | .................... | .................... |
| 31277 | MT439293.1 \|Severe acute respiratory syndrome coro | .................... | .................... | ..................... | .................... | .................... | .................... |
| 31278 | MT439295.1 \|Severe acute respiratory syndrome coro | .................... | .................... | ..................... | .................... | .................... | .................... |
| 31279 | MT439296.1 \|Severe acute respiratory syndrome coro | .................... | .................... | ..................... | .................... | .................... | .................... |
| 31280 | MT439297.1 \|Severe acute respiratory syndrome coro | .................... | .................... | ..................... | .................... | .................... | .................... |
| 31281 | MT439300.1 \|Severe acute respiratory syndrome coro | .................... | .................... | ..................... | .................... | .................... | .................... |
| 31282 | MT439302.1 \|Severe acute respiratory syndrome coro | .................... | .................... | ..................... | .................... | .................... | .................... |
| 31283 | MT439303.1 \|Severe acute respiratory syndrome coro | .................... | .................... | ..................... | .................... | .................... | .................... |
| 31284 | MT439307.1 \|Severe acute respiratory syndrome coro | .................... | .................... | ..................... | .................... | .................... | .................... |
| 31285 | MT439310.1 \|Severe acute respiratory syndrome coro | .................... | .................... | ..................... | .................... | .................... | .................... |
| 31286 | MT439311.1 \|Severe acute respiratory syndrome coro | .................... | .................... | ..................... | .................... | .................... | .................... |
| 31287 | MT439312.1 \|Severe acute respiratory syndrome coro | .................... | .................... | ..................... | .................... | .................... | .................... |
| 31288 | MT439314.1 \|Severe acute respiratory syndrome coro | .................... | .................... | ..................... | .................... | .................... | .................... |
| 31289 | MT439315.1 \|Severe acute respiratory syndrome coro | .................... | .................... | ..................... | .................... | .................... | .................... |
| 31290 | MT439316.1 \|Severe acute respiratory syndrome coro | .................... | .................... | ..................... | .................... | .................... | .................... |
| 31291 | MT439317.1 \|Severe acute respiratory syndrome coro | .................... | .................... | ..................... | .................... | .................... | .................... |
| 31292 | MT439318.1 \|Severe acute respiratory syndrome coro | .................... | .................... | ..................... | .................... | .................... | .................... |
| 31293 | MT434782.1 \|Severe acute respiratory syndrome coro | .................... | .................... | ..................... | .................... | .................... | .................... |
| 31294 | MT434783.1 \|Severe acute respiratory syndrome coro | .................... | .................... | ..................... | .................... | .................... | .................... |
| 31295 | MT434784.1 \|Severe acute respiratory syndrome coro | .................... | .................... | ..................... | .................... | .................... | .................... |
| 31296 | MT434785.1 \|Severe acute respiratory syndrome coro | .................... | .................... | ..................... | .................... | .................... | .................... |
| 31297 | MT434786.1 \|Severe acute respiratory syndrome coro | .................... | .................... | ..................... | .................... | .................... | .................... |
| 31298 | MT434787.1 \|Severe acute respiratory syndrome coro | .................... | .................... | ..................... | .................... | .................... | .................... |
| 31299 | MT434788.1 \|Severe acute respiratory syndrome coro | .................... | .................... | ..................... | .................... | .................... | .................... |
| 31300 | MT434789.1 \|Severe acute respiratory syndrome coro | .................... | .................... | ..................... | .................... | .................... | .................... |
| 31301 | MT434790.1 \|Severe acute respiratory syndrome coro | .................... | .................... | ..................... | .................... | .................... | .................... |
| 31302 | MT434791.1 \|Severe acute respiratory syndrome coro | .................... | .................... | ..................... | .................... | .................... | .................... |
| 31303 | MT434792.1 \|Severe acute respiratory syndrome coro | .................... | .................... | ..................... | .................... | .................... | .................... |
| 31304 | MT434793.1 \|Severe acute respiratory syndrome coro | .................... | .................... | ..................... | .................... | .................... | .................... |
| 31305 | MT434794.1 \|Severe acute respiratory syndrome coro | .................... | .................... | ..................... | .................... | .................... | .................... |
| 31306 | MT434795.1 \|Severe acute respiratory syndrome coro | .................... | .................... | ..................... | .................... | .................... | .................... |
| 31307 | MT434796.1 \|Severe acute respiratory syndrome coro | .................... | .................... | ..................... | .................... | .................... | .................... |
| 31308 | MT434797.1 \|Severe acute respiratory syndrome coro | .................... | .................... | ..................... | .................... | .................... | .................... |
| 31309 | MT434798.1 \|Severe acute respiratory syndrome coro | .................... | .................... | ..................... | .................... | .................... | .................... |
| 31310 | MT434799.1 \|Severe acute respiratory syndrome coro | .................... | .................... | ..................... | .................... | .................... | .................... |
| 31311 | MT434800.1 \|Severe acute respiratory syndrome coro | .................... | .................... | ..................... | .................... | .................... | .................... |
| 31312 | MT434801.1 \|Severe acute respiratory syndrome coro | .................... | .................... | ..................... | .................... | .................... | .................... |
| 31313 | MT434802.1 \|Severe acute respiratory syndrome coro | .................... | .................... | ..................... | .................... | .................... | .................... |
| 31314 | MT434803.1 \|Severe acute respiratory syndrome coro | .................... | .................... | ..................... | .................... | .................... | .................... |
| 31315 | MT434804.1 \|Severe acute respiratory syndrome coro | .................... | .................... | ..................... | .................... | .................... | .................... |
| 31316 | MT434805.1 \|Severe acute respiratory syndrome coro | .................... | .................... | ..................... | .................... | .................... | .................... |
| 31317 | MT434806.1 \|Severe acute respiratory syndrome coro | .................... | .................... | ..................... | .................... | .................... | .................... |
| 31318 | MT434807.1 \|Severe acute respiratory syndrome coro | .................... | .................... | ..................... | .................... | .................... | .................... |
| 31319 | MT434808.1 \|Severe acute respiratory syndrome coro | .................... | .................... | ..................... | .................... | .................... | .................... |
| 31320 | MT434809.1 \|Severe acute respiratory syndrome coro | .................... | .................... | ..................... | .................... | .................... | .................... |
| 31321 | MT434810.1 \|Severe acute respiratory syndrome coro | .................... | .................... | ..................... | .................... | .................... | .................... |
| 31322 | MT434811.1 \|Severe acute respiratory syndrome coro | .................... | .................... | ..................... | .................... | .................... | .................... |
| 31323 | MT434812.1 \|Severe acute respiratory syndrome coro | .................... | .................... | ..................... | .................... | .................... | .................... |
| 31324 | MT434813.1 \|Severe acute respiratory syndrome coro | .................... | .................... | ..................... | .................... | .................... | .................... |
| 31325 | MT434814.1 \|Severe acute respiratory syndrome coro | .................... | .................... | ..................... | .................... | .................... | .................... |
| 31326 | MT434815.1 \|Severe acute respiratory syndrome coro | .................... | .................... | ..................... | .................... | .................... | .................... |
| 31327 | MT434816.1 \|Severe acute respiratory syndrome coro | .................... | .................... | ..................... | .................... | .................... | .................... |
| 31328 | MT434817.1 \|Severe acute respiratory syndrome coro | .................... | .................... | ..................... | .................... | .................... | .................... |
| 31329 | MT435079.1 \|Severe acute respiratory syndrome coro | .................... | .................... | ..................... | .................... | .................... | .................... |
| 31330 | MT435080.1 \|Severe acute respiratory syndrome coro | .................... | .................... | ..................... | .................... | .................... | .................... |

| | | | | | | | | |
|---|---|---|---|---|---|---|---|---|
| 31331 | MT435081.1 | \|Severe acute respiratory syndrome coro | .................... | .................... | .................... | .................... | .................... | .................... |
| 31332 | MT435082.1 | \|Severe acute respiratory syndrome coro | .................... | .................... | .................... | .................... | .................... | .................... |
| 31333 | MT435083.1 | \|Severe acute respiratory syndrome coro | .................... | .................... | .................... | .................... | .................... | .................... |
| 31334 | MT435084.1 | \|Severe acute respiratory syndrome coro | .................... | .................... | .................... | .................... | .................... | .................... |
| 31335 | MT435085.1 | \|Severe acute respiratory syndrome coro | .................... | .A.................. | .................... | .................... | .................... | ..A................. |
| 31336 | MT435086.1 | \|Severe acute respiratory syndrome coro | .................... | .................... | .................... | .................... | .................... | .................... |
| 31337 | MT365028.1 | \|Severe acute respiratory syndrome coro | .................... | .................... | .................... | .................... | .................... | .................... |
| 31338 | MT365029.1 | \|Severe acute respiratory syndrome coro | .................... | .................... | .................... | .................... | .................... | .................... |
| 31339 | MT365030.1 | \|Severe acute respiratory syndrome coro | .................... | .A.................. | .................... | .................... | .................... | .................... |
| 31340 | MT365031.1 | \|Severe acute respiratory syndrome coro | .................... | .A.................. | .................... | .................... | .................... | .................... |
| 31341 | MT365032.1 | \|Severe acute respiratory syndrome coro | .................... | .................... | .................... | .................... | .................... | .................... |
| 31342 | MT428551.1 | \|Severe acute respiratory syndrome coro | .................... | .................... | .................... | .................... | .................... | .................... |
| 31343 | MT428552.1 | \|Severe acute respiratory syndrome coro | .................... | .................... | .................... | .................... | .................... | .................... |
| 31344 | MT428553.1 | \|Severe acute respiratory syndrome coro | .................... | .................... | .................... | .................... | .................... | .................... |
| 31345 | MT428554.1 | \|Severe acute respiratory syndrome coro | .................... | .................... | .................... | .................... | .................... | .................... |
| 31346 | MT429183.1 | \|Severe acute respiratory syndrome coro | .................... | .................... | .................... | .................... | .................... | .................... |
| 31347 | MT429184.1 | \|Severe acute respiratory syndrome coro | .................... | .................... | .................... | .................... | .................... | .................... |
| 31348 | MT429185.1 | \|Severe acute respiratory syndrome coro | .................... | .................... | .................... | .................... | .................... | .................... |
| 31349 | MT429186.1 | \|Severe acute respiratory syndrome coro | .................... | .................... | .................... | .................... | .................... | .................... |
| 31350 | MT429187.1 | \|Severe acute respiratory syndrome coro | .................... | .................... | .................... | .................... | .................... | .................... |
| 31351 | MT429188.1 | \|Severe acute respiratory syndrome coro | .................... | .................... | .................... | .................... | .................... | .................... |
| 31352 | MT429189.1 | \|Severe acute respiratory syndrome coro | .................... | .................... | .................... | .................... | .................... | .................... |
| 31353 | MT429190.1 | \|Severe acute respiratory syndrome coro | .................... | .................... | .................... | .................... | .................... | .................... |
| 31354 | MT429191.1 | \|Severe acute respiratory syndrome coro | .................... | .................... | .................... | .................... | .................... | .................... |
| 31355 | MT432195.1 | \|Severe acute respiratory syndrome coro | .................... | .................... | .................... | .................... | .................... | .................... |
| 31356 | MT318827.1 | \|Severe acute respiratory syndrome coro | .................... | .................... | .................... | .................... | .................... | .................... |
| 31357 | MT422806.1 | \|Severe acute respiratory syndrome coro | .................... | .................... | .................... | .................... | .................... | ...............A.... |
| 31358 | MT422807.1 | \|Severe acute respiratory syndrome coro | .................... | .................... | .................... | .................... | .................... | .................... |
| 31359 | MT215194.1 | \|Severe acute respiratory syndrome coro | .................... | .................... | .................... | .................... | .................... | .................... |
| 31360 | MT215195.1 | \|Severe acute respiratory syndrome coro | .................... | .................... | .................... | .................... | .................... | .................... |
| 31361 | MT270814.1 | \|Severe acute respiratory syndrome coro | .................... | .................... | .................... | .................... | .................... | .................... |
| 31362 | MT270815.1 | \|Severe acute respiratory syndrome coro | .................... | .................... | .................... | .................... | .................... | .................... |
| 31363 | MT276600.1 | \|Severe acute respiratory syndrome coro | .................... | .................... | .................... | .................... | .................... | .................... |
| 31364 | MT416725.2 | \|Severe acute respiratory syndrome coro | .................... | .................... | .................... | .................... | .................... | .................... |
| 31365 | MT416726.2 | \|Severe acute respiratory syndrome coro | .................... | .................... | .................... | .................... | .................... | .................... |
| 31366 | MT418880.1 | \|Severe acute respiratory syndrome coro | .................... | .................... | .................... | .................... | .................... | .................... |
| 31367 | MT418881.1 | \|Severe acute respiratory syndrome coro | .................... | .................... | .................... | .................... | .................... | .................... |
| 31368 | MT418882.1 | \|Severe acute respiratory syndrome coro | .................... | .................... | .................... | .................... | .................... | .................... |
| 31369 | MT418883.1 | \|Severe acute respiratory syndrome coro | .................... | .................... | .................... | .................... | .................... | .................... |
| 31370 | MT418884.1 | \|Severe acute respiratory syndrome coro | .................... | .................... | .................... | .................... | .................... | .................... |
| 31371 | MT418885.1 | \|Severe acute respiratory syndrome coro | .................... | .................... | .................... | .................... | .................... | .................... |
| 31372 | MT418886.1 | \|Severe acute respiratory syndrome coro | .................... | .................... | .................... | .................... | .................... | .................... |
| 31373 | MT418887.1 | \|Severe acute respiratory syndrome coro | .................... | .................... | .................... | .................... | .................... | .................... |
| 31374 | MT418888.1 | \|Severe acute respiratory syndrome coro | .................... | .................... | .................... | .................... | .................... | .................... |
| 31375 | MT418889.1 | \|Severe acute respiratory syndrome coro | .................... | .................... | .................... | .................... | .................... | .................... |
| 31376 | MT418890.1 | \|Severe acute respiratory syndrome coro | .................... | .................... | .................... | .................... | .................... | .................... |
| 31377 | MT418891.1 | \|Severe acute respiratory syndrome coro | .................... | .................... | .................... | .................... | .................... | .................... |
| 31378 | MT418892.1 | \|Severe acute respiratory syndrome coro | .................... | .................... | .................... | .................... | .................... | .................... |
| 31379 | MT419810.1 | \|Severe acute respiratory syndrome coro | .................... | .................... | .................... | .................... | .................... | .................... |
| 31380 | MT419812.1 | \|Severe acute respiratory syndrome coro | .................... | .................... | .................... | .................... | .................... | .................... |
| 31381 | MT419814.1 | \|Severe acute respiratory syndrome coro | .................... | .................... | .................... | .................... | .................... | .................... |
| 31382 | MT419815.1 | \|Severe acute respiratory syndrome coro | .................... | .................... | .................... | .................... | .................... | .................... |
| 31383 | MT419816.1 | \|Severe acute respiratory syndrome coro | .................... | .................... | .................... | .................... | .................... | .................... |
| 31384 | MT419818.1 | \|Severe acute respiratory syndrome coro | .................... | .................... | .................... | .................... | .................... | .................... |
| 31385 | MT419820.1 | \|Severe acute respiratory syndrome coro | .................... | .................... | .................... | .................... | .................... | .................... |
| 31386 | MT419829.1 | \|Severe acute respiratory syndrome coro | .................... | .................... | .................... | .................... | .................... | .................... |
| 31387 | MT419830.1 | \|Severe acute respiratory syndrome coro | .................... | .................... | .................... | .................... | .................... | .................... |
| 31388 | MT419831.1 | \|Severe acute respiratory syndrome coro | .................... | .................... | .................... | .................... | .................... | .................... |
| 31389 | MT419832.1 | \|Severe acute respiratory syndrome coro | .................... | .................... | .................... | .................... | .................... | .................... |
| 31390 | MT419834.1 | \|Severe acute respiratory syndrome coro | .................... | .................... | .................... | .................... | .................... | .................... |
| 31391 | MT419835.1 | \|Severe acute respiratory syndrome coro | .................... | .................... | .................... | .................... | .................... | .................... |
| 31392 | MT419836.1 | \|Severe acute respiratory syndrome coro | .................... | .................... | .................... | .................... | .................... | .................... |
| 31393 | MT419837.1 | \|Severe acute respiratory syndrome coro | .................... | .................... | .................... | .................... | .................... | .................... |
| 31394 | MT419839.1 | \|Severe acute respiratory syndrome coro | .................... | .................... | .................... | .................... | .................... | .................... |
| 31395 | MT419841.1 | \|Severe acute respiratory syndrome coro | .................... | .................... | .................... | .................... | .................... | .................... |
| 31396 | MT419842.1 | \|Severe acute respiratory syndrome coro | .................... | .................... | .................... | .................... | .................... | .................... |
| 31397 | MT419846.1 | \|Severe acute respiratory syndrome coro | .................... | .................... | .................... | .................... | .................... | .................... |
| 31398 | MT419847.1 | \|Severe acute respiratory syndrome coro | .................... | .................... | .................... | .................... | .................... | .................... |
| 31399 | MT419851.1 | \|Severe acute respiratory syndrome coro | .................... | .................... | .................... | .................... | .................... | .................... |
| 31400 | MT419853.1 | \|Severe acute respiratory syndrome coro | .................... | .................... | .................... | .................... | .................... | .................... |
| 31401 | MT419854.1 | \|Severe acute respiratory syndrome coro | .................... | .................... | .................... | .................... | .................... | .................... |
| 31402 | MT419860.1 | \|Severe acute respiratory syndrome coro | .................... | .................... | .................... | .................... | .................... | .................... |
| 31403 | MT412134.1 | \|Severe acute respiratory syndrome coro | .................... | .................... | .................... | .................... | .................... | .................... |
| 31404 | MT412155.1 | \|Severe acute respiratory syndrome coro | .................... | .................... | .................... | .................... | .................... | .................... |
| 31405 | MT412170.1 | \|Severe acute respiratory syndrome coro | .................... | .................... | .................... | .................... | .................... | .................... |
| 31406 | MT412173.1 | \|Severe acute respiratory syndrome coro | .................... | .................... | .................... | .................... | .................... | .................... |
| 31407 | MT412176.1 | \|Severe acute respiratory syndrome coro | .................... | .................... | .................... | .................... | .................... | .................... |
| 31408 | MT412177.1 | \|Severe acute respiratory syndrome coro | .................... | .................... | .................... | .................... | .................... | .................... |

| | | | | | | | | |
|---|---|---|---|---|---|---|---|---|
| 31409 | MT412178.1 | \|Severe acute respiratory syndrome coro | .................... | .................... | .................... | .................... | .................... | .................... |
| 31410 | MT412226.1 | \|Severe acute respiratory syndrome coro | .................... | .................... | .................... | .................... | .................... | .................... |
| 31411 | MT412227.1 | \|Severe acute respiratory syndrome coro | .................... | .................... | .................... | .................... | .................... | .................... |
| 31412 | MT412228.1 | \|Severe acute respiratory syndrome coro | .................... | .................... | .................... | .................... | .................... | .................... |
| 31413 | MT412229.1 | \|Severe acute respiratory syndrome coro | .................... | .................... | .................... | .................... | .................... | .................... |
| 31414 | MT412230.1 | \|Severe acute respiratory syndrome coro | .................... | .................... | .................... | .................... | .................... | .................... |
| 31415 | MT412233.1 | \|Severe acute respiratory syndrome coro | .................... | .................... | .................... | .................... | .................... | .................... |
| 31416 | MT412234.1 | \|Severe acute respiratory syndrome coro | .................... | .................... | .................... | .................... | .................... | .................... |
| 31417 | MT412235.1 | \|Severe acute respiratory syndrome coro | .................... | .................... | .................... | .................... | .................... | .................... |
| 31418 | MT412236.1 | \|Severe acute respiratory syndrome coro | .................... | .................... | .................... | .................... | .................... | .................... |
| 31419 | MT412237.1 | \|Severe acute respiratory syndrome coro | .................... | .................... | .................... | .................... | .................... | .................... |
| 31420 | MT412238.1 | \|Severe acute respiratory syndrome coro | .................... | .................... | .................... | .................... | .................... | .................... |
| 31421 | MT412239.1 | \|Severe acute respiratory syndrome coro | .................... | .................... | .................... | .................... | .................... | .................... |
| 31422 | MT412240.1 | \|Severe acute respiratory syndrome coro | .................... | .................... | .................... | .................... | .................... | .................... |
| 31423 | MT412241.1 | \|Severe acute respiratory syndrome coro | .................... | .................... | .................... | .................... | .................... | .................... |
| 31424 | MT412242.1 | \|Severe acute respiratory syndrome coro | .................... | .................... | .................... | .................... | .................... | .................... |
| 31425 | MT412243.1 | \|Severe acute respiratory syndrome coro | .................... | .................... | .................... | .................... | .................... | .................... |
| 31426 | MT412244.1 | \|Severe acute respiratory syndrome coro | .................... | .................... | .................... | .................... | .................... | .................... |
| 31427 | MT412245.1 | \|Severe acute respiratory syndrome coro | .................... | .................... | .................... | .................... | .................... | .................... |
| 31428 | MT412246.1 | \|Severe acute respiratory syndrome coro | .................... | .................... | .................... | .................... | .................... | .................... |
| 31429 | MT412247.1 | \|Severe acute respiratory syndrome coro | .................... | .................... | .................... | .................... | .................... | .................... |
| 31430 | MT412248.1 | \|Severe acute respiratory syndrome coro | .................... | .................... | .................... | .................... | .................... | .................... |
| 31431 | MT412249.1 | \|Severe acute respiratory syndrome coro | .................... | .................... | .................... | .................... | .................... | .................... |
| 31432 | MT412250.1 | \|Severe acute respiratory syndrome coro | .................... | .................... | .................... | .................... | .................... | .................... |
| 31433 | MT412251.1 | \|Severe acute respiratory syndrome coro | .................... | .................... | .................... | .................... | .................... | .................... |
| 31434 | MT412252.1 | \|Severe acute respiratory syndrome coro | .................... | .................... | .................... | .................... | .................... | .................... |
| 31435 | MT412253.1 | \|Severe acute respiratory syndrome coro | .................... | .................... | .................... | .................... | .................... | .................... |
| 31436 | MT412254.1 | \|Severe acute respiratory syndrome coro | .................... | .................... | .................... | .................... | .................... | .................... |
| 31437 | MT412255.1 | \|Severe acute respiratory syndrome coro | .................... | .................... | .................... | .................... | .................... | .................... |
| 31438 | MT412256.1 | \|Severe acute respiratory syndrome coro | .................... | .................... | .................... | .................... | .................... | .................... |
| 31439 | MT412257.1 | \|Severe acute respiratory syndrome coro | .................... | .................... | .................... | .................... | .................... | .................... |
| 31440 | MT412258.1 | \|Severe acute respiratory syndrome coro | .................... | .................... | .................... | .................... | .................... | .................... |
| 31441 | MT412259.1 | \|Severe acute respiratory syndrome coro | .................... | .................... | .................... | .................... | .................... | .................... |
| 31442 | MT412261.1 | \|Severe acute respiratory syndrome coro | .................... | .................... | .................... | .................... | .................... | .................... |
| 31443 | MT412262.1 | \|Severe acute respiratory syndrome coro | .................... | .................... | .................... | .................... | .................... | .................... |
| 31444 | MT412263.1 | \|Severe acute respiratory syndrome coro | .................... | .................... | .................... | .................... | .................... | .................... |
| 31445 | MT412264.1 | \|Severe acute respiratory syndrome coro | .................... | .................... | .................... | .................... | .................... | .................... |
| 31446 | MT412267.1 | \|Severe acute respiratory syndrome coro | .................... | .................... | .................... | .................... | .................... | .................... |
| 31447 | MT412268.1 | \|Severe acute respiratory syndrome coro | .................... | .................... | .................... | .................... | .................... | .................... |
| 31448 | MT412269.1 | \|Severe acute respiratory syndrome coro | .................... | .................... | .................... | .................... | .................... | .................... |
| 31449 | MT412271.1 | \|Severe acute respiratory syndrome coro | .................... | .................... | .................... | .................... | .................... | .................... |
| 31450 | MT412272.1 | \|Severe acute respiratory syndrome coro | .................... | .................... | .................... | .................... | .................... | .................... |
| 31451 | MT412273.1 | \|Severe acute respiratory syndrome coro | .................... | .................... | .................... | .................... | .................... | .................... |
| 31452 | MT412274.1 | \|Severe acute respiratory syndrome coro | .................... | .................... | .................... | .................... | .................... | .................... |
| 31453 | MT412275.1 | \|Severe acute respiratory syndrome coro | .................... | .................... | .................... | .................... | .................... | .................... |
| 31454 | MT412276.1 | \|Severe acute respiratory syndrome coro | .................... | .................... | .................... | .................... | .................... | .................... |
| 31455 | MT412277.1 | \|Severe acute respiratory syndrome coro | .................... | .................... | .................... | .................... | .................... | .................... |
| 31456 | MT412278.1 | \|Severe acute respiratory syndrome coro | .................... | .................... | .................... | .................... | .................... | .................... |
| 31457 | MT412279.1 | \|Severe acute respiratory syndrome coro | .................... | .................... | .................... | .................... | .................... | .................... |
| 31458 | MT412280.1 | \|Severe acute respiratory syndrome coro | .................... | .................... | .................... | .................... | .................... | .................... |
| 31459 | MT412281.1 | \|Severe acute respiratory syndrome coro | .................... | .................... | .................... | .................... | .................... | .................... |
| 31460 | MT412283.1 | \|Severe acute respiratory syndrome coro | .................... | .................... | .................... | .................... | .................... | .................... |
| 31461 | MT412284.1 | \|Severe acute respiratory syndrome coro | .................... | .................... | .................... | .................... | .................... | .................... |
| 31462 | MT412285.1 | \|Severe acute respiratory syndrome coro | .................... | .................... | .................... | .................... | .................... | .................... |
| 31463 | MT412286.1 | \|Severe acute respiratory syndrome coro | .................... | .................... | .................... | .................... | .................... | .................... |
| 31464 | MT412287.1 | \|Severe acute respiratory syndrome coro | .................... | .................... | .................... | .................... | .................... | .................... |
| 31465 | MT412288.1 | \|Severe acute respiratory syndrome coro | .................... | .................... | .................... | .................... | .................... | .................... |
| 31466 | MT412289.1 | \|Severe acute respiratory syndrome coro | .................... | .................... | .................... | .................... | .................... | .................... |
| 31467 | MT412290.1 | \|Severe acute respiratory syndrome coro | .................... | .................... | .................... | .................... | .................... | .................... |
| 31468 | MT412291.1 | \|Severe acute respiratory syndrome coro | .................... | .................... | .................... | .................... | .................... | .................... |
| 31469 | MT412292.1 | \|Severe acute respiratory syndrome coro | .................... | .................... | .................... | .................... | .................... | .................... |
| 31470 | MT412293.1 | \|Severe acute respiratory syndrome coro | .................... | .................... | .................... | .................... | .................... | .................... |
| 31471 | MT412294.1 | \|Severe acute respiratory syndrome coro | .................... | .................... | .................... | .................... | .................... | .................... |
| 31472 | MT412295.1 | \|Severe acute respiratory syndrome coro | .................... | .................... | .................... | .................... | .................... | .................... |
| 31473 | MT412296.1 | \|Severe acute respiratory syndrome coro | .................... | .................... | .................... | .................... | .................... | .................... |
| 31474 | MT412297.1 | \|Severe acute respiratory syndrome coro | .................... | .................... | .................... | .................... | .................... | .................... |
| 31475 | MT412298.1 | \|Severe acute respiratory syndrome coro | .................... | .................... | .................... | .................... | .................... | .................... |
| 31476 | MT412299.1 | \|Severe acute respiratory syndrome coro | .................... | .................... | .................... | .................... | .................... | .................... |
| 31477 | MT412300.1 | \|Severe acute respiratory syndrome coro | .................... | .................... | .................... | .................... | .................... | .................... |
| 31478 | MT412301.1 | \|Severe acute respiratory syndrome coro | .................... | .................... | .................... | .................... | .................... | .................... |
| 31479 | MT412302.1 | \|Severe acute respiratory syndrome coro | .................... | .................... | .................... | .................... | .................... | .................... |
| 31480 | MT412303.1 | \|Severe acute respiratory syndrome coro | .................... | .................... | .................... | .................... | .................... | .................... |
| 31481 | MT412304.1 | \|Severe acute respiratory syndrome coro | .................... | .................... | .................... | .................... | .................... | .................... |
| 31482 | MT412305.1 | \|Severe acute respiratory syndrome coro | .................... | .................... | .................... | .................... | .................... | .................... |
| 31483 | MT412306.1 | \|Severe acute respiratory syndrome coro | .................... | .................... | .................... | .................... | .................... | .................... |
| 31484 | MT412307.1 | \|Severe acute respiratory syndrome coro | .................... | .................... | .................... | .................... | .................... | .................... |
| 31485 | MT412308.1 | \|Severe acute respiratory syndrome coro | .................... | .................... | .................... | .................... | .................... | .................... |
| 31486 | MT412309.1 | \|Severe acute respiratory syndrome coro | .................... | .................... | .................... | .................... | .................... | .................... |

| | | | | | | | | |
|---|---|---|---|---|---|---|---|---|
| 31487 | MT412310.1 | \|Severe acute respiratory syndrome coro | ···················· | ···················· | ···················· | ···················· | ···················· | ···················· |
| 31488 | MT412311.1 | \|Severe acute respiratory syndrome coro | ···················· | ···················· | ···················· | ···················· | ···················· | ···················· |
| 31489 | MT412312.1 | \|Severe acute respiratory syndrome coro | ···················· | ···················· | ···················· | ···················· | ···················· | ···················· |
| 31490 | MT412313.1 | \|Severe acute respiratory syndrome coro | ···················· | ···················· | ···················· | ···················· | ···················· | ···················· |
| 31491 | MT412314.1 | \|Severe acute respiratory syndrome coro | ···················· | ···················· | ···················· | ···················· | ···················· | ···················· |
| 31492 | MT412316.1 | \|Severe acute respiratory syndrome coro | ···················· | ···················· | ···················· | ···················· | ···················· | ···················· |
| 31493 | MT412317.1 | \|Severe acute respiratory syndrome coro | ···················· | ···················· | ···················· | ···················· | ···················· | ···················· |
| 31494 | MT412318.1 | \|Severe acute respiratory syndrome coro | ···················· | ···················· | ···················· | ···················· | ···················· | ···················· |
| 31495 | MT412319.1 | \|Severe acute respiratory syndrome coro | ···················· | ···················· | ···················· | ···················· | ···················· | ···················· |
| 31496 | MT412321.1 | \|Severe acute respiratory syndrome coro | ···················· | ···················· | ···················· | ···················· | ···················· | ···················· |
| 31497 | MT412322.1 | \|Severe acute respiratory syndrome coro | ···················· | ···················· | ···················· | ···················· | ···················· | ···················· |
| 31498 | MT412323.1 | \|Severe acute respiratory syndrome coro | ···················· | ···················· | ···················· | ···················· | ···················· | ···················· |
| 31499 | MT412324.1 | \|Severe acute respiratory syndrome coro | ···················· | ···················· | ···················· | ···················· | ···················· | ···················· |
| 31500 | MT412325.1 | \|Severe acute respiratory syndrome coro | ···················· | ···················· | ···················· | ···················· | ···················· | ···················· |
| 31501 | MT412326.1 | \|Severe acute respiratory syndrome coro | ···················· | ···················· | ···················· | ···················· | ···················· | ···················· |
| 31502 | MT412327.1 | \|Severe acute respiratory syndrome coro | ···················· | ···················· | ···················· | ···················· | ···················· | ···················· |
| 31503 | MT412328.1 | \|Severe acute respiratory syndrome coro | ···················· | ···················· | ···················· | ···················· | ···················· | ···················· |
| 31504 | MT412329.1 | \|Severe acute respiratory syndrome coro | ···················· | ···················· | ···················· | ···················· | ···················· | ···················· |
| 31505 | MT412330.1 | \|Severe acute respiratory syndrome coro | ···················· | ···················· | ···················· | ···················· | ···················· | ···················· |
| 31506 | MT412331.1 | \|Severe acute respiratory syndrome coro | ···················· | ···················· | ···················· | ···················· | ···················· | ···················· |
| 31507 | MT412332.1 | \|Severe acute respiratory syndrome coro | ···················· | ···················· | ···················· | ···················· | ···················· | ···················· |
| 31508 | MT412333.1 | \|Severe acute respiratory syndrome coro | ···················· | ···················· | ···················· | ···················· | ···················· | ···················· |
| 31509 | MT412334.1 | \|Severe acute respiratory syndrome coro | ···················· | ···················· | ···················· | ···················· | ···················· | ···················· |
| 31510 | MT412336.1 | \|Severe acute respiratory syndrome coro | ···················· | ···················· | ···················· | ···················· | ···················· | ···················· |
| 31511 | MT412337.1 | \|Severe acute respiratory syndrome coro | ···················· | ···················· | ···················· | ···················· | ···················· | ···················· |
| 31512 | MT415320.1 | \|Severe acute respiratory syndrome coro | ···················· | ···················· | ···················· | ···················· | ···················· | ···················· |
| 31513 | MT415321.1 | \|Severe acute respiratory syndrome coro | ···················· | ···················· | ···················· | ···················· | ···················· | ···················· |
| 31514 | MT415322.1 | \|Severe acute respiratory syndrome coro | ···················· | ···················· | ···················· | ···················· | ···················· | ···················· |
| 31515 | MT415323.1 | \|Severe acute respiratory syndrome coro | ···················· | ···················· | ···················· | ···················· | ···················· | ···················· |
| 31516 | MT415894.1 | \|Severe acute respiratory syndrome coro | ···················· | ···················· | ···················· | ···················· | ···················· | ···················· |
| 31517 | MT415895.1 | \|Severe acute respiratory syndrome coro | ···················· | ···················· | ···················· | ···················· | ···················· | ···················· |
| 31518 | MT415896.1 | \|Severe acute respiratory syndrome coro | ···················· | ···················· | ···················· | ···················· | ···················· | ···················· |
| 31519 | MT415897.1 | \|Severe acute respiratory syndrome coro | ···················· | ···················· | ···················· | ···················· | ···················· | ···················· |
| 31520 | MT415898.1 | \|Severe acute respiratory syndrome coro | ···················· | ···················· | ···················· | ···················· | ···················· | ···················· |
| 31521 | MT415899.1 | \|Severe acute respiratory syndrome coro | ···················· | ···················· | ···················· | ···················· | ···················· | ···················· |
| 31522 | MT415900.1 | \|Severe acute respiratory syndrome coro | ···················· | ···················· | ···················· | ···················· | ···················· | ···················· |
| 31523 | MT415901.1 | \|Severe acute respiratory syndrome coro | ···················· | ···················· | ···················· | ···················· | ···················· | ···················· |
| 31524 | MT415902.1 | \|Severe acute respiratory syndrome coro | ···················· | ···················· | ···················· | ···················· | ···················· | ···················· |
| 31525 | MT415903.1 | \|Severe acute respiratory syndrome coro | ···················· | ···················· | ···················· | ···················· | ···················· | ···················· |
| 31526 | MT415904.1 | \|Severe acute respiratory syndrome coro | ···················· | ···················· | ···················· | ···················· | ···················· | ···················· |
| 31527 | MT415905.1 | \|Severe acute respiratory syndrome coro | ···················· | ···················· | ···················· | ···················· | ···················· | ···················· |
| 31528 | MT415906.1 | \|Severe acute respiratory syndrome coro | ···················· | ···················· | ···················· | ···················· | ···················· | ···················· |
| 31529 | MT415907.1 | \|Severe acute respiratory syndrome coro | ···················· | ···················· | ···················· | ···················· | ···················· | ···················· |
| 31530 | MT415908.1 | \|Severe acute respiratory syndrome coro | ···················· | ···················· | ···················· | ···················· | ···················· | ···················· |
| 31531 | MT415909.1 | \|Severe acute respiratory syndrome coro | ···················· | ···················· | ···················· | ···················· | ···················· | ···················· |
| 31532 | MT415910.1 | \|Severe acute respiratory syndrome coro | ···················· | ···················· | ···················· | ···················· | ···················· | ···················· |
| 31533 | MT415911.1 | \|Severe acute respiratory syndrome coro | ···················· | ···················· | ···················· | ···················· | ···················· | ···················· |
| 31534 | MT415912.1 | \|Severe acute respiratory syndrome coro | ···················· | ···················· | ···················· | ···················· | ···················· | ···················· |
| 31535 | MT394528.1 | \|Severe acute respiratory syndrome coro | ···················· | ···················· | ···················· | ···················· | ···················· | ···················· |
| 31536 | MT394529.1 | \|Severe acute respiratory syndrome coro | ···················· | ···················· | ···················· | ···················· | ···················· | ···················· |
| 31537 | MT394530.1 | \|Severe acute respiratory syndrome coro | ···················· | ···················· | ···················· | ···················· | ···················· | ···················· |
| 31538 | MT394531.1 | \|Severe acute respiratory syndrome coro | ···················· | ···················· | ···················· | ···················· | ···················· | ···················· |
| 31539 | MT396241.1 | \|Severe acute respiratory syndrome coro | ···················· | ···················· | ···················· | ···················· | ···················· | ···················· |
| 31540 | MT396266.1 | \|Severe acute respiratory syndrome coro | ···················· | ···················· | ···················· | ···················· | ···················· | ···················· |
| 31541 | MT380728.1 | \|Severe acute respiratory syndrome coro | ···················· | ···················· | ···················· | ···················· | ···················· | ···················· |
| 31542 | MT380729.1 | \|Severe acute respiratory syndrome coro | ···················· | ···················· | ···················· | ···················· | ···················· | ···················· |
| 31543 | MT380730.1 | \|Severe acute respiratory syndrome coro | ···················· | ···················· | ···················· | ···················· | ···················· | ···················· |
| 31544 | MT380731.1 | \|Severe acute respiratory syndrome coro | ···················· | ···················· | ···················· | ···················· | ···················· | ···················· |
| 31545 | MT380732.1 | \|Severe acute respiratory syndrome coro | ···················· | ···················· | ···················· | ···················· | ···················· | ···················· |
| 31546 | MT380733.1 | \|Severe acute respiratory syndrome coro | ···················· | ···················· | ···················· | ···················· | ···················· | ···················· |
| 31547 | MT380734.1 | \|Severe acute respiratory syndrome coro | ···················· | ···················· | ···················· | ···················· | ···················· | ···················· |
| 31548 | MT385414.1 | \|Severe acute respiratory syndrome coro | ···················· | ···················· | ···················· | ···················· | ···················· | ···················· |
| 31549 | MT385415.1 | \|Severe acute respiratory syndrome coro | ···················· | ···················· | ···················· | ···················· | ···················· | ···················· |
| 31550 | MT385416.1 | \|Severe acute respiratory syndrome coro | ···················· | ···················· | ···················· | ···················· | ···················· | ···················· |
| 31551 | MT385417.1 | \|Severe acute respiratory syndrome coro | ···················· | ···················· | ···················· | ···················· | ···················· | ···················· |
| 31552 | MT385418.1 | \|Severe acute respiratory syndrome coro | ···················· | ···················· | ···················· | ···················· | ···················· | ···················· |
| 31553 | MT385419.1 | \|Severe acute respiratory syndrome coro | ···················· | ···················· | ···················· | ···················· | ···················· | ···················· |
| 31554 | MT385420.1 | \|Severe acute respiratory syndrome coro | ···················· | ···················· | ···················· | ···················· | ···················· | ···················· |
| 31555 | MT385421.1 | \|Severe acute respiratory syndrome coro | ···················· | ···················· | ···················· | ···················· | ···················· | ···················· |
| 31556 | MT385422.1 | \|Severe acute respiratory syndrome coro | ···················· | ···················· | ···················· | ···················· | ···················· | ···················· |
| 31557 | MT385423.1 | \|Severe acute respiratory syndrome coro | ···················· | ···················· | ···················· | ···················· | ···················· | ···················· |
| 31558 | MT385424.1 | \|Severe acute respiratory syndrome coro | ···················· | ···················· | ···················· | ···················· | ···················· | ···················· |
| 31559 | MT385425.1 | \|Severe acute respiratory syndrome coro | ···················· | ···················· | ···················· | ···················· | ···················· | ···················· |
| 31560 | MT385426.1 | \|Severe acute respiratory syndrome coro | ···················· | ···················· | ···················· | ···················· | ···················· | ···················· |
| 31561 | MT385427.1 | \|Severe acute respiratory syndrome coro | ···················· | ···················· | ···················· | ···················· | ···················· | ···················· |
| 31562 | MT385428.1 | \|Severe acute respiratory syndrome coro | ···················· | ·A·················· | ···················· | ···················· | ···················· | ···················· |
| 31563 | MT385429.1 | \|Severe acute respiratory syndrome coro | ···················· | ···················· | ···················· | ···················· | ···················· | ···················· |
| 31564 | MT385430.1 | \|Severe acute respiratory syndrome coro | ···················· | ···················· | ···················· | ···················· | ···················· | ···················· |

| | | | | | | | | |
|---|---|---|---|---|---|---|---|---|
| 31565 | MT385431.1 | Severe acute respiratory syndrome coro | ···················· | ···················· | ····················· | ···················· | ···················· | ···················· |
| 31566 | MT385432.1 | Severe acute respiratory syndrome coro | ···················· | ···················· | ····················· | ···················· | ···················· | ···················· |
| 31567 | MT385433.1 | Severe acute respiratory syndrome coro | ···················· | ···················· | ····················· | ···················· | ···················· | ···················· |
| 31568 | MT385434.1 | Severe acute respiratory syndrome coro | ···················· | ···················· | ····················· | ···················· | ···················· | ···················· |
| 31569 | MT385435.1 | Severe acute respiratory syndrome coro | ···················· | ···················· | ····················· | ···················· | ···················· | ···················· |
| 31570 | MT385436.1 | Severe acute respiratory syndrome coro | ···················· | ···················· | ····················· | ···················· | ···················· | ···················· |
| 31571 | MT385437.1 | Severe acute respiratory syndrome coro | ···················· | ···················· | ····················· | ···················· | ···················· | ···················· |
| 31572 | MT385438.1 | Severe acute respiratory syndrome coro | ···················· | ···················· | ····················· | ···················· | ···················· | ···················· |
| 31573 | MT385439.1 | Severe acute respiratory syndrome coro | ···················· | ···················· | ····················· | ···················· | ···················· | ···················· |
| 31574 | MT385440.1 | Severe acute respiratory syndrome coro | ···················· | ···A················ | ····················· | ···················· | ···················· | ···················· |
| 31575 | MT385441.1 | Severe acute respiratory syndrome coro | ···················· | ···················· | ····················· | ···················· | ···················· | ···················· |
| 31576 | MT385442.1 | Severe acute respiratory syndrome coro | ···················· | ···················· | ····················· | ···················· | ···················· | ···················· |
| 31577 | MT385443.1 | Severe acute respiratory syndrome coro | ···················· | ···················· | ····················· | ···················· | ···················· | ···················· |
| 31578 | MT385444.1 | Severe acute respiratory syndrome coro | ···················· | ···················· | ····················· | ···················· | ···················· | ···················· |
| 31579 | MT385445.1 | Severe acute respiratory syndrome coro | ···················· | ···················· | ····················· | ···················· | ···················· | ···················· |
| 31580 | MT385446.1 | Severe acute respiratory syndrome coro | ···················· | ···················· | ····················· | ···················· | ···················· | ···················· |
| 31581 | MT385447.1 | Severe acute respiratory syndrome coro | ···················· | ···················· | ····················· | ···················· | ···················· | ···················· |
| 31582 | MT385448.1 | Severe acute respiratory syndrome coro | ···················· | ···················· | ····················· | ···················· | ···················· | ···················· |
| 31583 | MT385449.1 | Severe acute respiratory syndrome coro | ···················· | ···················· | ····················· | ···················· | ···················· | ···················· |
| 31584 | MT385450.1 | Severe acute respiratory syndrome coro | ···················· | ···················· | ····················· | ···················· | ···················· | ···················· |
| 31585 | MT385451.1 | Severe acute respiratory syndrome coro | ···················· | ···················· | ····················· | ···················· | ···················· | ···················· |
| 31586 | MT385452.1 | Severe acute respiratory syndrome coro | ···················· | ···················· | ····················· | ···················· | ···················· | ···················· |
| 31587 | MT385453.1 | Severe acute respiratory syndrome coro | ···················· | ···················· | ····················· | ···················· | ···················· | ···················· |
| 31588 | MT385454.1 | Severe acute respiratory syndrome coro | ···················· | ···················· | ····················· | ···················· | ···················· | ···················· |
| 31589 | MT385455.1 | Severe acute respiratory syndrome coro | ···················· | ···················· | ····················· | ···················· | ···················· | ···················· |
| 31590 | MT385456.1 | Severe acute respiratory syndrome coro | ···················· | ···················· | ····················· | ···················· | ···················· | ···················· |
| 31591 | MT385457.1 | Severe acute respiratory syndrome coro | ···················· | ···················· | ····················· | ···················· | ···················· | ···················· |
| 31592 | MT385458.1 | Severe acute respiratory syndrome coro | ···················· | ···················· | ····················· | ···················· | ···················· | ···················· |
| 31593 | MT385459.1 | Severe acute respiratory syndrome coro | ···················· | ···················· | ····················· | ···················· | ···················· | ···················· |
| 31594 | MT385460.1 | Severe acute respiratory syndrome coro | ···················· | ···················· | ····················· | ···················· | ···················· | ···················· |
| 31595 | MT385461.1 | Severe acute respiratory syndrome coro | ···················· | ···················· | ····················· | ···················· | ···················· | ···················· |
| 31596 | MT385462.1 | Severe acute respiratory syndrome coro | ···················· | ···················· | ····················· | ···················· | ···················· | ···················· |
| 31597 | MT385463.1 | Severe acute respiratory syndrome coro | ···················· | ···················· | ····················· | ···················· | ···················· | ···················· |
| 31598 | MT385464.1 | Severe acute respiratory syndrome coro | ···················· | ···················· | ····················· | ···················· | ···················· | ···················· |
| 31599 | MT385465.1 | Severe acute respiratory syndrome coro | ···················· | ···················· | ····················· | ···················· | ···················· | ···················· |
| 31600 | MT385466.1 | Severe acute respiratory syndrome coro | ···················· | ···················· | ····················· | ···················· | ···················· | ···················· |
| 31601 | MT385467.1 | Severe acute respiratory syndrome coro | ···················· | ···················· | ····················· | ···················· | ···················· | ···················· |
| 31602 | MT385468.1 | Severe acute respiratory syndrome coro | ···················· | ···················· | ····················· | ···················· | ···················· | ···················· |
| 31603 | MT385469.1 | Severe acute respiratory syndrome coro | ···················· | ···················· | ····················· | ···················· | ···················· | ···················· |
| 31604 | MT385470.1 | Severe acute respiratory syndrome coro | ···················· | ···················· | ····················· | ···················· | ···················· | ···················· |
| 31605 | MT385471.1 | Severe acute respiratory syndrome coro | ···················· | ···················· | ····················· | ···················· | ···················· | ···················· |
| 31606 | MT385472.1 | Severe acute respiratory syndrome coro | ···················· | ···················· | ····················· | ···················· | ···················· | ···················· |
| 31607 | MT385473.1 | Severe acute respiratory syndrome coro | ···················· | ···················· | ····················· | ···················· | ···················· | ···················· |
| 31608 | MT385474.1 | Severe acute respiratory syndrome coro | ···················· | ···················· | ····················· | ···················· | ···················· | ···················· |
| 31609 | MT385475.1 | Severe acute respiratory syndrome coro | ···················· | ···················· | ····················· | ···················· | ···················· | ···················· |
| 31610 | MT385476.1 | Severe acute respiratory syndrome coro | ···················· | ···················· | ····················· | ···················· | ···················· | ···················· |
| 31611 | MT385477.1 | Severe acute respiratory syndrome coro | ···················· | ···················· | ····················· | ···················· | ···················· | ···················· |
| 31612 | MT385478.1 | Severe acute respiratory syndrome coro | ···················· | ···················· | ····················· | ···················· | ···················· | ···················· |
| 31613 | MT385479.1 | Severe acute respiratory syndrome coro | ···················· | ···················· | ····················· | ···················· | ···················· | ···················· |
| 31614 | MT385482.1 | Severe acute respiratory syndrome coro | ···················· | ···················· | ····················· | ···················· | ···················· | ···················· |
| 31615 | MT385485.1 | Severe acute respiratory syndrome coro | ···················· | ···················· | ····················· | ···················· | ···················· | ···················· |
| 31616 | MT186683.1 | Severe acute respiratory syndrome coro | ···················· | ···················· | ····················· | ···················· | ···················· | ···················· |
| 31617 | MT252677.1 | Severe acute respiratory syndrome coro | ···················· | ···················· | ····················· | ···················· | ···················· | ···················· |
| 31618 | MT252678.1 | Severe acute respiratory syndrome coro | ···················· | ···················· | ····················· | ···················· | ···················· | ···················· |
| 31619 | MT252679.1 | Severe acute respiratory syndrome coro | ···················· | ···················· | ····················· | ···················· | ···················· | ···················· |
| 31620 | MT252680.1 | Severe acute respiratory syndrome coro | ···················· | ···················· | ····················· | ···················· | ···················· | ···················· |
| 31621 | MT252681.1 | Severe acute respiratory syndrome coro | ···················· | ···················· | ····················· | ···················· | ···················· | ···················· |
| 31622 | MT252682.1 | Severe acute respiratory syndrome coro | ···················· | ···················· | ····················· | ···················· | ···················· | ···················· |
| 31623 | MT252683.1 | Severe acute respiratory syndrome coro | ···················· | ···················· | ····················· | ···················· | ···················· | ···················· |
| 31624 | MT252684.1 | Severe acute respiratory syndrome coro | ···················· | ···················· | ····················· | ···················· | ···················· | ···················· |
| 31625 | MT252685.1 | Severe acute respiratory syndrome coro | ···················· | ···················· | ····················· | ···················· | ···················· | ···················· |
| 31626 | MT252686.1 | Severe acute respiratory syndrome coro | ···················· | ···················· | ····················· | ···················· | ···················· | ···················· |
| 31627 | MT252687.1 | Severe acute respiratory syndrome coro | ···················· | ···················· | ····················· | ···················· | ···················· | ···················· |
| 31628 | MT252688.1 | Severe acute respiratory syndrome coro | ···················· | ···················· | ····················· | ···················· | ···················· | ···················· |
| 31629 | MT252689.1 | Severe acute respiratory syndrome coro | ···················· | ···················· | ····················· | ···················· | ···················· | ···················· |
| 31630 | MT252690.1 | Severe acute respiratory syndrome coro | ···················· | ···················· | ····················· | ···················· | ···················· | ···················· |
| 31631 | MT252691.1 | Severe acute respiratory syndrome coro | ···················· | ···················· | ····················· | ···················· | ···················· | ···················· |
| 31632 | MT252693.1 | Severe acute respiratory syndrome coro | ···················· | ···················· | ····················· | ···················· | ···················· | ···················· |
| 31633 | MT252694.1 | Severe acute respiratory syndrome coro | ···················· | ···················· | ····················· | ···················· | ···················· | ···················· |
| 31634 | MT252695.1 | Severe acute respiratory syndrome coro | ···················· | ···················· | ····················· | ···················· | ···················· | ···················· |
| 31635 | MT252696.1 | Severe acute respiratory syndrome coro | ···················· | ···················· | ····················· | ···················· | ···················· | ···················· |
| 31636 | MT252697.1 | Severe acute respiratory syndrome coro | ···················· | ···················· | ····················· | ···················· | ···················· | ···················· |
| 31637 | MT252698.1 | Severe acute respiratory syndrome coro | ···················· | ···················· | ····················· | ···················· | ···················· | ···················· |
| 31638 | MT252699.1 | Severe acute respiratory syndrome coro | ···················· | ···················· | ····················· | ···················· | ···················· | ···················· |
| 31639 | MT252700.1 | Severe acute respiratory syndrome coro | ···················· | ···················· | ····················· | ···················· | ···················· | ···················· |
| 31640 | MT252701.1 | Severe acute respiratory syndrome coro | ···················· | ···················· | ····················· | ···················· | ···················· | ···················· |
| 31641 | MT252702.1 | Severe acute respiratory syndrome coro | ···················· | ···················· | ····················· | ···················· | ···················· | ···················· |
| 31642 | MT252703.1 | Severe acute respiratory syndrome coro | ···················· | ···················· | ····················· | ···················· | ···················· | ···················· |

```
31643 MT252704.1 |Severe acute respiratory syndrome coro    ....................  ....................  .....................  ....................  ....................  ....................
31644 MT252705.1 |Severe acute respiratory syndrome coro    ....................  ....................  .....................  ....................  ....................  ....................
31645 MT252706.1 |Severe acute respiratory syndrome coro    ....................  ....................  .....................  ....................  ....................  ....................
31646 MT252707.1 |Severe acute respiratory syndrome coro    ....................  ....................  .....................  ....................  ....................  ....................
31647 MT252708.1 |Severe acute respiratory syndrome coro    ....................  ....................  .....................  ....................  ....................  ....................
31648 MT252709.1 |Severe acute respiratory syndrome coro    ....................  ....................  .....................  ....................  ....................  ....................
31649 MT252710.1 |Severe acute respiratory syndrome coro    ....................  ....................  .....................  ....................  ....................  ....................
31650 MT252712.1 |Severe acute respiratory syndrome coro    ....................  ....................  .....................  ....................  ....................  ....................
31651 MT252713.1 |Severe acute respiratory syndrome coro    ....................  ....................  .....................  ....................  ....................  ....................
31652 MT252715.1 |Severe acute respiratory syndrome coro    ....................  ....................  .....................  ....................  ....................  ....................
31653 MT252716.1 |Severe acute respiratory syndrome coro    ....................  ....................  .....................  ....................  ....................  ....................
31654 MT252717.1 |Severe acute respiratory syndrome coro    ....................  ....................  .....................  ....................  ....................  ....................
31655 MT252719.1 |Severe acute respiratory syndrome coro    ....................  ....................  .....................  ....................  ....................  ....................
31656 MT252721.1 |Severe acute respiratory syndrome coro    ....................  ....................  .....................  ....................  ....................  ....................
31657 MT252723.1 |Severe acute respiratory syndrome coro    ....................  ....................  .....................  ....................  ....................  ....................
31658 MT252725.1 |Severe acute respiratory syndrome coro    ....................  ....................  .....................  ....................  ....................  ....................
31659 MT252726.1 |Severe acute respiratory syndrome coro    ....................  ....................  .....................  ....................  ....................  ....................
31660 MT252728.1 |Severe acute respiratory syndrome coro    ....................  ....................  .....................  ....................  ....................  ....................
31661 MT252729.1 |Severe acute respiratory syndrome coro    ....................  ....................  .....................  ....................  ....................  ....................
31662 MT252730.1 |Severe acute respiratory syndrome coro    ....................  ....................  .....................  ....................  ....................  ....................
31663 MT252733.1 |Severe acute respiratory syndrome coro    ....................  ....................  .....................  ....................  ....................  ....................
31664 MT252735.1 |Severe acute respiratory syndrome coro    ....................  ....................  .....................  ....................  ....................  ....................
31665 MT252736.1 |Severe acute respiratory syndrome coro    ....................  ....................  .....................  ....................  ....................  ....................
31666 MT252737.1 |Severe acute respiratory syndrome coro    ....................  ....................  .....................  ....................  ....................  ....................
31667 MT252738.1 |Severe acute respiratory syndrome coro    ....................  ....................  .....................  ....................  ....................  ....................
31668 MT252739.1 |Severe acute respiratory syndrome coro    ....................  ....................  .....................  ....................  ....................  ....................
31669 MT252740.1 |Severe acute respiratory syndrome coro    ....................  ....................  .....................  ....................  ....................  ....................
31670 MT252741.1 |Severe acute respiratory syndrome coro    ....................  ....................  .....................  ....................  ....................  ....................
31671 MT252742.1 |Severe acute respiratory syndrome coro    ....................  ....................  .....................  ....................  ....................  ....................
31672 MT252745.1 |Severe acute respiratory syndrome coro    ....................  ....................  .....................  ....................  ....................  ....................
31673 MT252746.1 |Severe acute respiratory syndrome coro    ....................  ....................  .....................  ....................  ....................  ....................
31674 MT252747.1 |Severe acute respiratory syndrome coro    ....................  ....................  .....................  ....................  ....................  ....................
31675 MT252748.1 |Severe acute respiratory syndrome coro    ....................  ....................  .....................  ....................  ....................  ....................
31676 MT252749.1 |Severe acute respiratory syndrome coro    ....................  ....................  .....................  ....................  ....................  ....................
31677 MT252756.1 |Severe acute respiratory syndrome coro    ....................  ....................  .....................  ....................  ....................  ....................
31678 MT252757.1 |Severe acute respiratory syndrome coro    ....................  ....................  .....................  ....................  ....................  ....................
31679 MT252758.1 |Severe acute respiratory syndrome coro    ....................  ....................  .....................  ....................  ....................  ....................
31680 MT252761.1 |Severe acute respiratory syndrome coro    ....................  ....................  .....................  ....................  ....................  ....................
31681 MT252763.1 |Severe acute respiratory syndrome coro    ....................  ....................  .....................  ....................  ....................  ....................
31682 MT252764.1 |Severe acute respiratory syndrome coro    ....................  ....................  .....................  ....................  ....................  ....................
31683 MT252767.1 |Severe acute respiratory syndrome coro    ....................  ....................  .....................  ....................  ....................  ....................
31684 MT252768.1 |Severe acute respiratory syndrome coro    ....................  ....................  .....................  ....................  ....................  ....................
31685 MT252769.1 |Severe acute respiratory syndrome coro    ....................  ....................  .....................  ....................  ....................  ....................
31686 MT252770.1 |Severe acute respiratory syndrome coro    ....................  ....................  .....................  ....................  ....................  ....................
31687 MT252772.1 |Severe acute respiratory syndrome coro    ....................  ....................  .....................  ....................  ....................  ....................
31688 MT252773.1 |Severe acute respiratory syndrome coro    ....................  ....................  .....................  ....................  ....................  ....................
31689 MT252774.1 |Severe acute respiratory syndrome coro    ....................  ....................  .....................  ....................  ....................  ....................
31690 MT252775.1 |Severe acute respiratory syndrome coro    ....................  ....................  .....................  ....................  ....................  ....................
31691 MT252778.1 |Severe acute respiratory syndrome coro    ....................  ....................  .....................  ....................  ....................  ....................
31692 MT252779.1 |Severe acute respiratory syndrome coro    ....................  ....................  .....................  ....................  ....................  ....................
31693 MT252780.1 |Severe acute respiratory syndrome coro    ....................  ....................  .....................  ....................  ....................  ....................
31694 MT252781.1 |Severe acute respiratory syndrome coro    ....................  ....................  .....................  ....................  ....................  ....................
31695 MT252782.1 |Severe acute respiratory syndrome coro    ....................  ....................  .....................  ....................  ....................  ....................
31696 MT252783.1 |Severe acute respiratory syndrome coro    ....................  ....................  .....................  ....................  ....................  ....................
31697 MT252784.1 |Severe acute respiratory syndrome coro    ....................  ....................  .....................  ....................  ....................  ....................
31698 MT252785.1 |Severe acute respiratory syndrome coro    ....................  ....................  .....................  ....................  ....................  ....................
31699 MT252787.1 |Severe acute respiratory syndrome coro    ....................  ....................  .....................  ....................  ....................  ....................
31700 MT252788.1 |Severe acute respiratory syndrome coro    ....................  ....................  .....................  ....................  ....................  ....................
31701 MT252792.1 |Severe acute respiratory syndrome coro    ....................  ....................  .....................  ....................  ....................  ....................
31702 MT252793.1 |Severe acute respiratory syndrome coro    ....................  ....................  .....................  ....................  ....................  ....................
31703 MT252794.1 |Severe acute respiratory syndrome coro    ....................  ....................  .....................  ....................  ....................  ....................
31704 MT252795.1 |Severe acute respiratory syndrome coro    ....................  ....................  .....................  ....................  ....................  ....................
31705 MT252797.1 |Severe acute respiratory syndrome coro    ....................  ....................  .....................  ....................  ....................  ....................
31706 MT252798.1 |Severe acute respiratory syndrome coro    ....................  ....................  .....................  ....................  ....................  ....................
31707 MT252799.1 |Severe acute respiratory syndrome coro    ....................  ....................  .....................  ....................  ....................  ....................
31708 MT252800.1 |Severe acute respiratory syndrome coro    ....................  ....................  .....................  ....................  ....................  ....................
31709 MT252801.1 |Severe acute respiratory syndrome coro    ....................  ....................  .....................  ....................  ....................  ....................
31710 MT252802.1 |Severe acute respiratory syndrome coro    ....................  ....................  .....................  ....................  ....................  ....................
31711 MT252803.1 |Severe acute respiratory syndrome coro    ....................  ....................  .....................  ....................  ....................  ....................
31712 MT252804.1 |Severe acute respiratory syndrome coro    ....................  ....................  .....................  ....................  ....................  ....................
31713 MT252805.1 |Severe acute respiratory syndrome coro    ....................  ....................  .....................  ....................  ....................  ....................
31714 MT252806.1 |Severe acute respiratory syndrome coro    ....................  ....................  .....................  ....................  ....................  ....................
31715 MT252807.1 |Severe acute respiratory syndrome coro    ....................  ....................  .....................  ....................  ....................  ....................
31716 MT252809.1 |Severe acute respiratory syndrome coro    ....................  ....................  .....................  ....................  ....................  ....................
31717 MT252810.1 |Severe acute respiratory syndrome coro    ....................  ....................  .....................  ....................  ....................  ....................
31718 MT252811.1 |Severe acute respiratory syndrome coro    ....................  ....................  .....................  ....................  ....................  ....................
31719 MT252822.1 |Severe acute respiratory syndrome coro    ....................  ....................  .....................  ....................  ....................  ....................
31720 MT252823.1 |Severe acute respiratory syndrome coro    ....................  ....................  .....................  ....................  ....................  ....................
```

| | | | | | | | | | |
|---|---|---|---|---|---|---|---|---|---|
| 31721 | MT339043.1 | Severe acute respiratory syndrome coro | .................... | .................... | .................... | .................... | .................... | .................... | |
| 31722 | MT365033.1 | Severe acute respiratory syndrome coro | .................... | .................... | .................... | .................... | .................... | .................... | |
| 31723 | MT374101.1 | Severe acute respiratory syndrome coro | .................... | .................... | .................... | .................... | .................... | .................... | |
| 31724 | MT374102.1 | Severe acute respiratory syndrome coro | .................... | .................... | .................... | .................... | .................... | .................... | |
| 31725 | MT374103.1 | Severe acute respiratory syndrome coro | .................... | .................... | .................... | .................... | .................... | .................... | |
| 31726 | MT374104.1 | Severe acute respiratory syndrome coro | .................... | .................... | .................... | .................... | .................... | .................... | |
| 31727 | MT374105.1 | Severe acute respiratory syndrome coro | .................... | .................... | .................... | .................... | .................... | .................... | |
| 31728 | MT374106.1 | Severe acute respiratory syndrome coro | .................... | .................... | .................... | .................... | .................... | .................... | |
| 31729 | MT374107.1 | Severe acute respiratory syndrome coro | .................... | .................... | .................... | .................... | .................... | .................... | |
| 31730 | MT374108.1 | Severe acute respiratory syndrome coro | .................... | .................... | .................... | .................... | .................... | .................... | |
| 31731 | MT374109.1 | Severe acute respiratory syndrome coro | .................... | .................... | .................... | .................... | .................... | .................... | |
| 31732 | MT374110.1 | Severe acute respiratory syndrome coro | .................... | .................... | .................... | .................... | .................... | .................... | |
| 31733 | MT374111.1 | Severe acute respiratory syndrome coro | .................... | .................... | .................... | .................... | .................... | .................... | |
| 31734 | MT374112.1 | Severe acute respiratory syndrome coro | .................... | .................... | .................... | .................... | .................... | .................... | |
| 31735 | MT374113.1 | Severe acute respiratory syndrome coro | .................... | .................... | .................... | .................... | .................... | .................... | |
| 31736 | MT374114.1 | Severe acute respiratory syndrome coro | .................... | .................... | .................... | .................... | .................... | .................... | |
| 31737 | MT374115.1 | Severe acute respiratory syndrome coro | .................... | .................... | .................... | .................... | .................... | .................... | |
| 31738 | MT374116.1 | Severe acute respiratory syndrome coro | .................... | .................... | .................... | .................... | .................... | .................... | |
| 31739 | MT375428.1 | Severe acute respiratory syndrome coro | .................... | .................... | .................... | .................... | .................... | .................... | |
| 31740 | MT375429.1 | Severe acute respiratory syndrome coro | .................... | .................... | .................... | .................... | .................... | .................... | |
| 31741 | MT375430.1 | Severe acute respiratory syndrome coro | .................... | .................... | .................... | .................... | .................... | .................... | |
| 31742 | MT375431.1 | Severe acute respiratory syndrome coro | .................... | .................... | .................... | .................... | .................... | .................... | |
| 31743 | MT375432.1 | Severe acute respiratory syndrome coro | .................... | .................... | .................... | .................... | .................... | .................... | |
| 31744 | MT375433.1 | Severe acute respiratory syndrome coro | .................... | .................... | .................... | .................... | .................... | .................... | |
| 31745 | MT375434.1 | Severe acute respiratory syndrome coro | .................... | .................... | .................... | .................... | .................... | .................... | |
| 31746 | MT375435.1 | Severe acute respiratory syndrome coro | .................... | .................... | .................... | .................... | .................... | .................... | |
| 31748 | MT375437.1 | Severe acute respiratory syndrome coro | .................... | .................... | .................... | .................... | .................... | .................... | |
| 31749 | MT375438.1 | Severe acute respiratory syndrome coro | .................... | .................... | .................... | .................... | .................... | .................... | |
| 31750 | MT375440.1 | Severe acute respiratory syndrome coro | .................... | .................... | .................... | .................... | .................... | .................... | |
| 31751 | MT375441.1 | Severe acute respiratory syndrome coro | .................... | .................... | .................... | .................... | .................... | .................... | |
| 31752 | MT375442.1 | Severe acute respiratory syndrome coro | .................... | .................... | .................... | .................... | .................... | .................... | |
| 31753 | MT375443.1 | Severe acute respiratory syndrome coro | .................... | .................... | .................... | .................... | .................... | .................... | |
| 31754 | MT375444.1 | Severe acute respiratory syndrome coro | .................... | .................... | .................... | .................... | .................... | .................... | |
| 31755 | MT375445.1 | Severe acute respiratory syndrome coro | .................... | .................... | .................... | .................... | .................... | .................... | |
| 31756 | MT375446.1 | Severe acute respiratory syndrome coro | .................... | .................... | .................... | .................... | .................... | .................... | |
| 31757 | MT375447.1 | Severe acute respiratory syndrome coro | .................... | .................... | .................... | .................... | .................... | .................... | |
| 31759 | MT375449.1 | Severe acute respiratory syndrome coro | .................... | .................... | .................... | .................... | .................... | .................... | |
| 31760 | MT375450.1 | Severe acute respiratory syndrome coro | .................... | .................... | .................... | .................... | .................... | .................... | |
| 31761 | MT375451.1 | Severe acute respiratory syndrome coro | .................... | .................... | .................... | .................... | .................... | .................... | |
| 31762 | MT375452.1 | Severe acute respiratory syndrome coro | .................... | .................... | .................... | .................... | .................... | .................... | |
| 31763 | MT375453.1 | Severe acute respiratory syndrome coro | .................... | .................... | .................... | .................... | .................... | .................... | |
| 31764 | MT375454.1 | Severe acute respiratory syndrome coro | .................... | .................... | .................... | .................... | .................... | .................... | |
| 31765 | MT375455.1 | Severe acute respiratory syndrome coro | .................... | .................... | .................... | .................... | .................... | .................... | |
| 31766 | MT375456.1 | Severe acute respiratory syndrome coro | .................... | .................... | .................... | .................... | .................... | .................... | |
| 31767 | MT375457.1 | Severe acute respiratory syndrome coro | .................... | .................... | .................... | .................... | .................... | .................... | |
| 31768 | MT375458.1 | Severe acute respiratory syndrome coro | .................... | .................... | .................... | .................... | .................... | .................... | |
| 31769 | MT375459.1 | Severe acute respiratory syndrome coro | .................... | .................... | .................... | .................... | .................... | .................... | |
| 31770 | MT375460.1 | Severe acute respiratory syndrome coro | .................... | .................... | .................... | .................... | .................... | .................... | |
| 31771 | MT375461.1 | Severe acute respiratory syndrome coro | .................... | .................... | .................... | .................... | .................... | .................... | |
| 31772 | MT375462.1 | Severe acute respiratory syndrome coro | .................... | .................... | .................... | .................... | .................... | .................... | |
| 31773 | MT375463.1 | Severe acute respiratory syndrome coro | .................... | .................... | .................... | .................... | .................... | .................... | |
| 31774 | MT375465.1 | Severe acute respiratory syndrome coro | .................... | .................... | .................... | .................... | .................... | .................... | |
| 31775 | MT375466.1 | Severe acute respiratory syndrome coro | .................... | .................... | .................... | .................... | .................... | .................... | |
| 31776 | MT375467.1 | Severe acute respiratory syndrome coro | .................... | .................... | .................... | .................... | .................... | .................... | |
| 31777 | MT375468.1 | Severe acute respiratory syndrome coro | .................... | .................... | .................... | .................... | .................... | .................... | |
| 31779 | MT375470.1 | Severe acute respiratory syndrome coro | .................... | .................... | .................... | .................... | .................... | .................... | |
| 31780 | MT375471.1 | Severe acute respiratory syndrome coro | .................... | ...................A | .................... | .................... | .................... | .................... | 1 |
| 31781 | MT375472.1 | Severe acute respiratory syndrome coro | .................... | .................... | .................... | .................... | .................... | .................... | |
| 31782 | MT375474.1 | Severe acute respiratory syndrome coro | .................... | .................... | .................... | .................... | .................... | .................... | |
| 31783 | MT375476.1 | Severe acute respiratory syndrome coro | .................... | .................... | .................... | .................... | .................... | .................... | |
| 31784 | MT375477.1 | Severe acute respiratory syndrome coro | .................... | .................... | .................... | .................... | .................... | .................... | |
| 31785 | MT375478.1 | Severe acute respiratory syndrome coro | .................... | .................... | .................... | .................... | .................... | .................... | |
| 31786 | MT375480.1 | Severe acute respiratory syndrome coro | .................... | .................... | .................... | .................... | .................... | .................... | |
| 31787 | MT375481.1 | Severe acute respiratory syndrome coro | .................... | .................... | .................... | .................... | .................... | .................... | |
| 31788 | MT375482.1 | Severe acute respiratory syndrome coro | .................... | .................... | .................... | .................... | .................... | .................... | |
| 31789 | MT375483.1 | Severe acute respiratory syndrome coro | .................... | .................... | .................... | .................... | .................... | .................... | |
| 31790 | MT370516.1 | Severe acute respiratory syndrome coro | .................... | .................... | .................... | .................... | .................... | .................... | |
| 31791 | MT370518.1 | Severe acute respiratory syndrome coro | .................... | .................... | .................... | .................... | .................... | .................... | |
| 31792 | MT370831.1 | Severe acute respiratory syndrome coro | .................... | .................... | .................... | .................... | .................... | .................... | |
| 31793 | MT370833.1 | Severe acute respiratory syndrome coro | .................... | .................... | .................... | .................... | .................... | .................... | |
| 31794 | MT370834.1 | Severe acute respiratory syndrome coro | .................... | ...................A | .................... | .................... | .................... | .................... | 1 |
| 31795 | MT370835.1 | Severe acute respiratory syndrome coro | .................... | .................... | .................... | .................... | .................... | .................... | |
| 31796 | MT370836.1 | Severe acute respiratory syndrome coro | .................... | .................... | .................... | .................... | .................... | .................... | |
| 31797 | MT370837.1 | Severe acute respiratory syndrome coro | .................... | .................... | .................... | .................... | .................... | .................... | |
| 31798 | MT370838.1 | Severe acute respiratory syndrome coro | .................... | .................... | .................... | .................... | .................... | .................... | |
| 31799 | MT370839.1 | Severe acute respiratory syndrome coro | .................... | .................... | .................... | .................... | .................... | .................... | |
| 31800 | MT370841.1 | Severe acute respiratory syndrome coro | .................... | .................... | .................... | .................... | .................... | .................... | |
| 31801 | MT370842.1 | Severe acute respiratory syndrome coro | .................... | .................... | .................... | .................... | .................... | .................... | |

| | | | | | | |
|---|---|---|---|---|---|---|
| 31802 MT370843.1 \|Severe acute respiratory syndrome coro | ···················· | ···················· | ····················· | ···················· | ···················· | ···················· |
| 31803 MT370844.1 \|Severe acute respiratory syndrome coro | ···················· | ···················· | ····················· | ···················· | ···················· | ···················· |
| 31804 MT370845.1 \|Severe acute respiratory syndrome coro | ···················· | ···················· | ····················· | ···················· | ···················· | ···················· |
| 31805 MT370846.1 \|Severe acute respiratory syndrome coro | ···················· | ···················· | ····················· | ···················· | ···················· | ···················· |
| 31806 MT370848.1 \|Severe acute respiratory syndrome coro | ···················· | ···················· | ····················· | ···················· | ···················· | ···················· |
| 31807 MT370849.1 \|Severe acute respiratory syndrome coro | ···················· | ···················· | ····················· | ···················· | ···················· | ···················· |
| 31808 MT370850.1 \|Severe acute respiratory syndrome coro | ···················· | ···················· | ····················· | ···················· | ···················· | ···················· |
| 31809 MT370851.1 \|Severe acute respiratory syndrome coro | ···················· | ···················· | ····················· | ···················· | ···················· | ···················· |
| 31810 MT370852.1 \|Severe acute respiratory syndrome coro | ···················· | ·············A······ | ····················· | ···················· | ···················· | ···················· |
| 31811 MT370853.1 \|Severe acute respiratory syndrome coro | ···················· | ···················· | ····················· | ···················· | ···················· | ···················· |
| 31812 MT370854.1 \|Severe acute respiratory syndrome coro | ···················· | ···················· | ····················· | ···················· | ···················· | ···················· |
| 31813 MT370856.1 \|Severe acute respiratory syndrome coro | ···················· | ···················· | ····················· | ···················· | ···················· | ···················· |
| 31814 MT370857.1 \|Severe acute respiratory syndrome coro | ···················· | ···················· | ····················· | ···················· | ···················· | ···················· |
| 31815 MT370858.1 \|Severe acute respiratory syndrome coro | ···················· | ······A············· | ····················· | ···················· | ···················· | ···················· |
| 31816 MT370859.1 \|Severe acute respiratory syndrome coro | ···················· | ···················· | ····················· | ···················· | ···················· | ···················· |
| 31817 MT370860.1 \|Severe acute respiratory syndrome coro | ···················· | ···················· | ····················· | ···················· | ···················· | ···················· |
| 31818 MT370861.1 \|Severe acute respiratory syndrome coro | ···················· | ···················· | ····················· | ···················· | ···················· | ···················· |
| 31819 MT370862.1 \|Severe acute respiratory syndrome coro | ···················· | ···················· | ····················· | ···················· | ···················· | ···················· |
| 31820 MT370863.1 \|Severe acute respiratory syndrome coro | ···················· | ···················· | ····················· | ···················· | ···················· | ···················· |
| 31821 MT370864.1 \|Severe acute respiratory syndrome coro | ···················· | ···················· | ····················· | ···················· | ···················· | ···················· |
| 31822 MT370865.1 \|Severe acute respiratory syndrome coro | ···················· | ···················· | ····················· | ···················· | ···················· | ···················· |
| 31823 MT370866.1 \|Severe acute respiratory syndrome coro | ···················· | ·············A······ | ····················· | ···················· | ···················· | ···················· |
| 31824 MT370868.1 \|Severe acute respiratory syndrome coro | ···················· | ···················· | ····················· | ···················· | ···················· | ···················· |
| 31825 MT370869.1 \|Severe acute respiratory syndrome coro | ···················· | ···················· | ····················· | ···················· | ···················· | ···················· |
| 31826 MT370870.1 \|Severe acute respiratory syndrome coro | ···················· | ···················· | ····················· | ···················· | ···················· | ···················· |
| 31827 MT370871.1 \|Severe acute respiratory syndrome coro | ···················· | ···················· | ····················· | ···················· | ···················· | ···················· |
| 31828 MT370872.1 \|Severe acute respiratory syndrome coro | ···················· | ···················· | ····················· | ···················· | ···················· | ···················· |
| 31829 MT370874.1 \|Severe acute respiratory syndrome coro | ···················· | ···················· | ····················· | ···················· | ···················· | ···················· |
| 31830 MT370875.1 \|Severe acute respiratory syndrome coro | ···················· | ···················· | ····················· | ···················· | ···················· | ···················· |
| 31831 MT370876.1 \|Severe acute respiratory syndrome coro | ···················· | ···················· | ····················· | ···················· | ···················· | ···················· |
| 31832 MT370877.1 \|Severe acute respiratory syndrome coro | ···················· | ···················· | ····················· | ···················· | ···················· | ···················· |
| 31833 MT370878.1 \|Severe acute respiratory syndrome coro | ···················· | ···················· | ····················· | ···················· | ···················· | ···················· |
| 31834 MT370880.1 \|Severe acute respiratory syndrome coro | ···················· | ···················· | ····················· | ···················· | ···················· | ···················· |
| 31835 MT370881.1 \|Severe acute respiratory syndrome coro | ···················· | ···················· | ····················· | ···················· | ···················· | ···················· |
| 31836 MT370882.1 \|Severe acute respiratory syndrome coro | ···················· | ···················· | ····················· | ···················· | ···················· | ···················· |
| 31837 MT370883.1 \|Severe acute respiratory syndrome coro | ···················· | ···················· | ····················· | ···················· | ···················· | ···················· |
| 31838 MT370885.1 \|Severe acute respiratory syndrome coro | ···················· | ···················· | ····················· | ···················· | ···················· | ···················· |
| 31839 MT370886.1 \|Severe acute respiratory syndrome coro | ···················· | ···················· | ····················· | ···················· | ···················· | ···················· |
| 31840 MT370888.1 \|Severe acute respiratory syndrome coro | ···················· | ···················· | ····················· | ···················· | ···················· | ···················· |
| 31841 MT370889.1 \|Severe acute respiratory syndrome coro | ···················· | ···················· | ····················· | ···················· | ···················· | ···················· |
| 31842 MT370890.1 \|Severe acute respiratory syndrome coro | ···················· | ···················· | ····················· | ···················· | ···················· | ···················· |
| 31843 MT370891.1 \|Severe acute respiratory syndrome coro | ···················· | ···················· | ····················· | ···················· | ···················· | ···················· |
| 31844 MT370892.1 \|Severe acute respiratory syndrome coro | ···················· | ···················· | ····················· | ···················· | ···················· | ···················· |
| 31845 MT370894.1 \|Severe acute respiratory syndrome coro | ···················· | ···················· | ····················· | ···················· | ···················· | ···················· |
| 31846 MT370895.1 \|Severe acute respiratory syndrome coro | ···················· | ···················· | ····················· | ···················· | ···················· | ···················· |
| 31847 MT370896.1 \|Severe acute respiratory syndrome coro | ···················· | ···················· | ····················· | ···················· | ···················· | ···················· |
| 31848 MT370897.1 \|Severe acute respiratory syndrome coro | ···················· | ···················· | ····················· | ···················· | ···················· | ···················· |
| 31849 MT370898.1 \|Severe acute respiratory syndrome coro | ···················· | ···················· | ····················· | ···················· | ···················· | ···················· |
| 31850 MT370899.1 \|Severe acute respiratory syndrome coro | ···················· | ···················· | ····················· | ···················· | ···················· | ···················· |
| 31851 MT370901.1 \|Severe acute respiratory syndrome coro | ···················· | ···················· | ····················· | ···················· | ···················· | ···················· |
| 31852 MT370902.1 \|Severe acute respiratory syndrome coro | ···················· | ···················· | ····················· | ···················· | ···················· | ···················· |
| 31853 MT370903.1 \|Severe acute respiratory syndrome coro | ···················· | ·············A······ | ····················· | ···················· | ···················· | ···················· |
| 31854 MT370904.1 \|Severe acute respiratory syndrome coro | ···················· | ···················· | ····················· | ···················· | ···················· | ···················· |
| 31855 MT370905.1 \|Severe acute respiratory syndrome coro | ···················· | ···················· | ····················· | ···················· | ···················· | ···················· |
| 31856 MT370906.1 \|Severe acute respiratory syndrome coro | ···················· | ···················· | ····················· | ···················· | ···················· | ···················· |
| 31857 MT370907.1 \|Severe acute respiratory syndrome coro | ···················· | ···················· | ····················· | ···················· | ···················· | ···················· |
| 31858 MT370908.1 \|Severe acute respiratory syndrome coro | ···················· | ···················· | ····················· | ···················· | ···················· | ···················· |
| 31859 MT370909.1 \|Severe acute respiratory syndrome coro | ···················· | ···················· | ····················· | ···················· | ···················· | ···················· |
| 31860 MT370911.1 \|Severe acute respiratory syndrome coro | ···················· | ···················· | ····················· | ···················· | ···················· | ···················· |
| 31861 MT370912.1 \|Severe acute respiratory syndrome coro | ···················· | ···················· | ····················· | ···················· | ···················· | ···················· |
| 31862 MT370913.1 \|Severe acute respiratory syndrome coro | ···················· | ···················· | ····················· | ···················· | ···················· | ···················· |
| 31863 MT370914.1 \|Severe acute respiratory syndrome coro | ···················· | ···················· | ····················· | ···················· | ···················· | ···················· |
| 31864 MT370915.1 \|Severe acute respiratory syndrome coro | ···················· | ···················· | ····················· | ···················· | ···················· | ···················· |
| 31865 MT370916.1 \|Severe acute respiratory syndrome coro | ···················· | ······A············· | ····················· | ···················· | ···················· | ···················· |
| 31866 MT370917.1 \|Severe acute respiratory syndrome coro | ···················· | ···················· | ····················· | ···················· | ···················· | ···················· |
| 31867 MT370919.1 \|Severe acute respiratory syndrome coro | ···················· | ···················· | ····················· | ···················· | ···················· | ···················· |
| 31868 MT370920.1 \|Severe acute respiratory syndrome coro | ···················· | ···················· | ····················· | ···················· | ···················· | ···················· |
| 31869 MT370925.1 \|Severe acute respiratory syndrome coro | ···················· | ···················· | ····················· | ···················· | ···················· | ···················· |
| 31870 MT370927.1 \|Severe acute respiratory syndrome coro | ···················· | ···················· | ····················· | ···················· | ···················· | ···················· |
| 31871 MT370930.1 \|Severe acute respiratory syndrome coro | ···················· | ···················· | ····················· | ···················· | ···················· | ···················· |
| 31872 MT370931.1 \|Severe acute respiratory syndrome coro | ···················· | ···················· | ····················· | ···················· | ···················· | ···················· |
| 31873 MT370932.1 \|Severe acute respiratory syndrome coro | ···················· | ···················· | ····················· | ···················· | ···················· | ···················· |
| 31874 MT370933.1 \|Severe acute respiratory syndrome coro | ···················· | ···················· | ····················· | ···················· | ···················· | ···················· |
| 31875 MT370934.1 \|Severe acute respiratory syndrome coro | ···················· | ···················· | ····················· | ···················· | ···················· | ···················· |
| 31876 MT370935.1 \|Severe acute respiratory syndrome coro | ···················· | ···················· | ····················· | ···················· | ···················· | ···················· |
| 31877 MT370937.1 \|Severe acute respiratory syndrome coro | ···················· | ···················· | ····················· | ···················· | ···················· | ···················· |
| 31878 MT370939.1 \|Severe acute respiratory syndrome coro | ···················· | ···················· | ····················· | ···················· | ···················· | ···················· |
| 31879 MT370940.1 \|Severe acute respiratory syndrome coro | ···················· | ···················· | ····················· | ···················· | ···················· | ···················· |

31880 MT370942.1 |Severe acute respiratory syndrome coro ···················· ···················· ···················· ···················· ···················· ····················
31881 MT370944.1 |Severe acute respiratory syndrome coro ···················· ···················· ···················· ···················· ···················· ····················
31882 MT370946.1 |Severe acute respiratory syndrome coro ···················· ···················· ···················· ···················· ···················· ····················
31883 MT370947.1 |Severe acute respiratory syndrome coro ···················· ···················· ···················· ···················· ···················· ····················
31884 MT370948.1 |Severe acute respiratory syndrome coro ···················· ···················· ···················· ···················· ···················· ····················
31885 MT370949.1 |Severe acute respiratory syndrome coro ···················· ···················· ···················· ···················· ···················· ····················
31886 MT370950.1 |Severe acute respiratory syndrome coro ···················· ···················· ···················· ···················· ···················· ····················
31887 MT370952.1 |Severe acute respiratory syndrome coro ···················· ···················· ···················· ···················· ···················· ····················
31888 MT370953.1 |Severe acute respiratory syndrome coro ···················· ···················· ···················· ···················· ···················· ····················
31889 MT370954.1 |Severe acute respiratory syndrome coro ···················· ···················· ···················· ···················· ···················· ····················
31890 MT370956.1 |Severe acute respiratory syndrome coro ···················· ···················· ···················· ···················· ···················· ····················
31891 MT370957.1 |Severe acute respiratory syndrome coro ···················· ···················· ···················· ···················· ···················· ····················
31892 MT370958.1 |Severe acute respiratory syndrome coro ···················· ···················· ···················· ···················· ···················· ····················
31893 MT370960.1 |Severe acute respiratory syndrome coro ···················· ···················· ···················· ···················· ···················· ····················
31894 MT370961.1 |Severe acute respiratory syndrome coro ···················· ···················· ···················· ···················· ···················· ····················
31895 MT370962.1 |Severe acute respiratory syndrome coro ···················· ···················· ···················· ···················· ···················· ····················
31896 MT370964.1 |Severe acute respiratory syndrome coro ···················· ···················· ···················· ···················· ···················· ····················
31897 MT370965.1 |Severe acute respiratory syndrome coro ···················· ···················· ···················· ···················· ···················· ····················
31898 MT370966.1 |Severe acute respiratory syndrome coro ···················· ···················· ···················· ···················· ···················· ····················
31899 MT370967.1 |Severe acute respiratory syndrome coro ···················· ···················· ···················· ···················· ···················· ····················
31900 MT370970.1 |Severe acute respiratory syndrome coro ···················· ···················· ···················· ···················· ···················· ····················
31901 MT370971.1 |Severe acute respiratory syndrome coro ···T················ ···················· ···················· ···················· ···················· ····················
31902 MT370972.1 |Severe acute respiratory syndrome coro ···················· ···················· ···················· ···················· ···················· ····················
31903 MT370975.1 |Severe acute respiratory syndrome coro ···················· ···················· ···················· ···················· ···················· ····················
31904 MT370976.1 |Severe acute respiratory syndrome coro ···················· ···················· ···················· ···················· ···················· ····················
31905 MT370977.1 |Severe acute respiratory syndrome coro ···················· ···················· ···················· ···················· ···················· ····················
31906 MT370979.1 |Severe acute respiratory syndrome coro ···················· ···················· ···················· ···················· ···················· ····················
31907 MT370980.1 |Severe acute respiratory syndrome coro ···················· ···················· ···················· ···················· ···················· ····················
31908 MT370981.1 |Severe acute respiratory syndrome coro ···················· ···················· ···················· ···················· ···················· ····················
31909 MT370984.1 |Severe acute respiratory syndrome coro ···················· ···················· ···················· ···················· ···················· ····················
31910 MT370985.1 |Severe acute respiratory syndrome coro ···················· ···················· ···················· ···················· ···················· ····················
31911 MT370986.1 |Severe acute respiratory syndrome coro ···················· ···················· ···················· ···················· ···················· ····················
31912 MT370987.1 |Severe acute respiratory syndrome coro ···················· ···················· ···················· ···················· ···················· ····················
31913 MT370988.1 |Severe acute respiratory syndrome coro ···················· ···················· ···················· ···················· ···················· ····················
31914 MT370991.1 |Severe acute respiratory syndrome coro ···················· ···················· ···················· ···················· ···················· ····················
31915 MT370993.1 |Severe acute respiratory syndrome coro ···················· ···················· ···················· ···················· ···················· ····················
31916 MT370994.1 |Severe acute respiratory syndrome coro ···················· ···················· ···················· ···················· ···················· ····················
31917 MT370996.1 |Severe acute respiratory syndrome coro ···················· ···················· ···················· ···················· ···················· ····················
31918 MT370997.1 |Severe acute respiratory syndrome coro ···················· ···················· ···················· ···················· ···················· ····················
31919 MT370998.1 |Severe acute respiratory syndrome coro ···················· ···················· ···················· ···················· ···················· ····················
31920 MT371000.1 |Severe acute respiratory syndrome coro ···················· ···················· ···················· ···················· ···················· ····················
31921 MT371001.1 |Severe acute respiratory syndrome coro ···················· ···················· ···················· ···················· ···················· ····················
31922 MT371002.1 |Severe acute respiratory syndrome coro ···················· ···················· ···················· ···················· ···················· ····················
31923 MT371006.1 |Severe acute respiratory syndrome coro ···················· ···················· ···················· ···················· ···················· ····················
31924 MT371007.1 |Severe acute respiratory syndrome coro ···················· ···················· ···················· ···················· ···················· ····················
31925 MT371008.1 |Severe acute respiratory syndrome coro ···················· ···················· ···················· ···················· ···················· ····················
31926 MT371009.1 |Severe acute respiratory syndrome coro ···················· ···················· ···················· ···················· ···················· ····················
31927 MT371012.1 |Severe acute respiratory syndrome coro ···················· ···················· ···················· ···················· ···················· ····················
31928 MT371013.1 |Severe acute respiratory syndrome coro ···················· ···················· ···················· ···················· ···················· ····················
31929 MT371015.1 |Severe acute respiratory syndrome coro ···················· ···················· ···················· ···················· ···················· ····················
31930 MT371016.1 |Severe acute respiratory syndrome coro ···················· ···················· ···················· ···················· ···················· ····················
31931 MT371017.1 |Severe acute respiratory syndrome coro ···················· ···················· ···················· ···················· ···················· ····················
31932 MT371018.1 |Severe acute respiratory syndrome coro ···················· ···················· ···················· ···················· ···················· ····················
31933 MT371019.1 |Severe acute respiratory syndrome coro ···················· ···················· ···················· ···················· ···················· ····················
31934 MT371022.1 |Severe acute respiratory syndrome coro ···················· ···················· ···················· ···················· ···················· ····················
31935 MT371023.1 |Severe acute respiratory syndrome coro ···················· ···················· ···················· ···················· ···················· ····················
31936 MT371026.1 |Severe acute respiratory syndrome coro ···················· ···················· ···················· ···················· ···················· ····················
31937 MT371027.1 |Severe acute respiratory syndrome coro ···················· ···················· ···················· ···················· ···················· ····················
31938 MT371028.1 |Severe acute respiratory syndrome coro ···················· ···················· ···················· ···················· ···················· ····················
31939 MT371029.1 |Severe acute respiratory syndrome coro ···················· ···················· ···················· ···················· ···················· ····················
31940 MT371032.1 |Severe acute respiratory syndrome coro ···················· ···················· ···················· ···················· ···················· ····················
31941 MT371033.1 |Severe acute respiratory syndrome coro ···················· ···················· ···················· ···················· ···················· ····················
31942 MT371034.1 |Severe acute respiratory syndrome coro ···················· ···················· ···················· ···················· ···················· ····················
31943 MT371035.1 |Severe acute respiratory syndrome coro ···················· ···················· ···················· ···················· ···················· ····················
31944 MT371036.1 |Severe acute respiratory syndrome coro ···················· ···················· ···················· ···················· ···················· ····················
31945 MT371038.1 |Severe acute respiratory syndrome coro ···················· ·······C············ ···················· ···················· ···················· ····················
31946 MT371047.1 |Severe acute respiratory syndrome coro ···················· ···················· ···················· ···················· ···················· ····················
31947 MT371048.1 |Severe acute respiratory syndrome coro ···················· ···················· ···················· ···················· ···················· ····················
31948 MT371049.1 |Severe acute respiratory syndrome coro ···················· ···················· ···················· ···················· ···················· ····················
31949 MT371050.1 |Severe acute respiratory syndrome coro ···················· ···················· ···················· ···················· ···················· ····················
31950 MT371568.1 |Severe acute respiratory syndrome coro ···················· ···················· ···················· ···················· ···················· ····················
31951 MT371569.1 |Severe acute respiratory syndrome coro ···················· ···················· ···················· ···················· ···················· ···············A····
31952 MT371570.1 |Severe acute respiratory syndrome coro ···················· ···················· ···················· ···················· ···················· ····················
31953 MT371571.1 |Severe acute respiratory syndrome coro ···················· ···················· ···················· ···················· ···················· ····················
31954 MT371572.1 |Severe acute respiratory syndrome coro ···················· ···················· ···················· ···················· ···················· ···············A····
31955 MT371573.1 |Severe acute respiratory syndrome coro ···················· ···················· ···················· ···················· ···················· ····················
31956 MT371574.1 |Severe acute respiratory syndrome coro ···················· ···················· ···················· ···················· ···················· ····················
31957 MT372480.1 |Severe acute respiratory syndrome coro ···················· ···················· ···················· ···················· ···················· ····················

| | | | | | | | | | |
|---|---|---|---|---|---|---|---|---|---|
| 31958 | MT372481.1 | \|Severe acute respiratory syndrome coro | ···················· | ···················· | ···················· | ···················· | ···················· | ···················· | |
| 31959 | MT372482.1 | \|Severe acute respiratory syndrome coro | ···················· | ···················· | ···················· | ···················· | ···················· | ·T··ATC·T·A········· | 1 |
| 31960 | LC542976.1 | \|Severe acute respiratory syndrome coro | ···················· | ···················· | ···················· | ···················· | ···················· | ···················· | |
| 31961 | LC542809.1 | \|Severe acute respiratory syndrome coro | ···················· | ···················· | ···················· | ···················· | ···················· | ···················· | |
| 31962 | MT114412.1 | \|Severe acute respiratory syndrome coro | ···················· | ···················· | ···················· | ···················· | ···················· | ···················· | |
| 31963 | MT114413.1 | \|Severe acute respiratory syndrome coro | ···················· | ···················· | ···················· | ···················· | ···················· | ···················· | |
| 31964 | MT114414.1 | \|Severe acute respiratory syndrome coro | ···················· | ···················· | ···················· | ···················· | ···················· | ···················· | |
| 31965 | MT114415.1 | \|Severe acute respiratory syndrome coro | ···················· | ···················· | ···················· | ···················· | ···················· | ···················· | |
| 31966 | MT114416.1 | \|Severe acute respiratory syndrome coro | ···················· | ···················· | ···················· | ···················· | ···················· | ···················· | |
| 31967 | MT114417.1 | \|Severe acute respiratory syndrome coro | ···················· | ···················· | ···················· | ···················· | ···················· | ···················· | |
| 31968 | MT114418.1 | \|Severe acute respiratory syndrome coro | ···················· | ···················· | ···················· | ···················· | ···················· | ···················· | |
| 31969 | MT114419.1 | \|Severe acute respiratory syndrome coro | ···················· | ···················· | ···················· | ···················· | ···················· | ···················· | |
| 31970 | MT230904.1 | \|Severe acute respiratory syndrome coro | ···················· | ···················· | ···················· | ···················· | ···················· | ···················· | |
| 31971 | MT358401.1 | \|Severe acute respiratory syndrome coro | ···················· | ···················· | ···················· | ···················· | ···················· | ···················· | |
| 31972 | MT358402.1 | \|Severe acute respiratory syndrome coro | ···················· | ···················· | ···················· | ···················· | ···················· | ···················· | |
| 31973 | MT358637.1 | \|Severe acute respiratory syndrome coro | ···················· | ···················· | ···················· | ···················· | ···················· | ···················· | |
| 31974 | MT358644.1 | \|Severe acute respiratory syndrome coro | ···················· | ···················· | ···················· | ···················· | ···················· | ···················· | |
| 31975 | MT358645.1 | \|Severe acute respiratory syndrome coro | ···················· | ···················· | ···················· | ···················· | ···················· | ···················· | |
| 31976 | MT358646.1 | \|Severe acute respiratory syndrome coro | ···················· | ···················· | ···················· | ···················· | ···················· | ···················· | |
| 31977 | MT358647.1 | \|Severe acute respiratory syndrome coro | ···················· | ···················· | ···················· | ···················· | ···················· | ···················· | |
| 31978 | MT358648.1 | \|Severe acute respiratory syndrome coro | ···················· | ···················· | ···················· | ···················· | ···················· | ···················· | |
| 31979 | MT358649.1 | \|Severe acute respiratory syndrome coro | ···················· | ···················· | ···················· | ···················· | ···················· | ···················· | |
| 31980 | MT358650.1 | \|Severe acute respiratory syndrome coro | ···················· | ···················· | ···················· | ···················· | ···················· | ···················· | |
| 31981 | MT358651.1 | \|Severe acute respiratory syndrome coro | ···················· | ···················· | ···················· | ···················· | ···················· | ···················· | |
| 31982 | MT358652.1 | \|Severe acute respiratory syndrome coro | ···················· | ···················· | ···················· | ···················· | ···················· | ···················· | |
| 31983 | MT358653.1 | \|Severe acute respiratory syndrome coro | ···················· | ···················· | ···················· | ···················· | ···················· | ···················· | |
| 31984 | MT358654.1 | \|Severe acute respiratory syndrome coro | ···················· | ···················· | ···················· | ···················· | ···················· | ···················· | |
| 31985 | MT358655.1 | \|Severe acute respiratory syndrome coro | ···················· | ···················· | ···················· | ···················· | ···················· | ···················· | |
| 31986 | MT358656.1 | \|Severe acute respiratory syndrome coro | ···················· | ···················· | ···················· | ···················· | ···················· | ···················· | |
| 31987 | MT358657.1 | \|Severe acute respiratory syndrome coro | ···················· | ···················· | ···················· | ···················· | ···················· | ···················· | |
| 31988 | MT358658.1 | \|Severe acute respiratory syndrome coro | ···················· | ···················· | ···················· | ···················· | ···················· | ···················· | |
| 31989 | MT358659.1 | \|Severe acute respiratory syndrome coro | ···················· | ···················· | ···················· | ···················· | ···················· | ···················· | |
| 31990 | MT358660.1 | \|Severe acute respiratory syndrome coro | ···················· | ···················· | ···················· | ···················· | ···················· | ···················· | |
| 31991 | MT358661.1 | \|Severe acute respiratory syndrome coro | ···················· | ···················· | ···················· | ···················· | ···················· | ···················· | |
| 31992 | MT358662.1 | \|Severe acute respiratory syndrome coro | ···················· | ···················· | ···················· | ···················· | ···················· | ···················· | |
| 31993 | MT358663.1 | \|Severe acute respiratory syndrome coro | ···················· | ···················· | ···················· | ···················· | ···················· | ···················· | |
| 31994 | MT358664.1 | \|Severe acute respiratory syndrome coro | ···················· | ···················· | ···················· | ···················· | ···················· | ···················· | |
| 31995 | MT358665.1 | \|Severe acute respiratory syndrome coro | ···················· | ···················· | ···················· | ···················· | ···················· | ···················· | |
| 31996 | MT358666.1 | \|Severe acute respiratory syndrome coro | ···················· | ···················· | ···················· | ···················· | ···················· | ···················· | |
| 31997 | MT358667.1 | \|Severe acute respiratory syndrome coro | ···················· | ···················· | ···················· | ···················· | ···················· | ···················· | |
| 31998 | MT358668.1 | \|Severe acute respiratory syndrome coro | ···················· | ···················· | ···················· | ···················· | ···················· | ···················· | |
| 31999 | MT358669.1 | \|Severe acute respiratory syndrome coro | ···················· | ···················· | ···················· | ···················· | ···················· | ···················· | |
| 32000 | MT358670.1 | \|Severe acute respiratory syndrome coro | ···················· | ···················· | ···················· | ···················· | ···················· | ···················· | |
| 32001 | MT358671.1 | \|Severe acute respiratory syndrome coro | ···················· | ···················· | ···················· | ···················· | ···················· | ···················· | |
| 32002 | MT358672.1 | \|Severe acute respiratory syndrome coro | ···················· | ···················· | ···················· | ···················· | ···················· | ···················· | |
| 32004 | MT358674.1 | \|Severe acute respiratory syndrome coro | ···················· | ···················· | ···················· | ···················· | ···················· | ···················· | |
| 32005 | MT358675.1 | \|Severe acute respiratory syndrome coro | ···················· | ···················· | ···················· | ···················· | ···················· | ···················· | |
| 32006 | MT358677.1 | \|Severe acute respiratory syndrome coro | ···················· | ···················· | ···················· | ···················· | ···················· | ···················· | |
| 32007 | MT358678.1 | \|Severe acute respiratory syndrome coro | ···················· | ···················· | ···················· | ···················· | ···················· | ···················· | |
| 32008 | MT358679.1 | \|Severe acute respiratory syndrome coro | ···················· | ···················· | ···················· | ···················· | ···················· | ···················· | |
| 32009 | MT358680.1 | \|Severe acute respiratory syndrome coro | ···················· | ···················· | ···················· | ···················· | ···················· | ···················· | |
| 32010 | MT358681.1 | \|Severe acute respiratory syndrome coro | ···················· | ···················· | ···················· | ···················· | ···················· | ···················· | |
| 32011 | MT358682.1 | \|Severe acute respiratory syndrome coro | ···················· | ···················· | ···················· | ···················· | ···················· | ···················· | |
| 32012 | MT358683.1 | \|Severe acute respiratory syndrome coro | ···················· | ···················· | ···················· | ···················· | ···················· | ···················· | |
| 32013 | MT358684.1 | \|Severe acute respiratory syndrome coro | ···················· | ···················· | ···················· | ···················· | ···················· | ···················· | |
| 32014 | MT358685.1 | \|Severe acute respiratory syndrome coro | ···················· | ···················· | ···················· | ···················· | ···················· | ···················· | |
| 32015 | MT358687.1 | \|Severe acute respiratory syndrome coro | ···················· | ···················· | ···················· | ···················· | ···················· | ···················· | |
| 32016 | MT358688.1 | \|Severe acute respiratory syndrome coro | ···················· | ···················· | ···················· | ···················· | ···················· | ···················· | |
| 32017 | MT358689.1 | \|Severe acute respiratory syndrome coro | ···················· | ···················· | ···················· | ···················· | ···················· | ···················· | |
| 32018 | MT358690.1 | \|Severe acute respiratory syndrome coro | ···················· | ···················· | ···················· | ···················· | ···················· | ···················· | |
| 32019 | MT358691.1 | \|Severe acute respiratory syndrome coro | ···················· | ···················· | ···················· | ···················· | ···················· | ···················· | |
| 32020 | MT358692.1 | \|Severe acute respiratory syndrome coro | ···················· | ···················· | ···················· | ···················· | ···················· | ···················· | |
| 32021 | MT358693.1 | \|Severe acute respiratory syndrome coro | ···················· | ···················· | ···················· | ···················· | ···················· | ···················· | |
| 32022 | MT358694.1 | \|Severe acute respiratory syndrome coro | ···················· | ···················· | ···················· | ···················· | ···················· | ···················· | |
| 32023 | MT358695.1 | \|Severe acute respiratory syndrome coro | ···················· | ···················· | ···················· | ···················· | ···················· | ···················· | |
| 32024 | MT358696.1 | \|Severe acute respiratory syndrome coro | ···················· | ···················· | ···················· | ···················· | ···················· | ···················· | |
| 32025 | MT358697.1 | \|Severe acute respiratory syndrome coro | ···················· | ···················· | ···················· | ···················· | ···················· | ···················· | |
| 32026 | MT358698.1 | \|Severe acute respiratory syndrome coro | ···················· | ···················· | ···················· | ···················· | ···················· | ···················· | |
| 32027 | MT358699.1 | \|Severe acute respiratory syndrome coro | ···················· | ···················· | ···················· | ···················· | ···················· | ···················· | |
| 32028 | MT358700.1 | \|Severe acute respiratory syndrome coro | ···················· | ···················· | ···················· | ···················· | ···················· | ···················· | |
| 32029 | MT358701.1 | \|Severe acute respiratory syndrome coro | ···················· | ···················· | ···················· | ···················· | ···················· | ···················· | |
| 32030 | MT358702.1 | \|Severe acute respiratory syndrome coro | ···················· | ···················· | ···················· | ···················· | ···················· | ···················· | |
| 32031 | MT358703.1 | \|Severe acute respiratory syndrome coro | ···················· | ···················· | ···················· | ···················· | ···················· | ···················· | |
| 32032 | MT358705.1 | \|Severe acute respiratory syndrome coro | ···················· | ···················· | ···················· | ···················· | ···················· | ···················· | |
| 32033 | MT358706.1 | \|Severe acute respiratory syndrome coro | ···················· | ···················· | ···················· | ···················· | ···················· | ···················· | |
| 32034 | MT358707.1 | \|Severe acute respiratory syndrome coro | ···················· | ···················· | ···················· | ···················· | ···················· | ···················· | |
| 32035 | MT358708.1 | \|Severe acute respiratory syndrome coro | ···················· | ···················· | ···················· | ···················· | ···················· | ···················· | |
| 32036 | MT358709.1 | \|Severe acute respiratory syndrome coro | ···················· | ···················· | ···················· | ···················· | ···················· | ···················· | |

| | | | | | | | | | |
|---|---|---|---|---|---|---|---|---|---|
| 32037 | MT358710.1 | |Severe acute respiratory syndrome coro | ···················· | ···················· | ···················· | ···················· | ···················· | ···················· | |
| 32038 | MT358711.1 | |Severe acute respiratory syndrome coro | ···················· | ···················· | ···················· | ···················· | ···················· | ···················· | |
| 32039 | MT358712.1 | |Severe acute respiratory syndrome coro | ···················· | ···················· | ···················· | ···················· | ···················· | ···················· | |
| 32040 | MT358713.1 | |Severe acute respiratory syndrome coro | ···················· | ···················· | ···················· | ···················· | ···················· | ···················· | |
| 32041 | MT358714.1 | |Severe acute respiratory syndrome coro | ···················· | ···················· | ···················· | ···················· | ···················· | ···················· | |
| 32042 | MT358715.1 | |Severe acute respiratory syndrome coro | ···················· | ···················· | ···················· | ···················· | ···················· | ···················· | |
| 32043 | MT358716.1 | |Severe acute respiratory syndrome coro | ···················· | ···················· | ···················· | ···················· | ···················· | ···················· | |
| 32044 | MT358717.1 | |Severe acute respiratory syndrome coro | ···················· | ···················· | ···················· | ···················· | ···················· | ···················· | |
| 32045 | MT358718.1 | |Severe acute respiratory syndrome coro | ···················· | ···················· | ···················· | ···················· | ···················· | ···················· | |
| 32046 | MT358719.1 | |Severe acute respiratory syndrome coro | ···················· | ···················· | ···················· | ···················· | ···················· | ···················· | |
| 32047 | MT358720.1 | |Severe acute respiratory syndrome coro | ···················· | ···················· | ···················· | ···················· | ···················· | ···················· | |
| 32048 | MT358721.1 | |Severe acute respiratory syndrome coro | ···················· | ···················· | ···················· | ···················· | ···················· | ···················· | |
| 32049 | MT358722.1 | |Severe acute respiratory syndrome coro | ···················· | ···················· | ···················· | ···················· | ···················· | ···················· | |
| 32050 | MT358723.1 | |Severe acute respiratory syndrome coro | ···················· | ···················· | ···················· | ···················· | ···················· | ···················· | |
| 32051 | MT358724.1 | |Severe acute respiratory syndrome coro | ···················· | ···················· | ···················· | ···················· | ···················· | ···················· | |
| 32052 | MT358725.1 | |Severe acute respiratory syndrome coro | ···················· | ···················· | ···················· | ···················· | ···················· | ···················· | |
| 32053 | MT358726.1 | |Severe acute respiratory syndrome coro | ···················· | ···················· | ···················· | ···················· | ···················· | ···················· | |
| 32054 | MT358727.1 | |Severe acute respiratory syndrome coro | ···················· | ···················· | ···················· | ···················· | ···················· | ···················· | |
| 32055 | MT358728.1 | |Severe acute respiratory syndrome coro | ···················· | ···················· | ···················· | ···················· | ···················· | ···················· | |
| 32056 | MT358729.1 | |Severe acute respiratory syndrome coro | ···················· | ···················· | ···················· | ···················· | ···················· | ···················· | |
| 32057 | MT358730.1 | |Severe acute respiratory syndrome coro | ···················· | ···················· | ···················· | ···················· | ···················· | ···················· | |
| 32058 | MT358731.1 | |Severe acute respiratory syndrome coro | ···················· | ···················· | ···················· | ···················· | ···················· | ···················· | |
| 32059 | MT358732.1 | |Severe acute respiratory syndrome coro | ···················· | ···················· | ···················· | ···················· | ···················· | ···················· | |
| 32060 | MT358733.1 | |Severe acute respiratory syndrome coro | ···················· | ···················· | ···················· | ···················· | ···················· | ···················· | |
| 32061 | MT358734.1 | |Severe acute respiratory syndrome coro | ···················· | ···················· | ···················· | ···················· | ···················· | ···················· | |
| 32062 | MT358735.1 | |Severe acute respiratory syndrome coro | ···················· | ···················· | ···················· | ···················· | ···················· | ···················· | |
| 32063 | MT358736.1 | |Severe acute respiratory syndrome coro | ···················· | ···················· | ···················· | ···················· | ···················· | ···················· | |
| 32064 | MT358737.1 | |Severe acute respiratory syndrome coro | ···················· | ···················· | ···················· | ···················· | ···················· | ···················· | |
| 32065 | MT358738.1 | |Severe acute respiratory syndrome coro | ···················· | ···················· | ···················· | ···················· | ···················· | ···················· | |
| 32066 | MT358739.1 | |Severe acute respiratory syndrome coro | ···················· | ···················· | ···················· | ···················· | ···················· | ···················· | |
| 32067 | MT358740.1 | |Severe acute respiratory syndrome coro | ···················· | ···················· | ···················· | ···················· | ···················· | ···················· | |
| 32068 | MT358741.1 | |Severe acute respiratory syndrome coro | ···················· | ···················· | ···················· | ···················· | ···················· | ···················· | |
| 32069 | MT358742.1 | |Severe acute respiratory syndrome coro | ···················· | ···················· | ···················· | ···················· | ···················· | ···················· | |
| 32070 | MT358743.1 | |Severe acute respiratory syndrome coro | ···················· | ···················· | ···················· | ···················· | ···················· | ···················· | |
| 32071 | MT358744.1 | |Severe acute respiratory syndrome coro | ···················· | ···················· | ···················· | ···················· | ···················· | ···················· | |
| 32072 | MT358745.1 | |Severe acute respiratory syndrome coro | ···················· | ···················· | ···················· | ···················· | ···················· | ··AT····C··A········ | 1 |
| 32073 | MT358746.1 | |Severe acute respiratory syndrome coro | ···················· | ···················· | ···················· | ···················· | ···················· | ···················· | |
| 32074 | MT358747.1 | |Severe acute respiratory syndrome coro | ···················· | ···················· | ···················· | ···················· | ···················· | ···················· | |
| 32075 | MT358748.1 | |Severe acute respiratory syndrome coro | ···················· | ···················· | ···················· | ···················· | ···················· | ···················· | |
| 32076 | MT359865.1 | |Severe acute respiratory syndrome coro | ···················· | ···················· | ···················· | ···················· | ···················· | ···················· | |
| 32077 | MT359866.1 | |Severe acute respiratory syndrome coro | ···················· | ···················· | ···················· | ···················· | ···················· | ···················· | |
| 32078 | MT350236.1 | |Severe acute respiratory syndrome coro | ···················· | ···················· | ···················· | ···················· | ···················· | ···················· | |
| 32079 | MT350237.1 | |Severe acute respiratory syndrome coro | ···················· | ···················· | ···················· | ···················· | ···················· | ···················· | |
| 32080 | MT350238.1 | |Severe acute respiratory syndrome coro | ···················· | ···················· | ···················· | ···················· | ···················· | ···················· | |
| 32081 | MT350239.1 | |Severe acute respiratory syndrome coro | ···················· | ···················· | ···················· | ···················· | ···················· | ···················· | |
| 32082 | MT350240.1 | |Severe acute respiratory syndrome coro | ···················· | ···················· | ···················· | ···················· | ···················· | ···················· | |
| 32083 | MT350241.1 | |Severe acute respiratory syndrome coro | ···················· | ···················· | ···················· | ···················· | ···················· | ···················· | |
| 32084 | MT350242.1 | |Severe acute respiratory syndrome coro | ···················· | ···················· | ···················· | ···················· | ···················· | ···················· | |
| 32085 | MT350243.1 | |Severe acute respiratory syndrome coro | ···················· | ···················· | ···················· | ···················· | ···················· | ···················· | |
| 32086 | MT350244.1 | |Severe acute respiratory syndrome coro | ···················· | ···················· | ···················· | ···················· | ···················· | ···················· | |
| 32087 | MT350245.1 | |Severe acute respiratory syndrome coro | ···················· | ···················· | ···················· | ···················· | ···················· | ···················· | |
| 32088 | MT350246.1 | |Severe acute respiratory syndrome coro | ···················· | ···················· | ···················· | ··············A····· | ···················· | ···················· | |
| 32089 | MT350247.1 | |Severe acute respiratory syndrome coro | ···················· | ···················· | ···················· | ···················· | ···················· | ···················· | |
| 32090 | MT350248.1 | |Severe acute respiratory syndrome coro | ···················· | ···················· | ···················· | ···················· | ···················· | ···················· | |
| 32091 | MT350249.1 | |Severe acute respiratory syndrome coro | ···················· | ···················· | ···················· | ···················· | ···················· | ···················· | |
| 32092 | MT350250.1 | |Severe acute respiratory syndrome coro | ···················· | ···················· | ···················· | ···················· | ···················· | ···················· | |
| 32093 | MT350251.1 | |Severe acute respiratory syndrome coro | ···················· | ···················· | ···················· | ···················· | ···················· | ···················· | |
| 32094 | MT350252.1 | |Severe acute respiratory syndrome coro | ···················· | ···················· | ···················· | ···················· | ···················· | ···················· | |
| 32095 | MT350253.1 | |Severe acute respiratory syndrome coro | ···················· | ···················· | ···················· | ···················· | ···················· | ···················· | |
| 32096 | MT350254.1 | |Severe acute respiratory syndrome coro | ···················· | ···················· | ···················· | ···················· | ···················· | ···················· | |
| 32097 | MT350255.1 | |Severe acute respiratory syndrome coro | ···················· | ···················· | ···················· | ···················· | ···················· | ···················· | |
| 32098 | MT350256.1 | |Severe acute respiratory syndrome coro | ···················· | ···················· | ···················· | ···················· | ···················· | ···················· | |
| 32099 | MT350257.1 | |Severe acute respiratory syndrome coro | ···················· | ···················· | ···················· | ···················· | ···················· | ···················· | |
| 32100 | MT350263.1 | |Severe acute respiratory syndrome coro | ···················· | ···················· | ···················· | ···················· | ···················· | ···················· | |
| 32101 | MT350264.1 | |Severe acute respiratory syndrome coro | ···················· | ···················· | ···················· | ···················· | ···················· | ···················· | |
| 32102 | MT350265.1 | |Severe acute respiratory syndrome coro | ···················· | ···················· | ···················· | ···················· | ···················· | ···················· | |
| 32103 | MT350266.1 | |Severe acute respiratory syndrome coro | ···················· | ···················· | ···················· | ···················· | ···················· | ···················· | |
| 32104 | MT350267.1 | |Severe acute respiratory syndrome coro | ···················· | ···················· | ···················· | ···················· | ···················· | ···················· | |
| 32105 | MT350268.1 | |Severe acute respiratory syndrome coro | ···················· | ···················· | ···················· | ···················· | ···················· | ···················· | |
| 32106 | MT350269.1 | |Severe acute respiratory syndrome coro | ···················· | ···················· | ···················· | ···················· | ···················· | ···················· | |
| 32107 | MT350270.1 | |Severe acute respiratory syndrome coro | ···················· | ···················· | ···················· | ···················· | ···················· | ···················· | |
| 32108 | MT350271.1 | |Severe acute respiratory syndrome coro | ···················· | ···················· | ···················· | ···················· | ···················· | ···················· | |
| 32109 | MT350272.1 | |Severe acute respiratory syndrome coro | ···················· | ···················· | ···················· | ···················· | ···················· | ···················· | |
| 32110 | MT350273.1 | |Severe acute respiratory syndrome coro | ···················· | ···················· | ···················· | ···················· | ···················· | ···················· | |
| 32111 | MT350274.1 | |Severe acute respiratory syndrome coro | ···················· | ···················· | ···················· | ···················· | ···················· | ···················· | |
| 32112 | MT350275.1 | |Severe acute respiratory syndrome coro | ···················· | ···················· | ···················· | ···················· | ···················· | ···················· | |
| 32113 | MT350276.1 | |Severe acute respiratory syndrome coro | ···················· | ···················· | ···················· | ···················· | ···················· | ···················· | |
| 32114 | MT350277.1 | |Severe acute respiratory syndrome coro | ···················· | ···················· | ···················· | ···················· | ···················· | ···················· | |

```
32115 MT350278.1 |Severe acute respiratory syndrome coro    ....................  ....................  ....................  ....................  ....................  ....................
32116 MT350279.1 |Severe acute respiratory syndrome coro    ....................  ....................  ....................  ....................  ....................  ....................
32117 MT350280.1 |Severe acute respiratory syndrome coro    ....................  ....................  ....................  ....................  ....................  ....................
32118 MT350282.1 |Severe acute respiratory syndrome coro    ....................  ....................  ....................  ....................  ....................  ....................
32119 MT344944.1 |Severe acute respiratory syndrome coro    ....................  ....................  ....................  ....................  ....................  ....................
32120 MT344945.1 |Severe acute respiratory syndrome coro    ....................  ....................  ....................  ....................  ....................  ....................
32121 MT344946.1 |Severe acute respiratory syndrome coro    ....................  ....................  ....................  ....................  ....................  ....................
32122 MT344947.1 |Severe acute respiratory syndrome coro    ....................  ....................  ....................  ....................  ....................  ....................
32123 MT344948.1 |Severe acute respiratory syndrome coro    ....................  .A..................  ....................  ....................  ....................  ....................
32124 MT344949.1 |Severe acute respiratory syndrome coro    ....................  ....................  ....................  ....................  ....................  ....................
32125 MT344950.1 |Severe acute respiratory syndrome coro    ....................  ....................  ....................  ....................  ....................  ....................
32126 MT344951.1 |Severe acute respiratory syndrome coro    ....................  ....................  ....................  ....................  ....................  ....................
32127 MT344952.1 |Severe acute respiratory syndrome coro    ....................  ....................  ....................  ....................  ....................  ....................
32128 MT344953.1 |Severe acute respiratory syndrome coro    ....................  ....................  ....................  ....................  ....................  ....................
32129 MT344954.1 |Severe acute respiratory syndrome coro    ....................  ....................  ....................  ....................  ....................  ....................
32130 MT344955.1 |Severe acute respiratory syndrome coro    ....................  ....................  ....................  ....................  ....................  ....................
32131 MT344956.1 |Severe acute respiratory syndrome coro    ....................  ....................  ....................  ....................  ....................  ....................
32132 MT344957.1 |Severe acute respiratory syndrome coro    ....................  ....................  ....................  ....................  ....................  ....................
32133 MT344958.1 |Severe acute respiratory syndrome coro    ....................  ....................  ....................  ....................  ....................  ....................
32134 MT344959.1 |Severe acute respiratory syndrome coro    ....................  ....................  ....................  ....................  ....................  ....................
32135 MT344960.1 |Severe acute respiratory syndrome coro    ....................  ....................  ....................  ....................  ....................  ....................
32136 MT344961.1 |Severe acute respiratory syndrome coro    ....................  ....................  ....................  ....................  ....................  ....................
32137 MT344962.1 |Severe acute respiratory syndrome coro    ....................  ....................  ....................  ....................  ....................  ....................
32138 MT344963.1 |Severe acute respiratory syndrome coro    ....................  ....................  ....................  ....................  ....................  ....................
32139 MT345798.1 |Severe acute respiratory syndrome coro    ....................  ....................  ....................  ....................  ....................  ....................
32140 MT345799.1 |Severe acute respiratory syndrome coro    ....................  ....................  ....................  ....................  ....................  ....................
32141 MT345801.1 |Severe acute respiratory syndrome coro    ....................  ....................  ....................  ....................  ....................  ....................
32142 MT345802.1 |Severe acute respiratory syndrome coro    ....................  ....................  ....................  ....................  ....................  ....................
32143 MT345803.1 |Severe acute respiratory syndrome coro    ....................  ....................  ....................  ....................  ....................  ....................
32144 MT345805.1 |Severe acute respiratory syndrome coro    ....................  ....................  ....................  ....................  ....................  ....................
32145 MT345806.1 |Severe acute respiratory syndrome coro    ....................  ....................  ....................  ....................  ....................  ....................
32146 MT345808.1 |Severe acute respiratory syndrome coro    ....................  ....................  ....................  ....................  ....................  ....................
32147 MT345809.1 |Severe acute respiratory syndrome coro    ....................  ....................  ....................  ....................  ....................  ....................
32148 MT345811.1 |Severe acute respiratory syndrome coro    ....................  ....................  ....................  ....................  ....................  ....................
32149 MT345812.1 |Severe acute respiratory syndrome coro    ....................  ....................  ....................  ....................  ....................  ....................
32150 MT345814.1 |Severe acute respiratory syndrome coro    ....................  ....................  ....................  ....................  ....................  ....................
32151 MT345815.1 |Severe acute respiratory syndrome coro    ....................  ....................  ....................  ....................  ....................  ....................
32152 MT345816.1 |Severe acute respiratory syndrome coro    ....................  ....................  ....................  ....................  ....................  ....................
32153 MT345817.1 |Severe acute respiratory syndrome coro    ....................  ....................  ....................  ....................  ....................  ....................
32154 MT345819.1 |Severe acute respiratory syndrome coro    ....................  ....................  ....................  ....................  ....................  ....................
32155 MT345821.1 |Severe acute respiratory syndrome coro    ....................  ....................  ....................  ....................  ....................  ....................
32156 MT345822.1 |Severe acute respiratory syndrome coro    ....................  ....................  ....................  ....................  ....................  ....................
32157 MT345823.1 |Severe acute respiratory syndrome coro    ....................  ....................  ....................  ....................  ....................  ....................
32158 MT345824.1 |Severe acute respiratory syndrome coro    ....................  ....................  ....................  ....................  ....................  ....................
32159 MT345825.1 |Severe acute respiratory syndrome coro    ....................  ....................  ....................  ....................  ....................  ....................
32160 MT345826.1 |Severe acute respiratory syndrome coro    ....................  ....................  ....................  ....................  ....................  ....................
32161 MT345827.1 |Severe acute respiratory syndrome coro    ....................  ....................  ....................  ....................  ....................  ....................
32162 MT345828.1 |Severe acute respiratory syndrome coro    ....................  ....................  ....................  ....................  ....................  ....................
32163 MT345829.1 |Severe acute respiratory syndrome coro    ....................  ....................  ....................  ....................  ....................  ....................
32164 MT345830.1 |Severe acute respiratory syndrome coro    ....................  ....................  ....................  ....................  ....................  ....................
32165 MT345832.1 |Severe acute respiratory syndrome coro    ....................  ....................  ....................  ....................  ....................  ....................
32166 MT345833.1 |Severe acute respiratory syndrome coro    ....................  ....................  ....................  ....................  ....................  ....................
32167 MT345834.1 |Severe acute respiratory syndrome coro    ....................  ....................  ....................  ....................  ....................  ....................
32168 MT345835.1 |Severe acute respiratory syndrome coro    ....................  ....................  ....................  ....................  ....................  ....................
32169 MT345836.1 |Severe acute respiratory syndrome coro    ....................  ....................  ....................  ....................  ....................  ....................
32170 MT345838.1 |Severe acute respiratory syndrome coro    ....................  ....................  ....................  ....................  ....................  ....................
32171 MT345840.1 |Severe acute respiratory syndrome coro    ....................  ....................  ....................  ....................  ....................  ....................
32172 MT345841.1 |Severe acute respiratory syndrome coro    ....................  ....................  ....................  ....................  ....................  ....................
32173 MT345842.1 |Severe acute respiratory syndrome coro    ....................  ....................  ....................  ....................  ....................  ....................
32174 MT345843.1 |Severe acute respiratory syndrome coro    ....................  ....................  ....................  ....................  ....................  ....................
32175 MT345844.1 |Severe acute respiratory syndrome coro    ....................  ....................  ....................  ....................  ....................  ....................
32176 MT345846.1 |Severe acute respiratory syndrome coro    ....................  ....................  ....................  ....................  ....................  ....................
32177 MT345847.1 |Severe acute respiratory syndrome coro    ....................  ....................  ....................  ....................  ....................  ....................
32178 MT345848.1 |Severe acute respiratory syndrome coro    ....................  ....................  ....................  ....................  ....................  ....................
32179 MT345849.1 |Severe acute respiratory syndrome coro    ....................  ....................  ....................  ....................  ....................  ....................
32181 MT345851.1 |Severe acute respiratory syndrome coro    ....................  ....................  ....................  ....................  ....................  ....................
32182 MT345852.1 |Severe acute respiratory syndrome coro    ....................  ....................  ....................  ....................  ....................  ....................
32183 MT345853.1 |Severe acute respiratory syndrome coro    ....................  ....................  ....................  ....................  ....................  ....................
32184 MT345854.1 |Severe acute respiratory syndrome coro    ....................  ....................  ....................  ....................  ....................  ....................
32185 MT345855.1 |Severe acute respiratory syndrome coro    ....................  ....................  ....................  ....................  ....................  ....................
32186 MT345856.1 |Severe acute respiratory syndrome coro    ....................  ....................  ....................  ....................  ....................  ....................
32187 MT345857.1 |Severe acute respiratory syndrome coro    ....................  ....................  ....................  ....................  ....................  ....................
32188 MT345858.1 |Severe acute respiratory syndrome coro    ....................  ....................  ....................  ....................  ....................  ....................
32189 MT345859.1 |Severe acute respiratory syndrome coro    ....................  ....................  ....................  ....................  ....................  ....................
32190 MT345860.1 |Severe acute respiratory syndrome coro    ....................  ....................  ....................  ....................  ....................  ....................
32191 MT345861.1 |Severe acute respiratory syndrome coro    ....................  ....................  ....................  ....................  ....................  ....................
32192 MT345862.1 |Severe acute respiratory syndrome coro    ....................  ....................  ....................  ....................  ....................  ....................
32193 MT345865.1 |Severe acute respiratory syndrome coro    ....................  ....................  ....................  ....................  ....................  ....................
```

```
32194 MT345866.1 |Severe acute respiratory syndrome coro    ....................  ....................  .....................  ....................  ....................  ....................
32195 MT345867.1 |Severe acute respiratory syndrome coro    ....................  ....................  .....................  ....................  ....................  ....................
32196 MT345868.1 |Severe acute respiratory syndrome coro    ....................  ....................  .....................  ....................  ....................  ....................
32197 MT345869.1 |Severe acute respiratory syndrome coro    ....................  ....................  .....................  ....................  ....................  ....................
32198 MT345870.1 |Severe acute respiratory syndrome coro    ....................  ....................  .....................  ....................  ....................  ....................
32199 MT345871.1 |Severe acute respiratory syndrome coro    ....................  ....................  .....................  ....................  ....................  ....................
32200 MT345872.1 |Severe acute respiratory syndrome coro    ....................  ....................  .....................  ....................  ....................  ....................
32201 MT345873.1 |Severe acute respiratory syndrome coro    ....................  ....................  .....................  ....................  ....................  ....................
32202 MT345874.1 |Severe acute respiratory syndrome coro    ....................  ....................  .....................  ....................  ....................  ....................
32203 MT345875.1 |Severe acute respiratory syndrome coro    ....................  ....................  .....................  ....................  ....................  ....................
32204 MT345876.1 |Severe acute respiratory syndrome coro    ....................  ....................  .....................  ....................  ....................  ....................
32205 MT345877.1 |Severe acute respiratory syndrome coro    ....................  ....................  .....................  ....................  ....................  ....................
32206 MT345878.1 |Severe acute respiratory syndrome coro    ....................  ....................  .....................  ....................  ....................  ....................
32207 MT345879.1 |Severe acute respiratory syndrome coro    ....................  ....................  .....................  ....................  ....................  ....................
32208 MT345880.1 |Severe acute respiratory syndrome coro    ....................  ....................  .....................  ....................  ....................  ....................
32209 MT345881.1 |Severe acute respiratory syndrome coro    ....................  ....................  .....................  ....................  ....................  ....................
32210 MT345882.1 |Severe acute respiratory syndrome coro    ....................  ....................  .....................  ....................  ....................  ....................
32211 MT345883.1 |Severe acute respiratory syndrome coro    ....................  ....................  .....................  ....................  ....................  ....................
32212 MT345884.1 |Severe acute respiratory syndrome coro    ....................  ....................  .....................  ....................  ....................  ....................
32213 MT345885.1 |Severe acute respiratory syndrome coro    ....................  ....................  .....................  ....................  ....................  ....................
32214 MT345886.1 |Severe acute respiratory syndrome coro    ....................  ....................  .....................  ....................  ....................  ....................
32215 MT345887.1 |Severe acute respiratory syndrome coro    ....................  ....................  .....................  ....................  ....................  ....................
32216 MT345888.1 |Severe acute respiratory syndrome coro    ....................  ....................  .....................  ....................  ....................  ....................
32217 MT339039.1 |Severe acute respiratory syndrome coro    ....................  ....................  .....................  ....................  ....................  ....................
32218 MT339040.1 |Severe acute respiratory syndrome coro    ....................  ....................  .....................  ....................  ....................  ....................
32219 MT339041.1 |Severe acute respiratory syndrome coro    ....................  ....................  .....................  ....................  ....................  ....................
32220 MT334523.1 |Severe acute respiratory syndrome coro    ....................  ....................  .....................  ....................  ....................  ....................
32221 MT334526.1 |Severe acute respiratory syndrome coro    ....................  ....................  .....................  ....................  ....................  ....................
32222 MT334529.1 |Severe acute respiratory syndrome coro    ....................  ....................  .....................  ....................  ....................  ....................
32223 MT334531.1 |Severe acute respiratory syndrome coro    ....................  ....................  .....................  ....................  ....................  ....................
32224 MT334533.1 |Severe acute respiratory syndrome coro    ....................  ....................  .....................  ....................  ....................  ....................
32225 MT334534.1 |Severe acute respiratory syndrome coro    ....................  ....................  .....................  ....................  ....................  ....................
32226 MT334535.1 |Severe acute respiratory syndrome coro    ....................  ....................  .....................  ....................  ....................  ....................
32227 MT334536.1 |Severe acute respiratory syndrome coro    ....................  ....................  .....................  ....................  ....................  ....................
32228 MT334537.1 |Severe acute respiratory syndrome coro    ....................  ....................  .....................  ....................  ....................  ....................
32229 MT334538.1 |Severe acute respiratory syndrome coro    ....................  ....................  .....................  ....................  ....................  ....................
32230 MT334539.1 |Severe acute respiratory syndrome coro    ....................  ....................  .....................  ....................  ....................  ....................
32231 MT334540.1 |Severe acute respiratory syndrome coro    ....................  ....................  .....................  ....................  ....................  ....................
32232 MT334541.1 |Severe acute respiratory syndrome coro    ....................  ....................  .....................  ....................  ....................  ....................
32233 MT334542.1 |Severe acute respiratory syndrome coro    ....................  ....................  .....................  ....................  ....................  ....................
32234 MT334544.1 |Severe acute respiratory syndrome coro    ....................  ....................  .....................  ....................  ....................  ....................
32235 MT334546.1 |Severe acute respiratory syndrome coro    ....................  ....................  .....................  ....................  ....................  ....................
32236 MT334547.1 |Severe acute respiratory syndrome coro    ....................  ....................  .....................  ....................  ....................  ....................
32237 MT334557.1 |Severe acute respiratory syndrome coro    ....................  ....................  .....................  ....................  ....................  ....................
32238 MT334561.1 |Severe acute respiratory syndrome coro    ....................  ....................  .....................  ....................  ....................  ....................
32239 MT324062.1 |Severe acute respiratory syndrome coro    ....................  ....................  .....................  ....................  ....................  ....................
32240 MT324684.1 |Severe acute respiratory syndrome coro    ....................  ....................  .....................  ....................  ....................  ....................
32241 MT325561.1 |Severe acute respiratory syndrome coro    ....................  ....................  .....................  ....................  ....................  ....................
32242 MT325562.1 |Severe acute respiratory syndrome coro    ....................  ....................  .....................  ....................  ....................  ....................
32243 MT325563.1 |Severe acute respiratory syndrome coro    ....................  ....................  .....................  ....................  ....................  ....................
32244 MT325564.1 |Severe acute respiratory syndrome coro    ....................  ....................  .....................  ....................  ....................  ....................
32245 MT325565.1 |Severe acute respiratory syndrome coro    ....................  ....................  .....................  ....................  ....................  ....................
32246 MT325566.1 |Severe acute respiratory syndrome coro    ....................  ....................  .....................  ....................  ....................  ....................
32247 MT325567.1 |Severe acute respiratory syndrome coro    ....................  ....................  .....................  ....................  ....................  ....................
32248 MT325568.1 |Severe acute respiratory syndrome coro    ....................  ....................  .....................  ....................  ....................  ....................
32249 MT325569.1 |Severe acute respiratory syndrome coro    ....................  ....................  .....................  ....................  ....................  ....................
32250 MT325570.1 |Severe acute respiratory syndrome coro    ....................  ....................  .....................  ....................  ....................  ....................
32251 MT325571.1 |Severe acute respiratory syndrome coro    ....................  ....................  .....................  ....................  ....................  ....................
32252 MT325572.1 |Severe acute respiratory syndrome coro    ....................  ....................  .....................  ....................  ....................  ....................
32253 MT325573.1 |Severe acute respiratory syndrome coro    ....................  ....................  .....................  ....................  ....................  ....................
32254 MT325574.1 |Severe acute respiratory syndrome coro    ....................  ....................  .....................  ....................  ....................  ....................
32255 MT325575.1 |Severe acute respiratory syndrome coro    ....................  ....................  .....................  ....................  ....................  ....................
32256 MT325576.1 |Severe acute respiratory syndrome coro    ....................  ....................  .....................  ....................  ....................  ....................
32257 MT325577.1 |Severe acute respiratory syndrome coro    ....................  ....................  .....................  ....................  ....................  ....................
32258 MT325578.1 |Severe acute respiratory syndrome coro    ....................  ....................  .....................  ....................  ....................  ....................
32259 MT325579.1 |Severe acute respiratory syndrome coro    ....................  ....................  .....................  ....................  ....................  ....................
32260 MT325580.1 |Severe acute respiratory syndrome coro    ....................  ....................  .....................  ....................  ....................  ....................
32261 MT325581.1 |Severe acute respiratory syndrome coro    ....................  ....................  .....................  ....................  ....................  ....................
32262 MT325582.1 |Severe acute respiratory syndrome coro    ....................  ....................  .....................  ....................  ....................  ....................
32263 MT325583.1 |Severe acute respiratory syndrome coro    ....................  ....................  .....................  ....................  ....................  ....................
32264 MT325584.1 |Severe acute respiratory syndrome coro    ....................  ....................  .....................  ....................  ....................  ....................
32265 MT325585.1 |Severe acute respiratory syndrome coro    ....................  ....................  .....................  ....................  ....................  ....................
32266 MT325586.1 |Severe acute respiratory syndrome coro    ....................  ....................  .....................  ....................  ....................  ....................
32267 MT325587.1 |Severe acute respiratory syndrome coro    ....................  ....................  .....................  ....................  ....................  ....................
32268 MT325588.1 |Severe acute respiratory syndrome coro    ....................  ....................  .....................  ....................  ....................  ....................
32269 MT325589.1 |Severe acute respiratory syndrome coro    ....................  ....................  .....................  ....................  ....................  ....................
32270 MT325590.1 |Severe acute respiratory syndrome coro    ....................  ....................  .....................  ....................  ....................  ....................
32271 MT325591.1 |Severe acute respiratory syndrome coro    ....................  ....................  .....................  ....................  ....................  ....................
```

```
32272 MT325592.1 |Severe acute respiratory syndrome coro    ....................  ....................  ....................  ....................  ....................  ....................
32273 MT325593.1 |Severe acute respiratory syndrome coro    ....................  ....................  ....................  ....................  ....................  ....................
32274 MT325594.1 |Severe acute respiratory syndrome coro    ....................  ....................  ....................  ....................  ....................  ....................
32275 MT325595.1 |Severe acute respiratory syndrome coro    ....................  ....................  ....................  ....................  ....................  ....................
32276 MT325596.1 |Severe acute respiratory syndrome coro    ....................  ....................  ....................  ....................  ....................  ....................
32277 MT325597.1 |Severe acute respiratory syndrome coro    ....................  ....................  ....................  ....................  ....................  ....................
32278 MT325598.1 |Severe acute respiratory syndrome coro    ....................  ....................  ....................  ....................  ....................  ....................
32279 MT325599.1 |Severe acute respiratory syndrome coro    ....................  ....................  ....................  ....................  ....................  ....................
32280 MT325600.1 |Severe acute respiratory syndrome coro    ....................  ....................  ....................  ....................  ....................  ....................
32281 MT325601.1 |Severe acute respiratory syndrome coro    ....................  ....................  ....................  ....................  ....................  ....................
32282 MT325602.1 |Severe acute respiratory syndrome coro    ....................  ....................  ....................  ....................  ....................  ....................
32283 MT325603.1 |Severe acute respiratory syndrome coro    ....................  ....................  ....................  ....................  ....................  ....................
32284 MT325604.1 |Severe acute respiratory syndrome coro    ....................  ....................  ....................  ....................  ....................  ....................
32285 MT325605.1 |Severe acute respiratory syndrome coro    ....................  ....................  ....................  ....................  ....................  ....................
32286 MT325606.1 |Severe acute respiratory syndrome coro    ....................  ....................  ....................  ....................  ....................  ....................
32287 MT325607.1 |Severe acute respiratory syndrome coro    ....................  ....................  ....................  ....................  ....................  ....................
32288 MT325608.1 |Severe acute respiratory syndrome coro    ....................  ....................  ....................  ....................  ....................  ....................
32289 MT325609.1 |Severe acute respiratory syndrome coro    ....................  ....................  ....................  ....................  ....................  ....................
32290 MT325610.1 |Severe acute respiratory syndrome coro    ....................  ....................  ....................  ....................  ....................  ....................
32291 MT325611.1 |Severe acute respiratory syndrome coro    ....................  ....................  ....................  ....................  ....................  ....................
32292 MT325612.1 |Severe acute respiratory syndrome coro    ....................  ....................  ....................  ....................  ....................  ....................
32293 MT325613.1 |Severe acute respiratory syndrome coro    ....................  ....................  ....................  ....................  ....................  ....................
32294 MT325614.1 |Severe acute respiratory syndrome coro    ....................  ....................  ....................  ....................  ....................  ....................
32295 MT325615.1 |Severe acute respiratory syndrome coro    ....................  ....................  ....................  ....................  ....................  ....................
32296 MT325616.1 |Severe acute respiratory syndrome coro    ....................  ....................  ....................  ....................  ....................  ....................
32297 MT325617.1 |Severe acute respiratory syndrome coro    ....................  ....................  ....................  ....................  ....................  ....................
32298 MT325618.1 |Severe acute respiratory syndrome coro    ....................  ....................  ....................  ....................  ....................  ....................
32299 MT325619.1 |Severe acute respiratory syndrome coro    ....................  ....................  ....................  ....................  ....................  ....................
32300 MT325620.1 |Severe acute respiratory syndrome coro    ....................  ....................  ....................  ....................  ....................  ....................
32301 MT325621.1 |Severe acute respiratory syndrome coro    ....................  ....................  ....................  ....................  ....................  ....................
32302 MT325622.1 |Severe acute respiratory syndrome coro    ....................  ....................  ....................  ....................  ....................  ....................
32303 MT325623.1 |Severe acute respiratory syndrome coro    ....................  ....................  ....................  ....................  ....................  ....................
32304 MT325624.1 |Severe acute respiratory syndrome coro    ....................  ....................  ....................  ....................  ....................  ....................
32305 MT325625.1 |Severe acute respiratory syndrome coro    ....................  ....................  ....................  ....................  ....................  ....................
32306 MT325626.1 |Severe acute respiratory syndrome coro    ....................  ....................  ....................  ....................  ....................  ....................
32307 MT325627.1 |Severe acute respiratory syndrome coro    ....................  ....................  ....................  ....................  ....................  ....................
32308 MT325628.1 |Severe acute respiratory syndrome coro    ....................  ....................  ....................  ....................  ....................  ....................
32309 MT325629.1 |Severe acute respiratory syndrome coro    ....................  ....................  ....................  ....................  ....................  ....................
32310 MT325630.1 |Severe acute respiratory syndrome coro    ....................  ....................  ....................  ....................  ....................  ....................
32311 MT325631.1 |Severe acute respiratory syndrome coro    ....................  ....................  ....................  ....................  ....................  ....................
32312 MT325632.1 |Severe acute respiratory syndrome coro    ....................  ....................  ....................  ....................  ....................  ....................
32313 MT325633.1 |Severe acute respiratory syndrome coro    ....................  ....................  ....................  ....................  ....................  ....................
32314 MT325634.1 |Severe acute respiratory syndrome coro    ....................  ....................  ....................  ....................  ....................  ....................
32315 MT325635.1 |Severe acute respiratory syndrome coro    ....................  ....................  ....................  ....................  ....................  ....................
32316 MT325636.1 |Severe acute respiratory syndrome coro    ....................  ....................  ....................  ....................  ....................  ....................
32317 MT325637.1 |Severe acute respiratory syndrome coro    ....................  ....................  ....................  ....................  ....................  ....................
32318 MT325638.1 |Severe acute respiratory syndrome coro    ....................  ....................  ....................  ....................  ....................  ....................
32319 MT325639.1 |Severe acute respiratory syndrome coro    ....................  ....................  ....................  ....................  ....................  ....................
32320 MT325640.1 |Severe acute respiratory syndrome coro    ....................  ....................  ....................  ....................  ....................  ....................
32321 MT326023.1 |Severe acute respiratory syndrome coro    ....................  ....................  ....................  ....................  ....................  ....................
32322 MT326024.1 |Severe acute respiratory syndrome coro    ....................  ....................  ....................  ....................  ....................  ....................
32323 MT326027.1 |Severe acute respiratory syndrome coro    ....................  ....................  ....................  ....................  ....................  ....................
32324 MT326028.1 |Severe acute respiratory syndrome coro    ....................  ....................  ....................  ....................  ....................  ....................
32325 MT326029.1 |Severe acute respiratory syndrome coro    ....................  ....................  ....................  ....................  ....................  ....................
32326 MT326030.1 |Severe acute respiratory syndrome coro    ....................  ....................  ....................  ....................  ....................  ....................
32327 MT326035.1 |Severe acute respiratory syndrome coro    ....................  ....................  ....................  ....................  ....................  ....................
32328 MT326036.1 |Severe acute respiratory syndrome coro    ....................  ....................  ....................  ....................  ....................  ....................
32329 MT326039.1 |Severe acute respiratory syndrome coro    ....................  ....................  ....................  ....................  ....................  ....................
32330 MT326040.1 |Severe acute respiratory syndrome coro    ....................  ....................  ....................  ....................  ....................  ....................
32331 MT326041.1 |Severe acute respiratory syndrome coro    ....................  ....................  ....................  ....................  ....................  ....................
32332 MT326042.1 |Severe acute respiratory syndrome coro    ....................  ....................  ....................  ....................  ....................  ....................
32333 MT326046.1 |Severe acute respiratory syndrome coro    ....................  ....................  ....................  ....................  ....................  ....................
32334 MT326048.1 |Severe acute respiratory syndrome coro    ....................  ....................  ....................  ....................  ....................  ....................
32335 MT326049.1 |Severe acute respiratory syndrome coro    ....................  ....................  ....................  ....................  ....................  ....................
32336 MT326051.1 |Severe acute respiratory syndrome coro    ....................  ....................  ....................  ....................  ....................  ....................
32337 MT326052.1 |Severe acute respiratory syndrome coro    ....................  ....................  ....................  ....................  ....................  ....................
32338 MT326053.1 |Severe acute respiratory syndrome coro    ....................  ....................  ....................  ....................  ....................  ....................
32339 MT326054.1 |Severe acute respiratory syndrome coro    ....................  ....................  ....................  ....................  ....................  ....................
32340 MT326055.1 |Severe acute respiratory syndrome coro    ....................  ....................  ....................  ....................  ....................  ....................
32341 MT326056.1 |Severe acute respiratory syndrome coro    ....................  ....................  ....................  ....................  ....................  ....................
32342 MT326058.1 |Severe acute respiratory syndrome coro    ....................  ....................  ....................  ....................  ....................  ....................
32343 MT326061.1 |Severe acute respiratory syndrome coro    ....................  ....................  ....................  ....................  ....................  ....................
32344 MT326063.1 |Severe acute respiratory syndrome coro    ....................  ....................  ....................  ....................  ....................  ....................
32345 MT326065.1 |Severe acute respiratory syndrome coro    ....................  ....................  ....................  ....................  ....................  ....................
32346 MT326066.1 |Severe acute respiratory syndrome coro    ....................  ....................  ....................  ....................  ....................  ....................
32347 MT326067.1 |Severe acute respiratory syndrome coro    ....................  ....................  ....................  ....................  ....................  ....................
32348 MT326068.1 |Severe acute respiratory syndrome coro    ....................  ....................  ....................  ....................  ....................  ....................
32349 MT326069.1 |Severe acute respiratory syndrome coro    ....................  ....................  ....................  ....................  ....................  ....................
```

| | | | | | | | | |
|---|---|---|---|---|---|---|---|---|
| 32350 | MT326070.1 | \|Severe acute respiratory syndrome coro | .................... | .................... | ..................... | .................... | .................... | .................... |
| 32351 | MT326071.1 | \|Severe acute respiratory syndrome coro | .................... | .................... | ..................... | .................... | .................... | .................... |
| 32352 | MT326074.1 | \|Severe acute respiratory syndrome coro | .................... | .................... | ..................... | .................... | .................... | .................... |
| 32353 | MT326075.1 | \|Severe acute respiratory syndrome coro | .................... | .................... | ..................... | .................... | .................... | .................... |
| 32354 | MT326076.1 | \|Severe acute respiratory syndrome coro | .................... | .................... | ..................... | .................... | .................... | .................... |
| 32355 | MT326078.1 | \|Severe acute respiratory syndrome coro | .................... | .................... | ..................... | .................... | .................... | .................... |
| 32356 | MT326081.1 | \|Severe acute respiratory syndrome coro | .................... | .................... | ..................... | .................... | .................... | .................... |
| 32357 | MT326082.1 | \|Severe acute respiratory syndrome coro | .................... | .................... | ..................... | .................... | .................... | .................... |
| 32358 | MT326083.1 | \|Severe acute respiratory syndrome coro | .................... | .................... | ..................... | .................... | .................... | .................... |
| 32359 | MT326084.1 | \|Severe acute respiratory syndrome coro | .................... | .................... | ..................... | .................... | .................... | .................... |
| 32360 | MT326085.1 | \|Severe acute respiratory syndrome coro | .................... | .................... | ..................... | .................... | .................... | .................... |
| 32361 | MT326086.1 | \|Severe acute respiratory syndrome coro | .................... | .................... | ..................... | .................... | .................... | .................... |
| 32362 | MT326087.1 | \|Severe acute respiratory syndrome coro | .................... | .................... | ..................... | .................... | .................... | .................... |
| 32363 | MT326088.1 | \|Severe acute respiratory syndrome coro | .................... | .................... | ..................... | .................... | .................... | .................... |
| 32364 | MT326089.1 | \|Severe acute respiratory syndrome coro | .................... | .................... | ..................... | .................... | .................... | .................... |
| 32365 | MT326090.1 | \|Severe acute respiratory syndrome coro | .................... | .................... | ..................... | .................... | .................... | .................... |
| 32366 | MT326091.1 | \|Severe acute respiratory syndrome coro | .................... | .................... | ..................... | .................... | .................... | .................... |
| 32367 | MT326092.1 | \|Severe acute respiratory syndrome coro | .................... | .................... | ..................... | .................... | .................... | .................... |
| 32368 | MT326093.1 | \|Severe acute respiratory syndrome coro | .................... | .................... | ..................... | .................... | .................... | .................... |
| 32369 | MT326094.1 | \|Severe acute respiratory syndrome coro | .................... | .................... | ..................... | .................... | .................... | .................... |
| 32370 | MT326095.1 | \|Severe acute respiratory syndrome coro | .................... | .................... | ..................... | .................... | .................... | .................... |
| 32371 | MT326096.1 | \|Severe acute respiratory syndrome coro | .................... | .................... | ..................... | .................... | .................... | .................... |
| 32372 | MT326097.1 | \|Severe acute respiratory syndrome coro | .................... | .................... | ..................... | .................... | .................... | .................... |
| 32373 | MT326099.1 | \|Severe acute respiratory syndrome coro | .................... | .................... | ..................... | .................... | .................... | .................... |
| 32374 | MT326100.1 | \|Severe acute respiratory syndrome coro | .................... | .................... | ..................... | .................... | .................... | .................... |
| 32375 | MT326103.1 | \|Severe acute respiratory syndrome coro | .................... | .................... | ..................... | .................... | .................... | .................... |
| 32376 | MT326104.1 | \|Severe acute respiratory syndrome coro | .................... | .................... | ..................... | .................... | .................... | .................... |
| 32377 | MT326105.1 | \|Severe acute respiratory syndrome coro | .................... | .................... | ..................... | .................... | .................... | .................... |
| 32378 | MT326106.1 | \|Severe acute respiratory syndrome coro | .................... | .................... | ..................... | .................... | .................... | .................... |
| 32379 | MT326111.1 | \|Severe acute respiratory syndrome coro | .................... | .................... | ..................... | .................... | .................... | .................... |
| 32380 | MT326112.1 | \|Severe acute respiratory syndrome coro | .................... | .................... | ..................... | .................... | .................... | .................... |
| 32381 | MT326113.1 | \|Severe acute respiratory syndrome coro | .................... | .................... | ..................... | .................... | .................... | .................... |
| 32382 | MT326116.1 | \|Severe acute respiratory syndrome coro | .................... | .................... | ..................... | .................... | .................... | .................... |
| 32383 | MT326117.1 | \|Severe acute respiratory syndrome coro | .................... | .................... | ..................... | .................... | .................... | .................... |
| 32384 | MT326118.1 | \|Severe acute respiratory syndrome coro | .................... | .................... | ..................... | .................... | .................... | .................... |
| 32385 | MT326119.1 | \|Severe acute respiratory syndrome coro | .................... | .................... | ..................... | .................... | .................... | .................... |
| 32386 | MT326120.1 | \|Severe acute respiratory syndrome coro | .................... | .................... | ..................... | .................... | .................... | .................... |
| 32387 | MT326121.1 | \|Severe acute respiratory syndrome coro | .................... | .................... | ..................... | .................... | .................... | .................... |
| 32388 | MT326123.1 | \|Severe acute respiratory syndrome coro | .................... | .................... | ..................... | .................... | .................... | .................... |
| 32389 | MT326124.1 | \|Severe acute respiratory syndrome coro | .................... | .................... | ..................... | .................... | .................... | .................... |
| 32390 | MT326125.1 | \|Severe acute respiratory syndrome coro | .................... | .................... | ..................... | .................... | .................... | .................... |
| 32391 | MT326127.1 | \|Severe acute respiratory syndrome coro | .................... | .................... | ..................... | .................... | .................... | .................... |
| 32392 | MT326128.1 | \|Severe acute respiratory syndrome coro | .................... | .................... | ..................... | .................... | .................... | .................... |
| 32393 | MT326129.1 | \|Severe acute respiratory syndrome coro | .................... | .................... | ..................... | .................... | .................... | .................... |
| 32394 | MT326130.1 | \|Severe acute respiratory syndrome coro | .................... | .................... | ..................... | .................... | .................... | .................... |
| 32395 | MT326132.1 | \|Severe acute respiratory syndrome coro | .................... | .................... | ..................... | .................... | .................... | .................... |
| 32396 | MT326133.1 | \|Severe acute respiratory syndrome coro | .................... | .................... | ..................... | .................... | .................... | .................... |
| 32397 | MT326134.1 | \|Severe acute respiratory syndrome coro | .................... | .................... | ..................... | .................... | .................... | .................... |
| 32398 | MT326135.1 | \|Severe acute respiratory syndrome coro | .................... | .................... | ..................... | .................... | .................... | .................... |
| 32399 | MT326136.1 | \|Severe acute respiratory syndrome coro | .................... | .................... | ..................... | .................... | .................... | .................... |
| 32400 | MT326137.1 | \|Severe acute respiratory syndrome coro | .................... | .................... | ..................... | .................... | .................... | .................... |
| 32401 | MT326138.1 | \|Severe acute respiratory syndrome coro | .................... | .................... | ..................... | .................... | .................... | .................... |
| 32402 | MT326140.1 | \|Severe acute respiratory syndrome coro | .................... | .................... | ..................... | .................... | .................... | .................... |
| 32403 | MT326148.1 | \|Severe acute respiratory syndrome coro | .................... | .................... | ..................... | .................... | .................... | .................... |
| 32404 | MT326149.1 | \|Severe acute respiratory syndrome coro | .................... | .................... | ..................... | .................... | .................... | .................... |
| 32405 | MT326150.1 | \|Severe acute respiratory syndrome coro | .................... | .................... | ..................... | .................... | .................... | .................... |
| 32406 | MT326151.1 | \|Severe acute respiratory syndrome coro | .................... | .................... | ..................... | .................... | .................... | .................... |
| 32407 | MT326152.1 | \|Severe acute respiratory syndrome coro | .................... | .................... | ..................... | .................... | .................... | .................... |
| 32408 | MT326153.1 | \|Severe acute respiratory syndrome coro | .................... | .................... | ..................... | .................... | .................... | .................... |
| 32409 | MT326154.1 | \|Severe acute respiratory syndrome coro | .................... | .................... | ..................... | .................... | .................... | .................... |
| 32410 | MT326158.1 | \|Severe acute respiratory syndrome coro | .................... | .................... | ..................... | .................... | .................... | .................... |
| 32411 | MT326159.1 | \|Severe acute respiratory syndrome coro | .................... | .................... | ..................... | .................... | .................... | .................... |
| 32412 | MT326160.1 | \|Severe acute respiratory syndrome coro | .................... | .................... | ..................... | .................... | .................... | .................... |
| 32413 | MT326161.1 | \|Severe acute respiratory syndrome coro | .................... | .................... | ..................... | .................... | .................... | .................... |
| 32414 | MT326162.1 | \|Severe acute respiratory syndrome coro | .................... | .................... | ..................... | .................... | .................... | .................... |
| 32415 | MT326164.1 | \|Severe acute respiratory syndrome coro | .................... | .................... | ..................... | .................... | .................... | .................... |
| 32416 | MT326166.1 | \|Severe acute respiratory syndrome coro | .................... | .................... | ..................... | .................... | .................... | .................... |
| 32417 | MT326167.1 | \|Severe acute respiratory syndrome coro | .................... | .................... | ..................... | .................... | .................... | .................... |
| 32418 | MT326169.1 | \|Severe acute respiratory syndrome coro | .................... | .................... | ..................... | .................... | .................... | .................... |
| 32419 | MT326173.1 | \|Severe acute respiratory syndrome coro | .................... | .................... | ..................... | .................... | .................... | .................... |
| 32420 | MT326178.1 | \|Severe acute respiratory syndrome coro | .................... | .................... | ..................... | .................... | .................... | .................... |
| 32421 | MT326180.1 | \|Severe acute respiratory syndrome coro | .................... | .................... | ..................... | .................... | .................... | .................... |
| 32422 | MT326181.1 | \|Severe acute respiratory syndrome coro | .................... | .................... | ..................... | .................... | .................... | .................... |
| 32423 | MT326184.1 | \|Severe acute respiratory syndrome coro | .................... | .................... | ..................... | .................... | .................... | .................... |
| 32424 | MT326185.1 | \|Severe acute respiratory syndrome coro | .................... | .................... | ..................... | .................... | .................... | .................... |
| 32425 | MT326187.1 | \|Severe acute respiratory syndrome coro | .................... | .................... | ..................... | .................... | .................... | .................... |
| 32426 | MT326190.1 | \|Severe acute respiratory syndrome coro | .................... | .................... | ..................... | .................... | .................... | .................... |
| 32427 | MT326191.1 | \|Severe acute respiratory syndrome coro | .................... | .................... | ..................... | .................... | .................... | .................... |

32428 MT327745.1 |Severe acute respiratory syndrome coro ···················· ···················· ···················· ···················· ···················· ····················
32429 MT328032.1 |Severe acute respiratory syndrome coro ···················· ···················· ···················· ···················· ···················· ····················
32430 MT328033.1 |Severe acute respiratory syndrome coro ···················· ···················· ···················· ···················· ···················· ····················
32431 MT328034.1 |Severe acute respiratory syndrome coro ···················· ···················· ···················· ···················· ···················· ····················
32432 MT328035.1 |Severe acute respiratory syndrome coro ···················· ···················· ···················· ···················· ···················· ····················
32433 MT039874.1 |Severe acute respiratory syndrome coro ···················· ···················· ···················· ···················· ···················· ····················
32434 MT077125.1 |Severe acute respiratory syndrome coro ···················· ···················· ···················· ···················· ···················· ····················
32435 MT322394.1 |Severe acute respiratory syndrome coro ···················· ···················· ···················· ···················· ···················· ····················
32436 MT322395.1 |Severe acute respiratory syndrome coro ···················· ···················· ···················· ···················· ···················· ····················
32437 MT322396.1 |Severe acute respiratory syndrome coro ···················· ···················· ···················· ···················· ···················· ····················
32438 MT322397.1 |Severe acute respiratory syndrome coro ···················· ···················· ···················· ···················· ···················· ····················
32439 MT322398.1 |Severe acute respiratory syndrome coro ···················· ···················· ···················· ···················· ···················· ····················
32440 MT322400.1 |Severe acute respiratory syndrome coro ···················· ···················· ···················· ···················· ···················· ····················
32441 MT322401.1 |Severe acute respiratory syndrome coro ···················· ···················· ···················· ···················· ···················· ····················
32442 MT322402.1 |Severe acute respiratory syndrome coro ···················· ···················· ···················· ···················· ···················· ····················
32443 MT322403.1 |Severe acute respiratory syndrome coro ···················· ···················· ···················· ···················· ···················· ····················
32444 MT322404.1 |Severe acute respiratory syndrome coro ···················· ···················· ···················· ···················· ···················· ····················
32445 MT322405.1 |Severe acute respiratory syndrome coro ···················· ···················· ···················· ···················· ···················· ····················
32446 MT322406.1 |Severe acute respiratory syndrome coro ···················· ···················· ···················· ···················· ···················· ··A·················
32447 MT322407.1 |Severe acute respiratory syndrome coro ···················· ···················· ···················· ···················· ···················· ····················
32448 MT322408.1 |Severe acute respiratory syndrome coro ···················· ···················· ···················· ···················· ···················· ····················
32449 MT322409.1 |Severe acute respiratory syndrome coro ···················· ···················· ···················· ···················· ···················· ····················
32450 MT322410.1 |Severe acute respiratory syndrome coro ···················· ···················· ···················· ···················· ···················· ····················
32451 MT322411.1 |Severe acute respiratory syndrome coro ···················· ···················· ···················· ···················· ···················· ····················
32452 MT322412.1 |Severe acute respiratory syndrome coro ···················· ···················· ···················· ···················· ···················· ····················
32453 MT322413.1 |Severe acute respiratory syndrome coro ···················· ···················· ···················· ···················· ···················· ····················
32454 MT322414.1 |Severe acute respiratory syndrome coro ···················· ···················· ···················· ···················· ···················· ····················
32455 MT322415.1 |Severe acute respiratory syndrome coro ···················· ···················· ···················· ···················· ···················· ····················
32456 MT322416.1 |Severe acute respiratory syndrome coro ···················· ···················· ···················· ···················· ···················· ····················
32457 MT322417.1 |Severe acute respiratory syndrome coro ···················· ···················· ···················· ···················· ···················· ····················
32458 MT322418.1 |Severe acute respiratory syndrome coro ···················· ···················· ···················· ···················· ···················· ····················
32459 MT322419.1 |Severe acute respiratory syndrome coro ···················· ···················· ···················· ···················· ···················· ····················
32460 MT322420.1 |Severe acute respiratory syndrome coro ···················· ···················· ···················· ···················· ···················· ····················
32461 MT322421.1 |Severe acute respiratory syndrome coro ···················· ···················· ···················· ···················· ···················· ····················
32462 MT322422.1 |Severe acute respiratory syndrome coro ···················· ···················· ···················· ···················· ···················· ····················
32463 MT322423.1 |Severe acute respiratory syndrome coro ···················· ···················· ···················· ···················· ···················· ····················
32464 MT322424.1 |Severe acute respiratory syndrome coro ···················· ···················· ···················· ···················· ···················· ····················
32465 MT320538.2 |Severe acute respiratory syndrome coro ···················· ···················· ···················· ···················· ···················· ····················
32466 MT320891.2 |Severe acute respiratory syndrome coro ···················· ···················· ···················· ···················· ···················· ····················
32467 MT308702.1 |Severe acute respiratory syndrome coro ···················· ···················· ···················· ···················· ···················· ····················
32468 MT308703.1 |Severe acute respiratory syndrome coro ···················· ···················· ···················· ···················· ···················· ····················
32469 MT308704.1 |Severe acute respiratory syndrome coro ···················· ···················· ···················· ···················· ···················· ····················
32470 MT300186.2 |Severe acute respiratory syndrome coro ···················· ···················· ···················· ···················· ···················· ····················
32471 MT304474.1 |Severe acute respiratory syndrome coro ···················· ···················· ···················· ···················· ···················· ····················
32472 MT304475.1 |Severe acute respiratory syndrome coro ···················· ···················· ···················· ···················· ···················· ····················
32473 MT304476.1 |Severe acute respiratory syndrome coro ···················· ···················· ···················· ···················· ···················· ····················
32474 MT304477.1 |Severe acute respiratory syndrome coro ···················· ···················· ···················· ···················· ···················· ····················
32475 MT304478.1 |Severe acute respiratory syndrome coro ···················· ···················· ···················· ···················· ···················· ····················
32476 MT304479.1 |Severe acute respiratory syndrome coro ···················· ···················· ···················· ···················· ···················· ····················
32477 MT304480.1 |Severe acute respiratory syndrome coro ···················· ···················· ···················· ···················· ···················· ····················
32478 MT304481.1 |Severe acute respiratory syndrome coro ···················· ···················· ···················· ···················· ···················· ····················
32479 MT304482.1 |Severe acute respiratory syndrome coro ···················· ···················· ···················· ···················· ···················· ····················
32480 MT304483.1 |Severe acute respiratory syndrome coro ···················· ···················· ···················· ···················· ···················· ····················
32481 MT304484.1 |Severe acute respiratory syndrome coro ···················· ···················· ···················· ···················· ···················· ····················
32482 MT304485.1 |Severe acute respiratory syndrome coro ···················· ···················· ···················· ···················· ···················· ····················
32483 MT304486.1 |Severe acute respiratory syndrome coro ···················· ···················· ···················· ···················· ···················· ····················
32484 MT304487.1 |Severe acute respiratory syndrome coro ···················· ···················· ···················· ···················· ···················· ····················
32485 MT304488.1 |Severe acute respiratory syndrome coro ···················· ···················· ···················· ···················· ···················· ····················
32486 MT304489.1 |Severe acute respiratory syndrome coro ···················· ···················· ···················· ···················· ···················· ····················
32487 MT304490.1 |Severe acute respiratory syndrome coro ···················· ···················· ···················· ···················· ···················· ····················
32488 MT304491.1 |Severe acute respiratory syndrome coro ···················· ···················· ···················· ···················· ···················· ····················
32489 MT281577.1 |Severe acute respiratory syndrome coro ···················· ···················· ···················· ···················· ···················· ····················
32490 MT291826.1 |Severe acute respiratory syndrome coro ···················· ···················· ···················· ···················· ···················· ····················
32491 MT291827.1 |Severe acute respiratory syndrome coro ···················· ···················· ···················· ···················· ···················· ····················
32492 MT291828.1 |Severe acute respiratory syndrome coro ···················· ···················· ···················· ···················· ···················· ····················
32493 MT291829.1 |Severe acute respiratory syndrome coro ···················· ···················· ···················· ···················· ···················· ····················
32494 MT291830.1 |Severe acute respiratory syndrome coro ···················· ···················· ···················· ···················· ···················· ····················
32495 MT291831.1 |Severe acute respiratory syndrome coro ···················· ···················· ···················· ···················· ···················· ····················
32496 MT291832.1 |Severe acute respiratory syndrome coro ···················· ···················· ···················· ···················· ···················· ····················
32497 MT291833.1 |Severe acute respiratory syndrome coro ···················· ···················· ···················· ···················· ···················· ····················
32498 MT291834.1 |Severe acute respiratory syndrome coro ···················· ···················· ···················· ···················· ···················· ····················
32499 MT291835.2 |Severe acute respiratory syndrome coro ···················· ···················· ···················· ···················· ···················· ····················
32500 MT291836.1 |Severe acute respiratory syndrome coro ···················· ···················· ···················· ···················· ···················· ····················
32501 MT292569.1 |Severe acute respiratory syndrome coro ···················· ···················· ···················· ···················· ···················· ····················
32502 MT292570.1 |Severe acute respiratory syndrome coro ···················· ···················· ···················· ···················· ···················· ····················
32503 MT292571.1 |Severe acute respiratory syndrome coro ···················· ···················· ···················· ···················· ···················· ····················
32504 MT292572.1 |Severe acute respiratory syndrome coro ···················· ···················· ···················· ···················· ···················· ····················
32505 MT292573.1 |Severe acute respiratory syndrome coro ···················· ···················· ···················· ···················· ···················· ····················

| | | | | | | | | |
|---|---|---|---|---|---|---|---|---|
| 32506 | MT292574.1 | \|Severe acute respiratory syndrome coro | .................... | .................... | .................... | .................... | .................... | .................... |
| 32507 | MT292575.1 | \|Severe acute respiratory syndrome coro | .................... | .................... | .................... | .................... | .................... | .................... |
| 32508 | MT293156.1 | \|Severe acute respiratory syndrome coro | .................... | .................... | .................... | .................... | .................... | .................... |
| 32509 | MT293158.1 | \|Severe acute respiratory syndrome coro | .................... | .................... | .................... | .................... | .................... | .................... |
| 32510 | MT293159.1 | \|Severe acute respiratory syndrome coro | .................... | .................... | .................... | .................... | .................... | .................... |
| 32511 | MT293160.1 | \|Severe acute respiratory syndrome coro | .................... | .................... | .................... | .................... | .................... | .................... |
| 32512 | MT293161.1 | \|Severe acute respiratory syndrome coro | .................... | .................... | .................... | .................... | .................... | .................... |
| 32513 | MT293162.1 | \|Severe acute respiratory syndrome coro | .................... | .................... | .................... | .................... | .................... | .................... |
| 32514 | MT293163.1 | \|Severe acute respiratory syndrome coro | .................... | .................... | .................... | .................... | .................... | .................... |
| 32515 | MT293164.1 | \|Severe acute respiratory syndrome coro | .................... | .................... | .................... | .................... | .................... | .................... |
| 32516 | MT293165.1 | \|Severe acute respiratory syndrome coro | .................... | .................... | .................... | .................... | .................... | .................... |
| 32517 | MT293166.1 | \|Severe acute respiratory syndrome coro | .................... | .................... | .................... | .................... | .................... | .................... |
| 32518 | MT293167.1 | \|Severe acute respiratory syndrome coro | .................... | .................... | .................... | .................... | .................... | .................... |
| 32519 | MT293168.1 | \|Severe acute respiratory syndrome coro | .................... | .................... | .................... | .................... | .................... | .................... |
| 32520 | MT293169.1 | \|Severe acute respiratory syndrome coro | .................... | .................... | .................... | .................... | .................... | .................... |
| 32521 | MT293170.1 | \|Severe acute respiratory syndrome coro | .................... | .................... | .................... | .................... | .................... | .................... |
| 32522 | MT293171.1 | \|Severe acute respiratory syndrome coro | .................... | .................... | .................... | .................... | .................... | .................... |
| 32523 | MT293172.1 | \|Severe acute respiratory syndrome coro | .................... | .................... | .................... | .................... | .................... | .................... |
| 32524 | MT293173.1 | \|Severe acute respiratory syndrome coro | .................... | .................... | .................... | .................... | .................... | .................... |
| 32525 | MT293174.1 | \|Severe acute respiratory syndrome coro | .................... | .................... | .................... | .................... | .................... | .................... |
| 32526 | MT293175.1 | \|Severe acute respiratory syndrome coro | .................... | .................... | .................... | .................... | .................... | .................... |
| 32527 | MT293176.1 | \|Severe acute respiratory syndrome coro | .................... | .................... | .................... | .................... | .................... | .................... |
| 32528 | MT293177.1 | \|Severe acute respiratory syndrome coro | .................... | .................... | .................... | .................... | .................... | .................... |
| 32529 | MT293178.1 | \|Severe acute respiratory syndrome coro | .................... | .................... | .................... | .................... | .................... | .................... |
| 32530 | MT293179.1 | \|Severe acute respiratory syndrome coro | .................... | .................... | .................... | .................... | .................... | .................... |
| 32531 | MT293180.1 | \|Severe acute respiratory syndrome coro | .................... | .................... | .................... | .................... | .................... | .................... |
| 32532 | MT293181.1 | \|Severe acute respiratory syndrome coro | .................... | .................... | .................... | .................... | .................... | .................... |
| 32533 | MT293182.1 | \|Severe acute respiratory syndrome coro | .................... | .................... | .................... | .................... | .................... | .................... |
| 32534 | MT293183.1 | \|Severe acute respiratory syndrome coro | .................... | .................... | .................... | .................... | .................... | .................... |
| 32535 | MT293184.1 | \|Severe acute respiratory syndrome coro | .................... | .................... | .................... | .................... | .................... | .................... |
| 32536 | MT293185.1 | \|Severe acute respiratory syndrome coro | .................... | .................... | .................... | .................... | .................... | .................... |
| 32537 | MT293186.1 | \|Severe acute respiratory syndrome coro | .................... | .................... | .................... | .................... | .................... | .................... |
| 32538 | MT293187.1 | \|Severe acute respiratory syndrome coro | .................... | .................... | .................... | .................... | .................... | .................... |
| 32539 | MT293188.1 | \|Severe acute respiratory syndrome coro | .................... | .................... | .................... | .................... | .................... | .................... |
| 32540 | MT293189.1 | \|Severe acute respiratory syndrome coro | .................... | .................... | .................... | .................... | .................... | .................... |
| 32541 | MT293190.1 | \|Severe acute respiratory syndrome coro | .................... | .................... | .................... | .................... | .................... | .................... |
| 32542 | MT293191.1 | \|Severe acute respiratory syndrome coro | .................... | .................... | .................... | .................... | .................... | .................... |
| 32543 | MT293192.1 | \|Severe acute respiratory syndrome coro | .................... | .................... | .................... | .................... | .................... | .................... |
| 32544 | MT293194.1 | \|Severe acute respiratory syndrome coro | .................... | .................... | .................... | .................... | .................... | .................... |
| 32545 | MT293195.1 | \|Severe acute respiratory syndrome coro | .................... | .................... | .................... | .................... | .................... | .................... |
| 32546 | MT293196.1 | \|Severe acute respiratory syndrome coro | .................... | .................... | .................... | .................... | .................... | .................... |
| 32547 | MT293197.1 | \|Severe acute respiratory syndrome coro | .................... | .................... | .................... | .................... | .................... | .................... |
| 32548 | MT293198.1 | \|Severe acute respiratory syndrome coro | .................... | .................... | .................... | .................... | .................... | .................... |
| 32549 | MT293199.1 | \|Severe acute respiratory syndrome coro | .................... | .................... | .................... | .................... | .................... | .................... |
| 32550 | MT293200.1 | \|Severe acute respiratory syndrome coro | .................... | .................... | .................... | .................... | .................... | .................... |
| 32551 | MT293201.1 | \|Severe acute respiratory syndrome coro | .................... | .................... | .................... | .................... | .................... | .................... |
| 32552 | MT293202.1 | \|Severe acute respiratory syndrome coro | .................... | .................... | .................... | .................... | .................... | .................... |
| 32553 | MT293204.1 | \|Severe acute respiratory syndrome coro | .................... | .................... | .................... | .................... | .................... | .................... |
| 32554 | MT293205.1 | \|Severe acute respiratory syndrome coro | .................... | .................... | .................... | .................... | .................... | .................... |
| 32555 | MT293206.1 | \|Severe acute respiratory syndrome coro | .................... | .................... | .................... | .................... | .................... | .................... |
| 32556 | MT293207.1 | \|Severe acute respiratory syndrome coro | .................... | .................... | .................... | .................... | .................... | .................... |
| 32557 | MT293208.1 | \|Severe acute respiratory syndrome coro | .................... | .................... | .................... | .................... | .................... | .................... |
| 32558 | MT293209.1 | \|Severe acute respiratory syndrome coro | .................... | .................... | .................... | .................... | .................... | .................... |
| 32559 | MT293210.1 | \|Severe acute respiratory syndrome coro | .................... | .................... | .................... | .................... | .................... | .................... |
| 32560 | MT293211.1 | \|Severe acute respiratory syndrome coro | .................... | .................... | .................... | .................... | .................... | .................... |
| 32561 | MT293212.1 | \|Severe acute respiratory syndrome coro | .................... | .................... | .................... | .................... | .................... | .................... |
| 32562 | MT293213.1 | \|Severe acute respiratory syndrome coro | .................... | .................... | .................... | .................... | .................... | .................... |
| 32563 | MT293215.1 | \|Severe acute respiratory syndrome coro | .................... | .................... | .................... | .................... | .................... | .................... |
| 32564 | MT293216.1 | \|Severe acute respiratory syndrome coro | .................... | .................... | .................... | .................... | .................... | .................... |
| 32565 | MT293218.1 | \|Severe acute respiratory syndrome coro | .................... | .................... | .................... | .................... | .................... | .................... |
| 32566 | MT293219.1 | \|Severe acute respiratory syndrome coro | .................... | .................... | .................... | .................... | .................... | .................... |
| 32567 | MT293220.1 | \|Severe acute respiratory syndrome coro | .................... | .................... | .................... | .................... | .................... | .................... |
| 32568 | MT293222.1 | \|Severe acute respiratory syndrome coro | .................... | .................... | .................... | .................... | .................... | .................... |
| 32569 | MT293224.1 | \|Severe acute respiratory syndrome coro | .................... | .................... | .................... | .................... | .................... | .................... |
| 32570 | MT293225.1 | \|Severe acute respiratory syndrome coro | .................... | .................... | .................... | .................... | .................... | .................... |
| 32571 | MT295464.1 | \|Severe acute respiratory syndrome coro | .................... | .................... | .................... | .................... | .................... | .................... |
| 32572 | MT295465.1 | \|Severe acute respiratory syndrome coro | .................... | .................... | .................... | .................... | .................... | .................... |
| 32573 | MT276323.1 | \|Severe acute respiratory syndrome coro | .................... | .................... | .................... | .................... | .................... | .................... |
| 32574 | MT276324.1 | \|Severe acute respiratory syndrome coro | .................... | .................... | .................... | .................... | .................... | .................... |
| 32575 | MT276325.2 | \|Severe acute respiratory syndrome coro | .................... | .................... | .................... | .................... | .................... | .................... |
| 32576 | MT276326.2 | \|Severe acute respiratory syndrome coro | .................... | .................... | .................... | .................... | .................... | .................... |
| 32577 | MT276327.1 | \|Severe acute respiratory syndrome coro | .................... | .................... | .................... | .................... | .................... | .................... |
| 32578 | MT276328.2 | \|Severe acute respiratory syndrome coro | .................... | .................... | .................... | .................... | .................... | .................... |
| 32579 | MT276329.1 | \|Severe acute respiratory syndrome coro | .................... | .................... | .................... | .................... | .................... | .................... |
| 32580 | MT276330.2 | \|Severe acute respiratory syndrome coro | .................... | .................... | .................... | .................... | .................... | .................... |
| 32581 | MT276597.1 | \|Severe acute respiratory syndrome coro | .................... | .................... | .................... | .................... | .................... | .................... |
| 32582 | MT276598.1 | \|Severe acute respiratory syndrome coro | .................... | .................... | .................... | .................... | .................... | .................... |
| 32583 | MT262896.1 | \|Severe acute respiratory syndrome coro | .................... | .................... | .................... | .................... | .................... | .................... |

```
32584 MT262897.1 |Severe acute respiratory syndrome coro    ····················  ····················  ····················  ····················  ····················  ····················
32585 MT262898.1 |Severe acute respiratory syndrome coro    ····················  ····················  ····················  ····················  ····················  ····················
32586 MT262899.1 |Severe acute respiratory syndrome coro    ····················  ····················  ····················  ····················  ····················  ····················
32587 MT262900.1 |Severe acute respiratory syndrome coro    ····················  ····················  ····················  ····················  ····················  ····················
32588 MT262901.1 |Severe acute respiratory syndrome coro    ····················  ····················  ····················  ····················  ····················  ····················
32589 MT262902.1 |Severe acute respiratory syndrome coro    ····················  ····················  ····················  ····················  ····················  ····················
32590 MT262903.1 |Severe acute respiratory syndrome coro    ····················  ····················  ····················  ····················  ····················  ····················
32591 MT262904.1 |Severe acute respiratory syndrome coro    ····················  ····················  ····················  ····················  ····················  ····················
32592 MT262905.1 |Severe acute respiratory syndrome coro    ····················  ····················  ····················  ····················  ····················  ····················
32593 MT262906.1 |Severe acute respiratory syndrome coro    ····················  ····················  ····················  ····················  ····················  ····················
32594 MT262907.1 |Severe acute respiratory syndrome coro    ····················  ····················  ····················  ····················  ····················  ····················
32595 MT262908.1 |Severe acute respiratory syndrome coro    ····················  ····················  ····················  ····················  ····················  ····················
32596 MT262909.1 |Severe acute respiratory syndrome coro    ····················  ····················  ····················  ····················  ····················  ····················
32597 MT262910.1 |Severe acute respiratory syndrome coro    ····················  ····················  ····················  ····················  ····················  ····················
32598 MT262911.1 |Severe acute respiratory syndrome coro    ····················  ····················  ····················  ····················  ····················  ····················
32599 MT262912.1 |Severe acute respiratory syndrome coro    ····················  ····················  ····················  ····················  ····················  ····················
32600 MT262913.1 |Severe acute respiratory syndrome coro    ····················  ····················  ····················  ····················  ····················  ····················
32601 MT262914.1 |Severe acute respiratory syndrome coro    ····················  ····················  ····················  ····················  ····················  ····················
32602 MT262915.1 |Severe acute respiratory syndrome coro    ····················  ····················  ····················  ····················  ····················  ····················
32603 MT262916.1 |Severe acute respiratory syndrome coro    ····················  ····················  ····················  ····················  ····················  ····················
32604 MT262993.1 |Severe acute respiratory syndrome coro    ····················  ····················  ····················  ····················  ····················  ····················
32605 MT263074.1 |Severe acute respiratory syndrome coro    ····················  ····················  ····················  ····················  ····················  ····················
32606 MT263381.1 |Severe acute respiratory syndrome coro    ····················  ····················  ····················  ····················  ····················  ····················
32607 MT263382.1 |Severe acute respiratory syndrome coro    ····················  ····················  ····················  ····················  ····················  ····················
32608 MT263383.1 |Severe acute respiratory syndrome coro    ····················  ····················  ····················  ····················  ····················  ····················
32609 MT263384.1 |Severe acute respiratory syndrome coro    ····················  ····················  ····················  ····················  ····················  ····················
32610 MT263386.1 |Severe acute respiratory syndrome coro    ····················  ····················  ····················  ····················  ····················  ····················
32611 MT263387.1 |Severe acute respiratory syndrome coro    ····················  ····················  ····················  ····················  ····················  ····················
32612 MT263388.1 |Severe acute respiratory syndrome coro    ····················  ····················  ····················  ····················  ····················  ····················
32613 MT263390.1 |Severe acute respiratory syndrome coro    ····················  ····················  ····················  ····················  ····················  ····················
32614 MT263391.1 |Severe acute respiratory syndrome coro    ····················  ····················  ····················  ····················  ····················  ····················
32615 MT263392.1 |Severe acute respiratory syndrome coro    ····················  ····················  ····················  ····················  ····················  ····················
32616 MT263394.1 |Severe acute respiratory syndrome coro    ····················  ····················  ····················  ····················  ····················  ····················
32617 MT263395.1 |Severe acute respiratory syndrome coro    ····················  ····················  ····················  ····················  ····················  ····················
32618 MT263396.1 |Severe acute respiratory syndrome coro    ····················  ····················  ····················  ····················  ····················  ····················
32619 MT263398.1 |Severe acute respiratory syndrome coro    ····················  ····················  ····················  ····················  ····················  ····················
32620 MT263399.1 |Severe acute respiratory syndrome coro    ····················  ····················  ····················  ····················  ····················  ····················
32621 MT263400.1 |Severe acute respiratory syndrome coro    ····················  ····················  ····················  ····················  ····················  ····················
32622 MT263402.1 |Severe acute respiratory syndrome coro    ····················  ····················  ····················  ····················  ····················  ····················
32623 MT263403.1 |Severe acute respiratory syndrome coro    ····················  ····················  ····················  ····················  ····················  ····················
32624 MT263404.1 |Severe acute respiratory syndrome coro    ····················  ····················  ····················  ····················  ····················  ····················
32625 MT263405.1 |Severe acute respiratory syndrome coro    ····················  ····················  ····················  ····················  ····················  ····················
32626 MT263406.1 |Severe acute respiratory syndrome coro    ····················  ····················  ····················  ····················  ····················  ····················
32627 MT263408.1 |Severe acute respiratory syndrome coro    ····················  ····················  ····················  ····················  ····················  ····················
32628 MT263410.1 |Severe acute respiratory syndrome coro    ····················  ····················  ····················  ····················  ····················  ····················
32629 MT263411.1 |Severe acute respiratory syndrome coro    ····················  ····················  ····················  ····················  ····················  ····················
32630 MT263412.1 |Severe acute respiratory syndrome coro    ····················  ····················  ····················  ····················  ····················  ····················
32631 MT263413.1 |Severe acute respiratory syndrome coro    ····················  ····················  ····················  ····················  ····················  ····················
32632 MT263414.1 |Severe acute respiratory syndrome coro    ····················  ····················  ····················  ····················  ····················  ····················
32633 MT263415.1 |Severe acute respiratory syndrome coro    ····················  ····················  ····················  ····················  ····················  ····················
32634 MT263416.1 |Severe acute respiratory syndrome coro    ····················  ····················  ····················  ····················  ····················  ····················
32635 MT263417.1 |Severe acute respiratory syndrome coro    ····················  ····················  ····················  ····················  ····················  ····················
32636 MT263418.1 |Severe acute respiratory syndrome coro    ····················  ····················  ····················  ····················  ····················  ····················
32637 MT263419.1 |Severe acute respiratory syndrome coro    ····················  ····················  ····················  ····················  ····················  ····················
32638 MT263420.1 |Severe acute respiratory syndrome coro    ····················  ····················  ····················  ····················  ····················  ····················
32639 MT263421.1 |Severe acute respiratory syndrome coro    ····················  ····················  ····················  ····················  ····················  ····················
32640 MT263422.1 |Severe acute respiratory syndrome coro    ····················  ····················  ····················  ····················  ····················  ····················
32641 MT263423.1 |Severe acute respiratory syndrome coro    ····················  ····················  ····················  ····················  ····················  ····················
32642 MT263424.1 |Severe acute respiratory syndrome coro    ····················  ····················  ····················  ····················  ····················  ····················
32643 MT263425.1 |Severe acute respiratory syndrome coro    ····················  ····················  ····················  ····················  ····················  ····················
32644 MT263426.1 |Severe acute respiratory syndrome coro    ····················  ····················  ····················  ····················  ····················  ····················
32645 MT263428.1 |Severe acute respiratory syndrome coro    ····················  ····················  ····················  ····················  ····················  ····················
32646 MT263429.1 |Severe acute respiratory syndrome coro    ····················  ·A··················  ····················  ····················  ····················  ····················
32647 MT263430.1 |Severe acute respiratory syndrome coro    ····················  ····················  ····················  ····················  ····················  ····················
32648 MT263431.1 |Severe acute respiratory syndrome coro    ····················  ····················  ····················  ····················  ····················  ····················
32649 MT263432.1 |Severe acute respiratory syndrome coro    ····················  ····················  ····················  ····················  ····················  ····················
32650 MT263433.1 |Severe acute respiratory syndrome coro    ····················  ····················  ····················  ····················  ····················  ····················
32651 MT263434.1 |Severe acute respiratory syndrome coro    ····················  ····················  ····················  ····················  ····················  ····················
32652 MT263435.1 |Severe acute respiratory syndrome coro    ····················  ····················  ····················  ····················  ····················  ····················
32653 MT263436.1 |Severe acute respiratory syndrome coro    ····················  ····················  ····················  ····················  ····················  ····················
32654 MT263437.1 |Severe acute respiratory syndrome coro    ····················  ····················  ····················  ····················  ····················  ····················
32655 MT263438.1 |Severe acute respiratory syndrome coro    ····················  ····················  ····················  ····················  ····················  ····················
32656 MT263439.1 |Severe acute respiratory syndrome coro    ····················  ····················  ····················  ····················  ····················  ····················
32657 MT263440.1 |Severe acute respiratory syndrome coro    ····················  ····················  ····················  ····················  ····················  ····················
32658 MT263441.1 |Severe acute respiratory syndrome coro    ····················  ····················  ····················  ····················  ····················  ····················
32659 MT263442.1 |Severe acute respiratory syndrome coro    ····················  ····················  ····················  ····················  ····················  ····················
32660 MT263443.1 |Severe acute respiratory syndrome coro    ····················  ····················  ····················  ····················  ····················  ····················
32661 MT263444.1 |Severe acute respiratory syndrome coro    ····················  ····················  ····················  ····················  ····················  ····················
```

```
32662 MT263445.1 |Severe acute respiratory syndrome coro    ....................  ....................  .....................  ....................  ....................  ....................
32663 MT263446.1 |Severe acute respiratory syndrome coro    ....................  ....................  .....................  ....................  ....................  ....................
32664 MT263447.1 |Severe acute respiratory syndrome coro    ....................  ....................  .....................  ....................  ....................  ....................
32665 MT263448.1 |Severe acute respiratory syndrome coro    ....................  ....................  .....................  ....................  ....................  ....................
32666 MT263449.1 |Severe acute respiratory syndrome coro    ....................  ....................  .....................  ....................  ....................  ....................
32667 MT263450.1 |Severe acute respiratory syndrome coro    ....................  ....................  .....................  ....................  ....................  ....................
32668 MT263451.1 |Severe acute respiratory syndrome coro    ....................  ....................  .....................  ....................  ....................  ....................
32669 MT263452.1 |Severe acute respiratory syndrome coro    ....................  ....................  .....................  ....................  ....................  ....................
32670 MT263453.1 |Severe acute respiratory syndrome coro    ....................  ....................  .....................  ....................  ....................  ....................
32671 MT263454.1 |Severe acute respiratory syndrome coro    ....................  ....................  .....................  ....................  ....................  ....................
32672 MT263455.1 |Severe acute respiratory syndrome coro    ....................  ....................  .....................  ....................  ....................  ....................
32673 MT263456.1 |Severe acute respiratory syndrome coro    ....................  ....................  .....................  ....................  ....................  ....................
32674 MT263457.1 |Severe acute respiratory syndrome coro    ....................  ....................  .....................  ....................  ....................  ....................
32675 MT263458.1 |Severe acute respiratory syndrome coro    ....................  ....................  .....................  ....................  ....................  ....................
32676 MT263459.1 |Severe acute respiratory syndrome coro    ....................  ....................  .....................  ....................  ....................  ....................
32677 MT263462.1 |Severe acute respiratory syndrome coro    ....................  ....................  .....................  ....................  ....................  ....................
32678 MT263463.1 |Severe acute respiratory syndrome coro    ....................  ....................  .....................  ....................  ....................  ....................
32679 MT263464.1 |Severe acute respiratory syndrome coro    ....................  ....................  .....................  ....................  ....................  ....................
32680 MT263465.1 |Severe acute respiratory syndrome coro    ....................  ....................  .....................  ....................  ....................  ....................
32681 MT263467.1 |Severe acute respiratory syndrome coro    ....................  ....................  .....................  ....................  ....................  ....................
32682 MT263468.1 |Severe acute respiratory syndrome coro    ....................  ....................  .....................  ....................  ....................  ....................
32683 MT263469.1 |Severe acute respiratory syndrome coro    ....................  ....................  .....................  ....................  ....................  ....................
32684 MT256924.2 |Severe acute respiratory syndrome coro    ....................  ....................  .....................  ....................  ....................  ....................
32685 MT258377.1 |Severe acute respiratory syndrome coro    ....................  ....................  .....................  ....................  ....................  ....................
32686 MT258378.1 |Severe acute respiratory syndrome coro    ....................  ....................  .....................  ....................  ....................  ....................
32687 MT258379.1 |Severe acute respiratory syndrome coro    ....................  ....................  .....................  ....................  ....................  ....................
32688 MT258380.1 |Severe acute respiratory syndrome coro    ....................  ....................  .....................  ....................  ....................  ....................
32689 MT258381.1 |Severe acute respiratory syndrome coro    ....................  ....................  .....................  ....................  ....................  ....................
32690 MT258382.1 |Severe acute respiratory syndrome coro    ....................  ....................  .....................  ....................  ....................  ....................
32691 MT258383.1 |Severe acute respiratory syndrome coro    ....................  ....................  .....................  ....................  ....................  ....................
32692 MT259226.1 |Severe acute respiratory syndrome coro    ....................  ....................  .....................  ....................  ....................  ....................
32693 MT259227.1 |Severe acute respiratory syndrome coro    ....................  ....................  .....................  ....................  ....................  ....................
32694 MT259228.1 |Severe acute respiratory syndrome coro    ....................  ....................  .....................  ....................  ....................  ....................
32695 MT259229.1 |Severe acute respiratory syndrome coro    ....................  ....................  .....................  ....................  ....................  ....................
32696 MT259230.1 |Severe acute respiratory syndrome coro    ....................  ....................  .....................  ....................  ....................  ....................
32697 MT259231.1 |Severe acute respiratory syndrome coro    ....................  ....................  .....................  ....................  ....................  ....................
32698 MT259235.1 |Severe acute respiratory syndrome coro    ....................  ....................  .....................  ....................  ....................  ....................
32699 MT259236.1 |Severe acute respiratory syndrome coro    ....................  ....................  .....................  ....................  ....................  ....................
32700 MT259237.1 |Severe acute respiratory syndrome coro    ....................  ....................  .....................  ....................  ....................  ....................
32701 MT259239.1 |Severe acute respiratory syndrome coro    ....................  ....................  .....................  ....................  ....................  ....................
32702 MT259240.1 |Severe acute respiratory syndrome coro    ....................  ....................  .....................  ....................  ....................  ....................
32703 MT259241.1 |Severe acute respiratory syndrome coro    ....................  ....................  .....................  ....................  ....................  ....................
32704 MT259243.1 |Severe acute respiratory syndrome coro    ....................  ....................  .....................  ....................  ....................  ....................
32705 MT259244.1 |Severe acute respiratory syndrome coro    ....................  ....................  .....................  ....................  ....................  ....................
32706 MT259245.1 |Severe acute respiratory syndrome coro    ....................  ....................  .....................  ....................  ....................  ....................
32707 MT259246.1 |Severe acute respiratory syndrome coro    ....................  ....................  .....................  ....................  ....................  ....................
32708 MT259247.1 |Severe acute respiratory syndrome coro    ....................  ....................  .....................  ....................  ....................  ....................
32709 MT259248.1 |Severe acute respiratory syndrome coro    ....................  ....................  .....................  ....................  ....................  ....................
32710 MT259249.1 |Severe acute respiratory syndrome coro    ....................  ....................  .....................  ....................  ....................  ....................
32711 MT259250.1 |Severe acute respiratory syndrome coro    ....................  ....................  .....................  ....................  ....................  ....................
32712 MT259251.1 |Severe acute respiratory syndrome coro    ....................  ....................  .....................  ....................  ....................  ....................
32713 MT259252.1 |Severe acute respiratory syndrome coro    ....................  ....................  .....................  ....................  ....................  ....................
32714 MT259253.1 |Severe acute respiratory syndrome coro    ....................  ....................  .....................  ....................  ....................  ....................
32715 MT259254.1 |Severe acute respiratory syndrome coro    ....................  ....................  .....................  ....................  ....................  ....................
32716 MT259256.1 |Severe acute respiratory syndrome coro    ....................  ....................  .....................  ....................  ....................  ....................
32717 MT259257.1 |Severe acute respiratory syndrome coro    ....................  ....................  .....................  ....................  ....................  ....................
32718 MT259258.1 |Severe acute respiratory syndrome coro    ....................  ....................  .....................  ....................  ....................  ....................
32719 MT259260.1 |Severe acute respiratory syndrome coro    ....................  ....................  .....................  ....................  ....................  ....................
32720 MT259261.1 |Severe acute respiratory syndrome coro    ....................  ....................  .....................  ....................  ....................  ....................
32721 MT259262.1 |Severe acute respiratory syndrome coro    ....................  ....................  .....................  ....................  ....................  ....................
32722 MT259263.1 |Severe acute respiratory syndrome coro    ....................  ....................  .....................  ....................  ....................  ....................
32723 MT259264.1 |Severe acute respiratory syndrome coro    ....................  ....................  .....................  ....................  ....................  ....................
32724 MT259266.1 |Severe acute respiratory syndrome coro    ....................  ....................  .....................  ....................  ....................  ....................
32725 MT259267.1 |Severe acute respiratory syndrome coro    ....................  ....................  .....................  ....................  ....................  ....................
32726 MT259268.1 |Severe acute respiratory syndrome coro    ....................  ....................  .....................  ....................  ....................  ....................
32727 MT259269.1 |Severe acute respiratory syndrome coro    ....................  ....................  .....................  ....................  ....................  ....................
32728 MT259271.1 |Severe acute respiratory syndrome coro    ....................  ....................  .....................  ....................  ....................  ....................
32729 MT259273.1 |Severe acute respiratory syndrome coro    ....................  ....................  .....................  ....................  ....................  ....................
32730 MT259274.1 |Severe acute respiratory syndrome coro    ....................  ....................  .....................  ....................  ....................  ....................
32731 MT259275.1 |Severe acute respiratory syndrome coro    ....................  ....................  .....................  ....................  ....................  ....................
32732 MT259277.1 |Severe acute respiratory syndrome coro    ....................  ....................  .....................  ....................  ....................  ....................
32733 MT259278.1 |Severe acute respiratory syndrome coro    ....................  ....................  .....................  ....................  ....................  ....................
32734 MT259280.1 |Severe acute respiratory syndrome coro    ....................  ....................  .....................  ....................  ....................  ....................
32735 MT259281.1 |Severe acute respiratory syndrome coro    ....................  ....................  .....................  ....................  ....................  ....................
32736 MT259282.1 |Severe acute respiratory syndrome coro    ....................  ....................  .....................  ....................  ....................  ....................
32737 MT259284.1 |Severe acute respiratory syndrome coro    ....................  ....................  .....................  ....................  ....................  ....................
32738 MT259285.1 |Severe acute respiratory syndrome coro    ....................  ....................  .....................  ....................  ....................  ....................
32739 MT259286.1 |Severe acute respiratory syndrome coro    ....................  ....................  .....................  ....................  ....................  ....................
```

| | | | | | | | | |
|---|---|---|---|---|---|---|---|---|
| 32740 | MT259287.1 | \|Severe acute respiratory syndrome coro | .................... | .................... | ..................... | .................... | .................... | .................... |
| 32741 | LC534418.1 | \|Severe acute respiratory syndrome coro | .................... | .................... | ..................... | .................... | .................... | .................... |
| 32742 | LC534419.1 | \|Severe acute respiratory syndrome coro | .................... | .................... | ..................... | .................... | .................... | .................... |
| 32743 | MT251972.1 | \|Severe acute respiratory syndrome coro | .................... | .................... | ..................... | .................... | .................... | .................... |
| 32744 | MT251973.1 | \|Severe acute respiratory syndrome coro | .................... | .................... | ..................... | .................... | .................... | .................... |
| 32745 | MT251974.1 | \|Severe acute respiratory syndrome coro | .................... | .................... | ..................... | .................... | .................... | .................... |
| 32746 | MT251975.1 | \|Severe acute respiratory syndrome coro | .................... | .................... | ..................... | .................... | .................... | .................... |
| 32747 | MT251976.1 | \|Severe acute respiratory syndrome coro | .................... | .................... | ..................... | .................... | .................... | .................... |
| 32748 | MT251977.1 | \|Severe acute respiratory syndrome coro | .................... | .................... | ..................... | .................... | .................... | .................... |
| 32749 | MT251978.1 | \|Severe acute respiratory syndrome coro | .................... | .................... | ..................... | .................... | .................... | .................... |
| 32750 | MT251979.1 | \|Severe acute respiratory syndrome coro | .................... | .................... | ..................... | .................... | .................... | .................... |
| 32751 | MT251980.1 | \|Severe acute respiratory syndrome coro | .................... | .................... | ..................... | .................... | .................... | .................... |
| 32752 | MT253696.1 | \|Severe acute respiratory syndrome coro | .................... | .................... | ..................... | .................... | .................... | .................... |
| 32753 | MT253697.1 | \|Severe acute respiratory syndrome coro | .................... | .................... | ..................... | .................... | .................... | .................... |
| 32754 | MT253698.1 | \|Severe acute respiratory syndrome coro | .................... | .................... | ..................... | .................... | .................... | .................... |
| 32755 | MT253699.1 | \|Severe acute respiratory syndrome coro | .................... | .................... | ..................... | .................... | .................... | .................... |
| 32756 | MT253700.1 | \|Severe acute respiratory syndrome coro | .................... | .................... | ..................... | .................... | .................... | .................... |
| 32757 | MT253701.1 | \|Severe acute respiratory syndrome coro | .................... | .................... | ..................... | .................... | .................... | .................... |
| 32758 | MT253702.1 | \|Severe acute respiratory syndrome coro | .................... | .................... | ..................... | .................... | .................... | .................... |
| 32759 | MT253703.1 | \|Severe acute respiratory syndrome coro | .................... | .................... | ..................... | .................... | .................... | .................... |
| 32760 | MT253704.1 | \|Severe acute respiratory syndrome coro | .................... | .................... | ..................... | .................... | .................... | .................... |
| 32761 | MT253705.1 | \|Severe acute respiratory syndrome coro | .................... | .................... | ..................... | .................... | .................... | .................... |
| 32762 | MT253706.1 | \|Severe acute respiratory syndrome coro | .................... | .................... | ..................... | .................... | .................... | .................... |
| 32763 | MT253707.1 | \|Severe acute respiratory syndrome coro | .................... | .................... | ..................... | .................... | .................... | .................... |
| 32764 | MT253708.1 | \|Severe acute respiratory syndrome coro | .................... | .................... | ..................... | .................... | .................... | .................... |
| 32765 | MT253709.1 | \|Severe acute respiratory syndrome coro | .................... | .................... | ..................... | .................... | .................... | .................... |
| 32766 | MT253710.1 | \|Severe acute respiratory syndrome coro | .................... | .................... | ..................... | .................... | .................... | .................... |
| 32767 | MT233526.1 | \|Severe acute respiratory syndrome coro | .................... | .................... | ..................... | .................... | .................... | .................... |
| 32768 | MT246449.1 | \|Severe acute respiratory syndrome coro | .................... | .................... | ..................... | .................... | .................... | .................... |
| 32769 | MT246450.1 | \|Severe acute respiratory syndrome coro | .................... | .................... | ..................... | .................... | .................... | .................... |
| 32770 | MT246451.1 | \|Severe acute respiratory syndrome coro | .................... | .................... | ..................... | .................... | .................... | .................... |
| 32771 | MT246452.1 | \|Severe acute respiratory syndrome coro | .................... | .................... | ..................... | .................... | .................... | .................... |
| 32772 | MT246453.1 | \|Severe acute respiratory syndrome coro | .................... | .................... | ..................... | .................... | .................... | .................... |
| 32773 | MT246454.1 | \|Severe acute respiratory syndrome coro | .................... | .................... | ..................... | .................... | .................... | .................... |
| 32774 | MT246455.1 | \|Severe acute respiratory syndrome coro | .................... | .................... | ..................... | .................... | .................... | .................... |
| 32775 | MT246456.1 | \|Severe acute respiratory syndrome coro | .................... | .................... | ..................... | .................... | .................... | .................... |
| 32776 | MT246457.1 | \|Severe acute respiratory syndrome coro | .................... | .................... | ..................... | .................... | .................... | .................... |
| 32777 | MT246458.1 | \|Severe acute respiratory syndrome coro | .................... | .................... | ..................... | .................... | .................... | .................... |
| 32778 | MT246459.1 | \|Severe acute respiratory syndrome coro | .................... | .................... | ..................... | .................... | .................... | .................... |
| 32779 | MT246460.1 | \|Severe acute respiratory syndrome coro | .................... | .................... | ..................... | .................... | .................... | .................... |
| 32780 | MT246461.1 | \|Severe acute respiratory syndrome coro | .................... | .................... | ..................... | .................... | .................... | .................... |
| 32781 | MT246462.1 | \|Severe acute respiratory syndrome coro | .................... | .................... | ..................... | .................... | .................... | .................... |
| 32782 | MT246463.1 | \|Severe acute respiratory syndrome coro | .................... | .................... | ..................... | .................... | .................... | .................... |
| 32783 | MT246464.1 | \|Severe acute respiratory syndrome coro | .................... | .................... | ..................... | .................... | .................... | .................... |
| 32784 | MT246466.1 | \|Severe acute respiratory syndrome coro | .................... | .................... | ..................... | .................... | .................... | .................... |
| 32785 | MT246467.1 | \|Severe acute respiratory syndrome coro | .................... | .................... | ..................... | .................... | .................... | .................... |
| 32786 | MT246468.1 | \|Severe acute respiratory syndrome coro | .................... | .................... | ..................... | .................... | .................... | .................... |
| 32787 | MT246469.1 | \|Severe acute respiratory syndrome coro | .................... | .................... | ..................... | .................... | .................... | .................... |
| 32788 | MT246470.1 | \|Severe acute respiratory syndrome coro | .................... | .................... | ..................... | .................... | .................... | .................... |
| 32789 | MT246471.1 | \|Severe acute respiratory syndrome coro | .................... | .................... | ..................... | .................... | .................... | .................... |
| 32790 | MT246472.1 | \|Severe acute respiratory syndrome coro | .................... | .................... | ..................... | .................... | .................... | .................... |
| 32791 | MT246473.1 | \|Severe acute respiratory syndrome coro | .................... | .................... | ..................... | .................... | .................... | .................... |
| 32792 | MT246474.1 | \|Severe acute respiratory syndrome coro | .................... | .................... | ..................... | .................... | .................... | .................... |
| 32793 | MT246475.1 | \|Severe acute respiratory syndrome coro | .................... | .................... | ..................... | .................... | .................... | .................... |
| 32794 | MT246476.1 | \|Severe acute respiratory syndrome coro | .................... | .................... | ..................... | .................... | .................... | .................... |
| 32795 | MT246477.1 | \|Severe acute respiratory syndrome coro | .................... | .................... | ..................... | .................... | .................... | .................... |
| 32796 | MT246478.1 | \|Severe acute respiratory syndrome coro | .................... | .................... | ..................... | .................... | .................... | .................... |
| 32797 | MT246479.1 | \|Severe acute respiratory syndrome coro | .................... | .................... | ..................... | .................... | .................... | .................... |
| 32798 | MT246480.1 | \|Severe acute respiratory syndrome coro | .................... | .................... | ..................... | .................... | .................... | .................... |
| 32799 | MT246481.1 | \|Severe acute respiratory syndrome coro | .................... | .................... | ..................... | .................... | .................... | .................... |
| 32800 | MT246482.1 | \|Severe acute respiratory syndrome coro | .................... | .................... | ..................... | .................... | .................... | .................... |
| 32801 | MT246484.1 | \|Severe acute respiratory syndrome coro | .................... | .................... | ..................... | .................... | .................... | .................... |
| 32802 | MT246485.1 | \|Severe acute respiratory syndrome coro | .................... | .................... | ..................... | .................... | .................... | .................... |
| 32803 | MT246486.1 | \|Severe acute respiratory syndrome coro | .................... | .................... | ..................... | .................... | .................... | .................... |
| 32804 | MT246487.1 | \|Severe acute respiratory syndrome coro | .................... | .................... | ..................... | .................... | .................... | .................... |
| 32805 | MT246488.1 | \|Severe acute respiratory syndrome coro | .................... | .................... | ..................... | .................... | .................... | .................... |
| 32806 | MT246489.1 | \|Severe acute respiratory syndrome coro | .................... | .................... | ..................... | .................... | .................... | .................... |
| 32807 | MT246490.1 | \|Severe acute respiratory syndrome coro | .................... | .................... | ..................... | .................... | .................... | .................... |
| 32808 | MT246667.1 | \|Severe acute respiratory syndrome coro | .................... | .................... | ..................... | .................... | .................... | .................... |
| 32809 | MT240479.1 | \|Severe acute respiratory syndrome coro | .................... | .................... | ..................... | .................... | .................... | .................... |
| 32810 | MT233519.1 | \|Severe acute respiratory syndrome coro | .................... | .................... | ..................... | .................... | .................... | .................... |
| 32811 | MT233522.1 | \|Severe acute respiratory syndrome coro | .................... | .................... | ..................... | .................... | .................... | .................... |
| 32812 | MT233523.1 | \|Severe acute respiratory syndrome coro | .................... | .................... | ..................... | .................... | .................... | .................... |
| 32813 | MT226610.1 | \|Severe acute respiratory syndrome coro | .................... | .................... | ..................... | .................... | .................... | .................... |
| 32814 | MT198652.2 | \|Severe acute respiratory syndrome coro | .................... | .................... | ..................... | .................... | .................... | .................... |
| 32815 | MT192759.1 | \|Severe acute respiratory syndrome coro | .................... | .................... | ..................... | .................... | .................... | .................... |
| 32816 | MT192765.1 | \|Severe acute respiratory syndrome coro | .................... | .................... | ..................... | .................... | .................... | .................... |
| 32817 | MT192772.1 | \|Severe acute respiratory syndrome coro | .................... | .................... | ..................... | .................... | .................... | .................... |

| | | | | | | | | |
|---|---|---|---|---|---|---|---|---|
| 32818 | MT192773.1 | \|Severe acute respiratory syndrome coro | .................... | .................... | .................... | .................... | .................... | .................... |
| 32819 | MT188339.1 | \|Severe acute respiratory syndrome coro | .................... | .................... | .................... | .................... | .................... | ...............A.... |
| 32820 | MT188340.1 | \|Severe acute respiratory syndrome coro | .................... | .................... | .................... | .................... | .................... | .................... |
| 32821 | MT188341.1 | \|Severe acute respiratory syndrome coro | .................... | .................... | .................... | .................... | .................... | .................... |
| 32822 | MT184907.2 | \|Severe acute respiratory syndrome coro | .................... | .................... | .................... | .................... | .................... | .................... |
| 32823 | MT184908.2 | \|Severe acute respiratory syndrome coro | .................... | .................... | .................... | .................... | .................... | .................... |
| 32824 | MT184909.2 | \|Severe acute respiratory syndrome coro | .................... | .................... | .................... | .................... | .................... | .................... |
| 32825 | MT184910.1 | \|Severe acute respiratory syndrome coro | .................... | .................... | .................... | .................... | .................... | .................... |
| 32826 | MT184911.1 | \|Severe acute respiratory syndrome coro | .................... | .................... | .................... | .................... | .................... | .................... |
| 32827 | MT184912.1 | \|Severe acute respiratory syndrome coro | .................... | .................... | .................... | .................... | .................... | .................... |
| 32828 | MT184913.1 | \|Severe acute respiratory syndrome coro | .................... | .................... | .................... | .................... | .................... | .................... |
| 32829 | MT163716.1 | \|Severe acute respiratory syndrome coro | .................... | .................... | .................... | .................... | .................... | .................... |
| 32830 | MT163717.1 | \|Severe acute respiratory syndrome coro | .................... | .................... | .................... | .................... | .................... | .................... |
| 32831 | MT163718.1 | \|Severe acute respiratory syndrome coro | .................... | .................... | .................... | .................... | .................... | .................... |
| 32832 | MT163719.1 | \|Severe acute respiratory syndrome coro | .................... | .................... | .................... | .................... | .................... | .................... |
| 32833 | MT163720.1 | \|Severe acute respiratory syndrome coro | .................... | .................... | .................... | .................... | .................... | .................... |
| 32834 | MT066156.1 | \|Severe acute respiratory syndrome coro | .................... | .................... | .................... | .................... | .................... | .................... |
| 32835 | MT121215.1 | \|Severe acute respiratory syndrome coro | .................... | .................... | .................... | .................... | .................... | .................... |
| 32836 | MT159705.2 | \|Severe acute respiratory syndrome coro | .................... | .................... | .................... | .................... | .................... | .................... |
| 32837 | MT159706.2 | \|Severe acute respiratory syndrome coro | .................... | .................... | .................... | .................... | .................... | .................... |
| 32838 | MT159707.2 | \|Severe acute respiratory syndrome coro | .................... | .................... | .................... | .................... | .................... | .................... |
| 32839 | MT159708.2 | \|Severe acute respiratory syndrome coro | .................... | .................... | .................... | .................... | .................... | .................... |
| 32840 | MT159709.2 | \|Severe acute respiratory syndrome coro | .................... | .................... | .................... | .................... | .................... | .................... |
| 32841 | MT159710.2 | \|Severe acute respiratory syndrome coro | .................... | .................... | .................... | .................... | .................... | .................... |
| 32842 | MT159711.2 | \|Severe acute respiratory syndrome coro | .................... | .................... | .................... | .................... | .................... | .................... |
| 32843 | MT159712.2 | \|Severe acute respiratory syndrome coro | .................... | .................... | .................... | .................... | .................... | .................... |
| 32844 | MT159713.2 | \|Severe acute respiratory syndrome coro | .................... | .................... | .................... | .................... | .................... | .................... |
| 32845 | MT159714.2 | \|Severe acute respiratory syndrome coro | .................... | .................... | .................... | .................... | .................... | .................... |
| 32846 | MT159715.2 | \|Severe acute respiratory syndrome coro | .................... | .................... | .................... | .................... | .................... | .................... |
| 32847 | MT159716.2 | \|Severe acute respiratory syndrome coro | .................... | .................... | .................... | .................... | .................... | .................... |
| 32848 | MT159717.2 | \|Severe acute respiratory syndrome coro | .................... | .................... | .................... | .................... | .................... | .................... |
| 32849 | MT159718.2 | \|Severe acute respiratory syndrome coro | .................... | .................... | .................... | .................... | .................... | .................... |
| 32850 | MT159719.2 | \|Severe acute respiratory syndrome coro | .................... | .................... | .................... | .................... | .................... | .................... |
| 32851 | MT159720.2 | \|Severe acute respiratory syndrome coro | .................... | .................... | .................... | .................... | .................... | .................... |
| 32852 | MT159721.2 | \|Severe acute respiratory syndrome coro | .................... | .................... | .................... | .................... | .................... | .................... |
| 32853 | MT159722.2 | \|Severe acute respiratory syndrome coro | .................... | .................... | .................... | .................... | .................... | .................... |
| 32854 | LC529905.1 | \|Severe acute respiratory syndrome coro | .................... | .................... | .................... | .................... | .................... | .................... |
| 32855 | MT012098.1 | \|Severe acute respiratory syndrome coro | .................... | .................... | .................... | .................... | .................... | .................... |
| 32856 | MT050493.1 | \|Severe acute respiratory syndrome coro | .................... | .................... | .................... | .................... | .................... | .................... |
| 32857 | MT152824.1 | \|Severe acute respiratory syndrome coro | .................... | .................... | .................... | .................... | .................... | .................... |
| 32858 | MT135041.1 | \|Severe acute respiratory syndrome coro | .................... | .................... | .................... | .................... | .................... | .................... |
| 32859 | MT135042.1 | \|Severe acute respiratory syndrome coro | .................... | .................... | .................... | .................... | .................... | .................... |
| 32860 | MT135043.1 | \|Severe acute respiratory syndrome coro | .................... | .................... | .................... | .................... | .................... | .................... |
| 32861 | MT135044.1 | \|Severe acute respiratory syndrome coro | .................... | .................... | .................... | .................... | .................... | .................... |
| 32862 | MT126808.1 | \|Severe acute respiratory syndrome coro | .................... | .................... | .................... | .................... | .................... | .................... |
| 32863 | LC528232.1 | \|Severe acute respiratory syndrome coro | .................... | .................... | .................... | .................... | .................... | .................... |
| 32864 | LC528233.1 | \|Severe acute respiratory syndrome coro | .................... | .................... | .................... | .................... | .................... | .................... |
| 32865 | MT123290.1 | \|Severe acute respiratory syndrome coro | .................... | .................... | .................... | .................... | .................... | .................... |
| 32866 | MT123291.2 | \|Severe acute respiratory syndrome coro | .................... | .................... | .................... | .................... | .................... | .................... |
| 32867 | MT123292.2 | \|Severe acute respiratory syndrome coro | .................... | .................... | .................... | .................... | .................... | .................... |
| 32868 | MT123293.2 | \|Severe acute respiratory syndrome coro | .................... | .................... | .................... | .................... | .................... | .................... |
| 32869 | MT118835.1 | \|Severe acute respiratory syndrome coro | .................... | .................... | .................... | .................... | .................... | .................... |
| 32870 | MT106052.1 | \|Severe acute respiratory syndrome coro | .................... | .................... | .................... | .................... | .................... | .................... |
| 32871 | MT106053.1 | \|Severe acute respiratory syndrome coro | .................... | .................... | .................... | .................... | .................... | .................... |
| 32872 | MT106054.1 | \|Severe acute respiratory syndrome coro | .................... | .................... | .................... | .................... | .................... | .................... |
| 32873 | MT093571.1 | \|Severe acute respiratory syndrome coro | .................... | .................... | .................... | .................... | .................... | .................... |
| 32874 | MT093631.2 | \|Severe acute respiratory syndrome coro | .................... | .................... | .................... | .................... | .................... | .................... |
| 32875 | MT072688.1 | \|Severe acute respiratory syndrome coro | .................... | .................... | .................... | .................... | .................... | .................... |
| 32876 | MT066175.1 | \|Severe acute respiratory syndrome coro | .................... | .................... | .................... | .................... | .................... | .................... |
| 32877 | MT066176.1 | \|Severe acute respiratory syndrome coro | .................... | .................... | .................... | .................... | .................... | .................... |
| 32878 | MT044257.1 | \|Severe acute respiratory syndrome coro | .................... | .................... | .................... | .................... | .................... | .................... |
| 32879 | MT044258.1 | \|Severe acute respiratory syndrome coro | .................... | .................... | .................... | .................... | .................... | .................... |
| 32880 | MT049951.1 | \|Severe acute respiratory syndrome coro | .................... | .................... | .................... | .................... | .................... | .................... |
| 32881 | MT039873.1 | \|Severe acute respiratory syndrome coro | .................... | .................... | .................... | .................... | .................... | .................... |
| 32882 | MT039887.1 | \|Severe acute respiratory syndrome coro | .................... | .................... | .................... | .................... | .................... | .................... |
| 32883 | MT039888.1 | \|Severe acute respiratory syndrome coro | .................... | .A.................. | .................... | .................... | .................... | .................... |
| 32884 | MT039890.1 | \|Severe acute respiratory syndrome coro | .................... | .................... | .................... | ...........T........ | .................... | .................... |
| 32885 | MT027062.1 | \|Severe acute respiratory syndrome coro | .................... | .A.................. | .................... | .................... | .................... | .................... |
| 32886 | MT027063.1 | \|Severe acute respiratory syndrome coro | .................... | .A.................. | .................... | .................... | .................... | .................... |
| 32887 | MT027064.1 | \|Severe acute respiratory syndrome coro | .................... | .................... | .................... | .................... | .................... | .................... |
| 32888 | MT019529.1 | \|Severe acute respiratory syndrome coro | .................... | .................... | .................... | .................... | .................... | .................... |
| 32889 | MT019530.1 | \|Severe acute respiratory syndrome coro | .................... | .................... | .................... | .................... | .................... | .................... |
| 32890 | MT019531.1 | \|Severe acute respiratory syndrome coro | .................... | .................... | .................... | .................... | .................... | .................... |
| 32891 | MT019532.1 | \|Severe acute respiratory syndrome coro | .................... | .................... | .................... | .................... | .................... | .................... |
| 32892 | MT019533.1 | \|Severe acute respiratory syndrome coro | .................... | .................... | .................... | .................... | .................... | .................... |
| 32893 | MT020781.2 | \|Severe acute respiratory syndrome coro | .................... | .................... | .................... | .................... | .................... | .................... |
| 32894 | MT020880.1 | \|Severe acute respiratory syndrome coro | .................... | .................... | .................... | .................... | .................... | .................... |
| 32895 | MT020881.1 | \|Severe acute respiratory syndrome coro | .................... | .................... | .................... | .................... | .................... | .................... |

```
32896 MT007544.1 |Severe acute respiratory syndrome coro    ····················  ····················  ····················  ····················  ····················  ····················
32897 MN996527.1 |Severe acute respiratory syndrome coro    ····················  ····················  ····················  ····················  ····················  ····················
32898 MN996528.1 |Severe acute respiratory syndrome coro    ····················  ····················  ····················  ····················  ····················  ····················
32899 MN996529.1 |Severe acute respiratory syndrome coro    ····················  ····················  ····················  ····················  ····················  ····················
32900 MN996530.1 |Severe acute respiratory syndrome coro    ····················  ····················  ····················  ····················  ····················  ····················
32901 MN996531.1 |Severe acute respiratory syndrome coro    ····················  ····················  ····················  ····················  ····················  ····················
32902 MN988668.1 |Severe acute respiratory syndrome coro    ····················  ····················  ····················  ····················  ····················  ····················
32903 MN988669.1 |Severe acute respiratory syndrome coro    ····················  ····················  ····················  ····················  ····················  ····················
32904 MN994467.1 |Severe acute respiratory syndrome coro    ····················  ····················  ····················  ····················  ····················  ····················
32905 MN994468.1 |Severe acute respiratory syndrome coro    ····················  ····················  ····················  ····················  ····················  ····················
32906 MN997409.1 |Severe acute respiratory syndrome coro    ····················  ····················  ····················  ····················  ····················  ····················
32907 MN988713.1 |Severe acute respiratory syndrome coro    ····················  ·R··················  ····················  ····················  ····················  ····················
32908 MN938384.1 |Severe acute respiratory syndrome coro    ····················  ····················  ····················  ····················  ····················  ····················
32909 MN975262.1 |Severe acute respiratory syndrome coro    ····················  ····················  ····················  ····················  ····················  ····················
32910 MN985325.1 |Severe acute respiratory syndrome coro    ····················  ····················  ····················  ····················  ····················  ····················
32911 MN908947.3 |Severe acute respiratory syndrome coro    ····················  ····················  ····················  ····················  ····················  ····················
```

**Removed from analysis**

| A | B | C | D | E | F | G | H | I | J | K |
|---|---|---|---|---|---|---|---|---|---|---|
| Database ID | NCBI Description | GCTGCAATCGTGCTACAACT | TGAACTGTTGCGACTACGTG | TTCGGAAGAGACAGGTACGTT | CACACAATCGAAGCGCAGTA | GCTGGTGCTGCAGCTTATTA | AGGGTCAAGTGCACAGTCTA | Incomplete? | Poor quality? | |
| 350 | MW454488.1 \|Severe acute respiratory syndrome coro | ···················· | ···················· | ···················· | ···················· | NNNNNNNNNNNNNNNNNNNN | NNNNNNNNN··········· | | 1 | |
| 523 | MW454668.1 \|Severe acute respiratory syndrome coro | ···················· | ···················· | ···················· | ···················· | ···················· | ··············NNNNNN | | 1 | |
| 543 | MW454689.1 \|Severe acute respiratory syndrome coro | ···················· | ···················· | ···················· | ···················· | NNNNNNNNNNNNNNNNNNNN | ···················· | | 1 | |
| 782 | MW446219.1 \|Severe acute respiratory syndrome coro | ···················· | ···················· | ···················· | ···················· | ··NNNN·············· | ···················· | | 1 | |
| 994 | MW433767.1 \|Severe acute respiratory syndrome coro | ···················· | ···················· | ···················· | ···················· | ···················· | NNNNNNNNNNNNNNNNNNNN | | 1 | |
| 2088 | MW403701.1 \|Severe acute respiratory syndrome coro | ···················· | ···················· | ···················· | ·········NNNNNNNNNNN | ···················· | ···················· | | 1 | |
| 2186 | MW390866.1 \|Severe acute respiratory syndrome coro | ···················· | ···················· | ···················· | ···················· | ·····NNNNNN········· | ···················· | | 1 | |
| 2720 | MW332829.1 \|Severe acute respiratory syndrome coro | ···················· | ···················· | ···················· | ···················· | NNNNNNNNNNNNNNNNNNNN | ···················· | | 1 | |
| 2984 | MW320729.1 \|Severe acute respiratory syndrome coro | ···················· | ···················· | ···················· | ···················· | NNNNNNNNNNNNNNNNNNNN | ···················· | | 1 | |
| 2992 | MW320747.1 \|Severe acute respiratory syndrome coro | ···················· | ···················· | ···················· | ···················· | NNNNNNNNNNNNNNNNNNNN | NNNNNNNNNNNNNNNNNNNN | | 1 | |
| 2994 | MW320750.1 \|Severe acute respiratory syndrome coro | ···················· | ···················· | ···················· | ···················· | NNNNNNNNNNNNNNNNNNNN | ···················· | | 1 | |
| 3009 | MW320771.1 \|Severe acute respiratory syndrome coro | ···················· | ···················· | ···················· | ···················· | N···········S······· | NNNN················ | | 1 | |
| 3054 | MW320864.1 \|Severe acute respiratory syndrome coro | ···················· | ·····R·············· | ···················· | ···················· | ···N················ | NNNNN··············· | | 1 | |
| 3066 | MW320881.1 \|Severe acute respiratory syndrome coro | ···················· | ···················· | ···················· | ···················· | NNNNNNNNNNNNNNNNNNNN | ···················· | | 1 | |
| 3069 | MW320887.1 \|Severe acute respiratory syndrome coro | ···················· | ···················· | ···················· | ···················· | NNNNNNNNNNNNNNNNNN·· | ···················· | | 1 | |
| 3072 | MW320891.1 \|Severe acute respiratory syndrome coro | ···················· | ···················· | ···················· | ···················· | NNNNNNNNNNNNNNNNNNNN | ···················· | | 1 | |
| 3082 | MW320908.1 \|Severe acute respiratory syndrome coro | ···················· | ···················· | ···················· | ···················· | NNNNNNNNNNNNNNNNN··· | ···················· | | 1 | |
| 3085 | MW320913.1 \|Severe acute respiratory syndrome coro | ···················· | ···················· | ···················· | ···················· | NNNNNNNNNNNN········ | NNNN················ | | 1 | |
| 3090 | MW320926.1 \|Severe acute respiratory syndrome coro | ···················· | ···················· | ···················· | ···················· | NNNNNNNNNNNNNNNNNNNN | NN·················· | | 1 | |
| 3093 | MW320930.1 \|Severe acute respiratory syndrome coro | ···················· | ···················· | ···················· | ···················· | NNNNNNNNNNNNNNNNNNNN | NNNNNNNNN··········· | | 1 | |
| 3094 | MW320931.1 \|Severe acute respiratory syndrome coro | ···················· | ···················· | ···················· | ···················· | NNNNNNNNNNNNNNNNNNNN | ··············NNNNNN | | 1 | |
| 3097 | MW320936.1 \|Severe acute respiratory syndrome coro | ···················· | ···················· | ···················· | ···················· | NNNNNNNNNNNNNNNNNNNN | ···················· | | 1 | |
| 3108 | MW320955.1 \|Severe acute respiratory syndrome coro | ···················· | ·····A·············· | ···················· | ···················· | NNNNNNNNNNNNNNNNNNNN | ·····N·············· | | 1 | |
| 3111 | MW320965.1 \|Severe acute respiratory syndrome coro | ···················· | ···················· | ···················· | ···················· | NNNNNNNNNNNNNNNNNNNN | ···················· | | 1 | |
| 3113 | MW320967.1 \|Severe acute respiratory syndrome coro | ···················· | ···················· | ···················· | ···················· | NNNNNNNNNNNNNNNNNNNN | ···················· | | 1 | |
| 3121 | MW320977.1 \|Severe acute respiratory syndrome coro | ···················· | ···················· | ···················· | ···················· | NNNNNNNNNNNN········ | ···················· | | 1 | |
| 3133 | MW320994.1 \|Severe acute respiratory syndrome coro | ···················· | ···················· | ···················· | ···················· | NNNNNNNNNNNNNNNNNNNN | ·············S······ | | 1 | |
| 3137 | MW321002.1 \|Severe acute respiratory syndrome coro | ···················· | ···················· | ···················· | ···················· | NNNNNNNNNNNNNNNNNNNN | ···················· | | 1 | |
| 3152 | MW321037.1 \|Severe acute respiratory syndrome coro | ···················· | ···················· | ···················· | ···················· | NNNNNNN············· | ···················· | | 1 | |
| 3153 | MW321040.1 \|Severe acute respiratory syndrome coro | ···················· | ···················· | ···················· | ···················· | NNNN················ | ···················· | | 1 | |
| 3157 | MW321045.1 \|Severe acute respiratory syndrome coro | ···················· | ···················· | ···················· | ···················· | NNNNNNNNNNNNNNNNN··· | NNNNNNNNN··········· | | 1 | |
| 3160 | MW321049.1 \|Severe acute respiratory syndrome coro | ···················· | ···················· | ···················· | ···················· | ···················· | NNNNNNNNNNNNNNN····· | | 1 | |
| 3172 | MW321065.1 \|Severe acute respiratory syndrome coro | ···················· | ···················· | ···················· | ···················· | NNNNNNNNNNNNNNNNN··· | ···················· | | 1 | |
| 3178 | MW321072.1 \|Severe acute respiratory syndrome coro | ···················· | ···················· | ···················· | ···················· | NNNNNNNNNNNNNNNNNNNN | ···················· | | 1 | |
| 3179 | MW321079.1 \|Severe acute respiratory syndrome coro | ···················· | ···················· | ···················· | ···················· | NNNNNNNNNNNNNNNNNNNN | NNNNN·······NNN····· | | 1 | |
| 3185 | MW321087.1 \|Severe acute respiratory syndrome coro | ···················· | ···················· | ···················· | ···················· | NNNNNNNNNNNNNNNNNNNN | ···················· | | 1 | |
| 3188 | MW321092.1 \|Severe acute respiratory syndrome coro | ···················· | ···················· | ···················· | ···················· | NNNNNNNNNNNNNNNNNNNN | ···················· | | 1 | |
| 3190 | MW321095.1 \|Severe acute respiratory syndrome coro | ···················· | ···················· | ···················· | ···················· | NNNNNNNNNNNNNNNNNNNN | NNNNNNNNNNNNNNNNNNNN | | 1 | |
| 3197 | MW321110.1 \|Severe acute respiratory syndrome coro | ···················· | ···················· | ···················· | ···················· | NNNNNNNNNNNNNNNNNNNN | NNNN··········NNNNNN | | 1 | |
| 3204 | MW321118.1 \|Severe acute respiratory syndrome coro | ···················· | ···················· | ···················· | ···················· | NNNNNNNNNNNNNNNNNNNN | ···················· | | 1 | |
| 3221 | MW321149.1 \|Severe acute respiratory syndrome coro | ···················· | ···················· | ···················· | ···················· | NNNNNNNN············ | ···················· | | 1 | |
| 3225 | MW321158.1 \|Severe acute respiratory syndrome coro | ···················· | ···················· | ···················· | ···················· | NNNNNNNNNNNNNNNNNNNN | ···················· | | 1 | |
| 3232 | MW321172.1 \|Severe acute respiratory syndrome coro | ···················· | ···················· | ···················· | ···················· | NNNNNNNNNNNNNNNNNNNN | ···················· | | 1 | |
| 3255 | MW321218.1 \|Severe acute respiratory syndrome coro | ···················· | ···················· | ···················· | ···················· | NNNNN··············· | ···················· | | 1 | |
| 3270 | MW321248.1 \|Severe acute respiratory syndrome coro | ···················· | ···················· | ···················· | ···················· | NNNNNNNNNNNNNNNNNNNN | ···················· | | 1 | |
| 3274 | MW321253.1 \|Severe acute respiratory syndrome coro | ···················· | ···················· | ···················· | ···················· | NNNNNNNNNNNNNNNNNNNN | NNNNNNNNNNNNNNN····· | | 1 | |
| 3275 | MW321257.1 \|Severe acute respiratory syndrome coro | ···················· | ···················· | ···················· | ···················· | NNNNNNNNNNNNNNNNNNNN | ···················· | | 1 | |
| 3276 | MW321258.1 \|Severe acute respiratory syndrome coro | ···················· | ···················· | ···················· | ···················· | ···················· | NNNNNNNNNNNNNNN····· | | 1 | |
| 3286 | MW321274.1 \|Severe acute respiratory syndrome coro | ···················· | ···················· | ···················· | ···················· | NNNNNNNNNNNNNNNNNNNN | NNNN················ | | 1 | |
| 3289 | MW321277.1 \|Severe acute respiratory syndrome coro | ···················· | ···················· | ···················· | ···················· | NNNNNNNNNNNNNNNNNNNN | ···················· | | 1 | |
| 3291 | MW321280.1 \|Severe acute respiratory syndrome coro | ···················· | ···················· | ···················· | ···················· | NNNNNNNNNNNNNNNNNNNN | ···················· | | 1 | |
| 3294 | MW321284.1 \|Severe acute respiratory syndrome coro | ···················· | ···················· | ···················· | ···················· | NNNNNNNNNNNNNNNNNNNN | ···················· | | 1 | |
| 3300 | MW321296.1 \|Severe acute respiratory syndrome coro | ···················· | ···················· | ···················· | ···················· | NNNNNNNNNNNNNNNNNNNN | ···················· | | 1 | |
| 3305 | MW321310.1 \|Severe acute respiratory syndrome coro | ···················· | ···················· | ···················· | ···················· | NNNN················ | ···················· | | 1 | |
| 3312 | MW321328.1 \|Severe acute respiratory syndrome coro | ···················· | ···················· | ···················· | ···················· | ···················· | NNNNNNNNNNNNNNNNN··· | | 1 | |
| 3317 | MW321342.1 \|Severe acute respiratory syndrome coro | ···················· | ···················· | ···················· | ···················· | ···················· | ·····NNNN··········· | | 1 | |
| 3326 | MW321364.1 \|Severe acute respiratory syndrome coro | ···················· | ···················· | ···················· | ···················· | NNNNNNNNNNNNNNNNNNNN | ···················· | | 1 | |
| 3327 | MW321366.1 \|Severe acute respiratory syndrome coro | ···················· | ···················· | ···················· | ···················· | NNNNNNNNNNNNNNNNNNNN | ···················· | | 1 | |
| 3334 | MW321376.1 \|Severe acute respiratory syndrome coro | ···················· | ···················· | ···················· | ···················· | NNNNNNN············· | NNNNNNNNNNNNNNN····· | | 1 | |
| 3352 | MW321413.1 \|Severe acute respiratory syndrome coro | ···················· | ···················· | ···················· | ···················· | NNNNNNNNNNNNNNNNNNNN | ···················· | | 1 | |
| 3353 | MW321414.1 \|Severe acute respiratory syndrome coro | ···················· | ···················· | ···················· | ···················· | ················NNNN | ···················· | | 1 | |
| 3360 | MW321428.1 \|Severe acute respiratory syndrome coro | ···················· | ···················· | ···················· | ···················· | NNNNNNNNNNNNNNNNNNNN | ···················· | | 1 | |
| 3363 | MW321432.1 \|Severe acute respiratory syndrome coro | ···················· | ···················· | ···················· | ···················· | NNNNNNN············· | ···················· | | 1 | |
| 3403 | MW309478.1 \|Severe acute respiratory syndrome coro | ···················· | ···················· | ···················· | ···················· | NNNNNNNNNNNNNNNNNNNN | ···················· | | 1 | |
| 3404 | MW309481.1 \|Severe acute respiratory syndrome coro | ···················· | ···················· | ···················· | ···················· | NNNNNNNNNNNNNNNNNNNN | NNNNN·······NNNNN·NN | | 1 | |
| 3406 | MW309483.1 \|Severe acute respiratory syndrome coro | ···················· | ···················· | ···················· | ···················· | NNNNNNNNNNNN········ | ···················· | | 1 | |
| 3408 | MW309487.1 \|Severe acute respiratory syndrome coro | ···················· | ···················· | ···················· | ···················· | ··NNNNNNNNNNNNNNNNNN | ···················· | | 1 | |

| | | | | | | | | | |
|---|---|---|---|---|---|---|---|---|---|
| 3478 | MW306321.1 | \|Severe acute respiratory syndrome coro | ···················· | ·A·················· | ···················· | ···················· | NNNN················ | ···················· | 1 |
| 3479 | MW306322.1 | \|Severe acute respiratory syndrome coro | ···················· | ·A·················· | ···················· | ···················· | NNNNNNNNNNNNNNNN···· | ···················· | 1 |
| 3483 | MW306327.1 | \|Severe acute respiratory syndrome coro | ···················· | ·A·················· | ···················· | ···················· | NNNNNNNNN··········· | ···················· | 1 |
| 3511 | MW306359.1 | \|Severe acute respiratory syndrome coro | ···················· | ·A·················· | ···················· | ···················· | NNNNNNNNNNNNNNNNNNNN | ···················· | 1 |
| 3516 | MW306364.1 | \|Severe acute respiratory syndrome coro | ···················· | NNNNNNNNNNNNNNNNNNNN | ···················· | ···················· | ···················· | ···················· | 1 |
| 3545 | MW306405.1 | \|Severe acute respiratory syndrome coro | ···················· | ·A·················· | ···················· | ···················· | ···················· | NNNNNNNNNNNNNNN····· | 1 |
| 3694 | MW286553.1 | \|Severe acute respiratory syndrome coro | ···················· | ···················· | ···················· | ···················· | NNNN················ | ···················· | 1 |
| 3701 | MW286580.1 | \|Severe acute respiratory syndrome coro | ···················· | ···················· | ···················· | ···················· | NNNNNNNNNNNNNNNNNNNN | ···················· | 1 |
| 3715 | MW286605.1 | \|Severe acute respiratory syndrome coro | ···················· | ···················· | ···················· | ···················· | NNNNNNNNNNNNNNNNNNNN | ···················· | 1 |
| 3868 | MW276254.1 | \|Severe acute respiratory syndrome coro | ···················· | ···················· | ···················· | ···················· | ··NNNNNNNNNNNNNNNNNN | ···················· | 1 |
| 3950 | MW276360.1 | \|Severe acute respiratory syndrome coro | ···················· | ···················· | ···················· | ···················· | NNNNNNNNNNNN·NNNNNNN | ···················· | 1 |
| 3973 | MW276385.1 | \|Severe acute respiratory syndrome coro | ···················· | ·A·················· | ···················· | ···················· | NNNN················ | ···················· | 1 |
| 3974 | MW276386.1 | \|Severe acute respiratory syndrome coro | ···················· | ·A·················· | ···················· | ···················· | NNNN················ | ··NNNNNNNNNNNNN····· | 1 |
| 4019 | MW276436.1 | \|Severe acute respiratory syndrome coro | ···················· | ·A·················· | ···················· | ···················· | NNNNNNNNNNN········· | NN·······NNNNNN····· | 1 |
| 4085 | MW276517.1 | \|Severe acute respiratory syndrome coro | ···················· | ···················· | ···················· | ···················· | NNNNNNNNNNNNNNNN···· | ···················· | 1 |
| 4087 | MW276520.1 | \|Severe acute respiratory syndrome coro | ···················· | ···················· | ···················· | ···················· | NNNNNNNNNNNNNNNNNNNN | ·NNN····NNNNNNN····· | 1 |
| 4089 | MW276522.1 | \|Severe acute respiratory syndrome coro | ···················· | ···················· | ···················· | ···················· | NNNNNNNNNNNNNNNNNNNN | NNNNNNNNNNNNNNN····· | 1 |
| 4090 | MW276523.1 | \|Severe acute respiratory syndrome coro | ···················· | ···················· | ···················· | ···················· | NNNNNNNNNNNNNNNNNNNN | NNNN···············N | 1 |
| 4092 | MW276525.1 | \|Severe acute respiratory syndrome coro | ···················· | ···················· | ···················· | ···················· | ·····NNNN··········· | ···················· | 1 |
| 4107 | MW276547.1 | \|Severe acute respiratory syndrome coro | ···················· | ···················· | ···················· | ···················· | NNNNNNNNNNNNNN······ | ···················· | 1 |
| 4108 | MW276548.1 | \|Severe acute respiratory syndrome coro | ···················· | ···················· | ···················· | ···················· | ···················N | NNNNNNNNNNNNNNNNNNNN | 1 |
| 4125 | MW276565.1 | \|Severe acute respiratory syndrome coro | ···················· | ·A·················· | ···················· | ···················· | NNNNNNNNNNNNNNNNNNNN | ···················· | 1 |
| 4146 | MW276588.1 | \|Severe acute respiratory syndrome coro | ···················· | ·A·················· | ···················· | ···················· | NNNNNNNNNNNNNNNNNNNN | ···················· | 1 |
| 4148 | MW276590.1 | \|Severe acute respiratory syndrome coro | ···················· | ·A·················· | ···················· | ···················· | NNNNNNNNNNNNNNNNNNNN | ···················· | 1 |
| 4158 | MW276605.1 | \|Severe acute respiratory syndrome coro | ···················· | ···················· | ···················· | ···················· | NNNNNNNNNNNNNN······ | ···················· | 1 |
| 4180 | MW276632.1 | \|Severe acute respiratory syndrome coro | ···················· | ·A·················· | ···················· | ···················· | NNNNNNNNNNNNNNNNNNNN | ···················· | 1 |
| 4184 | MW276637.1 | \|Severe acute respiratory syndrome coro | ···················· | ···················· | ···················· | ···················· | NNNNNNNNNNNNNNNNN··· | ···················· | 1 |
| 4188 | MW276642.1 | \|Severe acute respiratory syndrome coro | ···················· | ···················· | ···················· | ···················· | NNNNNNNNNNNNNNNNN··· | ···················· | 1 |
| 4205 | MW276665.1 | \|Severe acute respiratory syndrome coro | ···················· | ···················· | ···················· | ···················· | NNNNNNNNNNNNNNNNNNNN | ···················· | 1 |
| 4208 | MW276670.1 | \|Severe acute respiratory syndrome coro | ···················· | ·A·················· | ···················· | ···················· | NNNNNNNNNNNNNNNNNNNN | ···················· | 1 |
| 4265 | MW276746.1 | \|Severe acute respiratory syndrome coro | ···················· | ···················· | ···················· | ···················· | ······NNNNNNNN······ | ···················· | 1 |
| 4275 | MW276757.1 | \|Severe acute respiratory syndrome coro | ···················· | ···················· | ···················· | ···················· | ···················· | NNNNNNNNN··········· | 1 |
| 4276 | MW276759.1 | \|Severe acute respiratory syndrome coro | ···················· | ···················· | ···················· | ···················· | ···················· | NNNNNNNNNNNNNNN····· | 1 |
| 4295 | MW276781.1 | \|Severe acute respiratory syndrome coro | ···················· | ···················· | ···················· | ···················· | NNNNNNNNNNNNNNNNNNNN | ···················· | 1 |
| 4380 | MW276887.1 | \|Severe acute respiratory syndrome coro | ···················· | ···················· | ···················· | ···················· | ···················· | NNNN················ | 1 |
| 4381 | MW276888.1 | \|Severe acute respiratory syndrome coro | ···················· | ···················· | ···················· | ···················· | ·····NNNNNNNNNNNNNNN | ···················· | 1 |
| 4403 | MW276923.1 | \|Severe acute respiratory syndrome coro | ···················· | ···················· | ···················· | ···················· | NNNNNNNNNNNNNNNNNNNN | ···················· | 1 |
| 4429 | MW276954.1 | \|Severe acute respiratory syndrome coro | ···················· | ···················· | ···················· | ···················· | ······NNNNNNNNNNNNNN | ···················· | 1 |
| 4483 | MW277030.1 | \|Severe acute respiratory syndrome coro | ···················· | ···················· | ···················· | ···················· | NNNNNNNNNNNNNNNNNNNN | ···················· | 1 |
| 4485 | MW277032.1 | \|Severe acute respiratory syndrome coro | ···················· | ···················· | ···················· | ···················· | NNNNNNNNNNNNNNNNNNNN | ···················· | 1 |
| 4491 | MW277040.1 | \|Severe acute respiratory syndrome coro | ···················· | ···················· | ···················· | ···················· | ················NNNN | ···················· | 1 |
| 4563 | MW277144.1 | \|Severe acute respiratory syndrome coro | ···················· | ···················· | ···················· | ···················· | NNNNNNNNNNNNNNNNNNNN | ···················· | 1 |
| 4566 | MW277148.1 | \|Severe acute respiratory syndrome coro | ···················· | ···················· | ···················· | ···················· | NNNNNNNNNNNNNNNNNNNN | ···················· | 1 |
| 4567 | MW277149.1 | \|Severe acute respiratory syndrome coro | ···················· | ······A············· | ···················· | ···················· | NNNN················ | ···················· | 1 |
| 4602 | MW277196.1 | \|Severe acute respiratory syndrome coro | ···················· | ······A············· | ···················· | ···················· | NNNNNNNNNNNNNNNNNNNN | NNNNNNNNNNNNNNNNNNNN | 1 |
| 4695 | MW277315.1 | \|Severe acute respiratory syndrome coro | ···················· | ···················· | ···················· | ···················· | NNNNNNNNNNNN········ | ···················· | 1 |
| 4750 | MW277387.1 | \|Severe acute respiratory syndrome coro | ···················· | ···················· | ···················· | ···················· | NNNNNNNNNNNNNNN····· | ···················· | 1 |
| 4766 | MW277405.1 | \|Severe acute respiratory syndrome coro | ···················· | ···················· | ···················· | ···················· | NNNNNNNNNNNNNNNNNNNN | ···················· | 1 |
| 4789 | MW277435.1 | \|Severe acute respiratory syndrome coro | ···················· | ···················· | ···················· | ···················· | NNNN················ | ···················· | 1 |
| 4820 | MW277475.1 | \|Severe acute respiratory syndrome coro | ···················· | ···················· | ···················· | ···················· | NNNNNNNNNNNNNNNNNNNN | ···················· | 1 |
| 4849 | MW277509.1 | \|Severe acute respiratory syndrome coro | ···················· | ···················· | ···················· | ···················· | NNNNNNNNNNNNNNNNNNNN | ···················· | 1 |
| 4918 | MW277603.1 | \|Severe acute respiratory syndrome coro | ···················· | ···················· | ···················· | ···················· | NNNNNNNNNNNNNNNNNNNN | ···················· | 1 |
| 6056 | MW206400.1 | \|Severe acute respiratory syndrome coro | ···················· | ·············A······ | ···················· | ···················· | NNNNNNNNNNNNNNNNNNNN | ···················· | 1 |
| 6057 | MW206402.1 | \|Severe acute respiratory syndrome coro | ···················· | ·············A······ | ···················· | ···················· | NNNN················ | ···················· | 1 |
| 6071 | MW206453.1 | \|Severe acute respiratory syndrome coro | ···················· | ···················· | ···················· | ···················· | NNNNNNNNNNNNNNNNNNNN | ···················· | 1 |
| 6073 | MW206456.1 | \|Severe acute respiratory syndrome coro | ···················· | ···················· | ···················· | ···················· | NNNNNNNNNNNNNNNNNNNN | ···················· | 1 |
| 6074 | MW206458.1 | \|Severe acute respiratory syndrome coro | ···················· | ···················· | ···················· | ···················· | NNNNNNNNNNNNNNNNNNNN | ···················· | 1 |
| 6083 | MW206488.1 | \|Severe acute respiratory syndrome coro | ···················· | ···················· | ···················· | ···················· | NNNNNNNNNNNNNNNNNNNN | ···················· | 1 |
| 6098 | MW206515.1 | \|Severe acute respiratory syndrome coro | ···················· | ···················· | ···················· | ···················· | NNNNNNNNNNNNNNNNNNNN | ···················· | 1 |
| 6122 | MW206553.1 | \|Severe acute respiratory syndrome coro | ···················· | ···················· | ···················· | ···················· | ········NNNNNNNNNNNN | ···················· | 1 |
| 6125 | MW206559.1 | \|Severe acute respiratory syndrome coro | ···················· | ···················· | ···················· | ···················· | NNNNNNNNNNNNNNNNNNNN | ···················· | 1 |
| 6148 | MW206609.1 | \|Severe acute respiratory syndrome coro | ···················· | ···················· | ···················· | ···················· | NNNNNNNNNNNNNNNNNNNN | ···················· | 1 |
| 7767 | MW192063.1 | \|Severe acute respiratory syndrome coro | ···················· | ·A·················· | ···················· | ···················· | ··NNNNNNNNNN········ | ···················· | 1 |
| 7916 | MW183949.1 | \|Severe acute respiratory syndrome coro | ···················· | ···················· | ···················· | ···················· | NNNNNNNNNNN········· | ···················· | 1 |
| 7930 | MW183969.1 | \|Severe acute respiratory syndrome coro | ···················· | ···················· | ···················· | ···················· | NNNNNNNNNNN········· | ···················· | 1 |
| 7949 | MW183992.1 | \|Severe acute respiratory syndrome coro | ···················· | ···················· | ···················· | ···················· | NNNNNN·············· | ···················· | 1 |
| 7961 | MW184005.1 | \|Severe acute respiratory syndrome coro | ···················· | ···················· | ···················· | ···················· | NNNNNNNNNNNNNNNNNNNN | ···················· | 1 |
| 8161 | MW184267.1 | \|Severe acute respiratory syndrome coro | ···················· | ···················· | ···················· | ···················· | NNNNNNNNNNNNNN······ | ···················· | 1 |
| 8178 | MW184284.1 | \|Severe acute respiratory syndrome coro | ···················· | ···················· | ···················· | ···················· | NNNNNNNNNNNNNNNNNNNN | ···················· | 1 |
| 8255 | MW184383.1 | \|Severe acute respiratory syndrome coro | ···················· | ···················· | ···················· | ···················· | ···················· | NNNN················ | 1 |
| 8266 | MW184397.1 | \|Severe acute respiratory syndrome coro | ···················· | ···················· | ···················· | ···················· | NNNNNNNNNNNNNNNNNNNN | ···················· | 1 |
| 8271 | MW184403.1 | \|Severe acute respiratory syndrome coro | ···················· | ···················· | ···················· | ···················· | NNNNNNNNNNNNNNNNNNNN | ···················· | 1 |
| 8303 | MW184440.1 | \|Severe acute respiratory syndrome coro | ···················· | ···················· | ···················· | ···················· | NNNNNNNNNNNN········ | ···················· | 1 |
| 8312 | MW184460.1 | \|Severe acute respiratory syndrome coro | ···················· | ···················· | ···················· | ···················· | NNNNNNNNNNNNNNNNNNNN | ··················NN | 1 |
| 8331 | MW184486.1 | \|Severe acute respiratory syndrome coro | ···················· | ···················· | ···················· | ···················· | NNNNNNNNNNNNNNNNNNNN | ···················· | 1 |
| 8383 | MW184559.1 | \|Severe acute respiratory syndrome coro | ···················· | ···················· | ···················· | ···················· | ·NNN················ | NNNNNNNNNNNNNNNNNNNN | 1 |
| 8406 | MW184587.1 | \|Severe acute respiratory syndrome coro | ···················· | ···················· | ···················· | ···················· | NNNNNNNNNNNNNNNNNNNN | ·NNNNNNNN·····NNNNNN | 1 |

| | | | | | | | | | |
|---|---|---|---|---|---|---|---|---|---|
| 8420 | MW184607.1 | \|Severe acute respiratory syndrome coro | ···················· | ···················· | ···················· | ···················· | NNNNNNNNNNNNNNNNNNNN | ···················· | 1 |
| 8425 | MW184613.1 | \|Severe acute respiratory syndrome coro | ···················· | ···················· | ···················· | NNNNNNNNNNNNNNNNNNNN | ···················· | ···················· | 1 |
| 8438 | MW184630.1 | \|Severe acute respiratory syndrome coro | ···················· | ···················· | ···················· | ···················· | ···················· | NNNNNNNNNNNNNNN····· | 1 |
| 8451 | MW184645.1 | \|Severe acute respiratory syndrome coro | ···················· | ···················· | ···················· | ···················· | NNNNNNNNNNNNNNNNNNNN | ···················· | 1 |
| 8455 | MW184649.1 | \|Severe acute respiratory syndrome coro | ···················· | ···················· | ···················· | ···················· | ···················· | NNNNNNNNNNNNNNNNNNNN | 1 |
| 8572 | MW184814.1 | \|Severe acute respiratory syndrome coro | ···················· | ···················· | ···················· | ···················· | ···················· | NNNNNNNNNNNNNNN····· | 1 |
| 8574 | MW184816.1 | \|Severe acute respiratory syndrome coro | ···················· | ···················· | ···················· | ···················· | ···················· | NNNNNNNNNNNNNNN····· | 1 |
| 8599 | MW184848.1 | \|Severe acute respiratory syndrome coro | ···················· | ···················· | ···················· | ···················· | NNNNNNNNNNNNNNNNNNNN | ···················· | 1 |
| 8632 | MW184889.1 | \|Severe acute respiratory syndrome coro | ···················· | ······A············· | ···················· | ···················· | NNNNNNNNN··········· | ···················· | 1 |
| 8668 | MW184936.1 | \|Severe acute respiratory syndrome coro | ···················· | ···················· | ···················· | ···················· | NNNNNNNNNNNNNNNNNNNN | ···················· | 1 |
| 8751 | MW185050.1 | \|Severe acute respiratory syndrome coro | ···················· | ···················· | ···················· | ···················· | NNNNNNNNNNNN········ | ···················· | 1 |
| 8776 | MW185084.1 | \|Severe acute respiratory syndrome coro | ···················· | ···················· | ···················· | ···················· | NNNNNNNNNNNNNNNNNNNN | ···················· | 1 |
| 8790 | MW185103.1 | \|Severe acute respiratory syndrome coro | ···················· | ···················· | ···················· | ···················· | NNNNNNNNNNNNNNNNN··· | ···················· | 1 |
| 8791 | MW185104.1 | \|Severe acute respiratory syndrome coro | ···················· | ······A············· | ···················· | ···················· | NNNNNNNNNNNNNNNNNNNN | ···················· | 1 |
| 8807 | MW185125.1 | \|Severe acute respiratory syndrome coro | ···················· | ···················· | ···················· | ···················· | NNNNNNNNNNNNNNNNNNNN | ···················· | 1 |
| 8854 | MW185187.1 | \|Severe acute respiratory syndrome coro | ···················· | ···················· | ···················· | ···················· | NNNNNNNNNNNNNNNNNNNN | ···················· | 1 |
| 8861 | MW185197.1 | \|Severe acute respiratory syndrome coro | ···················· | ···················· | ···················· | ···················· | NNNNNNNNNNNNNNNNNNNN | ···················· | 1 |
| 8864 | MW185200.1 | \|Severe acute respiratory syndrome coro | ···················· | ···················· | ···················· | ···················· | NNNNNNNNNNNNNNNNNNNN | ···················· | 1 |
| 8886 | MW185232.1 | \|Severe acute respiratory syndrome coro | ···················· | ···················· | ···················· | ···················· | NNNN················ | ···················· | 1 |
| 8926 | MW185286.1 | \|Severe acute respiratory syndrome coro | ···················· | ···················· | ···················· | ···················· | NNNNNNNNNNNNNNNNNNNN | ···················· | 1 |
| 8953 | MW185324.1 | \|Severe acute respiratory syndrome coro | ···················· | ···················· | ···················· | ···················· | NNNNNNNNNNNNNNNNNNNN | ···················· | 1 |
| 8980 | MW185366.1 | \|Severe acute respiratory syndrome coro | ···················· | ···················· | ···················· | ···················· | NNNNNNNNNNNNNNNNNNNN | ···················· | 1 |
| 9023 | MW185422.1 | \|Severe acute respiratory syndrome coro | ···················· | ···················· | ···················· | ···················· | NNNNNNNNNNNNNNNNN··· | ···················· | 1 |
| 9081 | MW185509.1 | \|Severe acute respiratory syndrome coro | ···················· | ···················· | ···················· | NNNNNNNNNNNNNNNNNNNN | ···················· | ···················· | 1 |
| 9280 | MW185774.1 | \|Severe acute respiratory syndrome coro | ···················· | ···················· | ···················· | ···················· | NNNNNNNNNNNNNN······ | ···················· | 1 |
| 9314 | MW173446.1 | \|Severe acute respiratory syndrome coro | NNNNNNNNNNNNNNNNNNNN | ···················· | ···················· | ···················· | ···················· | ···················· | 1 |
| 9341 | MW166067.1 | \|Severe acute respiratory syndrome coro | ···················· | ···················· | ···················· | ···················· | NNNNNNNNNNNNNNNNNNNN | N··················· | 1 |
| 9344 | MW166071.1 | \|Severe acute respiratory syndrome coro | ···················· | ···················· | ···················· | ···················· | NNNNNNNNNNNNNNNNNN·· | ···················· | 1 |
| 9360 | MW166095.1 | \|Severe acute respiratory syndrome coro | ···················· | ···················· | ···················· | ···················· | NNNNNNNNNNNNNNNNN··· | ···················· | 1 |
| 9465 | MW153041.1 | \|Severe acute respiratory syndrome coro | ···················· | ···················· | ···················· | ···················· | NNNNNNNNNNNNNNNNNNNN | NNNN················ | 1 |
| 9466 | MW153043.1 | \|Severe acute respiratory syndrome coro | ···················· | ···················· | ···················· | ···················· | NNNNNNNNNNNNNNNNNNNN | ···················· | 1 |
| 9468 | MW153049.1 | \|Severe acute respiratory syndrome coro | ···················· | ···················· | ···················· | ···················· | ·NNNNNN············· | ···················· | 1 |
| 9470 | MW153052.1 | \|Severe acute respiratory syndrome coro | ···················· | ···················· | ···················· | ···················· | ·············NNNN··· | NNNNNNNNN··········· | 1 |
| 9471 | MW153058.1 | \|Severe acute respiratory syndrome coro | ···················· | ···················· | ···················· | ···················· | NNNNNNNNNNNNNNNNNNNN | ·······NNNNNNNNNNNNN | 1 |
| 9472 | MW153059.1 | \|Severe acute respiratory syndrome coro | ···················· | ···················· | ···················· | ···················· | NNNNNNNNNNNNNNNNNNNN | NNNNNNNNNNNNNNN···NN | 1 |
| 9477 | MW153077.1 | \|Severe acute respiratory syndrome coro | ···················· | ···················· | ···················· | ···················· | NNNNNNNNNNNNNNNNNNNN | NNNNNNNNNNNNNNNNNNNN | 1 |
| 9480 | MW153080.1 | \|Severe acute respiratory syndrome coro | ···················· | ···················· | ···················· | ···················· | NNNNNNNNNNNNNNNNNNNN | ···············NNNNN | 1 |
| 10387 | MW154128.1 | \|Severe acute respiratory syndrome coro | ···················· | ···················· | ···················· | ···················· | NNNNNNNNNNNNNNNNNNNN | ···················· | 1 |
| 10978 | MW154855.1 | \|Severe acute respiratory syndrome coro | ···················· | ···················· | ···················· | ···················· | NNNNNNNNNNNNNNNN···· | ···················· | 1 |
| 10982 | MW154859.1 | \|Severe acute respiratory syndrome coro | ···················· | ···················· | ···················· | ···················· | NNNNNNNNNNNNNNN····· | ····Y··············· | 1 |
| 11038 | MW154936.1 | \|Severe acute respiratory syndrome coro | ···················· | ···················· | ···················· | ···················· | NNNNNNNNNNNNNNNNNNNN | ···················· | 1 |
| 11096 | MW155007.1 | \|Severe acute respiratory syndrome coro | ···················· | ···················· | ···················· | ···················· | ···················· | NNNNNNNNNNNNNNN····· | 1 |
| 11097 | MW155008.1 | \|Severe acute respiratory syndrome coro | ···················· | ···················· | ···················· | ···················· | ···················· | NNNNNNNNNNNNNNNNNNNN | 1 |
| 11107 | MW155020.1 | \|Severe acute respiratory syndrome coro | ···················· | ···················· | ···················· | ···················· | NNNNNNNNNNNNNNNNNNNN | ···················· | 1 |
| 11109 | MW155028.1 | \|Severe acute respiratory syndrome coro | ···················· | ···················· | ···················· | ···················· | NNNNNNNNNNNNNNNNNNNN | ···················· | 1 |
| 11353 | MW155362.1 | \|Severe acute respiratory syndrome coro | ···················· | ···················· | ···················· | ···················· | ···················· | ·············NNNNNNN | 1 |
| 11380 | MW155402.1 | \|Severe acute respiratory syndrome coro | ···················· | ···················· | ···················· | ···················· | ···················· | NNNNNNNNNNNNNNNNN··· | 1 |
| 11550 | MW155633.1 | \|Severe acute respiratory syndrome coro | ···················· | ···················· | ···················· | ···················· | NNNNNNNNNNNNNNNNN··· | ···················· | 1 |
| 11573 | MW155658.1 | \|Severe acute respiratory syndrome coro | ···················· | ···················· | ···················· | ···················· | ···················· | NNNNNNNNNNNNNNN····· | 1 |
| 11645 | MW155742.1 | \|Severe acute respiratory syndrome coro | ···················· | ···················· | ···················· | ···················· | NNNNNNNNNNNNNNNNNNNN | ···················· | 1 |
| 11655 | MW155759.1 | \|Severe acute respiratory syndrome coro | ···················· | ···················· | ···················· | ···················· | NNNNNNNNNN·········· | ···················· | 1 |
| 11706 | MW155822.1 | \|Severe acute respiratory syndrome coro | ···················· | ···················· | ···················· | ···················· | NNNNNNNNNNNNNNNNNNNN | ···················· | 1 |
| 12049 | MW156272.1 | \|Severe acute respiratory syndrome coro | ···················· | ···················· | ···················· | ···················· | ··NNNNNNNNNN········ | ···················· | 1 |
| 12054 | MW156278.1 | \|Severe acute respiratory syndrome coro | ···················· | ···················· | ···················· | ···················· | ···················· | NNNNNNNN············ | 1 |
| 12071 | MW156296.1 | \|Severe acute respiratory syndrome coro | ···················· | ···················· | ···················· | ···················· | ···················· | NNNNNNNNNNNNNNN····· | 1 |
| 12105 | MW156337.1 | \|Severe acute respiratory syndrome coro | ···················· | ···················· | ···················· | ···················· | NNNNNNNNNNNNNNNNNNNN | ···················· | 1 |
| 12290 | MW156557.1 | \|Severe acute respiratory syndrome coro | ···················· | ···················· | ···················· | ···················· | NNNNNNNNNNNNNNNNNNNN | NNNNNNNNNNNNNNNN···· | 1 |
| 12326 | MW156597.1 | \|Severe acute respiratory syndrome coro | ···················· | ···················· | ···················· | ···················· | NNNNNNNNNNNNNNNNNNNN | N··················· | 1 |
| 12379 | MW156660.1 | \|Severe acute respiratory syndrome coro | ···················· | ···················· | ···················· | ···················· | NNNNNNNNNNNNNNNNNNNN | ···················· | 1 |
| 12380 | MW156661.1 | \|Severe acute respiratory syndrome coro | ···················· | ···················· | ···················· | ···················· | NNNNNNNNNNNNNNNNNNNN | ···················· | 1 |
| 12386 | MW156668.1 | \|Severe acute respiratory syndrome coro | ···················· | ···················· | ···················· | ···················· | ··NNNNNNNNNN········ | NNNNNNNNNNNNNNN····· | 1 |
| 12392 | MW156677.1 | \|Severe acute respiratory syndrome coro | ···················· | ···················· | ···················· | ···················· | NNNNNNNNNNNNNNNNNNNN | ···················· | 1 |
| 12504 | MW156838.1 | \|Severe acute respiratory syndrome coro | ···················· | ···················· | ···················· | ···················· | NNNNNNNNNNNNNNNNNNNN | ···················· | 1 |
| 12544 | MW156892.1 | \|Severe acute respiratory syndrome coro | ···················· | ···················· | ···················· | ···················· | ···················N | NNNNNNNNN··········· | 1 |
| 12664 | MW157070.1 | \|Severe acute respiratory syndrome coro | ···················· | ···················· | ···················· | ···················· | NNNNNNNNNNNNNNNNN··· | ···················· | 1 |
| 12670 | MW157079.1 | \|Severe acute respiratory syndrome coro | ···················· | ···················· | ···················· | ···················· | NNNNNNNNNNNNNNNNNNNN | ···················· | 1 |
| 12675 | MW157086.1 | \|Severe acute respiratory syndrome coro | ···················· | ···················· | ···················· | ···················· | NNNNNNNNNNNNNNNNNNNN | NNNN················ | 1 |
| 12728 | MW157178.1 | \|Severe acute respiratory syndrome coro | ···················· | ···················· | ···················· | ···················· | ···················· | NNNNNNNNNNNNNNNNNNNN | 1 |
| 12839 | MW134237.1 | \|Severe acute respiratory syndrome coro | ···················· | ·A·················· | ···················· | ···················· | NNNNNNNNNNNN········ | ···················· | 1 |
| 14634 | MW067695.1 | \|Severe acute respiratory syndrome coro | ···················· | ···················· | ···················· | ···················· | ···NNNNNNNNNNNNNNNNN | ···················· | 1 |
| 14840 | MW056112.1 | \|Severe acute respiratory syndrome coro | ···················· | ···················· | ···················· | ···················· | NNNNNNNNNNNNNNNNNNNN | ···················· | 1 |
| 14843 | MW056116.1 | \|Severe acute respiratory syndrome coro | ···················· | ···················· | ···················· | ···················· | NNNNNNNNNNNNNNNNNNNN | ···················· | 1 |
| 14853 | MW056135.1 | \|Severe acute respiratory syndrome coro | ···················· | ···················· | ···················· | ···················· | NNNNNNNNNNNNNNNNNNNN | ···················· | 1 |
| 14872 | MW056171.1 | \|Severe acute respiratory syndrome coro | ···················· | ···················· | ···················· | ···················· | ·············NNNN··· | ···················· | 1 |
| 15050 | MW040617.1 | \|Severe acute respiratory syndrome coro | ···················· | ···················· | ···················· | ···················· | NNNNNNNNNNNNNNNNNNNN | ···················· | 1 |
| 15067 | MW040639.1 | \|Severe acute respiratory syndrome coro | ···················· | ···················· | ···················· | ···················· | NNNNNNN············· | NNN················· | 1 |
| 15077 | MW040649.1 | \|Severe acute respiratory syndrome coro | ···················· | ···················· | ·········T·········· | ···················· | NNNNNNNNNNNNNNNNNNNN | NNNNNNNNNNNNNNN····· | 1 |

| | | | | | | | | | |
|---|---|---|---|---|---|---|---|---|---|
| 15091 | MW040669.1 | \|Severe acute respiratory syndrome coro | ···················· | ···················· | ···················· | ···················· | ···················· | NNNNN··············· | 1 |
| 15345 | MW035992.1 | \|Severe acute respiratory syndrome coro | ···················· | ···················· | ···················· | ···················· | NNNN················ | ···················· | 1 |
| 15640 | MW023435.1 | \|Severe acute respiratory syndrome coro | ···················· | ···················· | ···················· | ···················· | ··NNNNNNNNNNNNNNNNNN | ···················· | 1 |
| 15656 | MW023459.1 | \|Severe acute respiratory syndrome coro | ···················· | ···················· | ···················· | ···················· | NNNNNNNNNNNNNNNNNNNN | ···················· | 1 |
| 15666 | MW023480.1 | \|Severe acute respiratory syndrome coro | ···················· | ···················· | ···················· | ···················· | NNNNNNNNNNNNNN······ | ···················· | 1 |
| 15668 | MW023483.1 | \|Severe acute respiratory syndrome coro | ···················· | ···················· | ···················· | ···················· | NNNNNNNNNNNNNNNNNNNN | ···················· | 1 |
| 15681 | MW023523.1 | \|Severe acute respiratory syndrome coro | ···················· | ···················· | ···················· | ···················· | NNNNNNNNNNNNNNNNNNNN | ···················· | 1 |
| 15683 | MW023532.1 | \|Severe acute respiratory syndrome coro | ···················· | ···················· | ···················· | ···················· | NNNNNNNNNNNNNNNNNN·· | NNNNNNNNNNNNNNN····· | 1 |
| 15688 | MW023555.1 | \|Severe acute respiratory syndrome coro | ···················· | ···················· | ···················· | ···················· | NNNNNNNNNNNNNNNNNNNN | ··········NNN······· | 1 |
| 15689 | MW023558.1 | \|Severe acute respiratory syndrome coro | ···················· | ···················· | ···················· | ···················· | NNNNNNNNNNNNNNNNNNNN | NNNNNNNNNNNNNNN····· | 1 |
| 15690 | MW023562.1 | \|Severe acute respiratory syndrome coro | ···················· | ···················· | ···················· | ···················· | NNNNNNNNNNNNNNNNNNNN | ···················· | 1 |
| 16287 | MT981421.1 | \|Severe acute respiratory syndrome coro | ···················· | ···················· | ···················· | ···················· | ·NNNNNNNN··········· | ···················· | 1 |
| 16340 | MT966079.1 | \|Severe acute respiratory syndrome coro | ···················· | ···················· | ···················· | ···················· | NNNNNNNNNNNNNNNNNNNN | ···················· | 1 |
| 16341 | MT966081.1 | \|Severe acute respiratory syndrome coro | ···················· | ···················· | ···················· | ···················· | NNNN············NNNN | NNN·············NN·· | 1 |
| 16353 | MT966100.1 | \|Severe acute respiratory syndrome coro | ···················· | ···················· | ···················· | ···················· | NNNNNNNNNNNNNNNNNNNN | NNNNNNNNNNNNNN······ | 1 |
| 16359 | MT966107.1 | \|Severe acute respiratory syndrome coro | ···················· | ···················· | ···················· | ···················· | ·NNNNNNNNNNNNNNNNNNN | NNNNNNNNNNNNNNNNNNNN | 1 |
| 16360 | MT966108.1 | \|Severe acute respiratory syndrome coro | ···················· | ···················· | ···················· | ···················· | NNNNNNNNNNNNNNNNNNNN | ···················· | 1 |
| 16367 | MT966140.1 | \|Severe acute respiratory syndrome coro | ···················· | ···················· | ···················· | ···················· | ·NNNNNNNNNNNNN······ | ···················· | 1 |
| 16379 | MT966158.1 | \|Severe acute respiratory syndrome coro | ···················· | ···················· | ···················· | ···················· | NNNNNNNNNNNNNNNNNNNN | ···················· | 1 |
| 16381 | MT966160.1 | \|Severe acute respiratory syndrome coro | ···················· | ···················· | ···················· | ···················· | NNNNNNNNNNNNNNNNNNNN | NNNNNNNNNNNNNNN····· | 1 |
| 16386 | MT966165.1 | \|Severe acute respiratory syndrome coro | ···················· | ···················· | ···················· | ···················· | ···················· | NNNNNNNNNNNNNNNNNNNN | 1 |
| 16398 | MT966185.1 | \|Severe acute respiratory syndrome coro | ···················· | ···················· | ···················· | ···················· | NNNNNNNNNNNNNNNNNNNN | ···················· | 1 |
| 16399 | MT966186.1 | \|Severe acute respiratory syndrome coro | ···················· | ···················· | ···················· | ···················· | NNNNNNNNNNNNNNNNNNNN | ···················· | 1 |
| 16426 | MT966229.1 | \|Severe acute respiratory syndrome coro | ···················· | ···················· | ···················· | ···················· | NNNNNNNNNNNNNNNNNNNN | NNNNNNNNNNNNNN···NNN | 1 |
| 16431 | MT966237.1 | \|Severe acute respiratory syndrome coro | ···················· | ···················· | ···················· | ···················· | ·NNNNNNNNNNNNN······ | ···················· | 1 |
| 16444 | MT966253.1 | \|Severe acute respiratory syndrome coro | ···················· | ···················· | ···················· | ···················· | NNNNNNNNNNNNNNNNNNNN | NNNNNNNNNNNN········ | 1 |
| 16557 | MT969355.1 | \|Severe acute respiratory syndrome coro | ···················· | ···················· | ···················· | ···················· | NNNNNNNNNNNNNNNNNNNN | ···················· | 1 |
| 16576 | MT969378.1 | \|Severe acute respiratory syndrome coro | ···················· | ···················· | ···················· | ···················· | NNNNNNNNNNNNNNNNNNNN | NNNN···NNNNNNNNNN··· | 1 |
| 16600 | MT969405.1 | \|Severe acute respiratory syndrome coro | ···················· | ···················· | ···················· | ···················· | NNNNNNNNNNNNNNNNNNNN | NNNNNNNNNNNNNNNNNNNN | 1 |
| 16603 | MT969410.1 | \|Severe acute respiratory syndrome coro | ···················· | ···················· | ···················· | ···················· | NNNNNNNNNNNNNNNNNNNN | ·············NNNNNNN | 1 |
| 16606 | MT969413.1 | \|Severe acute respiratory syndrome coro | ···················· | ···················· | ···················· | ···················· | NNNN················ | ···················· | 1 |
| 16609 | MT969416.1 | \|Severe acute respiratory syndrome coro | ···················· | ···················· | ···················· | ···················· | NNNNNNNNNNNNNNNNNNNN | ···················· | 1 |
| 16626 | MT969434.1 | \|Severe acute respiratory syndrome coro | ···················· | ···················· | ···················· | ···················· | ···················· | NNNN················ | 1 |
| 16635 | MT969446.1 | \|Severe acute respiratory syndrome coro | ···················· | ···················· | ···················· | ···················· | NNNNNNNNNNNNNNNNNNNN | ···················· | 1 |
| 16639 | MT969450.1 | \|Severe acute respiratory syndrome coro | ···················· | ···················· | ···················· | ···················· | NNNNNNNNNNNNNNNN···· | ···················· | 1 |
| 16642 | MT969453.1 | \|Severe acute respiratory syndrome coro | ···················· | ···················· | ···················· | ···················· | NNNNNNNNNNNNNNNNNNN· | ··K················· | 1 |
| 16643 | MT969454.1 | \|Severe acute respiratory syndrome coro | ···················· | ···················· | ···················· | ···················· | NNNNNNNNNNNNNNNNNNNN | ···················· | 1 |
| 16644 | MT969455.1 | \|Severe acute respiratory syndrome coro | ···················· | ···················· | ···················· | ···················· | ·NNN···NNNNNNNNNNNNN | ···················· | 1 |
| 16645 | MT969456.1 | \|Severe acute respiratory syndrome coro | ···················· | ···················· | ···················· | ···················· | NNNNNNNNNNNNNNNNNNNN | ···················· | 1 |
| 16650 | MT969462.1 | \|Severe acute respiratory syndrome coro | ···················· | ···················· | ···················· | ···················· | ··············NNNNNN | ···················· | 1 |
| 16666 | MT969481.1 | \|Severe acute respiratory syndrome coro | ···················· | ···················· | ···················· | ···················· | ·NNNNNNNNNNNNNNNNNNN | ···················· | 1 |
| 16681 | MT969498.1 | \|Severe acute respiratory syndrome coro | ···················· | ···················· | ···················· | ···················· | NNNN················ | ···················· | 1 |
| 16696 | MT969513.1 | \|Severe acute respiratory syndrome coro | ···················· | ···················· | ···················· | ···················· | NNNNNNNNNNNNNNNNNNNN | ···················· | 1 |
| 16701 | MT969518.1 | \|Severe acute respiratory syndrome coro | ···················· | ···················· | ···················· | ···················· | NNNNNNNNNNNNNNNNNNNN | ···················· | 1 |
| 16715 | MT969534.1 | \|Severe acute respiratory syndrome coro | ···················· | ···················· | ···················· | ···················· | ··NNNNNNNNNNNNNNNNNN | ··············NNNNNN | 1 |
| 16723 | MT969543.1 | \|Severe acute respiratory syndrome coro | ···················· | ···················· | ···················· | ···················· | ···················· | NNNN················ | 1 |
| 16729 | MT969549.1 | \|Severe acute respiratory syndrome coro | ···················· | ···················· | ···················· | ···················· | NNNNNNNNNNNNNN······ | ···················· | 1 |
| 16804 | MT969624.1 | \|Severe acute respiratory syndrome coro | ···················· | ···················· | ···················· | ···················· | ··········NNNNNNNNNN | ···················· | 1 |
| 16809 | MT969630.1 | \|Severe acute respiratory syndrome coro | ···················· | ···················· | ···················· | ···················· | NNNN················ | ···················· | 1 |
| 16810 | MT969631.1 | \|Severe acute respiratory syndrome coro | ···················· | ···················· | ···················· | ···················· | NNNNNNNNNNNNNNNNNNNN | ···················· | 1 |
| 16814 | MT969637.1 | \|Severe acute respiratory syndrome coro | ···················· | ···················· | ···················· | ···················· | NNNNNNNNNNNNNNNNNNNN | ···················· | 1 |
| 16821 | MT969645.1 | \|Severe acute respiratory syndrome coro | ···················· | ···················· | ···················· | ···················· | NNNN················ | ···················· | 1 |
| 16846 | MT969670.1 | \|Severe acute respiratory syndrome coro | ···················· | ···················· | ···················· | ···················· | NNNN················ | ···················· | 1 |
| 16854 | MT969678.1 | \|Severe acute respiratory syndrome coro | ···················· | ···················· | ···················· | ···················· | ········NNNN········ | ···················· | 1 |
| 16856 | MT969680.1 | \|Severe acute respiratory syndrome coro | ···················· | ···················· | ···················· | ···················· | ···················· | NNNN··NNNNNNNNNNNNNN | 1 |
| 16858 | MT969682.1 | \|Severe acute respiratory syndrome coro | ···················· | ···················· | ···················· | ···················· | ···················· | NNNNNNNNNNNNNNN····· | 1 |
| 16867 | MT969692.1 | \|Severe acute respiratory syndrome coro | ···················· | ···················· | ···················· | ···················· | NNNNNNNNNNNNNN······ | ···················· | 1 |
| 16882 | MT969707.1 | \|Severe acute respiratory syndrome coro | ···················· | ···················· | ···················· | ···················· | NNNNNNNNNNNNNNNNNNNN | ···················· | 1 |
| 16898 | MT969724.1 | \|Severe acute respiratory syndrome coro | ···················· | ···················· | ···················· | ···················· | ··NNNNNNNNNNNNNNNNNN | ···················· | 1 |
| 16915 | MT969742.1 | \|Severe acute respiratory syndrome coro | ···················· | ···················· | ···················· | ···················· | ·············NNNNNNN | ···················· | 1 |
| 16928 | MT969755.1 | \|Severe acute respiratory syndrome coro | ···················· | ···················· | ···················· | ···················· | NNNNNNNNNNNNNNNNNNNN | ···················· | 1 |
| 16946 | MT969773.1 | \|Severe acute respiratory syndrome coro | ···················· | ···················· | ···················· | ···················· | NNNN················ | ···················· | 1 |
| 16956 | MT969783.1 | \|Severe acute respiratory syndrome coro | ···················· | ···················· | ···················· | ···················· | ·NNNNNNNNNNNNNNNNNNN | ···················· | 1 |
| 17005 | MT969833.1 | \|Severe acute respiratory syndrome coro | ···················· | ···················· | ···················· | ···················· | ·NNNNNNNNNNN········ | ···················· | 1 |
| 17012 | MT969840.1 | \|Severe acute respiratory syndrome coro | ···················· | ···················· | ···················· | ···················· | NNNNNNNNNNNNNNNNNNNN | ···················· | 1 |
| 17029 | MT969860.1 | \|Severe acute respiratory syndrome coro | ···················· | ···················· | ···················· | ···················· | NNNNNNNNNNNNNNNNNNNN | NNNNNNNNNNNN········ | 1 |
| 17034 | MT969866.1 | \|Severe acute respiratory syndrome coro | ···················· | ···················· | ···················· | ···················· | NNNN················ | ···················· | 1 |
| 17036 | MT969869.1 | \|Severe acute respiratory syndrome coro | ···················· | ···················· | ···················· | ···················· | ···················· | NNNNNNNNNNNNNNN····· | 1 |
| 17111 | MT969946.1 | \|Severe acute respiratory syndrome coro | ···················· | ···················· | ···················· | ···················· | NNNNNNNNNNNNNNNNNNNN | ···················· | 1 |
| 17188 | MT970081.1 | \|Severe acute respiratory syndrome coro | ···················· | ···················· | ···················· | ···················· | NNNNNNNNNNNNNNNNNNNN | ···················· | 1 |
| 17189 | MT970082.1 | \|Severe acute respiratory syndrome coro | ···················· | ···················· | ···················· | ···················· | NNNN················ | ···················· | 1 |
| 17190 | MT970084.1 | \|Severe acute respiratory syndrome coro | ···················· | ···················· | ···················· | ···················· | ········NNNNNNNNNNNN | ···················· | 1 |
| 17218 | MT970173.1 | \|Severe acute respiratory syndrome coro | ···················· | ···················· | ···················· | ···················· | NNNNNNNNNNN········· | ···················· | 1 |
| 17270 | MT970225.1 | \|Severe acute respiratory syndrome coro | ···················· | ···················· | ···················· | ···················· | NNNNNNNNNNNNNNNNNNNN | ···················· | 1 |
| 17319 | MT970275.1 | \|Severe acute respiratory syndrome coro | ···················· | ···················· | ···················· | ···················· | NNNNNNNNNNNNNNNNNNNN | ···················· | 1 |
| 17330 | MT970286.1 | \|Severe acute respiratory syndrome coro | ···················· | ···················· | ···················· | ···················· | NNNN················ | ···················· | 1 |
| 17374 | MT970339.1 | \|Severe acute respiratory syndrome coro | ···················· | ···················· | ···················· | ···················· | NNNN················ | ···················· | 1 |

17380 MT970345.1 |Severe acute respiratory syndrome coro ···················· ···················· ···················· ···················· NNNNNNNNNNNNNNNNNNNN ···················· 1
18414 MT971455.1 |Severe acute respiratory syndrome coro ···················· ···················· ···················· ···················· ···················· NNNNNNNNNNNNNNNNNNNN 1
18526 MT971586.1 |Severe acute respiratory syndrome coro ···················· ···················· ···················· ···················· NNNNNNNNNNNNNNNNNNNN ···················· 1
18576 MT971651.1 |Severe acute respiratory syndrome coro ···················· ···················· ···················· ···················· NNNNNNNNNNNNNNN····· ···················· 1
18595 MT971672.1 |Severe acute respiratory syndrome coro ···················· ···················· ···················· ···················· ···················· NNNNNNNNNNNNNNNNNNNN 1
18991 MT972100.1 |Severe acute respiratory syndrome coro ···················· ···················· ···················· ···················· NNNNNNNNNNNNNNNNNNNN NNNNNNNNN··········· 1
19005 MT972125.1 |Severe acute respiratory syndrome coro ···················· ···················· ···················· ···················· NNNNNNNNNNNNNNNNNNNN NNNNNNNN············ 1
19009 MT972131.1 |Severe acute respiratory syndrome coro ···················· ···················· ···················· ···················· NNNNNNNNNNNNNNNNNNNN ···················· 1
19044 MT972175.1 |Severe acute respiratory syndrome coro ···················· ···················· ···················· ···················· ···················· NNNNNNNNNN·········· 1
19045 MT972176.1 |Severe acute respiratory syndrome coro ···················· ···················· ···················· ···················· NNNNNNNNNNNNNNNNNNNN ···················· 1
19046 MT972177.1 |Severe acute respiratory syndrome coro ···················· ···················· ···················· ···················· ···················· NNNNNNNNN··········· 1
19071 MT972206.1 |Severe acute respiratory syndrome coro ···················· ···················· ···················· ···················· ··NNNNNNNNNNNNNNNNNN NNNNNNN············· 1
19077 MT972213.1 |Severe acute respiratory syndrome coro ···················· ···················· ···················· ···················· ··NNNNNNNNNNNNNNNNNN NNNNNNNNNNNNNNN····N 1
19140 MT972283.1 |Severe acute respiratory syndrome coro ···················· ···················· ···················· ···················· NNNNN··············· ···················· 1
19193 MT972340.1 |Severe acute respiratory syndrome coro ···················· ···················· ···················· ···················· NNNNNNNNNNNNNNNNNNNN ···················· 1
19273 MT972426.1 |Severe acute respiratory syndrome coro ···················· ···················· ···················· ···················· NNN················· NNNNNNNNNNNNNNNNNNNN 1
19295 MT972450.1 |Severe acute respiratory syndrome coro ···················· ···················· ···················· ···················· NNNNNNNNNNNNNNNNNNNN NNNNNNNNK··········· 1
19326 MT972481.1 |Severe acute respiratory syndrome coro ···················· ···················· ···················· ···················· NNNNNNNNNNNNNN······ ···················· 1
19335 MT972490.1 |Severe acute respiratory syndrome coro ···················· ···················· ···················· ···················· NNNNNNNNNNNNNNNNNNNN ···················· 1
19362 MT972519.1 |Severe acute respiratory syndrome coro ···················· ···················· ···················· ···················· NNNN················ ···················· 1
19372 MT972529.1 |Severe acute respiratory syndrome coro ···················· ···················· ···················· ···················· NNNNNNNNNNNNNNNNNNNN ···················· 1
19373 MT972530.1 |Severe acute respiratory syndrome coro ···················· ···················· ···················· ···················· NNNNNNNNNNNNNNNNNNNN ···················· 1
19381 MT972539.1 |Severe acute respiratory syndrome coro ···················· ···················· ···················· ···················· NNNN················ ···················· 1
19401 MT972561.1 |Severe acute respiratory syndrome coro ···················· ···················· ···················· ···················· NNNNNNNNNNNN········ ···················· 1
19413 MT972575.1 |Severe acute respiratory syndrome coro ···················· ···················· ···················· ···················· NNNN················ ···················· 1
19429 MT972596.1 |Severe acute respiratory syndrome coro ···················· ···················· ···················· ···················· NNNN················ ···················· 1
19433 MT972605.1 |Severe acute respiratory syndrome coro ···················· ···················· ···················· ···················· ··········GT·NNNNNNN ···················· 1
19436 MT972609.1 |Severe acute respiratory syndrome coro ···················· ···················· ···················· ···················· NNNN················ ···················· 1
19442 MT972616.1 |Severe acute respiratory syndrome coro ···················· ···················· ···················· ···················· NNNNNNNNNNNNNNNNNNNN ···················· 1
19453 MT972632.1 |Severe acute respiratory syndrome coro ···················· ···················· ···················· ···················· NNNN················ ···················· 1
19466 MT972651.1 |Severe acute respiratory syndrome coro ···················· ···················· ···················· ···················· NNNN················ ···················· 1
19473 MT972658.1 |Severe acute respiratory syndrome coro ···················· ···················· ···················· ···················· NNNNNNNNNNNNNNNNNNNN ···················· 1
19475 MT972662.1 |Severe acute respiratory syndrome coro ···················· ···················· ···················· ···················· NNNNNNNNNNNNNNNNNNNN ···················· 1
19479 MT972667.1 |Severe acute respiratory syndrome coro ···················· ···················· ···················· ···················· NNNNNNNNNNNNNNNNN··· ···················· 1
19483 MT972671.1 |Severe acute respiratory syndrome coro ···················· ···················· ···················· ···················· NNNNNNNNNNNNNNNNNNNN ···················· 1
19502 MT972700.1 |Severe acute respiratory syndrome coro ···················· ···················· ···················· ···················· NNNN················ ···················· 1
19508 MT972706.1 |Severe acute respiratory syndrome coro ···················· ···················· ···················· ···················· ·············NNNNNNN ···················· 1
19516 MT972714.1 |Severe acute respiratory syndrome coro ···················· ···················· ···················· ···················· NNNN················ ···················· 1
19520 MT972718.1 |Severe acute respiratory syndrome coro ···················· ···················· ···················· ···················· NNNNNNNNNNNNNNNNN··· ···················· 1
19521 MT972719.1 |Severe acute respiratory syndrome coro ···················· ···················· ···················· ···················· NNNN················ ···················· 1
19523 MT972721.1 |Severe acute respiratory syndrome coro ···················· ···················· ···················· ···················· NNNNNNNNNNNNNNNNNNNN NNNNNNNNNNNNNNN····· 1
19524 MT972722.1 |Severe acute respiratory syndrome coro ···················· ···················· ···················· ···················· NNNN················ ···················· 1
19528 MT972728.1 |Severe acute respiratory syndrome coro ···················· ···················· ···················· ···················· NNNNNNNNNNNNNNNNNNNN ················NNNN 1
19529 MT972729.1 |Severe acute respiratory syndrome coro ···················· ···················· ···················· ···················· NNNNNNNNNNNNNNNNNNNN NNNNNNNNNNNNNNNN···· 1
19531 MT972732.1 |Severe acute respiratory syndrome coro ···················· ···················· ···················· ···················· NNNNNNNNNNNNNNNNNNNN ···················· 1
19543 MT972745.1 |Severe acute respiratory syndrome coro ···················· ···················· ···················· ···················· NNNN················ ···················· 1
19545 MT972748.1 |Severe acute respiratory syndrome coro ···················· ···················· ···················· ···················· NNNN················ ···················· 1
19548 MT972752.1 |Severe acute respiratory syndrome coro ···················· ···················· ···················· ···················· NNNN················ ···················· 1
19552 MT972758.1 |Severe acute respiratory syndrome coro ···················· ···················· ···················· ···················· NNNN················ ···················· 1
19728 MT972955.1 |Severe acute respiratory syndrome coro ···················· ···················· ···················· ···················· NNNNNNNNNNNNNNNNNNNN ···················· 1
19944 MT973207.1 |Severe acute respiratory syndrome coro ···················· ···················· ···················· ···················· NNNNNNNNNNNNNNNNNNNN ···················· 1
20027 MT973305.1 |Severe acute respiratory syndrome coro ···················· ···················· ···················· ···················· ··NNNNNNNNNNN······· ···················· 1
20033 MT973312.1 |Severe acute respiratory syndrome coro ···················· ···················· ···················· ···················· NNNNNNNNNNNNNNNNNNNN ···················· 1
20039 MT973318.1 |Severe acute respiratory syndrome coro ···················· ···················· ···················· ···················· NNNN················ ···················· 1
20043 MT973322.1 |Severe acute respiratory syndrome coro ···················· ···················· ···················· ···················· NNNN················ ···················· 1
20062 MT973344.1 |Severe acute respiratory syndrome coro ···················· ···················· ···················· ···················· NNNNNNNNNNNNNNNNNNNN ···················· 1
20065 MT973353.1 |Severe acute respiratory syndrome coro ···················· ···················· ···················· ···················· NNNNNNNNNNNNNNNN···· ···················· 1
20066 MT973356.1 |Severe acute respiratory syndrome coro ···················· ···················· ···················· ···················· NNNNNNNNNNNNNNNNNNNN ···················· 1
20067 MT973358.1 |Severe acute respiratory syndrome coro ···················· ···················· ···················· ···················· ·NNNNNNNNNNNNNNNNNNN NNNNNNNNN··········N 1
20071 MT973376.1 |Severe acute respiratory syndrome coro ···················· ···················· ···················· ···················· NNNNNNNNNNNNNNNNNNNN NNNNNNNNN··········· 1
20072 MT973378.1 |Severe acute respiratory syndrome coro ···················· ···················· ···················· ···················· NNNNNNNNNNNNNNNNNNNN NNNN················ 1
20074 MT973381.1 |Severe acute respiratory syndrome coro ···················· ···················· ···················· ···················· NNNNNNNNNNNNNNNNNNNN ···················· 1
20075 MT973389.1 |Severe acute respiratory syndrome coro ···················· ···················· ···················· ···················· NNNNNNNNNNNNNNNNNNNN ···················· 1
20076 MT973391.1 |Severe acute respiratory syndrome coro ···················· ···················· ···················· ···················· NNNNNNNNNNNNNNNNNNNN ···················· 1
20077 MT973395.1 |Severe acute respiratory syndrome coro ···················· ···················· ···················· ···················· NNNNNNNNNNNNNNNNNNNN ···········NNNNNNNNN 1
20078 MT973396.1 |Severe acute respiratory syndrome coro ···················· ···················· ···················· ···················· ···················· NNNNNNNNN··········· 1
20079 MT973402.1 |Severe acute respiratory syndrome coro ···················· ···················· ···················· ···················· NNNNNNNNNNNNNNNNNNNN NNNNNNNNN··········· 1
20081 MT973419.1 |Severe acute respiratory syndrome coro ···················· ···················· ···················· ···················· NNN····NNNNNNNNNNNNN ···················· 1
20082 MT973421.1 |Severe acute respiratory syndrome coro ···················· ···················· ···················· ···················· NNNNNNNNNNNNNNNNNNNN ·················NNN 1
20083 MT973424.1 |Severe acute respiratory syndrome coro ···················· ···················· ···················· ···················· NNNNNNNNNNNNNNNNNNNN NNNNNNNNN··········· 1
20087 MT973435.1 |Severe acute respiratory syndrome coro ···················· ···················· ···················· ···················· NNNNNNNNNNNNNNNNNNNN ···················· 1
20088 MT973436.1 |Severe acute respiratory syndrome coro ···················· ···················· ···················· ···················· NNNN················ ···················· 1
20092 MT973484.1 |Severe acute respiratory syndrome coro ···················· ···················· ···················· ···················· ···················· NNNNNNNNNNNNNNN····· 1
20146 MT956698.1 |Severe acute respiratory syndrome coro ···················· ···················· ···················· ···················· NNNNNNNNNNNNNNNNNNNN ···················· 1
20149 MT956701.1 |Severe acute respiratory syndrome coro ···················· ···················· ···················· ···················· NNNNNNNNNNNNNNNNNNNN ···················· 1
20156 MT956712.1 |Severe acute respiratory syndrome coro ···················· ···················· ···················· ···················· NNNNNNT············· ···················· 1
20176 MT956736.1 |Severe acute respiratory syndrome coro ···················· ···················· ···················· ···················· ···················· NNNNN······NNNN·NNNN 1

| | | | | | | | | | |
|---|---|---|---|---|---|---|---|---|---|
| 20197 | MT956758.1 | Severe acute respiratory syndrome coro | ···················· | ···················· | ····················· | ···················· | NNNNNNNNNNNNNNNNNNNN | ············NNNNNNNN | 1 |
| 20199 | MT956760.1 | Severe acute respiratory syndrome coro | ···················· | ···················· | ····················· | ···················· | NNNNNN·············· | ···················· | 1 |
| 20206 | MT956772.1 | Severe acute respiratory syndrome coro | ···················· | ···················· | ····················· | ···················· | NNNNNNNNNNNNNNNNNNNN | NNNNNN···········NNN | 1 |
| 20208 | MT956775.1 | Severe acute respiratory syndrome coro | ···················· | ···················· | ····················· | ···················· | NNNNNNNNNNNNN······· | NNNNNNNNNNN········· | 1 |
| 20211 | MT956783.1 | Severe acute respiratory syndrome coro | ···················· | ···················· | ····················· | ···················· | NNNNNNNNNNNNNNNN···· | ···················· | 1 |
| 20234 | MT956814.1 | Severe acute respiratory syndrome coro | ···················· | ···················· | ····················· | ···················· | ········NNNNNNNNNNNN | ···················· | 1 |
| 20295 | MT958257.1 | Severe acute respiratory syndrome coro | ···················· | ·A·················· | ····················· | ···················· | ··············NNNNNN | ···················· | 1 |
| 20807 | MT929128.1 | Severe acute respiratory syndrome coro | ···················· | ···················· | ····················· | ···················· | NNNNNNNNNNNNNNNNNNNN | ···················· | 1 |
| 20916 | LR877965.1 | Severe acute respiratory syndrome coro | ···················· | ···················· | ····················· | ···················· | NNNNNNNNNNNNNNNN···· | ···················· | 1 |
| 20918 | LR877967.1 | Severe acute respiratory syndrome coro | ···················· | ···················· | ····················· | ···················· | NNNNNNNNN··········· | ···················· | 1 |
| 20931 | LR877980.1 | Severe acute respiratory syndrome coro | ···················· | ···················· | ····················· | ···················· | NNNNNNNNNNNNNNNNNNNN | ···················· | 1 |
| 20932 | LR877981.1 | Severe acute respiratory syndrome coro | ···················· | ···················· | ····················· | ···················· | NNNNNNNNNNNN········ | ···················· | 1 |
| 20934 | LR877983.1 | Severe acute respiratory syndrome coro | ···················· | ···················· | ····················· | ···················· | NNNNNNNNNNNNNNNNNNNN | NNNNNNNNNNNNNNNNNNNN | 1 |
| 20949 | LR877998.1 | Severe acute respiratory syndrome coro | ···················· | ···················· | ····················· | ···················· | NNNNNN·············· | ···················· | 1 |
| 21009 | LR878058.1 | Severe acute respiratory syndrome coro | ···················· | ···················· | ····················· | ···················· | NNNNNNNNN··········· | ···················· | 1 |
| 21010 | LR878059.1 | Severe acute respiratory syndrome coro | ···················· | ···················· | ····················· | ···················· | NNNNNNNNNNNNNN······ | ···················· | 1 |
| 21020 | LR878069.1 | Severe acute respiratory syndrome coro | ···················· | ···················· | ····················· | ···················· | NNNNNNNNNNNNNNNN···· | ···················· | 1 |
| 21080 | LR878129.1 | Severe acute respiratory syndrome coro | ···················· | ···················· | ····················· | ···················· | NNNNNNNN············ | ···················· | 1 |
| 21082 | LR878131.1 | Severe acute respiratory syndrome coro | NNNNNNNNNNNNNN······ | ···················· | ····················· | ···················· | ···················· | ···················· | 1 |
| 21105 | LR878154.1 | Severe acute respiratory syndrome coro | ···················· | ···················· | ····················· | ···················· | NNNNNN·············· | ···················· | 1 |
| 21179 | LR878228.1 | Severe acute respiratory syndrome coro | NNNNNNNNNNNNNNNNNNN· | ···················· | NNNNNNNNNNNNNNNNNNNNN | NNNNNNNNNNNNNNNNNNNN | NNNNNNNNNNNNNNNNNNNN | NNNNNNNNNNNNNNNNNNNN | 1 |
| 21182 | LR878231.1 | Severe acute respiratory syndrome coro | ···················· | ···················· | ····················· | ···················· | ···················· | NNNNNNNNNNNNNNNNNNNN | 1 |
| 21183 | LR878232.1 | Severe acute respiratory syndrome coro | NNNNNNNNNNNNNNN····· | ···················· | ····················· | ···················· | ···················· | ···················· | 1 |
| 21184 | LR878233.1 | Severe acute respiratory syndrome coro | ···················· | ···················· | ····················· | ···················· | ···················· | NNNNNNNNNNNNNNNNNNNN | 1 |
| 21186 | LR878235.1 | Severe acute respiratory syndrome coro | ···················· | ···················· | ····················· | ···················· | ···················· | NNNNNNNNNNNNNNNNNN·· | 1 |
| 21188 | LR878237.1 | Severe acute respiratory syndrome coro | ···················· | ···················· | ····················· | ···················· | ···················· | NNNNNNNNNNNNNNNNNNNN | 1 |
| 21190 | LR878239.1 | Severe acute respiratory syndrome coro | ···················· | ···················· | ····················· | ···················· | ···················· | NNNNNNNNNNNNNNNNNNNN | 1 |
| 21191 | LR878240.1 | Severe acute respiratory syndrome coro | NNNNNNNNNN·········· | ···················· | NNNNNNNNNNNNNNNNNNNNN | NNNNNNNNNNNNNNNNNNNN | ···················· | ···················· | 1 |
| 21193 | LR878242.1 | Severe acute respiratory syndrome coro | ···················· | ···················· | ····················· | ···················· | ···················· | NNNNNNNNNNNNNNNNNNNN | 1 |
| 21197 | LR878246.1 | Severe acute respiratory syndrome coro | ···················· | ···················· | ····················· | ···················· | ···················· | NNNNNNNNNNNNNNNNNNNN | 1 |
| 21208 | LR878257.1 | Severe acute respiratory syndrome coro | ···················· | ···················· | NNNNNNNNNNNNNNNNNNNNN | NNNNNNNNNNNNNNNNNNNN | NNNNNNNNNNNNNNNNNNNN | NNNNNNNNNNNNNNNNNNNN | 1 |
| 21250 | LR878299.1 | Severe acute respiratory syndrome coro | ···················· | ···················· | ····················· | ···················· | ···················· | NNNNNNNNNNNNNNNNNNNN | 1 |
| 21468 | MT913111.1 | Severe acute respiratory syndrome coro | ···················· | ···················A | ····················· | ···················· | ··NNNNNNNNNNNNNNNNNN | ···················· | 1 |
| 21722 | MT893662.1 | Severe acute respiratory syndrome coro | ···················· | ···················· | ····················· | ···················· | NNNNNNNNNNNNNNNNNNNN | ···················· | 1 |
| 21728 | MT893671.1 | Severe acute respiratory syndrome coro | ···················· | ···················· | ····················· | ···················· | NNNN················ | ···················· | 1 |
| 21754 | MT890211.1 | Severe acute respiratory syndrome coro | ···················· | ···················· | ····················· | ···················· | NNNNNNNNNNNNNNNNNNNN | NNNNNNNNNNNNNNNN···· | 1 |
| 21756 | MT890235.1 | Severe acute respiratory syndrome coro | ···················· | ···················· | ····················· | ···················· | NNNNNNN············· | NNNNNNN·····NNN····· | 1 |
| 21757 | MT890236.1 | Severe acute respiratory syndrome coro | ···················· | ···················· | ····················· | ···················· | NNNNNNNNNNNNNNNNNNNN | NNNNNNNNNNNNNNN····· | 1 |
| 21758 | MT890238.1 | Severe acute respiratory syndrome coro | ···················· | ···················· | ····················· | ···················· | NNNNNNNNNNNNNNNNNNNN | ···················· | 1 |
| 21759 | MT890240.1 | Severe acute respiratory syndrome coro | ···················· | ···················· | ····················· | ···················· | NNNNNNN············· | ···················· | 1 |
| 21760 | MT890243.1 | Severe acute respiratory syndrome coro | ···················· | ···················· | ····················· | ···················· | NNNNNNNNNNNNNNNNNNNN | ···················· | 1 |
| 21762 | MT890279.1 | Severe acute respiratory syndrome coro | ···················· | ···················· | ····················· | ···················· | NNNNNNNNNNNNNNNNNNNN | ···················· | 1 |
| 21764 | MT890281.1 | Severe acute respiratory syndrome coro | ···················· | ···················· | ····················· | ···················· | NNNNNNNNNNNNNNNNNNNN | ···················· | 1 |
| 21931 | MT886351.1 | Severe acute respiratory syndrome coro | ···················· | ···················· | ····················· | ···················· | ···················· | NNNNNNNN············ | 1 |
| 22005 | LR877184.1 | Severe acute respiratory syndrome coro | -------------------- | -------------------- | --------------------- | -------------------- | -------------------- | -------------------- | 1 |
| 22144 | MT873205.1 | Severe acute respiratory syndrome coro | NNNNNNNNNNNNNNNNNNNN | ···················· | ····················· | ···················· | ··················NN | ···················· | 1 |
| 22534 | MT846433.1 | Severe acute respiratory syndrome coro | ···················· | ···················· | ····················· | ···················· | NNNNNNNNNNNNNNNNNNNN | ···················· | 1 |
| 22547 | MT846450.1 | Severe acute respiratory syndrome coro | ···················· | ···················· | ····················· | ···················· | ·········NNN······NN | NNNNNNNNNNN········· | 1 |
| 22550 | MT846454.1 | Severe acute respiratory syndrome coro | ···················· | ···················· | ····················· | ···················· | NNNNNNNNNNNNNN·····N | ···················· | 1 |
| 23110 | MT831393.1 | Severe acute respiratory syndrome coro | ···················· | ···················· | ····················· | ···················· | ···················· | NNNNNNNNNNNNNNNNNNNN | 1 |
| 23916 | MT834616.1 | Severe acute respiratory syndrome coro | ···················· | ···················· | ····················· | ···················· | ········NNNNNNNNNNN· | ···················· | 1 |
| 23955 | MT834668.1 | Severe acute respiratory syndrome coro | ···················· | ···················· | ····················· | ···················· | NNNNNNNNNNNNNNNNNNNN | ···················· | 1 |
| 23960 | MT834675.1 | Severe acute respiratory syndrome coro | ···················· | ···················· | ····················· | ···················· | NNNNNNNNNNNN········ | ···················· | 1 |
| 23983 | MT834730.1 | Severe acute respiratory syndrome coro | ···················· | ···················· | ····················· | ···················· | NNNNNNNNNNNN········ | ···················· | 1 |
| 24582 | MT810706.1 | Severe acute respiratory syndrome coro | ···················· | ·A·················· | ····················· | ···················· | ···········NNN···NNN | ···················· | 1 |
| 24588 | MT810713.1 | Severe acute respiratory syndrome coro | ···················· | ·A·················· | ····················· | ···················· | NNNNNNNNNNNNNNNNNNNN | ···················· | 1 |
| 25135 | MT811639.1 | Severe acute respiratory syndrome coro | ···················· | ···················· | ····················· | ···················· | NNNNNNN············· | ···················· | 1 |
| 25510 | MT791885.1 | Severe acute respiratory syndrome coro | ···················· | ···················· | ····················· | ···················· | NNNNNNNNNNNNNNNNNNNN | ···················· | 1 |
| 25940 | MT757006.1 | Severe acute respiratory syndrome coro | ···················· | ···················· | ····················· | ···················· | NNNNNNNNN··········· | ···················· | 1 |
| 25965 | MT757051.1 | Severe acute respiratory syndrome coro | ···················· | ···················· | ····················· | ···················· | NNNNNNNNNNNNNNNNNNNN | ···················· | 1 |
| 25989 | MT757087.1 | Severe acute respiratory syndrome coro | ···················· | ···················· | ····················· | ···················· | NNNNNNNNNNNNNNNNNNNN | ···················· | 1 |
| 25990 | MT757094.1 | Severe acute respiratory syndrome coro | ···················· | ···················· | ····················· | ···················· | NNNNNNNNNNNN········ | ···················· | 1 |
| 26011 | MT745630.1 | Severe acute respiratory syndrome coro | ···················· | ···················· | ····················· | ···················· | NNNN················ | ···················· | 1 |
| 26049 | MT745679.1 | Severe acute respiratory syndrome coro | ···················· | ···················· | ····················· | ···················· | NNNNNNNNNNNNNNNNNNNN | ···················· | 1 |
| 26070 | MT745700.1 | Severe acute respiratory syndrome coro | ···················· | ···················· | ····················· | ···················· | NNNN················ | ···················· | 1 |
| 26092 | MT745722.1 | Severe acute respiratory syndrome coro | ···················· | ···················· | ····················· | ···················· | NNNNNNNNNNNNNNNNNNNN | ···················· | 1 |
| 26104 | MT745734.1 | Severe acute respiratory syndrome coro | ···················· | ···················· | ····················· | ···················· | NNNNNNNNNNNNNNNNNNNN | ···················· | 1 |
| 26115 | MT745745.1 | Severe acute respiratory syndrome coro | ···················· | ···················· | ····················· | ···················· | NNNN················ | ···················· | 1 |
| 26936 | MT703969.1 | Severe acute respiratory syndrome coro | ···················· | ···················· | ····················· | ···················· | NNNNNNNNNNNNNNNNNNNN | ···················· | 1 |
| 26939 | MT703976.1 | Severe acute respiratory syndrome coro | ···················· | ···················· | ····················· | ···················· | NNNNNNNNNNNNNNNNNNNN | ···················· | 1 |
| 27538 | MT641656.1 | Severe acute respiratory syndrome coro | ···················· | ···················· | ····················· | ···················· | NNNNNNN············· | ···················· | 1 |
| 27540 | MT641662.1 | Severe acute respiratory syndrome coro | ···················· | ···················· | ····················· | ···················· | ···················· | NNNNNNNNN··········· | 1 |
| 27551 | MT641679.1 | Severe acute respiratory syndrome coro | ···················· | ···················· | ····················· | ···················· | NNNNNNNNNNNNNNNNNNNN | ···················· | 1 |
| 27558 | MT641697.1 | Severe acute respiratory syndrome coro | ···················· | ···················· | ····················· | ···················· | NNNNNNNNNNNNNNNNNNNN | NNNN················ | 1 |
| 27564 | MT641722.1 | Severe acute respiratory syndrome coro | ···················· | ···················· | ····················· | ···················· | ···················· | ········NNNNNNNNNNNN | 1 |
| 27565 | MT641723.1 | Severe acute respiratory syndrome coro | ···················· | ···················· | ····················· | ···················· | NNNNNNNNNNNNNNNNNNNN | ···················· | 1 |
| 27566 | MT641724.1 | Severe acute respiratory syndrome coro | ···················· | ···················· | ····················· | ···················· | NNNNNNNNNNNNNNNN···· | ···················· | 1 |

| | | | | | | | | | |
|---|---|---|---|---|---|---|---|---|---|
| 27596 | MT641763.1 | \|Severe acute respiratory syndrome coro | ···················· | ···················· | ···················· | ···················· | NNNNNNNNNNNN········ | NNNNNNNNNNNNNNNNN··· | 1 |
| 27600 | MT641768.1 | \|Severe acute respiratory syndrome coro | ···················· | ···················· | ···················· | ···················· | NNNNNNNNNNNNNNNNNNNN | ···················· | 1 |
| 27603 | MT641777.1 | \|Severe acute respiratory syndrome coro | ···················· | ···················· | ···················· | ···················· | NNNNNNNNNNNNNNNNNNNN | NNNNNNNNNNNNNNNNNNNN | 1 |
| 28161 | MT627925.1 | \|Severe acute respiratory syndrome coro | ···················· | ·NNNNNNNNNNNNNNNNNNN | ···················· | ···················· | ···················· | ···················· | 1 |
| 28502 | MT632564.1 | \|Severe acute respiratory syndrome coro | ···················· | ···················· | ···················· | ···················· | NNNNNNNNNNNNNNNNNNNN | ···················· | 1 |
| 29042 | MT612109.1 | \|Severe acute respiratory syndrome coro | ···················· | ···················· | ···················· | ···················· | NN·················· | NNNN···NNNNNNNN····· | 1 |
| 29043 | MT612115.1 | \|Severe acute respiratory syndrome coro | ···················· | ···················· | ···················· | ···················· | NNNNNNNNNNNNNNNNNNNN | NNNNNNNNNNNNNNNNNNNN | 1 |
| 29044 | MT612117.1 | \|Severe acute respiratory syndrome coro | ···················· | ···················· | ···················· | ···················· | NNNNNNN············· | NNNN·M·············· | 1 |
| 29045 | MT612131.1 | \|Severe acute respiratory syndrome coro | ···················· | ···················· | ···················· | ···················· | ·NNNNNNNNNNNNNNNNNNN | ···················· | 1 |
| 29046 | MT612134.1 | \|Severe acute respiratory syndrome coro | ···················· | ···················· | ···················· | ···················· | NNNNNNNNNNNNNNNNNNNN | ···················· | 1 |
| 29048 | MT612141.1 | \|Severe acute respiratory syndrome coro | ···················· | ···················· | ···················· | ···················· | NNNNNNNNNNNNNNNNNNNN | ···················· | 1 |
| 29049 | MT612144.1 | \|Severe acute respiratory syndrome coro | ···················· | ···················· | ···················· | ···················· | NNNNNNNNNNNNNNNNNNNN | NNNN················ | 1 |
| 29050 | MT612145.1 | \|Severe acute respiratory syndrome coro | ···················· | ···················· | ···················· | ···················· | NNNNNNNNNNNNNNNNNNNN | ···················· | 1 |
| 29051 | MT612147.1 | \|Severe acute respiratory syndrome coro | ···················· | ···················· | ···················· | ·······S············ | NNNNNNNNNNNNNNNNNNNN | ·········R·········· | 1 |
| 29052 | MT612150.1 | \|Severe acute respiratory syndrome coro | ···················· | ···················· | ···················· | ···················· | NNNNNNNNNNNN········ | ···················· | 1 |
| 29055 | MT612155.1 | \|Severe acute respiratory syndrome coro | ···················· | ···················· | ···················· | ···················· | NNNNNNNNNNNNNNNNNNNN | ···················· | 1 |
| 29056 | MT612162.1 | \|Severe acute respiratory syndrome coro | ···················· | ···················· | ···················· | ···················· | NNNNNNNNNNNNNNNNNNNN | NNNNNNNNNNN········· | 1 |
| 29057 | MT612167.1 | \|Severe acute respiratory syndrome coro | ···················· | ···················· | ···················· | ···················· | NNNNNNNNNNNNNNNNNNNN | NNNNNNNNNNNNNNN····· | 1 |
| 29059 | MT612173.1 | \|Severe acute respiratory syndrome coro | ···················· | ···················· | ···················· | ···················· | NNNNNNNNNNNNNNNNNNNN | ·····M·············· | 1 |
| 29060 | MT612183.1 | \|Severe acute respiratory syndrome coro | ···················· | ···················· | ···················· | ···················· | NNNNNNNNNNNNNNNNNNNN | ···················· | 1 |
| 29061 | MT612198.1 | \|Severe acute respiratory syndrome coro | ···················· | ···················· | ···················· | ···················· | NNNNNNNNNNNNNNNNNNNN | NNNN················ | 1 |
| 29062 | MT612246.1 | \|Severe acute respiratory syndrome coro | ···················· | ···················· | ···················· | ···················· | NNNNNNNNNNNNNNNNNNNN | ·K·················· | 1 |
| 29063 | MT612251.1 | \|Severe acute respiratory syndrome coro | ···················· | ···················· | ···················· | ···················· | NNNNNNNNNNNNNNNN···· | NNNNNNNNN··········· | 1 |
| 29065 | MT612281.1 | \|Severe acute respiratory syndrome coro | ···················· | ···················· | ···················· | ···················· | NNNNNNNNNNNNNNNNNNNN | ···················· | 1 |
| 29501 | MT520423.1 | \|Severe acute respiratory syndrome coro | ···················· | ···················· | ···················· | ···················· | ·············NNNNNNN | ···················· | 1 |
| 30883 | MT451100.1 | \|Severe acute respiratory syndrome coro | ···················· | ···················· | ···················· | ···················· | NNNNNNNNNNNN········ | ···················· | 1 |
| 31034 | MT451324.1 | \|Severe acute respiratory syndrome coro | ···················· | ···················· | ···················· | ···················· | NNNNNNNNNNNNNNNNNNNN | ···················· | 1 |
| 31037 | MT451356.1 | \|Severe acute respiratory syndrome coro | ···················· | ···················· | ···················· | ···················· | NNNNNNN············· | NNNN················ | 1 |
| 31038 | MT451358.1 | \|Severe acute respiratory syndrome coro | ···················· | ···················· | ··K················· | ···················· | NNNNNNN············· | ···················· | 1 |
| 31039 | MT451366.1 | \|Severe acute respiratory syndrome coro | ···················· | ···················· | ···················· | ···················· | NNNNNNNNNNNNNNNNNNNN | NNNN··········NNNNNN | 1 |
| 31042 | MT451376.1 | \|Severe acute respiratory syndrome coro | ···················· | ···················· | ···················· | ···················· | NNNNNNNNNNNNNNNNNNNN | ···················· | 1 |
| 31043 | MT451386.1 | \|Severe acute respiratory syndrome coro | ···················· | ···················· | ··K················· | ···················· | NNNNNNN···········NN | NNNNNNNNN··········· | 1 |
| 31044 | MT451389.1 | \|Severe acute respiratory syndrome coro | ···················· | ···················· | ···················· | ···················· | NNNNNNN···W········· | NNNNNNNNN··········· | 1 |
| 31045 | MT451391.1 | \|Severe acute respiratory syndrome coro | ···················· | ···················· | ···················· | ···················· | NNNNNNNNNNNNNNNNNNNN | NNNNNNNNNNNNNNNN···· | 1 |
| 31048 | MT451411.1 | \|Severe acute respiratory syndrome coro | ···················· | ···················· | ···················· | ···················· | NNNNNNN············· | ···················· | 1 |
| 31049 | MT451412.1 | \|Severe acute respiratory syndrome coro | ···················· | ···················· | ···················· | ···················· | NNNNNNNNNNNNNNNNNNNN | NNNNNNN········NNNNN | 1 |
| 31050 | MT451415.1 | \|Severe acute respiratory syndrome coro | ···················· | ···················· | ···················· | ···················· | NNNNNNNNNNNNNNNNNNNN | ··K················· | 1 |
| 31051 | MT451434.1 | \|Severe acute respiratory syndrome coro | ···················· | ···················· | ···················· | ···················· | NNNNNNNNNNNNNNNNNNNN | ···················· | 1 |
| 31053 | MT451439.1 | \|Severe acute respiratory syndrome coro | ···················· | ···················· | ···················· | ···················· | NNNNNNNNNNNNNNNNNNNN | ···················· | 1 |
| 31059 | MT451452.1 | \|Severe acute respiratory syndrome coro | ···················· | ···················· | ···················· | ···················· | NNNNNNNNNNNNNNNNNNNN | ···················· | 1 |
| 31062 | MT451466.1 | \|Severe acute respiratory syndrome coro | ···················· | ···················· | ···················· | ···················· | NNNNNNN············· | ···················· | 1 |
| 31064 | MT451476.1 | \|Severe acute respiratory syndrome coro | ···················· | ···················· | ···················· | ···················· | NNNNNNNNNNNNNNNNNNNN | NNNN················ | 1 |
| 31065 | MT451478.1 | \|Severe acute respiratory syndrome coro | ···················· | ···················· | ···················· | ···················· | NNNNNNNNNNNNNNNNNNNN | NNNN················ | 1 |
| 31066 | MT451488.1 | \|Severe acute respiratory syndrome coro | ···················· | ···················· | ···················· | ···················· | ···················· | NNNN················ | 1 |
| 31067 | MT451496.1 | \|Severe acute respiratory syndrome coro | ···················· | ···················· | ···················· | ···················· | NNNNNNNNNNNNNNNNNNNN | NNNN················ | 1 |
| 31068 | MT451549.1 | \|Severe acute respiratory syndrome coro | ···················· | ···················· | ···················· | ···················· | NNNNNNN············· | N··················· | 1 |
| 31070 | MT451578.1 | \|Severe acute respiratory syndrome coro | ···················· | ···················· | ···················· | ···················· | NNNNNNNNNNNNNNNNNNNN | ···················· | 1 |
| 31074 | MT451584.1 | \|Severe acute respiratory syndrome coro | ···················· | ···················· | ···················· | ···················· | NNNNNNN············· | ···················· | 1 |
| 31079 | MT451599.1 | \|Severe acute respiratory syndrome coro | ···················· | ···················· | ···················· | ···················· | NNNNNNN············· | NNNNNN·············· | 1 |
| 31080 | MT451601.1 | \|Severe acute respiratory syndrome coro | ···················· | ···················· | ···················· | ···················· | NNNNNNNNNNNNNNNNNNNN | NNNNNNNNNNNNNNN····· | 1 |
| 31083 | MT451604.1 | \|Severe acute respiratory syndrome coro | ···················· | ···················· | ···················· | ···················· | NNNNNNNNNNNNNNNNNNNN | ···················· | 1 |
| 31086 | MT451609.1 | \|Severe acute respiratory syndrome coro | ···················· | ···················· | ···················· | ···················· | NNNNNNNNNNN········· | ···················· | 1 |
| 31088 | MT451614.1 | \|Severe acute respiratory syndrome coro | ···················· | ···················· | ···················· | ···················· | NNNNNNNNNNNNNNNNNNNN | ·················M·· | 1 |
| 31089 | MT451616.1 | \|Severe acute respiratory syndrome coro | ···················· | ···················· | ···················· | ···················· | ·NNNNNN············· | ···················· | 1 |
| 31092 | MT451651.1 | \|Severe acute respiratory syndrome coro | ···················· | ···················· | ···················· | ···················· | NNNNNNNNNNNNNNNNNNNN | ···················· | 1 |
| 31093 | MT451653.1 | \|Severe acute respiratory syndrome coro | ···················· | ···················· | ···················· | ···················· | NNNNNNNNNNNNNNNNNNNN | NNNNNNNNN···NNNNNNNN | 1 |
| 31095 | MT451657.1 | \|Severe acute respiratory syndrome coro | ···················· | ···················· | ···················· | ···················· | NNNNNNN············· | ···················· | 1 |
| 31101 | MT451680.1 | \|Severe acute respiratory syndrome coro | ···················· | ···················· | ···················· | ···················· | NNNNNNN············· | ···················· | 1 |
| 31102 | MT451684.1 | \|Severe acute respiratory syndrome coro | ···················· | ···················· | ···················· | ···················· | NNNNNNN············· | NNN················· | 1 |
| 31103 | MT451692.1 | \|Severe acute respiratory syndrome coro | ···················· | ···················· | ···················· | ···················· | NNNNNNN············· | ···················· | 1 |
| 31104 | MT451694.1 | \|Severe acute respiratory syndrome coro | ···················· | ···················· | ···················· | ···················· | NNNNNNNNNNNNNNNNNNNN | NNNNNNNNNNNNNNN····· | 1 |
| 31105 | MT451696.1 | \|Severe acute respiratory syndrome coro | ···················· | ···················· | ···················· | ···················· | NNNNNNNNNNNNNNNNNNNN | ···················· | 1 |
| 31106 | MT451704.1 | \|Severe acute respiratory syndrome coro | ···················· | ···················· | ···················· | ···················· | NNNNNNNNNNNNNNNNNNNN | NNNNNNNNN··········· | 1 |
| 31108 | MT451710.1 | \|Severe acute respiratory syndrome coro | ···················· | ···················· | ···················· | ···················· | NNNNNNNNNNNNNNNNNNNN | ···················· | 1 |
| 31113 | MT451718.1 | \|Severe acute respiratory syndrome coro | ···················· | ···················· | ···················· | ···················· | NNNNNNNNNNNNNNNNNNNN | NNNNNNNNNNNNN······· | 1 |
| 31114 | MT451723.1 | \|Severe acute respiratory syndrome coro | ···················· | ···················· | ···················· | ···················· | NNNNNNN············· | ··············N····· | 1 |
| 31117 | MT451730.1 | \|Severe acute respiratory syndrome coro | ···················· | ···················· | ···················· | ···················· | NNNNNNN············· | NNNNNNNNN··········· | 1 |
| 31119 | MT451734.1 | \|Severe acute respiratory syndrome coro | ···················· | ···················· | ···················· | ···················· | NNNNNNNNNNNNNNNNNNNN | NNNN················ | 1 |
| 31120 | MT451736.1 | \|Severe acute respiratory syndrome coro | ···················· | ···················· | ···················· | ···················· | NNNNNNN············· | ···················· | 1 |
| 31121 | MT451737.1 | \|Severe acute respiratory syndrome coro | ···················· | ···················· | ···················· | ···················· | NNNNNNNNNNNNNNNNNNNN | ···················· | 1 |
| 31122 | MT451740.1 | \|Severe acute respiratory syndrome coro | ···················· | ···················· | ···················· | ···················· | NNNNNNN············· | ···················· | 1 |
| 31123 | MT451742.1 | \|Severe acute respiratory syndrome coro | ···················· | ···················· | ···················· | ···················· | NNNNNNN············· | ···················· | 1 |
| 31124 | MT451748.1 | \|Severe acute respiratory syndrome coro | ···················· | ···················· | ···················· | ···················· | NNNNNNNNNNNNNNNNNNNN | NNN················· | 1 |
| 31125 | MT451749.1 | \|Severe acute respiratory syndrome coro | ···················· | ···················· | ···················· | ···················· | NNNNNNNNNNNNNNNNNNNN | ······M············· | 1 |
| 31128 | MT451755.1 | \|Severe acute respiratory syndrome coro | ···················· | ···················· | ···················· | ···················· | NNNNNNNNNNNNNNNNNNNN | ···················· | 1 |
| 31130 | MT451771.1 | \|Severe acute respiratory syndrome coro | ···················· | ···················· | ···················· | ···················· | ···················· | NNNN················ | 1 |
| 31132 | MT451776.1 | \|Severe acute respiratory syndrome coro | ···················· | ···················· | ···················· | ···················· | ···················· | NNNN················ | 1 |

| | | | | | | | | |
|---|---|---|---|---|---|---|---|---|
| 31134 | MT451779.1 \|Severe acute respiratory syndrome coro | ···················· | ···················· | ···················· | ···················· | NNNNNNN············· | ···················· | 1 |
| 31135 | MT451783.1 \|Severe acute respiratory syndrome coro | ···················· | ···················· | ···················· | ···················· | NNNNNNNNN··········· | NNNN················ | 1 |
| 31747 | MT375436.1 \|Severe acute respiratory syndrome coro | ···················· | ···················· | ···················· | ···················· | NNNNNNNNNNNNNNNNNNNN | ···················· | 1 |
| 31758 | MT375448.1 \|Severe acute respiratory syndrome coro | ···················· | ···················· | ···················· | ···················· | NNNNNNNNNNNNNNNNNNNN | ···················· | 1 |
| 31778 | MT375469.1 \|Severe acute respiratory syndrome coro | ···················· | ···················· | ···················· | ···················· | ···················· | ·NNNNNNNNNNNNNNNNNNN | 1 |
| 32003 | MT358673.1 \|Severe acute respiratory syndrome coro | ···················· | ···················A | ···················· | ···················· | N·············NNN··· | ···················· | 1 |
| 32180 | MT345850.1 \|Severe acute respiratory syndrome coro | ···················· | ···················· | ···················· | ···················· | ········NNNNNNNNNN·· | ···················· | 1 |
