## Supplementary material for "Development and Implementation of a scalable and versatile test for COVID-19 diagnostics in rural communities": Sup_Table_XI

Pool raw data

| A | C | D | E | F | G | H | I | J | K | L |
|---|---|---|---|---|---|---|---|---|---|---|
| | Sample 1 | | | | | | | | | |
| Pool name | Original test date | Plate number | Sample | $Ct_{RPP30}$ | $Ct_N$ | $Ct_E$ | $Ct_S$ | Result | cut-off N | cut-off RPP30 |
| 1N | | | | 29.19 | N/A | N/A | 44.51 | Negative | 35.59 | 34.67 |
| 2N | | | | 29.20 | N/A | N/A | N/A | Negative | 35.59 | 34.67 |
| 3N | | | | 29.73 | N/A | N/A | N/A | Negative | 35.59 | 34.67 |
| 4N | | | | 29.80 | N/A | N/A | N/A | Negative | 35.59 | 34.67 |
| 5N | | | | 29.73 | N/A | N/A | N/A | Negative | 35.59 | 34.67 |
| 6N | | | | 29.98 | N/A | N/A | N/A | Negative | 35.59 | 34.67 |
| 7N | | | | 29.13 | N/A | N/A | N/A | Negative | 35.59 | 34.67 |
| 8N | | | | 28.92 | N/A | N/A | 39.89 | Negative | 35.55 | 35.75 |
| 9N | | | | 31.06 | N/A | N/A | N/A | Negative | 35.59 | 34.67 |
| 10N | | | | 29.97 | N/A | N/A | N/A | Negative | 35.59 | 34.67 |
| 11N | | | | 29.37 | N/A | N/A | 43.98 | Negative | 35.60 | 35.06 |
| 12N | | | | 29.96 | N/A | N/A | N/A | Negative | 35.60 | 35.06 |
| 13N | | | | 31.63 | N/A | N/A | N/A | Negative | 35.60 | 35.06 |
| 14N | | | | 29.81 | N/A | N/A | N/A | Negative | 35.60 | 35.06 |
| 15N | | | | 28.87 | N/A | N/A | N/A | Negative | 35.60 | 35.06 |
| 16N | | | | 29.23 | N/A | N/A | N/A | Negative | 35.60 | 35.06 |
| 17N | | | | 29.97 | N/A | N/A | N/A | Negative | 35.60 | 35.06 |
| 18N | | | | 30.02 | N/A | N/A | N/A | Negative | 35.60 | 35.06 |
| 19N | | | | 29.96 | N/A | N/A | N/A | Negative | 35.60 | 35.06 |
| 20N | | | | 29.95 | N/A | N/A | N/A | Negative | 35.60 | 35.06 |
| 1P | | | | 29.62 | 13.39 | 14.94 | 15.31 | Positive | 35.87 | 36.02 |
| 2P | | | | 30.67 | 17.24 | 17.99 | 17.77 | Positive | 35.87 | 36.02 |
| 3P | | | | 30.61 | 18.55 | 20.04 | 20.17 | Positive | 35.87 | 36.02 |
| 4P | | | | 31.02 | 22.28 | 22.27 | 22.07 | Positive | 36.09 | 35.73 |
| 5P | | | | 31.22 | 17.15 | 18.22 | 18.54 | Positive | 36.09 | 35.73 |
| 6P | | | | 29.15 | 17.18 | 18.54 | 18.88 | Positive | 36.09 | 35.73 |
| 7P | | | | 29.99 | 17.05 | 19.17 | 19.53 | Positive | 35.97 | 36.32 |
| 8P | | | | 32.00 | 22.00 | 23.00 | 23.30 | Positive | 35.47 | 36.52 |
| 9P | | | | 29.13 | 16.78 | 18.44 | 19.05 | Positive | 36.14 | 36.22 |
| 10P | | | | 29.02 | 24.75 | 25.29 | 25.51 | Positive | 35.53 | 35.84 |
| 11P | | | | 30.89 | 15.28 | 17.27 | 17.78 | Positive | 35.88 | 34.58 |
| 12P | | | | 30.88 | 17.33 | 18.76 | 19.30 | Positive | 35.40 | 35.38 |
| 13P | | | | 27.15 | 22.54 | 22.26 | 21.56 | Positive | 35.40 | 35.38 |
| 14P | | | | 31.85 | 15.96 | 17.44 | 17.86 | Positive | 35.40 | 35.38 |
| 15P | | | | 30.25 | 15.27 | 16.31 | 16.54 | Positive | 35.40 | 35.38 |
| 16P | | | | 31.57 | 20.15 | 21.46 | 22.22 | Positive | 35.40 | 35.38 |
| 17P | | | | 29.94 | 20.48 | 21.76 | 22.11 | Positive | 35.40 | 35.38 |
| 18P | | | | 30.03 | 17.27 | 18.81 | 19.38 | Positive | 35.40 | 35.38 |
| 19P | | | | 29.23 | 16.83 | 17.94 | 17.89 | Positive | 35.40 | 35.38 |
| 20P | | | | 30.28 | 19.54 | 20.68 | 20.97 | Positive | 35.47 | 35.39 |
| 21P | | | | 31.28 | 16.16 | 17.94 | 18.29 | Positive | 35.47 | 35.39 |
| 22P | | | | 30.00 | 29.20 | 28.80 | 29.00 | Positive | 35.89 | 36.32 |

| A | M | N | O | P | Q | R | S | T | U | V |
|---|---|---|---|---|---|---|---|---|---|---|
| | Sample 2 | | | | | | | | | |
| Pool name | Original test date | Plate number | Sample | $Ct_{RPP}$ | $Ct_N$ | $Ct_E$ | $Ct_S$ | Result | cut-off N | cut-off RPP30 |
| 1N | | | | 28.94 | N/A | N/A | N/A | Negative | 35.59 | 34.67 |
| 2N | | | | 28.87 | N/A | N/A | N/A | Negative | 35.59 | 34.67 |
| 3N | | | | 29.48 | N/A | N/A | N/A | Negative | 35.59 | 34.67 |
| 4N | | | | 29.48 | N/A | N/A | N/A | Negative | 35.59 | 34.67 |
| 5N | | | | 30.36 | N/A | N/A | N/A | Negative | 35.59 | 34.67 |
| 6N | | | | 30.14 | N/A | N/A | N/A | Negative | 35.59 | 34.67 |
| 7N | | | | 28.96 | N/A | N/A | N/A | Negative | 35.59 | 34.67 |
| 8N | | | | 31.16 | 43.59 | N/A | N/A | Negative | 35.59 | 34.67 |
| 9N | | | | 29.30 | N/A | N/A | N/A | Negative | 35.59 | 34.67 |
| 10N | | | | 30.28 | 38.02 | N/A | N/A | Negative | 35.55 | 35.75 |
| 11N | | | | 30.20 | N/A | N/A | N/A | Negative | 35.60 | 35.06 |
| 12N | | | | 30.01 | 40.05 | N/A | N/A | Negative | 35.60 | 35.06 |
| 13N | | | | 30.16 | N/A | N/A | N/A | Negative | 35.60 | 35.06 |
| 14N | | | | 30.22 | N/A | N/A | N/A | Negative | 35.60 | 35.06 |
| 15N | | | | 29.46 | N/A | N/A | 37.69 | Negative | 35.60 | 35.06 |
| 16N | | | | 26.20 | N/A | N/A | 43.98 | Negative | 35.60 | 35.06 |
| 17N | | | | 29.48 | 36.31 | N/A | N/A | Negative | 35.60 | 35.06 |
| 18N | | | | 28.88 | N/A | N/A | N/A | Negative | 35.60 | 35.06 |
| 19N | | | | 29.63 | N/A | N/A | N/A | Negative | 35.60 | 35.06 |
| 20N | | | | 30.24 | 38.69 | N/A | N/A | Negative | 35.60 | 35.06 |
| 1P | | | | 28.94 | N/A | N/A | N/A | Negative | 35.59 | 34.67 |
| 2P | | | | 28.87 | N/A | N/A | N/A | Negative | 35.59 | 34.67 |
| 3P | | | | 29.48 | N/A | N/A | N/A | Negative | 35.59 | 34.67 |
| 4P | | | | 29.48 | N/A | N/A | N/A | Negative | 35.59 | 34.67 |
| 5P | | | | 30.35 | N/A | N/A | N/A | Negative | 35.59 | 34.67 |
| 6P | | | | 31.05 | N/A | N/A | N/A | Negative | 35.59 | 34.67 |
| 7P | | | | 30.36 | N/A | N/A | N/A | Negative | 35.59 | 34.67 |
| 8P | | | | 30.14 | N/A | N/A | N/A | Negative | 35.59 | 34.67 |
| 9P | | | | 29.94 | N/A | N/A | N/A | Negative | 35.59 | 34.67 |
| 10P | | | | 30.45 | N/A | N/A | N/A | Negative | 35.59 | 34.67 |
| 11P | | | | 28.96 | N/A | N/A | N/A | Negative | 35.59 | 34.67 |
| 12P | | | | 31.16 | 43.59 | N/A | N/A | Negative | 35.59 | 34.67 |
| 13P | | | | 29.30 | N/A | N/A | N/A | Negative | 35.59 | 34.67 |
| 14P | | | | 30.20 | N/A | N/A | N/A | Negative | 35.59 | 34.67 |
| 15P | | | | 30.71 | N/A | N/A | N/A | Negative | 35.59 | 34.67 |
| 16P | | | | 30.07 | N/A | N/A | N/A | Negative | 35.55 | 35.75 |
| 17P | | | | 28.88 | 39.95 | N/A | 37.85 | Negative | 35.55 | 35.75 |
| 18P | | | | 29.60 | N/A | N/A | N/A | Negative | 35.55 | 35.75 |
| 19P | | | | 29.27 | 39.31 | 42.68 | N/A | Negative | 35.55 | 35.75 |
| 20P | | | | 29.67 | N/A | N/A | 36.29 | Negative | 35.55 | 35.75 |
| 21P | | | | 30.28 | 38.02 | N/A | N/A | Negative | 35.55 | 35.75 |
| 22P | | | | 29.85 | N/A | N/A | 38.69 | Negative | 35.55 | 35.75 |

| A | W | X | Y | Z | AA | AB | AC | AD | AE | AF |
|---|---|---|---|---|---|---|---|---|---|---|
| | Pool | | | | | | | | | |
| Pool name | Test date | Plate number | Sample | $Ct_{RPP}$ | $Ct_N$ | $Ct_E$ | $Ct_S$ | Result | cut-off N | cut-off RPP30 |
| 1N | | | 1N | 29.72 | N/A | N/A | 41.24 | Negative | 35.54 | 36.06 |
| 2N | | | 2N | 29.97 | 38.77 | N/A | N/A | Negative | 35.54 | 36.06 |
| 3N | | | 3N | 31.05 | N/A | N/A | N/A | Negative | 35.54 | 36.06 |
| 4N | | | 4N | 30.23 | N/A | N/A | N/A | Negative | 35.54 | 36.06 |
| 5N | | | 5N | 30.77 | N/A | N/A | N/A | Negative | 35.97 | 34.87 |
| 6N | | | 6N | 30.51 | 37.11 | N/A | N/A | Negative | 35.97 | 34.87 |
| 7N | | | 7N | 29.18 | N/A | N/A | 42.43 | Negative | 36.42 | 36.49 |
| 8N | | | 8N | 30.27 | 42.24 | N/A | 41.85 | Negative | 36.42 | 36.49 |
| 9N | | | 9N | 30.22 | 37.15 | N/A | N/A | Negative | 36.42 | 36.49 |
| 10N | | | 10N | 29.25 | N/A | N/A | N/A | Negative | 36.10 | 34.84 |
| 11N | | | 11N | 29.89 | N/A | N/A | N/A | Negative | 36.15 | 35.56 |
| 12N | | | 12N | 29.62 | N/A | N/A | N/A | Negative | 36.15 | 35.56 |
| 13N | | | 13N | 30.01 | N/A | N/A | N/A | Negative | 36.15 | 35.56 |
| 14N | | | 14N | 29.62 | N/A | 42.02 | N/A | Negative | 36.15 | 35.56 |
| 15N | | | 15N | 28.94 | N/A | N/A | N/A | Negative | 36.15 | 35.56 |
| 16N | | | 16N | 29.27 | 40.62 | N/A | N/A | Negative | 36.15 | 35.56 |
| 17N | | | 17N | 30.19 | N/A | N/A | N/A | Negative | 36.15 | 35.56 |
| 18N | | | 18N | 29.86 | N/A | N/A | N/A | Negative | 36.15 | 35.56 |
| 19N | | | 19N | 29.81 | N/A | N/A | N/A | Negative | 36.15 | 35.56 |
| 20N | | | 20N | 30.13 | N/A | N/A | N/A | Negative | 36.15 | 35.56 |
| 1P | | | 1P | 29.94 | 14.52 | 16.19 | 16.66 | Positive | 36.42 | 36.49 |
| 2P | | | 2P | 30.12 | 18.21 | 19.05 | 18.82 | Positive | 36.42 | 36.49 |
| 3P | | | 3P | 30.86 | 21.31 | 23.07 | 23.23 | Positive | 36.42 | 36.49 |
| 4P | | | 4P | 30.11 | 22.92 | 23.08 | 22.91 | Positive | 36.42 | 36.49 |
| 5P | | | 5P | 31.23 | 18.43 | 19.50 | 19.91 | Positive | 36.42 | 36.49 |
| 6P | | | 6P | 29.85 | 18.07 | 19.24 | 19.56 | Positive | 36.42 | 36.49 |
| 7P | | | 7P | 31.25 | 18.92 | 20.93 | 21.63 | Positive | 36.42 | 36.49 |
| 8P | | | 8P | 31.16 | 22.81 | 24.44 | 24.68 | Positive | 36.42 | 36.49 |
| 9P | | | 9P | 29.81 | 17.82 | 19.52 | 20.00 | Positive | 36.42 | 36.49 |
| 10P | | | 10P | 28.40 | 23.86 | 24.85 | 25.31 | Positive | 36.42 | 36.49 |
| 11P | | | 11P | 29.84 | 15.23 | 17.08 | 17.49 | Positive | 36.42 | 36.49 |
| 12P | | | 12P | 30.53 | 16.46 | 17.89 | 18.58 | Positive | 36.42 | 36.49 |
| 13P | | | 13P | 27.19 | 22.65 | 22.55 | 22.27 | Positive | 36.42 | 36.49 |
| 14P | | | 14P | 31.05 | 16.85 | 18.43 | 19.18 | Positive | 36.42 | 36.49 |
| 15P | | | 15P | 30.34 | 16.42 | 18.03 | 18.30 | Positive | 36.42 | 36.49 |
| 16P | | | 16P | 30.95 | 19.36 | 20.91 | 21.34 | Positive | 35.97 | 34.87 |
| 17P | | | 17P | 29.95 | 20.58 | 22.05 | 22.22 | Positive | 35.97 | 34.87 |
| 18P | | | 18P | 30.35 | 18.37 | 20.15 | 20.58 | Positive | 35.97 | 34.87 |
| 19P | | | 19P | 30.37 | 17.99 | 19.47 | 19.71 | Positive | 35.97 | 34.87 |
| 20P | | | 20P | 30.39 | 20.47 | 21.37 | 21.57 | Positive | 35.97 | 34.87 |
| 21P | | | 21P | 29.13 | 17.74 | 19.55 | 19.75 | Positive | 35.97 | 34.87 |
| 22P | | | 22P | 30.81 | 29.40 | 29.74 | 29.99 | Positive | 35.97 | 34.87 |

**Positives**

| A | B | C | D | E | F | G | H | I | J | K | L | M |
|---|---|---|---|---|---|---|---|---|---|---|---|---|
| | $Ct_{RPP}$ | | | $Ct_N$ | | | $Ct_E$ | | | $Ct_S$ | | |
| **Pool name** | **Individual** | **Pool** | **Ct diffrence** | **Individual** | **Pool** | **Ct diffrence** | **Individual** | **Pool** | **Ct diffrence** | **Individual** | **Pool** | **Ct diffrence** |
| 1P | 29.62 | 29.94 | 0.32 | 13.39 | 14.52 | 1.13 | 14.94 | 16.19 | 1.24 | 15.31 | 16.66 | 1.34 |
| 2P | 30.67 | 30.12 | -0.55 | 17.24 | 18.21 | 0.97 | 17.99 | 19.05 | 1.06 | 17.77 | 18.82 | 1.05 |
| 3P | 30.61 | 30.86 | 0.25 | 18.55 | 21.31 | 2.76 | 20.04 | 23.07 | 3.03 | 20.17 | 23.23 | 3.06 |
| 4P | 31.02 | 30.11 | -0.90 | 22.28 | 22.92 | 0.65 | 22.27 | 23.08 | 0.81 | 22.07 | 22.91 | 0.84 |
| 5P | 31.22 | 31.23 | 0.01 | 17.15 | 18.43 | 1.29 | 18.22 | 19.50 | 1.27 | 18.54 | 19.91 | 1.37 |
| 6P | 29.15 | 29.85 | 0.70 | 17.18 | 18.07 | 0.89 | 18.54 | 19.24 | 0.70 | 18.88 | 19.56 | 0.68 |
| 7P | 29.99 | 31.25 | 1.26 | 17.05 | 18.92 | 1.87 | 19.17 | 20.93 | 1.76 | 19.53 | 21.63 | 2.10 |
| 8P | 32.00 | 31.16 | -0.84 | 22.00 | 22.81 | 0.81 | 23.00 | 24.44 | 1.44 | 23.30 | 24.68 | 1.38 |
| 9P | 29.13 | 29.81 | 0.68 | 16.78 | 17.82 | 1.04 | 18.44 | 19.52 | 1.08 | 19.05 | 20.00 | 0.96 |
| 10P | 29.02 | 28.40 | -0.63 | 24.75 | 23.86 | -0.89 | 25.29 | 24.85 | -0.44 | 25.51 | 25.31 | -0.20 |
| 11P | 30.89 | 29.84 | -1.05 | 15.28 | 15.23 | -0.06 | 17.27 | 17.08 | -0.20 | 17.78 | 17.49 | -0.30 |
| 12P | 30.88 | 30.53 | -0.35 | 17.33 | 16.46 | -0.87 | 18.76 | 17.89 | -0.88 | 19.30 | 18.58 | -0.71 |
| 13P | 27.15 | 27.19 | 0.04 | 22.54 | 22.65 | 0.11 | 22.26 | 22.55 | 0.28 | 21.56 | 22.27 | 0.71 |
| 14P | 31.85 | 31.05 | -0.80 | 15.96 | 16.85 | 0.90 | 17.44 | 18.43 | 1.00 | 17.86 | 19.18 | 1.32 |
| 15P | 30.25 | 30.34 | 0.08 | 15.27 | 16.42 | 1.16 | 16.31 | 18.03 | 1.72 | 16.54 | 18.30 | 1.76 |
| 16P | 31.57 | 30.95 | -0.62 | 20.15 | 19.36 | -0.79 | 21.46 | 20.91 | -0.55 | 22.22 | 21.34 | -0.88 |
| 17P | 29.94 | 29.95 | 0.02 | 20.48 | 20.58 | 0.10 | 21.76 | 22.05 | 0.29 | 22.11 | 22.22 | 0.11 |
| 18P | 30.03 | 30.35 | 0.33 | 17.27 | 18.37 | 1.10 | 18.81 | 20.15 | 1.34 | 19.38 | 20.58 | 1.20 |
| 19P | 29.23 | 30.37 | 1.14 | 16.83 | 17.99 | 1.15 | 17.94 | 19.47 | 1.54 | 17.89 | 19.71 | 1.82 |
| 20P | 30.28 | 30.39 | 0.10 | 19.54 | 20.47 | 0.93 | 20.68 | 21.37 | 0.69 | 20.97 | 21.57 | 0.60 |
| 21P | 31.28 | 29.13 | -2.16 | 16.16 | 17.74 | 1.58 | 17.94 | 19.55 | 1.61 | 18.29 | 19.75 | 1.47 |
| 22P | 30.00 | 30.81 | 0.81 | 29.20 | 29.40 | 0.20 | 28.80 | 29.74 | 0.94 | 29.00 | 29.99 | 0.99 |

Regression

| A | B | C | D | E | F | G |
|---|---|---|---|---|---|---|
| | $Ct_N$ | | $Ct_E$ | | $Ct_S$ | |
| **Pool name** | **Individual** | **Pool** | **Individual** | **Pool** | **Individual** | **Pool** |
| 1P | 13.39 | 14.52 | 14.94 | 16.19 | 15.31 | 16.66 |
| 2P | 17.24 | 18.21 | 17.99 | 19.05 | 17.77 | 18.82 |
| 3P | 18.55 | 21.31 | 20.04 | 23.07 | 20.17 | 23.23 |
| 4P | 22.28 | 22.92 | 22.27 | 23.08 | 22.07 | 22.91 |
| 5P | 17.15 | 18.43 | 18.22 | 19.50 | 18.54 | 19.91 |
| 6P | 17.18 | 18.07 | 18.54 | 19.24 | 18.88 | 19.56 |
| 7P | 17.05 | 18.92 | 19.17 | 20.93 | 19.53 | 21.63 |
| 8P | 22.00 | 22.81 | 23.00 | 24.44 | 23.30 | 24.68 |
| 9P | 16.78 | 17.82 | 18.44 | 19.52 | 19.05 | 20.00 |
| 10P | 24.75 | 23.86 | 25.29 | 24.85 | 25.51 | 25.31 |
| 11P | 15.28 | 15.23 | 17.27 | 17.08 | 17.78 | 17.49 |
| 12P | 17.33 | 16.46 | 18.76 | 17.89 | 19.30 | 18.58 |
| 13P | 22.54 | 22.65 | 22.26 | 22.55 | 21.56 | 22.27 |
| 14P | 15.96 | 16.85 | 17.44 | 18.43 | 17.86 | 19.18 |
| 15P | 15.27 | 16.42 | 16.31 | 18.03 | 16.54 | 18.30 |
| 16P | 20.15 | 19.36 | 21.46 | 20.91 | 22.22 | 21.34 |
| 17P | 20.48 | 20.58 | 21.76 | 22.05 | 22.11 | 22.22 |
| 18P | 17.27 | 18.37 | 18.81 | 20.15 | 19.38 | 20.58 |
| 19P | 16.83 | 17.99 | 17.94 | 19.47 | 17.89 | 19.71 |
| 20P | 19.54 | 20.47 | 20.68 | 21.37 | 20.97 | 21.57 |
| 21P | 16.16 | 17.74 | 17.94 | 19.55 | 18.29 | 19.75 |
| 22P | 29.20 | 29.40 | 28.80 | 29.74 | 29.00 | 29.99 |

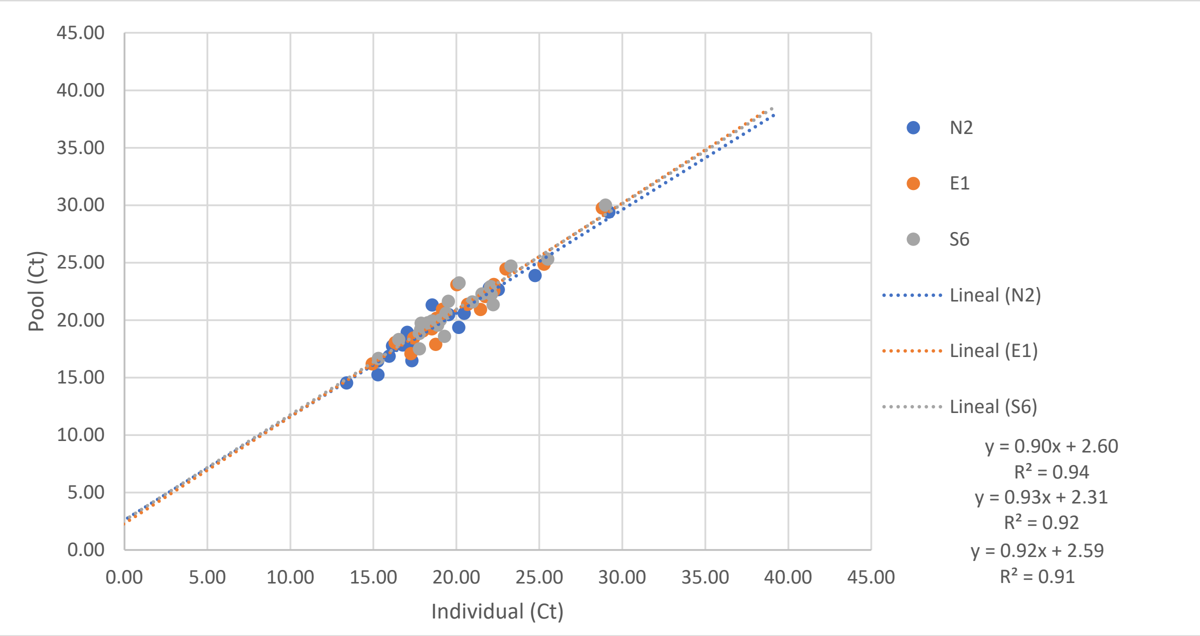


| | Pool | | | Individual | | |
|---|---|---|---|---|---|---|
| | LOD | 95% CI low | 95% CI upp | LOD | 95% CI low | 95% CI upper |
| N | 37.29 | 36.91 | 37.67 | 38.54 | 38.12 | 38.97 |
| E | 37.29 | 36.91 | 37.67 | 37.61 | 37.20 | 38.02 |
| S | 37.29 | 36.91 | 37.67 | 37.72 | 37.30 | 38.13 |

For a pool which is at the limit of detection, the individual sample would have had a Ct around 38.54 for N2 based on the regression analysis. Near the limit of detection, the regression analysis indicated this slight increase in Ct due to pooling when n=2, this is likely due to little change in Cts due to pooling two samples.
