## Supplementary material for "Development and Implementation of a scalable and versatile test for COVID-19 diagnostics in rural communities": Sup_Table_XII

**Pool raw data**

| B | C | D | E | F | G | H | I | J | K | L |
|---|---|---|---|---|---|---|---|---|---|---|
| | Sample 1 | | | | | | | | | |
| Pool name | Original test date | Plate number | Sample | $Ct_{RPP}$ | $Ct_{N}$ | $Ct_{E}$ | $Ct_{S}$ | Result | cut-off N | cut-off RPP30 |
| 1N | | | | 29.49 | 40.66 | N/A | N/A | Negative | 35.59 | 34.67 |
| 2N | | | | 29.02 | 40.24 | 37.58 | N/A | Negative | 35.59 | 34.67 |
| 3N | | | | 29.13 | N/A | N/A | N/A | Negative | 35.59 | 34.67 |
| 4N | | | | 31.06 | N/A | N/A | N/A | Negative | 35.59 | 34.67 |
| 5N | | | | 30.59 | N/A | N/A | N/A | Negative | 35.59 | 34.67 |
| 6N | | | | 28.84 | N/A | N/A | N/A | Negative | 35.59 | 34.67 |
| 7N | | | | 30.09 | N/A | N/A | N/A | Negative | 35.59 | 34.67 |
| 8N | | | | 29.50 | N/A | N/A | N/A | Negative | 35.59 | 34.67 |
| 9N | | | | 30.05 | N/A | N/A | N/A | Negative | 35.59 | 34.67 |
| 10N | | | | 29.97 | N/A | N/A | N/A | Negative | 35.59 | 34.67 |
| 11N | | | | 29.37 | N/A | N/A | 43.98 | Negative | 35.60 | 35.06 |
| 12N | | | | 29.96 | N/A | N/A | N/A | Negative | 35.60 | 35.06 |
| 13N | | | | 31.63 | N/A | N/A | N/A | Negative | 35.60 | 35.06 |
| 14N | | | | 29.81 | N/A | N/A | N/A | Negative | 35.60 | 35.06 |
| 15N | | | | 28.87 | N/A | N/A | N/A | Negative | 35.60 | 35.06 |
| 16N | | | | 29.23 | N/A | N/A | N/A | Negative | 35.60 | 35.06 |
| 17N | | | | 29.97 | N/A | N/A | N/A | Negative | 35.60 | 35.06 |
| 18N | | | | 30.02 | N/A | N/A | N/A | Negative | 35.60 | 35.06 |
| 19N | | | | 29.96 | N/A | N/A | N/A | Negative | 35.60 | 35.06 |
| 20N | | | | 29.95 | N/A | N/A | N/A | Negative | 35.60 | 35.06 |
| 1P | | | | 29.62 | 13.39 | 14.94 | 15.31 | Positive | 35.87 | 36.02 |
| 2P | | | | 30.67 | 17.24 | 17.99 | 17.77 | Positive | 35.87 | 36.02 |
| 3P | | | | 30.61 | 18.55 | 20.04 | 20.17 | Positive | 35.87 | 36.02 |
| 4P | | | | 31.02 | 22.28 | 22.27 | 22.07 | Positive | 36.09 | 35.73 |
| 5P | | | | 31.22 | 17.15 | 18.22 | 18.54 | Positive | 36.09 | 35.73 |
| 6P | | | | 29.15 | 17.18 | 18.54 | 18.88 | Positive | 36.09 | 35.73 |
| 7P | | | | 29.99 | 17.05 | 19.17 | 19.53 | Positive | 35.97 | 36.32 |
| 8P | | | | 32.00 | 22.00 | 23.00 | 23.30 | Positive | 35.47 | 36.52 |
| 9P | | | | 29.13 | 16.78 | 18.44 | 19.05 | Positive | 36.14 | 36.22 |
| 10P | | | | 29.02 | 24.75 | 25.29 | 25.51 | Positive | 35.53 | 35.84 |
| 11P | | | | 30.89 | 15.28 | 17.27 | 17.78 | Positive | 35.88 | 34.58 |
| 12P | | | | 30.88 | 17.33 | 18.76 | 19.30 | Positive | 35.40 | 35.38 |
| 13P | | | | 27.15 | 22.54 | 22.26 | 21.56 | Positive | 35.40 | 35.38 |
| 14P | | | | 31.85 | 15.96 | 17.44 | 17.86 | Positive | 35.40 | 35.38 |
| 15P | | | | 30.25 | 15.27 | 16.31 | 16.54 | Positive | 35.40 | 35.38 |
| 16P | | | | 31.57 | 20.15 | 21.46 | 22.22 | Positive | 35.40 | 35.38 |
| 17P | | | | 29.94 | 20.48 | 21.76 | 22.11 | Positive | 35.40 | 35.38 |
| 18P | | | | 30.03 | 17.27 | 18.81 | 19.38 | Positive | 35.40 | 35.38 |
| 19P | | | | 29.23 | 16.83 | 17.94 | 17.89 | Positive | 35.40 | 35.38 |
| 20P | | | | 30.28 | 19.54 | 20.68 | 20.97 | Positive | 35.47 | 35.39 |
| 21P | | | | 31.28 | 16.16 | 17.94 | 18.29 | Positive | 35.47 | 35.39 |
| 22P | | | | 30.00 | 29.20 | 28.80 | 29.00 | Positive | 35.89 | 36.32 |

| B | M | N | O | P | Q | R | S | T | U | V |
|---|---|---|---|---|---|---|---|---|---|---|
| | Sample 2 | | | | | | | | | |
| Pool name | Original test date | Plate number | Sample | $Ct_{RPP}$ | $Ct_{N}$ | $Ct_{E}$ | $Ct_{S}$ | Result | cut-off N | cut-off RPP30 |
| 1N | | | | 28.76 | 37.60 | N/A | N/A | Negative | 35.81 | 35.08 |
| 2N | | | | 29.04 | 38.01 | 42.29 | 39.06 | Negative | 35.81 | 35.08 |
| 3N | | | | 31.13 | N/A | N/A | N/A | Negative | 35.81 | 35.08 |
| 4N | | | | 30.86 | N/A | 42.50 | N/A | Negative | 35.81 | 35.08 |
| 5N | | | | 30.76 | N/A | N/A | 38.68 | Negative | 35.81 | 35.08 |
| 6N | | | | 31.01 | N/A | N/A | N/A | Negative | 35.81 | 35.08 |
| 7N | | | | 31.47 | N/A | N/A | N/A | Negative | 35.81 | 35.08 |
| 8N | | | | 30.12 | N/A | N/A | N/A | Negative | 35.81 | 35.08 |
| 9N | | | | 31.92 | N/A | N/A | N/A | Negative | 35.81 | 35.08 |
| 10N | | | | 31.13 | N/A | N/A | N/A | Negative | 35.81 | 35.08 |
| 11N | | | | 30.20 | N/A | N/A | N/A | Negative | 35.60 | 35.06 |
| 12N | | | | 30.01 | 40.05 | N/A | N/A | Negative | 35.60 | 35.06 |
| 13N | | | | 30.16 | N/A | N/A | N/A | Negative | 35.60 | 35.06 |
| 14N | | | | 30.22 | N/A | N/A | N/A | Negative | 35.60 | 35.06 |
| 15N | | | | 29.46 | N/A | N/A | 37.69 | Negative | 35.60 | 35.06 |
| 16N | | | | 26.20 | N/A | N/A | 43.98 | Negative | 35.60 | 35.06 |
| 17N | | | | 29.48 | 36.31 | N/A | N/A | Negative | 35.60 | 35.06 |
| 18N | | | | 28.88 | N/A | N/A | N/A | Negative | 35.60 | 35.06 |
| 19N | | | | 29.63 | N/A | N/A | N/A | Negative | 35.60 | 35.06 |
| 20N | | | | 30.24 | 38.69 | N/A | N/A | Negative | 35.60 | 35.06 |
| 1P | | | | 29.20 | N/A | N/A | N/A | Negative | 35.81 | 35.08 |
| 2P | | | | 28.06 | N/A | N/A | N/A | Negative | 35.81 | 35.08 |
| 3P | | | | 29.21 | N/A | N/A | 44.93 | Negative | 35.81 | 35.08 |
| 4P | | | | 29.94 | N/A | N/A | N/A | Negative | 35.81 | 35.08 |
| 5P | | | | 28.76 | 37.60 | N/A | N/A | Negative | 35.81 | 35.08 |
| 6P | | | | 29.04 | 38.01 | 42.29 | 39.06 | Negative | 35.81 | 35.08 |
| 7P | | | | 30.11 | N/A | N/A | N/A | Negative | 35.81 | 35.08 |
| 8P | | | | 29.75 | N/A | N/A | 41.95 | Negative | 35.81 | 35.08 |
| 9P | | | | 29.36 | N/A | N/A | N/A | Negative | 35.81 | 35.08 |
| 10P | | | | 30.27 | N/A | N/A | N/A | Negative | 35.81 | 35.08 |
| 11P | | | | 31.13 | N/A | N/A | N/A | Negative | 35.81 | 35.08 |
| 12P | | | | 31.11 | N/A | N/A | 41.61 | Negative | 35.81 | 35.08 |
| 13P | | | | 30.86 | N/A | 42.50 | N/A | Negative | 35.81 | 35.08 |
| 14P | | | | 30.76 | N/A | N/A | 38.68 | Negative | 35.81 | 35.08 |
| 15P | | | | 31.01 | N/A | N/A | N/A | Negative | 35.81 | 35.08 |
| 16P | | | | 31.47 | N/A | N/A | N/A | Negative | 35.81 | 35.08 |
| 17P | | | | 30.12 | N/A | N/A | N/A | Negative | 35.81 | 35.08 |
| 18P | | | | 31.92 | N/A | N/A | N/A | Negative | 35.81 | 35.08 |
| 19P | | | | 31.07 | N/A | N/A | N/A | Negative | 35.81 | 35.08 |
| 20P | | | | 30.21 | N/A | N/A | 37.06 | Negative | 35.81 | 35.08 |
| 21P | | | | 30.75 | N/A | N/A | N/A | Negative | 35.81 | 35.08 |
| 22P | | | | 30.11 | N/A | 44.99 | N/A | Negative | 35.81 | 35.08 |

| B | W | X | Y | Z | AA | AB | AC | AD | AE | AF |
|---|---|---|---|---|---|---|---|---|---|---|
| | Sample 3 | | | | | | | | | |
| Pool name | Original test date | Plate number | Sample | $Ct_{RPP}$ | $Ct_{N}$ | $Ct_{E}$ | $Ct_{S}$ | Result | cut-off N | cut-off RPP30 |
| 1N | | | | 28.98 | N/A | N/A | N/A | Negative | 35.81 | 35.08 |
| 2N | | | | 30.07 | N/A | 43.27 | 44.51 | Negative | 35.81 | 35.08 |
| 3N | | | | 29.72 | N/A | N/A | N/A | Negative | 35.81 | 35.08 |
| 4N | | | | 29.47 | 41.01 | N/A | N/A | Negative | 35.81 | 35.08 |
| 5N | | | | 29.38 | N/A | N/A | N/A | Negative | 35.81 | 35.08 |
| 6N | | | | 28.99 | 41.86 | N/A | N/A | Negative | 35.81 | 35.08 |
| 7N | | | | 29.25 | N/A | N/A | N/A | Negative | 35.81 | 35.08 |
| 8N | | | | 29.48 | N/A | N/A | N/A | Negative | 35.81 | 35.08 |
| 9N | | | | 29.22 | N/A | N/A | N/A | Negative | 35.81 | 35.08 |
| 10N | | | | 29.72 | N/A | N/A | N/A | Negative | 35.59 | 34.67 |
| 11N | | | | 31.13 | N/A | N/A | N/A | Negative | 35.60 | 35.06 |
| 12N | | | | 29.75 | N/A | N/A | N/A | Negative | 35.60 | 35.06 |
| 13N | | | | 30.14 | N/A | N/A | N/A | Negative | 35.60 | 35.06 |
| 14N | | | | 30.18 | 44.59 | N/A | N/A | Negative | 35.60 | 35.06 |
| 15N | | | | 28.86 | N/A | N/A | 40.91 | Negative | 35.60 | 35.06 |
| 16N | | | | 30.26 | N/A | N/A | N/A | Negative | 35.60 | 35.06 |
| 17N | | | | 30.37 | N/A | N/A | 40.47 | Negative | 35.60 | 35.06 |
| 18N | | | | 29.55 | N/A | N/A | N/A | Negative | 35.60 | 35.06 |
| 19N | | | | 30.03 | N/A | N/A | N/A | Negative | 35.52 | 34.96 |
| 20N | | | | 32.18 | N/A | N/A | N/A | Negative | 35.52 | 34.96 |
| 1P | | | | 34.10 | N/A | N/A | N/A | Negative | 35.81 | 35.08 |
| 2P | | | | 33.04 | N/A | N/A | N/A | Negative | 35.81 | 35.08 |
| 3P | | | | 29.10 | N/A | N/A | N/A | Negative | 35.81 | 35.08 |
| 4P | | | | 29.38 | N/A | N/A | N/A | Negative | 35.81 | 35.08 |
| 5P | | | | 28.98 | N/A | N/A | N/A | Negative | 35.81 | 35.08 |
| 6P | | | | 30.07 | N/A | 43.27 | 44.51 | Negative | 35.81 | 35.08 |
| 7P | | | | 28.95 | N/A | N/A | N/A | Negative | 35.81 | 35.08 |
| 8P | | | | 28.93 | N/A | N/A | 44.70 | Negative | 35.81 | 35.08 |
| 9P | | | | 30.07 | 43.32 | N/A | N/A | Negative | 35.81 | 35.08 |
| 10P | | | | 29.38 | N/A | N/A | 38.52 | Negative | 35.81 | 35.08 |
| 11P | | | | 29.72 | N/A | N/A | N/A | Negative | 35.81 | 35.08 |
| 12P | | | | 28.55 | N/A | 40.18 | 41.41 | Negative | 35.81 | 35.08 |
| 13P | | | | 29.47 | 41.01 | N/A | N/A | Negative | 35.81 | 35.08 |
| 14P | | | | 29.38 | N/A | N/A | N/A | Negative | 35.81 | 35.08 |
| 15P | | | | 28.99 | 41.86 | N/A | N/A | Negative | 35.81 | 35.08 |
| 16P | | | | 29.25 | N/A | N/A | N/A | Negative | 35.81 | 35.08 |
| 17P | | | | 29.48 | N/A | N/A | N/A | Negative | 35.81 | 35.08 |
| 18P | | | | 29.22 | N/A | N/A | N/A | Negative | 35.81 | 35.08 |
| 19P | | | | 29.46 | N/A | N/A | N/A | Negative | 35.59 | 34.67 |
| 20P | | | | 30.79 | N/A | N/A | N/A | Negative | 35.59 | 34.67 |
| 21P | | | | 29.53 | N/A | N/A | N/A | Negative | 35.59 | 34.67 |
| 22P | | | | 29.96 | N/A | N/A | 42.90 | Negative | 35.59 | 34.67 |

| B | AG | AH | AI | AJ | AK | AL | AM | AN | AO | AP |
|---|---|---|---|---|---|---|---|---|---|---|
| | Pool | | | | | | | | | |
| Pool name | Test date | Plate number | Sample | $Ct_{RPP}$ | $Ct_{N}$ | $Ct_{E}$ | $Ct_{S}$ | Result | cut-off N | cut-off RPP30 |
| 1N | | | | 29.40 | 37.25 | N/A | N/A | Negative | 36.08 | 35.95 |
| 2N | | | | 29.66 | N/A | 42.87 | 42.20 | Negative | 36.08 | 35.95 |
| 3N | | | | 29.64 | 37.13 | N/A | 36.76 | Negative | 36.08 | 35.95 |
| 4N | | | | 30.05 | N/A | 44.93 | 44.80 | Negative | 36.08 | 35.95 |
| 5N | | | | 29.65 | 37.90 | N/A | N/A | Negative | 36.08 | 35.95 |
| 6N | | | | 30.67 | N/A | N/A | N/A | Negative | 36.08 | 35.95 |
| 7N | | | | 31.04 | 37.52 | N/A | N/A | Negative | 36.08 | 35.95 |
| 8N | | | | 29.84 | 36.22 | N/A | 38.27 | Negative | 36.08 | 35.95 |
| 9N | | | | 29.79 | N/A | N/A | N/A | Negative | 36.10 | 34.84 |
| 10N | | | | 31.14 | N/A | N/A | N/A | Negative | 36.10 | 34.84 |
| 11N | | | | 29.83 | N/A | N/A | 40.18 | Negative | 36.15 | 35.56 |
| 12N | | | | 29.48 | N/A | N/A | N/A | Negative | 36.15 | 35.56 |
| 13N | | | | 30.46 | N/A | N/A | N/A | Negative | 36.15 | 35.56 |
| 14N | | | | 30.11 | N/A | N/A | N/A | Negative | 36.15 | 35.56 |
| 15N | | | | 29.32 | N/A | N/A | N/A | Negative | 36.15 | 35.56 |
| 16N | | | | 29.88 | N/A | N/A | N/A | Negative | 36.15 | 35.56 |
| 17N | | | | 29.78 | N/A | N/A | N/A | Negative | 36.15 | 35.56 |
| 18N | | | | 29.60 | N/A | N/A | N/A | Negative | 36.15 | 35.56 |
| 19N | | | | 30.03 | N/A | N/A | N/A | Negative | 36.15 | 35.56 |
| 20N | | | | 30.12 | N/A | N/A | N/A | Negative | 36.15 | 35.56 |
| 1P | | | | 29.66 | 15.21 | 16.09 | 16.89 | Positive | 36.08 | 35.95 |
| 2P | | | | 29.83 | 18.22 | 18.52 | 18.65 | Positive | 36.08 | 35.95 |
| 3P | | | | 31.34 | 21.62 | 22.81 | 23.21 | Positive | 36.08 | 35.95 |
| 4P | | | | 31.78 | 23.63 | 23.67 | 23.57 | Positive | 36.08 | 35.95 |
| 5P | | | | 30.01 | 18.84 | 19.88 | 20.23 | Positive | 36.08 | 35.95 |
| 6P | | | | 29.64 | 18.71 | 19.63 | 20.10 | Positive | 36.08 | 35.95 |
| 7P | | | | 30.74 | 18.52 | 20.06 | 20.94 | Positive | 36.08 | 35.95 |
| 8P | | | | 30.63 | 23.69 | 24.80 | 25.28 | Positive | 36.08 | 35.95 |
| 9P | | | | 29.30 | 18.04 | 19.19 | 20.01 | Positive | 36.08 | 35.95 |
| 10P | | | | 29.36 | 24.88 | 25.64 | 26.38 | Positive | 36.08 | 35.95 |
| 11P | | | | 30.88 | 16.00 | 17.49 | 18.67 | Positive | 36.08 | 35.95 |
| 12P | | | | 31.38 | 17.91 | 19.03 | 19.89 | Positive | 36.08 | 35.95 |
| 13P | | | | 28.31 | 23.54 | 23.50 | 23.32 | Positive | 36.08 | 35.95 |
| 14P | | | | 30.42 | 16.05 | 17.09 | 17.86 | Positive | 36.08 | 35.95 |
| 15P | | | | 30.23 | 17.07 | 18.10 | 18.75 | Positive | 36.08 | 35.95 |
| 16P | | | | 31.58 | 20.74 | 22.05 | 22.97 | Positive | 36.08 | 35.95 |
| 17P | | | | 30.17 | 21.70 | 22.80 | 23.48 | Positive | 36.08 | 35.95 |
| 18P | | | | 30.09 | 17.77 | 19.30 | 20.21 | Positive | 36.08 | 35.95 |
| 19P | | | | 30.47 | 17.97 | 19.10 | 19.44 | Positive | 36.08 | 35.95 |
| 20P | | | | 30.77 | 21.28 | 22.08 | 22.53 | Positive | 36.08 | 35.95 |
| 21P | | | | 31.06 | 18.31 | 20.25 | 20.70 | Positive | 35.19 | 34.96 |
| 22P | | | | 31.15 | 30.92 | 30.70 | 30.94 | Positive | 35.19 | 34.96 |

**Positives**

| A | B | C | D | E | F | G | H | I | J | K | L | M |
|---|---|---|---|---|---|---|---|---|---|---|---|---|
| | $Ct_{RPP}$ | | | $Ct_N$ | | | $Ct_E$ | | | $Ct_S$ | | |
| **Pool name** | **Individual** | **Pool** | **Ct diffrence** | **Individual** | **Pool** | **Ct diffrence** | **Individual** | **Pool** | **Ct diffrence** | **Individual** | **Pool** | **Ct diffrence** |
| 1P | 29.62 | 29.66 | 0.04 | 13.39 | 15.21 | 1.81 | 14.94 | 16.09 | 1.14 | 15.31 | 16.89 | 1.58 |
| 2P | 30.67 | 29.83 | -0.83 | 17.24 | 18.22 | 0.98 | 17.99 | 18.52 | 0.53 | 17.77 | 18.65 | 0.88 |
| 3P | 30.61 | 31.34 | 0.73 | 18.55 | 21.62 | 3.07 | 20.04 | 22.81 | 2.77 | 20.17 | 23.21 | 3.04 |
| 4P | 31.02 | 31.78 | 0.76 | 22.28 | 23.63 | 1.35 | 22.27 | 23.67 | 1.40 | 22.07 | 23.57 | 1.50 |
| 5P | 31.22 | 30.01 | -1.21 | 17.15 | 18.84 | 1.69 | 18.22 | 19.88 | 1.66 | 18.54 | 20.23 | 1.69 |
| 6P | 29.15 | 29.64 | 0.49 | 17.18 | 18.71 | 1.53 | 18.54 | 19.63 | 1.09 | 18.88 | 20.10 | 1.22 |
| 7P | 29.99 | 30.74 | 0.75 | 17.05 | 18.52 | 1.47 | 19.17 | 20.06 | 0.89 | 19.53 | 20.94 | 1.42 |
| 8P | 32.00 | 30.63 | -1.37 | 22.00 | 23.69 | 1.69 | 23.00 | 24.80 | 1.80 | 23.30 | 25.28 | 1.98 |
| 9P | 29.13 | 29.30 | 0.17 | 16.78 | 18.04 | 1.26 | 18.44 | 19.19 | 0.75 | 19.05 | 20.01 | 0.96 |
| 10P | 29.02 | 29.36 | 0.34 | 24.75 | 24.88 | 0.13 | 25.29 | 25.64 | 0.35 | 25.51 | 26.38 | 0.87 |
| 11P | 30.89 | 30.88 | -0.01 | 15.28 | 16.00 | 0.71 | 17.27 | 17.49 | 0.22 | 17.78 | 18.67 | 0.89 |
| 12P | 30.88 | 31.38 | 0.50 | 17.33 | 17.91 | 0.58 | 18.76 | 19.03 | 0.27 | 19.30 | 19.89 | 0.59 |
| 13P | 27.15 | 28.31 | 1.16 | 22.54 | 23.54 | 1.00 | 22.26 | 23.50 | 1.24 | 21.56 | 23.32 | 1.76 |
| 14P | 31.85 | 30.42 | -1.43 | 15.96 | 16.05 | 0.09 | 17.44 | 17.09 | -0.34 | 17.86 | 17.86 | 0.00 |
| 15P | 30.25 | 30.23 | -0.02 | 15.27 | 17.07 | 1.80 | 16.31 | 18.10 | 1.79 | 16.54 | 18.75 | 2.21 |
| 16P | 31.57 | 31.58 | 0.01 | 20.15 | 20.74 | 0.59 | 21.46 | 22.05 | 0.59 | 22.22 | 22.97 | 0.75 |
| 17P | 29.94 | 30.17 | 0.24 | 20.48 | 21.70 | 1.21 | 21.76 | 22.80 | 1.04 | 22.11 | 23.48 | 1.37 |
| 18P | 30.03 | 30.09 | 0.06 | 17.27 | 17.77 | 0.50 | 18.81 | 19.30 | 0.49 | 19.38 | 20.21 | 0.83 |
| 19P | 29.23 | 30.47 | 1.24 | 16.83 | 17.97 | 1.13 | 17.94 | 19.10 | 1.16 | 17.89 | 19.44 | 1.55 |
| 20P | 30.28 | 30.77 | 0.49 | 19.54 | 21.28 | 1.73 | 20.68 | 22.08 | 1.40 | 20.97 | 22.53 | 1.56 |
| 21P | 31.28 | 31.06 | -0.23 | 16.16 | 18.31 | 2.15 | 17.94 | 20.25 | 2.31 | 18.29 | 20.70 | 2.42 |
| 22P | 30.00 | 31.15 | 1.15 | 29.20 | 30.92 | 1.72 | 28.80 | 30.70 | 1.90 | 29.00 | 30.94 | 1.94 |

**Regression**

| A | B | C | D | E | F | G |
|---|---|---|---|---|---|---|
| | $Ct_N$ | | $Ct_E$ | | $Ct_S$ | |
| **Pool name** | **Individual** | **Pool** | **Individual** | **Pool** | **Individual** | **Pool** |
| 1P | 13.39 | 15.21 | 14.94 | 16.09 | 15.31 | 16.89 |
| 2P | 17.24 | 18.22 | 17.99 | 18.52 | 17.77 | 18.65 |
| 3P | 18.55 | 21.62 | 20.04 | 22.81 | 20.17 | 23.21 |
| 4P | 22.28 | 23.63 | 22.27 | 23.67 | 22.07 | 23.57 |
| 5P | 17.15 | 18.84 | 18.22 | 19.88 | 18.54 | 20.23 |
| 6P | 17.18 | 18.71 | 18.54 | 19.63 | 18.88 | 20.10 |
| 7P | 17.05 | 18.52 | 19.17 | 20.06 | 19.53 | 20.94 |
| 8P | 22.00 | 23.69 | 23.00 | 24.80 | 23.30 | 25.28 |
| 9P | 16.78 | 18.04 | 18.44 | 19.19 | 19.05 | 20.01 |
| 10P | 24.75 | 24.88 | 25.29 | 25.64 | 25.51 | 26.38 |
| 11P | 15.28 | 16.00 | 17.27 | 17.49 | 17.78 | 18.67 |
| 12P | 17.33 | 17.91 | 18.76 | 19.03 | 19.30 | 19.89 |
| 13P | 22.54 | 23.54 | 22.26 | 23.50 | 21.56 | 23.32 |
| 14P | 15.96 | 16.05 | 17.44 | 17.09 | 17.86 | 17.86 |
| 15P | 15.27 | 17.07 | 16.31 | 18.10 | 16.54 | 18.75 |
| 16P | 20.15 | 20.74 | 21.46 | 22.05 | 22.22 | 22.97 |
| 17P | 20.48 | 21.70 | 21.76 | 22.80 | 22.11 | 23.48 |
| 18P | 17.27 | 17.77 | 18.81 | 19.30 | 19.38 | 20.21 |
| 19P | 16.83 | 17.97 | 17.94 | 19.10 | 17.89 | 19.44 |
| 20P | 19.54 | 21.28 | 20.68 | 22.08 | 20.97 | 22.53 |
| 21P | 16.16 | 18.31 | 17.94 | 20.25 | 18.29 | 20.70 |
| 22P | 29.20 | 30.92 | 28.80 | 30.70 | 29.00 | 30.94 |

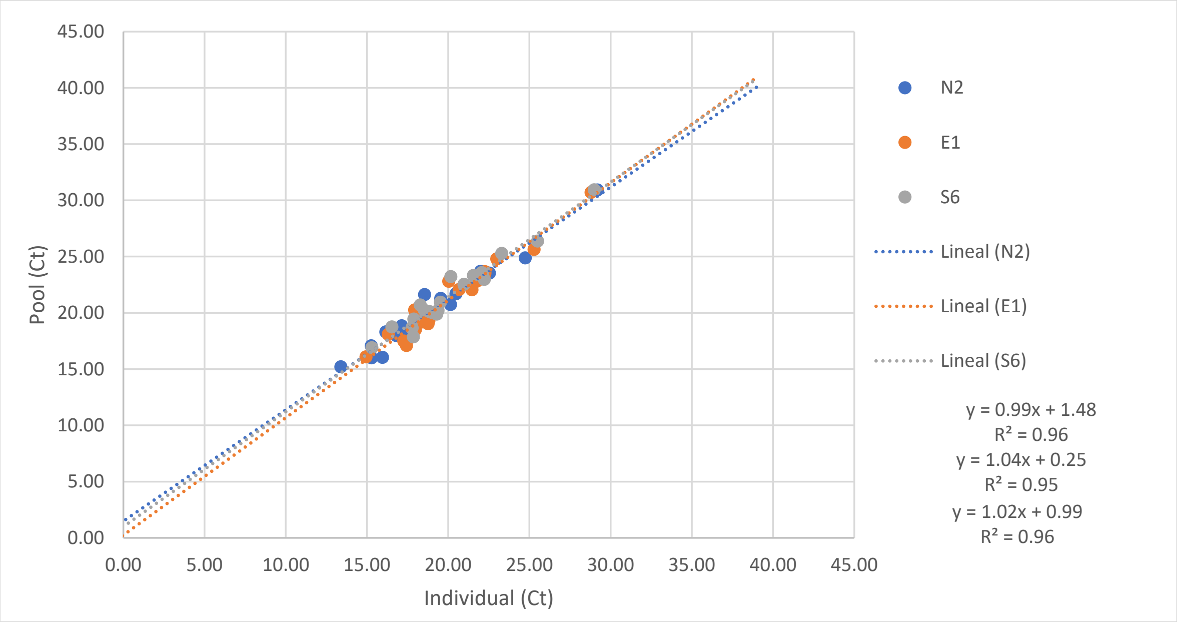


| | Pool | | | Individual | | |
|---|---|---|---|---|---|---|
| | LOD | 95% CI low | 95% CI upp | LOD | 95% CI low | 95% CI upper |
| N | 37.29 | 36.91 | 37.67 | 36.17 | 35.79 | 36.56 |
| E | 37.29 | 36.91 | 37.67 | 35.62 | 35.25 | 35.98 |
| S | 37.29 | 36.91 | 37.67 | 35.59 | 35.22 | 35.96 |

For a pool which is at the limit of detection, the individual sample would have had a Ct around 36.17 for N2. This means that during the pooling procedure samples with Cts between 36.17 and 37.29 would have been detected using individual testing but will not be due to pooling.
